# Vector-derived Cadherin Mimicry in Pemphigus Vulgaris: A Proposed Model Linking HLA-DRB1*04:02/14:01 Genotype with Environmental Exposure

**DOI:** 10.1101/2025.08.06.25333158

**Authors:** Bayram Toraman, Burak Kaan Kasap, Hande Ermis Akkus, Deniz Aksu Arica, Savas Yayli

**Author notes:** **Corresponding author: Bayram TORAMAN**, Faculty of Medicine, Department of Medical Biology, Karadeniz Technical University, Trabzon, Turkey.

## Abstract

Pemphigus vulgaris (PV) is a life-threatening autoimmune blistering disease caused by pathogenic autoantibodies targeting the desmosomal cadherins desmoglein 3 (DSG3) and desmoglein 1 (DSG1), essential for cell–cell adhesion. Although the role of these autoantibodies in disease pathogenesis is well established, the mechanisms initiating the autoimmune response remain unclear. A strong genetic association has been identified between PV and specific HLA class II alleles, particularly *HLA-DRB1*04:02* and *\*14:01*, which may facilitate the presentation of desmoglein-derived peptides to autoreactive CD4+ T cells. Environmental factors are also thought to contribute, with molecular mimicry being a leading hypothesis—whereby foreign antigens resembling host proteins trigger cross-reactive immune responses. In this study, we analyzed *HLA-DRB1* allele frequencies in Turkish PV patients and controls, confirming strong associations with *04:02 and 14:01. Notably, we found that the heterozygous *HLA-DRB104:02/14:01* genotype confers an approximately 100-fold increased risk for PV. Building on this, we propose a novel model: cadherin-like proteins from the salivary glands of mosquitoes or similar blood-feeding insects may structurally mimic DSG3/DSG1. In our model—termed vector-derived cadherin mimicry (VCM)—pattern recognition, rather than primary sequence identity (i.e., shared surface charge/topology patterns) is key to the mimicry mechanism. Repeated exposure to such antigens in genetically susceptible individuals may contribute to tolerance breakdown and disease initiation. Using HLA genotyping, peptide binding prediction, structural modeling, and molecular dynamics simulations, we provide preliminary in silico evidence supporting the VCM hypothesis as a potential trigger for PV-specific autoimmunity.

## Introduction

Autoimmune diseases arise when immune tolerance to self-antigens is disrupted, often through complex interactions between genetic susceptibility and environmental factors. Pemphigus vulgaris (PV) represents a prototypical model for studying organ-specific autoimmunity, particularly due to its well-characterized autoantigen targets and genetic associations such as HLA [1,2]. PV is a life-threatening autoimmune blistering disease characterized by the production of pathogenic autoantibodies targeting desmoglein 3 (DSG3) and desmoglein 1 (DSG1) which are critical desmosomal cadherins essential for intercellular adhesion [3,4]. The binding of these autoantibodies to keratinocyte interface surfaces leads to the loss of cell-cell adhesion (acantholysis), resulting in intraepidermal blister formation. Although the role of autoantibodies in disease pathogenesis is well established, the mechanisms underlying the initial loss of tolerance to desmogleins remain incompletely understood [5].

In PV, defects in both central and peripheral tolerance mechanisms are believed to contribute to disease onset. Central tolerance occurs in the thymus for T cells and in the bone marrow for B cells [6,7]. In the thymus, autoreactive T cells may escape deletion due to insufficient expression or presentation of desmoglein-derived peptides. Similarly, developing B cells in the bone marrow that recognize self-antigens with high affinity may escape clonal deletion or receptor editing [7]. Peripheral tolerance mechanisms, such as anergy, suppression by regulatory T cells (Tregs), and activation-induced cell death, may fail, thereby allowing autoreactive lymphocytes to escape deletion and become activated in the periphery [3–5,7]. Moreover, epitope spreading, a process in which immune responses expand from the initial epitope to others on the same or different proteins, has been proposed as a contributing mechanism in the chronic progression of PV [5,6]. This mechanism refers to the process whereby the immune system, initially targeting a specific epitope of an autoantigen, begins to recognize and respond to additional epitopes either within the same molecule (intramolecular spreading) or across different but related molecules (intermolecular spreading) [6,8,9]. In PV, this is exemplified by the progression of immune responses from DSG3 to DSG1 epitopes, reflecting a clinical shift from mucosal-limited disease to the mucocutaneous form, which often correlates with increased disease severity and resistance to therapy [8].

A strong association between particular HLA class II alleles, particularly *HLA-DRB1*04:02* and *DRB1*14:01*, and susceptibility to PV has been reported in multiple populations [10–12]. These alleles are believed to facilitate the presentation of DSG3-derived peptides to autoreactive CD4+ T cells, thereby contributing to disease initiation. In addition to genetic predisposition, environmental factors are thought to play a contributory role in triggering or perpetuating the autoimmune response. One prevailing hypothesis is molecular mimicry, wherein amino acid sequence similarities between microbial or environmental antigens and host proteins result in cross-reactive immune responses [1,3,5,8,13]. Endemic pemphigus foliaceus (fogo selvagem) in Brazil is one such example where sandfly (*Lutzomyia longipalpis*) salivary antigen (LJM11) has been implicated as a potential trigger due to its sequence resemblance to DSG1 [13,14]. Among suspected hematophagous vectors, exposure to black fly bites (*Simulium nigrimanum*) was shown to correlate with anti-desmoglein titers in an epidemiological study[15]. However, the applicability of such a mechanism to PV, and particularly to DSG3-specific autoimmunity, has not been clearly demonstrated.

In this study, we conducted a genetic association study to evaluate *HLA-DRB1* allele frequencies in Turkish PV patients and matched controls. Our findings revealed a strong association between PV and the *HLA-DRB1*04:02* and *DRB1*14:01* alleles, consistent with previous reports in other populations. Building on these results, we propose a novel hypothesis: cadherin-like proteins from the salivary glands of mosquitoes or similar hematophagous vectors (e.g., sandflies) may structurally mimic DSG3 or DSG1. We emphasize that in our hypothesis, pattern similarity— rather than primary amino acid sequence similarity —is the key factor driving molecular mimicry. Chronic exposure to these antigens in genetically susceptible individuals could contribute to the breakdown of tolerance and trigger PV-specific autoimmunity. To explore the plausibility of this <u>v</u>ector-derived pattern (<u>c</u>adherin) <u>m</u>imicry model (VCM) in the pathogenesis of PV, we employed a combination of HLA genotyping, peptide-binding predictions, structural modeling, and molecular dynamics analyses.

## Materials and Methods

### Study Population

This case–control study included patients diagnosed with PV who were followed up at the Bullous Diseases outpatient clinic of the Department of Dermatology and Venereology, Faculty of Medicine, Karadeniz Technical University (KTU). All PV diagnoses were confirmed based on clinical presentation, histopathological findings, direct immunofluorescence, and/or ELISA (Enzyme-Linked Immunosorbent Assay). Only patients who had no history of other autoimmune diseases and who provided written informed consent were included. A total of 86 PV patients aged between 21 and 81 years were enrolled in the study. The control group consisted of 200 healthy individuals with no known autoimmune diseases, matched to the patient group in terms of age and sex distribution, and who also provided written informed consent to participate. For both groups, 5–9 ml of peripheral blood was collected from everyone into EDTA (ethylenediaminetetraacetic acid)-containing tubes for further analysis. Genomic DNA (gDNA) was extracted from peripheral blood samples of all participants using the MiniFavorPrep™ Blood/Cultured Cells Genomic DNA Extraction kit (Favorgen, Australia) according to the manufacturer’s instructions. This study was approved by the Ethics Committee of the Faculty of Medicine, KTU, under protocol number 2022/51.

### Determination of *HLA-DRB1* alleles and Test Statistics

*HLA-DRB1* allele typing was performed using the PCR-SSOP (sequence-specific oligonucleotide probe) method combined with the Luminex platform. This technique is based on the hybridization of PCR-amplified target sequences (amplicons), generated using gene-specific primers, with short oligonucleotide probes specific to each *HLA-DRB1* allele. The resulting biotinylated PCR products were hybridized to microspheres, each conjugated with allele-specific oligonucleotide probes and distinguishable by unique spectral properties. Hybridized products were then detected by a fluorescence signal generated through the binding of streptavidin–phycoerythrin conjugate to the biotin label, indicating the presence of the target allele [16,17]. Genotyping was performed on a Luminex LABScan3D system (One Lambda, USA) capable of multiplex analysis, and the data were analyzed using HLA Fusion software. The commercially available One Lambda SSP^®^ HLA Typing kit (Thermo Fisher Scientific) was used. This validated kit, widely adopted in clinical research, provides high specificity and reliability for the identification of *HLA-DRB1* alleles within the human HLA class II region.

Power analysis was performed using Power for Genetic Association Analyses (PGA, version 2.0) software [18], and the number of patients and controls required to achieve 80% statistical power was calculated. The Hardy-Weinberg equilibrium of *HLA-DRB1* alleles in the control group was tested using the GENEPOP [19]. Statistical analyses of *HLA-DRB1* allele and genotype frequencies were conducted using Pearson’s Chi-Square Test of Independence, while Fisher’s Exact Test was applied for contingency tables with expected cell counts <5, using the RStudio platform (v2024.04.2). Statistical significance was assessed using the Benjamini–Hochberg false discovery rate (FDR) correction, with an adjusted p-value (p_adj) <0.05 considered statistically significant.

### Sequence/Pattern Similarity Analysis and Prediction of Ectodomain Antigenic Peptides

To identify candidate cadherin-like proteins with potential for molecular mimicry, we performed BLASTP searches against the VectorBase database (https://vectorbase.org/vectorbase/app) using the EC1 domain of human DSG3 as the query sequence. We employed PSI-BLAST to increase sensitivity for detecting structurally conserved and evolutionarily related sequences. PSI-BLAST utilize position-specific scoring matrices to capture deeper pattern-level homology not evident from linear sequence identity alone. The top-scoring result was a cadherin-like protein from *Aedes albopictus*, annotated based on homology to *Aedes aegypti* (protein: AAEL000597; Gene symbol: LOC5564848/ neural-cadherin) (**Suppl. File**). This protein, with a predicted length of 1569 amino acids, was selected for further analysis based on its high similarity score and cadherin domain architecture. From the BLAST alignment, we extracted the homologous region to human DSG3 ectodomain 1 (hDSG3 EC1) and converted it to FASTA format. This sequence was subjected to MHC class II peptide-binding predictions using the IEDB MHC-II binding tool (www.iedb.org), applying the same parameters used for hDSG3. Based on our genetic association findings, *HLA-DRB1*04:02* and *HLA-DRB1*14:01* were selected as the MHC class II alleles of interest. We selected candidate antigenic peptides for both hDSG3 and *A. albopictus* EC1 domain based on the following criteria: (1) strong predicted binding affinity to *HLA-DRB1*04:02* and *HLA-DRB1*14:01*, (2) a high combined immunogenicity score, and (3) the highest sequence similarity to the hDSG3 EC1 amino acid sequence, as determined by multiple sequence alignment with the predicted EC1 domain of *A. albopictus* homologous region [20,21].

### Structural Modeling, Molecular Dynamics Simulations and Binding Free Energy Calculations

Structural modeling of both the EC1 domain peptide of DSG3 and the cadherin-like protein peptide derived from *A. albopictus* was performed using AlfaFold3 and AlfaFoldMultimer tool [22,23] using Colab platform [24]. Representations of the molecular models are executed by UCSF ChimeraX (v1.10) [25]. All-atom molecular dynamics (MD) simulations were performed using GROMACS 2022.4 [26] with the AMBER99SB-ILDN force field. Systems were solvated with TIP3P water and neutralized with counterions. After energy minimization, simulations were equilibrated in NVT and NPT ensembles followed by a 100 ns production run. The simulations were carried out at 298.15 K and 1 atm using the V-rescale thermostat and Parrinello-Rahman barostat, respectively. The binding free energy (ΔG_binding) of the peptide–protein complexes was calculated using the gmx_MMPBSA v1.5.7 tool [27,28], which implements the MM/GBSA method. The last 10 frames of the production trajectory were extracted using trjconv after applying periodic boundary condition (PBC) correction. The input files included; md.tpr: topology file, md.xtc: PBC-corrected trajectory, index.ndx: index file with peptide (group 18) and receptor (group 17). The calculations were performed using the igb=5 implicit solvent model (GB-OBC II), and per-residue decomposition was enabled to identify the key binding contributors. Temperature was set to 298.15 K, and salt concentration to 0.15 M. Final binding energy results were reported from the FINAL_RESULTS_MMPBSA.dat output file. Detailed protocol steps and parameters are provided in **Suppl. File.**

## Results

### *HLA-DRB1* alleles and Genotypes Test Statistics

In controls, *HLA-DRB1* alleles were in Hardy-Weinberg equilibrium. A total of 33 different *HLA-DRB1* alleles were identified among the study groups (**Table 1**). The 10 most frequently observed *HLA-DRB1* alleles in PV patients and the control group are presented in **Table 1**. The alleles with the highest frequency in the PV group were *HLA-DRB1*04:02* at 40%, followed by *DRB1*14:01* at 18%. In the control group, the most frequently observed alleles were *HLA-DRB1*11:01, 11:04,* and *07:01* at 9%, respectively. The frequencies and test statistics of *HLA-DRB1* alleles observed in PV patients and controls are summarized in **Table 2**. Carriers of the *HLA-DRB1*04:02* allele exhibited a significantly increased risk of developing PV, with an odds ratio (OR) of 12.95 (95% CI: 7.40-24.57), indicating nearly a 13-fold higher susceptibility compared to non-carriers. Similarly, the *HLA-DRB1*14:01* allele was significantly more frequent among PV patients than controls (p = 1.23×10⁻⁵), and carriers of this allele showed a 4.55-fold increased risk of PV (OR: 4.55; 95% CI: 2.35–8.74).

**Table 1.** The top 10 most frequently observed *HLA-DRB1* alleles in PV patients and controls.

| **HLA-DRB1\*** | **PV (n)** | **%** | **HLA-DRB1\*** | **Control (n)** | **%** |
| --- | --- | --- | --- | --- | --- |
| **04:02** | 70 | 40.70 | 11:01 | 39 | 9 |
| **14:01** | 31 | 18.02 | 11:04 | 39 | 9 |
| 11:04 | 9 | 5 | 07:01 | 37 | 9 |
| 01:01 | 7 | 4 | 15:01 | 33 | 8 |
| 04:03 | 7 | 4 | 03:01 | 26 | 6 |
| 07:01 | 7 | 4 | 16:01 | 26 | 6 |
| 13:02 | 7 | 4 | 01:01 | 23 | 5 |
| 11:01 | 6 | 3 | 13:01 | 19 | 4 |
| 15:01 | 6 | 3 | **04:02** | 18 | 4.50 |
| 03:01 | 5 | 2 | **14:01** | 17 | 4.25 |
| **Total** | **155** | **87.72** |  | **277** | **64.75** |

n: number of alleles

**Table 2.** *HLA-DRB1* alleles and chi-square test statistics.

| **HLA-DRB1\*** | **Control (n)** | **PV (n)** | **p** | **adj_p** | **OR** | **95% CI** |
| --- | --- | --- | --- | --- | --- | --- |
| 04:02 | 18 | 70 | $2.03 \times 10^{-24}$ | **$6.71 \times 10^{-23}$** | 12.95 | 7.40-24.57 |
| 14:01 | 17 | 31 | $7.51 \times 10^{-7}$ | **$1.23 \times 10^{-5}$** | 4.55 | 2.35-8.74 |
| 16:01 | 26 | 0 | $9.75 \times 10^{-5}$ | **0.001** | 0.03 | 0-0.31 |
| 11:01 | 39 | 6 | 0.00 | **0.037** | 0.33 | 0.10-0.75 |
| 14:04 | 0 | 4 | 0.00 | 0.06 | 20.19 | 1.46-∞ |
| 15:01 | 33 | 6 | 0.02 | 0.10 | 0.40 | 0.12-0.91 |
| 13:03 | 12 | 0 | 0.02 | 0.10 | 0.08 | 0-0.77 |
| 07:01 | 37 | 7 | 0.03 | 0.13 | 0.41 | 0.17-0.97 |
| 09:01 | 8 | 0 | 0.06 | 0.22 | 0.12 | 0-1.27 |
| 04:01 | 16 | 1 | 0.07 | 0.23 | 0.19 | 0.029-1.14 |
| 11:04 | 39 | 9 | 0.07 | 0.23 | 0.50 | 0.22-1.09 |
| 08:01 | 0 | 1 | 0.09 | 0.25 | 6.61 | 0.40-∞ |
| 11:11 | 0 | 1 | 0.09 | 0.25 | 6.61 | 0.40-∞ |
| 03:01 | 26 | 5 | 0.12 | 0.28 | 0.43 | 0.16-1.23 |
| 04:05 | 6 | 0 | 0.18 | 0.37 | 0.16 | 0-1.86 |
| 04:08 | 5 | 0 | 0.18 | 0.37 | 0.19 | 0-1.86 |
| 15:02 | 12 | 1 | 0.24 | 0.37 | 0.25 | 0.03-1.62 |
| 10:01 | 11 | 2 | 0.24 | 0.44 | 0.46 | 0.03-1.64 |
| 04:04 | 4 | 4 | 0.26 | 0.46 | 2.22 | 0.40-12.06 |
| 04:03 | 9 | 7 | 0.30 | 0.49 | 1.76 | 0.59-5.10 |
| 04:07 | 3 | 0 | 0.31 | 0.49 | 0.31 | 0-3.34 |
| 13:02 | 12 | 7 | 0.46 | 0.69 | 1.3 | 0.51-3.98 |
| 13:01 | 19 | 6 | 0.51 | 0.74 | 0.72 | 0.21-1.72 |
| 01:01 | 23 | 7 | 0.55 | 0.76 | 0.68 | 0.27-1.67 |
| 08:04 | 2 | 1 | 0.59 | 0.78 | 1.31 | 0.15-30.33 |
| 01:02 | 3 | 2 | 1 | 1 | 1.57 | 0.1-7.80 |
| 04:06 | 1 | 0 | 1 | 1 | 0.72 | 0-11.78 |
| 13:05 | 1 | 0 | 1 | 1 | 0.72 | 0-11.78 |
| 16:02 | 1 | 0 | 1 | 1 | 0.72 | 0-11.78 |
| 08:02 | 2 | 0 | 1 | 1 | 0.43 | 0-11.70 |
| 08:03 | 7 | 3 | 1 | 1 | 1.02 | 0.24-4.15 |
| 11:03 | 6 | 1 | 1 | 1 | 0.50 | 0.07-4.01 |
| 12:01 | 2 | 0 | 1 | 1 | 0.43 | 0-11.71 |

n: number of alleles, adj p: adjusted p value, OR: risk ratio, CI: confidence interval

After showing that the *HLA-DRB1*04:02* and *HLA-DRB1*14:01* alleles significantly increased the risk of developing PV, genotype test statistics were performed to examine the distribution of the co-occurrence of these two alleles (heterozygosity) between patients and controls. Genotype frequency is the frequency of individuals with a particular DRB1 genotype identified in the PV and control groups. The relevant genotype data and test statistics are presented in **Table 3**.

**Table 3.** The most frequently observed genotypes and Fisher’s exact test statistics in PV patients and controls in the study.

| **DRB1**<br>**Genotype** | **Control**<br>**(n)** | **PV**<br>**(n)** | **a\*** | **b\*** | **c\*** | **d\*** | **p** | **adj_p** | **OR** | **%95 CI** |
| --- | --- | --- | --- | --- | --- | --- | --- | --- | --- | --- |
| 04:02/14:01 | 0 | 20 | 20.5 | 0.5 | 71.5 | 200.5 | $1.63 \times 10^{-11}$ | $2.21 \times 10^{-9}$ | 114.97 | 6.86 – 1925.82 |
| 04:02/04:03 | 0 | 6 | 6.5 | 0.5 | 85.5 | 200.5 | 0.0008 | **0.0374** | 30.48 | 1.69 – 547.22 |
| 04:02/13:02 | 0 | 6 | 6.5 | 0.5 | 85.5 | 200.5 | 0.0008 | **0.0374** | 30.48 | 1.69 – 547.22 |
| 01:01/04:02 | 0 | 5 | 5.5 | 0.5 | 86.5 | 200.5 | 0.0027 | 0.0934 | 25.49 | 1.39 – 466.2 |
| 04:02/04:04 | 0 | 3 | 3.5 | 0.5 | 88.5 | 200.5 | 0.0298 | 0.6670 | 15.85 | 0.81 – 310.28 |
| 04:02/13:01 | 0 | 4 | 4.5 | 0.5 | 87.5 | 200.5 | 0.0091 | 0.2465 | 20.62 | 1.09 – 387.21 |
| 11:04/11:04 | 12 | 1 | 1.5 | 12.5 | 90.5 | 188.5 | 0.0700 | 1 | 0.24 | 0.04 – 1.38 |
| 15:01/15:01 | 7 | 0 | 0.5 | 7.5 | 91.5 | 193.5 | 0.1027 | 1 | 0.14 | 0.007 – 2.49 |
| 07:01/11:01 | 5 | 0 | 0.5 | 5.5 | 91.5 | 195.5 | 0.3294 | 1 | 0.19 | 0.01 – 3.55 |

\* 0.5 was added to the cell frequencies as part of the Haldane-Anscombe correction. Therefore, the upper limit of the confidence interval was calculated as infinity. n: number of genotype, adj\_p: corrected p-value, OR: Risk ratio, CI: confidence interval

A noteworthy observation is that the heterozygous genotype *HLA-DRB1*04:02/14:01* was detected exclusively in PV patients (**Table 3**). Individuals carrying this genotype exhibited a remarkably elevated risk of developing PV, a 115-fold increase compared to non-carriers. This finding underscores the strong genotype-related association in a complex disease such as PV and highlights PV as a striking example of a rare autoimmune disorder in which disease susceptibility is significantly influenced by specific HLA genotype.

### *HLA-DRB1*04:02* and *DRB1*14:01* Specific DSG3 EC1 Ectodomain Antigenic Peptide Prediction and Multiple Sequence Alignment

We utilized peptide-binding predictions generated by the IEDB MHC-II binding tool, which identified 19 unique peptide sequences within hDSG3 EC1 and 167 within the *A. albopictus* EC1-like domain (Aa_p) (**Suppl. Table S1**). From these, we selected candidate antigenic peptides for both hDSG3 and Aa_p EC1-like domain based on the criteria mentioned before (**Figure 1**).

**Figure 1.**
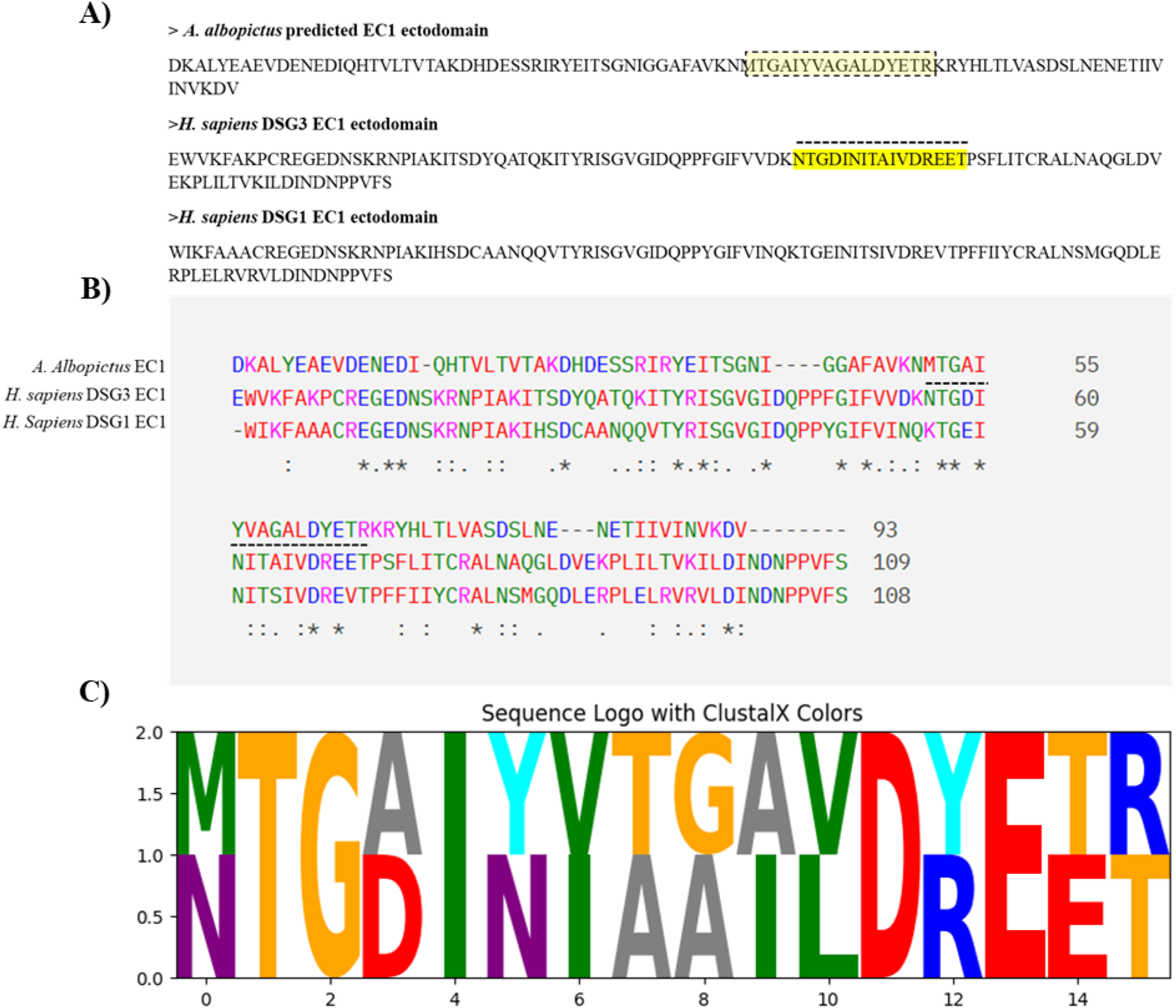
Comparative analysis of the EC1 domains of hDSG3, hDSG1 and *A. albopictus* cadherin-like protein reveals a conserved antigenic motif. **(A)** Schematic representation of the EC1 domain of hDSG3, hDSG1 and *A. albopictus*, showing the most antigenic 16-mer peptides identified using the IEDB MHC-II binding prediction tool (highlighted in yellow). **(B)** Multiple sequence alignment of hDSG3, hDSG1 and EC1 domains with the *A. albopictus* EC1-like domain. The highlighted regions correspond to the predicted peptides shown in panel A. Alignment was performed using Clustal Omega, and conserved residues are indicated below the sequences. **(C)** Sequence logo comparing the selected peptides from Aa_P (MTGAIYVAGALDYETR) and hDSG3 (NTGDINITAIVDREET). The height of each letter reflects the relative frequency and conservation of the residue at that position. Coloring follows the ClustalX scheme, with residues colored based on their physicochemical properties: hydrophobic (green), small/hydroxyl (orange), acidic (red), basic (blue), polar (cyan), aromatic (magenta), and others (gray)—sequence logo generated using the Python-based Logomaker library (v0.8) in Google Colab.

### Structural Models and Binding Free Energy

We modelled both the hDSG3 EC1 domain and the *A. albopictus* EC1-like domain using AlphaFold3 [22], and visualized the resulting structures with ChimeraX (**Figure 2A, B**). Structural alignment revealed a high degree of similarity in domain architecture and secondary structure elements between the two models (**Suppl. File, Figure 2C)**. Then, we performed the predicted antigenic peptides mentioned above that docked with *HLA-DRB1*04:02* based class II receptor by AlfaFold Multimer [23]. The receptors and corresponding ligand complexes are shown in **Figure 3**.

**Figure 2:**
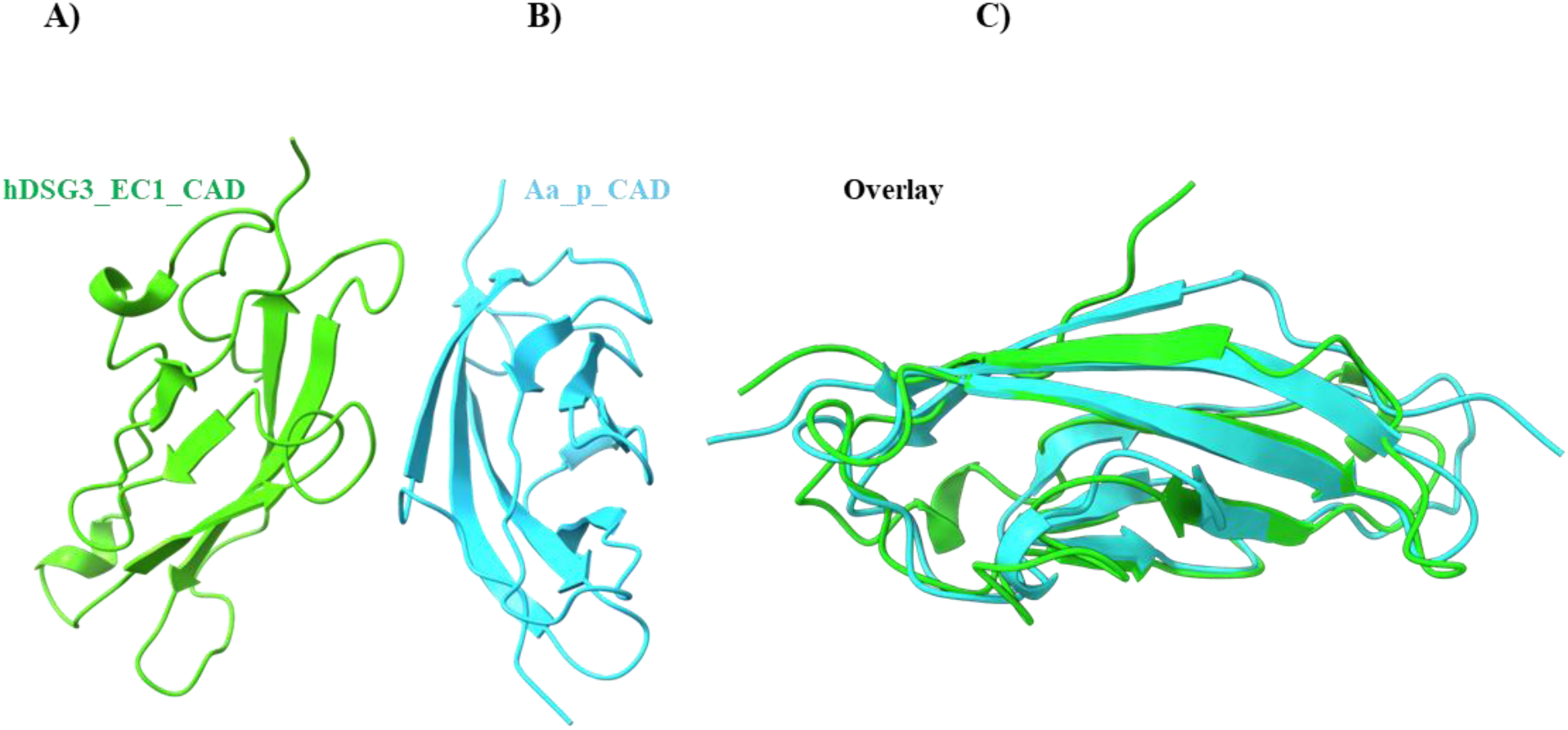
Structural comparison of hDSG3 EC1 domain (green) and *A. albopictus*-derived CAD-like domain (cyan). The right panel shows structural alignment, highlighting high β-strand conservation. Ribbon representations were generated using UCSF ChimeraX.

**Figure 3:**
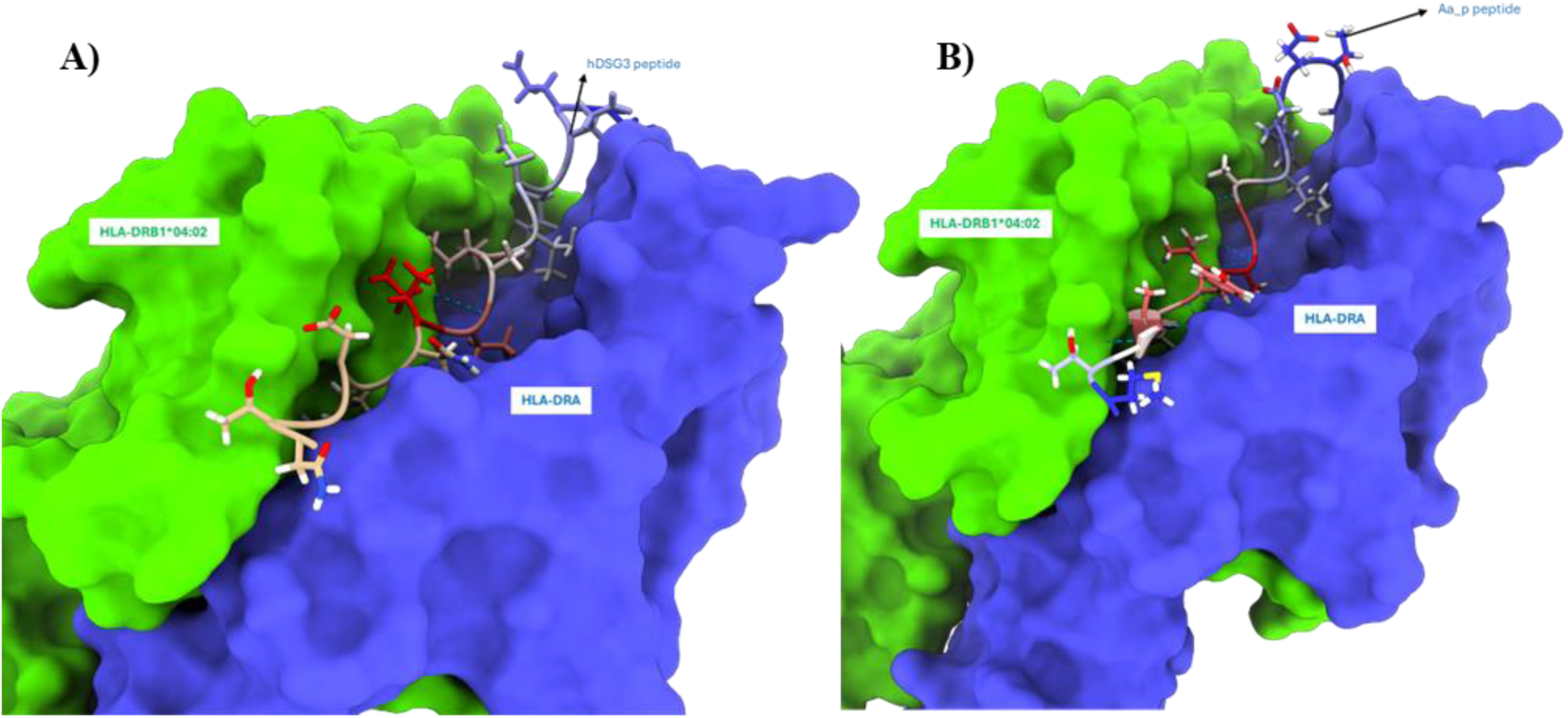
Surface representation of the HLA-DR heterodimer (HLA-DRA: blue, HLA-DRB1-HLA-DRB1*04:02: green) bound to the hDSG3-derived peptide (A) and Aa_P peptide (B) (stick model). Labels indicate the α and β chains and the bound peptide, which is accommodated within the peptide-binding groove.

After predicting and visualizing the binding structures, we performed binding free energy calculations for both structures using the gmx_MMPBSA tool (**Suppl. File**) [29] Even if binding poses of two peptides seem very similar, their total binding free energies are very different (**Table 4**).

**Table 4.** Binding free energies of both complexes (kcal/mol)

| Component | Mean_hDSG3 | Mean_Aa_P | Difference (Aa_P - hDSG3) |
| --- | --- | --- | --- |
| VDWAALS | -66.79 | -109.68 | -42.89 |
| EEL | 38.06 | 36.93 | -1.13 |
| EGB | 3.54 | -0.03 | -3.57 |
| ESURF | -9.85 | -14.72 | -4.87 |
| GGAS | -28.73 | -72.74 | -44.01 |
| GSOLV | -6.31 | -14.75 | -8.44 |
| **TOTAL** | **-35.04** | **-87.5** | **-52.46 kcal/mol** |

VDWAALS: Van der Waals; EEL: Electrostatic; EGB: GB solvent effect ESURF: Solvent surface energy; GGAS: Internal energy (VDW + EEL) GSOLV: Solvation energy (EGB + ESURF); TOTAL: Bonding $\Delta G$

## Discussion

Pemphigus vulgaris (PV) is a potentially life-threatening autoimmune blistering disease, and the underlying causes of autoantibody formation against desmogleins remain largely unknown. The disease is thought to result from a combination of genetic predisposition and environmental risk factors, with human leukocyte antigen (HLA) class II genes representing the strongest known genetic contributors. Multiple independent cohort studies have consistently implicated specific *HLA-DRB1* alleles, particularly *HLA-DRB1*04:02* and *HLA-DRB1*14:01*, as significant risk factors for PV [30–33]. The prevalence of *HLA-DRB1*04:02* has been reported to be as high as 92% among Ashkenazi Jewish patients [34] and between 33% and 85% in other PV populations of diverse ethnic origins [30]. Similarly, the *HLA-DRB1*14:01* allele has been observed in 28–44% of patients in different cohorts [30] In our study, *HLA-DRB1*04:02* and *HLA-DRB1*14:01* were present in 40.7% and 18% of PV patients, respectively, findings that are broadly consistent with previous reports. Notably, the frequency of *HLA-DRB1*14:01* in our population was lower than in many other populations reported to date (**Table 1**). Conversely, *HLA-DRB1*16:01* and *HLA-DRB1*11:01* alleles appeared to have a protective effect against PV in our cohort (**Table 2**).

Previous studies investigating HLA associations in PV have predominantly relied on allele- or haplotype-based analyses [35]. To the best of our knowledge, this is the first study to report that the heterozygous genotype *HLA-DRB1*04:02/14:01* confers an approximately 100-fold increased risk for PV (OR: 114.97; 95% CI: 6.86-1925.82), although the wide confidence interval warrants cautious interpretation (**Table 3**). Unlike typical genetic association studies that focus on common bi-allelic SNPs and report allele or genotype frequencies[36], *HLA-DRB1* is highly polymorphic, with hundreds of distinct alleles. This diversity results in numerous rare genotype combinations, limiting statistical power in small to moderate-sized cohorts. Consequently, most studies have emphasized allele- or haplotype-level associations rather than genotype-level effects [37]. Despite relatively low genotype frequencies, our study provides the first evidence that a specific heterozygous combination confers markedly higher PV risk than either allele alone. These results underscore the importance of genotype-level analyses in highly polymorphic loci such as *HLA-DRB1*, mainly when supported by adequate sample sizes. Collectively, our findings reinforce the central role of *HLA-DRB1* alleles and genotypes in PV susceptibility. However, these findings also emphasize that while specific HLA variants may be mostly necessary for disease development, they are unlikely to be sufficient in the absence of additional environmental triggers. A variety of environmental and microbial factors—including diet, medications, and infections—have been proposed to initiate PV [38,39]. Nevertheless, none of these factors has yet been conclusively demonstrated to explain the molecular mechanisms underlying the initial break in immune tolerance. In studies on endemic pemphigus foliaceus (Fogo selvagem), a possible causal relationship has been suggested between exposure to salivary antigens of hematophagous insects, such as sandflies and blackflies, and the development of autoantibodies against DSG1[15,40,41]

In line with this concept, and to evaluate the plausibility of our proposed Vector-derived Cadherin Mimicry (VCM) hypothesis, we employed a multidisciplinary approach combining HLA genotyping, peptide-binding prediction (**Figure 1, Suppl. File**), structural modeling (**Figures 2 and 3**), and molecular dynamics simulations (**Table 4**). These complementary analyses allowed us to investigate whether cadherin-like proteins from hematophagous vectors could exhibit sufficient structural and immunological similarity to hDSG3 to trigger PV-specific autoimmunity in individuals carrying *HLA-DRB1*04:02* or *HLA-DRB1*14:01* potentially.

We propose that with repeated vector bites, cadherin-like proteins of epithelial cells from the salivary glands may be introduced into host tissues, serving as environmental antigens that initiate cross-reactive immune responses in genetically predisposed individuals (**Figure 4**). The salivary glands of *A. albopictus* consist of a single layer of secretory epithelial cells, typically cuboidal or columnar, organized into tubular acini and ducts [42] These cells synthesize and secrete proteins essential for blood feeding and express cadherins at their apicolateral membranes to maintain epithelial integrity [43,44]. Mechanical disruption during feeding, natural cellular turnover, or lysis induced by apoptosis, necrosis, or infection (e.g., viral or protozoal) could result in the release of membrane-associated cadherin-like proteins into saliva and, consequently into host tissues (**Figure 4**).

**Figure 4.**
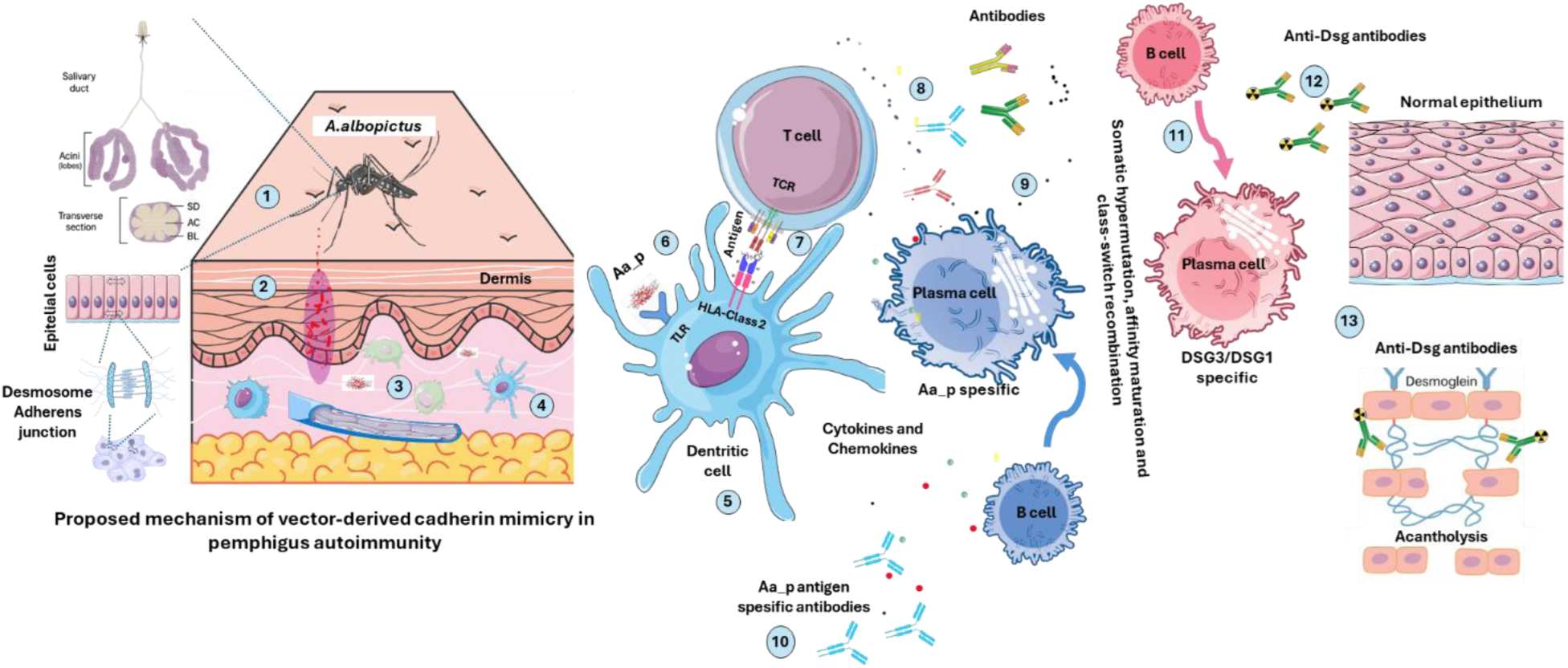
Proposed model of Vector-derived Cadherin Mimicry (VCM) hypothesis in PV. Figure was generated by using https://bioicons.com/ and https://bioart.niaid.nih.gov/. **1.** *A. albopictus* mosquito introduces salivary proteins into the host skin during blood feeding; **2.** Deposition of vector-derived cadherin-like peptides and other salivary molecules into the dermis; **3.** Uptake and processing of foreign peptides by local antigen-presenting cells (APCs); **4.** Migration of activated dendritic cells towards draining lymph nodes while presenting processed peptides; **5.** Innate immune activation via Toll-like receptors (TLRs) and other pattern recognition receptors on APCs; **6.** Formation of peptide–HLA class II complexes with high affinity to risk alleles (*HLA-DRB1*04:02, *14:01*); **7.** Recognition of peptide–HLA complex by cross-reactive T cell receptors (TCRs), initiating T cell activation; **8.** Provision of costimulatory signals by activated helper T cells to autoreactive B cells; **9.** Activation and clonal expansion of B cells recognizing hDSG3-mimicking peptides (Aa_P); **10.** Differentiation of autoreactive B cells into antibody-producing plasma cells; **11.** Production of pathogenic anti-DSG3 autoantibodies causing loss of keratinocyte adhesion and blister formation in PV **12.** Autoantibody binding to desmoglein in the epidermis. Pathogenic IgG autoantibodies penetrate the epidermis and bind to desmosomal cadherins (DSG3 and DSG1) on keratinocyte surfaces, disrupting intercellular adhesion. **13.** Acantholysis process. Autoantibody binding interferes with desmoglein-mediated cell–cell junctions, leading to keratin filament retraction, loss of cohesion between keratinocytes, and subsequent intraepidermal blister formation (acantholysis).

### Proposed Mechanism: From Vector-Derived Antibody Response to Desmoglein-Targeted Autoimmunity

In genetically susceptible individuals (e.g., carriers of *HLA-DRB1*04:02* or *\*14:01*), repeated exposure to cadherin-like peptides in the saliva of *A. albopictus* or other hematophagous insects (e.g., sandflies) may elicit a subclinical antibody response against these foreign antigens. Due to sequence and structural similarity with hDSG3 and hDSG1, certain B and T cell clones may exhibit molecular mimicry and cross-reactivity. Over time, somatic hypermutation, affinity maturation, and class-switch recombination [45] within germinal centers may enhance the specificity and binding strength of these B cell receptors, leading to increasing recognition of self-antigens (DSG3/DSG1) [46]. Initial tissue damage may expose additional cryptic desmoglein epitopes, which are further processed and presented by antigen-presenting cells, thereby promoting epitope spreading. This process could drive clonal expansion of autoreactive B cells and transition the immune response from vector-derived peptides to sustained autoimmunity. Ultimately, persistent autoantibody production against DSG3 and DSG1 would disrupt keratinocyte adhesion, resulting in the characteristic blistering observed in PV (**Figure 4)**.

To test our VCM hypothesis, at least in silico, we employed a combination of generating HLA-DRA/DRB1*04:02 complexes, peptide-binding predictions, structural modeling, and molecular dynamics analyses (**Table 4**). Our analysis revealed that the Aa_P peptide binds with greater affinity than the hDSG3 EC1-derived peptide. These data provide preliminary support for the VCM hypothesis, although experimental validation is required. For experimental validation, an appropriate animal model is essential. Humanized mice expressing the *HLA-DRB1*04:02* allele can be utilized, as previously demonstrated by Eming et al. in a preclinical mouse model of PV [47] The VCM hypothesis could be directly tested by immunizing these mice with both recombinant hDSG3 ectodomains and *A. albopictus*–derived cadherin-like (CAD-like) peptides (**Figure 1A** and **Figure 2B**).

Mosquitoes and other hematophagous insects are already known vectors for numerous viral, bacterial, and protozoan diseases, as well as triggers of allergic reactions [48]. If experimentally confirmed, our hypothesis would represent the first evidence linking an autoimmune disease such as PV to vector-derived molecular mimicry and autoimmunity. This finding could have significant implications for public health by identifying genetically susceptible individuals (e.g., *HLA-DRB1*04:02* carriers) who may benefit from preventive measures aimed at reducing exposure to such environmental triggers. Although some studies have proposed a possible association between exposure to sandfly salivary antigens (e.g., LJM17, LJM11, and PpSP32) and the development of anti-DSG1 autoantibodies in endemic pemphigus foliaceus (Fogo selvagem), these findings remain primarily epidemiological [15,40,49]. The molecular mimicry mechanisms suggested in such cases have not been strongly supported by structural or immunological evidence. Notably, the antigens identified in these studies do not appear to elicit immunodominant or pathogenic antibody responses capable of inducing acantholysis. In contrast, our proposed VCM hypothesis in PV is supported by peptide–HLA binding predictions, structural homology modeling, and preliminary energetic data suggesting cross-reactive potential between cadherin-like vector peptides and hDSG3. These findings provide a more mechanistically grounded framework for investigating how vector-derived proteins may contribute to autoimmune pathogenesis in genetically susceptible individuals.

In conclusion, to the best our knowledge, this is the first study to report that the heterozygous genotype *HLA-DRB1*04:02/14:01* confers an approximately 100-fold increased risk for PV. Our study highlights a previously unexplored link between genetic susceptibility to PV and repeated environmental exposure to vector-derived cadherin-like proteins. By integrating immunogenetic data, structural modeling, and molecular dynamics simulations, we provide preliminary in silico evidence supporting the plausibility of the VCM hypothesis. If validated experimentally, this model would represent a novel paradigm in autoimmune disease pathogenesis—one in which hematophagous insect vectors serve not only as transmitters of infectious agents but also as potential initiators of autoimmune processes through molecular mimicry. These insights may pave the way for identifying at-risk individuals based on their HLA genotype and implementing preventive strategies to minimize environmental exposure to such molecular mimics.

## Data Availability

All data produced in the present study are available upon reasonable request to the authors

## Acknowledgments

We would like to thank the families who participated in this study. We would also like to thank the Karadeniz Technical University Scientific Research Projects Coordination Unit for their support.

