## Supplementary material for "Vector-derived Cadherin Mimicry in Pemphigus Vulgaris: A Proposed Model Linking HLA-DRB1*04:02/14:01 Genotype with Environmental Exposure": Suppl. File: Supplemantary_File.pdf

BLASTP 2.10.1+

Reference: Stephen F. Altschul, Thomas L. Madden, Alejandro A. Schaffer, Jinghui Zhang, Zheng Zhang, Webb Miller, and David J. Lipman (1997), "Gapped BLAST and PSI-BLAST: a new generation of protein database search programs", Nucleic Acids Res. 25:3389-3402.

Reference for composition-based statistics: Alejandro A. Schaffer, L. Aravind, Thomas L. Madden, Sergei Shavirin, John L. Spouge, Yuri I. Wolf, Eugene V. Koonin, and Stephen F. Altschul (2001), "Improving the accuracy of PSI-BLAST protein database searches with composition-based statistics and other refinements", Nucleic Acids Res. 29:2994-3005.

Database: /eupath/data/apiSiteFilesStaging/VectorBase/49/real/webServices/VectorBase/release-CURRENT/AaegyptiLVP_AGWG/blast/AaegyptiLVP_AGWGAnnotatedProteins;
/eupath/data/apiSiteFilesStaging/VectorBase/49/real/webServices/VectorBase/release-CURRENT/AalbimanusSTECLA2020/blast/AalbimanusSTECLA2020AnnotatedProteins;
/eupath/data/apiSiteFilesStaging/VectorBase/49/real/webServices/VectorBase/release-CURRENT/AalbimanusSTECLA/blast/AalbimanusSTECLAAnnotatedProteins;
/eupath/data/apiSiteFilesStaging/VectorBase/49/real/webServices/VectorBase/release-CURRENT/AalbopictusC6-36/blast/AalbopictusC6-36AnnotatedProteins;
/eupath/data/apiSiteFilesStaging/VectorBase/49/real/webServices/VectorBase/release-CURRENT/AalbopictusFoshan/blast/AalbopictusFoshanAnnotatedProteins;
/eupath/data/apiSiteFilesStaging/VectorBase/49/real/webServices/VectorBase/release-CURRENT/AalbopictusFoshanFPA/blast/AalbopictusFoshanFPAAnnotatedProteins;
/eupath/data/apiSiteFilesStaging/VectorBase/49/real/webServices/VectorBase/release-CURRENT/AaquasalisAaquGF1/blast/AaquasalisAaquGF1AnnotatedProteins;
/eupath/data/apiSiteFilesStaging/VectorBase/49/real/webServices/VectorBase/release-CURRENT/AarabiensisDONGOLA2021/blast/AarabiensisDONGOLA2021AnnotatedProteins;
/eupath/data/apiSiteFilesStaging/VectorBase/49/real/webServices/VectorBase/release-CURRENT/AarabiensisDongola/blast/AarabiensisDongolaAnnotatedProteins;
/eupath/data/apiSiteFilesStaging/VectorBase/49/real/webServices/VectorBase/release-CURRENT/AatroparvusEBRO/blast/AatroparvusEBROAnnotatedProteins;
/eupath/data/apiSiteFilesStaging/VectorBase/49/real/webServices/VectorBase/release-CURRENT/AbellatorAbelBR1/blast/AbellatorAbelBR1AnnotatedProteins;
/eupath/data/apiSiteFilesStaging/VectorBase/49/real/webServices/VectorBase/release-CURRENT/AchristyiACHKN1017/blast/AchristyiACHKN1017AnnotatedProteins;
/eupath/data/apiSiteFilesStaging/VectorBase/49/real/webServices/VectorBase/release-CURRENT/AcoluzziiAcolN3/blast/AcoluzziiAcolN3AnnotatedProteins;
/eupath/data/apiSiteFilesStaging/VectorBase/49/real/webServices/VectorBase/release-CURRENT/AcoluzziiMOPTI/blast/AcoluzziiMOPTIAnnotatedProteins;
/eupath/data/apiSiteFilesStaging/VectorBase/49/real/webServices/VectorBase/release-CURRENT/AcoluzziiMali-NIH/blast/AcoluzziiMali-NIHAnnotatedProteins;
/eupath/data/apiSiteFilesStaging/VectorBase/49/real/webServices/VectorBase/release-CURRENT/AcoluzziiNgousso/blast/AcoluzziiNgoussoAnnotatedProteins;

/eupath/data/apiSiteFilesStaging/VectorBase/49/real/webServices/Vect
orBase/release-
CURRENT/AcoustaniAcouGA1/blast/AcoustaniAcouGA1AnnotatedProteins;
/eupath/data/apiSiteFilesStaging/VectorBase/49/real/webServices/Vect
orBase/release-
CURRENT/AcruziiAcruBR1/blast/AcruziiAcruBR1AnnotatedProteins;
/eupath/data/apiSiteFilesStaging/VectorBase/49/real/webServices/Vect
orBase/release-CURRENT/AculicifaciesA-37/blast/AculicifaciesA-
37AnnotatedProteins;
/eupath/data/apiSiteFilesStaging/VectorBase/49/real/webServices/Vect
orBase/release-
CURRENT/AdarlingiAdarGF1/blast/AdarlingiAdarGF1AnnotatedProteins;
/eupath/data/apiSiteFilesStaging/VectorBase/49/real/webServices/Vect
orBase/release-
CURRENT/AdarlingiCoari/blast/AdarlingiCoariAnnotatedProteins;
/eupath/data/apiSiteFilesStaging/VectorBase/49/real/webServices/Vect
orBase/release-
CURRENT/AdirusWRAIR2/blast/AdirusWRAIR2AnnotatedProteins;
/eupath/data/apiSiteFilesStaging/VectorBase/49/real/webServices/Vect
orBase/release-
CURRENT/AepiroticusEpiroticus2/blast/AepiroticusEpiroticus2Annotated
Proteins;
/eupath/data/apiSiteFilesStaging/VectorBase/49/real/webServices/Vect
orBase/release-
CURRENT/AfarautiFAR1/blast/AfarautiFAR1AnnotatedProteins;
/eupath/data/apiSiteFilesStaging/VectorBase/49/real/webServices/Vect
orBase/release-
CURRENT/AfunestusAfunGA1/blast/AfunestusAfunGA1AnnotatedProteins;
/eupath/data/apiSiteFilesStaging/VectorBase/49/real/webServices/Vect
orBase/release-
CURRENT/AfunestusFUMOZ/blast/AfunestusFUMOZAnnotatedProteins;
/eupath/data/apiSiteFilesStaging/VectorBase/49/real/webServices/Vect
orBase/release-
CURRENT/AgambiaeIfakara/blast/AgambiaeIfakaraAnnotatedProteins;
/eupath/data/apiSiteFilesStaging/VectorBase/49/real/webServices/Vect
orBase/release-
CURRENT/AgambiaePEST/blast/AgambiaePESTAnnotatedProteins;
/eupath/data/apiSiteFilesStaging/VectorBase/49/real/webServices/Vect
orBase/release-
CURRENT/Amaculatusmaculatus3/blast/Amaculatusmaculatus3AnnotatedProt
eins;
/eupath/data/apiSiteFilesStaging/VectorBase/49/real/webServices/Vect
orBase/release-
CURRENT/AmaculipalpisAmacGA1/blast/AmaculipalpisAmacGA1AnnotatedProt
eins;
/eupath/data/apiSiteFilesStaging/VectorBase/49/real/webServices/Vect
orBase/release-
CURRENT/AmarshalliiAmarGA1/blast/AmarshalliiAmarGA1AnnotatedProteins
;
/eupath/data/apiSiteFilesStaging/VectorBase/49/real/webServices/Vect
orBase/release-
CURRENT/AmelasCM1001059_A/blast/AmelasCM1001059_AAnnotatedProteins;
/eupath/data/apiSiteFilesStaging/VectorBase/49/real/webServices/Vect
orBase/release-
CURRENT/AmerusMAF2021/blast/AmerusMAF2021AnnotatedProteins;
/eupath/data/apiSiteFilesStaging/VectorBase/49/real/webServices/Vect
orBase/release-CURRENT/AmerusMAF/blast/AmerusMAFAnnotatedProteins;
/eupath/data/apiSiteFilesStaging/VectorBase/49/real/webServices/Vect
orBase/release-
CURRENT/AminimusMINIMUS1/blast/AminimusMINIMUS1AnnotatedProteins;
/eupath/data/apiSiteFilesStaging/VectorBase/49/real/webServices/Vect
orBase/release-
CURRENT/AmouchetiAmouCM1/blast/AmouchetiAmouCM1AnnotatedProteins;
/eupath/data/apiSiteFilesStaging/VectorBase/49/real/webServices/Vect
orBase/release-
CURRENT/AniliAnilCM1/blast/AniliAnilCM1AnnotatedProteins;
/eupath/data/apiSiteFilesStaging/VectorBase/49/real/webServices/Vect
orBase/release-
CURRENT/AquadriannulatusSANGWE/blast/AquadriannulatusSANGWEAnnotated
Proteins;
/eupath/data/apiSiteFilesStaging/VectorBase/49/real/webServices/Vect

```
orBase/release-
CURRENT/AsinensisChina/blast/AsinensisChinaAnnotatedProteins;
/eupath/data/apiSiteFilesStaging/VectorBase/49/real/webServices/Vect
orBase/release-
CURRENT/AsinensisSINENSIS/blast/AsinensisSINENSISAnnotatedProteins;
/eupath/data/apiSiteFilesStaging/VectorBase/49/real/webServices/Vect
orBase/release-
CURRENT/AstephensiIndian/blast/AstephensiIndianAnnotatedProteins;
/eupath/data/apiSiteFilesStaging/VectorBase/49/real/webServices/Vect
orBase/release-CURRENT/AstephensiSDA-500/blast/AstephensiSDA-
500AnnotatedProteins;
/eupath/data/apiSiteFilesStaging/VectorBase/49/real/webServices/Vect
orBase/release-
CURRENT/AstephensiUCISS2018/blast/AstephensiUCISS2018AnnotatedProtei
ns;
/eupath/data/apiSiteFilesStaging/VectorBase/49/real/webServices/Vect
orBase/release-
CURRENT/AziemanniAzieGA1/blast/AziemanniAzieGA1AnnotatedProteins;
/eupath/data/apiSiteFilesStaging/VectorBase/49/real/webServices/Vect
orBase/release-
CURRENT/CquinquefasciatusJHB2020/blast/CquinquefasciatusJHB2020Annot
atedProteins;
/eupath/data/apiSiteFilesStaging/VectorBase/49/real/webServices/Vect
orBase/release-
CURRENT/CquinquefasciatusJohannesburg/blast/CquinquefasciatusJohanne
sburgAnnotatedProteins;
/eupath/data/apiSiteFilesStaging/VectorBase/49/real/webServices/Vect
orBase/release-
CURRENT/ScyaneusScyaPA1/blast/ScyaneusScyaPA1AnnotatedProteins
           854,451 sequences; 547,869,893 total letters

Query=
Length=566
                                                                      Score     E
Sequences producing significant alignments:                          (Bits)  Value

AALF007356-PA | transcript=AALF007356-RA | gene=AALF007356 | orga...  149     6e-36
ASIC022140-PA | transcript=ASIC022140-RA | gene=ASIC022140 | orga...  147     2e-35
AALB20_033171.P66527 | transcript=AALB20_033171.R66527 | gene=AAL...  147     2e-35
AATE014116-PA | transcript=AATE014116-RA | gene=AATE014116 | orga...  147     2e-35
AFUN2_006054.P9868 | transcript=AFUN2_006054.R9868 | gene=AFUN2_0...  147     2e-35
AALB20_033171.P66512 | transcript=AALB20_033171.R66512 | gene=AAL...  147     2e-35
AALB20_033171.P66523 | transcript=AALB20_033171.R66523 | gene=AAL...  147     2e-35
ACMO_007424.P13504 | transcript=ACMO_007424.R13504 | gene=ACMO_00...  147     2e-35
ADAR2_001930.P21642 | transcript=ADAR2_001930.R21642 | gene=ADAR2...  147     2e-35
AMAC_003346.P3008 | transcript=AMAC_003346.R3008 | gene=AMAC_0033...  147     2e-35
AALB20_033171.P66516 | transcript=AALB20_033171.R66516 | gene=AAL...  147     2e-35
AMEM21_003341.P6430 | transcript=AMEM21_003341.R6430 | gene=AMEM2...  147     2e-35
ASTEI20_040830.P55056 | transcript=ASTEI20_040830.R55056 | gene=A...  147     2e-35
AGAMI1_007196.P14118 | transcript=AGAMI1_007196.R14118 | gene=AGA...  147     2e-35
ACON2_034794.P62462 | transcript=ACON2_034794.R62462 | gene=ACON2...  147     2e-35
AAQUA_001764.P1522 | transcript=AAQUA_001764.R1522 | gene=AAQUA_0...  147     2e-35
AARA21_009090.P17043 | transcript=AARA21_009090.R17043 | gene=AAR...  147     2e-35
ADAR2_001930.P21645 | transcript=ADAR2_001930.R21645 | gene=ADAR2...  147     2e-35
AMEM21_003341.P6434 | transcript=AMEM21_003341.R6434 | gene=AMEM2...  147     2e-35
AARA21_009090.P17046 | transcript=AARA21_009090.R17046 | gene=AAR...  147     2e-35
AARA21_009090.P17034 | transcript=AARA21_009090.R17034 | gene=AAR...  147     2e-35
AMEM21_003341.P6428 | transcript=AMEM21_003341.R6428 | gene=AMEM2...  147     3e-35
AFUN2_006054.P9870 | transcript=AFUN2_006054.R9870 | gene=AFUN2_0...  147     3e-35
ASTEI20_040830.P55060 | transcript=ASTEI20_040830.R55060 | gene=A...  147     3e-35
AGAMI1_007196.P14115 | transcript=AGAMI1_007196.R14115 | gene=AGA...  147     3e-35
ACON2_034794.P62473 | transcript=ACON2_034794.R62473 | gene=ACON2...  147     3e-35
ACMO_007424.P13500 | transcript=ACMO_007424.R13500 | gene=ACMO_00...  147     3e-35
AGAMI1_007196.P14116 | transcript=AGAMI1_007196.R14116 | gene=AGA...  147     3e-35
ACON2_034794.P62470 | transcript=ACON2_034794.R62470 | gene=ACON2...  147     3e-35
ACON029589-PB | transcript=ACON029589-RB | gene=ACON029589 | orga...  147     3e-35
AALB20_033171.P66493 | transcript=AALB20_033171.R66493 | gene=AAL...  147     3e-35
AMEM21_003341.P6419 | transcript=AMEM21_003341.R6419 | gene=AMEM2...  147     3e-35
ACMO_007424.P13494 | transcript=ACMO_007424.R13494 | gene=ACMO_00...  147     3e-35
```

```
ASTEI20_040830.P55059 | transcript=ASTEI20_040830.R55059 | gene=A...  147      3e-35
AMEM21_003341.P6424 | transcript=AMEM21_003341.R6424 | gene=AMEM2...  147      3e-35
ACMO_007424.P13508 | transcript=ACMO_007424.R13508 | gene=ACMO_00...  147      3e-35
AARA21_009090.P17035 | transcript=AARA21_009090.R17035 | gene=AAR...  147      3e-35
AGAP029589-PB | transcript=AGAP029589-RB | gene=AGAP029589 | orga...  147      3e-35
AMAC_003346.P2999 | transcript=AMAC_003346.R2999 | gene=AMAC_0033...  147      3e-35
AMAC_003346.P3007 | transcript=AMAC_003346.R3007 | gene=AMAC_0033...  147      3e-35
ASTEI20_040830.P55062 | transcript=ASTEI20_040830.R55062 | gene=A...  147      3e-35
ASTEI20_040830.P55052 | transcript=ASTEI20_040830.R55052 | gene=A...  147      3e-35
ACMO_007424.P13496 | transcript=ACMO_007424.R13496 | gene=ACMO_00...  147      3e-35
AALB20_033171.P66511 | transcript=AALB20_033171.R66511 | gene=AAL...  147      3e-35
AARA21_009090.P17045 | transcript=AARA21_009090.R17045 | gene=AAR...  147      3e-35
AARA21_009090.P17036 | transcript=AARA21_009090.R17036 | gene=AAR...  147      3e-35
ADAR2_001930.P21644 | transcript=ADAR2_001930.R21644 | gene=ADAR2...  147      3e-35
ADAR2_001930.P21647 | transcript=ADAR2_001930.R21647 | gene=ADAR2...  147      3e-35
AALB20_033171.P66510 | transcript=AALB20_033171.R66510 | gene=AAL...  147      3e-35
AMAC_003346.P3003 | transcript=AMAC_003346.R3003 | gene=AMAC_0033...  147      3e-35
ASTEI20_040830.P55068 | transcript=ASTEI20_040830.R55068 | gene=A...  147      3e-35
AFUN2_006054.P9879 | transcript=AFUN2_006054.R9879 | gene=AFUN2_0...  147      3e-35
AFUN2_006054.P9862 | transcript=AFUN2_006054.R9862 | gene=AFUN2_0...  147      3e-35
AMEM21_003341.P6433 | transcript=AMEM21_003341.R6433 | gene=AMEM2...  146      4e-35
AGAMI1_007196.P14110 | transcript=AGAMI1_007196.R14110 | gene=AGA...  146      4e-35
ACMO_007424.P13509 | transcript=ACMO_007424.R13509 | gene=ACMO_00...  146      4e-35
ACON2_034794.P62457 | transcript=ACON2_034794.R62457 | gene=ACON2...  146      4e-35
AAEL000597-PB | transcript=AAEL000597-RB | gene=AAEL000597 | orga...  146      4e-35
AALFPA_070705.P41263 | transcript=AALFPA_070705.R41263 | gene=AAL...  146      5e-35
AALC636_024560.P34471 | transcript=AALC636_024560.R34471 | gene=A...  146      5e-35
SCYAPA1_024866.P32618 | transcript=SCYAPA1_024866.R32618 | gene=S...  145      6e-35
CQUJHB012679.P19663 | transcript=CQUJHB012679.R19663 | gene=CQUJH...  145      7e-35
ASTEI02004-PA | transcript=ASTEI02004-RA | gene=ASTEI02004 | orga...  145      1e-34
ADAR2_001930.P21641 | transcript=ADAR2_001930.R21641 | gene=ADAR2...  144      1e-34
AALB20_033171.P66492 | transcript=AALB20_033171.R66492 | gene=AAL...  144      1e-34
AFAF019387-PA | transcript=AFAF019387-RA | gene=AFAF019387 | orga...  144      1e-34
AARA017833-PA | transcript=AARA017833-RA | gene=AARA017833 | orga...  145      1e-34
AARA21_009090.P17039 | transcript=AARA21_009090.R17039 | gene=AAR...  145      1e-34
ACMO_007424.P13503 | transcript=ACMO_007424.R13503 | gene=ACMO_00...  145      1e-34
ADAR2_001930.P21643 | transcript=ADAR2_001930.R21643 | gene=ADAR2...  145      1e-34
ANILCM1_038625.P14110 | transcript=ANILCM1_038625.R14110 | gene=A...  144      1e-34
AARA21_009090.P17044 | transcript=AARA21_009090.R17044 | gene=AAR...  144      1e-34
AARA21_009090.P17042 | transcript=AARA21_009090.R17042 | gene=AAR...  144      1e-34
AARA21_009090.P17038 | transcript=AARA21_009090.R17038 | gene=AAR...  144      1e-34
AARA21_009090.P17037 | transcript=AARA21_009090.R17037 | gene=AAR...  144      1e-34
AARA21_009090.P17033 | transcript=AARA21_009090.R17033 | gene=AAR...  144      1e-34
AALB20_033171.P66528 | transcript=AALB20_033171.R66528 | gene=AAL...  144      1e-34
AALB20_033171.P66526 | transcript=AALB20_033171.R66526 | gene=AAL...  144      1e-34
AALB20_033171.P66524 | transcript=AALB20_033171.R66524 | gene=AAL...  144      1e-34
AALB20_033171.P66522 | transcript=AALB20_033171.R66522 | gene=AAL...  144      1e-34
AALB20_033171.P66521 | transcript=AALB20_033171.R66521 | gene=AAL...  144      1e-34
AALB20_033171.P66520 | transcript=AALB20_033171.R66520 | gene=AAL...  144      1e-34
AALB20_033171.P66518 | transcript=AALB20_033171.R66518 | gene=AAL...  144      1e-34
AALB20_033171.P66515 | transcript=AALB20_033171.R66515 | gene=AAL...  144      1e-34
AALB20_033171.P66514 | transcript=AALB20_033171.R66514 | gene=AAL...  144      1e-34
AALB20_033171.P66509 | transcript=AALB20_033171.R66509 | gene=AAL...  144      1e-34
AALB20_033171.P66508 | transcript=AALB20_033171.R66508 | gene=AAL...  144      1e-34
AALB20_033171.P66507 | transcript=AALB20_033171.R66507 | gene=AAL...  144      1e-34
AALB20_033171.P66505 | transcript=AALB20_033171.R66505 | gene=AAL...  144      1e-34
AALB20_033171.P66503 | transcript=AALB20_033171.R66503 | gene=AAL...  144      1e-34
AALB20_033171.P66502 | transcript=AALB20_033171.R66502 | gene=AAL...  144      1e-34
AALB20_033171.P66500 | transcript=AALB20_033171.R66500 | gene=AAL...  144      1e-34
AALB20_033171.P66499 | transcript=AALB20_033171.R66499 | gene=AAL...  144      1e-34
AALB20_033171.P66497 | transcript=AALB20_033171.R66497 | gene=AAL...  144      1e-34
AALB20_033171.P66496 | transcript=AALB20_033171.R66496 | gene=AAL...  144      1e-34
AALB20_033171.P66495 | transcript=AALB20_033171.R66495 | gene=AAL...  144      1e-34
AALB20_033171.P66494 | transcript=AALB20_033171.R66494 | gene=AAL...  144      1e-34
AALB20_033171.P66491 | transcript=AALB20_033171.R66491 | gene=AAL...  144      1e-34
AALB20_033171.P66490 | transcript=AALB20_033171.R66490 | gene=AAL...  144      1e-34
ACMO_007424.P13499 | transcript=ACMO_007424.R13499 | gene=ACMO_00...  144      2e-34

>AALF007356-PA | transcript=AALF007356-RA | gene=AALF007356 |
organism=Aedes_albopictus_Foshan | gene_product=cadherin [Source:Projected
```

from Aedes aegypti (AAEL000597) VB Community Annotation] | transcript_product=cadherin [Source:Projected from Aedes aegypti (AAEL000597) VB Community Annotation] | location=JXUM01S001733:79044-136584(+) | protein_length=1569 | sequence_SO=supercontig | SO=protein_coding_gene | is_pseudo=false
Length=1569

```
 Score = 149 bits (375),  Expect = 6e-36, Method: Compositional matrix adjust.
 Identities = 132/456 (29%), Positives = 218/456 (48%), Gaps = 47/456 (10%)

Query  14   DNSKRNPIAKITSDYQATQKITYRISGVGID--QPPFGIFVVDKNTGDINITAIVDREET  71
            D   R  +     D    Q I Y ++G GID   P    F +++ +GDI +   +DR++
Sbjct  11   DGQNRKQVTATDGDKDRPQNIVYFLTGQGIDPDNPANSKFDINRTSGDIFVLKPLDRDQP  70

Query  72   ---PSFLITCRALNAQGLDVEKPLILTVKILDINDNPPVFSQQIFMGEIEENSASNSLVM  128
               P +  T  A +  G  +     + V + DINDN P+F Q ++ G + EN  +  +VM
Sbjct  71   NGRPQWRFTVFAQDEGGEGLVGYADVQVNLKDINDNAPIFPQGVYFGNVTENGTAGMVVM  130

Query  129  ILNATDADEPNH-LNSKIAFKI---VSQEPAGTPMFLLSRNTGEVRTLTNSLDREQASSY  184
             + A D D+PN   N+K+ + I   V +E  G+P+F +  +TG ++T    LDRE+   Y
Sbjct  131  TMTAVDYDDPNEGTNAKLIYSIEKNVIEEETGSPIFEIEADTGVIKTAVCCLDRERTPDY  190

Query  185  RLVVSGADKDGEGLSTQCECNIKVKDVNDNFPMFRDSQYSARIEENILSS--ELLRFQVT  242
             + V     DG GL      +I+VKD+ND  P F   ++   ++E   ++  E+    VT
Sbjct  191  SIQVVAM--DGGGLKGTGTASIRVKDINDMPPQFTKDEWFTEVDETDGTNLPEMPILTVT  248

Query  243  DLDEEYTDNWLAVYFFTSGNEGNWFEIQTDPRTNEGI--LKVVKALDYE-QLQS--VKLS  297
              DE+ T+ +       SG   + F +    R N+G   LK+V+ LDYE QLQS    +
Sbjct  249  VHDEDETNKFQYKVIDNSGYGADKFTMV---RNNDGTGSLKIVQPLDYEDQLQSNGFRFR  305

Query  298  IAVKNKAEFHQSVISRYRVQSTPVTIQVINVREGIAFRPASKTFTVQKGISSKKLVDYIL  357
            I V +K E + +   +Y V  + V +++ ++ +    +P  +   ++  +     V   L
Sbjct  306  IQVNDKGEDNDN--DKYHVAYSWVVVKLRDINDN---KPQFERPNIEVSVYENADVGKTL  360

Query  358  GTYQAIDEDTNKAASNVKYVMGRNDGGYLMIDSKTAEIKFVKNMNRDSTFIVNKTITAEV  417
             T++A D D     S V Y + R         S   + +F  ++N++ T  + + +  EV
Sbjct  361  ETFKATDPDQG-GRSKVSYAIDR---------SSDRQRQF--SINQEGTVTIQRQLDREV  408

Query  418  --------LAIDE-YTGKTSTGTVYVRVPDFNDNCP  444
                    LAID+    KT+T T+ V V D NDN P
Sbjct  409  TPRHQVKILAIDDGIPPKTATATLTVIVQDINDNPP  444

 Score = 53.1 bits (126),  Expect = 2e-05, Method: Compositional matrix adjust.
 Identities = 70/306 (23%), Positives = 125/306 (41%), Gaps = 30/306 (10%)

Query  20   PIAKIT-SDYQATQKITYRI---SGVGIDQPPFGIFVVDKNTGDINITAIVDRE---ETP  72
            PI  +T  D   T K  Y++   SG G D+  F +   +  TG + I   +D E   ++
Sbjct  243  PILTVTVHDEDETNKFQYKVIDNSGYGADK--FTMVRNNDGTGSLKIVQPLDYEDQLQSN  300

Query  73   SFLITCRALNAQGLDVEKPLI------LTVKILDINDNPPVFSQQIFMGEIEENSASNSL  126
             F    + +N +G D +          + VK+ DINDN P F +      + EN+
Sbjct  301  GFRFRIQ-VNDKGEDNDNDKYHVAYSWVVVKLRDINDNKPQFERPNIEVSVYENADVGKT  359

Query  127  VMILNATDADEPNHLNSKIAFKIVSQEPAGTPMFLLSRNTGEVRTLTNSLDREQASSYRL  186
            +    ATD D+      SK+++ I   + +       S N     T+   LDRE    +++
Sbjct  360  LETFKATDPDQGGR--SKVSYAI---DRSSDRQRQFSINQEGTVTIQRQLDREVTPRHQV  414

Query  187  VVSGADKDGEGLSTQCECNIKVKDVNDNFPMFRDSQYSARIEENILSSELLRFQVTDLDE  246
             +   D      +      + V+D+NDN P F    Y   + E++   +++    TD D+
Sbjct  415  KILAIDDGIPPKTATATLTVIVQDINDNPPKFL-KDYRPVLPEHVPPRKVVEILATDDDD  473

Query  247  EYTDNWLAVYFFTSGNEGNW----FEIQTDPRTNEG----ILKVVKALDYEQLQSVKLSI  298
                N     F       +     F+++ D +   G    I+  +++ D EQ +   + I
Sbjct  474  RSKSNGPPFQFRLDPGADDIIRASFKVEQDQKGANGDGMAIVSSLRSFDREQQKEYLIPI  533

Query  299  AVKNKA  304
             +K+
Sbjct  534  VIKDHG  539
```

```
 Score = 43.9 bits (102),  Expect = 0.011, Method: Compositional matrix adjust.
 Identities = 48/199 (24%), Positives = 73/199 (37%), Gaps = 19/199 (10%)

Query  27   DYQATQKITYRISGVGIDQPPFGIFVVDKNTGDINITAIVDREETPSFLITCRALNAQGL  86
            D     K++Y I      Q  F I       G + I   +DRE TP   +   A++
Sbjct  369  DQGGRSKVSYAIDRSSDRQRQFSI----NQEGTVTIQRQLDREVTPRHQVKILAIDDGIP  424

Query  87   DVEKPLILTVKILDINDNPPVFSQQIFMGEIEENSASNSLVMILNATDADEPNHLNSKIA  146
                   LTV + DINDNPP F +  +   + E+     +V IL   D D
Sbjct  425  PKTATATLTVIVQDINDNPPKFLKD-YRPVLPEHVPPRKVVEILATDDDDRSKSNGPPFQ  483

Query  147  FKIVSQEPAGTPMFLLSRNTGEVRTLTN-----------SLDREQASSYRLVVSGADKDG  195
            F++   +P    +   S    + +   N           S DREQ   Y + +   D
Sbjct  484  FRL---DPGADDIIRASFKVEQDQKGANGDGMAIVSSLRSFDREQQKEYLIPIVIKDHGN  540

Query  196  EGLSTQCECNIKVKDVNDN  214
              ++      + + DVNDN
Sbjct  541  PAMTGTSTLTVVIGDVNDN  559

>ASIC022140-PA | transcript=ASIC022140-RA | gene=ASIC022140 |
organism=Anopheles_sinensis_China | gene_product=unspecified
product | transcript_product=unspecified product |
location=KE525421:1028812-1084117(-)
| protein_length=1709 | sequence_SO=supercontig
| SO=protein_coding_gene | is_pseudo=false
Length=1709

 Score = 147 bits (371),  Expect = 2e-35, Method: Compositional matrix adjust.
 Identities = 128/446 (29%), Positives = 217/446 (49%), Gaps = 36/446 (8%)

Query  20   PIAKITS---DYQATQKITYRISGVGID--QPPFGIFVVDKNTGDINITAIVDREET---  71
            PI ++T+   D    Q I Y ++G GID   P    F +++ TG+I +   +DR++
Sbjct  149  PIMQVTATDGDKDRPQNIVYFLTGQGIDPDNPANSKFDINRTTGEIFVLKPLDRDQPNGR  208

Query  72   PSFLITCRALNAQGLDVEKPLILTVKILDINDNPPVFSQQIFMGEIEENSASNSLVMILN  131
            P +  T  A +  G  +     + V + DINDN P+F Q ++ G + EN  +  +VM +
Sbjct  209  PQWRFTVFAQDEGGEGLVGYADVQVNLKDINDNAPIFPQGVYFGNVTENGTAGMVVMTMT  268

Query  132  ATDADEPNH-LNSKIAFKI---VSQEPAGTPMFLLSRNTGEVRTLTNSLDREQASSYRLV  187
            A D D+PN   N+K+ + I   V +E  G+P+F +  +TG ++T    LDRE+   Y +
Sbjct  269  AVDYDDPNEGTNAKLIYSIEKNVIEEETGSPIFEIEADTGVIKTAVCCLDRERTPDYSIQ  328

Query  188  VSGADKDGEGLSTQCECNIKVKDVNDNFPMFRDSQYSARIEENILSS--ELLRFQVTDLD  245
            V     DG GL      +I+VKD+ND  P F   ++   ++E   ++  E+    VT  D
Sbjct  329  VVAM--DGGGLKGTGTASIRVKDINDMPPQFTKDEWFTEVDETDGTNLPEMPILTVTVHD  386

Query  246  EEYTDNWLAVYFFTSGNEGNWFEIQTDPRTNEGI--LKVVKALDYE---QLQSVKLSIAV  300
            E+ T+ +        SG   + F +    R N+G   LK+V+ LDYE   Q    +  I V
Sbjct  387  EDETNKFQYKVIDNSGYGADKFTMV---RNNDGTGSLKIVQPLDYEDPLQSNGFRFRIQV  443

Query  301  KNKAEFHQSVISRYRVQSTPVTIQVINVREGIAFRPASKTFTVQKGISSKKLVDYILGTY  360
             +K E + +   +Y V  + V +++ ++ +    +P  +   ++  +     V   L T+
Sbjct  444  NDKGEDNDN--DKYHVAYSWVVVKLRDINDN---KPQFERPNIEVSVYENADVGKTLETF  498

Query  361  QAIDEDTNKAASNVKYVMGR-NDGGYLMIDSKTAEIKFVKNMNRDSTFIVNKTITAEVLA  419
            +A D D     S V Y + R +D        ++   +  +N++R+    V       ++LA
Sbjct  499  KATDPDQG-GKSKVSYAIDRSSDRQRQFSINQEGTVTIQRNLDRE----VTPRHQVKILA  553

Query  420  IDE-YTGKTSTGTVYYRVPDFNDNCP  444
            ID+    KT+T T+ V V D NDN P
Sbjct  554  IDDGIPPKTATATLTVIVQDINDNPP  579

 Score = 87.4 bits (215),  Expect = 4e-16, Method: Compositional matrix adjust.
 Identities = 60/209 (29%), Positives = 100/209 (48%), Gaps = 13/209 (6%)

Query  27   DYQATQKITYRISGVGIDQPPFGIFVVDKNTGDINITAIVDREETPSFLITCRALNAQGL  86
            D+  + +I Y I+   I     G F V   TG I +   +D E    + +T  A ++
Sbjct  57   DHDESSRIRYEITSGNIG----GAFAVKNMTGAIYVAGALDYETRKRYHLTLVASDSLN-  111
```

```
Query  87   DVEKPLILTVKILDINDNPPVFSQQIFMGEIEENSASNSLVMILNATDADE--PNHLNSK  144
              E   I+ + + D+ND PP F QQ++   ++E  A    +M + ATD D+  P ++
Sbjct  112  --ENETIIVINVKDVNDMPPSFPQQVYERTMDEELAVPFPIMQVTATDGDKDRPQNIVYF  169

Query  145  IAFKIVSQEPAGTPMFLLSRNTGEVRTLTNSLDREQASS---YRLVVSGADKDGEGLSTQ  201
            +  + + +       F ++R TGE+  L   LDR+Q +    +R  V   D+ GEGL
Sbjct  170  LTGQGIDPDNPANSKFDINRTTGEIFVL-KPLDRDQPNGRPQWRFTVFAQDEGGEGLVGY  228

Query  202  CECNIKVKDVNDNFPMFRDSQYSARIEEN  230
             +  + +KD+NDN P+F    Y   + EN
Sbjct  229  ADVQVNLKDINDNAPIFPQGVYFGNVTEN  257

 Score = 72.8 bits (177),  Expect = 2e-11, Method: Compositional matrix adjust.
 Identities = 92/365 (25%), Positives = 154/365 (42%), Gaps = 38/365 (10%)

Query  93   ILTVKILDINDNPPVFSQQIFMGEIEENSASNSLVMILNATDADEPNHLNSKIAFKIVSQ  152
             + + I D NDNPP F + ++  E++EN      V+ + A D DE    +S+I ++I S
Sbjct  16   FIRIGIADKNDNPPYFDKALYEAEVDENEDIQHTVLTVTAKDHDE----SSRIRYEITSG  71

Query  153  EPAGTPMFLLSRNTGEVRTLTNSLDREQASSYRLVVSGADKDGEGLSTQCECNIKVKDVN  212
               G   F +   TG +  +  +LD E    Y L +  +D   E  +       I VKDVN
Sbjct  72   NIGGA--FAVKNMTGAI-YVAGALDYETRKRYHLTLVASDSLNENETIIV---INVKDVN  125

Query  213  DNFPMFRDSQYSARIEENI-LSSELLRFQVTDLDEEYTDNWLAVYFFTSG--NEGNWFEI  269
            D  P F    Y   ++E + +   +++   TD D++   N   VYF T    +  N
Sbjct  126  DMPPSFPQQVYERTMDEELAVPFPIMQVTATDGDKDRPQN--IVYFLTGQGIDPDNPANS  183

Query  270  QTDPRTNEGILKVVKALDYEQLQS-VKLSIAVKNKAEFHQSVISRYRVQSTPVTIQVINV  328
            + D     G + V+K LD +Q     +     V  + E  + ++    VQ     + + ++
Sbjct  184  KFDINRTTGEIFVLKPLDRDQPNGRPQWRFTVFAQDEGGEGLVGYADVQ-----VNLKDI  238

Query  329  REGIAFRPASKTFTVQKGISSKKLVDYILGTYQAIDEDTNKAASNVK--YVMGRN-----  381
             +     P    F     ++        ++ T  A+D D     +N K  Y + +N
Sbjct  239  NDNAPIFPQGVYF---GNVTENGTAGMVVMTMTAVDYDDPNEGTNAKLIYSIEKNVIEEE  295

Query  382  -DGGYLMIDSKTAEIK-FVKNMNRDSTFIVNKTITAEVLAIDEYTGKTSTGTVYVRVPDF  439
                   I++ T  IK  V  ++R+ T        + +V+A+D   G   TGT  +RV D
Sbjct  296  TGSPIFEIEADTGVIKTAVCCLDRERT----PDYSIQVVAMDG-GGLKGTGTASIRVKDI  350

Query  440  NDNCP  444
            ND  P
Sbjct  351  NDMPP  355

 Score = 53.5 bits (127),  Expect = 1e-05, Method: Compositional matrix adjust.
 Identities = 70/306 (23%), Positives = 126/306 (41%), Gaps = 30/306 (10%)

Query  20   PIAKIT-SDYQATQKITYRI---SGVGIDQPPFGIFVVDKNTGDINITAIVDRE---ETP  72
            PI  +T  D   T K  Y++   SG G D+  F +   +  TG + I   +D E   ++
Sbjct  378  PILTVTVHDEDETNKFQYKVIDNSGYGADK--FTMVRNNDGTGSLKIVQPLDYEDPLQSN  435

Query  73   SFLITCRALNAQGLDVEKPLI------LTVKILDINDNPPVFSQQIFMGEIEENSASNSL  126
             F    + +N +G D +          + VK+ DINDN P F +      + EN+
Sbjct  436  GFRFRIQ-VNDKGEDNDNDKYHVAYSWVVVKLRDINDNKPQFERPNIEVSVYENADVGKT  494

Query  127  VMILNATDADEPNHLNSKIAFKIVSQEPAGTPMFLLSRNTGEVRTLTNSLDREQASSYRL  186
            +    ATD D+     SK+++ I   + +        S N     T+  +LDRE    +++
Sbjct  495  LETFKATDPDQGG--KSKVSYAI---DRSSDRQRQFSINQEGTVTIQRNLDREVTPRHQV  549

Query  187  VVSGADKDGEGLSTQCECNIKVKDVNDNFPMFRDSQYSARIEENILSSELLRFQVTDLDE  246
             +   D       +       + V+D+NDN P F    Y   + E++   +++    TD D+
Sbjct  550  KILAIDDGIPPKTATATLTVIVQDINDNPPKFL-KDYRPVLPEHVPPRKVVEILATDDDD  608

Query  247  EYTDNWLAVYFFTSGNEGNW----FEIQTDPRTNEG----ILKVVKALDYEQLQSVKLSI  298
                N     F       +     F+++ D +   G    I+  +++ D EQ +   + I
Sbjct  609  RSKSNGPPFQFRLDPGADDIIRASFKVEQDQKGANGDGMAIVSSLRSFDREQQKEYLIPI  668

Query  299  AVKNKA  304
             +K+
```

```
Sbjct  669  VIKDHG  674

 Score = 44.3 bits (103),  Expect = 0.010, Method: Compositional matrix adjust.
 Identities = 48/199 (24%), Positives = 73/199 (37%), Gaps = 19/199 (10%)

Query  27   DYQATQKITYRISGVGIDQPPFGIFVVDKNTGDINITAIVDREETPSFLITCRALNAQGL  86
            D     K++Y I      Q  F I       G + I   +DRE TP   +   A++
Sbjct  504  DQGGKSKVSYAIDRSSDRQRQFSI----NQEGTVTIQRNLDREVTPRHQVKILAIDDGIP  559

Query  87   DVEKPLILTVKILDINDNPPVFSQQIFMGEIEENSASNSLVMILNATDADEPNHLNSKIA  146
                   LTV + DINDNPP F +  +   + E+     +V IL   D D
Sbjct  560  PKTATATLTVIVQDINDNPPKFLKD-YRPVLPEHVPPRKVVEILATDDDDRSKSNGPPFQ  618

Query  147  FKIVSQEPAGTPMFLLSRNTGEVRTLTN-----------SLDREQASSYRLVVSGADKDG  195
            F++   +P    +   S    + +   N           S DREQ   Y + +   D
Sbjct  619  FRL---DPGADDIIRASFKVEQDQKGANGDGMAIVSSLRSFDREQQKEYLIPIVIKDHGN  675

Query  196  EGLSTQCECNIKVKDVNDN  214
              ++      + + DVNDN
Sbjct  676  PAMTGTSTLTVVIGDVNDN  694

>AALB20_033171.P66527 | transcript=AALB20_033171.R66527 | gene=AALB20_033171
| organism=Anopheles_albimanus_STECLA_2020 |
gene_product=neural-cadherin, transcript variant X38 |
transcript_product=neural-cadherin,
transcript variant X35 | location=CM024292:75037560-75208393(+)
| protein_length=3122 | sequence_SO=chromosome
| SO=protein_coding_gene | is_pseudo=false
Length=3122

 Score = 147 bits (371),  Expect = 2e-35, Method: Compositional matrix adjust.
 Identities = 128/446 (29%), Positives = 217/446 (49%), Gaps = 36/446 (8%)

Query  20    PIAKITS---DYQATQKITYRISGVGID--QPPFGIFVVDKNTGDINITAIVDREET---  71
             PI ++T+   D    Q I Y ++G GID   P    F +++ TG+I +   +DR++
Sbjct  1562  PIMQVTATDGDKDRPQNIVYFLTGQGIDPDNPANSKFDINRTTGEIFVLKPLDRDQPNGR  1621

Query  72    PSFLITCRALNAQGLDVEKPLILTVKILDINDNPPVFSQQIFMGEIEENSASNSLVMILN  131
             P +  T  A +  G  +     + V + DINDN P+F Q ++ G + EN  +  +VM +
Sbjct  1622  PQWRFTVFAQDEGGEGLVGYADVQVNLKDINDNAPIFPQGVYFGNVTENGTAGMVVMTMT  1681

Query  132   ATDADEPNH-LNSKIAFKI---VSQEPAGTPMFLLSRNTGEVRTLTNSLDREQASSYRLV  187
             A D D+PN   N+K+ + I   V +E  G+P+F +  +TG ++T    LDRE+   Y +
Sbjct  1682  AVDYDDPNEGTNAKLIYSIEKNVIEEETGSPIFEIEADTGVIKTAVCCLDRERTPDYSIQ  1741

Query  188   VSGADKDGEGLSTQCECNIKVKDVNDNFPMFRDSQYSARIEENILSS--ELLRFQVTDLD  245
             V     DG GL      +I+VKD+ND  P F   ++   ++E   ++  E+    VT  D
Sbjct  1742  VVAM--DGGGLKGTGTASIRVKDINDMPPQFTKDEWFTEVDETDGTNLPEMPILTVTVHD  1799

Query  246   EEYTDNWLAVYFFTSGNEGNWFEIQTDPRTNEGI--LKVVKALDYE---QLQSVKLSIAV  300
             E+ T+ +       SG   + F +    R N+G   LK+V+ LDYE   Q    +  I V
Sbjct  1800  EDETNKFQYKVIDNSGYGADKFTMV---RNNDGTGSLKIVQPLDYEDPLQSNGFRFRIQV  1856

Query  301   KNKAEFHQSVISRYRVQSTPVTIQVINVREGIAFRPASKTFTVQKGISSKKLVDYILGTY  360
              +K E + +   +Y V  + V +++ ++ +    +P  +   ++  +     V   L T+
Sbjct  1857  NDKGEDNDN--DKYHVAYSWVVVKLRDINDN---KPQFERPNIEVSVYENADVGKTLETF  1911

Query  361   QAIDEDTNKAASNVKYVMGR-NDGGYLMIDSKTAEIKFVKNMNRDSTFIVNKTITAEVLA  419
             +A D D     S V Y + R +D       ++   +   +N++R+    V      ++LA
Sbjct  1912  KATDPDQG-GKSKVSYAIDRSSDRQRQFSINQEGTVTIQRNLDRE----VTPRHQVKILA  1966

Query  420   IDE-YTGKTSTGTVYVRVPDFNDNCP  444
             ID+    KT+T T+ V V D NDN P
Sbjct  1967  IDDGIPPKTATATLTVIVQDINDNPP  1992

 Score = 80.5 bits (197),  Expect = 6e-14, Method: Compositional matrix adjust.
 Identities = 107/424 (25%), Positives = 178/424 (42%), Gaps = 51/424 (12%)
```

```
Query  53    VDKNTGDINITAIVDREETPSFLITCRALNAQGLDVEKPLILTVKILDINDNPPVFSQ-Q  111
             +D++  ++N+TA+ D         +C  +N          ++ V I D+NDN PVF
Sbjct  950   LDRDKYELNVTAMDDG--------SC-CVNGDATIHTSTAVVVVFITDVNDNKPVFKDCG  1000

Query  112   IFMGEIEENSASNSLVMILNATDADEPNHLNSKIAFKIVSQEPAGTPMFLLSRNTGEVRT  171
              +  ++EE + + S V+ + ATD D+   +N ++ + IV Q        F +   TGEV T
Sbjct  1001  TYYPKVEEGAPNGSPVIKVQATDEDK--GVNGQVKYSIVQQPNQKGTKFTVDEETGEVST  1058

Query  172   LTNSLDRE--QASSYRLVVSGADKDGEGLSTQCECNIKVKDVNDNFPMFRDSQYSARIEE  229
                  DRE         + V   D+    L   C    +++ DVNDN P+F   +Y   +++
Sbjct  1059  -NKVFDREGDDGKFVSVTVKATDQGEPSLEGVCSFTVEITDVNDNPPLFDRQKYVENVKQ  1117

Query  230   NI-LSSELLRFQVTDLDEEYTDNWLAVYFFT---SGNEGNWFEIQTDPRTNEGILKVVKA  285
             +  + + +LR  V+  DE+  +N   VY  +   S N+  +FEIQ  P +   +LK  K
Sbjct  1118  DASIGTNILR--VSASDEDADNNGAIVYTLSAPHSPNDLEYFEIQ--PESGWIVLK--KP  1171

Query  286   LDYEQLQSVKLSIAVKNK------AEFHQSVISRYRVQSTPV----TIQVINVREGIAFR  335
             LD E+    +L +   ++      A+    +    R    P+         I++RE +
Sbjct  1172  LDRERY---RLKVYASDRGDPPSTADVDVELDVVDRNNKPPIWDRNIYGPIHIRENVTVG  1228

Query  336   PASKTFTVQKGISSKKLVDYILGTYQAIDEDTNKAASNVKYVMGRNDGGYLMIDSKTAEI  395
                       GI     V Y L          TNK   +  Y+  R D G         A+I
Sbjct  1229  TVVTAIKASSGIEGNPTVFYRL--MPGSTAQTNKL--HTFYLQQRPDNG-----DTWADI  1279

Query  396   KFVKNMNRDSTFIVNKTITAEVLAIDEYTGKTSTGTVYYRVPDFNDNCPTAV-LEKDAVC  454
             K    ++ ++    N TI  E     +     S  TVY+++ D ND  P     E++ V
Sbjct  1280  KVNHPLDFETIKEYNLTIRVENNGAQQL---ASEATVYIQLEDVNDEIPLFTEREQETVL  1336

Query  455   SSSP  458
                P
Sbjct  1337  EGEP  1340

 Score = 79.3 bits (194),  Expect = 1e-13, Method: Compositional matrix adjust.
 Identities = 104/419 (25%), Positives = 173/419 (41%), Gaps = 51/419 (12%)

Query  50    IFVVDKNTGDINITAIVDREETPSFLITCRAL----------NAQGLDVEKPLILTVKIL  99
              F ++  +G+I    + DRE+  ++ +   A            N Q   V K   + + I
Sbjct  1377  FFEINLQSGEIFTKVVFDREKQGAYALEVEARDGAPSARPNSNQQPNSVTK--FIRIGIA  1434

Query  100   DINDNPPVFSQQIFMGEIEENSASNSLVMILNATDADEPNHLNSKIAFKIVSQEPAGTPM  159
             D NDNPP F + ++  E++EN       V+ + A D DE    +S+I ++I S    G
Sbjct  1435  DKNDNPPYFDKALYEAEVDENEDIQHTVLTVTAKDHDE----SSRIRYEITSGNIGGA--  1488

Query  160   FLLSRNTGEVRTLTNSLDREQASSYRLVVSGADKDGEGLSTQCECNIKVKDVNDNFPMFR  219
             F +   TG +  +  +LD E     Y L +  +D   E  +       I VKDVND  P F
Sbjct  1489  FAVKNMTGAI-YVAGALDYETRKRYHLTLVASDSLNENATI---IEIHVKDVNDMPPTFS  1544

Query  220   DSQYSARI--EENILSSELLRFQVTDLDEEYTDNWLAVYFFTSG--NEGNWFEIQTDPRT  275
                Y A    E+ +    +++   TD D++   N   VYF T    +  N    + D
Sbjct  1545  QQIYEATQDEEQTVTDRPIMQVTATDGDKDRPQN--IVYFLTGQGIDPDNPANSKFDINR  1602

Query  276   NEGILKVVKALDYEQLQS-VKLSIAVKNKAEFHQSVISRYRVQSTPVTIQVINVREGIAF  334
               G + V+K LD +Q      +     V  + E  + ++     VQ      + + ++ +
Sbjct  1603  TTGEIFVLKPLDRDQPNGRPQWRFTVFAQDEGGEGLVGYADVQ-----VNLKDINDNAPI  1657

Query  335   RPASKTFTVQKGISSKKLVDYILGTYQAIDEDTNKAASNVK--YVMGRN------DGGYL  386
              P    F     ++        ++ T  A+D D     +N K  Y + +N
Sbjct  1658  FPQGVYF---GNVTENGTAGMVVMTMTAVDYDDPNEGTNAKLIYSIEKNVIEEETGSPIF  1714

Query  387   MIDSKTAEIK-FVKNMNRDSTFIVNKTITAEVLAIDEYTGKTSTGTVYYRVPDFNDNCP  444
              I++ T  IK  V  ++R+ T         + +V+A+D  G   TGT  +RV D ND  P
Sbjct  1715  EIEADTGVIKTAVCCLDRERT----PDYSIQVVAMDG-GGLKGTGTASIRVKDINDMPP  1768

 Score = 68.6 bits (166),  Expect = 4e-10, Method: Compositional matrix adjust.
 Identities = 88/361 (24%), Positives = 157/361 (43%), Gaps = 36/361 (10%)

Query  51    FVVDKNTGDINITAIVDRE-ETPSFL-ITCRALNAQGLDVEKPLILTVKILDINDNPPVF  108
             F VD+ TG+++    + DRE +    F+ +T +A +      +E      TV+I D+NDNPP+F
Sbjct  1047  FTVDEETGEVSTNKVFDREGDDGKFVSVTVKATDQGEPSLEGVCSFTVEITDVNDNPPLF  1106
```

```
Query  109   SQQIFMGEIEENSASNSLVMILNATDADEPNHLNSKIAFKIVS-QEPAGTPMFLLSRNTG  167
              +Q ++  ++++++  + ++ ++A+D D  N  N  I + + +   P     F +   +G
Sbjct  1107  DRQKYVENVKQDASIGTNILRVSASDEDADN--NGAIVYTLSAPHSPNDLEYFEIQPESG  1164

Query  168   EVRTLTNSLDREQASSYRLVVSGADKDGEGLSTQCECNIKVKDVNDNFPMFRDSQYSA--  225
              +  L   LDRE+   YRL V  +D+ G+  ST            +N P   D
Sbjct  1165  WI-VLKKPLDRER---YRLKVYASDR-GDPPSTADVDVELDVVDRNNKPPIWDRNIYGPI  1219

Query  226   RIEENILSSELLRFQVTDLDEEYTDNWLAVYFFTSG-----NEGNWFEIQTDPRTNE--G  278
              I EN+    ++             N    Y    G     N+ + F +Q  P   +
Sbjct  1220  HIRENVTVGTVV--TAIKASSGIEGNPTVFYRLMPGSTAQTNKLHTFYLQQRPDNGDTWA  1277

Query  279   ILKVVKALDYEQLQSVKLSIAVKNKAEFHQSVISRYRVQSTPVTIQVINVREGIAFRPAS  338
              +KV   LD+E ++   L+I V+N         ++       V IQ+ +V + I
Sbjct  1278  DIKVNHPLDFETIKEYNLTIRVENNG-------AQQLASEATVYIQLEDVNDEIPLFTER  1330

Query  339   KTFTVQKGISSKKLVDYILGTYQAIDEDTNKAASNVKYVM---GRNDG-GYLMIDSKTAE  394
             +  TV +G      V  +     AID+D     + V Y +    RN+G  +  I+ ++ E
Sbjct  1331  EQETVLEGEPIGTKVTQV----NAIDKDGTFPNNQVYYYIVDSPRNEGKDFFEINLQSGE  1386

Query  395   I  395
             I
Sbjct  1387  I  1387

 Score = 55.8 bits (133),  Expect = 3e-06, Method: Compositional matrix adjust.
 Identities = 49/158 (31%), Positives = 78/158 (49%), Gaps = 24/158 (15%)

Query  102   NDNPPVFSQQIFMGEIEENSASNSLVMILNATDADEPNHLNSKIAFKIVSQEPAGTP--M  159
             ND  P FSQQ++   ++EN+  N+LV  + A+D D  N     + F  V     GT
Sbjct  880   NDEEPKFSQQVYTPNVDENAGPNTLVTTVVASDKDGDN-----VRFGFVG---GGTSSGQ  931

Query  160   FLLSRNTGEVRTLTN--SLDREQASSYRLVVSGADK-------DGEGLSTQCECNIKVKD  210
             F++   TG +R  +   SLDR++   Y L V+  D        D    ++     + + D
Sbjct  932   FVIEEITGVIRLHSKAISLDRDK---YELNVTAMDDGSCCVNGDATIHTSTAVVVVFITD  988

Query  211   VNDNFPMFRD-SQYSARIEENILS-SELLRFQVTDLDE  246
             VNDN P+F+D   Y  ++EE   + S +++ Q TD D+
Sbjct  989   VNDNKPVFKDCGTYYPKVEEGAPNGSPVIKVQATDEDK  1026

 Score = 53.9 bits (128),  Expect = 1e-05, Method: Compositional matrix adjust.
 Identities = 70/306 (23%), Positives = 126/306 (41%), Gaps = 30/306 (10%)

Query  20    PIAKIT-SDYQATQKITYRI---SGVGIDQPPFGIFVVDKNTGDINITAIVDRE---ETP  72
             PI  +T  D   T K  Y++   SG G D+  F +   +  TG + I   +D E   ++
Sbjct  1791  PILTVTVHDEDETNKFQYKVIDNSGYGADK--FTMVRNNDGTGSLKIVQPLDYEDPLQSN  1848

Query  73    SFLITCRALNAQGLDVEKPLI------LTVKILDINDNPPVFSQQIFMGEIEENSASNSL  126
              F    + +N +G D +          + VK+ DINDN P F +      + EN+
Sbjct  1849  GFRFRIQ-VNDKGEDNDNDKYHVAYSWVVVKLRDINDNKPQFERPNIEVSVYENADVGKT  1907

Query  127   VMILNATDADEPNHLNSKIAFKIVSQEPAGTPMFLLSRNTGEVRTLTNSLDREQASSYRL  186
             +    ATD D+      SK+++ I   + +       S N     T+  +LDRE    +++
Sbjct  1908  LETFKATDPDQGG--KSKVSYAI---DRSSDRQRQFSINQEGTVTIQRNLDREVTPRHQV  1962

Query  187   VVSGADKDGEGLSTQCECNIKVKDVNDNFPMFRDSQYSARIEENILSSELLRFQVTDLDE  246
              +   D      +      + V+D+NDN P F    Y   + E++   +++    TD D+
Sbjct  1963  KILAIDDGIPPKTATATLTVIVQDINDNPPKFL-KDYRPVLPEHVPPRKVVEILATDDDD  2021

Query  247   EYTDNWLAVYFFTSGNEGNW----FEIQTDPRTNEG----ILKVVKALDYEQLQSVKLSI  298
                 N     F       +     F+++ D +   G    I+  +++ D EQ +   + I
Sbjct  2022  RSKSNGPPFQFRLDPGADDIIRASFKVEQDQKGANGDGMAIVSSLRSFDREQQKEYLIPI  2081

Query  299   AVKNKA  304
              +K+
Sbjct  2082  VIKDHG  2087

 Score = 52.0 bits (123),  Expect = 5e-05, Method: Compositional matrix adjust.
```

```
 Identities = 96/398 (24%), Positives = 163/398 (41%), Gaps = 54/398 (14%)

Query  106   PVFSQQIFMGEIEENSASNSLVMILNATDADEPNHLNSKIAFKIVSQ-EPAGTPMFLLSR  164
             P F    +  EI EN   +S ++ + A      +  + +I + + +Q + AGT  F +
Sbjct  677   PQFYMPSYEAEIPENQKKDSDIISVKAK-----SFADREIRYTLKAQGQGAGT--FNIGP  729

Query  165   NTGEVRTLTNSLDRE---QASSYRLVVSGADKDGEGLSTQCECNIKVKDVNDNFPMFRDS  221
              +G V+ L   LD E   Q   Y LVV+ A +D  G ST  E  I+V DVNDN P F
Sbjct  730   TSGIVK-LAKELDFEDLRQPHVYSLVVT-ATEDSGGFSTSVELTIRVTDVNDNAPKFELP  787

Query  222   QYSAR-IEENI-LSSELLRFQVTDLDEEYTDNWLAVYFFTSGNEGNWFEIQTDPRTNEGI  279
              Y A  ++E+I + + +L+ +  D D         + +  S +    F + ++     GI
Sbjct  788   DYQAHNVDEDIQIGTSILKVKAMDADSGAN---AEIEYLVSDDH---FAVDSN-----GI  836

Query  280   LKVVKALDYEQLQS-VKLSIAVKNKAEFHQSVISRYRVQSTPVTIQVINVREGIAFRPAS  338
             +   K LD +   +  +  +  K+K E  ++  +  RV +           E   F
Sbjct  837   IVNNKQLDADNNNAYYEFVVTAKDKGEPAKTGTATVRVYTKNKN------DEEPKFSQQV  890

Query  339   KTFTVQKGISSKKLVDYILGTYQAIDEDTNKAASNVK--YVMGRNDGGYLMIDSKTAEIK  396
              T  V +      LV  ++         ++K   NV+  +V G    G  +I+  T  I+
Sbjct  891   YTPNVDENAGPNTLVTTVVA--------SDKDGDNVRFGFVGGGTSSGQFVIEEITGVIR  942

Query  397   FVK---NMNRDSTFI-VNKTITAEVLAIDEYTGKTSTGTVYVRVPDFNDN------CPTA  446
                   +++RD   + V            + T  TST  V V + D NDN      C T
Sbjct  943   LHSKAISLDRDKYELNVTAMDDGSCCVNGDATIHTSTAVVVVFITDVNDNKPVFKDCGTY  1002

Query  447   VLE-KDAVCSSSPSVVVSARTLNNRYTGPYTFALEDQP  483
               + ++   + SP + V A   +    G   +++  QP
Sbjct  1003  YPKVEEGAPNGSPVIKVQATDEDKGVNGQVKYSIVQQP  1040

 Score = 44.7 bits (104),  Expect = 0.008, Method: Compositional matrix adjust.
 Identities = 48/199 (24%), Positives = 73/199 (37%), Gaps = 19/199 (10%)

Query  27    DYQATQKITYRISGVGIDQPPFGIFVVDKNTGDINITAIVDREETPSFLITCRALNAQGL  86
             D     K++Y I      Q  F I       G + I   +DRE TP   +   A++
Sbjct  1917  DQGGKSKVSYAIDRSSDRQRQFSI----NQEGTVTIQRNLDREVTPRHQVKILAIDDGIP  1972

Query  87    DVEKPLILTVKILDINDNPPVFSQQIFMGEIEENSASNSLVMILNATDADEPNHLNSKIA  146
                    LTV + DINDNPP F +  +   + E+     +V IL   D D
Sbjct  1973  PKTATATLTVIVQDINDNPPKFLKD-YRPVLPEHVPPRKVVEILATDDDDRSKSNGPPFQ  2031

Query  147   FKIVSQEPAGTPMFLLSRNTGEVRTLTN-----------SLDREQASSYRLVVSGADKDG  195
             F++   +P    +   S    + +   N           S DREQ   Y + +   D
Sbjct  2032  FRL---DPGADDIIRASFKVEQDQKGANGDGMAIVSSLRSFDREQQKEYLIPIVIKDHGN  2088

Query  196   EGLSTQCECNIKVKDVNDN  214
               ++      + + DVNDN
Sbjct  2089  PAMTGTSTLTVVIGDVNDN  2107

 Score = 40.8 bits (94),  Expect = 0.11, Method: Compositional matrix adjust.
 Identities = 55/242 (23%), Positives = 102/242 (42%), Gaps = 22/242 (9%)

Query  4    KFAKPCREGE--DNSKRNP-IAKITSDYQATQKITYRISGVGIDQPPFGIFVVDKNTGDI  60
            +F  P  E E  +N K++  I  + +   A ++I Y +   G      G F +   +G +
Sbjct  678  QFYMPSYEAEIPENQKKDSDIISVKAKSFADREIRYTLKAQGQGA---GTFNIGPTSGIV  734

Query  61   NITAIVDREE--TPSF--LITCRALNAQGLDVEKPLILTVKILDINDNPPVFSQQIFMGE  116
             +   +D E+   P    L+     ++ G      + LT+++ D+NDN P F    +
Sbjct  735  KLAKELDFEDLRQPHVYSLVVTATEDSGGFSTS--VELTIRVTDVNDNAPKFELPDYQAH  792

Query  117  -IEENSASNSLVMILNATDADEPNHLNSKIAFKIVSQEPAGTPMFLLSRNTGEVRTLTNS  175
             ++E+    + ++ + A DAD  +  N++I + +VS +      F +  N   V
Sbjct  793  NVDEDIQIGTSILKVKAMDAD--SGANAEIEY-LVSDD-----HFAVDSNGIIVNNKQLD  844

Query  176  LDREQASSYRLVVSGADKDGEGLSTQCECNIKVKDVNDNFPMFRDSQYSARIEENILSSE  235
             D   A  Y  VV+  DK     +      +  K+ ND  P F    Y+  ++EN   +
Sbjct  845  ADNNNA-YYEFVVTAKDKGEPAKTGTATVRVYTKNKNDEEPKFSQQVYTPNVDENAGPNT  903

Query  236  LL  237
```

```
             L+
Sbjct  904  LV  905

 Score = 39.3 bits (90),  Expect = 0.30, Method: Compositional matrix adjust.
 Identities = 55/225 (24%), Positives = 88/225 (39%), Gaps = 24/225 (11%)

Query  75   LITCRALNAQGLDVEKPLILTVKILDINDNPPVFSQQIF--MGEIEENSASNSLVMILNA  132
            +IT +  NA GL       + + + D+ND PP F  +       ++ N+  N+ V  L A
Sbjct  539  IITNQGHNA-GLKYTDNQRVIIHVKDVNDEPPYFINRPLPMQAVVQLNAPPNTPVFTLQA  597

Query  133  TDADEPNHLNSKIAFKIVSQEPAGTPMFLLSRNTGEVRTLTNSLDREQASSYRLVVSGAD  192
             D D     +  I + IV     G   F +   +G VRT    L  +    Y L V   D
Sbjct  598  RDPD----TDHNIHYFIVRDRTGG--RFEVDERSGVVRTRGTDL-FQLDMEYVLYVKAED  650

Query  193  KDGE----GLSTQCECNIKVKDVNDNFPMFRDSQYSARIEEN-ILSSELLRFQVTDLDEE  247
            ++G+       +  E  + +       P F    Y A I EN    S+++  +     +
Sbjct  651  QNGKVDDRRFQSTPEERLSIVG-GKRAPQFYMPSYEAEIPENQKKDSDIISVKA----KS  705

Query  248  YTDNWLAVYFFTSGNEGNWFEIQTDPRTNEGILKVVKALDYEQLQ  292
            + D  +       G     F I        GI+K+ K LD+E L+
Sbjct  706  FADREIRYTLKAQGQGAGTFNIGP----TSGIVKLAKELDFEDLR  746

>AATE014116-PA | transcript=AATE014116-RA | gene=AATE014116 |
organism=Anopheles_atroparvus_EBRO | gene_product=unspecified
product | transcript_product=unspecified product |
location=AatrE3_2R:13173248-13226058(-)
| protein_length=1803 | sequence_SO=chromosome
| SO=protein_coding_gene | is_pseudo=false
Length=1803

 Score = 147 bits (371),  Expect = 2e-35, Method: Compositional matrix adjust.
 Identities = 128/446 (29%), Positives = 217/446 (49%), Gaps = 36/446 (8%)

Query  20   PIAKITS---DYQATQKITYRISGVGID--QPPFGIFVVDKNTGDINITAIVDREET---  71
            PI ++T+   D    Q I Y ++G GID   P    F +++ TG+I +   +DR++
Sbjct  170  PIMQVTATDGDKDRPQNIVYFLTGQGIDPDNPANSKFDINRTTGEIFVLKPLDRDQPNGR  229

Query  72   PSFLITCRALNAQGLDVEKPLILTVKILDINDNPPVFSQQIFMGEIEENSASNSLVMILN  131
            P +  T  A +  G  +     + V + DINDN P+F Q ++ G + EN  +  +VM +
Sbjct  230  PQWRFTVFAQDEGGEGLVGYADVQVNLKDINDNAPIFPQGVYFGNVTENGTAGMVVMTMT  289

Query  132  ATDADEPNH-LNSKIAFKI---VSQEPAGTPMFLLSRNTGEVRTLTNSLDREQASSYRLV  187
            A D D+PN   N+K+ + I   V +E  G+P+F +  +TG ++T    LDRE+   Y +
Sbjct  290  AVDYDDPNEGTNAKLIYSIEKNVIEEETGSPIFEIEADTGVIKTAVCCLDRERTPDYSIQ  349

Query  188  VSGADKDGEGLSTQCECNIKVKDVNDNFPMFRDSQYSARIEENILSS--ELLRFQVTDLD  245
            V     DG GL       +I+VKD+ND  P F   ++   ++E   ++  E+    VT  D
Sbjct  350  VVAM--DGGGLKGTGTASIRVKDINDMPPQFTKDEWFTEVDETDGTNLPEMPILTVTVHD  407

Query  246  EEYTDNWLAVYFFTSGNEGNWFEIQTDPRTNEGI--LKVVKALDYE---QLQSVKLSIAV  300
            E+ T+ +         SG   + F +    R N+G   LK+V+ LDYE    Q    +  I V
Sbjct  408  EDETNKFQYKVIDNSGYGADKFTMV---RNNDGTGSLKIVQPLDYEDPLQSNGFRFRIQV  464

Query  301  KNKAEFHQSVISRYRVQSTPVTIQVINVREGIAFRPASKTFTVQKGISSKKLVDYILGTY  360
             +K E + +   +Y V  + V +++ ++ +    +P  +   ++  +      V   L T+
Sbjct  465  NDKGEDNDN--DKYHVAYSWVVVKLRDINDN---KPQFERPNIEVSVYENADVGKTLETF  519

Query  361  QAIDEDTNKAASNVKYVMGR-NDGGYLMIDSKTAEIKFVKNMNRDSTFIVNKTITAEVLA  419
            +A D D     S V Y + R +D        ++    +    +N++R+    V      ++LA
Sbjct  520  KATDPDQG-GKSKVSYAIDRSSDRQRQFSINQEGTVTIQRNLDRE----VTPRHQVKILA  574

Query  420  IDE-YTGKTSTGTVYVRVPDFNDNCP  444
            ID+    KT+T T+ V V D NDN P
Sbjct  575  IDDGIPPKTATATLTVIVQDINDNPP  600

 Score = 75.9 bits (185),  Expect = 2e-12, Method: Compositional matrix adjust.
 Identities = 57/192 (30%), Positives = 89/192 (46%), Gaps = 10/192 (5%)
```

```
Query  44   DQPPFGIFVVDKNTGDINITAIVDREETPSFLITCRALNAQGLDVEKPLILTVKILDIND  103
            D PP    V ++ T    IT   DR      L     L A     E   I+ + + D+ND
Sbjct  92   DNPP----VFERPTYRTQITEEDDRNLPKRVLQYHLTLVASDSLNENETIIVISVKDVND  147

Query  104  NPPVFSQQIFMGEIEENSASNSLVMILNATDADE--PNHLNSKIAFKIVSQEPAGTPMFL  161
             PP F QQ++   ++E  A    +M + ATD D+  P ++   +  + +  +      F
Sbjct  148  MPPSFPQQVYERTMDEELAVPFPIMQVTATDGDKDRPQNIVYFLTGQGIDPDNPANSKFD  207

Query  162  LSRNTGEVRTLTNSLDREQASS---YRLVVSGADKDGEGLSTQCECNIKVKDVNDNFPMF  218
            ++R TGE+  L   LDR+Q +    +R  V   D+ GEGL    +  + +KD+NDN P+F
Sbjct  208  INRTTGEIFVL-KPLDRDQPNGRPQWRFTVFAQDEGGEGLVGYADVQVNLKDINDNAPIF  266

Query  219  RDSQYSARIEEN  230
                Y   + EN
Sbjct  267  PQGVYFGNVTEN  278

 Score = 53.5 bits (127),  Expect = 1e-05, Method: Compositional matrix adjust.
 Identities = 70/306 (23%), Positives = 126/306 (41%), Gaps = 30/306 (10%)

Query  20   PIAKIT-SDYQATQKITYRI---SGVGIDQPPFGIFVVDKNTGDINITAIVDRE---ETP  72
            PI  +T  D   T K  Y++   SG G D+  F +   +  TG + I   +D E   ++
Sbjct  399  PILTVTVHDEDETNKFQYKVIDNSGYGADK--FTMVRNNDGTGSLKIVQPLDYEDPLQSN  456

Query  73   SFLITCRALNAQGLDVEKPLI------LTVKILDINDNPPVFSQQIFMGEIEENSASNSL  126
             F    + +N +G D +          + VK+ DINDN P F +      + EN+
Sbjct  457  GFRFRIQ-VNDKGEDNDNDKYHVAYSWVVVKLRDINDNKPQFERPNIEVSVYENADVGKT  515

Query  127  VMILNATDADEPNHLNSKIAFKIVSQEPAGTPMFLLSRNTGEVRTLTNSLDREQASSYRL  186
            +    ATD D+     SK+++ I   + +       S N     T+  +LDRE    +++
Sbjct  516  LETFKATDPDQGG--KSKVSYAI---DRSSDRQRQFSINQEGTVTIQRNLDREVTPRHQV  570

Query  187  VVSGADKDGEGLSTQCECNIKVKDVNDNFPMFRDSQYSARIEENILSSELLRFQVTDLDE  246
             +   D      +      + V+D+NDN P F    Y   + E++   +++    TD D+
Sbjct  571  KILAIDDGIPPKTATATLTVIVQDINDNPPKFL-KDYRPVLPEHVPPRKVVEILATDDDD  629

Query  247  EYTDNWLAVYFFTSGNEGNW----FEIQTDPRTNEG----ILKVVKALDYEQLQSVKLSI  298
                N     F       +     F+++ D +   G    I+  +++ D EQ +   + I
Sbjct  630  RSKSNGPPFQFRLDPGADDIIRASFKVEQDQKGANGDGMAIVSSLRSFDREQQKEYLIPI  689

Query  299  AVKNKA  304
             +K+
Sbjct  690  VIKDHG  695

 Score = 50.4 bits (119),  Expect = 1e-04, Method: Compositional matrix adjust.
 Identities = 40/134 (30%), Positives = 64/134 (48%), Gaps = 13/134 (10%)

Query  112  IFMGEIEENSASNSLVMILNATDADEPNHLNSKIAFKIVSQEPAGTPMFLLSRNTGEVRT  171
            ++  E++EN      V+ + A D DE    +S+I ++I S    G   F +   TG +
Sbjct  1    LYEAEVDENEDIQHTVLTVTAKDHDE----SSRIRYEITSGNIGGA--FAVKNMTGAI-Y  53

Query  172  LTNSLDREQASSYRLVVSGADKDGEGLSTQCECNIKVKDVNDNFPMFRDSQYSARIEENI  231
            +  +LD E    Y L ++ +D   E  +T     I VKDVNDN P+F    Y  +I E
Sbjct  54   VAGALDYETRKRYELRLAASDNLKENYTTVV---IHVKDVNDNPPVFERPTYRTQITEED  110

Query  232  ---LSSELLRFQVT  242
               L   +L++ +T
Sbjct  111  DRNLPKRVLQYHLT  124

 Score = 43.9 bits (102),  Expect = 0.011, Method: Compositional matrix adjust.
 Identities = 48/199 (24%), Positives = 73/199 (37%), Gaps = 19/199 (10%)

Query  27   DYQATQKITYRISGVGIDQPPFGIFVVDKNTGDINITAIVDREETPSFLITCRALNAQGL  86
            D     K++Y I      Q  F I       G + I   +DRE TP   +   A++
Sbjct  525  DQGGKSKVSYAIDRSSDRQRQFSI----NQEGTVTIQRNLDREVTPRHQVKILAIDDGIP  580

Query  87   DVEKPLILTVKILDINDNPPVFSQQIFMGEIEENSASNSLVMILNATDADEPNHLNSKIA  146
                   LTV + DINDNPP F +  +   + E+     +V IL   D D
Sbjct  581  PKTATATLTVIVQDINDNPPKFLKD-YRPVLPEHVPPRKVVEILATDDDDRSKSNGPPFQ  639
```

```
Query  147  FKIVSQEPAGTPMFLLSRNTGEVRTLTN-----------SLDREQASSYRLVVSGADKDG  195
            F++   +P    +   S    + +   N           S DREQ   Y + +   D
Sbjct  640  FRL---DPGADDIIRASFKVEQDQKGANGDGMAIVSSLRSFDREQQKEYLIPIVIKDHGN  696

Query  196  EGLSTQCECNIKVKDVNDN  214
              ++      + + DVNDN
Sbjct  697  PAMTGTSTLTVVIGDVNDN  715
```

Score = 36.6 bits (83),  Expect = 1.9, Method: Compositional matrix adjust.
Identities = 61/252 (24%), Positives = 103/252 (41%), Gaps = 28/252 (11%)

```
Query  206  IKVKDVNDNFPMFRDSQYSARIEENI-LSSELLRFQVTDLDEEYTDNWLAVYFFTSG--N  262
            I VKDVND  P F    Y   ++E + +   +++   TD D++   N   VYF T    +
Sbjct  140  ISVKDVNDMPPSFPQQVYERTMDEELAVPFPIMQVTATDGDKDRPQN--IVYFLTGQGID  197

Query  263  EGNWFEIQTDPRTNEGILKVVKALDYEQLQS-VKLSIAVKNKAEFHQSVISRYRVQSTPV  321
              N    + D     G + V+K LD +Q     +    V  + E  + ++    VQ
Sbjct  198  PDNPANSKFDINRTTGEIFVLKPLDRDQPNGRPQWRFTVFAQDEGGEGLVGYADVQ----  253

Query  322  TIQVINVREGIAFRPASKTFTVQKGISSKKLVDYILGTYQAIDEDTNKAASNVK--YVMG  379
             + + ++ +     P    F     ++       ++ T  A+D D     +N K  Y +
Sbjct  254  -VNLKDINDNAPIFPQGVYF---GNVTENGTAGMVVMTMTAVDYDDPNEGTNAKLIYSIE  309

Query  380  RN------DGGYLMIDSKTAEIK-FVKNMNRDSTFIVNKTITAEVLAIDEYTGKTSTGTV  432
            +N            I++ T  IK  V  ++R+ T       + +V+A+D   G   TGT
Sbjct  310  KNVIEEETGSPIFEIEADTGVIKTAVCCLDRERT----PDYSIQVVAMDG-GGLKGTGTA  364

Query  433  YVRVPDFNDNCP  444
             +RV D ND  P
Sbjct  365  SIRVKDINDMPP  376
```

>AFUN2_006054.P9868 | transcript=AFUN2_006054.R9868 | gene=AFUN2_006054
| organism=Anopheles_funestus_AfunGA1 | gene_product=neural-cadherin,
transcript variant X17, neural-cadherin,
transcript variant X22, neural-cadherin, transcript variant
X12, neural-cadherin, transcript variant X1, neural-cadherin,
transcript variant X15, neural-cadherin, transcript variant
X2, neural-cadherin, transcript variant X23, neural-cadherin,
transcript variant X26, neural-cadherin, transcript variant
X24, neural-cadherin, transcript variant X7, neural-cadherin,
transcript variant X6, neural-cadherin, transcript variant
X18, neural-cadherin, transcript variant X3, neural-cadherin,
transcript variant X9, neural-cadherin, transcript variant
X14, neural-cadherin, transcript variant X4, neural-cadherin,
transcript variant X25, neural-cadherin, transcript
variant X21, neural-cadherin, transcript variant X20, neural-cadherin,
transcript variant X19, neural-cadherin, transcript
variant X13, neural-cadherin, transcript variant X11, neural-cadherin,
transcript variant X16, neural-cadherin, transcript
variant X5, neural-cadherin, transcript variant X10,
neural-cadherin, transcript variant X8 | transcript_product=neural-cadherin,
transcript variant X23 | location=AfunGA1_2RL:82372181-82559663(-)
| protein_length=3108 | sequence_SO=chromosome
| SO=protein_coding_gene | is_pseudo=false
Length=3108

Score = 147 bits (371),  Expect = 2e-35, Method: Compositional matrix adjust.
Identities = 128/446 (29%), Positives = 217/446 (49%), Gaps = 36/446 (8%)

```
Query  20    PIAKITS---DYQATQKITYRISGVGID--QPPFGIFVVDKNTGDINITAIVDREET---  71
             PI ++T+   D    Q I Y ++G GID   P    F +++ TG+I +   +DR++
Sbjct  1549  PIMQVTATDGDKDRPQNIVYFLTGQGIDPDNPANSKFDINRTTGEIFVLKPLDRDQPNGR  1608

Query  72    PSFLITCRALNAQGLDVEKPLILTVKILDINDNPPVFSQQIFMGEIEENSASNSLVMILN  131
             P +  T  A +  G  +     + V + DINDN P+F Q ++ G + EN  +  +VM +
Sbjct  1609  PQWRFTVFAQDEGGEGLVGYADVQVNLKDINDNAPIFPQGVYFGNVTENGTAGMVVMTMT  1668

Query  132   ATDADEPNH-LNSKIAFKI---VSQEPAGTPMFLLSRNTGEVRTLTNSLDREQASSYRLV  187
```

```
             A D D+PN   N+K+ + I   V +E  G+P+F +  +TG ++T    LDRE+   Y +
Sbjct  1669  AVDYDDPNEGTNAKLIYSIEKNVIEEETGSPIFEIEADTGVIKTAVCCLDRERTPDYSIQ  1728

Query  188   VSGADKDGEGLSTQCECNIKVKDVNDNFPMFRDSQYSARIEENILSS--ELLRFQVTDLD  245
             V     DG GL      +I+VKD+ND  P F   ++   ++E   ++  E+    VT  D
Sbjct  1729  VVAM--DGGGLKGTGTASIRVKDINDMPPQFTKDEWFTEVDETDGTNLPEMPILTVTVHD  1786

Query  246   EEYTDNWLAVYFFTSGNEGNWFEIQTDPRTNEGI--LKVVKALDYE---QLQSVKLSIAV  300
             E+ T+ +        SG    + F +    R N+G   LK+V+ LDYE   Q    +  I V
Sbjct  1787  EDETNKFQYKVIDNSGYGADKFTMV---RNNDGTGSLKIVQPLDYEDPLQSNGFRFRIQV  1843

Query  301   KNKAEFHQSVISRYRVQSTPVTIQVINVREGIAFRPASKTFTVQKGISSKKLVDYILGTY  360
              +K E + +   +Y V  + V +++ ++ +    +P  +   ++  +      V   L T+
Sbjct  1844  NDKGEDNDN--DKYHVAYSWVVVKLRDINDN---KPQFERPNIEVSVYENADVGKTLETF  1898

Query  361   QAIDEDTNKAASNVKYVMGR-NDGGYLMIDSKTAEIKFVKNMNRDSTFIVNKTITAEVLA  419
             +A D D     S V Y + R +D        ++    +   +N++R+    V       ++LA
Sbjct  1899  KATDPDQG-GKSKVSYAIDRSSDRQRQFSINQEGTVTIQRNLDRE----VTPRHQVKILA  1953

Query  420   IDE-YTGKTSTGTVYYVRVPDFNDNCP  444
             ID+    KT+T T+ V V D NDN P
Sbjct  1954  IDDGIPPKTATATLTVIVQDINDNPP  1979

 Score = 78.6 bits (192),  Expect = 2e-13, Method: Compositional matrix adjust.
 Identities = 106/424 (25%), Positives = 180/424 (42%), Gaps = 51/424 (12%)

Query  53    VDKNTGDINITAIVDREETPSFLITCRALNAQGLDVEKPLILTVKILDINDNPPVFSQ-Q  111
             +D++  ++N+TA+ D          +C  +N          ++ V I D+NDN PVF
Sbjct  938   LDRDKYELNVTAMDDG---------SC-CVNGDATIHTSTAVVVVFITDVNDNKPVFKDCG  988

Query  112   IFMGEIEENSASNSLVMILNATDADEPNHLNSKIAFKIVSQEPAGTPMFLLSRNTGEVRT  171
              +  ++EE + + S V+ + ATD D+    +N ++ + IV Q        F +   TGEV T
Sbjct  989   TYYPKVEEGAPNGSPVIKVQATDEDK--GVNGQVKYSIVQQPNQKGTKFTVDEETGEVST  1046

Query  172   LTNSLDRE--QASSYRLVVSGADKDGEGLSTQCECNIKVKDVNDNFPMFRDSQYSARIEE  229
                  DRE          + V   D+     L   C    +++ DVNDN P+F    +Y   +++
Sbjct  1047  -NKVFDREGDDGKFVSVTVKATDQGEPSLEGVCSFTVEITDVNDNPPLFDRQKYVENVKQ  1105

Query  230   NI-LSSELLRFQVTDLDEEYTDNWLAVYFFT---SGNEGNWFEIQTDPRTNEGILKVVKA  285
             +  + + +LR  V+  DE+  +N   VY  +   S N+  +FEIQ  P +   +LK  K
Sbjct  1106  DASIGTNILR--VSASDEDADNNGAIVYTLSAPHSPNDLEYFEIQ--PESGWIVLK--KP  1159

Query  286   LDYEQLQSVKLSIAVKNK------AEFHQSVISRYRVQSTPVTIQV----INVREGIAFR  335
             LD E+    +L +   ++       A+    +    R    P+  +     I++RE +
Sbjct  1160  LDRERY---RLKVYASDRGDPPSTADVDVELDVVDRNNKPPIWDRTIYGPIHIRENVTVG  1216

Query  336   PASKTFTVQKGISSKKLVDYILGTYQAIDEDTNKAASNVKYVMGRNDGGYLMIDSKTAEI  395
                 +     GI     V Y L          TNK   +  Y+  R D G         A+I
Sbjct  1217  TVVTSVKASSGIEGNPTVFYRL--MPGSTAQTNKL--HTFYLQQRPDNG-----DTWADI  1267

Query  396   KFVKNMNRDSTFIVNKTITAEVLAIDEYTGKTSTGTVYYVRVPDFNDNCPTAV-LEKDAVC  454
             K    ++ ++    N T+  E     +     S  TVY+++ D ND  P     E++ V
Sbjct  1268  KVNHPLDFETIKEYNLTVRVENNGAQQL---ASEATVYIQLEDVNDEIPLFTEREQETVL  1324

Query  455   SSSP  458
                P
Sbjct  1325  EGEP  1328

 Score = 77.0 bits (188),  Expect = 8e-13, Method: Compositional matrix adjust.
 Identities = 103/418 (25%), Positives = 174/418 (42%), Gaps = 50/418 (12%)

Query  50    IFVVDKNTGDINITAIVDREETPSFLITCRAL----------NAQGLDVEKPLILTVKIL  99
              F ++  +G+I    + DRE+  ++ +   A            N Q   V K   + + I
Sbjct  1365  FFEINLQSGEIFTKVVFDREKQGAYALEVEARDGAPSARPNSNQQPNSVTK--FIRIGIA  1422

Query  100   DINDNPPVFSQQIFMGEIEENSASNSLVMILNATDADEPNHLNSKIAFKIVSQEPAGTPM  159
             D NDNPP F + ++  E++EN       V+ + A D DE    +S+I ++I S    G
Sbjct  1423  DKNDNPPYFDKALYEAEVDENEDIQHTVLTVTAKDHDE----SSRIRYEITSGNIGGA--  1476
```

```
Query  160   FLLSRNTGEVRTLTNSLDREQASSYRLVVSGADKDGEGLSTQCECNIKVKDVNDNFPMFR  219
             F +   TG +  +  +LD E    Y L +  +D   E  +       I VKDVND  P F
Sbjct  1477  FAVKNMTGAI-YVAGALDYETRKRYHLTLVASDSLNENETIIV---INVKDVNDMPPSFP  1532

Query  220   DSQYSARIEENI-LSSELLRFQVTDLDEEYTDNWLAVYFFTSG--NEGNWFEIQTDPRTN  276
                Y   ++E + +   +++   TD D++   N   VYF T    +  N    + D
Sbjct  1533  QQVYERTMDEELAVPFPIMQVTATDGDKDRPQN--IVYFLTGQGIDPDNPANSKFDINRT  1590

Query  277   EGILKVVKALDYEQLQS-VKLSIAVKNKAEFHQSVISRYRVQSTPVTIQVINVREGIAFR  335
              G + V+K LD +Q     +    V  + E  + ++    VQ      + + ++ +
Sbjct  1591  TGEIFVLKPLDRDQPNGRPQWRFTVFAQDEGGEGLVGYADVQ-----VNLKDINDNAPIF  1645

Query  336   PASKTFTVQKGISSKKLVDYILGTYQAIDEDTNKAASNVK--YVMGRN------DGGYLM  387
             P    F      ++        ++ T  A+D D     +N K  Y + +N
Sbjct  1646  PQGVYF---GNVTENGTAGMVVMTMTAVDYDDPNEGTNAKLIYSIEKNVIEEETGSPIFE  1702

Query  388   IDSKTAEIK-FVKNMNRDSTFIVNKTITAEVLAIDEYTGKTSTGTVYVRVPDFNDNCP  444
             I++ T  IK  V  ++R+ T        + +V+A+D   G   TGT  +RV D ND  P
Sbjct  1703  IEADTGVIKTAVCCLDRERT----PDYSIQVVAMDG-GGLKGTGTASIRVKDINDMPP  1755

 Score = 65.5 bits (158),  Expect = 3e-09, Method: Compositional matrix adjust.
 Identities = 86/361 (24%), Positives = 155/361 (43%), Gaps = 36/361 (10%)

Query  51    FVVDKNTGDINITAIVDRE-ETPSFL-ITCRALNAQGLDVEKPLILTVKILDINDNPPVF  108
             F VD+ TG+++    + DRE +    F+ +T +A +      +E      TV+I D+NDNPP+F
Sbjct  1035  FTVDEETGEVSTNKVFDREGDDGKFVSVTVKATDQGEPSLEGVCSFTVEITDVNDNPPLF  1094

Query  109   SQQIFMGEIEENSASNSLVMILNATDADEPNHLNSKIAFKIVS-QEPAGTPMFLLSRNTG  167
              +Q ++  ++++++  + ++ ++A+D D  N  N  I + + +   P     F +   +G
Sbjct  1095  DRQKYVENVKQDASIGTNILRVSASDEDADN--NGAIVYTLSAPHSPNDLEYFEIQPESG  1152

Query  168   EVRTLTNSLDREQASSYRLVVSGADKDGEGLSTQCECNIKVKDVNDNFPMFRDSQYSARI  227
              +  L   LDRE+   YRL V  +D+ G+  ST              +N P   D      I
Sbjct  1153  WI-VLKKPLDRER---YRLKVYASDR-GDPPSTADVDVELDVVDRNNKPPIWDRTIYGPI  1207

Query  228   EENILSSELLRFQVTDLDEEYTDNWLAVYFFTSG-----NEGNWFEIQTDPRTNE--GIL  280
                   +                 N    Y    G     N+ + F +Q  P  +    +
Sbjct  1208  HIRENVTVGTVVTSVKASSGIEGNPTVFYRLMPGSTAQTNKLHTFYLQQRPDNGDTWADI  1267

Query  281   KVVKALDYEQLQSVKLSIAVKNKAEFHQSVISRYRVQSTPVTIQVINVREGIAFRPASKT  340
             KV   LD+E ++   L++ V+N          ++        V IQ+ +V + I      +
Sbjct  1268  KVNHPLDFETIKEYNLTVRVENNG-------AQQLASEATVYIQLEDVNDEIPLFTEREQ  1320

Query  341   FTVQKG--ISSKKLVDYILGTYQAIDEDTNKAASNVKYVM---GRNDG-GYLMIDSKTAE  394
              TV +G  I +K      +    AID+D     + V Y +    RN+G  +  I+ ++ E
Sbjct  1321  ETVLEGEPIGTK------VTQVNAIDKDGTFPNNQVYYYIVDSPRNEGKDFFEINLQSGE  1374

Query  395   I  395
             I
Sbjct  1375  I  1375

 Score = 55.8 bits (133),  Expect = 3e-06, Method: Compositional matrix adjust.
 Identities = 49/158 (31%), Positives = 78/158 (49%), Gaps = 24/158 (15%)

Query  102   NDNPPVFSQQIFMGEIEENSASNSLVMILNATDADEPNHLNSKIAFKIVSQEPAGTP--M  159
             ND  P FSQQ++   ++EN+  N+LV  + A+D D  N     + F  V     GT
Sbjct  868   NDEEPKFSQQVYTPNVDENAGPNTLVTTVVASDKDGDN-----VRFGFVG---GGTSSGQ  919

Query  160   FLLSRNTGEVRTLTN--SLDREQASSYRLVVSGADK-------DGEGLSTQCECNIKVKD  210
             F++   TG +R  +   SLDR++   Y L V+  D         D    ++     + + D
Sbjct  920   FVIEEITGVIRLHSKAISLDRDK---YELNVTAMDDGSCCVNGDATIHTSTAVVVVFITD  976

Query  211   VNDNFPMFRD-SQYSARIEENILS-SELLRFQVTDLDE  246
             VNDN P+F+D   Y  ++EE   + S +++ Q TD D+
Sbjct  977   VNDNKPVFKDCGTYYPKVEEGAPNGSPVIKVQATDEDK  1014

 Score = 53.9 bits (128),  Expect = 1e-05, Method: Compositional matrix adjust.
 Identities = 70/306 (23%), Positives = 126/306 (41%), Gaps = 30/306 (10%)
```

```
Query  20    PIAKIT-SDYQATQKITYRI---SGVGIDQPPFGIFVVDKNTGDINITAIVDRE---ETP  72
             PI  +T  D   T K  Y++   SG G D+  F +   +  TG + I   +D E   ++
Sbjct  1778  PILTVTVHDEDETNKFQYKVIDNSGYGADK--FTMVRNNDGTGSLKIVQPLDYEDPLQSN  1835

Query  73    SFLITCRALNAQGLDVEKPLI------LTVKILDINDNPPVFSQQIFMGEIEENSASNSL  126
              F    + +N +G D +          + VK+ DINDN P F +      + EN+
Sbjct  1836  GFRFRIQ-VNDKGEDNDNDKYHVAYSWVVVKLRDINDNKPQFERPNIEVSVYENADVGKT  1894

Query  127   VMILNATDADEPNHLNSKIAFKIVSQEPAGTPMFLLSRNTGEVRTLTNSLDREQASSYRL  186
             +    ATD D+     SK+++ I   + +       S N     T+  +LDRE    +++
Sbjct  1895  LETFKATDPDQGG--KSKVSYAI---DRSSDRQRQFSINQEGTVTIQRNLDREVTPRHQV  1949

Query  187   VVSGADKDGEGLSTQCECNIKVKDVNDNFPMFRDSQYSARIEENILSSELLRFQVTDLDE  246
              +   D       +       + V+D+NDN P F    Y   + E++   +++    TD D+
Sbjct  1950  KILAIDDGIPPKTATATLTVIVQDINDNPPKFL-KDYRPVLPEHVPPRKVVEILATDDDD  2008

Query  247   EYTDNWLAVYFFTSGNEGNW----FEIQTDPRTNEG----ILKVVKALDYEQLQSVKLSI  298
                 N     F        +     F+++ D +   G    I+  +++ D EQ +   + I
Sbjct  2009  RSKSNGPPFQFRLDPGADDIIRASFKVEQDQKGANGDGMAIVSSLRSFDREQQKEYLIPI  2068

Query  299   AVKNKA  304
              +K+
Sbjct  2069  VIKDHG  2074

 Score = 52.8 bits (125),  Expect = 3e-05, Method: Compositional matrix adjust.
 Identities = 97/398 (24%), Positives = 163/398 (41%), Gaps = 54/398 (14%)

Query  106   PVFSQQIFMGEIEENSASNSLVMILNATDADEPNHLNSKIAFKIVSQ-EPAGTPMFLLSR  164
             P F    +  EI EN   +S ++ + A       +  + +I + + +Q + AGT  F +
Sbjct  665   PQFYMPSYEAEIPENQKKDSDIISVKAK-----SFADREIRYTLKAQGQGAGT--FNIGP  717

Query  165   NTGEVRTLTNSLDRE---QASSYRLVVSGADKDGEGLSTQCECNIKVKDVNDNFPMFRDS  221
              +G V+ L   LD E   Q   Y LVV+ A +D  G ST  E  I+V DVNDN P F
Sbjct  718   TSGIVK-LAKELDFEDLRQPHVYSLVVT-ATEDSGGFSTSVELTIRVTDVNDNAPKFELP  775

Query  222   QYSAR-IEENI-LSSELLRFQVTDLDEEYTDNWLAVYFFTSGNEGNWFEIQTDPRTNEGI  279
              Y A  ++E+I L + +L+ +  D D          + +  S +    F + ++     GI
Sbjct  776   DYQAHNVDEDIPLGTSILKVKAMDADSGAN---AEIEYLVSDDH---FAVDSN-----GI  824

Query  280   LKVVKALDYEQLQS-VKLSIAVKNKAEFHQSVISRYRVQSTPVTIQVINVREGIAFRPAS  338
             +   K LD +   +  +  +  K+K E  ++  +  RV +             E   F
Sbjct  825   IVNNKQLDADNNNAYYEFVVTAKDKGEPAKTGTATVRVYTKNKN------DEEPKFSQQV  878

Query  339   KTFTVQKGISSKKLVDYILGTYQAIDEDTNKAASNVK--YVMGRNDGGYLMIDSKTAEIK  396
              T  V +      LV  ++          ++K   NV+  +V G    G  +I+  T  I+
Sbjct  879   YTPNVDENAGPNTLVTTVVA--------SDKDGDNVRFGFVGGGTSSGQFVIEEITGVIR  930

Query  397   FVK---NMNRDSTFI-VNKTITAEVLAIDEYTGKTSTGTVYVRVPDFNDN------CPTA  446
                   +++RD   + V              + T  TST  V V + D NDN       C T
Sbjct  931   LHSKAISLDRDKYELNVTAMDDGSCCVNGDATIHTSTAVVVVFITDVNDNKPVFKDCGTY  990

Query  447   VLE-KDAVCSSSPSVVVSARTLNNRYTGPYTFALEDQP  483
               + ++   + SP + V A   +    G   +++  QP
Sbjct  991   YPKVEEGAPNGSPVIKVQATDEDKGVNGQVKYSIVQQP  1028

 Score = 44.7 bits (104),  Expect = 0.008, Method: Compositional matrix adjust.
 Identities = 48/199 (24%), Positives = 73/199 (37%), Gaps = 19/199 (10%)

Query  27    DYQATQKITYRISGVGIDQPPFGIFVVDKNTGDINITAIVDREETPSFLITCRALNAQGL  86
             D     K++Y I       Q  F I        G + I   +DRE TP   +    A++
Sbjct  1904  DQGGKSKVSYAIDRSSDRQRQFSI----NQEGTVTIQRNLDREVTPRHQVKILAIDDGIP  1959

Query  87    DVEKPLILTVKILDINDNPPVFSQQIFMGEIEENSASNSLVMILNATDADEPNHLNSKIA  146
                    LTV + DINDNPP F +  +    + E+     +V IL   D D
Sbjct  1960  PKTATATLTVIVQDINDNPPKFLKD-YRPVLPEHVPPRKVVEILATDDDDRSKSNGPPFQ  2018

Query  147   FKIVSQEPAGTPMFLLSRNTGEVRTLTN-----------SLDREQASSYRLVVSGADKDG  195
             F++   +P    +   S    + +   N           S DREQ   Y + +   D
```

```
Sbjct  2019  FRL---DPGADDIIRASFKVEQDQKGANGDGMAIVSSLRSFDREQQKEYLIPIVIKDHGN  2075

Query  196   EGLSTQCECNIKVKDVNDN  214
               ++      + + DVNDN
Sbjct  2076  PAMTGTSTLTVVIGDVNDN  2094

 Score = 40.8 bits (94),  Expect = 0.11, Method: Compositional matrix adjust.
 Identities = 54/235 (23%), Positives = 99/235 (42%), Gaps = 22/235 (9%)

Query  4    KFAKPCREGE--DNSKRNP-IAKITSDYQATQKITYRISGVGIDQPPFGIFVVDKNTGDI  60
            +F  P  E E  +N K++  I  + +   A ++I Y +   G      G F +   +G +
Sbjct  666  QFYMPSYEAEIPENQKKDSDIISVKAKSFADREIRYTLKAQGQGA---GTFNIGPTSGIV  722

Query  61   NITAIVDREE--TPSF--LITCRALNAQGLDVEKPLILTVKILDINDNPPVFSQQIFMGE  116
             +   +D E+   P    L+     ++ G      + LT+++ D+NDN P F    +
Sbjct  723  KLAKELDFEDLRQPHVYSLVVTATEDSGGFSTS--VELTIRVTDVNDNAPKFELPDYQAH  780

Query  117  -IEENSASNSLVMILNATDADEPNHLNSKIAFKIVSQEPAGTPMFLLSRNTGEVRTLTNS  175
             ++E+    + ++ + A DAD  +  N++I + +VS +      F +  N   V
Sbjct  781  NVDEDIPLGTSILKVKAMDAD--SGANAEIEY-LVSDD-----HFAVDSNGIIVNNKQLD  832

Query  176  LDREQASSYRLVVSGADKDGEGLSTQCECNIKVKDVNDNFPMFRDSQYSARIEEN  230
             D   A  Y  VV+  DK     +      +  K+ ND  P F    Y+  ++EN
Sbjct  833  ADNNNA-YYEFVVTAKDKGEPAKTGTATVRVYTKNKNDEEPKFSQQVYTPNVDEN  886

 Score = 39.3 bits (90),  Expect = 0.29, Method: Compositional matrix adjust.
 Identities = 89/389 (23%), Positives = 149/389 (38%), Gaps = 62/389 (16%)

Query  75   LITCRALNAQGLDVEKPLILTVKILDINDNPPVFSQQIF--MGEIEENSASNSLVMILNA  132
            +IT +  NA GL       + + + D+ND PP F  +       ++ N+  N+ V  L A
Sbjct  527  IITNQGHNA-GLKYTDNQRVIIHVKDVNDEPPYFINRPLPMQAVVQLNAPPNTPVFTLQA  585

Query  133  TDADEPNHLNSKIAFKIVSQEPAGTPMFLLSRNTGEVRTLTNSLDREQASSYRLVVSGAD  192
             D D     +  I + IV     G   F +   +G VRT    L  +    Y L V   D
Sbjct  586  RDPD----TDHNIHYFIVRDRTGG--RFEVDERSGVVRTRGTDL-FQLDMEYVLYVKAED  638

Query  193  KDGE----GLSTQCECNIKVKDVNDNFPMFRDSQYSARIEEN-ILSSELLRFQVTDLDEE  247
            ++G+       +  E  + +       P F    Y A I EN    S+++  +     +
Sbjct  639  QNGKVDDRRFQSTPEERLSIVG-GKRAPQFYMPSYEAEIPENQKKDSDIISVKA----KS  693

Query  248  YTDNWLAVYFFTSGNEGNWFEIQTDPRTNEGILKVVKALDYEQLQSVKLS----IAVKNK  303
            + D  +       G     F I        GI+K+ K LD+E L+   +      A ++
Sbjct  694  FADREIRYTLKAQGQGAGTFNIGP----TSGIVKLAKELDFEDLRQPHVYSLVVTATEDS  749

Query  304  AEFHQSVISRYRVQSTPVTIQVINVREGIAFRPASKTFTVQKGISSKKLVDYILGT----  359
              F  SV          +TI+V +V +       +  F +    +     D  LGT
Sbjct  750  GGFSTSV---------ELTIRVTDVNDN------APKFELPDYQAHNVDEDIPLGTSILK  794

Query  360  YQAIDEDTNKAASNVKYVMGRN----DGGYLMIDSKTAEIKFVKNMNRDSTFIVNKTITA  415
             +A+D D+  A + ++Y++  +    D   +++++K  +     N N    F+V
Sbjct  795  VKAMDADSG-ANAEIEYLVSDDHFAVDSNGIIVNNKQLD---ADNNNAYYEFVVTAKDKG  850

Query  416  EVLAIDEYTGKTSTGTVYVRVPDFNDNCP  444
            E         KT T TV V   + ND  P
Sbjct  851  E-------PAKTGTATVRVYTKNKNDEEP  872

>AALB20_033171.P66512 | transcript=AALB20_033171.R66512 | gene=AALB20_033171
| organism=Anopheles_albimanus_STECLA_2020 |
gene_product=neural-cadherin, transcript variant X38 |
transcript_product=neural-cadherin,
transcript variant X25 | location=CM024292:75037560-75208393(+)
| protein_length=3130 | sequence_SO=chromosome
| SO=protein_coding_gene | is_pseudo=false
Length=3130

 Score = 147 bits (371),  Expect = 2e-35, Method: Compositional matrix adjust.
 Identities = 128/446 (29%), Positives = 217/446 (49%), Gaps = 36/446 (8%)
```

```
Query  20    PIAKITS---DYQATQKITYRISGVGID--QPPFGIFVVDKNTGDINITAIVDREET---  71
             PI ++T+   D    Q I Y ++G GID   P    F +++ TG+I +   +DR++
Sbjct  1570  PIMQVTATDGDKDRPQNIVYFLTGQGIDPDNPANSKFDINRTTGEIFVLKPLDRDQPNGR  1629

Query  72    PSFLITCRALNAQGLDVEKPLILTVKILDINDNPPVFSQQIFMGEIEENSASNSLVMILN  131
             P +  T  A +  G  +     + V + DINDN P+F Q ++ G + EN   +  +VM +
Sbjct  1630  PQWRFTVFAQDEGGEGLVGYADVQVNLKDINDNAPIFPQGVYFGNVTENGTAGMVVMTMT  1689

Query  132   ATDADEPNH-LNSKIAFKI---VSQEPAGTPMFLLSRNTGEVRTLTNSLDREQASSYRLV  187
             A D D+PN   N+K+ + I   V +E  G+P+F +  +TG ++T    LDRE+   Y +
Sbjct  1690  AVDYDDPNEGTNAKLIYSIEKNVIEEETGSPIFEIEADTGVIKTAVCCLDRERTPDYSIQ  1749

Query  188   VSGADKDGEGLSTQCECNIKVKDVNDNFPMFRDSQYSARIEENILSS--ELLRFQVTDLD  245
             V     DG GL       +I+VKD+ND  P F   ++   ++E   ++  E+    VT  D
Sbjct  1750  VVAM--DGGGLKGTGTASIRVKDINDMPPQFTKDEWFTEVDETDGTNLPEMPILTVTVHD  1807

Query  246   EEYTDNWLAVYFFTSGNEGNWFEIQTDPRTNEGI--LKVVKALDYE---QLQSVKLSIAV  300
             E+ T+ +        SG    + F +    R N+G   LK+V+ LDYE   Q     +  I V
Sbjct  1808  EDETNKFQYKVIDNSGYGADKFTMV---RNNDGTGSLKIVQPLDYEDPLQSNGFRFRIQV  1864

Query  301   KNKAEFHQSVISRYRVQSTPVTIQVINVREGIAFRPASKTFTVQKGISSKKLVDYILGTY  360
              +K E + +    +Y V   + V +++ ++ +     +P  +    ++  +       V   L T+
Sbjct  1865  NDKGEDNDN--DKYHVAYSWVVVKLRDINDN---KPQFERPNIEVSVYENADVGKTLETF  1919

Query  361   QAIDEDTNKAASNVKYVMGR-NDGGYLMIDSKTAEIKFVKNMNRDSTFIVNKTITAEVLA  419
             +A D D     S V Y + R +D        ++    +    +N++R+    V       ++LA
Sbjct  1920  KATDPDQG-GKSKVSYAIDRSSDRQRQFSINQEGTVTIQRNLDRE----VTPRHQVKILA  1974

Query  420   IDE-YTGKTSTGTVYYVRVPDFNDNCP  444
             ID+    KT+T T+ V V D NDN P
Sbjct  1975  IDDGIPPKTATATLTVIVQDINDNPP  2000

 Score = 79.3 bits (194),  Expect = 1e-13, Method: Compositional matrix adjust.
 Identities = 104/419 (25%), Positives = 173/419 (41%), Gaps = 51/419 (12%)

Query  50    IFVVDKNTGDINITAIVDREETPSFLITCRAL----------NAQGLDVEKPLILTVKIL  99
              F ++   +G+I    + DRE+  ++ +   A           N Q   V K   + + I
Sbjct  1385  FFEINLQSGEIFTKVVFDREKQGAYALEVEARDGAPSARPNSNQQPNSVTK--FIRIGIA  1442

Query  100   DINDNPPVFSQQIFMGEIEENSASNSLVMILNATDADEPNHLNSKIAFKIVSQEPAGTPM  159
             D NDNPP F + ++  E++EN      V+ + A D DE    +S+I ++I S    G
Sbjct  1443  DKNDNPPYFDKALYEAEVDENEDIQHTVLTVTAKDHDE----SSRIRYEITSGNIGGA--  1496

Query  160   FLLSRNTGEVRTLTNSLDREQASSYRLVVSGADKDGEGLSTQCECNIKVKDVNDNFPMFR  219
             F +   TG +  +  +LD E     Y L +  +D   E  +       I VKDVND  P F
Sbjct  1497  FAVKNMTGAI-YVAGALDYETRKRYHLTLVASDSLNENATI---IEIHVKDVNDMPPTFS  1552

Query  220   DSQYSARI--EENILSSELLRFQVTDLDEEYTDNWLAVYFFTSG--NEGNWFEIQTDPRT  275
                Y A    E+ +    +++    TD D++   N   VYF T    +  N    + D
Sbjct  1553  QQIYEATQDEEQTVTDRPIMQVTATDGDKDRPQN--IVYFLTGQGIDPDNPANSKFDINR  1610

Query  276   NEGILKVVKALDYEQLQS-VKLSIAVKNKAEFHQSVISRYRVQSTPVTIQVINVREGIAF  334
               G + V+K LD +Q      +    V  + E  + ++    VQ     + + ++ +
Sbjct  1611  TTGEIFVLKPLDRDQPNGRPQWRFTVFAQDEGGEGLVGYADVQ-----VNLKDINDNAPI  1665

Query  335   RPASKTFTVQKGISSKKLVDYILGTYQAIDEDTNKAASNVK--YVMGRN------DGGYL  386
              P    F     ++        ++ T  A+D D     +N K  Y + +N
Sbjct  1666  FPQGVYF---GNVTENGTAGMVVMTMTAVDYDDPNEGTNAKLIYSIEKNVIEEETGSPIF  1722

Query  387   MIDSKTAEIK-FVKNMNRDSTFIVNKTITAEVLAIDEYTGKTSTGTVYYVRVPDFNDNCP  444
              I++ T  IK  V  ++R+ T        + +V+A+D   G   TGT  +RV D ND  P
Sbjct  1723  EIEADTGVIKTAVCCLDRERT----PDYSIQVVAMDG-GGLKGTGTASIRVKDINDMPP  1776

 Score = 75.9 bits (185),  Expect = 2e-12, Method: Compositional matrix adjust.
 Identities = 108/429 (25%), Positives = 178/429 (41%), Gaps = 53/429 (12%)

Query  53    VDKNTGDINITAIVDREETPSFLITCRALNAQGLDVEKPLILTVKILDINDNPPVFSQ-Q  111
             +D++  ++N+TA+ D           +C  +N            ++ V I D+NDN PVF
Sbjct  950   LDRDKYELNVTAMDDG--------SC-CVNGDATIHTSTAVVVVFITDVNDNKPVFKDCG  1000
```

```
Query  112   IFMGEIEENSASNSLVMILNATDADEPNHLNSKIAFKIVSQEPAGTPMFLLSRNTGEVRT  171
              +  ++EE + + S V+ + ATD D+   +N ++ + IV Q       F +   TGEV T
Sbjct  1001  TYYPKVEEGAPNGSPVIKVQATDEDK--GVNGQVKYSIVQQPNQKGTKFTVDEETGEVST  1058

Query  172   LTNSLDRE--QASSYRLVVSGADKDGEGLSTQCECNIKVKDVNDNFPMFRDSQYSARIEE  229
                   DRE        + V   D+    L   C   +++ DVNDN P+F   +Y   +++
Sbjct  1059  -NKVFDREGDDGKFVSVTVKATDQGEPSLEGVCSFTVEITDVNDNPPLFDRQKYVENVKQ  1117

Query  230   NI-LSSELLRFQVTDLDEEYTDNWLAVYFFT---SGNEGNWFEIQTDPRTNEGILKVVKA  285
             +  + + +LR  V+  DE+  +N   VY  +   S N+  +FEIQ  P +   +LK  K
Sbjct  1118  DASIGTNILR--VSASDEDADNNGAIVYTLSAPHSPNDLEYFEIQ--PESGWIVLK--KP  1171

Query  286   LD-YEQLQSVKLSIAVK----------NKAEFHQSVISRYRVQSTPV----TIQVINVRE  330
             LD    + SV+    +K          + A+    +    R    P+        I++RE
Sbjct  1172  LDGCYSVSSVRERYRLKVYASDRGDPPSTADVDVELDVVDRNNKPPIWDRNIYGPIHIRE  1231

Query  331   GIAFRPASKTFTVQKGISSKKLVDYILGTYQAIDEDTNKAASNVKYVMGRNDGGYLMIDS  390
              +             GI     V Y L         TNK   +  Y+  R D G
Sbjct  1232  NVTVGTVVTAIKASSGIEGNPTVFYRL--MPGSTAQTNKL--HTFYLQQRPDNG-----D  1282

Query  391   KTAEIKFVKNMNRDSTFIVNKTITAEVLAIDEYTGKTSTGTVYVRVPDFNDNCPTAV-LE  449
               A+IK    ++ ++    N TI  E     +     S  TVY+++ D ND  P     E
Sbjct  1283  TWADIKVNHPLDFETIKEYNLTIRVENNGAQQL---ASEATVYIQLEDVNDEIPLFTERE  1339

Query  450   KDAVCSSSP  458
             ++ V    P
Sbjct  1340  QETVLEGEP  1348

 Score = 61.2 bits (147),  Expect = 6e-08, Method: Compositional matrix adjust.
 Identities = 87/366 (24%), Positives = 156/366 (43%), Gaps = 38/366 (10%)

Query  51    FVVDKNTGDINITAIVDRE-ETPSFL-ITCRALNAQGLDVEKPLILTVKILDINDNPPVF  108
             F VD+ TG+++   + DRE +   F+ +T +A +     +E     TV+I D+NDNPP+F
Sbjct  1047  FTVDEETGEVSTNKVFDREGDDGKFVSVTVKATDQGEPSLEGVCSFTVEITDVNDNPPLF  1106

Query  109   SQQIFMGEIEENSASNSLVMILNATDADEPNHLNSKIAFKIVS-QEPAGTPMFLLSRNTG  167
              +Q ++  ++++++  + ++ ++A+D D  N  N  I + + +   P     F +   +G
Sbjct  1107  DRQKYVENVKQDASIGTNILRVSASDEDADN--NGAIVYTLSAPHSPNDLEYFEIQPESG  1164

Query  168   EVRTLTNSLDREQASS-----YRLVVSGADKDGEGLSTQCECNIKVKDVNDNFPMFRDSQ  222
              +  L   LD   + S     YRL V  +D+ G+  ST            +N P   D
Sbjct  1165  WI-VLKKPLDGCYSVSSVRERYRLKVYASDR-GDPPSTADVDVELDVVDRNNKPPIWDRN  1222

Query  223   YSA--RIEENILSSELLRFQVTDLDEEYTDNWLAVYFFTSG-----NEGNWFEIQTDPRT  275
                   I EN+    ++             N    Y    G     N+ + F +Q  P
Sbjct  1223  IYGPIHIRENVTVGTVV--TAIKASSGIEGNPTVFYRLMPGSTAQTNKLHTFYLQQRPDN  1280

Query  276   NE--GILKVVKALDYEQLQSVKLSIAVKNKAEFHQSVISRYRVQSTPVTIQVINVREGIA  333
              +    +KV   LD+E ++   L+I V+N         ++       V IQ+ +V + I
Sbjct  1281  GDTWADIKVNHPLDFETIKEYNLTIRVENNG-------AQQLASEATVYIQLEDVNDEIP  1333

Query  334   FRPASKTFTVQKGISSKKLVDYILGTYQAIDEDTNKAASNVKYVM---GRNDG-GYLMID  389
                  +  TV +G      V  +     AID+D     + V Y +    RN+G  +  I+
Sbjct  1334  LFTEREQETVLEGEPIGTKVTQV----NAIDKDGTFPNNQVYYYIVDSPRNEGKDFFEIN  1389

Query  390   SKTAEI  395
              ++ EI
Sbjct  1390  LQSGEI  1395

 Score = 55.8 bits (133),  Expect = 3e-06, Method: Compositional matrix adjust.
 Identities = 49/158 (31%), Positives = 78/158 (49%), Gaps = 24/158 (15%)

Query  102   NDNPPVFSQQIFMGEIEENSASNSLVMILNATDADEPNHLNSKIAFKIVSQEPAGTP--M  159
             ND  P FSQQ++   ++EN+  N+LV  + A+D D  N     + F  V     GT
Sbjct  880   NDEEPKFSQQVYTPNVDENAGPNTLVTTVVASDKDGDN-----VRFGFVG---GGTSSGQ  931

Query  160   FLLSRNTGEVRTLTN--SLDREQASSYRLVVSGADK-------DGEGLSTQCECNIKVKD  210
             F++   TG +R  +   SLDR++   Y L V+  D        D    ++     + + D
```

```
Sbjct  932   FVIEEITGVIRLHSKAISLDRDK---YELNVTAMDDGSCCVNGDATIHTSTAVVVVFITD  988

Query  211   VNDNFPMFRD-SQYSARIEENILS-SELLRFQVTDLDE  246
             VNDN P+F+D   Y  ++EE   + S +++ Q TD D+
Sbjct  989   VNDNKPVFKDCGTYYPKVEEGAPNGSPVIKVQATDEDK  1026

 Score = 53.9 bits (128),  Expect = 1e-05, Method: Compositional matrix adjust.
 Identities = 70/306 (23%), Positives = 126/306 (41%), Gaps = 30/306 (10%)

Query  20    PIAKIT-SDYQATQKITYRI---SGVGIDQPPFGIFVVDKNTGDINITAIVDRE---ETP  72
             PI  +T  D   T K  Y++   SG G D+  F +   +  TG + I   +D E   ++
Sbjct  1799  PILTVTVHDEDETNKFQYKVIDNSGYGADK--FTMVRNNDGTGSLKIVQPLDYEDPLQSN  1856

Query  73    SFLITCRALNAQGLDVEKPLI------LTVKILDINDNPPVFSQQIFMGEIEENSASNSL  126
              F    + +N +G D +           + VK+ DINDN P F +       + EN+
Sbjct  1857  GFRFRIQ-VNDKGEDNDNDKYHVAYSWVVVKLRDINDNKPQFERPNIEVSVYENADVGKT  1915

Query  127   VMILNATDADEPNHLNSKIAFKIVSQEPAGTPMFLLSRNTGEVRTLTNSLDREQASSYRL  186
             +    ATD D+      SK+++ I   + +        S N      T+  +LDRE    +++
Sbjct  1916  LETFKATDPDQGG--KSKVSYAI---DRSSDRQRQFSINQEGTVTIQRNLDREVTPRHQV  1970

Query  187   VVSGADKDGEGLSTQCECNIKVKDVNDNFPMFRDSQYSARIEENILSSELLRFQVTDLDE  246
              +    D       +       + V+D+NDN P F    Y    + E++   +++    TD D+
Sbjct  1971  KILAIDDGIPPKTATATLTVIVQDINDNPPKFL-KDYRPVLPEHVPPRKVVEILATDDDD  2029

Query  247   EYTDNWLAVYFFTSGNEGNW----FEIQTDPRTNEG----ILKVVKALDYEQLQSVKLSI  298
                 N     F        +     F+++ D +   G     I+  +++ D EQ +   + I
Sbjct  2030  RSKSNGPPFQFRLDPGADDIIRASFKVEQDQKGANGDGMAIVSSLRSFDREQQKEYLIPI  2089

Query  299   AVKNKA  304
              +K+
Sbjct  2090  VIKDHG  2095

 Score = 51.6 bits (122),  Expect = 5e-05, Method: Compositional matrix adjust.
 Identities = 96/398 (24%), Positives = 163/398 (41%), Gaps = 54/398 (14%)

Query  106   PVFSQQIFMGEIEENSASNSLVMILNATDADEPNHLNSKIAFKIVSQ-EPAGTPMFLLSR  164
             P F    +  EI EN   +S ++ + A       +  + +I + + +Q + AGT  F +
Sbjct  677   PQFYMPSYEAEIPENQKKDSDIISVKAK-----SFADREIRYTLKAQGQGAGT--FNIGP  729

Query  165   NTGEVRTLTNSLDRE---QASSYRLVVSGADKDGEGLSTQCECNIKVKDVNDNFPMFRDS  221
              +G V+ L   LD E   Q   Y LVV+ A +D  G ST  E  I+V DVNDN P F
Sbjct  730   TSGIVK-LAKELDFEDLRQPHVYSLVVT-ATEDSGGFSTSVELTIRVTDVNDNAPKFELP  787

Query  222   QYSAR-IEENI-LSSELLRFQVTDLDEEYTDNWLAVYFFTSGNEGNWFEIQTDPRTNEGI  279
              Y A  ++E+I + + +L+ +  D D          + +  S +    F + ++     GI
Sbjct  788   DYQAHNVDEDIQIGTSILKVKAMDADSGAN---AEIEYLVSDDH---FAVDSN-----GI  836

Query  280   LKVVKALDYEQLQSV-KLSIAVKNKAEFHQSVISRYRVQSTPVTIQVINVREGIAFRPAS  338
             +   K LD +   +  +  +  K+K E  ++  +  RV +             E   F
Sbjct  837   IVNNKQLDADNNNAYYEFVVTAKDKGEPAKTGTATVRVYTKNKN------DEEPKFSQQV  890

Query  339   KTFTVQKGISSKKLVDYILGTYQAIDEDTNKAASNVKY--VMGRNDGGYLMIDSKTAEIK  396
              T  V +       LV  ++ +         +K   NV++  V G    G  +I+  T  I+
Sbjct  891   YTPNVDENAGPNTLVTTVVAS--------DKDGDNVRFGFVGGGTSSGQFVIEEITGVIR  942

Query  397   FVK---NMNRDSTFI-VNKTITAEVLAIDEYTGKTSTGTVYVRVPDFNDN------CPTA  446
                   +++RD   + V               + T  TST  V V + D NDN      C T
Sbjct  943   LHSKAISLDRDKYELNVTAMDDGSCCVNGDATIHTSTAVVVVFITDVNDNKPVFKDCGTY  1002

Query  447   VLE-KDAVCSSSPSVVVSARTLNNRYTGPYTFALEDQP  483
              + ++   + SP + V A   +   G   +++   QP
Sbjct  1003  YPKVEEGAPNGSPVIKVQATDEDKGVNGQVKYSIVQQP  1040

 Score = 44.7 bits (104),  Expect = 0.008, Method: Compositional matrix adjust.
 Identities = 48/199 (24%), Positives = 73/199 (37%), Gaps = 19/199 (10%)

Query  27    DYQATQKITYRISGVGIDQPPFGIFVVDKNTGDINITAIVDREETPSFLITCRALNAQGL  86
```

```
             D     K++Y I      Q  F I       G + I   +DRE TP   +   A++
Sbjct  1925  DQGGKSKVSYAIDRSSDRQRQFSI----NQEGTVTIQRNLDREVTPRHQVKILAIDDGIP  1980

Query  87    DVEKPLILTVKILDINDNPPVFSQQIFMGEIEENSASNSLVMILNATDADEPNHLNSKIA  146
                    LTV + DINDNPP F +  +   + E+     +V IL   D D
Sbjct  1981  PKTATATLTVIVQDINDNPPKFLKD-YRPVLPEHVPPRKVVEILATDDDDRSKSNGPPFQ  2039

Query  147   FKIVSQEPAGTPMFLLSRNTGEVRTLTN-----------SLDREQASSYRLVVSGADKDG  195
             F++   +P    +   S    + +   N           S DREQ   Y + +   D
Sbjct  2040  FRL---DPGADDIIRASFKVEQDQKGANGDGMAIVSSLRSFDREQQKEYLIPIVIKDHGN  2096

Query  196   EGLSTQCECNIKVKDVNDN  214
               ++      + + DVNDN
Sbjct  2097  PAMTGTSTLTVVIGDVNDN  2115

 Score = 40.8 bits (94),  Expect = 0.12, Method: Compositional matrix adjust.
 Identities = 55/242 (23%), Positives = 102/242 (42%), Gaps = 22/242 (9%)

Query  4    KFAKPCREGE--DNSKRNP-IAKITSDYQATQKITYRISGVGIDQPPFGIFVVDKNTGDI  60
            +F  P  E E  +N K++  I  + +   A ++I Y +   G      G F +   +G +
Sbjct  678  QFYMPSYEAEIPENQKKDSDIISVKAKSFADREIRYTLKAQGQGA---GTFNIGPTSGIV  734

Query  61   NITAIVDREE--TPSF--LITCRALNAQGLDVEKPLILTVKILDINDNPPVFSQQIFMGE  116
             +   +D E+   P    L+     ++ G      + LT+++ D+NDN P F    +
Sbjct  735  KLAKELDFEDLRQPHVYSLVVTATEDSGGFSTS--VELTIRVTDVNDNAPKFELPDYQAH  792

Query  117  -IEENSASNSLVMILNATDADEPNHLNSKIAFKIVSQEPAGTPMFLLSRNTGEVRTLTNS  175
             ++E+    + ++ + A DAD  +  N++I + +VS +      F +  N   V
Sbjct  793  NVDEDIQIGTSILKVKAMDAD--SGANAEIEY-LVSDD-----HFAVDSNGIIVNNKQLD  844

Query  176  LDREQASSYRLVVSGADKDGEGLSTQCECNIKVKDVNDNFPMFRDSQYSARIEENILSSE  235
             D   A  Y  VV+  DK      +      +  K+ ND   P F    Y+  ++EN   +
Sbjct  845  ADNNNA-YYEFVVTAKDKGEPAKTGTATVRVYTKNKNDEEPKFSQQVYTPNVDENAGPNT  903

Query  236  LL  237
            L+
Sbjct  904  LV  905

 Score = 39.3 bits (90),  Expect = 0.32, Method: Compositional matrix adjust.
 Identities = 55/225 (24%), Positives = 88/225 (39%), Gaps = 24/225 (11%)

Query  75   LITCRALNAQGLDVEKPLILTVKILDINDNPPVFSQQIF--MGEIEENSASNSLVMILNA  132
            +IT +  NA GL       + + + D+ND PP F  +       ++ N+  N+ V  L A
Sbjct  539  IITNQGHNA-GLKYTDNQRVIIHVKDVNDEPPYFINRPLPMQAVVQLNAPPNTPVFTLQA  597

Query  133  TDADEPNHLNSKIAFKIVSQEPAGTPMFLLSRNTGEVRTLTNSLDREQASSYRLVVSGAD  192
             D D     +  I + IV     G   F +   +G VRT    L  +    Y L V   D
Sbjct  598  RDPD----TDHNIHYFIVRDRTGG--RFEVDERSGVVRTRGTDL-FQLDMEYVLYVKAED  650

Query  193  KDGE----GLSTQCECNIKVKDVNDNFPMFRDSQYSARIEEN-ILSSELLRFQVTDLDEE  247
            ++G+       +  E  + +       P F    Y A I EN    S+++  +     +
Sbjct  651  QNGKVDDRRFQSTPEERLSIVG-GKRAPQFYMPSYEAEIPENQKKDSDIISVKA----KS  705

Query  248  YTDNWLAVYFFTSGNEGNWFEIQTDPRTNEGILKVVKALDYEQLQ  292
            + D  +       G     F I         GI+K+ K LD+E L+
Sbjct  706  FADREIRYTLKAQGQGAGTFNIG----PTSGIVKLAKELDFEDLR  746

>AALB20_033171.P66523 | transcript=AALB20_033171.R66523 | gene=AALB20_033171
| organism=Anopheles_albimanus_STECLA_2020 |
gene_product=neural-cadherin, transcript variant X38 |
transcript_product=neural-cadherin,
transcript variant X28 | location=CM024292:75037560-75208393(+)
| protein_length=3126 | sequence_SO=chromosome
| SO=protein_coding_gene | is_pseudo=false
Length=3126

 Score = 147 bits (371),  Expect = 2e-35, Method: Compositional matrix adjust.
 Identities = 128/446 (29%), Positives = 217/446 (49%), Gaps = 36/446 (8%)
```

```
Query  20    PIAKITS---DYQATQKITYRISGVGID--QPPFGIFVVDKNTGDINITAIVDREET---  71
             PI ++T+   D    Q I Y ++G GID   P    F +++ TG+I +    +DR++
Sbjct  1566  PIMQVTATDGDKDRPQNIVYFLTGQGIDPDNPANSKFDINRTTGEIFVLKPLDRDQPNGR  1625

Query  72    PSFLITCRALNAQGLDVEKPLILTVKILDINDNPPVFSQQIFMGEIEENSASNSLVMILN  131
             P +  T  A +  G  +      + V + DINDN P+F Q ++ G + EN  +  +VM +
Sbjct  1626  PQWRFTVFAQDEGGEGLVGYADVQVNLKDINDNAPIFPQGVYFGNVTENGTAGMVVMTMT  1685

Query  132   ATDADEPNH-LNSKIAFKI---VSQEPAGTPMFLLSRNTGEVRTLTNSLDREQASSYRLV  187
             A D D+PN   N+K+ + I   V +E  G+P+F +  +TG ++T    LDRE+   Y +
Sbjct  1686  AVDYDDPNEGTNAKLIYSIEKNVIEEETGSPIFEIEADTGVIKTAVCCLDRERTPDYSIQ  1745

Query  188   VSGADKDGEGLSTQCECNIKVKDVNDNFPMFRDSQYSARIEENILSS--ELLRFQVTDLD  245
             V     DG GL       +I+VKD+ND  P F   ++   ++E   ++  E+    VT  D
Sbjct  1746  VVAM--DGGGLKGTGTASIRVKDINDMPPQFTKDEWFTEVDETDGTNLPEMPILTVTVHD  1803

Query  246   EEYTDNWLAVYFFTSGNEGNWFEIQTDPRTNEGI--LKVVKALDYE---QLQSVKLSIAV  300
             E+ T+ +        SG    + F +    R N+G   LK+V+ LDYE   Q    +  I V
Sbjct  1804  EDETNKFQYKVIDNSGYGADKFTMV---RNNDGTGSLKIVQPLDYEDPLQSNGFRFRIQV  1860

Query  301   KNKAEFHQSVISRYRVQSTPVTIQVINVREGIAFRPASKTFTVQKGISSKKLVDYILGTY  360
              +K E + +   +Y V  + V +++ ++ +    +P  +   ++  +      V   L T+
Sbjct  1861  NDKGEDNDN--DKYHVAYSWVVVKLRDINDN---KPQFERPNIEVSVYENADVGKTLETF  1915

Query  361   QAIDEDTNKAASNVKYVMGR-NDGGYLMIDSKTAEIKFVKNMNRDSTFIVNKTITAEVLA  419
             +A D D     S V Y + R +D        ++   +   +N++R+    V       ++LA
Sbjct  1916  KATDPDQG-GKSKVSYAIDRSSDRQRQFSINQEGTVTIQRNLDRE----VTPRHQVKILA  1970

Query  420   IDE-YTGKTSTGTVYVRVPDFNDNCP  444
             ID+    KT+T T+ V V D NDN P
Sbjct  1971  IDDGIPPKTATATLTVIVQDINDNPP  1996

 Score = 79.3 bits (194),  Expect = 1e-13, Method: Compositional matrix adjust.
 Identities = 104/419 (25%), Positives = 173/419 (41%), Gaps = 51/419 (12%)

Query  50    IFVVDKNTGDINITAIVDREETPSFLITCRAL----------NAQGLDVEKPLILTVKIL  99
              F ++  +G+I    + DRE+  ++ +   A            N Q   V K   + + I
Sbjct  1381  FFEINLQSGEIFTKVVFDREKQGAYALEVEARDGAPSARPNSNQQPNSVTK--FIRIGIA  1438

Query  100   DINDNPPVFSQQIFMGEIEENSASNSLVMILNATDADEPNHLNSKIAFKIVSQEPAGTPM  159
             D NDNPP F + ++  E++EN       V+ + A D DE    +S+I ++I S    G
Sbjct  1439  DKNDNPPYFDKALYEAEVDENEDIQHTVLTVTAKDHDE----SSRIRYEITSGNIGGA--  1492

Query  160   FLLSRNTGEVRTLTNSLDREQASSYRLVVSGADKDGEGLSTQCECNIKVKDVNDNFPMFR  219
             F +   TG +  +  +LD E    Y L +  +D   E  +       I VKDVND  P F
Sbjct  1493  FAVKNMTGAI-YVAGALDYETRKRYHLTLVASDSLNENATI---IEIHVKDVNDMPPTFS  1548

Query  220   DSQYSARI--EENILSSELLRFQVTDLDEEYTDNWLAVYFFTSG--NEGNWFEIQTDPRT  275
                Y A    E+ +    +++   TD D++   N   VYF T    +  N    + D
Sbjct  1549  QQIYEATQDEEQTVTDRPIMQVTATDGDKDRPQN--IVYFLTGQGIDPDNPANSKFDINR  1606

Query  276   NEGILKVVKALDYEQLQS-VKLSIAVKNKAEFHQSVISRYRVQSTPVTIQVINVREGIAF  334
               G + V+K LD +Q     +     V  + E  + ++   VQ      + + ++ +
Sbjct  1607  TTGEIFVLKPLDRDQPNGRPQWRFTVFAQDEGGEGLVGYADVQ-----VNLKDINDNAPI  1661

Query  335   RPASKTFTVQKGISSKKLVDYILGTYQAIDEDTNKAASNVK--YVMGRN------DGGYL  386
              P    F      ++        ++ T  A+D D      +N K  Y + +N
Sbjct  1662  FPQGVYF---GNVTENGTAGMVVMTMTAVDYDDPNEGTNAKLIYSIEKNVIEEETGSPIF  1718

Query  387   MIDSKTAEIK-FVKNMNRDSTFIVNKTITAEVLAIDEYTGKTSTGTVYVRVPDFNDNCP  444
              I++ T  IK  V  ++R+ T         + +V+A+D   G  TGT  +RV D ND  P
Sbjct  1719  EIEADTGVIKTAVCCLDRERT----PDYSIQVVAMDG-GGLKGTGTASIRVKDINDMPP  1772

 Score = 77.8 bits (190),  Expect = 5e-13, Method: Compositional matrix adjust.
 Identities = 107/426 (25%), Positives = 179/426 (42%), Gaps = 51/426 (12%)

Query  53    VDKNTGDINITAIVDREETPSFLITCRALNAQGLDVEKPLILTVKILDINDNPPVFSQ-Q  111
             +D++  ++N+TA+ D          +C  +N           ++ V I D+NDN PVF
```

```
Sbjct  950   LDRDKYELNVTAMDDG--------SC-CVNGDATIHTSTAVVVVFITDVNDNKPVFKDCG  1000

Query  112   IFMGEIEENSASNSLVMILNATDADEPNHLNSKIAFKIVSQEPAGTPMFLLSRNTGEVRT  171
              +  ++EE + + S V+ + ATD D+   +N ++ + IV Q       F +   TGEV T
Sbjct  1001  TYYPKVEEGAPNGSPVIKVQATDEDK--GVNGQVKYSIVQQPNQKGTKFTVDEETGEVST  1058

Query  172   LTNSLDRE--QASSYRLVVSGADKDGEGLSTQCECNIKVKDVNDNFPMFRDSQYSARIEE  229
                   DRE        + V   D+    L   C   +++ DVNDN P+F   +Y   +++
Sbjct  1059  -NKVFDREGDDGKFVSVTVKATDQGEPSLEGVCSFTVEITDVNDNPPLFDRQKYVENVKQ  1117

Query  230   NI-LSSELLRFQVTDLDEEYTDNWLAVYFFT---SGNEGNWFEIQTDPRTNEGILKVVKA  285
             +  + + +LR  V+  DE+  +N   VY  +   S N+  +FEIQ  P +   +LK  K
Sbjct  1118  DASIGTNILR--VSASDEDADNNGAIVYTLSAPHSPNDLEYFEIQ--PESGWIVLK--KP  1171

Query  286   LD--YEQLQSVKLSIAVKNK------AEFHQSVISRYRVQSTPV----TIQVINVREGIA  333
             LD  Y + +  +L +   ++      A+    +    R    P+        I++RE +
Sbjct  1172  LDGCYSR-ERYRLKVYASDRGDPPSTADVDVELDVVDRNNKPPIWDRNIYGPIHIRENVT  1230

Query  334   FRPASKTFTVQKGISSKKLVDYILGTYQAIDEDTNKAASNVKYVMGRNDGGYLMIDSKTA  393
                         GI     V Y L         TNK   +  Y+  R D G        A
Sbjct  1231  VGTVVTAIKASSGIEGNPTVFYRL--MPGSTAQTNKL--HTFYLQQRPDNG-----DTWA  1281

Query  394   EIKFVKNMNRDSTFIVNKTITAEVLAIDEYTGKTSTGTVYYRVPDFNDNCPTAV-LEKDA  452
             +IK    ++ ++    N TI  E     +     S  TVY+++ D ND  P     E++
Sbjct  1282  DIKVNHPLDFETIKEYNLTIRVENNGAQQL---ASEATVYIQLEDVNDEIPLFTEREQET  1338

Query  453   VCSSSP  458
             V    P
Sbjct  1339  VLEGEP  1344

 Score = 63.5 bits (153),  Expect = 1e-08, Method: Compositional matrix adjust.
 Identities = 86/364 (24%), Positives = 156/364 (43%), Gaps = 38/364 (10%)

Query  51    FVVDKNTGDINITAIVDRE-ETPSFL-ITCRALNAQGLDVEKPLILTVKILDINDNPPVF  108
             F VD+ TG+++   + DRE +   F+ +T +A +     +E     TV+I D+NDNPP+F
Sbjct  1047  FTVDEETGEVSTNKVFDREGDDGKFVSVTVKATDQGEPSLEGVCSFTVEITDVNDNPPLF  1106

Query  109   SQQIFMGEIEENSASNSLVMILNATDADEPNHLNSKIAFKIVS-QEPAGTPMFLLSRNTG  167
              +Q ++  ++++++  + ++ ++A+D D  N  N  I + + +   P     F +   +G
Sbjct  1107  DRQKYVENVKQDASIGTNILRVSASDEDADN--NGAIVYTLSAPHSPNDLEYFEIQPESG  1164

Query  168   EV---RTLTNSLDREQASSYRLVVSGADKDGEGLSTQCECNIKVKDVNDNFPMFRDSQYS  224
              +   + L     RE+   YRL V  +D+ G+  ST            +N P   D
Sbjct  1165  WIVLKKPLDGCYSRER---YRLKVYASDR-GDPPSTADVDVELDVVDRNNKPPIWDRNIY  1220

Query  225   A--RIEENILSSELLRFQVTDLDEEYTDNWLAVYFFTSG-----NEGNWFEIQTDPRTNE  277
                 I EN+    ++             N    Y    G     N+ + F +Q  P   +
Sbjct  1221  GPIHIRENVTVGTVV--TAIKASSGIEGNPTVFYRLMPGSTAQTNKLHTFYLQQRPDNGD  1278

Query  278   --GILKVVKALDYEQLQSVKLSIAVKNKAEFHQSVISRYRVQSTPVTIQVINVREGIAFR  335
                 +KV   LD+E ++   L+I V+N         ++       V IQ+ +V + I
Sbjct  1279  TWADIKVNHPLDFETIKEYNLTIRVENNG-------AQQLASEATVYIQLEDVNDEIPLF  1331

Query  336   PASKTFTVQKGISSKKLVDYILGTYQAIDEDTNKAASNVKYVM---GRNDG-GYLMIDSK  391
                +  TV +G      V  +     AID+D     + V Y +    RN+G  +  I+ +
Sbjct  1332  TEREQETVLEGEPIGTKVTQV----NAIDKDGTFPNNQVYYYIVDSPRNEGKDFFEINLQ  1387

Query  392   TAEI  395
             + EI
Sbjct  1388  SGEI  1391

 Score = 55.8 bits (133),  Expect = 3e-06, Method: Compositional matrix adjust.
 Identities = 49/158 (31%), Positives = 78/158 (49%), Gaps = 24/158 (15%)

Query  102   NDNPPVFSQQIFMGEIEENSASNSLVMILNATDADEPNHLNSKIAFKIVSQEPAGTP--M  159
             ND  P FSQQ++   ++EN+  N+LV  + A+D D  N     + F  V     GT
Sbjct  880   NDEEPKFSQQVYTPNVDENAGPNTLVTTVVASDKDGDN-----VRFGFVG---GGTSSGQ  931

Query  160   FLLSRNTGEVRTLTN--SLDREQASSYRLVVSGADK-------DGEGLSTQCECNIKVKD  210
```

```
                F++   TG +R  +   SLDR++   Y L V+  D        D    ++     + + D
Sbjct  932   FVIEEITGVIRLHSKAISLDRDK---YELNVTAMDDGSCCVNGDATIHTSTAVVVVFITD  988

Query  211   VNDNFPMFRD-SQYSARIEENILS-SELLRFQVTDLDE  246
             VNDN P+F+D   Y  ++EE   + S +++ Q TD D+
Sbjct  989   VNDNKPVFKDCGTYYPKVEEGAPNGSPVIKVQATDEDK  1026

 Score = 53.9 bits (128),  Expect = 1e-05, Method: Compositional matrix adjust.
 Identities = 70/306 (23%), Positives = 126/306 (41%), Gaps = 30/306 (10%)

Query  20    PIAKIT-SDYQATQKITYRI---SGVGIDQPPFGIFVVDKNTGDINITAIVDRE---ETP  72
             PI  +T  D   T K  Y++   SG G D+  F +   +  TG + I   +D E   ++
Sbjct  1795  PILTVTVHDEDETNKFQYKVIDNSGYGADK--FTMVRNNDGTGSLKIVQPLDYEDPLQSN  1852

Query  73    SFLITCRALNAQGLDVEKPLI------LTVKILDINDNPPVFSQQIFMGEIEENSASNSL  126
              F    + +N +G D +           + VK+ DINDN P F +       + EN+
Sbjct  1853  GFRFRIQ-VNDKGEDNDNDKYHVAYSWVVVKLRDINDNKPQFERPNIEVSVYENADVGKT  1911

Query  127   VMILNATDADEPNHLNSKIAFKIVSQEPAGTPMFLLSRNTGEVRTLTNSLDREQASSYRL  186
             +    ATD D+     SK+++ I   + +       S N     T+  +LDRE    +++
Sbjct  1912  LETFKATDPDQGG--KSKVSYAI---DRSSDRQRQFSINQEGTVTIQRNLDREVTPRHQV  1966

Query  187   VVSGADKDGEGLSTQCECNIKVKDVNDNFPMFRDSQYSARIEENILSSELLRFQVTDLDE  246
              +   D      +      + V+D+NDN P F    Y   + E++   +++    TD D+
Sbjct  1967  KILAIDDGIPPKTATATLTVIVQDINDNPPKFL-KDYRPVLPEHVPPRKVVEILATDDDD  2025

Query  247   EYTDNWLAVYFFTSGNEGNW----FEIQTDPRTNEG----ILKVVKALDYEQLQSVKLSI  298
                 N     F       +     F+++ D +   G    I+  +++ D EQ +   + I
Sbjct  2026  RSKSNGPPFFQFRLDPGADDIIRASFKVEQDQKGANGDGMAIVSSLRSFDREQQKEYLIPI  2085

Query  299   AVKNKA  304
              +K+
Sbjct  2086  VIKDHG  2091

 Score = 51.6 bits (122),  Expect = 5e-05, Method: Compositional matrix adjust.
 Identities = 96/398 (24%), Positives = 163/398 (41%), Gaps = 54/398 (14%)

Query  106   PVFSQQIFMGEIEENSASNSLVMILNATDADEPNHLNSKIAFKIVSQ-EPAGTPMFLLSR  164
             P F    +  EI EN   +S ++ + A       +  + +I + + +Q + AGT  F +
Sbjct  677   PQFYMPSYEAEIPENQKKDSDIISVKAK-----SFADREIRYTLKAQGQGAGT--FNIGP  729

Query  165   NTGEVRTLTNSLDRE---QASSYRLVVSGADKDGEGLSTQCECNIKVKDVNDNFPMFRDS  221
              +G V+ L   LD E   Q   Y LVV+ A +D  G ST  E  I+V DVNDN P F
Sbjct  730   TSGIVK-LAKELDFEDLRQPHVYSLVVT-ATEDSGGFSTSVELTIRVTDVNDNAPKFELP  787

Query  222   QYSAR-IEENI-LSSELLRFQVTDLDEEYTDNWLAVYFFTSGNEGNWFEIQTDPRTNEGI  279
              Y A  ++E+I + + +L+ +  D D          + +  S +    F + ++     GI
Sbjct  788   DYQAHNVDEDIQIGTSILKVKAMDADSGAN---AEIEYLVSDDH---FAVDSN-----GI  836

Query  280   LKVVKALDYEQLQSV-KLSIAVKNKAEFHQSVISRYRVQSTPVTIQVINVREGIAFRPAS  338
             +   K LD +   +  +  K+K E  ++  +  RV +            E   F
Sbjct  837   IVNNKQLDADNNNAYYEFVVTAKDKGEPAKTGTATVRVYTKNKN------DEEPKFSQQV  890

Query  339   KTFTVQKGISSKKLVDYILGTYQAIDEDTNKAASNVKY--VMGRNDGGYLMIDSKTAEIK  396
              T  V +       LV  ++ +         +K   NV++  V G    G  +I+  T  I+
Sbjct  891   YTPNVDENAGPNTLVTTVVAS--------DKDGDNVRFGFVGGGTSSGQFVIEEITGVIR  942

Query  397   FVK---NMNRDSTFI-VNKTITAEVLAIDEYTGKTSTGTVYVRVPDFNDN------CPTA  446
                   +++RD   + V              + T  TST  V V + D NDN      C T
Sbjct  943   LHSKAISLDRDKYELNVTAMDDGSCCVNGDATIHTSTAVVVVFITDVNDNKPVFKDCGTY  1002

Query  447   VLE-KDAVCSSSPSVVVSARTLNNRYTGPYTFALEDQP  483
              + ++   + SP + V A   +    G   +++   QP
Sbjct  1003  YPKVEEGAPNGSPVIKVQATDEDKGVNGQVKYSIVQQP  1040

 Score = 44.7 bits (104),  Expect = 0.008, Method: Compositional matrix adjust.
 Identities = 48/199 (24%), Positives = 73/199 (37%), Gaps = 19/199 (10%)
```

```
Query  27    DYQATQKITYRISGVGIDQPPFGIFVVDKNTGDINITAIVDREETPSFLITCRALNAQGL  86
             D     K++Y I      Q  F I       G + I   +DRE TP   +   A++
Sbjct  1921  DQGGKSKVSYAIDRSSDRQRQFSI----NQEGTVTIQRNLDREVTPRHQVKILAIDDGIP  1976

Query  87    DVEKPLILTVKILDINDNPPVFSQQIFMGEIEENSASNSLVMILNATDADEPNHLNSKIA  146
                    LTV + DINDNPP F +  +   + E+     +V IL   D D
Sbjct  1977  PKTATATLTVIVQDINDNPPKFLKD-YRPVLPEHVPPRKVVEILATDDDDRSKSNGPPFQ  2035

Query  147   FKIVSQEPAGTPMFLLSRNTGEVRTLTN-----------SLDREQASSYRLVVSGADKDG  195
             F++   +P    +   S    + +   N           S DREQ   Y + +   D
Sbjct  2036  FRL---DPGADDIIRASFKVEQDQKGANGDGMAIVSSLRSFDREQQKEYLIPIVIKDHGN  2092

Query  196   EGLSTQCECNIKVKDVNDN  214
               ++      + + DVNDN
Sbjct  2093  PAMTGTSTLTVVIGDVNDN  2111

 Score = 40.8 bits (94),  Expect = 0.11, Method: Compositional matrix adjust.
 Identities = 55/242 (23%), Positives = 102/242 (42%), Gaps = 22/242 (9%)

Query  4    KFAKPCREGE--DNSKRNP-IAKITSDYQATQKITYRISGVGIDQPPFGIFVVDKNTGDI  60
            +F  P  E E  +N K++  I  + +   A ++I Y +   G      G F +   +G +
Sbjct  678  QFYMPSYEAEIPENQKKDSDIISVKAKSFADREIRYTLKAQGQGA---GTFNIGPTSGIV  734

Query  61   NITAIVDREE--TPSF--LITCRALNAQGLDVEKPLILTVKILDINDNPPVFSQQIFMGE  116
             +   +D E+   P    L+     ++ G      + LT+++ D+NDN P F    +
Sbjct  735  KLAKELDFEDLRQPHVYSLVVTATEDSGGFSTS--VELTIRVTDVNDNAPKFELPDYQAH  792

Query  117  -IEENSASNSLVMILNATDADEPNHLNSKIAFKIVSQEPAGTPMFLLSRNTGEVRTLTNS  175
             ++E+    + ++ + A DAD  +  N++I + +VS +      F +  N   V
Sbjct  793  NVDEDIQIGTSILKVKAMDAD--SGANAEIEY-LVSDD-----HFAVDSNGIIVNNKQLD  844

Query  176  LDREQASSYRLVVSGADKDGEGLSTQCECNIKVKDVNDNFPMFRDSQYSARIEENILSSE  235
             D   A  Y  VV+  DK      +      +  K+ ND  P F    Y+  ++EN   +
Sbjct  845  ADNNNA-YYEFVVTAKDKGEPAKTGTATVRVYTKNKNDEEPKFSQQVYTPNVDENAGPNT  903

Query  236  LL  237
            L+
Sbjct  904  LV  905

 Score = 39.3 bits (90),  Expect = 0.30, Method: Compositional matrix adjust.
 Identities = 55/225 (24%), Positives = 88/225 (39%), Gaps = 24/225 (11%)

Query  75   LITCRALNAQGLDVEKPLILTVKILDINDNPPVFSQQIF--MGEIEENSASNSLVMILNA  132
            +IT +  NA GL       + + + D+ND PP F  +       ++ N+  N+ V  L A
Sbjct  539  IITNQGHNA-GLKYTDNQRVIIHVKDVNDEPPYFINRPLPMQAVVQLNAPPNTPVFTLQA  597

Query  133  TDADEPNHLNSKIAFKIVSQEPAGTPMFLLSRNTGEVRTLTNSLDREQASSYRLVVSGAD  192
             D D     +  I + IV     G   F +   +G VRT    L  +    Y L V   D
Sbjct  598  RDPD----TDHNIHYFIVRDRTGG--RFEVDERSGVVRTRGTDL-FQLDMEYVLYVKAED  650

Query  193  KDGE----GLSTQCECNIKVKDVNDNFPMFRDSQYSARIEEN-ILSSELLRFQVTDLDEE  247
            ++G+       +  E  + +       P F    Y A I EN    S+++  +     +
Sbjct  651  QNGKVDDRRFQSTPEERLSIVG-GKRAPQFYMPSYEAEIPENQKKDSDIISVKA----KS  705

Query  248  YTDNWLAVYFFTSGNEGNWFEIQTDPRTNEGILKVVKALDYEQLQ  292
            + D  +       G     F I        GI+K+ K LD+E L+
Sbjct  706  FADREIRYTLKAQGQGAGTFNIG----PTSGIVKLAKELDFEDLR  746

>ACMO_007424.P13504 | transcript=ACMO_007424.R13504 | gene=ACMO_007424
| organism=Anopheles_coluzzii_MOPTI | gene_product=neural-cadherin,
transcript variant X4 | transcript_product=neural-cadherin,
transcript variant X16 | location=CM029349:40739311-40945968(+)
| protein_length=3098 | sequence_SO=chromosome
| SO=protein_coding_gene | is_pseudo=false
Length=3098

 Score = 147 bits (371),  Expect = 2e-35, Method: Compositional matrix adjust.
 Identities = 128/446 (29%), Positives = 217/446 (49%), Gaps = 36/446 (8%)
```

```
Query  20    PIAKITS---DYQATQKITYRISGVGID--QPPFGIFVVDKNTGDINITAIVDREET---  71
             PI ++T+   D    Q I Y ++G GID   P    F +++ TG+I +    +DR++
Sbjct  1539  PIMQVTATDGDKDRPQNIVYFLTGQGIDPDNPANSKFDINRTTGEIFVLKPLDRDQPNGR  1598

Query  72    PSFLITCRALNAQGLDVEKPLILTVKILDINDNPPVFSQQIFMGEIEENSASNSLVMILN  131
             P +  T  A +  G  +      + V + DINDN P+F Q ++ G + EN  +  +VM +
Sbjct  1599  PQWRFTVFAQDEGGEGLVGYADVQVNLKDINDNAPIFPQGVYFGNVTENGTAGMVVMTMT  1658

Query  132   ATDADEPNH-LNSKIAFKI---VSQEPAGTPMFLLSRNTGEVRTLTNSLDREQASSYRLV  187
             A D D+PN   N+K+ + I   V +E  G+P+F +  +TG ++T    LDRE+   Y +
Sbjct  1659  AVDYDDPNEGTNAKLIYSIEKNVIEEETGSPIFEIEADTGVIKTAVCCLDRERTPDYSIQ  1718

Query  188   VSGADKDGEGLSTQCECNIKVKDVNDNFPMFRDSQYSARIEENILSS--ELLRFQVTDLD  245
             V     DG GL       +I+VKD+ND  P F   ++   ++E   ++  E+    VT  D
Sbjct  1719  VVAM--DGGGLKGTGTASIRVKDINDMPPQFTKDEWFTEVDETDGTNLPEMPILTVTVHD  1776

Query  246   EEYTDNWLAVYFFTSGNEGNWFEIQTDPRTNEGI--LKVVKALDYE---QLQSVKLSIAV  300
             E+ T+ +        SG    + F +    R N+G   LK+V+ LDYE   Q    +  I V
Sbjct  1777  EDETNKFQYKVIDNSGYGADKFTMV---RNNDGTGSLKIVQPLDYEDPLQSNGFRFRIQV  1833

Query  301   KNKAEFHQSVISRYRVQSTPVTIQVINVREGIAFRPASKTFTVQKGISSKKLVDYILGTY  360
              +K E + +   +Y V  + V +++ ++ +    +P  +   ++  +     V   L T+
Sbjct  1834  NDKGEDNDN--DKYHVAYSWVVVKLRDINDN---KPQFERPNIEVSVYENADVGKTLETF  1888

Query  361   QAIDEDTNKAASNVKYVMGR-NDGGYLMIDSKTAEIKFVKNMNRDSTFIVNKTITAEVLA  419
             +A D D     S V Y + R +D        ++   +   +N++R+    V       ++LA
Sbjct  1889  KATDPDQG-GKSKVSYAIDRSSDRQRQFSINQEGTVTIQRNLDRE----VTPRHQVKILA  1943

Query  420   IDE-YTGKTSTGTVYYVRVPDFNDNCP  444
             ID+    KT+T T+ V V D NDN P
Sbjct  1944  IDDGIPPKTATATLTVIVQDINDNPP  1969

 Score = 79.0 bits (193),  Expect = 2e-13, Method: Compositional matrix adjust.
 Identities = 106/424 (25%), Positives = 180/424 (42%), Gaps = 51/424 (12%)

Query  53    VDKNTGDINITAIVDREETPSFLITCRALNAQGLDVEKPLILTVKILDINDNPPVFSQ-Q  111
             +D++  ++N+TA+ D          +C  +N           ++ V I D+NDN PVF
Sbjct  928   LDRDKYELNVTAMDDG--------SC-CVNGDATIHTSTAVVVVFITDVNDNKPVFKDCG  978

Query  112   IFMGEIEENSASNSLVMILNATDADEPNHLNSKIAFKIVSQEPAGTPMFLLSRNTGEVRT  171
              +  ++EE + + S V+ + ATD D+   +N ++ + IV Q       F +   TGEV T
Sbjct  979   TYYPKVEEGAPNGSPVIKVQATDEDK--GVNGQVKYSIVQQPNQKGTKFTVDEETGEVST  1036

Query  172   LTNSLDRE--QASSYRLVVSGADKDGEGLSTQCECNIKVKDVNDNFPMFRDSQYSARIEE  229
                 DRE         + V   D+    L   C   +++ DVNDN P+F   +Y   +++
Sbjct  1037  -NKVFDREGDDGKFVSVTVKATDQGEPSLEGVCSFTVEITDVNDNPPLFDRQKYVENVKQ  1095

Query  230   NI-LSSELLRFQVTDLDEEYTDNWLAVYFFT---SGNEGNWFEIQTDPRTNEGILKVVKA  285
             +  + + +LR  V+  DE+  +N   VY  +   S N+  +FEIQ  P +   +LK  K
Sbjct  1096  DASIGTNILR--VSASDEDADNNGAIVYTLSAPHSPNDLEYFEIQ--PESGWIVLK--KP  1149

Query  286   LDYEQLQSVKLSIAVKNK------AEFHQSVISRYRVQSTPVTIQV----INVREGIAFR  335
             LD E+    +L +   ++      A+     +    R    P+  +     I++RE +
Sbjct  1150  LDRERY---RLKVYASDRGDPPSTADVDVELDVVDRNNKPPIWDRTIYGPIHIRENVTVG  1206

Query  336   PASKTFTVQKGISSKKLVDYILGTYQAIDEDTNKAASNVKYVMGRNDGGYLMIDSKTAEI  395
                +      GI      V Y L          TNK   +  Y+  R D G         A+I
Sbjct  1207  TVVTSVKASSGIEGNPTVFYRL--MPGSTAQTNKL--HTFYLQQRPDNG-----DTWADI  1257

Query  396   KFVKNMNRDSTFIVNKTITAEVLAIDEYTGKTSTGTVYYVRVPDFNDNCPTAV-LEKDAVC  454
             K    ++ ++    N T+  E      +     S  TVY+++ D ND  P      E++ V
Sbjct  1258  KVNHPLDFETIKEYNLTVRVENNGAQQL---ASEATVYIQLEDVNDEIPLFTEREQETVL  1314

Query  455   SSSP  458
                P
Sbjct  1315  EGEP  1318

 Score = 76.3 bits (186),  Expect = 1e-12, Method: Compositional matrix adjust.
```

```
 Identities = 103/418 (25%), Positives = 174/418 (42%), Gaps = 50/418 (12%)

Query  50    IFVVDKNTGDINITAIVDREETPSFLITCRAL----------NAQGLDVEKPLILTVKIL  99
              F ++  +G+I    + DRE+  ++ +   A           N Q   V K   + + I
Sbjct  1355  FFEINLQSGEIFTKVVFDREKQGAYALEVEARDGAPSARPNSNQQPNSVTK--FIRIGIA  1412

Query  100   DINDNPPVFSQQIFMGEIEENSASNSLVMILNATDADEPNHLNSKIAFKIVSQEPAGTPM  159
             D NDNPP F + ++  E++EN      V+ + A D DE    +S+I ++I S    G
Sbjct  1413  DKNDNPPYFDKALYEAEVDENEDIQHTVLTVTAKDHDE----SSRIRYEITSGNIGGA--  1466

Query  160   FLLSRNTGEVRTLTNSLDREQASSYRLVVSGADKDGEGLSTQCECNIKVKDVNDNFPMFR  219
             F +   TG +  +  +LD E    Y L +  +D   E  +      I VKDVND  P F
Sbjct  1467  FAVKNMTGAI-YVAGALDYETRKRYYLTLVASDSLNENETIIV---INVKDVNDMPPSFP  1522

Query  220   DSQYSARIEENI-LSSELLRFQVTDLDEEYTDNWLAVYFFTSG--NEGNWFEIQTDPRTN  276
                Y   ++E + +   +++   TD D++   N   VYF T    +  N    + D
Sbjct  1523  QQVYERTMDEELAVPFPIMQVTATDGDKDRPQN--IVYFLTGQGIDPDNPANSKFDINRT  1580

Query  277   EGILKVVKALDYEQLQS-VKLSIAVKNKAEFHQSVISRYRVQSTPVTIQVINVREGIAFR  335
              G + V+K LD +Q     +    V  + E  + ++    VQ     + + ++ +
Sbjct  1581  TGEIFVLKPLDRDQPNGRPQWRFTVFAQDEGGEGLVGYADVQ-----VNLKDINDNAPIF  1635

Query  336   PASKTFTVQKGISSKKLVDYILGTYQAIDEDTNKAASNVK--YVMGRN------DGGYLM  387
             P    F     ++       ++ T  A+D D     +N K  Y + +N
Sbjct  1636  PQGVYF---GNVTENGTAGMVVMTMTAVDYDDPNEGTNAKLIYSIEKNVIEEETGSPIFE  1692

Query  388   IDSKTAEIK-FVKNMNRDSTFIVNKTITAEVLAIDEYTGKTSTGTVYVRVPDFNDNCP  444
             I++ T  IK  V  ++R+ T       + +V+A+D   G   TGT  +RV D ND  P
Sbjct  1693  IEADTGVIKTAVCCLDRERT----PDYSIQVVAMDG-GGLKGTGTASIRVKDINDMPP  1745

 Score = 65.9 bits (159),  Expect = 3e-09, Method: Compositional matrix adjust.
 Identities = 85/359 (24%), Positives = 153/359 (43%), Gaps = 32/359 (9%)

Query  51    FVVDKNTGDINITAIVDRE-ETPSFL-ITCRALNAQGLDVEKPLILTVKILDINDNPPVF  108
             F VD+ TG+++   + DRE +   F+ +T +A +     +E     TV+I D+NDNPP+F
Sbjct  1025  FTVDEETGEVSTNKVFDREGDDGKFVSVTVKATDQGEPSLEGVCSFTVEITDVNDNPPLF  1084

Query  109   SQQIFMGEIEENSASNSLVMILNATDADEPNHLNSKIAFKIVS-QEPAGTPMFLLSRNTG  167
              +Q ++  ++++++  + ++ ++A+D D  N  N  I + + +   P     F +   +G
Sbjct  1085  DRQKYVENVKQDASIGTNILRVSASDEDADN--NGAIVYTLSAPHSPNDLEYFEIQPESG  1142

Query  168   EVRTLTNSLDREQASSYRLVVSGADKDGEGLSTQCECNIKVKDVNDNFPMFRDSQYSARI  227
              +  L   LDRE+   YRL V  +D+ G+  ST            +N P   D      I
Sbjct  1143  WI-VLKKPLDRER---YRLKVYASDR-GDPPSTADVDVELDVVDRNNKPPIWDRTIYGPI  1197

Query  228   EENILSSELLRFQVTDLDEEYTDNWLAVYFFTSG-----NEGNWFEIQTDPRTNE--GIL  280
                   +                N    Y    G     N+ + F +Q  P   +    +
Sbjct  1198  HIRENVTVGTVVTSVKASSGIEGNPTVFYRLMPGSTAQTNKLHTFYLQQRPDNGDTWADI  1257

Query  281   KVVKALDYEQLQSVKLSIAVKNKAEFHQSVISRYRVQSTPVTIQVINVREGIAFRPASKT  340
             KV   LD+E ++   L++ V+N         ++        V IQ+ +V + I      +
Sbjct  1258  KVNHPLDFETIKEYNLTVRVENNG-------AQQLASEATVYIQLEDVNDEIPLFTEREQ  1310

Query  341   FTVQKGISSKKLVDYILGTYQAIDEDTNKAASNVKYVM---GRNDG-GYLMIDSKTAEI  395
              TV +G      V  +     AID+D     + V Y +    RN+G  +  I+ ++ EI
Sbjct  1311  ETVLEGEPIGTKVTQV----NAIDKDGTFPNNQVYYYIVDSPRNEGKDFFEINLQSGEI  1365

 Score = 55.8 bits (133),  Expect = 3e-06, Method: Compositional matrix adjust.
 Identities = 49/158 (31%), Positives = 78/158 (49%), Gaps = 24/158 (15%)

Query  102   NDNPPVFSQQIFMGEIEENSASNSLVMILNATDADEPNHLNSKIAFKIVSQEPAGTP--M  159
             ND  P FSQQ++   ++EN+  N+LV  + A+D D  N     + F  V     GT
Sbjct  858   NDEEPKFSQQVYTPNVDENAGPNTLVTTVVASDKDGDN-----VRFGFVG---GGTSSGQ  909

Query  160   FLLSRNTGEVRTLTN--SLDREQASSYRLVVSGADK-------DGEGLSTQCECNIKVKD  210
             F++   TG +R  +   SLDR++   Y L V+  D        D    ++     + + D
Sbjct  910   FVIEEITGVIRLHSKAISLDRDK---YELNVTAMDDGSCCVNGDATIHTSTAVVVVFITD  966

Query  211   VNDNFPMFRD-SQYSARIEENILS-SELLRFQVTDLDE  246
```

```
               VNDN P+F+D   Y  ++EE   + S +++ Q TD D+
Sbjct  967     VNDNKPVFKDCGTYYPKVEEGAPNGSPVIKVQATDEDK  1004

 Score = 53.9 bits (128),  Expect = 1e-05, Method: Compositional matrix adjust.
 Identities = 70/306 (23%), Positives = 126/306 (41%), Gaps = 30/306 (10%)

Query  20    PIAKIT-SDYQATQKITYRI---SGVGIDQPPFGIFVVDKNTGDINITAIVDRE---ETP  72
             PI  +T  D   T K  Y++   SG G D+  F +   +  TG + I   +D E   ++
Sbjct  1768  PILTVTVHDEDETNKFQYKVIDNSGYGADK--FTMVRNNDGTGSLKIVQPLDYEDPLQSN  1825

Query  73    SFLITCRALNAQGLDVEKPLI------LTVKILDINDNPPVFSQQIFMGEIEENSASNSL  126
              F    + +N +G D +           + VK+ DINDN P F +      + EN+
Sbjct  1826  GFRFRIQ-VNDKGEDNDNDKYHVAYSWVVVKLRDINDNKPQFERPNIEVSVYENADVGKT  1884

Query  127   VMILNATDADEPNHLNSKIAFKIVSQEPAGTPMFLLSRNTGEVRTLTNSLDREQASSYRL  186
             +    ATD D+      SK+++ I    + +        S N      T+  +LDRE    +++
Sbjct  1885  LETFKATDPDQGG--KSKVSYAI---DRSSDRQRQFSINQEGTVTIQRNLDREVTPRHQV  1939

Query  187   VVSGADKDGEGLSTQCECNIKVKDVNDNFPMFRDSQYSARIEENILSSELLRFQVTDLDE  246
              +   D       +       + V+D+NDN P F    Y   + E++   +++    TD D+
Sbjct  1940  KILAIDDGIPPKTATATLTVIVQDINDNPPKFL-KDYRPVLPEHVPPRKVVEILATDDDD  1998

Query  247   EYTDNWLAVYFFTSGNEGNW----FEIQTDPRTNEG----ILKVVKALDYEQLQSVKLSI  298
                 N     F        +     F+++ D +   G    I+  +++ D EQ +   + I
Sbjct  1999  RSKSNGPPFQFRLDPGADDIIRASFKVEQDQKGANGDGMAIVSSLRSFDREQQKEYLIPI  2058

Query  299   AVKNKA  304
              +K+
Sbjct  2059  VIKDHG  2064

 Score = 52.0 bits (123),  Expect = 4e-05, Method: Compositional matrix adjust.
 Identities = 97/398 (24%), Positives = 163/398 (41%), Gaps = 54/398 (14%)

Query  106   PVFSQQIFMGEIEENSASNSLVMILNATDADEPNHLNSKIAFKIVSQ-EPAGTPMFLLSR  164
             P F    +  EI EN   +S ++ + A       +  + +I + + +Q + AGT  F +
Sbjct  655   PQFYMPSYEAEIPENQKKDSDIISVKAK-----SFADREIRYTLKAQGQGAGT--FNIGP  707

Query  165   NTGEVRTLTNSLDRE---QASSYRLVVSGADKDGEGLSTQCECNIKVKDVNDNFPMFRDS  221
              +G V+ L   LD E   Q   Y LVV+ A +D  G ST  E  I+V DVNDN P F
Sbjct  708   TSGIVK-LAKELDFEDLRQPHVYSLVVT-ATEDSGGFSTSVELTIRVTDVNDNAPKFELP  765

Query  222   QYSAR-IEENI-LSSELLRFQVTDLDEEYTDNWLAVYFFTSGNEGNWFEIQTDPRTNEGI  279
              Y A  ++E+I L + +L+ +  D D          + +  S +    F + ++     GI
Sbjct  766   DYQAHNVDEDIPLGTSILKVKAMDSDSGAN---AEIEYLVSDDH---FAVDSN-----GI  814

Query  280   LKVVKALDYEQLQS-VKLSIAVKNKAEFHQSVISRYRVQSTPVTIQVINVREGIAFRPAS  338
             +   K LD +   +  +  +  K+K E  ++  +  RV +             E   F
Sbjct  815   IVNNKQLDADNNNAYYEFVVTAKDKGEPAKTGTATVRVYTKNKN------DEEPKFSQQV  868

Query  339   KTFTVQKGISSKKLVDYILGTYQAIDEDTNKAASNVK--YVMGRNDGGYLMIDSKTAEIK  396
              T  V +       LV  ++          ++K    NV+  +V G    G  +I+  T  I+
Sbjct  869   YTPNVDENAGPNTLVTTVVA--------SDKDGDNVRFGFVGGGTSSGQFVIEEITGVIR  920

Query  397   FVK---NMNRDSTFI-VNKTITAEVLAIDEYTGKTSTGTVYVRVPDFNDN------CPTA  446
                   +++RD   + V              + T  TST  V V + D NDN      C T
Sbjct  921   LHSKAISLDRDKYELNVTAMDDGSCCVNGDATIHTSTAVVVVFITDVNDNKPVFKDCGTY  980

Query  447   VLE-KDAVCSSSPSVVVSARTLNNRYTGPYTFALEDQP  483
              + ++   + SP + V A   +    G   +++  QP
Sbjct  981   YPKVEEGAPNGSPVIKVQATDEDKGVNGQVKYSIVQQP  1018

 Score = 44.7 bits (104),  Expect = 0.008, Method: Compositional matrix adjust.
 Identities = 48/199 (24%), Positives = 73/199 (37%), Gaps = 19/199 (10%)

Query  27    DYQATQKITYRISGVGIDQPPFGIFVVDKNTGDINITAIVDREETPSFLITCRALNAQGL  86
             D     K++Y I       Q  F I        G + I   +DRE TP   +    A++
Sbjct  1894  DQGGKSKVSYAIDRSSDRQRQFSI----NQEGTVTIQRNLDREVTPRHQVKILAIDDGIP  1949
```

```
Query  87    DVEKPLILTVKILDINDNPPVFSQQIFMGEIEENSASNSLVMILNATDADEPNHLNSKIA  146
                    LTV + DINDNPP F +  +   + E+     +V IL   D D
Sbjct  1950  PKTATATLTVIVQDINDNPPKFLKD-YRPVLPEHVPPRKVVEILATDDDDRSKSNGPPFQ  2008

Query  147   FKIVSQEPAGTPMFLLSRNTGEVRTLTN-----------SLDREQASSYRLVVSGADKDG  195
             F++   +P    +   S    + +   N           S DREQ   Y + +   D
Sbjct  2009  FRL---DPGADDIIRASFKVEQDQKGANGDGMAIVSSLRSFDREQQKEYLIPIVIKDHGN  2065

Query  196   EGLSTQCECNIKVKDVNDN  214
               ++      + + DVNDN
Sbjct  2066  PAMTGTSTLTVVIGDVNDN  2084

 Score = 40.0 bits (92),  Expect = 0.20, Method: Compositional matrix adjust.
 Identities = 89/389 (23%), Positives = 149/389 (38%), Gaps = 62/389 (16%)

Query  75   LITCRALNAQGLDVEKPLILTVKILDINDNPPVFSQQIF--MGEIEENSASNSLVMILNA  132
            +IT +  NA GL       + + + D+ND PP F  +       ++ N+  N+ V  L A
Sbjct  517  IITNQGHNA-GLKYTDNQRVIIHVKDVNDEPPYFINRPLPMQAVVQLNAPPNTPVFTLQA  575

Query  133  TDADEPNHLNSKIAFKIVSQEPAGTPMFLLSRNTGEVRTLTNSLDREQASSYRLVVSGAD  192
             D D     +  I + IV     G   F +   +G VRT    L  +    Y L V   D
Sbjct  576  RDPD----TDHNIHYFIVRDRTGG--RFEVDERSGVVRTRGTDL-FQLDMEYVLYVKAED  628

Query  193  KDGE----GLSTQCECNIKVKDVNDNFPMFRDSQYSARIEEN-ILSSELLRFQVTDLDEE  247
            ++G+        +  E  + +       P F    Y A I EN    S+++  +     +
Sbjct  629  QNGKVDDRRFQSTPEERLSIVG-GKRAPQFYMPSYEAEIPENQKKDSDIISVKA----KS  683

Query  248  YTDNWLAVYFFTSGNEGNWFEIQTDPRTNEGILKVVKALDYEQLQSVKLS----IAVKNK  303
            + D  +       G     F I        GI+K+ K LD+E L+   +      A ++
Sbjct  684  FADREIRYTLKAQGQGAGTFNIGP----TSGIVKLAKELDFEDLRQPHVYSLVVTATEDS  739

Query  304  AEFHQSVISRYRVQSTPVTIQVINVREGIAFRPASKTFTVQKGISSKKLVDYILGT----  359
              F  SV          +TI+V +V +       +  F +    +     D  LGT
Sbjct  740  GGFSTSV---------ELTIRVTDVNDN------APKFELPDYQAHNVDEDIPLGTSILK  784

Query  360  YQAIDEDTNKAASNVKYVMGRN----DGGYLMIDSKTAEIKFVKNMNRDSTFIVNKTITA  415
             +A+D D+  A + ++Y++  +    D   +++++K  +     N N    F+V
Sbjct  785  VKAMDSDSG-ANAEIEYLVSDDHFAVDSNGIIVNNKQLD---ADNNNAYYEFVVTAKDKG  840

Query  416  EVLAIDEYTGKTSTGTVYVRVPDFNDNCP  444
            E         KT T TV V   + ND  P
Sbjct  841  E-------PAKTGTATVRVYTKNKNDEEP  862

 Score = 39.7 bits (91),  Expect = 0.27, Method: Compositional matrix adjust.
 Identities = 54/242 (22%), Positives = 102/242 (42%), Gaps = 22/242 (9%)

Query  4    KFAKPCREGE--DNSKRNP-IAKITSDYQATQKITYRISGVGIDQPPFGIFVVDKNTGDI  60
            +F  P  E E  +N K++  I + +    A ++I Y +   G      G F +   +G +
Sbjct  656  QFYMPSYEAEIPENQKKDSDIISVKAKSFADREIRYTLKAQGQGA---GTFNIGPTSGIV  712

Query  61   NITAIVDREE--TPSF--LITCRALNAQGLDVEKPLILTVKILDINDNPPVFSQQIFMGE  116
             +   +D E+   P    L+     ++ G      + LT+++ D+NDN P F    +
Sbjct  713  KLAKELDFEDLRQPHVYSLVVTATEDSGGFSTS--VELTIRVTDVNDNAPKFELPDYQAH  770

Query  117  -IEENSASNSLVMILNATDADEPNHLNSKIAFKIVSQEPAGTPMFLLSRNTGEVRTLTNS  175
             ++E+    + ++ + A D+D  +  N++I + +VS +      F +  N   V
Sbjct  771  NVDEDIPLGTSILKVKAMDSD--SGANAEIEY-LVSDD-----HFAVDSNGIIVNNKQLD  822

Query  176  LDREQASSYRLVVSGADKDGEGLSTQCECNIKVKDVNDNFPMFRDSQYSARIEENILSSE  235
             D   A  Y  VV+  DK     +      +  K+ ND  P F    Y+  ++EN   +
Sbjct  823  ADNNNA-YYEFVVTAKDKGEPAKTGTATVRVYTKNKNDEEPKFSQQVYTPNVDENAGPNT  881

Query  236  LL  237
            L+
Sbjct  882  LV  883

>ADAR2_001930.P21642 | transcript=ADAR2_001930.R21642 | gene=ADAR2_001930
| organism=Anopheles_darlingi_AdarGF1 | gene_product=neural-cadherin,
```

```
transcript variant X8 | transcript_product=neural-cadherin,
transcript variant X8 | location=AdarGF1_2RL:74991964-75171550(+)
| protein_length=3129 | sequence_SO=chromosome
| SO=protein_coding_gene | is_pseudo=false
Length=3129

 Score = 147 bits (371),  Expect = 2e-35, Method: Compositional matrix adjust.
 Identities = 128/446 (29%), Positives = 217/446 (49%), Gaps = 36/446 (8%)

Query  20    PIAKITS---DYQATQKITYRISGVGID--QPPFGIFVVDKNTGDINITAIVDREET---  71
             PI ++T+   D    Q I Y ++G GID   P    F +++ TG+I +   +DR++
Sbjct  1569  PIMQVTATDGDKDRPQNIVYFLTGQGIDPDNPANSKFDINRTTGEIFVLKPLDRDQPNGR  1628

Query  72    PSFLITCRALNAQGLDVEKPLILTVKILDINDNPPVFSQQIFMGEIEENSASNSLVMILN  131
             P +  T  A +  G  +     + V + DINDN P+F Q ++ G + EN  +  +VM +
Sbjct  1629  PQWRFTVFAQDEGGEGLVGYADVQVNLKDINDNAPIFPQGVYFGNVTENGTAGMVVMTMT  1688

Query  132   ATDADEPNH-LNSKIAFKI---VSQEPAGTPMFLLSRNTGEVRTLTNSLDREQASSYRLV  187
             A D D+PN   N+K+ + I   V +E  G+P+F +  +TG ++T    LDRE+   Y +
Sbjct  1689  AVDYDDPNEGTNAKLIYSIEKNVIEEETGSPIFEIEADTGVIKTAVCCLDRERTPDYSIQ  1748

Query  188   VSGADKDGEGLSTQCECNIKVKDVNDNFPMFRDSQYSARIEENILSS--ELLRFQVTDLD  245
             V     DG GL      +I+VKD+ND  P F   ++   ++E   ++  E+    VT  D
Sbjct  1749  VVAM--DGGGLKGTGTASIRVKDINDMPPQFTKDEWFTEVDETDGTNLPEMPILTVTVHD  1806

Query  246   EEYTDNWLAVYFFTSGNEGNWFEIQTDPRTNEGI--LKVVKALDYE---QLQSVKLSIAV  300
             E+ T+ +       SG   + F +    R N+G   LK+V+ LDYE   Q    +  I V
Sbjct  1807  EDETNKFQYKVIDNSGYGADKFTMV---RNNDGTGSLKIVQPLDYEDPLQSNGFRFRIQV  1863

Query  301   KNKAEFHQSVISRYRVQSTPVTIQVINVREGIAFRPASKTFTVQKGISSKKLVDYILGTY  360
              +K E + +   +Y V  + V +++ ++ +    +P  +   ++  +     V   L T+
Sbjct  1864  NDKGEDNDN--DKYHVAYSWVVVKLRDINDN---KPQFERPNIEVSVYENADVGKTLETF  1918

Query  361   QAIDEDTNKAASNVKYVMGR-NDGGYLMIDSKTAEIKFVKNMNRDSTFIVNKTITAEVLA  419
             +A D D     S V Y + R +D       ++    +   +N++R+    V      ++LA
Sbjct  1919  KATDPDQG-GKSKVSYAIDRSSDRQRQFSINQEGTVTIQRNLDRE----VTPRHQVKILA  1973

Query  420   IDE-YTGKTSTGTVYVRVPDFNDNCP  444
             ID+    KT+T T+ V V D NDN P
Sbjct  1974  IDDGIPPKTATATLTVIVQDINDNPP  1999

 Score = 80.5 bits (197),  Expect = 7e-14, Method: Compositional matrix adjust.
 Identities = 107/424 (25%), Positives = 178/424 (42%), Gaps = 51/424 (12%)

Query  53    VDKNTGDINITAIVDREETPSFLITCRALNAQGLDVEKPLILTVKILDINDNPPVFSQ-Q  111
             +D++  ++N+TA+ D         +C  +N          ++ V I D+NDN PVF
Sbjct  957   LDRDKYELNVTAMDDG--------SC-CVNGDATIHTSTAVVVVFITDVNDNKPVFKDCG  1007

Query  112   IFMGEIEENSASNSLVMILNATDADEPNHLNSKIAFKIVSQEPAGTPMFLLSRNTGEVRT  171
              +  ++EE + + S V+ + ATD D+   +N ++ + IV Q       F +   TGEV T
Sbjct  1008  TYYPKVEEGAPNGSPVIKVQATDEDK--GVNGQVKYSIVQQPNQKGTKFTVDEETGEVST  1065

Query  172   LTNSLDRE--QASSYRLVVSGADKDGEGLSTQCECNIKVKDVNDNFPMFRDSQYSARIEE  229
                  DRE        + V   D+    L   C   +++ DVNDN P+F   +Y   +++
Sbjct  1066  -NKVFDREGDDGKFVSVTVKATDQGEPSLEGVCSFTVEITDVNDNPPLFDRQKYVENVKQ  1124

Query  230   NI-LSSELLRFQVTDLDEEYTDNWLAVYFFT---SGNEGNWFEIQTDPRTNEGILKVVKA  285
             +  + + +LR  V+  DE+  +N   VY  +   S N+  +FEIQ  P +   +LK  K
Sbjct  1125  DASIGTNILR--VSASDEDADNNGAIVYTLSAPHSPNDLEYFEIQ--PESGWIVLK--KP  1178

Query  286   LDYEQLQSVKLSIAVKNK------AEFHQSVISRYRVQSTPV----TIQVINVREGIAFR  335
             LD E+    +L +   ++      A+    +    R    P+        I++RE +
Sbjct  1179  LDRERY---RLKVYASDRGDPPSTADVDVELDVVDRNNKPPIWDRNIYGPIHIRENVTVG  1235

Query  336   PASKTFTVQKGISSKKLVDYILGTYQAIDEDTNKAASNVKYVMGRNDGGYLMIDSKTAEI  395
                       GI     V Y L         TNK   +  Y+  R D G        A+I
Sbjct  1236  TVVTAIKASSGIEGNPTVFYRL--MPGSTAQTNKL--HTFYLQQRPDNG-----DTWADI  1286

Query  396   KFVKNMNRDSTFIVNKTITAEVLAIDEYTGKTSTGTVYVRVPDFNDNCPTAV-LEKDAVC  454
             K    ++ ++    N TI  E     +     S  TVY+++ D ND  P     E++ V
```

```
Sbjct  1287  KVNHPLDFETIKEYNLTIRVENNGAQQL---ASEATVYIQLEDVNDEIPLFTEREQETVL  1343

Query  455   SSSP  458
                P
Sbjct  1344  EGEP  1347

 Score = 77.4 bits (189),  Expect = 6e-13, Method: Compositional matrix adjust.
 Identities = 103/419 (25%), Positives = 173/419 (41%), Gaps = 51/419 (12%)

Query  50    IFVVDKNTGDINITAIVDREETPSFLITCRAL----------NAQGLDVEKPLILTVKIL  99
              F ++   +G+I    + DRE+  ++ +   A           N Q   V K   + + I
Sbjct  1384  FFEINLQSGEIFTKVVFDREKQGAYALEVEARDGAPSARPNSNQQPNSVTK--FIRIGIA  1441

Query  100   DINDNPPVFSQQIFMGEIEENSASNSLVMILNATDADEPNHLNSKIAFKIVSQEPAGTPM  159
             D NDNPP F + ++  E++EN       V+ + A D DE    +S+I ++I S    G
Sbjct  1442  DKNDNPPYFDKALYEAEVDENEDIQHTVLTVTAKDHDE----SSRIRYEITSGNIGGA--  1495

Query  160   FLLSRNTGEVRTLTNSLDREQASSYRLVVSGADKDGEGLSTQCECNIKVKDVNDNFPMFR  219
             F +   TG +  +  +LD E    Y L +  +D   E     +    I VKDVND  P F
Sbjct  1496  FAVKNMTGAI-YVAGALDYETRKRYHLTLVASDSLNE---NETIIEIHVKDVNDMPPTFS  1551

Query  220   DSQY--SARIEENILSSELLRFQVTDLDEEYTDNWLAVYFFTSG--NEGNWFEIQTDPRT  275
                Y  +   E+ +    +++   TD D++   N   VYF T    +  N    + D
Sbjct  1552  QQVYETTQDEEQTVTDRPIMQVTATDGDKDRPQN--IVYFLTGQGIDPDNPANSKFDINR  1609

Query  276   NEGILKVVKALDYEQLQS-VKLSIAVKNKAEFHQSVISRYRVQSTPVTIQVINVREGIAF  334
               G + V+K LD +Q      +    V  + E  + ++    VQ     + + ++ +
Sbjct  1610  TTGEIFVLKPLDRDQPNGRPQWRFTVFAQDEGGEGLVGYADVQ-----VNLKDINDNAPI  1664

Query  335   RPASKTFTVQKGISSKKLVDYILGTYQAIDEDTNKAASNVK--YVMGRN------DGGYL  386
              P    F     ++        ++ T  A+D D     +N K  Y + +N
Sbjct  1665  FPQGVYF---GNVTENGTAGMVVMTMTAVDYDDPNEGTNAKLIYSIEKNVIEEETGSPIF  1721

Query  387   MIDSKTAEIK-FVKNMNRDSTFIVNKTITAEVLAIDEYTGKTSTGTVYVRVPDFNDNCP  444
              I++ T  IK  V  ++R+ T        + +V+A+D   G   TGT  +RV D ND  P
Sbjct  1722  EIEADTGVIKTAVCCLDRERT----PDYSIQVVAMDG-GGLKGTGTASIRVKDINDMPP  1775

 Score = 68.6 bits (166),  Expect = 4e-10, Method: Compositional matrix adjust.
 Identities = 88/361 (24%), Positives = 157/361 (43%), Gaps = 36/361 (10%)

Query  51    FVVDKNTGDINITAIVDRE-ETPSFL-ITCRALNAQGLDVEKPLILTVKILDINDNPPVF  108
             F VD+ TG+++    + DRE +   F+ +T +A +     +E     TV+I D+NDNPP+F
Sbjct  1054  FTVDEETGEVSTNKVFDREGDDGKFVSVTVKATDQGEPSLEGVCSFTVEITDVNDNPPLF  1113

Query  109   SQQIFMGEIEENSASNSLVMILNATDADEPNHLNSKIAFKIVS-QEPAGTPMFLLSRNTG  167
              +Q ++  ++++++  + ++ ++A+D D  N  N  I + + +   P     F +   +G
Sbjct  1114  DRQKYVENVKQDASIGTNILRVSASDEDADN--NGAIVYTLSAPHSPNDLEYFEIQPESG  1171

Query  168   EVRTLTNSLDREQASSYRLVVSGADKDGEGLSTQCECNIKVKDVNDNFPMFRDSQYSA--  225
              +  L   LDRE+   YRL V  +D+ G+  ST              +N P   D
Sbjct  1172  WI-VLKKPLDRER---YRLKVYASDR-GDPPSTADVDVELDVVDRNNKPPIWDRNIYGPI  1226

Query  226   RIEENILSSELLRFQVTDLDEEYTDNWLAVYFFTSG-----NEGNWFEIQTDPRTNE--G  278
              I EN+    ++               N    Y    G     N+ + F +Q  P   +
Sbjct  1227  HIRENVTVGTVV--TAIKASSGIEGNPTVFYRLMPGSTAQTNKLHTFYLQQRPDNGDTWA  1284

Query  279   ILKVVKALDYEQLQSVKLSIAVKNKAEFHQSVISRYRVQSTPVTIQVINVREGIAFRPAS  338
              +KV   LD+E ++   L+I V+N           ++        V IQ+ +V + I
Sbjct  1285  DIKVNHPLDFETIKEYNLTIRVENNG-------AQQLASEATVYIQLEDVNDEIPLFTER  1337

Query  339   KTFTVQKGISSKKLVDYILGTYQAIDEDTNKAASNVKYVM---GRNDG-GYLMIDSKTAE  394
             +  TV +G      V  +      AID+D      + V Y +      RN+G  +  I+ ++ E
Sbjct  1338  EQETVLEGEPIGTKVTQV----NAIDKDGTFPNNQVYYYIVDSPRNEGKDFFEINLQSGE  1393

Query  395   I  395
             I
Sbjct  1394  I  1394
```

```
 Score = 55.8 bits (133),  Expect = 3e-06, Method: Compositional matrix adjust.
 Identities = 49/158 (31%), Positives = 78/158 (49%), Gaps = 24/158 (15%)

Query  102   NDNPPVFSQQIFMGEIEENSASNSLVMILNATDADEPNHLNSKIAFKIVSQEPAGTP--M  159
             ND  P FSQQ++   ++EN+  N+LV  + A+D D  N     + F  V     GT
Sbjct  887   NDEEPKFSQQVYTPNVDENAGPNTLVTTVVASDKDGDN-----VRFGFVG---GGTSSGQ  938

Query  160   FLLSRNTGEVRTLTN--SLDREQASSYRLVVSGADK-------DGEGLSTQCECNIKVKD  210
             F++   TG +R  +   SLDR++   Y L V+  D        D    ++     + + D
Sbjct  939   FVIEEITGVIRLHSKAISLDRDK---YELNVTAMDDGSCCVNGDATIHTSTAVVVVFITD  995

Query  211   VNDNFPMFRD-SQYSARIEENILS-SELLRFQVTDLDE  246
             VNDN P+F+D   Y  ++EE   + S +++ Q TD D+
Sbjct  996   VNDNKPVFKDCGTYYPKVEEGAPNGSPVIKVQATDEDK  1033

 Score = 53.9 bits (128),  Expect = 1e-05, Method: Compositional matrix adjust.
 Identities = 70/306 (23%), Positives = 126/306 (41%), Gaps = 30/306 (10%)

Query  20    PIAKIT-SDYQATQKITYRI---SGVGIDQPPFGIFVVDKNTGDINITAIVDRE---ETP  72
             PI  +T  D   T K  Y++   SG G D+  F +   +  TG + I   +D E   ++
Sbjct  1798  PILTVTVHDEDETNKFQYKVIDNSGYGADK--FTMVRNNDGTGSLKIVQPLDYEDPLQSN  1855

Query  73    SFLITCRALNAQGLDVEKPLI------LTVKILDINDNPPVFSQQIFMGEIEENSASNSL  126
              F    + +N +G D +          + VK+ DINDN P F +      + EN+
Sbjct  1856  GFRFRIQ-VNDKGEDNDNDKYHVAYSWVVVKLRDINDNKPQFERPNIEVSVYENADVGKT  1914

Query  127   VMILNATDADEPNHLNSKIAFKIVSQEPAGTPMFLLSRNTGEVRTLTNSLDREQASSYRL  186
             +    ATD D+     SK+++ I   + +       S N     T+  +LDRE    +++
Sbjct  1915  LETFKATDPDQGG--KSKVSYAI---DRSSDRQRQFSINQEGTVTIQRNLDREVTPRHQV  1969

Query  187   VVSGADKDGEGLSTQCECNIKVKDVNDNFPMFRDSQYSARIEENILSSELLRFQVTDLDE  246
              +   D      +      + V+D+NDN P F    Y   + E++   +++    TD D+
Sbjct  1970  KILAIDDGIPPKTATATLTVIVQDINDNPPKFL-KDYRPVLPEHVPPRKVVEILATDDDD  2028

Query  247   EYTDNWLAVYFFTSGNEGNW----FEIQTDPRTNEG----ILKVVKALDYEQLQSVKLSI  298
                 N     F        +     F+++ D +   G    I+  +++ D EQ +   + I
Sbjct  2029  RSKSNGPPFQFRLDPGADDIIRASFKVEQDQKGANGDGMAIVSSLRSFDREQQKEYLIPI  2088

Query  299   AVKNKA  304
              +K+
Sbjct  2089  VIKDHG  2094

 Score = 52.8 bits (125),  Expect = 2e-05, Method: Compositional matrix adjust.
 Identities = 97/398 (24%), Positives = 163/398 (41%), Gaps = 54/398 (14%)

Query  106   PVFSQQIFMGEIEENSASNSLVMILNATDADEPNHLNSKIAFKIVSQ-EPAGTPMFLLSR  164
             P F    +  EI EN   +S ++ + A      +  + +I + + +Q + AGT  F +
Sbjct  684   PQFYMPSYEAEIPENQKKDSDIISVKAK-----SFADREIRYTLKAQGQGAGT--FNIGP  736

Query  165   NTGEVRTLTNSLDRE---QASSYRLVVSGADKDGEGLSTQCECNIKVKDVNDNFPMFRDS  221
              +G V+ L   LD E   Q   Y LVV+ A +D  G ST  E  I+V DVNDN P F
Sbjct  737   TSGIVK-LAKELDFEDLRQPHVYSLVVT-ATEDSGGFSTSVELTIRVTDVNDNAPKFELP  794

Query  222   QYSAR-IEENI-LSSELLRFQVTDLDEEYTDNWLAVYFFTSGNEGNWFEIQTDPRTNEGI  279
              Y A  ++E+I L + +L+ +  D D         + +  S +    F + ++     GI
Sbjct  795   DYQAHNVDEDIPLGTSILKVKAMDADSGAN---AEIEYLVSDDH---FAVDSN-----GI  843

Query  280   LKVVKALDYEQLQS-VKLSIAVKNKAEFHQSVISRYRVQSTPVTIQVINVREGIAFRPAS  338
             +   K LD +   +  +  +  K+K E  ++  +  RV +           E   F
Sbjct  844   IVNNKQLDADNNNAYYEFVVTAKDKGEPAKTGTATVRVYTKNKN------DEEPKFSQQV  897

Query  339   KTFTVQKGISSKKLVDYILGTYQAIDEDTNKAASNVK--YVMGRNDGGYLMIDSKTAEIK  396
              T  V +      LV  ++         ++K   NV+  +V G    G  +I+  T  I+
Sbjct  898   YTPNVDENAGPNTLVTTVVA--------SDKDGDNVRFGFVGGGTSSGQFVIEEITGVIR  949

Query  397   FVK---NMNRDSTFI-VNKTITAEVLAIDEYTGKTSTGTVYVRVPDFNDN------CPTA  446
                   +++RD   + V            + T  TST  V V + D NDN      C T
Sbjct  950   LHSKAISLDRDKYELNVTAMDDGSCCVNGDATIHTSTAVVVVFITDVNDNKPVFKDCGTY  1009
```

```
Query  447   VLE-KDAVCSSSPSVVVSARTLNNRYTGPYTFALEDQP  483
               + ++   + SP + V A   +    G   +++  QP
Sbjct  1010  YPKVEEGAPNGSPVIKVQATDEDKGVNGQVKYSIVQQP  1047

 Score = 44.7 bits (104),  Expect = 0.008, Method: Compositional matrix adjust.
 Identities = 48/199 (24%), Positives = 73/199 (37%), Gaps = 19/199 (10%)

Query  27    DYQATQKITYRISGVGIDQPPFGIFVVDKNTGDINITAIVDREETPSFLITCRALNAQGL  86
             D     K++Y I      Q  F I       G + I   +DRE TP   +   A++
Sbjct  1924  DQGGKSKVSYAIDRSSDRQRQFSI----NQEGTVTIQRNLDREVTPRHQVKILAIDDGIP  1979

Query  87    DVEKPLILTVKILDINDNPPVFSQQIFMGEIEENSASNSLVMILNATDADEPNHLNSKIA  146
                    LTV + DINDNPP F +  +   + E+     +V IL   D D
Sbjct  1980  PKTATATLTVIVQDINDNPPKFLKD-YRPVLPEHVPPRKVVEILATDDDDRSKSNGPPFQ  2038

Query  147   FKIVSQEPAGTPMFLLSRNTGEVRTLTN-----------SLDREQASSYRLVVSGADKDG  195
             F++   +P    +   S    + +   N           S DREQ   Y + +   D
Sbjct  2039  FRL---DPGADDIIRASFKVEQDQKGANGDGMAIVSSLRSFDREQQKEYLIPIVIKDHGN  2095

Query  196   EGLSTQCECNIKVKDVNDN  214
               ++      + + DVNDN
Sbjct  2096  PAMTGTSTLTVVIGDVNDN  2114

 Score = 40.8 bits (94),  Expect = 0.11, Method: Compositional matrix adjust.
 Identities = 55/242 (23%), Positives = 102/242 (42%), Gaps = 22/242 (9%)

Query  4     KFAKPCREGE--DNSKRNP-IAKITSDYQATQKITYRISGVGIDQPPFGIFVVDKNTGDI  60
            +F  P  E E  +N K++  I  + +   A ++I Y +   G      G F +   +G +
Sbjct  685  QFYMPSYEAEIPENQKKDSDIISVKAKSFADREIRYTLKAQGQGA---GTFNIGPTSGIV  741

Query  61   NITAIVDREE--TPSF--LITCRALNAQGLDVEKPLILTVKILDINDNPPVFSQQIFMGE  116
             +   +D E+   P    L+     ++ G      + LT+++ D+NDN P F    +
Sbjct  742  KLAKELDFEDLRQPHVYSLVVTATEDSGGFSTS--VELTIRVTDVNDNAPKFELPDYQAH  799

Query  117  -IEENSASNSLVMILNATDADEPNHLNSKIAFKIVSQEPAGTPMFLLSRNTGEVRTLTNS  175
             ++E+    + ++ + A DAD  +  N++I + +VS +      F +  N   V
Sbjct  800  NVDEDIPLGTSILKVKAMDAD--SGANAEIEY-LVSDD-----HFAVDSNGIIVNNKQLD  851

Query  176  LDREQASSYRLVVSGADKDGEGLSTQCECNIKVKDVNDNFPMFRDSQYSARIEENILSSE  235
             D   A  Y  VV+  DK     +      +  K+ ND  P F    Y+  ++EN   +
Sbjct  852  ADNNNA-YYEFVVTAKDKGEPAKTGTATVRVYTKNKNDEEPKFSQQVYTPNVDENAGPNT  910

Query  236  LL  237
            L+
Sbjct  911  LV  912

 Score = 39.7 bits (91),  Expect = 0.25, Method: Compositional matrix adjust.
 Identities = 89/389 (23%), Positives = 149/389 (38%), Gaps = 62/389 (16%)

Query  75   LITCRALNAQGLDVEKPLILTVKILDINDNPPVFSQQIF--MGEIEENSASNSLVMILNA  132
            +IT +  NA GL       + + + D+ND PP F  +        ++ N+  N+ V  L A
Sbjct  546  IITNQGHNA-GLKYTDNQRVIIHVKDVNDEPPYFINRPLPMQAVVQLNAPPNTPVFTLQA  604

Query  133  TDADEPNHLNSKIAFKIVSQEPAGTPMFLLSRNTGEVRTLTNSLDREQASSYRLVVSGAD  192
             D D     +  I + IV     G   F +   +G VRT    L  +    Y L V   D
Sbjct  605  RDPD----TDHNIHYFIVRDRTGG--RFEVDERSGVVRTRGTDL-FQLDMEYVLYVKAED  657

Query  193  KDGE----GLSTQCECNIKVKDVNDNFPMFRDSQYSARIEEN-ILSSELLRFQVTDLDEE  247
            ++G+       +  E  + +       P F    Y A I EN    S+++  +     +
Sbjct  658  QNGKVDDRRFQSTPEERLSIVG-GKRAPQFYMPSYEAEIPENQKKDSDIISVKA----KS  712

Query  248  YTDNWLAVYFFTSGNEGNWFEIQTDPRTNEGILKVVKALDYEQLQSVKLS----IAVKNK  303
            + D  +       G     F I        GI+K+ K LD+E L+   +      A ++
Sbjct  713  FADREIRYTLKAQGQGAGTFNIGP----TSGIVKLAKELDFEDLRQPHVYSLVVTATEDS  768

Query  304  AEFHQSVISRYRVQSTPVTIQVINVREGIAFRPASKTFTVQKGISSKKLVDYILGT----  359
              F  SV          +TI+V +V +       +  F +    +     D  LGT
Sbjct  769  GGFSTSV---------ELTIRVTDVNDN------APKFELPDYQAHNVDEDIPLGTSILK  813
```

```
Query  360  YQAIDEDTNKAASNVKYVMGRN----DGGYLMIDSKTAEIKFVKNMNRDSTFIVNKTITA  415
             +A+D D+  A + ++Y++  +    D   +++++K  +     N N    F+V
Sbjct  814  VKAMDADSG-ANAEIEYLVSDDHFAVDSNGIIVNNKQLD---ADNNNAYYEFVVTAKDKG  869

Query  416  EVLAIDEYTGKTSTGTVYVRVPDFNDNCP  444
            E         KT T TV V   + ND  P
Sbjct  870  E-------PAKTGTATVRVYTKNKNDEEP  891

>AMAC_003346.P3008 | transcript=AMAC_003346.R3008 | gene=AMAC_003346
| organism=Anopheles_maculipalpis_AmacGA1 | gene_product=neural-cadherin,
transcript variant X12 | transcript_product=neural-cadherin,
transcript variant X10 | location=AmacGA1_3RL:17033394-17218514(+)
| protein_length=3106 | sequence_SO=chromosome
| SO=protein_coding_gene | is_pseudo=false
Length=3106

 Score = 147 bits (371),  Expect = 2e-35, Method: Compositional matrix adjust.
 Identities = 128/446 (29%), Positives = 217/446 (49%), Gaps = 36/446 (8%)

Query  20    PIAKITS---DYQATQKITYRISGVGID--QPPFGIFVVDKNTGDINITAIVDREET---  71
             PI ++T+   D    Q I Y ++G GID   P    F +++ TG+I +   +DR++
Sbjct  1547  PIMQVTATDGDKDRPQNIVYFLTGQGIDPDNPANSKFDINRTTGEIFVLKPLDRDQPNGR  1606

Query  72    PSFLITCRALNAQGLDVEKPLILTVKILDINDNPPVFSQQIFMGEIEENSASNSLVMILN  131
             P +  T  A +  G  +     + V + DINDN P+F Q ++ G + EN  +  +VM +
Sbjct  1607  PQWRFTVFAQDEGGEGLVGYADVQVNLKDINDNAPIFPQGVYFGNVTENGTAGMVVMTMT  1666

Query  132   ATDADEPNH-LNSKIAFKI---VSQEPAGTPMFLLSRNTGEVRTLTNSLDREQASSYRLV  187
             A D D+PN   N+K+ + I   V +E  G+P+F +  +TG ++T    LDRE+   Y +
Sbjct  1667  AVDYDDPNEGTNAKLIYSIEKNVIEEETGSPIFEIEADTGVIKTAVCCLDRERTPDYSIQ  1726

Query  188   VSGADKDGEGLSTQCECNIKVKDVNDNFPMFRDSQYSARIEENILSS--ELLRFQVTDLD  245
             V     DG GL      +I+VKD+ND  P F   ++   ++E   ++  E+    VT  D
Sbjct  1727  VVAM--DGGGLKGTGTASIRVKDINDMPPQFTKDEWFTEVDETDGTNLPEMPILTVTVHD  1784

Query  246   EEYTDNWLAVYFFTSGNEGNWFEIQTDPRTNEGI--LKVVKALDYE---QLQSVKLSIAV  300
             E+ T+ +       SG   + F +    R N+G   LK+V+ LDYE   Q    +  I V
Sbjct  1785  EDETNKFQYKVIDNSGYGADKFTMV---RNNDGTGSLKIVQPLDYEDPLQSNGFRFRIQV  1841

Query  301   KNKAEFHQSVISRYRVQSTPVTIQVINVREGIAFRPASKTFTVQKGISSKKLVDYILGTY  360
              +K E + +   +Y V  + V +++ ++ +    +P  +   ++  +     V   L T+
Sbjct  1842  NDKGEDNDN--DKYHVAYSWVVVKLRDINDN---KPQFERPNIEVSVYENADVGKTLETF  1896

Query  361   QAIDEDTNKAASNVKYVMGR-NDGGYLMIDSKTAEIKFVKNMNRDSTFIVNKTITAEVLA  419
             +A D D     S V Y + R +D       ++   +   +N++R+    V      ++LA
Sbjct  1897  KATDPDQG-GKSKVSYAIDRSSDRQRQFSINQEGTVTIQRNLDRE----VTPRHQVKILA  1951

Query  420   IDE-YTGKTSTGTVYVRVPDFNDNCP  444
             ID+    KT+T T+ V V D NDN P
Sbjct  1952  IDDGIPPKTATATLTVIVQDINDNPP  1977

 Score = 79.0 bits (193),  Expect = 2e-13, Method: Compositional matrix adjust.
 Identities = 106/424 (25%), Positives = 180/424 (42%), Gaps = 51/424 (12%)

Query  53    VDKNTGDINITAIVDREETPSFLITCRALNAQGLDVEKPLILTVKILDINDNPPVFSQ-Q  111
             +D++  ++N+TA+ D         +C  +N          ++ V I D+NDN PVF
Sbjct  936   LDRDKYELNVTAMDDG--------SC-CVNGDATIHTSTAVVVVFITDVNDNKPVFKDCG  986

Query  112   IFMGEIEENSASNSLVMILNATDADEPNHLNSKIAFKIVSQEPAGTPMFLLSRNTGEVRT  171
              +  ++EE + + S V+ + ATD D+   +N ++ + IV Q       F +   TGEV T
Sbjct  987   TYYPKVEEGAPNGSPVIKVQATDEDK--GVNGQVKYSIVQQPNQKGTKFTVDEETGEVST  1044

Query  172   LTNSLDRE--QASSYRLVVSGADKDGEGLSTQCECNIKVKDVNDNFPMFRDSQYSARIEE  229
                  DRE        + V   D+    L   C   +++ DVNDN P+F   +Y   +++
Sbjct  1045  -NKVFDREGDDGKFVSVTVKATDQGEPSLEGVCSFTVEITDVNDNPPLFDRQKYVENVKQ  1103

Query  230   NI-LSSELLRFQVTDLDEEYTDNWLAVYFFT---SGNEGNWFEIQTDPRTNEGILKVVKA  285
             +  + + +LR  V+  DE+ +N    VY  +   S N+  +FEIQ  P +   +LK  K
```

```
Sbjct  1104  DASIGTNILR--VSASDEDADNNGAIVYTLSAPHSPNDLEYFEIQ--PESGWIVLK--KP  1157

Query  286   LDYEQLQSVKLSIAVKNK------AEFHQSVISRYRVQSTPVTIQV----INVREGIAFR  335
             LD E+    +L +   ++      A+    +    R    P+  +     I++RE +
Sbjct  1158  LDRERY---RLKVYASDRGDPPSTADVDVELDVVDRNNKPPIWDRTIYGPIHIRENVTVG  1214

Query  336   PASKTFTVQKGISSKKLVDYILGTYQAIDEDTNKAASNVKYVMGRNDGGYLMIDSKTAEI  395
                 +     GI     V Y L         TNK   +  Y+  R D G        A+I
Sbjct  1215  TVVTSVKASSGIEGNPTVFYRL--MPGSTAQTNKL--HTFYLQQRPDNG-----DTWADI  1265

Query  396   KFVKNMNRDSTFIVNKTITAEVLAIDEYTGKTSTGTVYYRVPDFNDNCPTAV-LEKDAVC  454
             K    ++ ++    N T+  E     +     S  TVY+++ D ND  P     E++ V
Sbjct  1266  KVNHPLDFETIKEYNLTVRVENNGAQQL---ASEATVYIQLEDVNDEIPLFTEREQETVL  1322

Query  455   SSSP  458
                P
Sbjct  1323  EGEP  1326

 Score = 77.0 bits (188),  Expect = 8e-13, Method: Compositional matrix adjust.
 Identities = 103/418 (25%), Positives = 174/418 (42%), Gaps = 50/418 (12%)

Query  50    IFVVDKNTGDINITAIVDREETPSFLITCRAL----------NAQGLDVEKPLILTVKIL  99
              F ++  +G+I    + DRE+  ++ +   A           N Q   V K   + + I
Sbjct  1363  FFEINLQSGEIFTKVVFDREKQGAYALEVEARDGAPSARPNSNQQPNSVTK--FIRIGIA  1420

Query  100   DINDNPPVFSQQIFMGEIEENSASNSLVMILNATDADEPNHLNSKIAFKIVSQEPAGTPM  159
             D NDNPP F + ++  E++EN      V+ + A D DE    +S+I ++I S    G
Sbjct  1421  DKNDNPPYFDKALYEAEVDENEDIQHTVLTVTAKDHDE----SSRIRYEITSGNIGGA--  1474

Query  160   FLLSRNTGEVRTLTNSLDREQASSYRLVVSGADKDGEGLSTQCECNIKVKDVNDNFPMFR  219
             F +   TG +  +  +LD E    Y L +  +D   E  +      I VKDVND  P F
Sbjct  1475  FAVKNMTGAI-YVAGALDYETRKRYHLTLVASDSLNENETIIV---INVKDVNDMPPSFP  1530

Query  220   DSQYSARIEENI-LSSELLRFQVTDLDEEYTDNWLAVYFFTSG--NEGNWFEIQTDPRTN  276
                Y   ++E + +   +++   TD D++   N   VYF T    +  N    + D
Sbjct  1531  QQVYERTMDEELAVPFPIMQVTATDGDKDRPQN--IVYFLTGQGIDPDNPANSKFDINRT  1588

Query  277   EGILKVVKALDYEQLQS-VKLSIAVKNKAEFHQSVISRYRVQSTPVTIQVINVREGIAFR  335
              G + V+K LD +Q     +    V  + E  + ++    VQ     + + ++ +
Sbjct  1589  TGEIFVLKPLDRDQPNGRPQWRFTVFAQDEGGEGLVGYADVQ-----VNLKDINDNAPIF  1643

Query  336   PASKTFTVQKGISSKKLVDYILGTYQAIDEDTNKAASNVK--YVMGRN------DGGYLM  387
             P    F     ++       ++ T  A+D D     +N K  Y + +N
Sbjct  1644  PQGVYF---GNVTENGTAGMVVMTMTAVDYDDPNEGTNAKLIYSIEKNVIEEETGSPIFE  1700

Query  388   IDSKTAEIK-FVKNMNRDSTFIVNKTITAEVLAIDEYTGKTSTGTVYYRVPDFNDNCP  444
             I++ T  IK  V  ++R+ T       + +V+A+D   G   TGT  +RV D ND  P
Sbjct  1701  IEADTGVIKTAVCCLDRERT----PDYSIQVVAMDG-GGLKGTGTASIRVKDINDMPP  1753

 Score = 65.5 bits (158),  Expect = 3e-09, Method: Compositional matrix adjust.
 Identities = 85/359 (24%), Positives = 153/359 (43%), Gaps = 32/359 (9%)

Query  51    FVVDKNTGDINITAIVDRE-ETPSFL-ITCRALNAQGLDVEKPLILTVKILDINDNPPVF  108
             F VD+ TG+++   + DRE +   F+ +T +A +     +E     TV+I D+NDNPP+F
Sbjct  1033  FTVDEETGEVSTNKVFDREGDDGKFVSVTVKATDQGEPSLEGVCSFTVEITDVNDNPPLF  1092

Query  109   SQQIFMGEIEENSASNSLVMILNATDADEPNHLNSKIAFKIVS-QEPAGTPMFLLSRNTG  167
              +Q ++  ++++++  + ++ ++A+D D  N  N  I + + +   P     F +   +G
Sbjct  1093  DRQKYVENVKQDASIGTNILRVSASDEDADN--NGAIVYTLSAPHSPNDLEYFEIQPESG  1150

Query  168   EVRTLTNSLDREQASSYRLVVSGADKDGEGLSTQCECNIKVKDVNDNFPMFRDSQYSARI  227
              +  L   LDRE+   YRL V  +D+ G+  ST            +N P   D      I
Sbjct  1151  WI-VLKKPLDRER---YRLKVYASDR-GDPPSTADVDVELDVVDRNNKPPIWDRTIYGPI  1205

Query  228   EENILSSELLRFQVTDLDEEYTDNWLAVYFFTSG-----NEGNWFEIQTDPRTNE--GIL  280
                   +                N    Y    G     N+ + F +Q  P   +    +
Sbjct  1206  HIRENVTVGTVVTSVKASSGIEGNPTVFYRLMPGSTAQTNKLHTFYLQQRPDNGDTWADI  1265

Query  281   KVVKALDYEQLQSVKLSIAVKNKAEFHQSVISRYRVQSTPVTIQVINVREGIAFRPASKT  340
```

```
             KV   LD+E ++   L++ V+N         ++       V IQ+ +V + I      +
Sbjct  1266  KVNHPLDFETIKEYNLTVRVENNG-------AQQLASEATVYIQLEDVNDEIPLFTEREQ  1318

Query  341   FTVQKGISSKKLVDYILGTYQAIDEDTNKAASNVKYVM---GRNDG-GYLMIDSKTAEI  395
              TV +G      V  +     AID+D     + V Y +    RN+G  +  I+ ++ EI
Sbjct  1319  ETVLEGEPIGTKVTQV----NAIDKDGTFPNNQVYYYIVDSPRNEGKDFFEINLQSGEI  1373

 Score = 55.8 bits (133),  Expect = 3e-06, Method: Compositional matrix adjust.
 Identities = 49/158 (31%), Positives = 78/158 (49%), Gaps = 24/158 (15%)

Query  102   NDNPPVFSQQIFMGEIEENSASNSLVMILNATDADEPNHLNSKIAFKIVSQEPAGTP--M  159
             ND  P FSQQ++   ++EN+  N+LV  + A+D D  N     + F  V     GT
Sbjct  866   NDEEPKFSQQVYTPNVDENAGPNTLVTTVVASDKDGDN-----VRFGFVG---GGTSSGQ  917

Query  160   FLLSRNTGEVRTLTN--SLDREQASSYRLVVSGADK-------DGEGLSTQCECNIKVKD  210
             F++   TG +R  +   SLDR++   Y L V+  D        D    ++     + + D
Sbjct  918   FVIEEITGVIRLHSKAISLDRDK---YELNVTAMDDGSCCVNGDATIHTSTAVVVVFITD  974

Query  211   VNDNFPMFRD-SQYSARIEENILS-SELLRFQVTDLDE  246
             VNDN P+F+D   Y  ++EE   + S +++ Q TD D+
Sbjct  975   VNDNKPVFKDCGTYYPKVEEGAPNGSPVIKVQATDEDK  1012

 Score = 53.9 bits (128),  Expect = 1e-05, Method: Compositional matrix adjust.
 Identities = 70/306 (23%), Positives = 126/306 (41%), Gaps = 30/306 (10%)

Query  20    PIAKIT-SDYQATQKITYRI---SGVGIDQPPFGIFVVDKNTGDINITAIVDRE---ETP  72
             PI  +T  D   T K  Y++   SG G D+  F +   +  TG + I   +D E   ++
Sbjct  1776  PILTVTVHDEDETNKFQYKVIDNSGYGADK--FTMVRNNDGTGSLKIVQPLDYEDPLQSN  1833

Query  73    SFLITCRALNAQGLDVEKPLI------LTVKILDINDNPPVFSQQIFMGEIEENSASNSL  126
              F    + +N +G D +          + VK+ DINDN P F +      + EN+
Sbjct  1834  GFRFRIQ-VNDKGEDNDNDKYHVAYSWVVVKLRDINDNKPQFERPNIEVSVYENADVGKT  1892

Query  127   VMILNATDADEPNHLNSKIAFKIVSQEPAGTPMFLLSRNTGEVRTLTNSLDREQASSYRL  186
             +    ATD D+     SK+++ I   + +       S N     T+  +LDRE    +++
Sbjct  1893  LETFKATDPDQGG--KSKVSYAI---DRSSDRQRQFSINQEGTVTIQRNLDREVTPRHQV  1947

Query  187   VVSGADKDGEGLSTQCECNIKVKDVNDNFPMFRDSQYSARIEENILSSELLRFQVTDLDE  246
              +   D      +      + V+D+NDN P F    Y   + E++   +++    TD D+
Sbjct  1948  KILAIDDGIPPKTATATLTVIVQDINDNPPKFL-KDYRPVLPEHVPPRKVVEILATDDDD  2006

Query  247   EYTDNWLAVYFFTSGNEGNW----FEIQTDPRTNEG----ILKVVKALDYEQLQSVKLSI  298
                 N     F        +    F+++ D +   G    I+ +++  D EQ  +  + I
Sbjct  2007  RSKSNGPPFQFRLDPGADDIIRASFKVEQDQKGANGDGMAIVSSLRSFDREQQKEYLIPI  2066

Query  299   AVKNKA  304
              +K+
Sbjct  2067  VIKDHG  2072

 Score = 52.8 bits (125),  Expect = 2e-05, Method: Compositional matrix adjust.
 Identities = 97/398 (24%), Positives = 163/398 (41%), Gaps = 54/398 (14%)

Query  106   PVFSQQIFMGEIEENSASNSLVMILNATDADEPNHLNSKIAFKIVSQ-EPAGTPMFLLSR  164
             P F    +  EI EN   +S ++ + A      +  + +I + + +Q + AGT  F +
Sbjct  663   PQFYMPSYEAEIPENQKKDSDIISVKAK-----SFADREIRYTLKAQGQGAGT--FNIGP  715

Query  165   NTGEVRTLTNSLDRE---QASSYRLVVSGADKDGEGLSTQCECNIKVKDVNDNFPMFRDS  221
              +G V+ L   LD E   Q   Y LVV+ A +D  G ST  E  I+V DVNDN P F
Sbjct  716   TSGIVK-LAKELDFEDLRQPHVYSLVVT-ATEDSGGFSTSVELTIRVTDVNDNAPKFELP  773

Query  222   QYSAR-IEENI-LSSELLRFQVTDLDEEYTDNWLAVYFFTSGNEGNWFEIQTDPRTNEGI  279
              Y A  ++E+I L + +L+ +  D D         + +  S +    F + ++     GI
Sbjct  774   DYQAHNVDEDIPLGTSILKVKAMDADSGAN---AEIEYLVSDDH---FAVDSN-----GI  822

Query  280   LKVVKALDYEQLQS-VKLSIAVKNKAEFHQSVISRYRVQSTPVTIQVINVREGIAFRPAS  338
             +   K LD +   +  +  +  K+K E  ++  +  RV +           E   F
Sbjct  823   IVNNKQLDADNNNAYYEFVVTAKDKGEPAKTGTATVRVYTKNKN------DEEPKFSQQV  876
```

```
Query  339   KTFTVQKGISSKKLVDYILGTYQAIDEDTNKAASNVK--YVMGRNDGGYLMIDSKTAEIK  396
              T  V +      LV  ++         ++K   NV+  +V G    G  +I+  T  I+
Sbjct  877   YTPNVDENAGPNTLVTTVVA--------SDKDGDNVRFGFVGGGTSSGQFVIEEITGVIR  928

Query  397   FVK---NMNRDSTFI-VNKTITAEVLAIDEYTGKTSTGTVYYRVPDFNDN------CPTA  446
                   +++RD   + V            + T  TST  V V + D NDN      C T
Sbjct  929   LHSKAISLDRDKYELNVTAMDDGSCCVNGDATIHTSTAVVVVFITDVNDNKPVFKDCGTY  988

Query  447   VLE-KDAVCSSSPSVVVSARTLNNRYTGPYTFALEDQP  483
               + ++    + SP + V A   +    G   +++  QP
Sbjct  989   YPKVEEGAPNGSPVIKVQATDEDKGVNGQVKYSIVQQP  1026

 Score = 44.7 bits (104),  Expect = 0.008, Method: Compositional matrix adjust.
 Identities = 48/199 (24%), Positives = 73/199 (37%), Gaps = 19/199 (10%)

Query  27    DYQATQKITYRISGVGIDQPPFGIFVVDKNTGDINITAIVDREETPSFLITCRALNAQGL  86
             D     K++Y I      Q  F I       G + I   +DRE TP    +   A++
Sbjct  1902  DQGGKSKVSYAIDRSSDRQRQFSI----NQEGTVTIQRNLDREVTPRHQVKILAIDDGIP  1957

Query  87    DVEKPLILTVKILDINDNPPVFSQQIFMGEIEENSASNSLVMILNATDADEPNHLNSKIA  146
                    LTV + DINDNPP F +  +   + E+     +V IL   D D
Sbjct  1958  PKTATATLTVIVQDINDNPPKFLKD-YRPVLPEHVPPRKVVEILATDDDDRSKSNGPPFQ  2016

Query  147   FKIVSQEPAGTPMFLLSRNTGEVRTLTN-----------SLDREQASSYRLVVSGADKDG  195
             F++   +P    +   S    + +   N           S DREQ   Y + +   D
Sbjct  2017  FRL---DPGADDIIRASFKVEQDQKGANGDGMAIVSSLRSFDREQQKEYLIPIVIKDHGN  2073

Query  196   EGLSTQCECNIKVKDVNDN  214
               ++      + + DVNDN
Sbjct  2074  PAMTGTSTLTVVIGDVNDN  2092

 Score = 40.8 bits (94),  Expect = 0.11, Method: Compositional matrix adjust.
 Identities = 55/242 (23%), Positives = 102/242 (42%), Gaps = 22/242 (9%)

Query  4    KFAKPCREGE--DNSKRNP-IAKITSDYQATQKITYRISGVGIDQPPFGIFVVDKNTGDI  60
            +F  P  E E  +N K++  I  + +   A ++I Y +   G      G F +   +G +
Sbjct  664  QFYMPSYEAEIPENQKKDSDIISVKAKSFADREIRYTLKAQGQGA---GTFNIGPTSGIV  720

Query  61   NITAIVDREE--TPSF--LITCRALNAQGLDVEKPLILTVKILDINDNPPVFSQQIFMGE  116
             +   +D E+   P    L+     ++ G      + LT+++ D+NDN P F    +
Sbjct  721  KLAKELDFEDLRQPHVYSLVVTATEDSGGFSTS--VELTIRVTDVNDNAPKFELPDYQAH  778

Query  117  -IEENSASNSLVMILNATDADEPNHLNSKIAFKIVSQEPAGTPMFLLSRNTGEVRTLTNS  175
             ++E+    + ++ + A DAD  +  N++I + +VS +      F +  N   V
Sbjct  779  NVDEDIPLGTSILKVKAMDAD--SGANAEIEY-LVSDD-----HFAVDSNGIIVNNKQLD  830

Query  176  LDREQASSYRLVVSGADKDGEGLSTQCECNIKVKDVNDNFPMFRDSQYSARIEENILSSE  235
             D   A  Y  VV+  DK      +      +  K+ ND  P F    Y+  ++EN   +
Sbjct  831  ADNNNA-YYEFVVTAKDKGEPAKTGTATVRVYTKNKNDEEPKFSQQVYTPNVDENAGPNT  889

Query  236  LL  237
            L+
Sbjct  890  LV  891

 Score = 39.7 bits (91),  Expect = 0.23, Method: Compositional matrix adjust.
 Identities = 89/389 (23%), Positives = 149/389 (38%), Gaps = 62/389 (16%)

Query  75   LITCRALNAQGLDVEKPLILTVKILDINDNPPVFSQQIF--MGEIEENSASNSLVMILNA  132
            +IT +  NA GL        + + + D+ND PP F  +       ++ N+  N+ V  L A
Sbjct  525  IITNQGHNA-GLKYTDNQRVIIHVKDVNDEPPYFINRPLPMQAVVQLNAPPNTPVFTLQA  583

Query  133  TDADEPNHLNSKIAFKIVSQEPAGTPMFLLSRNTGEVRTLTNSLDREQASSYRLVVSGAD  192
             D D     +  I + IV     G   F +   +G VRT    L  +    Y L V   D
Sbjct  584  RDPD----TDHNIHYFIVRDRTGG--RFEVDERSGVVRTRGTDL-FQLDMEYVLYVKAED  636

Query  193  KDGE----GLSTQCECNIKVKDVNDNFPMFRDSQYSARIEEN-ILSSELLRFQVTDLDEE  247
            ++G+       +  E  + +       P F    Y A I EN    S+++  +     +
Sbjct  637  QNGKVDDRRFQSTPEERLSIVG-GKRAPQFYMPSYEAEIPENQKKDSDIISVKA----KS  691
```

```
Query  248  YTDNWLAVYFFTSGNEGNWFEIQTDPRTNEGILKVVKALDYEQLQSVKLS----IAVKNK  303
            + D  +       G     F I        GI+K+ K LD+E L+   +      A ++
Sbjct  692  FADREIRYTLKAQGQGAGTFNIGP----TSGIVKLAKELDFEDLRQPHVYSLVVTATEDS  747

Query  304  AEFHQSVISRYRVQSTPVTIQVINVREGIAFRPASKTFTVQKGISSKKLVDYILGT----  359
              F  SV          +TI+V +V +       +  F +    +     D  LGT
Sbjct  748  GGFSTSV---------ELTIRVTDVNDN------APKFELPDYQAHNVDEDIPLGTSILK  792

Query  360  YQAIDEDTNKAASNVKYVMGRN----DGGYLMIDSKTAEIKFVKNMNRDSTFIVNKTITA  415
             +A+D D+  A + ++Y++  +    D   +++++K  +     N N    F+V
Sbjct  793  VKAMDADSG-ANAEIEYLVSDDHFAVDSNGIIVNNKQLD---ADNNNAYYEFVVTAKDKG  848

Query  416  EVLAIDEYTGKTSTGTVYVRVPDFNDNCP  444
            E         KT T TV V   + ND  P
Sbjct  849  E-------PAKTGTATVRVYTKNKNDEEP  870

>AALB20_033171.P66516 | transcript=AALB20_033171.R66516 | gene=AALB20_033171
| organism=Anopheles_albimanus_STECLA_2020 |
gene_product=neural-cadherin, transcript variant X38 |
transcript_product=neural-cadherin,
transcript variant X33 | location=CM024292:75037560-75208393(+)
| protein_length=3123 | sequence_SO=chromosome
| SO=protein_coding_gene | is_pseudo=false
Length=3123

 Score = 147 bits (371),  Expect = 2e-35, Method: Compositional matrix adjust.
 Identities = 128/446 (29%), Positives = 217/446 (49%), Gaps = 36/446 (8%)

Query  20    PIAKITS---DYQATQKITYRISGVGID--QPPFGIFVVDKNTGDINITAIVDREET---  71
             PI ++T+   D    Q I Y ++G GID   P    F +++ TG+I +   +DR++
Sbjct  1563  PIMQVTATDGDKDRPQNIVYFLTGQGIDPDNPANSKFDINRTTGEIFVLKPLDRDQPNGR  1622

Query  72    PSFLITCRALNAQGLDVEKPLILTVKILDINDNPPVFSQQIFMGEIEENSASNSLVMILN  131
             P +  T  A +  G  +     + V + DINDN P+F Q ++ G + EN  +  +VM +
Sbjct  1623  PQWRFTVFAQDEGGEGLVGYADVQVNLKDINDNAPIFPQGVYFGNVTENGTAGMVVMTMT  1682

Query  132   ATDADEPNH-LNSKIAFKI---VSQEPAGTPMFLLSRNTGEVRTLTNSLDREQASSYRLV  187
             A D D+PN   N+K+ + I   V +E  G+P+F +  +TG ++T    LDRE+   Y +
Sbjct  1683  AVDYDDPNEGTNAKLIYSIEKNVIEEETGSPIFEIEADTGVIKTAVCCLDRERTPDYSIQ  1742

Query  188   VSGADKDGEGLSTQCECNIKVKDVNDNFPMFRDSQYSARIEENILSS--ELLRFQVTDLD  245
             V     DG GL      +I+VKD+ND  P F   ++   ++E   ++  E+    VT  D
Sbjct  1743  VVAM--DGGGLKGTGTASIRVKDINDMPPQFTKDEWFTEVDETDGTNLPEMPILTVTVHD  1800

Query  246   EEYTDNWLAVYFFTSGNEGNWFEIQTDPRTNEGI--LKVVKALDYE---QLQSVKLSIAV  300
             E+ T+ +       SG   + F +    R N+G   LK+V+ LDYE   Q    +  I V
Sbjct  1801  EDETNKFQYKVIDNSGYGADKFTMV---RNNDGTGSLKIVQPLDYEDPLQSNGFRFRIQV  1857

Query  301   KNKAEFHQSVISRYRVQSTPVTIQVINVREGIAFRPASKTFTVQKGISSKKLVDYILGTY  360
              +K E + +   +Y V  + V +++ ++ +    +P  +   ++  +     V   L T+
Sbjct  1858  NDKGEDNDN--DKYHVAYSWVVVKLRDINDN---KPQFERPNIEVSVYENADVGKTLETF  1912

Query  361   QAIDEDTNKAASNVKYVMGR-NDGGYLMIDSKTAEIKFVKNMNRDSTFIVNKTITAEVLA  419
             +A D D     S V Y + R +D       ++   +   +N++R+    V      ++LA
Sbjct  1913  KATDPDQG-GKSKVSYAIDRSSDRQRQFSINQEGTVTIQRNLDRE----VTPRHQVKILA  1967

Query  420   IDE-YTGKTSTGTVYVRVPDFNDNCP  444
             ID+    KT+T T+ V V D NDN P
Sbjct  1968  IDDGIPPKTATATLTVIVQDINDNPP  1993

 Score = 79.3 bits (194),  Expect = 2e-13, Method: Compositional matrix adjust.
 Identities = 104/419 (25%), Positives = 173/419 (41%), Gaps = 51/419 (12%)

Query  50    IFVVDKNTGDINITAIVDREETPSFLITCRAL----------NAQGLDVEKPLILTVKIL  99
              F ++  +G+I    + DRE+  ++ +   A           N Q   V K   + + I
Sbjct  1378  FFEINLQSGEIFTKVVFDREKQGAYALEVEARDGAPSARPNSNQQPNSVTK--FIRIGIA  1435

Query  100   DINDNPPVFSQQIFMGEIEENSASNSLVMILNATDADEPNHLNSKIAFKIVSQEPAGTPM  159
```

```
                 D NDNPP F + ++  E++EN      V+ + A D DE    +S+I ++I S    G
Sbjct  1436  DKNDNPPYFDKALYEAEVDENEDIQHTVLTVTAKDHDE----SSRIRYEITSGNIGGA--  1489

Query  160   FLLSRNTGEVRTLTNSLDREQASSYRLVVSGADKDGEGLSTQCECNIKVKDVNDNFPMFR  219
             F +   TG +  +  +LD E    Y L +  +D   E  +       I VKDVND  P F
Sbjct  1490  FAVKNMTGAI-YVAGALDYETRKRYHLTLVASDSLNENATI---IEIHVKDVNDMPPTFS  1545

Query  220   DSQYSARI--EENILSSELLRFQVTDLDEEYTDNWLAVYFFTSG--NEGNWFEIQTDPRT  275
                Y A    E+ +    +++   TD D++   N   VYF T    +  N    + D
Sbjct  1546  QQIYEATQDEEQTVTDRPIMQVTATDGDKDRPQN--IVYFLTGQGIDPDNPANSKFDINR  1603

Query  276   NEGILKVVKALDYEQLQS-VKLSIAVKNKAEFHQSVISRYRVQSTPVTIQVINVREGIAF  334
               G + V+K LD +Q     +    V  + E  + ++    VQ     + + ++ +
Sbjct  1604  TTGEIFVLKPLDRDQPNGRPQWRFTVFAQDEGGEGLVGYADVQ-----VNLKDINDNAPI  1658

Query  335   RPASKTFFTVQKGISSKKLVDYILGTYQAIDEDTNKAASNVK--YVMGRN------DGGYL  386
              P    F     ++        ++ T  A+D D     +N K  Y + +N
Sbjct  1659  FPQGVYF---GNVTENGTAGMVVMTMTAVDYDDPNEGTNAKLIYSIEKNVIEEETGSPIF  1715

Query  387   MIDSKTAEIK-FVKNMNRDSTFIVNKTITAEVLAIDEYTGKTSTGTVYVRVPDFNDNCP  444
              I++ T  IK  V  ++R+ T        + +V+A+D   G   TGT  +RV D ND  P
Sbjct  1716  EIEADTGVIKTAVCCLDRERT----PDYSIQVVAMDG-GGLKGTGTASIRVKDINDMPP  1769

 Score = 73.9 bits (180),  Expect = 6e-12, Method: Compositional matrix adjust.
 Identities = 90/362 (25%), Positives = 166/362 (46%), Gaps = 37/362 (10%)

Query  51    FVVDKNTGDINITAIVDRE-ETPSFL-ITCRALNAQGLDVEKPLILTVKILDINDNPPVF  108
             F VD+ TG+++   + DRE +   F+ +T +A +     +E     TV+I D+NDNPP+F
Sbjct  1047  FTVDEETGEVSTNKVFDREGDDGKFVSVTVKATDQGEPSLEGVCSFTVEITDVNDNPPLF  1106

Query  109   SQQIFMGEIEENSASNSLVMILNATDADEPNHLNSKIAFKIVS-QEPAGTPMFLLSRNTG  167
              +Q ++  ++++++  + ++ ++A+D D  N  N  I + + +   P     F +   +G
Sbjct  1107  DRQKYVENVKQDASIGTNILRVSASDEDADN--NGAIVYTLSAPHSPNDLEYFEIQPESG  1164

Query  168   EVRTLTNSLDREQASSYRLVVSGADKDGEGLSTQCECNIKVKDVNDNFPMFRDSQYSARI  227
              +  L   LDRE   +Y+L      DK    LS   E  I V D  +N P++ ++     I
Sbjct  1165  WI-VLKKPLDRE---TYKLEAMAQDKGYPPLSRTVEVQIDVVDRANNPPVWDNTGVYGPI  1220

Query  228   --EENI-LSSELLRFQVTDLDEEYTDNWLAVYFFTSG-----NEGNWFEIQTDPRTNE--  277
               +EN+ +  +++  + +   E    N    Y    G     N+ + F +Q  P   +
Sbjct  1221  YVKENMPVGGKVVSIKASSGIE---GNPTVFYRLMPGSTAQTNKLHTFYLQQRPDNGDTW  1277

Query  278   GILKVVKALDYEQLQSVKLSIAVKNKAEFHQSVISRYRVQSTPVTIQVINVREGIAFRPA  337
               +KV   LD+E ++   L+I V+N          ++        V IQ+ +V + I
Sbjct  1278  ADIKVNHPLDFETIKEYNLTIRVENNG-------AQQLASEATVYIQLEDVNDEIPLFTE  1330

Query  338   SKTFTVQKGISSKKLVDYILGTYQAIDEDTNKAASNVKYVM---GRNDG-GYLMIDSKTA  393
              +  TV +G       V  +     AID+D      + V Y +    RN+G  +  I+ ++
Sbjct  1331  REQETVLEGEPIGTKVTQV----NAIDKDGTFPNNQVYYYIVDSPRNEGKDFFEINLQSG  1386

Query  394   EI  395
             EI
Sbjct  1387  EI  1388

 Score = 55.8 bits (133),  Expect = 3e-06, Method: Compositional matrix adjust.
 Identities = 49/158 (31%), Positives = 78/158 (49%), Gaps = 24/158 (15%)

Query  102   NDNPPVFSQQIFMGEIEENSASNSLVMILNATDADEPNHLNSKIAFKIVSQEPAGTP--M  159
             ND  P FSQQ++    ++EN+  N+LV  + A+D D  N     + F  V     GT
Sbjct  880   NDEEPKFSQQVYTPNVDENAGPNTLVTTVVASDKDGDN-----VRFGFVG---GGTSSGQ  931

Query  160   FLLSRNTGEVRTLTN--SLDREQASSYRLVVSGADK-------DGEGLSTQCECNIKVKD  210
             F++   TG +R  +   SLDR++    Y L V+  D         D    ++       + D
Sbjct  932   FVIEEITGVIRLHSKAISLDRDK---YELNVTAMDDGSCCVNGDATIHTSTAVVVVFITD  988

Query  211   VNDNFPMFRD-SQYSARIEENILS-SELLRFQVTDLDE  246
             VNDN P+F+D   Y  ++EE   + S +++ Q TD D+
Sbjct  989   VNDNKPVFKDCGTYYPKVEEGAPNGSPVIKVQATDEDK  1026
```

```
 Score = 53.9 bits (128),  Expect = 1e-05, Method: Compositional matrix adjust.
 Identities = 70/306 (23%), Positives = 126/306 (41%), Gaps = 30/306 (10%)

Query  20    PIAKIT-SDYQATQKITYRI---SGVGIDQPPFGIFVVDKNTGDINITAIVDRE---ETP  72
             PI  +T  D   T K  Y++   SG G D+  F +   +  TG + I   +D E   ++
Sbjct  1792  PILTVTVHDEDETNKFQYKVIDNSGYGADK--FTMVRNNDGTGSLKIVQPLDYEDPLQSN  1849

Query  73    SFLITCRALNAQGLDVEKPLI------LTVKILDINDNPPVFSQQIFMGEIEENSASNSL  126
              F    + +N +G D +          + VK+ DINDN P F +      + EN+
Sbjct  1850  GFRFRIQ-VNDKGEDNDNDKYHVAYSWVVVKLRDINDNKPQFERPNIEVSVYENADVGKT  1908

Query  127   VMILNATDADEPNHLNSKIAFKIVSQEPAGTPMFLLSRNTGEVRTLTNSLDREQASSYRL  186
             +    ATD D+     SK+++ I   + +        S N     T+  +LDRE    +++
Sbjct  1909  LETFKATDPDQGG--KSKVSYAI---DRSSDRQRQFSINQEGTVTIQRNLDREVTPRHQV  1963

Query  187   VVSGADKDGEGLSTQCECNIKVKDVNDNFPMFRDSQYSARIEENILSSELLRFQVTDLDE  246
              +   D      +      + V+D+NDN P F    Y   + E++   +++    TD D+
Sbjct  1964  KILAIDDGIPPKTATATLTVIVQDINDNPPKFL-KDYRPVLPEHVPPRKVVEILATDDDD  2022

Query  247   EYTDNWLAVYFFTSGNEGNW----FEIQTDPRTNEG----ILKVVKALDYEQLQSVKLSI  298
                 N     F       +     F+++ D +   G    I+  +++ D EQ +   + I
Sbjct  2023  RSKSNGPPFQFRLDPGADDIIRASFKVEQDQKGANGDGMAIVSSLRSFDREQQKEYLIPI  2082

Query  299   AVKNKA  304
              +K+
Sbjct  2083  VIKDHG  2088

 Score = 51.6 bits (122),  Expect = 5e-05, Method: Compositional matrix adjust.
 Identities = 96/398 (24%), Positives = 163/398 (41%), Gaps = 54/398 (14%)

Query  106   PVFSQQIFMGEIEENSASNSLVMILNATDADEPNHLNSKIAFKIVSQ-EPAGTPMFLLSR  164
             P F    +  EI EN   +S ++ + A      +  + +I + + +Q + AGT  F +
Sbjct  677   PQFYMPSYEAEIPENQKKDSDIISVKAK-----SFADREIRYTLKAQGQGAGT--FNIGP  729

Query  165   NTGEVRTLTNSLDRE---QASSYRLVVSGADKDGEGLSTQCECNIKVKDVNDNFPMFRDS  221
              +G V+ L   LD E   Q   Y LVV+ A +D  G ST  E  I+V DVNDN P F
Sbjct  730   TSGIVK-LAKELDFEDLRQPHVYSLVVT-ATEDSGGFSTSVELTIRVTDVNDNAPKFELP  787

Query  222   QYSAR-IEENI-LSSELLRFQVTDLDEEYTDNWLAVYFFTSGNEGNWFEIQTDPRTNEGI  279
              Y A  ++E+I + + +L+ +  D D         + +  S +    F + ++     GI
Sbjct  788   DYQAHNVDEDIQIGTSILKVKAMDADSGAN---AEIEYLVSDDH---FAVDSN-----GI  836

Query  280   LKVVKALDYEQLQS-VKLSIAVKNKAEFHQSVISRYRVQSTPVTIQVINVREGIAFRPAS  338
             +   K LD +   +  +  +  K+K E  ++  +  RV +           E   F
Sbjct  837   IVNNKQLDADNNNAYYEFVVTAKDKGEPAKTGTATVRVYTKNKN------DEEPKFSQQV  890

Query  339   KTFTVQKGISSKKLVDYILGTYQAIDEDTNKAASNVK--YVMGRNDGGYLMIDSKTAEIK  396
              T  V +      LV  ++         ++K   NV+  +V G    G  +I+  T  I+
Sbjct  891   YTPNVDENAGPNTLVTTVVA--------SDKDGDNVRFGFVGGGTSSGQFVIEEITGVIR  942

Query  397   FVK---NMNRDSTFI-VNKTITAEVLAIDEYTGKTSTGTVYVRVPDFNDN------CPTA  446
                   +++RD   + V            + T  TST  V V + D NDN      C T
Sbjct  943   LHSKAISLDRDKYELNVTAMDDGSCCVNGDATIHTSTAVVVVFITDVNDNKPVFKDCGTY  1002

Query  447   VLE-KDAVCSSSPSVVVSARTLNNRYTGPYTFALEDQP  483
               + ++   + SP + V A    +    G   +++  QP
Sbjct  1003  YPKVEEGAPNGSPVIKVQATDEDKGVNGQVKYSIVQQP  1040

 Score = 44.3 bits (103),  Expect = 0.009, Method: Compositional matrix adjust.
 Identities = 48/199 (24%), Positives = 73/199 (37%), Gaps = 19/199 (10%)

Query  27    DYQATQKITYRISGVGIDQPPFGIFVVDKNTGDINITAIVDREETPSFLITCRALNAQGL  86
             D     K++Y I      Q  F I       G + I   +DRE TP   +   A++
Sbjct  1918  DQGGKSKVSYAIDRSSDRQRQFSI----NQEGTVTIQRNLDREVTPRHQVKILAIDDGIP  1973

Query  87    DVEKPLILTVKILDINDNPPVFSQQIFMGEIEENSASNSLVMILNATDADEPNHLNSKIA  146
                    LTV + DINDNPP F +  +   + E+     +V IL   D D
Sbjct  1974  PKTATATLTVIVQDINDNPPKFLKD-YRPVLPEHVPPRKVVEILATDDDDRSKSNGPPFQ  2032
```

```
Query  147   FKIVSQEPAGTPMFLLSRNTGEVRTLTN-----------SLDREQASSYRLVVSGADKDG  195
             F++   +P    +   S    + +   N           S DREQ   Y + +   D
Sbjct  2033  FRL---DPGADDIIRASFKVEQDQKGANGDGMAIVSSLRSFDREQQKEYLIPIVIKDHGN  2089

Query  196   EGLSTQCECNIKVKDVNDN  214
               ++      + + DVNDN
Sbjct  2090  PAMTGTSTLTVVIGDVNDN  2108

 Score = 40.8 bits (94),  Expect = 0.12, Method: Compositional matrix adjust.
 Identities = 55/242 (23%), Positives = 102/242 (42%), Gaps = 22/242 (9%)

Query  4    KFAKPCREGE--DNSKRNP-IAKITSDYQATQKITYRISGVGIDQPPFGIFVVDKNTGDI  60
            +F  P  E E  +N K++  I  + +   A ++I Y +   G      G F +   +G +
Sbjct  678  QFYMPSYEAEIPENQKKDSDIISVKAKSFADREIRYTLKAQGQGA---GTFNIGPTSGIV  734

Query  61   NITAIVDREE--TPSF--LITCRALNAQGLDVEKPLILTVKILDINDNPPVFSQQIFMGE  116
             +   +D E+   P    L+     ++ G      + LT+++ D+NDN P F    +
Sbjct  735  KLAKELDFEDLRQPHVYSLVVTATEDSGGFSTS--VELTIRVTDVNDNAPKFELPDYQAH  792

Query  117  -IEENSASNSLVMILNATDADEPNHLNSKIAFKIVSQEPAGTPMFLLSRNTGEVRTLTNS  175
             ++E+    + ++ + A DAD  +  N++I + +VS +      F +  N   V
Sbjct  793  NVDEDIQIGTSILKVKAMDAD--SGANAEIEY-LVSDD-----HFAVDSNGIIVNNKQLD  844

Query  176  LDREQASSYRLVVSGADKDGEGLSTQCECNIKVKDVNDNFPMFRDSQYSARIEENILSSE  235
             D   A  Y  VV+  DK     +      +  K+ ND  P F    Y+  ++EN   +
Sbjct  845  ADNNNA-YYEFVVTAKDKGEPAKTGTATVRVYTKNKNDEEPKFSQQVYTPNVDENAGPNT  903

Query  236  LL  237
            L+
Sbjct  904  LV  905

 Score = 39.3 bits (90),  Expect = 0.33, Method: Compositional matrix adjust.
 Identities = 55/225 (24%), Positives = 88/225 (39%), Gaps = 24/225 (11%)

Query  75   LITCRALNAQGLDVEKPLILTVKILDINDNPPVFSQQIF--MGEIEENSASNSLVMILNA  132
            +IT +  NA GL       + + + D+ND PP F  +       ++ N+  N+ V  L A
Sbjct  539  IITNQGHNA-GLKYTDNQRVIIHVKDVNDEPPYFINRPLPMQAVVQLNAPPNTPVFTLQA  597

Query  133  TDADEPNHLNSKIAFKIVSQEPAGTPMFLLSRNTGEVRTLTNSLDREQASSYRLVVSGAD  192
             D D     +  I + IV     G   F +   +G VRT    L  +    Y L V   D
Sbjct  598  RDPD----TDHNIHYFIVRDRTGG--RFEVDERSGVVRTRGTDL-FQLDMEYVLYVKAED  650

Query  193  KDGE----GLSTQCECNIKVKDVNDNFPMFRDSQYSARIEEN-ILSSELLRFQVTDLDEE  247
            ++G+        +  E  + +       P F    Y A I EN    S+++  +     +
Sbjct  651  QNGKVDDRRFQSTPEERLSIVG-GKRAPQFYMPSYEAEIPENQKKDSDIISVKA----KS  705

Query  248  YTDNWLAVYFFTSGNEGNWFEIQTDPRTNEGILKVVKALDYEQLQ  292
            + D  +       G     F I        GI+K+ K LD+E L+
Sbjct  706  FADREIRYTLKAQGQGAGTFNIGP----TSGIVKLAKELDFEDLR  746

>AMEM21_003341.P6430 | transcript=AMEM21_003341.R6430 | gene=AMEM21_003341
| organism=Anopheles_merus_MAF_2021 | gene_product=neural-cadherin,
transcript variant X1 | transcript_product=neural-cadherin,
transcript variant X11 | location=CM029766:40289848-40498951(+)
| protein_length=3102 | sequence_SO=chromosome
| SO=protein_coding_gene | is_pseudo=false
Length=3102

 Score = 147 bits (371),  Expect = 2e-35, Method: Compositional matrix adjust.
 Identities = 128/446 (29%), Positives = 217/446 (49%), Gaps = 36/446 (8%)

Query  20    PIAKITS---DYQATQKITYRISGVGID--QPPFGIFVVDKNTGDINITAIVDREET---  71
             PI ++T+   D    Q I Y ++G GID   P    F +++ TG+I +   +DR++
Sbjct  1543  PIMQVTATDGDKDRPQNIVYFLTGQGIDPDNPANSKFDINRTTGEIFVLKPLDRDQPNGR  1602

Query  72    PSFLITCRALNAQGLDVEKPLILTVKILDINDNPPVFSQQIFMGEIEENSASNSLVMILN  131
             P +  T  A +  G  +     + V + DINDN P+F Q ++ G + EN  +  +VM +
```

```
Sbjct  1603  PQWRFTVFAQDEGGEGLVGYADVQVNLKDINDNAPIFPQGVYFGNVTENGTAGMVVMTMT  1662

Query  132   ATDADEPNH-LNSKIAFKI---VSQEPAGTPMFLLSRNTGEVRTLTNSLDREQASSYRLV  187
             A D D+PN   N+K+ + I   V +E  G+P+F +  +TG ++T    LDRE+   Y +
Sbjct  1663  AVDYDDPNEGTNAKLIYSIEKNVIEEETGSPIFEIEADTGVIKTAVCCLDRERTPDYSIQ  1722

Query  188   VSGADKDGEGLSTQCECNIKVKDVNDNFPMFRDSQYSARIEENILSS--ELLRFQVTDLD  245
             V     DG GL      +I+VKD+ND  P F   ++   ++E   ++  E+    VT  D
Sbjct  1723  VVAM--DGGGLKGTGTASIRVKDINDMPPQFTKDEWFTEVDETDGTNLPEMPILTVTVHD  1780

Query  246   EEYTDNWLAVYFFTSGNEGNWFEIQTDPRTNEGI--LKVVKALDYE---QLQSVKLSIAV  300
             E+ T+ +        SG   + F +    R N+G   LK+V+ LDYE   Q    +  I V
Sbjct  1781  EDETNKFQYKVIDNSGYGADKFTMV---RNNDGTGSLKIVQPLDYEDPLQSNGFRFRIQV  1837

Query  301   KNKAEFHQSVISRYRVQSTPVTIQVINVREGIAFRPASKTFTVQKGISSKKLVDYILGTY  360
              +K E + +   +Y V  + V +++ ++ +    +P  +   ++  +      V   L T+
Sbjct  1838  NDKGEDNDN--DKYHVAYSWVVVKLRDINDN---KPQFERPNIEVSVYENADVGKTLETF  1892

Query  361   QAIDEDTNKAASNVKYVMGR-NDGGYLMIDSKTAEIKFVKNMNRDSTFIVNKTITAEVLA  419
             +A D D     S V Y + R +D        ++    +   +N++R+    V       ++LA
Sbjct  1893  KATDPDQG-GKSKVSYAIDRSSDRQRQFSINQEGTVTIQRNLDRE----VTPRHQVKILA  1947

Query  420   IDE-YTGKTSTGTVYVRVPDFNDNCP  444
             ID+    KT+T T+ V V D NDN P
Sbjct  1948  IDDGIPPKTATATLTVIVQDINDNPP  1973

 Score = 76.3 bits (186),  Expect = 1e-12, Method: Compositional matrix adjust.
 Identities = 103/418 (25%), Positives = 174/418 (42%), Gaps = 50/418 (12%)

Query  50    IFVVDKNTGDINITAIVDREETPSFLITCRAL----------NAQGLDVEKPLILTVKIL  99
              F ++  +G+I    + DRE+  ++ +   A           N Q   V K   + + I
Sbjct  1359  FFEINLQSGEIFTKVVFDREKQGAYALEVEARDGAPSARPNSNQQPNSVTK--FIRIGIA  1416

Query  100   DINDNPPVFSQQIFMGEIEENSASNSLVMILNATDADEPNHLNSKIAFKIVSQEPAGTPM  159
             D NDNPP F + ++  E++EN       V+ + A D DE    +S+I ++I S    G
Sbjct  1417  DKNDNPPYFDKALYEAEVDENEDIQHTVLTVTAKDHDE----SSRIRYEITSGNIGGA--  1470

Query  160   FLLSRNTGEVRTLTNSLDREQASSYRLVVSGADKDGEGLSTQCECNIKVKDVNDNFPMFR  219
             F +   TG +  +  +LD E    Y L +  +D   E  +       I VKDVND  P F
Sbjct  1471  FAVKNMTGAI-YVAGALDYETRKRYYLTLVASDSLNENETIIV---INVKDVNDMPPSFP  1526

Query  220   DSQYSARIEENI-LSSELLRFQVTDLDEEYTDNWLAVYFFTSG--NEGNWFEIQTDPRTN  276
                Y   ++E + +   +++   TD D++   N   VYF T    +  N    + D
Sbjct  1527  QQVYERTMDEELAVPFPIMQVTATDGDKDRPQN--IVYFLTGQGIDPDNPANSKFDINRT  1584

Query  277   EGILKVVKALDYEQLQS-VKLSIAVKNKAEFHQSVISRYRVQSTPVTIQVINVREGIAFR  335
              G + V+K LD +Q     +    V  + E  + ++    VQ     + + ++ +
Sbjct  1585  TGEIFVLKPLDRDQPNGRPQWRFTVFAQDEGGEGLVGYADVQ-----VNLKDINDNAPIF  1639

Query  336   PASKTFTVQKGISSKKLVDYILGTYQAIDEDTNKAASNVK--YVMGRN------DGGYLM  387
             P    F     ++        ++ T  A+D D     +N K  Y + +N
Sbjct  1640  PQGVYF---GNVTENGTAGMVVMTMTAVDYDDPNEGTNAKLIYSIEKNVIEEETGSPIFE  1696

Query  388   IDSKTAEIK-FVKNMNRDSTFIVNKTITAEVLAIDEYTGKTSTGTVYVRVPDFNDNCP  444
             I++ T  IK  V  ++R+ T       + +V+A+D   G   TGT  +RV D ND  P
Sbjct  1697  IEADTGVIKTAVCCLDRERT----PDYSIQVVAMDG-GGLKGTGTASIRVKDINDMPP  1749

 Score = 75.9 bits (185),  Expect = 2e-12, Method: Compositional matrix adjust.
 Identities = 106/426 (25%), Positives = 181/426 (42%), Gaps = 51/426 (12%)

Query  53    VDKNTGDINITAIVDREETPSFLITCRALNAQGLDVEKPLILTVKILDINDNPPVFSQ-Q  111
             +D++  ++N+TA+ D          +C  +N           ++ V I D+NDN PVF
Sbjct  928   LDRDKYELNVTAMDDG--------SC-CVNGDATIHTSTAVVVVFITDVNDNKPVFKDCG  978

Query  112   IFMGEIEENSASNSLVMILNATDADEPNHLNSKIAFKIVSQEPAGTPMFLLSRNTGEVRT  171
              +  ++EE + + S V+ + ATD D+   +N ++ + IV Q        F +   TGEV T
Sbjct  979   TYYPKVEEGAPNGSPVIKVQATDEDK--GVNGQVKYSIVQQPNQKGTKFTVDEETGEVST  1036

Query  172   LTNSLDRE--QASSYRLVVSGADKDGEGLSTQCECNIKVKDVNDNFPMFRDSQYSARIEE  229
```

```
                  DRE        + V   D+    L   C   +++ DVNDN P+F   +Y   +++
Sbjct  1037  -NKVFDREGDDGKFVSVTVKATDQGEPSLEGVCSFTVEITDVNDNPPLFDRQKYVENVKQ  1095

Query  230   NI-LSSELLRFQVTDLDEEYTDNWLAVYFFT---SGNEGNWFEIQTDPRTNEGILKVVKA  285
             +  + + +LR  V+  DE+  +N   VY  +   S N+  +FEIQ  P +   +LK  K
Sbjct  1096  DASIGTNILR--VSASDEDADNNGAIVYTLSAPHSPNDLEYFEIQ--PESGWIVLK--KP  1149

Query  286   LD--YEQLQSVKLSIAVKNK------AEFHQSVISRYRVQSTPVTIQV----INVREGIA  333
             LD  Y + +  +L +   ++      A+    +    R    P+  +     I++RE +
Sbjct  1150  LDGCYSR-ERYRLKVYASDRGDPPSTADVDVELDVVDRNNKPPIWDRTIYGPIHIRENVT  1208

Query  334   FRPASKTFTVQKGISSKKLVDYILGTYQAIDEDTNKAASNVKYVMGRNDGGYLMIDSKTA  393
                   +     GI     V Y L         TNK   +  Y+  R D G        A
Sbjct  1209  VGTVVTSVKASSGIEGNPTVFYRL--MPGSTAQTNKL--HTFYLQQRPDNG-----DTWA  1259

Query  394   EIKFVKNMNRDSTFIVNKTITAEVLAIDEYTGKTSTGTVYVRVPDFNDNCPTAV-LEKDA  452
             +IK    ++ ++    N T+  E     +     S  TVY+++ D ND  P     E++
Sbjct  1260  DIKVNHPLDFETIKEYNLTVRVENNGAQQL---ASEATVYIQLEDVNDEIPLFTEREQET  1316

Query  453   VCSSSP  458
             V    P
Sbjct  1317  VLEGEP  1322

 Score = 60.8 bits (146),  Expect = 9e-08, Method: Compositional matrix adjust.
 Identities = 83/362 (23%), Positives = 152/362 (42%), Gaps = 34/362 (9%)

Query  51    FVVDKNTGDINITAIVDRE-ETPSFL-ITCRALNAQGLDVEKPLILTVKILDINDNPPVF  108
             F VD+ TG+++   + DRE +   F+ +T +A +     +E     TV+I D+NDNPP+F
Sbjct  1025  FTVDEETGEVSTNKVFDREGDDGKFVSVTVKATDQGEPSLEGVCSFTVEITDVNDNPPLF  1084

Query  109   SQQIFMGEIEENSASNSLVMILNATDADEPNHLNSKIAFKIVS-QEPAGTPMFLLSRNTG  167
              +Q ++  ++++++  + ++ ++A+D D  N  N  I + + +   P     F +   +G
Sbjct  1085  DRQKYVENVKQDASIGTNILRVSASDEDADN--NGAIVYTLSAPHSPNDLEYFEIQPESG  1142

Query  168   EV---RTLTNSLDREQASSYRLVVSGADKDGEGLSTQCECNIKVKDVNDNFPMFRDSQYS  224
              +   + L     RE+   YRL V  +D+ G+  ST            +N P   D
Sbjct  1143  WIVLKKPLDGCYSRER---YRLKVYASDR-GDPPSTADVDVELDVVDRNNKPPIWDRTIY  1198

Query  225   ARIEENILSSELLRFQVTDLDEEYTDNWLAVYFFTSG-----NEGNWFEIQTDPRTNE--  277
               I      +                N    Y    G     N+ + F +Q  P   +
Sbjct  1199  GPIHIRENVTVGTVVTSVKASSGIEGNPTVFYRLMPGSTAQTNKLHTFYLQQRPDNGDTW  1258

Query  278   GILKVVKALDYEQLQSVKLSIAVKNKAEFHQSVISRYRVQSTPVTIQVINVREGIAFRPA  337
               +KV   LD+E ++   L++ V+N         ++       V IQ+ +V + I
Sbjct  1259  ADIKVNHPLDFETIKEYNLTVRVENNG-------AQQLASEATVYIQLEDVNDEIPLFTE  1311

Query  338   SKTFTVQKGISSKKLVDYILGTYQAIDEDTNKAASNVKYVM---GRNDG-GYLMIDSKTA  393
              +  TV +G      V  +     AID+D     + V Y +    RN+G  +  I+ ++
Sbjct  1312  REQETVLEGEPIGTKVTQV----NAIDKDGTFPNNQVYYYIVDSPRNEGKDFFEINLQSG  1367

Query  394   EI  395
             EI
Sbjct  1368  EI  1369

 Score = 55.8 bits (133),  Expect = 3e-06, Method: Compositional matrix adjust.
 Identities = 49/158 (31%), Positives = 78/158 (49%), Gaps = 24/158 (15%)

Query  102   NDNPPVFSQQIFMGEIEENSASNSLVMILNATDADEPNHLNSKIAFKIVSQEPAGTP--M  159
             ND  P FSQQ++   ++EN+  N+LV  + A+D D  N     + F  V     GT
Sbjct  858   NDEEPKFSQQVYTPNVDENAGPNTLVTTVVASDKDGDN-----VRFGFVG---GGTSSGQ  909

Query  160   FLLSRNTGEVRTLTN--SLDREQASSYRLVVSGADK-------DGEGLSTQCECNIKVKD  210
             F++   TG +R  +   SLDR++   Y L V+  D        D    ++     + + D
Sbjct  910   FVIEEITGVIRLHSKAISLDRDK---YELNVTAMDDGSCCVNGDATIHTSTAVVVVFITD  966

Query  211   VNDNFPMFRD-SQYSARIEENILS-SELLRFQVTDLDE  246
             VNDN P+F+D   Y  ++EE   + S +++ Q TD D+
Sbjct  967   VNDNKPVFKDCGTYYPKVEEGAPNGSPVIKVQATDEDK  1004
```

```
 Score = 53.9 bits (128),  Expect = 1e-05, Method: Compositional matrix adjust.
 Identities = 70/306 (23%), Positives = 126/306 (41%), Gaps = 30/306 (10%)

Query  20    PIAKIT-SDYQATQKITYRI---SGVGIDQPPFGIFVVDKNTGDINITAIVDRE---ETP  72
             PI  +T  D   T K  Y++   SG G D+  F +   +  TG + I   +D E   ++
Sbjct  1772  PILTVTVHDEDETNKFQYKVIDNSGYGADK--FTMVRNNDGTGSLKIVQPLDYEDPLQSN  1829

Query  73    SFLITCRALNAQGLDVEKPLI------LTVKILDINDNPPVFSQQIFMGEIEENSASNSL  126
              F    + +N +G D +           + VK+ DINDN P F +       + EN+
Sbjct  1830  GFRFRIQ-VNDKGEDNDNDKYHVAYSWVVVKLRDINDNKPQFERPNIEVSVYENADVGKT  1888

Query  127   VMILNATDADEPNHLNSKIAFKIVSQEPAGTPMFLLSRNTGEVRTLTNSLDREQASSYRL  186
             +    ATD D+     SK+++ I   + +        S N     T+  +LDRE    +++
Sbjct  1889  LETFKATDPDQGG--KSKVSYAI---DRSSDRQRQFSINQEGTVTIQRNLDREVTPRHQV  1943

Query  187   VVSGADKDGEGLSTQCECNIKVKDVNDNFPMFRDSQYSARIEENILSSELLRFQVTDLDE  246
              +   D      +      + V+D+NDN P F    Y   + E++   +++    TD D+
Sbjct  1944  KILAIDDGIPPKTATATLTVIVQDINDNPPKFL-KDYRPVLPEHVPPRKVVEILATDDDD  2002

Query  247   EYTDNWLAVYFFTSGNEGNW----FEIQTDPRTNEG----ILKVVKALDYEQLQSVKLSI  298
                 N     F       +     F+++ D +   G    I+  +++ D EQ +   + I
Sbjct  2003  RSKSNGPPFQFRLDPGADDIIRASFKVEQDQKGANGDGMAIVSSLRSFDREQQKEYLIPI  2062

Query  299   AVKNKA  304
              +K+
Sbjct  2063  VIKDHG  2068

 Score = 52.0 bits (123),  Expect = 4e-05, Method: Compositional matrix adjust.
 Identities = 97/398 (24%), Positives = 163/398 (41%), Gaps = 54/398 (14%)

Query  106   PVFSQQIFMGEIEENSASNSLVMILNATDADEPNHLNSKIAFKIVSQ-EPAGTPMFLLSR  164
             P F    +  EI EN   +S ++ + A       +  + +I + + +Q + AGT  F +
Sbjct  655   PQFYMPSYEAEIPENQKKDSDIISVKAK-----SFADREIRYTLKAQGQGAGT--FNIGP  707

Query  165   NTGEVRTLTNSLDRE---QASSYRLVVSGADKDGEGLSTQCECNIKVKDVNDNFPMFRDS  221
              +G V+ L   LD E   Q   Y LVV+ A +D  G ST  E  I+V DVNDN P F
Sbjct  708   TSGIVK-LAKELDFEDLRQPHVYSLVVT-ATEDSGGFSTSVELTIRVTDVNDNAPKFELP  765

Query  222   QYSAR-IEENI-LSSELLRFQVTDLDEEYTDNWLAVYFFTSGNEGNWFEIQTDPRTNEGI  279
              Y A  ++E+I L + +L+ +  D D          + +  S +    F + ++     GI
Sbjct  766   DYQAHNVDEDIPLGTSILKVKAMDSDSGAN---AEIEYLVSDDH---FAVDSN-----GI  814

Query  280   LKVVKALDYEQLQS-VKLSIAVKNKAEFHQSVISRYRVQSTPVTIQVINVREGIAFRPAS  338
             +   K LD +   +  +  + K+K E  ++  +  RV +            E  F
Sbjct  815   IVNNKQLDADNNNAYYEFVVTAKDKGEPAKTGTATVRVYTKNKN------DEEPKFSQQV  868

Query  339   KTFTVQKGISSKKLVDYILGTYQAIDEDTNKAASNVK--YVMGRNDGGYLMIDSKTAEIK  396
              T  V +       LV  ++          ++K   NV+  +V G    G  +I+  T  I+
Sbjct  869   YTPNVDENAGPNTLVTTVVA--------SDKDGDNVRFGFVGGGTSSGQFVIEEITGVIR  920

Query  397   FVK---NMNRDSTFI-VNKTITAEVLAIDEYTGKTSTGTVYVYRVPDFNDN------CPTA  446
                   +++RD   + V              + T  TST  V V + D NDN      C T
Sbjct  921   LHSKAISLDRDKYELNVTAMDDGSCCVNGDATIHTSTAVVVVFITDVNDNKPVFKDCGTY  980

Query  447   VLE-KDAVCSSSPSVVVSARTLNNRYTGPYTFALEDQP  483
               + ++   + SP + V A   +    G   +++  QP
Sbjct  981   YPKVEEGAPNGSPVIKVQATDEDKGVNGQVKYSIVQQP  1018

 Score = 44.7 bits (104),  Expect = 0.008, Method: Compositional matrix adjust.
 Identities = 48/199 (24%), Positives = 73/199 (37%), Gaps = 19/199 (10%)

Query  27    DYQATQKITYRISGVGIDQPPFGIFVVDKNTGDINITAIVDREETPSFLITCRALNAQGL  86
             D     K++Y I       Q  F I        G + I   +DRE TP   +    A++
Sbjct  1898  DQGGKSKVSYAIDRSSDRQRQFSI----NQEGTVTIQRNLDREVTPRHQVKILAIDDGIP  1953

Query  87    DVEKPLILTVKILDINDNPPVFSQQIFMGEIEENSASNSLVMILNATDADEPNHLNSKIA  146
                    LTV + DINDNPP F +  +    + E+     +V IL   D D
Sbjct  1954  PKTATATLTVIVQDINDNPPKFLKD-YRPVLPEHVPPRKVVEILATDDDDRSKSNGPPFQ  2012
```

```
Query  147   FKIVSQEPAGTPMFLLSRNTGEVRTLTN-----------SLDREQASSYRLVVSGADKDG  195
             F++   +P    +   S    + +   N           S DREQ   Y + +   D
Sbjct  2013  FRL---DPGADDIIRASFKVEQDQKGANGDMAIVSSLRSFDREQQKEYLIPIVIKDHGN  2069

Query  196   EGLSTQCECNIKVKDVNDN  214
               ++      + + DVNDN
Sbjct  2070  PAMTGTSTLTVVIGDVNDN  2088

 Score = 40.0 bits (92),  Expect = 0.21, Method: Compositional matrix adjust.
 Identities = 89/389 (23%), Positives = 149/389 (38%), Gaps = 62/389 (16%)

Query  75   LITCRALNAQGLDVEKPLILTVKILDINDNPPVFSQQIF--MGEIEENSASNSLVMILNA  132
            +IT +  NA GL       + + + D+ND PP F    +     ++ N+  N+ V  L A
Sbjct  517  IITNQGHNA-GLKYTDNQRVIIHVKDVNDEPPYFINRPLPMQAVVQLNAPPNTPVFTLQA  575

Query  133  TDADEPNHLNSKIAFKIVSQEPAGTPMFLLSRNTGEVRTLTNSLDREQASSYRLVVSGAD  192
             D D     +  I + IV     G   F +   +G VRT    L  +    Y L V   D
Sbjct  576  RDPD----TDHNIHYFIVRDRTGG--RFEVDERSGVVRTRGTDL-FQLDMEYVLYVKAED  628

Query  193  KDGE----GLSTQCECNIKVKDVNDNFPMFRDSQYSARIEEN-ILSSELLRFQVTDLDEE  247
            ++G+       +  E  + +       P F    Y A I EN    S+++  +     +
Sbjct  629  QNGKVDDRRFQSTPEERLSIVG-GKRAPQFYMPSYEAEIPENQKKDSDIISVKA----KS  683

Query  248  YTDNWLAVYFFTSGNEGNWFEIQTDPRTNEGILKVVKALDYEQLQSVKLS----IAVKNK  303
            + D  +       G     F I        GI+K+ K LD+E L+   +      A ++
Sbjct  684  FADREIRYTLKAQGQGAGTFNIGP----TSGIVKLAKELDFEDLRQPHVYSLVVTATEDS  739

Query  304  AEFHQSVISRYRVQSTPVTIQVINVREGIAFRPASKTFTVQKGISSKKLVDYILGT----  359
              F  SV          +TI+V +V +       +  F +    +     D  LGT
Sbjct  740  GGFSTSV---------ELTIRVTDVNDN------APKFELPDYQAHNVDEDIPLGTSILK  784

Query  360  YQAIDEDTNKAASNVKYVMGRN----DGGYLMIDSKTAEIKFVKNMNRDSTFIVNKTITA  415
             +A+D D+  A + ++Y++  +    D   +++++K  +     N N    F+V
Sbjct  785  VKAMDSDSG-ANAEIEYLVSDDHFAVDSNGIIVNNKQLD---ADNNNAYYEFVVTAKDKG  840

Query  416  EVLAIDEYTGKTSTGTVYVRVPDFNDNCP  444
            E         KT T TV V   + ND  P
Sbjct  841  E-------PAKTGTATVRVYTKNKNDEEP  862

 Score = 39.7 bits (91),  Expect = 0.28, Method: Compositional matrix adjust.
 Identities = 54/242 (22%), Positives = 102/242 (42%), Gaps = 22/242 (9%)

Query  4    KFAKPCREGE--DNSKRNP-IAKITSDYQATQKITYRISGVGIDQPPFGIFVVDKNTGDI  60
            +F  P  E E  +N K++  I  + +   A ++I Y +   G      G F +   +G +
Sbjct  656  QFYMPSYEAEIPENQKKDSDIISVKAKSFADREIRYTLKAQGQGA---GTFNIGPTSGIV  712

Query  61   NITAIVDREE--TPSF--LITCRALNAQGLDVEKPLILTVKILDINDNPPVFSQQIFMGE  116
             +   +D E+   P    L+     ++ G      + LT+++ D+NDN P F    +
Sbjct  713  KLAKELDFEDLRQPHVYSLVVTATEDSGGFSTS--VELTIRVTDVNDNAPKFELPDYQAH  770

Query  117  -IEENSASNSLVMILNATDADEPNHLNSKIAFKIVSQEPAGTPMFLLSRNTGEVRTLTNS  175
             ++E+    + ++ + A D+D  +  N++I + +VS +      F +  N   V
Sbjct  771  NVDEDIPLGTSILKVKAMDSD--SGANAEIEY-LVSDD-----HFAVDSNGIIVNNKQLD  822

Query  176  LDREQASSYRLVVSGADKDGEGLSTQCECNIKVKDVNDNFPMFRDSQYSARIEENILSSE  235
             D   A  Y  VV+  DK     +      +  K+ ND  P F    Y+  ++EN   +
Sbjct  823  ADNNNA-YYEFVVTAKDKGEPAKTGTATVRVYTKNKNDEEPKFSQQVYTPNVDENAGPNT  881

Query  236  LL  237
            L+
Sbjct  882  LV  883

>ASTEI20_040830.P55056 | transcript=ASTEI20_040830.R55056 | gene=ASTEI20_040830
| organism=Anopheles_stephensi_UCISS2018 |
gene_product=neural-cadherin, transcript variant X5 |
transcript_product=neural-cadherin,
transcript variant X8 | location=CM023249:57079885-57269506(-)
```

```
| protein_length=3111 | sequence_SO=chromosome
| SO=protein_coding_gene | is_pseudo=false
Length=3111

 Score = 147 bits (371),  Expect = 2e-35, Method: Compositional matrix adjust.
 Identities = 128/446 (29%), Positives = 217/446 (49%), Gaps = 36/446 (8%)

Query  20    PIAKITS---DYQATQKITYRISGVGID--QPPFGIFVVDKNTGDINITAIVDREET---  71
             PI ++T+   D    Q I Y ++G GID   P    F +++ TG+I +   +DR++
Sbjct  1552  PIMQVTATDGDKDRPQNIVYFLTGQGIDPDNPANSKFDINRTTGEIFVLKPLDRDQPNGR  1611

Query  72    PSFLITCRALNAQGLDVEKPLILTVKILDINDNPPVFSQQIFMGEIEENSASNSLVMILN  131
             P +  T  A +  G  +     + V + DINDN P+F Q ++ G + EN  +  +VM +
Sbjct  1612  PQWRFTVFAQDEGGEGLVGYADVQVNLKDINDNAPIFPQGVYFGNVTENGTAGMVVMTMT  1671

Query  132   ATDADEPNH-LNSKIAFKI---VSQEPAGTPMFLLSRNTGEVRTLTNSLDREQASSYRLV  187
             A D D+PN   N+K+ + I   V +E  G+P+F +  +TG ++T    LDRE+   Y +
Sbjct  1672  AVDYDDPNEGTNAKLIYSIEKNVIEEETGSPIFEIEADTGVIKTAVCCLDRERTPDYSIQ  1731

Query  188   VSGADKDGEGLSTQCECNIKVKDVNDNFPMFRDSQYSARIEENILSS--ELLRFQVTDLD  245
             V     DG GL      +I+VKD+ND  P F   ++   ++E   ++  E+    VT  D
Sbjct  1732  VVAM--DGGGLKGTGTASIRVKDINDMPPQFTKDEWFTEVDETDGTNLPEMPILTVTVHD  1789

Query  246   EEYTDNWLAVYFFTSGNEGNWFEIQTDPRTNEGI--LKVVKALDYE---QLQSVKLSIAV  300
             E+ T+ +       SG   + F +    R N+G   LK+V+ LDYE   Q    +  I V
Sbjct  1790  EDETNKFQYKVIDNSGYGADKFTMV---RNNDGTGSLKIVQPLDYEDPLQSNGFRFRIQV  1846

Query  301   KNKAEFHQSVISRYRVQSTPVTIQVINVREGIAFRPASKTFTVQKGISSKKLVDYILGTY  360
              +K E + +   +Y V  + V +++ ++ +    +P  +   ++  +     V   L T+
Sbjct  1847  NDKGEDNDN--DKYHVAYSWVVVKLRDINDN---KPQFERPNIEVSVYENADVGKTLETF  1901

Query  361   QAIDEDTNKAASNVKYVMGR-NDGGYLMIDSKTAEIKFVKNMNRDSTFIVNKTITAEVLA  419
             +A D D     S V Y + R +D       ++   +   +N++R+    V      ++LA
Sbjct  1902  KATDPDQG-GKSKVSYAIDRSSDRQRQFSINQEGTVTIQRNLDRE----VTPRHQVKILA  1956

Query  420   IDE-YTGKTSTGTVYVRVPDFNDNCP  444
             ID+    KT+T T+ V V D NDN P
Sbjct  1957  IDDGIPPKTATATLTVIVQDINDNPP  1982

 Score = 77.0 bits (188),  Expect = 9e-13, Method: Compositional matrix adjust.
 Identities = 103/418 (25%), Positives = 174/418 (42%), Gaps = 50/418 (12%)

Query  50    IFVVDKNTGDINITAIVDREETPSFLITCRAL----------NAQGLDVEKPLILTVKIL  99
              F ++  +G+I    + DRE+  ++ +   A           N Q   V K   + + I
Sbjct  1368  FFEINLQSGEIFTKVVFDREKQGAYALEVEARDGAPSARPNSNQQPNSVTK--FIRIGIA  1425

Query  100   DINDNPPVFSQQIFMGEIEENSASNSLVMILNATDADEPNHLNSKIAFKIVSQEPAGTPM  159
             D NDNPP F + ++  E++EN      V+ + A D DE    +S+I ++I S    G
Sbjct  1426  DKNDNPPYFDKALYEAEVDENEDIQHTVLTVTAKDHDE----SSRIRYEITSGNIGGA--  1479

Query  160   FLLSRNTGEVRTLTNSLDREQASSYRLVVSGADKDGEGLSTQCECNIKVKDVNDNFPMFR  219
             F +   TG +  +  +LD E    Y L +  +D   E  +      I VKDVND  P F
Sbjct  1480  FAVKNMTGAI-YVAGALDYETRKRYHLTLVASDSLNENETIIV---INVKDVNDMPPSFP  1535

Query  220   DSQYSARIEENI-LSSELLRFQVTDLDEEYTDNWLAVYFFTSG--NEGNWFEIQTDPRTN  276
                Y   ++E + +   +++   TD D++   N   VYF T    +  N    + D
Sbjct  1536  QQVYERTMDEELAVPFPIMQVTATDGDKDRPQN--IVYFLTGQGIDPDNPANSKFDINRT  1593

Query  277   EGILKVVKALDYEQLQS-VKLSIAVKNKAEFHQSVISRYRVQSTPVTIQVINVREGIAFR  335
              G + V+K LD +Q     +    V  + E  + ++    VQ     + + ++ +
Sbjct  1594  TGEIFVLKPLDRDQPNGRPQWRFTVFAQDEGGEGLVGYADVQ-----VNLKDINDNAPIF  1648

Query  336   PASKTFTVQKGISSKKLVDYILGTYQAIDEDTNKAASNVK--YVMGRN------DGGYLM  387
             P    F     ++       ++ T  A+D D     +N K  Y + +N
Sbjct  1649  PQGVYF---GNVTENGTAGMVVMTMTAVDYDDPNEGTNAKLIYSIEKNVIEEETGSPIFE  1705

Query  388   IDSKTAEIK-FVKNMNRDSTFIVNKTITAEVLAIDEYTGKTSTGTVYVRVPDFNDNCP  444
             I++ T  IK  V  ++R+ T       + +V+A+D   G   TGT  +RV D ND  P
Sbjct  1706  IEADTGVIKTAVCCLDRERT----PDYSIQVVAMDG-GGLKGTGTASIRVKDINDMPP  1758
```

```
 Score = 75.9 bits (185),  Expect = 2e-12, Method: Compositional matrix adjust.
 Identities = 106/426 (25%), Positives = 181/426 (42%), Gaps = 51/426 (12%)

Query  53    VDKNTGDINITAIVDREETPSFLITCRALNAQGLDVEKPLILTVKILDINDNPPVFSQ-Q  111
             +D++  ++N+TA+ D         +C  +N          ++ V I D+NDN PVF
Sbjct  937   LDRDKYELNVTAMDDG--------SC-CVNGDATIHTSTAVVVVFITDVNDNKPVFKDCG  987

Query  112   IFMGEIEENSASNSLVMILNATDADEPNHLNSKIAFKIVSQEPAGTPMFLLSRNTGEVRT  171
              +  ++EE + + S V+ + ATD D+   +N ++ + IV Q       F +   TGEV T
Sbjct  988   TYYPKVEEGAPNGSPVIKVQATDEDK--GVNGQVKYSIVQQPNQKGTKFTVDEETGEVST  1045

Query  172   LTNSLDRE--QASSYRLVVSGADKDGEGLSTQCECNIKVKDVNDNFPMFRDSQYSARIEE  229
                  DRE        + V   D+    L   C   +++ DVNDN P+F   +Y   +++
Sbjct  1046  -NKVFDREGDDGKFVSVTVKATDQGEPSLEGVCSFTVEITDVNDNPPLFDRQKYVENVKQ  1104

Query  230   NI-LSSELLRFQVTDLDEEYTDNWLAVYFFT---SGNEGNWFEIQTDPRTNEGILKVVKA  285
             +  + + +LR  V+  DE+  +N   VY  +   S N+  +FEIQ  P +   +LK  K
Sbjct  1105  DASIGTNILR--VSASDEDADNNGAIVYTLSAPHSPNDLEYFEIQ--PESGWIVLK--KP  1158

Query  286   LD--YEQLQSVKLSIAVKNK------AEFHQSVISRYRVQSTPVTIQV----INVREGIA  333
             LD  Y + +  +L +   ++      A+    +    R    P+  +     I++RE +
Sbjct  1159  LDGCYSR-ERYRLKVYASDRGDPPSTADVDVELDVVDRNNKPPIWDRTIYGPIHIRENVT  1217

Query  334   FRPASKTFTVQKGISSKKLVDYILGTYQAIDEDTNKAASNVKYVMGRNDGGYLMIDSKTA  393
                   +     GI     V Y L         TNK   +  Y+  R D G        A
Sbjct  1218  VGTVVTSVKASSGIEGNPTVFYRL--MPGSTAQTNKL--HTFYLQQRPDNG-----DTWA  1268

Query  394   EIKFVKNMNRDSTFIVNKTITAEVLAIDEYTGKTSTGTVYVRVPDFNDNCPTAV-LEKDA  452
             +IK    ++ ++    N T+  E     +     S  TVY+++ D ND  P     E++
Sbjct  1269  DIKVNHPLDFETIKEYNLTVRVENNGAQQL---ASEATVYIQLEDVNDEIPLFTEREQET  1325

Query  453   VCSSSP  458
             V    P
Sbjct  1326  VLEGEP  1331

 Score = 60.5 bits (145),  Expect = 1e-07, Method: Compositional matrix adjust.
 Identities = 83/362 (23%), Positives = 152/362 (42%), Gaps = 34/362 (9%)

Query  51    FVVDKNTGDINITAIVDRE-ETPSFL-ITCRALNAQGLDVEKPLILTVKILDINDNPPVF  108
             F VD+ TG+++   + DRE +   F+ +T +A +     +E     TV+I D+NDNPP+F
Sbjct  1034  FTVDEETGEVSTNKVFDREGDDGKFVSVTVKATDQGEPSLEGVCSFTVEITDVNDNPPLF  1093

Query  109   SQQIFMGEIEENSASNSLVMILNATDADEPNHLNSKIAFKIVS-QEPAGTPMFLLSRNTG  167
              +Q ++  ++++++  + ++ ++A+D D  N  N  I + + +   P     F +   +G
Sbjct  1094  DRQKYVENVKQDASIGTNILRVSASDEDADN--NGAIVYTLSAPHSPNDLEYFEIQPESG  1151

Query  168   EV---RTLTNSLDREQASSYRLVVSGADKDGEGLSTQCECNIKVKDVNDNFPMFRDSQYS  224
              +   + L     RE+   YRL V  +D+ G+  ST            +N P   D
Sbjct  1152  WIVLKKPLDGCYSRER---YRLKVYASDR-GDPPSTADVDVELDVVDRNNKPPIWDRTIY  1207

Query  225   ARIEENILSSELLRFQVTDLDEEYTDNWLAVYFFTSG-----NEGNWFEIQTDPRTNE--  277
               I      +                N    Y    G     N+ + F +Q  P   +
Sbjct  1208  GPIHIRENVTVGTVVTSVKASSGIEGNPTVFYRLMPGSTAQTNKLHTFYLQQRPDNGDTW  1267

Query  278   GILKVVKALDYEQLQSVKLSIAVKNKAEFHQSVISRYRVQSTPVTIQVINVREGIAFRPA  337
               +KV   LD+E ++   L++ V+N         ++       V IQ+ +V + I
Sbjct  1268  ADIKVNHPLDFETIKEYNLTVRVENNG-------AQQLASEATVYIQLEDVNDEIPLFTE  1320

Query  338   SKTFTVQKGISSKKLVDYILGTYQAIDEDTNKAASNVKYVM---GRNDG-GYLMIDSKTA  393
              +  TV +G      V  +     AID+D     + V Y +    RN+G  +  I+ ++
Sbjct  1321  REQETVLEGEPIGTKVTQV----NAIDKDGTFPNNQVYYYIVDSPRNEGKDFFEINLQSG  1376

Query  394   EI  395
             EI
Sbjct  1377  EI  1378

 Score = 55.8 bits (133),  Expect = 3e-06, Method: Compositional matrix adjust.
 Identities = 49/158 (31%), Positives = 78/158 (49%), Gaps = 24/158 (15%)
```

```
Query  102   NDNPPVFSQQIFMGEIEENSASNSLVMILNATDADEPNHLNSKIAFKIVSQEPAGTP--M  159
             ND  P FSQQ++   ++EN+  N+LV  + A+D D  N     + F  V     GT
Sbjct  867   NDEEPKFSQQVYTPNVDENAGPNTLVTTVVASDKDGDN-----VRFGFVG---GGTSSGQ  918

Query  160   FLLSRNTGEVRTLTN--SLDREQASSYRLVVSGADK-------DGEGLSTQCECNIKVKD  210
             F++   TG +R  +   SLDR++   Y L V+  D        D    ++     + + D
Sbjct  919   FVIEEITGVIRLHSKAISLDRDK---YELNVTAMDDGSCCVNGDATIHTSTAVVVVFITD  975

Query  211   VNDNFPMFRD-SQYSARIEENILS-SELLRFQVTDLDE  246
             VNDN P+F+D   Y  ++EE   + S +++ Q TD D+
Sbjct  976   VNDNKPVFKDCGTYYPKVEEGAPNGSPVIKVQATDEDK  1013

  Score = 53.9 bits (128),  Expect = 1e-05, Method: Compositional matrix adjust.
  Identities = 70/306 (23%), Positives = 126/306 (41%), Gaps = 30/306 (10%)

Query  20    PIAKIT-SDYQATQKITYRI---SGVGIDQPPFGIFVVDKNTGDINITAIVDRE---ETP  72
             PI  +T  D   T K  Y++   SG G D+  F +   +  TG + I   +D E   ++
Sbjct  1781  PILTVTVHDEDETNKFQYKVIDNSGYGADK--FTMVRNNDGTGSLKIVQPLDYEDPLQSN  1838

Query  73    SFLITCRALNAQGLDVEKPLI------LTVKILDINDNPPVFSQQIFMGEIEENSASNSL  126
              F    + +N +G D +          + VK+ DINDN P F +      + EN+
Sbjct  1839  GFRFRIQ-VNDKGEDNDNDKYHVAYSWVVVKLRDINDNKPQFERPNIEVSVYENADVGKT  1897

Query  127   VMILNATDADEPNHLNSKIAFKIVSQEPAGTPMFLLSRNTGEVRTLTNSLDREQASSYRL  186
             +    ATD D+     SK+++ I   + +       S N     T+  +LDRE    +++
Sbjct  1898  LETFKATDPDQGG--KSKVSYAI---DRSSDRQRQFSINQEGTVTIQRNLDREVTPRHQV  1952

Query  187   VVSGADKDGEGLSTQCECNIKVKDVNDNFPMFRDSQYSARIEENILSSELLRFQVTDLDE  246
              +   D      +      + V+D+NDN P F    Y   + E++   +++    TD D+
Sbjct  1953  KILAIDDGIPPKTATATLTVIVQDINDNPPKFL-KDYRPVLPEHVPPRKVVEILATDDDD  2011

Query  247   EYTDNWLAVYFFTSGNEGNW----FEIQTDPRTNEG----ILKVVKALDYEQLQSVKLSI  298
                 N     F       +     F+++ D +   G    I+  +++ D EQ +   + I
Sbjct  2012  RSKSNGPPFQFRLDPGADDIIRASFKVEQDQKGANGDGMAIVSSLRSFDREQQKEYLIPI  2071

Query  299   AVKNKA  304
              +K+
Sbjct  2072  VIKDHG  2077

  Score = 52.4 bits (124),  Expect = 3e-05, Method: Compositional matrix adjust.
  Identities = 97/398 (24%), Positives = 163/398 (41%), Gaps = 54/398 (14%)

Query  106   PVFSQQIFMGEIEENSASNSLVMILNATDADEPNHLNSKIAFKIVSQ-EPAGTPMFLLSR  164
             P F    +  EI EN   +S ++ + A      +  + +I + + +Q + AGT  F +
Sbjct  664   PQFYMPSYEAEIPENQKKDSDIISVKAK-----SFADREIRYTLKAQGQGAGT--FNIGP  716

Query  165   NTGEVRTLTNSLDRE---QASSYRLVVSGADKDGEGLSTQCECNIKVKDVNDNFPMFRDS  221
              +G V+ L   LD E   Q   Y LVV+ A +D  G ST  E  I+V DVNDN P F
Sbjct  717   TSGIVK-LAKELDFEDLRQPHVYSLVVT-ATEDSGGFSTSVELTIRVTDVNDNAPKFELP  774

Query  222   QYSAR-IEENI-LSSELLRFQVTDLDEEYTDNWLAVYFFTSGNEGNWFEIQTDPRTNEGI  279
              Y A  ++E+I L + +L+ +  D D         + +  S +    F + ++     GI
Sbjct  775   DYQAHNVDEDIPLGTSILKVKAMDADSGAN---AEIEYLVSDDH---FAVDSN-----GI  823

Query  280   LKVVKALDYEQLQS-VKLSIAVKNKAEFHQSVISRYRVQSTPVTIQVINVREGIAFRPAS  338
             +   K LD +   +  +  +  K+K E  ++  +  RV +           E   F
Sbjct  824   IVNNKQLDADNNNAYYEFVVTAKDKGEPAKTGTATVRVYTKNKN------DEEPKFSQQV  877

Query  339   KTFTVQKGISSKKLVDYILGTYQAIDEDTNKAASNVK--YVMGRNDGGYLMIDSKTAEIK  396
              T  V +      LV  ++         ++K   NV+  +V G    G  +I+  T  I+
Sbjct  878   YTPNVDENAGPNTLVTTVVA--------SDKDGDNVRFGFVGGGTSSGQFVIEEITGVIR  929

Query  397   FVK---NMNRDSTFI-VNKTITAEVLAIDEYTGKTSTGTVYVRVPDFNDN------CPTA  446
                   +++RD   + V            + T  TST  V V + D NDN      C T
Sbjct  930   LHSKAISLDRDKYELNVTAMDDGSCCVNGDATIHTSTAVVVVFITDVNDNKPVFKDCGTY  989

Query  447   VLE-KDAVCSSSPSVVVSARTLNNRYTGPYTFALEDQP  483
               + ++   + SP + V A   +    G   +++  QP
```

```
Sbjct  990   YPKVEEGAPNGSPVIKVQATDEDKGVNGQVKYSIVQQP  1027

 Score = 44.7 bits (104),  Expect = 0.008, Method: Compositional matrix adjust.
 Identities = 48/199 (24%), Positives = 73/199 (37%), Gaps = 19/199 (10%)

Query  27    DYQATQKITYRISGVGIDQPPFGIFVVDKNTGDINITAIVDREETPSFLITCRALNAQGL  86
             D     K++Y I      Q  F I       G + I   +DRE TP   +   A++
Sbjct  1907  DQGGKSKVSYAIDRSSDRQRQFSI----NQEGTVTIQRNLDREVTPRHQVKILAIDDGIP  1962

Query  87    DVEKPLILTVKILDINDNPPVFSQQIFMGEIEENSASNSLVMILNATDADEPNHLNSKIA  146
                    LTV + DINDNPP F +  +   + E+     +V IL   D D
Sbjct  1963  PKTATATLTVIVQDINDNPPKFLKD-YRPVLPEHVPPRKVVEILATDDDDRSKSNGPPFQ  2021

Query  147   FKIVSQEPAGTPMFLLSRNTGEVRTLTN-----------SLDREQASSYRLVVSGADKDG  195
             F++   +P    +   S    + +   N           S DREQ   Y + +   D
Sbjct  2022  FRL---DPGADDIIRASFKVEQDQKGANGDGMAIVSSLRSFDREQQKEYLIPIVIKDHGN  2078

Query  196   EGLSTQCECNIKVKDVNDN  214
               ++      + + DVNDN
Sbjct  2079  PAMTGTSTLTVVIGDVNDN  2097

 Score = 40.8 bits (94),  Expect = 0.11, Method: Compositional matrix adjust.
 Identities = 55/242 (23%), Positives = 102/242 (42%), Gaps = 22/242 (9%)

Query  4    KFAKPCREGE--DNSKRNP-IAKITSDYQATQKITYRISGVGIDQPPFGIFVVDKNTGDI  60
            +F  P  E E  +N K++  I  + +   A ++I Y +   G      G F +   +G +
Sbjct  665  QFYMPSYEAEIPENQKKDSDIISVKAKSFADREIRYTLKAQGQGA---GTFNIGPTSGIV  721

Query  61   NITAIVDREE--TPSF--LITCRALNAQGLDVEKPLILTVKILDINDNPPVFSQQIFMGE  116
             +   +D E+   P    L+     ++ G      + LT+++ D+NDN P F    +
Sbjct  722  KLAKELDFEDLRQPHVYSLVVTATEDSGGFSTS--VELTIRVTDVNDNAPKFELPDYQAH  779

Query  117  -IEENSASNSLVMILNATDADEPNHLNSKIAFKIVSQEPAGTPMFLLSRNTGEVRTLTNS  175
             ++E+    + ++ + A DAD  +  N++I + +VS +      F +  N   V
Sbjct  780  NVDEDIPLGTSILKVKAMDAD--SGANAEIEY-LVSDD-----HFAVDSNGIIVNNKQLD  831

Query  176  LDREQASSYRLVVSGADKDGEGLSTQCECNIKVKDVNDNFPMFRDSQYSARIEENILSSE  235
             D   A  Y  VV+  DK     +      +  K+ ND  P F    Y+  ++EN   +
Sbjct  832  ADNNNA-YYEFVVTAKDKGEPAKTGTATVRVYTKNKNDEEPKFSQQVYTPNVDENAGPNT  890

Query  236  LL  237
            L+
Sbjct  891  LV  892

 Score = 39.3 bits (90),  Expect = 0.29, Method: Compositional matrix adjust.
 Identities = 89/389 (23%), Positives = 149/389 (38%), Gaps = 62/389 (16%)

Query  75   LITCRALNAQGLDVEKPLILTVKILDINDNPPVFSQQIF--MGEIEENSASNSLVMILNA  132
            +IT +  NA GL       + + + D+ND PP F  +       ++ N+  N+ V  L A
Sbjct  526  IITNQGHNA-GLKYTDNQRVIIHVKDVNDEPPYFINRPLPMQAVVQLNAPPNTPVFTLQA  584

Query  133  TDADEPNHLNSKIAFKIVSQEPAGTPMFLLSRNTGEVRTLTNSLDREQASSYRLVVSGAD  192
             D D     +  I + IV     G   F +   +G VRT    L  +    Y L V   D
Sbjct  585  RDPD----TDHNIHYFIVRDRTGG--RFEVDERSGVVRTRGTDL-FQLDMEYVLYVKAED  637

Query  193  KDGE----GLSTQCECNIKVKDVNDNFPMFRDSQYSARIEEN-ILSSELLRFQVTDLDEE  247
            ++G+       +  E  + +       P F    Y A I EN    S+++  +     +
Sbjct  638  QNGKVDDRRFQSTPEERLSIVG-GKRAPQFYMPSYEAEIPENQKKDSDIISVKA----KS  692

Query  248  YTDNWLAVYFFTSGNEGNWFEIQTDPRTNEGILKVVKALDYEQLQSVKLS----IAVKNK  303
            + D  +       G     F I        GI+K+ K LD+E L+   +      A ++
Sbjct  693  FADREIRYTLKAQGQGAGTFNIGP----TSGIVKLAKELDFEDLRQPHVYSLVVTATEDS  748

Query  304  AEFHQSVISRYRVQSTPVTIQVINVREGIAFRPASKTFTVQKGISSKKLVDYILGT----  359
              F  SV          +TI+V +V +       +  F +    +     D  LGT
Sbjct  749  GGFSTSV---------ELTIRVTDVNDN------APKFELPDYQAHNVDEDIPLGTSILK  793

Query  360  YQAIDEDTNKAASNVKYVMGRN----DGGYLMIDSKTAEIKFVKNMNRDSTFIVNKTITA  415
```

```
             +A+D D+  A + ++Y++  +    D   +++++K  +     N N    F+V
Sbjct  794  VKAMDADSG-ANAEIEYLVSDDHFAVDSNGIIVNNKQLD---ADNNNAYYEFVVTAKDKG  849

Query  416  EVLAIDEYTGKTSTGTVYVRVPDFNDNCP  444
            E         KT T TV V   + ND  P
Sbjct  850  E-------PAKTGTATVRVYTKNKNDEEP  871

>AGAMI1_007196.P14118 | transcript=AGAMI1_007196.R14118 | gene=AGAMI1_007196
| organism=Anopheles_gambiae_Ifakara | gene_product=neural-cadherin
| transcript_product=neural-cadherin,
transcript variant X17 | location=AgamI1_3RL:41252608-41455387(+)
| protein_length=3098 | sequence_SO=chromosome | SO=protein_coding_gene
| is_pseudo=false
Length=3098

 Score = 147 bits (371),  Expect = 2e-35, Method: Compositional matrix adjust.
 Identities = 128/446 (29%), Positives = 217/446 (49%), Gaps = 36/446 (8%)

Query  20    PIAKITS---DYQATQKITYRISGVGID--QPPFGIFVVDKNTGDINITAIVDREET---  71
             PI ++T+   D    Q I Y ++G GID   P    F +++ TG+I +   +DR++
Sbjct  1539  PIMQVTATDGDKDRPQNIVYFLTGQGIDPDNPANSKFDINRTTGEIFVLKPLDRDQPNGR  1598

Query  72    PSFLITCRALNAQGLDVEKPLILTVKILDINDNPPVFSQQIFMGEIEENSASNSLVMILN  131
             P +  T  A +  G  +     + V + DINDN P+F Q ++ G + EN  +  +VM +
Sbjct  1599  PQWRFTVFAQDEGGEGLVGYADVQVNLKDINDNAPIFPQGVYFGNVTENGTAGMVVMTMT  1658

Query  132   ATDADEPNH-LNSKIAFKI---VSQEPAGTPMFLLSRNTGEVRTLTNSLDREQASSYRLV  187
             A D D+PN   N+K+ + I   V +E  G+P+F +  +TG ++T    LDRE+   Y +
Sbjct  1659  AVDYDDPNEGTNAKLIYSIEKNVIEEETGSPIFEIEADTGVIKTAVCCLDRERTPDYSIQ  1718

Query  188   VSGADKDGEGLSTQCECNIKVKDVNDNFPMFRDSQYSARIEENILSS--ELLRFQVTDLD  245
             V     DG GL      +I+VKD+ND  P F   ++   ++E   ++  E+    VT  D
Sbjct  1719  VVAM--DGGGLKGTGTASIRVKDINDMPPQFTKDEWFTEVDETDGTNLPEMPILTVTVHD  1776

Query  246   EEYTDNWLAVYFFTSGNEGNWFEIQTDPRTNEGI--LKVVKALDYE---QLQSVKLSIAV  300
             E+ T+ +       SG   + F +    R N+G   LK+V+ LDYE   Q    +  I V
Sbjct  1777  EDETNKFQYKVIDNSGYGADKFTMV---RNNDGTGSLKIVQPLDYEDPLQSNGFRFRIQV  1833

Query  301   KNKAEFHQSVISRYRVQSTPVTIQVINVREGIAFRPASKTFTVQKGISSKKLVDYILGTY  360
              +K E + +   +Y V  + V +++ ++ +    +P  +   ++  +     V   L T+
Sbjct  1834  NDKGEDNDN--DKYHVAYSWVVVKLRDINDN---KPQFERPNIEVSVYENADVGKTLETF  1888

Query  361   QAIDEDTNKAASNVKYVMGR-NDGGYLMIDSKTAEIKFVKNMNRDSTFIVNKTITAEVLA  419
             +A D D     S V Y + R +D       ++   +   +N++R+    V      ++LA
Sbjct  1889  KATDPDQG-GKSKVSYAIDRSSDRQRQFSINQEGTVTIQRNLDRE----VTPRHQVKILA  1943

Query  420   IDE-YTGKTSTGTVYVRVPDFNDNCP  444
             ID+    KT+T T+ V V D NDN P
Sbjct  1944  IDDGIPPKTATATLTVIVQDINDNPP  1969

 Score = 79.0 bits (193),  Expect = 2e-13, Method: Compositional matrix adjust.
 Identities = 106/424 (25%), Positives = 180/424 (42%), Gaps = 51/424 (12%)

Query  53    VDKNTGDINITAIVDREETPSFLITCRALNAQGLDVEKPLILTVKILDINDNPPVFSQ-Q  111
             +D++  ++N+TA+ D         +C  +N          ++ V I D+NDN PVF
Sbjct  928   LDRDKYELNVTAMDDG--------SC-CVNGDATIHTSTAVVVVFITDVNDNKPVFKDCG  978

Query  112   IFMGEIEENSASNSLVMILNATDADEPNHLNSKIAFKIVSQEPAGTPMFLLSRNTGEVRT  171
              +  ++EE + + S V+ + ATD D+   +N ++ + IV Q       F +   TGEV T
Sbjct  979   TYYPKVEEGAPNGSPVIKVQATDEDK--GVNGQVKYSIVQQPNQKGTKFTVDEETGEVST  1036

Query  172   LTNSLDRE--QASSYRLVVSGADKDGEGLSTQCECNIKVKDVNDNFPMFRDSQYSARIEE  229
                  DRE        + V   D+    L   C   +++ DVNDN P+F   +Y   +++
Sbjct  1037  -NKVFDREGDDGKFVSVTVKATDQGEPSLEGVCSFTVEITDVNDNPPLFDRQKYVENVKQ  1095

Query  230   NI-LSSELLRFQVTDLDEEYTDNWLAVYFFT---SGNEGNWFEIQTDPRTNEGILKVVKA  285
             +  + + +LR  V+  DE+  +N   VY  +   S N+  +FEIQ  P +   +LK  K
Sbjct  1096  DASIGTNILR--VSASDEDADNNGAIVYTLSAPHSPNDLEYFEIQ--PESGWIVLK--KP  1149
```

```
Query  286   LDYEQLQSVKLSIAVKNK------AEFHQSVISRYRVQSTPVTIQV----INVREGIAFR  335
             LD E+    +L +   ++       A+    +    R    P+  +     I++RE +
Sbjct  1150  LDRERY---RLKVYASDRGDPPSTADVDVELDVVDRNNKPPIWDRTIYGPIHIRENVTVG  1206

Query  336   PASKTFTVQKGISSKKLVDYILGTYQAIDEDTNKAASNVKYVMGRNDGGYLMIDSKTAEI  395
                 +     GI     V Y L          TNK    +  Y+  R D G         A+I
Sbjct  1207  TVVTSVKASSGIEGNPTVFYRL--MPGSTAQTNKL--HTFYLQQRPDNG-----DTWADI  1257

Query  396   KFVKNMNRDSTFIVNKTITAEVLAIDEYTGKTSTGTVYYRVPDFNDNCPTAV-LEKDAVC  454
             K    ++ ++    N T+  E     +     S  TVY+++ D ND  P     E++ V
Sbjct  1258  KVNHPLDFETIKEYNLTVRVENNGAQQL---ASEATVYIQLEDVNDEIPLFTEREQETVL  1314

Query  455   SSSP  458
                P
Sbjct  1315  EGEP  1318

 Score = 76.3 bits (186),  Expect = 1e-12, Method: Compositional matrix adjust.
 Identities = 103/418 (25%), Positives = 174/418 (42%), Gaps = 50/418 (12%)

Query  50    IFVVDKNTGDINITAIVDREETPSFLITCRAL----------NAQGLDVEKPLILTVKIL  99
              F ++  +G+I    + DRE+  ++ +   A           N Q   V K   + + I
Sbjct  1355  FFEINLQSGEIFTKVVFDREKQGAYALEVEARDGAPSARPNSNQQPNSVTK--FIRIGIA  1412

Query  100   DINDNPPVFSQQIFMGEIEENSASNSLVMILNATDADEPNHLNSKIAFKIVSQEPAGTPM  159
             D NDNPP F + ++  E++EN      V+ + A D DE    +S+I ++I S    G
Sbjct  1413  DKNDNPPYFDKALYEAEVDENEDIQHTVLTVTAKDHDE----SSRIRYEITSGNIGGA--  1466

Query  160   FLLSRNTGEVRTLTNSLDREQASSYRLVVSGADKDGEGLSTQCECNIKVKDVNDNFPMFR  219
             F +   TG +  +  +LD E    Y L +  +D   E  +      I VKDVND  P F
Sbjct  1467  FAVKNMTGAI-YVAGALDYETRKRYYLTLVASDSLNENETIIV---INVKDVNDMPPSFP  1522

Query  220   DSQYSARIEENI-LSSELLRFQVTDLDEEYTDNWLAVYFFTSG--NEGNWFEIQTDPRTN  276
                Y   ++E + +   +++   TD D++   N   VYF T    +  N    + D
Sbjct  1523  QQVYERTMDEELAVPFPIMQVTATDGDKDRPQN--IVYFLTGQGIDPDNPANSKFDINRT  1580

Query  277   EGILKVVKALDYEQLQS-VKLSIAVKNKAEFHQSVISRYRVQSTPVTIQVINVREGIAFR  335
              G + V+K LD +Q     +     V  + E  + ++      VQ     + + ++ +
Sbjct  1581  TGEIFVLKPLDRDQPNGRPQWRFTVFAQDEGGEGLVGYADVQ-----VNLKDINDNAPIF  1635

Query  336   PASKTFTVQKGISSKKLVDYILGTYQAIDEDTNKAASNVK--YVMGRN------DGGYLM  387
             P    F     ++        ++ T  A+D D     +N K  Y + +N
Sbjct  1636  PQGVYF---GNVTENGTAGMVVMTMTAVDYDDPNEGTNAKLIYSIEKNVIEEETGSPIFE  1692

Query  388   IDSKTAEIK-FVKNMNRDSTFIVNKTITAEVLAIDEYTGKTSTGTVYYRVPDFNDNCP  444
             I++ T  IK  V  ++R+ T       + +V+A+D   G   TGT  +RV D ND  P
Sbjct  1693  IEADTGVIKTAVCCLDRERT----PDYSIQVVAMDG-GGLKGTGTASIRVKDINDMPP  1745

 Score = 65.5 bits (158),  Expect = 3e-09, Method: Compositional matrix adjust.
 Identities = 85/359 (24%), Positives = 153/359 (43%), Gaps = 32/359 (9%)

Query  51    FVVDKNTGDINITAIVDRE-ETPSFL-ITCRALNAQGLDVEKPLILTVKILDINDNPPVF  108
             F VD+ TG+++   + DRE +   F+ +T +A +     +E     TV+I D+NDNPP+F
Sbjct  1025  FTVDEETGEVSTNKVFDREGDDGKFVSVTVKATDQGEPSLEGVCSFTVEITDVNDNPPLF  1084

Query  109   SQQIFMGEIEENSASNSLVMILNATDADEPNHLNSKIAFKIVS-QEPAGTPMFLLSRNTG  167
              +Q ++  ++++++  + ++ ++A+D D  N  N  I + + +   P     F +   +G
Sbjct  1085  DRQKYVENVKQDASIGTNILRVSASDEDADN--NGAIVYTLSAPHSPNDLEYFEIQPESG  1142

Query  168   EVRTLTNSLDREQASSYRLVVSGADKDGEGLSTQCECNIKVKDVNDNFPMFRDSQYSARI  227
              +  L   LDRE+   YRL V  +D+ G+  ST              +N P   D      I
Sbjct  1143  WI-VLKKPLDRER---YRLKVYASDR-GDPPSTADVDVELDVVDRNNKPPIWDRTIYGPI  1197

Query  228   EENILSSELLRFQVTDLDEEYTDNWLAVYFFTSG-----NEGNWFEIQTDPRTNE--GIL  280
                  +                 N    Y    G     N+ + F +Q  P    +    +
Sbjct  1198  HIRENVTVGTVVTSVKASSGIEGNPTVFYRLMPGSTAQTNKLHTFYLQQRPDNGDTWADI  1257

Query  281   KVVKALDYEQLQSVKLSIAVKNKAEFHQSVISRYRVQSTPVTIQVINVREGIAFRPASKT  340
             KV   LD+E ++   L++ V+N          ++        V IQ+ +V + I       +
Sbjct  1258  KVNHPLDFETIKEYNLTVRVENNG-------AQQLASEATVYIQLEDVNDEIPLFTEREQ  1310
```

```
Query  341   FTVQKGISSKKLVDYILGTYQAIDEDTNKAASNVKYVM---GRNDG-GYLMIDSKTAEI  395
              TV +G      V  +     AID+D     + V Y +    RN+G  +  I+ ++ EI
Sbjct  1311  ETVLEGEPIGTKVTQV----NAIDKDGTFPNNQVYYYIVDSPRNEGKDFFEINLQSGEI  1365

 Score = 55.8 bits (133),  Expect = 3e-06, Method: Compositional matrix adjust.
 Identities = 49/158 (31%), Positives = 78/158 (49%), Gaps = 24/158 (15%)

Query  102   NDNPPVFSQQIFMGEIEENSASNSLVMILNATDADEPNHLNSKIAFKIVSQEPAGTP--M  159
             ND  P FSQQ++   ++EN+  N+LV  + A+D D  N     + F  V     GT
Sbjct  858   NDEEPKFSQQVYTPNVDENAGPNTLVTTVVASDKDGDN-----VRFGFVG---GGTSSGQ  909

Query  160   FLLSRNTGEVRTLTN--SLDREQASSYRLVVSGADK-------DGEGLSTQCECNIKVKD  210
             F++   TG +R  +   SLDR++   Y L V+  D         D    ++     + + D
Sbjct  910   FVIEEITGVIRLHSKAISLDRDK---YELNVTAMDDGSCCVNGDATIHTSTAVVVVFITD  966

Query  211   VNDNFPMFRD-SQYSARIEENILS-SELLRFQVTDLDE  246
             VNDN P+F+D   Y  ++EE   + S +++ Q TD D+
Sbjct  967   VNDNKPVFKDCGTYYPKVEEGAPNGSPVIKVQATDEDK  1004

 Score = 53.9 bits (128),  Expect = 1e-05, Method: Compositional matrix adjust.
 Identities = 70/306 (23%), Positives = 126/306 (41%), Gaps = 30/306 (10%)

Query  20    PIAKIT-SDYQATQKITYRI---SGVGIDQPPFGIFVVDKNTGDINITAIVDRE---ETP  72
             PI  +T  D   T K  Y++   SG G D+  F +   +  TG + I   +D E   ++
Sbjct  1768  PILTVTVHDEDETNKFQYKVIDNSGYGADK--FTMVRNNDGTGSLKIVQPLDYEDPLQSN  1825

Query  73    SFLITCRALNAQGLDVEKPLI------LTVKILDINDNPPVFSQQIFMGEIEENSASNSL  126
              F    + +N +G D +           + VK+ DINDN P F +       + EN+
Sbjct  1826  GFRFRIQ-VNDKGEDNDNDKYHVAYSWVVVKLRDINDNKPQFERPNIEVSVYENADVGKT  1884

Query  127   VMILNATDADEPNHLNSKIAFKIVSQEPAGTPMFLLSRNTGEVRTLTNSLDREQASSYRL  186
             +    ATD D+     SK+++ I    + +        S N      T+  +LDRE    +++
Sbjct  1885  LETFKATDPDQGG--KSKVSYAI---DRSSDRQRQFSINQEGTVTIQRNLDREVTPRHQV  1939

Query  187   VVSGADKDGEGLSTQCECNIKVKDVNDNFPMFRDSQYSARIEENILSSELLRFQVTDLDE  246
              +   D      +      + V+D+NDN P F    Y   + E++   +++    TD D+
Sbjct  1940  KILAIDDGIPPKTATATLTVIVQDINDNPPKFL-KDYRPVLPEHVPPRKVVEILATDDDD  1998

Query  247   EYTDNWLAVYFFTSGNEGNW----FEIQTDPRTNEG----ILKVVKALDYEQLQSVKLSI  298
                 N     F       +     F+++ D +   G    I+  +++ D EQ +   + I
Sbjct  1999  RSKSNGPPFQFRLDPGADDIIRASFKVEQDQKGANGDGMAIVSSLRSFDREQQKEYLIPI  2058

Query  299   AVKNKA  304
              +K+
Sbjct  2059  VIKDHG  2064

 Score = 52.0 bits (123),  Expect = 4e-05, Method: Compositional matrix adjust.
 Identities = 97/398 (24%), Positives = 163/398 (41%), Gaps = 54/398 (14%)

Query  106   PVFSQQIFMGEIEENSASNSLVMILNATDADEPNHLNSKIAFKIVSQ-EPAGTPMFLLSR  164
             P F    +  EI EN   +S ++ + A      +  + +I + + +Q + AGT  F +
Sbjct  655   PQFYMPSYEAEIPENQKKDSDIISVKAK-----SFADREIRYTLKAQGQGAGT--FNIGP  707

Query  165   NTGEVRTLTNSLDRE---QASSYRLVVSGADKDGEGLSTQCECNIKVKDVNDNFPMFRDS  221
              +G V+ L   LD E   Q   Y LVV+ A +D  G ST  E  I+V DVNDN P F
Sbjct  708   TSGIVK-LAKELDFEDLRQPHVYSLVVT-ATEDSGGFSTSVELTIRVTDVNDNAPKFELP  765

Query  222   QYSAR-IEENI-LSSELLRFQVTDLDEEYTDNWLAVYFFTSGNEGNWFEIQTDPRTNEGI  279
              Y A  ++E+I L + +L+ +  D D          + +  S +    F + ++     GI
Sbjct  766   DYQAHNVDEDIPLGTSILKVKAMDSDSGAN---AEIEYLVSDDH---FAVDSN-----GI  814

Query  280   LKVVKALDYEQLQS-VKLSIAVKNKAEFHQSVISRYRVQSTPVTIQVINVREGIAFRPAS  338
             +   K LD +   +  +  +  K+K E  ++  +  RV +             E   F
Sbjct  815   IVNNKQLDADNNNAYYEFVVTAKDKGEPAKTGTATVRVYTKNKN------DEEPKFSQQV  868

Query  339   KTFTVQKGISSKKLVDYILGTYQAIDEDTNKAASNVK--YVMGRNDGGYLMIDSKTAEIK  396
              T V +      LV  ++          ++K   NV+  +V G    G +I+  T  I+
```

```
Sbjct  869   YTPNVDENAGPNTLVTTVVA--------SDKDGDNVRFGFVGGGTSSGQFVIEEITGVIR  920

Query  397   FVK---NMNRDSTFI-VNKTITAEVLAIDEYTGKTSTGTVYVRVPDFNDN------CPTA  446
                   +++RD   + V            + T  TST  V V + D NDN      C T
Sbjct  921   LHSKAISLDRDKYELNVTAMDDGSCCVNGDATIHTSTAVVVVFITDVNDNKPVFKDCGTY  980

Query  447   VLE-KDAVCSSSPSVVVSARTLNNRYTGPYTFALEDQP  483
               + ++   + SP + V A   +    G   +++  QP
Sbjct  981   YPKVEEGAPNGSPVIKVQATDEDKGVNGQVKYSIVQQP  1018

 Score = 44.7 bits (104),  Expect = 0.008, Method: Compositional matrix adjust.
 Identities = 48/199 (24%), Positives = 73/199 (37%), Gaps = 19/199 (10%)

Query  27    DYQATQKITYRISGVGIDQPPFGIFVVDKNTGDINITAIVDREETPSFLITCRALNAQGL  86
             D     K++Y I      Q  F I       G + I   +DRE TP   +   A++
Sbjct  1894  DQGGKSKVSYAIDRSSDRQRQFSI----NQEGTVTIQRNLDREVTPRHQVKILAIDDGIP  1949

Query  87    DVEKPLILTVKILDINDNPPVFSQQIFMGEIEENSASNSLVMILNATDADEPNHLNSKIA  146
                    LTV + DINDNPP F +  +   + E+     +V IL   D D
Sbjct  1950  PKTATATLTVIVQDINDNPPKFLKD-YRPVLPEHVPPRKVVEILATDDDDRSKSNGPPFQ  2008

Query  147   FKIVSQEPAGTPMFLLSRNTGEVRTLTN-----------SLDREQASSYRLVVSGADKDG  195
             F++   +P    +   S    + +   N           S DREQ   Y + +   D
Sbjct  2009  FRL---DPGADDIIRASFKVEQDQKGANGDGMAIVSSLRSFDREQQKEYLIPIVIKDHGN  2065

Query  196   EGLSTQCECNIKVKDVNDN  214
               ++      + + DVNDN
Sbjct  2066  PAMTGTSTLTVVIGDVNDN  2084

 Score = 40.0 bits (92),  Expect = 0.20, Method: Compositional matrix adjust.
 Identities = 89/389 (23%), Positives = 149/389 (38%), Gaps = 62/389 (16%)

Query  75    LITCRALNAQGLDVEKPLILTVKILDINDNPPVFSQQIF--MGEIEENSASNSLVMILNA  132
             +IT +  NA GL       + + + D+ND PP F  +        ++ N+  N+ V  L A
Sbjct  517   IITNQGHNA-GLKYTDNQRVIIHVKDVNDEPPYFINRPLPMQAVVQLNAPPNTPVFTLQA  575

Query  133   TDADEPNHLNSKIAFKIVSQEPAGTPMFLLSRNTGEVRTLTNSLDREQASSYRLVVSGAD  192
              D D     +  I + IV     G   F +   +G VRT    L  +    Y L V   D
Sbjct  576   RDPD----TDHNIHYFIVRDRTGG--RFEVDERSGVVRTRGTDL-FQLDMEYVLYVKAED  628

Query  193   KDGE----GLSTQCECNIKVKDVNDNFPMFRDSQYSARIEEN-ILSSELLRFQVTDLDEE  247
             ++G+       +  E  + +       P F    Y A I EN    S+++  +     +
Sbjct  629   QNGKVDDRRFQSTPEERLSIVG-GKRAPQFYMPSYEAEIPENQKKDSDIISVKA----KS  683

Query  248   YTDNWLAVYFFTSGNEGNWFEIQTDPRTNEGILKVVKALDYEQLQSVKLS----IAVKNK  303
             + D  +       G     F I        GI+K+ K LD+E L+   +      A ++
Sbjct  684   FADREIRYTLKAQGQGAGTFNIGP----TSGIVKLAKELDFEDLRQPHVYSLVVTATEDS  739

Query  304   AEFHQSVISRYRVQSTPVTIQVINVREGIAFRPASKTFTVQKGISSKKLVDYILGT----  359
               F  SV          +TI+V +V +       +  F +    +     D  LGT
Sbjct  740   GGFSTSV---------ELTIRVTDVNDN------APKFELPDYQAHNVDEDIPLGTSILK  784

Query  360   YQAIDEDTNKAASNVKYVMGRN----DGGYLMIDSKTAEIKFVKNMNRDSTFIVNKTITA  415
              +A+D D+  A + ++Y++  +    D   +++++K  +     N N    F+V
Sbjct  785   VKAMDSDSG-ANAEIEYLVSDDHFAVDSNGIIVNNKQLD---ADNNNAYYEFVVTAKDKG  840

Query  416   EVLAIDEYTGKTSTGTVYVRVPDFNDNCP  444
             E         KT T TV V   + ND  P
Sbjct  841   E-------PAKTGTATVRVYTKNKNDEEP  862

 Score = 39.7 bits (91),  Expect = 0.28, Method: Compositional matrix adjust.
 Identities = 54/242 (22%), Positives = 102/242 (42%), Gaps = 22/242 (9%)

Query  4     KFAKPCREGE--DNSKRNP-IAKITSDYQATQKITYRISGVGIDQPPFGIFVVDKNTGDI  60
             +F  P  E E  +N K++  I  + +   A ++I Y +   G      G F +   +G +
Sbjct  656   QFYMPSYEAEIPENQKKDSDIISVKAKSFADREIRYTLKAQGQGA---GTFNIGPTSGIV  712

Query  61    NITAIVDREE--TPSF--LITCRALNAQGLDVEKPLILTVKILDINDNPPVFSQQIFMGE  116
```

```
               +   +D E+   P    L+     ++ G      + LT+++ D+NDN P F    +
Sbjct  713  KLAKELDFEDLRQPHVYSLVVTATEDSGGFSTS--VELTIRVTDVNDNAPKFELPDYQAH  770

Query  117  -IEENSASNSLVMILNATDADEPNHLNSKIAFKIVSQEPAGTPMFLLSRNTGEVRTLTNS  175
             ++E+    + ++ + A D+D  +  N++I + +VS +      F +  N   V
Sbjct  771  NVDEDIPLGTSILKVKAMDSD--SGANAEIEY-LVSDD-----HFAVDSNGIIVNNKQLD  822

Query  176  LDREQASSYRLVVSGADKDGEGLSTQCECNIKVKDVNDNFPMFRDSQYSARIEENILSSE  235
             D   A  Y  VV+  DK     +       +  K+ ND  P F    Y+  ++EN   +
Sbjct  823  ADNNNA-YYEFVVTAKDKGEPAKTGTATVRVYTKNKNDEEPKFSQQVYTPNVDENAGPNT  881

Query  236  LL  237
            L+
Sbjct  882  LV  883

>ACON2_034794.P62462 | transcript=ACON2_034794.R62462 | gene=ACON2_034794
| organism=Anopheles_coluzzii_AcolN3 | gene_product=neural-cadherin,
transcript variant X15, neural-cadherin,
transcript variant X13, neural-cadherin, transcript variant
X8, neural-cadherin, transcript variant X1, neural-cadherin,
transcript variant X3, neural-cadherin, transcript variant
X10, neural-cadherin, transcript variant X16, neural-cadherin,
transcript variant X2, neural-cadherin, transcript variant
X12, neural-cadherin, transcript variant X4, neural-cadherin,
transcript variant X7, neural-cadherin, transcript variant
X9, neural-cadherin, transcript variant X5, neural-cadherin,
transcript variant X17, neural-cadherin, transcript variant
X14, neural-cadherin, transcript variant X11, neural-cadherin,
transcript variant X6 | transcript_product=neural-cadherin,
transcript variant X16 | location=AcolN3_3RL:40912217-41119468(+)
| protein_length=3098 | sequence_SO=chromosome
| SO=protein_coding_gene | is_pseudo=false
Length=3098

 Score = 147 bits (371),  Expect = 2e-35, Method: Compositional matrix adjust.
 Identities = 128/446 (29%), Positives = 217/446 (49%), Gaps = 36/446 (8%)

Query  20    PIAKITS---DYQATQKITYRISGVGID--QPPFGIFVVDKNTGDINITAIVDREET---  71
             PI ++T+   D    Q I Y ++G GID   P    F +++ TG+I +   +DR++
Sbjct  1539  PIMQVTATDGDKDRPQNIVYFLTGQGIDPDNPANSKFDINRTTGEIFVLKPLDRDQPNGR  1598

Query  72    PSFLITCRALNAQGLDVEKPLILTVKILDINDNPPVFSQQIFMGEIEENSASNSLVMILN  131
             P +  T  A +  G  +     + V + DINDN P+F Q ++ G + EN  +  +VM +
Sbjct  1599  PQWRFTVFAQDEGGEGLVGYADVQVNLKDINDNAPIFPQGVYFGNVTENGTAGMVVMTMT  1658

Query  132   ATDADEPNH-LNSKIAFKI---VSQEPAGTPMFLLSRNTGEVRTLTNSLDREQASSYRLV  187
             A D D+PN   N+K+ + I   V +E  G+P+F +  +TG ++T    LDRE+   Y +
Sbjct  1659  AVDYDDPNEGTNAKLIYSIEKNVIEEETGSPIFEIEADTGVIKTAVCCLDRERTPDYSIQ  1718

Query  188   VSGADKDGEGLSTQCECNIKVKDVNDNFPMFRDSQYSARIEENILSS--ELLRFQVTDLD  245
             V     DG GL      +I+VKD+ND  P F   ++   ++E   ++  E+    VT  D
Sbjct  1719  VVAM--DGGGLKGTGTASIRVKDINDMPPQFTKDEWFTEVDETDGTNLPEMPILTVTVHD  1776

Query  246   EEYTDNWLAVYFFTSGNEGNWFEIQTDPRTNEGI--LKVVKALDYE---QLQSVKLSIAV  300
             E+ T+ +        SG    + F +    R N+G   LK+V+ LDYE   Q    +  I V
Sbjct  1777  EDETNKFQYKVIDNSGYGADKFTMV---RNNDGTGSLKIVQPLDYEDPLQSNGFRFRIQV  1833

Query  301   KNKAEFHQSVISRYRVQSTPVTIQVINVREGIAFRPASKTFTVQKGISSKKLVDYILGTY  360
              +K E + +   +Y V  + V +++ ++ +    +P  +   ++  +      V   L T+
Sbjct  1834  NDKGEDNDN--DKYHVAYSWVVVKLRDINDN---KPQFERPNIEVSVYENADVGKTLETF  1888

Query  361   QAIDEDTNKAASNVKYVMGR-NDGGYLMIDSKTAEIKFVKNMNRDSTFIVNKTITAEVLA  419
             +A D D     S V Y + R +D       ++    +   +N++R+    V       ++LA
Sbjct  1889  KATDPDQG-GKSKVSYAIDRSSDRQRQFSINQEGTVTIQRNLDRE----VTPRHQVKILA  1943

Query  420   IDE-YTGKTSTGTVYVRVPDFNDNCP  444
             ID+    KT+T T+ V V D NDN P
Sbjct  1944  IDDGIPPKTATATLTVIVQDINDNPP  1969
```

```
 Score = 79.0 bits (193),  Expect = 2e-13, Method: Compositional matrix adjust.
 Identities = 106/424 (25%), Positives = 180/424 (42%), Gaps = 51/424 (12%)

Query  53    VDKNTGDINITAIVDREETPSFLITCRALNAQGLDVEKPLILTVKILDINDNPPVFSQ-Q  111
             +D++  ++N+TA+ D         +C  +N          ++ V I D+NDN PVF
Sbjct  928   LDRDKYELNVTAMDDG--------SC-CVNGDATIHTSTAVVVVFITDVNDNKPVFKDCG  978

Query  112   IFMGEIEENSASNSLVMILNATDADEPNHLNSKIAFKIVSQEPAGTPMFLLSRNTGEVRT  171
              +  ++EE + + S V+ + ATD D+   +N ++ + IV Q       F +   TGEV T
Sbjct  979   TYYPKVEEGAPNGSPVIKVQATDEDK--GVNGQVKYSIVQQPNQKGTKFTVDEETGEVST  1036

Query  172   LTNSLDRE--QASSYRLVVSGADKDGEGLSTQCECNIKVKDVNDNFPMFRDSQYSARIEE  229
                  DRE        + V   D+    L   C   +++ DVNDN P+F   +Y   +++
Sbjct  1037  -NKVFDREGDDGKFVSVTVKATDQGEPSLEGVCSFTVEITDVNDNPPLFDRQKYVENVKQ  1095

Query  230   NI-LSSELLRFQVTDLDEEYTDNWLAVYFFT---SGNEGNWFEIQTDPRTNEGILKVVKA  285
             +  + + +LR  V+  DE+  +N   VY  +   S N+  +FEIQ  P +   +LK  K
Sbjct  1096  DASIGTNILR--VSASDEDADNNGAIVYTLSAPHSPNDLEYFEIQ--PESGWIVLK--KP  1149

Query  286   LDYEQLQSVKLSIAVKNK------AEFHQSVISRYRVQSTPVTIQV----INVREGIAFR  335
             LD E+    +L +   ++      A+    +    R    P+  +     I++RE +
Sbjct  1150  LDRERY---RLKVYASDRGDPPSTADVDVELDVVDRNNKPPIWDRTIYGPIHIRENVTVG  1206

Query  336   PASKTFTVQKGISSKKLVDYILGTYQAIDEDTNKAASNVKYVMGRNDGGYLMIDSKTAEI  395
                 +     GI     V Y L         TNK   +  Y+  R D G        A+I
Sbjct  1207  TVVTSVKASSGIEGNPTVFYRL--MPGSTAQTNKL--HTFYLQQRPDNG-----DTWADI  1257

Query  396   KFVKNMNRDSTFIVNKTITAEVLAIDEYTGKTSTGTVYVRVPDFNDNCPTAV-LEKDAVC  454
             K    ++ ++    N T+  E     +     S  TVY+++ D ND  P     E++ V
Sbjct  1258  KVNHPLDFETIKEYNLTVRVENNGAQQL---ASEATVYIQLEDVNDEIPLFTEREQETVL  1314

Query  455   SSSP  458
                P
Sbjct  1315  EGEP  1318

 Score = 76.3 bits (186),  Expect = 1e-12, Method: Compositional matrix adjust.
 Identities = 103/418 (25%), Positives = 174/418 (42%), Gaps = 50/418 (12%)

Query  50    IFVVDKNTGDINITAIVDREETPSFLITCRAL----------NAQGLDVEKPLILTVKIL  99
              F ++  +G+I    + DRE+  ++ +   A           N Q   V K   + + I
Sbjct  1355  FFEINLQSGEIFTKVVFDREKQGAYALEVEARDGAPSARPNSNQQPNSVTK--FIRIGIA  1412

Query  100   DINDNPPVFSQQIFMGEIEENSASNSLVMILNATDADEPNHLNSKIAFKIVSQEPAGTPM  159
             D NDNPP F + ++  E++EN      V+ + A D DE    +S+I ++I S    G
Sbjct  1413  DKNDNPPYFDKALYEAEVDENEDIQHTVLTVTAKDHDE----SSRIRYEITSGNIGGA--  1466

Query  160   FLLSRNTGEVRTLTNSLDREQASSYRLVVSGADKDGEGLSTQCECNIKVKDVNDNFPMFR  219
             F +   TG +  +  +LD E    Y L +  +D   E  +      I VKDVND  P F
Sbjct  1467  FAVKNMTGAI-YVAGALDYETRKRYYLTLVASDSLNENETIIV---INVKDVNDMPPSFP  1522

Query  220   DSQYSARIEENI-LSSELLRFQVTDLDEEYTDNWLAVYFFTSG--NEGNWFEIQTDPRTN  276
                Y   ++E + +   +++   TD D++   N   VYF T    +  N    + D
Sbjct  1523  QQVYERTMDEELAVPFPIMQVTATDGDKDRPQN--IVYFLTGQGIDPDNPANSKFDINRT  1580

Query  277   EGILKVVKALDYEQLQS-VKLSIAVKNKAEFHQSVISRYRVQSTPVTIQVINVREGIAFR  335
              G + V+K LD +Q     +    V  + E  + ++    VQ     + + ++ +
Sbjct  1581  TGEIFVLKPLDRDQPNGRPQWRFTVFAQDEGGEGLVGYADVQ-----VNLKDINDNAPIF  1635

Query  336   PASKTFTVQKGISSKKLVDYILGTYQAIDEDTNKAASNVK--YVMGRN------DGGYLM  387
             P    F     ++        ++ T  A+D D     +N K  Y + +N
Sbjct  1636  PQGVYF---GNVTENGTAGMVVMTMTAVDYDDPNEGTNAKLIYSIEKNVIEEETGSPIFE  1692

Query  388   IDSKTAEIK-FVKNMNRDSTFIVNKTITAEVLAIDEYTGKTSTGTVYVRVPDFNDNCP  444
             I++ T  IK  V  ++R+ T       + +V+A+D   G   TGT  +RV D ND  P
Sbjct  1693  IEADTGVIKTAVCCLDRERT----PDYSIQVVAMDG-GGLKGTGTASIRVKDINDMPP  1745

 Score = 65.5 bits (158),  Expect = 3e-09, Method: Compositional matrix adjust.
 Identities = 85/359 (24%), Positives = 153/359 (43%), Gaps = 32/359 (9%)

```

```
Query  51    FVVDKNTGDINITAIVDRE-ETPSFL-ITCRALNAQGLDVEKPLILTVKILDINDNPPVF  108
             F VD+ TG+++   + DRE +   F+ +T +A +     +E     TV+I D+NDNPP+F
Sbjct  1025  FTVDEETGEVSTNKVFDREGDDGKFVSVTVKATDQGEPSLEGVCSFTVEITDVNDNPPLF  1084

Query  109   SQQIFMGEIEENSASNSLVMILNATDADEPNHLNSKIAFKIVS-QEPAGTPMFLLSRNTG  167
              +Q ++  ++++++  + ++ ++A+D D  N  N  I + + +   P     F +   +G
Sbjct  1085  DRQKYVENVKQDASIGTNILRVSASDEDADN--NGAIVYTLSAPHSPNDLEYFEIQPESG  1142

Query  168   EVRTLTNSLDREQASSYRLVVSGADKDGEGLSTQCECNIKVKDVNDNFPMFRDSQYSARI  227
              +  L   LDRE+   YRL V  +D+ G+  ST            +N P   D      I
Sbjct  1143  WI-VLKKPLDRER---YRLKVYASDR-GDPPSTADVDVELDVVDRNNKPPIWDRTIYGPI  1197

Query  228   EENILSSELLRFQVTDLDEEYTDNWLAVYFFTSG-----NEGNWFEIQTDPRTNE--GIL  280
                   +                N    Y    G     N+ + F +Q  P   +    +
Sbjct  1198  HIRENVTVGTVVTSVKASSGIEGNPTVFYRLMPGSTAQTNKLHTFYLQQRPDNGDTWADI  1257

Query  281   KVVKALDYEQLQSVKLSIAVKNKAEFHQSVISRYRVQSTPVTIQVINVREGIAFRPASKT  340
             KV   LD+E ++   L++ V+N         ++       V IQ+ +V + I      +
Sbjct  1258  KVNHPLDFETIKEYNLTVRVENNG-------AQQLASEATVYIQLEDVNDEIPLFTEREQ  1310

Query  341   FTVQKGISSKKLVDYILGTYQAIDEDTNKAASNVKYVM---GRNDG-GYLMIDSKTAEI  395
              TV +G      V  +     AID+D     + V Y +    RN+G  +  I+ ++ EI
Sbjct  1311  ETVLEGEPIGTKVTQV----NAIDKDGTFPNNQVYYYIVDSPRNEGKDFFEINLQSGEI  1365

 Score = 55.8 bits (133),  Expect = 3e-06, Method: Compositional matrix adjust.
 Identities = 49/158 (31%), Positives = 78/158 (49%), Gaps = 24/158 (15%)

Query  102   NDNPPVFSQQIFMGEIEENSASNSLVMILNATDADEPNHLNSKIAFKIVSQEPAGTP--M  159
             ND  P FSQQ++   ++EN+  N+LV  + A+D D  N     + F  V     GT
Sbjct  858   NDEEPKFSQQVYTPNVDENAGPNTLVTTVVASDKDGDN-----VRFGFVG---GGTSSGQ  909

Query  160   FLLSRNTGEVRTLTN--SLDREQASSYRLVVSGADK-------DGEGLSTQCECNIKVKD  210
             F++   TG +R  +   SLDR++   Y L V+  D        D    ++     + + D
Sbjct  910   FVIEEITGVIRLHSKAISLDRDK---YELNVTAMDDGSCCVNGDATIHTSTAVVVVFITD  966

Query  211   VNDNFPMFRD-SQYSARIEENILS-SELLRFQVTDLDE  246
             VNDN P+F+D   Y  ++EE   + S +++ Q TD D+
Sbjct  967   VNDNKPVFKDCGTYYPKVEEGAPNGSPVIKVQATDEDK  1004

 Score = 53.9 bits (128),  Expect = 1e-05, Method: Compositional matrix adjust.
 Identities = 70/306 (23%), Positives = 126/306 (41%), Gaps = 30/306 (10%)

Query  20    PIAKIT-SDYQATQKITYRI---SGVGIDQPPFGIFVVDKNTGDINITAIVDRE---ETP  72
             PI  +T  D   T K  Y++   SG G D+  F +   +  TG + I   +D E   ++
Sbjct  1768  PILTVTVHDEDETNKFQYKVIDNSGYGADK--FTMVRNNDGTGSLKIVQPLDYEDPLQSN  1825

Query  73    SFLITCRALNAQGLDVEKPLI------LTVKILDINDNPPVFSQQIFMGEIEENSASNSL  126
              F    + +N +G D +          + VK+ DINDN P F +      + EN+
Sbjct  1826  GFRFRIQ-VNDKGEDNDNDKYHVAYSWVVVKLRDINDNKPQFERPNIEVSVYENADVGKT  1884

Query  127   VMILNATDADEPNHLNSKIAFKIVSQEPAGTPMFLLSRNTGEVRTLTNSLDREQASSYRL  186
             +    ATD D+     SK+++ I   + +       S N     T+  +LDRE    +++
Sbjct  1885  LETFKATDPDQGG--KSKVSYAI---DRSSDRQRQFSINQEGTVTIQRNLDREVTPRHQV  1939

Query  187   VVSGADKDGEGLSTQCECNIKVKDVNDNFPMFRDSQYSARIEENILSSELLRFQVTDLDE  246
              +   D      +      + V+D+NDN P F    Y   + E++   +++    TD D+
Sbjct  1940  KILAIDDGIPPKTATATLTVIVQDINDNPPKFL-KDYRPVLPEHVPPRKVVEILATDDDD  1998

Query  247   EYTDNWLAVYFFTSGNEGNW----FEIQTDPRTNEG----ILKVVKALDYEQLQSVKLSI  298
                 N     F       +     F+++ D +   G    I+  +++ D EQ +   + I
Sbjct  1999  RSKSNGPPFQFRLDPGADDIIRASFKVEQDQKGANGDGMAIVSSLRSFDREQQKEYLIPI  2058

Query  299   AVKNKA  304
              +K+
Sbjct  2059  VIKDHG  2064

 Score = 52.0 bits (123),  Expect = 4e-05, Method: Compositional matrix adjust.
 Identities = 97/398 (24%), Positives = 163/398 (41%), Gaps = 54/398 (14%)
```

```
Query  106   PVFSQQIFMGEIEENSASNSLVMILNATDADEPNHLNSKIAFKIVSQ-EPAGTPMFLLSR  164
             P F    +  EI EN   +S ++ + A      +  + +I + + +Q + AGT  F +
Sbjct  655   PQFYMPSYEAEIPENQKKDSDIISVKAK-----SFADREIRYTLKAQGQGAGT--FNIGP  707

Query  165   NTGEVRTLTNSLDRE---QASSYRLVVSGADKDGEGLSTQCECNIKVKDVNDNFPMFRDS  221
              +G V+ L   LD E   Q   Y LVV+ A +D  G ST  E  I+V DVNDN P F
Sbjct  708   TSGIVK-LAKELDFEDLRQPHVYSLVVT-ATEDSGGFSTSVELTIRVTDVNDNAPKFELP  765

Query  222   QYSAR-IEENI-LSSELLRFQVTDLDEEYTDNWLAVYFFTSGNEGNWFEIQTDPRTNEGI  279
              Y A  ++E+I L + +L+ +  D D          + +  S +    F + ++     GI
Sbjct  766   DYQAHNVDEDIPLGTSILKVKAMDSDSGAN---AEIEYLVSDDH---FAVDSN-----GI  814

Query  280   LKVVKALDYEQLQS-VKLSIAVKNKAEFHQSVISRYRVQSTPVTIQVINVREGIAFRPAS  338
             +   K LD +   +  +  +  K+K E  ++  +  RV +             E   F
Sbjct  815   IVNNKQLDADNNNAYYEFVVTAKDKGEPAKTGTATVRVYTKNKN------DEEPKFSQQV  868

Query  339   KTFTVQKGISSKKLVDYILGTYQAIDEDTNKAASNVK--YVMGRNDGGYLMIDSKTAEIK  396
              T  V +       LV  ++          ++K   NV+  +V G    G  +I+  T  I+
Sbjct  869   YTPNVDENAGPNTLVTTVVA--------SDKDGDNVRFGFVGGGTSSGQFVIEEITGVIR  920

Query  397   FVK---NMNRDSTFI-VNKTITAEVLAIDEYTGKTSTGTVYVRVPDFNDN------CPTA  446
                   +++RD   + V              + T  TST  V V + D NDN      C T
Sbjct  921   LHSKAISLDRDKYELNVTAMDDGSCCVNGDATIHTSTAVVVVFITDVNDNKPVFKDCGTY  980

Query  447   VLE-KDAVCSSSPSVVVSARTLNNRYTGPYTFALEDQP  483
               + ++   + SP + V A    +    G   +++  QP
Sbjct  981   YPKVEEGAPNGSPVIKVQATDEDKGVNGQVKYSIVQQP  1018

 Score = 44.7 bits (104),  Expect = 0.008, Method: Compositional matrix adjust.
 Identities = 48/199 (24%), Positives = 73/199 (37%), Gaps = 19/199 (10%)

Query  27    DYQATQKITYRISGVGIDQPPFGIFVVDKNTGDINITAIVDREETPSFLITCRALNAQGL  86
             D     K++Y I      Q  F I        G + I   +DRE TP   +    A++
Sbjct  1894  DQGGKSKVSYAIDRSSDRQRQFSI----NQEGTVTIQRNLDREVTPRHQVKILAIDDGIP  1949

Query  87    DVEKPLILTVKILDINDNPPVFSQQIFMGEIEENSASNSLVMILNATDADEPNHLNSKIA  146
                     LTV + DINDNPP F +  +   + E+     +V IL   D D
Sbjct  1950  PKTATATLTVIVQDINDNPPKFLKD-YRPVLPEHVPPRKVVEILATDDDDRSKSNGPPFQ  2008

Query  147   FKIVSQEPAGTPMFLLSRNTGEVRTLTN-----------SLDREQASSYRLVVSGADKDG  195
             F++   +P    +   S    + +   N           S DREQ   Y + +   D
Sbjct  2009  FRL---DPGADDIIRASFKVEQDQKGANGDGMAIVSSLRSFDREQQKEYLIPIVIKDHGN  2065

Query  196   EGLSTQCECNIKVKDVNDN  214
               ++       + + DVNDN
Sbjct  2066  PAMTGTSTLTVVIGDVNDN  2084

 Score = 40.0 bits (92),  Expect = 0.20, Method: Compositional matrix adjust.
 Identities = 89/389 (23%), Positives = 149/389 (38%), Gaps = 62/389 (16%)

Query  75   LITCRALNAQGLDVEKPLILTVKILDINDNPPVFSQQIF--MGEIEENSASNSLVMILNA  132
            +IT +  NA GL       + + + D+ND PP F  +       ++ N+  N+ V  L A
Sbjct  517  IITNQGHNA-GLKYTDNQRVIIHVKDVNDEPPYFINRPLPMQAVVQLNAPPNTPVFTLQA  575

Query  133  TDADEPNHLNSKIAFKIVSQEPAGTPMFLLSRNTGEVRTLTNSLDREQASSYRLVVSGAD  192
             D D      +  I + IV     G   F +   +G VRT    L  +     Y L V   D
Sbjct  576  RDPD----TDHNIHYFIVRDRTGG--RFEVDERSGVVRTRGTDL-FQLDMEYVLYVKAED  628

Query  193  KDGE----GLSTQCECNIKVKDVNDNFPMFRDSQYSARIEEN-ILSSELLRFQVTDLDEE  247
            ++G+         +  E  + +        P F    Y A I EN    S+++  +     +
Sbjct  629  QNGKVDDRRFQSTPEERLSIVG-GKRAPQFYMPSYEAEIPENQKKDSDIISVKA----KS  683

Query  248  YTDNWLAVYFFTSGNEGNWFEIQTDPRTNEGILKVVKALDYEQLQSVKLS----IAVKNK  303
            + D  +        G     F I          GI+K+ K LD+E L+    +       A ++
Sbjct  684  FADREIRYTLKAQGQGAGTFNIGP----TSGIVKLAKELDFEDLRQPHVYSLVVTATEDS  739

Query  304  AEFHQSVISRYRVQSTPVTIQVINVREGIAFRPASKTFTVQKGISSKKLVDYILGT----  359
              F  SV           +TI+V +V +        +  F +     +      D  LGT
```

```
Sbjct  740  GGFSTSV---------ELTIRVTDVNDN------APKFELPDYQAHNVDEDIPLGTSILK  784

Query  360  YQAIDEDTNKAASNVKYVMGRN----DGGYLMIDSKTAEIKFVKNMNRDSTFIVNKTITA  415
             +A+D D+  A + ++Y++  +    D   +++++K  +     N N    F+V
Sbjct  785  VKAMDSDSG-ANAEIEYLVSDDHFAVDSNGIIVNNKQLD---ADNNNAYYEFVVTAKDKG  840

Query  416  EVLAIDEYTGKTSTGTVYVRVPDFNDNCP  444
            E         KT T TV V   + ND  P
Sbjct  841  E-------PAKTGTATVRVYTKNKNDEEP  862

 Score = 39.7 bits (91),  Expect = 0.28, Method: Compositional matrix adjust.
 Identities = 54/242 (22%), Positives = 102/242 (42%), Gaps = 22/242 (9%)

Query  4    KFAKPCREGE--DNSKRNP-IAKITSDYQATQKITYRISGVGIDQPPFGIFVVDKNTGDI  60
            +F  P  E E  +N K++  I  + +   A ++I Y +   G      G F +   +G +
Sbjct  656  QFYMPSYEAEIPENQKKDSDIISVKAKSFADREIRYTLKAQGQGA---GTFNIGPTSGIV  712

Query  61   NITAIVDREE--TPSF--LITCRALNAQGLDVEKPLILTVKILDINDNPPVFSQQIFMGE  116
             +   +D E+   P    L+      ++ G      + LT+++ D+NDN P F    +
Sbjct  713  KLAKELDFEDLRQPHVYSLVVTATEDSGGFSTS--VELTIRVTDVNDNAPKFELPDYQAH  770

Query  117  -IEENSASNSLVMILNATDADEPNHLNSKIAFKIVSQEPAGTPMFLLSRNTGEVRTLTNS  175
             ++E+    + ++ + A D+D  +  N++I + +VS +      F +  N   V
Sbjct  771  NVDEDIPLGTSILKVKAMDSD--SGANAEIEY-LVSDD-----HFAVDSNGIIVNNKQLD  822

Query  176  LDREQASSYRLVVSGADKDGEGLSTQCECNIKVKDVNDNFPMFRDSQYSARIEENILSSE  235
             D   A  Y  VV+  DK      +      +  K+ ND  P F    Y+  ++EN   +
Sbjct  823  ADNNNA-YYEFVVTAKDKGEPAKTGTATVRVYTKNKNDEEPKFSQQVYTPNVDENAGPNT  881

Query  236  LL  237
            L+
Sbjct  882  LV  883

>AAQUA_001764.P1522 | transcript=AAQUA_001764.R1522 | gene=AAQUA_001764
| organism=Anopheles_aquasalis_AaquGF1 | gene_product=neural-cadherin
| transcript_product=neural-cadherin | location=AaquGF1_2RL:61671047-61852165(+)
| protein_length=3119
| sequence_SO=chromosome | SO=protein_coding_gene | is_pseudo=false
Length=3119

 Score = 147 bits (371),  Expect = 2e-35, Method: Compositional matrix adjust.
 Identities = 128/446 (29%), Positives = 217/446 (49%), Gaps = 36/446 (8%)

Query  20    PIAKITS---DYQATQKITYRISGVGID--QPPFGIFVVDKNTGDINITAIVDREET---  71
             PI ++T+   D    Q I Y ++G GID   P    F +++ TG+I +   +DR++
Sbjct  1559  PIMQVTATDGDKDRPQNIVYFLTGQGIDPDNPANSKFDINRTTGEIFVLKPLDRDQPNGR  1618

Query  72    PSFLITCRALNAQGLDVEKPLILTVKILDINDNPPVFSQQIFMGEIEENSASNSLVMILN  131
             P +  T  A +  G  +     + V + DINDN P+F Q ++ G + EN  +  +VM +
Sbjct  1619  PQWRFTVFAQDEGGEGLVGYADVQVNLKDINDNAPIFPQGVYFGNVTENGTAGMVVMTMT  1678

Query  132   ATDADEPNH-LNSKIAFKI---VSQEPAGTPMFLLSRNTGEVRTLTNSLDREQASSYRLV  187
             A D D+PN   N+K+ + I   V +E  G+P+F +  +TG ++T    LDRE+   Y +
Sbjct  1679  AVDYDDPNEGTNAKLIYSIEKNVIEEETGSPIFEIEADTGVIKTAVCCLDRERTPDYSIQ  1738

Query  188   VSGADKDGEGLSTQCECNIKVKDVNDNFPMFRDSQYSARIEENILSS--ELLRFQVTDLD  245
             V      DG GL      +I+VKD+ND  P F   ++   ++E   ++  E+    VT  D
Sbjct  1739  VVAM--DGGGLKGTGTASIRVKDINDMPPQFTKDEWFTEVDETDGTNLPEMPILTVTVHD  1796

Query  246   EEYTDNWLAVYFFTSGNEGNWFEIQTDPRTNEGI--LKVVKALDYE---QLQSVKLSIAV  300
             E+ T+ +        SG   + F +    R N+G   LK+V+ LDYE   Q    +  I V
Sbjct  1797  EDETNKFQYKVIDNSGYGADKFTMV---RNNDGTGSLKIVQPLDYEDPLQSNGFRFRIQV  1853

Query  301   KNKAEFHQSVISRYRVQSTPVTIQVINVREGIAFRPASKTFTVQKGISSKKLVDYILGTY  360
              +K E + +   +Y V  + V +++ ++ +    +P  +   ++  +     V   L T+
Sbjct  1854  NDKGEDNDN--DKYHVAYSWVVVKLRDINDN---KPQFERPNIEVSVYENADVGKTLETF  1908

Query  361   QAIDEDTNKAASNVKYVMGR-NDGGYLMIDSKTAEIKFVKNMNRDSTFIVNKTITAEVLA  419
             +A D D     S V Y + R +D       ++    +  +N++R+    V      ++LA
```

```
Sbjct  1909  KATDPDQG-GKSKVSYAIDRSSDRQRQFSINQEGTVTIQRNLDRE----VTPRHQVKILA  1963

Query  420   IDE-YTGKTSTGTVYYVRVPDFNDNCP  444
             ID+    KT+T T+ V V D NDN P
Sbjct  1964  IDDGIPPKTATATLTVIVQDINDNPP  1989

 Score = 77.0 bits (188),  Expect = 8e-13, Method: Compositional matrix adjust.
 Identities = 103/419 (25%), Positives = 173/419 (41%), Gaps = 51/419 (12%)

Query  50    IFVVDKNTGDINITAIVDREETPSFLITCRAL----------NAQGLDVEKPLILTVKIL  99
              F ++  +G+I    + DRE+  ++ +   A           N Q   V K   + + I
Sbjct  1374  FFEINLQSGEIFTKVVFDREKQGAYALEVEARDGAPSARPNSNQQPNSVTK--FIRIGIA  1431

Query  100   DINDNPPVFSQQIFMGEIEENSASNSLVMILNATDADEPNHLNSKIAFKIVSQEPAGTPM  159
             D NDNPP F + ++  E++EN       V+ + A D DE    +S+I ++I S    G
Sbjct  1432  DKNDNPPYFDKALYEAEVDENEDIQHTVLTVTAKDHDE----SSRIRYEITSGNIGGA--  1485

Query  160   FLLSRNTGEVRTLTNSLDREQASSYRLVVSGADKDGEGLSTQCECNIKVKDVNDNFPMFR  219
             F +   TG +  +  +LD E    Y L +  +D   E    +    I VKDVND  P F
Sbjct  1486  FAVKNMTGAI-YVAGALDYETRKRYHLTLVASDSLNE---NETIIEIHVKDVNDMPPTFS  1541

Query  220   DSQY--SARIEENILSSELLRFQVTDLDEEYTDNWLAVYFFTSG--NEGNWFEIQTDPRT  275
                Y  +   E+ +    +++   TD D++   N   VYF T    +  N    + D
Sbjct  1542  QQIYETTQDEEQTVTDRPIMQVTATDGDKDRPQN--IVYFLTGQGIDPDNPANSKFDINR  1599

Query  276   NEGILKVVKALDYEQLQS-VKLSIAVKNKAEFHQSVISRYRVQSTPVTIQVINVREGIAF  334
              G + V+K LD +Q     +    V  + E  + ++    VQ     + + ++ +
Sbjct  1600  TTGEIFVLKPLDRDQPNGRPQWRFTVFAQDEGGEGLVGYADVQ-----VNLKDINDNAPI  1654

Query  335   RPASKTFTVQKGISSKKLVDYILGTYQAIDEDTNKAASNVK--YVMGRN------DGGYL  386
              P    F     ++        ++ T  A+D D     +N K  Y + +N
Sbjct  1655  FPQGVYF---GNVTENGTAGMVVMTMTAVDYDDPNEGTNAKLIYSIEKNVIEEETGSPIF  1711

Query  387   MIDSKTAEIK-FVKNMNRDSTFIVNKTITAEVLAIDEYTGKTSTGTVYYVRVPDFNDNCP  444
              I++ T  IK  V  ++R+ T        + +V+A+D   G   TGT  +RV D ND  P
Sbjct  1712  EIEADTGVIKTAVCCLDRERT----PDYSIQVVAMDG-GGLKGTGTASIRVKDINDMPP  1765

 Score = 73.9 bits (180),  Expect = 7e-12, Method: Compositional matrix adjust.
 Identities = 90/362 (25%), Positives = 166/362 (46%), Gaps = 37/362 (10%)

Query  51    FVVDKNTGDINITAIVDRE-ETPSFL-ITCRALNAQGLDVEKPLILTVKILDINDNPPVF  108
             F VD+ TG+++    + DRE +   F+ +T +A +     +E     TV+I D+NDNPP+F
Sbjct  1043  FTVDEETGEVSTNKVFDREGDDGKFVSVTVKATDQGEPSLEGVCSFTVEITDVNDNPPLF  1102

Query  109   SQQIFMGEIEENSASNSLVMILNATDADEPNHLNSKIAFKIVS-QEPAGTPMFLLSRNTG  167
              +Q ++  ++++++  + ++ ++A+D D  N  N  I + + +   P     F +   +G
Sbjct  1103  DRQKYVENVKQDASIGTNILRVSASDEDADN--NGAIVYTLSAPHSPNDLEYFEIQPESG  1160

Query  168   EVRTLTNSLDREQASSYRLVVSGADKDGEGLSTQCECNIKVKDVNDNFPMFRDSQYSARI  227
              +  L   LDRE   +Y+L      DK    LS   E  I V D  +N P++ ++      I
Sbjct  1161  WI-VLKKPLDRE---TYKLEAMAQDKGYPPLSRTVEVQIDVVDRANNPPVWDNTGVYGPI  1216

Query  228   --EENI-LSSELLRFQVTDLDEEYTDNWLAVYFFTSG-----NEGNWFEIQTDPRTNE--  277
                +EN+ +  +++  + +   E    N    Y    G     N+ + F +Q  P   +
Sbjct  1217  YVKENMPVGGKVVSIKASSGIE---GNPTVFYRLMPGSTAQTNKLHTFYLQQRPDNGDTW  1273

Query  278   GILKVVKALDYEQLQSVKLSIAVKNKAEFHQSVISRYRVQSTPVTIQVINVREGIAFRPA  337
               +KV   LD+E ++   L+I V+N          ++        V IQ+ +V + I
Sbjct  1274  ADIKVNHPLDFETIKEYNLTIRVENNG-------AQQLASEATVYIQLEDVNDEIPLFTE  1326

Query  338   SKTFTVQKGISSKKLVDYILGTYQAIDEDTNKAASNVKYVM---GRNDG-GYLMIDSKTA  393
              +  TV +G         V  +     AID+D      + V Y +    RN+G  +  I+ ++
Sbjct  1327  REQETVLEGEPIGTKVTQV----NAIDKDGTFPNNQVYYYIVDSPRNEGKDFFEINLQSG  1382

Query  394   EI  395
             EI
Sbjct  1383  EI  1384
```

```
 Score = 55.8 bits (133),  Expect = 3e-06, Method: Compositional matrix adjust.
 Identities = 49/158 (31%), Positives = 78/158 (49%), Gaps = 24/158 (15%)

Query  102   NDNPPVFSQQIFMGEIEENSASNSLVMILNATDADEPNHLNSKIAFKIVSQEPAGTP--M  159
             ND  P FSQQ++   ++EN+  N+LV  + A+D D  N     + F  V     GT
Sbjct  876   NDEEPKFSQQVYTPNVDENAGPNTLVTTVVASDKDGDN-----VRFGFVG---GGTSSGQ  927

Query  160   FLLSRNTGEVRTLTN--SLDREQASSYRLVVSGADK-------DGEGLSTQCECNIKVKD  210
             F++   TG +R  +   SLDR++   Y L V+  D        D    ++     + + D
Sbjct  928   FVIEEITGVIRLHSKAISLDRDK---YELNVTAMDDGSCCVNGDATIHTSTAVVVVFITD  984

Query  211   VNDNFPMFRD-SQYSARIEENILS-SELLRFQVTDLDE  246
             VNDN P+F+D   Y  ++EE   + S +++ Q TD D+
Sbjct  985   VNDNKPVFKDCGTYYPKVEEGAPNGSPVIKVQATDEDK  1022

 Score = 53.9 bits (128),  Expect = 1e-05, Method: Compositional matrix adjust.
 Identities = 70/306 (23%), Positives = 126/306 (41%), Gaps = 30/306 (10%)

Query  20    PIAKIT-SDYQATQKITYRI---SGVGIDQPPFGIFVVDKNTGDINITAIVDRE---ETP  72
             PI  +T  D   T K  Y++   SG G D+  F +   +  TG + I   +D E   ++
Sbjct  1788  PILTVTVHDEDETNKFQYKVIDNSGYGADK--FTMVRNNDGTGSLKIVQPLDYEDPLQSN  1845

Query  73    SFLITCRALNAQGLDVEKPLI------LTVKILDINDNPPVFSQQIFMGEIEENSASNSL  126
              F    + +N +G D +           + VK+ DINDN P F +      + EN+
Sbjct  1846  GFRFRIQ-VNDKGEDNDNDKYHVAYSWVVVKLRDINDNKPQFERPNIEVSVYENADVGKT  1904

Query  127   VMILNATDADEPNHLNSKIAFKIVSQEPAGTPMFLLSRNTGEVRTLTNSLDREQASSYRL  186
             +    ATD D+     SK+++ I   + +       S N     T+  +LDRE    +++
Sbjct  1905  LETFKATDPDQGG--KSKVSYAI---DRSSDRQRQFSINQEGTVTIQRNLDREVTPRHQV  1959

Query  187   VVSGADKDGEGLSTQCECNIKVKDVNDNFPMFRDSQYSARIEENILSSELLRFQVTDLDE  246
              +   D      +      + V+D+NDN P F    Y   + E++   +++    TD D+
Sbjct  1960  KILAIDDGIPPKTATATLTVIVQDINDNPPKFL-KDYRPVLPEHVPPRKVVEILATDDDD  2018

Query  247   EYTDNWLAVYFFTSGNEGNW----FEIQTDPRTNEG----ILKVVKALDYEQLQSVKLSI  298
                 N     F       +     F+++ D +   G    I+  +++ D EQ +   + I
Sbjct  2019  RSKSNGPPFQFRLDPGADDIIRASFKVEQDQKGANGDGMAIVSSLRSFDREQQKEYLIPI  2078

Query  299   AVKNKA  304
              +K+
Sbjct  2079  VIKDHG  2084

 Score = 52.8 bits (125),  Expect = 2e-05, Method: Compositional matrix adjust.
 Identities = 97/398 (24%), Positives = 163/398 (41%), Gaps = 54/398 (14%)

Query  106   PVFSQQIFMGEIEENSASNSLVMILNATDADEPNHLNSKIAFKIVSQ-EPAGTPMFLLSR  164
             P F    +  EI EN   +S ++ + A      +  + +I + + +Q + AGT  F +
Sbjct  673   PQFYMPSYEAEIPENQKKDSDIISVKAK-----SFADREIRYTLKAQGQGAGT--FNIGP  725

Query  165   NTGEVRTLTNSLDRE---QASSYRLVVSGADKDGEGLSTQCECNIKVKDVNDNFPMFRDS  221
              +G V+ L   LD E   Q   Y LVV+ A +D  G ST  E  I+V DVNDN P F
Sbjct  726   TSGIVK-LAKELDFEDLRQPHVYSLVVT-ATEDSGGFSTSVELTIRVTDVNDNAPKFELP  783

Query  222   QYSAR-IEENI-LSSELLRFQVTDLDEEYTDNWLAVYFFTSGNEGNWFEIQTDPRTNEGI  279
              Y A  ++E+I L + +L+ +  D D         + +  S +    F + ++     GI
Sbjct  784   DYQAHNVDEDIPLGTSILKVKAMDADSGAN---AEIEYLVSDDH---FAVDSN-----GI  832

Query  280   LKVVKALDYEQLQS-VKLSIAVKNKAEFHQSVISRYRVQSTPVTIQVINVREGIAFRPAS  338
             +   K LD +   +  +  +  K+K E  ++  +  RV +           E   F
Sbjct  833   IVNNKQLDADNNNAYYEFVVTAKDKGEPAKTGTATVRVYTKNKN------DEEPKFSQQV  886

Query  339   KTFTVQKGISSKKLVDYILGTYQAIDEDTNKAASNVK--YVMGRNDGGYLMIDSKTAEIK  396
              T  V +      LV  ++         ++K   NV+  +V G    G  +I+  T  I+
Sbjct  887   YTPNVDENAGPNTLVTTVVA--------SDKDGDNVRFGFVGGGTSSGQFVIEEITGVIR  938

Query  397   FVK---NMNRDSTFI-VNKTITAEVLAIDEYTGKTSTGTVYVRVPDFNDN------CPTA  446
                   +++RD   + V            + T  TST  V V + D NDN      C T
Sbjct  939   LHSKAISLDRDKYELNVTAMDDGSCCVNGDATIHTSTAVVVVFITDVNDNKPVFKDCGTY  998
```

```
Query  447   VLE-KDAVCSSSPSVVVSARTLNNRYTGPYTFALEDQP  483
               + ++   + SP + V A   +    G    +++ QP
Sbjct  999   YPKVEEGAPNGSPVIKVQATDEDKGVNGQVKYSIVQQP  1036

 Score = 44.3 bits (103),  Expect = 0.009, Method: Compositional matrix adjust.
 Identities = 48/199 (24%), Positives = 73/199 (37%), Gaps = 19/199 (10%)

Query  27    DYQATQKITYRISGVGIDQPPFGIFVVDKNTGDINITAIVDREETPSFLITCRALNAQGL  86
             D     K++Y I      Q  F I       G + I   +DRE TP   +   A++
Sbjct  1914  DQGGKSKVSYAIDRSSDRQRQFSI----NQEGTVTIQRNLDREVTPRHQVKILAIDDGIP  1969

Query  87    DVEKPLILTVKILDINDNPPVFSQQIFMGEIEENSASNSLVMILNATDADEPNHLNSKIA  146
                    LTV + DINDNPP F +  +   + E+     +V IL   D D
Sbjct  1970  PKTATATLTVIVQDINDNPPKFLKD-YRPVLPEHVPPRKVVEILATDDDDRSKSNGPPFQ  2028

Query  147   FKIVSQEPAGTPMFLLSRNTGEVRTLTN-----------SLDREQASSYRLVVSGADKDG  195
             F++   +P    +   S    + +   N           S DREQ   Y + +   D
Sbjct  2029  FRL---DPGADDIIRASFKVEQDQKGANGDGMAIVSSLRSFDREQQKEYLIPIVIKDHGN  2085

Query  196   EGLSTQCECNIKVKDVNDN  214
               ++      + + DVNDN
Sbjct  2086  PAMTGTSTLTVVIGDVNDN  2104

 Score = 40.8 bits (94),  Expect = 0.12, Method: Compositional matrix adjust.
 Identities = 55/242 (23%), Positives = 102/242 (42%), Gaps = 22/242 (9%)

Query  4    KFAKPCREGE--DNSKRNP-IAKITSDYQATQKITYRISGVGIDQPPFGIFVVDKNTGDI  60
            +F  P  E E  +N K++  I  + +   A ++I Y +   G      G F +   +G +
Sbjct  674  QFYMPSYEAEIPENQKKDSDIISVKAKSFADREIRYTLKAQGQGA---GTFNIGPTSGIV  730

Query  61   NITAIVDREE--TPSF--LITCRALNAQGLDVEKPLILTVKILDINDNPPVFSQQIFMGE  116
             +   +D E+   P    L+     ++ G      + LT+++ D+NDN P F    +
Sbjct  731  KLAKELDFEDLRQPHVYSLVVTATEDSGGFSTS--VELTIRVTDVNDNAPKFELPDYQAH  788

Query  117  -IEENSASNSLVMILNATDADEPNHLNSKIAFKIVSQEPAGTPMFLLSRNTGEVRTLTNS  175
             ++E+    + ++ + A DAD  +  N++I + +VS +      F +  N   V
Sbjct  789  NVDEDIPLGTSILKVKAMDAD--SGANAEIEY-LVSDD-----HFAVDSNGIIVNNKQLD  840

Query  176  LDREQASSYRLVVSGADKDGEGLSTQCECNIKVKDVNDNFPMFRDSQYSARIEENILSSE  235
             D   A  Y  VV+  DK     +      +  K+ ND  P F    Y+  ++EN   +
Sbjct  841  ADNNNA-YYEFVVTAKDKGEPAKTGTATVRVYTKNKNDEEPKFSQQVYTPNVDENAGPNT  899

Query  236  LL  237
            L+
Sbjct  900  LV  901

 Score = 39.3 bits (90),  Expect = 0.30, Method: Compositional matrix adjust.
 Identities = 89/389 (23%), Positives = 149/389 (38%), Gaps = 62/389 (16%)

Query  75   LITCRALNAQGLDVEKPLILTVKILDINDNPPVFSQQIF--MGEIEENSASNSLVMILNA  132
            +IT +  NA GL       + + + D+ND PP F  +       ++ N+  N+ V  L A
Sbjct  535  IITNQGHNA-GLKYTDNQRVIIHVKDVNDEPPYFINRPLPMQAVVQLNAPPNTPVFTLQA  593

Query  133  TDADEPNHLNSKIAFKIVSQEPAGTPMFLLSRNTGEVRTLTNSLDREQASSYRLVVSGAD  192
             D D     +  I + IV     G   F +   +G VRT    L  +    Y L V   D
Sbjct  594  RDPD----TDHNIHYFIVRDRTGG--RFEVDERSGVVRTRGTDL-FQLDMEYVLYVKAED  646

Query  193  KDGE----GLSTQCECNIKVKDVNDNFPMFRDSQYSARIEEN-ILSSELLRFQVTDLDEE  247
            ++G+       +  E  + +       P F    Y A I EN    S+++  +     +
Sbjct  647  QNGKVDDRRFQSTPEERLSIVG-GKRAPQFYMPSYEAEIPENQKKDSDIISVKA----KS  701

Query  248  YTDNWLAVYFFTSGNEGNWFEIQTDPRTNEGILKVVKALDYEQLQSVKLS----IAVKNK  303
            + D  +       G     F I        GI+K+ K LD+E L+   +      A ++
Sbjct  702  FADREIRYTLKAQGQGAGTFNIGP----TSGIVKLAKELDFEDLRQPHVYSLVVTATEDS  757

Query  304  AEFHQSVISRYRVQSTPVTIQVINVREGIAFRPASKTFTVQKGISSKKLVDYILGT----  359
              F  SV          +TI+V +V +       +  F +    +     D  LGT
Sbjct  758  GGFSTSV---------ELTIRVTDVNDN------APKFELPDYQAHNVDEDIPLGTSILK  802
```

```
Query  360  YQAIDEDTNKAASNVKYVMGRN----DGGYLMIDSKTAEIKFVKNMNRDSTFIVNKTITA  415
             +A+D D+  A + ++Y++  +    D   +++++K  +     N N    F+V
Sbjct  803  VKAMDADSG-ANAEIEYLVSDDHFAVDSNGIIVNNKQLD---ADNNNAYYEFVVTAKDKG  858

Query  416  EVLAIDEYTGKTSTGTVYVRVPDFNDNCP  444
            E         KT T TV V   + ND  P
Sbjct  859  E-------PAKTGTATVRVYTKNKNDEEP  880

>AARA21_009090.P17043 | transcript=AARA21_009090.R17043 | gene=AARA21_009090
| organism=Anopheles_arabiensis_DONGOLA_2021
| gene_product=neural-cadherin, transcript variant X5 |
transcript_product=neural-cadherin,
transcript variant X8 | location=CM029353:40466514-40666848(+)
| protein_length=3102 | sequence_SO=chromosome
| SO=protein_coding_gene | is_pseudo=false
Length=3102

 Score = 147 bits (371),  Expect = 2e-35, Method: Compositional matrix adjust.
 Identities = 128/446 (29%), Positives = 217/446 (49%), Gaps = 36/446 (8%)

Query  20    PIAKITS---DYQATQKITYRISGVGID--QPPFGIFVVDKNTGDINITAIVDREET---  71
             PI ++T+   D    Q I Y ++G GID   P    F +++ TG+I +   +DR++
Sbjct  1543  PIMQVTATDGDKDRPQNIVYFLTGQGIDPDNPANSKFDINRTTGEIFVLKPLDRDQPNGR  1602

Query  72    PSFLITCRALNAQGLDVEKPLILTVKILDINDNPPVFSQQIFMGEIEENSASNSLVMILN  131
             P +  T  A +  G  +     + V + DINDN P+F Q ++ G + EN  +  +VM +
Sbjct  1603  PQWRFTVFAQDEGGEGLVGYADVQVNLKDINDNAPIFPQGVYFGNVTENGTAGMVVMTMT  1662

Query  132   ATDADEPNH-LNSKIAFKI---VSQEPAGTPMFLLSRNTGEVRTLTNSLDREQASSYRLV  187
             A D D+PN   N+K+ + I   V +E  G+P+F +  +TG ++T    LDRE+   Y +
Sbjct  1663  AVDYDDPNEGTNAKLIYSIEKNVIEEETGSPIFEIEADTGVIKTAVCCLDRERTPDYSIQ  1722

Query  188   VSGADKDGEGLSTQCECNIKVKDVNDNFPMFRDSQYSARIEENILSS--ELLRFQVTDLD  245
             V     DG GL      +I+VKD+ND  P F   ++   ++E   ++  E+    VT  D
Sbjct  1723  VVAM--DGGGLKGTGTASIRVKDINDMPPQFTKDEWFTEVDETDGTNLPEMPILTVTVHD  1780

Query  246   EEYTDNWLAVYFFTSGNEGNWFEIQTDPRTNEGI--LKVVKALDYE---QLQSVKLSIAV  300
             E+ T+ +       SG    + F +    R N+G   LK+V+ LDYE   Q    +  I V
Sbjct  1781  EDETNKFQYKVIDNSGYGADKFTMV---RNNDGTGSLKIVQPLDYEDPLQSNGFRFRIQV  1837

Query  301   KNKAEFHQSVISRYRVQSTPVTIQVINVREGIAFRPASKTFTVQKGISSKKLVDYILGTY  360
              +K E + +   +Y V  + V +++ ++ +    +P  +   ++  +     V   L T+
Sbjct  1838  NDKGEDNDN--DKYHVAYSWVVVKLRDINDN---KPQFERPNIEVSVYENADVGKTLETF  1892

Query  361   QAIDEDTNKAASNVKYVMGR-NDGGYLMIDSKTAEIKFVKNMNRDSTFIVNKTITAEVLA  419
             +A D D     S V Y + R +D       ++   +   +N++R+    V      ++LA
Sbjct  1893  KATDPDQG-GKSKVSYAIDRSSDRQRQFSINQEGTVTIQRNLDRE----VTPRHQVKILA  1947

Query  420   IDE-YTGKTSTGTVYVRVPDFNDNCP  444
             ID+    KT+T T+ V V D NDN P
Sbjct  1948  IDDGIPPKTATATLTVIVQDINDNPP  1973

 Score = 76.3 bits (186),  Expect = 1e-12, Method: Compositional matrix adjust.
 Identities = 103/418 (25%), Positives = 174/418 (42%), Gaps = 50/418 (12%)

Query  50    IFVVDKNTGDINITAIVDREETPSFLITCRAL----------NAQGLDVEKPLILTVKIL  99
              F ++  +G+I    + DRE+  ++ +   A           N Q   V K   + + I
Sbjct  1359  FFEINLQSGEIFTKVVFDREKQGAYALEVEARDGAPSARPNSNQQPNSVTK--FIRIGIA  1416

Query  100   DINDNPPVFSQQIFMGEIEENSASNSLVMILNATDADEPNHLNSKIAFKIVSQEPAGTPM  159
             D NDNPP F + ++  E++EN      V+ + A D DE    +S+I ++I S    G
Sbjct  1417  DKNDNPPYFDKALYEAEVDENEDIQHTVLTVTAKDHDE----SSRIRYEITSGNIGGA--  1470

Query  160   FLLSRNTGEVRTLTNSLDREQASSYRLVVSGADKDGEGLSTQCECNIKVKDVNDNFPMFR  219
             F +   TG +  +  +LD E    Y L +  +D   E  +      I VKDVND  P F
Sbjct  1471  FAVKNMTGAI-YVAGALDYETRKRYYLTLVASDSLNENETIIV---INVKDVNDMPPSFP  1526

Query  220   DSQYSARIEENI-LSSELLRFQVTDLDEEYTDNWLAVYFFTSG--NEGNWFEIQTDPRTN  276
```

```
                Y   ++E + +   +++   TD D++   N   VYF T    +  N    + D
Sbjct  1527  QQVYERTMDEELAVPFPIMQVTATDGDKDRPQN--IVYFLTGQGIDPDNPANSKFDINRT  1584

Query  277   EGILKVVKALDYEQLQS-VKLSIAVKNKAEFHQSVISRYRVQSTPVTIQVINVREGIAFR  335
              G + V+K LD +Q     +    V  + E  + ++    VQ     + + ++ +
Sbjct  1585  TGEIFVLKPLDRDQPNGRPQWRFTVFAQDEGGEGLVGYADVQ-----VNLKDINDNAPIF  1639

Query  336   PASKTFTVQKGISSKKLVDYILGTYQAIDEDTNKAASNVK--YVMGRN------DGGYLM  387
             P    F     ++       ++ T  A+D D     +N K  Y + +N
Sbjct  1640  PQGVYF---GNVTENGTAGMVVMTMTAVDYDDPNEGTNAKLIYSIEKNVIEEETGSPIFE  1696

Query  388   IDSKTAEIK-FVKNMNRDSTFIVNKTITAEVLAIDEYTGKTSTGTVYVRVPDFNDNCP  444
             I++ T  IK  V  ++R+ T       + +V+A+D   G   TGT  +RV D ND  P
Sbjct  1697  IEADTGVIKTAVCCLDRERT----PDYSIQVVAMDG-GGLKGTGTASIRVKDINDMPP  1749

 Score = 76.3 bits (186),  Expect = 2e-12, Method: Compositional matrix adjust.
 Identities = 106/426 (25%), Positives = 181/426 (42%), Gaps = 51/426 (12%)

Query  53    VDKNTGDINITAIVDREETPSFLITCRALNAQGLDVEKPLILTVKILDINDNPPVFSQ-Q  111
             +D++  ++N+TA+ D         +C  +N          ++ V I D+NDN PVF
Sbjct  928   LDRDKYELNVTAMDDG--------SC-CVNGDATIHTSTAVVVVFITDVNDNKPVFKDCG  978

Query  112   IFMGEIEENSASNSLVMILNATDADEPNHLNSKIAFKIVSQEPAGTPMFLLSRNTGEVRT  171
              +  ++EE + + S V+ + ATD D+   +N ++ + IV Q       F +   TGEV T
Sbjct  979   TYYPKVEEGAPNGSPVIKVQATDEDK--GVNGQVKYSIVQQPNQKGTKFTVDEETGEVST  1036

Query  172   LTNSLDRE--QASSYRLVVSGADKDGEGLSTQCECNIKVKDVNDNFPMFRDSQYSARIEE  229
                  DRE        + V   D+    L   C   +++ DVNDN P+F   +Y   +++
Sbjct  1037  -NKVFDREGDDGKFVSVTVKATDQGEPSLEGVCSFTVEITDVNDNPPLFDRQKYVENVKQ  1095

Query  230   NI-LSSELLRFQVTDLDEEYTDNWLAVYFFT---SGNEGNWFEIQTDPRTNEGILKVVKA  285
             +  + + +LR  V+  DE+  +N   VY  +   S N+  +FEIQ  P +   +LK  K
Sbjct  1096  DASIGTNILR--VSASDEDADNNGAIVYTLSAPHSPNDLEYFEIQ--PESGWIVLK--KP  1149

Query  286   LD--YEQLQSVKLSIAVKNK------AEFHQSVISRYRVQSTPVTIQV----INVREGIA  333
             LD  Y + +  +L +   ++      A+    +    R    P+  +     I++RE +
Sbjct  1150  LDGCYSR-ERYRLKVYASDRGDPPSTADVDVELDVVDRNNKPPIWDRTIYGPIHIRENVT  1208

Query  334   FRPASKTFTVQKGISSKKLVDYILGTYQAIDEDTNKAASNVKYVMGRNDGGYLMIDSKTA  393
                   +     GI     V Y L         TNK   +  Y+  R D G        A
Sbjct  1209  VGTVVTSVKASSGIEGNPTVFYRL--MPGSTAQTNKL--HTFYLQQRPDNG-----DTWA  1259

Query  394   EIKFVKNMNRDSTFIVNKTITAEVLAIDEYTGKTSTGTVYVRVPDFNDNCPTAV-LEKDA  452
             +IK    ++ ++    N T+  E     +     S  TVY+++ D ND  P     E++
Sbjct  1260  DIKVNHPLDFETIKEYNLTVRVENNGAQQL---ASEATVYIQLEDVNDEIPLFTEREQET  1316

Query  453   VCSSSP  458
             V    P
Sbjct  1317  VLEGEP  1322

 Score = 60.8 bits (146),  Expect = 9e-08, Method: Compositional matrix adjust.
 Identities = 83/362 (23%), Positives = 152/362 (42%), Gaps = 34/362 (9%)

Query  51    FVVDKNTGDINITAIVDRE-ETPSFL-ITCRALNAQGLDVEKPLILTVKILDINDNPPVF  108
             F VD+ TG+++    + DRE +   F+ +T +A +     +E     TV+I D+NDNPP+F
Sbjct  1025  FTVDEETGEVSTNKVFDREGDDGKFVSVTVKATDQGEPSLEGVCSFTVEITDVNDNPPLF  1084

Query  109   SQQIFMGEIEENSASNSLVMILNATDADEPNHLNSKIAFKIVS-QEPAGTPMFLLSRNTG  167
              +Q ++  ++++++  + ++ ++A+D D  N  N  I + + +   P     F +   +G
Sbjct  1085  DRQKYVENVKQDASIGTNILRVSASDEDADN--NGAIVYTLSAPHSPNDLEYFEIQPESG  1142

Query  168   EV---RTLTNSLDREQASSYRLVVSGADKDGEGLSTQCECNIKVKDVNDNFPMFRDSQYS  224
              +   + L     RE+   YRL V  +D+ G+  ST            +N P   D
Sbjct  1143  WIVLKKPLDGCYSRER---YRLKVYASDR-GDPPSTADVDVELDVVDRNNKPPIWDRTIY  1198

Query  225   ARIEENILSSELLRFQVTDLDEEYTDNWLAVYFFTSG-----NEGNWFEIQTDPRTNE--  277
               I      +                N    Y    G     N+ + F +Q  P   +
Sbjct  1199  GPIHIRENVTVGTVVTSVKASSGIEGNPTVFYRLMPGSTAQTNKLHTFYLQQRPDNGDTW  1258
```

```
Query  278   GILKVVKALDYEQLQSVKLSIAVKNKAEFHQSVISRYRVQSTPVTIQVINVREGIAFRPA  337
               +KV   LD+E ++   L++ V+N         ++       V IQ+ +V + I
Sbjct  1259  ADIKVNHPLDFETIKEYNLTVRVENNG-------AQQLASEATVYIQLEDVNDEIPLFTE  1311

Query  338   SKTFTVQKGISSKKLVDYILGTYQAIDEDTNKAASNVKYVM---GRNDG-GYLMIDSKTA  393
              +  TV +G      V  +     AID+D     + V Y +    RN+G  +  I+ ++
Sbjct  1312  REQETVLEGEPIGTKVTQV----NAIDKDGTFPNNQVYYYIVDSPRNEGKDFFEINLQSG  1367

Query  394   EI  395
             EI
Sbjct  1368  EI  1369

 Score = 55.8 bits (133),  Expect = 3e-06, Method: Compositional matrix adjust.
 Identities = 49/158 (31%), Positives = 78/158 (49%), Gaps = 24/158 (15%)

Query  102   NDNPPVFSQQIFMGEIEENSASNSLVMILNATDADEPNHLNSKIAFKIVSQEPAGTP--M  159
             ND  P FSQQ++   ++EN+  N+LV  + A+D D  N     + F  V     GT
Sbjct  858   NDEEPKFSQQVYTPNVDENAGPNTLVTTVVASDKDGDN-----VRFGFVG---GGTSSGQ  909

Query  160   FLLSRNTGEVRTLTN--SLDREQASSYRLVVSGADK-------DGEGLSTQCECNIKVKD  210
             F++   TG +R  +   SLDR++   Y L V+  D        D    ++      + + D
Sbjct  910   FVIEEITGVIRLHSKAISLDRDK---YELNVTAMDDGSCCVNGDATIHTSTAVVVVFITD  966

Query  211   VNDNFPMFRD-SQYSARIEENILS-SELLRFQVTDLDE  246
             VNDN P+F+D   Y  ++EE   + S +++ Q TD D+
Sbjct  967   VNDNKPVFKDCGTYYPKVEEGAPNGSPVIKVQATDEDK  1004

 Score = 53.9 bits (128),  Expect = 1e-05, Method: Compositional matrix adjust.
 Identities = 70/306 (23%), Positives = 126/306 (41%), Gaps = 30/306 (10%)

Query  20    PIAKIT-SDYQATQKITYRI---SGVGIDQPPFGIFVVDKNTGDINITAIVDRE---ETP  72
             PI  +T  D   T K  Y++   SG G D+  F +   +  TG + I   +D E   ++
Sbjct  1772  PILTVTVHDEDETNKFQYKVIDNSGYGADK--FTMVRNNDGTGSLKIVQPLDYEDPLQSN  1829

Query  73    SFLITCRALNAQGLDVEKPLI------LTVKILDINDNPPVFSQQIFMGEIEENSASNSL  126
              F    + +N +G D +          + VK+ DINDN P F +      + EN+
Sbjct  1830  GFRFRIQ-VNDKGEDNDNDKYHVAYSWVVVKLRDINDNKPQFERPNIEVSVYENADVGKT  1888

Query  127   VMILNATDADEPNHLNSKIAFKIVSQEPAGTPMFLLSRNTGEVRTLTNSLDREQASSYRL  186
             +    ATD D+     SK+++ I   + +       S N     T+  +LDRE    +++
Sbjct  1889  LETFKATDPDQGG--KSKVSYAI---DRSSDRQRQFSINQEGTVTIQRNLDREVTPRHQV  1943

Query  187   VVSGADKDGEGLSTQCECNIKVKDVNDNFPMFRDSQYSARIEENILSSELLRFQVTDLDE  246
              +   D      +      + V+D+NDN P F    Y   + E++   +++    TD D+
Sbjct  1944  KILAIDDGIPPKTATATLTVIVQDINDNPPKFL-KDYRPVLPEHVPPRKVVEILATDDDD  2002

Query  247   EYTDNWLAVYFFTSGNEGNW----FEIQTDPRTNEG----ILKVVKALDYEQLQSVKLSI  298
                 N     F       +     F+++ D +   G    I+  +++ D EQ +   + I
Sbjct  2003  RSKSNGPPFQFRLDPGADDIIRASFKVEQDQKGANGDGMAIVSSLRSFDREQQKEYLIPI  2062

Query  299   AVKNKA  304
              +K+
Sbjct  2063  VIKDHG  2068

 Score = 52.0 bits (123),  Expect = 4e-05, Method: Compositional matrix adjust.
 Identities = 97/398 (24%), Positives = 163/398 (41%), Gaps = 54/398 (14%)

Query  106   PVFSQQIFMGEIEENSASNSLVMILNATDADEPNHLNSKIAFKIVSQ-EPAGTPMFLLSR  164
             P F    +  EI EN   +S ++ + A      +  + +I + + +Q + AGT  F +
Sbjct  655   PQFYMPSYEAEIPENQKKDSDIISVKAK-----SFADREIRYTLKAQGQGAGT--FNIGP  707

Query  165   NTGEVRTLTNSLDRE---QASSYRLVVSGADKDGEGLSTQCECNIKVKDVNDNFPMFRDS  221
              +G V+ L   LD E   Q   Y LVV+ A +D  G ST  E  I+V DVNDN P F
Sbjct  708   TSGIVK-LAKELDFEDLRQPHVYSLVVT-ATEDSGGFSTSVELTIRVTDVNDNAPKFELP  765

Query  222   QYSAR-IEENI-LSSELLRFQVTDLDEEYTDNWLAVYFFTSGNEGNWFEIQTDPRTNEGI  279
              Y A  ++E+I L + +L+ +  D D         + +  S +    F + ++     GI
Sbjct  766   DYQAHNVDEDIPLGTSILKVKAMDSDSGAN---AEIEYLVSDDH---FAVDSN-----GI  814
```

```
Query  280   LKVVKALDYEQLQS-VKLSIAVKNKAEFHQSVISRYRVQSTPVTIQVINVREGIAFRPAS  338
             +   K LD +   +  +  +  K+K E  ++  +  RV +           E   F
Sbjct  815   IVNNKQLDADNNNAYYEFVVTAKDKGEPAKTGTATVRVYTKNKN------DEEPKFSQQV  868

Query  339   KTFTVQKGISSKKLVDYILGTYQAIDEDTNKAASNVK--YVMGRNDGGYLMIDSKTAEIK  396
              T  V +      LV  ++         ++K   NV+  +V G    G  +I+  T  I+
Sbjct  869   YTPNVDENAGPNTLVTTVVA--------SDKDGDNVRFGFVGGGTSSGQFVIEEITGVIR  920

Query  397   FVK---NMNRDSTFI-VNKTITAEVLAIDEYTGKTSTGTVYVRVPDFNDN------CPTA  446
                   +++RD   + V            + T  TST  V V + D NDN      C T
Sbjct  921   LHSKAISLDRDKYELNVTAMDDGSCCVNGDATIHTSTAVVVVFITDVNDNKPVFKDCGTY  980

Query  447   VLE-KDAVCSSSPSVVVSARTLNNRYTGPYTFALEDQP  483
               + ++   + SP + V A   +    G   +++  QP
Sbjct  981   YPKVEEGAPNGSPVIKVQATDEDKGVNGQVKYSIVQQP  1018

 Score = 44.7 bits (104),  Expect = 0.008, Method: Compositional matrix adjust.
 Identities = 48/199 (24%), Positives = 73/199 (37%), Gaps = 19/199 (10%)

Query  27    DYQATQKITYRISGVGIDQPPFGIFVVDKNTGDINITAIVDREETPSFLITCRALNAQGL  86
             D     K++Y I      Q  F I       G + I   +DRE TP   +   A++
Sbjct  1898  DQGGKSKVSYAIDRSSDRQRQFSI----NQEGTVTIQRNLDREVTPRHQVKILAIDDGIP  1953

Query  87    DVEKPLILTVKILDINDNPPVFSQQIFMGEIEENSASNSLVMILNATDADEPNHLNSKIA  146
                    LTV + DINDNPP F +  +   + E+     +V IL   D D
Sbjct  1954  PKTATATLTVIVQDINDNPPKFLKD-YRPVLPEHVPPRKVVEILATDDDDRSKSNGPPFQ  2012

Query  147   FKIVSQEPAGTPMFLLSRNTGEVRTLTN-----------SLDREQASSYRLVVSGADKDG  195
             F++   +P    +   S    + +   N           S DREQ   Y + +   D
Sbjct  2013  FRL---DPGADDIIRASFKVEQDQKGANGDGMAIVSSLRSFDREQQKEYLIPIVIKDHGN  2069

Query  196   EGLSTQCECNIKVKDVNDN  214
               ++      + + DVNDN
Sbjct  2070  PAMTGTSTLTVVIGDVNDN  2088

 Score = 40.0 bits (92),  Expect = 0.21, Method: Compositional matrix adjust.
 Identities = 89/389 (23%), Positives = 149/389 (38%), Gaps = 62/389 (16%)

Query  75   LITCRALNAQGLDVEKPLILTVKILDINDNPPVFSQQIF--MGEIEENSASNSLVMILNA  132
            +IT +  NA GL       + + + D+ND PP F  +       ++ N+  N+ V  L A
Sbjct  517  IITNQGHNA-GLKYTDNQRVIIHVKDVNDEPPYFINRPLPMQAVVQLNAPPNTPVFTLQA  575

Query  133  TDADEPNHLNSKIAFKIVSQEPAGTPMFLLSRNTGEVRTLTNSLDREQASSYRLVVSGAD  192
             D D     +  I + IV     G   F +   +G VRT    L  +    Y L V   D
Sbjct  576  RDPD----TDHNIHYFIVRDRTGG--RFEVDERSGVVRTRGTDL-FQLDMEYVLYVKAED  628

Query  193  KDGE----GLSTQCECNIKVKDVNDNFPMFRDSQYSARIEEN-ILSSELLRFQVTDLDEE  247
            ++G+       +  E  + +       P F    Y A I EN    S+++  +     +
Sbjct  629  QNGKVDDRRFQSTPEERLSIVG-GKRAPQFYMPSYEAEIPENQKKDSDIISVKA----KS  683

Query  248  YTDNWLAVYFFTSGNEGNWFEIQTDPRTNEGILKVVKALDYEQLQSVKLS----IAVKNK  303
            + D  +       G     F I        GI+K+ K LD+E L+   +      A ++
Sbjct  684  FADREIRYTLKAQGQGAGTFNIGP----TSGIVKLAKELDFEDLRQPHVYSLVVTATEDS  739

Query  304  AEFHQSVISRYRVQSTPVTIQVINVREGIAFRPASKTFTVQKGISSKKLVDYILGT----  359
              F  SV          +TI+V +V +       +  F +    +     D  LGT
Sbjct  740  GGFSTSV---------ELTIRVTDVNDN------APKFELPDYQAHNVDEDIPLGTSILK  784

Query  360  YQAIDEDTNKAASNVKYVMGRN----DGGYLMIDSKTAEIKFVKNMNRDSTFIVNKTITA  415
             +A+D D+  A + ++Y++  +    D   +++++K  +     N N    F+V
Sbjct  785  VKAMDSDSG-ANAEIEYLVSDDHFAVDSNGIIVNNKQLD---ADNNNAYYEFVVTAKDKG  840

Query  416  EVLAIDEYTGKTSTGTVYVRVPDFNDNCP  444
            E         KT T TV V   + ND  P
Sbjct  841  E-------PAKTGTATVRVYTKNKNDEEP  862

 Score = 39.7 bits (91),  Expect = 0.28, Method: Compositional matrix adjust.
```

```
 Identities = 54/242 (22%), Positives = 102/242 (42%), Gaps = 22/242 (9%)

Query  4    KFAKPCREGE--DNSKRNP-IAKITSDYQATQKITYRISGVGIDQPPFGIFVVDKNTGDI  60
            +F  P  E E  +N K++  I  + +   A ++I Y +   G      G F +   +G +
Sbjct  656  QFYMPSYEAEIPENQKKDSDIISVKAKSFADREIRYTLKAQGQGA---GTFNIGPTSGIV  712

Query  61   NITAIVDREE--TPSF--LITCRALNAQGLDVEKPLILTVKILDINDNPPVFSQQIFMGE  116
             +   +D E+    P    L+      ++ G       + LT+++ D+NDN P F    +
Sbjct  713  KLAKELDFEDLRQPHVYSLVVTATEDSGGFSTS--VELTIRVTDVNDNAPKFELPDYQAH  770

Query  117  -IEENSASNSLVMILNATDADEPNHLNSKIAFKIVSQEPAGTPMFLLSRNTGEVRTLTNS  175
             ++E+    + ++ + A D+D  +  N++I + +VS +      F +  N   V
Sbjct  771  NVDEDIPLGTSILKVKAMDSD--SGANAEIEY-LVSDD-----HFAVDSNGIIVNNKQLD  822

Query  176  LDREQASSYRLVVSGADKDGEGLSTQCECNIKVKDVNDNFPMFRDSQYSARIEENILSSE  235
             D   A  Y  VV+  DK      +      +  K+ ND  P F    Y+  ++EN   +
Sbjct  823  ADNNNA-YYEFVVTAKDKGEPAKTGTATVRVYTKNKNDEEPKFSQQVYTPNVDENAGPNT  881

Query  236  LL  237
            L+
Sbjct  882  LV  883

>ADAR2_001930.P21645 | transcript=ADAR2_001930.R21645 | gene=ADAR2_001930
| organism=Anopheles_darlingi_AdarGF1 | gene_product=neural-cadherin,
transcript variant X4 | transcript_product=neural-cadherin,
transcript variant X4 | location=AdarGF1_2RL:74991964-75171550(+)
| protein_length=3130 | sequence_SO=chromosome
| SO=protein_coding_gene | is_pseudo=false
Length=3130

 Score = 147 bits (371),  Expect = 2e-35, Method: Compositional matrix adjust.
 Identities = 128/446 (29%), Positives = 217/446 (49%), Gaps = 36/446 (8%)

Query  20    PIAKITS---DYQATQKITYRISGVGID--QPPFGIFVVDKNTGDINITAIVDREET---  71
             PI ++T+    D    Q I Y ++G GID   P    F +++ TG+I +   +DR++
Sbjct  1570  PIMQVTATDGDKDRPQNIVYFLTGQGIDPDNPANSKFDINRTTGEIFVLKPLDRDQPNGR  1629

Query  72    PSFLITCRALNAQGLDVEKPLILTVKILDINDNPPVFSQQIFMGEIEENSASNSLVMILN  131
             P +  T  A +  G  +      + V + DINDN P+F Q ++ G + EN  +  +VM +
Sbjct  1630  PQWRFTVFAQDEGGEGLVGYADVQVNLKDINDNAPIFPQGVYFGNVTENGTAGMVVMTMT  1689

Query  132   ATDADEPNH-LNSKIAFKI---VSQEPAGTPMFLLSRNTGEVRTLTNSLDREQASSYRLV  187
             A D D+PN   N+K+ + I   V +E  G+P+F +  +TG ++T    LDRE+   Y +
Sbjct  1690  AVDYDDPNEGTNAKLIYSIEKNVIEEETGSPIFEIEADTGVIKTAVCCLDRERTPDYSIQ  1749

Query  188   VSGADKDGEGLSTQCECNIKVKDVNDNFPMFRDSQYSARIEENILSS--ELLRFQVTDLD  245
             V      DG GL       +I+VKD+ND  P F    ++    ++E    ++  E+    VT  D
Sbjct  1750  VVAM--DGGGLKGTGTASIRVKDINDMPPQFTKDEWFTEVDETDGTNLPEMPILTVTVHD  1807

Query  246   EEYTDNWLAVYFFTSGNEGNWFEIQTDPRTNEGI--LKVVKALDYE---QLQSVKLSIAV  300
             E+ T+ +        SG    + F +     R N+G   LK+V+ LDYE   Q    +  I V
Sbjct  1808  EDETNKFQYKVIDNSGYGADKFTMV---RNNDGTGSLKIVQPLDYEDPLQSNGFRFRIQV  1864

Query  301   KNKAEFHQSVISRYRVQSTPVTIQVINVREGIAFRPASKTFTVQKGISSKKLVDYILGTY  360
              +K E + +   +Y V  + V +++ ++ +    +P  +   ++  +      V   L T+
Sbjct  1865  NDKGEDNDN--DKYHVAYSWVVVKLRDINDN---KPQFERPNIEVSVYENADVGKTLETF  1919

Query  361   QAIDEDTNKAASNVKYVMGR-NDGGYLMIDSKTAEIKFVKNMNRDSTFIVNKTITAEVLA  419
             +A D D      S V Y + R +D        ++   +   +N++R+    V       ++LA
Sbjct  1920  KATDPDQG-GKSKVSYAIDRSSDRQRQFSINQEGTVTIQRNLDRE----VTPRHQVKILA  1974

Query  420   IDE-YTGKTSTGTVYVRVPDFNDNCP  444
             ID+    KT+T T+ V V D NDN P
Sbjct  1975  IDDGIPPKTATATLTVIVQDINDNPP  2000

 Score = 77.4 bits (189),  Expect = 6e-13, Method: Compositional matrix adjust.
 Identities = 103/419 (25%), Positives = 173/419 (41%), Gaps = 51/419 (12%)

Query  50    IFVVDKNTGDINITAIVDREETPSFLITCRAL----------NAQGLDVEKPLILTVKIL  99
```

```
                 F ++  +G+I    + DRE+  ++ +   A           N Q   V K   + + I
Sbjct  1385  FFEINLQSGEIFTKVVFDREKQGAYALEVEARDGAPSARPNSNQQPNSVTK--FIRIGIA  1442

Query  100   DINDNPPVFSQQIFMGEIEENSASNSLVMILNATDADEPNHLNSKIAFKIVSQEPAGTPM  159
             D NDNPP F + ++  E++EN      V+ + A D DE    +S+I ++I S    G
Sbjct  1443  DKNDNPPYFDKALYEAEVDENEDIQHTVLTVTAKDHDE----SSRIRYEITSGNIGGA--  1496

Query  160   FLLSRNTGEVRTLTNSLDREQASSYRLVVSGADKDGEGLSTQCECNIKVKDVNDNFPMFR  219
             F +   TG +  +  +LD E    Y L +  +D   E    +    I VKDVND  P F
Sbjct  1497  FAVKNMTGAI-YVAGALDYETRKRYHLTLVASDSLNE---NETIIEIHVKDVNDMPPTFS  1552

Query  220   DSQY--SARIEENILSSELLRFQVTDLDEEYTDNWLAVYFFTSG--NEGNWFEIQTDPRT  275
                Y  +   E+ +    +++   TD D++   N   VYF T    +  N    + D
Sbjct  1553  QQVYETTQDEEQTVTDRPIMQVTATDGDKDRPQN--IVYFLTGQGIDPDNPANSKFDINR  1610

Query  276   NEGILKVVKALDYEQLQS-VKLSIAVKNKAEFHQSVISRYRVQSTPVTIQVINVREGIAF  334
               G + V+K LD +Q     +    V  + E  + ++    VQ     + + ++ +
Sbjct  1611  TTGEIFVLKPLDRDQPNGRPQWRFTVFAQDEGGEGLVGYADVQ-----VNLKDINDNAPI  1665

Query  335   RPASKTFTVQKGISSKKLVDYILGTYQAIDEDTNKAASNVK--YVMGRN------DGGYL  386
              P    F     ++        ++ T  A+D D     +N K  Y + +N
Sbjct  1666  FPQGVYF---GNVTENGTAGMVVMTMTAVDYDDPNEGTNAKLIYSIEKNVIEEETGSPIF  1722

Query  387   MIDSKTAEIK-FVKNMNRDSTFIVNKTITAEVLAIDEYTGKTSTGTVYYVRVPDFNDNCP  444
              I++ T  IK  V  ++R+ T        + +V+A+D   G   TGT  +RV D ND  P
Sbjct  1723  EIEADTGVIKTAVCCLDRERT----PDYSIQVVAMDG-GGLKGTGTASIRVKDINDMPP  1776

 Score = 73.9 bits (180),  Expect = 6e-12, Method: Compositional matrix adjust.
 Identities = 90/362 (25%), Positives = 166/362 (46%), Gaps = 37/362 (10%)

Query  51    FVVDKNTGDINITAIVDRE-ETPSFL-ITCRALNAQGLDVEKPLILTVKILDINDNPPVF  108
             F VD+ TG+++   + DRE +   F+ +T +A +     +E     TV+I D+NDNPP+F
Sbjct  1054  FTVDEETGEVSTNKVFDREGDDGKFVSVTVKATDQGEPSLEGVCSFTVEITDVNDNPPLF  1113

Query  109   SQQIFMGEIEENSASNSLVMILNATDADEPNHLNSKIAFKIVS-QEPAGTPMFLLSRNTG  167
              +Q ++  ++++++  + ++ ++A+D D  N  N  I + + +  P     F +   +G
Sbjct  1114  DRQKYVENVKQDASIGTNILRVSASDEDADN--NGAIVYTLSAPHSPNDLEYFEIQPESG  1171

Query  168   EVRTLTNSLDREQASSYRLVVSGADKDGEGLSTQCECNIKVKDVNDNFPMFRDSQYSARI  227
              +  L   LDRE   +Y+L      DK    LS   E  I V D  +N P++ ++     I
Sbjct  1172  WI-VLKKPLDRE---TYKLEAMAQDKGYPPLSRTVEVQIDVVDRANNPPVWDNTGVYGPI  1227

Query  228   --EENI-LSSELLRFQVTDLDEEYTDNWLAVYFFTSG-----NEGNWFEIQTDPRTNE--  277
               +EN+ +  +++  + +   E    N    Y    G     N+ + F +Q   P   +
Sbjct  1228  YVKENMPVGGKVVSIKASSGIE---GNPTVFYRLMPGSTAQTNKLHTFYLQQRPDNGDTW  1284

Query  278   GILKVVKALDYEQLQSVKLSIAVKNKAEFHQSVISRYRVQSTPVTIQVINVREGIAFRPA  337
               +KV   LD+E ++   L+I V+N          ++       V IQ+ +V + I
Sbjct  1285  ADIKVNHPLDFETIKEYNLTIRVENNG-------AQQLASEATVYIQLEDVNDEIPLFTE  1337

Query  338   SKTFTVQKGISSKKLVDYILGTYQAIDEDTNKAASNVKYVM---GRNDG-GYLMIDSKTA  393
              +  TV +G      V  +     AID+D     + V Y +    RN+G  +  I+ ++
Sbjct  1338  REQETVLEGEPIGTKVTQV----NAIDKDGTFPNNQVYYYIVDSPRNEGKDFFEINLQSG  1393

Query  394   EI  395
             EI
Sbjct  1394  EI  1395

 Score = 55.8 bits (133),  Expect = 3e-06, Method: Compositional matrix adjust.
 Identities = 49/158 (31%), Positives = 78/158 (49%), Gaps = 24/158 (15%)

Query  102   NDNPPVFSQQIFMGEIEENSASNSLVMILNATDADEPNHLNSKIAFKIVSQEPAGTP--M  159
             ND  P FSQQ++   ++EN+  N+LV  + A+D D  N      + F  V     GT
Sbjct  887   NDEEPKFSQQVYTPNVDENAGPNTLVTTVVASDKDGDN-----VRFGFVG---GGTSSGQ  938

Query  160   FLLSRNTGEVRTLTN--SLDREQASSYRLVVSGADK-------DGEGLSTQCECNIKVKD  210
             F++    TG +R  +   SLDR++    Y L V+  D         D    ++     + + D
Sbjct  939   FVIEEITGVIRLHSKAISLDRDK---YELNVTAMDDGSCCVNGDATIHTSTAVVVVFITD  995
```

```
Query  211   VNDNFPMFRD-SQYSARIEENILS-SELLRFQVTDLDE  246
             VNDN P+F+D   Y  ++EE   + S +++ Q TD D+
Sbjct  996   VNDNKPVFKDCGTYYPKVEEGAPNGSPVIKVQATDEDK  1033

 Score = 53.9 bits (128),  Expect = 1e-05, Method: Compositional matrix adjust.
 Identities = 70/306 (23%), Positives = 126/306 (41%), Gaps = 30/306 (10%)

Query  20    PIAKIT-SDYQATQKITYRI---SGVGIDQPPFGIFVVDKNTGDINITAIVDRE---ETP  72
             PI  +T  D   T K  Y++   SG G D+  F +   +  TG + I   +D E   ++
Sbjct  1799  PILTVTVHDEDETNKFQYKVIDNSGYGADK--FTMVRNNDGTGSLKIVQPLDYEDPLQSN  1856

Query  73    SFLITCRALNAQGLDVEKPLI------LTVKILDINDNPPVFSQQIFMGEIEENSASNSL  126
              F    + +N +G D +          + VK+ DINDN P F +      + EN+
Sbjct  1857  GFRFRIQ-VNDKGEDNDNDKYHVAYSWVVVKLRDINDNKPQFERPNIEVSVYENADVGKT  1915

Query  127   VMILNATDADEPNHLNSKIAFKIVSQEPAGTPMFLLSRNTGEVRTLTNSLDREQASSYRL  186
             +    ATD D+     SK+++ I   + +       S N     T+  +LDRE    +++
Sbjct  1916  LETFKATDPDQGG--KSKVSYAI---DRSSDRQRQFSINQEGTVTIQRNLDREVTPRHQV  1970

Query  187   VVSGADKDGEGLSTQCECNIKVKDVNDNFPMFRDSQYSARIEENILSSELLRFQVTDLDE  246
              +   D      +      + V+D+NDN P F    Y   + E++   +++    TD D+
Sbjct  1971  KILAIDDGIPPKTATATLTVIVQDINDNPPKFL-KDYRPVLPEHVPPRKVVEILATDDDD  2029

Query  247   EYTDNWLAVYFFTSGNEGNW----FEIQTDPRTNEG----ILKVVKALDYEQLQSVKLSI  298
                 N     F       +     F+++ D +   G    I+  +++ D EQ +   + I
Sbjct  2030  RSKSNGPPFQFRLDPGADDIIRASFKVEQDQKGANGDGMAIVSSLRSFDREQQKEYLIPI  2089

Query  299   AVKNKA  304
              +K+
Sbjct  2090  VIKDHG  2095

 Score = 52.8 bits (125),  Expect = 2e-05, Method: Compositional matrix adjust.
 Identities = 97/398 (24%), Positives = 163/398 (41%), Gaps = 54/398 (14%)

Query  106   PVFSQQIFMGEIEENSASNSLVMILNATDADEPNHLNSKIAFKIVSQ-EPAGTPMFLLSR  164
             P F    +  EI EN   +S ++ + A      +  + +I + + +Q + AGT  F +
Sbjct  684   PQFYMPSYEAEIPENQKKDSDIISVKAK-----SFADREIRYTLKAQGQGAGT--FNIGP  736

Query  165   NTGEVRTLTNSLDRE---QASSYRLVVSGADKDGEGLSTQCECNIKVKDVNDNFPMFRDS  221
              +G V+ L   LD E   Q   Y LVV+ A +D  G ST  E  I+V DVNDN P F
Sbjct  737   TSGIVK-LAKELDFEDLRQPHVYSLVVT-ATEDSGGFSTSVELTIRVTDVNDNAPKFELP  794

Query  222   QYSAR-IEENI-LSSELLRFQVTDLDEEYTDNWLAVYFFTSGNEGNWFEIQTDPRTNEGI  279
              Y A  ++E+I L + +L+ +  D D         + +  S +    F + ++     GI
Sbjct  795   DYQAHNVDEDIPLGTSILKVKAMDADSGAN---AEIEYLVSDDH---FAVDSN-----GI  843

Query  280   LKVVKALDYEQLQS-VKLSIAVKNKAEFHQSVISRYRVQSTPVTIQVINVREGIAFRPAS  338
             +   K LD +   +  +  +  K+K E  ++  +  RV +           E   F
Sbjct  844   IVNNKQLDADNNNAYYEFVVTAKDKGEPAKTGTATVRVYTKNKN------DEEPKFSQQV  897

Query  339   KTFTVQKGISSKKLVDYILGTYQAIDEDTNKAASNVK--YVMGRNDGGYLMIDSKTAEIK  396
              T  V +      LV  ++         ++K   NV+  +V G    G  +I+  T  I+
Sbjct  898   YTPNVDENAGPNTLVTTVVA--------SDKDGDNVRFGFVGGGTSSGQFVIEEITGVIR  949

Query  397   FVK---NMNRDSTFI-VNKTITAEVLAIDEYTGKTSTGTVYVRVPDFNDN------CPTA  446
                   +++RD   + V            + T  TST  V V + D NDN      C T
Sbjct  950   LHSKAISLDRDKYELNVTAMDDGSCCVNGDATIHTSTAVVVVFITDVNDNKPVFKDCGTY  1009

Query  447   VLE-KDAVCSSSPSVVVSARTLNNRYTGPYTFALEDQP  483
               + ++   + SP + V A   +    G   +++  QP
Sbjct  1010  YPKVEEGAPNGSPVIKVQATDEDKGVNGQVKYSIVQQP  1047

 Score = 44.7 bits (104),  Expect = 0.008, Method: Compositional matrix adjust.
 Identities = 48/199 (24%), Positives = 73/199 (37%), Gaps = 19/199 (10%)

Query  27    DYQATQKITYRISGVGIDQPPFGIFVVDKNTGDINITAIVDREETPSFLITCRALNAQGL  86
             D     K++Y I      Q  F I       G + I   +DRE TP   +   A++
Sbjct  1925  DQGGKSKVSYAIDRSSDRQRQFSI----NQEGTVTIQRNLDREVTPRHQVKILAIDDGIP  1980
```

```
Query  87    DVEKPLILTVKILDINDNPPVFSQQIFMGEIEENSASNSLVMILNATDADEPNHLNSKIA  146
                    LTV + DINDNPP F +  +   + E+     +V IL   D D
Sbjct  1981  PKTATATLTVIVQDINDNPPKFLKD-YRPVLPEHVPPRKVVEILATDDDDRSKSNGPPFQ  2039

Query  147   FKIVSQEPAGTPMFLLSRNTGEVRTLTN-----------SLDREQASSYRLVVSGADKDG  195
             F++   +P    +   S    + +   N           S DREQ   Y + +   D
Sbjct  2040  FRL---DPGADDIIRASFKVEQDQKGANGDGMAIVSSLRSFDREQQKEYLIPIVIKDHGN  2096

Query  196   EGLSTQCECNIKVKDVNDN  214
               ++      + + DVNDN
Sbjct  2097  PAMTGTSTLTVVIGDVNDN  2115

 Score = 40.8 bits (94),  Expect = 0.11, Method: Compositional matrix adjust.
 Identities = 55/242 (23%), Positives = 102/242 (42%), Gaps = 22/242 (9%)

Query  4    KFAKPCREGE--DNSKRNP-IAKITSDYQATQKITYRISGVGIDQPPFGIFVVDKNTGDI  60
            +F  P  E E  +N K++  I  + +   A ++I Y +   G      G F +   +G +
Sbjct  685  QFYMPSYEAEIPENQKKDSDIISVKAKSFADREIRYTLKAQGQGA---GTFNIGPTSGIV  741

Query  61   NITAIVDREE--TPSF--LITCRALNAQGLDVEKPLILTVKILDINDNPPVFSQQIFMGE  116
             +   +D E+   P    L+     ++ G      + LT+++ D+NDN P F    +
Sbjct  742  KLAKELDFEDLRQPHVYSLVVTATEDSGGFSTS--VELTIRVTDVNDNAPKFELPDYQAH  799

Query  117  -IEENSASNSLVMILNATDADEPNHLNSKIAFKIVSQEPAGTPMFLLSRNTGEVRTLTNS  175
             ++E+    + ++ + A DAD  +  N++I + +VS +      F +  N   V
Sbjct  800  NVDEDIPLGTSILKVKAMDAD--SGANAEIEY-LVSDD-----HFAVDSNGIIVNNKQLD  851

Query  176  LDREQASSYRLVVSGADKDGEGLSTQCECNIKVKDVNDNFPMFRDSQYSARIEENILSSE  235
             D   A  Y  VV+  DK     +      +  K+ ND  P F    Y+  ++EN   +
Sbjct  852  ADNNNA-YYEFVVTAKDKGEPAKTGTATVRVYTKNKNDEEPKFSQQVYTPNVDENAGPNT  910

Query  236  LL  237
            L+
Sbjct  911  LV  912

 Score = 39.7 bits (91),  Expect = 0.27, Method: Compositional matrix adjust.
 Identities = 89/389 (23%), Positives = 149/389 (38%), Gaps = 62/389 (16%)

Query  75   LITCRALNAQGLDVEKPLILTVKILDINDNPPVFSQQIF--MGEIEENSASNSLVMILNA  132
            +IT +  NA GL       + + + D+ND PP F  +       ++ N+  N+ V  L A
Sbjct  546  IITNQGHNA-GLKYTDNQRVIIHVKDVNDEPPYFINRPLPMQAVVQLNAPPNTPVFTLQA  604

Query  133  TDADEPNHLNSKIAFKIVSQEPAGTPMFLLSRNTGEVRTLTNSLDREQASSYRLVVSGAD  192
             D D     +  I + IV     G   F +   +G VRT    L  +    Y L V   D
Sbjct  605  RDPD----TDHNIHYFIVRDRTGG--RFEVDERSGVVRTRGTDL-FQLDMEYVLYVKAED  657

Query  193  KDGE----GLSTQCECNIKVKDVNDNFPMFRDSQYSARIEEN-ILSSELLRFQVTDLDEE  247
            ++G+       +  E  + +       P F    Y A I EN    S+++  +     +
Sbjct  658  QNGKVDDRRFQSTPEERLSIVG-GKRAPQFYMPSYEAEIPENQKKDSDIISVKA----KS  712

Query  248  YTDNWLAVYFFTSGNEGNWFEIQTDPRTNEGILKVVKALDYEQLQSVKLS----IAVKNK  303
            + D  +       G     F I        GI+K+ K LD+E L+   +      A ++
Sbjct  713  FADREIRYTLKAQGQGAGTFNIGP----TSGIVKLAKELDFEDLRQPHVYSLVVTATEDS  768

Query  304  AEFHQSVISRYRVQSTPVTIQVINVREGIAFRPASKTFTVQKGISSKKLVDYILGT----  359
              F  SV          +TI+V +V +       +  F +    +     D  LGT
Sbjct  769  GGFSTSV---------ELTIRVTDVNDN------APKFELPDYQAHNVDEDIPLGTSILK  813

Query  360  YQAIDEDTNKAASNVKYVMGRN----DGGYLMIDSKTAEIKFVKNMNRDSTFIVNKTITA  415
             +A+D D+  A + ++Y++  +    D   +++++K  +     N N    F+V
Sbjct  814  VKAMDADSG-ANAEIEYLVSDDHFAVDSNGIIVNNKQLD---ADNNNAYYEFVVTAKDKG  869

Query  416  EVLAIDEYTGKTSTGTVYVRVPDFNDNCP  444
            E         KT T TV V   + ND  P
Sbjct  870  E-------PAKTGTATVRVYTKNKNDEEP  891

>AMEM21_003341.P6434 | transcript=AMEM21_003341.R6434 | gene=AMEM21_003341
```

```
| organism=Anopheles_merus_MAF_2021 | gene_product=neural-cadherin,
transcript variant X1 | transcript_product=neural-cadherin,
transcript variant X18 | location=CM029766:40289848-40498951(+)
| protein_length=3098 | sequence_SO=chromosome
| SO=protein_coding_gene | is_pseudo=false
Length=3098

 Score = 147 bits (371),  Expect = 2e-35, Method: Compositional matrix adjust.
 Identities = 128/446 (29%), Positives = 217/446 (49%), Gaps = 36/446 (8%)

Query  20    PIAKITS---DYQATQKITYRISGVGID--QPPFGIFVVDKNTGDINITAIVDREET---  71
             PI ++T+   D    Q I Y ++G GID   P    F +++ TG+I +    +DR++
Sbjct  1539  PIMQVTATDGDKDRPQNIVYFLTGQGIDPDNPANSKFDINRTTGEIFVLKPLDRDQPNGR  1598

Query  72    PSFLITCRALNAQGLDVEKPLILTVKILDINDNPPVFSQQIFMGEIEENSASNSLVMILN  131
             P +  T  A +  G  +      + V + DINDN P+F Q ++ G + EN  +  +VM +
Sbjct  1599  PQWRFTVFAQDEGGEGLVGYADVQVNLKDINDNAPIFPQGVYFGNVTENGTAGMVVMTMT  1658

Query  132   ATDADEPNH-LNSKIAFKI---VSQEPAGTPMFLLSRNTGEVRTLTNSLDREQASSYRLV  187
             A D D+PN   N+K+ + I   V +E  G+P+F +  +TG ++T    LDRE+   Y +
Sbjct  1659  AVDYDDPNEGTNAKLIYSIEKNVIEEETGSPIFEIEADTGVIKTAVCCLDRERTPDYSIQ  1718

Query  188   VSGADKDGEGLSTQCECNIKVKDVNDNFPMFRDSQYSARIEENILSS--ELLRFQVTDLD  245
             V      DG GL       +I+VKD+ND  P F   ++   ++E   ++  E+    VT  D
Sbjct  1719  VVAM--DGGGLKGTGTASIRVKDINDMPPQFTKDEWFTEVDETDGTNLPEMPILTVTVHD  1776

Query  246   EEYTDNWLAVYFFTSGNEGNWFEIQTDPRTNEGI--LKVVKALDYE---QLQSVKLSIAV  300
             E+ T+ +        SG    + F +    R N+G   LK+V+ LDYE   Q    +  I V
Sbjct  1777  EDETNKFQYKVIDNSGYGADKFTMV---RNNDGTGSLKIVQPLDYEDPLQSNGFRFRIQV  1833

Query  301   KNKAEFHQSVISRYRVQSTPVTIQVINVREGIAFRPASKTFTVQKGISSKKLVDYILGTY  360
              +K E + +   +Y V  + V +++ ++ +    +P  +   ++  +      V   L T+
Sbjct  1834  NDKGEDNDN--DKYHVAYSWVVVKLRDINDN---KPQFERPNIEVSVYENADVGKTLETF  1888

Query  361   QAIDEDTNKAASNVKYVMGR-NDGGYLMIDSKTAEIKFVKNMNRDSTFIVNKTITAEVLA  419
             +A D D     S V Y + R +D        ++   +   +N++R+    V       ++LA
Sbjct  1889  KATDPDQG-GKSKVSYAIDRSSDRQRQFSINQEGTVTIQRNLDRE----VTPRHQVKILA  1943

Query  420   IDE-YTGKTSTGTVYVRVPDFNDNCP  444
             ID+    KT+T T+ V V D NDN P
Sbjct  1944  IDDGIPPKTATATLTVIVQDINDNPP  1969

 Score = 76.6 bits (187),  Expect = 1e-12, Method: Compositional matrix adjust.
 Identities = 89/361 (25%), Positives = 167/361 (46%), Gaps = 36/361 (10%)

Query  51    FVVDKNTGDINITAIVDRE-ETPSFL-ITCRALNAQGLDVEKPLILTVKILDINDNPPVF  108
             F VD+ TG+++    + DRE +   F+ +T +A +      +E      TV+I D+NDNPP+F
Sbjct  1025  FTVDEETGEVSTNKVFDREGDDGKFVSVTVKATDQGEPSLEGVCSFTVEITDVNDNPPLF  1084

Query  109   SQQIFMGEIEENSASNSLVMILNATDADEPNHLNSKIAFKIVS-QEPAGTPMFLLSRNTG  167
              +Q ++  ++++++  + ++ ++A+D D  N  N  I + + +   P     F +   +G
Sbjct  1085  DRQKYVENVKQDASIGTNILRVSASDEDADN--NGAIVYTLSAPHSPNDLEYFEIQPESG  1142

Query  168   EVRTLTNSLDREQASSYRLVVSGADKDGEGLSTQCECNIKVKDVNDNFPMFRDSQYSA-R  226
              +  L   LDRE   +Y+L      DK    LS   E  I V D  +N P++ ++ Y
Sbjct  1143  WI-VLKKPLDRE---TYKLEAMAQDKGYPPLSRTVEVQIDVVDRANNPPVWDNTVYGPIY  1198

Query  227   IEENI-LSSELLRFQVTDLDEEYTDNWLAVYFFTSG-----NEGNWFEIQTDPRTNE--G  278
             ++EN+ +  +++  + +   E    N    Y    G     N+ + F +Q  P   +
Sbjct  1199  VKENMPVGGKVVSIKASSGIE---GNPTVFYRLMPGSTAQTNKLHTFYLQQRPDNGDTWA  1255

Query  279   ILKVVKALDYEQLQSVKLSIAVKNKAEFHQSVISRYRVQSTPVTIQVINVREGIAFRPAS  338
              +KV   LD+E ++   L++ V+N          ++        V IQ+ +V + I
Sbjct  1256  DIKVNHPLDFETIKEYNLTVRVENNG-------AQQLASEATVYIQLEDVNDEIPLFTER  1308

Query  339   KTFTVQKGISSKKLVDYILGTYQAIDEDTNKAASNVKYVM---GRNDG-GYLMIDSKTAE  394
             +  TV +G       V  +      AID+D       + V Y +    RN+G  +  I+ ++ E
Sbjct  1309  EQETVLEGEPIGTKVTQV----NAIDKDGTFPNNQVYYYIVDSPRNEGKDFFEINLQSGE  1364

Query  395   I  395
```

```
               I
Sbjct  1365  I  1365

 Score = 76.3 bits (186),  Expect = 1e-12, Method: Compositional matrix adjust.
 Identities = 103/418 (25%), Positives = 174/418 (42%), Gaps = 50/418 (12%)

Query  50    IFVVDKNTGDINITAIVDREETPSFLITCRAL----------NAQGLDVEKPLILTVKIL  99
              F ++  +G+I    + DRE+  ++ +   A           N Q   V K   + + I
Sbjct  1355  FFEINLQSGEIFTKVVFDREKQGAYALEVEARDGAPSARPNSNQQPNSVTK--FIRIGIA  1412

Query  100   DINDNPPVFSQQIFMGEIEENSASNSLVMILNATDADEPNHLNSKIAFKIVSQEPAGTPM  159
             D NDNPP F + ++  E++EN      V+ + A D DE    +S+I ++I S    G
Sbjct  1413  DKNDNPPYFDKALYEAEVDENEDIQHTVLTVTAKDHDE----SSRIRYEITSGNIGGA--  1466

Query  160   FLLSRNTGEVRTLTNSLDREQASSYRLVVSGADKDGEGLSTQCECNIKVKDVNDNFPMFR  219
             F +   TG +  +  +LD E    Y L +  +D   E  +       I VKDVND  P F
Sbjct  1467  FAVKNMTGAI-YVAGALDYETRKRYYLTLVASDSLNENETIIV---INVKDVNDMPPSFP  1522

Query  220   DSQYSARIEENI-LSSELLRFQVTDLDEEYTDNWLAVYFFTSG--NEGNWFEIQTDPRTN  276
                Y   ++E + +   +++   TD D++   N   VYF T    +  N    + D
Sbjct  1523  QQVYERTMDEELAVPFPIMQVTATDGDKDRPQN--IVYFLTGQGIDPDNPANSKFDINRT  1580

Query  277   EGILKVVKALDYEQLQS-VKLSIAVKNKAEFHQSVISRYRVQSTPVTIQVINVREGIAFR  335
              G + V+K LD +Q      +    V  + E  + ++    VQ     + + ++ +
Sbjct  1581  TGEIFVLKPLDRDQPNGRPQWRFTVFAQDEGGEGLVGYADVQ-----VNLKDINDNAPIF  1635

Query  336   PASKTFTVQKGISSKKLVDYILGTYQAIDEDTNKAASNVK--YVMGRN------DGGYLM  387
             P    F     ++        ++ T  A+D D     +N K  Y + +N
Sbjct  1636  PQGVYF---GNVTENGTAGMVVMTMTAVDYDDPNEGTNAKLIYSIEKNVIEEETGSPIFE  1692

Query  388   IDSKTAEIK-FVKNMNRDSTFIVNKTITAEVLAIDEYTGKTSTGTVYVRVPDFNDNCP  444
             I++ T  IK  V  ++R+ T        + +V+A+D   G   TGT  +RV D ND  P
Sbjct  1693  IEADTGVIKTAVCCLDRERT----PDYSIQVVAMDG-GGLKGTGTASIRVKDINDMPP  1745

 Score = 55.8 bits (133),  Expect = 3e-06, Method: Compositional matrix adjust.
 Identities = 49/158 (31%), Positives = 78/158 (49%), Gaps = 24/158 (15%)

Query  102   NDNPPVFSQQIFMGEIEENSASNSLVMILNATDADEPNHLNSKIAFKIVSQEPAGTP--M  159
             ND  P FSQQ++   ++EN+  N+LV  + A+D D  N     + F  V     GT
Sbjct  858   NDEEPKFSQQVYTPNVDENAGPNTLVTTVVASDKDGDN-----VRFGFVG---GGTSSGQ  909

Query  160   FLLSRNTGEVRTLTN--SLDREQASSYRLVVSGADK-------DGEGLSTQCECNIKVKD  210
             F++   TG +R  +   SLDR++   Y L V+  D         D    ++     + + D
Sbjct  910   FVIEEITGVIRLHSKAISLDRDK---YELNVTAMDDGSCCVNGDATIHTSTAVVVVFITD  966

Query  211   VNDNFPMFRD-SQYSARIEENILS-SELLRFQVTDLDE  246
             VNDN P+F+D   Y  ++EE   + S +++ Q TD D+
Sbjct  967   VNDNKPVFKDCGTYYPKVEEGAPNGSPVIKVQATDEDK  1004

 Score = 53.9 bits (128),  Expect = 1e-05, Method: Compositional matrix adjust.
 Identities = 70/306 (23%), Positives = 126/306 (41%), Gaps = 30/306 (10%)

Query  20    PIAKIT-SDYQATQKITYRI---SGVGIDQPPFGIFVVDKNTGDINITAIVDRE---ETP  72
             PI  +T  D   T K  Y++   SG G D+  F +    +  TG + I   +D E   ++
Sbjct  1768  PILTVTVHDEDETNKFQYKVIDNSGYGADK--FTMVRNNDGTGSLKIVQPLDYEDPLQSN  1825

Query  73    SFLITCRALNAQGLDVEKPLI------LTVKILDINDNPPVFSQQIFMGEIEENSASNSL  126
              F    + +N +G D +            + VK+ DINDN P F +      + EN+
Sbjct  1826  GFRFRIQ-VNDKGEDNDNDKYHVAYSWVVVKLRDINDNKPQFERPNIEVSVYENADVGKT  1884

Query  127   VMILNATDADEPNHLNSKIAFKIVSQEPAGTPMFLLSRNTGEVRTLTNSLDREQASSYRL  186
             +    ATD D+      SK+++ I    + +        S N     T+  +LDRE    +++
Sbjct  1885  LETFKATDPDQGG--KSKVSYAI---DRSSDRQRQFSINQEGTVTIQRNLDREVTPRHQV  1939

Query  187   VVSGADKDGEGLSTQCECNIKVKDVNDNFPMFRDSQYSARIEENILSSELLRFQVTDLDE  246
              +   D      +       + V+D+NDN P F    Y   + E++   +++    TD D+
Sbjct  1940  KILAIDDGIPPKTATATLTVIVQDINDNPPKFL-KDYRPVLPEHVPPRKVVEILATDDDD  1998
```

```
Query  247   EYTDNWLAVYFFTSGNEGNW----FEIQTDPRTNEG----ILKVVKALDYEQLQSVKLSI  298
                 N     F       +     F+++ D +   G    I+  +++ D EQ +   + I
Sbjct  1999  RSKSNGPPFQFRLDPGADDIIRASFKVEQDQKGANGDGMAIVSSLRSFDREQQKEYLIPI  2058

Query  299   AVKNKA  304
              +K+
Sbjct  2059  VIKDHG  2064

 Score = 52.0 bits (123),  Expect = 4e-05, Method: Compositional matrix adjust.
 Identities = 97/398 (24%), Positives = 163/398 (41%), Gaps = 54/398 (14%)

Query  106   PVFSQQIFMGEIEENSASNSLVMILNATDADEPNHLNSKIAFKIVSQ-EPAGTPMFLLSR  164
             P F    +  EI EN   +S ++ + A      +  + +I + + +Q + AGT  F +
Sbjct  655   PQFYMPSYEAEIPENQKKDSDIISVKAK-----SFADREIRYTLKAQGQGAGT--FNIGP  707

Query  165   NTGEVRTLTNSLDRE---QASSYRLVVSGADKDGEGLSTQCECNIKVKDVNDNFPMFRDS  221
              +G V+ L   LD E   Q   Y LVV+ A +D  G ST  E  I+V DVNDN P F
Sbjct  708   TSGIVK-LAKELDFEDLRQPHVYSLVVT-ATEDSGGFSTSVELTIRVTDVNDNAPKFELP  765

Query  222   QYSAR-IEENI-LSSELLRFQVTDLDEEYTDNWLAVYFFTSGNEGNWFEIQTDPRTNEGI  279
              Y A  ++E+I L + +L+ +  D D         + +  S +    F + ++     GI
Sbjct  766   DYQAHNVDEDIPLGTSILKVKAMDSDSGAN---AEIEYLVSDDH---FAVDSN-----GI  814

Query  280   LKVVKALDYEQLQS-VKLSIAVKNKAEFHQSVISRYRVQSTPVTIQVINVREGIAFRPAS  338
             +   K LD +   +  +  K+K E  ++  +  RV +           E   F
Sbjct  815   IVNNKQLDADNNNAYYEFVVTAKDKGEPAKTGTATVRVYTKNKN------DEEPKFSQQV  868

Query  339   KTFTVQKGISSKKLVDYILGTYQAIDEDTNKAASNVK--YVMGRNDGGYLMIDSKTAEIK  396
              T  V +      LV  ++         ++K   NV+  +V G    G  +I+  T  I+
Sbjct  869   YTPNVDENAGPNTLVTTVVA--------SDKDGDNVRFGFVGGGTSSGQFVIEEITGVIR  920

Query  397   FVK---NMNRDSTFI-VNKTITAEVLAIDEYTGKTSTGTVYVRVPDFNDN------CPTA  446
                   +++RD   + V            + T  TST  V V + D NDN      C T
Sbjct  921   LHSKAISLDRDKYELNVTAMDDGSCCVNGDATIHTSTAVVVVFITDVNDNKPVFKDCGTY  980

Query  447   VLE-KDAVCSSSPSVVVSARTLNNRYTGPYTFALEDQP  483
               + ++   + SP + V A   +    G   +++  QP
Sbjct  981   YPKVEEGAPNGSPVIKVQATDEDKGVNGQVKYSIVQQP  1018

 Score = 44.3 bits (103),  Expect = 0.008, Method: Compositional matrix adjust.
 Identities = 48/199 (24%), Positives = 73/199 (37%), Gaps = 19/199 (10%)

Query  27    DYQATQKITYRISGVGIDQPPFGIFVVDKNTGDINITAIVDREETPSFLITCRALNAQGL  86
             D     K++Y I      Q  F I       G + I   +DRE TP   +   A++
Sbjct  1894  DQGGKSKVSYAIDRSSDRQRQFSI----NQEGTVTIQRNLDREVTPRHQVKILAIDDGIP  1949

Query  87    DVEKPLILTVKILDINDNPPVFSQQIFMGEIEENSASNSLVMILNATDADEPNHLNSKIA  146
                    LTV + DINDNPP F +  +   + E+     +V IL   D D
Sbjct  1950  PKTATATLTVIVQDINDNPPKFLKD-YRPVLPEHVPPRKVVEILATDDDDRSKSNGPPFQ  2008

Query  147   FKIVSQEPAGTPMFLLSRNTGEVRTLTN-----------SLDREQASSYRLVVSGADKDG  195
             F++   +P    +   S    + +   N           S DREQ   Y + +   D
Sbjct  2009  FRL---DPGADDIIRASFKVEQDQKGANGDGMAIVSSLRSFDREQQKEYLIPIVIKDHGN  2065

Query  196   EGLSTQCECNIKVKDVNDN  214
               ++      + + DVNDN
Sbjct  2066  PAMTGTSTLTVVIGDVNDN  2084

 Score = 39.7 bits (91),  Expect = 0.24, Method: Compositional matrix adjust.
 Identities = 89/389 (23%), Positives = 149/389 (38%), Gaps = 62/389 (16%)

Query  75   LITCRALNAQGLDVEKPLILTVKILDINDNPPVFSQQIF--MGEIEENSASNSLVMILNA  132
            +IT +  NA GL       + + + D+ND PP F  +       ++ N+  N+ V  L A
Sbjct  517  IITNQGHNA-GLKYTDNQRVIIHVKDVNDEPPYFINRPLPMQAVVQLNAPPNTPVFTLQA  575

Query  133  TDADEPNHLNSKIAFKIVSQEPAGTPMFLLSRNTGEVRTLTNSLDREQASSYRLVVSGAD  192
             D D     +  I + IV     G   F +   +G VRT    L  +    Y L V   D
Sbjct  576  RDPD----TDHNIHYFIVRDRTGG--RFEVDERSGVVRTRGTDL-FQLDMEYVLYVKAED  628
```

```
Query  193  KDGE----GLSTQCECNIKVKDVNDNFPMFRDSQYSARIEEN-ILSSELLRFQVTDLDEE  247
            ++G+       +  E  + +       P F    Y A I EN    S+++  +     +
Sbjct  629  QNGKVDDRRFQSTPEERLSIVG-GKRAPQFYMPSYEAEIPENQKKDSDIISVKA----KS  683

Query  248  YTDNWLAVYFFTSGNEGNWFEIQTDPRTNEGILKVVKALDYEQLQSVKLS----IAVKNK  303
            + D  +       G     F I        GI+K+ K LD+E L+   +      A ++
Sbjct  684  FADREIRYTLKAQGQGAGTFNIGP----TSGIVKLAKELDFEDLRQPHVYSLVVTATEDS  739

Query  304  AEFHQSVISRYRVQSTPVTIQVINVREGIAFRPASKTFTVQKGISSKKLVDYILGT----  359
              F  SV          +TI+V +V +       +  F +    +     D  LGT
Sbjct  740  GGFSTSV---------ELTIRVTDVNDN------APKFELPDYQAHNVDEDIPLGTSILK  784

Query  360  YQAIDEDTNKAASNVKYVMGRN----DGGYLMIDSKTAEIKFVKNMNRDSTFIVNKTITA  415
             +A+D D+  A + ++Y++  +    D   +++++K  +     N N    F+V
Sbjct  785  VKAMDSDSG-ANAEIEYLVSDDHFAVDSNGIIVNNKQLD---ADNNNAYYEFVVTAKDKG  840

Query  416  EVLAIDEYTGKTSTGTVYVRVPDFNDNCP  444
            E         KT T TV V   + ND  P
Sbjct  841  E-------PAKTGTATVRVYTKNKNDEEP  862

 Score = 39.7 bits (91),  Expect = 0.28, Method: Compositional matrix adjust.
 Identities = 53/235 (23%), Positives = 99/235 (42%), Gaps = 22/235 (9%)

Query  4    KFAKPCREGE--DNSKRNP-IAKITSDYQATQKITYRISGVGIDQPPFGIFVVDKNTGDI  60
            +F  P  E E  +N K++  I  + +   A ++I Y +   G      G F +   +G +
Sbjct  656  QFYMPSYEAEIPENQKKDSDIISVKAKSFADREIRYTLKAQGQGA---GTFNIGPTSGIV  712

Query  61   NITAIVDREE--TPSF--LITCRALNAQGLDVEKPLILTVKILDINDNPPVFSQQIFMGE  116
             +   +D E+   P    L+      ++ G      + LT+++ D+NDN P F    +
Sbjct  713  KLAKELDFEDLRQPHVYSLVVTATEDSGGFSTS--VELTIRVTDVNDNAPKFELPDYQAH  770

Query  117  -IEENSASNSLVMILNATDADEPNHLNSKIAFKIVSQEPAGTPMFLLSRNTGEVRTLTNS  175
             ++E+    + ++ + A D+D  +  N++I + +VS +      F +  N   V
Sbjct  771  NVDEDIPLGTSILKVKAMDSD--SGANAEIEY-LVSDD-----HFAVDSNGIIVNNKQLD  822

Query  176  LDREQASSYRLVVSGADKDGEGLSTQCECNIKVKDVNDNFPMFRDSQYSARIEEN  230
             D   A  Y  VV+  DK     +      +  K+ ND  P F    Y+  ++EN
Sbjct  823  ADNNNA-YYEFVVTAKDKGEPAKTGTATVRVYTKNKNDEEPKFSQQVYTPNVDEN  876

>AARA21_009090.P17046 | transcript=AARA21_009090.R17046 | gene=AARA21_009090
| organism=Anopheles_arabiensis_DONGOLA_2021
| gene_product=neural-cadherin, transcript variant X5 |
transcript_product=neural-cadherin,
transcript variant X16 | location=CM029353:40466514-40666848(+)
| protein_length=3098 |
sequence_SO=chromosome | SO=protein_coding_gene | is_pseudo=false
Length=3098

 Score = 147 bits (371),  Expect = 2e-35, Method: Compositional matrix adjust.
 Identities = 128/446 (29%), Positives = 217/446 (49%), Gaps = 36/446 (8%)

Query  20    PIAKITS---DYQATQKITYRISGVGID--QPPFGIFVVDKNTGDINITAIVDREET---  71
             PI ++T+   D    Q I Y ++G GID   P    F +++ TG+I +   +DR++
Sbjct  1539  PIMQVTATDGDKDRPQNIVYFLTGQGIDPDNPANSKFDINRTTGEIFVLKPLDRDQPNGR  1598

Query  72    PSFLITCRALNAQGLDVEKPLILTVKILDINDNPPVFSQQIFMGEIEENSASNSLVMILN  131
             P +  T  A +  G  +     + V + DINDN P+F Q ++ G + EN  +  +VM +
Sbjct  1599  PQWRFTVFAQDEGGEGLVGYADVQVNLKDINDNAPIFPQGVYFGNVTENGTAGMVVMTMT  1658

Query  132   ATDADEPNH-LNSKIAFKI---VSQEPAGTPMFLLSRNTGEVRTLTNSLDREQASSYRLV  187
             A D D+PN   N+K+ + I   V +E  G+P+F +  +TG ++T    LDRE+   Y +
Sbjct  1659  AVDYDDPNEGTNAKLIYSIEKNVIEEETGSPIFEIEADTGVIKTAVCCLDRERTPDYSIQ  1718

Query  188   VSGADKDGEGLSTQCECNIKVKDVNDNFPMFRDSQYSARIEENILSS--ELLRFQVTDLD  245
             V     DG GL      +I+VKD+ND  P F   ++   ++E   ++  E+    VT  D
Sbjct  1719  VVAM--DGGGLKGTGTASIRVKDINDMPPQFTKDEWFTEVDETDGTNLPEMPILTVTVHD  1776

Query  246   EEYTDNWLAVYFFTSGNEGNWFEIQTDPRTNEGI--LKVVKALDYE---QLQSVKLSIAV  300
```

```
             E+ T+ +       SG   + F +    R N+G   LK+V+ LDYE   Q    +  I V
Sbjct  1777  EDETNKFQYKVIDNSGYGADKFTMV---RNNDGTGSLKIVQPLDYEDPLQSNGFRFRIQV  1833

Query  301   KNKAEFHQSVISRYRVQSTPVTIQVINVREGIAFRPASKTFTVQKGISSKKLVDYILGTY  360
              +K E + +   +Y V  + V +++ ++ +    +P  +   ++  +     V   L T+
Sbjct  1834  NDKGEDNDN--DKYHVAYSWVVVKLRDINDN---KPQFERPNIEVSVYENADVGKTLETF  1888

Query  361   QAIDEDTNKAASNVKYVMGR-NDGGYLMIDSKTAEIKFVKNMNRDSTFIVNKTITAEVLA  419
             +A D D     S V Y + R +D       ++   +   +N++R+    V      ++LA
Sbjct  1889  KATDPDQG-GKSKVSYAIDRSSDRQRQFSINQEGTVTIQRNLDRE----VTPRHQVKILA  1943

Query  420   IDE-YTGKTSTGTVYYRVPDFNDNCP  444
             ID+    KT+T T+ V V D NDN P
Sbjct  1944  IDDGIPPKTATATLTVIVQDINDNPP  1969

 Score = 79.0 bits (193),  Expect = 2e-13, Method: Compositional matrix adjust.
 Identities = 106/424 (25%), Positives = 180/424 (42%), Gaps = 51/424 (12%)

Query  53    VDKNTGDINITAIVDREETPSFLITCRALNAQGLDVEKPLILTVKILDINDNPPVFSQ-Q  111
             +D++  ++N+TA+ D          +C  +N           ++ V I D+NDN PVF
Sbjct  928   LDRDKYELNVTAMDDG--------SC-CVNGDATIHTSTAVVVVFITDVNDNKPVFKDCG  978

Query  112   IFMGEIEENSASNSLVMILNATDADEPNHLNSKIAFKIVSQEPAGTPMFLLSRNTGEVRT  171
              +  ++EE + + S V+ + ATD D+   +N ++ + IV Q       F +   TGEV T
Sbjct  979   TYYPKVEEGAPNGSPVIKVQATDEDK--GVNGQVKYSIVQQPNQKGTKFTVDEETGEVST  1036

Query  172   LTNSLDRE--QASSYRLVVSGADKDGEGLSTQCECNIKVKDVNDNFPMFRDSQYSARIEE  229
                  DRE        + V   D+    L   C   +++ DVNDN P+F   +Y   +++
Sbjct  1037  -NKVFDREGDDGKFVSVTVKATDQGEPSLEGVCSFTVEITDVNDNPPLFDRQKYVENVKQ  1095

Query  230   NI-LSSELLRFQVTDLDEEYTDNWLAVYFFT---SGNEGNWFEIQTDPRTNEGILKVVKA  285
             +  + + +LR  V+  DE+  +N   VY  +   S N+  +FEIQ  P +   +LK  K
Sbjct  1096  DASIGTNILR--VSASDEDADNNGAIVYTLSAPHSPNDLEYFEIQ--PESGWIVLK--KP  1149

Query  286   LDYEQLQSVKLSIAVKNK------AEFHQSVISRYRVQSTPVTIQV----INVREGIAFR  335
             LD E+    +L +   ++       A+    +    R    P+  +      I++RE +
Sbjct  1150  LDRERY---RLKVYASDRGDPPSTADVDVELDVVDRNNKPPIWDRTIYGPIHIRENVTVG  1206

Query  336   PASKTFTVQKGISSKKLVDYILGTYQAIDEDTNKAASNVKYVMGRNDGGYLMIDSKTAEI  395
                 +     GI     V Y L          TNK   +  Y+  R D G        A+I
Sbjct  1207  TVVTSVKASSGIEGNPTVFYRL--MPGSTAQTNKL--HTFYLQQRPDNG-----DTWADI  1257

Query  396   KFVKNMNRDSTFIVNKTITAEVLAIDEYTGKTSTGTVYYRVPDFNDNCPTAV-LEKDAVC  454
             K    ++ ++    N T+  E      +      S  TVY+++ D ND  P     E++ V
Sbjct  1258  KVNHPLDFETIKEYNLTVRVENNGAQQL---ASEATVYIQLEDVNDEIPLFTEREQETVL  1314

Query  455   SSSP  458
                P
Sbjct  1315  EGEP  1318

 Score = 75.9 bits (185),  Expect = 2e-12, Method: Compositional matrix adjust.
 Identities = 103/418 (25%), Positives = 174/418 (42%), Gaps = 50/418 (12%)

Query  50    IFVVDKNTGDINITAIVDREETPSFLITCRAL----------NAQGLDVEKPLILTVKIL  99
              F ++  +G+I    + DRE+  ++ +   A           N Q   V K   + + I
Sbjct  1355  FFEINLQSGEIFTKVVFDREKQGAYALEVEARDGAPSARPNSNQQPNSVTK--FIRIGIA  1412

Query  100   DINDNPPVFSQQIFMGEIEENSASNSLVMILNATDADEPNHLNSKIAFKIVSQEPAGTPM  159
             D NDNPP F + ++  E++EN      V+ + A D DE    +S+I ++I S    G
Sbjct  1413  DKNDNPPYFDKALYEAEVDENEDIQHTVLTVTAKDHDE----SSRIRYEITSGNIGGA--  1466

Query  160   FLLSRNTGEVRTLTNSLDREQASSYRLVVSGADKDGEGLSTQCECNIKVKDVNDNFPMFR  219
             F +   TG +  +  +LD E    Y L +  +D   E  +       I VKDVND  P F
Sbjct  1467  FAVKNMTGAI-YVAGALDYETRKRYYLTLVASDSLNENETIIV---INVKDVNDMPPSFP  1522

Query  220   DSQYSARIEENI-LSSELLRFQVTDLDEEYTDNWLAVYFFTSG--NEGNWFEIQTDPRTN  276
                Y   ++E + +   +++   TD D++   N   VYF T    +  N    + D
Sbjct  1523  QQVYERTMDEELAVPFPIMQVTATDGDKDRPQN--IVYFLTGQGIDPDNPANSKFDINRT  1580
```

```
Query  277   EGILKVVKALDYEQLQS-VKLSIAVKNKAEFHQSVISRYRVQSTPVTIQVINVREGIAFR  335
              G + V+K LD +Q     +    V  + E  + ++    VQ     + + ++ +
Sbjct  1581  TGEIFVLKPLDRDQPNGRPQWRFTVFAQDEGGEGLVGYADVQ-----VNLKDINDNAPIF  1635

Query  336   PASKTFTVQKGISSKKLVDYILGTYQAIDEDTNKAASNVK--YVMGRN------DGGYLM  387
             P    F     ++        ++ T  A+D D     +N K  Y + +N
Sbjct  1636  PQGVYF---GNVTENGTAGMVVMTMTAVDYDDPNEGTNAKLIYSIEKNVIEEETGSPIFE  1692

Query  388   IDSKTAEIK-FVKNMNRDSTFIVNKTITAEVLAIDEYTGKTSTGTVYVRVPDFNDNCP  444
             I++ T  IK  V  ++R+ T        + +V+A+D   G   TGT  +RV D ND  P
Sbjct  1693  IEADTGVIKTAVCCLDRERT----PDYSIQVVAMDG-GGLKGTGTASIRVKDINDMPP  1745

 Score = 65.5 bits (158),  Expect = 3e-09, Method: Compositional matrix adjust.
 Identities = 85/359 (24%), Positives = 153/359 (43%), Gaps = 32/359 (9%)

Query  51    FVVDKNTGDINITAIVDRE-ETPSFL-ITCRALNAQGLDVEKPLILTVKILDINDNPPVF  108
             F VD+ TG+++    + DRE +    F+ +T +A +      +E      TV+I D+NDNPP+F
Sbjct  1025  FTVDEETGEVSTNKVFDREGDDGKFVSVTVKATDQGEPSLEGVCSFTVEITDVNDNPPLF  1084

Query  109   SQQIFMGEIEENSASNSLVMILNATDADEPNHLNSKIAFKIVS-QEPAGTPMFLLSRNTG  167
              +Q ++  ++++++  + ++ ++A+D D  N  N  I + + +   P     F +   +G
Sbjct  1085  DRQKYVENVKQDASIGTNILRVSASDEDADN--NGAIVYTLSAPHSPNDLEYFEIQPESG  1142

Query  168   EVRTLTNSLDREQASSYRLVVSGADKDGEGLSTQCECNIKVKDVNDNFPMFRDSQYSARI  227
              +  L   LDRE+   YRL V  +D+ G+  ST              +N P   D      I
Sbjct  1143  WI-VLKKPLDRER---YRLKVYASDR-GDPPSTADVDVELDVVDRNNKPPIWDRTIYGPI  1197

Query  228   EENILSSELLRFQVTDLDEEYTDNWLAVYFFTSG-----NEGNWFEIQTDPRTNE--GIL  280
                   +                 N    Y    G     N+ + F +Q  P   +    +
Sbjct  1198  HIRENVTVGTVVTSVKASSGIEGNPTVFYRLMPGSTAQTNKLHTFYLQQRPDNGDTWADI  1257

Query  281   KVVKALDYEQLQSVKLSIAVKNKAEFHQSVISRYRVQSTPVTIQVINVREGIAFRPASKT  340
             KV   LD+E ++   L++ V+N           ++        V IQ+ +V + I       +
Sbjct  1258  KVNHPLDFETIKEYNLTVRVENNG-------AQQLASEATVYIQLEDVNDEIPLFTEREQ  1310

Query  341   FTVQKGISSKKLVDYILGTYQAIDEDTNKAASNVKYVM---GRNDG-GYLMIDSKTAEI  395
              TV +G       V  +     AID+D     + V Y +    RN+G  +  I+ ++ EI
Sbjct  1311  ETVLEGEPIGTKVTQV----NAIDKDGTFPNNQVYYYIVDSPRNEGKDFFEINLQSGEI  1365

 Score = 55.8 bits (133),  Expect = 3e-06, Method: Compositional matrix adjust.
 Identities = 49/158 (31%), Positives = 78/158 (49%), Gaps = 24/158 (15%)

Query  102   NDNPPVFSQQIFMGEIEENSASNSLVMILNATDADEPNHLNSKIAFKIVSQEPAGTP--M  159
             ND  P FSQQ++    ++EN+  N+LV  + A+D D  N      + F  V     GT
Sbjct  858   NDEEPKFSQQVYTPNVDENAGPNTLVTTVVASDKDGDN-----VRFGFVG---GGTSSGQ  909

Query  160   FLLSRNTGEVRTLTN--SLDREQASSYRLVVSGADK-------DGEGLSTQCECNIKVKD  210
             F++   TG +R  +   SLDR++    Y L V+  D          D    ++     + + D
Sbjct  910   FVIEEITGVIRLHSKAISLDRDK---YELNVTAMDDGSCCVNGDATIHTSTAVVVVFITD  966

Query  211   VNDNFPMFRD-SQYSARIEENILS-SELLRFQVTDLDE  246
             VNDN P+F+D   Y  ++EE   + S +++ Q TD D+
Sbjct  967   VNDNKPVFKDCGTYYPKVEEGAPNGSPVIKVQATDEDK  1004

 Score = 53.9 bits (128),  Expect = 1e-05, Method: Compositional matrix adjust.
 Identities = 70/306 (23%), Positives = 126/306 (41%), Gaps = 30/306 (10%)

Query  20    PIAKIT-SDYQATQKITYRI---SGVGIDQPPFGIFVVDKNTGDINITAIVDRE---ETP  72
             PI  +T  D    T K  Y++    SG G D+  F +    +  TG + I    +D E   ++
Sbjct  1768  PILTVTVHDEDETNKFQYKVIDNSGYGADK--FTMVRNNDGTGSLKIVQPLDYEDPLQSN  1825

Query  73    SFLITCRALNAQGLDVEKPLI------LTVKILDINDNPPVFSQQIFMGEIEENSASNSL  126
              F    + +N +G D +          + VK+ DINDN P F +       + EN+
Sbjct  1826  GFRFRIQ-VNDKGEDNDNDKYHVAYSWVVVKLRDINDNKPQFERPNIEVSVYENADVGKT  1884

Query  127   VMILNATDADEPNHLNSKIAFKIVSQEPAGTPMFLLSRNTGEVRTLTNSLDREQASSYRL  186
             +    ATD D+      SK+++  I    + +        S N      T+  +LDRE    +++
Sbjct  1885  LETFKATDPDQGG--KSKVSYAI---DRSSDRQRQFSINQEGTVTIQRNLDREVTPRHQV  1939
```

```
Query  187   VVSGADKDGEGLSTQCECNIKVKDVNDNFPMFRDSQYSARIEENILSSELLRFQVTDLDE  246
              +   D      +      + V+D+NDN P F    Y   + E++  +++     TD D+
Sbjct  1940  KILAIDDGIPPKTATATLTVIVQDINDNPPKFL-KDYRPVLPEHVPPRKVVEILATDDDD  1998

Query  247   EYTDNWLAVYFFTSGNEGNW----FEIQTDPRTNEG----ILKVVKALDYEQLQSVKLSI  298
                 N     F       +     F+++ D +   G    I+  +++ D EQ +   + I
Sbjct  1999  RSKSNGPPFQFRLDPGADDIIRASFKVEQDQKGANGDGMAIVSSLRSFDREQQKEYLIPI  2058

Query  299   AVKNKA  304
              +K+
Sbjct  2059  VIKDHG  2064

 Score = 52.0 bits (123),  Expect = 4e-05, Method: Compositional matrix adjust.
 Identities = 97/398 (24%), Positives = 163/398 (41%), Gaps = 54/398 (14%)

Query  106   PVFSQQIFMGEIEENSASNSLVMILNATDADEPNHLNSKIAFKIVSQ-EPAGTPMFLLSR  164
             P F    +  EI EN   +S ++ + A       +  + +I + + +Q + AGT  F +
Sbjct  655   PQFYMPSYEAEIPENQKKDSDIISVKAK-----SFADREIRYTLKAQGQGAGT--FNIGP  707

Query  165   NTGEVRTLTNSLDRE---QASSYRLVVSGADKDGEGLSTQCECNIKVKDVNDNFPMFRDS  221
              +G V+ L   LD E   Q   Y LVV+ A +D  G ST  E  I+V DVNDN P F
Sbjct  708   TSGIVK-LAKELDFEDLRQPHVYSLVVT-ATEDSGGFSTSVELTIRVTDVNDNAPKFELP  765

Query  222   QYSAR-IEENI-LSSELLRFQVTDLDEEYTDNWLAVYFFTSGNEGNWFEIQTDPRTNEGI  279
              Y A  ++E+I L + +L+ +    D D         + +  S +    F + ++     GI
Sbjct  766   DYQAHNVDEDIPLGTSILKVKAMDSDSGAN---AEIEYLVSDDH---FAVDSN-----GI  814

Query  280   LKVVKALDYEQLQS-VKLSIAVKNKAEFHQSVISRYRVQSTPVTIQVINVREGIAFRPAS  338
             +   K LD +   +  +  +  K+K E  ++   +  RV +           E   F
Sbjct  815   IVNNKQLDADNNNAYYEFVVTAKDKGEPAKTGTATVRVYTKNKN------DEEPKFSQQV  868

Query  339   KTFTVQKGISSKKLVDYILGTYQAIDEDTNKAASNVK--YVMGRNDGGYLMIDSKTAEIK  396
              T  V +      LV  ++         ++K    NV+  +V G     G  +I+  T  I+
Sbjct  869   YTPNVDENAGPNTLVTTVVA--------SDKDGDNVRFGFVGGGTSSGQFVIEEITGVIR  920

Query  397   FVK---NMNRDSTFI-VNKTITAEVLAIDEYTGKTSTGTVYVRVPDFNDN------CPTA  446
                   +++RD   + V            + T  TST  V V + D NDN      C T
Sbjct  921   LHSKAISLDRDKYELNVTAMDDGSCCVNGDATIHTSTAVVVVFITDVNDNKPVFKDCGTY  980

Query  447   VLE-KDAVCSSSPSVVVSARTLNNRYTGPYTFALEDQP  483
               + ++   + SP + V A    +    G  +++   QP
Sbjct  981   YPKVEEGAPNGSPVIKVQATDEDKGVNGQVKYSIVQQP  1018

 Score = 44.7 bits (104),  Expect = 0.008, Method: Compositional matrix adjust.
 Identities = 48/199 (24%), Positives = 73/199 (37%), Gaps = 19/199 (10%)

Query  27    DYQATQKITYRISGVGIDQPPFGIFVVDKNTGDINITAIVDREETPSFLITCRALNAQGL  86
             D     K++Y I      Q  F I       G + I   +DRE TP   +   A++
Sbjct  1894  DQGGKSKVSYAIDRSSDRQRQFSI----NQEGTVTIQRNLDREVTPRHQVKILAIDDGIP  1949

Query  87    DVEKPLILTVKILDINDNPPVFSQQIFMGEIEENSASNSLVMILNATDADEPNHLNSKIA  146
                    LTV + DINDNPP F +  +   + E+     +V IL   D D
Sbjct  1950  PKTATATLTVIVQDINDNPPKFLKD-YRPVLPEHVPPRKVVEILATDDDDRSKSNGPPFQ  2008

Query  147   FKIVSQEPAGTPMFLLSRNTGEVRTLTN-----------SLDREQASSYRLVVSGADKDG  195
             F++   +P    +   S    + +   N           S DREQ   Y + +   D
Sbjct  2009  FRL---DPGADDIIRASFKVEQDQKGANGDGMAIVSSLRSFDREQQKEYLIPIVIKDHGN  2065

Query  196   EGLSTQCECNIKVKDVNDN  214
               ++      + + DVNDN
Sbjct  2066  PAMTGTSTLTVVIGDVNDN  2084

 Score = 40.0 bits (92),  Expect = 0.21, Method: Compositional matrix adjust.
 Identities = 89/389 (23%), Positives = 149/389 (38%), Gaps = 62/389 (16%)

Query  75   LITCRALNAQGLDVEKPLILTVKILDINDNPPVFSQQIF--MGEIEENSASNSLVMILNA  132
            +IT +  NA GL       + + + D+ND PP F  +       ++ N+  N+ V  L A
```

```
Sbjct  517  IITNQGHNA-GLKYTDNQRVIIHVKDVNDEPPYFINRPLPMQAVVQLNAPPNTPVFTLQA  575

Query  133  TDADEPNHLNSKIAFKIVSQEPAGTPMFLLSRNTGEVRTLTNSLDREQASSYRLVVSGAD  192
             D D     +  I + IV     G   F +    +G VRT    L  +     Y L V   D
Sbjct  576  RDPD----TDHNIHYFIVRDRTGG--RFEVDERSGVVRTRGTDL-FQLDMEYVLYVKAED  628

Query  193  KDGE----GLSTQCECNIKVKDVNDNFPMFRDSQYSARIEEN-ILSSELLRFQVTDLDEE  247
            ++G+        +  E  + +        P F    Y A I EN    S+++  +     +
Sbjct  629  QNGKVDDRRFQSTPEERLSIVG-GKRAPQFYMPSYEAEIPENQKKDSDIISVKA----KS  683

Query  248  YTDNWLAVYFFTSGNEGNWFEIQTDPRTNEGILKVVKALDYEQLQSVKLS----IAVKNK  303
            + D  +        G     F I         GI+K+ K LD+E L+   +      A ++
Sbjct  684  FADREIRYTLKAQGQGAGTFNIGP----TSGIVKLAKELDFEDLRQPHVYSLVVTATEDS  739

Query  304  AEFHQSVISRYRVQSTPVTIQVINVREGIAFRPASKTFTVQKGISSKKLVDYILGT----  359
              F  SV           +TI+V +V +        +  F +    +     D  LGT
Sbjct  740  GGFSTSV---------ELTIRVTDVNDN------APKFELPDYQAHNVDEDIPLGTSILK  784

Query  360  YQAIDEDTNKAASNVKYVMGRN----DGGYLMIDSKTAEIKFVKNMNRDSTFIVNKTITA  415
             +A+D D+  A + ++Y++  +    D   +++++K  +     N N    F+V
Sbjct  785  VKAMDSDSG-ANAEIEYLVSDDHFAVDSNGIIVNNKQLD---ADNNNAYYEFVVTAKDKG  840

Query  416  EVLAIDEYTGKTSTGTVYVRVPDFNDNCP  444
            E         KT T TV V   + ND  P
Sbjct  841  E-------PAKTGTATVRVYTKNKNDEEP  862

 Score = 39.7 bits (91),  Expect = 0.28, Method: Compositional matrix adjust.
 Identities = 54/242 (22%), Positives = 102/242 (42%), Gaps = 22/242 (9%)

Query  4    KFAKPCREGE--DNSKRNP-IAKITSDYQATQKITYRISGVGIDQPPFGIFVVDKNTGDI  60
            +F  P  E E  +N K++  I  + +   A ++I Y +   G       G F +   +G +
Sbjct  656  QFYMPSYEAEIPENQKKDSDIISVKAKSFADREIRYTLKAQGQGA---GTFNIGPTSGIV  712

Query  61   NITAIVDREE--TPSF--LITCRALNAQGLDVEKPLILTVKILDINDNPPVFSQQIFMGE  116
             +   +D E+   P    L+      ++ G       + LT+++ D+NDN P F    +
Sbjct  713  KLAKELDFEDLRQPHVYSLVVTATEDSGGFSTS--VELTIRVTDVNDNAPKFELPDYQAH  770

Query  117  -IEENSASNSLVMILNATDADEPNHLNSKIAFKIVSQEPAGTPMFLLSRNTGEVRTLTNS  175
             ++E+    + ++ + A D+D  +  N++I + +VS +      F +  N   V
Sbjct  771  NVDEDIPLGTSILKVKAMDSD--SGANAEIEY-LVSDD-----HFAVDSNGIIVNNKQLD  822

Query  176  LDREQASSYRLVVSGADKDGEGLSTQCECNIKVKDVNDNFPMFRDSQYSARIEENILSSE  235
             D   A  Y  VV+  DK     +      +  K+ ND  P F    Y+  ++EN   +
Sbjct  823  ADNNNA-YYEFVVTAKDKGEPAKTGTATVRVYTKNKNDEEPKFSQQVYTPNVDENAGPNT  881

Query  236  LL  237
            L+
Sbjct  882  LV  883

>AARA21_009090.P17034 | transcript=AARA21_009090.R17034 | gene=AARA21_009090
| organism=Anopheles_arabiensis_DONGOLA_2021
| gene_product=neural-cadherin, transcript variant X5 |
transcript_product=neural-cadherin,
transcript variant X14 | location=CM029353:40466514-40666848(+)
| protein_length=3099 |
sequence_SO=chromosome | SO=protein_coding_gene | is_pseudo=false
Length=3099

 Score = 147 bits (371),  Expect = 2e-35, Method: Compositional matrix adjust.
 Identities = 128/446 (29%), Positives = 217/446 (49%), Gaps = 36/446 (8%)

Query  20    PIAKITS---DYQATQKITYRISGVGID--QPPFGIFVVDKNTGDINITAIVDREET---  71
              PI ++T+    D    Q I Y ++G GID    P    F +++ TG+I +    +DR++
Sbjct  1539  PIMQVTATDGDKDRPQNIVYFLTGQGIDPDNPANSKFDINRTTGEIFVLKPLDRDQPNGR  1598

Query  72    PSFLITCRALNAQGLDVEKPLILTVKILDINDNPPVFSQQIFMGEIEENSASNSLVMILN  131
              P +  T  A +  G  +       + V + DINDN P+F Q ++ G + EN  +  +VM +
Sbjct  1599  PQWRFTVFAQDEGGEGLVGYADVQVNLKDINDNAPIFPQGVYFGNVTENGTAGMVVMTMT  1658
```

```
Query  132   ATDADEPNH-LNSKIAFKI---VSQEPAGTPMFLLSRNTGEVRTLTNSLDREQASSYRLV  187
             A D D+PN   N+K+ + I   V +E  G+P+F +  +TG ++T    LDRE+   Y +
Sbjct  1659  AVDYDDPNEGTNAKLIYSIEKNVIEEETGSPIFEIEADTGVIKTAVCCLDRERTPDYSIQ  1718

Query  188   VSGADKDGEGLSTQCECNIKVKDVNDNFPMFRDSQYSARIEENILSS--ELLRFQVTDLD  245
             V     DG GL      +I+VKD+ND  P F   ++   ++E   ++  E+    VT  D
Sbjct  1719  VVAM--DGGGLKGTGTASIRVKDINDMPPQFTKDEWFTEVDETDGTNLPEMPILTVTVHD  1776

Query  246   EEYTDNWLAVYFFTSGNEGNWFEIQTDPRTNEGI--LKVVKALDYE---QLQSVKLSIAV  300
             E+ T+ +       SG   + F +    R N+G   LK+V+ LDYE   Q     +  I V
Sbjct  1777  EDETNKFQYKVIDNSGYGADKFTMV---RNNDGTGSLKIVQPLDYEDPLQSNGFRFRIQV  1833

Query  301   KNKAEFHQSVISRYRVQSTPVTIQVINVREGIAFRPASKTFTVQKGISSKKLVDYILGTY  360
              +K E + +   +Y V  + V +++ ++ +    +P  +   ++  +     V   L T+
Sbjct  1834  NDKGEDNDN--DKYHVAYSWVVVKLRDINDN---KPQFERPNIEVSVYENADVGKTLETF  1888

Query  361   QAIDEDTNKAASNVKYVMGR-NDGGYLMIDSKTAEIKFVKNMNRDSTFIVNKTITAEVLA  419
             +A D D     S V Y + R +D       ++   +   +N++R+    V      ++LA
Sbjct  1889  KATDPDQG-GKSKVSYAIDRSSDRQRQFSINQEGTVTIQRNLDRE----VTPRHQVKILA  1943

Query  420   IDE-YTGKTSTGTVYVRVPDFNDNCP  444
             ID+    KT+T T+ V V D NDN P
Sbjct  1944  IDDGIPPKTATATLTVIVQDINDNPP  1969

 Score = 78.6 bits (192),  Expect = 3e-13, Method: Compositional matrix adjust.
 Identities = 106/424 (25%), Positives = 180/424 (42%), Gaps = 51/424 (12%)

Query  53    VDKNTGDINITAIVDREETPSFLITCRALNAQGLDVEKPLILTVKILDINDNPPVFSQ-Q  111
             +D++  ++N+TA+ D         +C  +N          ++ V I D+NDN PVF
Sbjct  928   LDRDKYELNVTAMDDG--------SC-CVNGDATIHTSTAVVVVFITDVNDNKPVFKDCG  978

Query  112   IFMGEIEENSASNSLVMILNATDADEPNHLNSKIAFKIVSQEPAGTPMFLLSRNTGEVRT  171
              +  ++EE + + S V+ + ATD D+   +N ++ + IV Q       F +   TGEV T
Sbjct  979   TYYPKVEEGAPNGSPVIKVQATDEDK--GVNGQVKYSIVQQPNQKGTKFTVDEETGEVST  1036

Query  172   LTNSLDRE--QASSYRLVVSGADKDGEGLSTQCECNIKVKDVNDNFPMFRDSQYSARIEE  229
                  DRE        + V   D+    L   C   +++ DVNDN P+F   +Y   +++
Sbjct  1037  -NKVFDREGDDGKFVSVTVKATDQGEPSLEGVCSFTVEITDVNDNPPLFDRQKYVENVKQ  1095

Query  230   NI-LSSELLRFQVTDLDEEYTDNWLAVYFFT---SGNEGNWFEIQTDPRTNEGILKVVKA  285
             +  + + +LR  V+  DE+  +N   VY  +   S N+  +FEIQ  P +   +LK  K
Sbjct  1096  DASIGTNILR--VSASDEDADNNGAIVYTLSAPHSPNDLEYFEIQ--PESGWIVLK--KP  1149

Query  286   LDYEQLQSVKLSIAVKNK------AEFHQSVISRYRVQSTPVTIQV----INVREGIAFR  335
             LD E+    +L +   ++      A+    +    R    P+  +     I++RE +
Sbjct  1150  LDRERY---RLKVYASDRGDPPSTADVDVELDVVDRNNKPPIWDRTIYGPIHIRENVTVG  1206

Query  336   PASKTFTVQKGISSKKLVDYILGTYQAIDEDTNKAASNVKYVMGRNDGGYLMIDSKTAEI  395
                 +     GI     V Y L         TNK   +  Y+  R D G        A+I
Sbjct  1207  TVVTSVKASSGIEGNPTVFYRL--MPGSTAQTNKL--HTFYLQQRPDNG-----DTWADI  1257

Query  396   KFVKNMNRDSTFIVNKTITAEVLAIDEYTGKTSTGTVYYRVPDFNDNCPTAV-LEKDAVC  454
             K    ++ ++    N T+  E     +     S  TVY+++ D ND  P     E++ V
Sbjct  1258  KVNHPLDFETIKEYNLTVRVENNGAQQL---ASEATVYIQLEDVNDEIPLFTEREQETVL  1314

Query  455   SSSP  458
                P
Sbjct  1315  EGEP  1318

 Score = 75.9 bits (185),  Expect = 2e-12, Method: Compositional matrix adjust.
 Identities = 103/418 (25%), Positives = 174/418 (42%), Gaps = 50/418 (12%)

Query  50    IFVVDKNTGDINITAIVDREETPSFLITCRAL----------NAQGLDVEKPLILTVKIL  99
              F ++  +G+I    + DRE+  ++ +   A           N Q   V K   + + I
Sbjct  1355  FFEINLQSGEIFTKVVFDREKQGAYALEVEARDGAPSARPNSNQQPNSVTK--FIRIGIA  1412

Query  100   DINDNPPVFSQQIFMGEIEENSASNSLVMILNATDADEPNHLNSKIAFKIVSQEPAGTPM  159
             D NDNPP F + ++  E++EN      V+ + A D DE    +S+I ++I S    G
Sbjct  1413  DKNDNPPYFDKALYEAEVDENEDIQHTVLTVTAKDHDE----SSRIRYEITSGNIGGA--  1466
```

```
Query  160   FLLSRNTGEVRTLTNSLDREQASSYRLVVSGADKDGEGLSTQCECNIKVKDVNDNFPMFR  219
             F +   TG +  +  +LD E    Y L +  +D   E  +      I VKDVND  P F
Sbjct  1467  FAVKNMTGAI-YVAGALDYETRKRYYLTLVASDSLNENETIIV---INVKDVNDMPPSFP  1522

Query  220   DSQYSARIEENI-LSSELLRFQVTDLDEEYTDNWLAVYFFTSG--NEGNWFEIQTDPRTN  276
                Y   ++E + +   +++   TD D++   N   VYF T    +  N    + D
Sbjct  1523  QQVYERTMDEELAVPFPIMQVTATDGDKDRPQN--IVYFLTGQGIDPDNPANSKFDINRT  1580

Query  277   EGILKVVKALDYEQLQS-VKLSIAVKNKAEFHQSVISRYRVQSTPVTIQVINVREGIAFR  335
              G + V+K LD +Q     +    V  + E  + ++    VQ     + + ++ +
Sbjct  1581  TGEIFVLKPLDRDQPNGRPQWRFTVFAQDEGGEGLVGYADVQ-----VNLKDINDNAPIF  1635

Query  336   PASKTFTVQKGISSKKLVDYILGTYQAIDEDTNKAASNVK--YVMGRN------DGGYLM  387
             P    F     ++        ++ T  A+D D     +N K  Y + +N
Sbjct  1636  PQGVYF---GNVTENGTAGMVVMTMTAVDYDDPNEGTNAKLIYSIEKNVIEEETGSPIFE  1692

Query  388   IDSKTAEIK-FVKNMNRDSTFIVNKTITAEVLAIDEYTGKTSTGTVYVRVPDFNDNCP  444
             I++ T  IK  V  ++R+ T       + +V+A+D   G   TGT  +RV D ND  P
Sbjct  1693  IEADTGVIKTAVCCLDRERT----PDYSIQVVAMDG-GGLKGTGTASIRVKDINDMPP  1745

 Score = 65.5 bits (158),  Expect = 3e-09, Method: Compositional matrix adjust.
 Identities = 85/359 (24%), Positives = 153/359 (43%), Gaps = 32/359 (9%)

Query  51    FVVDKNTGDINITAIVDRE-ETPSFL-ITCRALNAQGLDVEKPLILTVKILDINDNPPVF  108
             F VD+ TG+++   + DRE +   F+ +T +A +     +E     TV+I D+NDNPP+F
Sbjct  1025  FTVDEETGEVSTNKVFDREGDDGKFVSVTVKATDQGEPSLEGVCSFTVEITDVNDNPPLF  1084

Query  109   SQQIFMGEIEENSASNSLVMILNATDADEPNHLNSKIAFKIVS-QEPAGTPMFLLSRNTG  167
              +Q ++  ++++++  + ++ ++A+D D  N  N  I + + +   P     F +   +G
Sbjct  1085  DRQKYVENVKQDASIGTNILRVSASDEDADN--NGAIVYTLSAPHSPNDLEYFEIQPESG  1142

Query  168   EVRTLTNSLDREQASSYRLVVSGADKDGEGLSTQCECNIKVKDVNDNFPMFRDSQYSARI  227
              +  L   LDRE+   YRL V  +D+ G+  ST            +N P   D      I
Sbjct  1143  WI-VLKKPLDRER---YRLKVYASDR-GDPPSTADVDVELDVVDRNNKPPIWDRTIYGPI  1197

Query  228   EENILSSELLRFQVTDLDEEYTDNWLAVYFFTSG-----NEGNWFEIQTDPRTNE--GIL  280
                   +                N    Y    G     N+ + F +Q  P   +    +
Sbjct  1198  HIRENVTVGTVVTSVKASSGIEGNPTVFYRLMPGSTAQTNKLHTFYLQQRPDNGDTWADI  1257

Query  281   KVVKALDYEQLQSVKLSIAVKNKAEFHQSVISRYRVQSTPVTIQVINVREGIAFRPASKT  340
             KV   LD+E ++   L++ V+N          ++       V IQ+ +V + I      +
Sbjct  1258  KVNHPLDFETIKEYNLTVRVENNG-------AQQLASEATVYIQLEDVNDEIPLFTEREQ  1310

Query  341   FTVQKGISSKKLVDYILGTYQAIDEDTNKAASNVKYVM---GRNDG-GYLMIDSKTAEI  395
              TV +G      V  +     AID+D     + V Y +    RN+G  +  I+ ++ EI
Sbjct  1311  ETVLEGEPIGTKVTQV----NAIDKDGTFPNNQVYYYIVDSPRNEGKDFFEINLQSGEI  1365

 Score = 55.8 bits (133),  Expect = 3e-06, Method: Compositional matrix adjust.
 Identities = 49/158 (31%), Positives = 78/158 (49%), Gaps = 24/158 (15%)

Query  102   NDNPPVFSQQIFMGEIEENSASNSLVMILNATDADEPNHLNSKIAFKIVSQEPAGTP--M  159
             ND  P FSQQ++   ++EN+  N+LV  + A+D D  N     + F  V     GT
Sbjct  858   NDEEPKFSQQVYTPNVDENAGPNTLVTTVVASDKDGDN-----VRFGFVG---GGTSSGQ  909

Query  160   FLLSRNTGEVRTLTN--SLDREQASSYRLVVSGADK-------DGEGLSTQCECNIKVKD  210
             F++   TG +R  +   SLDR++   Y L V+  D        D    ++     + + D
Sbjct  910   FVIEEITGVIRLHSKAISLDRDK---YELNVTAMDDGSCCVNGDATIHTSTAVVVVFITD  966

Query  211   VNDNFPMFRD-SQYSARIEENILS-SELLRFQVTDLDE  246
             VNDN P+F+D   Y  ++EE   + S +++ Q TD D+
Sbjct  967   VNDNKPVFKDCGTYYPKVEEGAPNGSPVIKVQATDEDK  1004

 Score = 53.9 bits (128),  Expect = 1e-05, Method: Compositional matrix adjust.
 Identities = 70/306 (23%), Positives = 126/306 (41%), Gaps = 30/306 (10%)

Query  20    PIAKIT-SDYQATQKITYRI---SGVGIDQPPFGIFVVDKNTGDINITAIVDRE---ETP  72
             PI  +T  D   T K  Y++   SG G D+  F +   +  TG + I   +D E   ++
```

```
Sbjct  1768  PILTVTVHDEDETNKFQYKVIDNSGYGADK--FTMVRNNDGTGSLKIVQPLDYEDPLQSN  1825

Query  73    SFLITCRALNAQGLDVEKPLI------LTVKILDINDNPPVFSQQIFMGEIEENSASNSL  126
              F    + +N +G D +          + VK+ DINDN P F +      + EN+
Sbjct  1826  GFRFRIQ-VNDKGEDNDNDKYHVAYSWVVVKLRDINDNKPQFERPNIEVSVYENADVGKT  1884

Query  127   VMILNATDADEPNHLNSKIAFKIVSQEPAGTPMFLLSRNTGEVRTLTNSLDREQASSYRL  186
             +    ATD D+     SK+++ I   + +       S N     T+  +LDRE    +++
Sbjct  1885  LETFKATDPDQGG--KSKVSYAI---DRSSDRQRQFSINQEGTVTIQRNLDREVTPRHQV  1939

Query  187   VVSGADKDGEGLSTQCECNIKVKDVNDNFPMFRDSQYSARIEENILSSELLRFQVTDLDE  246
              +   D      +      + V+D+NDN P F    Y   + E++   +++    TD D+
Sbjct  1940  KILAIDDGIPPKTATATLTVIVQDINDNPPKFL-KDYRPVLPEHVPPRKVVEILATDDDD  1998

Query  247   EYTDNWLAVYFFTSGNEGNW----FEIQTDPRTNEG----ILKVVKALDYEQLQSVKLSI  298
                 N     F       +     F+++ D +   G    I+  +++ D EQ +   + I
Sbjct  1999  RSKSNGPPFQFRLDPGADDIIRASFKVEQDQKGANGDGMAIVSSLRSFDREQQKEYLIPI  2058

Query  299   AVKNKA  304
              +K+
Sbjct  2059  VIKDHG  2064

 Score = 52.0 bits (123),  Expect = 4e-05, Method: Compositional matrix adjust.
 Identities = 97/398 (24%), Positives = 163/398 (41%), Gaps = 54/398 (14%)

Query  106   PVFSQQIFMGEIEENSASNSLVMILNATDADEPNHLNSKIAFKIVSQ-EPAGTPMFLLSR  164
             P F    +  EI EN   +S ++ + A      +  + +I + + +Q + AGT  F +
Sbjct  655   PQFYMPSYEAEIPENQKKDSDIISVKAK-----SFADREIRYTLKAQGQGAGT--FNIGP  707

Query  165   NTGEVRTLTNSLDRE---QASSYRLVVSGADKDGEGLSTQCECNIKVKDVNDNFPMFRDS  221
              +G V+ L   LD E   Q   Y LVV+ A +D  G ST  E  I+V DVNDN P F
Sbjct  708   TSGIVK-LAKELDFEDLRQPHVYSLVVT-ATEDSGGFSTSVELTIRVTDVNDNAPKFELP  765

Query  222   QYSAR-IEENI-LSSELLRFQVTDLDEEYTDNWLAVYFFTSGNEGNWFEIQTDPRTNEGI  279
              Y A  ++E+I L + +L+ +  D D         + +  S +    F + ++     GI
Sbjct  766   DYQAHNVDEDIPLGTSILKVKAMDSDSGAN---AEIEYLVSDDH---FAVDSN-----GI  814

Query  280   LKVVKALDYEQLQS-VKLSIAVKNKAEFHQSVISRYRVQSTPVTIQVINVREGIAFRPAS  338
             +   K LD +   +  +  +  K+K E  ++  +  RV +           E   F
Sbjct  815   IVNNKQLDADNNNAYYEFVVTAKDKGEPAKTGTATVRVYTKNKN------DEEPKFSQQV  868

Query  339   KTFTVQKGISSKKLVDYILGTYQAIDEDTNKAASNVK--YVMGRNDGGYLMIDSKTAEIK  396
              T  V +      LV  ++         ++K   NV+  +V G    G  +I+  T  I+
Sbjct  869   YTPNVDENAGPNTLVTTVVA--------SDKDGDNVRFGFVGGGTSSGQFVIEEITGVIR  920

Query  397   FVK---NMNRDSTFI-VNKTITAEVLAIDEYTGKTSTGTVYVRVPDFNDN------CPTA  446
                   +++RD   + V            + T  TST  V V + D NDN      C T
Sbjct  921   LHSKAISLDRDKYELNVTAMDDGSCCVNGDATIHTSTAVVVVFITDVNDNKPVFKDCGTY  980

Query  447   VLE-KDAVCSSSPSVVVSARTLNNRYTGPYTFALEDQP  483
               + ++   + SP + V A   +    G   +++  QP
Sbjct  981   YPKVEEGAPNGSPVIKVQATDEDKGVNGQVKYSIVQQP  1018

 Score = 44.3 bits (103),  Expect = 0.009, Method: Compositional matrix adjust.
 Identities = 48/199 (24%), Positives = 73/199 (37%), Gaps = 19/199 (10%)

Query  27    DYQATQKITYRISGVGIDQPPFGIFVVDKNTGDINITAIVDREETPSFLITCRALNAQGL  86
             D     K++Y I      Q  F I       G + I   +DRE TP   +   A++
Sbjct  1894  DQGGKSKVSYAIDRSSDRQRQFSI----NQEGTVTIQRNLDREVTPRHQVKILAIDDGIP  1949

Query  87    DVEKPLILTVKILDINDNPPVFSQQIFMGEIEENSASNSLVMILNATDADEPNHLNSKIA  146
                    LTV + DINDNPP F +  +   + E+     +V IL   D D
Sbjct  1950  PKTATATLTVIVQDINDNPPKFLKD-YRPVLPEHVPPRKVVEILATDDDDRSKSNGPPFQ  2008

Query  147   FKIVSQEPAGTPMFLLSRNTGEVRTLTN-----------SLDREQASSYRLVVSGADKDG  195
             F++   +P    +   S    + +   N           S DREQ    Y + +   D
Sbjct  2009  FRL---DPGADDIIRASFKVEQDQKGANGDGMAIVSSLRSFDREQQKEYLIPIVIKDHGN  2065

Query  196   EGLSTQCECNIKVKDVNDN  214
```

```
               ++      + + DVNDN
Sbjct  2066  PAMTGTSTLTVVIGDVNDN  2084

 Score = 39.7 bits (91),  Expect = 0.27, Method: Compositional matrix adjust.
 Identities = 89/389 (23%), Positives = 149/389 (38%), Gaps = 62/389 (16%)

Query  75   LITCRALNAQGLDVEKPLILTVKILDINDNPPVFSQQIF--MGEIEENSASNSLVMILNA  132
            +IT +  NA GL       + + + D+ND PP F  +       ++ N+  N+ V  L A
Sbjct  517  IITNQGHNA-GLKYTDNQRVIIHVKDVNDEPPYFINRPLPMQAVVQLNAPPNTPVFTLQA  575

Query  133  TDADEPNHLNSKIAFKIVSQEPAGTPMFLLSRNTGEVRTLTNSLDREQASSYRLVVSGAD  192
             D D     +  I + IV     G   F +   +G VRT    L  +    Y L V   D
Sbjct  576  RDPD----TDHNIHYFIVRDRTGG--RFEVDERSGVVRTRGTDL-FQLDMEYVLYVKAED  628

Query  193  KDGE----GLSTQCECNIKVKDVNDNFPMFRDSQYSARIEEN-ILSSELLRFQVTDLDEE  247
            ++G+       +  E  + +       P F    Y A I EN    S+++  +      +
Sbjct  629  QNGKVDDRRFQSTPEERLSIVG-GKRAPQFYMPSYEAEIPENQKKDSDIISVKA----KS  683

Query  248  YTDNWLAVYFFTSGNEGNWFEIQTDPRTNEGILKVVKALDYEQLQSVKLS----IAVKNK  303
            + D  +       G     F I        GI+K+ K LD+E L+   +      A ++
Sbjct  684  FADREIRYTLKAQGQGAGTFNIGP----TSGIVKLAKELDFEDLRQPHVYSLVVTATEDS  739

Query  304  AEFHQSVISRYRVQSTPVTIQVINVREGIAFRPASKTFTVQKGISSKKLVDYILGT----  359
              F  SV          +TI+V +V +       +  F +    +     D  LGT
Sbjct  740  GGFSTSV---------ELTIRVTDVNDN------APKFELPDYQAHNVDEDIPLGTSILK  784

Query  360  YQAIDEDTNKAASNVKYVMGRN----DGGYLMIDSKTAEIKFVKNMNRDSTFIVNKTITA  415
             +A+D D+  A + ++Y++  +    D   +++++K  +     N N    F+V
Sbjct  785  VKAMDSDSG-ANAEIEYLVSDDHFAVDSNGIIVNNKQLD---ADNNNAYYEFVVTAKDKG  840

Query  416  EVLAIDEYTGKTSTGTVYVRVPDFNDNCP  444
            E         KT T TV V   + ND  P
Sbjct  841  E-------PAKTGTATVRVYTKNKNDEEP  862

 Score = 39.3 bits (90),  Expect = 0.30, Method: Compositional matrix adjust.
 Identities = 54/242 (22%), Positives = 102/242 (42%), Gaps = 22/242 (9%)

Query  4    KFAKPCREGE--DNSKRNP-IAKITSDYQATQKITYRISGVGIDQPPFGIFVVDKNTGDI  60
            +F  P  E E  +N K++  I  + +   A ++I Y +   G      G F +   +G +
Sbjct  656  QFYMPSYEAEIPENQKKDSDIISVKAKSFADREIRYTLKAQGQGA---GTFNIGPTSGIV  712

Query  61   NITAIVDREE--TPSF--LITCRALNAQGLDVEKPLILTVKILDINDNPPVFSQQIFMGE  116
             +   +D E+   P    L+     ++ G      + LT+++ D+NDN P F    +
Sbjct  713  KLAKELDFEDLRQPHVYSLVVTATEDSGGFSTS--VELTIRVTDVNDNAPKFELPDYQAH  770

Query  117  -IEENSASNSLVMILNATDADEPNHLNSKIAFKIVSQEPAGTPMFLLSRNTGEVRTLTNS  175
             ++E+    + ++ + A D+D  +  N++I + +VS +      F +  N   V
Sbjct  771  NVDEDIPLGTSILKVKAMDSD--SGANAEIEY-LVSDD-----HFAVDSNGIIVNNKQLD  822

Query  176  LDREQASSYRLVVSGADKDGEGLSTQCECNIKVKDVNDNFPMFRDSQYSARIEENILSSE  235
             D   A  Y  VV+  DK     +      +  K+ ND  P F    Y+  ++EN   +
Sbjct  823  ADNNNA-YYEFVVTAKDKGEPAKTGTATVRVYTKNKNDEEPKFSQQVYTPNVDENAGPNT  881

Query  236  LL  237
            L+
Sbjct  882  LV  883

>AMEM21_003341.P6428 | transcript=AMEM21_003341.R6428 | gene=AMEM21_003341
| organism=Anopheles_merus_MAF_2021 | gene_product=neural-cadherin,
transcript variant X1 | transcript_product=neural-cadherin,
transcript variant X19 | location=CM029766:40289848-40498951(+)
| protein_length=3098 | sequence_SO=chromosome
| SO=protein_coding_gene | is_pseudo=false
Length=3098

 Score = 147 bits (371),  Expect = 3e-35, Method: Compositional matrix adjust.
 Identities = 128/446 (29%), Positives = 217/446 (49%), Gaps = 36/446 (8%)

```

```
Query  20    PIAKITS---DYQATQKITYRISGVGID--QPPFGIFVVDKNTGDINITAIVDREET---  71
             PI ++T+   D    Q I Y ++G GID   P    F +++ TG+I +   +DR++
Sbjct  1539  PIMQVTATDGDKDRPQNIVYFLTGQGIDPDNPANSKFDINRTTGEIFVLKPLDRDQPNGR  1598

Query  72    PSFLITCRALNAQGLDVEKPLILTVKILDINDNPPVFSQQIFMGEIEENSASNSLVMILN  131
             P +  T  A +  G  +     + V + DINDN P+F Q ++ G + EN  +  +VM +
Sbjct  1599  PQWRFTVFAQDEGGEGLVGYADVQVNLKDINDNAPIFPQGVYFGNVTENGTAGMVVMTMT  1658

Query  132   ATDADEPNH-LNSKIAFKI---VSQEPAGTPMFLLSRNTGEVRTLTNSLDREQASSYRLV  187
             A D D+PN   N+K+ + I   V +E  G+P+F +  +TG ++T    LDRE+   Y +
Sbjct  1659  AVDYDDPNEGTNAKLIYSIEKNVIEEETGSPIFEIEADTGVIKTAVCCLDRERTPDYSIQ  1718

Query  188   VSGADKDGEGLSTQCECNIKVKDVNDNFPMFRDSQYSARIEENILSS--ELLRFQVTDLD  245
             V     DG GL      +I+VKD+ND  P F   ++   ++E   ++  E+    VT  D
Sbjct  1719  VVAM--DGGGLKGTGTASIRVKDINDMPPQFTKDEWFTEVDETDGTNLPEMPILTVTVHD  1776

Query  246   EEYTDNWLAVYFFTSGNEGNWFEIQTDPRTNEGI--LKVVKALDYE---QLQSVKLSIAV  300
             E+ T+ +       SG    + F +    R N+G   LK+V+ LDYE   Q    +  I V
Sbjct  1777  EDETNKFQYKVIDNSGYGADKFTMV---RNNDGTGSLKIVQPLDYEDPLQSNGFRFRIQV  1833

Query  301   KNKAEFHQSVISRYRVQSTPVTIQVINVREGIAFRPASKTFTVQKGISSKKLVDYILGTY  360
              +K E + +   +Y V  + V +++ ++ +    +P  +   ++  +     V   L T+
Sbjct  1834  NDKGEDNDN--DKYHVAYSWVVVKLRDINDN---KPQFERPNIEVSVYENADVGKTLETF  1888

Query  361   QAIDEDTNKAASNVKYVMGR-NDGGYLMIDSKTAEIKFVKNMNRDSTFIVNKTITAEVLA  419
             +A D D     S V Y + R +D        ++   +   +N++R+    V      ++LA
Sbjct  1889  KATDPDQG-GKSKVSYAIDRSSDRQRQFSINQEGTVTIQRNLDRE----VTPRHQVKILA  1943

Query  420   IDE-YTGKTSTGTVYVRVPDFNDNCP  444
             ID+    KT+T T+ V V D NDN P
Sbjct  1944  IDDGIPPKTATATLTVIVQDINDNPP  1969

 Score = 79.0 bits (193),  Expect = 2e-13, Method: Compositional matrix adjust.
 Identities = 106/424 (25%), Positives = 180/424 (42%), Gaps = 51/424 (12%)

Query  53    VDKNTGDINITAIVDREETPSFLITCRALNAQGLDVEKPLILTVKILDINDNPPVFSQ-Q  111
             +D++  ++N+TA+ D         +C  +N           ++ V I D+NDN PVF
Sbjct  928   LDRDKYELNVTAMDDG--------SC-CVNGDATIHTSTAVVVVFITDVNDNKPVFKDCG  978

Query  112   IFMGEIEENSASNSLVMILNATDADEPNHLNSKIAFKIVSQEPAGTPMFLLSRNTGEVRT  171
              +  ++EE + + S V+ + ATD D+   +N ++ + IV Q       F +   TGEV T
Sbjct  979   TYYPKVEEGAPNGSPVIKVQATDEDK--GVNGQVKYSIVQQPNQKGTKFTVDEETGEVST  1036

Query  172   LTNSLDRE--QASSYRLVVSGADKDGEGLSTQCECNIKVKDVNDNFPMFRDSQYSARIEE  229
                  DRE        + V   D+    L   C   +++ DVNDN P+F    +Y   +++
Sbjct  1037  -NKVFDREGDDGKFVSVTVKATDQGEPSLEGVCSFTVEITDVNDNPPLFDRQKYVENVKQ  1095

Query  230   NI-LSSELLRFQVTDLDEEYTDNWLAVYFFT---SGNEGNWFEIQTDPRTNEGILKVVKA  285
             +  + + +LR  V+  DE+  +N   VY  +   S N+  +FEIQ  P +   +LK  K
Sbjct  1096  DASIGTNILR--VSASDEDADNNGAIVYTLSAPHSPNDLEYFEIQ--PESGWIVLK--KP  1149

Query  286   LDYEQLQSVKLSIAVKNK------AEFHQSVISRYRVQSTPVTIQV----INVREGIAFR  335
             LD E+    +L +   ++      A+    +    R    P+  +     I++RE +
Sbjct  1150  LDRERY---RLKVYASDRGDPPSTADVDVELDVVDRNNKPPIWDRTIYGPIHIRENVTVG  1206

Query  336   PASKTFTVQKGISSKKLVDYILGTYQAIDEDTNKAASNVKYVMGRNDGGYLMIDSKTAEI  395
                 +     GI     V Y L         TNK   +  Y+  R D G        A+I
Sbjct  1207  TVVTSVKASSGIEGNPTVFYRL--MPGSTAQTNKL--HTFYLQQRPDNG-----DTWADI  1257

Query  396   KFVKNMNRDSTFIVNKTITAEVLAIDEYTGKTSTGTVYVRVPDFNDNCPTAV-LEKDAVC  454
             K    ++ ++    N T+  E     +     S  TVY+++ D ND  P     E++ V
Sbjct  1258  KVNHPLDFETIKEYNLTVRVENNGAQQL---ASEATVYIQLEDVNDEIPLFTEREQETVL  1314

Query  455   SSSP  458
                P
Sbjct  1315  EGEP  1318

 Score = 76.3 bits (186),  Expect = 1e-12, Method: Compositional matrix adjust.
 Identities = 103/418 (25%), Positives = 174/418 (42%), Gaps = 50/418 (12%)
```

```
Query  50    IFVVDKNTGDINITAIVDREETPSFLITCRAL----------NAQGLDVEKPLILTVKIL  99
              F ++  +G+I    + DRE+  ++ +   A           N Q   V K   + + I
Sbjct  1355  FFEINLQSGEIFTKVVFDREKQGAYALEVEARDGAPSARPNSNQQPNSVTK--FIRIGIA  1412

Query  100   DINDNPPVFSQQIFMGEIEENSASNSLVMILNATDADEPNHLNSKIAFKIVSQEPAGTPM  159
             D NDNPP F + ++  E++EN      V+ + A D DE    +S+I ++I S    G
Sbjct  1413  DKNDNPPYFDKALYEAEVDENEDIQHTVLTVTAKDHDE----SSRIRYEITSGNIGGA--  1466

Query  160   FLLSRNTGEVRTLTNSLDREQASSYRLVVSGADKDGEGLSTQCECNIKVKDVNDNFPMFR  219
             F +   TG +  +  +LD E    Y L +  +D   E  +      I VKDVND  P F
Sbjct  1467  FAVKNMTGAI-YVAGALDYETRKRYYLTLVASDSLNENETIIV---INVKDVNDMPPSFP  1522

Query  220   DSQYSARIEENI-LSSELLRFQVTDLDEEYTDNWLAVYFFTSG--NEGNWFEIQTDPRTN  276
                Y   ++E + +   +++   TD D++   N   VYF T    +  N    + D
Sbjct  1523  QQVYERTMDEELAVPFPIMQVTATDGDKDRPQN--IVYFLTGQGIDPDNPANSKFDINRT  1580

Query  277   EGILKVVKALDYEQLQS-VKLSIAVKNKAEFHQSVISRYRVQSTPVTIQVINVREGIAFR  335
              G + V+K LD +Q     +    V  + E  + ++    VQ     + + ++ +
Sbjct  1581  TGEIFVLKPLDRDQPNGRPQWRFTVFAQDEGGEGLVGYADVQ-----VNLKDINDNAPIF  1635

Query  336   PASKTFTVQKGISSKKLVDYILGTYQAIDEDTNKAASNVK--YVMGRN------DGGYLM  387
             P    F     ++       ++ T  A+D D     +N K  Y + +N
Sbjct  1636  PQGVYF---GNVTENGTAGMVVMTMTAVDYDDPNEGTNAKLIYSIEKNVIEEETGSPIFE  1692

Query  388   IDSKTAEIK-FVKNMNRDSTFIVNKTITAEVLAIDEYTGKTSTGTVYVRVPDFNDNCP  444
             I++ T  IK  V  ++R+ T       + +V+A+D   G   TGT  +RV D ND  P
Sbjct  1693  IEADTGVIKTAVCCLDRERT----PDYSIQVVAMDG-GGLKGTGTASIRVKDINDMPP  1745

 Score = 65.9 bits (159),  Expect = 2e-09, Method: Compositional matrix adjust.
 Identities = 85/359 (24%), Positives = 153/359 (43%), Gaps = 32/359 (9%)

Query  51    FVVDKNTGDINITAIVDRE-ETPSFL-ITCRALNAQGLDVEKPLILTVKILDINDNPPVF  108
             F VD+ TG+++   + DRE +   F+ +T +A +     +E     TV+I D+NDNPP+F
Sbjct  1025  FTVDEETGEVSTNKVFDREGDDGKFVSVTVKATDQGEPSLEGVCSFTVEITDVNDNPPLF  1084

Query  109   SQQIFMGEIEENSASNSLVMILNATDADEPNHLNSKIAFKIVS-QEPAGTPMFLLSRNTG  167
              +Q ++  ++++++  + ++ ++A+D D  N  N  I + + +   P     F +   +G
Sbjct  1085  DRQKYVENVKQDASIGTNILRVSASDEDADN--NGAIVYTLSAPHSPNDLEYFEIQPESG  1142

Query  168   EVRTLTNSLDREQASSYRLVVSGADKDGEGLSTQCECNIKVKDVNDNFPMFRDSQYSARI  227
              +  L   LDRE+   YRL V  +D+ G+  ST            +N P   D      I
Sbjct  1143  WI-VLKKPLDRER---YRLKVYASDR-GDPPSTADVDVELDVVDRNNKPPIWDRTIYGPI  1197

Query  228   EENILSSELLRFQVTDLDEEYTDNWLAVYFFTSG-----NEGNWFEIQTDPRTNE--GIL  280
                   +                N    Y    G     N+ + F +Q  P   +    +
Sbjct  1198  HIRENVTVGTVVTSVKASSGIEGNPTVFYRLMPGSTAQTNKLHTFYLQQRPDNGDTWADI  1257

Query  281   KVVKALDYEQLQSVKLSIAVKNKAEFHQSVISRYRVQSTPVTIQVINVREGIAFRPASKT  340
             KV   LD+E ++   L++ V+N         ++       V IQ+ +V + I      +
Sbjct  1258  KVNHPLDFETIKEYNLTVRVENNG-------AQQLASEATVYIQLEDVNDEIPLFTEREQ  1310

Query  341   FTVQKGISSKKLVDYILGTYQAIDEDTNKAASNVKYVM---GRNDG-GYLMIDSKTAEI  395
              TV +G      V  +     AID+D     + V Y +    RN+G  +  I+ ++ EI
Sbjct  1311  ETVLEGEPIGTKVTQV----NAIDKDGTFPNNQVYYYIVDSPRNEGKDFFEINLQSGEI  1365

 Score = 55.8 bits (133),  Expect = 3e-06, Method: Compositional matrix adjust.
 Identities = 49/158 (31%), Positives = 78/158 (49%), Gaps = 24/158 (15%)

Query  102   NDNPPVFSQQIFMGEIEENSASNSLVMILNATDADEPNHLNSKIAFKIVSQEPAGTP--M  159
             ND  P FSQQ++   ++EN+  N+LV  + A+D D  N     + F  V     GT
Sbjct  858   NDEEPKFSQQVYTPNVDENAGPNTLVTTVVASDKDGDN-----VRFGFVG---GGTSSGQ  909

Query  160   FLLSRNTGEVRTLTN--SLDREQASSYRLVVSGADK-------DGEGLSTQCECNIKVKD  210
             F++   TG +R  +   SLDR++   Y L V+  D        D    ++     + + D
Sbjct  910   FVIEEITGVIRLHSKAISLDRDK---YELNVTAMDDGSCCVNGDATIHTSTAVVVVFITD  966

Query  211   VNDNFPMFRD-SQYSARIEENILS-SELLRFQVTDLDE  246
             VNDN P+F+D   Y  ++EE   + S +++ Q TD D+
```

```
Sbjct  967   VNDNKPVFKDCGTYYPKVEEGAPNGSPVIKVQATDEDK  1004

 Score = 53.9 bits (128),  Expect = 1e-05, Method: Compositional matrix adjust.
 Identities = 70/306 (23%), Positives = 126/306 (41%), Gaps = 30/306 (10%)

Query  20    PIAKIT-SDYQATQKITYRI---SGVGIDQPPFGIFVVDKNTGDINITAIVDRE---ETP  72
             PI  +T  D   T K  Y++   SG G D+  F +   +  TG + I   +D E   ++
Sbjct  1768  PILTVTVHDEDETNKFQYKVIDNSGYGADK--FTMVRNNDGTGSLKIVQPLDYEDPLQSN  1825

Query  73    SFLITCRALNAQGLDVEKPLI------LTVKILDINDNPPVFSQQIFMGEIEENSASNSL  126
              F    + +N +G D +           + VK+ DINDN P F +      + EN+
Sbjct  1826  GFRFRIQ-VNDKGEDNDNDKYHVAYSWVVVKLRDINDNKPQFERPNIEVSVYENADVGKT  1884

Query  127   VMILNATDADEPNHLNSKIAFKIVSQEPAGTPMFLLSRNTGEVRTLTNSLDREQASSYRL  186
             +    ATD D+      SK+++ I   + +        S N     T+  +LDRE    +++
Sbjct  1885  LETFKATDPDQGG--KSKVSYAI---DRSSDRQRQFSINQEGTVTIQRNLDREVTPRHQV  1939

Query  187   VVSGADKDGEGLSTQCECNIKVKDVNDNFPMFRDSQYSARIEENILSSELLRFQVTDLDE  246
              +   D      +       + V+D+NDN P F    Y   + E++   +++    TD D+
Sbjct  1940  KILAIDDGIPPKTATATLTVIVQDINDNPPKFL-KDYRPVLPEHVPPRKVVEILATDDDD  1998

Query  247   EYTDNWLAVYFFTSGNEGNW----FEIQTDPRTNEG----ILKVVKALDYEQLQSVKLSI  298
                N     F       +     F+++ D +   G    I+  +++ D EQ +   + I
Sbjct  1999  RSKSNGPPFQFRLDPGADDIIRASFKVEQDQKGANGDGMAIVSSLRSFDREQQKEYLIPI  2058

Query  299   AVKNKA  304
              +K+
Sbjct  2059  VIKDHG  2064

 Score = 52.0 bits (123),  Expect = 4e-05, Method: Compositional matrix adjust.
 Identities = 97/398 (24%), Positives = 163/398 (41%), Gaps = 54/398 (14%)

Query  106   PVFSQQIFMGEIEENSASNSLVMILNATDADEPNHLNSKIAFKIVSQ-EPAGTPMFLLSR  164
             P F    +  EI EN   +S ++ + A       +  + +I + + +Q + AGT  F +
Sbjct  655   PQFYMPSYEAEIPENQKKDSDIISVKAK-----SFADREIRYTLKAQGQGAGT--FNIGP  707

Query  165   NTGEVRTLTNSLDRE---QASSYRLVVSGADKDGEGLSTQCECNIKVKDVNDNFPMFRDS  221
              +G V+ L   LD E   Q   Y LVV+ A +D  G ST  E  I+V DVNDN P F
Sbjct  708   TSGIVK-LAKELDFEDLRQPHVYSLVVT-ATEDSGGFSTSVELTIRVTDVNDNAPKFELP  765

Query  222   QYSAR-IEENI-LSSELLRFQVTDLDEEYTDNWLAVYFFTSGNEGNWFEIQTDPRTNEGI  279
              Y A  ++E+I L + +L+ +  D D          + +  S +    F + ++     GI
Sbjct  766   DYQAHNVDEDIPLGTSILKVKAMDSDSGAN---AEIEYLVSDDH---FAVDSN-----GI  814

Query  280   LKVVKALDYEQLQS-VKLSIAVKNKAEFHQSVISRYRVQSTPVTIQVINVREGIAFRPAS  338
             +    K LD +    +  +  K+K E  ++  +  RV +            E   F
Sbjct  815   IVNNKQLDADNNNAYYEFVVTAKDKGEPAKTGTATVRVYTKNKN------DEEPKFSQQV  868

Query  339   KTFTVQKGISSKKLVDYILGTYQAIDEDTNKAASNVK--YVMGRNDGGYLMIDSKTAEIK  396
              T  V +      LV  ++         ++K   NV+  +V G    G  +I+  T  I+
Sbjct  869   YTPNVDENAGPNTLVTTVVA--------SDKDGDNVRFGFVGGGTSSGQFVIEEITGVIR  920

Query  397   FVK---NMNRDSTFI-VNKTITAEVLAIDEYTGKTSTGTVYYVRVPDFNDN------CPTA  446
                   +++RD   + V             + T  TST  V V + D NDN      C T
Sbjct  921   LHSKAISLDRDKYELNVTAMDDGSCCVNGDATIHTSTAVVVVFITDVNDNKPVFKDCGTY  980

Query  447   VLE-KDAVCSSSSPSVVVSARTLNNRYTGPYTFALEDQP  483
               + ++   + SP + V A   +    G   +++  QP
Sbjct  981   YPKVEEGAPNGSPVIKVQATDEDKGVNGQVKYSIVQQP  1018

 Score = 44.7 bits (104),  Expect = 0.008, Method: Compositional matrix adjust.
 Identities = 48/199 (24%), Positives = 73/199 (37%), Gaps = 19/199 (10%)

Query  27    DYQATQKITYRISGVGIDQPPFGIFVVDKNTGDINITAIVDREETPSFLITCRALNAQGL  86
             D     K++Y I       Q  F I        G + I   +DRE TP    +   A++
Sbjct  1894  DQGGKSKVSYAIDRSSDRQRQFSI----NQEGTVTIQRNLDREVTPRHQVKILAIDDGIP  1949

Query  87    DVEKPLILTVKILDINDNPPVFSQQIFMGEIEENSASNSLVMILNATDADEPNHLNSKIA  146
```

```
                    LTV + DINDNPP F +  +   + E+     +V IL   D D
Sbjct  1950  PKTATATLTVIVQDINDNPPKFLKD-YRPVLPEHVPPRKVVEILATDDDDRSKSNGPPFQ  2008

Query  147   FKIVSQEPAGTPMFLLSRNTGEVRTLTN-----------SLDREQASSYRLVVSGADKDG  195
             F++   +P    +   S    + +   N           S DREQ   Y + +   D
Sbjct  2009  FRL---DPGADDIIRASFKVEQDQKGANGDGMAIVSSLRSFDREQQKEYLIPIVIKDHGN  2065

Query  196   EGLSTQCECNIKVKDVNDN  214
               ++      + + DVNDN
Sbjct  2066  PAMTGTSTLTVVIGDVNDN  2084

 Score = 40.0 bits (92),  Expect = 0.21, Method: Compositional matrix adjust.
 Identities = 89/389 (23%), Positives = 149/389 (38%), Gaps = 62/389 (16%)

Query  75   LITCRALNAQGLDVEKPLILTVKILDINDNPPVFSQQIF--MGEIEENSASNSLVMILNA  132
            +IT +  NA GL       + + + D+ND PP F  +       ++ N+  N+ V  L A
Sbjct  517  IITNQGHNA-GLKYTDNQRVIIHVKDVNDEPPYFINRPLPMQAVVQLNAPPNTPVFTLQA  575

Query  133  TDADEPNHLNSKIAFKIVSQEPAGTPMFLLSRNTGEVRTLTNSLDREQASSYRLVVSGAD  192
             D D     +  I + IV     G   F +   +G VRT   L  +    Y L V   D
Sbjct  576  RDPD----TDHNIHYFIVRDRTGG--RFEVDERSGVVRTRGTDL-FQLDMEYVLYVKAED  628

Query  193  KDGE----GLSTQCECNIKVKDVNDNFPMFRDSQYSARIEEN-ILSSELLRFQVTDLDEE  247
            ++G+        +  E  + +       P F    Y A I EN    S+++  +     +
Sbjct  629  QNGKVDDRRFQSTPEERLSIVG-GKRAPQFYMPSYEAEIPENQKKDSDIISVKA----KS  683

Query  248  YTDNWLAVYFFTSGNEGNWFEIQTDPRTNEGILKVVKALDYEQLQSVKLS----IAVKNK  303
            + D  +        G     F I        GI+K+ K LD+E L+   +      A ++
Sbjct  684  FADREIRYTLKAQGQGAGTFNIGP----TSGIVKLAKELDFEDLRQPHVYSLVVTATEDS  739

Query  304  AEFHQSVISRYRVQSTPVTIQVINVREGIAFRPASKTFTVQKGISSKKLVDYILGT----  359
              F  SV          +TI+V +V +       +  F +    +     D  LGT
Sbjct  740  GGFSTSV---------ELTIRVTDVNDN------APKFELPDYQAHNVDEDIPLGTSILK  784

Query  360  YQAIDEDTNKAASNVKYVMGRN----DGGYLMIDSKTAEIKFVKNMNRDSTFIVNKTITA  415
             +A+D D+  A + ++Y++  +    D   +++++K  +     N N    F+V
Sbjct  785  VKAMDSDSG-ANAEIEYLVSDDHFAVDSNGIIVNNKQLD---ADNNNAYYEFVVTAKDKG  840

Query  416  EVLAIDEYTGKTSTGTVYVRVPDFNDNCP  444
            E         KT T TV V   + ND  P
Sbjct  841  E-------PAKTGTATVRVYTKNKNDEEP  862

 Score = 39.7 bits (91),  Expect = 0.27, Method: Compositional matrix adjust.
 Identities = 54/242 (22%), Positives = 102/242 (42%), Gaps = 22/242 (9%)

Query  4    KFAKPCREGE--DNSKRNP-IAKITSDYQATQKITYRISGVGIDQPPFGIFVVDKNTGDI  60
            +F  P  E E  +N K++  I  + +  A ++I Y +   G      G F +  +G +
Sbjct  656  QFYMPSYEAEIPENQKKDSDIISVKAKSFADREIRYTLKAQGQGA---GTFNIGPTSGIV  712

Query  61   NITAIVDREE--TPSF--LITCRALNAQGLDVEKPLILTVKILDINDNPPVFSQQIFMGE  116
             +   +D E+   P    L+     ++ G      + LT+++ D+NDN P F    +
Sbjct  713  KLAKELDFEDLRQPHVYSLVVTATEDSGGFSTS--VELTIRVTDVNDNAPKFELPDYQAH  770

Query  117  -IEENSASNSLVMILNATDADEPNHLNSKIAFKIVSQEPAGTPMFLLSRNTGEVRTLTNS  175
             ++E+    + ++ + A D+D  +  N++I + +VS +     F +  N   V
Sbjct  771  NVDEDIPLGTSILKVKAMDSD--SGANAEIEY-LVSDD-----HFAVDSNGIIVNNKQLD  822

Query  176  LDREQASSYRLVVSGADKDGEGLSTQCECNIKVKDVNDNFPMFRDSQYSARIEENILSSE  235
             D   A  Y  VV+ DK     +      +  K+ ND  P F    Y+  ++EN   +
Sbjct  823  ADNNNA-YYEFVVTAKDKGEPAKTGTATVRVYTKNKNDEEPKFSQQVYTPNVDENAGPNT  881

Query  236  LL  237
            L+
Sbjct  882  LV  883

>AFUN2_006054.P9870 | transcript=AFUN2_006054.R9870 | gene=AFUN2_006054
| organism=Anopheles_funestus_AfunGA1 | gene_product=neural-cadherin,
transcript variant X17, neural-cadherin,
```

```
transcript variant X22, neural-cadherin, transcript variant
X12, neural-cadherin, transcript variant X1, neural-cadherin,
transcript variant X15, neural-cadherin, transcript variant
X2, neural-cadherin, transcript variant X23, neural-cadherin,
transcript variant X26, neural-cadherin, transcript variant
X24, neural-cadherin, transcript variant X7, neural-cadherin,
transcript variant X6, neural-cadherin, transcript variant
X18, neural-cadherin, transcript variant X3, neural-cadherin,
transcript variant X9, neural-cadherin, transcript variant
X14, neural-cadherin, transcript variant X4, neural-cadherin,
transcript variant X25, neural-cadherin, transcript
variant X21, neural-cadherin, transcript variant X20, neural-cadherin,
transcript variant X19, neural-cadherin, transcript
variant X13, neural-cadherin, transcript variant X11, neural-cadherin,
transcript variant X16, neural-cadherin, transcript
variant X5, neural-cadherin, transcript variant X10,
neural-cadherin, transcript variant X8 | transcript_product=neural-cadherin,
transcript variant X24 | location=AfunGA1_2RL:82372181-82559663(-)
| protein_length=3108 | sequence_SO=chromosome
| SO=protein_coding_gene | is_pseudo=false
Length=3108

 Score = 147 bits (371),  Expect = 3e-35, Method: Compositional matrix adjust.
 Identities = 128/446 (29%), Positives = 217/446 (49%), Gaps = 36/446 (8%)

Query  20    PIAKITS---DYQATQKITYRISGVGID--QPPFGIFVVDKNTGDINITAIVDREET---  71
             PI ++T+   D    Q I Y ++G GID   P    F +++ TG+I +   +DR++
Sbjct  1549  PIMQVTATDGDKDRPQNIVYFLTGQGIDPDNPANSKFDINRTTGEIFVLKPLDRDQPNGR  1608

Query  72    PSFLITCRALNAQGLDVEKPLILTVKILDINDNPPVFSQQIFMGEIEENSASNSLVMILN  131
             P +  T  A +  G  +     + V + DINDN P+F Q ++ G + EN  +  +VM +
Sbjct  1609  PQWRFTVFAQDEGGEGLVGYADVQVNLKDINDNAPIFPQGVYFGNVTENGTAGMVVMTMT  1668

Query  132   ATDADEPNH-LNSKIAFKI---VSQEPAGTPMFLLSRNTGEVRTLTNSLDREQASSYRLV  187
             A D D+PN   N+K+ + I   V +E  G+P+F +  +TG ++T    LDRE+   Y +
Sbjct  1669  AVDYDDPNEGTNAKLIYSIEKNVIEEETGSPIFEIEADTGVIKTAVCCLDRERTPDYSIQ  1728

Query  188   VSGADKDGEGLSTQCECNIKVKDVNDNFPMFRDSQYSARIEENILSS--ELLRFQVTDLD  245
             V     DG GL      +I+VKD+ND  P F   ++   ++E   ++  E+    VT  D
Sbjct  1729  VVAM--DGGGLKGTGTASIRVKDINDMPPQFTKDEWFTEVDETDGTNLPEMPILTVTVHD  1786

Query  246   EEYTDNWLAVYFFTSGNEGNWFEIQTDPRTNEGI--LKVVKALDYE---QLQSVKLSIAV  300
             E+ T+ +       SG   + F +    R N+G   LK+V+ LDYE   Q    +  I V
Sbjct  1787  EDETNKFQYKVIDNSGYGADKFTMV---RNNDGTGSLKIVQPLDYEDPLQSNGFRFRIQV  1843

Query  301   KNKAEFHQSVISRYRVQSTPVTIQVINVREGIAFRPASKTFTVQKGISSKKLVDYILGTY  360
              +K E + +   +Y V  + V +++ ++ +    +P  +   ++  +     V   L T+
Sbjct  1844  NDKGEDNDN--DKYHVAYSWVVVKLRDINDN---KPQFERPNIEVSVYENADVGKTLETF  1898

Query  361   QAIDEDTNKAASNVKYVMGR-NDGGYLMIDSKTAEIKFVKNMNRDSTFIVNKTITAEVLA  419
             +A D D     S V Y + R +D       ++   +   +N++R+    V      ++LA
Sbjct  1899  KATDPDQG-GKSKVSYAIDRSSDRQRQFSINQEGTVTIQRNLDRE----VTPRHQVKILA  1953

Query  420   IDE-YTGKTSTGTVYVRVPDFNDNCP  444
             ID+    KT+T T+ V V D NDN P
Sbjct  1954  IDDGIPPKTATATLTVIVQDINDNPP  1979

 Score = 76.6 bits (187),  Expect = 9e-13, Method: Compositional matrix adjust.
 Identities = 103/418 (25%), Positives = 174/418 (42%), Gaps = 50/418 (12%)

Query  50    IFVVDKNTGDINITAIVDREETPSFLITCRAL----------NAQGLDVEKPLILTVKIL  99
              F ++  +G+I    + DRE+  ++ +   A            N Q   V K   + + I
Sbjct  1365  FFEINLQSGEIFTKVVFDREKQGAYALEVEARDGAPSARPNSNQQPNSVTK--FIRIGIA  1422

Query  100   DINDNPPVFSQQIFMGEIEENSASNSLVMILNATDADEPNHLNSKIAFKIVSQEPAGTPM  159
             D NDNPP F + ++  E++EN      V+ + A D DE    +S+I ++I S    G
Sbjct  1423  DKNDNPPYFDKALYEAEVDENEDIQHTVLTVTAKDHDE----SSRIRYEITSGNIGGA--  1476

Query  160   FLLSRNTGEVRTLTNSLDREQASSYRLVVSGADKDGEGLSTQCECNIKVKDVNDNFPMFR  219
             F +   TG +  +  +LD E    Y L +  +D   E  +      I VKDVND  P F
```

```
Sbjct  1477  FAVKNMTGAI-YVAGALDYETRKRYHLTLVASDSLNENETIIV---INVKDVNDMPPSFP  1532

Query  220   DSQYSARIEENI-LSSELLRFQVTDLDEEYTDNWLAVYFFTSG--NEGNWFEIQTDPRTN  276
                Y   ++E + +   +++   TD D++   N   VYF T    +  N    + D
Sbjct  1533  QQVYERTMDEELAVPFPIMQVTATDGDKDRPQN--IVYFLTGQGIDPDNPANSKFDINRT  1590

Query  277   EGILKVVKALDYEQLQS-VKLSIAVKNKAEFHQSVISRYRVQSTPVTIQVINVREGIAFR  335
              G + V+K LD +Q      +     V  + E  + ++     VQ      + + ++ +
Sbjct  1591  TGEIFVLKPLDRDQPNGRPQWRFTVFAQDEGGEGLVGYADVQ-----VNLKDINDNAPIF  1645

Query  336   PASKTFTVQKGISSKKLVDYILGTYQAIDEDTNKAASNVK--YVMGRN------DGGYLM  387
             P    F     ++        ++ T  A+D D      +N K  Y + +N
Sbjct  1646  PQGVYF---GNVTENGTAGMVVMTMTAVDYDDPNEGTNAKLIYSIEKNVIEEETGSPIFE  1702

Query  388   IDSKTAEIK-FVKNMNRDSTFIVNKTITAEVLAIDEYTGKTSTGTVYVRVPDFNDNCP  444
             I++ T  IK  V  ++R+ T        + +V+A+D    G   TGT  +RV D ND  P
Sbjct  1703  IEADTGVIKTAVCCLDRERT----PDYSIQVVAMDG-GGLKGTGTASIRVKDINDMPP  1755

 Score = 76.3 bits (186),  Expect = 1e-12, Method: Compositional matrix adjust.
 Identities = 90/363 (25%), Positives = 169/363 (47%), Gaps = 40/363 (11%)

Query  51    FVVDKNTGDINITAIVDRE-ETPSFL-ITCRALNAQGLDVEKPLILTVKILDINDNPPVF  108
             F VD+ TG+++    + DRE +   F+ +T +A +      +E      TV+I D+NDNPP+F
Sbjct  1035  FTVDEETGEVSTNKVFDREGDDGKFVSVTVKATDQGEPSLEGVCSFTVEITDVNDNPPLF  1094

Query  109   SQQIFMGEIEENSASNSLVMILNATDADEPNHLNSKIAFKIVS-QEPAGTPMFLLSRNTG  167
              +Q ++  ++++++  + ++ ++A+D D  N  N  I + + + +   P     F +   +G
Sbjct  1095  DRQKYVENVKQDASIGTNILRVSASDEDADN--NGAIVYTLSAPHSPNDLEYFEIQPESG  1152

Query  168   EVRTLTNSLDREQASSYRLVVSGADKDGEGLSTQCECNIKVKDVNDNFPMFRDSQYSA-R  226
              +  L   LDRE   +Y+L      DK     LS   E  I V D  +N P++ ++ Y
Sbjct  1153  WI-VLKKPLDRE---TYKLEAMAQDKGYPPLSRTVEVQIDVVDRANNPPVWDNTVYGPIY  1208

Query  227   IEENI-LSSELLRFQVTDLDEEYTDNWLAVYFFTSG-----NEGNWFEIQTDPRTNE--G  278
             ++EN+ +  +++  + +   E    N    Y    G     N+ + F +Q  P   +
Sbjct  1209  VKENMPVGGKVVSIKASSGIE---GNPTVFYRLMPGSTAQTNKLHTFYLQQRPDNGDTWA  1265

Query  279   ILKVVKALDYEQLQSVKLSIAVKNKAEFHQSVISRYRVQSTPVTIQVINVREGIAFRPAS  338
              +KV   LD+E ++   L++ V+N          ++        V IQ+ +V + I
Sbjct  1266  DIKVNHPLDFETIKEYNLTVRVENNG-------AQQLASEATVYIQLEDVNDEIPLFTER  1318

Query  339   KTFTVQKG--ISSKKLVDYILGTYQAIDEDTNKAASNVKYVM---GRNDG-GYLMIDSKT  392
             +  TV +G  I +K      +    AID+D      + V Y +    RN+G  +  I+ ++
Sbjct  1319  EQETVLEGEPIGTK------VTQVNAIDKDGTFPNNQVYYYIVDSPRNEGKDFFEINLQS  1372

Query  393   AEI  395
              EI
Sbjct  1373  GEI  1375

 Score = 55.8 bits (133),  Expect = 3e-06, Method: Compositional matrix adjust.
 Identities = 49/158 (31%), Positives = 78/158 (49%), Gaps = 24/158 (15%)

Query  102   NDNPPVFSQQIFMGEIEENSASNSLVMILNATDADEPNHLNSKIAFKIVSQEPAGTP--M  159
             ND  P FSQQ++   ++EN+  N+LV  + A+D D  N      + F  V     GT
Sbjct  868   NDEEPKFSQQVYTPNVDENAGPNTLVTTVVASDKDGDN-----VRFGFVG---GGTSSGQ  919

Query  160   FLLSRNTGEVRTLTN--SLDREQASSYRLVVSGADK-------DGEGLSTQCECNIKVKD  210
             F++   TG +R  +   SLDR++   Y L V+  D         D    ++     + + D
Sbjct  920   FVIEEITGVIRLHSKAISLDRDK---YELNVTAMDDGSCCVNGDATIHTSTAVVVVFITD  976

Query  211   VNDNFPMFRD-SQYSARIEENILS-SELLRFQVTDLDE  246
             VNDN P+F+D   Y  ++EE   + S +++ Q TD D+
Sbjct  977   VNDNKPVFKDCGTYYPKVEEGAPNGSPVIKVQATDEDK  1014

 Score = 53.9 bits (128),  Expect = 1e-05, Method: Compositional matrix adjust.
 Identities = 70/306 (23%), Positives = 126/306 (41%), Gaps = 30/306 (10%)

Query  20    PIAKIT-SDYQATQKITYRI---SGVGIDQPPFGIFVVDKNTGDINITAIVDRE---ETP  72
```

```
              PI  +T  D   T K  Y++   SG G D+  F +   +  TG + I   +D E   ++
Sbjct  1778  PILTVTVHDEDETNKFQYKVIDNSGYGADK--FTMVRNNDGTGSLKIVQPLDYEDPLQSN  1835

Query  73    SFLITCRALNAQGLDVEKPLI------LTVKILDINDNPPVFSQQIFMGEIEENSASNSL  126
               F    + +N +G D +           + VK+ DINDN P F +      + EN+
Sbjct  1836  GFRFRIQ-VNDKGEDNDNDKYHVAYSWVVVKLRDINDNKPQFERPNIEVSVYENADVGKT  1894

Query  127   VMILNATDADEPNHLNSKIAFKIVSQEPAGTPMFLLSRNTGEVRTLTNSLDREQASSYRL  186
              +    ATD D+     SK+++ I    + +        S N     T+  +LDRE    +++
Sbjct  1895  LETFKATDPDQGG--KSKVSYAI---DRSSDRQRQFSINQEGTVTIQRNLDREVTPRHQV  1949

Query  187   VVSGADKDGEGLSTQCECNIKVKDVNDNFPMFRDSQYSARIEENILSSELLRFQVTDLDE  246
               +    D       +       + V+D+NDN P F    Y    + E++    +++     TD D+
Sbjct  1950  KILAIDDGIPPKTATATLTVIVQDINDNPPKFL-KDYRPVLPEHVPPRKVVEILATDDDD  2008

Query  247   EYTDNWLAVYFFTSGNEGNW----FEIQTDPRTNEG----ILKVVKALDYEQLQSVKLSI  298
                  N     F        +     F+++ D +    G     I+  +++ D EQ +    + I
Sbjct  2009  RSKSNGPPFQFRLDPGADDIIRASFKVEQDQKGANGDGMAIVSSLRSFDREQQKEYLIPI  2068

Query  299   AVKNKA  304
               +K+
Sbjct  2069  VIKDHG  2074

 Score = 52.4 bits (124),  Expect = 3e-05, Method: Compositional matrix adjust.
 Identities = 97/398 (24%), Positives = 163/398 (41%), Gaps = 54/398 (14%)

Query  106   PVFSQQIFMGEIEENSASNSLVMILNATDADEPNHLNSKIAFKIVSQ-EPAGTPMFLLSR  164
              P F    +  EI EN   +S ++ + A       +  + +I + + + +Q + AGT  F +
Sbjct  665   PQFYMPSYEAEIPENQKKDSDIISVKAK-----SFADREIRYTLKAQGQGAGT--FNIGP  717

Query  165   NTGEVRTLTNSLDRE---QASSYRLVVSGADKDGEGLSTQCECNIKVKDVNDNFPMFRDS  221
               +G V+ L   LD E   Q   Y LVV+ A +D  G ST  E  I+V DVNDN P F
Sbjct  718   TSGIVK-LAKELDFEDLRQPHVYSLVVT-ATEDSGGFSTSVELTIRVTDVNDNAPKFELP  775

Query  222   QYSAR-IEENI-LSSELLRFQVTDLDEEYTDNWLAVYFFTSGNEGNWFEIQTDPRTNEGI  279
               Y A  ++E+I L + +L+ +  D D          + +  S +    F + ++      GI
Sbjct  776   DYQAHNVDEDIPLGTSILKVKAMDADSGAN---AEIEYLVSDDH---FAVDSN-----GI  824

Query  280   LKVVKALDYEQLQS-VKLSIAVKNKAEFHQSVISRYRVQSTPVTIQVINVREGIAFRPAS  338
              +   K LD +   +  +  +  K+K E  ++  +  RV +              E   F
Sbjct  825   IVNNKQLDADNNNAYYEFVVTAKDKGEPAKTGTATVRVYTKNKN------DEEPKFSQQV  878

Query  339   KTFTVQKGISSKKLVDYILGTYQAIDEDTNKAASNVK--YVMGRNDGGYLMIDSKTAEIK  396
               T  V +      LV  ++          ++K   NV+  +V G    G  +I+  T  I+
Sbjct  879   YTPNVDENAGPNTLVTTVVA--------SDKDGDNVRFGFVGGGTSSGQFVIEEITGVIR  930

Query  397   FVK---NMNRDSTFI-VNKTITAEVLAIDEYTGKTSTGTVYVRVPDFNDN------CPTA  446
                    +++RD   + V               + T  TST  V V + D NDN      C T
Sbjct  931   LHSKAISLDRDKYELNVTAMDDGSCCVNGDATIHTSTAVVVVFITDVNDNKPVFKDCGTY  990

Query  447   VLE-KDAVCSSSPSVVVSARTLNNRYTGPYTFALEDQP  483
                + ++    + SP + V A    +    G    +++   QP
Sbjct  991   YPKVEEGAPNGSPVIKVQATDEDKGVNGQVKYSIVQQP  1028

 Score = 44.3 bits (103),  Expect = 0.009, Method: Compositional matrix adjust.
 Identities = 48/199 (24%), Positives = 73/199 (37%), Gaps = 19/199 (10%)

Query  27    DYQATQKITYRISGVGIDQPPFGIFVVDKNTGDINITAIVDREETPSFLITCRALNAQGL  86
              D     K++Y I      Q  F I        G + I   +DRE TP   +   A++
Sbjct  1904  DQGGKSKVSYAIDRSSDRQRQFSI----NQEGTVTIQRNLDREVTPRHQVKILAIDDGIP  1959

Query  87    DVEKPLILTVKILDINDNPPVFSQQIFMGEIEENSASNSLVMILNATDADEPNHLNSKIA  146
                      LTV + DINDNPP F +  +    + E+     +V IL   D D
Sbjct  1960  PKTATATLTVIVQDINDNPPKFLKD-YRPVLPEHVPPRKVVEILATDDDDRSKSNGPPFQ  2018

Query  147   FKIVSQEPAGTPMFLLSRNTGEVRTLTN-----------SLDREQASSYRLVVSGADKDG  195
              F++    +P     +   S     + +   N             S DREQ    Y + +    D
Sbjct  2019  FRL---DPGADDIIRASFKVEQDQKGANGDGMAIVSSLRSFDREQQKEYLIPIVIKDHGN  2075
```

```
Query  196   EGLSTQCECNIKVKDVNDN  214
               ++      + + DVNDN
Sbjct  2076  PAMTGTSTLTVVIGDVNDN  2094

 Score = 40.8 bits (94),  Expect = 0.11, Method: Compositional matrix adjust.
 Identities = 54/235 (23%), Positives = 99/235 (42%), Gaps = 22/235 (9%)

Query  4    KFAKPCREGE--DNSKRNP-IAKITSDYQATQKITYRISGVGIDQPPFGIFVVDKNTGDI  60
            +F  P  E E  +N K++  I  + +   A ++I Y +   G      G F +   +G +
Sbjct  666  QFYMPSYEAEIPENQKKDSDIISVKAKSFADREIRYTLKAQGQGA---GTFNIGPTSGIV  722

Query  61   NITAIVDREE--TPSF--LITCRALNAQGLDVEKPLILTVKILDINDNPPVFSQQIFMGE  116
             +   +D E+   P    L+     ++ G      + LT+++ D+NDN P F    +
Sbjct  723  KLAKELDFEDLRQPHVYSLVVTATEDSGGFSTS--VELTIRVTDVNDNAPKFELPDYQAH  780

Query  117  -IEENSASNSLVMILNATDADEPNHLNSKIAFKIVSQEPAGTPMFLLSRNTGEVRTLTNS  175
             ++E+    + ++ + A DAD  +  N++I + +VS +      F +  N   V
Sbjct  781  NVDEDIPLGTSILKVKAMDAD--SGANAEIEY-LVSDD-----HFAVDSNGIIVNNKQLD  832

Query  176  LDREQASSYRLVVSGADKDGEGLSTQCECNIKVKDVNDNFPMFRDSQYSARIEEN  230
             D   A  Y  VV+  DK     +      +  K+ ND  P F    Y+  ++EN
Sbjct  833  ADNNNA-YYEFVVTAKDKGEPAKTGTATVRVYTKNKNDEEPKFSQQVYTPNVDEN  886

 Score = 39.3 bits (90),  Expect = 0.31, Method: Compositional matrix adjust.
 Identities = 89/389 (23%), Positives = 149/389 (38%), Gaps = 62/389 (16%)

Query  75   LITCRALNAQGLDVEKPLILTVKILDINDNPPVFSQQIF--MGEIEENSASNSLVMILNA  132
            +IT +  NA GL       + + + D+ND PP F  +       ++ N+  N+ V  L A
Sbjct  527  IITNQGHNA-GLKYTDNQRVIIHVKDVNDEPPYFINRPLPMQAVVQLNAPPNTPVFTLQA  585

Query  133  TDADEPNHLNSKIAFKIVSQEPAGTPMFLLSRNTGEVRTLTNSLDREQASSYRLVVSGAD  192
             D D     +  I + IV     G   F +   +G VRT    L  +    Y L V   D
Sbjct  586  RDPD----TDHNIHYFIVRDRTGG--RFEVDERSGVVRTRGTDL-FQLDMEYVLYVKAED  638

Query  193  KDGE----GLSTQCECNIKVKDVNDNFPMFRDSQYSARIEEN-ILSSELLRFQVTDLDEE  247
            ++G+       +  E  + +       P F    Y A I EN    S+++  +     +
Sbjct  639  QNGKVDDRRFQSTPEERLSIVG-GKRAPQFYMPSYEAEIPENQKKDSDIISVKA----KS  693

Query  248  YTDNWLAVYFFTSGNEGNWFEIQTDPRTNEGILKVVKALDYEQLQSVKLS----IAVKNK  303
            + D  +       G     F I        GI+K+ K LD+E L+   +      A ++
Sbjct  694  FADREIRYTLKAQGQGAGTFNIGP----TSGIVKLAKELDFEDLRQPHVYSLVVTATEDS  749

Query  304  AEFHQSVISRYRVQSTPVTIQVINVREGIAFRPASKTFTVQKGISSKKLVDYILGT----  359
              F  SV          +TI+V +V +       +  F +    +     D  LGT
Sbjct  750  GGFSTSV---------ELTIRVTDVNDN------APKFELPDYQAHNVDEDIPLGTSILK  794

Query  360  YQAIDEDTNKAASNVKYVMGRN----DGGYLMIDSKTAEIKFVKNMNRDSTFIVNKTITA  415
             +A+D D+  A + ++Y++  +    D   +++++K  +     N N    F+V
Sbjct  795  VKAMDADSG-ANAEIEYLVSDDHFAVDSNGIIVNNKQLD---ADNNNAYYEFVVTAKDKG  850

Query  416  EVLAIDEYTGKTSTGTVYVRVPDFNDNCP  444
            E         KT T TV V   + ND  P
Sbjct  851  E-------PAKTGTATVRVYTKNKNDEEP  872

>ASTEI20_040830.P55060 | transcript=ASTEI20_040830.R55060 | gene=ASTEI20_040830
| organism=Anopheles_stephensi_UCISS2018 |
gene_product=neural-cadherin, transcript variant X5 |
transcript_product=neural-cadherin,
transcript variant X13 | location=CM023249:57079885-57269506(-)
| protein_length=3108 | sequence_SO=chromosome
| SO=protein_coding_gene | is_pseudo=false
Length=3108

 Score = 147 bits (370),  Expect = 3e-35, Method: Compositional matrix adjust.
 Identities = 128/446 (29%), Positives = 217/446 (49%), Gaps = 36/446 (8%)

Query  20    PIAKITS---DYQATQKITYRISGVGID--QPPFGIFVVDKNTGDINITAIVDREET---  71
             PI ++T+   D    Q I Y ++G GID   P    F +++ TG+I +   +DR++
```

```
Sbjct  1548  PIMQVTATDGDKDRPQNIVYFLTGQGIDPDNPANSKFDINRTTGEIFVLKPLDRDQPNGR  1607

Query  72    PSFLITCRALNAQGLDVEKPLILTVKILDINDNPPVFSQQIFMGEIEENSASNSLVMILN  131
             P +  T  A +  G  +     + V + DINDN P+F Q ++ G + EN  +  +VM +
Sbjct  1608  PQWRFTVFAQDEGGEGLVGYADVQVNLKDINDNAPIFPQGVYFGNVTENGTAGMVVMTMT  1667

Query  132   ATDADEPNH-LNSKIAFKI---VSQEPAGTPMFLLSRNTGEVRTLTNSLDREQASSYRLV  187
             A D D+PN   N+K+ + I   V +E  G+P+F +  +TG ++T    LDRE+   Y +
Sbjct  1668  AVDYDDPNEGTNAKLIYSIEKNVIEEETGSPIFEIEADTGVIKTAVCCLDRERTPDYSIQ  1727

Query  188   VSGADKDGEGLSTQCECNIKVKDVNDNFPMFRDSQYSARIEENILSS--ELLRFQVTDLD  245
             V     DG GL      +I+VKD+ND  P F   ++    ++E   ++  E+    VT  D
Sbjct  1728  VVAM--DGGGLKGTGTASIRVKDINDMPPQFTKDEWFTEVDETDGTNLPEMPILTVTVHD  1785

Query  246   EEYTDNWLAVYFFTSGNEGNWFEIQTDPRTNEGI--LKVVKALDYE---QLQSVKLSIAV  300
             E+ T+ +        SG    + F +    R N+G   LK+V+ LDYE   Q    +  I V
Sbjct  1786  EDETNKFQYKVIDNSGYGADKFTMV---RNNDGTGSLKIVQPLDYEDPLQSNGFRFRIQV  1842

Query  301   KNKAEFHQSVISRYRVQSTPVTIQVINVREGIAFRPASKTFTVQKGISSKKLVDYILGTY  360
              +K E + +   +Y V  + V +++ ++ +    +P  +   ++  +      V   L T+
Sbjct  1843  NDKGEDNDN--DKYHVAYSWVVVKLRDINDN---KPQFERPNIEVSVYENADVGKTLETF  1897

Query  361   QAIDEDTNKAASNVKYVMGR-NDGGYLMIDSKTAEIKFVKNMNRDSTFIVNKTITAEVLA  419
             +A D D     S V Y + R +D        ++    +   +N++R+    V       ++LA
Sbjct  1898  KATDPDQG-GKSKVSYAIDRSSDRQRQFSINQEGTVTIQRNLDRE----VTPRHQVKILA  1952

Query  420   IDE-YTGKTSTGTVYYVRVPDFNDNCP  444
             ID+    KT+T T+ V V D NDN P
Sbjct  1953  IDDGIPPKTATATLTVIVQDINDNPP  1978

 Score = 78.6 bits (192),  Expect = 3e-13, Method: Compositional matrix adjust.
 Identities = 106/424 (25%), Positives = 180/424 (42%), Gaps = 51/424 (12%)

Query  53    VDKNTGDINITAIVDREETPSFLITCRALNAQGLDVEKPLILTVKILDINDNPPVFSQ-Q  111
             +D++  ++N+TA+ D          +C  +N           ++ V I D+NDN PVF
Sbjct  937   LDRDKYELNVTAMDDG--------SC-CVNGDATIHTSTAVVVVFITDVNDNKPVFKDCG  987

Query  112   IFMGEIEENSASNSLVMILNATDADEPNHLNSKIAFKIVSQEPAGTPMFLLSRNTGEVRT  171
              +  ++EE + + S V+ + ATD D+   +N ++ + IV Q       F +   TGEV T
Sbjct  988   TYYPKVEEGAPNGSPVIKVQATDEDK--GVNGQVKYSIVQQPNQKGTKFTVDEETGEVST  1045

Query  172   LTNSLDRE--QASSYRLVVSGADKDGEGLSTQCECNIKVKDVNDNFPMFRDSQYSARIEE  229
                  DRE         + V   D+    L   C    +++ DVNDN P+F    +Y   +++
Sbjct  1046  -NKVFDREGDDGKFVSVTVKATDQGEPSLEGVCSFTVEITDVNDNPPLFDRQKYVENVKQ  1104

Query  230   NI-LSSELLRFQVTDLDEEYTDNWLAVYFFT---SGNEGNWFEIQTDPRTNEGILKVVKA  285
             +  + + +LR  V+  DE+  +N   VY  +   S N+  +FEIQ  P +   +LK  K
Sbjct  1105  DASIGTNILR--VSASDEDADNNGAIVYTLSAPHSPNDLEYFEIQ--PESGWIVLK--KP  1158

Query  286   LDYEQLQSVKLSIAVKNK------AEFHQSVISRYRVQSTPVTIQV----INVREGIAFR  335
             LD E+     +L +   ++      A+     +     R     P+  +     I++RE +
Sbjct  1159  LDRERY---RLKVYASDRGDPPSTADVDVELDVVDRNNKPPIWDRTIYGPIHIRENVTVG  1215

Query  336   PASKTFTVQKGISSKKLVDYILGTYQAIDEDTNKAASNVKYVMGRNDGGYLMIDSKTAEI  395
                 +      GI      V Y L            TNK    +  Y+  R D G         A+I
Sbjct  1216  TVVTSVKASSGIEGNPTVFYRL--MPGSTAQTNKL--HTFYLQQRPDNG-----DTWADI  1266

Query  396   KFVKNMNRDSTFIVNKTITAEVLAIDEYTGKTSTGTVYYVRVPDFNDNCPTAV-LEKDAVC  454
             K    ++ ++    N T+  E      +     S  TVY+++ D ND  P     E++ V
Sbjct  1267  KVNHPLDFETIKEYNLTVRVENNGAQQL---ASEATVYIQLEDVNDEIPLFTEREQETVL  1323

Query  455   SSSP  458
                P
Sbjct  1324  EGEP  1327

 Score = 76.6 bits (187),  Expect = 1e-12, Method: Compositional matrix adjust.
 Identities = 103/418 (25%), Positives = 174/418 (42%), Gaps = 50/418 (12%)

Query  50    IFVVDKNTGDINITAIVDREETPSFLITCRAL----------NAQGLDVEKPLILTVKIL  99
```

```
              F ++  +G+I    + DRE+  ++ +   A           N Q   V K   + + I
Sbjct  1364  FFEINLQSGEIFTKVVFDREKQGAYALEVEARDGAPSARPNSNQQPNSVTK--FIRIGIA  1421

Query  100   DINDNPPVFSQQIFMGEIEENSASNSLVMILNATDADEPNHLNSKIAFKIVSQEPAGTPM  159
             D NDNPP F + ++  E++EN      V+ + A D DE    +S+I ++I S    G
Sbjct  1422  DKNDNPPYFDKALYEAEVDENEDIQHTVLTVTAKDHDE----SSRIRYEITSGNIGGA--  1475

Query  160   FLLSRNTGEVRTLTNSLDREQASSYRLVVSGADKDGEGLSTQCECNIKVKDVNDNFPMFR  219
             F +   TG +  +  +LD E    Y L +  +D   E  +      I VKDVND  P F
Sbjct  1476  FAVKNMTGAI-YVAGALDYETRKRYHLTLVASDSLNENETIIV---INVKDVNDMPPSFP  1531

Query  220   DSQYSARIEENI-LSSELLRFQVTDLDEEYTDNWLAVYFFTSG--NEGNWFEIQTDPRTN  276
                Y   ++E + +   +++   TD D++   N   VYF T    +  N    + D
Sbjct  1532  QQVYERTMDEELAVPFPIMQVTATDGDKDRPQN--IVYFLTGQGIDPDNPANSKFDINRT  1589

Query  277   EGILKVVKALDYEQLQS-VKLSIAVKNKAEFHQSVISRYRVQSTPVTIQVINVREGIAFR  335
              G + V+K LD +Q     +    V  + E  + ++    VQ     + + ++ +
Sbjct  1590  TGEIFVLKPLDRDQPNGRPQWRFTVFAQDEGGEGLVGYADVQ-----VNLKDINDNAPIF  1644

Query  336   PASKTFTVQKGISSKKLVDYILGTYQAIDEDTNKAASNVK--YVMGRN------DGGYLM  387
             P    F     ++       ++ T  A+D D     +N K  Y + +N
Sbjct  1645  PQGVYF---GNVTENGTAGMVVMTMTAVDYDDPNEGTNAKLIYSIEKNVIEEETGSPIFE  1701

Query  388   IDSKTAEIK-FVKNMNRDSTFIVNKTITAEVLAIDEYTGKTSTGTVYVRVPDFNDNCP  444
             I++ T  IK  V  ++R+ T       + +V+A+D   G   TGT  +RV D ND  P
Sbjct  1702  IEADTGVIKTAVCCLDRERT----PDYSIQVVAMDG-GGLKGTGTASIRVKDINDMPP  1754

 Score = 65.5 bits (158),  Expect = 3e-09, Method: Compositional matrix adjust.
 Identities = 85/359 (24%), Positives = 153/359 (43%), Gaps = 32/359 (9%)

Query  51    FVVDKNTGDINITAIVDRE-ETPSFL-ITCRALNAQGLDVEKPLILTVKILDINDNPPVF  108
             F VD+ TG+++   + DRE +   F+ +T +A +     +E     TV+I D+NDNPP+F
Sbjct  1034  FTVDEETGEVSTNKVFDREGDDGKFVSVTVKATDQGEPSLEGVCSFTVEITDVNDNPPLF  1093

Query  109   SQQIFMGEIEENSASNSLVMILNATDADEPNHLNSKIAFKIVS-QEPAGTPMFLLSRNTG  167
              +Q ++  ++++++  + ++ ++A+D D  N  N  I + + +   P     F +   +G
Sbjct  1094  DRQKYVENVKQDASIGTNILRVSASDEDADN--NGAIVYTLSAPHSPNDLEYFEIQPESG  1151

Query  168   EVRTLTNSLDREQASSYRLVVSGADKDGEGLSTQCECNIKVKDVNDNFPMFRDSQYSARI  227
              +  L   LDRE+   YRL V  +D+ G+  ST            +N P   D      I
Sbjct  1152  WI-VLKKPLDRER---YRLKVYASDR-GDPPSTADVDVELDVVDRNNKPPIWDRTIYGPI  1206

Query  228   EENILSSELLRFQVTDLDEEYTDNWLAVYFFTSG-----NEGNWFEIQTDPRTNE--GIL  280
                   +                N    Y    G     N+ + F +Q  P   +    +
Sbjct  1207  HIRENVTVGTVVTSVKASSGIEGNPTVFYRLMPGSTAQTNKLHTFYLQQRPDNGDTWADI  1266

Query  281   KVVKALDYEQLQSVKLSIAVKNKAEFHQSVISRYRVQSTPVTIQVINVREGIAFRPASKT  340
             KV   LD+E ++   L++ V+N         ++       V IQ+ +V + I      +
Sbjct  1267  KVNHPLDFETIKEYNLTVRVENNG-------AQQLASEATVYIQLEDVNDEIPLFTEREQ  1319

Query  341   FTVQKGISSKKLVDYILGTYQAIDEDTNKAASNVKYVM---GRNDG-GYLMIDSKTAEI  395
              TV +G      V  +     AID+D     + V Y +    RN+G  +  I+ ++ EI
Sbjct  1320  ETVLEGEPIGTKVTQV----NAIDKDGTFPNNQVYYYIVDSPRNEGKDFFEINLQSGEI  1374

 Score = 55.8 bits (133),  Expect = 3e-06, Method: Compositional matrix adjust.
 Identities = 49/158 (31%), Positives = 78/158 (49%), Gaps = 24/158 (15%)

Query  102   NDNPPVFSQQIFMGEIEENSASNSLVMILNATDADEPNHLNSKIAFKIVSQEPAGTP--M  159
             ND  P FSQQ++   ++EN+  N+LV  + A+D D  N     + F  V     GT
Sbjct  867   NDEEPKFSQQVYTPNVDENAGPNTLVTTVVASDKDGDN-----VRFGFVG---GGTSSGQ  918

Query  160   FLLSRNTGEVRTLTN--SLDREQASSYRLVVSGADK-------DGEGLSTQCECNIKVKD  210
             F++   TG +R  +   SLDR++   Y L V+  D        D    ++     + + D
Sbjct  919   FVIEEITGVIRLHSKAISLDRDK---YELNVTAMDDGSCCVNGDATIHTSTAVVVVFITD  975

Query  211   VNDNFPMFRD-SQYSARIEENILS-SELLRFQVTDLDE  246
             VNDN P+F+D   Y  ++EE   + S +++ Q TD D+
Sbjct  976   VNDNKPVFKDCGTYYPKVEEGAPNGSPVIKVQATDEDK  1013
```

```
 Score = 53.5 bits (127),  Expect = 1e-05, Method: Compositional matrix adjust.
 Identities = 70/306 (23%), Positives = 126/306 (41%), Gaps = 30/306 (10%)

Query  20    PIAKIT-SDYQATQKITYRI---SGVGIDQPPFGIFVVDKNTGDINITAIVDRE---ETP  72
             PI  +T  D   T K  Y++   SG G D+  F +   +  TG + I   +D E   ++
Sbjct  1777  PILTVTVHDEDETNKFQYKVIDNSGYGADK--FTMVRNNDGTGSLKIVQPLDYEDPLQSN  1834

Query  73    SFLITCRALNAQGLDVEKPLI------LTVKILDINDNPPVFSQQIFMGEIEENSASNSL  126
              F    + +N +G D +           + VK+ DINDN P F +      + EN+
Sbjct  1835  GFRFRIQ-VNDKGEDNDNDKYHVAYSWVVVKLRDINDNKPQFERPNIEVSVYENADVGKT  1893

Query  127   VMILNATDADEPNHLNSKIAFKIVSQEPAGTPMFLLSRNTGEVRTLTNSLDREQASSYRL  186
             +    ATD D+     SK+++ I   + +        S N     T+  +LDRE    +++
Sbjct  1894  LETFKATDPDQGG--KSKVSYAI---DRSSDRQRQFSINQEGTVTIQRNLDREVTPRHQV  1948

Query  187   VVSGADKDGEGLSTQCECNIKVKDVNDNFPMFRDSQYSARIEENILSSELLRFQVTDLDE  246
              +   D      +      + V+D+NDN P F    Y   + E++   +++    TD D+
Sbjct  1949  KILAIDDGIPPKTATATLTVIVQDINDNPPKFL-KDYRPVLPEHVPPRKVVEILATDDDD  2007

Query  247   EYTDNWLAVYFFTSGNEGNW----FEIQTDPRTNEG----ILKVVKALDYEQLQSVKLSI  298
                 N     F       +     F+++ D +   G    I+  +++ D EQ +   + I
Sbjct  2008  RSKSNGPPFQFRLDPGADDIIRASFKVEQDQKGANGDGMAIVSSLRSFDREQQKEYLIPI  2067

Query  299   AVKNKA  304
              +K+
Sbjct  2068  VIKDHG  2073

 Score = 52.8 bits (125),  Expect = 3e-05, Method: Compositional matrix adjust.
 Identities = 97/398 (24%), Positives = 163/398 (41%), Gaps = 54/398 (14%)

Query  106   PVFSQQIFMGEIEENSASNSLVMILNATDADEPNHLNSKIAFKIVSQ-EPAGTPMFLLSR  164
             P F    +  EI EN   +S ++ + A      +  + +I + + +Q + AGT  F +
Sbjct  664   PQFYMPSYEAEIPENQKKDSDIISVKAK-----SFADREIRYTLKAQGQGAGT--FNIGP  716

Query  165   NTGEVRTLTNSLDRE---QASSYRLVVSGADKDGEGLSTQCECNIKVKDVNDNFPMFRDS  221
              +G V+ L    LD E   Q    Y LVV+ A +D  G ST  E  I+V DVNDN P F
Sbjct  717   TSGIVK-LAKELDFEDLRQPHVYSLVVT-ATEDSGGFSTSVELTIRVTDVNDNAPKFELP  774

Query  222   QYSAR-IEENI-LSSELLRFQVTDLDEEYTDNWLAVYFFTSGNEGNWFEIQTDPRTNEGI  279
              Y A  ++E+I L + +L+ +  D D          + +  S +    F + ++     GI
Sbjct  775   DYQAHNVDEDIPLGTSILKVKAMDADSGAN---AEIEYLVSDDH---FAVDSN-----GI  823

Query  280   LKVVKALDYEQLQS-VKLSIAVKNKAEFHQSVISRYRVQSTPVTIQVINVREGIAFRPAS  338
             +   K LD +   +  +  + K+K E  ++  +  RV +             E  F
Sbjct  824   IVNNKQLDADNNNAYYEFVVTAKDKGEPAKTGTATVRVYTKNKN------DEEPKFSQQV  877

Query  339   KTFTVQKGISSKKLVDYILGTYQAIDEDTNKAASNVK--YVMGRNDGGYLMIDSKTAEIK  396
              T  V +       LV  ++          ++K   NV+  +V G    G  +I+  T  I+
Sbjct  878   YTPNVDENAGPNTLVTTVVA--------SDKDGDNVRFGFVGGGTSSGQFVIEEITGVIR  929

Query  397   FVK---NMNRDSTFI-VNKTITAEVLAIDEYTGKTSTGTVYVRVPDFNDN------CPTA  446
                   +++RD   + V              + T  TST  V V + D NDN      C T
Sbjct  930   LHSKAISLDRDKYELNVTAMDDGSCCVNGDATIHTSTAVVVVFITDVNDNKPVFKDCGTY  989

Query  447   VLE-KDAVCSSSPSVVVSARTLNNRYTGPYTFALEDQP  483
               + ++   + SP + V A   +    G   +++  QP
Sbjct  990   YPKVEEGAPNGSPVIKVQATDEDKGVNGQVKYSIVQQP  1027

 Score = 44.3 bits (103),  Expect = 0.009, Method: Compositional matrix adjust.
 Identities = 48/199 (24%), Positives = 73/199 (37%), Gaps = 19/199 (10%)

Query  27    DYQATQKITYRISGVGIDQPPFGIFVVDKNTGDINITAIVDREETPSFLITCRALNAQGL  86
             D     K++Y I      Q  F I        G + I   +DRE TP   +   A++
Sbjct  1903  DQGGKSKVSYAIDRSSDRQRQFSI----NQEGTVTIQRNLDREVTPRHQVKILAIDDGIP  1958

Query  87    DVEKPLILTVKILDINDNPPVFSQQIFMGEIEENSASNSLVMILNATDADEPNHLNSKIA  146
                    LTV + DINDNPP F +  +   + E+     +V IL   D D
Sbjct  1959  PKTATATLTVIVQDINDNPPKFLKD-YRPVLPEHVPPRKVVEILATDDDDRSKSNGPPFQ  2017
```

```
Query  147   FKIVSQEPAGTPMFLLSRNTGEVRTLTN-----------SLDREQASSYRLVVSGADKDG  195
             F++   +P    +   S    + +   N           S DREQ   Y + +   D
Sbjct  2018  FRL---DPGADDIIRASFKVEQDQKGANGDGMAIVSSLRSFDREQQKEYLIPIVIKDHGN  2074

Query  196   EGLSTQCECNIKVKDVNDN  214
               ++       + + DVNDN
Sbjct  2075  PAMTGTSTLTVVIGDVNDN  2093

 Score = 40.8 bits (94),  Expect = 0.13, Method: Compositional matrix adjust.
 Identities = 55/242 (23%), Positives = 102/242 (42%), Gaps = 22/242 (9%)

Query  4    KFAKPCREGE--DNSKRNP-IAKITSDYQATQKITYRISGVGIDQPPFGIFVVDKNTGDI  60
            +F  P  E E  +N K++  I  + +   A ++I Y +   G      G F +   +G +
Sbjct  665  QFYMPSYEAEIPENQKKDSDIISVKAKSFADREIRYTLKAQGQGA---GTFNIGPTSGIV  721

Query  61   NITAIVDREE--TPSF--LITCRALNAQGLDVEKPLILTVKILDINDNPPVFSQQIFMGE  116
             +   +D E+    P    L+     ++ G       + LT+++ D+NDN P F    +
Sbjct  722  KLAKELDFEDLRQPHVYSLVVTATEDSGGFSTS--VELTIRVTDVNDNAPKFELPDYQAH  779

Query  117  -IEENSASNSLVMILNATDADEPNHLNSKIAFKIVSQEPAGTPMFLLSRNTGEVRTLTNS  175
             ++E+    + ++ + A DAD  +  N++I + +VS +      F +  N   V
Sbjct  780  NVDEDIPLGTSILKVKAMDAD--SGANAEIEY-LVSDD-----HFAVDSNGIIVNNKQLD  831

Query  176  LDREQASSYRLVVSGADKDGEGLSTQCECNIKVKDVNDNFPMFRDSQYSARIEENILSSE  235
             D   A  Y  VV+  DK     +       +  K+ ND  P F    Y+  ++EN   +
Sbjct  832  ADNNNA-YYEFVVTAKDKGEPAKTGTATVRVYTKNKNDEEPKFSQQVYTPNVDENAGPNT  890

Query  236  LL  237
            L+
Sbjct  891  LV  892

 Score = 39.3 bits (90),  Expect = 0.31, Method: Compositional matrix adjust.
 Identities = 89/389 (23%), Positives = 149/389 (38%), Gaps = 62/389 (16%)

Query  75   LITCRALNAQGLDVEKPLILTVKILDINDNPPVFSQQIF--MGEIEENSASNSLVMILNA  132
            +IT +  NA GL       + + + D+ND PP F  +       ++ N+  N+ V  L A
Sbjct  526  IITNQGHNA-GLKYTDNQRVIIHVKDVNDEPPYFINRPLPMQAVVQLNAPPNTPVFTLQA  584

Query  133  TDADEPNHLNSKIAFKIVSQEPAGTPMFLLSRNTGEVRTLTNSLDREQASSYRLVVSGAD  192
             D D     +  I + IV     G   F +   +G VRT    L  +    Y L V   D
Sbjct  585  RDPD----TDHNIHYFIVRDRTGG--RFEVDERSGVVRTRGTDL-FQLDMEYVLYVKAED  637

Query  193  KDGE----GLSTQCECNIKVKDVNDNFPMFRDSQYSARIEEN-ILSSELLRFQVTDLDEE  247
            ++G+        +  E  + +       P F    Y A I EN    S+++  +     +
Sbjct  638  QNGKVDDRRFQSTPEERLSIVG-GKRAPQFYMPSYEAEIPENQKKDSDIISVKA----KS  692

Query  248  YTDNWLAVYFFTSGNEGNWFEIQTDPRTNEGILKVVKALDYEQLQSVKLS----IAVKNK  303
            + D  +       G     F I         GI+K+ K LD+E L+   +      A ++
Sbjct  693  FADREIRYTLKAQGQGAGTFNIGP----TSGIVKLAKELDFEDLRQPHVYSLVVTATEDS  748

Query  304  AEFHQSVISRYRVQSTPVTIQVINVREGIAFRPASKTFTVQKGISSKKLVDYILGT----  359
              F  SV          +TI+V +V +       +  F +    +     D  LGT
Sbjct  749  GGFSTSV---------ELTIRVTDVNDN------APKFELPDYQAHNVDEDIPLGTSILK  793

Query  360  YQAIDEDTNKAASNVKYVMGRN----DGGYLMIDSKTAEIKFVKNMNRDSTFIVNKTITA  415
             +A+D D+  A + ++Y++  +    D    +++++K  +     N N    F+V
Sbjct  794  VKAMDADSG-ANAEIEYLVSDDHFAVDSNGIIVNNKQLD---ADNNNAYYEFVVTAKDKG  849

Query  416  EVLAIDEYTGKTSTGTVYVRVPDFNDNCP  444
            E          KT T TV V   + ND  P
Sbjct  850  E-------PAKTGTATVRVYTKNKNDEEP  871

>AGAMI1_007196.P14115 | transcript=AGAMI1_007196.R14115 | gene=AGAMI1_007196
| organism=Anopheles_gambiae_Ifakara | gene_product=neural-cadherin
| transcript_product=neural-cadherin,
transcript variant X16 | location=AgamI1_3RL:41252608-41455387(+)
| protein_length=3098 | sequence_SO=chromosome | SO=protein_coding_gene
```

```
| is_pseudo=false
Length=3098

 Score = 147 bits (370),  Expect = 3e-35, Method: Compositional matrix adjust.
 Identities = 128/446 (29%), Positives = 217/446 (49%), Gaps = 36/446 (8%)

Query  20    PIAKITS---DYQATQKITYRISGVGID--QPPFGIFVVDKNTGDINITAIVDREET---  71
             PI ++T+   D    Q I Y ++G GID   P    F +++ TG+I +    +DR++
Sbjct  1539  PIMQVTATDGDKDRPQNIVYFLTGQGIDPDNPANSKFDINRTTGEIFVLKPLDRDQPNGR  1598

Query  72    PSFLITCRALNAQGLDVEKPLILTVKILDINDNPPVFSQQIFMGEIEENSASNSLVMILN  131
             P +  T  A +  G  +     + V + DINDN P+F Q ++ G + EN  +  +VM +
Sbjct  1599  PQWRFTVFAQDEGGEGLVGYADVQVNLKDINDNAPIFPQGVYFGNVTENGTAGMVVMTMT  1658

Query  132   ATDADEPNH-LNSKIAFKI---VSQEPAGTPMFLLSRNTGEVRTLTNSLDREQASSYRLV  187
             A D D+PN   N+K+ + I   V +E  G+P+F +  +TG ++T    LDRE+   Y +
Sbjct  1659  AVDYDDPNEGTNAKLIYSIEKNVIEEETGSPIFEIEADTGVIKTAVCCLDRERTPDYSIQ  1718

Query  188   VSGADKDGEGLSTQCECNIKVKDVNDNFPMFRDSQYSARIEENILSS--ELLRFQVTDLD  245
             V      DG GL       +I+VKD+ND  P F   ++   ++E   ++  E+    VT  D
Sbjct  1719  VVAM--DGGGLKGTGTASIRVKDINDMPPQFTKDEWFTEVDETDGTNLPEMPILTVTVHD  1776

Query  246   EEYTDNWLAVYFFTSGNEGNWFEIQTDPRTNEGI--LKVVKALDYE---QLQSVKLSIAV  300
             E+ T+ +        SG    + F +    R N+G   LK+V+ LDYE   Q    +  I V
Sbjct  1777  EDETNKFQYKVIDNSGYGADKFTMV---RNNDGTGSLKIVQPLDYEDPLQSNGFRFRIQV  1833

Query  301   KNKAEFHQSVISRYRVQSTPVTIQVINVREGIAFRPASKTFTVQKGISSKKLVDYILGTY  360
              +K E + +   +Y V  + V +++ ++ +    +P  +   ++  +      V   L T+
Sbjct  1834  NDKGEDNDN--DKYHVAYSWVVVKLRDINDN---KPQFERPNIEVSVYENADVGKTLETF  1888

Query  361   QAIDEDTNKAASNVKYVMGR-NDGGYLMIDSKTAEIKFVKNMNRDSTFIVNKTITAEVLA  419
             +A D D     S V Y + R +D        ++   +   +N++R+    V       ++LA
Sbjct  1889  KATDPDQG-GKSKVSYAIDRSSDRQRQFSINQEGTVTIQRNLDRE----VTPRHQVKILA  1943

Query  420   IDE-YTGKTSTGTVYVRVPDFNDNCP  444
             ID+    KT+T T+ V V D NDN P
Sbjct  1944  IDDGIPPKTATATLTVIVQDINDNPP  1969

 Score = 76.3 bits (186),  Expect = 1e-12, Method: Compositional matrix adjust.
 Identities = 89/361 (25%), Positives = 167/361 (46%), Gaps = 36/361 (10%)

Query  51    FVVDKNTGDINITAIVDRE-ETPSFL-ITCRALNAQGLDVEKPLILTVKILDINDNPPVF  108
             F VD+ TG+++    + DRE +   F+ +T +A +      +E      TV+I D+NDNPP+F
Sbjct  1025  FTVDEETGEVSTNKVFDREGDDGKFVSVTVKATDQGEPSLEGVCSFTVEITDVNDNPPLF  1084

Query  109   SQQIFMGEIEENSASNSLVMILNATDADEPNHLNSKIAFKIVS-QEPAGTPMFLLSRNTG  167
              +Q ++  ++++++  + ++ ++A+D D  N  N  I + + +   P     F +   +G
Sbjct  1085  DRQKYVENVKQDASIGTNILRVSASDEDADN--NGAIVYTLSAPHSPNDLEYFEIQPESG  1142

Query  168   EVRTLTNSLDREQASSYRLVVSGADKDGEGLSTQCECNIKVKDVNDNFPMFRDSQYSA-R  226
              +  L   LDRE   +Y+L      DK    LS   E  I V D  +N P++ ++ Y
Sbjct  1143  WI-VLKKPLDRE---TYKLEAMAQDKGYPPLSRTVEVQIDVVDRANNPPVWDNTVYGPIY  1198

Query  227   IEENI-LSSELLRFQVTDLDEEYTDNWLAVYFFTSG-----NEGNWFEIQTDPRTNE--G  278
             ++EN+ +  +++  + +   E    N    Y    G     N+ + F +Q  P   +
Sbjct  1199  VKENMPVGGKVVSIKASSGIE---GNPTVFYRLMPGSTAQTNKLHTFYLQQRPDNGDTWA  1255

Query  279   ILKVVKALDYEQLQSVKLSIAVKNKAEFHQSVISRYRVQSTPVTIQVINVREGIAFRPAS  338
              +KV   LD+E ++   L++ V+N           ++        V IQ+ +V + I
Sbjct  1256  DIKVNHPLDFETIKEYNLTVRVENNG-------AQQLASEATVYIQLEDVNDEIPLFTER  1308

Query  339   KTFTVQKGISSKKLVDYILGTYQAIDEDTNKAASNVKYVM---GRNDG-GYLMIDSKTAE  394
             +  TV +G       V  +      AID+D      + V Y +    RN+G  +  I+ ++ E
Sbjct  1309  EQETVLEGEPIGTKVTQV----NAIDKDGTFPNNQVYYYIVDSPRNEGKDFFEINLQSGE  1364

Query  395   I  395
             I
Sbjct  1365  I  1365
```

```
 Score = 75.9 bits (185),  Expect = 2e-12, Method: Compositional matrix adjust.
 Identities = 103/418 (25%), Positives = 174/418 (42%), Gaps = 50/418 (12%)

Query  50    IFVVDKNTGDINITAIVDREETPSFLITCRAL----------NAQGLDVEKPLILTVKIL  99
              F ++  +G+I    + DRE+  ++ +   A           N Q   V K   + + I
Sbjct  1355  FFEINLQSGEIFTKVVFDREKQGAYALEVEARDGAPSARPNSNQQPNSVTK--FIRIGIA  1412

Query  100   DINDNPPVFSQQIFMGEIEENSASNSLVMILNATDADEPNHLNSKIAFKIVSQEPAGTPM  159
             D NDNPP F + ++  E++EN      V+ + A D DE    +S+I ++I S    G
Sbjct  1413  DKNDNPPYFDKALYEAEVDENEDIQHTVLTVTAKDHDE----SSRIRYEITSGNIGGA--  1466

Query  160   FLLSRNTGEVRTLTNSLDREQASSYRLVVSGADKDGEGLSTQCECNIKVKDVNDNFPMFR  219
             F +   TG +  +  +LD E    Y L +  +D   E  +      I VKDVND  P F
Sbjct  1467  FAVKNMTGAI-YVAGALDYETRKRYYLTLVASDSLNENETIIV---INVKDVNDMPPSFP  1522

Query  220   DSQYSARIEENI-LSSELLRFQVTDLDEEYTDNWLAVYFFTSG--NEGNWFEIQTDPRTN  276
                Y   ++E + +   +++   TD D++   N   VYF T    +  N    + D
Sbjct  1523  QQVYERTMDEELAVPFPIMQVTATDGDKDRPQN--IVYFLTGQGIDPDNPANSKFDINRT  1580

Query  277   EGILKVVKALDYEQLQS-VKLSIAVKNKAEFHQSVISRYRVQSTPVTIQVINVREGIAFR  335
              G + V+K LD +Q     +    V  + E  + ++    VQ     + + ++ +
Sbjct  1581  TGEIFVLKPLDRDQPNGRPQWRFTVFAQDEGGEGLVGYADVQ-----VNLKDINDNAPIF  1635

Query  336   PASKTFTVQKGISSKKLVDYILGTYQAIDEDTNKAASNVK--YVMGRN------DGGYLM  387
             P    F     ++       ++ T  A+D D     +N K  Y + +N
Sbjct  1636  PQGVYF---GNVTENGTAGMVVMTMTAVDYDDPNEGTNAKLIYSIEKNVIEEETGSPIFE  1692

Query  388   IDSKTAEIK-FVKNMNRDSTFIVNKTITAEVLAIDEYTGKTSTGTVYVRVPDFNDNCP  444
             I++ T  IK  V  ++R+ T       + +V+A+D   G   TGT  +RV D ND  P
Sbjct  1693  IEADTGVIKTAVCCLDRERT----PDYSIQVVAMDG-GGLKGTGTASIRVKDINDMPP  1745

 Score = 55.8 bits (133),  Expect = 3e-06, Method: Compositional matrix adjust.
 Identities = 49/158 (31%), Positives = 78/158 (49%), Gaps = 24/158 (15%)

Query  102   NDNPPVFSQQIFMGEIEENSASNSLVMILNATDADEPNHLNSKIAFKIVSQEPAGTP--M  159
             ND  P FSQQ++   ++EN+  N+LV  + A+D D  N     + F  V     GT
Sbjct  858   NDEEPKFSQQVYTPNVDENAGPNTLVTTVVASDKDGDN-----VRFGFVG---GGTSSGQ  909

Query  160   FLLSRNTGEVRTLTN--SLDREQASSYRLVVSGADK-------DGEGLSTQCECNIKVKD  210
             F++   TG +R  +   SLDR++   Y L V+  D        D    ++     + + D
Sbjct  910   FVIEEITGVIRLHSKAISLDRDK---YELNVTAMDDGSCCVNGDATIHTSTAVVVVFITD  966

Query  211   VNDNFPMFRD-SQYSARIEENILS-SELLRFQVTDLDE  246
             VNDN P+F+D   Y  ++EE   + S +++ Q TD D+
Sbjct  967   VNDNKPVFKDCGTYYPKVEEGAPNGSPVIKVQATDEDK  1004

 Score = 53.9 bits (128),  Expect = 1e-05, Method: Compositional matrix adjust.
 Identities = 70/306 (23%), Positives = 126/306 (41%), Gaps = 30/306 (10%)

Query  20    PIAKIT-SDYQATQKITYRI---SGVGIDQPPFGIFVVDKNTGDINITAIVDRE---ETP  72
             PI  +T  D   T K  Y++   SG G D+  F +   +  TG + I   +D E   ++
Sbjct  1768  PILTVTVHDEDETNKFQYKVIDNSGYGADK--FTMVRNNDGTGSLKIVQPLDYEDPLQSN  1825

Query  73    SFLITCRALNAQGLDVEKPLI------LTVKILDINDNPPVFSQQIFMGEIEENSASNSL  126
              F    + +N +G D +          + VK+ DINDN P F +       + EN+
Sbjct  1826  GFRFRIQ-VNDKGEDNDNDKYHVAYSWVVVKLRDINDNKPQFERPNIEVSVYENADVGKT  1884

Query  127   VMILNATDADEPNHLNSKIAFKIVSQEPAGTPMFLLSRNTGEVRTLTNSLDREQASSYRL  186
             +    ATD D+      SK+++ I   + +       S N     T+  +LDRE    +++
Sbjct  1885  LETFKATDPDQGG--KSKVSYAI---DRSSDRQRQFSINQEGTVTIQRNLDREVTPRHQV  1939

Query  187   VVSGADKDGEGLSTQCECNIKVKDVNDNFPMFRDSQYSARIEENILSSELLRFQVTDLDE  246
              +   D      +      + V+D+NDN P F    Y   + E++   +++    TD D+
Sbjct  1940  KILAIDDGIPPKTATATLTVIVQDINDNPPKFL-KDYRPVLPEHVPPRKVVEILATDDDD  1998

Query  247   EYTDNWLAVYFFTSGNEGNW----FEIQTDPRTNEG----ILKVVKALDYEQLQSVKLSI  298
                 N     F       +     F+++ D +   G    I+  +++ D EQ +   + I
Sbjct  1999  RSKSNGPPFQFRLDPGADDIIRASFKVEQDQKGANGDGMAIVSSLRSFDREQQKEYLIPI  2058
```

```
Query  299   AVKNKA  304
              +K+
Sbjct  2059  VIKDHG  2064

 Score = 52.0 bits (123),  Expect = 4e-05, Method: Compositional matrix adjust.
 Identities = 97/398 (24%), Positives = 163/398 (41%), Gaps = 54/398 (14%)

Query  106   PVFSQQIFMGEIEENSASNSLVMILNATDADEPNHLNSKIAFKIVSQ-EPAGTPMFLLSR  164
             P F    +  EI EN   +S ++ + A      +  + +I + + +Q + AGT  F +
Sbjct  655   PQFYMPSYEAEIPENQKKDSDIISVKAK-----SFADREIRYTLKAQGQGAGT--FNIGP  707

Query  165   NTGEVRTLTNSLDRE---QASSYRLVVSGADKDGEGLSTQCECNIKVKDVNDNFPMFRDS  221
              +G V+ L   LD E   Q   Y LVV+ A +D  G ST  E  I+V DVNDN P F
Sbjct  708   TSGIVK-LAKELDFEDLRQPHVYSLVVT-ATEDSGGFSTSVELTIRVTDVNDNAPKFELP  765

Query  222   QYSAR-IEENI-LSSELLRFQVTDLDEEYTDNWLAVYFFTSGNEGNWFEIQTDPRTNEGI  279
              Y A  ++E+I L + +L+ +  D D         + +  S +    F + ++     GI
Sbjct  766   DYQAHNVDEDIPLGTSILKVKAMDSDSGAN---AEIEYLVSDDH---FAVDSN-----GI  814

Query  280   LKVVKALDYEQLQS-VKLSIAVKNKAEFHQSVISRYRVQSTPVTIQVINVREGIAFRPAS  338
             +   K LD +   +  +  +  K+K E  ++  +  RV +           E   F
Sbjct  815   IVNNKQLDADNNNAYYEFVVTAKDKGEPAKTGTATVRVYTKNKN------DEEPKFSQQV  868

Query  339   KTFTVQKGISSKKLVDYILGTYQAIDEDTNKAASNVK--YVMGRNDGGYLMIDSKTAEIK  396
              T  V +      LV  ++         ++K   NV+  +V G    G  +I+  T  I+
Sbjct  869   YTPNVDENAGPNTLVTTVVA--------SDKDGDNVRFGFVGGGTSSGQFVIEEITGVIR  920

Query  397   FVK---NMNRDSTFI-VNKTITAEVLAIDEYTGKTSTGTVYVRVPDFNDN------CPTA  446
                   +++RD   + V            + T  TST  V V + D NDN      C T
Sbjct  921   LHSKAISLDRDKYELNVTAMDDGSCCVNGDATIHTSTAVVVVFITDVNDNKPVFKDCGTY  980

Query  447   VLE-KDAVCSSSPSVVVSARTLNNRYTGPYTFALEDQP  483
               + ++   + SP + V A   +    G   +++  QP
Sbjct  981   YPKVEEGAPNGSPVIKVQATDEDKGVNGQVKYSIVQQP  1018

 Score = 44.7 bits (104),  Expect = 0.008, Method: Compositional matrix adjust.
 Identities = 48/199 (24%), Positives = 73/199 (37%), Gaps = 19/199 (10%)

Query  27    DYQATQKITYRISGVGIDQPPFGIFVVDKNTGDINITAIVDREETPSFLITCRALNAQGL  86
             D     K++Y I      Q  F I       G + I   +DRE TP   +   A++
Sbjct  1894  DQGGKSKVSYAIDRSSDRQRQFSI----NQEGTVTIQRNLDREVTPRHQVKILAIDDGIP  1949

Query  87    DVEKPLILTVKILDINDNPPVFSQQIFMGEIEENSASNSLVMILNATDADEPNHLNSKIA  146
                    LTV + DINDNPP F +  +   + E+     +V IL   D D
Sbjct  1950  PKTATATLTVIVQDINDNPPKFLKD-YRPVLPEHVPPRKVVEILATDDDDRSKSNGPPFQ  2008

Query  147   FKIVSQEPAGTPMFLLSRNTGEVRTLTN-----------SLDREQASSYRLVVSGADKDG  195
             F++   +P    +   S    + +   N           S DREQ   Y + +   D
Sbjct  2009  FRL---DPGADDIIRASFKVEQDQKGANGDGMAIVSSLRSFDREQQKEYLIPIVIKDHGN  2065

Query  196   EGLSTQCECNIKVKDVNDN  214
               ++      + + DVNDN
Sbjct  2066  PAMTGTSTLTVVIGDVNDN  2084

 Score = 39.7 bits (91),  Expect = 0.22, Method: Compositional matrix adjust.
 Identities = 89/389 (23%), Positives = 149/389 (38%), Gaps = 62/389 (16%)

Query  75   LITCRALNAQGLDVEKPLILTVKILDINDNPPVFSQQIF--MGEIEENSASNSLVMILNA  132
            +IT +  NA GL       + + + D+ND PP F  +       ++ N+  N+ V  L A
Sbjct  517  IITNQGHNA-GLKYTDNQRVIIHVKDVNDEPPYFINRPLPMQAVVQLNAPPNTPVFTLQA  575

Query  133  TDADEPNHLNSKIAFKIVSQEPAGTPMFLLSRNTGEVRTLTNSLDREQASSYRLVVSGAD  192
             D D     +  I + IV     G   F +   +G VRT    L  +    Y L V   D
Sbjct  576  RDPD----TDHNIHYFIVRDRTGG--RFEVDERSGVVRTRGTDL-FQLDMEYVLYVKAED  628

Query  193  KDGE----GLSTQCECNIKVKDVNDNFPMFRDSQYSARIEEN-ILSSELLRFQVTDLDEE  247
            ++G+       +  E  + +       P F    Y A I EN    S+++  +     +
Sbjct  629  QNGKVDDRRFQSTPEERLSIVG-GKRAPQFYMPSYEAEIPENQKKDSDIISVKA----KS  683
```

```
Query  248  YTDNWLAVYFFTSGNEGNWFEIQTDPRTNEGILKVVKALDYEQLQSVKLS----IAVKNK  303
            + D  +       G     F I        GI+K+ K LD+E L+   +      A ++
Sbjct  684  FADREIRYTLKAQGQGAGTFNIGP----TSGIVKLAKELDFEDLRQPHVYSLVVTATEDS  739

Query  304  AEFHQSVISRYRVQSTPVTIQVINVREGIAFRPASKTFTVQKGISSKKLVDYILGT----  359
              F  SV          +TI+V +V +       +  F +    +     D  LGT
Sbjct  740  GGFSTSV---------ELTIRVTDVNDN------APKFELPDYQAHNVDEDIPLGTSILK  784

Query  360  YQAIDEDTNKAASNVKYVMGRN----DGGYLMIDSKTAEIKFVKNMNRDSTFIVNKTITA  415
             +A+D D+  A + ++Y++  +    D   +++++K  +     N N    F+V
Sbjct  785  VKAMDSDSG-ANAEIEYLVSDDHFAVDSNGIIVNNKQLD---ADNNNAYYEFVVTAKDKG  840

Query  416  EVLAIDEYTGKTSTGTVYVRVPDFNDNCP  444
            E         KT T TV V   + ND  P
Sbjct  841  E-------PAKTGTATVRVYTKNKNDEEP  862

 Score = 39.7 bits (91),  Expect = 0.27, Method: Compositional matrix adjust.
 Identities = 54/242 (22%), Positives = 102/242 (42%), Gaps = 22/242 (9%)

Query  4    KFAKPCREGE--DNSKRNP-IAKITSDYQATQKITYRISGVGIDQPPFGIFVVDKNTGDI  60
            +F  P  E E  +N K++  I  + +   A ++I Y +   G      G F +   +G +
Sbjct  656  QFYMPSYEAEIPENQKKDSDIISVKAKSFADREIRYTLKAQGQGA---GTFNIGPTSGIV  712

Query  61   NITAIVDREE--TPSF--LITCRALNAQGLDVEKPLILTVKILDINDNPPVFSQQIFMGE  116
             +   +D E+   P    L+     ++ G      + LT+++ D+NDN P F    +
Sbjct  713  KLAKELDFEDLRQPHVYSLVVTATEDSGGFSTS--VELTIRVTDVNDNAPKFELPDYQAH  770

Query  117  -IEENSASNSLVMILNATDADEPNHLNSKIAFKIVSQEPAGTPMFLLSRNTGEVRTLTNS  175
             ++E+    + ++ + A D+D  +  N++I + +VS +      F +  N   V
Sbjct  771  NVDEDIPLGTSILKVKAMDSD--SGANAEIEY-LVSDD-----HFAVDSNGIIVNNKQLD  822

Query  176  LDREQASSYRLVVSGADKDGEGLSTQCECNIKVKDVNDNFPMFRDSQYSARIEENILSSE  235
             D   A  Y  VV+  DK     +      +  K+ ND  P F    Y+  ++EN   +
Sbjct  823  ADNNNA-YYEFVVTAKDKGEPAKTGTATVRVYTKNKNDEEPKFSQQVYTPNVDENAGPNT  881

Query  236  LL  237
            L+
Sbjct  882  LV  883

>ACON2_034794.P62473 | transcript=ACON2_034794.R62473 | gene=ACON2_034794
| organism=Anopheles_coluzzii_AcolN3 | gene_product=neural-cadherin,
transcript variant X15, neural-cadherin,
transcript variant X13, neural-cadherin, transcript variant
X8, neural-cadherin, transcript variant X1, neural-cadherin,
transcript variant X3, neural-cadherin, transcript variant
X10, neural-cadherin, transcript variant X16, neural-cadherin,
transcript variant X2, neural-cadherin, transcript variant
X12, neural-cadherin, transcript variant X4, neural-cadherin,
transcript variant X7, neural-cadherin, transcript variant
X9, neural-cadherin, transcript variant X5, neural-cadherin,
transcript variant X17, neural-cadherin, transcript variant
X14, neural-cadherin, transcript variant X11, neural-cadherin,
transcript variant X6 | transcript_product=neural-cadherin,
transcript variant X15 | location=AcolN3_3RL:40912217-41119468(+)
| protein_length=3098 | sequence_SO=chromosome
| SO=protein_coding_gene | is_pseudo=false
Length=3098

 Score = 147 bits (370),  Expect = 3e-35, Method: Compositional matrix adjust.
 Identities = 128/446 (29%), Positives = 217/446 (49%), Gaps = 36/446 (8%)

Query  20    PIAKITS---DYQATQKITYRISGVGID--QPPFGIFVVDKNTGDINITAIVDREET---  71
             PI ++T+   D    Q I Y ++G GID   P    F +++ TG+I +   +DR++
Sbjct  1539  PIMQVTATDGDKDRPQNIVYFLTGQGIDPDNPANSKFDINRTTGEIFVLKPLDRDQPNGR  1598

Query  72    PSFLITCRALNAQGLDVEKPLILTVKILDINDNPPVFSQQIFMGEIEENSASNSLVMILN  131
             P +  T  A +  G  +     + V +  DINDN P+F Q ++ G + EN  +  +VM +
Sbjct  1599  PQWRFTVFAQDEGGEGLVGYADVQVNLKDINDNAPIFPQGVYFGNVTENGTAGMVVMTMT  1658
```

```
Query  132   ATDADEPNH-LNSKIAFKI---VSQEPAGTPMFLLSRNTGEVRTLTNSLDREQASSYRLV  187
             A D D+PN   N+K+ + I   V +E  G+P+F +  +TG ++T    LDRE+   Y +
Sbjct  1659  AVDYDDPNEGTNAKLIYSIEKNVIEEETGSPIFEIEADTGVIKTAVCCLDRERTPDYSIQ  1718

Query  188   VSGADKDGEGLSTQCECNIKVKDVNDNFPMFRDSQYSARIEENILSS--ELLRFQVTDLD  245
             V      DG GL       +I+VKD+ND  P F   ++   ++E   ++  E+    VT  D
Sbjct  1719  VVAM--DGGGLKGTGTASIRVKDINDMPPQFTKDEWFTEVDETDGTNLPEMPILTVTVHD  1776

Query  246   EEYTDNWLAVYFFTSGNEGNWFEIQTDPRTNEGI--LKVVKALDYE---QLQSVKLSIAV  300
             E+ T+ +        SG    + F +    R N+G   LK+V+ LDYE   Q    +  I V
Sbjct  1777  EDETNKFQYKVIDNSGYGADKFTMV---RNNDGTGSLKIVQPLDYEDPLQSNGFRFRIQV  1833

Query  301   KNKAEFHQSVISRYRVQSTPVTIQVINVREGIAFRPASKTFTVQKGISSKKLVDYILGTY  360
              +K E + +   +Y V  + V +++ ++ +    +P  +   ++  +     V   L T+
Sbjct  1834  NDKGEDNDN--DKYHVAYSWVVVKLRDINDN---KPQFERPNIEVSVYENADVGKTLETF  1888

Query  361   QAIDEDTNKAASNVKYVMGR-NDGGYLMIDSKTAEIKFVKNMNRDSTFIVNKTITAEVLA  419
             +A D D     S V Y + R +D        ++   +   +N++R+    V       ++LA
Sbjct  1889  KATDPDQG-GKSKVSYAIDRSSDRQRQFSINQEGTVTIQRNLDRE----VTPRHQVKILA  1943

Query  420   IDE-YTGKTSTGTVYVRVPDFNDNCP  444
             ID+    KT+T T+ V V D NDN P
Sbjct  1944  IDDGIPPKTATATLTVIVQDINDNPP  1969

 Score = 76.3 bits (186),  Expect = 1e-12, Method: Compositional matrix adjust.
 Identities = 89/361 (25%), Positives = 167/361 (46%), Gaps = 36/361 (10%)

Query  51    FVVDKNTGDINITAIVDRE-ETPSFL-ITCRALNAQGLDVEKPLILTVKILDINDNPPVF  108
             F VD+ TG+++    + DRE +   F+ +T +A +      +E      TV+I D+NDNPP+F
Sbjct  1025  FTVDEETGEVSTNKVFDREGDDGKFVSVTVKATDQGEPSLEGVCSFTVEITDVNDNPPLF  1084

Query  109   SQQIFMGEIEENSASNSLVMILNATDADEPNHLNSKIAFKIVS-QEPAGTPMFLLSRNTG  167
              +Q ++  ++++++  + ++ ++A+D D  N  N  I + + +   P     F +   +G
Sbjct  1085  DRQKYVENVKQDASIGTNILRVSASDEDADN--NGAIVYTLSAPHSPNDLEYFEIQPESG  1142

Query  168   EVRTLTNSLDREQASSYRLVVSGADKDGEGLSTQCECNIKVKDVNDNFPMFRDSQYSA-R  226
              +  L   LDRE   +Y+L     DK    LS   E  I V D  +N P++ ++ Y
Sbjct  1143  WI-VLKKPLDRE---TYKLEAMAQDKGYPPLSRTVEVQIDVVDRANNPPVWDNTVYGPIY  1198

Query  227   IEENI-LSSELLRFQVTDLDEEYTDNWLAVYFFTSG-----NEGNWFEIQTDPRTNE--G  278
             ++EN+ +  +++  + +   E    N    Y    G     N+ + F +Q  P   +
Sbjct  1199  VKENMPVGGKVVSIKASSGIE---GNPTVFYRLMPGSTAQTNKLHTFYLQQRPDNGDTWA  1255

Query  279   ILKVVKALDYEQLQSVKLSIAVKNKAEFHQSVISRYRVQSTPVTIQVINVREGIAFRPAS  338
              +KV   LD+E ++   L++ V+N          ++        V IQ+ +V + I
Sbjct  1256  DIKVNHPLDFETIKEYNLTVRVENNG-------AQQLASEATVYIQLEDVNDEIPLFTER  1308

Query  339   KTFTVQKGISSKKLVDYILGTYQAIDEDTNKAASNVKYVM---GRNDG-GYLMIDSKTAE  394
             +  TV +G      V  +     AID+D     + V Y +    RN+G  +  I+ ++ E
Sbjct  1309  EQETVLEGEPIGTKVTQV----NAIDKDGTFPNNQVYYYIVDSPRNEGKDFFEINLQSGE  1364

Query  395   I  395
             I
Sbjct  1365  I  1365

 Score = 75.9 bits (185),  Expect = 2e-12, Method: Compositional matrix adjust.
 Identities = 103/418 (25%), Positives = 174/418 (42%), Gaps = 50/418 (12%)

Query  50    IFVVDKNTGDINITAIVDREETPSFLITCRAL----------NAQGLDVEKPLILTVKIL  99
              F ++  +G+I     + DRE+  ++ +   A             N Q   V K   + + I
Sbjct  1355  FFEINLQSGEIFTKVVFDREKQGAYALEVEARDGAPSARPNSNQQPNSVTK--FIRIGIA  1412

Query  100   DINDNPPVFSQQIFMGEIEENSASNSLVMILNATDADEPNHLNSKIAFKIVSQEPAGTPM  159
             D NDNPP F + ++  E++EN       V+ + A D DE    +S+I ++I S    G
Sbjct  1413  DKNDNPPYFDKALYEAEVDENEDIQHTVLTVTAKDHDE----SSRIRYEITSGNIGGA--  1466

Query  160   FLLSRNTGEVRTLTNSLDREQASSYRLVVSGADKDGEGLSTQCECNIKVKDVNDNFPMFR  219
             F +    TG +  +  +LD E    Y L +  +D   E  +       I VKDVND  P F
```

```
Sbjct  1467   FAVKNMTGAI-YVAGALDYETRKRYYLTLVASDSLNENETIIV---INVKDVNDMPPSFP  1522

Query  220    DSQYSARIEENI-LSSELLRFQVTDLDEEYTDNWLAVYFFTSG--NEGNWFEIQTDPRTN  276
                 Y   ++E + +   +++   TD D++   N   VYF T    +  N    + D
Sbjct  1523   QQVYERTMDEELAVPFPIMQVTATDGDKDRPQN--IVYFLTGQGIDPDNPANSKFDINRT  1580

Query  277    EGILKVVKALDYEQLQS-VKLSIAVKNKAEFHQSVISRYRVQSTPVTIQVINVREGIAFR  335
               G + V+K LD +Q      +    V  + E  + ++    VQ     + + ++ +
Sbjct  1581   TGEIFVLKPLDRDQPNGRPQWRFTVFAQDEGGEGLVGYADVQ-----VNLKDINDNAPIF  1635

Query  336    PASKTFTVQKGISSKKLVDYILGTYQAIDEDTNKAASNVK--YVMGRN------DGGYLM  387
              P    F     ++        ++ T  A+D D     +N K  Y + +N
Sbjct  1636   PQGVYF---GNVTENGTAGMVVMTMTAVDYDDPNEGTNAKLIYSIEKNVIEEETGSPIFE  1692

Query  388    IDSKTAEIK-FVKNMNRDSTFIVNKTITAEVLAIDEYTGKTSTGTVYVRVPDFNDNCP  444
              I++ T  IK  V  ++R+ T        + +V+A+D   G   TGT  +RV D ND  P
Sbjct  1693   IEADTGVIKTAVCCLDRERT----PDYSIQVVAMDG-GGLKGTGTASIRVKDINDMPP  1745

 Score = 55.8 bits (133),  Expect = 3e-06, Method: Compositional matrix adjust.
 Identities = 49/158 (31%), Positives = 78/158 (49%), Gaps = 24/158 (15%)

Query  102    NDNPPVFSQQIFMGEIEENSASNSLVMILNATDADEPNHLNSKIAFKIVSQEPAGTP--M  159
              ND  P FSQQ++   ++EN+  N+LV  + A+D D  N      + F  V     GT
Sbjct  858    NDEEPKFSQQVYTPNVDENAGPNTLVTTVVASDKDGDN-----VRFGFVG---GGTSSGQ  909

Query  160    FLLSRNTGEVRTLTN--SLDREQASSYRLVVSGADK-------DGEGLSTQCECNIKVKD  210
              F++   TG +R  +   SLDR++   Y L V+  D         D    ++      + + D
Sbjct  910    FVIEEITGVIRLHSKAISLDRDK---YELNVTAMDDGSCCVNGDATIHTSTAVVVVFITD  966

Query  211    VNDNFPMFRD-SQYSARIEENILS-SELLRFQVTDLDE  246
              VNDN P+F+D   Y  ++EE   + S +++ Q TD D+
Sbjct  967    VNDNKPVFKDCGTYYPKVEEGAPNGSPVIKVQATDEDK  1004

 Score = 53.9 bits (128),  Expect = 1e-05, Method: Compositional matrix adjust.
 Identities = 70/306 (23%), Positives = 126/306 (41%), Gaps = 30/306 (10%)

Query  20     PIAKIT-SDYQATQKITYRI---SGVGIDQPPFGIFVVDKNTGDINITAIVDRE---ETP  72
              PI  +T  D   T K  Y++   SG G D+  F +    +  TG + I   +D E   ++
Sbjct  1768   PILTVTVHDEDETNKFQYKVIDNSGYGADK--FTMVRNNDGTGSLKIVQPLDYEDPLQSN  1825

Query  73     SFLITCRALNAQGLDVEKPLI------LTVKILDINDNPPVFSQQIFMGEIEENSASNSL  126
               F    + +N +G D +           + VK+ DINDN P F +      + EN+
Sbjct  1826   GFRFRIQ-VNDKGEDNDNDKYHVAYSWVVVKLRDINDNKPQFERPNIEVSVYENADVGKT  1884

Query  127    VMILNATDADEPNHLNSKIAFKIVSQEPAGTPMFLLSRNTGEVRTLTNSLDREQASSYRL  186
              +    ATD D+     SK+++ I    + +        S N     T+  +LDRE    +++
Sbjct  1885   LETFKATDPDQGG--KSKVSYAI---DRSSDRQRQFSINQEGTVTIQRNLDREVTPRHQV  1939

Query  187    VVSGADKDGEGLSTQCECNIKVKDVNDNFPMFRDSQYSARIEENILSSELLRFQVTDLDE  246
               +   D      +       + V+D+NDN P F    Y   + E++   +++    TD D+
Sbjct  1940   KILAIDDGIPPKTATATLTVIVQDINDNPPKFL-KDYRPVLPEHVPPRKVVEILATDDDD  1998

Query  247    EYTDNWLAVYFFTSGNEGNW----FEIQTDPRTNEG----ILKVVKALDYEQLQSVKLSI  298
                  N     F        +     F+++ D +   G    I+  +++ D EQ +   + I
Sbjct  1999   RSKSNGPPFQFRLDPGADDIIRASFKVEQDQKGANGDGMAIVSSLRSFDREQQKEYLIPI  2058

Query  299    AVKNKA  304
               +K+
Sbjct  2059   VIKDHG  2064

 Score = 52.0 bits (123),  Expect = 4e-05, Method: Compositional matrix adjust.
 Identities = 97/398 (24%), Positives = 163/398 (41%), Gaps = 54/398 (14%)

Query  106    PVFSQQIFMGEIEENSASNSLVMILNATDADEPNHLNSKIAFKIVSQ-EPAGTPMFLLSR  164
              P F    +  EI EN   +S ++ + A       +  + +I + + +Q + AGT  F +
Sbjct  655    PQFYMPSYEAEIPENQKKDSDIISVKAK-----SFADREIRYTLKAQGQGAGT--FNIGP  707

Query  165    NTGEVRTLTNSLDRE---QASSYRLVVSGADKDGEGLSTQCECNIKVKDVNDNFPMFRDS  221
```

```
              +G V+ L   LD E   Q   Y LVV+ A +D  G ST  E  I+V DVNDN P F
Sbjct  708    TSGIVK-LAKELDFEDLRQPHVYSLVVT-ATEDSGGFSTSVELTIRVTDVNDNAPKFELP  765

Query  222    QYSAR-IEENI-LSSELLRFQVTDLDEEYTDNWLAVYFFTSGNEGNWFEIQTDPRTNEGI  279
               Y A  ++E+I L + +L+ +  D D         + +  S +    F + ++     GI
Sbjct  766    DYQAHNVDEDIPLGTSILKVKAMDSDSGAN---AEIEYLVSDDH---FAVDSN-----GI  814

Query  280    LKVVKALDYEQLQS-VKLSIAVKNKAEFHQSVISRYRVQSTPVTIQVINVREGIAFRPAS  338
              +   K LD +   +  +  +  K+K E  ++  +  RV +           E   F
Sbjct  815    IVNNKQLDADNNNAYYEFVVTAKDKGEPAKTGTATVRVYTKNKN------DEEPKFSQQV  868

Query  339    KTFTVQKGISSKKLVDYILGTYQAIDEDTNKAASNVK--YVMGRNDGGYLMIDSKTAEIK  396
               T  V +      LV  ++         ++K   NV+  +V G    G  +I+  T  I+
Sbjct  869    YTPNVDENAGPNTLVTTVVA--------SDKDGDNVRFGFVGGGTSSGQFVIEEITGVIR  920

Query  397    FVK---NMNRDSTFI-VNKTITAEVLAIDEYTGKTSTGTVYVRVPDFNDN------CPTA  446
                    +++RD   + V            + T  TST  V V + D NDN      C T
Sbjct  921    LHSKAISLDRDKYELNVTAMDDGSCCVNGDATIHTSTAVVVVFITDVNDNKPVFKDCGTY  980

Query  447    VLE-KDAVCSSSPSVVVSARTLNNRYTGPYTFALEDQP  483
                + ++   + SP + V A   +    G   +++  QP
Sbjct  981    YPKVEEGAPNGSPVIKVQATDEDKGVNGQVKYSIVQQP  1018

 Score = 44.7 bits (104),  Expect = 0.008, Method: Compositional matrix adjust.
 Identities = 48/199 (24%), Positives = 73/199 (37%), Gaps = 19/199 (10%)

Query  27     DYQATQKITYRISGVGIDQPPFGIFVVDKNTGDINITAIVDREETPSFLITCRALNAQGL  86
              D     K++Y I      Q  F I       G + I   +DRE TP   +   A++
Sbjct  1894   DQGGKSKVSYAIDRSSDRQRQFSI----NQEGTVTIQRNLDREVTPRHQVKILAIDDGIP  1949

Query  87     DVEKPLILTVKILDINDNPPVFSQQIFMGEIEENSASNSLVMILNATDADEPNHLNSKIA  146
                     LTV + DINDNPP F +  +   + E+     +V IL   D D
Sbjct  1950   PKTATATLTVIVQDINDNPPKFLKD-YRPVLPEHVPPRKVVEILATDDDDRSKSNGPPFQ  2008

Query  147    FKIVSQEPAGTPMFLLSRNTGEVRTLTN-----------SLDREQASSYRLVVSGADKDG  195
              F++   +P    +   S    + +   N           S DREQ   Y + +   D
Sbjct  2009   FRL---DPGADDIIRASFKVEQDQKGANGDGMAIVSSLRSFDREQQKEYLIPIVIKDHGN  2065

Query  196    EGLSTQCECNIKVKDVNDN  214
                ++      + + DVNDN
Sbjct  2066   PAMTGTSTLTVVIGDVNDN  2084

 Score = 39.7 bits (91),  Expect = 0.22, Method: Compositional matrix adjust.
 Identities = 89/389 (23%), Positives = 149/389 (38%), Gaps = 62/389 (16%)

Query  75   LITCRALNAQGLDVEKPLILTVKILDINDNPPVFSQQIF--MGEIEENSASNSLVMILNA  132
            +IT +  NA GL       + + + D+ND PP F  +       ++ N+  N+ V  L A
Sbjct  517  IITNQGHNA-GLKYTDNQRVIIHVKDVNDEPPYFINRPLPMQAVVQLNAPPNTPVFTLQA  575

Query  133  TDADEPNHLNSKIAFKIVSQEPAGTPMFLLSRNTGEVRTLTNSLDREQASSYRLVVSGAD  192
             D D     +  I + IV     G   F +   +G VRT    L  +    Y L V   D
Sbjct  576  RDPD----TDHNIHYFIVRDRTGG--RFEVDERSGVVRTRGTDL-FQLDMEYVLYVKAED  628

Query  193  KDGE----GLSTQCECNIKVKDVNDNFPMFRDSQYSARIEEN-ILSSELLRFQVTDLDEE  247
            ++G+       +  E  + +       P F    Y A I EN    S+++  +     +
Sbjct  629  QNGKVDDRRFQSTPEERLSIVG-GKRAPQFYMPSYEAEIPENQKKDSDIISVKA----KS  683

Query  248  YTDNWLAVYFFTSGNEGNWFEIQTDPRTNEGILKVVKALDYEQLQSVKLS----IAVKNK  303
            + D  +       G     F I        GI+K+ K LD+E L+   +      A ++
Sbjct  684  FADREIRYTLKAQGQGAGTFNIGP----TSGIVKLAKELDFEDLRQPHVYSLVVTATEDS  739

Query  304  AEFHQSVISRYRVQSTPVTIQVINVREGIAFRPASKTFTVQKGISSKKLVDYILGT----  359
              F  SV          +TI+V +V +       +  F +    +     D  LGT
Sbjct  740  GGFSTSV---------ELTIRVTDVNDN------APKFELPDYQAHNVDEDIPLGTSILK  784

Query  360  YQAIDEDTNKAASNVKYVMGRN----DGGYLMIDSKTAEIKFVKNMNRDSTFIVNKTITA  415
             +A+D D+  A + ++Y++  +    D   +++++K  +     N N    F+V
Sbjct  785  VKAMDSDSG-ANAEIEYLVSDDHFAVDSNGIIVNNKQLD---ADNNNAYYEFVVTAKDKG  840
```

```
Query  416  EVLAIDEYTGKTSTGTVYVRVPDFNDNCP  444
            E         KT T TV V   + ND  P
Sbjct  841  E-------PAKTGTATVRVYTKNKNDEEP  862

 Score = 39.7 bits (91),  Expect = 0.27, Method: Compositional matrix adjust.
 Identities = 54/242 (22%), Positives = 102/242 (42%), Gaps = 22/242 (9%)

Query  4    KFAKPCREGE--DNSKRNP-IAKITSDYQATQKITYRISGVGIDQPPFGIFVVDKNTGDI  60
            +F  P  E E  +N K++  I  + +   A ++I Y +   G      G F +   +G +
Sbjct  656  QFYMPSYEAEIPENQKKDSDIISVKAKSFADREIRYTLKAQGQGA---GTFNIGPTSGIV  712

Query  61   NITAIVDREE--TPSF--LITCRALNAQGLDVEKPLILTVKILDINDNPPVFSQQIFMGE  116
             +   +D E+   P    L+     ++ G       + LT+++ D+NDN P F    +
Sbjct  713  KLAKELDFEDLRQPHVYSLVVTATEDSGGFSTS--VELTIRVTDVNDNAPKFELPDYQAH  770

Query  117  -IEENSASNSLVMILNATDADEPNHLNSKIAFKIVSQEPAGTPMFLLSRNTGEVRTLTNS  175
             ++E+    + ++ + A D+D  +  N++I + +VS +     F +  N   V
Sbjct  771  NVDEDIPLGTSILKVKAMDSD--SGANAEIEY-LVSDD-----HFAVDSNGIIVNNKQLD  822

Query  176  LDREQASSYRLVVSGADKDGEGLSTQCECNIKVKDVNDNFPMFRDSQYSARIEENILSSE  235
             D   A  Y  VV+  DK     +      +  K+ ND  P F    Y+  ++EN   +
Sbjct  823  ADNNNA-YYEFVVTAKDKGEPAKTGTATVRVYTKNKNDEEPKFSQQVYTPNVDENAGPNT  881

Query  236  LL  237
            L+
Sbjct  882  LV  883

>ACMO_007424.P13500 | transcript=ACMO_007424.R13500 | gene=ACMO_007424
| organism=Anopheles_coluzzii_MOPTI | gene_product=neural-cadherin,
transcript variant X4 | transcript_product=neural-cadherin,
transcript variant X15 | location=CM029349:40739311-40945968(+)
| protein_length=3098 | sequence_SO=chromosome
| SO=protein_coding_gene | is_pseudo=false
Length=3098

 Score = 147 bits (370),  Expect = 3e-35, Method: Compositional matrix adjust.
 Identities = 128/446 (29%), Positives = 217/446 (49%), Gaps = 36/446 (8%)

Query  20    PIAKITS---DYQATQKITYRISGVGID--QPPFGIFVVDKNTGDINITAIVDREET---  71
             PI ++T+   D    Q I Y ++G GID   P    F +++ TG+I +   +DR++
Sbjct  1539  PIMQVTATDGDKDRPQNIVYFLTGQGIDPDNPANSKFDINRTTGEIFVLKPLDRDQPNGR  1598

Query  72    PSFLITCRALNAQGLDVEKPLILTVKILDINDNPPVFSQQIFMGEIEENSASNSLVMILN  131
             P +  T  A +  G  +     + V + DINDN P+F Q ++ G + EN  +  +VM +
Sbjct  1599  PQWRFTVFAQDEGGEGLVGYADVQVNLKDINDNAPIFPQGVYFGNVTENGTAGMVVMTMT  1658

Query  132   ATDADEPNH-LNSKIAFKI---VSQEPAGTPMFLLSRNTGEVRTLTNSLDREQASSYRLV  187
             A D D+PN   N+K+ + I   V +E  G+P+F +  +TG ++T    LDRE+   Y +
Sbjct  1659  AVDYDDPNEGTNAKLIYSIEKNVIEEETGSPIFEIEADTGVIKTAVCCLDRERTPDYSIQ  1718

Query  188   VSGADKDGEGLSTQCECNIKVKDVNDNFPMFRDSQYSARIEENILSS--ELLRFQVTDLD  245
             V      DG GL      +I+VKD+ND  P F   ++   ++E   ++  E+    VT  D
Sbjct  1719  VVAM--DGGGLKGTGTASIRVKDINDMPPQFTKDEWFTEVDETDGTNLPEMPILTVTVHD  1776

Query  246   EEYTDNWLAVYFFTSGNEGNWFEIQTDPRTNEGI--LKVVKALDYE---QLQSVKLSIAV  300
             E+ T+ +       SG   + F +    R N+G   LK+V+ LDYE   Q    +  I V
Sbjct  1777  EDETNKFQYKVIDNSGYGADKFTMV---RNNDGTGSLKIVQPLDYEDPLQSNGFRFRIQV  1833

Query  301   KNKAEFHQSVISRYRVQSTPVTIQVINVREGIAFRPASKTFTVQKGISSKKLVDYILGTY  360
              +K E + +   +Y V  + V +++ ++ +    +P  +   ++  +     V   L T+
Sbjct  1834  NDKGEDNDN--DKYHVAYSWVVVKLRDINDN---KPQFERPNIEVSVYENADVGKTLETF  1888

Query  361   QAIDEDTNKAASNVKYVMGR-NDGGYLMIDSKTAEIKFVKNMNRDSTFIVNKTITAEVLA  419
             +A D D     S V Y + R +D       ++   +   +N++R+    V      ++LA
Sbjct  1889  KATDPDQG-GKSKVSYAIDRSSDRQRQFSINQEGTVTIQRNLDRE----VTPRHQVKILA  1943

Query  420   IDE-YTGKTSTGTVYVRVPDFNDNCP  444
             ID+    KT+T T+ V V D NDN P
Sbjct  1944  IDDGIPPKTATATLTVIVQDINDNPP  1969
```

```
 Score = 76.3 bits (186),  Expect = 1e-12, Method: Compositional matrix adjust.
 Identities = 89/361 (25%), Positives = 167/361 (46%), Gaps = 36/361 (10%)

Query  51    FVVDKNTGDINITAIVDRE-ETPSFL-ITCRALNAQGLDVEKPLILTVKILDINDNPPVF  108
             F VD+ TG+++   + DRE +   F+ +T +A +     +E     TV+I D+NDNPP+F
Sbjct  1025  FTVDEETGEVSTNKVFDREGDDGKFVSVTVKATDQGEPSLEGVCSFTVEITDVNDNPPLF  1084

Query  109   SQQIFMGEIEENSASNSLVMILNATDADEPNHLNSKIAFKIVS-QEPAGTPMFLLSRNTG  167
              +Q ++  ++++++  + ++ ++A+D D  N  N  I + + +   P     F +   +G
Sbjct  1085  DRQKYVENVKQDASIGTNILRVSASDEDADN--NGAIVYTLSAPHSPNDLEYFEIQPESG  1142

Query  168   EVRTLTNSLDREQASSYRLVVSGADKDGEGLSTQCECNIKVKDVNDNFPMFRDSQYSA-R  226
              +  L   LDRE   +Y+L     DK    LS   E  I V D  +N P++ ++ Y
Sbjct  1143  WI-VLKKPLDRE---TYKLEAMAQDKGYPPLSRTVEVQIDVVDRANNPPVWDNTVYGPIY  1198

Query  227   IEENI-LSSELLRFQVTDLDEEYTDNWLAVYFFTSG-----NEGNWFEIQTDPRTNE--G  278
             ++EN+ +  +++  + +   E    N    Y    G     N+ + F +Q  P   +
Sbjct  1199  VKENMPVGGKVVSIKASSGIE---GNPTVFYRLMPGSTAQTNKLHTFYLQQRPDNGDTWA  1255

Query  279   ILKVVKALDYEQLQSVKLSIAVKNKAEFHQSVISRYRVQSTPVTIQVINVREGIAFRPAS  338
              +KV   LD+E ++   L++ V+N         ++       V IQ+ +V + I
Sbjct  1256  DIKVNHPLDFETIKEYNLTVRVENNG-------AQQLASEATVYIQLEDVNDEIPLFTER  1308

Query  339   KTFTVQKGISSKKLVDYILGTYQAIDEDTNKAASNVKYVM---GRNDG-GYLMIDSKTAE  394
             +  TV +G      V  +     AID+D     + V Y +    RN+G  +  I+ ++ E
Sbjct  1309  EQETVLEGEPIGTKVTQV----NAIDKDGTFPNNQVYYYIVDSPRNEGKDFFEINLQSGE  1364

Query  395   I  395
             I
Sbjct  1365  I  1365

 Score = 75.9 bits (185),  Expect = 2e-12, Method: Compositional matrix adjust.
 Identities = 103/418 (25%), Positives = 174/418 (42%), Gaps = 50/418 (12%)

Query  50    IFVVDKNTGDINITAIVDREETPSFLITCRAL----------NAQGLDVEKPLILTVKIL  99
              F ++  +G+I    + DRE+  ++ +   A           N Q   V K   + + I
Sbjct  1355  FFEINLQSGEIFTKVVFDREKQGAYALEVEARDGAPSARPNSNQQPNSVTK--FIRIGIA  1412

Query  100   DINDNPPVFSQQIFMGEIEENSASNSLVMILNATDADEPNHLNSKIAFKIVSQEPAGTPM  159
             D NDNPP F + ++  E++EN      V+ + A D DE    +S+I ++I S    G
Sbjct  1413  DKNDNPPYFDKALYEAEVDENEDIQHTVLTVTAKDHDE----SSRIRYEITSGNIGGA--  1466

Query  160   FLLSRNTGEVRTLTNSLDREQASSYRLVVSGADKDGEGLSTQCECNIKVKDVNDNFPMFR  219
             F +   TG +  +  +LD E    Y L +  +D   E  +      I VKDVND  P F
Sbjct  1467  FAVKNMTGAI-YVAGALDYETRKRYYLTLVASDSLNENETIIV---INVKDVNDMPPSFP  1522

Query  220   DSQYSARIEENI-LSSELLRFQVTDLDEEYTDNWLAVYFFTSG--NEGNWFEIQTDPRTN  276
                Y   ++E + +   +++   TD D++   N   VYF T    +  N    + D
Sbjct  1523  QQVYERTMDEELAVPFPIMQVTATDGDKDRPQN--IVYFLTGQGIDPDNPANSKFDINRT  1580

Query  277   EGILKVVKALDYEQLQS-VKLSIAVKNKAEFHQSVISRYRVQSTPVTIQVINVREGIAFR  335
              G + V+K LD +Q     +    V  + E  + ++    VQ     + + ++ +
Sbjct  1581  TGEIFVLKPLDRDQPNGRPQWRFTVFAQDEGGEGLVGYADVQ-----VNLKDINDNAPIF  1635

Query  336   PASKTFTVQKGISSKKLVDYILGTYQAIDEDTNKAASNVK--YVMGRN------DGGYLM  387
             P    F     ++       ++ T  A+D D     +N K  Y + +N
Sbjct  1636  PQGVYF---GNVTENGTAGMVVMTMTAVDYDDPNEGTNAKLIYSIEKNVIEEETGSPIFE  1692

Query  388   IDSKTAEIK-FVKNMNRDSTFIVNKTITAEVLAIDEYTGKTSTGTVYVRVPDFNDNCP  444
             I++ T  IK  V  ++R+ T       + +V+A+D   G   TGT  +RV D ND  P
Sbjct  1693  IEADTGVIKTAVCCLDRERT----PDYSIQVVAMDG-GGLKGTGTASIRVKDINDMPP  1745

 Score = 55.8 bits (133),  Expect = 3e-06, Method: Compositional matrix adjust.
 Identities = 49/158 (31%), Positives = 78/158 (49%), Gaps = 24/158 (15%)

Query  102   NDNPPVFSQQIFMGEIEENSASNSLVMILNATDADEPNHLNSKIAFKIVSQEPAGTP--M  159
             ND  P FSQQ++   ++EN+  N+LV  + A+D D  N     + F  V     GT
```

```
Sbjct  858   NDEEPKFSQQVYTPNVDENAGPNTLVTTVVASDKDGDN-----VRFGFVG---GGTSSGQ  909

Query  160   FLLSRNTGEVRTLTN--SLDREQASSYRLVVSGADK-------DGEGLSTQCECNIKVKD  210
             F++   TG +R  +   SLDR++   Y L V+  D        D    ++      + + D
Sbjct  910   FVIEEITGVIRLHSKAISLDRDK---YELNVTAMDDGSCCVNGDATIHTSTAVVVVFITD  966

Query  211   VNDNFPMFRD-SQYSARIEENILS-SELLRFQVTDLDE  246
             VNDN P+F+D   Y  ++EE   + S +++ Q TD D+
Sbjct  967   VNDNKPVFKDCGTYYPKVEEGAPNGSPVIKVQATDEDK  1004

 Score = 53.9 bits (128),  Expect = 1e-05, Method: Compositional matrix adjust.
 Identities = 70/306 (23%), Positives = 126/306 (41%), Gaps = 30/306 (10%)

Query  20    PIAKIT-SDYQATQKITYRI---SGVGIDQPPFGIFVVDKNTGDINITAIVDRE---ETP  72
             PI  +T  D   T K  Y++   SG G D+  F +   +  TG + I   +D E   ++
Sbjct  1768  PILTVTVHDEDETNKFQYKVIDNSGYGADK--FTMVRNNDGTGSLKIVQPLDYEDPLQSN  1825

Query  73    SFLITCRALNAQGLDVEKPLI------LTVKILDINDNPPVFSQQIFMGEIEENSASNSL  126
               F    + +N +G D +           + VK+ DINDN P F +       + EN+
Sbjct  1826  GFRFRIQ-VNDKGEDNDNDKYHVAYSWVVVKLRDINDNKPQFERPNIEVSVYENADVGKT  1884

Query  127   VMILNATDADEPNHLNSKIAFKIVSQEPAGTPMFLLSRNTGEVRTLTNSLDREQASSYRL  186
             +    ATD D+      SK+++ I   + +        S N     T+  +LDRE    +++
Sbjct  1885  LETFKATDPDQGG--KSKVSYAI---DRSSDRQRQFSINQEGTVTIQRNLDREVTPRHQV  1939

Query  187   VVSGADKDGEGLSTQCECNIKVKDVNDNFPMFRDSQYSARIEENILSSELLRFQVTDLDE  246
               +   D       +       + V+D+NDN P F    Y   + E++   +++    TD D+
Sbjct  1940  KILAIDDGIPPKTATATLTVIVQDINDNPPKFL-KDYRPVLPEHVPPRKVVEILATDDDD  1998

Query  247   EYTDNWLAVYFFTSGNEGNW----FEIQTDPRTNEG----ILKVVKALDYEQLQSVKLSI  298
                 N     F       +     F+++ D +   G    I+  +++ D EQ +   + I
Sbjct  1999  RSKSNGPPFQFRLDPGADDIIRASFKVEQDQKGANGDGMAIVSSLRSFDREQQKEYLIPI  2058

Query  299   AVKNKA  304
              +K+
Sbjct  2059  VIKDHG  2064

 Score = 52.0 bits (123),  Expect = 4e-05, Method: Compositional matrix adjust.
 Identities = 97/398 (24%), Positives = 163/398 (41%), Gaps = 54/398 (14%)

Query  106   PVFSQQIFMGEIEENSASNSLVMILNATDADEPNHLNSKIAFKIVSQ-EPAGTPMFLLSR  164
             P F    +  EI EN   +S ++ + A       +  + +I + + +Q + AGT  F +
Sbjct  655   PQFYMPSYEAEIPENQKKDSDIISVKAK-----SFADREIRYTLKAQGQGAGT--FNIGP  707

Query  165   NTGEVRTLTNSLDRE---QASSYRLVVSGADKDGEGLSTQCECNIKVKDVNDNFPMFRDS  221
              +G V+ L   LD E   Q   Y LVV+ A +D  G ST  E  I+V DVNDN P F
Sbjct  708   TSGIVK-LAKELDFEDLRQPHVYSLVVT-ATEDSGGFSTSVELTIRVTDVNDNAPKFELP  765

Query  222   QYSAR-IEENI-LSSELLRFQVTDLDEEYTDNWLAVYFFTSGNEGNWFEIQTDPRTNEGI  279
              Y A  ++E+I L + +L+ +  D D          + +  S +    F + ++     GI
Sbjct  766   DYQAHNVDEDIPLGTSILKVKAMDSDSGAN---AEIEYLVSDDH---FAVDSN-----GI  814

Query  280   LKVVKALDYEQLQS-VKLSIAVKNKAEFHQSVISRYRVQSTPVTIQVINVREGIAFRPAS  338
             +   K LD +   +  +  +  K+K E  ++  +  RV +             E   F
Sbjct  815   IVNNKQLDADNNNAYYEFVVTAKDKGEPAKTGTATVRVYTKNKN------DEEPKFSQQV  868

Query  339   KTFTVQKGISSKKLVDYILGTYQAIDEDTNKAASNVK--YVMGRNDGGYLMIDSKTAEIK  396
              T  V +       LV  ++          ++K   NV+  +V G    G  +I+  T  I+
Sbjct  869   YTPNVDENAGPNTLVTTVVA--------SDKDGDNVRFGFVGGGTSSGQFVIEEITGVIR  920

Query  397   FVK---NMNRDSTFI-VNKTITAEVLAIDEYTGKTSTGTVYVRVPDFNDN------CPTA  446
                   +++RD   + V             + T  TST  V V + D NDN      C T
Sbjct  921   LHSKAISLDRDKYELNVTAMDDGSCCVNGDATIHTSTAVVVVFITDVNDNKPVFKDCGTY  980

Query  447   VLE-KDAVCSSSPSVVVSARTLNNRYTGPYTFALEDQP  483
               + ++    + SP + V A     +    G   +++  QP
Sbjct  981   YPKVEEGAPNGSPVIKVQATDEDKGVNGQVKYSIVQQP  1018
```

```
 Score = 44.7 bits (104),  Expect = 0.008, Method: Compositional matrix adjust.
 Identities = 48/199 (24%), Positives = 73/199 (37%), Gaps = 19/199 (10%)

Query  27    DYQATQKITYRISGVGIDQPPFGIFVVDKNTGDINITAIVDREETPSFLITCRALNAQGL  86
             D     K++Y I      Q  F I       G + I   +DRE TP   +   A++
Sbjct  1894  DQGGKSKVSYAIDRSSDRQRQFSI----NQEGTVTIQRNLDREVTPRHQVKILAIDDGIP  1949

Query  87    DVEKPLILTVKILDINDNPPVFSQQIFMGEIEENSASNSLVMILNATDADEPNHLNSKIA  146
                    LTV + DINDNPP F +  +   + E+     +V IL   D D
Sbjct  1950  PKTATATLTVIVQDINDNPPKFLKD-YRPVLPEHVPPRKVVEILATDDDDRSKSNGPPFQ  2008

Query  147   FKIVSQEPAGTPMFLLSRNTGEVRTLTN-----------SLDREQASSYRLVVSGADKDG  195
             F++   +P    +   S    + +   N           S DREQ   Y + +   D
Sbjct  2009  FRL---DPGADDIIRASFKVEQDQKGANGDGMAIVSSLRSFDREQQKEYLIPIVIKDHGN  2065

Query  196   EGLSTQCECNIKVKDVNDN  214
               ++      + + DVNDN
Sbjct  2066  PAMTGTSTLTVVIGDVNDN  2084

 Score = 39.7 bits (91),  Expect = 0.23, Method: Compositional matrix adjust.
 Identities = 89/389 (23%), Positives = 149/389 (38%), Gaps = 62/389 (16%)

Query  75   LITCRALNAQGLDVEKPLILTVKILDINDNPPVFSQQIF--MGEIEENSASNSLVMILNA  132
            +IT +  NA GL       + + + D+ND PP F  +       ++ N+  N+ V  L A
Sbjct  517  IITNQGHNA-GLKYTDNQRVIIHVKDVNDEPPYFINRPLPMQAVVQLNAPPNTPVFTLQA  575

Query  133  TDADEPNHLNSKIAFKIVSQEPAGTPMFLLSRNTGEVRTLTNSLDREQASSYRLVVSGAD  192
             D D     +  I + IV     G   F +   +G VRT    L  +    Y L V   D
Sbjct  576  RDPD----TDHNIHYFIVRDRTGG--RFEVDERSGVVRTRGTDL-FQLDMEYVLYVKAED  628

Query  193  KDGE----GLSTQCECNIKVKDVNDNFPMFRDSQYSARIEEN-ILSSELLRFQVTDLDEE  247
            ++G+       +  E  +  + +       P F    Y A I EN    S+++  +     +
Sbjct  629  QNGKVDDRRFQSTPEERLSIVG-GKRAPQFYMPSYEAEIPENQKKDSDIISVKA----KS  683

Query  248  YTDNWLAVYFFTSGNEGNWFEIQTDPRTNEGILKVVKALDYEQLQSVKLS----IAVKNK  303
            + D  +       G     F I        GI+K+ K LD+E L+   +      A ++
Sbjct  684  FADREIRYTLKAQGQGAGTFNIGP----TSGIVKLAKELDFEDLRQPHVYSLVVTATEDS  739

Query  304  AEFHQSVISRYRVQSTPVTIQVINVREGIAFRPASKTFTVQKGISSKKLVDYILGT----  359
              F  SV          +TI+V +V +       +  F +    +     D  LGT
Sbjct  740  GGFSTSV---------ELTIRVTDVNDN------APKFELPDYQAHNVDEDIPLGTSILK  784

Query  360  YQAIDEDTNKAASNVKYVMGRN----DGGYLMIDSKTAEIKFVKNMNRDSTFIVNKTITA  415
             +A+D D+  A + ++Y++  +    D   +++++K  +     N N    F+V
Sbjct  785  VKAMDSDSG-ANAEIEYLVSDDHFAVDSNGIIVNNKQLD---ADNNNAYYEFVVTAKDKG  840

Query  416  EVLAIDEYTGKTSTGTVYVRVPDFNDNCP  444
            E         KT T TV V   + ND  P
Sbjct  841  E-------PAKTGTATVRVYTKNKNDEEP  862

 Score = 39.7 bits (91),  Expect = 0.28, Method: Compositional matrix adjust.
 Identities = 54/242 (22%), Positives = 102/242 (42%), Gaps = 22/242 (9%)

Query  4    KFAKPCREGE--DNSKRNP-IAKITSDYQATQKITYRISGVGIDQPPFGIFVVDKNTGDI  60
            +F  P  E E  +N K++  I  + +   A ++I Y +   G      G F +   +G +
Sbjct  656  QFYMPSYEAEIPENQKKDSDIISVKAKSFADREIRYTLKAQGQGA---GTFNIGPTSGIV  712

Query  61   NITAIVDREE--TPSF--LITCRALNAQGLDVEKPLILTVKILDINDNPPVFSQQIFMGE  116
             +   +D E+   P    L+     ++ G      + LT+++ D+NDN P F    +
Sbjct  713  KLAKELDFEDLRQPHVYSLVVTATEDSGGFSTS--VELTIRVTDVNDNAPKFELPDYQAH  770

Query  117  -IEENSASNSLVMILNATDADEPNHLNSKIAFKIVSQEPAGTPMFLLSRNTGEVRTLTNS  175
             ++E+    + ++ + A D+D  +  N++I + +VS +      F + N   V
Sbjct  771  NVDEDIPLGTSILKVKAMDSD--SGANAEIEY-LVSDD-----HFAVDSNGIIVNNKQLD  822

Query  176  LDREQASSYRLVVSGADKDGEGLSTQCECNIKVKDVNDNFPMFRDSQYSARIEENILSSE  235
             D   A  Y  VV+  DK     +      +  K+ ND  P F    Y+  ++EN   +
Sbjct  823  ADNNNA-YYEFVVTAKDKGEPAKTGTATVRVYTKNKNDEEPKFSQQVYTPNVDENAGPNT  881
```

```
Query  236  LL  237
            L+
Sbjct  882  LV  883

>AGAMI1_007196.P14116 | transcript=AGAMI1_007196.R14116 | gene=AGAMI1_007196
| organism=Anopheles_gambiae_Ifakara | gene_product=neural-cadherin
| transcript_product=neural-cadherin,
transcript variant X15 | location=AgamI1_3RL:41252608-41455387(+)
| protein_length=3099 | sequence_SO=chromosome | SO=protein_coding_gene
| is_pseudo=false
Length=3099

 Score = 147 bits (370),  Expect = 3e-35, Method: Compositional matrix adjust.
 Identities = 128/446 (29%), Positives = 217/446 (49%), Gaps = 36/446 (8%)

Query  20    PIAKITS---DYQATQKITYRISGVGID--QPPFGIFVVDKNTGDINITAIVDREET---  71
             PI ++T+   D    Q I Y ++G GID   P    F +++ TG+I +   +DR++
Sbjct  1539  PIMQVTATDGDKDRPQNIVYFLTGQGIDPDNPANSKFDINRTTGEIFVLKPLDRDQPNGR  1598

Query  72    PSFLITCRALNAQGLDVEKPLILTVKILDINDNPPVFSQQIFMGEIEENSASNSLVMILN  131
             P +  T  A +  G  +     + V + DINDN P+F Q ++ G + EN  +  +VM +
Sbjct  1599  PQWRFTVFAQDEGGEGLVGYADVQVNLKDINDNAPIFPQGVYFGNVTENGTAGMVVMTMT  1658

Query  132   ATDADEPNH-LNSKIAFKI---VSQEPAGTPMFLLSRNTGEVRTLTNSLDREQASSYRLV  187
             A D D+PN   N+K+ + I   V +E  G+P+F +  +TG ++T    LDRE+   Y +
Sbjct  1659  AVDYDDPNEGTNAKLIYSIEKNVIEEETGSPIFEIEADTGVIKTAVCCLDRERTPDYSIQ  1718

Query  188   VSGADKDGEGLSTQCECNIKVKDVNDNFPMFRDSQYSARIEENILSS--ELLRFQVTDLD  245
             V     DG GL       +I+VKD+ND  P F   ++   ++E   ++  E+    VT  D
Sbjct  1719  VVAM--DGGGLKGTGTASIRVKDINDMPPQFTKDEWFTEVDETDGTNLPEMPILTVTVHD  1776

Query  246   EEYTDNWLAVYFFTSGNEGNWFEIQTDPRTNEGI--LKVVKALDYE---QLQSVKLSIAV  300
             E+ T+ +       SG   + F +    R N+G   LK+V+ LDYE   Q    +  I V
Sbjct  1777  EDETNKFQYKVIDNSGYGADKFTMV---RNNDGTGSLKIVQPLDYEDPLQSNGFRFRIQV  1833

Query  301   KNKAEFHQSVISRYRVQSTPVTIQVINVREGIAFRPASKTFTVQKGISSKKLVDYILGTY  360
              +K E + +   +Y V  + V +++ ++ +    +P  +   ++  +     V   L T+
Sbjct  1834  NDKGEDNDN--DKYHVAYSWVVVKLRDINDN---KPQFERPNIEVSVYENADVGKTLETF  1888

Query  361   QAIDEDTNKAASNVKYVMGR-NDGGYLMIDSKTAEIKFVKNMNRDSTFIVNKTITAEVLA  419
             +A D D     S V Y + R +D       ++   +   +N++R+    V      ++LA
Sbjct  1889  KATDPDQG-GKSKVSYAIDRSSDRQRQFSINQEGTVTIQRNLDRE----VTPRHQVKILA  1943

Query  420   IDE-YTGKTSTGTVYVRVPDFNDNCP  444
             ID+    KT+T T+ V V D NDN P
Sbjct  1944  IDDGIPPKTATATLTVIVQDINDNPP  1969

 Score = 78.6 bits (192),  Expect = 2e-13, Method: Compositional matrix adjust.
 Identities = 106/424 (25%), Positives = 180/424 (42%), Gaps = 51/424 (12%)

Query  53    VDKNTGDINITAIVDREETPSFLITCRALNAQGLDVEKPLILTVKILDINDNPPVFSQ-Q  111
             +D++  ++N+TA+ D         +C  +N          ++ V I D+NDN PVF
Sbjct  928   LDRDKYELNVTAMDDG--------SC-CVNGDATIHTSTAVVVVFITDVNDNKPVFKDCG  978

Query  112   IFMGEIEENSASNSLVMILNATDADEPNHLNSKIAFKIVSQEPAGTPMFLLSRNTGEVRT  171
              +  ++EE + + S V+ + ATD D+   +N ++ + IV Q       F +   TGEV T
Sbjct  979   TYYPKVEEGAPNGSPVIKVQATDEDK--GVNGQVKYSIVQQPNQKGTKFTVDEETGEVST  1036

Query  172   LTNSLDRE--QASSYRLVVSGADKDGEGLSTQCECNIKVKDVNDNFPMFRDSQYSARIEE  229
                  DRE        + V   D+    L   C   +++ DVNDN P+F   +Y   +++
Sbjct  1037  -NKVFDREGDDGKFVSVTVKATDQGEPSLEGVCSFTVEITDVNDNPPLFDRQKYVENVKQ  1095

Query  230   NI-LSSELLRFQVTDLDEEYTDNWLAVYFFT---SGNEGNWFEIQTDPRTNEGILKVVKA  285
             +  + + +LR  V+  DE+  +N   VY  +   S N+  +FEIQ  P +   +LK  K
Sbjct  1096  DASIGTNILR--VSASDEDADNNGAIVYTLSAPHSPNDLEYFEIQ--PESGWIVLK--KP  1149

Query  286   LDYEQLQSVKLSIAVKNK------AEFHQSVISRYRVQSTPVTIQV----INVREGIAFR  335
             LD E+    +L +   ++      A+    +    R    P+  +     I++RE +
Sbjct  1150  LDRERY---RLKVYASDRGDPPSTADVDVELDVVDRNNKPPIWDRTIYGPIHIRENVTVG  1206
```

```
Query  336   PASKTFTVQKGISSKKLVDYILGTYQAIDEDTNKAASNVKYVMGRNDGGYLMIDSKTAEI  395
                 +     GI     V Y L         TNK   +  Y+  R D G        A+I
Sbjct  1207  TVVTSVKASSGIEGNPTVFYRL--MPGSTAQTNKL--HTFYLQQRPDNG-----DTWADI  1257

Query  396   KFVKNMNRDSTFIVNKTITAEVLAIDEYTGKTSTGTVYYRVPDFNDNCPTAV-LEKDAVC  454
             K    ++ ++    N T+  E     +      S  TVY+++ D ND  P     E++ V
Sbjct  1258  KVNHPLDFETIKEYNLTVRVENNGAQQL---ASEATVYIQLEDVNDEIPLFTEREQETVL  1314

Query  455   SSSP  458
                P
Sbjct  1315  EGEP  1318

 Score = 75.9 bits (185),  Expect = 2e-12, Method: Compositional matrix adjust.
 Identities = 103/418 (25%), Positives = 174/418 (42%), Gaps = 50/418 (12%)

Query  50    IFVVDKNTGDINITAIVDREETPSFLITCRAL----------NAQGLDVEKPLILTVKIL  99
              F ++  +G+I    + DRE+  ++ +   A           N Q   V K   + + I
Sbjct  1355  FFEINLQSGEIFTKVVFDREKQGAYALEVEARDGAPSARPNSNQQPNSVTK--FIRIGIA  1412

Query  100   DINDNPPVFSQQIFMGEIEENSASNSLVMILNATDADEPNHLNSKIAFKIVSQEPAGTPM  159
             D NDNPP F + ++  E++EN      V+ + A D DE    +S+I ++I S    G
Sbjct  1413  DKNDNPPYFDKALYEAEVDENEDIQHTVLTVTAKDHDE----SSRIRYEITSGNIGGA--  1466

Query  160   FLLSRNTGEVRTLTNSLDREQASSYRLVVSGADKDGEGLSTQCECNIKVKDVNDNFPMFR  219
             F +   TG +  +  +LD E    Y L +  +D   E  +      I VKDVND  P F
Sbjct  1467  FAVKNMTGAI-YVAGALDYETRKRYYLTLVASDSLNENETIIV---INVKDVNDMPPSFP  1522

Query  220   DSQYSARIEENI-LSSELLRFQVTDLDEEYTDNWLAVYFFTSG--NEGNWFEIQTDPRTN  276
                Y   ++E + +   +++   TD D++   N   VYF T    +  N    + D
Sbjct  1523  QQVYERTMDEELAVPFPIMQVTATDGDKDRPQN--IVYFLTGQGIDPDNPANSKFDINRT  1580

Query  277   EGILKVVKALDYEQLQS-VKLSIAVKNKAEFHQSVISRYRVQSTPVTIQVINVREGIAFR  335
              G + V+K LD +Q     +    V  + E  + ++    VQ     + + ++ +
Sbjct  1581  TGEIFVLKPLDRDQPNGRPQWRFTVFAQDEGGEGLVGYADVQ-----VNLKDINDNAPIF  1635

Query  336   PASKTFTVQKGISSKKLVDYILGTYQAIDEDTNKAASNVK--YVMGRN------DGGYLM  387
             P    F     ++       ++ T  A+D D     +N K  Y + +N
Sbjct  1636  PQGVYF---GNVTENGTAGMVVMTMTAVDYDDPNEGTNAKLIYSIEKNVIEEETGSPIFE  1692

Query  388   IDSKTAEIK-FVKNMNRDSTFIVNKTITAEVLAIDEYTGKTSTGTVYYRVPDFNDNCP  444
             I++ T  IK  V  ++R+ T       + +V+A+D   G   TGT  +RV D ND  P
Sbjct  1693  IEADTGVIKTAVCCLDRERT----PDYSIQVVAMDG-GGLKGTGTASIRVKDINDMPP  1745

 Score = 65.5 bits (158),  Expect = 3e-09, Method: Compositional matrix adjust.
 Identities = 85/359 (24%), Positives = 153/359 (43%), Gaps = 32/359 (9%)

Query  51    FVVDKNTGDINITAIVDRE-ETPSFL-ITCRALNAQGLDVEKPLILTVKILDINDNPPVF  108
             F VD+ TG+++   + DRE +   F+ +T +A +     +E     TV+I D+NDNPP+F
Sbjct  1025  FTVDEETGEVSTNKVFDREGDDGKFVSVTVKATDQGEPSLEGVCSFTVEITDVNDNPPLF  1084

Query  109   SQQIFMGEIEENSASNSLVMILNATDADEPNHLNSKIAFKIVS-QEPAGTPMFLLSRNTG  167
              +Q ++  ++++++  + ++ ++A+D D  N  N  I + + +   P     F +   +G
Sbjct  1085  DRQKYVENVKQDASIGTNILRVSASDEDADN--NGAIVYTLSAPHSPNDLEYFEIQPESG  1142

Query  168   EVRTLTNSLDREQASSYRLVVSGADKDGEGLSTQCECNIKVKDVNDNFPMFRDSQYSARI  227
              +  L   LDRE+   YRL V  +D+ G+  ST            +N P   D      I
Sbjct  1143  WI-VLKKPLDRER---YRLKVYASDR-GDPPSTADVDVELDVVDRNNKPPIWDRTIYGPI  1197

Query  228   EENILSSELLRFQVTDLDEEYTDNWLAVYFFTSG-----NEGNWFEIQTDPRTNE--GIL  280
                   +                N    Y    G     N+ + F +Q  P   +    +
Sbjct  1198  HIRENVTVGTVVTSVKASSGIEGNPTVFYRLMPGSTAQTNKLHTFYLQQRPDNGDTWADI  1257

Query  281   KVVKALDYEQLQSVKLSIAVKNKAEFHQSVISRYRVQSTPVTIQVINVREGIAFRPASKT  340
             KV   LD+E ++   L++ V+N         ++       V IQ+ +V + I      +
Sbjct  1258  KVNHPLDFETIKEYNLTVRVENNG-------AQQLASEATVYIQLEDVNDEIPLFTEREQ  1310

Query  341   FTVQKGISSKKLVDYILGTYQAIDEDTNKAASNVKYVM---GRNDG-GYLMIDSKTAEI  395
              TV +G      V  +     AID+D     + V Y +    RN+G  +  I+ ++ EI
```

```
Sbjct  1311   ETVLEGEPIGTKVTQV----NAIDKDGTFPNNQVYYYIVDSPRNEGKDFFEINLQSGEI  1365

 Score = 55.8 bits (133),  Expect = 3e-06, Method: Compositional matrix adjust.
 Identities = 49/158 (31%), Positives = 78/158 (49%), Gaps = 24/158 (15%)

Query  102    NDNPPVFSQQIFMGEIEENSASNSLVMILNATDADEPNHLNSKIAFKIVSQEPAGTP--M  159
              ND  P FSQQ++   ++EN+  N+LV  + A+D D  N     + F  V     GT
Sbjct  858    NDEEPKFSQQVYTPNVDENAGPNTLVTTVVASDKDGDN-----VRFGFVG---GGTSSGQ  909

Query  160    FLLSRNTGEVRTLTN--SLDREQASSYRLVVSGADK-------DGEGLSTQCECNIKVKD  210
              F++   TG +R  +   SLDR++   Y L V+  D        D    ++     + + D
Sbjct  910    FVIEEITGVIRLHSKAISLDRDK---YELNVTAMDDGSCCVNGDATIHTSTAVVVVFITD  966

Query  211    VNDNFPMFRD-SQYSARIEENILS-SELLRFQVTDLDE  246
              VNDN P+F+D   Y  ++EE   + S +++ Q TD D+
Sbjct  967    VNDNKPVFKDCGTYYPKVEEGAPNGSPVIKVQATDEDK  1004

 Score = 53.9 bits (128),  Expect = 1e-05, Method: Compositional matrix adjust.
 Identities = 70/306 (23%), Positives = 126/306 (41%), Gaps = 30/306 (10%)

Query  20     PIAKIT-SDYQATQKITYRI---SGVGIDQPPFGIFVVDKNTGDINITAIVDRE---ETP  72
              PI  +T  D   T K  Y++   SG G D+  F +   +  TG + I   +D E   ++
Sbjct  1768   PILTVTVHDEDETNKFQYKVIDNSGYGADK--FTMVRNNDGTGSLKIVQPLDYEDPLQSN  1825

Query  73     SFLITCRALNAQGLDVEKPLI------LTVKILDINDNPPVFSQQIFMGEIEENSASNSL  126
               F    + +N +G D +          + VK+ DINDN P F +      + EN+
Sbjct  1826   GFRFRIQ-VNDKGEDNDNDKYHVAYSWVVVKLRDINDNKPQFERPNIEVSVYENADVGKT  1884

Query  127    VMILNATDADEPNHLNSKIAFKIVSQEPAGTPMFLLSRNTGEVRTLTNSLDREQASSYRL  186
              +    ATD D+     SK+++ I   + +       S N     T+  +LDRE    +++
Sbjct  1885   LETFKATDPDQGG--KSKVSYAI---DRSSDRQRQFSINQEGTVTIQRNLDREVTPRHQV  1939

Query  187    VVSGADKDGEGLSTQCECNIKVKDVNDNFPMFRDSQYSARIEENILSSELLRFQVTDLDE  246
               +   D      +      + V+D+NDN P F    Y   + E++   +++    TD D+
Sbjct  1940   KILAIDDGIPPKTATATLTVIVQDINDNPPKFL-KDYRPVLPEHVPPRKVVEILATDDDD  1998

Query  247    EYTDNWLAVYFFTSGNEGNW----FEIQTDPRTNEG----ILKVVKALDYEQLQSVKLSI  298
                  N     F       +     F+++ D +   G    I+  +++ D EQ +   + I
Sbjct  1999   RSKSNGPPFQFRLDPGADDIIRASFKVEQDQKGANGDGMAIVSSLRSFDREQQKEYLIPI  2058

Query  299    AVKNKA  304
               +K+
Sbjct  2059   VIKDHG  2064

 Score = 52.0 bits (123),  Expect = 4e-05, Method: Compositional matrix adjust.
 Identities = 97/398 (24%), Positives = 163/398 (41%), Gaps = 54/398 (14%)

Query  106    PVFSQQIFMGEIEENSASNSLVMILNATDADEPNHLNSKIAFKIVSQ-EPAGTPMFLLSR  164
              P F    +  EI EN   +S ++ + A      +  + +I + + +Q + AGT  F +
Sbjct  655    PQFYMPSYEAEIPENQKKDSDIISVKAK-----SFADREIRYTLKAQGQGAGT--FNIGP  707

Query  165    NTGEVRTLTNSLDRE---QASSYRLVVSGADKDGEGLSTQCECNIKVKDVNDNFPMFRDS  221
               +G V+ L   LD E   Q   Y LVV+ A +D  G ST  E  I+V DVNDN P F
Sbjct  708    TSGIVK-LAKELDFEDLRQPHVYSLVVT-ATEDSGGFSTSVELTIRVTDVNDNAPKFELP  765

Query  222    QYSAR-IEENI-LSSELLRFQVTDLDEEYTDNWLAVYFFTSGNEGNWFEIQTDPRTNEGI  279
               Y A  ++E+I L + +L+ +  D D         + +  S +    F + ++     GI
Sbjct  766    DYQAHNVDEDIPLGTSILKVKAMDSDSGAN---AEIEYLVSDDH---FAVDSN-----GI  814

Query  280    LKVVKALDYEQLQS-VKLSIAVKNKAEFHQSVISRYRVQSTPVTIQVINVREGIAFRPAS  338
              +   K LD +   +  +  +  K+K E  ++  +  RV +           E   F
Sbjct  815    IVNNKQLDADNNNAYYEFVVTAKDKGEPAKTGTATVRVYTKNKN------DEEPKFSQQV  868

Query  339    KTFTVQKGISSKKLVDYILGTYQAIDEDTNKAASNVK--YVMGRNDGGYLMIDSKTAEIK  396
               T  V +      LV  ++         ++K   NV+  +V G    G  +I+  T  I+
Sbjct  869    YTPNVDENAGPNTLVTTVVA--------SDKDGDNVRFGFVGGGTSSGQFVIEEITGVIR  920

Query  397    FVK---NMNRDSTFI-VNKTITAEVLAIDEYTGKTSTGTVYVRVPDFNDN------CPTA  446
```

```
                   +++RD   + V            + T  TST  V V + D NDN      C T
Sbjct  921   LHSKAISLDRDKYELNVTAMDDGSCCVNGDATIHTSTAVVVVFITDVNDNKPVFKDCGTY  980

Query  447   VLE-KDAVCSSSPSVVVSARTLNNRYTGPYTFALEDQP  483
               + ++   + SP + V A   +    G   +++  QP
Sbjct  981   YPKVEEGAPNGSPVIKVQATDEDKGVNGQVKYSIVQQP  1018

 Score = 44.7 bits (104),  Expect = 0.008, Method: Compositional matrix adjust.
 Identities = 48/199 (24%), Positives = 73/199 (37%), Gaps = 19/199 (10%)

Query  27    DYQATQKITYRISGVGIDQPPFGIFVVDKNTGDINITAIVDREETPSFLITCRALNAQGL  86
             D     K++Y I      Q  F I       G + I   +DRE TP   +   A++
Sbjct  1894  DQGGKSKVSYAIDRSSDRQRQFSI----NQEGTVTIQRNLDREVTPRHQVKILAIDDGIP  1949

Query  87    DVEKPLILTVKILDINDNPPVFSQQIFMGEIEENSASNSLVMILNATDADEPNHLNSKIA  146
                    LTV + DINDNPP F +  +   + E+     +V IL   D D
Sbjct  1950  PKTATATLTVIVQDINDNPPKFLKD-YRPVLPEHVPPRKVVEILATDDDDRSKSNGPPFQ  2008

Query  147   FKIVSQEPAGTPMFLLSRNTGEVRTLTN-----------SLDREQASSYRLVVSGADKDG  195
             F++   +P    +   S    + +   N           S DREQ   Y + +   D
Sbjct  2009  FRL---DPGADDIIRASFKVEQDQKGANGDGMAIVSSLRSFDREQQKEYLIPIVIKDHGN  2065

Query  196   EGLSTQCECNIKVKDVNDN  214
               ++      + + DVNDN
Sbjct  2066  PAMTGTSTLTVVIGDVNDN  2084

 Score = 39.7 bits (91),  Expect = 0.28, Method: Compositional matrix adjust.
 Identities = 89/389 (23%), Positives = 149/389 (38%), Gaps = 62/389 (16%)

Query  75   LITCRALNAQGLDVEKPLILTVKILDINDNPPVFSQQIF--MGEIEENSASNSLVMILNA  132
            +IT +  NA GL       + + + D+ND PP F  +       ++ N+  N+ V  L A
Sbjct  517  IITNQGHNA-GLKYTDNQRVIIHVKDVNDEPPYFINRPLPMQAVVQLNAPPNTPVFTLQA  575

Query  133  TDADEPNHLNSKIAFKIVSQEPAGTPMFLLSRNTGEVRTLTNSLDREQASSYRLVVSGAD  192
             D D     +  I + IV     G   F +   +G VRT    L  +    Y L V   D
Sbjct  576  RDPD----TDHNIHYFIVRDRTGG--RFEVDERSGVVRTRGTDL-FQLDMEYVLYVKAED  628

Query  193  KDGE----GLSTQCECNIKVKDVNDNFPMFRDSQYSARIEEN-ILSSELLRFQVTDLDEE  247
            ++G+       +  E  + +       P F    Y A I EN    S+++  +     +
Sbjct  629  QNGKVDDRRFQSTPEERLSIVG-GKRAPQFYMPSYEAEIPENQKKDSDIISVKA----KS  683

Query  248  YTDNWLAVYFFTSGNEGNWFEIQTDPRTNEGILKVVKALDYEQLQSVKLS----IAVKNK  303
            + D  +       G     F I        GI+K+ K LD+E L+   +      A ++
Sbjct  684  FADREIRYTLKAQGQGAGTFNIGP----TSGIVKLAKELDFEDLRQPHVYSLVVTATEDS  739

Query  304  AEFHQSVISRYRVQSTPVTIQVINVREGIAFRPASKTFTVQKGISSKKLVDYILGT----  359
              F  SV          +TI+V +V +       +  F +    +     D  LGT
Sbjct  740  GGFSTSV---------ELTIRVTDVNDN------APKFELPDYQAHNVDEDIPLGTSILK  784

Query  360  YQAIDEDTNKAASNVKYVMGRN----DGGYLMIDSKTAEIKFVKNMNRDSTFIVNKTITA  415
             +A+D D+  A + ++Y++  +    D   +++++K  +     N N    F+V
Sbjct  785  VKAMDSDSG-ANAEIEYLVSDDHFAVDSNGIIVNNKQLD---ADNNNAYYEFVVTAKDKG  840

Query  416  EVLAIDEYTGKTSTGTVYVRVPDFNDNCP  444
            E         KT T TV V   + ND  P
Sbjct  841  E-------PAKTGTATVRVYTKNKNDEEP  862

 Score = 39.3 bits (90),  Expect = 0.29, Method: Compositional matrix adjust.
 Identities = 54/242 (22%), Positives = 102/242 (42%), Gaps = 22/242 (9%)

Query  4    KFAKPCREGE--DNSKRNP-IAKITSDYQATQKITYRISGVGIDQPPFGIFVVDKNTGDI  60
            +F  P  E E  +N K++  I  + +   A ++I Y +   G      G F +   +G +
Sbjct  656  QFYMPSYEAEIPENQKKDSDIISVKAKSFADREIRYTLKAQGQGA---GTFNIGPTSGIV  712

Query  61   NITAIVDREE--TPSF--LITCRALNAQGLDVEKPLILTVKILDINDNPPVFSQQIFMGE  116
             +   +D E+   P    L+     ++ G      + LT+++ D+NDN P F    +
Sbjct  713  KLAKELDFEDLRQPHVYSLVVTATEDSGGFSTS--VELTIRVTDVNDNAPKFELPDYQAH  770
```

```
Query  117  -IEENSASNSLVMILNATDADEPNHLNSKIAFKIVSQEPAGTPMFLLSRNTGEVRTLTNS  175
             ++E+    + ++ + A D+D  +  N++I + +VS +      F +  N   V
Sbjct  771  NVDEDIPLGTSILKVKAMDSD--SGANAEIEY-LVSDD-----HFAVDSNGIIVNNKQLD  822

Query  176  LDREQASSYRLVVSGADKDGEGLSTQCECNIKVKDVNDNFPMFRDSQYSARIEENILSSE  235
             D   A  Y  VV+  DK     +      +  K+ ND  P F    Y+  ++EN   +
Sbjct  823  ADNNNA-YYEFVVTAKDKGEPAKTGTATVRVYTKNKNDEEPKFSQQVYTPNVDENAGPNT  881

Query  236  LL  237
            L+
Sbjct  882  LV  883

>ACON2_034794.P62470 | transcript=ACON2_034794.R62470 | gene=ACON2_034794
| organism=Anopheles_coluzzii_AcolN3 | gene_product=neural-cadherin,
transcript variant X15, neural-cadherin,
transcript variant X13, neural-cadherin, transcript variant
X8, neural-cadherin, transcript variant X1, neural-cadherin,
transcript variant X3, neural-cadherin, transcript variant
X10, neural-cadherin, transcript variant X16, neural-cadherin,
transcript variant X2, neural-cadherin, transcript variant
X12, neural-cadherin, transcript variant X4, neural-cadherin,
transcript variant X7, neural-cadherin, transcript variant
X9, neural-cadherin, transcript variant X5, neural-cadherin,
transcript variant X17, neural-cadherin, transcript variant
X14, neural-cadherin, transcript variant X11, neural-cadherin,
transcript variant X6 | transcript_product=neural-cadherin,
transcript variant X14 | location=AcolN3_3RL:40912217-41119468(+)
| protein_length=3099 | sequence_SO=chromosome
| SO=protein_coding_gene | is_pseudo=false
Length=3099

 Score = 147 bits (370),  Expect = 3e-35, Method: Compositional matrix adjust.
 Identities = 128/446 (29%), Positives = 217/446 (49%), Gaps = 36/446 (8%)

Query  20    PIAKITS---DYQATQKITYRISGVGID--QPPFGIFVVDKNTGDINITAIVDREET---  71
             PI ++T+   D    Q I Y ++G GID   P    F +++ TG+I +   +DR++
Sbjct  1539  PIMQVTATDGDKDRPQNIVYFLTGQGIDPDNPANSKFDINRTTGEIFVLKPLDRDQPNGR  1598

Query  72    PSFLITCRALNAQGLDVEKPLILTVKILDINDNPPVFSQQIFMGEIEENSASNSLVMILN  131
             P +  T  A +  G  +     + V + DINDN P+F Q ++ G + EN  +  +VM +
Sbjct  1599  PQWRFTVFAQDEGGEGLVGYADVQVNLKDINDNAPIFPQGVYFGNVTENGTAGMVVMTMT  1658

Query  132   ATDADEPNH-LNSKIAFKI---VSQEPAGTPMFLLSRNTGEVRTLTNSLDREQASSYRLV  187
             A D D+PN   N+K+ + I   V +E  G+P+F +  +TG ++T    LDRE+   Y +
Sbjct  1659  AVDYDDPNEGTNAKLIYSIEKNVIEEETGSPIFEIEADTGVIKTAVCCLDRERTPDYSIQ  1718

Query  188   VSGADKDGEGLSTQCECNIKVKDVNDNFPMFRDSQYSARIEENILSS--ELLRFQVTDLD  245
             V      DG GL      +I+VKD+ND  P F   ++   ++E   ++  E+    VT  D
Sbjct  1719  VVAM--DGGGLKGTGTASIRVKDINDMPPQFTKDEWFTEVDETDGTNLPEMPILTVTVHD  1776

Query  246   EEYTDNWLAVYFFTSGNEGNWFEIQTDPRTNEGI--LKVVKALDYE---QLQSVKLSIAV  300
             E+ T+ +       SG   + F +    R N+G   LK+V+ LDYE   Q    +  I V
Sbjct  1777  EDETNKFQYKVIDNSGYGADKFTMV---RNNDGTGSLKIVQPLDYEDPLQSNGFRFRIQV  1833

Query  301   KNKAEFHQSVISRYRVQSTPVTIQVINVREGIAFRPASKTFTVQKGISSKKLVDYILGTY  360
              +K E + +   +Y V  + V +++ ++ +    +P  +   ++  +     V   L T+
Sbjct  1834  NDKGEDNDN--DKYHVAYSWVVVKLRDINDN---KPQFERPNIEVSVYENADVGKTLETF  1888

Query  361   QAIDEDTNKAASNVKYVMGR-NDGGYLMIDSKTAEIKFVKNMNRDSTFIVNKTITAEVLA  419
             +A D D     S V Y + R +D       ++   +   +N++R+    V      ++LA
Sbjct  1889  KATDPDQG-GKSKVSYAIDRSSDRQRQFSINQEGTVTIQRNLDRE----VTPRHQVKILA  1943

Query  420   IDE-YTGKTSTGTVYVRVPDFNDNCP  444
             ID+    KT+T T+ V V D NDN P
Sbjct  1944  IDDGIPPKTATATLTVIVQDINDNPP  1969

 Score = 78.6 bits (192),  Expect = 2e-13, Method: Compositional matrix adjust.
 Identities = 106/424 (25%), Positives = 180/424 (42%), Gaps = 51/424 (12%)
```

```
Query  53    VDKNTGDINITAIVDREETPSFLITCRALNAQGLDVEKPLILTVKILDINDNPPVFSQ-Q  111
             +D++  ++N+TA+ D         +C  +N          ++ V I D+NDN PVF
Sbjct  928   LDRDKYELNVTAMDDG--------SC-CVNGDATIHTSTAVVVVFITDVNDNKPVFKDCG  978

Query  112   IFMGEIEENSASNSLVMILNATDADEPNHLNSKIAFKIVSQEPAGTPMFLLSRNTGEVRT  171
              +  ++EE + + S V+ + ATD D+   +N ++ + IV Q       F +   TGEV T
Sbjct  979   TYYPKVEEGAPNGSPVIKVQATDEDK--GVNGQVKYSIVQQPNQKGTKFTVDEETGEVST  1036

Query  172   LTNSLDRE--QASSYRLVVSGADKDGEGLSTQCECNIKVKDVNDNFPMFRDSQYSARIEE  229
                  DRE        + V   D+    L   C   +++ DVNDN P+F   +Y   +++
Sbjct  1037  -NKVFDREGDDGKFVSVTVKATDQGEPSLEGVCSFTVEITDVNDNPPLFDRQKYVENVKQ  1095

Query  230   NI-LSSELLRFQVTDLDEEYTDNWLAVYFFT---SGNEGNWFEIQTDPRTNEGILKVVKA  285
             +  + + +LR  V+  DE+  +N   VY  +   S N+  +FEIQ  P +   +LK  K
Sbjct  1096  DASIGTNILR--VSASDEDADNNGAIVYTLSAPHSPNDLEYFEIQ--PESGWIVLK--KP  1149

Query  286   LDYEQLQSVKLSIAVKNK------AEFHQSVISRYRVQSTPVTIQV----INVREGIAFR  335
             LD E+    +L +   ++      A+    +    R    P+  +     I++RE +
Sbjct  1150  LDRERY---RLKVYASDRGDPPSTADVDVELDVVDRNNKPPIWDRTIYGPIHIRENVTVG  1206

Query  336   PASKTFTVQKGISSKKLVDYILGTYQAIDEDTNKAASNVKYVMGRNDGGYLMIDSKTAEI  395
                 +     GI     V Y L         TNK   +  Y+  R D G        A+I
Sbjct  1207  TVVTSVKASSGIEGNPTVFYRL--MPGSTAQTNKL--HTFYLQQRPDNG-----DTWADI  1257

Query  396   KFVKNMNRDSTFIVNKTITAEVLAIDEYTGKTSTGTVYYRVPDFNDNCPTAV-LEKDAVC  454
             K    ++ ++    N T+  E     +     S  TVY+++ D ND  P     E++ V
Sbjct  1258  KVNHPLDFETIKEYNLTVRVENNGAQQL---ASEATVYIQLEDVNDEIPLFTEREQETVL  1314

Query  455   SSSP  458
                P
Sbjct  1315  EGEP  1318

 Score = 75.9 bits (185),  Expect = 2e-12, Method: Compositional matrix adjust.
 Identities = 103/418 (25%), Positives = 174/418 (42%), Gaps = 50/418 (12%)

Query  50    IFVVDKNTGDINITAIVDREETPSFLITCRAL----------NAQGLDVEKPLILTVKIL  99
              F ++  +G+I    + DRE+  ++ +   A           N Q   V K   + + I
Sbjct  1355  FFEINLQSGEIFTKVVFDREKQGAYALEVEARDGAPSARPNSNQQPNSVTK--FIRIGIA  1412

Query  100   DINDNPPVFSQQIFMGEIEENSASNSLVMILNATDADEPNHLNSKIAFKIVSQEPAGTPM  159
             D NDNPP F + ++  E++EN      V+ + A D DE    +S+I ++I S    G
Sbjct  1413  DKNDNPPYFDKALYEAEVDENEDIQHTVLTVTAKDHDE----SSRIRYEITSGNIGGA--  1466

Query  160   FLLSRNTGEVRTLTNSLDREQASSYRLVVSGADKDGEGLSTQCECNIKVKDVNDNFPMFR  219
             F +   TG +  +  +LD E    Y L +  +D   E  +      I VKDVND  P F
Sbjct  1467  FAVKNMTGAI-YVAGALDYETRKRYYLTLVASDSLNENETIIV---INVKDVNDMPPSFP  1522

Query  220   DSQYSARIEENI-LSSELLRFQVTDLDEEYTDNWLAVYFFTSG--NEGNWFEIQTDPRTN  276
                Y   ++E + +   +++   TD D++   N   VYF T    +  N    + D
Sbjct  1523  QQVYERTMDEELAVPFPIMQVTATDGDKDRPQN--IVYFLTGQGIDPDNPANSKFDINRT  1580

Query  277   EGILKVVKALDYEQLQS-VKLSIAVKNKAEFHQSVISRYRVQSTPVTIQVINVREGIAFR  335
              G + V+K LD +Q     +    V  + E  + ++    VQ     + + ++ +
Sbjct  1581  TGEIFVLKPLDRDQPNGRPQWRFTVFAQDEGGEGLVGYADVQ-----VNLKDINDNAPIF  1635

Query  336   PASKTFTVQKGISSKKLVDYILGTYQAIDEDTNKAASNVK--YVMGRN------DGGYLM  387
             P    F     ++       ++ T  A+D D     +N K  Y + +N
Sbjct  1636  PQGVYF---GNVTENGTAGMVVMTMTAVDYDDPNEGTNAKLIYSIEKNVIEEETGSPIFE  1692

Query  388   IDSKTAEIK-FVKNMNRDSTFIVNKTITAEVLAIDEYTGKTSTGTVYYRVPDFNDNCP  444
             I++ T  IK  V  ++R+ T       + +V+A+D   G   TGT  +RV D ND  P
Sbjct  1693  IEADTGVIKTAVCCLDRERT----PDYSIQVVAMDG-GGLKGTGTASIRVKDINDMPP  1745

 Score = 65.5 bits (158),  Expect = 3e-09, Method: Compositional matrix adjust.
 Identities = 85/359 (24%), Positives = 153/359 (43%), Gaps = 32/359 (9%)

Query  51    FVVDKNTGDINITAIVDRE-ETPSFL-ITCRALNAQGLDVEKPLILTVKILDINDNPPVF  108
             F VD+ TG+++   + DRE +   F+ +T +A +     +E     TV+I D+NDNPP+F
Sbjct  1025  FTVDEETGEVSTNKVFDREGDDGKFVSVTVKATDQGEPSLEGVCSFTVEITDVNDNPPLF  1084
```

```
Query  109   SQQIFMGEIEENSASNSLVMILNATDADEPNHLNSKIAFKIVS-QEPAGTPMFLLSRNTG  167
              +Q ++  ++++++  + ++ ++A+D D  N  N  I + + +   P     F +   +G
Sbjct  1085  DRQKYVENVKQDASIGTNILRVSASDEDADN--NGAIVYTLSAPHSPNDLEYFEIQPESG  1142

Query  168   EVRTLTNSLDREQASSYRLVVSGADKDGEGLSTQCECNIKVKDVNDNFPMFRDSQYSARI  227
              +  L   LDRE+   YRL V  +D+ G+  ST            +N P   D      I
Sbjct  1143  WI-VLKKPLDRER---YRLKVYASDR-GDPPSTADVDVELDVVDRNNKPPIWDRTIYGPI  1197

Query  228   EENILSSELLRFQVTDLDEEYTDNWLAVYFFTSG-----NEGNWFEIQTDPRTNE--GIL  280
                   +                N    Y    G     N+ + F +Q  P   +    +
Sbjct  1198  HIRENVTVGTVVTSVKASSGIEGNPTVFYRLMPGSTAQTNKLHTFYLQQRPDNGDTWADI  1257

Query  281   KVVKALDYEQLQSVKLSIAVKNKAEFHQSVISRYRVQSTPVTIQVINVREGIAFRPASKT  340
             KV   LD+E ++   L++ V+N         ++       V IQ+ +V + I      +
Sbjct  1258  KVNHPLDFETIKEYNLTVRVENNG-------AQQLASEATVYIQLEDVNDEIPLFTEREQ  1310

Query  341   FTVQKGISSKKLVDYILGTYQAIDEDTNKAASNVKYVM---GRNDG-GYLMIDSKTAEI  395
              TV +G      V  +     AID+D     + V Y +    RN+G  +  I+ ++ EI
Sbjct  1311  ETVLEGEPIGTKVTQV----NAIDKDGTFPNNQVYYYIVDSPRNEGKDFFEINLQSGEI  1365

 Score = 55.8 bits (133),  Expect = 3e-06, Method: Compositional matrix adjust.
 Identities = 49/158 (31%), Positives = 78/158 (49%), Gaps = 24/158 (15%)

Query  102   NDNPPVFSQQIFMGEIEENSASNSLVMILNATDADEPNHLNSKIAFKIVSQEPAGTP--M  159
             ND  P FSQQ++   ++EN+  N+LV  + A+D D  N     + F  V     GT
Sbjct  858   NDEEPKFSQQVYTPNVDENAGPNTLVTTVVASDKDGDN-----VRFGFVG---GGTSSGQ  909

Query  160   FLLSRNTGEVRTLTN--SLDREQASSYRLVVSGADK-------DGEGLSTQCECNIKVKD  210
             F++   TG +R  +   SLDR++   Y L V+  D        D   ++      + + D
Sbjct  910   FVIEEITGVIRLHSKAISLDRDK---YELNVTAMDDGSCCVNGDATIHTSTAVVVVFITD  966

Query  211   VNDNFPMFRD-SQYSARIEENILS-SELLRFQVTDLDE  246
             VNDN P+F+D   Y  ++EE   + S +++ Q TD D+
Sbjct  967   VNDNKPVFKDCGTYYPKVEEGAPNGSPVIKVQATDEDK  1004

 Score = 53.9 bits (128),  Expect = 1e-05, Method: Compositional matrix adjust.
 Identities = 70/306 (23%), Positives = 126/306 (41%), Gaps = 30/306 (10%)

Query  20    PIAKIT-SDYQATQKITYRI---SGVGIDQPPFGIFVVDKNTGDINITAIVDRE---ETP  72
             PI  +T  D   T K  Y++   SG G D+  F +   +  TG + I   +D E   ++
Sbjct  1768  PILTVTVHDEDETNKFQYKVIDNSGYGADK--FTMVRNNDGTGSLKIVQPLDYEDPLQSN  1825

Query  73    SFLITCRALNAQGLDVEKPLI------LTVKILDINDNPPVFSQQIFMGEIEENSASNSL  126
              F    + +N +G D +          + VK+ DINDN P F +      + EN+
Sbjct  1826  GFRFRIQ-VNDKGEDNDNDKYHVAYSWVVVKLRDINDNKPQFERPNIEVSVYENADVGKT  1884

Query  127   VMILNATDADEPNHLNSKIAFKIVSQEPAGTPMFLLSRNTGEVRTLTNSLDREQASSYRL  186
             +    ATD D+     SK+++ I   + +       S N     T+  +LDRE    +++
Sbjct  1885  LETFKATDPDQGG--KSKVSYAI---DRSSDRQRQFSINQEGTVTIQRNLDREVTPRHQV  1939

Query  187   VVSGADKDGEGLSTQCECNIKVKDVNDNFPMFRDSQYSARIEENILSSELLRFQVTDLDE  246
              +   D      +      + V+D+NDN P F    Y   + E++   +++    TD D+
Sbjct  1940  KILAIDDGIPPKTATATLTVIVQDINDNPPKFL-KDYRPVLPEHVPPRKVVEILATDDDD  1998

Query  247   EYTDNWLAVYFFTSGNEGNW----FEIQTDPRTNEG----ILKVVKALDYEQLQSVKLSI  298
                 N     F       +     F+++ D +   G    I+  +++ D EQ +   + I
Sbjct  1999  RSKSNGPPFQFRLDPGADDIIRASFKVEQDQKGANGDGMAIVSSLRSFDREQQKEYLIPI  2058

Query  299   AVKNKA  304
              +K+
Sbjct  2059  VIKDHG  2064

 Score = 52.0 bits (123),  Expect = 4e-05, Method: Compositional matrix adjust.
 Identities = 97/398 (24%), Positives = 163/398 (41%), Gaps = 54/398 (14%)

Query  106   PVFSQQIFMGEIEENSASNSLVMILNATDADEPNHLNSKIAFKIVSQ-EPAGTPMFLLSR  164
             P F    +  EI EN   +S ++ + A      +  + +I + + +Q + AGT  F +
```

```
Sbjct  655   PQFYMPSYEAEIPENQKKDSDIISVKAK-----SFADREIRYTLKAQGQGAGT--FNIGP  707

Query  165   NTGEVRTLTNSLDRE---QASSYRLVVSGADKDGEGLSTQCECNIKVKDVNDNFPMFRDS  221
              +G V+ L   LD E   Q   Y LVV+ A +D  G ST  E  I+V DVNDN P F
Sbjct  708   TSGIVK-LAKELDFEDLRQPHVYSLVVT-ATEDSGGFSTSVELTIRVTDVNDNAPKFELP  765

Query  222   QYSAR-IEENI-LSSELLRFQVTDLDEEYTDNWLAVYFFTSGNEGNWFEIQTDPRTNEGI  279
              Y A  ++E+I L + +L+ +  D D         + +  S +    F + ++     GI
Sbjct  766   DYQAHNVDEDIPLGTSILKVKAMDSDSGAN---AEIEYLVSDDH---FAVDSN-----GI  814

Query  280   LKVVKALDYEQLQS-VKLSIAVKNKAEFHQSVISRYRVQSTPVTIQVINVREGIAFRPAS  338
             +   K LD +   +  +  +  K+K E  ++  +  RV +           E   F
Sbjct  815   IVNNKQLDADNNNAYYEFVVTAKDKGEPAKTGTATVRVYTKNKN------DEEPKFSQQV  868

Query  339   KTFTVQKGISSKKLVDYILGTYQAIDEDTNKAASNVK--YVMGRNDGGYLMIDSKTAEIK  396
              T  V +      LV  ++         ++K   NV+  +V G    G  +I+  T  I+
Sbjct  869   YTPNVDENAGPNTLVTTVVA--------SDKDGDNVRFGFVGGGTSSGQFVIEEITGVIR  920

Query  397   FVK---NMNRDSTFI-VNKTITAEVLAIDEYTGKTSTGTVYVRVPDFNDN------CPTA  446
                   +++RD   + V            + T  TST  V V + D NDN      C T
Sbjct  921   LHSKAISLDRDKYELNVTAMDDGSCCVNGDATIHTSTAVVVVFITDVNDNKPVFKDCGTY  980

Query  447   VLE-KDAVCSSSPSVVVSARTLNNRYTGPYTFALEDQP  483
               + ++   + SP + V A   +    G   +++  QP
Sbjct  981   YPKVEEGAPNGSPVIKVQATDEDKGVNGQVKYSIVQQP  1018

 Score = 44.7 bits (104),  Expect = 0.008, Method: Compositional matrix adjust.
 Identities = 48/199 (24%), Positives = 73/199 (37%), Gaps = 19/199 (10%)

Query  27    DYQATQKITYRISGVGIDQPPFGIFVVDKNTGDINITAIVDREETPSFLITCRALNAQGL  86
             D     K++Y I      Q  F I       G + I   +DRE TP   +   A++
Sbjct  1894  DQGGKSKVSYAIDRSSDRQRQFSI----NQEGTVTIQRNLDREVTPRHQVKILAIDDGIP  1949

Query  87    DVEKPLILTVKILDINDNPPVFSQQIFMGEIEENSASNSLVMILNATDADEPNHLNSKIA  146
                    LTV + DINDNPP F +  +   + E+     +V IL   D D
Sbjct  1950  PKTATATLTVIVQDINDNPPKFLKD-YRPVLPEHVPPRKVVEILATDDDDRSKSNGPPFQ  2008

Query  147   FKIVSQEPAGTPMFLLSRNTGEVRTLTN-----------SLDREQASSYRLVVSGADKDG  195
             F++   +P    +   S    + +   N           S DREQ   Y + +   D
Sbjct  2009  FRL---DPGADDIIRASFKVEQDQKGANGDGMAIVSSLRSFDREQQKEYLIPIVIKDHGN  2065

Query  196   EGLSTQCECNIKVKDVNDN  214
               ++      + + DVNDN
Sbjct  2066  PAMTGTSTLTVVIGDVNDN  2084

 Score = 39.7 bits (91),  Expect = 0.28, Method: Compositional matrix adjust.
 Identities = 89/389 (23%), Positives = 149/389 (38%), Gaps = 62/389 (16%)

Query  75   LITCRALNAQGLDVEKPLILTVKILDINDNPPVFSQQIF--MGEIEENSASNSLVMILNA  132
            +IT +  NA GL       + + + D+ND PP F  +       ++ N+  N+ V  L A
Sbjct  517  IITNQGHNA-GLKYTDNQRVIIHVKDVNDEPPYFINRPLPMQAVVQLNAPPNTPVFTLQA  575

Query  133  TDADEPNHLNSKIAFKIVSQEPAGTPMFLLSRNTGEVRTLTNSLDREQASSYRLVVSGAD  192
             D D     +  I + IV     G   F +   +G VRT    L  +    Y L V   D
Sbjct  576  RDPD----TDHNIHYFIVRDRTGG--RFEVDERSGVVRTRGTDL-FQLDMEYVLYVKAED  628

Query  193  KDGE----GLSTQCECNIKVKDVNDNFPMFRDSQYSARIEEN-ILSSELLRFQVTDLDEE  247
            ++G+       +  E  + +       P F    Y A I EN    S+++  +     +
Sbjct  629  QNGKVDDRRFQSTPEERLSIVG-GKRAPQFYMPSYEAEIPENQKKDSDIISVKA----KS  683

Query  248  YTDNWLAVYFFTSGNEGNWFEIQTDPRTNEGILKVVKALDYEQLQSVKLS----IAVKNK  303
            + D  +       G     F I        GI+K+ K LD+E L+   +      A ++
Sbjct  684  FADREIRYTLKAQGQGAGTFNIGP----TSGIVKLAKELDFEDLRQPHVYSLVVTATEDS  739

Query  304  AEFHQSVISRYRVQSTPVTIQVINVREGIAFRPASKTFTVQKGISSKKLVDYILGT----  359
              F  SV          +TI+V +V +       +  F +    +     D  LGT
Sbjct  740  GGFSTSV---------ELTIRVTDVNDN------APKFELPDYQAHNVDEDIPLGTSILK  784

Query  360  YQAIDEDTNKAASNVKYVMGRN----DGGYLMIDSKTAEIKFVKNMNRDSTFIVNKTITA  415
```

```
             +A+D D+  A + ++Y++  +    D   +++++K  +     N N    F+V
Sbjct  785  VKAMDSDSG-ANAEIEYLVSDDHFAVDSNGIIVNNKQLD---ADNNNAYYEFVVTAKDKG  840

Query  416  EVLAIDEYTGKTSTGTVYVRVPDFNDNCP  444
            E         KT T TV V   + ND  P
Sbjct  841  E-------PAKTGTATVRVYTKNKNDEEP  862

 Score = 39.3 bits (90),  Expect = 0.29, Method: Compositional matrix adjust.
 Identities = 54/242 (22%), Positives = 102/242 (42%), Gaps = 22/242 (9%)

Query  4    KFAKPCREGE--DNSKRNP-IAKITSDYQATQKITYRISGVGIDQPPFGIFVVDKNTGDI  60
            +F  P  E E  +N K++  I  + +   A ++I Y +   G      G F +   +G +
Sbjct  656  QFYMPSYEAEIPENQKKDSDIISVKAKSFADREIRYTLKAQGQGA---GTFNIGPTSGIV  712

Query  61   NITAIVDREE--TPSF--LITCRALNAQGLDVEKPLILTVKILDINDNPPVFSQQIFMGE  116
             +   +D E+   P    L+     ++ G      + LT+++ D+NDN P F    +
Sbjct  713  KLAKELDFEDLRQPHVYSLVVTATEDSGGFSTS--VELTIRVTDVNDNAPKFELPDYQAH  770

Query  117  -IEENSASNSLVMILNATDADEPNHLNSKIAFKIVSQEPAGTPMFLLSRNTGEVRTLTNS  175
             ++E+    + ++ + A D+D  +  N++I + +VS +      F +  N   V
Sbjct  771  NVDEDIPLGTSILKVKAMDSD--SGANAEIEY-LVSDD-----HFAVDSNGIIVNNKQLD  822

Query  176  LDREQASSYRLVVSGADKDGEGLSTQCECNIKVKDVNDNFPMFRDSQYSARIEENILSSE  235
             D   A  Y  VV+  DK     +      +  K+ ND  P F    Y+  ++EN   +
Sbjct  823  ADNNNA-YYEFVVTAKDKGEPAKTGTATVRVYTKNKNDEEPKFSQQVYTPNVDENAGPNT  881

Query  236  LL  237
            L+
Sbjct  882  LV  883

>ACON029589-PB | transcript=ACON029589-RB | gene=ACON029589 |
organism=Anopheles_coluzzii_Ngousso | gene_product=unspecified
product | transcript_product=unspecified product |
location=RWKB01000008:1791488-1991166(+)
| protein_length=3103 | sequence_SO=supercontig
| SO=protein_coding_gene | is_pseudo=false
Length=3103

 Score = 147 bits (370),  Expect = 3e-35, Method: Compositional matrix adjust.
 Identities = 128/446 (29%), Positives = 217/446 (49%), Gaps = 36/446 (8%)

Query  20    PIAKITS---DYQATQKITYRISGVGID--QPPFGIFVVDKNTGDINITAIVDREET---  71
             PI ++T+   D    Q I Y ++G GID   P    F +++ TG+I +   +DR++
Sbjct  1543  PIMQVTATDGDKDRPQNIVYFLTGQGIDPDNPANSKFDINRTTGEIFVLKPLDRDQPNGR  1602

Query  72    PSFLITCRALNAQGLDVEKPLILTVKILDINDNPPVFSQQIFMGEIEENSASNSLVMILN  131
             P +  T  A +  G  +     + V + DINDN P+F Q ++ G + EN  +  +VM +
Sbjct  1603  PQWRFTVFAQDEGGEGLVGYADVQVNLKDINDNAPIFPQGVYFGNVTENGTAGMVVMTMT  1662

Query  132   ATDADEPNH-LNSKIAFKI---VSQEPAGTPMFLLSRNTGEVRTLTNSLDREQASSYRLV  187
             A D D+PN   N+K+ + I   V +E  G+P+F +  +TG ++T    LDRE+   Y +
Sbjct  1663  AVDYDDPNEGTNAKLIYSIEKNVIEEETGSPIFEIEADTGVIKTAVCCLDRERTPDYSIQ  1722

Query  188   VSGADKDGEGLSTQCECNIKVKDVNDNFPMFRDSQYSARIEENILSS--ELLRFQVTDLD  245
             V     DG GL       +I+VKD+ND  P F   ++   ++E   ++  E+    VT  D
Sbjct  1723  VVAM--DGGGLKGTGTASIRVKDINDMPPQFTKDEWFTEVDETDGTNLPEMPILTVTVHD  1780

Query  246   EEYTDNWLAVYFFTSGNEGNWFEIQTDPRTNEGI--LKVVKALDYE---QLQSVKLSIAV  300
             E+ T+ +        SG    + F +    R N+G   LK+V+ LDYE   Q    +  I V
Sbjct  1781  EDETNKFQYKVIDNSGYGADKFTMV---RNNDGTGSLKIVQPLDYEDPLQSNGFRFRIQV  1837

Query  301   KNKAEFHQSVISRYRVQSTPVTIQVINVREGIAFRPASKTFTVQKGISSKKLVDYILGTY  360
              +K E + +   +Y V  + V +++ ++ +    +P  +   ++  +     V   L T+
Sbjct  1838  NDKGEDNDN--DKYHVAYSWVVVKLRDINDN---KPQFERPNIEVSVYENADVGKTLETF  1892

Query  361   QAIDEDTNKAASNVKYVMGR-NDGGYLMIDSKTAEIKFVKNMNRDSTFIVNKTITAEVLA  419
             +A D D     S V Y + R +D       ++   +   +N++R+    V      ++LA
Sbjct  1893  KATDPDQG-GKSKVSYAIDRSSDRQRQFSINQEGTVTIQRNLDRE----VTPRHQVKILA  1947
```

```
Query  420   IDE-YTGKTSTGTVYYVRVPDFNDNCP  444
             ID+    KT+T T+ V V D NDN P
Sbjct  1948  IDDGIPPKTATATLTVIVQDINDNPP  1973

 Score = 75.9 bits (185),  Expect = 2e-12, Method: Compositional matrix adjust.
 Identities = 106/426 (25%), Positives = 181/426 (42%), Gaps = 51/426 (12%)

Query  53    VDKNTGDINITAIVDREETPSFLITCRALNAQGLDVEKPLILTVKILDINDNPPVFSQ-Q  111
             +D++  ++N+TA+ D         +C  +N          ++ V I D+NDN PVF
Sbjct  928   LDRDKYELNVTAMDDG--------SC-CVNGDATIHTSTAVVVVFITDVNDNKPVFKDCG  978

Query  112   IFMGEIEENSASNSLVMILNATDADEPNHLNSKIAFKIVSQEPAGTPMFLLSRNTGEVRT  171
              +  ++EE + + S V+ + ATD D+   +N ++ + IV Q       F +   TGEV T
Sbjct  979   TYYPKVEEGAPNGSPVIKVQATDEDK--GVNGQVKYSIVQQPNQKGTKFTVDEETGEVST  1036

Query  172   LTNSLDRE--QASSYRLVVSGADKDGEGLSTQCECNIKVKDVNDNFPMFRDSQYSARIEE  229
                  DRE        + V   D+    L   C   +++ DVNDN P+F   +Y   +++
Sbjct  1037  -NKVFDREGDDGKFVSVTVKATDQGEPSLEGVCSFTVEITDVNDNPPLFDRQKYVENVKQ  1095

Query  230   NI-LSSELLRFQVTDLDEEYTDNWLAVYFFT---SGNEGNWFEIQTDPRTNEGILKVVKA  285
             +  + + +LR  V+  DE+  +N   VY  +   S N+  +FEIQ  P +   +LK  K
Sbjct  1096  DASIGTNILR--VSASDEDADNNGAIVYTLSAPHSPNDLEYFEIQ--PESGWIVLK--KP  1149

Query  286   LD--YEQLQSVKLSIAVKNK------AEFHQSVISRYRVQSTPVTIQV----INVREGIA  333
             LD  Y + +  +L +   ++      A+    +    R    P+  +     I++RE +
Sbjct  1150  LDGCYSR-ERYRLKVYASDRGDPPSTADVDVELDVVDRNNKPPIWDRTIYGPIHIRENVT  1208

Query  334   FRPASKTFTVQKGISSKKLVDYILGTYQAIDEDTNKAASNVKYVMGRNDGGYLMIDSKTA  393
                   +     GI     V Y L         TNK   +  Y+  R D G        A
Sbjct  1209  VGTVVTSVKASSGIEGNPTVFYRL--MPGSTAQTNKL--HTFYLQQRPDNG-----DTWA  1259

Query  394   EIKFVKNMNRDSTFIVNKTITAEVLAIDEYTGKTSTGTVYYVRVPDFNDNCPTAV-LEKDA  452
             +IK    ++ ++    N T+  E     +     S  TVY+++ D ND  P     E++
Sbjct  1260  DIKVNHPLDFETIKEYNLTVRVENNGAQQL---ASEATVYIQLEDVNDEIPLFTEREQET  1316

Query  453   VCSSSP  458
             V    P
Sbjct  1317  VLEGEP  1322

 Score = 75.9 bits (185),  Expect = 2e-12, Method: Compositional matrix adjust.
 Identities = 103/418 (25%), Positives = 174/418 (42%), Gaps = 50/418 (12%)

Query  50    IFVVDKNTGDINITAIVDREETPSFLITCRAL----------NAQGLDVEKPLILTVKIL  99
              F ++  +G+I    + DRE+  ++ +   A           N Q   V K   + + I
Sbjct  1359  FFEINLQSGEIFTKVVFDREKQGAYALEVEARDGAPSARPNSNQQPNSVTK--FIRIGIA  1416

Query  100   DINDNPPVFSQQIFMGEIEENSASNSLVMILNATDADEPNHLNSKIAFKIVSQEPAGTPM  159
             D NDNPP F + ++  E++EN      V+ + A D DE    +S+I ++I S    G
Sbjct  1417  DKNDNPPYFDKALYEAEVDENEDIQHTVLTVTAKDHDE----SSRIRYEITSGNIGGA--  1470

Query  160   FLLSRNTGEVRTLTNSLDREQASSYRLVVSGADKDGEGLSTQCECNIKVKDVNDNFPMFR  219
             F +   TG +  +  +LD E    Y L +  +D   E  +      I VKDVND  P F
Sbjct  1471  FAVKNMTGAI-YVAGALDYETRKRYYLTLVASDSLNENETIIV---INVKDVNDMPPSFP  1526

Query  220   DSQYSARIEENI-LSSELLRFQVTDLDEEYTDNWLAVYFFTSG--NEGNWFEIQTDPRTN  276
                Y   ++E + +   +++   TD D++   N   VYF T    +  N    + D
Sbjct  1527  QQVYERTMDEELAVPFPIMQVTATDGDKDRPQN--IVYFLTGQGIDPDNPANSKFDINRT  1584

Query  277   EGILKVVKALDYEQLQS-VKLSIAVKNKAEFHQSVISRYRVQSTPVTIQVINVREGIAFR  335
              G + V+K LD +Q     +    V  + E  + ++    VQ     + + ++ +
Sbjct  1585  TGEIFVLKPLDRDQPNGRPQWRFTVFAQDEGGEGLVGYADVQ-----VNLKDINDNAPIF  1639

Query  336   PASKTFTVQKGISSKKLVDYILGTYQAIDEDTNKAASNVK--YVMGRN------DGGYLM  387
             P    F     ++       ++ T  A+D D     +N K  Y + +N
Sbjct  1640  PQGVYF---GNVTENGTAGMVVMTMTAVDYDDPNEGTNAKLIYSIEKNVIEEETGSPIFE  1696

Query  388   IDSKTAEIK-FVKNMNRDSTFIVNKTITAEVLAIDEYTGKTSTGTVYYVRVPDFNDNCP  444
             I++ T  IK  V  ++R+ T       + +V+A+D   G   TGT  +RV D ND  P
Sbjct  1697  IEADTGVIKTAVCCLDRERT----PDYSIQVVAMDG-GGLKGTGTASIRVKDINDMPP  1749
```

```
 Score = 60.5 bits (145),  Expect = 1e-07, Method: Compositional matrix adjust.
 Identities = 83/362 (23%), Positives = 152/362 (42%), Gaps = 34/362 (9%)

Query  51    FVVDKNTGDINITAIVDRE-ETPSFL-ITCRALNAQGLDVEKPLILTVKILDINDNPPVF  108
             F VD+ TG+++   + DRE +   F+ +T +A +     +E     TV+I D+NDNPP+F
Sbjct  1025  FTVDEETGEVSTNKVFDREGDDGKFVSVTVKATDQGEPSLEGVCSFTVEITDVNDNPPLF  1084

Query  109   SQQIFMGEIEENSASNSLVMILNATDADEPNHLNSKIAFKIVS-QEPAGTPMFLLSRNTG  167
              +Q ++  ++++++  + ++ ++A+D D  N  N  I + + +   P     F +   +G
Sbjct  1085  DRQKYVENVKQDASIGTNILRVSASDEDADN--NGAIVYTLSAPHSPNDLEYFEIQPESG  1142

Query  168   EV---RTLTNSLDREQASSYRLVVSGADKDGEGLSTQCECNIKVKDVNDNFPMFRDSQYS  224
              +   + L     RE+   YRL V  +D+ G+  ST            +N P   D
Sbjct  1143  WIVLKKPLDGCYSRER---YRLKVYASDR-GDPPSTADVDVELDVVDRNNKPPIWDRTIY  1198

Query  225   ARIEENILSSELLRFQVTDLDEEYTDNWLAVYFFTSG-----NEGNWFEIQTDPRTNE--  277
               I      +                N    Y    G     N+ + F +Q  P   +
Sbjct  1199  GPIHIRENVTVGTVVTSVKASSGIEGNPTVFYRLMPGSTAQTNKLHTFYLQQRPDNGDTW  1258

Query  278   GILKVVKALDYEQLQSVKLSIAVKNKAEFHQSVISRYRVQSTPVTIQVINVREGIAFRPA  337
               +KV   LD+E ++   L++ V+N         ++       V IQ+ +V + I
Sbjct  1259  ADIKVNHPLDFETIKEYNLTVRVENNG-------AQQLASEATVYIQLEDVNDEIPLFTE  1311

Query  338   SKTFTVQKGISSKKLVDYILGTYQAIDEDTNKAASNVKYVM---GRNDG-GYLMIDSKTA  393
              +  TV +G      V  +     AID+D     + V Y +    RN+G  +  I+ ++
Sbjct  1312  REQETVLEGEPIGTKVTQV----NAIDKDGTFPNNQVYYYIVDSPRNEGKDFFEINLQSG  1367

Query  394   EI  395
             EI
Sbjct  1368  EI  1369

 Score = 55.8 bits (133),  Expect = 3e-06, Method: Compositional matrix adjust.
 Identities = 49/158 (31%), Positives = 78/158 (49%), Gaps = 24/158 (15%)

Query  102   NDNPPVFSQQIFMGEIEENSASNSLVMILNATDADEPNHLNSKIAFKIVSQEPAGTP--M  159
             ND  P FSQQ++   ++EN+  N+LV  + A+D D  N     + F  V     GT
Sbjct  858   NDEEPKFSQQVYTPNVDENAGPNTLVTTVVASDKDGDN-----VRFGFVG---GGTSSGQ  909

Query  160   FLLSRNTGEVRTLTN--SLDREQASSYRLVVSGADK-------DGEGLSTQCECNIKVKD  210
             F++   TG +R  +   SLDR++   Y L V+  D        D    ++     + + D
Sbjct  910   FVIEEITGVIRLHSKAISLDRDK---YELNVTAMDDGSCCVNGDATIHTSTAVVVVFITD  966

Query  211   VNDNFPMFRD-SQYSARIEENILS-SELLRFQVTDLDE  246
             VNDN P+F+D   Y  ++EE   + S +++ Q TD D+
Sbjct  967   VNDNKPVFKDCGTYYPKVEEGAPNGSPVIKVQATDEDK  1004

 Score = 53.9 bits (128),  Expect = 1e-05, Method: Compositional matrix adjust.
 Identities = 70/306 (23%), Positives = 126/306 (41%), Gaps = 30/306 (10%)

Query  20    PIAKIT-SDYQATQKITYRI---SGVGIDQPPFGIFVVDKNTGDINITAIVDRE---ETP  72
             PI  +T  D   T K  Y++   SG G D+  F +   +  TG + I   +D E   ++
Sbjct  1772  PILTVTVHDEDETNKFQYKVIDNSGYGADK--FTMVRNNDGTGSLKIVQPLDYEDPLQSN  1829

Query  73    SFLITCRALNAQGLDVEKPLI------LTVKILDINDNPPVFSQQIFMGEIEENSASNSL  126
              F    + +N +G D +          + VK+ DINDN P F +      + EN+
Sbjct  1830  GFRFRIQ-VNDKGEDNDNDKYHVAYSWVVVKLRDINDNKPQFERPNIEVSVYENADVGKT  1888

Query  127   VMILNATDADEPNHLNSKIAFKIVSQEPAGTPMFLLSRNTGEVRTLTNSLDREQASSYRL  186
             +    ATD D+     SK+++ I   + +       S N     T+  +LDRE    +++
Sbjct  1889  LETFKATDPDQGG--KSKVSYAI---DRSSDRQRQFSINQEGTVTIQRNLDREVTPRHQV  1943

Query  187   VVSGADKDGEGLSTQCECNIKVKDVNDNFPMFRDSQYSARIEENILSSELLRFQVTDLDE  246
              +   D      +      + V+D+NDN P F    Y   + E++   +++    TD D+
Sbjct  1944  KILAIDDGIPPKTATATLTVIVQDINDNPPKFL-KDYRPVLPEHVPPRKVVEILATDDDD  2002

Query  247   EYTDNWLAVYFFTSGNEGNW----FEIQTDPRTNEG----ILKVVKALDYEQLQSVKLSI  298
                 N     F       +     F+++ D +   G    I+  +++ D EQ +   + I
```

```
Sbjct  2003  RSKSNGPPFQFRLDPGADDIIRASFKVEQDQKGANGDGMAIVSSLRSFDREQQKEYLIPI  2062

Query  299   AVKNKA  304
              +K+
Sbjct  2063  VIKDHG  2068

 Score = 52.0 bits (123),  Expect = 4e-05, Method: Compositional matrix adjust.
 Identities = 97/398 (24%), Positives = 163/398 (41%), Gaps = 54/398 (14%)

Query  106   PVFSQQIFMGEIEENSASNSLVMILNATDADEPNHLNSKIAFKIVSQ-EPAGTPMFLLSR  164
             P F    +  EI EN   +S ++ + A      +  + +I + + +Q + AGT  F +
Sbjct  655   PQFYMPSYEAEIPENQKKDSDIISVKAK-----SFADREIRYTLKAQGQGAGT--FNIGP  707

Query  165   NTGEVRTLTNSLDRE---QASSYRLVVSGADKDGEGLSTQCECNIKVKDVNDNFPMFRDS  221
              +G V+ L   LD E   Q   Y LVV+ A +D  G ST  E  I+V DVNDN P F
Sbjct  708   TSGIVK-LAKELDFEDLRQPHVYSLVVT-ATEDSGGFSTSVELTIRVTDVNDNAPKFELP  765

Query  222   QYSAR-IEENI-LSSELLRFQVTDLDEEYTDNWLAVYFFTSGNEGNWFEIQTDPRTNEGI  279
              Y A  ++E+I L + +L+ +  D D         + +  S +    F + ++     GI
Sbjct  766   DYQAHNVDEDIPLGTSILKVKAMDSDSGAN---AEIEYLVSDDH---FAVDSN-----GI  814

Query  280   LKVVKALDYEQLQS-VKLSIAVKNKAEFHQSVISRYRVQSTPVTIQVINVREGIAFRPAS  338
             +   K LD +   +  +  +  K+K E  ++  +  RV +           E   F
Sbjct  815   IVNNKQLDADNNNAYYEFVVTAKDKGEPAKTGTATVRVYTKNKN------DEEPKFSQQV  868

Query  339   KTFTVQKGISSKKLVDYILGTYQAIDEDTNKAASNVK--YVMGRNDGGYLMIDSKTAEIK  396
              T  V +      LV  ++         ++K   NV+  +V G    G  +I+  T  I+
Sbjct  869   YTPNVDENAGPNTLVTTVVA--------SDKDGDNVRFGFVGGGTSSGQFVIEEITGVIR  920

Query  397   FVK---NMNRDSTFI-VNKTITAEVLAIDEYTGKTSTGTVYVRVPDFNDN------CPTA  446
                   +++RD   + V            + T  TST  V V + D NDN      C T
Sbjct  921   LHSKAISLDRDKYELNVTAMDDGSCCVNGDATIHTSTAVVVVFITDVNDNKPVFKDCGTY  980

Query  447   VLE-KDAVCSSSPSVVVSARTLNNRYTGPYTFALEDQP  483
               + ++   + SP + V A   +    G   +++  QP
Sbjct  981   YPKVEEGAPNGSPVIKVQATDEDKGVNGQVKYSIVQQP  1018

 Score = 44.3 bits (103),  Expect = 0.008, Method: Compositional matrix adjust.
 Identities = 48/199 (24%), Positives = 73/199 (37%), Gaps = 19/199 (10%)

Query  27    DYQATQKITYRISGVGIDQPPFGIFVVDKNTGDINITAIVDREETPSFLITCRALNAQGL  86
             D     K++Y I      Q  F I       G + I   +DRE TP   +   A++
Sbjct  1898  DQGGKSKVSYAIDRSSDRQRQFSI----NQEGTVTIQRNLDREVTPRHQVKILAIDDGIP  1953

Query  87    DVEKPLILTVKILDINDNPPVFSQQIFMGEIEENSASNSLVMILNATDADEPNHLNSKIA  146
                    LTV + DINDNPP F +  +   + E+     +V IL   D D
Sbjct  1954  PKTATATLTVIVQDINDNPPKFLKD-YRPVLPEHVPPRKVVEILATDDDDRSKSNGPPFQ  2012

Query  147   FKIVSQEPAGTPMFLLSRNTGEVRTLTN-----------SLDREQASSYRLVVSGADKDG  195
             F++   +P    +   S    + +   N           S DREQ   Y + +   D
Sbjct  2013  FRL---DPGADDIIRASFKVEQDQKGANGDGMAIVSSLRSFDREQQKEYLIPIVIKDHGN  2069

Query  196   EGLSTQCECNIKVKDVNDN  214
               ++      + + DVNDN
Sbjct  2070  PAMTGTSTLTVVIGDVNDN  2088

 Score = 39.7 bits (91),  Expect = 0.28, Method: Compositional matrix adjust.
 Identities = 89/389 (23%), Positives = 149/389 (38%), Gaps = 62/389 (16%)

Query  75   LITCRALNAQGLDVEKPLILTVKILDINDNPPVFSQQIF--MGEIEENSASNSLVMILNA  132
            +IT +  NA GL       + + + D+ND PP F  +       ++ N+  N+ V  L A
Sbjct  517  IITNQGHNA-GLKYTDNQRVIIHVKDVNDEPPYFINRPLPMQAVVQLNAPPNTPVFTLQA  575

Query  133  TDADEPNHLNSKIAFKIVSQEPAGTPMFLLSRNTGEVRTLTNSLDREQASSYRLVVSGAD  192
             D D     +  I + IV     G   F +   +G VRT    L  +    Y L V   D
Sbjct  576  RDPD----TDHNIHYFIVRDRTGG--RFEVDERSGVVRTRGTDL-FQLDMEYVLYVKAED  628

Query  193  KDGE----GLSTQCECNIKVKDVNDNFPMFRDSQYSAR-IEEN-ILSSELLRFQVTDLDEE  247
```

```
            ++G+       +  E  + +       P F    Y A I EN    S+++  +     +
Sbjct  629  QNGKVDDRRFQSTPEERLSIVG-GKRAPQFYMPSYEAEIPENQKKDSDIISVKA----KS  683

Query  248  YTDNWLAVYFFTSGNEGNWFEIQTDPRTNEGILKVVKALDYEQLQSVKLS----IAVKNK  303
            + D  +       G     F I        GI+K+ K LD+E L+    +      A ++
Sbjct  684  FADREIRYTLKAQGQGAGTFNIGP----TSGIVKLAKELDFEDLRQPHVYSLVVTATEDS  739

Query  304  AEFHQSVISRYRVQSTPVTIQVINVREGIAFRPASKTFTVQKGISSKKLVDYILGT----  359
              F  SV          +TI+V +V +       +  F +    +     D  LGT
Sbjct  740  GGFSTSV---------ELTIRVTDVNDN------APKFELPDYQAHNVDEDIPLGTSILK  784

Query  360  YQAIDEDTNKAASNVKYVMGRN----DGGYLMIDSKTAEIKFVKNMNRDSTFIVNKTITA  415
             +A+D D+  A + ++Y++  +    D   +++++K  +     N N    F+V
Sbjct  785  VKAMDSDSG-ANAEIEYLVSDDHFAVDSNGIIVNNKQLD---ADNNNAYYEFVVTAKDKG  840

Query  416  EVLAIDEYTGKTSTGTVYVRVPDFNDNCP  444
            E         KT T TV V   + ND  P
Sbjct  841  E-------PAKTGTATVRVYTKNKNDEEP  862

 Score = 39.3 bits (90),  Expect = 0.30, Method: Compositional matrix adjust.
 Identities = 54/242 (22%), Positives = 102/242 (42%), Gaps = 22/242 (9%)

Query  4    KFAKPCREGE--DNSKRNP-IAKITSDYQATQKITYRISGVGIDQPPFGIFVVDKNTGDI  60
            +F  P  E E  +N K++  I  + +   A ++I Y +   G      G F +   +G +
Sbjct  656  QFYMPSYEAEIPENQKKDSDIISVKAKSFADREIRYTLKAQGQGA---GTFNIGPTSGIV  712

Query  61   NITAIVDREE--TPSF--LITCRALNAQGLDVEKPLILTVKILDINDNPPVFSQQIFMGE  116
             +   +D E+   P    L+     ++ G      + LT+++ D+NDN P F    +
Sbjct  713  KLAKELDFEDLRQPHVYSLVVTATEDSGGFSTS--VELTIRVTDVNDNAPKFELPDYQAH  770

Query  117  -IEENSASNSLVMILNATDADEPNHLNSKIAFKIVSQEPAGTPMFLLSRNTGEVRTLTNS  175
             ++E+    + ++ + A D+D  +  N++I + +VS +      F +  N   V
Sbjct  771  NVDEDIPLGTSILKVKAMDSD--SGANAEIEY-LVSDD-----HFAVDSNGIIVNNKQLD  822

Query  176  LDREQASSYRLVVSGADKDGEGLSTQCECNIKVKDVNDNFPMFRDSQYSARIEENILSSE  235
             D   A  Y  VV+  DK     +      +  K+ ND  P F    Y+  ++EN   +
Sbjct  823  ADNNNA-YYEFVVTAKDKGEPAKTGTATVRVYTKNKNDEEPKFSQQVYTPNVDENAGPNT  881

Query  236  LL  237
            L+
Sbjct  882  LV  883

>AALB20_033171.P66493 | transcript=AALB20_033171.R66493 | gene=AALB20_033171
| organism=Anopheles_albimanus_STECLA_2020 |
gene_product=neural-cadherin, transcript variant X38 |
transcript_product=neural-cadherin,
transcript variant X36 | location=CM024292:75037560-75208393(+)
| protein_length=3122 | sequence_SO=chromosome
| SO=protein_coding_gene | is_pseudo=false
Length=3122

 Score = 147 bits (370),  Expect = 3e-35, Method: Compositional matrix adjust.
 Identities = 128/446 (29%), Positives = 217/446 (49%), Gaps = 36/446 (8%)

Query  20    PIAKITS---DYQATQKITYRISGVGID--QPPFGIFVVDKNTGDINITAIVDREET---  71
             PI ++T+   D    Q I Y ++G GID   P    F +++ TG+I +   +DR++
Sbjct  1563  PIMQVTATDGDKDRPQNIVYFLTGQGIDPDNPANSKFDINRTTGEIFVLKPLDRDQPNGR  1622

Query  72    PSFLITCRALNAQGLDVEKPLILTVKILDINDNPPVFSQQIFMGEIEENSASNSLVMILN  131
             P +  T  A +  G  +     + V + DINDN P+F Q ++ G + EN  +  +VM +
Sbjct  1623  PQWRFTVFAQDEGGEGLVGYADVQVNLKDINDNAPIFPQGVYFGNVTENGTAGMVVMTMT  1682

Query  132   ATDADEPNH-LNSKIAFKI---VSQEPAGTPMFLLSRNTGEVRTLTNSLDREQASSYRLV  187
             A D D+PN   N+K+ + I   V +E  G+P+F +  +TG ++T    LDRE+   Y +
Sbjct  1683  AVDYDDPNEGTNAKLIYSIEKNVIEEETGSPIFEIEADTGVIKTAVCCLDRERTPDYSIQ  1742

Query  188   VSGADKDGEGLSTQCECNIKVKDVNDNFPMFRDSQYSARIEENILSS--ELLRFQVTDLD  245
             V     DG GL      +I+VKD+ND  P F  ++    ++E   ++  E+    VT  D
Sbjct  1743  VVAM--DGGGLKGTGTASIRVKDINDMPPQFTKDEWFTEVDETDGTNLPEMPILTVTVHD  1800
```

```
Query  246    EEYTDNWLAVYFFTSGNEGNWFEIQTDPRTNEGI--LKVVKALDYE---QLQSVKLSIAV  300
              E+ T+ +       SG    + F +    R N+G   LK+V+ LDYE   Q    +  I V
Sbjct  1801   EDETNKFQYKVIDNSGYADKFTMV---RNNDGTGSLKIVQPLDYEDPLQSNGFRFRIQV  1857

Query  301    KNKAEFHQSVISRYRVQSTPVTIQVINVREGIAFRPASKTFTVQKGISSKKLVDYILGTY  360
               +K E + +   +Y V  + V +++ ++ +     +P  +   ++  +      V   L T+
Sbjct  1858   NDKGEDNDN--DKYHVAYSWVVVKLRDINDN---KPQFERPNIEVSVYENADVGKTLETF  1912

Query  361    QAIDEDTNKAASNVKYVMGR-NDGGYLMIDSKTAEIKFVKNMNRDSTFIVNKTITAEVLA  419
              +A D D     S V Y + R +D        ++   +   +N++R+    V       ++LA
Sbjct  1913   KATDPDQG-GKSKVSYAIDRSSDRQRQFSINQEGTVTIQRNLDRE----VTPRHQVKILA  1967

Query  420    IDE-YTGKTSTGTVYYRVPDFNDNCP  444
              ID+    KT+T T+ V V D NDN P
Sbjct  1968   IDDGIPPKTATATLTVIVQDINDNPP  1993

 Score = 79.3 bits (194),  Expect = 1e-13, Method: Compositional matrix adjust.
 Identities = 104/419 (25%), Positives = 173/419 (41%), Gaps = 51/419 (12%)

Query  50     IFVVDKNTGDINITAIVDREETPSFLITCRAL----------NAQGLDVEKPLILTVKIL  99
               F ++  +G+I    + DRE+  ++ +   A            N Q   V K   + + I
Sbjct  1378   FFEINLQSGEIFTKVVFDREKQGAYALEVEARDGAPSARPNSNQQPNSVTK--FIRIGIA  1435

Query  100    DINDNPPVFSQQIFMGEIEENSASNSLVMILNATDADEPNHLNSKIAFKIVSQEPAGTPM  159
              D NDNPP F + ++  E++EN       V+ + A D DE    +S+I ++I S    G
Sbjct  1436   DKNDNPPYFDKALYEAEVDENEDIQHTVLTVTAKDHDE----SSRIRYEITSGNIGGA--  1489

Query  160    FLLSRNTGEVRTLTNSLDREQASSYRLVVSGADKDGEGLSTQCECNIKVKDVNDNFPMFR  219
              F +   TG +  +  +LD E    Y L +  +D   E  +       I VKDVND  P F
Sbjct  1490   FAVKNMTGAI-YVAGALDYETRKRYHLTLVASDSLNENATI---IEIHVKDVNDMPPTFS  1545

Query  220    DSQYSARI--EENILSSELLRFQVTDLDEEYTDNWLAVYFFTSG--NEGNWFEIQTDPRT  275
                Y A    E+ +    +++   TD D++   N   VYF T    +  N    + D
Sbjct  1546   QQIYEATQDEEQTVTDRPIMQVTATDGDKDRPQN--IVYFLTGQGIDPDNPANSKFDINR  1603

Query  276    NEGILKVVKALDYEQLQS-VKLSIAVKNKAEFHQSVISRYRVQSTPVTIQVINVREGIAF  334
                G + V+K LD +Q      +     V  + E  + ++    VQ     + + ++ +
Sbjct  1604   TTGEIFVLKPLDRDQPNGRPQWRFTVFAQDEGGEGLVGYADVQ-----VNLKDINDNAPI  1658

Query  335    RPASKTFTVQKGISSKKLVDYILGTYQAIDEDTNKAASNVK--YVMGRN------DGGYL  386
               P    F     ++        ++ T  A+D D     +N K  Y + +N
Sbjct  1659   FPQGVYF---GNVTENGTAGMVVMTMTAVDYDDPNEGTNAKLIYSIEKNVIEEETGSPIF  1715

Query  387    MIDSKTAEIK-FVKNMNRDSTFIVNKTITAEVLAIDEYTGKTSTGTVYYRVPDFNDNCP  444
               I++ T  IK  V  ++R+ T        + +V+A+D   G   TGT  +RV D ND  P
Sbjct  1716   EIEADTGVIKTAVCCLDRERT----PDYSIQVVAMDG-GGLKGTGTASIRVKDINDMPP  1769

 Score = 73.9 bits (180),  Expect = 7e-12, Method: Compositional matrix adjust.
 Identities = 90/362 (25%), Positives = 166/362 (46%), Gaps = 37/362 (10%)

Query  51     FVVDKNTGDINITAIVDRE-ETPSFL-ITCRALNAQGLDVEKPLILTVKILDINDNPPVF  108
              F VD+ TG+++    + DRE +   F+ +T +A +      +E      TV+I D+NDNPP+F
Sbjct  1047   FTVDEETGEVSTNKVFDREGDDGKFVSVTVKATDQGEPSLEGVCSFTVEITDVNDNPPLF  1106

Query  109    SQQIFMGEIEENSASNSLVMILNATDADEPNHLNSKIAFKIVS-QEPAGTPMFLLSRNTG  167
               +Q ++  ++++++  + ++ ++A+D D  N  N  I + + +    P      F +   +G
Sbjct  1107   DRQKYVENVKQDASIGTNILRVSASDEDADN--NGAIVYTLSAPHSPNDLEYFEIQPESG  1164

Query  168    EVRTLTNSLDREQASSYRLVVSGADKDGEGLSTQCECNIKVKDVNDNFPMFRDSQYSARI  227
               +  L   LDRE    +Y+L       DK     LS   E  I V D  +N P++ ++      I
Sbjct  1165   WI-VLKKPLDRE---TYKLEAMAQDKGYPPLSRTVEVQIDVVDRANNPPVWDNTGVYGPI  1220

Query  228    --EENI-LSSELLRFQVTDLDEEYTDNWLAVYFFTSG-----NEGNWFEIQTDPRTNE--  277
                +EN+ +  +++  + +   E    N     Y    G       N+ + F +Q   P    +
Sbjct  1221   YVKENMPVGGKVVSIKASSGIE---GNPTVFYRLMPGSTAQTNKLHTFYLQQRPDNGDTW  1277

Query  278    GILKVVKALDYEQLQSVKLSIAVKNKAEFHQSVISRYRVQSTPVTIQVINVREGIAFRPA  337
                +KV   LD+E ++   L+I V+N          ++        V IQ+ +V + I
```

```
Sbjct  1278  ADIKVNHPLDFETIKEYNLTIRVENNG-------AQQLASEATVYIQLEDVNDEIPLFTE  1330

Query  338   SKTFTVQKGISSKKLVDYILGTYQAIDEDTNKAASNVKYVM---GRNDG-GYLMIDSKTA  393
              +  TV +G      V  +     AID+D     + V Y +    RN+G  +  I+ ++
Sbjct  1331  REQETVLEGEPIGTKVTQV----NAIDKDGTFPNNQVYYYIVDSPRNEGKDFFEINLQSG  1386

Query  394   EI  395
             EI
Sbjct  1387  EI  1388

 Score = 55.8 bits (133),  Expect = 3e-06, Method: Compositional matrix adjust.
 Identities = 49/158 (31%), Positives = 78/158 (49%), Gaps = 24/158 (15%)

Query  102   NDNPPVFSQQIFMGEIEENSASNSLVMILNATDADEPNHLNSKIAFKIVSQEPAGTP--M  159
             ND  P FSQQ++   ++EN+  N+LV  + A+D D  N     + F  V     GT
Sbjct  880   NDEEPKFSQQVYTPNVDENAGPNTLVTTVVASDKDGDN-----VRFGFVG---GGTSSGQ  931

Query  160   FLLSRNTGEVRTLTN--SLDREQASSYRLVVSGADK-------DGEGLSTQCECNIKVKD  210
             F++   TG +R  +   SLDR++   Y L V+  D        D    ++     + + D
Sbjct  932   FVIEEITGVIRLHSKAISLDRDK---YELNVTAMDDGSCCVNGDATIHTSTAVVVVFITD  988

Query  211   VNDNFPMFRD-SQYSARIEENILS-SELLRFQVTDLDE  246
             VNDN P+F+D   Y  ++EE   + S +++ Q TD D+
Sbjct  989   VNDNKPVFKDCGTYYPKVEEGAPNGSPVIKVQATDEDK  1026

 Score = 53.5 bits (127),  Expect = 1e-05, Method: Compositional matrix adjust.
 Identities = 70/306 (23%), Positives = 126/306 (41%), Gaps = 30/306 (10%)

Query  20    PIAKIT-SDYQATQKITYRI---SGVGIDQPPFGIFVVDKNTGDINITAIVDRE---ETP  72
             PI  +T  D   T K  Y++   SG G D+  F +   +  TG + I   +D E   ++
Sbjct  1792  PILTVTVHDEDETNKFQYKVIDNSGYGADK--FTMVRNNDGTGSLKIVQPLDYEDPLQSN  1849

Query  73    SFLITCRALNAQGLDVEKPLI------LTVKILDINDNPPVFSQQIFMGEIEENSASNSL  126
              F    + +N +G D +          + VK+ DINDN P F +      + EN+
Sbjct  1850  GFRFRIQ-VNDKGEDNDNDKYHVAYSWVVVKLRDINDNKPQFERPNIEVSVYENADVGKT  1908

Query  127   VMILNATDADEPNHLNSKIAFKIVSQEPAGTPMFLLSRNTGEVRTLTNSLDREQASSYRL  186
             +    ATD D+     SK+++ I   + +       S N     T+  +LDRE    +++
Sbjct  1909  LETFKATDPDQGG--KSKVSYAI---DRSSDRQRQFSINQEGTVTIQRNLDREVTPRHQV  1963

Query  187   VVSGADKDGEGLSTQCECNIKVKDVNDNFPMFRDSQYSARIEENILSSELLRFQVTDLDE  246
              +   D      +      + V+D+NDN P F    Y   + E++   +++    TD D+
Sbjct  1964  KILAIDDGIPPKTATATLTVIVQDINDNPPKFL-KDYRPVLPEHVPPRKVVEILATDDDD  2022

Query  247   EYTDNWLAVYFFTSGNEGNW----FEIQTDPRTNEG----ILKVVKALDYEQLQSVKLSI  298
                 N     F       +     F+++ D +   G    I+  +++ D EQ +   + I
Sbjct  2023  RSKSNGPPFQFRLDPGADDIIRASFKVEQDQKGANGDGMAIVSSLRSFDREQQKEYLIPI  2082

Query  299   AVKNKA  304
              +K+
Sbjct  2083  VIKDHG  2088

 Score = 51.6 bits (122),  Expect = 6e-05, Method: Compositional matrix adjust.
 Identities = 96/398 (24%), Positives = 163/398 (41%), Gaps = 54/398 (14%)

Query  106   PVFSQQIFMGEIEENSASNSLVMILNATDADEPNHLNSKIAFKIVSQ-EPAGTPMFLLSR  164
             P F    +  EI EN   +S ++ + A      +  + +I + + +Q + AGT  F +
Sbjct  677   PQFYMPSYEAEIPENQKKDSDIISVKAK-----SFADREIRYTLKAQGQGAGT--FNIGP  729

Query  165   NTGEVRTLTNSLDRE---QASSYRLVVSGADKDGEGLSTQCECNIKVKDVNDNFPMFRDS  221
              +G V+ L   LD E   Q   Y LVV+ A +D  G ST  E  I+V DVNDN P F
Sbjct  730   TSGIVK-LAKELDFEDLRQPHVYSLVVT-ATEDSGGFSTSVELTIRVTDVNDNAPKFELP  787

Query  222   QYSAR-IEENI-LSSELLRFQVTDLDEEYTDNWLAVYFFTSGNEGNWFEIQTDPRTNEGI  279
              Y A  ++E+I + + +L+ +  D D         + +  S +    F + ++     GI
Sbjct  788   DYQAHNVDEDIQIGTSILKVKAMDADSGAN---AEIEYLVSDDH---FAVDSN-----GI  836

Query  280   LKVVKALDYEQLQS-VKLSIAVKNKAEFHQSVISRYRVQSTPVTIQVINVREGIAFRPAS  338
```

```
             +   K LD +   +  +  +  K+K E  ++  +  RV +           E   F
Sbjct  837   IVNNKQLDADNNNAYYEFVVTAKDKGEPAKTGTATVRVYTKNKN------DEEPKFSQQV  890

Query  339   KTFTVQKGISSKKLVDYILGTYQAIDEDTNKAASNVK--YVMGRNDGGYLMIDSKTAEIK  396
              T  V +      LV  ++         ++K   NV+  +V G    G  +I+  T  I+
Sbjct  891   YTPNVDENAGPNTLVTTVVA--------SDKDGDNVRFGFVGGGTSSGQFVIEEITGVIR  942

Query  397   FVK---NMNRDSTFI-VNKTITAEVLAIDEYTGKTSTGTVYVRVPDFNDN------CPTA  446
                   +++RD   + V            + T  TST  V V + D NDN      C T
Sbjct  943   LHSKAISLDRDKYELNVTAMDDGSCCVNGDATIHTSTAVVVVFITDVNDNKPVFKDCGTY  1002

Query  447   VLE-KDAVCSSSPSVVVSARTLNNRYTGPYTFALEDQP  483
               + ++   + SP + V A   +    G   +++  QP
Sbjct  1003  YPKVEEGAPNGSPVIKVQATDEDKGVNGQVKYSIVQQP  1040

 Score = 44.3 bits (103),  Expect = 0.009, Method: Compositional matrix adjust.
 Identities = 48/199 (24%), Positives = 73/199 (37%), Gaps = 19/199 (10%)

Query  27    DYQATQKITYRISGVGIDQPPFGIFVVDKNTGDINITAIVDREETPSFLITCRALNAQGL  86
             D     K++Y I      Q  F I       G + I   +DRE TP   +   A++
Sbjct  1918  DQGGKSKVSYAIDRSSDRQRQFSI----NQEGTVTIQRNLDREVTPRHQVKILAIDDGIP  1973

Query  87    DVEKPLILTVKILDINDNPPVFSQQIFMGEIEENSASNSLVMILNATDADEPNHLNSKIA  146
                    LTV + DINDNPP F +  +   + E+     +V IL   D D
Sbjct  1974  PKTATATLTVIVQDINDNPPKFLKD-YRPVLPEHVPPRKVVEILATDDDDRSKSNGPPFQ  2032

Query  147   FKIVSQEPAGTPMFLLSRNTGEVRTLTN-----------SLDREQASSYRLVVSGADKDG  195
             F++   +P    +   S    + +   N           S DREQ   Y + +   D
Sbjct  2033  FRL---DPGADDIIRASFKVEQDQKGANGDGMAIVSSLRSFDREQQKEYLIPIVIKDHGN  2089

Query  196   EGLSTQCECNIKVKDVNDN  214
               ++      + + DVNDN
Sbjct  2090  PAMTGTSTLTVVIGDVNDN  2108

 Score = 40.8 bits (94),  Expect = 0.12, Method: Compositional matrix adjust.
 Identities = 54/235 (23%), Positives = 99/235 (42%), Gaps = 22/235 (9%)

Query  4    KFAKPCREGE--DNSKRNP-IAKITSDYQATQKITYRISGVGIDQPPFGIFVVDKNTGDI  60
            +F  P  E E  +N K++  I  + +   A ++I Y +   G      G F +   +G +
Sbjct  678  QFYMPSYEAEIPENQKKDSDIISVKAKSFADREIRYTLKAQGQGA---GTFNIGPTSGIV  734

Query  61   NITAIVDREE--TPSF--LITCRALNAQGLDVEKPLILTVKILDINDNPPVFSQQIFMGE  116
             +   +D E+   P    L+     ++ G      + LT+++ D+NDN P F    +
Sbjct  735  KLAKELDFEDLRQPHVYSLVVTATEDSGGFSTS--VELTIRVTDVNDNAPKFELPDYQAH  792

Query  117  -IEENSASNSLVMILNATDADEPNHLNSKIAFKIVSQEPAGTPMFLLSRNTGEVRTLTNS  175
             ++E+    + ++ + A DAD  +  N++I + +VS +      F +  N   V
Sbjct  793  NVDEDIQIGTSILKVKAMDAD--SGANAEIEY-LVSDD-----HFAVDSNGIIVNNKQLD  844

Query  176  LDREQASSYRLVVSGADKDGEGLSTQCECNIKVKDVNDNFPMFRDSQYSARIEEN  230
             D   A  Y  VV+  DK     +      +  K+ ND  P F    Y+  ++EN
Sbjct  845  ADNNNA-YYEFVVTAKDKGEPAKTGTATVRVYTKNKNDEEPKFSQQVYTPNVDEN  898

 Score = 39.3 bits (90),  Expect = 0.33, Method: Compositional matrix adjust.
 Identities = 55/225 (24%), Positives = 88/225 (39%), Gaps = 24/225 (11%)

Query  75   LITCRALNAQGLDVEKPLILTVKILDINDNPPVFSQQIF--MGEIEENSASNSLVMILNA  132
            +IT +  NA GL       + + + D+ND PP F  +       ++ N+  N+ V  L A
Sbjct  539  IITNQGHNA-GLKYTDNQRVIIHVKDVNDEPPYFINRPLPMQAVVQLNAPPNTPVFTLQA  597

Query  133  TDADEPNHLNSKIAFKIVSQEPAGTPMFLLSRNTGEVRTLTNSLDREQASSYRLVVSGAD  192
             D D     +  I + IV     G   F +   +G VRT    L  +    Y L V   D
Sbjct  598  RDPD----TDHNIHYFIVRDRTGG--RFEVDERSGVVRTRGTDL-FQLDMEYVLYVKAED  650

Query  193  KDGE----GLSTQCECNIKVKDVNDNFPMFRDSQYSARIEEN-ILSSELLRFQVTDLDEE  247
            ++G+       +  E  + +       P F    Y A I EN    S+++  +     +
Sbjct  651  QNGKVDDRRFQSTPEERLSIVG-GKRAPQFYMPSYEAEIPENQKKDSDIISVKA----KS  705
```

```
Query  248  YTDNWLAVYFFTSGNEGNWFEIQTDPRTNEGILKVVKALDYEQLQ  292
            + D  +       G     F I        GI+K+ K LD+E L+
Sbjct  706  FADREIRYTLKAQGQGAGTFNIGP----TSGIVKLAKELDFEDLR  746

>AMEM21_003341.P6419 | transcript=AMEM21_003341.R6419 | gene=AMEM21_003341
| organism=Anopheles_merus_MAF_2021 | gene_product=neural-cadherin,
transcript variant X1 | transcript_product=neural-cadherin,
transcript variant X17 | location=CM029766:40289848-40498951(+)
| protein_length=3099 | sequence_SO=chromosome
| SO=protein_coding_gene | is_pseudo=false
Length=3099

 Score = 147 bits (370),  Expect = 3e-35, Method: Compositional matrix adjust.
 Identities = 128/446 (29%), Positives = 217/446 (49%), Gaps = 36/446 (8%)

Query  20    PIAKITS---DYQATQKITYRISGVGID--QPPFGIFVVDKNTGDINITAIVDREET---  71
             PI ++T+   D    Q I Y ++G GID   P    F +++ TG+I +   +DR++
Sbjct  1539  PIMQVTATDGDKDRPQNIVYFLTGQGIDPDNPANSKFDINRTTGEIFVLKPLDRDQPNGR  1598

Query  72    PSFLITCRALNAQGLDVEKPLILTVKILDINDNPPVFSQQIFMGEIEENSASNSLVMILN  131
             P +  T  A +  G  +     + V + DINDN P+F Q ++ G + EN  +  +VM +
Sbjct  1599  PQWRFTVFAQDEGGEGLVGYADVQVNLKDINDNAPIFPQGVYFGNVTENGTAGMVVMTMT  1658

Query  132   ATDADEPNH-LNSKIAFKI---VSQEPAGTPMFLLSRNTGEVRTLTNSLDREQASSYRLV  187
             A D D+PN   N+K+ + I   V +E  G+P+F +  +TG ++T    LDRE+   Y +
Sbjct  1659  AVDYDDPNEGTNAKLIYSIEKNVIEEETGSPIFEIEADTGVIKTAVCCLDRERTPDYSIQ  1718

Query  188   VSGADKDGEGLSTQCECNIKVKDVNDNFPMFRDSQYSARIEENILSS--ELLRFQVTDLD  245
             V     DG GL      +I+VKD+ND  P F   ++   ++E   ++  E+    VT  D
Sbjct  1719  VVAM--DGGGLKGTGTASIRVKDINDMPPQFTKDEWFTEVDETDGTNLPEMPILTVTVHD  1776

Query  246   EEYTDNWLAVYFFTSGNEGNWFEIQTDPRTNEGI--LKVVKALDYE---QLQSVKLSIAV  300
             E+ T+ +       SG   + F +    R N+G   LK+V+ LDYE   Q    +  I V
Sbjct  1777  EDETNKFQYKVIDNSGYGADKFTMV---RNNDGTGSLKIVQPLDYEDPLQSNGFRFRIQV  1833

Query  301   KNKAEFHQSVISRYRVQSTPVTIQVINVREGIAFRPASKTFTVQKGISSKKLVDYILGTY  360
              +K E + +   +Y V  + V +++ ++ +    +P  +   ++  +     V   L T+
Sbjct  1834  NDKGEDNDN--DKYHVAYSWVVVKLRDINDN---KPQFERPNIEVSVYENADVGKTLETF  1888

Query  361   QAIDEDTNKAASNVKYVMGR-NDGGYLMIDSKTAEIKFVKNMNRDSTFIVNKTITAEVLA  419
             +A D D     S V Y + R +D       ++   +   +N++R+    V      ++LA
Sbjct  1889  KATDPDQG-GKSKVSYAIDRSSDRQRQFSINQEGTVTIQRNLDRE----VTPRHQVKILA  1943

Query  420   IDE-YTGKTSTGTVYVRVPDFNDNCP  444
             ID+    KT+T T+ V V D NDN P
Sbjct  1944  IDDGIPPKTATATLTVIVQDINDNPP  1969

 Score = 78.6 bits (192),  Expect = 2e-13, Method: Compositional matrix adjust.
 Identities = 106/424 (25%), Positives = 180/424 (42%), Gaps = 51/424 (12%)

Query  53    VDKNTGDINITAIVDREETPSFLITCRALNAQGLDVEKPLILTVKILDINDNPPVFSQ-Q  111
             +D++  ++N+TA+ D         +C  +N          ++ V I D+NDN PVF
Sbjct  928   LDRDKYELNVTAMDDG--------SC-CVNGDATIHTSTAVVVVFITDVNDNKPVFKDCG  978

Query  112   IFMGEIEENSASNSLVMILNATDADEPNHLNSKIAFKIVSQEPAGTPMFLLSRNTGEVRT  171
              +  ++EE + + S V+ + ATD D+   +N ++ + IV Q       F +   TGEV T
Sbjct  979   TYYPKVEEGAPNGSPVIKVQATDEDK--GVNGQVKYSIVQQPNQKGTKFTVDEETGEVST  1036

Query  172   LTNSLDRE--QASSYRLVVSGADKDGEGLSTQCECNIKVKDVNDNFPMFRDSQYSARIEE  229
                  DRE        + V   D+    L   C   +++ DVNDN P+F   +Y   +++
Sbjct  1037  -NKVFDREGDDGKFVSVTVKATDQGEPSLEGVCSFTVEITDVNDNPPLFDRQKYVENVKQ  1095

Query  230   NI-LSSELLRFQVTDLDEEYTDNWLAVYFFT---SGNEGNWFEIQTDPRTNEGILKVVKA  285
             +  + + +LR  V+  DE+  +N   VY  +   S N+  +FEIQ  P +   +LK  K
Sbjct  1096  DASIGTNILR--VSASDEDADNNGAIVYTLSAPHSPNDLEYFEIQ--PESGWIVLK--KP  1149

Query  286   LDYEQLQSVKLSIAVKNK------AEFHQSVISRYRVQSTPVTIQV----INVREGIAFR  335
             LD E+    +L +   ++      A+    +    R    P+  +     I++RE +
Sbjct  1150  LDRERY---RLKVYASDRGDPPSTADVDVELDVVDRNNKPPIWDRTIYGPIHIRENVTVG  1206
```

```
Query  336   PASKTFTVQKGISSKKLVDYILGTYQAIDEDTNKAASNVKYVMGRNDGGYLMIDSKTAEI  395
                 +     GI     V Y L         TNK    + Y+  R D G        A+I
Sbjct  1207  TVVTSVKASSGIEGNPTVFYRL--MPGSTAQTNKL--HTFYLQQRPDNG-----DTWADI  1257

Query  396   KFVKNMNRDSTFIVNKTITAEVLAIDEYTGKTSTGTVYYRVPDFNDNCPTAV-LEKDAVC  454
             K    ++ ++    N T+  E     +     S  TVY+++ D ND  P     E++ V
Sbjct  1258  KVNHPLDFETIKEYNLTVRVENNGAQQL---ASEATVYIQLEDVNDEIPLFTEREQETVL  1314

Query  455   SSSP  458
                P
Sbjct  1315  EGEP  1318

 Score = 75.9 bits (185),  Expect = 2e-12, Method: Compositional matrix adjust.
 Identities = 103/418 (25%), Positives = 174/418 (42%), Gaps = 50/418 (12%)

Query  50    IFVVDKNTGDINITAIVDREETPSFLITCRAL----------NAQGLDVEKPLILTVKIL  99
              F ++  +G+I    + DRE+  ++ +   A           N Q   V K   + + I
Sbjct  1355  FFEINLQSGEIFTKVVFDREKQGAYALEVEARDGAPSARPNSNQQPNSVTK--FIRIGIA  1412

Query  100   DINDNPPVFSQQIFMGEIEENSASNSLVMILNATDADEPNHLNSKIAFKIVSQEPAGTPM  159
             D NDNPP F + ++  E++EN      V+ + A D DE    +S+I ++I S    G
Sbjct  1413  DKNDNPPYFDKALYEAEVDENEDIQHTVLTVTAKDHDE----SSRIRYEITSGNIGGA--  1466

Query  160   FLLSRNTGEVRTLTNSLDREQASSYRLVVSGADKDGEGLSTQCECNIKVKDVNDNFPMFR  219
             F +   TG +  +  +LD E    Y L +  +D   E  +      I VKDVND  P F
Sbjct  1467  FAVKNMTGAI-YVAGALDYETRKRYYLTLVASDSLNENETIIV---INVKDVNDMPPSFP  1522

Query  220   DSQYSARIEENI-LSSELLRFQVTDLDEEYTDNWLAVYFFTSG--NEGNWFEIQTDPRTN  276
                Y   ++E + +   +++   TD D++   N   VYF T    +  N    + D
Sbjct  1523  QQVYERTMDEELAVPFPIMQVTATDGDKDRPQN--IVYFLTGQGIDPDNPANSKFDINRT  1580

Query  277   EGILKVVKALDYEQLQS-VKLSIAVKNKAEFHQSVISRYRVQSTPVTIQVINVREGIAFR  335
              G + V+K LD +Q     +    V  + E  + ++    VQ     + + ++ +
Sbjct  1581  TGEIFVLKPLDRDQPNGRPQWRFTVFAQDEGGEGLVGYADVQ-----VNLKDINDNAPIF  1635

Query  336   PASKTFTVQKGISSKKLVDYILGTYQAIDEDTNKAASNVK--YVMGRN------DGGYLM  387
             P    F     ++       ++ T  A+D D     +N K  Y + +N
Sbjct  1636  PQGVYF---GNVTENGTAGMVVMTMTAVDYDDPNEGTNAKLIYSIEKNVIEEETGSPIFE  1692

Query  388   IDSKTAEIK-FVKNMNRDSTFIVNKTITAEVLAIDEYTGKTSTGTVYYRVPDFNDNCP  444
             I++ T  IK  V  ++R+ T       + +V+A+D   G   TGT  +RV D ND  P
Sbjct  1693  IEADTGVIKTAVCCLDRERT----PDYSIQVVAMDG-GGLKGTGTASIRVKDINDMPP  1745

 Score = 65.5 bits (158),  Expect = 3e-09, Method: Compositional matrix adjust.
 Identities = 85/359 (24%), Positives = 153/359 (43%), Gaps = 32/359 (9%)

Query  51    FVVDKNTGDINITAIVDRE-ETPSFL-ITCRALNAQGLDVEKPLILTVKILDINDNPPVF  108
             F VD+ TG+++   + DRE +   F+ +T +A +     +E     TV+I D+NDNPP+F
Sbjct  1025  FTVDEETGEVSTNKVFDREGDDGKFVSVTVKATDQGEPSLEGVCSFTVEITDVNDNPPLF  1084

Query  109   SQQIFMGEIEENSASNSLVMILNATDADEPNHLNSKIAFKIVS-QEPAGTPMFLLSRNTG  167
              +Q ++  ++++++  + ++ ++A+D D  N  N  I + + +   P     F +   +G
Sbjct  1085  DRQKYVENVKQDASIGTNILRVSASDEDADN--NGAIVYTLSAPHSPNDLEYFEIQPESG  1142

Query  168   EVRTLTNSLDREQASSYRLVVSGADKDGEGLSTQCECNIKVKDVNDNFPMFRDSQYSARI  227
              +  L   LDRE+   YRL V  +D+ G+  ST            +N P   D      I
Sbjct  1143  WI-VLKKPLDRER---YRLKVYASDR-GDPPSTADVDVELDVVDRNNKPPIWDRTIYGPI  1197

Query  228   EENILSSELLRFQVTDLDEEYTDNWLAVYFFTSG-----NEGNWFEIQTDPRTNE--GIL  280
                   +                N    Y    G     N+ + F +Q  P   +    +
Sbjct  1198  HIRENVTVGTVVTSVKASSGIEGNPTVFYRLMPGSTAQTNKLHTFYLQQRPDNGDTWADI  1257

Query  281   KVVKALDYEQLQSVKLSIAVKNKAEFHQSVISRYRVQSTPVTIQVINVREGIAFRPASKT  340
             KV   LD+E ++   L++ V+N         ++       V IQ+ +V + I      +
Sbjct  1258  KVNHPLDFETIKEYNLTVRVENNG-------AQQLASEATVYIQLEDVNDEIPLFTEREQ  1310

Query  341   FTVQKGISSKKLVDYILGTYQAIDEDTNKAASNVKYVM---GRNDG-GYLMIDSKTAEI  395
              TV +G      V  +     AID+D     + V Y +    RN+G  +  I+ ++ EI
```

```
Sbjct  1311  ETVLEGEPIGTKVTQV----NAIDKDGTFPNNQVYYYIVDSPRNEGKDFFEINLQSGEI  1365

 Score = 55.8 bits (133),  Expect = 3e-06, Method: Compositional matrix adjust.
 Identities = 49/158 (31%), Positives = 78/158 (49%), Gaps = 24/158 (15%)

Query  102   NDNPPVFSQQIFMGEIEENSASNSLVMILNATDADEPNHLNSKIAFKIVSQEPAGTP--M  159
             ND  P FSQQ++   ++EN+  N+LV  + A+D D  N     + F  V     GT
Sbjct  858   NDEEPKFSQQVYTPNVDENAGPNTLVTTVVASDKDGDN-----VRFGFVG---GGTSSGQ  909

Query  160   FLLSRNTGEVRTLTN--SLDREQASSYRLVVSGADK-------DGEGLSTQCECNIKVKD  210
             F++    TG +R  +   SLDR++   Y L V+  D         D    ++     + + D
Sbjct  910   FVIEEITGVIRLHSKAISLDRDK---YELNVTAMDDGSCCVNGDATIHTSTAVVVVFITD  966

Query  211   VNDNFPMFRD-SQYSARIEENILS-SELLRFQVTDLDE  246
             VNDN P+F+D   Y  ++EE   + S +++ Q TD D+
Sbjct  967   VNDNKPVFKDCGTYYPKVEEGAPNGSPVIKVQATDEDK  1004

 Score = 53.9 bits (128),  Expect = 1e-05, Method: Compositional matrix adjust.
 Identities = 70/306 (23%), Positives = 126/306 (41%), Gaps = 30/306 (10%)

Query  20    PIAKIT-SDYQATQKITYRI---SGVGIDQPPFGIFVVDKNTGDINITAIVDRE---ETP  72
             PI  +T  D   T K  Y++   SG G D+  F +   +  TG + I   +D E   ++
Sbjct  1768  PILTVTVHDEDETNKFQYKVIDNSGYGADK--FTMVRNNDGTGSLKIVQPLDYEDPLQSN  1825

Query  73    SFLITCRALNAQGLDVEKPLI------LTVKILDINDNPPVFSQQIFMGEIEENSASNSL  126
              F    + +N +G D +           + VK+ DINDN P F +       + EN+
Sbjct  1826  GFRFRIQ-VNDKGEDNDNDKYHVAYSWVVVKLRDINDNKPQFERPNIEVSVYENADVGKT  1884

Query  127   VMILNATDADEPNHLNSKIAFKIVSQEPAGTPMFLLSRNTGEVRTLTNSLDREQASSYRL  186
             +    ATD D+      SK+++ I   + +        S N      T+  +LDRE    +++
Sbjct  1885  LETFKATDPDQGG--KSKVSYAI---DRSSDRQRQFSINQEGTVTIQRNLDREVTPRHQV  1939

Query  187   VVSGADKDGEGLSTQCECNIKVKDVNDNFPMFRDSQYSARIEENILSSELLRFQVTDLDE  246
              +   D      +       + V+D+NDN P F    Y   + E++   +++    TD D+
Sbjct  1940  KILAIDDGIPPKTATATLTVIVQDINDNPPKFL-KDYRPVLPEHVPPRKVVEILATDDDD  1998

Query  247   EYTDNWLAVYFFTSGNEGNW----FEIQTDPRTNEG----ILKVVKALDYEQLQSVKLSI  298
                 N     F       +     F+++ D +   G    I+  +++ D EQ +   + I
Sbjct  1999  RSKSNGPPFQFRLDPGADDIIRASFKVEQDQKGANGDGMAIVSSLRSFDREQQKEYLIPI  2058

Query  299   AVKNKA  304
              +K+
Sbjct  2059  VIKDHG  2064

 Score = 52.0 bits (123),  Expect = 4e-05, Method: Compositional matrix adjust.
 Identities = 97/398 (24%), Positives = 163/398 (41%), Gaps = 54/398 (14%)

Query  106   PVFSQQIFMGEIEENSASNSLVMILNATDADEPNHLNSKIAFKIVSQ-EPAGTPMFLLSR  164
             P F    +  EI EN   +S ++ + A       +  + +I + + +Q + AGT  F +
Sbjct  655   PQFYMPSYEAEIPENQKKDSDIISVKAK-----SFADREIRYTLKAQGQGAGT--FNIGP  707

Query  165   NTGEVRTLTNSLDRE---QASSYRLVVSGADKDGEGLSTQCECNIKVKDVNDNFPMFRDS  221
              +G V+ L   LD E   Q   Y LVV+ A +D  G ST  E  I+V DVNDN P F
Sbjct  708   TSGIVK-LAKELDFEDLRQPHVYSLVVT-ATEDSGGFSTSVELTIRVTDVNDNAPKFELP  765

Query  222   QYSAR-IEENI-LSSELLRFQVTDLDEEYTDNWLAVYFFTSGNEGNWFEIQTDPRTNEGI  279
              Y A  ++E+I L + +L+ +  D D          + +  S +    F + ++     GI
Sbjct  766   DYQAHNVDEDIPLGTSILKVKAMDSDSGAN---AEIEYLVSDDH---FAVDSN-----GI  814

Query  280   LKVVKALDYEQLQS-VKLSIAVKNKAEFHQSVISRYRVQSTPVTIQVINVREGIAFRPAS  338
             +   K LD +    +  +  K+K E  ++  +  RV +               E   F
Sbjct  815   IVNNKQLDADNNNAYYEFVVTAKDKGEPAKTGTATVRVYTKNKN------DEEPKFSQQV  868

Query  339   KTFTVQKGISSKKLVDYILGTYQAIDEDTNKAASNVK--YVMGRNDGGYLMIDSKTAEIK  396
              T  V +       LV  ++           ++K   NV+  +V G    G  +I+  T  I+
Sbjct  869   YTPNVDENAGPNTLVTTVVA--------SDKDGDNVRFGFVGGGTSSGQFVIEEITGVIR  920

Query  397   FVK---NMNRDSTFI-VNKTITAEVLAIDEYTGKTSTGTVYVRVPDFNDN------CPTA  446
```

```
                   +++RD   + V            + T  TST  V V + D NDN      C T
Sbjct  921   LHSKAISLDRDKYELNVTAMDDGSCCVNGDATIHTSTAVVVVFITDVNDNKPVFKDCGTY  980

Query  447   VLE-KDAVCSSSPSVVVSARTLNNRYTGPYTFALEDQP  483
               + ++   + SP + V A   +    G   +++  QP
Sbjct  981   YPKVEEGAPNGSPVIKVQATDEDKGVNGQVKYSIVQQP  1018

 Score = 44.3 bits (103),  Expect = 0.009, Method: Compositional matrix adjust.
 Identities = 48/199 (24%), Positives = 73/199 (37%), Gaps = 19/199 (10%)

Query  27    DYQATQKITYRISGVGIDQPPFGIFVVDKNTGDINITAIVDREETPSFLITCRALNAQGL  86
             D     K++Y I      Q  F I       G + I   +DRE TP   +   A++
Sbjct  1894  DQGGKSKVSYAIDRSSDRQRQFSI----NQEGTVTIQRNLDREVTPRHQVKILAIDDGIP  1949

Query  87    DVEKPLILTVKILDINDNPPVFSQQIFMGEIEENSASNSLVMILNATDADEPNHLNSKIA  146
                    LTV + DINDNPP F +  +   + E+     +V IL   D D
Sbjct  1950  PKTATATLTVIVQDINDNPPKFLKD-YRPVLPEHVPPRKVVEILATDDDDRSKSNGPPFQ  2008

Query  147   FKIVSQEPAGTPMFLLSRNTGEVRTLTN-----------SLDREQASSYRLVVSGADKDG  195
             F++   +P    +   S    + +   N           S DREQ   Y + +   D
Sbjct  2009  FRL---DPGADDIIRASFKVEQDQKGANGDGMAIVSSLRSFDREQQKEYLIPIVIKDHGN  2065

Query  196   EGLSTQCECNIKVKDVNDN  214
               ++      + + DVNDN
Sbjct  2066  PAMTGTSTLTVVIGDVNDN  2084

 Score = 39.7 bits (91),  Expect = 0.27, Method: Compositional matrix adjust.
 Identities = 89/389 (23%), Positives = 149/389 (38%), Gaps = 62/389 (16%)

Query  75   LITCRALNAQGLDVEKPLILTVKILDINDNPPVFSQQIF--MGEIEENSASNSLVMILNA  132
            +IT +  NA GL       + + + D+ND PP F  +       ++ N+  N+ V  L A
Sbjct  517  IITNQGHNA-GLKYTDNQRVIIHVKDVNDEPPYFINRPLPMQAVVQLNAPPNTPVFTLQA  575

Query  133  TDADEPNHLNSKIAFKIVSQEPAGTPMFLLSRNTGEVRTLTNSLDREQASSYRLVVSGAD  192
             D D     +  I + IV     G   F +   +G VRT    L  +    Y L V   D
Sbjct  576  RDPD----TDHNIHYFIVRDRTGG--RFEVDERSGVVRTRGTDL-FQLDMEYVLYVKAED  628

Query  193  KDGE----GLSTQCECNIKVKDVNDNFPMFRDSQYSARIEEN-ILSSELLRFQVTDLDEE  247
            ++G+       +  E  + +       P F    Y A I EN    S+++  +     +
Sbjct  629  QNGKVDDRRFQSTPEERLSIVG-GKRAPQFYMPSYEAEIPENQKKDSDIISVKA----KS  683

Query  248  YTDNWLAVYFFTSGNEGNWFEIQTDPRTNEGILKVVKALDYEQLQSVKLS----IAVKNK  303
            + D  +       G     F I        GI+K+ K LD+E L+   +      A ++
Sbjct  684  FADREIRYTLKAQGQGAGTFNIGP----TSGIVKLAKELDFEDLRQPHVYSLVVTATEDS  739

Query  304  AEFHQSVISRYRVQSTPVTIQVINVREGIAFRPASKTFTVQKGISSKKLVDYILGT----  359
              F  SV          +TI+V +V +       +  F +    +     D  LGT
Sbjct  740  GGFSTSV---------ELTIRVTDVNDN------APKFELPDYQAHNVDEDIPLGTSILK  784

Query  360  YQAIDEDTNKAASNVKYVMGRN----DGGYLMIDSKTAEIKFVKNMNRDSTFIVNKTITA  415
             +A+D D+  A + ++Y++  +    D   +++++K  +     N N    F+V
Sbjct  785  VKAMDSDSG-ANAEIEYLVSDDHFAVDSNGIIVNNKQLD---ADNNNAYYEFVVTAKDKG  840

Query  416  EVLAIDEYTGKTSTGTVYVRVPDFNDNCP  444
            E         KT T TV V   + ND  P
Sbjct  841  E-------PAKTGTATVRVYTKNKNDEEP  862

 Score = 39.3 bits (90),  Expect = 0.30, Method: Compositional matrix adjust.
 Identities = 54/242 (22%), Positives = 102/242 (42%), Gaps = 22/242 (9%)

Query  4    KFAKPCREGE--DNSKRNP-IAKITSDYQATQKITYRISGVGIDQPPFGIFVVDKNTGDI  60
            +F  P  E E  +N K++  I  + +   A ++I Y +   G      G F +   +G +
Sbjct  656  QFYMPSYEAEIPENQKKDSDIISVKAKSFADREIRYTLKAQGQGA---GTFNIGPTSGIV  712

Query  61   NITAIVDREE--TPSF--LITCRALNAQGLDVEKPLILTVKILDINDNPPVFSQQIFMGE  116
             +   +D E+   P    L+     ++ G      + LT+++ D+NDN P F    +
Sbjct  713  KLAKELDFEDLRQPHVYSLVVTATEDSGGFSTS--VELTIRVTDVNDNAPKFELPDYQAH  770
```

```
Query  117  -IEENSASNSLVMILNATDADEPNHLNSKIAFKIVSQEPAGTPMFLLSRNTGEVRTLTNS  175
             ++E+    + ++ + A D+D  +  N++I + +VS +      F +  N   V
Sbjct  771  NVDEDIPLGTSILKVKAMDSD--SGANAEIEY-LVSDD-----HFAVDSNGIIVNNKQLD  822

Query  176  LDREQASSYRLVVSGADKDGEGLSTQCECNIKVKDVNDNFPMFRDSQYSARIEENILSSE  235
             D   A  Y  VV+  DK      +       +  K+ ND  P F    Y+  ++EN   +
Sbjct  823  ADNNNA-YYEFVVTAKDKGEPAKTGTATVRVYTKNKNDEEPKFSQQVYTPNVDENAGPNT  881

Query  236  LL  237
            L+
Sbjct  882  LV  883

>ACMO_007424.P13494 | transcript=ACMO_007424.R13494 | gene=ACMO_007424
| organism=Anopheles_coluzzii_MOPTI | gene_product=neural-cadherin,
transcript variant X4 | transcript_product=neural-cadherin,
transcript variant X8 | location=CM029349:40739311-40945968(+)
| protein_length=3102 | sequence_SO=chromosome
| SO=protein_coding_gene | is_pseudo=false
Length=3102

 Score = 147 bits (370),  Expect = 3e-35, Method: Compositional matrix adjust.
 Identities = 128/446 (29%), Positives = 217/446 (49%), Gaps = 36/446 (8%)

Query  20    PIAKITS---DYQATQKITYRISGVGID--QPPFGIFVVDKNTGDINITAIVDREET---  71
             PI ++T+    D    Q I Y ++G GID    P    F +++ TG+I +    +DR++
Sbjct  1543  PIMQVTATDGDKDRPQNIVYFLTGQGIDPDNPANSKFDINRTTGEIFVLKPLDRDQPNGR  1602

Query  72    PSFLITCRALNAQGLDVEKPLILTVKILDINDNPPVFSQQIFMGEIEENSASNSLVMILN  131
             P +   T  A +  G  +     + V + DINDN P+F Q ++ G + EN  +  +VM +
Sbjct  1603  PQWRFTVFAQDEGGEGLVGYADVQVNLKDINDNAPIFPQGVYFGNVTENGTAGMVVMTMT  1662

Query  132   ATDADEPNH-LNSKIAFKI---VSQEPAGTPMFLLSRNTGEVRTLTNSLDREQASSYRLV  187
             A D D+PN   N+K+ + I   V +E  G+P+F +  +TG ++T    LDRE+   Y +
Sbjct  1663  AVDYDDPNEGTNAKLIYSIEKNVIEEETGSPIFEIEADTGVIKTAVCCLDRERTPDYSIQ  1722

Query  188   VSGADKDGEGLSTQCECNIKVKDVNDNFPMFRDSQYSARIEENILSS--ELLRFQVTDLD  245
             V      DG GL       +I+VKD+ND  P F   ++   ++E   ++  E+    VT  D
Sbjct  1723  VVAM--DGGGLKGTGTASIRVKDINDMPPQFTKDEWFTEVDETDGTNLPEMPILTVTVHD  1780

Query  246   EEYTDNWLAVYFFTSGNEGNWFEIQTDPRTNEGI--LKVVKALDYE---QLQSVKLSIAV  300
             E+ T+ +        SG    + F +    R N+G   LK+V+ LDYE   Q     +  I V
Sbjct  1781  EDETNKFQYKVIDNSGYGADKFTMV---RNNDGTGSLKIVQPLDYEDPLQSNGFRFRIQV  1837

Query  301   KNKAEFHQSVISRYRVQSTPVTIQVINVREGIAFRPASKTFTVQKGISSKKLVDYILGTY  360
              +K E + +   +Y V  + V +++ ++ +    +P  +   ++  +     V   L T+
Sbjct  1838  NDKGEDNDN--DKYHVAYSWVVVKLRDINDN---KPQFERPNIEVSVYENADVGKTLETF  1892

Query  361   QAIDEDTNKAASNVKYVMGR-NDGGYLMIDSKTAEIKFVKNMNRDSTFIVNKTITAEVLA  419
             +A D D     S V Y + R +D        ++   +   +N++R+    V       ++LA
Sbjct  1893  KATDPDQG-GKSKVSYAIDRSSDRQRQFSINQEGTVTIQRNLDRE----VTPRHQVKILA  1947

Query  420   IDE-YTGKTSTGTVYVRVPDFNDNCP  444
             ID+    KT+T T+ V V D NDN P
Sbjct  1948  IDDGIPPKTATATLTVIVQDINDNPP  1973

 Score = 76.3 bits (186),  Expect = 1e-12, Method: Compositional matrix adjust.
 Identities = 103/418 (25%), Positives = 174/418 (42%), Gaps = 50/418 (12%)

Query  50    IFVVDKNTGDINITAIVDREETPSFLITCRAL----------NAQGLDVEKPLILTVKIL  99
              F ++  +G+I    + DRE+  ++ +   A           N Q   V K   + + I
Sbjct  1359  FFEINLQSGEIFTKVVFDREKQGAYALEVEARDGAPSARPNSNQQPNSVTK--FIRIGIA  1416

Query  100   DINDNPPVFSQQIFMGEIEENSASNSLVMILNATDADEPNHLNSKIAFKIVSQEPAGTPM  159
             D NDNPP F + ++  E++EN       V+ + A D DE    +S+I ++I S    G
Sbjct  1417  DKNDNPPYFDKALYEAEVDENEDIQHTVLTVTAKDHDE----SSRIRYEITSGNIGGA--  1470

Query  160   FLLSRNTGEVRTLTNSLDREQASSYRLVVSGADKDGEGLSTQCECNIKVKDVNDNFPMFR  219
             F +   TG +  +  +LD E    Y L +  +D   E  +       I VKDVND  P F
Sbjct  1471  FAVKNMTGAI-YVAGALDYETRKRYYLTLVASDSLNENETIIV---INVKDVNDMPPSFP  1526
```

```
Query  220   DSQYSARIEENI-LSSELLRFQVTDLDEEYTDNWLAVYFFTSG--NEGNWFEIQTDPRTN  276
                Y   ++E + +   +++   TD D++   N   VYF T    +  N    + D
Sbjct  1527  QQVYERTMDEELAVPFPIMQVTATDGDKDRPQN--IVYFLTGQGIDPDNPANSKFDINRT  1584

Query  277   EGILKVVKALDYEQLQS-VKLSIAVKNKAEFHQSVISRYRVQSTPVTIQVINVREGIAFR  335
              G + V+K LD +Q     +    V  + E  + ++    VQ     + + ++ +
Sbjct  1585  TGEIFVLKPLDRDQPNGRPQWRFTVFAQDEGGEGLVGYADVQ-----VNLKDINDNAPIF  1639

Query  336   PASKTFTVQKGISSKKLVDYILGTYQAIDEDTNKAASNVK--YVMGRN------DGGYLM  387
             P    F     ++       ++ T  A+D D     +N K  Y + +N
Sbjct  1640  PQGVYF---GNVTENGTAGMVVMTMTAVDYDDPNEGTNAKLIYSIEKNVIEEETGSPIFE  1696

Query  388   IDSKTAEIK-FVKNMNRDSTFIVNKTITAEVLAIDEYTGKTSTGTVYVRVPDFNDNCP  444
             I++ T  IK  V  ++R+ T       + +V+A+D   G   TGT  +RV D ND  P
Sbjct  1697  IEADTGVIKTAVCCLDRERT----PDYSIQVVAMDG-GGLKGTGTASIRVKDINDMPP  1749

 Score = 75.9 bits (185),  Expect = 2e-12, Method: Compositional matrix adjust.
 Identities = 106/426 (25%), Positives = 181/426 (42%), Gaps = 51/426 (12%)

Query  53    VDKNTGDINITAIVDREETPSFLITCRALNAQGLDVEKPLILTVKILDINDNPPVFSQ-Q  111
             +D++  ++N+TA+ D         +C  +N          ++ V I D+NDN PVF
Sbjct  928   LDRDKYELNVTAMDDG--------SC-CVNGDATIHTSTAVVVVFITDVNDNKPVFKDCG  978

Query  112   IFMGEIEENSASNSLVMILNATDADEPNHLNSKIAFKIVSQEPAGTPMFLLSRNTGEVRT  171
              +  ++EE + + S V+ + ATD D+   +N ++ + IV Q       F +   TGEV T
Sbjct  979   TYYPKVEEGAPNGSPVIKVQATDEDK--GVNGQVKYSIVQQPNQKGTKFTVDEETGEVST  1036

Query  172   LTNSLDRE--QASSYRLVVSGADKDGEGLSTQCECNIKVKDVNDNFPMFRDSQYSARIEE  229
                  DRE        + V   D+    L   C   +++ DVNDN P+F   +Y   +++
Sbjct  1037  -NKVFDREGDDGKFVSVTVKATDQGEPSLEGVCSFTVEITDVNDNPPLFDRQKYVENVKQ  1095

Query  230   NI-LSSELLRFQVTDLDEEYTDNWLAVYFFT---SGNEGNWFEIQTDPRTNEGILKVVKA  285
             +  + + +LR  V+  DE+  +N   VY  +   S N+  +FEIQ  P +   +LK  K
Sbjct  1096  DASIGTNILR--VSASDEDADNNGAIVYTLSAPHSPNDLEYFEIQ--PESGWIVLK--KP  1149

Query  286   LD--YEQLQSVKLSIAVKNK------AEFHQSVISRYRVQSTPVTIQV----INVREGIA  333
             LD  Y + +  +L +   ++      A+    +    R    P+  +     I++RE +
Sbjct  1150  LDGCYSR-ERYRLKVYASDRGDPPSTADVDVELDVVDRNNKPPIWDRTIYGPIHIRENVT  1208

Query  334   FRPASKTFTVQKGISSKKLVDYILGTYQAIDEDTNKAASNVKYVMGRNDGGYLMIDSKTA  393
                   +     GI     V Y L         TNK   +  Y+  R D G        A
Sbjct  1209  VGTVVTSVKASSGIEGNPTVFYRL--MPGSTAQTNKL--HTFYLQQRPDNG-----DTWA  1259

Query  394   EIKFVKNMNRDSTFIVNKTITAEVLAIDEYTGKTSTGTVYVRVPDFNDNCPTAV-LEKDA  452
             +IK    ++ ++    N T+  E     +     S  TVY+++ D ND  P     E++
Sbjct  1260  DIKVNHPLDFETIKEYNLTVRVENNGAQQL---ASEATVYIQLEDVNDEIPLFTEREQET  1316

Query  453   VCSSSP  458
             V    P
Sbjct  1317  VLEGEP  1322

 Score = 60.5 bits (145),  Expect = 9e-08, Method: Compositional matrix adjust.
 Identities = 83/362 (23%), Positives = 152/362 (42%), Gaps = 34/362 (9%)

Query  51    FVVDKNTGDINITAIVDRE-ETPSFL-ITCRALNAQGLDVEKPLILTVKILDINDNPPVF  108
             F VD+ TG+++   + DRE +   F+ +T +A +     +E     TV+I D+NDNPP+F
Sbjct  1025  FTVDEETGEVSTNKVFDREGDDGKFVSVTVKATDQGEPSLEGVCSFTVEITDVNDNPPLF  1084

Query  109   SQQIFMGEIEENSASNSLVMILNATDADEPNHLNSKIAFKIVS-QEPAGTPMFLLSRNTG  167
              +Q ++  ++++++  + ++ ++A+D D  N  N  I + + +   P     F +   +G
Sbjct  1085  DRQKYVENVKQDASIGTNILRVSASDEDADN--NGAIVYTLSAPHSPNDLEYFEIQPESG  1142

Query  168   EV---RTLTNSLDREQASSYRLVVSGADKDGEGLSTQCECNIKVKDVNDNFPMFRDSQYS  224
              +   + L     RE+   YRL V  +D+ G+  ST            +N P   D
Sbjct  1143  WIVLKKPLDGCYSRER---YRLKVYASDR-GDPPSTADVDVELDVVDRNNKPPIWDRTIY  1198

Query  225   ARIEENILSSELLRFQVTDLDEEYTDNWLAVYFFTSG-----NEGNWFEIQTDPRTNE--  277
               I      +                N    Y    G     N+ + F +Q  P   +
```

```
Sbjct  1199  GPIHIRENVTVGTVVTSVKASSGIEGNPTVFYRLMPGSTAQTNKLHTFYLQQRPDNGDTW  1258

Query  278   GILKVVKALDYEQLQSVKLSIAVKNKAEFHQSVISRYRVQSTPVTIQVINVREGIAFRPA  337
               +KV   LD+E ++   L++ V+N         ++       V IQ+ +V + I
Sbjct  1259  ADIKVNHPLDFETIKEYNLTVRVENNG-------AQQLASEATVYIQLEDVNDEIPLFTE  1311

Query  338   SKTFTVQKGISSKKLVDYILGTYQAIDEDTNKAASNVKYVM---GRNDG-GYLMIDSKTA  393
              +  TV +G      V +      AID+D     + V Y +    RN+G  +  I+ ++
Sbjct  1312  REQETVLEGEPIGTKVTQV----NAIDKDGTFPNNQVYYYIVDSPRNEGKDFFEINLQSG  1367

Query  394   EI  395
             EI
Sbjct  1368  EI  1369

 Score = 55.8 bits (133),  Expect = 3e-06, Method: Compositional matrix adjust.
 Identities = 49/158 (31%), Positives = 78/158 (49%), Gaps = 24/158 (15%)

Query  102   NDNPPVFSQQIFMGEIEENSASNSLVMILNATDADEPNHLNSKIAFKIVSQEPAGTP--M  159
             ND  P FSQQ++   ++EN+  N+LV  + A+D D  N     + F  V     GT
Sbjct  858   NDEEPKFSQQVYTPNVDENAGPNTLVTTVVASDKDGDN-----VRFGFVG---GGTSSGQ  909

Query  160   FLLSRNTGEVRTLTN--SLDREQASSYRLVVSGADK-------DGEGLSTQCECNIKVKD  210
             F++   TG +R  +   SLDR++   Y L V+  D        D    ++     + + D
Sbjct  910   FVIEEITGVIRLHSKAISLDRDK---YELNVTAMDDGSCCVNGDATIHTSTAVVVVFITD  966

Query  211   VNDNFPMFRD-SQYSARIEENILS-SELLRFQVTDLDE  246
             VNDN P+F+D   Y  ++EE   + S +++ Q TD D+
Sbjct  967   VNDNKPVFKDCGTYYPKVEEGAPNGSPVIKVQATDEDK  1004

 Score = 53.9 bits (128),  Expect = 1e-05, Method: Compositional matrix adjust.
 Identities = 70/306 (23%), Positives = 126/306 (41%), Gaps = 30/306 (10%)

Query  20    PIAKIT-SDYQATQKITYRI---SGVGIDQPPFGIFVVDKNTGDINITAIVDRE---ETP  72
             PI  +T  D   T K  Y++   SG G D+  F +   +  TG + I   +D E   ++
Sbjct  1772  PILTVTVHDEDETNKFQYKVIDNSGYGADK--FTMVRNNDGTGSLKIVQPLDYEDPLQSN  1829

Query  73    SFLITCRALNAQGLDVEKPLI------LTVKILDINDNPPVFSQQIFMGEIEENSASNSL  126
              F    + +N +G D +          + VK+ DINDN P F +      + EN+
Sbjct  1830  GFRFRIQ-VNDKGEDNDNDKYHVAYSWVVVKLRDINDNKPQFERPNIEVSVYENADVGKT  1888

Query  127   VMILNATDADEPNHLNSKIAFKIVSQEPAGTPMFLLSRNTGEVRTLTNSLDREQASSYRL  186
             +    ATD D+     SK+++ I   + +       S N     T+  +LDRE    +++
Sbjct  1889  LETFKATDPDQGG--KSKVSYAI---DRSSDRQRQFSINQEGTVTIQRNLDREVTPRHQV  1943

Query  187   VVSGADKDGEGLSTQCECNIKVKDVNDNFPMFRDSQYSARIEENILSSELLRFQVTDLDE  246
              +   D      +      + V+D+NDN P F    Y   + E++   +++    TD D+
Sbjct  1944  KILAIDDGIPPKTATATLTVIVQDINDNPPKFL-KDYRPVLPEHVPPRKVVEILATDDDD  2002

Query  247   EYTDNWLAVYFFTSGNEGNW----FEIQTDPRTNEG----ILKVVKALDYEQLQSVKLSI  298
                 N     F       +     F+++ D +   G    I+  +++ D EQ +   + I
Sbjct  2003  RSKSNGPPFQFRLDPGADDIIRASFKVEQDQKGANGDGMAIVSSLRSFDREQQKEYLIPI  2062

Query  299   AVKNKA  304
              +K+
Sbjct  2063  VIKDHG  2068

 Score = 52.0 bits (123),  Expect = 4e-05, Method: Compositional matrix adjust.
 Identities = 97/398 (24%), Positives = 163/398 (41%), Gaps = 54/398 (14%)

Query  106   PVFSQQIFMGEIEENSASNSLVMILNATDADEPNHLNSKIAFKIVSQ-EPAGTPMFLLSR  164
             P F    +  EI EN   +S ++ + A      +  + +I + + +Q + AGT  F +
Sbjct  655   PQFYMPSYEAEIPENQKKDSDIISVKAK-----SFADREIRYTLKAQGQGAGT--FNIGP  707

Query  165   NTGEVRTLTNSLDRE---QASSYRLVVSGADKDGEGLSTQCECNIKVKDVNDNFPMFRDS  221
              +G V+ L   LD E   Q   Y LVV+ A +D  G ST  E  I+V DVNDN P F
Sbjct  708   TSGIVK-LAKELDFEDLRQPHVYSLVVT-ATEDSGGFSTSVELTIRVTDVNDNAPKFELP  765

Query  222   QYSAR-IEENI-LSSELLRFQVTDLDEEYTDNWLAVYFFTSGNEGNWFEIQTDPRTNEGI  279
```

```
              Y A  ++E+I L + +L+ +  D D         + +  S +    F + ++     GI
Sbjct  766   DYQAHNVDEDIPLGTSILKVKAMDSDSGAN---AEIEYLVSDDH---FAVDSN-----GI  814

Query  280   LKVVKALDYEQLQS-VKLSIAVKNKAEFHQSVISRYRVQSTPVTIQVINVREGIAFRPAS  338
             +   K LD +    +  +  +  K+K E  ++  +  RV +            E   F
Sbjct  815   IVNNKQLDADNNNAYYEFVVTAKDKGEPAKTGTATVRVYTKNKN------DEEPKFSQQV  868

Query  339   KTFTVQKGISSKKLVDYILGTYQAIDEDTNKAASNVK--YVMGRNDGGYLMIDSKTAEIK  396
              T  V +      LV  ++          ++K   NV+  +V G    G  +I+  T  I+
Sbjct  869   YTPNVDENAGPNTLVTTVVA--------SDKDGDNVRFGFVGGGTSSGQFVIEEITGVIR  920

Query  397   FVK---NMNRDSTFI-VNKTITAEVLAIDEYTGKTSTGTVYVRVPDFNDN------CPTA  446
                   +++RD   + V             + T  TST  V V + D NDN      C T
Sbjct  921   LHSKAISLDRDKYELNVTAMDDGSCCVNGDATIHTSTAVVVVFITDVNDNKPVFKDCGTY  980

Query  447   VLE-KDAVCSSSPSVVVSARTLNNRYTGPYTFALEDQP  483
               + ++   + SP + V A    +    G   +++  QP
Sbjct  981   YPKVEEGAPNGSPVIKVQATDEDKGVNGQVKYSIVQQP  1018

 Score = 44.7 bits (104),  Expect = 0.008, Method: Compositional matrix adjust.
 Identities = 48/199 (24%), Positives = 73/199 (37%), Gaps = 19/199 (10%)

Query  27    DYQATQKITYRISGVGIDQPPFGIFVVDKNTGDINITAIVDREETPSFLITCRALNAQGL  86
             D     K++Y I      Q  F I        G + I   +DRE TP   +    A++
Sbjct  1898  DQGGKSKVSYAIDRSSDRQRQFSI----NQEGTVTIQRNLDREVTPRHQVKILAIDDGIP  1953

Query  87    DVEKPLILTVKILDINDNPPVFSQQIFMGEIEENSASNSLVMILNATDADEPNHLNSKIA  146
                    LTV + DINDNPP F +  +    + E+     +V IL   D D
Sbjct  1954  PKTATATLTVIVQDINDNPPKFLKD-YRPVLPEHVPPRKVVEILATDDDDRSKSNGPPFQ  2012

Query  147   FKIVSQEPAGTPMFLLSRNTGEVRTLTN-----------SLDREQASSYRLVVSGADKDG  195
             F++    +P    +   S    + +   N           S DREQ    Y + +   D
Sbjct  2013  FRL---DPGADDIIRASFKVEQDQKGANGDGMAIVSSLRSFDREQQKEYLIPIVIKDHGN  2069

Query  196   EGLSTQCECNIKVKDVNDN  214
               ++      + + DVNDN
Sbjct  2070  PAMTGTSTLTVVIGDVNDN  2088

 Score = 40.0 bits (92),  Expect = 0.22, Method: Compositional matrix adjust.
 Identities = 89/389 (23%), Positives = 149/389 (38%), Gaps = 62/389 (16%)

Query  75   LITCRALNAQGLDVEKPLILTVKILDINDNPPVFSQQIF--MGEIEENSASNSLVMILNA  132
            +IT +  NA GL       + + + D+ND PP F  +        ++ N+  N+ V  L A
Sbjct  517  IITNQGHNA-GLKYTDNQRVIIHVKDVNDEPPYFINRPLPMQAVVQLNAPPNTPVFTLQA  575

Query  133  TDADEPNHLNSKIAFKIVSQEPAGTPMFLLSRNTGEVRTLTNSLDREQASSYRLVVSGAD  192
             D D     +  I + IV     G  F +   +G VRT    L  +    Y L V   D
Sbjct  576  RDPD----TDHNIHYFIVRDRTGG--RFEVDERSGVVRTRGTDL-FQLDMEYVLYVKAED  628

Query  193  KDGE----GLSTQCECNIKVKDVNDNFPMFRDSQYSARIEEN-ILSSELLRFQVTDLDEE  247
            ++G+        +  E  + +        P F    Y A I EN    S+++  +     +
Sbjct  629  QNGKVDDRRFQSTPEERLSIVG-GKRAPQFYMPSYEAEIPENQKKDSDIISVKA----KS  683

Query  248  YTDNWLAVYFFTSGNEGNWFEIQTDPRTNEGILKVVKALDYEQLQSVKLS----IAVKNK  303
            + D  +        G     F I          GI+K+ K LD+E L+    +      A ++
Sbjct  684  FADREIRYTLKAQGQGAGTFNIGP----TSGIVKLAKELDFEDLRQPHVYSLVVTATEDS  739

Query  304  AEFHQSVISRYRVQSTPVTIQVINVREGIAFRPASKTFTVQKGISSKKLVDYILGT----  359
              F  SV           +TI+V +V +        +  F +    +     D  LGT
Sbjct  740  GGFSTSV---------ELTIRVTDVNDN------APKFELPDYQAHNVDEDIPLGTSILK  784

Query  360  YQAIDEDTNKAASNVKYVMGRN----DGGYLMIDSKTAEIKFVKNMNRDSTFIVNKTITA  415
             +A+D D+  A + ++Y++  +    D   +++++K  +     N N    F+V
Sbjct  785  VKAMDSDSG-ANAEIEYLVSDDHFAVDSNGIIVNNKQLD---ADNNNAYYEFVVTAKDKG  840

Query  416  EVLAIDEYTGKTSTGTVYVRVPDFNDNCP  444
            E        KT T TV V   + ND  P
Sbjct  841  E-------PAKTGTATVRVYTKNKNDEEP  862
```

```
 Score = 39.7 bits (91),  Expect = 0.28, Method: Compositional matrix adjust.
 Identities = 54/242 (22%), Positives = 102/242 (42%), Gaps = 22/242 (9%)

Query  4    KFAKPCREGE--DNSKRNP-IAKITSDYQATQKITYRISGVGIDQPPFGIFVVDKNTGDI  60
            +F  P  E E  +N K++  I  + +   A ++I Y +   G      G F +   +G +
Sbjct  656  QFYMPSYEAEIPENQKKDSDIISVKAKSFADREIRYTLKAQGQGA---GTFNIGPTSGIV  712

Query  61   NITAIVDREE--TPSF--LITCRALNAQGLDVEKPLILTVKILDINDNPPVFSQQIFMGE  116
             +   +D E+   P    L+     ++ G      + LT+++ D+NDN P F    +
Sbjct  713  KLAKELDFEDLRQPHVYSLVVTATEDSGGFSTS--VELTIRVTDVNDNAPKFELPDYQAH  770

Query  117  -IEENSASNSLVMILNATDADEPNHLNSKIAFKIVSQEPAGTPMFLLSRNTGEVRTLTNS  175
             ++E+    + ++ + A D+D  +  N++I + +VS +      F +  N   V
Sbjct  771  NVDEDIPLGTSILKVKAMDSD--SGANAEIEY-LVSDD-----HFAVDSNGIIVNNKQLD  822

Query  176  LDREQASSYRLVVSGADKDGEGLSTQCECNIKVKDVNDNFPMFRDSQYSARIEENILSSE  235
             D   A  Y  VV+  DK     +      +  K+ ND  P F    Y+  ++EN   +
Sbjct  823  ADNNNA-YYEFVVTAKDKGEPAKTGTATVRVYTKNKNDEEPKFSQQVYTPNVDENAGPNT  881

Query  236  LL  237
            L+
Sbjct  882  LV  883

>ASTEI20_040830.P55059 | transcript=ASTEI20_040830.R55059 | gene=ASTEI20_040830
| organism=Anopheles_stephensi_UCISS2018 |
gene_product=neural-cadherin, transcript variant X5 |
transcript_product=neural-cadherin,
transcript variant X7 | location=CM023249:57079885-57269506(-)
| protein_length=3112 | sequence_SO=chromosome
| SO=protein_coding_gene | is_pseudo=false
Length=3112

 Score = 147 bits (370),  Expect = 3e-35, Method: Compositional matrix adjust.
 Identities = 128/446 (29%), Positives = 217/446 (49%), Gaps = 36/446 (8%)

Query  20    PIAKITS---DYQATQKITYRISGVGID--QPPFGIFVVDKNTGDINITAIVDREET---  71
             PI ++T+   D    Q I Y ++G GID   P    F +++ TG+I +   +DR++
Sbjct  1552  PIMQVTATDGDKDRPQNIVYFLTGQGIDPDNPANSKFDINRTTGEIFVLKPLDRDQPNGR  1611

Query  72    PSFLITCRALNAQGLDVEKPLILTVKILDINDNPPVFSQQIFMGEIEENSASNSLVMILN  131
             P +  T  A +  G  +     + V + DINDN P+F Q ++ G + EN  +  +VM +
Sbjct  1612  PQWRFTVFAQDEGGEGLVGYADVQVNLKDINDNAPIFPQGVYFGNVTENGTAGMVVMTMT  1671

Query  132   ATDADEPNH-LNSKIAFKI---VSQEPAGTPMFLLSRNTGEVRTLTNSLDREQASSYRLV  187
             A D D+PN   N+K+ + I   V +E  G+P+F +  +TG ++T    LDRE+   Y +
Sbjct  1672  AVDYDDPNEGTNAKLIYSIEKNVIEEETGSPIFEIEADTGVIKTAVCCLDRERTPDYSIQ  1731

Query  188   VSGADKDGEGLSTQCECNIKVKDVNDNFPMFRDSQYSARIEENILSS--ELLRFQVTDLD  245
             V     DG GL      +I+VKD+ND  P F   ++   ++E   ++  E+    VT  D
Sbjct  1732  VVAM--DGGGLKGTGTASIRVKDINDMPPQFTKDEWFTEVDETDGTNLPEMPILTVTVHD  1789

Query  246   EEYTDNWLAVYFFTSGNEGNWFEIQTDPRTNEGI--LKVVKALDYE---QLQSVKLSIAV  300
             E+ T+ +       SG   + F +    R N+G   LK+V+ LDYE   Q    +  I V
Sbjct  1790  EDETNKFQYKVIDNSGYGADKFTMV---RNNDGTGSLKIVQPLDYEDPLQSNGFRFRIQV  1846

Query  301   KNKAEFHQSVISRYRVQSTPVTIQVINVREGIAFRPASKTFTVQKGISSKKLVDYILGTY  360
              +K E + +   +Y V  + V +++ ++ +    +P  +   ++  +     V   L T+
Sbjct  1847  NDKGEDNDN--DKYHVAYSWVVVKLRDINDN---KPQFERPNIEVSVYENADVGKTLETF  1901

Query  361   QAIDEDTNKAASNVKYVMGR-NDGGYLMIDSKTAEIKFVKNMNRDSTFIVNKTITAEVLA  419
             +A D D     S V Y + R +D       ++   +   +N++R+    V      ++LA
Sbjct  1902  KATDPDQG-GKSKVSYAIDRSSDRQRQFSINQEGTVTIQRNLDRE----VTPRHQVKILA  1956

Query  420   IDE-YTGKTSTGTVYVRVPDFNDNCP  444
             ID+    KT+T T+ V V D NDN P
Sbjct  1957  IDDGIPPKTATATLTVIVQDINDNPP  1982

 Score = 76.6 bits (187),  Expect = 1e-12, Method: Compositional matrix adjust.
```

```
 Identities = 103/418 (25%), Positives = 174/418 (42%), Gaps = 50/418 (12%)

Query  50    IFVVDKNTGDINITAIVDREETPSFLITCRAL----------NAQGLDVEKPLILTVKIL  99
              F ++  +G+I    + DRE+  ++ +   A           N Q   V K   + + I
Sbjct  1368  FFEINLQSGEIFTKVVFDREKQGAYALEVEARDGAPSARPNSNQQPNSVTK--FIRIGIA  1425

Query  100   DINDNPPVFSQQIFMGEIEENSASNSLVMILNATDADEPNHLNSKIAFKIVSQEPAGTPM  159
             D NDNPP F + ++  E++EN      V+ + A D DE    +S+I ++I S    G
Sbjct  1426  DKNDNPPYFDKALYEAEVDENEDIQHTVLTVTAKDHDE----SSRIRYEITSGNIGGA--  1479

Query  160   FLLSRNTGEVRTLTNSLDREQASSYRLVVSGADKDGEGLSTQCECNIKVKDVNDNFPMFR  219
             F +    TG +  +  +LD E    Y L +  +D   E  +       I VKDVND  P F
Sbjct  1480  FAVKNMTGAI-YVAGALDYETRKRYHLTLVASDSLNENETIIV---INVKDVNDMPPSFP  1535

Query  220   DSQYSARIEENI-LSSELLRFQVTDLDEEYTDNWLAVYFFTSG--NEGNWFEIQTDPRTN  276
                Y   ++E + +   +++   TD D++   N   VYF T    +  N    + D
Sbjct  1536  QQVYERTMDEELAVPFPIMQVTATDGDKDRPQN--IVYFLTGQGIDPDNPANSKFDINRT  1593

Query  277   EGILKVVKALDYEQLQS-VKLSIAVKNKAEFHQSVISRYRVQSTPVTIQVINVREGIAFR  335
              G + V+K LD +Q      +     V  + E  + ++    VQ     + + ++ +
Sbjct  1594  TGEIFVLKPLDRDQPNGRPQWRFTVFAQDEGGEGLVGYADVQ-----VNLKDINDNAPIF  1648

Query  336   PASKTFTVQKGISSKKLVDYILGTYQAIDEDTNKAASNVK--YVMGRN------DGGYLM  387
             P    F     ++        ++ T  A+D D     +N K  Y + +N
Sbjct  1649  PQGVYF---GNVTENGTAGMVVMTMTAVDYDDPNEGTNAKLIYSIEKNVIEEETGSPIFE  1705

Query  388   IDSKTAEIK-FVKNMNRDSTFIVNKTITAEVLAIDEYTGKTSTGTVYVRVPDFNDNCP  444
             I++ T  IK  V  ++R+ T        + +V+A+D   G   TGT  +RV D ND  P
Sbjct  1706  IEADTGVIKTAVCCLDRERT----PDYSIQVVAMDG-GGLKGTGTASIRVKDINDMPP  1758

 Score = 75.9 bits (185),  Expect = 2e-12, Method: Compositional matrix adjust.
 Identities = 106/426 (25%), Positives = 181/426 (42%), Gaps = 51/426 (12%)

Query  53    VDKNTGDINITAIVDREETPSFLITCRALNAQGLDVEKPLILTVKILDINDNPPVFSQ-Q  111
             +D++  ++N+TA+ D          +C  +N            ++ V I D+NDN PVF
Sbjct  937   LDRDKYELNVTAMDDG--------SC-CVNGDATIHTSTAVVVVFITDVNDNKPVFKDCG  987

Query  112   IFMGEIEENSASNSLVMILNATDADEPNHLNSKIAFKIVSQEPAGTPMFLLSRNTGEVRT  171
              +  ++EE + + S V+ + ATD D+   +N ++ + IV Q        F +   TGEV T
Sbjct  988   TYYPKVEEGAPNGSPVIKVQATDEDK--GVNGQVKYSIVQQPNQKGTKFTVDEETGEVST  1045

Query  172   LTNSLDRE--QASSYRLVVSGADKDGEGLSTQCECNIKVKDVNDNFPMFRDSQYSARIEE  229
                   DRE         + V   D+     L   C    +++ DVNDN P+F    +Y   +++
Sbjct  1046  -NKVFDREGDDGKFVSVTVKATDQGEPSLEGVCSFTVEITDVNDNPPLFDRQKYVENVKQ  1104

Query  230   NI-LSSELLRFQVTDLDEEYTDNWLAVYFFT---SGNEGNWFEIQTDPRTNEGILKVVKA  285
             +  + + +LR  V+  DE+  +N   VY  +    S N+  +FEIQ  P +   +LK  K
Sbjct  1105  DASIGTNILR--VSASDEDADNNGAIVYTLSAPHSPNDLEYFEIQ--PESGWIVLK--KP  1158

Query  286   LD--YEQLQSVKLSIAVKNK------AEFHQSVISRYRVQSTPVTIQV----INVREGIA  333
             LD  Y + +  +L +   ++       A+    +    R    P+  +      I++RE +
Sbjct  1159  LDGCYSR-ERYRLKVYASDRGDPPSTADVDVELDVVDRNNKPPIWDRTIYGPIHIRENVT  1217

Query  334   FRPASKTFTVQKGISSKKLVDYILGTYQAIDEDTNKAASNVKYVMGRNDGGYLMIDSKTA  393
                     +      GI      V Y L           TNK    + Y+  R D G          A
Sbjct  1218  VGTVVTSVKASSGIEGNPTVFYRL--MPGSTAQTNKL--HTFYLQQRPDNG-----DTWA  1268

Query  394   EIKFVKNMNRDSTFIVNKTITAEVLAIDEYTGKTSTGTVYVRVPDFNDNCPTAV-LEKDA  452
             +IK    ++ ++    N T+  E      +      S  TVY+++ D ND  P      E++
Sbjct  1269  DIKVNHPLDFETIKEYNLTVRVENNGAQQL---ASEATVYIQLEDVNDEIPLFTEREQET  1325

Query  453   VCSSSP  458
             V   P
Sbjct  1326  VLEGEP  1331

 Score = 60.5 bits (145),  Expect = 1e-07, Method: Compositional matrix adjust.
 Identities = 83/362 (23%), Positives = 152/362 (42%), Gaps = 34/362 (9%)

Query  51    FVVDKNTGDINITAIVDRE-ETPSFL-ITCRALNAQGLDVEKPLILTVKILDINDNPPVF  108
```

```
               F VD+ TG+++   + DRE +   F+ +T +A +     +E     TV+I D+NDNPP+F
Sbjct  1034   FTVDEETGEVSTNKVFDREGDDGKFVSVTVKATDQGEPSLEGVCSFTVEITDVNDNPPLF  1093

Query  109    SQQIFMGEIEENSASNSLVMILNATDADEPNHLNSKIAFKIVS-QEPAGTPMFLLSRNTG  167
               +Q ++  ++++++  + ++ ++A+D D  N  N  I + + +   P     F +   +G
Sbjct  1094   DRQKYVENVKQDASIGTNILRVSASDEDADN--NGAIVYTLSAPHSPNDLEYFEIQPESG  1151

Query  168    EV---RTLTNSLDREQASSYRLVVSGADKDGEGLSTQCECNIKVKDVNDNFPMFRDSQYS  224
                +   + L     RE+   YRL V  +D+ G+  ST             +N P   D
Sbjct  1152   WIVLKKPLDGCYSRER---YRLKVYASDR-GDPPSTADVDVELDVVDRNNKPPIWDRTIY  1207

Query  225    ARIEENILSSELLRFQVTDLDEEYTDNWLAVYFFTSG-----NEGNWFEIQTDPRTNE--  277
                I      +                   N    Y    G     N+ + F +Q  P   +
Sbjct  1208   GPIHIRENVTVGTVVTSVKASSGIEGNPTVFYRLMPGSTAQTNKLHTFYLQQRPDNGDTW  1267

Query  278    GILKVVKALDYEQLQSVKLSIAVKNKAEFHQSVISRYRVQSTPVTIQVINVREGIAFRPA  337
                +KV   LD+E ++   L++ V+N          ++       V IQ+ +V + I
Sbjct  1268   ADIKVNHPLDFETIKEYNLTVRVENNG-------AQQLASEATVYIQLEDVNDEIPLFTE  1320

Query  338    SKTFTVQKGISSKKLVDYILGTYQAIDEDTNKAASNVKYVM---GRNDG-GYLMIDSKTA  393
                +  TV +G      V  +     AID+D     + V Y +    RN+G  +  I+ ++
Sbjct  1321   REQETVLEGEPIGTKVTQV----NAIDKDGTFPNNQVYYYIVDSPRNEGKDFFEINLQSG  1376

Query  394    EI  395
               EI
Sbjct  1377   EI  1378

 Score = 55.8 bits (133),  Expect = 3e-06, Method: Compositional matrix adjust.
 Identities = 49/158 (31%), Positives = 78/158 (49%), Gaps = 24/158 (15%)

Query  102    NDNPPVFSQQIFMGEIEENSASNSLVMILNATDADEPNHLNSKIAFKIVSQEPAGTP--M  159
               ND  P FSQQ++   ++EN+  N+LV  + A+D D  N     + F  V     GT
Sbjct  867    NDEEPKFSQQVYTPNVDENAGPNTLVTTVVASDKDGDN-----VRFGFVG---GGTSSGQ  918

Query  160    FLLSRNTGEVRTLTN--SLDREQASSYRLVVSGADK-------DGEGLSTQCECNIKVKD  210
               F++   TG +R  +   SLDR++   Y L V+  D         D   ++      + + D
Sbjct  919    FVIEEITGVIRLHSKAISLDRDK---YELNVTAMDDGSCCVNGDATIHTSTAVVVVFITD  975

Query  211    VNDNFPMFRD-SQYSARIEENILS-SELLRFQVTDLDE  246
               VNDN P+F+D   Y  ++EE   + S +++ Q TD D+
Sbjct  976    VNDNKPVFKDCGTYYPKVEEGAPNGSPVIKVQATDEDK  1013

 Score = 53.5 bits (127),  Expect = 1e-05, Method: Compositional matrix adjust.
 Identities = 70/306 (23%), Positives = 126/306 (41%), Gaps = 30/306 (10%)

Query  20     PIAKIT-SDYQATQKITYRI---SGVGIDQPPFGIFVVDKNTGDINITAIVDRE---ETP  72
               PI  +T  D   T K  Y++   SG G D+  F +    +  TG + I   +D E   ++
Sbjct  1781   PILTVTVHDEDETNKFQYKVIDNSGYGADK--FTMVRNNDGTGSLKIVQPLDYEDPLQSN  1838

Query  73     SFLITCRALNAQGLDVEKPLI------LTVKILDINDNPPVFSQQIFMGEIEENSASNSL  126
                F    + +N +G D +           + VK+ DINDN P F +      + EN+
Sbjct  1839   GFRFRIQ-VNDKGEDNDNDKYHVAYSWVVVKLRDINDNKPQFERPNIEVSVYENADVGKT  1897

Query  127    VMILNATDADEPNHLNSKIAFKIVSQEPAGTPMFLLSRNTGEVRTLTNSLDREQASSYRL  186
               +    ATD D+      SK+++ I    + +        S N      T+  +LDRE    +++
Sbjct  1898   LETFKATDPDQGG--KSKVSYAI---DRSSDRQRQFSINQEGTVTIQRNLDREVTPRHQV  1952

Query  187    VVSGADKDGEGLSTQCECNIKVKDVNDNFPMFRDSQYSARIEENILSSELLRFQVTDLDE  246
                +   D       +       + V+D+NDN P F     Y    + E++    +++     TD D+
Sbjct  1953   KILAIDDGIPPKTATATLTVIVQDINDNPPKFL-KDYRPVLPEHVPPRKVVEILATDDDD  2011

Query  247    EYTDNWLAVYFFTSGNEGNW----FEIQTDPRTNEG----ILKVVKALDYEQLQSVKLSI  298
                  N     F        +      F+++ D +   G     I+  +++ D EQ +    + I
Sbjct  2012   RSKSNGPPFQFRLDPGADDIIRASFKVEQDQKGANGDGMAIVSSLRSFDREQQKEYLIPI  2071

Query  299    AVKNKA  304
                +K+
Sbjct  2072   VIKDHG  2077
```

```
 Score = 52.4 bits (124),  Expect = 3e-05, Method: Compositional matrix adjust.
 Identities = 97/398 (24%), Positives = 163/398 (41%), Gaps = 54/398 (14%)

Query  106   PVFSQQIFMGEIEENSASNSLVMILNATDADEPNHLNSKIAFKIVSQ-EPAGTPMFLLSR  164
             P F    +  EI EN   +S ++ + A      +  + +I + + +Q + AGT  F +
Sbjct  664   PQFYMPSYEAEIPENQKKDSDIISVKAK-----SFADREIRYTLKAQGQGAGT--FNIGP  716

Query  165   NTGEVRTLTNSLDRE---QASSYRLVVSGADKDGEGLSTQCECNIKVKDVNDNFPMFRDS  221
              +G V+ L   LD E   Q   Y LVV+ A +D  G ST  E  I+V DVNDN P F
Sbjct  717   TSGIVK-LAKELDFEDLRQPHVYSLVVT-ATEDSGGFSTSVELTIRVTDVNDNAPKFELP  774

Query  222   QYSAR-IEENI-LSSELLRFQVTDLDEEYTDNWLAVYFFTSGNEGNWFEIQTDPRTNEGI  279
              Y A  ++E+I L + +L+ +  D D         + +  S +    F + ++     GI
Sbjct  775   DYQAHNVDEDIPLGTSILKVKAMDADSGAN---AEIEYLVSDDH---FAVDSN-----GI  823

Query  280   LKVVKALDYEQLQS-VKLSIAVKNKAEFHQSVISRYRVQSTPVTIQVINVREGIAFRPAS  338
             +   K LD +   +  +  +  K+K E  ++  +  RV +            E   F
Sbjct  824   IVNNKQLDADNNNAYYEFVVTAKDKGEPAKTGTATVRVYTKNKN------DEEPKFSQQV  877

Query  339   KTFTVQKGISSKKLVDYILGTYQAIDEDTNKAASNVK--YVMGRNDGGYLMIDSKTAEIK  396
              T  V +      LV  ++          ++K   NV+  +V G    G  +I+  T  I+
Sbjct  878   YTPNVDENAGPNTLVTTVVA--------SDKDGDNVRFGFVGGGTSSGQFVIEEITGVIR  929

Query  397   FVK---NMNRDSTFI-VNKTITAEVLAIDEYTGKTSTGTVYVRVPDFNDN------CPTA  446
                   +++RD   + V             + T  TST  V V + D NDN      C T
Sbjct  930   LHSKAISLDRDKYELNVTAMDDGSCCVNGDATIHTSTAVVVVFITDVNDNKPVFKDCGTY  989

Query  447   VLE-KDAVCSSSPSVVVSARTLNNRYTGPYTFALEDQP  483
               + ++   + SP + V A   +    G   +++  QP
Sbjct  990   YPKVEEGAPNGSPVIKVQATDEDKGVNGQVKYSIVQQP  1027

 Score = 44.3 bits (103),  Expect = 0.009, Method: Compositional matrix adjust.
 Identities = 48/199 (24%), Positives = 73/199 (37%), Gaps = 19/199 (10%)

Query  27    DYQATQKITYRISGVGIDQPPFGIFVVDKNTGDINITAIVDREETPSFLITCRALNAQGL  86
             D     K++Y I      Q  F I        G + I   +DRE TP    +   A++
Sbjct  1907  DQGGKSKVSYAIDRSSDRQRQFSI----NQEGTVTIQRNLDREVTPRHQVKILAIDDGIP  1962

Query  87    DVEKPLILTVKILDINDNPPVFSQQIFMGEIEENSASNSLVMILNATDADEPNHLNSKIA  146
                    LTV + DINDNPP F +  +   + E+     +V IL   D D
Sbjct  1963  PKTATATLTVIVQDINDNPPKFLKD-YRPVLPEHVPPRKVVEILATDDDDRSKSNGPPFQ  2021

Query  147   FKIVSQEPAGTPMFLLSRNTGEVRTLTN-----------SLDREQASSYRLVVSGADKDG  195
             F++   +P     +   S    + +   N           S DREQ   Y + +   D
Sbjct  2022  FRL---DPGADDIIRASFKVEQDQKGANGDGMAIVSSLRSFDREQQKEYLIPIVIKDHGN  2078

Query  196   EGLSTQCECNIKVKDVNDN  214
               ++      + + DVNDN
Sbjct  2079  PAMTGTSTLTVVIGDVNDN  2097

 Score = 40.4 bits (93),  Expect = 0.13, Method: Compositional matrix adjust.
 Identities = 54/235 (23%), Positives = 99/235 (42%), Gaps = 22/235 (9%)

Query  4    KFAKPCREGE--DNSKRNP-IAKITSDYQATQKITYRISGVGIDQPPFGIFVVDKNTGDI  60
            +F  P  E E  +N K++  I  + +   A ++I Y +   G      G F +   +G +
Sbjct  665  QFYMPSYEAEIPENQKKDSDIISVKAKSFADREIRYTLKAQGQGA---GTFNIGPTSGIV  721

Query  61   NITAIVDREE--TPSF--LITCRALNAQGLDVEKPLILTVKILDINDNPPVFSQQIFMGE  116
             +   +D E+   P    L+      ++ G       + LT+++ D+NDN P F    +
Sbjct  722  KLAKELDFEDLRQPHVYSLVVTATEDSGGFSTS--VELTIRVTDVNDNAPKFELPDYQAH  779

Query  117  -IEENSASNSLVMILNATDADEPNHLNSKIAFKIVSQEPAGTPMFLLSRNTGEVRTLTNS  175
             ++E+    + ++ + A DAD  +  N++I + +VS +       F +  N   V
Sbjct  780  NVDEDIPLGTSILKVKAMDAD--SGANAEIEY-LVSDD-----HFAVDSNGIIVNNKQLD  831

Query  176  LDREQASSYRLVVSGADKDGEGLSTQCECNIKVKDVNDNFPMFRDSQYSARIEEN  230
             D   A  Y  VV+  DK     +       +  K+ ND  P F    Y+  ++EN
Sbjct  832  ADNNNA-YYEFVVTAKDKGEPAKTGTATVRVYTKNKNDEEPKFSQQVYTPNVDEN  885
```

```
 Score = 39.3 bits (90),  Expect = 0.33, Method: Compositional matrix adjust.
 Identities = 89/389 (23%), Positives = 149/389 (38%), Gaps = 62/389 (16%)

Query  75   LITCRALNAQGLDVEKPLILTVKILDINDNPPVFSQQIF--MGEIEENSASNSLVMILNA  132
            +IT +  NA GL       + + + D+ND PP F  +       ++ N+  N+ V  L A
Sbjct  526  IITNQGHNA-GLKYTDNQRVIIHVKDVNDEPPYFINRPLPMQAVVQLNAPPNTPVFTLQA  584

Query  133  TDADEPNHLNSKIAFKIVSQEPAGTPMFLLSRNTGEVRTLTNSLDREQASSYRLVVSGAD  192
             D D     +  I + IV     G   F +   +G VRT    L  +    Y L V   D
Sbjct  585  RDPD----TDHNIHYFIVRDRTGG--RFEVDERSGVVRTRGTDL-FQLDMEYVLYVKAED  637

Query  193  KDGE----GLSTQCECNIKVKDVNDNFPMFRDSQYSARIEEN-ILSSELLRFQVTDLDEE  247
            ++G+       +  E  + +       P F    Y A I EN    S+++  +     +
Sbjct  638  QNGKVDDRRFQSTPEERLSIVG-GKRAPQFYMPSYEAEIPENQKKDSDIISVKA----KS  692

Query  248  YTDNWLAVYFFTSGNEGNWFEIQTDPRTNEGILKVVKALDYEQLQSVKLS----IAVKNK  303
            + D  +       G     F I        GI+K+ K LD+E L+   +      A ++
Sbjct  693  FADREIRYTLKAQGQGAGTFNIGP----TSGIVKLAKELDFEDLRQPHVYSLVVTATEDS  748

Query  304  AEFHQSVISRYRVQSTPVTIQVINVREGIAFRPASKTFTVQKGISSKKLVDYILGT----  359
              F  SV          +TI+V +V +       +  F +    +     D  LGT
Sbjct  749  GGFSTSV---------ELTIRVTDVNDN------APKFELPDYQAHNVDEDIPLGTSILK  793

Query  360  YQAIDEDTNKAASNVKYVMGRN----DGGYLMIDSKTAEIKFVKNMNRDSTFIVNKTITA  415
             +A+D D+  A + ++Y++  +    D   +++++K  +     N N    F+V
Sbjct  794  VKAMDADSG-ANAEIEYLVSDDHFAVDSNGIIVNNKQLD---ADNNNAYYEFVVTAKDKG  849

Query  416  EVLAIDEYTGKTSTGTVYVRVPDFNDNCP  444
            E         KT T TV V   + ND  P
Sbjct  850  E-------PAKTGTATVRVYTKNKNDEEP  871

>AMEM21_003341.P6424 | transcript=AMEM21_003341.R6424 | gene=AMEM21_003341
| organism=Anopheles_merus_MAF_2021 | gene_product=neural-cadherin,
transcript variant X1 | transcript_product=neural-cadherin,
transcript variant X10 | location=CM029766:40289848-40498951(+)
| protein_length=3103 | sequence_SO=chromosome
| SO=protein_coding_gene | is_pseudo=false
Length=3103

 Score = 147 bits (370),  Expect = 3e-35, Method: Compositional matrix adjust.
 Identities = 128/446 (29%), Positives = 217/446 (49%), Gaps = 36/446 (8%)

Query  20    PIAKITS---DYQATQKITYRISGVGID--QPPFGIFVVDKNTGDINITAIVDREET---  71
             PI ++T+   D    Q I Y ++G GID   P    F +++ TG+I +   +DR++
Sbjct  1543  PIMQVTATDGDKDRPQNIVYFLTGQGIDPDNPANSKFDINRTTGEIFVLKPLDRDQPNGR  1602

Query  72    PSFLITCRALNAQGLDVEKPLILTVKILDINDNPPVFSQQIFMGEIEENSASNSLVMILN  131
             P +  T  A +  G  +     + V + DINDN P+F Q ++ G + EN  +  +VM +
Sbjct  1603  PQWRFTVFAQDEGGEGLVGYADVQVNLKDINDNAPIFPQGVYFGNVTENGTAGMVVMTMT  1662

Query  132   ATDADEPNH-LNSKIAFKI---VSQEPAGTPMFLLSRNTGEVRTLTNSLDREQASSYRLV  187
             A D D+PN   N+K+ + I   V +E  G+P+F +  +TG ++T    LDRE+   Y +
Sbjct  1663  AVDYDDPNEGTNAKLIYSIEKNVIEEETGSPIFEIEADTGVIKTAVCCLDRERTPDYSIQ  1722

Query  188   VSGADKDGEGLSTQCECNIKVKDVNDNFPMFRDSQYSARIEENILSS--ELLRFQVTDLD  245
             V     DG GL      +I+VKD+ND  P F   ++   ++E   ++  E+    VT  D
Sbjct  1723  VVAM--DGGGLKGTGTASIRVKDINDMPPQFTKDEWFTEVDETDGTNLPEMPILTVTVHD  1780

Query  246   EEYTDNWLAVYFFTSGNEGNWFEIQTDPRTNEGI--LKVVKALDYE---QLQSVKLSIAV  300
             E+ T+ +       SG   + F +    R N+G   LK+V+ LDYE   Q    +  I V
Sbjct  1781  EDETNKFQYKVIDNSGYGADKFTMV---RNNDGTGSLKIVQPLDYEDPLQSNGFRFRIQV  1837

Query  301   KNKAEFHQSVISRYRVQSTPVTIQVINVREGIAFRPASKTFTVQKGISSKKLVDYILGTY  360
              +K E + +   +Y V  + V +++ ++ +    +P  +   ++  +     V   L T+
Sbjct  1838  NDKGEDNDN--DKYHVAYSWVVVKLRDINDN---KPQFERPNIEVSVYENADVGKTLETF  1892

Query  361   QAIDEDTNKAASNVKYVMGR-NDGGYLMIDSKTAEIKFVKNMNRDSTFIVNKTITAEVLA  419
             +A D D     S V Y + R +D       ++   +   +N++R+    V      ++LA
```

```
Sbjct  1893  KATDPDQG-GKSKVSYAIDRSSDRQRQFSINQEGTVTIQRNLDRE----VTPRHQVKILA  1947

Query  420   IDE-YTGKTSTGTVYVRVPDFNDNCP  444
             ID+    KT+T T+ V V D NDN P
Sbjct  1948  IDDGIPPKTATATLTVIVQDINDNPP  1973

 Score = 75.9 bits (185),  Expect = 2e-12, Method: Compositional matrix adjust.
 Identities = 106/426 (25%), Positives = 181/426 (42%), Gaps = 51/426 (12%)

Query  53    VDKNTGDINITAIVDREETPSFLITCRALNAQGLDVEKPLILTVKILDINDNPPVFSQ-Q  111
             +D++  ++N+TA+ D         +C  +N          ++ V I D+NDN PVF
Sbjct  928   LDRDKYELNVTAMDDG--------SC-CVNGDATIHTSTAVVVVFITDVNDNKPVFKDCG  978

Query  112   IFMGEIEENSASNSLVMILNATDADEPNHLNSKIAFKIVSQEPAGTPMFLLSRNTGEVRT  171
              +  ++EE + + S V+ + ATD D+   +N ++ + IV Q       F +   TGEV T
Sbjct  979   TYYPKVEEGAPNGSPVIKVQATDEDK--GVNGQVKYSIVQQPNQKGTKFTVDEETGEVST  1036

Query  172   LTNSLDRE--QASSYRLVVSGADKDGEGLSTQCECNIKVKDVNDNFPMFRDSQYSARIEE  229
                  DRE        + V   D+    L   C   +++ DVNDN P+F   +Y   +++
Sbjct  1037  -NKVFDREGDDGKFVSVTVKATDQGEPSLEGVCSFTVEITDVNDNPPLFDRQKYVENVKQ  1095

Query  230   NI-LSSELLRFQVTDLDEEYTDNWLAVYFFT---SGNEGNWFEIQTDPRTNEGILKVVKA  285
             +  + + +LR  V+  DE+  +N   VY  +   S N+  +FEIQ  P +   +LK  K
Sbjct  1096  DASIGTNILR--VSASDEDADNNGAIVYTLSAPHSPNDLEYFEIQ--PESGWIVLK--KP  1149

Query  286   LD--YEQLQSVKLSIAVKNK------AEFHQSVISRYRVQSTPVTIQV----INVREGIA  333
             LD  Y + +  +L +   ++      A+    +    R    P+  +     I++RE +
Sbjct  1150  LDGCYSR-ERYRLKVYASDRGDPPSTADVDVELDVVDRNNKPPIWDRTIYGPIHIRENVT  1208

Query  334   FRPASKTFTVQKGISSKKLVDYILGTYQAIDEDTNKAASNVKYVMGRNDGGYLMIDSKTA  393
                   +     GI     V Y L         TNK   +  Y+  R D G        A
Sbjct  1209  VGTVVTSVKASSGIEGNPTVFYRL--MPGSTAQTNKL--HTFYLQQRPDNG-----DTWA  1259

Query  394   EIKFVKNMNRDSTFIVNKTITAEVLAIDEYTGKTSTGTVYVRVPDFNDNCPTAV-LEKDA  452
             +IK    ++ ++    N T+  E     +     S  TVY+++ D ND  P     E++
Sbjct  1260  DIKVNHPLDFETIKEYNLTVRVENNGAQQL---ASEATVYIQLEDVNDEIPLFTEREQET  1316

Query  453   VCSSSP  458
             V    P
Sbjct  1317  VLEGEP  1322

 Score = 75.9 bits (185),  Expect = 2e-12, Method: Compositional matrix adjust.
 Identities = 103/418 (25%), Positives = 174/418 (42%), Gaps = 50/418 (12%)

Query  50    IFVVDKNTGDINITAIVDREETPSFLITCRAL----------NAQGLDVEKPLILTVKIL  99
              F ++  +G+I    + DRE+  ++ +   A           N Q   V K   + + I
Sbjct  1359  FFEINLQSGEIFTKVVFDREKQGAYALEVEARDGAPSARPNSNQQPNSVTK--FIRIGIA  1416

Query  100   DINDNPPVFSQQIFMGEIEENSASNSLVMILNATDADEPNHLNSKIAFKIVSQEPAGTPM  159
             D NDNPP F + ++  E++EN      V+ + A D DE    +S+I ++I S    G
Sbjct  1417  DKNDNPPYFDKALYEAEVDENEDIQHTVLTVTAKDHDE----SSRIRYEITSGNIGGA--  1470

Query  160   FLLSRNTGEVRTLTNSLDREQASSYRLVVSGADKDGEGLSTQCECNIKVKDVNDNFPMFR  219
             F +   TG +  +  +LD E    Y L +  +D   E  +      I VKDVND  P F
Sbjct  1471  FAVKNMTGAI-YVAGALDYETRKRYYLTLVASDSLNENETIIV---INVKDVNDMPPSFP  1526

Query  220   DSQYSARIEENI-LSSELLRFQVTDLDEEYTDNWLAVYFFTSG--NEGNWFEIQTDPRTN  276
                Y   ++E + +   +++   TD D++   N   VYF T    +  N    + D
Sbjct  1527  QQVYERTMDEELAVPFPIMQVTATDGDKDRPQN--IVYFLTGQGIDPDNPANSKFDINRT  1584

Query  277   EGILKVVKALDYEQLQS-VKLSIAVKNKAEFHQSVISRYRVQSTPVTIQVINVREGIAFR  335
              G + V+K LD +Q     +    V  + E  + ++    VQ     + + ++ +
Sbjct  1585  TGEIFVLKPLDRDQPNGRPQWRFTVFAQDEGGEGLVGYADVQ-----VNLKDINDNAPIF  1639

Query  336   PASKTFTVQKGISSKKLVDYILGTYQAIDEDTNKAASNVK--YVMGRN------DGGYLM  387
             P    F     ++       ++ T  A+D D     +N K  Y + +N
Sbjct  1640  PQGVYF---GNVTENGTAGMVVMTMTAVDYDDPNEGTNAKLIYSIEKNVIEEETGSPIFE  1696

Query  388   IDSKTAEIK-FVKNMNRDSTFIVNKTITAEVLAIDEYTGKTSTGTVYVRVPDFNDNCP  444
```

```
             I++ T  IK  V  ++R+ T       + +V+A+D   G   TGT  +RV D ND  P
Sbjct  1697  IEADTGVIKTAVCCLDRERT----PDYSIQVVAMDG-GGLKGTGTASIRVKDINDMPP  1749

 Score = 60.5 bits (145),  Expect = 1e-07, Method: Compositional matrix adjust.
 Identities = 83/362 (23%), Positives = 152/362 (42%), Gaps = 34/362 (9%)

Query  51    FVVDKNTGDINITAIVDRE-ETPSFL-ITCRALNAQGLDVEKPLILTVKILDINDNPPVF  108
             F VD+ TG+++    + DRE +    F+ +T +A +      +E      TV+I D+NDNPP+F
Sbjct  1025  FTVDEETGEVSTNKVFDREGDDGKFVSVTVKATDQGEPSLEGVCSFTVEITDVNDNPPLF  1084

Query  109   SQQIFMGEIEENSASNSLVMILNATDADEPNHLNSKIAFKIVS-QEPAGTPMFLLSRNTG  167
              +Q ++  ++++++  + ++ ++A+D D  N  N  I + + +   P     F +   +G
Sbjct  1085  DRQKYVENVKQDASIGTNILRVSASDEDADN--NGAIVYTLSAPHSPNDLEYFEIQPESG  1142

Query  168   EV---RTLTNSLDREQASSYRLVVSGADKDGEGLSTQCECNIKVKDVNDNFPMFRDSQYS  224
              +   + L     RE+   YRL V  +D+ G+  ST             +N P   D
Sbjct  1143  WIVLKKPLDGCYSRER---YRLKVYASDR-GDPPSTADVDVELDVVDRNNKPPIWDRTIY  1198

Query  225   ARIEENILSSELLRFQVTDLDEEYTDNWLAVYFFTSG-----NEGNWFEIQTDPRTNE--  277
               I      +                   N    Y    G     N+ + F +Q   P   +
Sbjct  1199  GPIHIRENVTVGTVVTSVKASSGIEGNPTVFYRLMPGSTAQTNKLHTFYLQQRPDNGDTW  1258

Query  278   GILKVVKALDYEQLQSVKLSIAVKNKAEFHQSVISRYRVQSTPVTIQVINVREGIAFRPA  337
               +KV   LD+E ++   L++ V+N          ++       V IQ+ +V + I
Sbjct  1259  ADIKVNHPLDFETIKEYNLTVRVENNG-------AQQLASEATVYIQLEDVNDEIPLFTE  1311

Query  338   SKTFTVQKGISSKKLVDYILGTYQAIDEDTNKAASNVKYVM---GRNDG-GYLMIDSKTA  393
              +  TV +G      V  +     AID+D      + V Y +    RN+G  +  I+ ++
Sbjct  1312  REQETVLEGEPIGTKVTQV----NAIDKDGTFPNNQVYYYIVDSPRNEGKDFFEINLQSG  1367

Query  394   EI  395
             EI
Sbjct  1368  EI  1369

 Score = 55.8 bits (133),  Expect = 3e-06, Method: Compositional matrix adjust.
 Identities = 49/158 (31%), Positives = 78/158 (49%), Gaps = 24/158 (15%)

Query  102   NDNPPVFSQQIFMGEIEENSASNSLVMILNATDADEPNHLNSKIAFKIVSQEPAGTP--M  159
             ND  P FSQQ++   ++EN+  N+LV  + A+D D  N     + F  V     GT
Sbjct  858   NDEEPKFSQQVYTPNVDENAGPNTLVTTVVASDKDGDN-----VRFGFVG---GGTSSGQ  909

Query  160   FLLSRNTGEVRTLTN--SLDREQASSYRLVVSGADK-------DGEGLSTQCECNIKVKD  210
             F++   TG +R  +   SLDR++   Y L V+  D         D    ++      + + D
Sbjct  910   FVIEEITGVIRLHSKAISLDRDK---YELNVTAMDDGSCCVNGDATIHTSTAVVVVFITD  966

Query  211   VNDNFPMFRD-SQYSARIEENILS-SELLRFQVTDLDE  246
             VNDN P+F+D   Y  ++EE   + S +++ Q TD D+
Sbjct  967   VNDNKPVFKDCGTYYPKVEEGAPNGSPVIKVQATDEDK  1004

 Score = 53.9 bits (128),  Expect = 1e-05, Method: Compositional matrix adjust.
 Identities = 70/306 (23%), Positives = 126/306 (41%), Gaps = 30/306 (10%)

Query  20    PIAKIT-SDYQATQKITYRI---SGVGIDQPPFGIFVVDKNTGDINITAIVDRE---ETP  72
             PI  +T  D   T K  Y++   SG G D+  F +   +  TG + I   +D E   ++
Sbjct  1772  PILTVTVHDEDETNKFQYKVIDNSGYGADK--FTMVRNNDGTGSLKIVQPLDYEDPLQSN  1829

Query  73    SFLITCRALNAQGLDVEKPLI------LTVKILDINDNPPVFSQQIFMGEIEENSASNSL  126
              F    + +N +G D +           + VK+ DINDN P F +      + EN+
Sbjct  1830  GFRFRIQ-VNDKGEDNDNDKYHVAYSWVVVKLRDINDNKPQFERPNIEVSVYENADVGKT  1888

Query  127   VMILNATDADEPNHLNSKIAFKIVSQEPAGTPMFLLSRNTGEVRTLTNSLDREQASSYRL  186
             +    ATD D+     SK+++ I   + +       S N     T+  +LDRE    +++
Sbjct  1889  LETFKATDPDQGG--KSKVSYAI---DRSSDRQRQFSINQEGTVTIQRNLDREVTPRHQV  1943

Query  187   VVSGADKDGEGLSTQCECNIKVKDVNDNFPMFRDSQYSARIEENILSSELLRFQVTDLDE  246
              +   D      +      + V+D+NDN P F    Y   + E++    +++    TD D+
Sbjct  1944  KILAIDDGIPPKTATATLTVIVQDINDNPPKFL-KDYRPVLPEHVPPRKVVEILATDDDD  2002
```

```
Query  247   EYTDNWLAVYFFTSGNEGNW----FEIQTDPRTNEG----ILKVVKALDYEQLQSVKLSI  298
                 N     F       +     F+++ D +   G    I+  +++ D EQ +   + I
Sbjct  2003  RSKSNGPPFQFRLDPGADDIIRASFKVEQDQKGANGDGMAIVSSLRSFDREQQKEYLIPI  2062

Query  299   AVKNKA  304
              +K+
Sbjct  2063  VIKDHG  2068

 Score = 52.0 bits (123),  Expect = 4e-05, Method: Compositional matrix adjust.
 Identities = 97/398 (24%), Positives = 163/398 (41%), Gaps = 54/398 (14%)

Query  106   PVFSQQIFMGEIEENSASNSLVMILNATDADEPNHLNSKIAFKIVSQ-EPAGTPMFLLSR  164
             P F    +  EI EN   +S ++ + A      +  + +I + + +Q + AGT  F +
Sbjct  655   PQFYMPSYEAEIPENQKKDSDIISVKAK-----SFADREIRYTLKAQGQGAGT--FNIGP  707

Query  165   NTGEVRTLTNSLDRE---QASSYRLVVSGADKDGEGLSTQCECNIKVKDVNDNFPMFRDS  221
              +G V+ L   LD E   Q   Y LVV+ A +D  G ST  E  I+V DVNDN P F
Sbjct  708   TSGIVK-LAKELDFEDLRQPHVYSLVVT-ATEDSGGFSTSVELTIRVTDVNDNAPKFELP  765

Query  222   QYSAR-IEENI-LSSELLRFQVTDLDEEYTDNWLAVYFFTSGNEGNWFEIQTDPRTNEGI  279
              Y A  ++E+I L + +L+ +  D D         + +  S +    F + ++     GI
Sbjct  766   DYQAHNVDEDIPLGTSILKVKAMDSDSGAN---AEIEYLVSDDH---FAVDSN-----GI  814

Query  280   LKVVKALDYEQLQS-VKLSIAVKNKAEFHQSVISRYRVQSTPVTIQVINVREGIAFRPAS  338
             +   K LD +   +  +  +  K+K E  ++  +  RV +           E   F
Sbjct  815   IVNNKQLDADNNNAYYEFVVTAKDKGEPAKTGTATVRVYTKNKN------DEEPKFSQQV  868

Query  339   KTFTVQKGISSKKLVDYILGTYQAIDEDTNKAASNVK--YVMGRNDGGYLMIDSKTAEIK  396
              T  V +      LV  ++         ++K   NV+  +V G    G  +I+  T  I+
Sbjct  869   YTPNVDENAGPNTLVTTVVA--------SDKDGDNVRFGFVGGGTSSGQFVIEEITGVIR  920

Query  397   FVK---NMNRDSTFI-VNKTITAEVLAIDEYTGKTSTGTVYVRVPDFNDN------CPTA  446
                   +++RD   + V            + T  TST  V V + D NDN      C T
Sbjct  921   LHSKAISLDRDKYELNVTAMDDGSCCVNGDATIHTSTAVVVVFITDVNDNKPVFKDCGTY  980

Query  447   VLE-KDAVCSSSPSVVVSARTLNNRYTGPYTFALEDQP  483
               + ++   + SP + V A   +    G   +++  QP
Sbjct  981   YPKVEEGAPNGSPVIKVQATDEDKGVNGQVKYSIVQQP  1018

 Score = 44.3 bits (103),  Expect = 0.009, Method: Compositional matrix adjust.
 Identities = 48/199 (24%), Positives = 73/199 (37%), Gaps = 19/199 (10%)

Query  27    DYQATQKITYRISGVGIDQPPFGIFVVDKNTGDINITAIVDREETPSFLITCRALNAQGL  86
             D     K++Y I      Q  F I       G + I   +DRE TP   +   A++
Sbjct  1898  DQGGKSKVSYAIDRSSDRQRQFSI----NQEGTVTIQRNLDREVTPRHQVKILAIDDGIP  1953

Query  87    DVEKPLILTVKILDINDNPPVFSQQIFMGEIEENSASNSLVMILNATDADEPNHLNSKIA  146
                    LTV + DINDNPP F +  +   + E+     +V IL   D D
Sbjct  1954  PKTATATLTVIVQDINDNPPKFLKD-YRPVLPEHVPPRKVVEILATDDDDRSKSNGPPFQ  2012

Query  147   FKIVSQEPAGTPMFLLSRNTGEVRTLTN-----------SLDREQASSYRLVVSGADKDG  195
             F++   +P    +   S    + +   N           S DREQ   Y + +   D
Sbjct  2013  FRL---DPGADDIIRASFKVEQDQKGANGDGMAIVSSLRSFDREQQKEYLIPIVIKDHGN  2069

Query  196   EGLSTQCECNIKVKDVNDN  214
               ++      + + DVNDN
Sbjct  2070  PAMTGTSTLTVVIGDVNDN  2088

 Score = 39.7 bits (91),  Expect = 0.28, Method: Compositional matrix adjust.
 Identities = 89/389 (23%), Positives = 149/389 (38%), Gaps = 62/389 (16%)

Query  75   LITCRALNAQGLDVEKPLILTVKILDINDNPPVFSQQIF--MGEIEENSASNSLVMILNA  132
            +IT +  NA GL       + + + D+ND PP F  +       ++ N+  N+ V  L A
Sbjct  517  IITNQGHNA-GLKYTDNQRVIIHVKDVNDEPPYFINRPLPMQAVVQLNAPPNTPVFTLQA  575

Query  133  TDADEPNHLNSKIAFKIVSQEPAGTPMFLLSRNTGEVRTLTNSLDREQASSYRLVVSGAD  192
             D D     +  I + IV     G   F +   +G VRT    L  +    Y L V   D
Sbjct  576  RDPD----TDHNIHYFIVRDRTGG--RFEVDERSGVVRTRGTDL-FQLDMEYVLYVKAED  628
```

```
Query  193  KDGE----GLSTQCECNIKVKDVNDNFPMFRDSQYSARIEEN-ILSSELLRFQVTDLDEE  247
            ++G+       +  E  + +       P F    Y A I EN    S+++  +     +
Sbjct  629  QNGKVDDRRFQSTPEERLSIVG-GKRAPQFYMPSYEAEIPENQKKDSDIISVKA----KS  683

Query  248  YTDNWLAVYFFTSGNEGNWFEIQTDPRTNEGILKVVKALDYEQLQSVKLS----IAVKNK  303
            + D  +       G     F I        GI+K+ K LD+E L+   +      A ++
Sbjct  684  FADREIRYTLKAQGQGAGTFNIGP----TSGIVKLAKELDFEDLRQPHVYSLVVTATEDS  739

Query  304  AEFHQSVISRYRVQSTPVTIQVINVREGIAFRPASKTFTVQKGISSKKLVDYILGT----  359
              F  SV          +TI+V +V +       +  F +    +     D  LGT
Sbjct  740  GGFSTSV---------ELTIRVTDVNDN------APKFELPDYQAHNVDEDIPLGTSILK  784

Query  360  YQAIDEDTNKAASNVKYVMGRN----DGGYLMIDSKTAEIKFVKNMNRDSTFIVNKTITA  415
             +A+D D+  A + ++Y++  +    D   +++++K  +     N N    F+V
Sbjct  785  VKAMDSDSG-ANAEIEYLVSDDHFAVDSNGIIVNNKQLD---ADNNNAYYEFVVTAKDKG  840

Query  416  EVLAIDEYTGKTSTGTVYVRVPDFNDNCP  444
            E         KT T TV V   + ND  P
Sbjct  841  E-------PAKTGTATVRVYTKNKNDEEP  862

 Score = 39.3 bits (90),  Expect = 0.32, Method: Compositional matrix adjust.
 Identities = 54/242 (22%), Positives = 102/242 (42%), Gaps = 22/242 (9%)

Query  4    KFAKPCREGE--DNSKRNP-IAKITSDYQATQKITYRISGVGIDQPPFGIFVVDKNTGDI  60
            +F  P  E E  +N K++  I  + +   A ++I Y +   G      G F +   +G +
Sbjct  656  QFYMPSYEAEIPENQKKDSDIISVKAKSFADREIRYTLKAQGQGA---GTFNIGPTSGIV  712

Query  61   NITAIVDREE--TPSF--LITCRALNAQGLDVEKPLILTVKILDINDNPPVFSQQIFMGE  116
             +   +D E+   P    L+     ++ G      + LT+++ D+NDN P F    +
Sbjct  713  KLAKELDFEDLRQPHVYSLVVTATEDSGGFSTS--VELTIRVTDVNDNAPKFELPDYQAH  770

Query  117  -IEENSASNSLVMILNATDADEPNHLNSKIAFKIVSQEPAGTPMFLLSRNTGEVRTLTNS  175
             ++E+    + ++ + A D+D  +  N++I + +VS +      F +  N   V
Sbjct  771  NVDEDIPLGTSILKVKAMDSD--SGANAEIEY-LVSDD-----HFAVDSNGIIVNNKQLD  822

Query  176  LDREQASSYRLVVSGADKDGEGLSTQCECNIKVKDVNDNFPMFRDSQYSARIEENILSSE  235
             D   A  Y  VV+  DK     +      +  K+ ND  P F    Y+  ++EN   +
Sbjct  823  ADNNNA-YYEFVVTAKDKGEPAKTGTATVRVYTKNKNDEEPKFSQQVYTPNVDENAGPNT  881

Query  236  LL  237
            L+
Sbjct  882  LV  883

>ACMO_007424.P13508 | transcript=ACMO_007424.R13508 | gene=ACMO_007424
| organism=Anopheles_coluzzii_MOPTI | gene_product=neural-cadherin,
transcript variant X4 | transcript_product=neural-cadherin,
transcript variant X7 | location=CM029349:40739311-40945968(+)
| protein_length=3103 | sequence_SO=chromosome
| SO=protein_coding_gene | is_pseudo=false
Length=3103

 Score = 147 bits (370),  Expect = 3e-35, Method: Compositional matrix adjust.
 Identities = 128/446 (29%), Positives = 217/446 (49%), Gaps = 36/446 (8%)

Query  20    PIAKITS---DYQATQKITYRISGVGID--QPPFGIFVVDKNTGDINITAIVDREET---  71
             PI ++T+   D    Q I Y ++G GID   P    F +++ TG+I +   +DR++
Sbjct  1543  PIMQVTATDGDKDRPQNIVYFLTGQGIDPDNPANSKFDINRTTGEIFVLKPLDRDQPNGR  1602

Query  72    PSFLITCRALNAQGLDVEKPLILTVKILDINDNPPVFSQQIFMGEIEENSASNSLVMILN  131
             P +  T  A +  G  +     + V + DINDN P+F Q ++ G + EN  +  +VM +
Sbjct  1603  PQWRFTVFAQDEGGEGLVGYADVQVNLKDINDNAPIFPQGVYFGNVTENGTAGMVVMTMT  1662

Query  132   ATDADEPNH-LNSKIAFKI---VSQEPAGTPMFLLSRNTGEVRTLTNSLDREQASSYRLV  187
             A D D+PN   N+K+ + I   V +E  G+P+F +  +TG ++T    LDRE+   Y +
Sbjct  1663  AVDYDDPNEGTNAKLIYSIEKNVIEEETGSPIFEIEADTGVIKTAVCCLDRERTPDYSIQ  1722

Query  188   VSGADKDGEGLSTQCECNIKVKDVNDNFPMFRDSQYSARIEENILSS--ELLRFQVTDLD  245
             V     DG GL       +I+VKD+ND  P F   ++   ++E   ++  E+    VT  D
```

```
Sbjct  1723  VVAM--DGGGLKGTGTASIRVKDINDMPPQFTKDEWFTEVDETDGTNLPEMPILTVTVHD  1780

Query  246   EEYTDNWLAVYFFTSGNEGNWFEIQTDPRTNEGI--LKVVKALDYE---QLQSVKLSIAV  300
             E+ T+ +       SG   + F +    R N+G   LK+V+ LDYE   Q    +  I V
Sbjct  1781  EDETNKFQYKVIDNSGYGADKFTMV---RNNDGTGSLKIVQPLDYEDPLQSNGFRFRIQV  1837

Query  301   KNKAEFHQSVISRYRVQSTPVTIQVINVREGIAFRPASKTFTVQKGISSKKLVDYILGTY  360
              +K E + +   +Y V  + V +++ ++ +    +P  +   ++  +     V   L T+
Sbjct  1838  NDKGEDNDN--DKYHVAYSWVVVKLRDINDN---KPQFERPNIEVSVYENADVGKTLETF  1892

Query  361   QAIDEDTNKAASNVKYVMGR-NDGGYLMIDSKTAEIKFVKNMNRDSTFIVNKTITAEVLA  419
             +A D D     S V Y + R +D       ++   +   +N++R+    V      ++LA
Sbjct  1893  KATDPDQG-GKSKVSYAIDRSSDRQRQFSINQEGTVTIQRNLDRE----VTPRHQVKILA  1947

Query  420   IDE-YTGKTSTGTVYVRVPDFNDNCP  444
             ID+    KT+T T+ V V D NDN P
Sbjct  1948  IDDGIPPKTATATLTVIVQDINDNPP  1973

 Score = 75.9 bits (185),  Expect = 2e-12, Method: Compositional matrix adjust.
 Identities = 106/426 (25%), Positives = 181/426 (42%), Gaps = 51/426 (12%)

Query  53    VDKNTGDINITAIVDREETPSFLITCRALNAQGLDVEKPLILTVKILDINDNPPVFSQ-Q  111
             +D++  ++N+TA+ D         +C  +N          ++ V I D+NDN PVF
Sbjct  928   LDRDKYELNVTAMDDG--------SC-CVNGDATIHTSTAVVVVFITDVNDNKPVFKDCG  978

Query  112   IFMGEIEENSASNSLVMILNATDADEPNHLNSKIAFKIVSQEPAGTPMFLLSRNTGEVRT  171
              +  ++EE + + S V+ + ATD D+   +N ++ + IV Q       F +   TGEV T
Sbjct  979   TYYPKVEEGAPNGSPVIKVQATDEDK--GVNGQVKYSIVQQPNQKGTKFTVDEETGEVST  1036

Query  172   LTNSLDRE--QASSYRLVVSGADKDGEGLSTQCECNIKVKDVNDNFPMFRDSQYSARIEE  229
                  DRE        + V   D+    L   C   +++ DVNDN P+F   +Y   +++
Sbjct  1037  -NKVFDREGDDGKFVSVTVKATDQGEPSLEGVCSFTVEITDVNDNPPLFDRQKYVENVKQ  1095

Query  230   NI-LSSELLRFQVTDLDEEYTDNWLAVYFFT---SGNEGNWFEIQTDPRTNEGILKVVKA  285
             +  + + +LR  V+  DE+  +N   VY  +   S N+  +FEIQ  P +   +LK  K
Sbjct  1096  DASIGTNILR--VSASDEDADNNGAIVYTLSAPHSPNDLEYFEIQ--PESGWIVLK--KP  1149

Query  286   LD--YEQLQSVKLSIAVKNK------AEFHQSVISRYRVQSTPVTIQV----INVREGIA  333
             LD  Y + +  +L +   ++      A+    +    R    P+  +     I++RE +
Sbjct  1150  LDGCYSR-ERYRLKVYASDRGDPPSTADVDVELDVVDRNNKPPIWDRTIYGPIHIRENVT  1208

Query  334   FRPASKTFTVQKGISSKKLVDYILGTYQAIDEDTNKAASNVKYVMGRNDGGYLMIDSKTA  393
                   +     GI     V Y L         TNK   +  Y+  R D G        A
Sbjct  1209  VGTVVTSVKASSGIEGNPTVFYRL--MPGSTAQTNKL--HTFYLQQRPDNG-----DTWA  1259

Query  394   EIKFVKNMNRDSTFIVNKTITAEVLAIDEYTGKTSTGTVYVRVPDFNDNCPTAV-LEKDA  452
             +IK    ++ ++     N T+  E     +     S  TVY+++ D ND  P     E++
Sbjct  1260  DIKVNHPLDFETIKEYNLTVRVENNGAQQL---ASEATVYIQLEDVNDEIPLFTEREQET  1316

Query  453   VCSSSP  458
             V    P
Sbjct  1317  VLEGEP  1322

 Score = 75.9 bits (185),  Expect = 2e-12, Method: Compositional matrix adjust.
 Identities = 103/418 (25%), Positives = 174/418 (42%), Gaps = 50/418 (12%)

Query  50    IFVVDKNTGDINITAIVDREETPSFLITCRAL----------NAQGLDVEKPLILTVKIL  99
              F ++  +G+I    + DRE+  ++ +   A           N Q   V K   + + I
Sbjct  1359  FFEINLQSGEIFTKVVFDREKQGAYALEVEARDGAPSARPNSNQQPNSVTK--FIRIGIA  1416

Query  100   DINDNPPVFSQQIFMGEIEENSASNSLVMILNATDADEPNHLNSKIAFKIVSQEPAGTPM  159
             D NDNPP F + ++  E++EN      V+ + A D DE    +S+I ++I S    G
Sbjct  1417  DKNDNPPYFDKALYEAEVDENEDIQHTVLTVTAKDHDE----SSRIRYEITSGNIGGA--  1470

Query  160   FLLSRNTGEVRTLTNSLDREQASSYRLVVSGADKDGEGLSTQCECNIKVKDVNDNFPMFR  219
             F +   TG +  +  +LD E    Y L +  +D   E  +       I VKDVND  P F
Sbjct  1471  FAVKNMTGAI-YVAGALDYETRKRYYLTLVASDSLNENETIIV---INVKDVNDMPPSFP  1526

Query  220   DSQYSARIEENI-LSSELLRFQVTDLDEEYTDNWLAVYFFTSG--NEGNWFEIQTDPRTN  276
```

```
                Y   ++E + +   +++   TD D++   N   VYF T    +  N    + D
Sbjct  1527  QQVYERTMDEELAVPFPIMQVTATDGDKDRPQN--IVYFLTGQGIDPDNPANSKFDINRT  1584

Query  277   EGILKVVKALDYEQLQS-VKLSIAVKNKAEFHQSVISRYRVQSTPVTIQVINVREGIAFR  335
              G + V+K LD +Q      +    V  + E  + ++    VQ     + + ++ +
Sbjct  1585  TGEIFVLKPLDRDQPNGRPQWRFTVFAQDEGGEGLVGYADVQ-----VNLKDINDNAPIF  1639

Query  336   PASKTFTVQKGISSKKLVDYILGTYQAIDEDTNKAASNVK--YVMGRN------DGGYLM  387
             P    F     ++        ++ T  A+D D     +N K  Y + +N
Sbjct  1640  PQGVYF---GNVTENGTAGMVVMTMTAVDYDDPNEGTNAKLIYSIEKNVIEEETGSPIFE  1696

Query  388   IDSKTAEIK-FVKNMNRDSTFIVNKTITAEVLAIDEYTGKTSTGTVYVRVPDFNDNCP  444
             I++ T  IK  V  ++R+ T       + +V+A+D   G   TGT  +RV D ND  P
Sbjct  1697  IEADTGVIKTAVCCLDRERT----PDYSIQVVAMDG-GGLKGTGTASIRVKDINDMPP  1749

 Score = 60.5 bits (145),  Expect = 1e-07, Method: Compositional matrix adjust.
 Identities = 83/362 (23%), Positives = 152/362 (42%), Gaps = 34/362 (9%)

Query  51    FVVDKNTGDINITAIVDRE-ETPSFL-ITCRALNAQGLDVEKPLILTVKILDINDNPPVF  108
             F VD+ TG+++   + DRE +   F+ +T +A +      +E      TV+I D+NDNPP+F
Sbjct  1025  FTVDEETGEVSTNKVFDREGDDGKFVSVTVKATDQGEPSLEGVCSFTVEITDVNDNPPLF  1084

Query  109   SQQIFMGEIEENSASNSLVMILNATDADEPNHLNSKIAFKIVS-QEPAGTPMFLLSRNTG  167
              +Q ++  ++++++  + ++ ++A+D D  N  N  I + + +   P     F +   +G
Sbjct  1085  DRQKYVENVKQDASIGTNILRVSASDEDADN--NGAIVYTLSAPHSPNDLEYFEIQPESG  1142

Query  168   EV---RTLTNSLDREQASSYRLVVSGADKDGEGLSTQCECNIKVKDVNDNFPMFRDSQYS  224
              +   + L     RE+   YRL V  +D+ G+  ST              +N P   D
Sbjct  1143  WIVLKKPLDGCYSRER---YRLKVYASDR-GDPPSTADVDVELDVVDRNNKPPIWDRTIY  1198

Query  225   ARIEENILSSELLRFQVTDLDEEYTDNWLAVYFFTSG-----NEGNWFEIQTDPRTNE--  277
               I      +                  N    Y    G     N+ + F +Q  P   +
Sbjct  1199  GPIHIRENVTVGTVVTSVKASSGIEGNPTVFYRLMPGSTAQTNKLHTFYLQQRPDNGDTW  1258

Query  278   GILKVVKALDYEQLQSVKLSIAVKNKAEFHQSVISRYRVQSTPVTIQVINVREGIAFRPA  337
               +KV   LD+E ++   L++ V+N          ++        V IQ+ +V + I
Sbjct  1259  ADIKVNHPLDFETIKEYNLTVRVENNG-------AQQLASEATVYIQLEDVNDEIPLFTE  1311

Query  338   SKTFTVQKGISSKKLVDYILGTYQAIDEDTNKAASNVKYVM---GRNDG-GYLMIDSKTA  393
              +  TV +G       V  +     AID+D     + V Y +    RN+G  +  I+ ++
Sbjct  1312  REQETVLEGEPIGTKVTQV----NAIDKDGTFPNNQVYYYIVDSPRNEGKDFFEINLQSG  1367

Query  394   EI  395
             EI
Sbjct  1368  EI  1369

 Score = 55.8 bits (133),  Expect = 3e-06, Method: Compositional matrix adjust.
 Identities = 49/158 (31%), Positives = 78/158 (49%), Gaps = 24/158 (15%)

Query  102   NDNPPVFSQQIFMGEIEENSASNSLVMILNATDADEPNHLNSKIAFKIVSQEPAGTP--M  159
             ND  P FSQQ++   ++EN+  N+LV  + A+D D  N     + F  V     GT
Sbjct  858   NDEEPKFSQQVYTPNVDENAGPNTLVTTVVASDKDGDN-----VRFGFVG---GGTSSGQ  909

Query  160   FLLSRNTGEVRTLTN--SLDREQASSYRLVVSGADK-------DGEGLSTQCECNIKVKD  210
             F++   TG +R  +   SLDR++    Y L V+  D         D    ++      + + D
Sbjct  910   FVIEEITGVIRLHSKAISLDRDK---YELNVTAMDDGSCCVNGDATIHTSTAVVVVFITD  966

Query  211   VNDNFPMFRD-SQYSARIEENILS-SELLRFQVTDLDE  246
             VNDN P+F+D   Y  ++EE   + S +++ Q TD D+
Sbjct  967   VNDNKPVFKDCGTYYPKVEEGAPNGSPVIKVQATDEDK  1004

 Score = 53.5 bits (127),  Expect = 1e-05, Method: Compositional matrix adjust.
 Identities = 70/306 (23%), Positives = 126/306 (41%), Gaps = 30/306 (10%)

Query  20    PIAKIT-SDYQATQKITYRI---SGVGIDQPPFGIFVVDKNTGDINITAIVDRE---ETP  72
             PI  +T  D   T K  Y++   SG G D+  F +    +  TG + I   +D E   ++
Sbjct  1772  PILTVTVHDEDETNKFQYKVIDNSGYGADK--FTMVRNNDGTGSLKIVQPLDYEDPLQSN  1829
```

```
Query  73    SFLITCRALNAQGLDVEKPLI------LTVKILDINDNPPVFSQQIFMGEIEENSASNSL  126
              F    + +N +G D +          + VK+ DINDN P F +      + EN+
Sbjct  1830  GFRFRIQ-VNDKGEDNDNDKYHVAYSWVVVKLRDINDNKPQFERPNIEVSVYENADVGKT  1888

Query  127   VMILNATDADEPNHLNSKIAFKIVSQEPAGTPMFLLSRNTGEVRTLTNSLDREQASSYRL  186
             +    ATD D+     SK+++ I   + +       S N     T+  +LDRE    +++
Sbjct  1889  LETFKATDPDQGG--KSKVSYAI---DRSSDRQRQFSINQEGTVTIQRNLDREVTPRHQV  1943

Query  187   VVSGADKDGEGLSTQCECNIKVKDVNDNFPMFRDSQYSARIEENILSSELLRFQVTDLDE  246
              +   D      +      + V+D+NDN P F    Y   + E++   +++    TD D+
Sbjct  1944  KILAIDDGIPPKTATATLTVIVQDINDNPPKFL-KDYRPVLPEHVPPRKVVEILATDDDD  2002

Query  247   EYTDNWLAVYFFTSGNEGNW----FEIQTDPRTNEG----ILKVVKALDYEQLQSVKLSI  298
                 N     F       +     F+++ D +   G    I+  +++ D EQ +   + I
Sbjct  2003  RSKSNGPPFQFRLDPGADDIIRASFKVEQDQKGANGDGMAIVSSLRSFDREQQKEYLIPI  2062

Query  299   AVKNKA  304
              +K+
Sbjct  2063  VIKDHG  2068

 Score = 52.0 bits (123),  Expect = 4e-05, Method: Compositional matrix adjust.
 Identities = 97/398 (24%), Positives = 163/398 (41%), Gaps = 54/398 (14%)

Query  106   PVFSQQIFMGEIEENSASNSLVMILNATDADEPNHLNSKIAFKIVSQ-EPAGTPMFLLSR  164
             P F    +  EI EN   +S ++ + A      +  + +I + + +Q + AGT  F +
Sbjct  655   PQFYMPSYEAEIPENQKKDSDIISVKAK-----SFADREIRYTLKAQGQGAGT--FNIGP  707

Query  165   NTGEVRTLTNSLDRE---QASSYRLVVSGADKDGEGLSTQCECNIKVKDVNDNFPMFRDS  221
              +G V+ L   LD E   Q   Y LVV+ A +D  G ST  E  I+V DVNDN P F
Sbjct  708   TSGIVK-LAKELDFEDLRQPHVYSLVVT-ATEDSGGFSTSVELTIRVTDVNDNAPKFELP  765

Query  222   QYSAR-IEENI-LSSELLRFQVTDLDEEYTDNWLAVYFFTSGNEGNWFEIQTDPRTNEGI  279
              Y A  ++E+I L + +L+ +  D D         + +  S +    F + ++     GI
Sbjct  766   DYQAHNVDEDIPLGTSILKVKAMDSDSGAN---AEIEYLVSDDH---FAVDSN-----GI  814

Query  280   LKVVKALDYEQLQS-VKLSIAVKNKAEFHQSVISRYRVQSTPVTIQVINVREGIAFRPAS  338
             +   K LD +   +  +  +  K+K E  ++  +  RV +           E   F
Sbjct  815   IVNNKQLDADNNNAYYEFVVTAKDKGEPAKTGTATVRVYTKNKN------DEEPKFSQQV  868

Query  339   KTFTVQKGISSKKLVDYILGTYQAIDEDTNKAASNVK--YVMGRNDGGYLMIDSKTAEIK  396
              T  V +      LV  ++         ++K   NV+  +V G    G  +I+  T  I+
Sbjct  869   YTPNVDENAGPNTLVTTVVA--------SDKDGDNVRFGFVGGGTSSGQFVIEEITGVIR  920

Query  397   FVK---NMNRDSTFI-VNKTITAEVLAIDEYTGKTSTGTVYVRVPDFNDN------CPTA  446
                   +++RD   + V            + T  TST  V V + D NDN      C T
Sbjct  921   LHSKAISLDRDKYELNVTAMDDGSCCVNGDATIHTSTAVVVVFITDVNDNKPVFKDCGTY  980

Query  447   VLE-KDAVCSSSPSVVVSARTLNNRYTGPYTFALEDQP  483
               + ++   + SP + V A   +    G   +++  QP
Sbjct  981   YPKVEEGAPNGSPVIKVQATDEDKGVNGQVKYSIVQQP  1018

 Score = 44.3 bits (103),  Expect = 0.009, Method: Compositional matrix adjust.
 Identities = 48/199 (24%), Positives = 73/199 (37%), Gaps = 19/199 (10%)

Query  27    DYQATQKITYRISGVGIDQPPFGIFVVDKNTGDINITAIVDREETPSFLITCRALNAQGL  86
             D     K++Y I      Q  F I       G + I   +DRE TP   +   A++
Sbjct  1898  DQGGKSKVSYAIDRSSDRQRQFSI----NQEGTVTIQRNLDREVTPRHQVKILAIDDGIP  1953

Query  87    DVEKPLILTVKILDINDNPPVFSQQIFMGEIEENSASNSLVMILNATDADEPNHLNSKIA  146
                    LTV + DINDNPP F +  +   + E+     +V IL   D D
Sbjct  1954  PKTATATLTVIVQDINDNPPKFLKD-YRPVLPEHVPPRKVVEILATDDDDRSKSNGPPFQ  2012

Query  147   FKIVSQEPAGTPMFLLSRNTGEVRTLTN-----------SLDREQASSYRLVVSGADKDG  195
             F++   +P    +   S    + +   N           S DREQ   Y + +   D
Sbjct  2013  FRL---DPGADDIIRASFKVEQDQKGANGDGMAIVSSLRSFDREQQKEYLIPIVIKDHGN  2069

Query  196   EGLSTQCECNIKVKDVNDN  214
               ++      + + DVNDN
Sbjct  2070  PAMTGTSTLTVVIGDVNDN  2088
```

```
 Score = 39.7 bits (91),  Expect = 0.29, Method: Compositional matrix adjust.
 Identities = 89/389 (23%), Positives = 149/389 (38%), Gaps = 62/389 (16%)

Query  75   LITCRALNAQGLDVEKPLILTVKILDINDNPPVFSQQIF--MGEIEENSASNSLVMILNA  132
            +IT +  NA GL       + + + D+ND PP F  +       ++ N+  N+ V  L A
Sbjct  517  IITNQGHNA-GLKYTDNQRVIIHVKDVNDEPPYFINRPLPMQAVVQLNAPPNTPVFTLQA  575

Query  133  TDADEPNHLNSKIAFKIVSQEPAGTPMFLLSRNTGEVRTLTNSLDREQASSYRLVVSGAD  192
             D D     +  I + IV     G   F +   +G VRT    L  +    Y L V   D
Sbjct  576  RDPD----TDHNIHYFIVRDRTGG--RFEVDERSGVVRTRGTDL-FQLDMEYVLYVKAED  628

Query  193  KDGE----GLSTQCECNIKVKDVNDNFPMFRDSQYSARIEEN-ILSSELLRFQVTDLDEE  247
            ++G+       +  E  + +       P F    Y A I EN    S+++  +     +
Sbjct  629  QNGKVDDRRFQSTPEERLSIVG-GKRAPQFYMPSYEAEIPENQKKDSDIISVKA----KS  683

Query  248  YTDNWLAVYFFTSGNEGNWFEIQTDPRTNEGILKVVKALDYEQLQSVKLS----IAVKNK  303
            + D  +       G     F I        GI+K+ K LD+E L+   +      A ++
Sbjct  684  FADREIRYTLKAQGQGAGTFNIGP----TSGIVKLAKELDFEDLRQPHVYSLVVTATEDS  739

Query  304  AEFHQSVISRYRVQSTPVTIQVINVREGIAFRPASKTFTVQKGISSKKLVDYILGT----  359
              F  SV          +TI+V +V +       +  F +    +     D  LGT
Sbjct  740  GGFSTSV---------ELTIRVTDVNDN------APKFELPDYQAHNVDEDIPLGTSILK  784

Query  360  YQAIDEDTNKAASNVKYVMGRN----DGGYLMIDSKTAEIKFVKNMNRDSTFIVNKTITA  415
             +A+D D+  A + ++Y++  +    D   +++++K  +     N N    F+V
Sbjct  785  VKAMDSDSG-ANAEIEYLVSDDHFAVDSNGIIVNNKQLD---ADNNNAYYEFVVTAKDKG  840

Query  416  EVLAIDEYTGKTSTGTVYVRVPDFNDNCP  444
            E         KT T TV V   + ND  P
Sbjct  841  E-------PAKTGTATVRVYTKNKNDEEP  862

 Score = 39.3 bits (90),  Expect = 0.30, Method: Compositional matrix adjust.
 Identities = 54/242 (22%), Positives = 102/242 (42%), Gaps = 22/242 (9%)

Query  4    KFAKPCREGE--DNSKRNP-IAKITSDYQATQKITYRISGVGIDQPPFGIFVVDKNTGDI  60
            +F  P  E E  +N K++  I  + +   A ++I Y +   G      G F +   +G +
Sbjct  656  QFYMPSYEAEIPENQKKDSDIISVKAKSFADREIRYTLKAQGQGA---GTFNIGPTSGIV  712

Query  61   NITAIVDREE--TPSF--LITCRALNAQGLDVEKPLILTVKILDINDNPPVFSQQIFMGE  116
             +   +D E+   P    L+     ++ G      + LT+++ D+NDN P F    +
Sbjct  713  KLAKELDFEDLRQPHVYSLVVTATEDSGGFSTS--VELTIRVTDVNDNAPKFELPDYQAH  770

Query  117  -IEENSASNSLVMILNATDADEPNHLNSKIAFKIVSQEPAGTPMFLLSRNTGEVRTLTNS  175
             ++E+    + ++ + A D+D  +  N++I + +VS +      F +  N   V
Sbjct  771  NVDEDIPLGTSILKVKAMDSD--SGANAEIEY-LVSDD-----HFAVDSNGIIVNNKQLD  822

Query  176  LDREQASSYRLVVSGADKDGEGLSTQCECNIKVKDVNDNFPMFRDSQYSARIEENILSSE  235
             D   A  Y  VV+  DK     +      +  K+ ND  P F    Y+  ++EN   +
Sbjct  823  ADNNNA-YYEFVVTAKDKGEPAKTGTATVRVYTKNKNDEEPKFSQQVYTPNVDENAGPNT  881

Query  236  LL  237
            L+
Sbjct  882  LV  883

>AARA21_009090.P17035 | transcript=AARA21_009090.R17035 | gene=AARA21_009090
| organism=Anopheles_arabiensis_DONGOLA_2021
| gene_product=neural-cadherin, transcript variant X5 |
transcript_product=neural-cadherin,
transcript variant X15 | location=CM029353:40466514-40666848(+)
| protein_length=3098 |
sequence_SO=chromosome | SO=protein_coding_gene | is_pseudo=false
Length=3098

 Score = 147 bits (370),  Expect = 3e-35, Method: Compositional matrix adjust.
 Identities = 128/446 (29%), Positives = 217/446 (49%), Gaps = 36/446 (8%)

Query  20    PIAKITS---DYQATQKITYRISGVGID--QPPFGIFVVDKNTGDINITAIVDREET---  71
```

```
             PI ++T+   D    Q I Y ++G GID   P    F +++ TG+I +   +DR++
Sbjct  1539  PIMQVTATDGDKDRPQNIVYFLTGQGIDPDNPANSKFDINRTTGEIFVLKPLDRDQPNGR  1598

Query  72    PSFLITCRALNAQGLDVEKPLILTVKILDINDNPPVFSQQIFMGEIEENSASNSLVMILN  131
             P +  T  A +  G  +     + V + DINDN P+F Q ++ G + EN  +  +VM +
Sbjct  1599  PQWRFTVFAQDEGGEGLVGYADVQVNLKDINDNAPIFPQGVYFGNVTENGTAGMVVMTMT  1658

Query  132   ATDADEPNH-LNSKIAFKI---VSQEPAGTPMFLLSRNTGEVRTLTNSLDREQASSYRLV  187
             A D D+PN   N+K+ + I   V +E  G+P+F +  +TG ++T    LDRE+   Y +
Sbjct  1659  AVDYDDPNEGTNAKLIYSIEKNVIEEETGSPIFEIEADTGVIKTAVCCLDRERTPDYSIQ  1718

Query  188   VSGADKDGEGLSTQCECNIKVKDVNDNFPMFRDSQYSARIEENILSS--ELLRFQVTDLD  245
             V     DG GL       +I+VKD+ND  P F   ++   ++E   ++  E+    VT  D
Sbjct  1719  VVAM--DGGGLKGTGTASIRVKDINDMPPQFTKDEWFTEVDETDGTNLPEMPILTVTVHD  1776

Query  246   EEYTDNWLAVYFFTSGNEGNWFEIQTDPRTNEGI--LKVVKALDYE---QLQSVKLSIAV  300
             E+ T+ +        SG    + F +    R N+G   LK+V+ LDYE   Q    +  I V
Sbjct  1777  EDETNKFQYKVIDNSGYGADKFTMV---RNNDGTGSLKIVQPLDYEDPLQSNGFRFRIQV  1833

Query  301   KNKAEFHQSVISRYRVQSTPVTIQVINVREGIAFRPASKTFTVQKGISSKKLVDYILGTY  360
              +K E + +   +Y V  + V +++ ++ +    +P  +   ++  +     V   L T+
Sbjct  1834  NDKGEDNDN--DKYHVAYSWVVVKLRDINDN---KPQFERPNIEVSVYENADVGKTLETF  1888

Query  361   QAIDEDTNKAASNVKYVMGR-NDGGYLMIDSKTAEIKFVKNMNRDSTFIVNKTITAEVLA  419
             +A D D     S V Y + R +D        ++   +   +N++R+    V       ++LA
Sbjct  1889  KATDPDQG-GKSKVSYAIDRSSDRQRQFSINQEGTVTIQRNLDRE----VTPRHQVKILA  1943

Query  420   IDE-YTGKTSTGTVYVRVPDFNDNCP  444
             ID+    KT+T T+ V V D NDN P
Sbjct  1944  IDDGIPPKTATATLTVIVQDINDNPP  1969

 Score = 76.3 bits (186),  Expect = 1e-12, Method: Compositional matrix adjust.
 Identities = 89/361 (25%), Positives = 167/361 (46%), Gaps = 36/361 (10%)

Query  51    FVVDKNTGDINITAIVDRE-ETPSFL-ITCRALNAQGLDVEKPLILTVKILDINDNPPVF  108
             F VD+ TG+++   + DRE +   F+ +T +A +     +E     TV+I D+NDNPP+F
Sbjct  1025  FTVDEETGEVSTNKVFDREGDDGKFVSVTVKATDQGEPSLEGVCSFTVEITDVNDNPPLF  1084

Query  109   SQQIFMGEIEENSASNSLVMILNATDADEPNHLNSKIAFKIVS-QEPAGTPMFLLSRNTG  167
              +Q ++  ++++++  + ++ ++A+D D  N  N  I + + +   P     F +   +G
Sbjct  1085  DRQKYVENVKQDASIGTNILRVSASDEDADN--NGAIVYTLSAPHSPNDLEYFEIQPESG  1142

Query  168   EVRTLTNSLDREQASSYRLVVSGADKDGEGLSTQCECNIKVKDVNDNFPMFRDSQYSA-R  226
              +  L   LDRE   +Y+L      DK    LS   E  I V D  +N P++ ++ Y
Sbjct  1143  WI-VLKKPLDRE---TYKLEAMAQDKGYPPLSRTVEVQIDVVDRANNPPVWDNTVYGPIY  1198

Query  227   IEENI-LSSELLRFQVTDLDEEYTDNWLAVYFFTSG-----NEGNWFEIQTDPRTNE--G  278
             ++EN+ +  +++  + +   E    N    Y    G     N+ + F +Q  P   +
Sbjct  1199  VKENMPVGGKVVSIKASSGIE---GNPTVFYRLMPGSTAQTNKLHTFYLQQRPDNGDTWA  1255

Query  279   ILKVVKALDYEQLQSVKLSIAVKNKAEFHQSVISRYRVQSTPVTIQVINVREGIAFRPAS  338
              +KV   LD+E ++   L++ V+N          ++        V IQ+ +V + I
Sbjct  1256  DIKVNHPLDFETIKEYNLTVRVENNG-------AQQLASEATVYIQLEDVNDEIPLFTER  1308

Query  339   KTFTVQKGISSKKLVDYILGTYQAIDEDTNKAASNVKYVM---GRNDG-GYLMIDSKTAE  394
             +   TV +G       V  +     AID+D     + V Y +    RN+G  +  I+ ++ E
Sbjct  1309  EQETVLEGEPIGTKVTQV----NAIDKDGTFPNNQVYYYIVDSPRNEGKDFFEINLQSGE  1364

Query  395   I  395
             I
Sbjct  1365  I  1365

 Score = 75.9 bits (185),  Expect = 2e-12, Method: Compositional matrix adjust.
 Identities = 103/418 (25%), Positives = 174/418 (42%), Gaps = 50/418 (12%)

Query  50    IFVVDKNTGDINITAIVDREETPSFLITCRAL----------NAQGLDVEKPLILTVKIL  99
              F ++  +G+I    + DRE+  ++ +   A            N Q   V K   + + I
Sbjct  1355  FFEINLQSGEIFTKVVFDREKQGAYALEVEARDGAPSARPNSNQQPNSVTK--FIRIGIA  1412
```

```
Query  100   DINDNPPVFSQQIFMGEIEENSASNSLVMILNATDADEPNHLNSKIAFKIVSQEPAGTPM  159
             D NDNPP F + ++  E++EN      V+ + A D DE    +S+I ++I S    G
Sbjct  1413  DKNDNPPYFDKALYEAEVDENEDIQHTVLTVTAKDHDE----SSRIRYEITSGNIGGA--  1466

Query  160   FLLSRNTGEVRTLTNSLDREQASSYRLVVSGADKDGEGLSTQCECNIKVKDVNDNFPMFR  219
             F +   TG +  +  +LD E    Y L +  +D   E  +       I VKDVND  P F
Sbjct  1467  FAVKNMTGAI-YVAGALDYETRKRYYLTLVASDSLNENETIIV---INVKDVNDMPPSFP  1522

Query  220   DSQYSARIEENI-LSSELLRFQVTDLDEEYTDNWLAVYFFTSG--NEGNWFEIQTDPRTN  276
                Y   ++E + +   +++   TD D++   N   VYF T    +  N    + D
Sbjct  1523  QQVYERTMDEELAVPFPIMQVTATDGDKDRPQN--IVYFLTGQGIDPDNPANSKFDINRT  1580

Query  277   EGILKVVKALDYEQLQS-VKLSIAVKNKAEFHQSVISRYRVQSTPVTIQVINVREGIAFR  335
              G + V+K LD +Q     +    V  + E  + ++    VQ     + + ++ +
Sbjct  1581  TGEIFVLKPLDRDQPNGRPQWRFTVFAQDEGGEGLVGYADVQ-----VNLKDINDNAPIF  1635

Query  336   PASKTFTVQKGISSKKLVDYILGTYQAIDEDTNKAASNVK--YVMGRN------DGGYLM  387
             P    F     ++        ++ T  A+D D     +N K  Y + +N
Sbjct  1636  PQGVYF---GNVTENGTAGMVVMTMTAVDYDDPNEGTNAKLIYSIEKNVIEEETGSPIFE  1692

Query  388   IDSKTAEIK-FVKNMNRDSTFIVNKTITAEVLAIDEYTGKTSTGTVYVRVPDFNDNCP  444
             I++ T  IK  V  ++R+ T        + +V+A+D   G   TGT  +RV D ND  P
Sbjct  1693  IEADTGVIKTAVCCLDRERT----PDYSIQVVAMDG-GGLKGTGTASIRVKDINDMPP  1745

 Score = 55.8 bits (133),  Expect = 3e-06, Method: Compositional matrix adjust.
 Identities = 49/158 (31%), Positives = 78/158 (49%), Gaps = 24/158 (15%)

Query  102   NDNPPVFSQQIFMGEIEENSASNSLVMILNATDADEPNHLNSKIAFKIVSQEPAGTP--M  159
             ND  P FSQQ++   ++EN+  N+LV  + A+D D  N      + F  V     GT
Sbjct  858   NDEEPKFSQQVYTPNVDENAGPNTLVTTVVASDKDGDN-----VRFGFVG---GGTSSGQ  909

Query  160   FLLSRNTGEVRTLTN--SLDREQASSYRLVVSGADK-------DGEGLSTQCECNIKVKD  210
             F++   TG +R  +   SLDR++   Y L V+  D         D    ++     + + D
Sbjct  910   FVIEEITGVIRLHSKAISLDRDK---YELNVTAMDDGSCCVNGDATIHTSTAVVVVFITD  966

Query  211   VNDNFPMFRD-SQYSARIEENILS-SELLRFQVTDLDE  246
             VNDN P+F+D   Y  ++EE   + S +++ Q TD D+
Sbjct  967   VNDNKPVFKDCGTYYPKVEEGAPNGSPVIKVQATDEDK  1004

 Score = 53.5 bits (127),  Expect = 1e-05, Method: Compositional matrix adjust.
 Identities = 70/306 (23%), Positives = 126/306 (41%), Gaps = 30/306 (10%)

Query  20    PIAKIT-SDYQATQKITYRI---SGVGIDQPPFGIFVVDKNTGDINITAIVDRE---ETP  72
             PI  +T  D   T K  Y++   SG G D+  F +   +  TG + I   +D E   ++
Sbjct  1768  PILTVTVHDEDETNKFQYKVIDNSGYGADK--FTMVRNNDGTGSLKIVQPLDYEDPLQSN  1825

Query  73    SFLITCRALNAQGLDVEKPLI------LTVKILDINDNPPVFSQQIFMGEIEENSASNSL  126
              F    + +N +G D +           + VK+ DINDN P F +       + EN+
Sbjct  1826  GFRFRIQ-VNDKGEDNDNDKYHVAYSWVVVKLRDINDNKPQFERPNIEVSVYENADVGKT  1884

Query  127   VMILNATDADEPNHLNSKIAFKIVSQEPAGTPMFLLSRNTGEVRTLTNSLDREQASSYRL  186
             +    ATD D+      SK+++ I   + +         S N      T+  +LDRE    +++
Sbjct  1885  LETFKATDPDQGG--KSKVSYAI---DRSSDRQRQFSINQEGTVTIQRNLDREVTPRHQV  1939

Query  187   VVSGADKDGEGLSTQCECNIKVKDVNDNFPMFRDSQYSARIEENILSSELLRFQVTDLDE  246
              +   D      +      + V+D+NDN P F    Y   + E++   +++    TD D+
Sbjct  1940  KILAIDDGIPPKTATATLTVIVQDINDNPPKFL-KDYRPVLPEHVPPRKVVEILATDDDD  1998

Query  247   EYTDNWLAVYFFTSGNEGNW----FEIQTDPRTNEG----ILKVVKALDYEQLQSVKLSI  298
                N     F       +     F+++ D +   G    I+  +++ D EQ +   + I
Sbjct  1999  RSKSNGPPFFQFRLDPGADDIIRASFKVEQDQKGANGDGMAIVSSLRSFDREQQKEYLIPI  2058

Query  299   AVKNKA  304
              +K+
Sbjct  2059  VIKDHG  2064

 Score = 52.0 bits (123),  Expect = 4e-05, Method: Compositional matrix adjust.
 Identities = 97/398 (24%), Positives = 163/398 (41%), Gaps = 54/398 (14%)
```

```
Query  106   PVFSQQIFMGEIEENSASNSLVMILNATDADEPNHLNSKIAFKIVSQ-EPAGTPMFLLSR  164
             P F    +  EI EN   +S ++ + A      +  + +I + + + +Q + AGT  F +
Sbjct  655   PQFYMPSYEAEIPENQKKDSDIISVKAK-----SFADREIRYTLKAQGQGAGT--FNIGP  707

Query  165   NTGEVRTLTNSLDRE---QASSYRLVVSGADKDGEGLSTQCECNIKVKDVNDNFPMFRDS  221
              +G V+ L   LD E   Q   Y LVV+ A +D  G ST  E  I+V DVNDN P F
Sbjct  708   TSGIVK-LAKELDFEDLRQPHVYSLVVT-ATEDSGGFSTSVELTIRVTDVNDNAPKFELP  765

Query  222   QYSAR-IEENI-LSSELLRFQVTDLDEEYTDNWLAVYFFTSGNEGNWFEIQTDPRTNEGI  279
              Y A  ++E+I L + +L+ +  D D         + +  S +    F + ++     GI
Sbjct  766   DYQAHNVDEDIPLGTSILKVKAMDSDSGAN---AEIEYLVSDDH---FAVDSN-----GI  814

Query  280   LKVVKALDYEQLQS-VKLSIAVKNKAEFHQSVISRYRVQSTPVTIQVINVREGIAFRPAS  338
             +   K LD +   +  +  +  K+K E  ++  +  RV +           E   F
Sbjct  815   IVNNKQLDADNNNAYYEFVVTAKDKGEPAKTGTATVRVYTKNKN------DEEPKFSQQV  868

Query  339   KTFTVQKGISSKKLVDYILGTYQAIDEDTNKAASNVK--YVMGRNDGGYLMIDSKTAEIK  396
              T  V +      LV  ++        ++K   NV+  +V G    G  +I+  T  I+
Sbjct  869   YTPNVDENAGPNTLVTTVVA--------SDKDGDNVRFGFVGGGTSSGQFVIEEITGVIR  920

Query  397   FVK---NMNRDSTFI-VNKTITAEVLAIDEYTGKTSTGTVYVRVPDFNDN------CPTA  446
                   +++RD   + V            + T  TST  V V + D NDN      C T
Sbjct  921   LHSKAISLDRDKYELNVTAMDDGSCCVNGDATIHTSTAVVVVFITDVNDNKPVFKDCGTY  980

Query  447   VLE-KDAVCSSSPSVVVSARTLNNRYTGPYTFALEDQP  483
               + ++   + SP + V A   +    G   +++  QP
Sbjct  981   YPKVEEGAPNGSPVIKVQATDEDKGVNGQVKYSIVQQP  1018

 Score = 44.7 bits (104),  Expect = 0.008, Method: Compositional matrix adjust.
 Identities = 48/199 (24%), Positives = 73/199 (37%), Gaps = 19/199 (10%)

Query  27    DYQATQKITYRISGVGIDQPPFGIFVVDKNTGDINITAIVDREETPSFLITCRALNAQGL  86
             D     K++Y I      Q  F I       G + I   +DRE TP   +   A++
Sbjct  1894  DQGGKSKVSYAIDRSSDRQRQFSI----NQEGTVTIQRNLDREVTPRHQVKILAIDDGIP  1949

Query  87    DVEKPLILTVKILDINDNPPVFSQQIFMGEIEENSASNSLVMILNATDADEPNHLNSKIA  146
                    LTV + DINDNPP F +  +   + E+     +V IL   D D
Sbjct  1950  PKTATATLTVIVQDINDNPPKFLKD-YRPVLPEHVPPRKVVEILATDDDDRSKSNGPPFQ  2008

Query  147   FKIVSQEPAGTPMFLLSRNTGEVRTLTN-----------SLDREQASSYRLVVSGADKDG  195
             F++   +P    +   S    + +   N           S DREQ    Y + +   D
Sbjct  2009  FRL---DPGADDIIRASFKVEQDQKGANGDGMAIVSSLRSFDREQQKEYLIPIVIKDHGN  2065

Query  196   EGLSTQCECNIKVKDVNDN  214
               ++      + + DVNDN
Sbjct  2066  PAMTGTSTLTVVIGDVNDN  2084

 Score = 39.7 bits (91),  Expect = 0.24, Method: Compositional matrix adjust.
 Identities = 89/389 (23%), Positives = 149/389 (38%), Gaps = 62/389 (16%)

Query  75   LITCRALNAQGLDVEKPLILTVKILDINDNPPVFSQQIF--MGEIEENSASNSLVMILNA  132
            +IT +  NA GL       + + + D+ND PP F  +       ++ N+  N+ V  L A
Sbjct  517  IITNQGHNA-GLKYTDNQRVIIHVKDVNDEPPYFINRPLPMQAVVQLNAPPNTPVFTLQA  575

Query  133  TDADEPNHLNSKIAFKIVSQEPAGTPMFLLSRNTGEVRTLTNSLDREQASSYRLVVSGAD  192
             D D     +  I + IV     G   F +   +G VRT    L  +    Y L V   D
Sbjct  576  RDPD----TDHNIHYFIVRDRTGG--RFEVDERSGVVRTRGTDL-FQLDMEYVLYVKAED  628

Query  193  KDGE----GLSTQCECNIKVKDVNDNFPMFRDSQYSARIEEN-ILSSELLRFQVTDLDEE  247
            ++G+       +  E  + +       P F    Y A I EN    S+++  +     +
Sbjct  629  QNGKVDDRRFQSTPEERLSIVG-GKRAPQFYMPSYEAEIPENQKKDSDIISVKA----KS  683

Query  248  YTDNWLAVYFFTSGNEGNWFEIQTDPRTNEGILKVVKALDYEQLQSVKLS----IAVKNK  303
            + D  +       G     F I        GI+K+ K LD+E L+   +      A ++
Sbjct  684  FADREIRYTLKAQGQGAGTFNIGP----TSGIVKLAKELDFEDLRQPHVYSLVVTATEDS  739

Query  304  AEFHQSVISRYRVQSTPVTIQVINVREGIAFRPASKTFTVQKGISSKKLVDYILGT----  359
              F  SV          +TI+V +V +       +  F +    +     D  LGT
```

```
Sbjct  740  GGFSTSV---------ELTIRVTDVNDN------APKFELPDYQAHNVDEDIPLGTSILK  784

Query  360  YQAIDEDTNKAASNVKYVMGRN----DGGYLMIDSKTAEIKFVKNMNRDSTFIVNKTITA  415
             +A+D D+  A + ++Y++  +    D   +++++K  +     N N    F+V
Sbjct  785  VKAMDSDSG-ANAEIEYLVSDDHFAVDSNGIIVNNKQLD---ADNNNAYYEFVVTAKDKG  840

Query  416  EVLAIDEYTGKTSTGTVYVRVPDFNDNCP  444
            E         KT T TV V   + ND  P
Sbjct  841  E-------PAKTGTATVRVYTKNKNDEEP  862

 Score = 39.7 bits (91),  Expect = 0.28, Method: Compositional matrix adjust.
 Identities = 54/242 (22%), Positives = 102/242 (42%), Gaps = 22/242 (9%)

Query  4    KFAKPCREGE--DNSKRNP-IAKITSDYQATQKITYRISGVGIDQPPFGIFVVDKNTGDI  60
            +F  P  E E  +N K++  I  + +   A ++I Y +   G      G F +   +G +
Sbjct  656  QFYMPSYEAEIPENQKKDSDIISVKAKSFADREIRYTLKAQGQGA---GTFNIGPTSGIV  712

Query  61   NITAIVDREE--TPSF--LITCRALNAQGLDVEKPLILTVKILDINDNPPVFSQQIFMGE  116
             +   +D E+   P    L+     ++ G       + LT+++ D+NDN P F    +
Sbjct  713  KLAKELDFEDLRQPHVYSLVVTATEDSGGFSTS--VELTIRVTDVNDNAPKFELPDYQAH  770

Query  117  -IEENSASNSLVMILNATDADEPNHLNSKIAFKIVSQEPAGTPMFLLSRNTGEVRTLTNS  175
             ++E+    + ++ + A D+D  +  N++I + +VS +      F +  N   V
Sbjct  771  NVDEDIPLGTSILKVKAMDSD--SGANAEIEY-LVSDD-----HFAVDSNGIIVNNKQLD  822

Query  176  LDREQASSYRLVVSGADKDGEGLSTQCECNIKVKDVNDNFPMFRDSQYSARIEENILSSE  235
             D   A  Y  VV+  DK     +        +  K+ ND  P F    Y+  ++EN   +
Sbjct  823  ADNNNA-YYEFVVTAKDKGEPAKTGTATVRVYTKNKNDEEPKFSQQVYTPNVDENAGPNT  881

Query  236  LL  237
            L+
Sbjct  882  LV  883

>AGAP029589-PB | transcript=AGAP029589-RB | gene=AGAP029589 |
organism=Anopheles_gambiae_PEST | gene_product=unspecified product
| transcript_product=unspecified product | location=AgamP4_3R:41575588-41776937(+)
| protein_length=3103 | sequence_SO=chromosome
| SO=protein_coding_gene | is_pseudo=false
Length=3103

 Score = 147 bits (370),  Expect = 3e-35, Method: Compositional matrix adjust.
 Identities = 128/446 (29%), Positives = 217/446 (49%), Gaps = 36/446 (8%)

Query  20    PIAKITS---DYQATQKITYRISGVGID--QPPFGIFVVDKNTGDINITAIVDREET---  71
             PI ++T+   D    Q I Y ++G GID   P    F +++ TG+I +   +DR++
Sbjct  1543  PIMQVTATDGDKDRPQNIVYFLTGQGIDPDNPANSKFDINRTTGEIFVLKPLDRDQPNGR  1602

Query  72    PSFLITCRALNAQGLDVEKPLILTVKILDINDNPPVFSQQIFMGEIEENSASNSLVMILN  131
             P +  T  A +  G  +     + V + DINDN P+F Q ++ G + EN  +  +VM +
Sbjct  1603  PQWRFTVFAQDEGGEGLVGYADVQVNLKDINDNAPIFPQGVYFGNVTENGTAGMVVMTMT  1662

Query  132   ATDADEPNH-LNSKIAFKI---VSQEPAGTPMFLLSRNTGEVRTLTNSLDREQASSYRLV  187
             A D D+PN   N+K+ + I   V +E  G+P+F +  +TG ++T    LDRE+   Y +
Sbjct  1663  AVDYDDPNEGTNAKLIYSIEKNVIEEETGSPIFEIEADTGVIKTAVCCLDRERTPDYSIQ  1722

Query  188   VSGADKDGEGLSTQCECNIKVKDVNDNFPMFRDSQYSARIEENILSS--ELLRFQVTDLD  245
             V      DG GL       +I+VKD+ND  P F   ++   ++E   ++  E+    VT  D
Sbjct  1723  VVAM--DGGGLKGTGTASIRVKDINDMPPQFTKDEWFTEVDETDGTNLPEMPILTVTVHD  1780

Query  246   EEYTDNWLAVYFFTSGNEGNWFEIQTDPRTNEGI--LKVVKALDYE---QLQSVKLSIAV  300
             E+ T+ +        SG    + F +    R N+G   LK+V+ LDYE    Q    + I V
Sbjct  1781  EDETNKFQYKVIDNSGYGADKFTMV---RNNDGTGSLKIVQPLDYEDPLQSNGFRFRIQV  1837

Query  301   KNKAEFHQSVISRYRVQSTPVTIQVINVREGIAFRPASKTFTVQKGISSKKLVDYILGTY  360
              +K E + +   +Y V  + V +++ ++ +    +P  +   ++  +      V   L T+
Sbjct  1838  NDKGEDNDN--DKYHVAYSWVVVKLRDINDN---KPQFERPNIEVSVYENADVGKTLETF  1892

Query  361   QAIDEDTNKAASNVKYVMGR-NDGGYLMIDSKTAEIKFVKNMNRDSTFIVNKTITAEVLA  419
             +A D D     S V Y + R +D        ++   +   +N++R+     V      ++LA
```

```
Sbjct  1893  KATDPDQG-GKSKVSYAIDRSSDRQRQFSINQEGTVTIQRNLDRE----VTPRHQVKILA  1947

Query  420   IDE-YTGKTSTGTVYVRVPDFNDNCP  444
             ID+    KT+T T+ V V D NDN P
Sbjct  1948  IDDGIPPKTATATLTVIVQDINDNPP  1973

 Score = 75.9 bits (185),  Expect = 2e-12, Method: Compositional matrix adjust.
 Identities = 103/418 (25%), Positives = 174/418 (42%), Gaps = 50/418 (12%)

Query  50    IFVVDKNTGDINITAIVDREETPSFLITCRAL----------NAQGLDVEKPLILTVKIL  99
              F ++  +G+I    + DRE+  ++ +   A           N Q   V K   + + I
Sbjct  1359  FFEINLQSGEIFTKVVFDREKQGAYALEVEARDGAPSARPNSNQQPNSVTK--FIRIGIA  1416

Query  100   DINDNPPVFSQQIFMGEIEENSASNSLVMILNATDADEPNHLNSKIAFKIVSQEPAGTPM  159
             D NDNPP F + ++  E++EN      V+ + A D DE    +S+I ++I S    G
Sbjct  1417  DKNDNPPYFDKALYEAEVDENEDIQHTVLTVTAKDHDE----SSRIRYEITSGNIGGA--  1470

Query  160   FLLSRNTGEVRTLTNSLDREQASSYRLVVSGADKDGEGLSTQCECNIKVKDVNDNFPMFR  219
             F +   TG +  +  +LD E    Y L +  +D   E  +      I VKDVND  P F
Sbjct  1471  FAVKNMTGAI-YVAGALDYETRKRYYLTLVASDSLNENETIIV---INVKDVNDMPPSFP  1526

Query  220   DSQYSARIEENI-LSSELLRFQVTDLDEEYTDNWLAVYFFTSG--NEGNWFEIQTDPRTN  276
                Y   ++E + +   +++   TD D++   N   VYF T    +  N    + D
Sbjct  1527  QQVYERTMDEELAVPFPIMQVTATDGDKDRPQN--IVYFLTGQGIDPDNPANSKFDINRT  1584

Query  277   EGILKVVKALDYEQLQS-VKLSIAVKNKAEFHQSVISRYRVQSTPVTIQVINVREGIAFR  335
              G + V+K LD +Q     +    V  + E  + ++    VQ     + + ++ +
Sbjct  1585  TGEIFVLKPLDRDQPNGRPQWRFTVFAQDEGGEGLVGYADVQ-----VNLKDINDNAPIF  1639

Query  336   PASKTFTVQKGISSKKLVDYILGTYQAIDEDTNKAASNVK--YVMGRN------DGGYLM  387
             P    F     ++       ++ T  A+D D     +N K  Y + +N
Sbjct  1640  PQGVYF---GNVTENGTAGMVVMTMTAVDYDDPNEGTNAKLIYSIEKNVIEEETGSPIFE  1696

Query  388   IDSKTAEIK-FVKNMNRDSTFIVNKTITAEVLAIDEYTGKTSTGTVYVRVPDFNDNCP  444
             I++ T  IK  V  ++R+ T       + +V+A+D   G   TGT  +RV D ND  P
Sbjct  1697  IEADTGVIKTAVCCLDRERT----PDYSIQVVAMDG-GGLKGTGTASIRVKDINDMPP  1749

 Score = 75.1 bits (183),  Expect = 3e-12, Method: Compositional matrix adjust.
 Identities = 109/429 (25%), Positives = 178/429 (41%), Gaps = 57/429 (13%)

Query  53    VDKNTGDINITAIVDREETPSFLITCRALNAQGLDVEKPLILTVKILDINDNPPVFSQ-Q  111
             +D++  ++N+TA+ D         +C  +N          ++ V I D+NDN PVF
Sbjct  928   LDRDKYELNVTAMDDG---------SC-CVNGDATIHTSTAVVVVFITDVNDNKPVFKDCG  978

Query  112   IFMGEIEENSASNSLVMILNATDADEPNHLNSKIAFKIVSQEPAGTPMFLLSRNTGEVRT  171
              +  ++EE + + S V+ + ATD D+   +N ++ + IV Q       F +   TGEV T
Sbjct  979   TYYPKVEEGAPNGSPVIKVQATDEDK--GVNGQVKYSIVQQPNQKGTKFTVDEETGEVST  1036

Query  172   LTNSLDRE--QASSYRLVVSGADKDGEGLSTQCECNIKVKDVNDNFPMFRDSQYSARIEE  229
                  DRE        + V   D+    L   C   +++ DVNDN P+F   +Y   +++
Sbjct  1037  -NKVFDREGDDGKFVSVTVKATDQGEPSLEGVCSFTVEITDVNDNPPLFDRQKYVENVKQ  1095

Query  230   NI-LSSELLRFQVTDLDEEYTDNWLAVYFFT---SGNEGNWFEIQTDPRTNEGILKVVKA  285
             +  + + +LR  V+  DE+  +N   VY  +   S N+  +FEIQ  P +   +LK  K
Sbjct  1096  DASIGTNILR--VSASDEDADNNGAIVYTLSAPHSPNDLEYFEIQ--PESGWIVLK--KP  1149

Query  286   LD--YEQLQSVKLSIAVKNK------AEFHQSVISRYRVQSTPVTIQV----INVREGIA  333
             LD  Y + +  +L +   ++      A+    +    R    P+  +     I++RE +
Sbjct  1150  LDGCYSR-ERYRLKVYASDRGDPPSTADVDVELDVVDRNNKPPIWDRTIYGPIHIRENVT  1208

Query  334   FRPASKTFTVQKGISSKKLVDYILGTYQAIDEDTNKAASNVKYVMGRNDGGYLMIDSKTA  393
                   +     GI     V Y L         TNK   +  Y+  R D G    D K
Sbjct  1209  VGTVVTSVKASSGIEGNPTVFYRL--MPGSTAQTNKL--HTFYLQQRPDNGDTWADIKV-  1263

Query  394   EIKFVKNMNRDSTFIVNKTITAEVLAIDEYTGK---TSTGTVYYVRVPDFNDNCPTAV-LE  449
                   N   D   I    +T  V    E  G     S  TVY+++ D ND  P     E
Sbjct  1264  ------NHPLDFEMIKEYNLTVRV----ENNGAQQLASEATVYIQLEDVNDEIPLFTERE  1313

Query  450   KDAVCSSSP  458
```

```
             ++ V    P
Sbjct  1314  QETVLEGEP  1322

 Score = 60.8 bits (146),  Expect = 8e-08, Method: Compositional matrix adjust.
 Identities = 83/362 (23%), Positives = 152/362 (42%), Gaps = 34/362 (9%)

Query  51    FVVDKNTGDINITAIVDRE-ETPSFL-ITCRALNAQGLDVEKPLILTVKILDINDNPPVF  108
             F VD+ TG+++   + DRE +   F+ +T +A +     +E     TV+I D+NDNPP+F
Sbjct  1025  FTVDEETGEVSTNKVFDREGDDGKFVSVTVKATDQGEPSLEGVCSFTVEITDVNDNPPLF  1084

Query  109   SQQIFMGEIEENSASNSLVMILNATDADEPNHLNSKIAFKIVS-QEPAGTPMFLLSRNTG  167
              +Q ++  ++++++  + ++ ++A+D D  N  N  I + + +   P     F +   +G
Sbjct  1085  DRQKYVENVKQDASIGTNILRVSASDEDADN--NGAIVYTLSAPHSPNDLEYFEIQPESG  1142

Query  168   EV---RTLTNSLDREQASSYRLVVSGADKDGEGLSTQCECNIKVKDVNDNFPMFRDSQYS  224
              +   + L     RE+   YRL V  +D+ G+  ST            +N P   D
Sbjct  1143  WIVLKKPLDGCYSRER---YRLKVYASDR-GDPPSTADVDVELDVVDRNNKPPIWDRTIY  1198

Query  225   ARIEENILSSELLRFQVTDLDEEYTDNWLAVYFFTSG-----NEGNWFEIQTDPRTNE--  277
               I      +                N    Y    G     N+ + F +Q  P   +
Sbjct  1199  GPIHIRENVTVGTVVTSVKASSGIEGNPTVFYRLMPGSTAQTNKLHTFYLQQRPDNGDTW  1258

Query  278   GILKVVKALDYEQLQSVKLSIAVKNKAEFHQSVISRYRVQSTPVTIQVINVREGIAFRPA  337
               +KV   LD+E ++   L++ V+N         ++       V IQ+ +V + I
Sbjct  1259  ADIKVNHPLDFEMIKEYNLTVRVENNG-------AQQLASEATVYIQLEDVNDEIPLFTE  1311

Query  338   SKTFTVQKGISSKKLVDYILGTYQAIDEDTNKAASNVKYVM---GRNDG-GYLMIDSKTA  393
              +  TV +G      V  +     AID+D      + V Y +    RN+G  +  I+ ++
Sbjct  1312  REQETVLEGEPIGTKVTQV----NAIDKDGTFPNNQVYYYIVDSPRNEGKDFFEINLQSG  1367

Query  394   EI  395
             EI
Sbjct  1368  EI  1369

 Score = 55.8 bits (133),  Expect = 3e-06, Method: Compositional matrix adjust.
 Identities = 49/158 (31%), Positives = 78/158 (49%), Gaps = 24/158 (15%)

Query  102   NDNPPVFSQQIFMGEIEENSASNSLVMILNATDADEPNHLNSKIAFKIVSQEPAGTP--M  159
             ND  P FSQQ++   ++EN+  N+LV  + A+D D  N     + F  V     GT
Sbjct  858   NDEEPKFSQQVYTPNVDENAGPNTLVTTVVASDKDGDN-----VRFGFVG---GGTSSGQ  909

Query  160   FLLSRNTGEVRTLTN--SLDREQASSYRLVVSGADK-------DGEGLSTQCECNIKVKD  210
             F++   TG +R  +   SLDR++   Y L V+  D        D    ++     + + D
Sbjct  910   FVIEEITGVIRLHSKAISLDRDK---YELNVTAMDDGSCCVNGDATIHTSTAVVVVFITD  966

Query  211   VNDNFPMFRD-SQYSARIEENILS-SELLRFQVTDLDE  246
             VNDN P+F+D   Y  ++EE   + S +++ Q TD D+
Sbjct  967   VNDNKPVFKDCGTYYPKVEEGAPNGSPVIKVQATDEDK  1004

 Score = 53.9 bits (128),  Expect = 1e-05, Method: Compositional matrix adjust.
 Identities = 70/306 (23%), Positives = 126/306 (41%), Gaps = 30/306 (10%)

Query  20    PIAKIT-SDYQATQKITYRI---SGVGIDQPPFGIFVVDKNTGDINITAIVDRE---ETP  72
             PI  +T  D   T K  Y++   SG G D+  F +   +  TG + I   +D E   ++
Sbjct  1772  PILTVTVHDEDETNKFQYKVIDNSGYGADK--FTMVRNNDGTGSLKIVQPLDYEDPLQSN  1829

Query  73    SFLITCRALNAQGLDVEKPLI------LTVKILDINDNPPVFSQQIFMGEIEENSASNSL  126
              F    + +N +G D +          + VK+ DINDN P F +      + EN+
Sbjct  1830  GFRFRIQ-VNDKGEDNDNDKYHVAYSWVVVKLRDINDNKPQFERPNIEVSVYENADVGKT  1888

Query  127   VMILNATDADEPNHLNSKIAFKIVSQEPAGTPMFLLSRNTGEVRTLTNSLDREQASSYRL  186
             +    ATD D+     SK+++ I   + +       S N     T+  +LDRE    +++
Sbjct  1889  LETFKATDPDQGG--KSKVSYAI---DRSSDRQRQFSINQEGTVTIQRNLDREVTPRHQV  1943

Query  187   VVSGADKDGEGLSTQCECNIKVKDVNDNFPMFRDSQYSARIEENILSSELLRFQVTDLDE  246
              +   D      +      + V+D+NDN P F    Y   + E++   +++    TD D+
Sbjct  1944  KILAIDDGIPPKTATATLTVIVQDINDNPPKFL-KDYRPVLPEHVPPRKVVEILATDDDD  2002
```

```
Query  247   EYTDNWLAVYFFTSGNEGNW----FEIQTDPRTNEG----ILKVVKALDYEQLQSVKLSI  298
                 N     F       +     F+++ D +   G    I+  +++ D EQ +   + I
Sbjct  2003  RSKSNGPPFQFRLDPGADDIIRASFKVEQDQKGANGDGMAIVSSLRSFDREQQKEYLIPI  2062

Query  299   AVKNKA  304
              +K+
Sbjct  2063  VIKDHG  2068

 Score = 52.0 bits (123),  Expect = 4e-05, Method: Compositional matrix adjust.
 Identities = 97/398 (24%), Positives = 163/398 (41%), Gaps = 54/398 (14%)

Query  106   PVFSQQIFMGEIEENSASNSLVMILNATDADEPNHLNSKIAFKIVSQ-EPAGTPMFLLSR  164
             P F    +  EI EN   +S ++ + A      +  + +I + + +Q + AGT  F +
Sbjct  655   PQFYMPSYEAEIPENQKKDSDIISVKAK-----SFADREIRYTLKAQGQGAGT--FNIGP  707

Query  165   NTGEVRTLTNSLDRE---QASSYRLVVSGADKDGEGLSTQCECNIKVKDVNDNFPMFRDS  221
              +G V+ L   LD E   Q   Y LVV+ A +D  G ST  E  I+V DVNDN P F
Sbjct  708   TSGIVK-LAKELDFEDLRQPHVYSLVVT-ATEDSGGFSTSVELTIRVTDVNDNAPKFELP  765

Query  222   QYSAR-IEENI-LSSELLRFQVTDLDEEYTDNWLAVYFFTSGNEGNWFEIQTDPRTNEGI  279
              Y A  ++E+I L + +L+ +  D D         + +  S +    F + ++     GI
Sbjct  766   DYQAHNVDEDIPLGTSILKVKAMDSDSGAN---AEIEYLVSDDH---FAVDSN-----GI  814

Query  280   LKVVKALDYEQLQS-VKLSIAVKNKAEFHQSVISRYRVQSTPVTIQVINVREGIAFRPAS  338
             +   K LD +   +  +  +  K+K E  ++  +  RV +           E   F
Sbjct  815   IVNNKQLDADNNNAYYEFVVTAKDKGEPAKTGTATVRVYTKNKN------DEEPKFSQQV  868

Query  339   KTFTVQKGISSKKLVDYILGTYQAIDEDTNKAASNVK--YVMGRNDGGYLMIDSKTAEIK  396
              T  V +      LV  ++         ++K   NV+  +V G    G  +I+  T  I+
Sbjct  869   YTPNVDENAGPNTLVTTVVA--------SDKDGDNVRFGFVGGGTSSGQFVIEEITGVIR  920

Query  397   FVK---NMNRDSTFI-VNKTITAEVLAIDEYTGKTSTGTVYVRVPDFNDN------CPTA  446
                   +++RD   + V            + T  TST  V V + D NDN      C T
Sbjct  921   LHSKAISLDRDKYELNVTAMDDGSCCVNGDATIHTSTAVVVVFITDVNDNKPVFKDCGTY  980

Query  447   VLE-KDAVCSSSPSVVVSARTLNNRYTGPYTFALEDQP  483
               + ++   + SP + V A    +   G   +++  QP
Sbjct  981   YPKVEEGAPNGSPVIKVQATDEDKGVNGQVKYSIVQQP  1018

 Score = 44.3 bits (103),  Expect = 0.008, Method: Compositional matrix adjust.
 Identities = 48/199 (24%), Positives = 73/199 (37%), Gaps = 19/199 (10%)

Query  27    DYQATQKITYRISGVGIDQPPFGIFVVDKNTGDINITAIVDREETPSFLITCRALNAQGL  86
             D     K++Y I      Q  F I       G + I   +DRE TP   +   A++
Sbjct  1898  DQGGKSKVSYAIDRSSDRQRQFSI----NQEGTVTIQRNLDREVTPRHQVKILAIDDGIP  1953

Query  87    DVEKPLILTVKILDINDNPPVFSQQIFMGEIEENSASNSLVMILNATDADEPNHLNSKIA  146
                    LTV + DINDNPP F +  +   + E+     +V IL   D D
Sbjct  1954  PKTATATLTVIVQDINDNPPKFLKD-YRPVLPEHVPPRKVVEILATDDDDRSKSNGPPFQ  2012

Query  147   FKIVSQEPAGTPMFLLSRNTGEVRTLTN-----------SLDREQASSYRLVVSGADKDG  195
             F++   +P    +   S    + +   N           S DREQ   Y + +   D
Sbjct  2013  FRL---DPGADDIIRASFKVEQDQKGANGDGMAIVSSLRSFDREQQKEYLIPIVIKDHGN  2069

Query  196   EGLSTQCECNIKVKDVNDN  214
               ++      + + DVNDN
Sbjct  2070  PAMTGTSTLTVVIGDVNDN  2088

 Score = 39.7 bits (91),  Expect = 0.28, Method: Compositional matrix adjust.
 Identities = 89/389 (23%), Positives = 149/389 (38%), Gaps = 62/389 (16%)

Query  75   LITCRALNAQGLDVEKPLILTVKILDINDNPPVFSQQIF--MGEIEENSASNSLVMILNA  132
            +IT +  NA GL       + + + D+ND PP F  +       ++ N+  N+ V  L A
Sbjct  517  IITNQGHNA-GLKYTDNQRVIIHVKDVNDEPPYFINRPLPMQAVVQLNAPPNTPVFTLQA  575

Query  133  TDADEPNHLNSKIAFKIVSQEPAGTPMFLLSRNTGEVRTLTNSLDREQASSYRLVVSGAD  192
             D D     +  I + IV     G   F +   +G VRT    L  +    Y L V   D
Sbjct  576  RDPD----TDHNIHYFIVRDRTGG--RFEVDERSGVVRTRGTDL-FQLDMEYVLYVKAED  628
```

```
Query  193  KDGE----GLSTQCECNIKVKDVNDNFPMFRDSQYSARIEEN-ILSSELLRFQVTDLDEE  247
            ++G+        +  E  + +       P F    Y A I EN    S+++  +     +
Sbjct  629  QNGKVDDRRFQSTPEERLSIVG-GKRAPQFYMPSYEAEIPENQKKDSDIISVKA----KS  683

Query  248  YTDNWLAVYFFTSGNEGNWFEIQTDPRTNEGILKVVKALDYEQLQSVKLS----IAVKNK  303
            + D  +        G     F I         GI+K+ K LD+E L+   +       A ++
Sbjct  684  FADREIRYTLKAQGQGAGTFNIGP----TSGIVKLAKELDFEDLRQPHVYSLVVTATEDS  739

Query  304  AEFHQSVISRYRVQSTPVTIQVINVREGIAFRPASKTFTVQKGISSKKLVDYILGT----  359
              F  SV           +TI+V +V +        +  F +    +     D  LGT
Sbjct  740  GGFSTSV---------ELTIRVTDVNDN------APKFELPDYQAHNVDEDIPLGTSILK  784

Query  360  YQAIDEDTNKAASNVKYVMGRN----DGGYLMIDSKTAEIKFVKNMNRDSTFIVNKTITA  415
             +A+D D+  A + ++Y++  +    D    +++++K  +     N N    F+V
Sbjct  785  VKAMDSDSG-ANAEIEYLVSDDHFAVDSNGIIVNNKQLD---ADNNNAYYEFVVTAKDKG  840

Query  416  EVLAIDEYTGKTSTGTVYVRVPDFNDNCP  444
            E          KT T TV V   + ND  P
Sbjct  841  E-------PAKTGTATVRVYTKNKNDEEP  862

 Score = 39.3 bits (90),  Expect = 0.30, Method: Compositional matrix adjust.
 Identities = 54/242 (22%), Positives = 102/242 (42%), Gaps = 22/242 (9%)

Query  4    KFAKPCREGE--DNSKRNP-IAKITSDYQATQKITYRISGVGIDQPPFGIFVVDKNTGDI  60
            +F  P  E E  +N K++  I  + +   A ++I Y +   G       G F +   +G +
Sbjct  656  QFYMPSYEAEIPENQKKDSDIISVKAKSFADREIRYTLKAQGQGA---GTFNIGPTSGIV  712

Query  61   NITAIVDREE--TPSF--LITCRALNAQGLDVEKPLILTVKILDINDNPPVFSQQIFMGE  116
             +   +D E+   P    L+     ++ G       + LT+++ D+NDN P F    +
Sbjct  713  KLAKELDFEDLRQPHVYSLVVTATEDSGGFSTS--VELTIRVTDVNDNAPKFELPDYQAH  770

Query  117  -IEENSASNSLVMILNATDADEPNHLNSKIAFKIVSQEPAGTPMFLLSRNTGEVRTLTNS  175
             ++E+    + ++ + A D+D  +  N++I + +VS +       F +  N   V
Sbjct  771  NVDEDIPLGTSILKVKAMDSD--SGANAEIEY-LVSDD-----HFAVDSNGIIVNNKQLD  822

Query  176  LDREQASSYRLVVSGADKDGEGLSTQCECNIKVKDVNDNFPMFRDSQYSARIEENILSSE  235
             D   A  Y  VV+  DK      +       +  K+ ND  P F    Y+  ++EN   +
Sbjct  823  ADNNNA-YYEFVVTAKDKGEPAKTGTATVRVYTKNKNDEEPKFSQQVYTPNVDENAGPNT  881

Query  236  LL  237
            L+
Sbjct  882  LV  883

>AMAC_003346.P2999 | transcript=AMAC_003346.R2999 | gene=AMAC_003346
| organism=Anopheles_maculipalpis_AmacGA1 | gene_product=neural-cadherin,
transcript variant X12 | transcript_product=neural-cadherin,
transcript variant X8 | location=AmacGA1_3RL:17033394-17218514(+)
| protein_length=3107 | sequence_SO=chromosome
| SO=protein_coding_gene | is_pseudo=false
Length=3107

 Score = 147 bits (370),  Expect = 3e-35, Method: Compositional matrix adjust.
 Identities = 128/446 (29%), Positives = 217/446 (49%), Gaps = 36/446 (8%)

Query  20    PIAKITS---DYQATQKITYRISGVGID--QPPFGIFVVDKNTGDINITAIVDREET---  71
              PI ++T+    D    Q I Y ++G GID   P    F +++ TG+I +   +DR++
Sbjct  1547  PIMQVTATDGDKDRPQNIVYFLTGQGIDPDNPANSKFDINRTTGEIFVLKPLDRDQPNGR  1606

Query  72    PSFLITCRALNAQGLDVEKPLILTVKILDINDNPPVFSQQIFMGEIEENSASNSLVMILN  131
              P +  T  A +  G  +      + V + DINDN P+F Q ++ G + EN  +  +VM +
Sbjct  1607  PQWRFTVFAQDEGGEGLVGYADVQVNLKDINDNAPIFPQGVYFGNVTENGTAGMVVMTMT  1666

Query  132   ATDADEPNH-LNSKIAFKI---VSQEPAGTPMFLLSRNTGEVRTLTNSLDREQASSYRLV  187
              A D D+PN   N+K+ + I   V +E  G+P+F +  +TG ++T    LDRE+   Y +
Sbjct  1667  AVDYDDPNEGTNAKLIYSIEKNVIEEETGSPIFEIEADTGVIKTAVCCLDRERTPDYSIQ  1726

Query  188   VSGADKDGEGLSTQCECNIKVKDVNDNFPMFRDSQYSARIEENILSS--ELLRFQVTDLD  245
              V      DG GL       +I+VKD+ND  P F   ++   ++E   ++  E+    VT  D
```

```
Sbjct  1727   VVAM--DGGGLKGTGTASIRVKDINDMPPQFTKDEWFTEVDETDGTNLPEMPILTVTVHD  1784

Query  246    EEYTDNWLAVYFFTSGNEGNWFEIQTDPRTNEGI--LKVVKALDYE---QLQSVKLSIAV  300
              E+ T+ +       SG   + F +    R N+G   LK+V+ LDYE   Q    +  I V
Sbjct  1785   EDETNKFQYKVIDNSGYGADKFTMV---RNNDGTGSLKIVQPLDYEDPLQSNGFRFRIQV  1841

Query  301    KNKAEFHQSVISRYRVQSTPVTIQVINVREGIAFRPASKTFTVQKGISSKKLVDYILGTY  360
               +K E + +   +Y V  + V +++ ++ +    +P  +   ++  +      V   L T+
Sbjct  1842   NDKGEDNDN--DKYHVAYSWVVVKLRDINDN---KPQFERPNIEVSVYENADVGKTLETF  1896

Query  361    QAIDEDTNKAASNVKYVMGR-NDGGYLMIDSKTAEIKFVKNMNRDSTFIVNKTITAEVLA  419
              +A D D     S V Y + R +D       ++    +   +N++R+    V      ++LA
Sbjct  1897   KATDPDQG-GKSKVSYAIDRSSDRQRQFSINQEGTVTIQRNLDRE----VTPRHQVKILA  1951

Query  420    IDE-YTGKTSTGTVYYRVPDFNDNCP  444
              ID+    KT+T T+ V V D NDN P
Sbjct  1952   IDDGIPPKTATATLTVIVQDINDNPP  1977

 Score = 78.6 bits (192),  Expect = 3e-13, Method: Compositional matrix adjust.
 Identities = 106/424 (25%), Positives = 180/424 (42%), Gaps = 51/424 (12%)

Query  53     VDKNTGDINITAIVDREETPSFLITCRALNAQGLDVEKPLILTVKILDINDNPPVFSQ-Q  111
              +D++  ++N+TA+ D          +C  +N           ++ V I D+NDN PVF
Sbjct  936    LDRDKYELNVTAMDDG--------SC-CVNGDATIHTSTAVVVVFITDVNDNKPVFKDCG  986

Query  112    IFMGEIEENSASNSLVMILNATDADEPNHLNSKIAFKIVSQEPAGTPMFLLSRNTGEVRT  171
               +  ++EE + + S V+ + ATD D+   +N ++ + IV Q       F +   TGEV T
Sbjct  987    TYYPKVEEGAPNGSPVIKVQATDEDK--GVNGQVKYSIVQQPNQKGTKFTVDEETGEVST  1044

Query  172    LTNSLDRE--QASSYRLVVSGADKDGEGLSTQCECNIKVKDVNDNFPMFRDSQYSARIEE  229
                   DRE         + V   D+    L   C   +++ DVNDN P+F   +Y   +++
Sbjct  1045   -NKVFDREGDDGKFVSVTVKATDQGEPSLEGVCSFTVEITDVNDNPPLFDRQKYVENVKQ  1103

Query  230    NI-LSSELLRFQVTDLDEEYTDNWLAVYFFT---SGNEGNWFEIQTDPRTNEGILKVVKA  285
              +  + + +LR  V+  DE+  +N   VY  +   S N+  +FEIQ  P +   +LK  K
Sbjct  1104   DASIGTNILR--VSASDEDADNNGAIVYTLSAPHSPNDLEYFEIQ--PESGWIVLK--KP  1157

Query  286    LDYEQLQSVKLSIAVKNK------AEFHQSVISRYRVQSTPVTIQV----INVREGIAFR  335
              LD E+    +L +   ++       A+    +    R    P+  +     I++RE +
Sbjct  1158   LDRERY---RLKVYASDRGDPPSTADVDVELDVVDRNNKPPIWDRTIYGPIHIRENVTVG  1214

Query  336    PASKTFTVQKGISSKKLVDYILGTYQAIDEDTNKAASNVKYVMGRNDGGYLMIDSKTAEI  395
                  +     GI     V Y L          TNK   +  Y+  R D G        A+I
Sbjct  1215   TVVTSVKASSGIEGNPTVFYRL--MPGSTAQTNKL--HTFYLQQRPDNG-----DTWADI  1265

Query  396    KFVKNMNRDSTFIVNKTITAEVLAIDEYTGKTSTGTVYYRVPDFNDNCPTAV-LEKDAVC  454
              K    ++ ++    N T+  E     +      S  TVY+++ D ND  P     E++ V
Sbjct  1266   KVNHPLDFETIKEYNLTVRVENNGAQQL---ASEATVYIQLEDVNDEIPLFTEREQETVL  1322

Query  455    SSSP  458
                P
Sbjct  1323   EGEP  1326

 Score = 76.6 bits (187),  Expect = 1e-12, Method: Compositional matrix adjust.
 Identities = 103/418 (25%), Positives = 174/418 (42%), Gaps = 50/418 (12%)

Query  50     IFVVDKNTGDINITAIVDREETPSFLITCRAL----------NAQGLDVEKPLILTVKIL  99
               F ++  +G+I    + DRE+  ++ +   A            N Q   V K   + + I
Sbjct  1363   FFEINLQSGEIFTKVVFDREKQGAYALEVEARDGAPSARPNSNQQPNSVTK--FIRIGIA  1420

Query  100    DINDNPPVFSQQIFMGEIEENSASNSLVMILNATDADEPNHLNSKIAFKIVSQEPAGTPM  159
              D NDNPP F + ++  E++EN      V+ + A D DE    +S+I ++I S    G
Sbjct  1421   DKNDNPPYFDKALYEAEVDENEDIQHTVLTVTAKDHDE----SSRIRYEITSGNIGGA--  1474

Query  160    FLLSRNTGEVRTLTNSLDREQASSYRLVVSGADKDGEGLSTQCECNIKVKDVNDNFPMFR  219
              F +   TG +  +  +LD E    Y L +  +D   E  +       I VKDVND  P F
Sbjct  1475   FAVKNMTGAI-YVAGALDYETRKRYHLTLVASDSLNENETIIV---INVKDVNDMPPSFP  1530

Query  220    DSQYSARIEENI-LSSELLRFQVTDLDEEYTDNWLAVYFFTSG--NEGNWFEIQTDPRTN  276
```

```
                Y   ++E + +   +++   TD D++   N   VYF T    +  N    + D
Sbjct  1531   QQVYERTMDEELAVPFPIMQVTATDGDKDRPQN--IVYFLTGQGIDPDNPANSKFDINRT  1588

Query  277    EGILKVVKALDYEQLQS-VKLSIAVKNKAEFHQSVISRYRVQSTPVTIQVINVREGIAFR  335
               G + V+K LD +Q     +    V  + E  + ++    VQ     + + ++ +
Sbjct  1589   TGEIFVLKPLDRDQPNGRPQWRFTVFAQDEGGEGLVGYADVQ-----VNLKDINDNAPIF  1643

Query  336    PASKTFTVQKGISSKKLVDYILGTYQAIDEDTNKAASNVK--YVMGRN------DGGYLM  387
              P    F     ++        ++ T  A+D D     +N K  Y + +N
Sbjct  1644   PQGVYF---GNVTENGTAGMVVMTMTAVDYDDPNEGTNAKLIYSIEKNVIEEETGSPIFE  1700

Query  388    IDSKTAEIK-FVKNMNRDSTFIVNKTITAEVLAIDEYTGKTSTGTVYVRVPDFNDNCP  444
              I++ T  IK  V  ++R+ T        + +V+A+D   G   TGT  +RV D ND  P
Sbjct  1701   IEADTGVIKTAVCCLDRERT----PDYSIQVVAMDG-GGLKGTGTASIRVKDINDMPP  1753

 Score = 65.5 bits (158),  Expect = 3e-09, Method: Compositional matrix adjust.
 Identities = 85/359 (24%), Positives = 153/359 (43%), Gaps = 32/359 (9%)

Query  51     FVVDKNTGDINITAIVDRE-ETPSFL-ITCRALNAQGLDVEKPLILTVKILDINDNPPVF  108
              F VD+ TG+++   + DRE +   F+ +T +A +      +E      TV+I D+NDNPP+F
Sbjct  1033   FTVDEETGEVSTNKVFDREGDDGKFVSVTVKATDQGEPSLEGVCSFTVEITDVNDNPPLF  1092

Query  109    SQQIFMGEIEENSASNSLVMILNATDADEPNHLNSKIAFKIVS-QEPAGTPMFLLSRNTG  167
               +Q ++  ++++++  + ++ ++A+D D  N  N  I + + +   P      F +   +G
Sbjct  1093   DRQKYVENVKQDASIGTNILRVSASDEDADN--NGAIVYTLSAPHSPNDLEYFEIQPESG  1150

Query  168    EVRTLTNSLDREQASSYRLVVSGADKDGEGLSTQCECNIKVKDVNDNFPMFRDSQYSARI  227
               +  L   LDRE+   YRL V  +D+ G+  ST             +N P   D      I
Sbjct  1151   WI-VLKKPLDRER---YRLKVYASDR-GDPPSTADVDVELDVVDRNNKPPIWDRTIYGPI  1205

Query  228    EENILSSELLRFQVTDLDEEYTDNWLAVYFFTSG-----NEGNWFEIQTDPRTNE--GIL  280
                   +                  N    Y    G     N+ + F +Q  P   +    +
Sbjct  1206   HIRENVTVGTVVTSVKASSGIEGNPTVFYRLMPGSTAQTNKLHTFYLQQRPDNGDTWADI  1265

Query  281    KVVKALDYEQLQSVKLSIAVKNKAEFHQSVISRYRVQSTPVTIQVINVREGIAFRPASKT  340
              KV   LD+E ++   L++ V+N          ++        V IQ+ +V + I      +
Sbjct  1266   KVNHPLDFETIKEYNLTVRVENNG-------AQQLASEATVYIQLEDVNDEIPLFTEREQ  1318

Query  341    FTVQKGISSKKLVDYILGTYQAIDEDTNKAASNVKYVM---GRNDG-GYLMIDSKTAEI  395
               TV +G       V  +     AID+D     + V Y +    RN+G  +  I+ ++ EI
Sbjct  1319   ETVLEGEPIGTKVTQV----NAIDKDGTFPNNQVYYYIVDSPRNEGKDFFEINLQSGEI  1373

 Score = 55.8 bits (133),  Expect = 3e-06, Method: Compositional matrix adjust.
 Identities = 49/158 (31%), Positives = 78/158 (49%), Gaps = 24/158 (15%)

Query  102    NDNPPVFSQQIFMGEIEENSASNSLVMILNATDADEPNHLNSKIAFKIVSQEPAGTP--M  159
              ND  P FSQQ++   ++EN+  N+LV  + A+D D  N     + F  V     GT
Sbjct  866    NDEEPKFSQQVYTPNVDENAGPNTLVTTVVASDKDGDN-----VRFGFVG---GGTSSGQ  917

Query  160    FLLSRNTGEVRTLTN--SLDREQASSYRLVVSGADK-------DGEGLSTQCECNIKVKD  210
              F++   TG +R  +   SLDR++   Y L V+  D         D    ++     + + D
Sbjct  918    FVIEEITGVIRLHSKAISLDRDK---YELNVTAMDDGSCCVNGDATIHTSTAVVVVFITD  974

Query  211    VNDNFPMFRD-SQYSARIEENILS-SELLRFQVTDLDE  246
              VNDN P+F+D   Y  ++EE   + S +++ Q TD D+
Sbjct  975    VNDNKPVFKDCGTYYPKVEEGAPNGSPVIKVQATDEDK  1012

 Score = 53.5 bits (127),  Expect = 1e-05, Method: Compositional matrix adjust.
 Identities = 70/306 (23%), Positives = 126/306 (41%), Gaps = 30/306 (10%)

Query  20     PIAKIT-SDYQATQKITYRI---SGVGIDQPPFGIFVVDKNTGDINITAIVDRE---ETP  72
              PI  +T  D   T K  Y++   SG G D+  F +   +  TG + I   +D E   ++
Sbjct  1776   PILTVTVHDEDETNKFQYKVIDNSGYGADK--FTMVRNNDGTGSLKIVQPLDYEDPLQSN  1833

Query  73     SFLITCRALNAQGLDVEKPLI------LTVKILDINDNPPVFSQQIFMGEIEENSASNSL  126
               F    + +N +G D +            + VK+ DINDN P F +       + EN+
Sbjct  1834   GFRFRIQ-VNDKGEDNDNDKYHVAYSWVVVKLRDINDNKPQFERPNIEVSVYENADVGKT  1892
```

```
Query  127   VMILNATDADEPNHLNSKIAFKIVSQEPAGTPMFLLSRNTGEVRTLTNSLDREQASSYRL  186
             +    ATD D+     SK+++ I   + +       S N     T+  +LDRE    +++
Sbjct  1893  LETFKATDPDQGG--KSKVSYAI---DRSSDRQRQFSINQEGTVTIQRNLDREVTPRHQV  1947

Query  187   VVSGADKDGEGLSTQCECNIKVKDVNDNFPMFRDSQYSARIEENILSSELLRFQVTDLDE  246
              +   D      +      + V+D+NDN P F    Y   + E++   +++    TD D+
Sbjct  1948  KILAIDDGIPPKTATATLTVIVQDINDNPPKFL-KDYRPVLPEHVPPRKVVEILATDDDD  2006

Query  247   EYTDNWLAVYFFTSGNEGNW----FEIQTDPRTNEG----ILKVVKALDYEQLQSVKLSI  298
                 N     F       +     F+++ D +   G    I+  +++ D EQ +   + I
Sbjct  2007  RSKSNGPPFQFRLDPGADDIIRASFKVEQDQKGANGDGMAIVSSLRSFDREQQKEYLIPI  2066

Query  299   AVKNKA  304
              +K+
Sbjct  2067  VIKDHG  2072

 Score = 52.4 bits (124),  Expect = 3e-05, Method: Compositional matrix adjust.
 Identities = 97/398 (24%), Positives = 163/398 (41%), Gaps = 54/398 (14%)

Query  106   PVFSQQIFMGEIEENSASNSLVMILNATDADEPNHLNSKIAFKIVSQ-EPAGTPMFLLSR  164
             P F    +  EI EN   +S ++ + A      +  + +I + + +Q + AGT  F +
Sbjct  663   PQFYMPSYEAEIPENQKKDSDIISVKAK-----SFADREIRYTLKAQGQGAGT--FNIGP  715

Query  165   NTGEVRTLTNSLDRE---QASSYRLVVSGADKDGEGLSTQCECNIKVKDVNDNFPMFRDS  221
              +G V+ L   LD E   Q   Y LVV+ A +D  G ST  E  I+V DVNDN P F
Sbjct  716   TSGIVK-LAKELDFEDLRQPHVYSLVVT-ATEDSGGFSTSVELTIRVTDVNDNAPKFELP  773

Query  222   QYSAR-IEENI-LSSELLRFQVTDLDEEYTDNWLAVYFFTSGNEGNWFEIQTDPRTNEGI  279
              Y A  ++E+I L + +L+ +  D D         + +  S +    F + ++     GI
Sbjct  774   DYQAHNVDEDIPLGTSILKVKAMDADSGAN---AEIEYLVSDDH---FAVDSN-----GI  822

Query  280   LKVVKALDYEQLQS-VKLSIAVKNKAEFHQSVISRYRVQSTPVTIQVINVREGIAFRPAS  338
             +   K LD +   +  +  +  K+K E  ++  +  RV +           E   F
Sbjct  823   IVNNKQLDADNNNAYYEFVVTAKDKGEPAKTGTATVRVYTKNKN------DEEPKFSQQV  876

Query  339   KTFTVQKGISSKKLVDYILGTYQAIDEDTNKAASNVK--YVMGRNDGGYLMIDSKTAEIK  396
              T  V +      LV  ++         ++K   NV+  +V G    G  +I+  T  I+
Sbjct  877   YTPNVDENAGPNTLVTTVVA--------SDKDGDNVRFGFVGGGTSSGQFVIEEITGVIR  928

Query  397   FVK---NMNRDSTFI-VNKTITAEVLAIDEYTGKTSTGTVYVRVPDFNDN------CPTA  446
                   +++RD   + V            + T  TST  V V + D NDN      C T
Sbjct  929   LHSKAISLDRDKYELNVTAMDDGSCCVNGDATIHTSTAVVVVFITDVNDNKPVFKDCGTY  988

Query  447   VLE-KDAVCSSSPSVVVSARTLNNRYTGPYTFALEDQP  483
               + ++   + SP + V A   +    G   +++  QP
Sbjct  989   YPKVEEGAPNGSPVIKVQATDEDKGVNGQVKYSIVQQP  1026

 Score = 44.3 bits (103),  Expect = 0.009, Method: Compositional matrix adjust.
 Identities = 48/199 (24%), Positives = 73/199 (37%), Gaps = 19/199 (10%)

Query  27    DYQATQKITYRISGVGIDQPPFGIFVVDKNTGDINITAIVDREETPSFLITCRALNAQGL  86
             D     K++Y I      Q  F I       G + I   +DRE TP   +   A++
Sbjct  1902  DQGGKSKVSYAIDRSSDRQRQFSI----NQEGTVTIQRNLDREVTPRHQVKILAIDDGIP  1957

Query  87    DVEKPLILTVKILDINDNPPVFSQQIFMGEIEENSASNSLVMILNATDADEPNHLNSKIA  146
                    LTV + DINDNPP F +  +   + E+     +V IL   D D
Sbjct  1958  PKTATATLTVIVQDINDNPPKFLKD-YRPVLPEHVPPRKVVEILATDDDDRSKSNGPPFQ  2016

Query  147   FKIVSQEPAGTPMFLLSRNTGEVRTLTN-----------SLDREQASSYRLVVSGADKDG  195
             F++   +P    +   S    + +   N           S DREQ   Y + +   D
Sbjct  2017  FRL---DPGADDIIRASFKVEQDQKGANGDGMAIVSSLRSFDREQQKEYLIPIVIKDHGN  2073

Query  196   EGLSTQCECNIKVKDVNDN  214
               ++      + + DVNDN
Sbjct  2074  PAMTGTSTLTVVIGDVNDN  2092

 Score = 40.4 bits (93),  Expect = 0.13, Method: Compositional matrix adjust.
 Identities = 54/235 (23%), Positives = 99/235 (42%), Gaps = 22/235 (9%)
```

```

Query  4    KFAKPCREGE--DNSKRNP-IAKITSDYQATQKITYRISGVGIDQPPFGIFVVDKNTGDI  60
            +F  P  E E  +N K++  I   + +   A ++I Y +   G      G F +   +G +
Sbjct  664  QFYMPSYEAEIPENQKKDSDIISVKAKSFADREIRYTLKAQGQGA---GTFNIGPTSGIV  720

Query  61   NITAIVDREE--TPSF--LITCRALNAQGLDVEKPLILTVKILDINDNPPVFSQQIFMGE  116
             +   +D E+   P    L+     ++ G       + LT+++ D+NDN P F    +
Sbjct  721  KLAKELDFEDLRQPHVYSLVVTATEDSGGFSTS--VELTIRVTDVNDNAPKFELPDYQAH  778

Query  117  -IEENSASNSLVMILNATDADEPNHLNSKIAFKIVSQEPAGTPMFLLSRNTGEVRTLTNS  175
             ++E+    + ++ + A DAD  +  N++I + +VS +      F +  N   V
Sbjct  779  NVDEDIPLGTSILKVKAMDAD--SGANAEIEY-LVSDD-----HFAVDSNGIIVNNKQLD  830

Query  176  LDREQASSYRLVVSGADKDGEGLSTQCECNIKVKDVNDNFPMFRDSQYSARIEEN  230
             D   A  Y  VV+  DK      +       +  K+ ND  P F    Y+  ++EN
Sbjct  831  ADNNNA-YYEFVVTAKDKGEPAKTGTATVRVYTKNKNDEEPKFSQQVYTPNVDEN  884

 Score = 39.3 bits (90),  Expect = 0.35, Method: Compositional matrix adjust.
 Identities = 89/389 (23%), Positives = 149/389 (38%), Gaps = 62/389 (16%)

Query  75   LITCRALNAQGLDVEKPLILTVKILDINDNPPVFSQQIF--MGEIEENSASNSLVMILNA  132
            +IT +  NA GL       + + + D+ND PP F  +        ++ N+  N+ V  L A
Sbjct  525  IITNQGHNA-GLKYTDNQRVIIHVKDVNDEPPYFINRPLPMQAVVQLNAPPNTPVFTLQA  583

Query  133  TDADEPNHLNSKIAFKIVSQEPAGTPMFLLSRNTGEVRTLTNSLDREQASSYRLVVSGAD  192
             D D     +  I + IV     G   F +   +G VRT    L  +    Y L V   D
Sbjct  584  RDPD----TDHNIHYFIVRDRTGG--RFEVDERSGVVRTRGTDL-FQLDMEYVLYVKAED  636

Query  193  KDGE----GLSTQCECNIKVKDVNDNFPMFRDSQYSARIEEN-ILSSELLRFQVTDLDEE  247
            ++G+        +  E  + +        P F    Y A I EN    S+++  +     +
Sbjct  637  QNGKVDDRRFQSTPEERLSIVG-GKRAPQFYMPSYEAEIPENQKKDSDIISVKA----KS  691

Query  248  YTDNWLAVYFFTSGNEGNWFEIQTDPRTNEGILKVVKALDYEQLQSVKLS----IAVKNK  303
            + D  +        G     F I          GI+K+ K LD+E L+   +      A ++
Sbjct  692  FADREIRYTLKAQGQGAGTFNIGP----TSGIVKLAKELDFEDLRQPHVYSLVVTATEDS  747

Query  304  AEFHQSVISRYRVQSTPVTIQVINVREGIAFRPASKTFTVQKGISSKKLVDYILGT----  359
              F  SV           +TI+V +V +       +  F +    +     D  LGT
Sbjct  748  GGFSTSV---------ELTIRVTDVNDN------APKFELPDYQAHNVDEDIPLGTSILK  792

Query  360  YQAIDEDTNKAASNVKYVMGRN----DGGYLMIDSKTAEIKFVKNMNRDSTFIVNKTITA  415
             +A+D D+  A + ++Y++  +    D   +++++K  +     N N    F+V
Sbjct  793  VKAMDADSG-ANAEIEYLVSDDHFAVDSNGIIVNNKQLD---ADNNNAYYEFVVTAKDKG  848

Query  416  EVLAIDEYTGKTSTGTVYVRVPDFNDNCP  444
            E         KT T TV V   + ND  P
Sbjct  849  E-------PAKTGTATVRVYTKNKNDEEP  870

>AMAC_003346.P3007 | transcript=AMAC_003346.R3007 | gene=AMAC_003346
| organism=Anopheles_maculipalpis_AmacGA1 | gene_product=neural-cadherin,
transcript variant X12 | transcript_product=neural-cadherin,
transcript variant X11 | location=AmacGA1_3RL:17033394-17218514(+)
| protein_length=3106 | sequence_SO=chromosome
| SO=protein_coding_gene | is_pseudo=false
Length=3106

 Score = 147 bits (370),  Expect = 3e-35, Method: Compositional matrix adjust.
 Identities = 128/446 (29%), Positives = 217/446 (49%), Gaps = 36/446 (8%)

Query  20    PIAKITS---DYQATQKITYRISGVGID--QPPFGIFVVDKNTGDINITAIVDREET---  71
             PI ++T+    D    Q I Y ++G GID    P    F +++ TG+I +    +DR++
Sbjct  1547  PIMQVTATDGDKDRPQNIVYFLTGQGIDPDNPANSKFDINRTTGEIFVLKPLDRDQPNGR  1606

Query  72    PSFLITCRALNAQGLDVEKPLILTVKILDINDNPPVFSQQIFMGEIEENSASNSLVMILN  131
             P +  T  A +  G  +      + V + DINDN P+F Q ++ G + EN  +  +VM +
Sbjct  1607  PQWRFTVFAQDEGGEGLVGYADVQVNLKDINDNAPIFPQGVYFGNVTENGTAGMVVMTMT  1666

Query  132   ATDADEPNH-LNSKIAFKI---VSQEPAGTPMFLLSRNTGEVRTLTNSLDREQASSYRLV  187
             A D D+PN   N+K+ + I   V +E  G+P+F +  +TG ++T    LDRE+   Y +
```

```
Sbjct  1667  AVDYDDPNEGTNAKLIYSIEKNVIEEETGSPIFEIEADTGVIKTAVCCLDRERTPDYSIQ  1726

Query  188   VSGADKDGEGLSTQCECNIKVKDVNDNFPMFRDSQYSARIEENILSS--ELLRFQVTDLD  245
             V     DG GL       +I+VKD+ND  P F   ++   ++E   ++  E+    VT  D
Sbjct  1727  VVAM--DGGGLKGTGTASIRVKDINDMPPQFTKDEWFTEVDETDGTNLPEMPILTVTVHD  1784

Query  246   EEYTDNWLAVYFFTSGNEGNWFEIQTDPRTNEGI--LKVVKALDYE---QLQSVKLSIAV  300
             E+ T+ +        SG    + F +     R N+G   LK+V+ LDYE   Q    +  I V
Sbjct  1785  EDETNKFQYKVIDNSGYGADKFTMV---RNNDGTGSLKIVQPLDYEDPLQSNGFRFRIQV  1841

Query  301   KNKAEFHQSVISRYRVQSTPVTIQVINVREGIAFRPASKTFTVQKGISSKKLVDYILGTY  360
              +K E + +    +Y V   + V +++ ++ +    +P  +   ++  +     V   L T+
Sbjct  1842  NDKGEDNDN--DKYHVAYSWVVVKLRDINDN---KPQFERPNIEVSVYENADVGKTLETF  1896

Query  361   QAIDEDTNKAASNVKYVMGR-NDGGYLMIDSKTAEIKFVKNMNRDSTFIVNKTITAEVLA  419
             +A D D     S V Y + R +D        ++    +   +N++R+    V       ++LA
Sbjct  1897  KATDPDQG-GKSKVSYAIDRSSDRQRQFSINQEGTVTIQRNLDRE----VTPRHQVKILA  1951

Query  420   IDE-YTGKTSTGTVYVRVPDFNDNCP  444
             ID+    KT+T T+ V V D NDN P
Sbjct  1952  IDDGIPPKTATATLTVIVQDINDNPP  1977

 Score = 77.0 bits (188),  Expect = 8e-13, Method: Compositional matrix adjust.
 Identities = 103/418 (25%), Positives = 174/418 (42%), Gaps = 50/418 (12%)

Query  50    IFVVDKNTGDINITAIVDREETPSFLITCRAL----------NAQGLDVEKPLILTVKIL  99
              F ++  +G+I    + DRE+  ++ +   A            N Q   V K   + + I
Sbjct  1363  FFEINLQSGEIFTKVVFDREKQGAYALEVEARDGAPSARPNSNQQPNSVTK--FIRIGIA  1420

Query  100   DINDNPPVFSQQIFMGEIEENSASNSLVMILNATDADEPNHLNSKIAFKIVSQEPAGTPM  159
             D NDNPP F + ++  E++EN       V+ + A D DE    +S+I ++I S    G
Sbjct  1421  DKNDNPPYFDKALYEAEVDENEDIQHTVLTVTAKDHDE----SSRIRYEITSGNIGGA--  1474

Query  160   FLLSRNTGEVRTLTNSLDREQASSYRLVVSGADKDGEGLSTQCECNIKVKDVNDNFPMFR  219
             F +   TG +  +  +LD E    Y L +  +D   E  +       I VKDVND  P F
Sbjct  1475  FAVKNMTGAI-YVAGALDYETRKRYHLTLVASDSLNENETIIV---INVKDVNDMPPSFP  1530

Query  220   DSQYSARIEENI-LSSELLRFQVTDLDEEYTDNWLAVYFFTSG--NEGNWFEIQTDPRTN  276
                Y   ++E + +   +++   TD D++   N   VYF T    +  N    + D
Sbjct  1531  QQVYERTMDEELAVPFPIMQVTATDGDKDRPQN--IVYFLTGQGIDPDNPANSKFDINRT  1588

Query  277   EGILKVVKALDYEQLQS-VKLSIAVKNKAEFHQSVISRYRVQSTPVTIQVINVREGIAFR  335
              G + V+K LD +Q     +    V  + E  + ++    VQ     + + ++ +
Sbjct  1589  TGEIFVLKPLDRDQPNGRPQWRFTVFAQDEGGEGLVGYADVQ-----VNLKDINDNAPIF  1643

Query  336   PASKTFTVQKGISSKKLVDYILGTYQAIDEDTNKAASNVK--YVMGRN------DGGYLM  387
             P    F     ++        ++ T  A+D D     +N K  Y + +N
Sbjct  1644  PQGVYF---GNVTENGTAGMVVMTMTAVDYDDPNEGTNAKLIYSIEKNVIEEETGSPIFE  1700

Query  388   IDSKTAEIK-FVKNMNRDSTFIVNKTITAEVLAIDEYTGKTSTGTVYVRVPDFNDNCP  444
             I++ T  IK  V  ++R+ T        + +V+A+D   G   TGT  +RV D ND  P
Sbjct  1701  IEADTGVIKTAVCCLDRERT----PDYSIQVVAMDG-GGLKGTGTASIRVKDINDMPP  1753

 Score = 76.3 bits (186),  Expect = 1e-12, Method: Compositional matrix adjust.
 Identities = 89/361 (25%), Positives = 167/361 (46%), Gaps = 36/361 (10%)

Query  51    FVVDKNTGDINITAIVDRE-ETPSFL-ITCRALNAQGLDVEKPLILTVKILDINDNPPVF  108
             F VD+ TG+++    + DRE +    F+ +T +A +     +E      TV+I D+NDNPP+F
Sbjct  1033  FTVDEETGEVSTNKVFDREGDDGKFVSVTVKATDQGEPSLEGVCSFTVEITDVNDNPPLF  1092

Query  109   SQQIFMGEIEENSASNSLVMILNATDADEPNHLNSKIAFKIVS-QEPAGTPMFLLSRNTG  167
              +Q ++  ++++++   + ++ ++A+D D  N  N  I + + +   P     F +   +G
Sbjct  1093  DRQKYVENVKQDASIGTNILRVSASDEDADN--NGAIVYTLSAPHSPNDLEYFEIQPESG  1150

Query  168   EVRTLTNSLDREQASSYRLVVSGADKDGEGLSTQCECNIKVKDVNDNFPMFRDSQYSA-R  226
              +  L    LDRE    +Y+L      DK     LS   E  I V D  +N P++ ++ Y
Sbjct  1151  WI-VLKKPLDRE---TYKLEAMAQDKGYPPLSRTVEVQIDVVDRANNPPVWDNTVYGPIY  1206

Query  227   IEENI-LSSELLRFQVTDLDEEYTDNWLAVYFFTSG-----NEGNWFEIQTDPRTNE--G  278
```

```
             ++EN+ +  +++  + +   E    N    Y    G     N+ + F +Q  P   +
Sbjct  1207  VKENMPVGGKVVSIKASSGIE---GNPTVFYRLMPGSTAQTNKLHTFYLQQRPDNGDTWA  1263

Query  279   ILKVVKALDYEQLQSVKLSIAVKNKAEFHQSVISRYRVQSTPVTIQVINVREGIAFRPAS  338
              +KV   LD+E ++   L++ V+N         ++       V IQ+ +V + I
Sbjct  1264  DIKVNHPLDFETIKEYNLTVRVENNG-------AQQLASEATVYIQLEDVNDEIPLFTER  1316

Query  339   KTFTVQKGISSKKLVDYILGTYQAIDEDTNKAASNVKYVM---GRNDG-GYLMIDSKTAE  394
             +  TV +G      V  +     AID+D     + V Y +    RN+G  +  I+ ++ E
Sbjct  1317  EQETVLEGEPIGTKVTQV----NAIDKDGTFPNNQVYYYIVDSPRNEGKDFFEINLQSGE  1372

Query  395   I  395
             I
Sbjct  1373  I  1373

 Score = 55.8 bits (133),  Expect = 3e-06, Method: Compositional matrix adjust.
 Identities = 49/158 (31%), Positives = 78/158 (49%), Gaps = 24/158 (15%)

Query  102   NDNPPVFSQQIFMGEIEENSASNSLVMILNATDADEPNHLNSKIAFKIVSQEPAGTP--M  159
             ND  P FSQQ++   ++EN+  N+LV  + A+D D  N     + F  V     GT
Sbjct  866   NDEEPKFSQQVYTPNVDENAGPNTLVTTVVASDKDGDN-----VRFGFVG---GGTSSGQ  917

Query  160   FLLSRNTGEVRTLTN--SLDREQASSYRLVVSGADK-------DGEGLSTQCECNIKVKD  210
             F++   TG +R  +   SLDR++   Y L V+  D        D    ++     + + D
Sbjct  918   FVIEEITGVIRLHSKAISLDRDK---YELNVTAMDDGSCCVNGDATIHTSTAVVVVFITD  974

Query  211   VNDNFPMFRD-SQYSARIEENILS-SELLRFQVTDLDE  246
             VNDN P+F+D   Y  ++EE   + S +++ Q TD D+
Sbjct  975   VNDNKPVFKDCGTYYPKVEEGAPNGSPVIKVQATDEDK  1012

 Score = 53.9 bits (128),  Expect = 1e-05, Method: Compositional matrix adjust.
 Identities = 70/306 (23%), Positives = 126/306 (41%), Gaps = 30/306 (10%)

Query  20    PIAKIT-SDYQATQKITYRI---SGVGIDQPPFGIFVVDKNTGDINITAIVDRE---ETP  72
             PI  +T  D   T K  Y++   SG G D+  F +   +  TG + I   +D E   ++
Sbjct  1776  PILTVTVHDEDETNKFQYKVIDNSGYGADK--FTMVRNNDGTGSLKIVQPLDYEDPLQSN  1833

Query  73    SFLITCRALNAQGLDVEKPLI------LTVKILDINDNPPVFSQQIFMGEIEENSASNSL  126
              F    + +N +G D +          + VK+ DINDN P F +      + EN+
Sbjct  1834  GFRFRIQ-VNDKGEDNDNDKYHVAYSWVVVKLRDINDNKPQFERPNIEVSVYENADVGKT  1892

Query  127   VMILNATDADEPNHLNSKIAFKIVSQEPAGTPMFLLSRNTGEVRTLTNSLDREQASSYRL  186
             +    ATD D+     SK+++ I   + +       S N     T+  +LDRE    +++
Sbjct  1893  LETFKATDPDQGG--KSKVSYAI---DRSSDRQRQFSINQEGTVTIQRNLDREVTPRHQV  1947

Query  187   VVSGADKDGEGLSTQCECNIKVKDVNDNFPMFRDSQYSARIEENILSSELLRFQVTDLDE  246
              +   D      +      + V+D+NDN P F    Y   + E++   +++    TD D+
Sbjct  1948  KILAIDDGIPPKTATATLTVIVQDINDNPPKFL-KDYRPVLPEHVPPRKVVEILATDDDD  2006

Query  247   EYTDNWLAVYFFTSGNEGNW----FEIQTDPRTNEG----ILKVVKALDYEQLQSVKLSI  298
                 N     F       +     F+++ D +   G    I+  +++ D EQ +   + I
Sbjct  2007  RSKSNGPPFQFRLDPGADDIIRASFKVEQDQKGANGDGMAIVSSLRSFDREQQKEYLIPI  2066

Query  299   AVKNKA  304
              +K+
Sbjct  2067  VIKDHG  2072

 Score = 52.4 bits (124),  Expect = 3e-05, Method: Compositional matrix adjust.
 Identities = 97/398 (24%), Positives = 163/398 (41%), Gaps = 54/398 (14%)

Query  106   PVFSQQIFMGEIEENSASNSLVMILNATDADEPNHLNSKIAFKIVSQ-EPAGTPMFLLSR  164
             P F    +  EI EN   +S ++ + A      +  + +I + + +Q + AGT  F +
Sbjct  663   PQFYMPSYEAEIPENQKKDSDIISVKAK-----SFADREIRYTLKAQGQGAGT--FNIGP  715

Query  165   NTGEVRTLTNSLDRE---QASSYRLVVSGADKDGEGLSTQCECNIKVKDVNDNFPMFRDS  221
              +G V+ L   LD E   Q   Y LVV+ A +D  G ST  E  I+V DVNDN P F
Sbjct  716   TSGIVK-LAKELDFEDLRQPHVYSLVVT-ATEDSGGFSTSVELTIRVTDVNDNAPKFELP  773
```

```
Query  222   QYSAR-IEENI-LSSELLRFQVTDLDEEYTDNWLAVYFFTSGNEGNWFEIQTDPRTNEGI  279
              Y A  ++E+I L + +L+ +  D D         + +  S +    F + ++     GI
Sbjct  774   DYQAHNVDEDIPLGTSILKVKAMDADSGAN---AEIEYLVSDDH---FAVDSN-----GI  822

Query  280   LKVVKALDYEQLQS-VKLSIAVKNKAEFHQSVISRYRVQSTPVTIQVINVREGIAFRPAS  338
             +   K LD +   +  +  +  K+K E  ++  +  RV +            E   F
Sbjct  823   IVNNKQLDADNNNAYYEFVVTAKDKGEPAKTGTATVRVYTKNKN------DEEPKFSQQV  876

Query  339   KTFTVQKGISSKKLVDYILGTYQAIDEDTNKAASNVK--YVMGRNDGGYLMIDSKTAEIK  396
              T  V +      LV  ++          ++K   NV+  +V G    G  +I+  T  I+
Sbjct  877   YTPNVDENAGPNTLVTTVVA--------SDKDGDNVRFGFVGGGTSSGQFVIEEITGVIR  928

Query  397   FVK---NMNRDSTFI-VNKTITAEVLAIDEYTGKTSTGTVYVRVPDFNDN------CPTA  446
                   +++RD   + V            + T  TST  V V + D NDN      C T
Sbjct  929   LHSKAISLDRDKYELNVTAMDDGSCCVNGDATIHTSTAVVVVFITDVNDNKPVFKDCGTY  988

Query  447   VLE-KDAVCSSSPSVVVSARTLNNRYTGPYTFALEDQP  483
               + ++   + SP + V A   +    G   +++  QP
Sbjct  989   YPKVEEGAPNGSPVIKVQATDEDKGVNGQVKYSIVQQP  1026

 Score = 44.3 bits (103),  Expect = 0.009, Method: Compositional matrix adjust.
 Identities = 48/199 (24%), Positives = 73/199 (37%), Gaps = 19/199 (10%)

Query  27    DYQATQKITYRISGVGIDQPPFGIFVVDKNTGDINITAIVDREETPSFLITCRALNAQGL  86
             D     K++Y I      Q  F I        G + I   +DRE TP   +    A++
Sbjct  1902  DQGGKSKVSYAIDRSSDRQRQFSI----NQEGTVTIQRNLDREVTPRHQVKILAIDDGIP  1957

Query  87    DVEKPLILTVKILDINDNPPVFSQQIFMGEIEENSASNSLVMILNATDADEPNHLNSKIA  146
                    LTV + DINDNPP F +  +    + E+      +V IL   D D
Sbjct  1958  PKTATATLTVIVQDINDNPPKFLKD-YRPVLPEHVPPRKVVEILATDDDDRSKSNGPPFQ  2016

Query  147   FKIVSQEPAGTPMFLLSRNTGEVRTLTN-----------SLDREQASSYRLVVSGADKDG  195
             F++   +P    +   S    + +   N            S DREQ   Y + +   D
Sbjct  2017  FRL---DPGADDIIRASFKVEQDQKGANGDGMAIVSSLRSFDREQQKEYLIPIVIKDHGN  2073

Query  196   EGLSTQCECNIKVKDVNDN  214
               ++      + + DVNDN
Sbjct  2074  PAMTGTSTLTVVIGDVNDN  2092

 Score = 40.8 bits (94),  Expect = 0.12, Method: Compositional matrix adjust.
 Identities = 55/242 (23%), Positives = 102/242 (42%), Gaps = 22/242 (9%)

Query  4    KFAKPCREGE--DNSKRNP-IAKITSDYQATQKITYRISGVGIDQPPFGIFVVDKNTGDI  60
            +F  P  E E  +N K++  I  + +   A ++I Y +   G      G F +   +G +
Sbjct  664  QFYMPSYEAEIPENQKKDSDIISVKAKSFADREIRYTLKAQGQGA---GTFNIGPTSGIV  720

Query  61   NITAIVDREE--TPSF--LITCRALNAQGLDVEKPLILTVKILDINDNPPVFSQQIFMGE  116
              +   +D E+   P    L+     ++ G       + LT+++ D+NDN P F    +
Sbjct  721  KLAKELDFEDLRQPHVYSLVVTATEDSGGFSTS--VELTIRVTDVNDNAPKFELPDYQAH  778

Query  117  -IEENSASNSLVMILNATDADEPNHLNSKIAFKIVSQEPAGTPMFLLSRNTGEVRTLTNS  175
             ++E+    + ++ + A DAD  +  N++I + +VS +      F +  N   V
Sbjct  779  NVDEDIPLGTSILKVKAMDAD--SGANAEIEY-LVSDD-----HFAVDSNGIIVNNKQLD  830

Query  176  LDREQASSYRLVVSGADKDGEGLSTQCECNIKVKDVNDNFPMFRDSQYSARIEENILSSE  235
             D   A  Y  VV+  DK     +      +  K+ ND  P F    Y+  ++EN   +
Sbjct  831  ADNNNA-YYEFVVTAKDKGEPAKTGTATVRVYTKNKNDEEPKFSQQVYTPNVDENAGPNT  889

Query  236  LL  237
            L+
Sbjct  890  LV  891

 Score = 39.3 bits (90),  Expect = 0.29, Method: Compositional matrix adjust.
 Identities = 89/389 (23%), Positives = 149/389 (38%), Gaps = 62/389 (16%)

Query  75   LITCRALNAQGLDVEKPLILTVKILDINDNPPVFSQQIF--MGEIEENSASNSLVMILNA  132
            +IT +  NA GL       + + + D+ND PP F  +        ++ N+  N+ V  L A
Sbjct  525  IITNQGHNA-GLKYTDNQRVIIHVKDVNDEPPYFINRPLPMQAVVQLNAPPNTPVFTLQA  583
```

```
Query  133  TDADEPNHLNSKIAFKIVSQEPAGTPMFLLSRNTGEVRTLTNSLDREQASSYRLVVSGAD  192
             D D     +  I + IV     G   F +   +G VRT    L  +     Y L V   D
Sbjct  584  RDPD----TDHNIHYFIVRDRTGG--RFEVDERSGVVRTRGTDL-FQLDMEYVLYVKAED  636

Query  193  KDGE----GLSTQCECNIKVKDVNDNFPMFRDSQYSARIEEN-ILSSELLRFQVTDLDEE  247
            ++G+        +  E  + +        P F    Y A I EN    S+++  +     +
Sbjct  637  QNGKVDDRRFQSTPEERLSIVG-GKRAPQFYMPSYEAEIPENQKKDSDIISVKA----KS  691

Query  248  YTDNWLAVYFFTSGNEGNWFEIQTDPRTNEGILKVVKALDYEQLQSVKLS----IAVKNK  303
            + D  +        G     F I         GI+K+ K LD+E L+   +       A ++
Sbjct  692  FADREIRYTLKAQGQGAGTFNIGP----TSGIVKLAKELDFEDLRQPHVYSLVVTATEDS  747

Query  304  AEFHQSVISRYRVQSTPVTIQVINVREGIAFRPASKTFTVQKGISSKKLVDYILGT----  359
              F  SV           +TI+V +V +        +  F +    +      D  LGT
Sbjct  748  GGFSTSV---------ELTIRVTDVNDN------APKFELPDYQAHNVDEDIPLGTSILK  792

Query  360  YQAIDEDTNKAASNVKYVMGRN----DGGYLMIDSKTAEIKFVKNMNRDSTFIVNKTITA  415
             +A+D D+  A + ++Y++  +    D   +++++K  +     N N    F+V
Sbjct  793  VKAMDADSG-ANAEIEYLVSDDHFAVDSNGIIVNNKQLD---ADNNNAYYEFVVTAKDKG  848

Query  416  EVLAIDEYTGKTSTGTVYVRVPDFNDNCP  444
            E          KT T TV V   + ND  P
Sbjct  849  E-------PAKTGTATVRVYTKNKNDEEP  870

>ASTEI20_040830.P55062 | transcript=ASTEI20_040830.R55062 | gene=ASTEI20_040830
| organism=Anopheles_stephensi_UCISS2018 |
gene_product=neural-cadherin, transcript variant X5 |
transcript_product=neural-cadherin,
transcript variant X16 | location=CM023249:57079885-57269506(-)
| protein_length=3107 | sequence_SO=chromosome
| SO=protein_coding_gene | is_pseudo=false
Length=3107

 Score = 147 bits (370),  Expect = 3e-35, Method: Compositional matrix adjust.
 Identities = 128/446 (29%), Positives = 217/446 (49%), Gaps = 36/446 (8%)

Query  20    PIAKITS---DYQATQKITYRISGVGID--QPPFGIFVVDKNTGDINITAIVDREET---  71
             PI ++T+    D    Q I Y ++G GID   P    F +++ TG+I +    +DR++
Sbjct  1548  PIMQVTATDGDKDRPQNIVYFLTGQGIDPDNPANSKFDINRTTGEIFVLKPLDRDQPNGR  1607

Query  72    PSFLITCRALNAQGLDVEKPLILTVKILDINDNPPVFSQQIFMGEIEENSASNSLVMILN  131
             P +  T  A +  G  +      + V + DINDN P+F Q ++ G + EN  +  +VM +
Sbjct  1608  PQWRFTVFAQDEGGEGLVGYADVQVNLKDINDNAPIFPQGVYFGNVTENGTAGMVVMTMT  1667

Query  132   ATDADEPNH-LNSKIAFKI---VSQEPAGTPMFLLSRNTGEVRTLTNSLDREQASSYRLV  187
             A D D+PN   N+K+ + I   V +E  G+P+F +  +TG ++T    LDRE+   Y +
Sbjct  1668  AVDYDDPNEGTNAKLIYSIEKNVIEEETGSPIFEIEADTGVIKTAVCCLDRERTPDYSIQ  1727

Query  188   VSGADKDGEGLSTQCECNIKVKDVNDNFPMFRDSQYSARIEENILSS--ELLRFQVTDLD  245
             V     DG GL       +I+VKD+ND  P F   ++   ++E   ++  E+    VT  D
Sbjct  1728  VVAM--DGGGLKGTGTASIRVKDINDMPPQFTKDEWFTEVDETDGTNLPEMPILTVTVHD  1785

Query  246   EEYTDNWLAVYFFTSGNEGNWFEIQTDPRTNEGI--LKVVKALDYE---QLQSVKLSIAV  300
             E+ T+ +        SG    + F +     R N+G   LK+V+ LDYE   Q     + I V
Sbjct  1786  EDETNKFQYKVIDNSGYGADKFTMV---RNNDGTGSLKIVQPLDYEDPLQSNGFRFRIQV  1842

Query  301   KNKAEFHQSVISRYRVQSTPVTIQVINVREGIAFRPASKTFTVQKGISSKKLVDYILGTY  360
              +K E + +   +Y V  + V +++ ++ +    +P  +   ++  +     V   L T+
Sbjct  1843  NDKGEDNDN--DKYHVAYSWVVVKLRDINDN---KPQFERPNIEVSVYENADVGKTLETF  1897

Query  361   QAIDEDTNKAASNVKYVMGR-NDGGYLMIDSKTAEIKFVKNMNRDSTFIVNKTITAEVLA  419
             +A D D     S V Y + R +D        ++    +   +N++R+     V     ++LA
Sbjct  1898  KATDPDQG-GKSKVSYAIDRSSDRQRQFSINQEGTVTIQRNLDRE----VTPRHQVKILA  1952

Query  420   IDE-YTGKTSTGTVYVRVPDFNDNCP  444
             ID+     KT+T T+ V V D NDN P
Sbjct  1953  IDDGIPPKTATATLTVIVQDINDNPP  1978
```

```
 Score = 76.6 bits (187),  Expect = 1e-12, Method: Compositional matrix adjust.
 Identities = 103/418 (25%), Positives = 174/418 (42%), Gaps = 50/418 (12%)

Query  50    IFVVDKNTGDINITAIVDREETPSFLITCRAL----------NAQGLDVEKPLILTVKIL  99
              F ++  +G+I    + DRE+  ++ +   A           N Q   V K   + + I
Sbjct  1364  FFEINLQSGEIFTKVVFDREKQGAYALEVEARDGAPSARPNSNQQPNSVTK--FIRIGIA  1421

Query  100   DINDNPPVFSQQIFMGEIEENSASNSLVMILNATDADEPNHLNSKIAFKIVSQEPAGTPM  159
             D NDNPP F + ++  E++EN      V+ + A D DE    +S+I ++I S    G
Sbjct  1422  DKNDNPPYFDKALYEAEVDENEDIQHTVLTVTAKDHDE----SSRIRYEITSGNIGGA--  1475

Query  160   FLLSRNTGEVRTLTNSLDREQASSYRLVVSGADKDGEGLSTQCECNIKVKDVNDNFPMFR  219
             F +   TG +  +  +LD E    Y L +  +D   E  +      I VKDVND  P F
Sbjct  1476  FAVKNMTGAI-YVAGALDYETRKRYHLTLVASDSLNENETIIV---INVKDVNDMPPSFP  1531

Query  220   DSQYSARIEENI-LSSELLRFQVTDLDEEYTDNWLAVYFFTSG--NEGNWFEIQTDPRTN  276
                Y   ++E + +   +++   TD D++   N   VYF T    +  N    + D
Sbjct  1532  QQVYERTMDEELAVPFPIMQVTATDGDKDRPQN--IVYFLTGQGIDPDNPANSKFDINRT  1589

Query  277   EGILKVVKALDYEQLQS-VKLSIAVKNKAEFHQSVISRYRVQSTPVTIQVINVREGIAFR  335
              G + V+K LD +Q     +    V  + E  + ++    VQ     + + ++ +
Sbjct  1590  TGEIFVLKPLDRDQPNGRPQWRFTVFAQDEGGEGLVGYADVQ-----VNLKDINDNAPIF  1644

Query  336   PASKTFTVQKGISSKKLVDYILGTYQAIDEDTNKAASNVK--YVMGRN------DGGYLM  387
             P    F     ++       ++ T  A+D D     +N K  Y + +N
Sbjct  1645  PQGVYF---GNVTENGTAGMVVMTMTAVDYDDPNEGTNAKLIYSIEKNVIEEETGSPIFE  1701

Query  388   IDSKTAEIK-FVKNMNRDSTFIVNKTITAEVLAIDEYTGKTSTGTVYVRVPDFNDNCP  444
             I++ T  IK  V  ++R+ T       + +V+A+D   G   TGT  +RV D ND  P
Sbjct  1702  IEADTGVIKTAVCCLDRERT----PDYSIQVVAMDG-GGLKGTGTASIRVKDINDMPP  1754

 Score = 76.3 bits (186),  Expect = 1e-12, Method: Compositional matrix adjust.
 Identities = 90/363 (25%), Positives = 169/363 (47%), Gaps = 40/363 (11%)

Query  51    FVVDKNTGDINITAIVDRE-ETPSFL-ITCRALNAQGLDVEKPLILTVKILDINDNPPVF  108
             F VD+ TG+++   + DRE +   F+ +T +A +     +E     TV+I D+NDNPP+F
Sbjct  1034  FTVDEETGEVSTNKVFDREGDDGKFVSVTVKATDQGEPSLEGVCSFTVEITDVNDNPPLF  1093

Query  109   SQQIFMGEIEENSASNSLVMILNATDADEPNHLNSKIAFKIVS-QEPAGTPMFLLSRNTG  167
              +Q ++  ++++++  + ++ ++A+D D  N  N  I + + +   P     F +   +G
Sbjct  1094  DRQKYVENVKQDASIGTNILRVSASDEDADN--NGAIVYTLSAPHSPNDLEYFEIQPESG  1151

Query  168   EVRTLTNSLDREQASSYRLVVSGADKDGEGLSTQCECNIKVKDVNDNFPMFRDSQYSA-R  226
              +  L   LDRE   +Y+L     DK    LS   E  I V D  +N P++ ++ Y
Sbjct  1152  WI-VLKKPLDRE---TYKLEAMAQDKGYPPLSRTVEVQIDVVDRANNPPVWDNTVYGPIY  1207

Query  227   IEENI-LSSELLRFQVTDLDEEYTDNWLAVYFFTSG-----NEGNWFEIQTDPRTNE--G  278
             ++EN+ +  +++  + +   E    N    Y    G     N+ + F +Q  P   +
Sbjct  1208  VKENMPVGGKVVSIKASSGIE---GNPTVFYRLMPGSTAQTNKLHTFYLQQRPDNGDTWA  1264

Query  279   ILKVVKALDYEQLQSVKLSIAVKNKAEFHQSVISRYRVQSTPVTIQVINVREGIAFRPAS  338
              +KV   LD+E ++   L++ V+N         ++       V IQ+ +V + I
Sbjct  1265  DIKVNHPLDFETIKEYNLTVRVENNG-------AQQLASEATVYIQLEDVNDEIPLFTER  1317

Query  339   KTFTVQKG--ISSKKLVDYILGTYQAIDEDTNKAASNVKYVM---GRNDG-GYLMIDSKT  392
             +  TV +G  I +K      +    AID+D     + V Y +    RN+G  +  I+ ++
Sbjct  1318  EQETVLEGEPIGTK------VTQVNAIDKDGTFPNNQVYYYIVDSPRNEGKDFFEINLQS  1371

Query  393   AEI  395
              EI
Sbjct  1372  GEI  1374

 Score = 55.8 bits (133),  Expect = 3e-06, Method: Compositional matrix adjust.
 Identities = 49/158 (31%), Positives = 78/158 (49%), Gaps = 24/158 (15%)

Query  102   NDNPPVFSQQIFMGEIEENSASNSLVMILNATDADEPNHLNSKIAFKIVSQEPAGTP--M  159
             ND  P FSQQ++   ++EN+  N+LV  + A+D D  N     + F  V     GT
Sbjct  867   NDEEPKFSQQVYTPNVDENAGPNTLVTTVVASDKDGDN-----VRFGFVG---GGTSSGQ  918
```

```
Query  160   FLLSRNTGEVRTLTN--SLDREQASSYRLVVSGADK-------DGEGLSTQCECNIKVKD  210
             F++   TG +R  +   SLDR++   Y L V+  D         D    ++      + + D
Sbjct  919   FVIEEITGVIRLHSKAISLDRDK---YELNVTAMDDGSCCVNGDATIHTSTAVVVVFITD  975

Query  211   VNDNFPMFRD-SQYSARIEENILS-SELLRFQVTDLDE  246
             VNDN P+F+D   Y  ++EE   + S +++ Q TD D+
Sbjct  976   VNDNKPVFKDCGTYYPKVEEGAPNGSPVIKVQATDEDK  1013

 Score = 53.5 bits (127),  Expect = 1e-05, Method: Compositional matrix adjust.
 Identities = 70/306 (23%), Positives = 126/306 (41%), Gaps = 30/306 (10%)

Query  20    PIAKIT-SDYQATQKITYRI---SGVGIDQPPFGIFVVDKNTGDINITAIVDRE---ETP  72
             PI  +T  D   T K  Y++   SG G D+  F +   +  TG + I   +D E   ++
Sbjct  1777  PILTVTVHDEDETNKFQYKVIDNSGYGADK--FTMVRNNDGTGSLKIVQPLDYEDPLQSN  1834

Query  73    SFLITCRALNAQGLDVEKPLI------LTVKILDINDNPPVFSQQIFMGEIEENSASNSL  126
              F    + +N +G D +           + VK+ DINDN P F +       + EN+
Sbjct  1835  GFRFRIQ-VNDKGEDNDNDKYHVAYSWVVVKLRDINDNKPQFERPNIEVSVYENADVGKT  1893

Query  127   VMILNATDADEPNHLNSKIAFKIVSQEPAGTPMFLLSRNTGEVRTLTNSLDREQASSYRL  186
             +    ATD D+      SK+++ I    + +        S N      T+  +LDRE    +++
Sbjct  1894  LETFKATDPDQGG--KSKVSYAI---DRSSDRQRQFSINQEGTVTIQRNLDREVTPRHQV  1948

Query  187   VVSGADKDGEGLSTQCECNIKVKDVNDNFPMFRDSQYSARIEENILSSELLRFQVTDLDE  246
              +   D      +       + V+D+NDN P F    Y   + E++   +++    TD D+
Sbjct  1949  KILAIDDGIPPKTATATLTVIVQDINDNPPKFL-KDYRPVLPEHVPPRKVVEILATDDDD  2007

Query  247   EYTDNWLAVYFFTSGNEGNW----FEIQTDPRTNEG----ILKVVKALDYEQLQSVKLSI  298
                 N     F       +     F+++ D +   G    I+  +++ D EQ +    + I
Sbjct  2008  RSKSNGPPFQFRLDPGADDIIRASFKVEQDQKGANGDGMAIVSSLRSFDREQQKEYLIPI  2067

Query  299   AVKNKA  304
              +K+
Sbjct  2068  VIKDHG  2073

 Score = 52.4 bits (124),  Expect = 3e-05, Method: Compositional matrix adjust.
 Identities = 97/398 (24%), Positives = 163/398 (41%), Gaps = 54/398 (14%)

Query  106   PVFSQQIFMGEIEENSASNSLVMILNATDADEPNHLNSKIAFKIVSQ-EPAGTPMFLLSR  164
             P F    +  EI EN   +S ++ + A       +  + +I + + +Q + AGT  F +
Sbjct  664   PQFYMPSYEAEIPENQKKDSDIISVKAK-----SFADREIRYTLKAQGQGAGT--FNIGP  716

Query  165   NTGEVRTLTNSLDRE---QASSYRLVVSGADKDGEGLSTQCECNIKVKDVNDNFPMFRDS  221
              +G V+ L   LD E   Q   Y LVV+ A +D  G ST  E  I+V DVNDN P F
Sbjct  717   TSGIVK-LAKELDFEDLRQPHVYSLVVT-ATEDSGGFSTSVELTIRVTDVNDNAPKFELP  774

Query  222   QYSAR-IEENI-LSSELLRFQVTDLDEEYTDNWLAVYFFTSGNEGNWFEIQTDPRTNEGI  279
              Y A  ++E+I L + +L+ +  D D          + +  S +    F + ++     GI
Sbjct  775   DYQAHNVDEDIPLGTSILKVKAMDADSGAN---AEIEYLVSDDH---FAVDSN-----GI  823

Query  280   LKVVKALDYEQLQS-VKLSIAVKNKAEFHQSVISRYRVQSTPVTIQVINVREGIAFRPAS  338
             +   K LD +   +  +  + K+K E  ++  +  RV +            E   F
Sbjct  824   IVNNKQLDADNNNAYYEFVVTAKDKGEPAKTGTATVRVYTKNKN------DEEPKFSQQV  877

Query  339   KTFTVQKGISSKKLVDYILGTYQAIDEDTNKAASNVK--YVMGRNDGGYLMIDSKTAEIK  396
              T  V +       LV  ++          ++K   NV+  +V G    G  +I+  T  I+
Sbjct  878   YTPNVDENAGPNTLVTTVVA--------SDKDGDNVRFGFVGGGTSSGQFVIEEITGVIR  929

Query  397   FVK---NMNRDSTFI-VNKTITAEVLAIDEYTGKTSTGTVYVRVPDFNDN------CPTA  446
                   +++RD   + V              + T  TST  V V + D NDN      C T
Sbjct  930   LHSKAISLDRDKYELNVTAMDDGSCCVNGDATIHTSTAVVVVFITDVNDNKPVFKDCGTY  989

Query  447   VLE-KDAVCSSSSPSVVVSARTLNNRYTGPYTFALEDQP  483
              + ++   + SP + V A    +    G    +++  QP
Sbjct  990   YPKVEEGAPNGSPVIKVQATDEDKGVNGQVKYSIVQQP  1027

 Score = 44.3 bits (103),  Expect = 0.008, Method: Compositional matrix adjust.
 Identities = 48/199 (24%), Positives = 73/199 (37%), Gaps = 19/199 (10%)
```

```
Query  27    DYQATQKITYRISGVGIDQPPFGIFVVDKNTGDINITAIVDREETPSFLITCRALNAQGL  86
             D     K++Y I      Q  F I       G + I   +DRE TP   +    A++
Sbjct  1903  DQGGKSKVSYAIDRSSDRQRQFSI----NQEGTVTIQRNLDREVTPRHQVKILAIDDGIP  1958

Query  87    DVEKPLILTVKILDINDNPPVFSQQIFMGEIEENSASNSLVMILNATDADEPNHLNSKIA  146
                    LTV + DINDNPP F +  +   + E+     +V IL   D D
Sbjct  1959  PKTATATLTVIVQDINDNPPKFLKD-YRPVLPEHVPPRKVVEILATDDDDRSKSNGPPFQ  2017

Query  147   FKIVSQEPAGTPMFLLSRNTGEVRTLTN-----------SLDREQASSYRLVVSGADKDG  195
             F++   +P    +   S    + +   N           S DREQ   Y + +   D
Sbjct  2018  FRL---DPGADDIIRASFKVEQDQKGANGDGMAIVSSLRSFDREQQKEYLIPIVIKDHGN  2074

Query  196   EGLSTQCECNIKVKDVNDN  214
               ++      + + DVNDN
Sbjct  2075  PAMTGTSTLTVVIGDVNDN  2093

 Score = 40.8 bits (94),  Expect = 0.12, Method: Compositional matrix adjust.
 Identities = 54/235 (23%), Positives = 99/235 (42%), Gaps = 22/235 (9%)

Query  4    KFAKPCREGE--DNSKRNP-IAKITSDYQATQKITYRISGVGIDQPPFGIFVVDKNTGDI  60
            +F  P  E E  +N K++  I  + +   A ++I Y +   G      G F +   +G +
Sbjct  665  QFYMPSYEAEIPENQKKDSDIISVKAKSFADREIRYTLKAQGQGA---GTFNIGPTSGIV  721

Query  61   NITAIVDREE--TPSF--LITCRALNAQGLDVEKPLILTVKILDINDNPPVFSQQIFMGE  116
             +   +D E+   P    L+     ++ G      + LT+++ D+NDN P F    +
Sbjct  722  KLAKELDFEDLRQPHVYSLVVTATEDSGGFSTS--VELTIRVTDVNDNAPKFELPDYQAH  779

Query  117  -IEENSASNSLVMILNATDADEPNHLNSKIAFKIVSQEPAGTPMFLLSRNTGEVRTLTNS  175
             ++E+    + ++ + A DAD  +  N++I + +VS +      F +  N   V
Sbjct  780  NVDEDIPLGTSILKVKAMDAD--SGANAEIEY-LVSDD-----HFAVDSNGIIVNNKQLD  831

Query  176  LDREQASSYRLVVSGADKDGEGLSTQCECNIKVKDVNDNFPMFRDSQYSARIEEN  230
             D   A  Y  VV+  DK     +      +  K+ ND  P F    Y+  ++EN
Sbjct  832  ADNNNA-YYEFVVTAKDKGEPAKTGTATVRVYTKNKNDEEPKFSQQVYTPNVDEN  885

 Score = 39.3 bits (90),  Expect = 0.34, Method: Compositional matrix adjust.
 Identities = 55/225 (24%), Positives = 88/225 (39%), Gaps = 24/225 (11%)

Query  75   LITCRALNAQGLDVEKPLILTVKILDINDNPPVFSQQIF--MGEIEENSASNSLVMILNA  132
            +IT +  NA GL       + + + D+ND PP F  +       ++ N+  N+ V  L A
Sbjct  526  IITNQGHNA-GLKYTDNQRVIIHVKDVNDEPPYFINRPLPMQAVVQLNAPPNTPVFTLQA  584

Query  133  TDADEPNHLNSKIAFKIVSQEPAGTPMFLLSRNTGEVRTLTNSLDREQASSYRLVVSGAD  192
             D D     +  I + IV     G   F +   +G VRT    L  +    Y L V   D
Sbjct  585  RDPD----TDHNIHYFIVRDRTGG--RFEVDERSGVVRTRGTDL-FQLDMEYVLYVKAED  637

Query  193  KDGE----GLSTQCECNIKVKDVNDNFPMFRDSQYSARIEEN-ILSSELLRFQVTDLDEE  247
            ++G+       +  E  + +       P F    Y A I EN    S+++  +     +
Sbjct  638  QNGKVDDRRFQSTPEERLSIVG-GKRAPQFYMPSYEAEIPENQKKDSDIISVKA----KS  692

Query  248  YTDNWLAVYFFTSGNEGNWFEIQTDPRTNEGILKVVKALDYEQLQ  292
            + D  +       G     F I        GI+K+ K LD+E L+
Sbjct  693  FADREIRYTLKAQGQGAGTFNIGP----TSGIVKLAKELDFEDLR  733

>ASTEI20_040830.P55052 | transcript=ASTEI20_040830.R55052 | gene=ASTEI20_040830
| organism=Anopheles_stephensi_UCISS2018 |
gene_product=neural-cadherin, transcript variant X5 |
transcript_product=neural-cadherin,
transcript variant X15 | location=CM023249:57079885-57269506(-)
| protein_length=3107 | sequence_SO=chromosome
| SO=protein_coding_gene | is_pseudo=false
Length=3107

 Score = 147 bits (370),  Expect = 3e-35, Method: Compositional matrix adjust.
 Identities = 128/446 (29%), Positives = 217/446 (49%), Gaps = 36/446 (8%)

Query  20    PIAKITS---DYQATQKITYRISGVGID--QPPFGIFVVDKNTGDINITAIVDREET---  71
```

```
             PI ++T+   D    Q I Y ++G GID   P    F +++ TG+I +   +DR++
Sbjct  1548  PIMQVTATDGDKDRPQNIVYFLTGQGIDPDNPANSKFDINRTTGEIFVLKPLDRDQPNGR  1607

Query  72    PSFLITCRALNAQGLDVEKPLILTVKILDINDNPPVFSQQIFMGEIEENSASNSLVMILN  131
             P +  T  A +  G  +     + V + DINDN P+F Q ++ G + EN  +  +VM +
Sbjct  1608  PQWRFTVFAQDEGGEGLVGYADVQVNLKDINDNAPIFPQGVYFGNVTENGTAGMVVMTMT  1667

Query  132   ATDADEPNH-LNSKIAFKI---VSQEPAGTPMFLLSRNTGEVRTLTNSLDREQASSYRLV  187
             A D D+PN   N+K+ + I   V +E  G+P+F +  +TG ++T    LDRE+   Y +
Sbjct  1668  AVDYDDPNEGTNAKLIYSIEKNVIEEETGSPIFEIEADTGVIKTAVCCLDRERTPDYSIQ  1727

Query  188   VSGADKDGEGLSTQCECNIKVKDVNDNFPMFRDSQYSARIEENILSS--ELLRFQVTDLD  245
             V     DG GL      +I+VKD+ND  P F   ++   ++E   ++  E+    VT  D
Sbjct  1728  VVAM--DGGGLKGTGTASIRVKDINDMPPQFTKDEWFTEVDETDGTNLPEMPILTVTVHD  1785

Query  246   EEYTDNWLAVYFFTSGNEGNWFEIQTDPRTNEGI--LKVVKALDYE---QLQSVKLSIAV  300
             E+ T+ +       SG   + F +    R N+G   LK+V+ LDYE   Q    +  I V
Sbjct  1786  EDETNKFQYKVIDNSGYGADKFTMV---RNNDGTGSLKIVQPLDYEDPLQSNGFRFRIQV  1842

Query  301   KNKAEFHQSVISRYRVQSTPVTIQVINVREGIAFRPASKTFTVQKGISSKKLVDYILGTY  360
              +K E + +   +Y V  + V +++ ++ +    +P  +   ++  +     V   L T+
Sbjct  1843  NDKGEDNDN--DKYHVAYSWVVVKLRDINDN---KPQFERPNIEVSVYENADVGKTLETF  1897

Query  361   QAIDEDTNKAASNVKYVMGR-NDGGYLMIDSKTAEIKFVKNMNRDSTFIVNKTITAEVLA  419
             +A D D     S V Y + R +D       ++   +   +N++R+    V      ++LA
Sbjct  1898  KATDPDQG-GKSKVSYAIDRSSDRQRQFSINQEGTVTIQRNLDRE----VTPRHQVKILA  1952

Query  420   IDE-YTGKTSTGTVYYVRVPDFNDNCP  444
             ID+    KT+T T+ V V D NDN P
Sbjct  1953  IDDGIPPKTATATLTVIVQDINDNPP  1978

 Score = 78.6 bits (192),  Expect = 3e-13, Method: Compositional matrix adjust.
 Identities = 106/424 (25%), Positives = 180/424 (42%), Gaps = 51/424 (12%)

Query  53    VDKNTGDINITAIVDREETPSFLITCRALNAQGLDVEKPLILTVKILDINDNPPVFSQ-Q  111
             +D++  ++N+TA+ D         +C  +N          ++ V I D+NDN PVF
Sbjct  937   LDRDKYELNVTAMDDG--------SC-CVNGDATIHTSTAVVVVFITDVNDNKPVFKDCG  987

Query  112   IFMGEIEENSASNSLVMILNATDADEPNHLNSKIAFKIVSQEPAGTPMFLLSRNTGEVRT  171
              +  ++EE + + S V+ + ATD D+   +N ++ + IV Q       F +   TGEV T
Sbjct  988   TYYPKVEEGAPNGSPVIKVQATDEDK--GVNGQVKYSIVQQPNQKGTKFTVDEETGEVST  1045

Query  172   LTNSLDRE--QASSYRLVVSGADKDGEGLSTQCECNIKVKDVNDNFPMFRDSQYSARIEE  229
                  DRE        + V   D+    L   C   +++ DVNDN P+F   +Y   +++
Sbjct  1046  -NKVFDREGDDGKFVSVTVKATDQGEPSLEGVCSFTVEITDVNDNPPLFDRQKYVENVKQ  1104

Query  230   NI-LSSELLRFQVTDLDEEYTDNWLAVYFFT---SGNEGNWFEIQTDPRTNEGILKVVKA  285
             +  + + +LR  V+  DE+  +N   VY  +   S N+  +FEIQ  P +   +LK  K
Sbjct  1105  DASIGTNILR--VSASDEDADNNGAIVYTLSAPHSPNDLEYFEIQ--PESGWIVLK--KP  1158

Query  286   LDYEQLQSVKLSIAVKNK------AEFHQSVISRYRVQSTPVTIQV----INVREGIAFR  335
             LD E+    +L +   ++      A+    +    R    P+  +     I++RE +
Sbjct  1159  LDRERY---RLKVYASDRGDPPSTADVDVELDVVDRNNKPPIWDRTIYGPIHIRENVTVG  1215

Query  336   PASKTFTVQKGISSKKLVDYILGTYQAIDEDTNKAASNVKYVMGRNDGGYLMIDSKTAEI  395
                 +     GI     V Y L         TNK   +  Y+  R D G        A+I
Sbjct  1216  TVVTSVKASSGIEGNPTVFYRL--MPGSTAQTNKL--HTFYLQQRPDNG-----DTWADI  1266

Query  396   KFVKNMNRDSTFIVNKTITAEVLAIDEYTGKTSTGTVYYVRVPDFNDNCPTAV-LEKDAVC  454
             K    ++ ++    N T+  E     +     S  TVY+++ D ND  P     E++ V
Sbjct  1267  KVNHPLDFETIKEYNLTVRVENNGAQQL---ASEATVYIQLEDVNDEIPLFTEREQETVL  1323

Query  455   SSSP  458
                P
Sbjct  1324  EGEP  1327

 Score = 76.6 bits (187),  Expect = 1e-12, Method: Compositional matrix adjust.
 Identities = 103/418 (25%), Positives = 174/418 (42%), Gaps = 50/418 (12%)
```

```
Query  50    IFVVDKNTGDINITAIVDREETPSFLITCRAL----------NAQGLDVEKPLILTVKIL  99
              F ++  +G+I    + DRE+  ++ +   A           N Q   V K   + + I
Sbjct  1364  FFEINLQSGEIFTKVVFDREKQGAYALEVEARDGAPSARPNSNQQPNSVTK--FIRIGIA  1421

Query  100   DINDNPPVFSQQIFMGEIEENSASNSLVMILNATDADEPNHLNSKIAFKIVSQEPAGTPM  159
             D NDNPP F + ++  E++EN      V+ + A D DE    +S+I ++I S    G
Sbjct  1422  DKNDNPPYFDKALYEAEVDENEDIQHTVLTVTAKDHDE----SSRIRYEITSGNIGGA--  1475

Query  160   FLLSRNTGEVRTLTNSLDREQASSYRLVVSGADKDGEGLSTQCECNIKVKDVNDNFPMFR  219
             F +   TG +  +  +LD E    Y L +  +D   E  +      I VKDVND  P F
Sbjct  1476  FAVKNMTGAI-YVAGALDYETRKRYHLTLVASDSLNENETIIV---INVKDVNDMPPSFP  1531

Query  220   DSQYSARIEENI-LSSELLRFQVTDLDEEYTDNWLAVYFFTSG--NEGNWFEIQTDPRTN  276
                Y   ++E + +   +++   TD D++   N   VYF T    +  N    + D
Sbjct  1532  QQVYERTMDEELAVPFPIMQVTATDGDKDRPQN--IVYFLTGQGIDPDNPANSKFDINRT  1589

Query  277   EGILKVVKALDYEQLQS-VKLSIAVKNKAEFHQSVISRYRVQSTPVTIQVINVREGIAFR  335
              G + V+K LD +Q     +    V  + E  + ++    VQ     + + ++ +
Sbjct  1590  TGEIFVLKPLDRDQPNGRPQWRFTVFAQDEGGEGLVGYADVQ-----VNLKDINDNAPIF  1644

Query  336   PASKTFTVQKGISSKKLVDYILGTYQAIDEDTNKAASNVK--YVMGRN------DGGYLM  387
             P    F     ++       ++ T  A+D D     +N K  Y + +N
Sbjct  1645  PQGVYF---GNVTENGTAGMVVMTMTAVDYDDPNEGTNAKLIYSIEKNVIEEETGSPIFE  1701

Query  388   IDSKTAEIK-FVKNMNRDSTFIVNKTITAEVLAIDEYTGKTSTGTVYVRVPDFNDNCP  444
             I++ T  IK  V  ++R+ T       + +V+A+D   G   TGT  +RV D ND  P
Sbjct  1702  IEADTGVIKTAVCCLDRERT----PDYSIQVVAMDG-GGLKGTGTASIRVKDINDMPP  1754

 Score = 65.5 bits (158),  Expect = 3e-09, Method: Compositional matrix adjust.
 Identities = 85/359 (24%), Positives = 153/359 (43%), Gaps = 32/359 (9%)

Query  51    FVVDKNTGDINITAIVDRE-ETPSFL-ITCRALNAQGLDVEKPLILTVKILDINDNPPVF  108
             F VD+ TG+++   + DRE +   F+ +T +A +     +E     TV+I D+NDNPP+F
Sbjct  1034  FTVDEETGEVSTNKVFDREGDDGKFVSVTVKATDQGEPSLEGVCSFTVEITDVNDNPPLF  1093

Query  109   SQQIFMGEIEENSASNSLVMILNATDADEPNHLNSKIAFKIVS-QEPAGTPMFLLSRNTG  167
              +Q ++  ++++++  + ++ ++A+D D  N  N  I + + +   P     F +   +G
Sbjct  1094  DRQKYVENVKQDASIGTNILRVSASDEDADN--NGAIVYTLSAPHSPNDLEYFEIQPESG  1151

Query  168   EVRTLTNSLDREQASSYRLVVSGADKDGEGLSTQCECNIKVKDVNDNFPMFRDSQYSARI  227
              +  L   LDRE+   YRL V  +D+ G+  ST            +N P   D      I
Sbjct  1152  WI-VLKKPLDRER---YRLKVYASDR-GDPPSTADVDVELDVVDRNNKPPIWDRTIYGPI  1206

Query  228   EENILSSELLRFQVTDLDEEYTDNWLAVYFFTSG-----NEGNWFEIQTDPRTNE--GIL  280
                   +                N    Y    G     N+ + F +Q   P   +    +
Sbjct  1207  HIRENVTVGTVVTSVKASSGIEGNPTVFYRLMPGSTAQTNKLHTFYLQQRPDNGDTWADI  1266

Query  281   KVVKALDYEQLQSVKLSIAVKNKAEFHQSVISRYRVQSTPVTIQVINVREGIAFRPASKT  340
             KV   LD+E ++   L++ V+N         ++       V IQ+ +V + I      +
Sbjct  1267  KVNHPLDFETIKEYNLTVRVENNG-------AQQLASEATVYIQLEDVNDEIPLFTEREQ  1319

Query  341   FTVQKGISSKKLVDYILGTYQAIDEDTNKAASNVKYVM---GRNDG-GYLMIDSKTAEI  395
              TV +G      V  +    AID+D     + V Y +    RN+G  +  I+ ++ EI
Sbjct  1320  ETVLEGEPIGTKVTQV----NAIDKDGTFPNNQVYYYIVDSPRNEGKDFFEINLQSGEI  1374

 Score = 55.8 bits (133),  Expect = 3e-06, Method: Compositional matrix adjust.
 Identities = 49/158 (31%), Positives = 78/158 (49%), Gaps = 24/158 (15%)

Query  102   NDNPPVFSQQIFMGEIEENSASNSLVMILNATDADEPNHLNSKIAFKIVSQEPAGTP--M  159
             ND  P FSQQ++   ++EN+  N+LV  + A+D D  N     + F  V     GT
Sbjct  867   NDEEPKFSQQVYTPNVDENAGPNTLVTTVVASDKDGDN-----VRFGFVG---GGTSSGQ  918

Query  160   FLLSRNTGEVRTLTN--SLDREQASSYRLVVSGADK-------DGEGLSTQCECNIKVKD  210
             F++   TG +R  +   SLDR++   Y L V+  D        D    ++     + + D
Sbjct  919   FVIEEITGVIRLHSKAISLDRDK---YELNVTAMDDGSCCVNGDATIHTSTAVVVVFITD  975

Query  211   VNDNFPMFRD-SQYSARIEENILS-SELLRFQVTDLDE  246
             VNDN P+F+D   Y  ++EE   + S +++ Q TD D+
Sbjct  976   VNDNKPVFKDCGTYYPKVEEGAPNGSPVIKVQATDEDK  1013
```

```
 Score = 53.9 bits (128),  Expect = 1e-05, Method: Compositional matrix adjust.
 Identities = 70/306 (23%), Positives = 126/306 (41%), Gaps = 30/306 (10%)

Query  20    PIAKIT-SDYQATQKITYRI---SGVGIDQPPFGIFVVDKNTGDINITAIVDRE---ETP  72
             PI  +T  D   T K  Y++   SG G D+  F +   +  TG + I   +D E   ++
Sbjct  1777  PILTVTVHDEDETNKFQYKVIDNSGYGADK--FTMVRNNDGTGSLKIVQPLDYEDPLQSN  1834

Query  73    SFLITCRALNAQGLDVEKPLI------LTVKILDINDNPPVFSQQIFMGEIEENSASNSL  126
              F    + +N +G D +           + VK+ DINDN P F +       + EN+
Sbjct  1835  GFRFRIQ-VNDKGEDNDNDKYHVAYSWVVVKLRDINDNKPQFERPNIEVSVYENADVGKT  1893

Query  127   VMILNATDADEPNHLNSKIAFKIVSQEPAGTPMFLLSRNTGEVRTLTNSLDREQASSYRL  186
             +    ATD D+     SK+++ I   + +        S N     T+  +LDRE    +++
Sbjct  1894  LETFKATDPDQGG--KSKVSYAI---DRSSDRQRQFSINQEGTVTIQRNLDREVTPRHQV  1948

Query  187   VVSGADKDGEGLSTQCECNIKVKDVNDNFPMFRDSQYSARIEENILSSELLRFQVTDLDE  246
              +   D      +      + V+D+NDN P F    Y   + E++   +++    TD D+
Sbjct  1949  KILAIDDGIPPKTATATLTVIVQDINDNPPKFL-KDYRPVLPEHVPPRKVVEILATDDDD  2007

Query  247   EYTDNWLAVYFFTSGNEGNW----FEIQTDPRTNEG----ILKVVKALDYEQLQSVKLSI  298
                 N     F       +     F+++ D +   G    I+  +++ D EQ +   + I
Sbjct  2008  RSKSNGPPFQFRLDPGADDIIRASFKVEQDQKGANGDGMAIVSSLRSFDREQQKEYLIPI  2067

Query  299   AVKNKA  304
              +K+
Sbjct  2068  VIKDHG  2073

 Score = 52.4 bits (124),  Expect = 3e-05, Method: Compositional matrix adjust.
 Identities = 97/398 (24%), Positives = 163/398 (41%), Gaps = 54/398 (14%)

Query  106   PVFSQQIFMGEIEENSASNSLVMILNATDADEPNHLNSKIAFKIVSQ-EPAGTPMFLLSR  164
             P F    +  EI EN   +S ++ + A      +  + +I + + +Q + AGT  F +
Sbjct  664   PQFYMPSYEAEIPENQKKDSDIISVKAK-----SFADREIRYTLKAQGQGAGT--FNIGP  716

Query  165   NTGEVRTLTNSLDRE---QASSYRLVVSGADKDGEGLSTQCECNIKVKDVNDNFPMFRDS  221
              +G V+ L   LD E   Q   Y LVV+ A +D  G ST  E  I+V DVNDN P F
Sbjct  717   TSGIVK-LAKELDFEDLRQPHVYSLVVT-ATEDSGGFSTSVELTIRVTDVNDNAPKFELP  774

Query  222   QYSAR-IEENI-LSSELLRFQVTDLDEEYTDNWLAVYFFTSGNEGNWFEIQTDPRTNEGI  279
              Y A  ++E+I L + +L+ +  D D         + +  S +    F + ++     GI
Sbjct  775   DYQAHNVDEDIPLGTSILKVKAMDADSGAN---AEIEYLVSDDH---FAVDSN-----GI  823

Query  280   LKVVKALDYEQLQS-VKLSIAVKNKAEFHQSVISRYRVQSTPVTIQVINVREGIAFRPAS  338
             +   K LD +   +  +  +  K+K E  ++  +  RV +           E   F
Sbjct  824   IVNNKQLDADNNNAYYEFVVTAKDKGEPAKTGTATVRVYTKNKN------DEEPKFSQQV  877

Query  339   KTFTVQKGISSKKLVDYILGTYQAIDEDTNKAASNVK--YVMGRNDGGYLMIDSKTAEIK  396
              T  V +      LV  ++         ++K   NV+  +V G    G  +I+  T  I+
Sbjct  878   YTPNVDENAGPNTLVTTVVA--------SDKDGDNVRFGFVGGGTSSGQFVIEEITGVIR  929

Query  397   FVK---NMNRDSTFI-VNKTITAEVLAIDEYTGKTSTGTVYVRVPDFNDN------CPTA  446
                   +++RD   + V            + T  TST  V V + D NDN      C T
Sbjct  930   LHSKAISLDRDKYELNVTAMDDGSCCVNGDATIHTSTAVVVVFITDVNDNKPVFKDCGTY  989

Query  447   VLE-KDAVCSSSPSVVVSARTLNNRYTGPYTFALEDQP  483
               + ++   + SP + V A    +    G   +++  QP
Sbjct  990   YPKVEEGAPNGSPVIKVQATDEDKGVNGQVKYSIVQQP  1027

 Score = 44.3 bits (103),  Expect = 0.009, Method: Compositional matrix adjust.
 Identities = 48/199 (24%), Positives = 73/199 (37%), Gaps = 19/199 (10%)

Query  27    DYQATQKITYRISGVGIDQPPFGIFVVDKNTGDINITAIVDREETPSFLITCRALNAQGL  86
             D     K++Y I      Q  F I       G + I   +DRE TP    +   A++
Sbjct  1903  DQGGKSKVSYAIDRSSDRQRQFSI----NQEGTVTIQRNLDREVTPRHQVKILAIDDGIP  1958

Query  87    DVEKPLILTVKILDINDNPPVFSQQIFMGEIEENSASNSLVMILNATDADEPNHLNSKIA  146
                    LTV + DINDNPP F +  +   + E+     +V IL   D D
```

```
Sbjct  1959  PKTATATLTVIVQDINDNPPKFLKD-YRPVLPEHVPPRKVVEILATDDDDRSKSNGPPFQ  2017

Query  147   FKIVSQEPAGTPMFLLSRNTGEVRTLTN-----------SLDREQASSYRLVVSGADKDG  195
             F++   +P    +   S    + +   N           S DREQ   Y + +   D
Sbjct  2018  FRL---DPGADDIIRASFKVEQDQKGANGDGMAIVSSLRSFDREQQKEYLIPIVIKDHGN  2074

Query  196   EGLSTQCECNIKVKDVNDN  214
               ++      + + DVNDN
Sbjct  2075  PAMTGTSTLTVVIGDVNDN  2093

 Score = 40.8 bits (94),  Expect = 0.12, Method: Compositional matrix adjust.
 Identities = 54/235 (23%), Positives = 99/235 (42%), Gaps = 22/235 (9%)

Query  4    KFAKPCREGE--DNSKRNP-IAKITSDYQATQKITYRISGVGIDQPPFGIFVVDKNTGDI  60
            +F  P  E E  +N K++  I  + +   A ++I Y +   G      G F +   +G +
Sbjct  665  QFYMPSYEAEIPENQKKDSDIISVKAKSFADREIRYTLKAQGQGA---GTFNIGPTSGIV  721

Query  61   NITAIVDREE--TPSF--LITCRALNAQGLDVEKPLILTVKILDINDNPPVFSQQIFMGE  116
             +   +D E+   P    L+     ++ G      + LT+++ D+NDN P F    +
Sbjct  722  KLAKELDFEDLRQPHVYSLVVTATEDSGGFSTS--VELTIRVTDVNDNAPKFELPDYQAH  779

Query  117  -IEENSASNSLVMILNATDADEPNHLNSKIAFKIVSQEPAGTPMFLLSRNTGEVRTLTNS  175
             ++E+    + ++ + A DAD  +  N++I + +VS +      F +  N   V
Sbjct  780  NVDEDIPLGTSILKVKAMDAD--SGANAEIEY-LVSDD-----HFAVDSNGIIVNNKQLD  831

Query  176  LDREQASSYRLVVSGADKDGEGLSTQCECNIKVKDVNDNFPMFRDSQYSARIEEN  230
             D   A  Y  VV+  DK     +      +  K+ ND  P F    Y+  ++EN
Sbjct  832  ADNNNA-YYEFVVTAKDKGEPAKTGTATVRVYTKNKNDEEPKFSQQVYTPNVDEN  885

 Score = 39.3 bits (90),  Expect = 0.33, Method: Compositional matrix adjust.
 Identities = 55/225 (24%), Positives = 88/225 (39%), Gaps = 24/225 (11%)

Query  75   LITCRALNAQGLDVEKPLILTVKILDINDNPPVFSQQIF--MGEIEENSASNSLVMILNA  132
            +IT +  NA GL       + + + D+ND PP F  +       ++ N+  N+ V  L A
Sbjct  526  IITNQGHNA-GLKYTDNQRVIIHVKDVNDEPPYFINRPLPMQAVVQLNAPPNTPVFTLQA  584

Query  133  TDADEPNHLNSKIAFKIVSQEPAGTPMFLLSRNTGEVRTLTNSLDREQASSYRLVVSGAD  192
             D D     +  I + IV     G   F +   +G VRT    L  +    Y L V   D
Sbjct  585  RDPD----TDHNIHYFIVRDRTGG--RFEVDERSGVVRTRGTDL-FQLDMEYVLYVKAED  637

Query  193  KDGE----GLSTQCECNIKVKDVNDNFPMFRDSQYSARIEEN-ILSSELLRFQVTDLDEE  247
            ++G+       +  E  + +       P F    Y A I EN    S+++  +     +
Sbjct  638  QNGKVDDRRFQSTPEERLSIVG-GKRAPQFYMPSYEAEIPENQKKDSDIISVKA----KS  692

Query  248  YTDNWLAVYFFTSGNEGNWFEIQTDPRTNEGILKVVKALDYEQLQ  292
            + D  +       G     F I        GI+K+ K LD+E L+
Sbjct  693  FADREIRYTLKAQGQGAGTFNIGP----TSGIVKLAKELDFEDLR  733

>ACMO_007424.P13496 | transcript=ACMO_007424.R13496 | gene=ACMO_007424
| organism=Anopheles_coluzzii_MOPTI | gene_product=neural-cadherin,
transcript variant X4 | transcript_product=neural-cadherin,
transcript variant X14 | location=CM029349:40739311-40945968(+)
| protein_length=3099 | sequence_SO=chromosome
| SO=protein_coding_gene | is_pseudo=false
Length=3099

 Score = 147 bits (370),  Expect = 3e-35, Method: Compositional matrix adjust.
 Identities = 128/446 (29%), Positives = 217/446 (49%), Gaps = 36/446 (8%)

Query  20    PIAKITS---DYQATQKITYRISGVGID--QPPFGIFVVDKNTGDINITAIVDREET---  71
             PI ++T+   D    Q I Y ++G GID   P    F +++ TG+I +   +DR++
Sbjct  1539  PIMQVTATDGDKDRPQNIVYFLTGQGIDPDNPANSKFDINRTTGEIFVLKPLDRDQPNGR  1598

Query  72    PSFLITCRALNAQGLDVEKPLILTVKILDINDNPPVFSQQIFMGEIEENSASNSLVMILN  131
             P +  T  A +  G  +     + V + DINDN P+F Q ++ G + EN  +  +VM +
Sbjct  1599  PQWRFTVFAQDEGGEGLVGYADVQVNLKDINDNAPIFPQGVYFGNVTENGTAGMVVMTMT  1658

Query  132   ATDADEPNH-LNSKIAFKI---VSQEPAGTPMFLLSRNTGEVRTLTNSLDREQASSYRLV  187
```

```
             A D D+PN   N+K+ + I   V +E  G+P+F +  +TG ++T    LDRE+   Y +
Sbjct  1659  AVDYDDPNEGTNAKLIYSIEKNVIEEETGSPIFEIEADTGVIKTAVCCLDRERTPDYSIQ  1718

Query  188   VSGADKDGEGLSTQCECNIKVKDVNDNFPMFRDSQYSARIEENILSS--ELLRFQVTDLD  245
             V     DG GL       +I+VKD+ND  P F   ++   ++E   ++  E+    VT  D
Sbjct  1719  VVAM--DGGGLKGTGTASIRVKDINDMPPQFTKDEWFTEVDETDGTNLPEMPILTVTVHD  1776

Query  246   EEYTDNWLAVYFFTSGNEGNWFEIQTDPRTNEGI--LKVVKALDYE---QLQSVKLSIAV  300
             E+ T+ +        SG    + F +    R N+G   LK+V+ LDYE   Q    +  I V
Sbjct  1777  EDETNKFQYKVIDNSGYGADKFTMV---RNNDGTGSLKIVQPLDYEDPLQSNGFRFRIQV  1833

Query  301   KNKAEFHQSVISRYRVQSTPVTIQVINVREGIAFRPASKTFTVQKGISSKKLVDYILGTY  360
              +K E + +   +Y V  + V +++ ++ +    +P  +   ++  +     V   L T+
Sbjct  1834  NDKGEDNDN--DKYHVAYSWVVVKLRDINDN---KPQFERPNIEVSVYENADVGKTLETF  1888

Query  361   QAIDEDTNKAASNVKYVMGR-NDGGYLMIDSKTAEIKFVKNMNRDSTFIVNKTITAEVLA  419
             +A D D     S V Y + R +D        ++    +   +N++R+    V       ++LA
Sbjct  1889  KATDPDQG-GKSKVSYAIDRSSDRQRQFSINQEGTVTIQRNLDRE----VTPRHQVKILA  1943

Query  420   IDE-YTGKTSTGTVYYVRVPDFNDNCP  444
             ID+    KT+T T+ V V D NDN P
Sbjct  1944  IDDGIPPKTATATLTVIVQDINDNPP  1969

 Score = 78.6 bits (192),  Expect = 3e-13, Method: Compositional matrix adjust.
 Identities = 106/424 (25%), Positives = 180/424 (42%), Gaps = 51/424 (12%)

Query  53    VDKNTGDINITAIVDREETPSFLITCRALNAQGLDVEKPLILTVKILDINDNPPVFSQ-Q  111
             +D++  ++N+TA+ D          +C  +N          ++ V I D+NDN PVF
Sbjct  928   LDRDKYELNVTAMDDG--------SC-CVNGDATIHTSTAVVVVFITDVNDNKPVFKDCG  978

Query  112   IFMGEIEENSASNSLVMILNATDADEPNHLNSKIAFKIVSQEPAGTPMFLLSRNTGEVRT  171
              +  ++EE + + S V+ + ATD D+   +N ++ + IV Q        F +   TGEV T
Sbjct  979   TYYPKVEEGAPNGSPVIKVQATDEDK--GVNGQVKYSIVQQPNQKGTKFTVDEETGEVST  1036

Query  172   LTNSLDRE--QASSYRLVVSGADKDGEGLSTQCECNIKVKDVNDNFPMFRDSQYSARIEE  229
                  DRE         + V   D+    L   C   +++ DVNDN P+F   +Y   +++
Sbjct  1037  -NKVFDREGDDGKFVSVTVKATDQGEPSLEGVCSFTVEITDVNDNPPLFDRQKYVENVKQ  1095

Query  230   NI-LSSELLRFQVTDLDEEYTDNWLAVYFFT---SGNEGNWFEIQTDPRTNEGILKVVKA  285
             +  + + +LR  V+  DE+  +N   VY  +   S N+  +FEIQ  P +   +LK  K
Sbjct  1096  DASIGTNILR--VSASDEDADNNGAIVYTLSAPHSPNDLEYFEIQ--PESGWIVLK--KP  1149

Query  286   LDYEQLQSVKLSIAVKNK------AEFHQSVISRYRVQSTPVTIQV----INVREGIAFR  335
             LD E+    +L +   ++      A+    +    R    P+  +     I++RE +
Sbjct  1150  LDRERY---RLKVYASDRGDPPSTADVDVELDVVDRNNKPPIWDRTIYGPIHIRENVTVG  1206

Query  336   PASKTFTVQKGISSKKLVDYILGTYQAIDEDTNKAASNVKYVMGRNDGGYLMIDSKTAEI  395
                 +     GI     V Y L          TNK   +  Y+  R D G         A+I
Sbjct  1207  TVVTSVKASSGIEGNPTVFYRL--MPGSTAQTNKL--HTFYLQQRPDNG-----DTWADI  1257

Query  396   KFVKNMNRDSTFIVNKTITAEVLAIDEYTGKTSTGTVYYVRVPDFNDNCPTAV-LEKDAVC  454
             K    ++ ++    N T+  E     +      S  TVY+++ D ND  P     E++ V
Sbjct  1258  KVNHPLDFETIKEYNLTVRVENNGAQQL---ASEATVYIQLEDVNDEIPLFTEREQETVL  1314

Query  455   SSSP  458
                P
Sbjct  1315  EGEP  1318

 Score = 75.5 bits (184),  Expect = 2e-12, Method: Compositional matrix adjust.
 Identities = 103/418 (25%), Positives = 174/418 (42%), Gaps = 50/418 (12%)

Query  50    IFVVDKNTGDINITAIVDREETPSFLITCRAL----------NAQGLDVEKPLILTVKIL  99
              F ++  +G+I    + DRE+  ++ +   A            N Q   V K   + + I
Sbjct  1355  FFEINLQSGEIFTKVVFDREKQGAYALEVEARDGAPSARPNSNQQPNSVTK--FIRIGIA  1412

Query  100   DINDNPPVFSQQIFMGEIEENSASNSLVMILNATDADEPNHLNSKIAFKIVSQEPAGTPM  159
             D NDNPP F + ++  E++EN       V+ + A D DE    +S+I ++I S    G
Sbjct  1413  DKNDNPPYFDKALYEAEVDENEDIQHTVLTVTAKDHDE----SSRIRYEITSGNIGGA--  1466
```

```
Query  160   FLLSRNTGEVRTLTNSLDREQASSYRLVVSGADKDGEGLSTQCECNIKVKDVNDNFPMFR  219
             F +   TG +  +  +LD E    Y L +  +D   E  +      I VKDVND  P F
Sbjct  1467  FAVKNMTGAI-YVAGALDYETRKRYYLTLVASDSLNENETIIV---INVKDVNDMPPSFP  1522

Query  220   DSQYSARIEENI-LSSELLRFQVTDLDEEYTDNWLAVYFFTSG--NEGNWFEIQTDPRTN  276
                Y   ++E + +   +++   TD D++   N   VYF T    +  N    + D
Sbjct  1523  QQVYERTMDEELAVPFPIMQVTATDGDKDRPQN--IVYFLTGQGIDPDNPANSKFDINRT  1580

Query  277   EGILKVVKALDYEQLQS-VKLSIAVKNKAEFHQSVISRYRVQSTPVTIQVINVREGIAFR  335
              G + V+K LD +Q     +    V  + E  + ++    VQ     + + ++ +
Sbjct  1581  TGEIFVLKPLDRDQPNGRPQWRFTVFAQDEGGEGLVGYADVQ-----VNLKDINDNAPIF  1635

Query  336   PASKTFTVQKGISSKKLVDYILGTYQAIDEDTNKAASNVK--YVMGRN------DGGYLM  387
             P    F     ++       ++ T  A+D D     +N K  Y + +N
Sbjct  1636  PQGVYF---GNVTENGTAGMVVMTMTAVDYDDPNEGTNAKLIYSIEKNVIEEETGSPIFE  1692

Query  388   IDSKTAEIK-FVKNMNRDSTFIVNKTITAEVLAIDEYTGKTSTGTVYVRVPDFNDNCP  444
             I++ T  IK  V  ++R+ T       + +V+A+D   G   TGT  +RV D ND  P
Sbjct  1693  IEADTGVIKTAVCCLDRERT----PDYSIQVVAMDG-GGLKGTGTASIRVKDINDMPP  1745

 Score = 65.5 bits (158),  Expect = 3e-09, Method: Compositional matrix adjust.
 Identities = 85/359 (24%), Positives = 153/359 (43%), Gaps = 32/359 (9%)

Query  51    FVVDKNTGDINITAIVDRE-ETPSFL-ITCRALNAQGLDVEKPLILTVKILDINDNPPVF  108
             F VD+ TG+++   + DRE +   F+ +T +A +     +E     TV+I D+NDNPP+F
Sbjct  1025  FTVDEETGEVSTNKVFDREGDDGKFVSVTVKATDQGEPSLEGVCSFTVEITDVNDNPPLF  1084

Query  109   SQQIFMGEIEENSASNSLVMILNATDADEPNHLNSKIAFKIVS-QEPAGTPMFLLSRNTG  167
              +Q ++  ++++++  + ++ ++A+D D  N  N  I + + +   P     F +   +G
Sbjct  1085  DRQKYVENVKQDASIGTNILRVSASDEDADN--NGAIVYTLSAPHSPNDLEYFEIQPESG  1142

Query  168   EVRTLTNSLDREQASSYRLVVSGADKDGEGLSTQCECNIKVKDVNDNFPMFRDSQYSARI  227
              +  L   LDRE+   YRL V  +D+ G+  ST            +N P   D      I
Sbjct  1143  WI-VLKKPLDRER---YRLKVYASDR-GDPPSTADVDVELDVVDRNNKPPIWDRTIYGPI  1197

Query  228   EENILSSELLRFQVTDLDEEYTDNWLAVYFFTSG-----NEGNWFEIQTDPRTNE--GIL  280
                   +                N    Y    G     N+ + F +Q  P   +    +
Sbjct  1198  HIRENVTVGTVVTSVKASSGIEGNPTVFYRLMPGSTAQTNKLHTFYLQQRPDNGDTWADI  1257

Query  281   KVVKALDYEQLQSVKLSIAVKNKAEFHQSVISRYRVQSTPVTIQVINVREGIAFRPASKT  340
             KV   LD+E ++   L++ V+N         ++       V IQ+ +V + I      +
Sbjct  1258  KVNHPLDFETIKEYNLTVRVENNG-------AQQLASEATVYIQLEDVNDEIPLFTEREQ  1310

Query  341   FTVQKGISSKKLVDYILGTYQAIDEDTNKAASNVKYVM---GRNDG-GYLMIDSKTAEI  395
              TV +G      V  +     AID+D     + V Y +    RN+G  +  I+ ++ EI
Sbjct  1311  ETVLEGEPIGTKVTQV----NAIDKDGTFPNNQVYYYIVDSPRNEGKDFFEINLQSGEI  1365

 Score = 55.8 bits (133),  Expect = 3e-06, Method: Compositional matrix adjust.
 Identities = 49/158 (31%), Positives = 78/158 (49%), Gaps = 24/158 (15%)

Query  102   NDNPPVFSQQIFMGEIEENSASNSLVMILNATDADEPNHLNSKIAFKIVSQEPAGTP--M  159
             ND  P FSQQ++   ++EN+  N+LV  + A+D D  N     + F  V     GT
Sbjct  858   NDEEPKFSQQVYTPNVDENAGPNTLVTTVVASDKDGDN-----VRFGFVG---GGTSSGQ  909

Query  160   FLLSRNTGEVRTLTN--SLDREQASSYRLVVSGADK-------DGEGLSTQCECNIKVKD  210
             F++   TG +R  +   SLDR++   Y L V+  D        D    ++      + + D
Sbjct  910   FVIEEITGVIRLHSKAISLDRDK---YELNVTAMDDGSCCVNGDATIHTSTAVVVVFITD  966

Query  211   VNDNFPMFRD-SQYSARIEENILS-SELLRFQVTDLDE  246
             VNDN P+F+D   Y  ++EE   + S +++ Q TD D+
Sbjct  967   VNDNKPVFKDCGTYYPKVEEGAPNGSPVIKVQATDEDK  1004

 Score = 53.5 bits (127),  Expect = 1e-05, Method: Compositional matrix adjust.
 Identities = 70/306 (23%), Positives = 126/306 (41%), Gaps = 30/306 (10%)

Query  20    PIAKIT-SDYQATQKITYRI---SGVGIDQPPFGIFVVDKNTGDINITAIVDRE---ETP  72
             PI  +T  D   T K  Y++   SG G D+  F +   +  TG + I   +D E   ++
Sbjct  1768  PILTVTVHDEDETNKFQYKVIDNSGYGADK--FTMVRNNDGTGSLKIVQPLDYEDPLQSN  1825
```

```
Query  73    SFLITCRALNAQGLDVEKPLI------LTVKILDINDNPPVFSQQIFMGEIEENSASNSL  126
              F    + +N +G D +          + VK+ DINDN P F +      + EN+
Sbjct  1826  GFRFRIQ-VNDKGEDNDNDKYHVAYSWVVVKLRDINDNKPQFERPNIEVSVYENADVGKT  1884

Query  127   VMILNATDADEPNHLNSKIAFKIVSQEPAGTPMFLLSRNTGEVRTLTNSLDREQASSYRL  186
             +    ATD D+     SK+++ I    + +        S N     T+  +LDRE    +++
Sbjct  1885  LETFKATDPDQGG--KSKVSYAI---DRSSDRQRQFSINQEGTVTIQRNLDREVTPRHQV  1939

Query  187   VVSGADKDGEGLSTQCECNIKVKDVNDNFPMFRDSQYSARIEENILSSELLRFQVTDLDE  246
              +   D       +       + V+D+NDN P F    Y   + E++   +++    TD D+
Sbjct  1940  KILAIDDGIPPKTATATLTVIVQDINDNPPKFL-KDYRPVLPEHVPPRKVVEILATDDDD  1998

Query  247   EYTDNWLAVYFFTSGNEGNW----FEIQTDPRTNEG----ILKVVKALDYEQLQSVKLSI  298
                 N     F       +     F+++ D +   G    I+  +++ D EQ +   + I
Sbjct  1999  RSKSNGPPFQFRLDPGADDIIRASFKVEQDQKGANGDGMAIVSSLRSFDREQQKEYLIPI  2058

Query  299   AVKNKA  304
              +K+
Sbjct  2059  VIKDHG  2064

 Score = 52.0 bits (123),  Expect = 5e-05, Method: Compositional matrix adjust.
 Identities = 97/398 (24%), Positives = 163/398 (41%), Gaps = 54/398 (14%)

Query  106   PVFSQQIFMGEIEENSASNSLVMILNATDADEPNHLNSKIAFKIVSQ-EPAGTPMFLLSR  164
             P F    +  EI EN   +S ++ + A       +  + +I + + +Q + AGT  F +
Sbjct  655   PQFYMPSYEAEIPENQKKDSDIISVKAK-----SFADREIRYTLKAQGQGAGT--FNIGP  707

Query  165   NTGEVRTLTNSLDRE---QASSYRLVVSGADKDGEGLSTQCECNIKVKDVNDNFPMFRDS  221
              +G V+ L   LD E   Q   Y LVV+ A +D  G ST  E  I+V DVNDN P F
Sbjct  708   TSGIVK-LAKELDFEDLRQPHVYSLVVT-ATEDSGGFSTSVELTIRVTDVNDNAPKFELP  765

Query  222   QYSAR-IEENI-LSSELLRFQVTDLDEEYTDNWLAVYFFTSGNEGNWFEIQTDPRTNEGI  279
              Y A  ++E+I L + +L+ +  D D          + +  S +    F + ++     GI
Sbjct  766   DYQAHNVDEDIPLGTSILKVKAMDSDSGAN---AEIEYLVSDDH---FAVDSN-----GI  814

Query  280   LKVVKALDYEQLQS-VKLSIAVKNKAEFHQSVISRYRVQSTPVTIQVINVREGIAFRPAS  338
             +   K LD +   +  +  +  K+K E  ++  +  RV +             E   F
Sbjct  815   IVNNKQLDADNNNAYYEFVVTAKDKGEPAKTGTATVRVYTKNKN------DEEPKFSQQV  868

Query  339   KTFTVQKGISSKKLVDYILGTYQAIDEDTNKAASNVK--YVMGRNDGGYLMIDSKTAEIK  396
              T  V +      LV  ++          ++K   NV+  +V G    G  +I+  T  I+
Sbjct  869   YTPNVDENAGPNTLVTTVVA--------SDKDGDNVRFGFVGGGTSSGQFVIEEITGVIR  920

Query  397   FVK---NMNRDSTFI-VNKTITAEVLAIDEYTGKTSTGTVYVRVPDFNDN------CPTA  446
                    +++RD   + V              + T  TST  V V + D NDN      C T
Sbjct  921   LHSKAISLDRDKYELNVTAMDDGSCCVNGDATIHTSTAVVVVFITDVNDNKPVFKDCGTY  980

Query  447   VLE-KDAVCSSSSPSVVVSARTLNNRYTGPYTFALEDQP  483
               + ++   + SP + V A   +    G   +++  QP
Sbjct  981   YPKVEEGAPNGSPVIKVQATDEDKGVNGQVKYSIVQQP  1018

 Score = 44.3 bits (103),  Expect = 0.009, Method: Compositional matrix adjust.
 Identities = 48/199 (24%), Positives = 73/199 (37%), Gaps = 19/199 (10%)

Query  27    DYQATQKITYRISGVGIDQPPFGIFVVDKNTGDINITAIVDREETPSFLITCRALNAQGL  86
             D     K++Y I       Q  F I        G + I   +DRE TP   +   A++
Sbjct  1894  DQGGKSKVSYAIDRSSDRQRQFSI----NQEGTVTIQRNLDREVTPRHQVKILAIDDGIP  1949

Query  87    DVEKPLILTVKILDINDNPPVFSQQIFMGEIEENSASNSLVMILNATDADEPNHLNSKIA  146
                    LTV + DINDNPP F +  +    + E+     +V IL   D D
Sbjct  1950  PKTATATLTVIVQDINDNPPKFLKD-YRPVLPEHVPPRKVVEILATDDDDRSKSNGPPFQ  2008

Query  147   FKIVSQEPAGTPMFLLSRNTGEVRTLTN-----------SLDREQASSYRLVVSGADKDG  195
             F++   +P    +   S    + +   N           S DREQ    Y + +   D
Sbjct  2009  FRL---DPGADDIIRASFKVEQDQKGANGDGMAIVSSLRSFDREQQKEYLIPIVIKDHGN  2065

Query  196   EGLSTQCECNIKVKDVNDN  214
               ++       + + DVNDN
```

```
Sbjct  2066  PAMTGTSTLTVVIGDVNDN  2084

 Score = 39.7 bits (91),  Expect = 0.29, Method: Compositional matrix adjust.
 Identities = 89/389 (23%), Positives = 149/389 (38%), Gaps = 62/389 (16%)

Query  75   LITCRALNAQGLDVEKPLILTVKILDINDNPPVFSQQIF--MGEIEENSASNSLVMILNA  132
            +IT +  NA GL       + + + D+ND PP F  +       ++ N+  N+ V  L A
Sbjct  517  IITNQGHNA-GLKYTDNQRVIIHVKDVNDEPPYFINRPLPMQAVVQLNAPPNTPVFTLQA  575

Query  133  TDADEPNHLNSKIAFKIVSQEPAGTPMFLLSRNTGEVRTLTNSLDREQASSYRLVVSGAD  192
             D D     +  I + IV     G   F +    +G VRT    L  +    Y L V   D
Sbjct  576  RDPD----TDHNIHYFIVRDRTGG--RFEVDERSGVVRTRGTDL-FQLDMEYVLYVKAED  628

Query  193  KDGE----GLSTQCECNIKVKDVNDNFPMFRDSQYSARIEEN-ILSSELLRFQVTDLDEE  247
            ++G+       +  E  + +       P F    Y A I EN    S+++  +     +
Sbjct  629  QNGKVDDRRFQSTPEERLSIVG-GKRAPQFYMPSYEAEIPENQKKDSDIISVKA----KS  683

Query  248  YTDNWLAVYFFTSGNEGNWFEIQTDPRTNEGILKVVKALDYEQLQSVKLS----IAVKNK  303
            + D  +        G     F I        GI+K+ K LD+E L+   +       A ++
Sbjct  684  FADREIRYTLKAQGQGAGTFNIGP----TSGIVKLAKELDFEDLRQPHVYSLVVTATEDS  739

Query  304  AEFHQSVISRYRVQSTPVTIQVINVREGIAFRPASKTFTVQKGISSKKLVDYILGT----  359
              F  SV           +TI+V +V +        +  F +    +     D  LGT
Sbjct  740  GGFSTSV---------ELTIRVTDVNDN------APKFELPDYQAHNVDEDIPLGTSILK  784

Query  360  YQAIDEDTNKAASNVKYVMGRN----DGGYLMIDSKTAEIKFVKNMNRDSTFIVNKTITA  415
             +A+D D+  A + ++Y++  +    D   +++++K  +     N N    F+V
Sbjct  785  VKAMDSDSG-ANAEIEYLVSDDHFAVDSNGIIVNNKQLD---ADNNNAYYEFVVTAKDKG  840

Query  416  EVLAIDEYTGKTSTGTVYVRVPDFNDNCP  444
            E         KT T TV V   + ND  P
Sbjct  841  E-------PAKTGTATVRVYTKNKNDEEP  862

 Score = 39.3 bits (90),  Expect = 0.30, Method: Compositional matrix adjust.
 Identities = 54/242 (22%), Positives = 102/242 (42%), Gaps = 22/242 (9%)

Query  4    KFAKPCREGE--DNSKRNP-IAKITSDYQATQKITYRISGVGIDQPPFGIFVVDKNTGDI  60
            +F  P  E E  +N K++  I  + +   A ++I Y +   G      G F +   +G +
Sbjct  656  QFYMPSYEAEIPENQKKDSDIISVKAKSFADREIRYTLKAQGQGA---GTFNIGPTSGIV  712

Query  61   NITAIVDREE--TPSF--LITCRALNAQGLDVEKPLILTVKILDINDNPPVFSQQIFMGE  116
             +   +D E+   P    L+     ++ G       + LT+++ D+NDN P F    +
Sbjct  713  KLAKELDFEDLRQPHVYSLVVTATEDSGGFSTS--VELTIRVTDVNDNAPKFELPDYQAH  770

Query  117  -IEENSASNSLVMILNATDADEPNHLNSKIAFKIVSQEPAGTPMFLLSRNTGEVRTLTNS  175
             ++E+    + ++ + A D+D  +  N++I + +VS +      F +  N   V
Sbjct  771  NVDEDIPLGTSILKVKAMDSD--SGANAEIEY-LVSDD-----HFAVDSNGIIVNNKQLD  822

Query  176  LDREQASSYRLVVSGADKDGEGLSTQCECNIKVKDVNDNFPMFRDSQYSARIEENILSSE  235
             D   A  Y  VV+  DK      +      +  K+ ND  P F    Y+  ++EN   +
Sbjct  823  ADNNNA-YYEFVVTAKDKGEPAKTGTATVRVYTKNKNDEEPKFSQQVYTPNVDENAGPNT  881

Query  236  LL  237
            L+
Sbjct  882  LV  883

>AALB20_033171.P66511 | transcript=AALB20_033171.R66511 | gene=AALB20_033171
| organism=Anopheles_albimanus_STECLA_2020 |
gene_product=neural-cadherin, transcript variant X38 |
transcript_product=neural-cadherin,
transcript variant X37 | location=CM024292:75037560-75208393(+)
| protein_length=3121 | sequence_SO=chromosome
| SO=protein_coding_gene | is_pseudo=false
Length=3121

 Score = 147 bits (370),  Expect = 3e-35, Method: Compositional matrix adjust.
 Identities = 128/446 (29%), Positives = 217/446 (49%), Gaps = 36/446 (8%)
```

```
Query  20    PIAKITS---DYQATQKITYRISGVGID--QPPFGIFVVDKNTGDINITAIVDREET---  71
             PI ++T+   D    Q I Y ++G GID   P    F +++ TG+I +   +DR++
Sbjct  1562  PIMQVTATDGDKDRPQNIVYFLTGQGIDPDNPANSKFDINRTTGEIFVLKPLDRDQPNGR  1621

Query  72    PSFLITCRALNAQGLDVEKPLILTVKILDINDNPPVFSQQIFMGEIEENSASNSLVMILN  131
             P +  T  A +  G  +      + V + DINDN P+F Q ++ G + EN  +  +VM +
Sbjct  1622  PQWRFTVFAQDEGGEGLVGYADVQVNLKDINDNAPIFPQGVYFGNVTENGTAGMVVMTMT  1681

Query  132   ATDADEPNH-LNSKIAFKI---VSQEPAGTPMFLLSRNTGEVRTLTNSLDREQASSYRLV  187
             A D D+PN   N+K+ + I   V +E  G+P+F +   +TG ++T    LDRE+   Y +
Sbjct  1682  AVDYDDPNEGTNAKLIYSIEKNVIEEETGSPIFEIEADTGVIKTAVCCLDRERTPDYSIQ  1741

Query  188   VSGADKDGEGLSTQCECNIKVKDVNDNFPMFRDSQYSARIEENILSS--ELLRFQVTDLD  245
             V      DG GL       +I+VKD+ND  P F    ++    ++E    ++  E+    VT  D
Sbjct  1742  VVAM--DGGGLKGTGTASIRVKDINDMPPQFTKDEWFTEVDETDGTNLPEMPILTVTVHD  1799

Query  246   EEYTDNWLAVYFFTSGNEGNWFEIQTDPRTNEGI--LKVVKALDYE---QLQSVKLSIAV  300
             E+ T+ +         SG    + F +     R N+G   LK+V+ LDYE   Q     +  I V
Sbjct  1800  EDETNKFQYKVIDNSGYGADKFTMV---RNNDGTGSLKIVQPLDYEDPLQSNGFRFRIQV  1856

Query  301   KNKAEFHQSVISRYRVQSTPVTIQVINVREGIAFRPASKTFTVQKGISSKKLVDYILGTY  360
              +K E + +    +Y V   + V +++ ++ +     +P  +    ++  +      V   L T+
Sbjct  1857  NDKGEDNDN--DKYHVAYSWVVVKLRDINDN---KPQFERPNIEVSVYENADVGKTLETF  1911

Query  361   QAIDEDTNKAASNVKYVMGR-NDGGYLMIDSKTAEIKFVKNMNRDSTFIVNKTITAEVLA  419
             +A D D      S V Y + R +D        ++    +    +N++R+    V        ++LA
Sbjct  1912  KATDPDQG-GKSKVSYAIDRSSDRQRQFSINQEGTVTIQRNLDRE----VTPRHQVKILA  1966

Query  420   IDE-YTGKTSTGTVYYVRVPDFNDNCP  444
             ID+    KT+T T+ V V D NDN P
Sbjct  1967  IDDGIPPKTATATLTVIVQDINDNPP  1992

 Score = 80.1 bits (196),  Expect = 8e-14, Method: Compositional matrix adjust.
 Identities = 107/424 (25%), Positives = 178/424 (42%), Gaps = 51/424 (12%)

Query  53    VDKNTGDINITAIVDREETPSFLITCRALNAQGLDVEKPLILTVKILDINDNPPVFSQ-Q  111
             +D++  ++N+TA+ D          +C  +N           ++ V I D+NDN PVF
Sbjct  950   LDRDKYELNVTAMDDG--------SC-CVNGDATIHTSTAVVVVFITDVNDNKPVFKDCG  1000

Query  112   IFMGEIEENSASNSLVMILNATDADEPNHLNSKIAFKIVSQEPAGTPMFLLSRNTGEVRT  171
              +  ++EE + + S V+ + ATD D+    +N ++ + IV Q        F +   TGEV T
Sbjct  1001  TYYPKVEEGAPNGSPVIKVQATDEDK--GVNGQVKYSIVQQPNQKGTKFTVDEETGEVST  1058

Query  172   LTNSLDRE--QASSYRLVVSGADKDGEGLSTQCECNIKVKDVNDNFPMFRDSQYSARIEE  229
                  DRE         + V   D+     L   C    +++ DVNDN P+F    +Y   +++
Sbjct  1059  -NKVFDREGDDGKFVSVTVKATDQGEPSLEGVCSFTVEITDVNDNPPLFDRQKYVENVKQ  1117

Query  230   NI-LSSELLRFQVTDLDEEYTDNWLAVYFFT---SGNEGNWFEIQTDPRTNEGILKVVKA  285
             +  + + +LR  V+  DE+  +N   VY  +   S N+  +FEIQ  P +   +LK  K
Sbjct  1118  DASIGTNILR--VSASDEDADNNGAIVYTLSAPHSPNDLEYFEIQ--PESGWIVLK--KP  1171

Query  286   LDYEQLQSVKLSIAVKNK------AEFHQSVISRYRVQSTPV----TIQVINVREGIAFR  335
             LD E+     +L +   ++       A+     +     R     P+          I++RE +
Sbjct  1172  LDRERY---RLKVYASDRGDPPSTADVDVELDVVDRNNKPPIWDRNIYGPIHIRENVTVG  1228

Query  336   PASKTFTVQKGISSKKLVDYILGTYQAIDEDTNKAASNVKYVMGRNDGGYLMIDSKTAEI  395
                       GI      V Y L          TNK    +  Y+  R D G          A+I
Sbjct  1229  TVVTAIKASSGIEGNPTVFYRL--MPGSTAQTNKL--HTFYLQQRPDNG-----DTWADI  1279

Query  396   KFVKNMNRDSTFIVNKTITAEVLAIDEYTGKTSTGTVYYVRVPDFNDNCPTAV-LEKDAVC  454
             K     ++ ++    N TI  E      +      S  TVY+++ D ND  P      E++ V
Sbjct  1280  KVNHPLDFETIKEYNLTIRVENNGAQQL---ASEATVYIQLEDVNDEIPLFTEREQETVL  1336

Query  455   SSSP  458
                P
Sbjct  1337  EGEP  1340

 Score = 79.0 bits (193),  Expect = 2e-13, Method: Compositional matrix adjust.
 Identities = 104/419 (25%), Positives = 173/419 (41%), Gaps = 51/419 (12%)
```

```
Query  50    IFVVDKNTGDINITAIVDREETPSFLITCRAL----------NAQGLDVEKPLILTVKIL  99
              F ++  +G+I    + DRE+  ++ +   A           N Q   V K   + + I
Sbjct  1377  FFEINLQSGEIFTKVVFDREKQGAYALEVEARDGAPSARPNSNQQPNSVTK--FIRIGIA  1434

Query  100   DINDNPPVFSQQIFMGEIEENSASNSLVMILNATDADEPNHLNSKIAFKIVSQEPAGTPM  159
             D NDNPP F + ++  E++EN       V+ + A D DE    +S+I ++I S    G
Sbjct  1435  DKNDNPPYFDKALYEAEVDENEDIQHTVLTVTAKDHDE----SSRIRYEITSGNIGGA--  1488

Query  160   FLLSRNTGEVRTLTNSLDREQASSYRLVVSGADKDGEGLSTQCECNIKVKDVNDNFPMFR  219
             F +   TG +  +  +LD E    Y L +  +D   E  +       I VKDVND  P F
Sbjct  1489  FAVKNMTGAI-YVAGALDYETRKRYHLTLVASDSLNENATI---IEIHVKDVNDMPPTFS  1544

Query  220   DSQYSARI--EENILSSELLRFQVTDLDEEYTDNWLAVYFFTSG--NEGNWFEIQTDPRT  275
                Y A    E+ +    +++   TD D++   N   VYF T    +  N    + D
Sbjct  1545  QQIYEATQDEEQTVTDRPIMQVTATDGDKDRPQN--IVYFLTGQGIDPDNPANSKFDINR  1602

Query  276   NEGILKVVKALDYEQLQS-VKLSIAVKNKAEFHQSVISRYRVQSTPVTIQVINVREGIAF  334
               G + V+K LD +Q     +    V  + E  + ++    VQ     + + ++ +
Sbjct  1603  TTGEIFVLKPLDRDQPNGRPQWRFTVFAQDEGGEGLVGYADVQ-----VNLKDINDNAPI  1657

Query  335   RPASKTFTVQKGISSKKLVDYILGTYQAIDEDTNKAASNVK--YVMGRN------DGGYL  386
              P    F     ++        ++ T  A+D D     +N K  Y + +N
Sbjct  1658  FPQGVYF---GNVTENGTAGMVVMTMTAVDYDDPNEGTNAKLIYSIEKNVIEEETGSPIF  1714

Query  387   MIDSKTAEIK-FVKNMNRDSTFIVNKTITAEVLAIDEYTGKTSTGTVYVRVPDFNDNCP  444
              I++ T  IK  V  ++R+ T        + +V+A+D   G   TGT  +RV D ND  P
Sbjct  1715  EIEADTGVIKTAVCCLDRERT----PDYSIQVVAMDG-GGLKGTGTASIRVKDINDMPP  1768

 Score = 68.2 bits (165),  Expect = 4e-10, Method: Compositional matrix adjust.
 Identities = 88/361 (24%), Positives = 157/361 (43%), Gaps = 36/361 (10%)

Query  51    FVVDKNTGDINITAIVDRE-ETPSFL-ITCRALNAQGLDVEKPLILTVKILDINDNPPVF  108
             F VD+ TG+++    + DRE +    F+ +T +A +     +E      TV+I D+NDNPP+F
Sbjct  1047  FTVDEETGEVSTNKVFDREGDDGKFVSVTVKATDQGEPSLEGVCSFTVEITDVNDNPPLF  1106

Query  109   SQQIFMGEIEENSASNSLVMILNATDADEPNHLNSKIAFKIVS-QEPAGTPMFLLSRNTG  167
              +Q ++  ++++++  + ++ ++A+D D  N  N  I + + +   P     F +   +G
Sbjct  1107  DRQKYVENVKQDASIGTNILRVSASDEDADN--NGAIVYTLSAPHSPNDLEYFEIQPESG  1164

Query  168   EVRTLTNSLDREQASSYRLVVSGADKDGEGLSTQCECNIKVKDVNDNFPMFRDSQYSA--  225
              +  L   LDRE+   YRL V  +D+ G+  ST             +N P   D
Sbjct  1165  WI-VLKKPLDRER---YRLKVYASDR-GDPPSTADVDVELDVVDRNNKPPIWDRNIYGPI  1219

Query  226   RIEENILSSELLRFQVTDLDEEYTDNWLAVYFFTSG-----NEGNWFEIQTDPRTNE--G  278
              I EN+    ++              N    Y    G     N+ + F +Q  P   +
Sbjct  1220  HIRENVTVGTVV--TAIKASSGIEGNPTVFYRLMPGSTAQTNKLHTFYLQQRPDNGDTWA  1277

Query  279   ILKVVKALDYEQLQSVKLSIAVKNKAEFHQSVISRYRVQSTPVTIQVINVREGIAFRPAS  338
              +KV   LD+E ++   L+I V+N          ++        V IQ+ +V + I
Sbjct  1278  DIKVNHPLDFETIKEYNLTIRVENNG-------AQQLASEATVYIQLEDVNDEIPLFTER  1330

Query  339   KTFTVQKGISSKKLVDYILGTYQAIDEDTNKAASNVKYVM---GRNDG-GYLMIDSKTAE  394
             +  TV +G       V  +     AID+D       + V Y +    RN+G  +  I+ ++ E
Sbjct  1331  EQETVLEGEPIGTKVTQV----NAIDKDGTFPNNQVYYYIVDSPRNEGKDFFEINLQSGE  1386

Query  395   I  395
             I
Sbjct  1387  I  1387

 Score = 55.8 bits (133),  Expect = 3e-06, Method: Compositional matrix adjust.
 Identities = 49/158 (31%), Positives = 78/158 (49%), Gaps = 24/158 (15%)

Query  102   NDNPPVFSQQIFMGEIEENSASNSLVMILNATDADEPNHLNSKIAFKIVSQEPAGTP--M  159
             ND  P FSQQ++   ++EN+  N+LV  + A+D D  N     + F  V     GT
Sbjct  880   NDEEPKFSQQVYTPNVDENAGPNTLVTTVVASDKDGDN-----VRFGFVG---GGTSSGQ  931

Query  160   FLLSRNTGEVRTLTN--SLDREQASSYRLVVSGADK-------DGEGLSTQCECNIKVKD  210
             F++   TG +R  +   SLDR++   Y L V+  D         D    ++     + + D
```

```
Sbjct  932   FVIEEITGVIRLHSKAISLDRDK---YELNVTAMDDGSCCVNGDATIHTSTAVVVVFITD  988

Query  211   VNDNFPMFRD-SQYSARIEENILS-SELLRFQVTDLDE  246
             VNDN P+F+D   Y  ++EE   + S +++ Q TD D+
Sbjct  989   VNDNKPVFKDCGTYYPKVEEGAPNGSPVIKVQATDEDK  1026

 Score = 53.5 bits (127),  Expect = 1e-05, Method: Compositional matrix adjust.
 Identities = 70/306 (23%), Positives = 126/306 (41%), Gaps = 30/306 (10%)

Query  20    PIAKIT-SDYQATQKITYRI---SGVGIDQPPFGIFVVDKNTGDINITAIVDRE---ETP  72
             PI  +T  D   T K  Y++   SG G D+  F +   +  TG + I   +D E   ++
Sbjct  1791  PILTVTVHDEDETNKFQYKVIDNSGYGADK--FTMVRNNDGTGSLKIVQPLDYEDPLQSN  1848

Query  73    SFLITCRALNAQGLDVEKPLI------LTVKILDINDNPPVFSQQIFMGEIEENSASNSL  126
              F    + +N +G D +          + VK+ DINDN P F +      + EN+
Sbjct  1849  GFRFRIQ-VNDKGEDNDNDKYHVAYSWVVVKLRDINDNKPQFERPNIEVSVYENADVGKT  1907

Query  127   VMILNATDADEPNHLNSKIAFKIVSQEPAGTPMFLLSRNTGEVRTLTNSLDREQASSYRL  186
             +    ATD D+     SK+++ I   + +       S N     T+  +LDRE    +++
Sbjct  1908  LETFKATDPDQGG--KSKVSYAI---DRSSDRQRQFSINQEGTVTIQRNLDREVTPRHQV  1962

Query  187   VVSGADKDGEGLSTQCECNIKVKDVNDNFPMFRDSQYSARIEENILSSELLRFQVTDLDE  246
              +   D      +      + V+D+NDN P F    Y   + E++   +++    TD D+
Sbjct  1963  KILAIDDGIPPKTATATLTVIVQDINDNPPKFL-KDYRPVLPEHVPPRKVVEILATDDDD  2021

Query  247   EYTDNWLAVYFFTSGNEGNW----FEIQTDPRTNEG----ILKVVKALDYEQLQSVKLSI  298
                 N     F       +     F+++ D +   G    I+  +++ D EQ +   + I
Sbjct  2022  RSKSNGPPFQFRLDPGADDIIRASFKVEQDQKGANGDGMAIVSSLRSFDREQQKEYLIPI  2081

Query  299   AVKNKA  304
              +K+
Sbjct  2082  VIKDHG  2087

 Score = 51.2 bits (121),  Expect = 7e-05, Method: Compositional matrix adjust.
 Identities = 96/398 (24%), Positives = 163/398 (41%), Gaps = 54/398 (14%)

Query  106   PVFSQQIFMGEIEENSASNSLVMILNATDADEPNHLNSKIAFKIVSQ-EPAGTPMFLLSR  164
             P F    +  EI EN   +S ++ + A      +  + +I + + +Q + AGT  F +
Sbjct  677   PQFYMPSYEAEIPENQKKDSDIISVKAK-----SFADREIRYTLKAQGQGAGT--FNIGP  729

Query  165   NTGEVRTLTNSLDRE---QASSYRLVVSGADKDGEGLSTQCECNIKVKDVNDNFPMFRDS  221
              +G V+ L   LD E   Q   Y LVV+ A +D  G ST  E  I+V DVNDN P F
Sbjct  730   TSGIVK-LAKELDFEDLRQPHVYSLVVT-ATEDSGGFSTSVELTIRVTDVNDNAPKFELP  787

Query  222   QYSAR-IEENI-LSSELLRFQVTDLDEEYTDNWLAVYFFTSGNEGNWFEIQTDPRTNEGI  279
              Y A  ++E+I + + +L+ +  D D         + +  S +    F + ++     GI
Sbjct  788   DYQAHNVDEDIQIGTSILKVKAMDADSGAN---AEIEYLVSDDH---FAVDSN-----GI  836

Query  280   LKVVKALDYEQLQS-VKLSIAVKNKAEFHQSVISRYRVQSTPVTIQVINVREGIAFRPAS  338
             +   K LD +   +  +  +  K+K E  ++  +  RV +           E   F
Sbjct  837   IVNNKQLDADNNNAYYEFVVTAKDKGEPAKTGTATVRVYTKNKN------DEEPKFSQQV  890

Query  339   KTFTVQKGISSKKLVDYILGTYQAIDEDTNKAASNVK--YVMGRNDGGYLMIDSKTAEIK  396
              T  V +      LV  ++         ++K   NV+  +V G    G  +I+  T  I+
Sbjct  891   YTPNVDENAGPNTLVTTVVA--------SDKDGDNVRFGFVGGGTSSGQFVIEEITGVIR  942

Query  397   FVK---NMNRDSTFI-VNKTITAEVLAIDEYTGKTSTGTVYVRVPDFNDN------CPTA  446
                   +++RD   + V            + T  TST  V V + D NDN      C T
Sbjct  943   LHSKAISLDRDKYELNVTAMDDGSCCVNGDATIHTSTAVVVVFITDVNDNKPVFKDCGTY  1002

Query  447   VLE-KDAVCSSSPSVVVSARTLNNRYTGPYTFALEDQP  483
               + ++   + SP + V A   +    G   +++  QP
Sbjct  1003  YPKVEEGAPNGSPVIKVQATDEDKGVNGQVKYSIVQQP  1040

 Score = 44.3 bits (103),  Expect = 0.009, Method: Compositional matrix adjust.
 Identities = 48/199 (24%), Positives = 73/199 (37%), Gaps = 19/199 (10%)

Query  27    DYQATQKITYRISGVGIDQPPFGIFVVDKNTGDINITAIVDREETPSFLITCRALNAQGL  86
```

```
             D     K++Y I      Q  F I       G + I   +DRE TP   +   A++
Sbjct  1917  DQGGKSKVSYAIDRSSDRQRQFSI----NQEGTVTIQRNLDREVTPRHQVKILAIDDGIP  1972

Query  87    DVEKPLILTVKILDINDNPPVFSQQIFMGEIEENSASNSLVMILNATDADEPNHLNSKIA  146
                    LTV + DINDNPP F +  +   + E+     +V IL   D D
Sbjct  1973  PKTATATLTVIVQDINDNPPKFLKD-YRPVLPEHVPPRKVVEILATDDDDRSKSNGPPFQ  2031

Query  147   FKIVSQEPAGTPMFLLSRNTGEVRTLTN-----------SLDREQASSYRLVVSGADKDG  195
             F++   +P    +   S    + +   N           S DREQ   Y + +   D
Sbjct  2032  FRL---DPGADDIIRASFKVEQDQKGANGDGMAIVSSLRSFDREQQKEYLIPIVIKDHGN  2088

Query  196   EGLSTQCECNIKVKDVNDN  214
               ++      + + DVNDN
Sbjct  2089  PAMTGTSTLTVVIGDVNDN  2107

 Score = 40.4 bits (93),  Expect = 0.13, Method: Compositional matrix adjust.
 Identities = 54/235 (23%), Positives = 99/235 (42%), Gaps = 22/235 (9%)

Query  4    KFAKPCREGE--DNSKRNP-IAKITSDYQATQKITYRISGVGIDQPPFGIFVVDKNTGDI  60
            +F  P  E E  +N K++  I  + +   A ++I Y +   G      G F +   +G +
Sbjct  678  QFYMPSYEAEIPENQKKDSDIISVKAKSFADREIRYTLKAQGQGA---GTFNIGPTSGIV  734

Query  61   NITAIVDREE--TPSF--LITCRALNAQGLDVEKPLILTVKILDINDNPPVFSQQIFMGE  116
             +   +D E+   P    L+     ++ G      + LT+++ D+NDN P F    +
Sbjct  735  KLAKELDFEDLRQPHVYSLVVTATEDSGGFSTS--VELTIRVTDVNDNAPKFELPDYQAH  792

Query  117  -IEENSASNSLVMILNATDADEPNHLNSKIAFKIVSQEPAGTPMFLLSRNTGEVRTLTNS  175
             ++E+    + ++ + A DAD  +  N++I + +VS +      F +  N   V
Sbjct  793  NVDEDIQIGTSILKVKAMDAD--SGANAEIEY-LVSDD-----HFAVDSNGIIVNNKQLD  844

Query  176  LDREQASSYRLVVSGADKDGEGLSTQCECNIKVKDVNDNFPMFRDSQYSARIEEN  230
             D   A  Y  VV+  DK     +      +  K+ ND  P F    Y+  ++EN
Sbjct  845  ADNNNA-YYEFVVTAKDKGEPAKTGTATVRVYTKNKNDEEPKFSQQVYTPNVDEN  898

 Score = 39.3 bits (90),  Expect = 0.35, Method: Compositional matrix adjust.
 Identities = 55/225 (24%), Positives = 88/225 (39%), Gaps = 24/225 (11%)

Query  75   LITCRALNAQGLDVEKPLILTVKILDINDNPPVFSQQIF--MGEIEENSASNSLVMILNA  132
            +IT +  NA GL       + + + D+ND PP F  +       ++ N+  N+ V  L A
Sbjct  539  IITNQGHNA-GLKYTDNQRVIIHVKDVNDEPPYFINRPLPMQAVVQLNAPPNTPVFTLQA  597

Query  133  TDADEPNHLNSKIAFKIVSQEPAGTPMFLLSRNTGEVRTLTNSLDREQASSYRLVVSGAD  192
             D D     +  I + IV     G   F +   +G VRT    L  +    Y L V   D
Sbjct  598  RDPD----TDHNIHYFIVRDRTGG--RFEVDERSGVVRTRGTDL-FQLDMEYVLYVKAED  650

Query  193  KDGE----GLSTQCECNIKVKDVNDNFPMFRDSQYSARIEEN-ILSSELLRFQVTDLDEE  247
            ++G+       +  E  + +       P F    Y A I EN    S+++  +     +
Sbjct  651  QNGKVDDRRFQSTPEERLSIVG-GKRAPQFYMPSYEAEIPENQKKDSDIISVKA----KS  705

Query  248  YTDNWLAVYFFTSGNEGNWFEIQTDPRTNEGILKVVKALDYEQLQ  292
            + D  +       G     F I        GI+K+ K LD+E L+
Sbjct  706  FADREIRYTLKAQGQGAGTFNIGP----TSGIVKLAKELDFEDLR  746

>AARA21_009090.P17045 | transcript=AARA21_009090.R17045 | gene=AARA21_009090
| organism=Anopheles_arabiensis_DONGOLA_2021
| gene_product=neural-cadherin, transcript variant X5 |
transcript_product=neural-cadherin,
transcript variant X13 | location=CM029353:40466514-40666848(+)
| protein_length=3099 |
sequence_SO=chromosome | SO=protein_coding_gene | is_pseudo=false
Length=3099

 Score = 147 bits (370),  Expect = 3e-35, Method: Compositional matrix adjust.
 Identities = 128/446 (29%), Positives = 217/446 (49%), Gaps = 36/446 (8%)

Query  20    PIAKITS---DYQATQKITYRISGVGID--QPPFGIFVVDKNTGDINITAIVDREET---  71
             PI ++T+   D    Q I Y ++G GID   P    F +++ TG+I +   +DR++
Sbjct  1539  PIMQVTATDGDKDRPQNIVYFLTGQGIDPDNPANSKFDINRTTGEIFVLKPLDRDQPNGR  1598
```

```
Query  72    PSFLITCRALNAQGLDVEKPLILTVKILDINDNPPVFSQQIFMGEIEENSASNSLVMILN  131
             P +  T  A +  G  +     + V + DINDN P+F Q ++ G + EN  +  +VM +
Sbjct  1599  PQWRFTVFAQDEGGEGLVGYADVQVNLKDINDNAPIFPQGVYFGNVTENGTAGMVVMTMT  1658

Query  132   ATDADEPNH-LNSKIAFKI---VSQEPAGTPMFLLSRNTGEVRTLTNSLDREQASSYRLV  187
             A D D+PN   N+K+ + I   V +E  G+P+F +  +TG ++T    LDRE+   Y +
Sbjct  1659  AVDYDDPNEGTNAKLIYSIEKNVIEEETGSPIFEIEADTGVIKTAVCCLDRERTPDYSIQ  1718

Query  188   VSGADKDGEGLSTQCECNIKVKDVNDNFPMFRDSQYSARIEENILSS--ELLRFQVTDLD  245
             V     DG GL       +I+VKD+ND  P F   ++   ++E   ++  E+    VT  D
Sbjct  1719  VVAM--DGGGLKGTGTASIRVKDINDMPPQFTKDEWFTEVDETDGTNLPEMPILTVTVHD  1776

Query  246   EEYTDNWLAVYFFTSGNEGNWFEIQTDPRTNEGI--LKVVKALDYE---QLQSVKLSIAV  300
             E+ T+ +        SG    + F +    R N+G   LK+V+ LDYE   Q    +  I V
Sbjct  1777  EDETNKFQYKVIDNSGYGADKFTMV---RNNDGTGSLKIVQPLDYEDPLQSNGFRFRIQV  1833

Query  301   KNKAEFHQSVISRYRVQSTPVTIQVINVREGIAFRPASKTFTVQKGISSKKLVDYILGTY  360
              +K E + +   +Y V  + V +++ ++ +    +P  +   ++  +      V   L T+
Sbjct  1834  NDKGEDNDN--DKYHVAYSWVVVKLRDINDN---KPQFERPNIEVSVYENADVGKTLETF  1888

Query  361   QAIDEDTNKAASNVKYVMGR-NDGGYLMIDSKTAEIKFVKNMNRDSTFIVNKTITAEVLA  419
             +A D D     S V Y + R +D        ++   +   +N++R+    V       ++LA
Sbjct  1889  KATDPDQG-GKSKVSYAIDRSSDRQRQFSINQEGTVTIQRNLDRE----VTPRHQVKILA  1943

Query  420   IDE-YTGKTSTGTVYVRVPDFNDNCP  444
             ID+    KT+T T+ V V D NDN P
Sbjct  1944  IDDGIPPKTATATLTVIVQDINDNPP  1969

 Score = 75.9 bits (185),  Expect = 2e-12, Method: Compositional matrix adjust.
 Identities = 89/361 (25%), Positives = 167/361 (46%), Gaps = 36/361 (10%)

Query  51    FVVDKNTGDINITAIVDRE-ETPSFL-ITCRALNAQGLDVEKPLILTVKILDINDNPPVF  108
             F VD+ TG+++    + DRE +   F+ +T +A +      +E      TV+I D+NDNPP+F
Sbjct  1025  FTVDEETGEVSTNKVFDREGDDGKFVSVTVKATDQGEPSLEGVCSFTVEITDVNDNPPLF  1084

Query  109   SQQIFMGEIEENSASNSLVMILNATDADEPNHLNSKIAFKIVS-QEPAGTPMFLLSRNTG  167
              +Q ++  ++++++  + ++ ++A+D D  N  N  I + + +   P     F +   +G
Sbjct  1085  DRQKYVENVKQDASIGTNILRVSASDEDADN--NGAIVYTLSAPHSPNDLEYFEIQPESG  1142

Query  168   EVRTLTNSLDREQASSYRLVVSGADKDGEGLSTQCECNIKVKDVNDNFPMFRDSQYSA-R  226
              +  L   LDRE   +Y+L      DK    LS   E  I V D  +N P++ ++ Y
Sbjct  1143  WI-VLKKPLDRE---TYKLEAMAQDKGYPPLSRTVEVQIDVVDRANNPPVWDNTVYGPIY  1198

Query  227   IEENI-LSSELLRFQVTDLDEEYTDNWLAVYFFTSG-----NEGNWFEIQTDPRTNE--G  278
             ++EN+ +  +++  + +    E    N    Y    G     N+ + F +Q  P   +
Sbjct  1199  VKENMPVGGKVVSIKASSGIE---GNPTVFYRLMPGSTAQTNKLHTFYLQQRPDNGDTWA  1255

Query  279   ILKVVKALDYEQLQSVKLSIAVKNKAEFHQSVISRYRVQSTPVTIQVINVREGIAFRPAS  338
              +KV   LD+E ++   L++ V+N          ++        V IQ+ +V + I
Sbjct  1256  DIKVNHPLDFETIKEYNLTVRVENNG-------AQQLASEATVYIQLEDVNDEIPLFTER  1308

Query  339   KTFTVQKGISSKKLVDYILGTYQAIDEDTNKAASNVKYVM---GRNDG-GYLMIDSKTAE  394
             +  TV +G       V  +      AID+D     + V Y +    RN+G  +  I+ ++ E
Sbjct  1309  EQETVLEGEPIGTKVTQV----NAIDKDGTFPNNQVYYYIVDSPRNEGKDFFEINLQSGE  1364

Query  395   I  395
             I
Sbjct  1365  I  1365

 Score = 75.5 bits (184),  Expect = 2e-12, Method: Compositional matrix adjust.
 Identities = 103/418 (25%), Positives = 174/418 (42%), Gaps = 50/418 (12%)

Query  50    IFVVDKNTGDINITAIVDREETPSFLITCRAL----------NAQGLDVEKPLILTVKIL  99
              F ++  +G+I    + DRE+  ++ +   A           N Q   V K   + + I
Sbjct  1355  FFEINLQSGEIFTKVVFDREKQGAYALEVEARDGAPSARPNSNQQPNSVTK--FIRIGIA  1412

Query  100   DINDNPPVFSQQIFMGEIEENSASNSLVMILNATDADEPNHLNSKIAFKIVSQEPAGTPM  159
             D NDNPP F + ++  E++EN       V+ + A D DE    +S+I ++I S    G
```

```
Sbjct  1413  DKNDNPPYFDKALYEAEVDENEDIQHTVLTVTAKDHDE----SSRIRYEITSGNIGGA--  1466

Query  160   FLLSRNTGEVRTLTNSLDREQASSYRLVVSGADKDGEGLSTQCECNIKVKDVNDNFPMFR  219
             F +   TG +  +  +LD E    Y L +  +D   E  +      I VKDVND  P F
Sbjct  1467  FAVKNMTGAI-YVAGALDYETRKRYYLTLVASDSLNENETIIV---INVKDVNDMPPSFP  1522

Query  220   DSQYSARIEENI-LSSELLRFQVTDLDEEYTDNWLAVYFFTSG--NEGNWFEIQTDPRTN  276
                Y   ++E + +   +++   TD D++   N   VYF T    +  N    + D
Sbjct  1523  QQVYERTMDEELAVPFPIMQVTATDGDKDRPQN--IVYFLTGQGIDPDNPANSKFDINRT  1580

Query  277   EGILKVVKALDYEQLQS-VKLSIAVKNKAEFHQSVISRYRVQSTPVTIQVINVREGIAFR  335
              G + V+K LD +Q     +    V  + E  + ++    VQ     + + ++ +
Sbjct  1581  TGEIFVLKPLDRDQPNGRPQWRFTVFAQDEGGEGLVGYADVQ-----VNLKDINDNAPIF  1635

Query  336   PASKTFTVQKGISSKKLVDYILGTYQAIDEDTNKAASNVK--YVMGRN------DGGYLM  387
             P    F     ++        ++ T  A+D D     +N K  Y + +N
Sbjct  1636  PQGVYF---GNVTENGTAGMVVMTMTAVDYDDPNEGTNAKLIYSIEKNVIEEETGSPIFE  1692

Query  388   IDSKTAEIK-FVKNMNRDSTFIVNKTITAEVLAIDEYTGKTSTGTVYVRVPDFNDNCP  444
             I++ T  IK  V  ++R+ T        + +V+A+D   G   TGT  +RV D ND  P
Sbjct  1693  IEADTGVIKTAVCCLDRERT----PDYSIQVVAMDG-GGLKGTGTASIRVKDINDMPP  1745

 Score = 55.8 bits (133),  Expect = 3e-06, Method: Compositional matrix adjust.
 Identities = 49/158 (31%), Positives = 78/158 (49%), Gaps = 24/158 (15%)

Query  102   NDNPPVFSQQIFMGEIEENSASNSLVMILNATDADEPNHLNSKIAFKIVSQEPAGTP--M  159
             ND  P FSQQ++   ++EN+  N+LV  + A+D D  N     + F  V     GT
Sbjct  858   NDEEPKFSQQVYTPNVDENAGPNTLVTTVVASDKDGDN-----VRFGFVG---GGTSSGQ  909

Query  160   FLLSRNTGEVRTLTN--SLDREQASSYRLVVSGADK-------DGEGLSTQCECNIKVKD  210
             F++   TG +R  +   SLDR++   Y L V+  D         D    ++     + + D
Sbjct  910   FVIEEITGVIRLHSKAISLDRDK---YELNVTAMDDGSCCVNGDATIHTSTAVVVVFITD  966

Query  211   VNDNFPMFRD-SQYSARIEENILS-SELLRFQVTDLDE  246
             VNDN P+F+D   Y  ++EE   + S +++ Q TD D+
Sbjct  967   VNDNKPVFKDCGTYYPKVEEGAPNGSPVIKVQATDEDK  1004

 Score = 53.5 bits (127),  Expect = 1e-05, Method: Compositional matrix adjust.
 Identities = 70/306 (23%), Positives = 126/306 (41%), Gaps = 30/306 (10%)

Query  20    PIAKIT-SDYQATQKITYRI---SGVGIDQPPFGIFVVDKNTGDINITAIVDRE---ETP  72
             PI  +T  D   T K  Y++   SG G D+  F +    +  TG + I   +D E   ++
Sbjct  1768  PILTVTVHDEDETNKFQYKVIDNSGYGADK--FTMVRNNDGTGSLKIVQPLDYEDPLQSN  1825

Query  73    SFLITCRALNAQGLDVEKPLI------LTVKILDINDNPPVFSQQIFMGEIEENSASNSL  126
              F    + +N +G D +          + VK+ DINDN P F +      + EN+
Sbjct  1826  GFRFRIQ-VNDKGEDNDNDKYHVAYSWVVVKLRDINDNKPQFERPNIEVSVYENADVGKT  1884

Query  127   VMILNATDADEPNHLNSKIAFKIVSQEPAGTPMFLLSRNTGEVRTLTNSLDREQASSYRL  186
             +    ATD D+     SK+++ I   + +        S N     T+  +LDRE    +++
Sbjct  1885  LETFKATDPDQGG--KSKVSYAI---DRSSDRQRQFSINQEGTVTIQRNLDREVTPRHQV  1939

Query  187   VVSGADKDGEGLSTQCECNIKVKDVNDNFPMFRDSQYSARIEENILSSELLRFQVTDLDE  246
              +   D      +       + V+D+NDN P F    Y   + E++   +++    TD D+
Sbjct  1940  KILAIDDGIPPKTATATLTVIVQDINDNPPKFL-KDYRPVLPEHVPPRKVVEILATDDDD  1998

Query  247   EYTDNWLAVYFFTSGNEGNW----FEIQTDPRTNEG----ILKVVKALDYEQLQSVKLSI  298
                 N     F       +     F+++ D +   G    I+  +++ D EQ +   + I
Sbjct  1999  RSKSNGPPFFQFRLDPGADDIIRASFKVEQDQKGANGDGMAIVSSLRSFDREQQKEYLIPI  2058

Query  299   AVKNKA  304
              +K+
Sbjct  2059  VIKDHG  2064

 Score = 52.0 bits (123),  Expect = 5e-05, Method: Compositional matrix adjust.
 Identities = 97/398 (24%), Positives = 163/398 (41%), Gaps = 54/398 (14%)

Query  106   PVFSQQIFMGEIEENSASNSLVMILNATDADEPNHLNSKIAFKIVSQ-EPAGTPMFLLSR  164
```

```
                P F    +  EI EN   +S ++ + A      +  + +I + + +Q + AGT  F +
Sbjct  655   PQFYMPSYEAEIPENQKKDSDIISVKAK-----SFADREIRYTLKAQGQGAGT--FNIGP  707

Query  165   NTGEVRTLTNSLDRE---QASSYRLVVSGADKDGEGLSTQCECNIKVKDVNDNFPMFRDS  221
              +G V+ L   LD E   Q   Y LVV+ A +D  G ST  E  I+V DVNDN P F
Sbjct  708   TSGIVK-LAKELDFEDLRQPHVYSLVVT-ATEDSGGFSTSVELTIRVTDVNDNAPKFELP  765

Query  222   QYSAR-IEENI-LSSELLRFQVTDLDEEYTDNWLAVYFFTSGNEGNWFEIQTDPRTNEGI  279
              Y A  ++E+I L + +L+ +  D D         + +  S +    F + ++     GI
Sbjct  766   DYQAHNVDEDIPLGTSILKVKAMDSDSGAN---AEIEYLVSDDH---FAVDSN-----GI  814

Query  280   LKVVKALDYEQLQS-VKLSIAVKNKAEFHQSVISRYRVQSTPVTIQVINVREGIAFRPAS  338
             +   K LD +   +  +  +  K+K E  ++  +  RV +             E  F
Sbjct  815   IVNNKQLDADNNNAYYEFVVTAKDKGEPAKTGTATVRVYTKNKN------DEEPKFSQQV  868

Query  339   KTFTVQKGISSKKLVDYILGTYQAIDEDTNKAASNVK--YVMGRNDGGYLMIDSKTAEIK  396
              T  V +      LV  ++          ++K   NV+  +V G    G  +I+  T  I+
Sbjct  869   YTPNVDENAGPNTLVTTVVA--------SDKDGDNVRFGFVGGGTSSGQFVIEEITGVIR  920

Query  397   FVK---NMNRDSTFI-VNKTITAEVLAIDEYTGKTSTGTVYVRVPDFNDN------CPTA  446
                  +++RD   + V              + T  TST  V V + D NDN      C T
Sbjct  921   LHSKAISLDRDKYELNVTAMDDGSCCVNGDATIHTSTAVVVVFITDVNDNKPVFKDCGTY  980

Query  447   VLE-KDAVCSSSPSVVVSARTLNNRYTGPYTFALEDQP  483
               + ++   + SP + V A   +    G   +++  QP
Sbjct  981   YPKVEEGAPNGSPVIKVQATDEDKGVNGQVKYSIVQQP  1018

 Score = 44.3 bits (103),  Expect = 0.009, Method: Compositional matrix adjust.
 Identities = 48/199 (24%), Positives = 73/199 (37%), Gaps = 19/199 (10%)

Query  27    DYQATQKITYRISGVGIDQPPFGIFVVDKNTGDINITAIVDREETPSFLITCRALNAQGL  86
             D     K++Y I      Q  F I       G + I   +DRE TP   +   A++
Sbjct  1894  DQGGKSKVSYAIDRSSDRQRQFSI----NQEGTVTIQRNLDREVTPRHQVKILAIDDGIP  1949

Query  87    DVEKPLILTVKILDINDNPPVFSQQIFMGEIEENSASNSLVMILNATDADEPNHLNSKIA  146
                    LTV + DINDNPP F +  +   + E+     +V IL   D D
Sbjct  1950  PKTATATLTVIVQDINDNPPKFLKD-YRPVLPEHVPPRKVVEILATDDDDRSKSNGPPFQ  2008

Query  147   FKIVSQEPAGTPMFLLSRNTGEVRTLTN-----------SLDREQASSYRLVVSGADKDG  195
             F++   +P    +   S    + +   N           S DREQ   Y + +   D
Sbjct  2009  FRL---DPGADDIIRASFKVEQDQKGANGDGMAIVSSLRSFDREQQKEYLIPIVIKDHGN  2065

Query  196   EGLSTQCECNIKVKDVNDN  214
               ++      + + DVNDN
Sbjct  2066  PAMTGTSTLTVVIGDVNDN  2084

 Score = 39.7 bits (91),  Expect = 0.29, Method: Compositional matrix adjust.
 Identities = 89/389 (23%), Positives = 149/389 (38%), Gaps = 62/389 (16%)

Query  75   LITCRALNAQGLDVEKPLILTVKILDINDNPPVFSQQIF--MGEIEENSASNSLVMILNA  132
            +IT +  NA GL       + + + D+ND PP F  +       ++ N+  N+ V  L A
Sbjct  517  IITNQGHNA-GLKYTDNQRVIIHVKDVNDEPPYFINRPLPMQAVVQLNAPPNTPVFTLQA  575

Query  133  TDADEPNHLNSKIAFKIVSQEPAGTPMFLLSRNTGEVRTLTNSLDREQASSYRLVVSGAD  192
             D D     +  I + IV     G  F +   +G VRT    L  +     Y L V   D
Sbjct  576  RDPD----TDHNIHYFIVRDRTGG--RFEVDERSGVVRTRGTDL-FQLDMEYVLYVKAED  628

Query  193  KDGE----GLSTQCECNIKVKDVNDNFPMFRDSQYSARIEEN-ILSSELLRFQVTDLDEE  247
            ++G+        +  E  + +        P F    Y A I EN    S+++  +     +
Sbjct  629  QNGKVDDRRFQSTPEERLSIVG-GKRAPQFYMPSYEAEIPENQKKDSDIISVKA----KS  683

Query  248  YTDNWLAVYFFTSGNEGNWFEIQTDPRTNEGILKVVKALDYEQLQSVKLS----IAVKNK  303
            + D  +         G     F I          GI+K+ K LD+E L+   +      A ++
Sbjct  684  FADREIRYTLKAQGQGAGTFNIGP----TSGIVKLAKELDFEDLRQPHVYSLVVTATEDS  739

Query  304  AEFHQSVISRYRVQSTPVTIQVINVREGIAFRPASKTFTVQKGISSKKLVDYILGT----  359
              F  SV          +TI+V +V +       +  F +    +     D  LGT
Sbjct  740  GGFSTSV---------ELTIRVTDVNDN------APKFELPDYQAHNVDEDIPLGTSILK  784
```

```
Query  360  YQAIDEDTNKAASNVKYVMGRN----DGGYLMIDSKTAEIKFVKNMNRDSTFIVNKTITA  415
             +A+D D+  A + ++Y++  +    D   +++++K  +     N N    F+V
Sbjct  785  VKAMDSDSG-ANAEIEYLVSDDHFAVDSNGIIVNNKQLD---ADNNNAYYEFVVTAKDKG  840

Query  416  EVLAIDEYTGKTSTGTVYVRVPDFNDNCP  444
            E         KT T TV V   + ND  P
Sbjct  841  E-------PAKTGTATVRVYTKNKNDEEP  862

 Score = 39.3 bits (90),  Expect = 0.29, Method: Compositional matrix adjust.
 Identities = 54/242 (22%), Positives = 102/242 (42%), Gaps = 22/242 (9%)

Query  4    KFAKPCREGE--DNSKRNP-IAKITSDYQATQKITYRISGVGIDQPPFGIFVVDKNTGDI  60
            +F  P  E E  +N K++  I  + +   A ++I Y +   G      G F +   +G +
Sbjct  656  QFYMPSYEAEIPENQKKDSDIISVKAKSFADREIRYTLKAQGQGA---GTFNIGPTSGIV  712

Query  61   NITAIVDREE--TPSF--LITCRALNAQGLDVEKPLILTVKILDINDNPPVFSQQIFMGE  116
             +   +D E+   P    L+     ++ G      + LT+++ D+NDN P F    +
Sbjct  713  KLAKELDFEDLRQPHVYSLVVTATEDSGGFSTS--VELTIRVTDVNDNAPKFELPDYQAH  770

Query  117  -IEENSASNSLVMILNATDADEPNHLNSKIAFKIVSQEPAGTPMFLLSRNTGEVRTLTNS  175
             ++E+    + ++ + A D+D  +  N++I + +VS +     F +  N   V
Sbjct  771  NVDEDIPLGTSILKVKAMDSD--SGANAEIEY-LVSDD-----HFAVDSNGIIVNNKQLD  822

Query  176  LDREQASSYRLVVSGADKDGEGLSTQCECNIKVKDVNDNFPMFRDSQYSARIEENILSSE  235
             D   A  Y  VV+  DK     +      +  K+ ND  P F    Y+  ++EN   +
Sbjct  823  ADNNNA-YYEFVVTAKDKGEPAKTGTATVRVYTKNKNDEEPKFSQQVYTPNVDENAGPNT  881

Query  236  LL  237
            L+
Sbjct  882  LV  883

>AARA21_009090.P17036 | transcript=AARA21_009090.R17036 | gene=AARA21_009090
| organism=Anopheles_arabiensis_DONGOLA_2021
| gene_product=neural-cadherin, transcript variant X5 |
transcript_product=neural-cadherin,
transcript variant X7 | location=CM029353:40466514-40666848(+)
| protein_length=3103 | sequence_SO=chromosome
| SO=protein_coding_gene | is_pseudo=false
Length=3103

 Score = 147 bits (370),  Expect = 3e-35, Method: Compositional matrix adjust.
 Identities = 128/446 (29%), Positives = 217/446 (49%), Gaps = 36/446 (8%)

Query  20    PIAKITS---DYQATQKITYRISGVGID--QPPFGIFVVDKNTGDINITAIVDREET---  71
             PI ++T+   D    Q I Y ++G GID   P    F +++ TG+I +   +DR++
Sbjct  1543  PIMQVTATDGDKDRPQNIVYFLTGQGIDPDNPANSKFDINRTTGEIFVLKPLDRDQPNGR  1602

Query  72    PSFLITCRALNAQGLDVEKPLILTVKILDINDNPPVFSQQIFMGEIEENSASNSLVMILN  131
             P +  T  A +  G  +     + V + DINDN P+F Q ++ G + EN  +  +VM +
Sbjct  1603  PQWRFTVFAQDEGGEGLVGYADVQVNLKDINDNAPIFPQGVYFGNVTENGTAGMVVMTMT  1662

Query  132   ATDADEPNH-LNSKIAFKI---VSQEPAGTPMFLLSRNTGEVRTLTNSLDREQASSYRLV  187
             A D D+PN   N+K+ + I   V +E  G+P+F +  +TG ++T    LDRE+   Y +
Sbjct  1663  AVDYDDPNEGTNAKLIYSIEKNVIEEETGSPIFEIEADTGVIKTAVCCLDRERTPDYSIQ  1722

Query  188   VSGADKDGEGLSTQCECNIKVKDVNDNFPMFRDSQYSARIEENILSS--ELLRFQVTDLD  245
             V     DG GL      +I+VKD+ND  P F   ++   ++E   ++  E+    VT  D
Sbjct  1723  VVAM--DGGGLKGTGTASIRVKDINDMPPQFTKDEWFTEVDETDGTNLPEMPILTVTVHD  1780

Query  246   EEYTDNWLAVYFFTSGNEGNWFEIQTDPRTNEGI--LKVVKALDYE---QLQSVKLSIAV  300
             E+ T+ +       SG   + F +    R N+G   LK+V+ LDYE   Q    +  I V
Sbjct  1781  EDETNKFQYKVIDNSGYGADKFTMV---RNNDGTGSLKIVQPLDYEDPLQSNGFRFRIQV  1837

Query  301   KNKAEFHQSVISRYRVQSTPVTIQVINVREGIAFRPASKTFTVQKGISSKKLVDYILGTY  360
              +K E + +   +Y V  + V +++ ++ +    +P  +   ++  +     V   L T+
Sbjct  1838  NDKGEDNDN--DKYHVAYSWVVVKLRDINDN---KPQFERPNIEVSVYENADVGKTLETF  1892

Query  361   QAIDEDTNKAASNVKYVMGR-NDGGYLMIDSKTAEIKFVKNMNRDSTFIVNKTITAEVLA  419
             +A D D     S V Y + R +D       ++   +   +N++R+    V      ++LA
```

```
Sbjct  1893  KATDPDQG-GKSKVSYAIDRSSDRQRQFSINQEGTVTIQRNLDRE----VTPRHQVKILA  1947

Query  420   IDE-YTGKTSTGTVYVRVPDFNDNCP  444
             ID+    KT+T T+ V V D NDN P
Sbjct  1948  IDDGIPPKTATATLTVIVQDINDNPP  1973

 Score = 75.9 bits (185),  Expect = 2e-12, Method: Compositional matrix adjust.
 Identities = 106/426 (25%), Positives = 181/426 (42%), Gaps = 51/426 (12%)

Query  53    VDKNTGDINITAIVDREETPSFLITCRALNAQGLDVEKPLILTVKILDINDNPPVFSQ-Q  111
             +D++  ++N+TA+ D         +C  +N          ++ V I D+NDN PVF
Sbjct  928   LDRDKYELNVTAMDDG--------SC-CVNGDATIHTSTAVVVVFITDVNDNKPVFKDCG  978

Query  112   IFMGEIEENSASNSLVMILNATDADEPNHLNSKIAFKIVSQEPAGTPMFLLSRNTGEVRT  171
              +  ++EE + + S V+ + ATD D+   +N ++ + IV Q       F +   TGEV T
Sbjct  979   TYYPKVEEGAPNGSPVIKVQATDEDK--GVNGQVKYSIVQQPNQKGTKFTVDEETGEVST  1036

Query  172   LTNSLDRE--QASSYRLVVSGADKDGEGLSTQCECNIKVKDVNDNFPMFRDSQYSARIEE  229
                  DRE        + V   D+    L   C   +++ DVNDN P+F   +Y   +++
Sbjct  1037  -NKVFDREGDDGKFVSVTVKATDQGEPSLEGVCSFTVEITDVNDNPPLFDRQKYVENVKQ  1095

Query  230   NI-LSSELLRFQVTDLDEEYTDNWLAVYFFT---SGNEGNWFEIQTDPRTNEGILKVVKA  285
             +  + + +LR  V+  DE+  +N   VY  +   S N+  +FEIQ  P +   +LK  K
Sbjct  1096  DASIGTNILR--VSASDEDADNNGAIVYTLSAPHSPNDLEYFEIQ--PESGWIVLK--KP  1149

Query  286   LD--YEQLQSVKLSIAVKNK------AEFHQSVISRYRVQSTPVTIQV----INVREGIA  333
             LD  Y + +  +L +   ++      A+    +    R    P+  +     I++RE +
Sbjct  1150  LDGCYSR-ERYRLKVYASDRGDPPSTADVDVELDVVDRNNKPPIWDRTIYGPIHIRENVT  1208

Query  334   FRPASKTFTVQKGISSKKLVDYILGTYQAIDEDTNKAASNVKYVMGRNDGGYLMIDSKTA  393
                   +     GI     V Y L         TNK   +  Y+  R D G        A
Sbjct  1209  VGTVVTSVKASSGIEGNPTVFYRL--MPGSTAQTNKL--HTFYLQQRPDNG-----DTWA  1259

Query  394   EIKFVKNMNRDSTFIVNKTITAEVLAIDEYTGKTSTGTVYVRVPDFNDNCPTAV-LEKDA  452
             +IK    ++ ++    N T+  E     +     S  TVY+++ D ND  P     E++
Sbjct  1260  DIKVNHPLDFETIKEYNLTVRVENNGAQQL---ASEATVYIQLEDVNDEIPLFTEREQET  1316

Query  453   VCSSSP  458
             V    P
Sbjct  1317  VLEGEP  1322

 Score = 75.5 bits (184),  Expect = 2e-12, Method: Compositional matrix adjust.
 Identities = 103/418 (25%), Positives = 174/418 (42%), Gaps = 50/418 (12%)

Query  50    IFVVDKNTGDINITAIVDREETPSFLITCRAL----------NAQGLDVEKPLILTVKIL  99
              F ++  +G+I    + DRE+  ++ +   A           N Q   V K   + + I
Sbjct  1359  FFEINLQSGEIFTKVVFDREKQGAYALEVEARDGAPSARPNSNQQPNSVTK--FIRIGIA  1416

Query  100   DINDNPPVFSQQIFMGEIEENSASNSLVMILNATDADEPNHLNSKIAFKIVSQEPAGTPM  159
             D NDNPP F + ++  E++EN      V+ + A D DE    +S+I ++I S    G
Sbjct  1417  DKNDNPPYFDKALYEAEVDENEDIQHTVLTVTAKDHDE----SSRIRYEITSGNIGGA--  1470

Query  160   FLLSRNTGEVRTLTNSLDREQASSYRLVVSGADKDGEGLSTQCECNIKVKDVNDNFPMFR  219
             F +   TG +  +  +LD E    Y L +  +D   E  +      I VKDVND  P F
Sbjct  1471  FAVKNMTGAI-YVAGALDYETRKRYYLTLVASDSLNENETIIV---INVKDVNDMPPSFP  1526

Query  220   DSQYSARIEENI-LSSELLRFQVTDLDEEYTDNWLAVYFFTSG--NEGNWFEIQTDPRTN  276
                Y   ++E + +   +++   TD D++   N   VYF T    +  N    + D
Sbjct  1527  QQVYERTMDEELAVPFPIMQVTATDGDKDRPQN--IVYFLTGQGIDPDNPANSKFDINRT  1584

Query  277   EGILKVVKALDYEQLQS-VKLSIAVKNKAEFHQSVISRYRVQSTPVTIQVINVREGIAFR  335
              G + V+K LD +Q     +    V  + E  + ++    VQ     + + ++ +
Sbjct  1585  TGEIFVLKPLDRDQPNGRPQWRFTVFAQDEGGEGLVGYADVQ-----VNLKDINDNAPIF  1639

Query  336   PASKTFTVQKGISSKKLVDYILGTYQAIDEDTNKAASNVK--YVMGRN------DGGYLM  387
             P    F     ++       ++ T  A+D D     +N K  Y + +N
Sbjct  1640  PQGVYF---GNVTENGTAGMVVMTMTAVDYDDPNEGTNAKLIYSIEKNVIEEETGSPIFE  1696

Query  388   IDSKTAEIK-FVKNMNRDSTFIVNKTITAEVLAIDEYTGKTSTGTVYVRVPDFNDNCP  444
```

```
             I++ T  IK  V  ++R+ T       + +V+A+D   G   TGT  +RV D ND  P
Sbjct  1697  IEADTGVIKTAVCCLDRERT----PDYSIQVVAMDG-GGLKGTGTASIRVKDINDMPP  1749

 Score = 60.5 bits (145),  Expect = 1e-07, Method: Compositional matrix adjust.
 Identities = 83/362 (23%), Positives = 152/362 (42%), Gaps = 34/362 (9%)

Query  51    FVVDKNTGDINITAIVDRE-ETPSFL-ITCRALNAQGLDVEKPLILTVKILDINDNPPVF  108
             F VD+ TG+++   + DRE +   F+ +T +A +     +E     TV+I D+NDNPP+F
Sbjct  1025  FTVDEETGEVSTNKVFDREGDDGKFVSVTVKATDQGEPSLEGVCSFTVEITDVNDNPPLF  1084

Query  109   SQQIFMGEIEENSASNSLVMILNATDADEPNHLNSKIAFKIVS-QEPAGTPMFLLSRNTG  167
              +Q ++  ++++++  + ++ ++A+D D  N  N  I + + +   P     F +   +G
Sbjct  1085  DRQKYVENVKQDASIGTNILRVSASDEDADN--NGAIVYTLSAPHSPNDLEYFEIQPESG  1142

Query  168   EV---RTLTNSLDREQASSYRLVVSGADKDGEGLSTQCECNIKVKDVNDNFPMFRDSQYS  224
              +   + L     RE+   YRL V  +D+ G+  ST             +N P   D
Sbjct  1143  WIVLKKPLDGCYSRER---YRLKVYASDR-GDPPSTADVDVELDVVDRNNKPPIWDRTIY  1198

Query  225   ARIEENILSSELLRFQVTDLDEEYTDNWLAVYFFTSG-----NEGNWFEIQTDPRTNE--  277
               I      +                N    Y    G     N+ + F +Q  P   +
Sbjct  1199  GPIHIRENVTVGTVVTSVKASSGIEGNPTVFYRLMPGSTAQTNKLHTFYLQQRPDNGDTW  1258

Query  278   GILKVVKALDYEQLQSVKLSIAVKNKAEFHQSVISRYRVQSTPVTIQVINVREGIAFRPA  337
               +KV   LD+E ++   L++ V+N          ++       V IQ+ +V + I
Sbjct  1259  ADIKVNHPLDFETIKEYNLTVRVENNG-------AQQLASEATVYIQLEDVNDEIPLFTE  1311

Query  338   SKTFTVQKGISSKKLVDYILGTYQAIDEDTNKAASNVKYVM---GRNDG-GYLMIDSKTA  393
              +  TV +G      V  +     AID+D     + V Y +    RN+G  +  I+ ++
Sbjct  1312  REQETVLEGEPIGTKVTQV----NAIDKDGTFPNNQVYYYIVDSPRNEGKDFFEINLQSG  1367

Query  394   EI  395
             EI
Sbjct  1368  EI  1369

 Score = 55.8 bits (133),  Expect = 3e-06, Method: Compositional matrix adjust.
 Identities = 49/158 (31%), Positives = 78/158 (49%), Gaps = 24/158 (15%)

Query  102   NDNPPVFSQQIFMGEIEENSASNSLVMILNATDADEPNHLNSKIAFKIVSQEPAGTP--M  159
             ND  P FSQQ++   ++EN+  N+LV  + A+D D  N     + F  V     GT
Sbjct  858   NDEEPKFSQQVYTPNVDENAGPNTLVTTVVASDKDGDN-----VRFGFVG---GGTSSGQ  909

Query  160   FLLSRNTGEVRTLTN--SLDREQASSYRLVVSGADK-------DGEGLSTQCECNIKVKD  210
             F++   TG +R  +   SLDR++   Y L V+  D         D    ++      + + D
Sbjct  910   FVIEEITGVIRLHSKAISLDRDK---YELNVTAMDDGSCCVNGDATIHTSTAVVVVFITD  966

Query  211   VNDNFPMFRD-SQYSARIEENILS-SELLRFQVTDLDE  246
             VNDN P+F+D   Y  ++EE   + S +++ Q TD D+
Sbjct  967   VNDNKPVFKDCGTYYPKVEEGAPNGSPVIKVQATDEDK  1004

 Score = 53.5 bits (127),  Expect = 1e-05, Method: Compositional matrix adjust.
 Identities = 70/306 (23%), Positives = 126/306 (41%), Gaps = 30/306 (10%)

Query  20    PIAKIT-SDYQATQKITYRI---SGVGIDQPPFGIFVVDKNTGDINITAIVDRE---ETP  72
             PI  +T  D   T K  Y++   SG G D+  F +   +  TG + I   +D E   ++
Sbjct  1772  PILTVTVHDEDETNKFQYKVIDNSGYGADK--FTMVRNNDGTGSLKIVQPLDYEDPLQSN  1829

Query  73    SFLITCRALNAQGLDVEKPLI------LTVKILDINDNPPVFSQQIFMGEIEENSASNSL  126
              F    + +N +G D +           + VK+ DINDN P F +      + EN+
Sbjct  1830  GFRFRIQ-VNDKGEDNDNDKYHVAYSWVVVKLRDINDNKPQFERPNIEVSVYENADVGKT  1888

Query  127   VMILNATDADEPNHLNSKIAFKIVSQEPAGTPMFLLSRNTGEVRTLTNSLDREQASSYRL  186
             +    ATD D+      SK+++ I   + +        S N     T+  +LDRE    +++
Sbjct  1889  LETFKATDPDQGG--KSKVSYAI---DRSSDRQRQFSINQEGTVTIQRNLDREVTPRHQV  1943

Query  187   VVSGADKDGEGLSTQCECNIKVKDVNDNFPMFRDSQYSARIEENILSSELLRFQVTDLDE  246
              +   D      +       + V+D+NDN P F    Y   + E++    +++    TD D+
Sbjct  1944  KILAIDDGIPPKTATATLTVIVQDINDNPPKFL-KDYRPVLPEHVPPRKVVEILATDDDD  2002
```

```
Query  247   EYTDNWLAVYFFTSGNEGNW----FEIQTDPRTNEG----ILKVVKALDYEQLQSVKLSI  298
                 N     F       +     F+++ D +   G    I+  +++ D EQ +   + I
Sbjct  2003  RSKSNGPPFQFRLDPGADDIIRASFKVEQDQKGANGDGMAIVSSLRSFDREQQKEYLIPI  2062

Query  299   AVKNKA  304
              +K+
Sbjct  2063  VIKDHG  2068

 Score = 52.0 bits (123),  Expect = 4e-05, Method: Compositional matrix adjust.
 Identities = 97/398 (24%), Positives = 163/398 (41%), Gaps = 54/398 (14%)

Query  106   PVFSQQIFMGEIEENSASNSLVMILNATDADEPNHLNSKIAFKIVSQ-EPAGTPMFLLSR  164
             P F    +  EI EN   +S ++ + A      +  + +I + + +Q + AGT  F +
Sbjct  655   PQFYMPSYEAEIPENQKKDSDIISVKAK-----SFADREIRYTLKAQGQGAGT--FNIGP  707

Query  165   NTGEVRTLTNSLDRE---QASSYRLVVSGADKDGEGLSTQCECNIKVKDVNDNFPMFRDS  221
              +G V+ L   LD E   Q   Y LVV+ A +D  G ST  E  I+V DVNDN P F
Sbjct  708   TSGIVK-LAKELDFEDLRQPHVYSLVVT-ATEDSGGFSTSVELTIRVTDVNDNAPKFELP  765

Query  222   QYSAR-IEENI-LSSELLRFQVTDLDEEYTDNWLAVYFFTSGNEGNWFEIQTDPRTNEGI  279
              Y A  ++E+I L + +L+ +  D D         + +  S +    F + ++     GI
Sbjct  766   DYQAHNVDEDIPLGTSILKVKAMDSDSGAN---AEIEYLVSDDH---FAVDSN-----GI  814

Query  280   LKVVKALDYEQLQS-VKLSIAVKNKAEFHQSVISRYRVQSTPVTIQVINVREGIAFRPAS  338
             +   K LD +   +  +  +  K+K E  ++  +  RV +           E   F
Sbjct  815   IVNNKQLDADNNNAYYEFVVTAKDKGEPAKTGTATVRVYTKNKN------DEEPKFSQQV  868

Query  339   KTFTVQKGISSKKLVDYILGTYQAIDEDTNKAASNVK--YVMGRNDGGYLMIDSKTAEIK  396
              T  V +      LV  ++         ++K   NV+  +V G    G  +I+  T  I+
Sbjct  869   YTPNVDENAGPNTLVTTVVA--------SDKDGDNVRFGFVGGGTSSGQFVIEEITGVIR  920

Query  397   FVK---NMNRDSTFI-VNKTITAEVLAIDEYTGKTSTGTVYVRVPDFNDN------CPTA  446
                   +++RD   + V            + T  TST  V V + D NDN      C T
Sbjct  921   LHSKAISLDRDKYELNVTAMDDGSCCVNGDATIHTSTAVVVVFITDVNDNKPVFKDCGTY  980

Query  447   VLE-KDAVCSSSPSVVVSARTLNNRYTGPYTFALEDQP  483
               + ++   + SP + V A   +    G   +++  QP
Sbjct  981   YPKVEEGAPNGSPVIKVQATDEDKGVNGQVKYSIVQQP  1018

 Score = 44.3 bits (103),  Expect = 0.009, Method: Compositional matrix adjust.
 Identities = 48/199 (24%), Positives = 73/199 (37%), Gaps = 19/199 (10%)

Query  27    DYQATQKITYRISGVGIDQPPFGIFVVDKNTGDINITAIVDREETPSFLITCRALNAQGL  86
             D     K++Y I      Q  F I       G + I   +DRE TP   +   A++
Sbjct  1898  DQGGKSKVSYAIDRSSDRQRQFSI----NQEGTVTIQRNLDREVTPRHQVKILAIDDGIP  1953

Query  87    DVEKPLILTVKILDINDNPPVFSQQIFMGEIEENSASNSLVMILNATDADEPNHLNSKIA  146
                    LTV + DINDNPP F +  +   + E+     +V IL   D D
Sbjct  1954  PKTATATLTVIVQDINDNPPKFLKD-YRPVLPEHVPPRKVVEILATDDDDRSKSNGPPFQ  2012

Query  147   FKIVSQEPAGTPMFLLSRNTGEVRTLTN-----------SLDREQASSYRLVVSGADKDG  195
             F++   +P    +   S    + +   N           S DREQ   Y + +   D
Sbjct  2013  FRL---DPGADDIIRASFKVEQDQKGANGDGMAIVSSLRSFDREQQKEYLIPIVIKDHGN  2069

Query  196   EGLSTQCECNIKVKDVNDN  214
               ++      + + DVNDN
Sbjct  2070  PAMTGTSTLTVVIGDVNDN  2088

 Score = 39.7 bits (91),  Expect = 0.28, Method: Compositional matrix adjust.
 Identities = 89/389 (23%), Positives = 149/389 (38%), Gaps = 62/389 (16%)

Query  75   LITCRALNAQGLDVEKPLILTVKILDINDNPPVFSQQIF--MGEIEENSASNSLVMILNA  132
            +IT +  NA GL       + + + D+ND PP F  +       ++ N+  N+ V  L A
Sbjct  517  IITNQGHNA-GLKYTDNQRVIIHVKDVNDEPPYFINRPLPMQAVVQLNAPPNTPVFTLQA  575

Query  133  TDADEPNHLNSKIAFKIVSQEPAGTPMFLLSRNTGEVRTLTNSLDREQASSYRLVVSGAD  192
             D D     +  I + IV     G   F +   +G VRT    L  +    Y L V   D
Sbjct  576  RDPD----TDHNIHYFIVRDRTGG--RFEVDERSGVVRTRGTDL-FQLDMEYVLYVKAED  628
```

```
Query  193  KDGE----GLSTQCECNIKVKDVNDNFPMFRDSQYSARIEEN-ILSSELLRFQVTDLDEE  247
            ++G+        +  E  + +        P F    Y A I EN    S+++  +     +
Sbjct  629  QNGKVDDRRFQSTPEERLSIVG-GKRAPQFYMPSYEAEIPENQKKDSDIISVKA----KS  683

Query  248  YTDNWLAVYFFTSGNEGNWFEIQTDPRTNEGILKVVKALDYEQLQSVKLS----IAVKNK  303
            + D  +        G     F I         GI+K+ K LD+E L+   +       A ++
Sbjct  684  FADREIRYTLKAQGQGAGTFNIGP----TSGIVKLAKELDFEDLRQPHVYSLVVTATEDS  739

Query  304  AEFHQSVISRYRVQSTPVTIQVINVREGIAFRPASKTFTVQKGISSKKLVDYILGT----  359
              F  SV           +TI+V +V +        +  F +    +     D  LGT
Sbjct  740  GGFSTSV---------ELTIRVTDVNDN------APKFELPDYQAHNVDEDIPLGTSILK  784

Query  360  YQAIDEDTNKAASNVKYVMGRN----DGGYLMIDSKTAEIKFVKNMNRDSTFIVNKTITA  415
             +A+D D+  A + ++Y++  +    D    +++++K  +     N N    F+V
Sbjct  785  VKAMDSDSG-ANAEIEYLVSDDHFAVDSNGIIVNNKQLD---ADNNNAYYEFVVTAKDKG  840

Query  416  EVLAIDEYTGKTSTGTVYVRVPDFNDNCP  444
            E          KT T TV V   + ND  P
Sbjct  841  E-------PAKTGTATVRVYTKNKNDEEP  862

 Score = 39.3 bits (90),  Expect = 0.30, Method: Compositional matrix adjust.
 Identities = 54/242 (22%), Positives = 102/242 (42%), Gaps = 22/242 (9%)

Query  4    KFAKPCREGE--DNSKRNP-IAKITSDYQATQKITYRISGVGIDQPPFGIFVVDKNTGDI  60
            +F  P  E E  +N K++  I  + +   A ++I Y +   G       G F +   +G +
Sbjct  656  QFYMPSYEAEIPENQKKDSDIISVKAKSFADREIRYTLKAQGQGA---GTFNIGPTSGIV  712

Query  61   NITAIVDREE--TPSF--LITCRALNAQGLDVEKPLILTVKILDINDNPPVFSQQIFMGE  116
             +   +D E+   P    L+      ++ G       + LT+++ D+NDN P F    +
Sbjct  713  KLAKELDFEDLRQPHVYSLVVTATEDSGGFSTS--VELTIRVTDVNDNAPKFELPDYQAH  770

Query  117  -IEENSASNSLVMILNATDADEPNHLNSKIAFKIVSQEPAGTPMFLLSRNTGEVRTLTNS  175
             ++E+    + ++ + A D+D  +  N++I + +VS +       F +  N   V
Sbjct  771  NVDEDIPLGTSILKVKAMDSD--SGANAEIEY-LVSDD-----HFAVDSNGIIVNNKQLD  822

Query  176  LDREQASSYRLVVSGADKDGEGLSTQCECNIKVKDVNDNFPMFRDSQYSARIEENILSSE  235
             D   A  Y  VV+  DK      +       +  K+ ND  P F    Y+  ++EN   +
Sbjct  823  ADNNNA-YYEFVVTAKDKGEPAKTGTATVRVYTKNKNDEEPKFSQQVYTPNVDENAGPNT  881

Query  236  LL  237
            L+
Sbjct  882  LV  883

>ADAR2_001930.P21644 | transcript=ADAR2_001930.R21644 | gene=ADAR2_001930
| organism=Anopheles_darlingi_AdarGF1 | gene_product=neural-cadherin,
transcript variant X9 | transcript_product=neural-cadherin,
transcript variant X9 | location=AdarGF1_2RL:74991964-75171550(+)
| protein_length=3128 | sequence_SO=chromosome
| SO=protein_coding_gene | is_pseudo=false
Length=3128

 Score = 147 bits (370),  Expect = 3e-35, Method: Compositional matrix adjust.
 Identities = 128/446 (29%), Positives = 217/446 (49%), Gaps = 36/446 (8%)

Query  20    PIAKITS---DYQATQKITYRISGVGID--QPPFGIFVVDKNTGDINITAIVDREET---  71
             PI ++T+    D    Q I Y ++G GID   P    F +++ TG+I +    +DR++
Sbjct  1569  PIMQVTATDGDKDRPQNIVYFLTGQGIDPDNPANSKFDINRTTGEIFVLKPLDRDQPNGR  1628

Query  72    PSFLITCRALNAQGLDVEKPLILTVKILDINDNPPVFSQQIFMGEIEENSASNSLVMILN  131
             P +  T  A +  G  +       + V + DINDN P+F Q ++ G + EN  +  +VM +
Sbjct  1629  PQWRFTVFAQDEGGEGLVGYADVQVNLKDINDNAPIFPQGVYFGNVTENGTAGMVVMTMT  1688

Query  132   ATDADEPNH-LNSKIAFKI---VSQEPAGTPMFLLSRNTGEVRTLTNSLDREQASSYRLV  187
             A D D+PN   N+K+ + I   V +E  G+P+F +  +TG ++T     LDRE+   Y +
Sbjct  1689  AVDYDDPNEGTNAKLIYSIEKNVIEEETGSPIFEIEADTGVIKTAVCCLDRERTPDYSIQ  1748

Query  188   VSGADKDGEGLSTQCECNIKVKDVNDNFPMFRDSQYSARIEENILSS--ELLRFQVTDLD  245
             V      DG GL       +I+VKD+ND  P F   ++   ++E   ++  E+     VT  D
```

```
Sbjct  1749   VVAM--DGGGLKGTGTASIRVKDINDMPPQFTKDEWFTEVDETDGTNLPEMPILTVTVHD  1806

Query  246    EEYTDNWLAVYFFTSGNEGNWFEIQTDPRTNEGI--LKVVKALDYE---QLQSVKLSIAV  300
              E+ T+ +       SG   + F +    R N+G   LK+V+ LDYE   Q     +  I V
Sbjct  1807   EDETNKFQYKVIDNSGYGADKFTMV---RNNDGTGSLKIVQPLDYEDPLQSNGFRFRIQV  1863

Query  301    KNKAEFHQSVISRYRVQSTPVTIQVINVREGIAFRPASKTFTVQKGISSKKLVDYILGTY  360
               +K E + +   +Y V  + V +++ ++ +    +P  +   ++  +     V   L T+
Sbjct  1864   NDKGEDNDN--DKYHVAYSWVVVKLRDINDN---KPQFERPNIEVSVYENADVGKTLETF  1918

Query  361    QAIDEDTNKAASNVKYVMGR-NDGGYLMIDSKTAEIKFVKNMNRDSTFIVNKTITAEVLA  419
              +A D D     S V Y + R +D       ++   +   +N++R+    V      ++LA
Sbjct  1919   KATDPDQG-GKSKVSYAIDRSSDRQRQFSINQEGTVTIQRNLDRE----VTPRHQVKILA  1973

Query  420    IDE-YTGKTSTGTVYVRVPDFNDNCP  444
              ID+    KT+T T+ V V D NDN P
Sbjct  1974   IDDGIPPKTATATLTVIVQDINDNPP  1999

 Score = 80.1 bits (196),  Expect = 7e-14, Method: Compositional matrix adjust.
 Identities = 107/424 (25%), Positives = 178/424 (42%), Gaps = 51/424 (12%)

Query  53     VDKNTGDINITAIVDREETPSFLITCRALNAQGLDVEKPLILTVKILDINDNPPVFSQ-Q  111
              +D++  ++N+TA+ D         +C  +N          ++ V I D+NDN PVF
Sbjct  957    LDRDKYELNVTAMDDG--------SC-CVNGDATIHTSTAVVVVFITDVNDNKPVFKDCG  1007

Query  112    IFMGEIEENSASNSLVMILNATDADEPNHLNSKIAFKIVSQEPAGTPMFLLSRNTGEVRT  171
               +  ++EE + + S V+ + ATD D+   +N ++ + IV Q       F +   TGEV T
Sbjct  1008   TYYPKVEEGAPNGSPVIKVQATDEDK--GVNGQVKYSIVQQPNQKGTKFTVDEETGEVST  1065

Query  172    LTNSLDRE--QASSYRLVVSGADKDGEGLSTQCECNIKVKDVNDNFPMFRDSQYSARIEE  229
                   DRE        + V   D+    L   C   +++ DVNDN P+F   +Y   +++
Sbjct  1066   -NKVFDREGDDGKFVSVTVKATDQGEPSLEGVCSFTVEITDVNDNPPLFDRQKYVENVKQ  1124

Query  230    NI-LSSELLRFQVTDLDEEYTDNWLAVYFFT---SGNEGNWFEIQTDPRTNEGILKVVKA  285
              +  + + +LR  V+  DE+  +N   VY  +   S N+  +FEIQ  P +   +LK  K
Sbjct  1125   DASIGTNILR--VSASDEDADNNGAIVYTLSAPHSPNDLEYFEIQ--PESGWIVLK--KP  1178

Query  286    LDYEQLQSVKLSIAVKNK------AEFHQSVISRYRVQSTPV----TIQVINVREGIAFR  335
              LD E+    +L +   ++      A+    +    R    P+        I++RE +
Sbjct  1179   LDRERY---RLKVYASDRGDPPSTADVDVELDVVDRNNKPPIWDRNIYGPIHIRENVTVG  1235

Query  336    PASKTFTVQKGISSKKLVDYILGTYQAIDEDTNKAASNVKYVMGRNDGGYLMIDSKTAEI  395
                        GI     V Y L         TNK   +  Y+  R D G        A+I
Sbjct  1236   TVVTAIKASSGIEGNPTVFYRL--MPGSTAQTNKL--HTFYLQQRPDNG-----DTWADI  1286

Query  396    KFVKNMNRDSTFIVNKTITAEVLAIDEYTGKTSTGTVYVRVPDFNDNCPTAV-LEKDAVC  454
              K    ++ ++    N TI  E     +     S  TVY+++ D ND  P     E++ V
Sbjct  1287   KVNHPLDFETIKEYNLTIRVENNGAQQL---ASEATVYIQLEDVNDEIPLFTEREQETVL  1343

Query  455    SSSP  458
                 P
Sbjct  1344   EGEP  1347

 Score = 77.4 bits (189),  Expect = 6e-13, Method: Compositional matrix adjust.
 Identities = 103/419 (25%), Positives = 173/419 (41%), Gaps = 51/419 (12%)

Query  50     IFVVDKNTGDINITAIVDREETPSFLITCRAL----------NAQGLDVEKPLILTVKIL  99
               F ++  +G+I    + DRE+  ++ +   A           N Q   V K   + + I
Sbjct  1384   FFEINLQSGEIFTKVVFDREKQGAYALEVEARDGAPSARPNSNQQPNSVTK--FIRIGIA  1441

Query  100    DINDNPPVFSQQIFMGEIEENSASNSLVMILNATDADEPNHLNSKIAFKIVSQEPAGTPM  159
              D NDNPP F + ++  E++EN      V+ + A D DE    +S+I ++I S    G
Sbjct  1442   DKNDNPPYFDKALYEAEVDENEDIQHTVLTVTAKDHDE----SSRIRYEITSGNIGGA--  1495

Query  160    FLLSRNTGEVRTLTNSLDREQASSYRLVVSGADKDGEGLSTQCECNIKVKDVNDNFPMFR  219
              F +   TG +  +  +LD E    Y L +  +D   E    +    I VKDVND  P F
Sbjct  1496   FAVKNMTGAI-YVAGALDYETRKRYHLTLVASDSLNE---NETIIEIHVKDVNDMPPTFS  1551

Query  220    DSQY--SARIEENILSSELLRFQVTDLDEEYTDNWLAVYFFTSG--NEGNWFEIQTDPRT  275
```

```
                Y  +   E+ +    +++   TD D++   N   VYF T    +  N    + D
Sbjct  1552  QQVYETTQDEEQTVTDRPIMQVTATDGDKDRPQN--IVYFLTGQGIDPDNPANSKFDINR  1609

Query  276   NEGILKVVKALDYEQLQS-VKLSIAVKNKAEFHQSVISRYRVQSTPVTIQVINVREGIAF  334
               G + V+K LD +Q     +    V  + E  + ++    VQ     + + ++ +
Sbjct  1610  TTGEIFVLKPLDRDQPNGRPQWRFTVFAQDEGGEGLVGYADVQ-----VNLKDINDNAPI  1664

Query  335   RPASKTFTVQKGISSKKLVDYILGTYQAIDEDTNKAASNVK--YVMGRN------DGGYL  386
              P    F     ++       ++ T  A+D D     +N K  Y + +N
Sbjct  1665  FPQGVYF---GNVTENGTAGMVVMTMTAVDYDDPNEGTNAKLIYSIEKNVIEEETGSPIF  1721

Query  387   MIDSKTAEIK-FVKNMNRDSTFIVNKTITAEVLAIDEYTGKTSTGTVYVRVPDFNDNCP  444
              I++ T  IK  V  ++R+ T       + +V+A+D   G   TGT  +RV D ND  P
Sbjct  1722  EIEADTGVIKTAVCCLDRERT----PDYSIQVVAMDG-GGLKGTGTASIRVKDINDMPP  1775

 Score = 68.2 bits (165),  Expect = 4e-10, Method: Compositional matrix adjust.
 Identities = 88/361 (24%), Positives = 157/361 (43%), Gaps = 36/361 (10%)

Query  51    FVVDKNTGDINITAIVDRE-ETPSFL-ITCRALNAQGLDVEKPLILTVKILDINDNPPVF  108
             F VD+ TG+++   + DRE +   F+ +T +A +     +E     TV+I D+NDNPP+F
Sbjct  1054  FTVDEETGEVSTNKVFDREGDDGKFVSVTVKATDQGEPSLEGVCSFTVEITDVNDNPPLF  1113

Query  109   SQQIFMGEIEENSASNSLVMILNATDADEPNHLNSKIAFKIVS-QEPAGTPMFLLSRNTG  167
              +Q ++  ++++++  + ++ ++A+D D  N  N  I + + +   P     F +   +G
Sbjct  1114  DRQKYVENVKQDASIGTNILRVSASDEDADN--NGAIVYTLSAPHSPNDLEYFEIQPESG  1171

Query  168   EVRTLTNSLDREQASSYRLVVSGADKDGEGLSTQCECNIKVKDVNDNFPMFRDSQYSA--  225
              +  L   LDRE+   YRL V  +D+ G+  ST            +N P   D
Sbjct  1172  WI-VLKKPLDRER---YRLKVYASDR-GDPPSTADVDVELDVVDRNNKPPIWDRNIYGPI  1226

Query  226   RIEENILSSELLRFQVTDLDEEYTDNWLAVYFFTSG-----NEGNWFEIQTDPRTNE--G  278
              I EN+    ++             N    Y    G     N+ + F +Q  P   +
Sbjct  1227  HIRENVTVGTVV--TAIKASSGIEGNPTVFYRLMPGSTAQTNKLHTFYLQQRPDNGDTWA  1284

Query  279   ILKVVKALDYEQLQSVKLSIAVKNKAEFHQSVISRYRVQSTPVTIQVINVREGIAFRPAS  338
              +KV   LD+E ++   L+I V+N         ++       V IQ+ +V + I
Sbjct  1285  DIKVNHPLDFETIKEYNLTIRVENNG-------AQQLASEATVYIQLEDVNDEIPLFTER  1337

Query  339   KTFTVQKGISSKKLVDYILGTYQAIDEDTNKAASNVKYVM---GRNDG-GYLMIDSKTAE  394
             +  TV +G      V  +     AID+D     + V Y +    RN+G  +  I+ ++ E
Sbjct  1338  EQETVLEGEPIGTKVTQV----NAIDKDGTFPNNQVYYYIVDSPRNEGKDFFEINLQSGE  1393

Query  395   I  395
             I
Sbjct  1394  I  1394

 Score = 55.8 bits (133),  Expect = 3e-06, Method: Compositional matrix adjust.
 Identities = 49/158 (31%), Positives = 78/158 (49%), Gaps = 24/158 (15%)

Query  102   NDNPPVFSQQIFMGEIEENSASNSLVMILNATDADEPNHLNSKIAFKIVSQEPAGTP--M  159
             ND  P FSQQ++   ++EN+  N+LV  + A+D D  N     + F  V     GT
Sbjct  887   NDEEPKFSQQVYTPNVDENAGPNTLVTTVVASDKDGDN-----VRFGFVG---GGTSSGQ  938

Query  160   FLLSRNTGEVRTLTN--SLDREQASSYRLVVSGADK-------DGEGLSTQCECNIKVKD  210
             F++   TG +R  +   SLDR++   Y L V+  D        D    ++     + + D
Sbjct  939   FVIEEITGVIRLHSKAISLDRDK---YELNVTAMDDGSCCVNGDATIHTSTAVVVVFITD  995

Query  211   VNDNFPMFRD-SQYSARIEENILS-SELLRFQVTDLDE  246
             VNDN P+F+D   Y  ++EE   + S +++ Q TD D+
Sbjct  996   VNDNKPVFKDCGTYYPKVEEGAPNGSPVIKVQATDEDK  1033

 Score = 53.9 bits (128),  Expect = 1e-05, Method: Compositional matrix adjust.
 Identities = 70/306 (23%), Positives = 126/306 (41%), Gaps = 30/306 (10%)

Query  20    PIAKIT-SDYQATQKITYRI---SGVGIDQPPFGIFVVDKNTGDINITAIVDRE---ETP  72
             PI  +T  D   T K  Y++   SG G D+  F +   +  TG + I   +D E   ++
Sbjct  1798  PILTVTVHDEDETNKFQYKVIDNSGYGADK--FTMVRNNDGTGSLKIVQPLDYEDPLQSN  1855
```

```
Query  73    SFLITCRALNAQGLDVEKPLI------LTVKILDINDNPPVFSQQIFMGEIEENSASNSL  126
              F    + +N +G D +          + VK+ DINDN P F +      + EN+
Sbjct  1856  GFRFRIQ-VNDKGEDNDNDKYHVAYSWVVVKLRDINDNKPQFERPNIEVSVYENADVGKT  1914

Query  127   VMILNATDADEPNHLNSKIAFKIVSQEPAGTPMFLLSRNTGEVRTLTNSLDREQASSYRL  186
             +    ATD D+     SK+++ I   + +       S N     T+  +LDRE    +++
Sbjct  1915  LETFKATDPDQGG--KSKVSYAI---DRSSDRQRQFSINQEGTVTIQRNLDREVTPRHQV  1969

Query  187   VVSGADKDGEGLSTQCECNIKVKDVNDNFPMFRDSQYSARIEENILSSELLRFQVTDLDE  246
              +   D      +      + V+D+NDN P F    Y   + E++   +++    TD D+
Sbjct  1970  KILAIDDGIPPKTATATLTVIVQDINDNPPKFL-KDYRPVLPEHVPPRKVVEILATDDDD  2028

Query  247   EYTDNWLAVYFFTSGNEGNW----FEIQTDPRTNEG----ILKVVKALDYEQLQSVKLSI  298
                 N     F       +     F+++ D +   G    I+ +++  D EQ +   + I
Sbjct  2029  RSKSNGPPFQFRLDPGADDIIRASFKVEQDQKGANGDGMAIVSSLRSFDREQQKEYLIPI  2088

Query  299   AVKNKA  304
              +K+
Sbjct  2089  VIKDHG  2094

 Score = 52.4 bits (124),  Expect = 3e-05, Method: Compositional matrix adjust.
 Identities = 97/398 (24%), Positives = 163/398 (41%), Gaps = 54/398 (14%)

Query  106   PVFSQQIFMGEIEENSASNSLVMILNATDADEPNHLNSKIAFKIVSQ-EPAGTPMFLLSR  164
             P F    +  EI EN   +S ++ + A      +  + +I + + +Q + AGT  F +
Sbjct  684   PQFYMPSYEAEIPENQKKDSDIISVKAK-----SFADREIRYTLKAQGQGAGT--FNIGP  736

Query  165   NTGEVRTLTNSLDRE---QASSYRLVVSGADKDGEGLSTQCECNIKVKDVNDNFPMFRDS  221
              +G V+ L   LD E   Q   Y LVV+ A +D  G ST  E  I+V DVNDN P F
Sbjct  737   TSGIVK-LAKELDFEDLRQPHVYSLVVT-ATEDSGGFSTSVELTIRVTDVNDNAPKFELP  794

Query  222   QYSAR-IEENI-LSSELLRFQVTDLDEEYTDNWLAVYFFTSGNEGNWFEIQTDPRTNEGI  279
              Y A  ++E+I L + +L+ +  D D         + +  S +    F + ++     GI
Sbjct  795   DYQAHNVDEDIPLGTSILKVKAMDADSGAN---AEIEYLVSDDH---FAVDSN-----GI  843

Query  280   LKVVKALDYEQLQS-VKLSIAVKNKAEFHQSVISRYRVQSTPVTIQVINVREGIAFRPAS  338
             +   K LD +   +  +  +  K+K E  ++  +  RV +           E   F
Sbjct  844   IVNNKQLDADNNNAYYEFVVTAKDKGEPAKTGTATVRVYTKNKN------DEEPKFSQQV  897

Query  339   KTFTVQKGISSKKLVDYILGTYQAIDEDTNKAASNVK--YVMGRNDGGYLMIDSKTAEIK  396
              T  V +      LV  ++         ++K   NV+  +V G    G  +I+  T  I+
Sbjct  898   YTPNVDENAGPNTLVTTVVA--------SDKDGDNVRFGFVGGGTSSGQFVIEEITGVIR  949

Query  397   FVK---NMNRDSTFI-VNKTITAEVLAIDEYTGKTSTGTVYVRVPDFNDN------CPTA  446
                   +++RD   + V            + T  TST  V V + D NDN      C T
Sbjct  950   LHSKAISLDRDKYELNVTAMDDGSCCVNGDATIHTSTAVVVVFITDVNDNKPVFKDCGTY  1009

Query  447   VLE-KDAVCSSSPSVVVSARTLNNRYTGPYTFALEDQP  483
               + ++   + SP + V A   +    G   +++  QP
Sbjct  1010  YPKVEEGAPNGSPVIKVQATDEDKGVNGQVKYSIVQQP  1047

 Score = 44.7 bits (104),  Expect = 0.008, Method: Compositional matrix adjust.
 Identities = 48/199 (24%), Positives = 73/199 (37%), Gaps = 19/199 (10%)

Query  27    DYQATQKITYRISGVGIDQPPFGIFVVDKNTGDINITAIVDREETPSFLITCRALNAQGL  86
             D     K++Y I      Q  F I       G + I   +DRE TP   +   A++
Sbjct  1924  DQGGKSKVSYAIDRSSDRQRQFSI----NQEGTVTIQRNLDREVTPRHQVKILAIDDGIP  1979

Query  87    DVEKPLILTVKILDINDNPPVFSQQIFMGEIEENSASNSLVMILNATDADEPNHLNSKIA  146
                    LTV + DINDNPP F +  +   + E+     +V IL   D D
Sbjct  1980  PKTATATLTVIVQDINDNPPKFLKD-YRPVLPEHVPPRKVVEILATDDDDRSKSNGPPFQ  2038

Query  147   FKIVSQEPAGTPMFLLSRNTGEVRTLTN-----------SLDREQASSYRLVVSGADKDG  195
             F++   +P    +   S    + +   N           S DREQ   Y + +   D
Sbjct  2039  FRL---DPGADDIIRASFKVEQDQKGANGDGMAIVSSLRSFDREQQKEYLIPIVIKDHGN  2095

Query  196   EGLSTQCECNIKVKDVNDN  214
               ++      + + DVNDN
Sbjct  2096  PAMTGTSTLTVVIGDVNDN  2114
```

```
 Score = 40.8 bits (94),  Expect = 0.12, Method: Compositional matrix adjust.
 Identities = 54/235 (23%), Positives = 99/235 (42%), Gaps = 22/235 (9%)

Query  4    KFAKPCREGE--DNSKRNP-IAKITSDYQATQKITYRISGVGIDQPPFGIFVVDKNTGDI  60
            +F  P  E E  +N K++  I  + +   A ++I Y +   G      G F +   +G +
Sbjct  685  QFYMPSYEAEIPENQKKDSDIISVKAKSFADREIRYTLKAQGQGA---GTFNIGPTSGIV  741

Query  61   NITAIVDREE--TPSF--LITCRALNAQGLDVEKPLILTVKILDINDNPPVFSQQIFMGE  116
             +   +D E+   P    L+     ++ G      + LT+++ D+NDN P F    +
Sbjct  742  KLAKELDFEDLRQPHVYSLVVTATEDSGGFSTS--VELTIRVTDVNDNAPKFELPDYQAH  799

Query  117  -IEENSASNSLVMILNATDADEPNHLNSKIAFKIVSQEPAGTPMFLLSRNTGEVRTLTNS  175
             ++E+    + ++ + A DAD  +  N++I + +VS +      F +  N   V
Sbjct  800  NVDEDIPLGTSILKVKAMDAD--SGANAEIEY-LVSDD-----HFAVDSNGIIVNNKQLD  851

Query  176  LDREQASSYRLVVSGADKDGEGLSTQCECNIKVKDVNDNFPMFRDSQYSARIEEN  230
             D   A  Y  VV+  DK     +      +  K+ ND  P F    Y+  ++EN
Sbjct  852  ADNNNA-YYEFVVTAKDKGEPAKTGTATVRVYTKNKNDEEPKFSQQVYTPNVDEN  905

 Score = 39.3 bits (90),  Expect = 0.31, Method: Compositional matrix adjust.
 Identities = 89/389 (23%), Positives = 149/389 (38%), Gaps = 62/389 (16%)

Query  75   LITCRALNAQGLDVEKPLILTVKILDINDNPPVFSQQIF--MGEIEENSASNSLVMILNA  132
            +IT +  NA GL       + + + D+ND PP F  +       ++ N+  N+ V  L A
Sbjct  546  IITNQGHNA-GLKYTDNQRVIIHVKDVNDEPPYFINRPLPMQAVVQLNAPPNTPVFTLQA  604

Query  133  TDADEPNHLNSKIAFKIVSQEPAGTPMFLLSRNTGEVRTLTNSLDREQASSYRLVVSGAD  192
             D D     +  I + IV     G   F +   +G VRT    L  +    Y L V   D
Sbjct  605  RDPD----TDHNIHYFIVRDRTGG--RFEVDERSGVVRTRGTDL-FQLDMEYVLYVKAED  657

Query  193  KDGE----GLSTQCECNIKVKDVNDNFPMFRDSQYSARIEEN-ILSSELLRFQVTDLDEE  247
            ++G+       +  E  + +       P F    Y A I EN    S+++  +     +
Sbjct  658  QNGKVDDRRFQSTPEERLSIVG-GKRAPQFYMPSYEAEIPENQKKDSDIISVKA----KS  712

Query  248  YTDNWLAVYFFTSGNEGNWFEIQTDPRTNEGILKVVKALDYEQLQSVKLS----IAVKNK  303
            + D  +       G     F I        GI+K+ K LD+E L+   +      A ++
Sbjct  713  FADREIRYTLKAQGQGAGTFNIGP----TSGIVKLAKELDFEDLRQPHVYSLVVTATEDS  768

Query  304  AEFHQSVISRYRVQSTPVTIQVINVREGIAFRPASKTFTVQKGISSKKLVDYILGT----  359
              F  SV          +TI+V +V +       +  F +    +     D  LGT
Sbjct  769  GGFSTSV---------ELTIRVTDVNDN------APKFELPDYQAHNVDEDIPLGTSILK  813

Query  360  YQAIDEDTNKAASNVKYVMGRN----DGGYLMIDSKTAEIKFVKNMNRDSTFIVNKTITA  415
             +A+D D+  A + ++Y++  +    D   +++++K  +     N N    F+V
Sbjct  814  VKAMDADSG-ANAEIEYLVSDDHFAVDSNGIIVNNKQLD---ADNNNAYYEFVVTAKDKG  869

Query  416  EVLAIDEYTGKTSTGTVYVRVPDFNDNCP  444
            E         KT T TV V   + ND  P
Sbjct  870  E-------PAKTGTATVRVYTKNKNDEEP  891

>ADAR2_001930.P21647 | transcript=ADAR2_001930.R21647 | gene=ADAR2_001930
| organism=Anopheles_darlingi_AdarGF1 | gene_product=neural-cadherin,
transcript variant X5 | transcript_product=neural-cadherin,
transcript variant X5 | location=AdarGF1_2RL:74991964-75171550(+)
| protein_length=3129 | sequence_SO=chromosome
| SO=protein_coding_gene | is_pseudo=false
Length=3129

 Score = 147 bits (370),  Expect = 3e-35, Method: Compositional matrix adjust.
 Identities = 128/446 (29%), Positives = 217/446 (49%), Gaps = 36/446 (8%)

Query  20    PIAKITS---DYQATQKITYRISGVGID--QPPFGIFVVDKNTGDINITAIVDREET---  71
             PI ++T+   D    Q I Y ++G GID   P    F +++ TG+I +   +DR++
Sbjct  1570  PIMQVTATDGDKDRPQNIVYFLTGQGIDPDNPANSKFDINRTTGEIFVLKPLDRDQPNGR  1629

Query  72    PSFLITCRALNAQGLDVEKPLILTVKILDINDNPPVFSQQIFMGEIEENSASNSLVMILN  131
             P +  T  A +  G  +     + V + DINDN P+F Q ++ G + EN  +  +VM +
```

```
Sbjct  1630  PQWRFTVFAQDEGGEGLVGYADVQVNLKDINDNAPIFPQGVYFGNVTENGTAGMVVMTMT  1689

Query  132   ATDADEPNH-LNSKIAFKI---VSQEPAGTPMFLLSRNTGEVRTLTNSLDREQASSYRLV  187
             A D D+PN   N+K+ + I   V +E  G+P+F +  +TG ++T    LDRE+   Y +
Sbjct  1690  AVDYDDPNEGTNAKLIYSIEKNVIEEETGSPIFEIEADTGVIKTAVCCLDRERTPDYSIQ  1749

Query  188   VSGADKDGEGLSTQCECNIKVKDVNDNFPMFRDSQYSARIEENILSS--ELLRFQVTDLD  245
             V     DG GL       +I+VKD+ND  P F   ++   ++E   ++  E+    VT  D
Sbjct  1750  VVAM--DGGGLKGTGTASIRVKDINDMPPQFTKDEWFTEVDETDGTNLPEMPILTVTVHD  1807

Query  246   EEYTDNWLAVYFFTSGNEGNWFEIQTDPRTNEGI--LKVVKALDYE---QLQSVKLSIAV  300
             E+ T+ +        SG   + F +    R N+G   LK+V+ LDYE   Q    +  I V
Sbjct  1808  EDETNKFQYKVIDNSGYGADKFTMV---RNNDGTGSLKIVQPLDYEDPLQSNGFRFRIQV  1864

Query  301   KNKAEFHQSVISRYRVQSTPVTIQVINVREGIAFRPASKTFTVQKGISSKKLVDYILGTY  360
              +K E + +   +Y V  + V +++ ++ +    +P  +   ++  +     V   L T+
Sbjct  1865  NDKGEDNDN--DKYHVAYSWVVVKLRDINDN---KPQFERPNIEVSVYENADVGKTLETF  1919

Query  361   QAIDEDTNKAASNVKYVMGR-NDGGYLMIDSKTAEIKFVKNMNRDSTFIVNKTITAEVLA  419
             +A D D     S V Y + R +D        ++   +   +N++R+    V       ++LA
Sbjct  1920  KATDPDQG-GKSKVSYAIDRSSDRQRQFSINQEGTVTIQRNLDRE----VTPRHQVKILA  1974

Query  420   IDE-YTGKTSTGTVYYRVPDFNDNCP  444
             ID+    KT+T T+ V V D NDN P
Sbjct  1975  IDDGIPPKTATATLTVIVQDINDNPP  2000

 Score = 77.4 bits (189),  Expect = 6e-13, Method: Compositional matrix adjust.
 Identities = 103/419 (25%), Positives = 173/419 (41%), Gaps = 51/419 (12%)

Query  50    IFVVDKNTGDINITAIVDREETPSFLITCRAL----------NAQGLDVEKPLILTVKIL  99
              F ++  +G+I    + DRE+  ++ +   A            N Q   V K   + + I
Sbjct  1385  FFEINLQSGEIFTKVVFDREKQGAYALEVEARDGAPSARPNSNQQPNSVTK--FIRIGIA  1442

Query  100   DINDNPPVFSQQIFMGEIEENSASNSLVMILNATDADEPNHLNSKIAFKIVSQEPAGTPM  159
             D NDNPP F + ++  E++EN       V+ + A D DE    +S+I ++I S    G
Sbjct  1443  DKNDNPPYFDKALYEAEVDENEDIQHTVLTVTAKDHDE----SSRIRYEITSGNIGGA--  1496

Query  160   FLLSRNTGEVRTLTNSLDREQASSYRLVVSGADKDGEGLSTQCECNIKVKDVNDNFPMFR  219
             F +   TG +  +  +LD E    Y L +  +D   E    +    I VKDVND  P F
Sbjct  1497  FAVKNMTGAI-YVAGALDYETRKRYHLTLVASDSLNE---NETIIEIHVKDVNDMPPTFS  1552

Query  220   DSQY--SARIEENILSSELLRFQVTDLDEEYTDNWLAVYFFTSG--NEGNWFEIQTDPRT  275
                Y  +   E+ +    +++   TD D++   N   VYF T    +  N    + D
Sbjct  1553  QQVYETTQDEEQTVTDRPIMQVTATDGDKDRPQN--IVYFLTGQGIDPDNPANSKFDINR  1610

Query  276   NEGILKVVKALDYEQLQS-VKLSIAVKNKAEFHQSVISRYRVQSTPVTIQVINVREGIAF  334
               G + V+K LD +Q      +     V  + E  + ++    VQ     + + ++ +
Sbjct  1611  TTGEIFVLKPLDRDQPNGRPQWRFTVFAQDEGGEGLVGYADVQ-----VNLKDINDNAPI  1665

Query  335   RPASKTFTVQKGISSKKLVDYILGTYQAIDEDTNKAASNVK--YVMGRN------DGGYL  386
              P    F     ++        ++ T  A+D D     +N K  Y + +N
Sbjct  1666  FPQGVYF---GNVTENGTAGMVVMTMTAVDYDDPNEGTNAKLIYSIEKNVIEEETGSPIF  1722

Query  387   MIDSKTAEIK-FVKNMNRDSTFIVNKTITAEVLAIDEYTGKTSTGTVYYRVPDFNDNCP  444
              I++ T  IK  V  ++R+ T        + +V+A+D   G   TGT  +RV D ND  P
Sbjct  1723  EIEADTGVIKTAVCCLDRERT----PDYSIQVVAMDG-GGLKGTGTASIRVKDINDMPP  1776

 Score = 73.9 bits (180),  Expect = 7e-12, Method: Compositional matrix adjust.
 Identities = 90/362 (25%), Positives = 166/362 (46%), Gaps = 37/362 (10%)

Query  51    FVVDKNTGDINITAIVDRE-ETPSFL-ITCRALNAQGLDVEKPLILTVKILDINDNPPVF  108
             F VD+ TG+++    + DRE +   F+ +T +A +     +E      TV+I D+NDNPP+F
Sbjct  1054  FTVDEETGEVSTNKVFDREGDDGKFVSVTVKATDQGEPSLEGVCSFTVEITDVNDNPPLF  1113

Query  109   SQQIFMGEIEENSASNSLVMILNATDADEPNHLNSKIAFKIVS-QEPAGTPMFLLSRNTG  167
              +Q ++  ++++++  + ++ ++A+D D  N  N  I + + +   P     F +    +G
Sbjct  1114  DRQKYVENVKQDASIGTNILRVSASDEDADN--NGAIVYTLSAPHSPNDLEYFEIQPESG  1171

Query  168   EVRTLTNSLDREQASSYRLVVSGADKDGEGLSTQCECNIKVKDVNDNFPMFRDSQYSARI  227
```

```
                 +  L   LDRE   +Y+L     DK    LS   E  I V D +N P++ ++     I
Sbjct  1172  WI-VLKKPLDRE---TYKLEAMAQDKGYPPLSRTVEVQIDVVDRANNPPVWDNTGVYGPI  1227

Query  228   --EENI-LSSELLRFQVTDLDEEYTDNWLAVYFFTSG-----NEGNWFEIQTDPRTNE--  277
               +EN+ +  +++  + +   E    N    Y    G     N+ + F +Q  P   +
Sbjct  1228  YVKENMPVGGKVVSIKASSGIE---GNPTVFYRLMPGSTAQTNKLHTFYLQQRPDNGDTW  1284

Query  278   GILKVVKALDYEQLQSVKLSIAVKNKAEFHQSVISRYRVQSTPVTIQVINVREGIAFRPA  337
               +KV   LD+E ++   L+I V+N         ++       V IQ+ +V + I
Sbjct  1285  ADIKVNHPLDFETIKEYNLTIRVENNG-------AQQLASEATVYIQLEDVNDEIPLFTE  1337

Query  338   SKTFTVQKGISSKKLVDYILGTYQAIDEDTNKAASNVKYVM---GRNDG-GYLMIDSKTA  393
              +  TV +G      V  +     AID+D     + V Y +    RN+G  +  I+ ++
Sbjct  1338  REQETVLEGEPIGTKVTQV----NAIDKDGTFPNNQVYYYIVDSPRNEGKDFFEINLQSG  1393

Query  394   EI  395
             EI
Sbjct  1394  EI  1395

 Score = 55.8 bits (133),  Expect = 3e-06, Method: Compositional matrix adjust.
 Identities = 49/158 (31%), Positives = 78/158 (49%), Gaps = 24/158 (15%)

Query  102   NDNPPVFSQQIFMGEIEENSASNSLVMILNATDADEPNHLNSKIAFKIVSQEPAGTP--M  159
             ND  P FSQQ++   ++EN+  N+LV  + A+D D  N     + F  V     GT
Sbjct  887   NDEEPKFSQQVYTPNVDENAGPNTLVTTVVASDKDGDN-----VRFGFVG---GGTSSGQ  938

Query  160   FLLSRNTGEVRTLTN--SLDREQASSYRLVVSGADK-------DGEGLSTQCECNIKVKD  210
             F++   TG +R  +   SLDR++   Y L V+  D        D    ++     + + D
Sbjct  939   FVIEEITGVIRLHSKAISLDRDK---YELNVTAMDDGSCCVNGDATIHTSTAVVVVFITD  995

Query  211   VNDNFPMFRD-SQYSARIEENILS-SELLRFQVTDLDE  246
             VNDN P+F+D   Y  ++EE   + S +++ Q TD D+
Sbjct  996   VNDNKPVFKDCGTYYPKVEEGAPNGSPVIKVQATDEDK  1033

 Score = 53.5 bits (127),  Expect = 1e-05, Method: Compositional matrix adjust.
 Identities = 70/306 (23%), Positives = 126/306 (41%), Gaps = 30/306 (10%)

Query  20    PIAKIT-SDYQATQKITYRI---SGVGIDQPPFGIFVVDKNTGDINITAIVDRE---ETP  72
             PI  +T  D   T K  Y++   SG G D+  F +   +  TG + I   +D E   ++
Sbjct  1799  PILTVTVHDEDETNKFQYKVIDNSGYGADK--FTMVRNNDGTGSLKIVQPLDYEDPLQSN  1856

Query  73    SFLITCRALNAQGLDVEKPLI------LTVKILDINDNPPVFSQQIFMGEIEENSASNSL  126
              F    + +N +G D +          + VK+ DINDN P F +      + EN+
Sbjct  1857  GFRFRIQ-VNDKGEDNDNDKYHVAYSWVVVKLRDINDNKPQFERPNIEVSVYENADVGKT  1915

Query  127   VMILNATDADEPNHLNSKIAFKIVSQEPAGTPMFLLSRNTGEVRTLTNSLDREQASSYRL  186
             +    ATD D+     SK+++ I   + +       S N     T+  +LDRE    +++
Sbjct  1916  LETFKATDPDQGG--KSKVSYAI---DRSSDRQRQFSINQEGTVTIQRNLDREVTPRHQV  1970

Query  187   VVSGADKDGEGLSTQCECNIKVKDVNDNFPMFRDSQYSARIEENILSSELLRFQVTDLDE  246
              +   D      +      + V+D+NDN P F    Y   + E++   +++    TD D+
Sbjct  1971  KILAIDDGIPPKTATATLTVIVQDINDNPPKFL-KDYRPVLPEHVPPRKVVEILATDDDD  2029

Query  247   EYTDNWLAVYFFTSGNEGNW----FEIQTDPRTNEG----ILKVVKALDYEQLQSVKLSI  298
                 N     F       +     F+++ D +   G    I+  +++ D EQ +   + I
Sbjct  2030  RSKSNGPPFQFRLDPGADDIIRASFKVEQDQKGANGDGMAIVSSLRSFDREQQKEYLIPI  2089

Query  299   AVKNKA  304
              +K+
Sbjct  2090  VIKDHG  2095

 Score = 52.4 bits (124),  Expect = 3e-05, Method: Compositional matrix adjust.
 Identities = 97/398 (24%), Positives = 163/398 (41%), Gaps = 54/398 (14%)

Query  106   PVFSQQIFMGEIEENSASNSLVMILNATDADEPNHLNSKIAFKIVSQ-EPAGTPMFLLSR  164
             P F    +  EI EN   +S ++ + A      +  + +I + + +Q + AGT  F +
Sbjct  684   PQFYMPSYEAEIPENQKKDSDIISVKAK-----SFADREIRYTLKAQGQGAGT--FNIGP  736
```

```
Query  165   NTGEVRTLTNSLDRE---QASSYRLVVSGADKDGEGLSTQCECNIKVKDVNDNFPMFRDS  221
              +G V+ L   LD E   Q   Y LVV+ A +D  G ST  E  I+V DVNDN P F
Sbjct  737   TSGIVK-LAKELDFEDLRQPHVYSLVVT-ATEDSGGFSTSVELTIRVTDVNDNAPKFELP  794

Query  222   QYSAR-IEENI-LSSELLRFQVTDLDEEYTDNWLAVYFFTSGNEGNWFEIQTDPRTNEGI  279
              Y A  ++E+I L + +L+ +  D D         + +  S +    F + ++     GI
Sbjct  795   DYQAHNVDEDIPLGTSILKVKAMDADSGAN---AEIEYLVSDDH---FAVDSN-----GI  843

Query  280   LKVVKALDYEQLQS-VKLSIAVKNKAEFHQSVISRYRVQSTPVTIQVINVREGIAFRPAS  338
             +   K LD +   +  +  +  K+K E  ++  +  RV +           E   F
Sbjct  844   IVNNKQLDADNNNAYYEFVVTAKDKGEPAKTGTATVRVYTKNKN------DEEPKFSQQV  897

Query  339   KTFTVQKGISSKKLVDYILGTYQAIDEDTNKAASNVK--YVMGRNDGGYLMIDSKTAEIK  396
              T  V +      LV  ++         ++K   NV+  +V G    G  +I+  T  I+
Sbjct  898   YTPNVDENAGPNTLVTTVVA--------SDKDGDNVRFGFVGGGTSSGQFVIEEITGVIR  949

Query  397   FVK---NMNRDSTFI-VNKTITAEVLAIDEYTGKTSTGTVYVRVPDFNDN------CPTA  446
                   +++RD   + V            + T  TST  V V + D NDN      C T
Sbjct  950   LHSKAISLDRDKYELNVTAMDDGSCCVNGDATIHTSTAVVVVFITDVNDNKPVFKDCGTY  1009

Query  447   VLE-KDAVCSSSPSVVVSARTLNNRYTGPYTFALEDQP  483
               + ++   + SP + V A   +    G   +++  QP
Sbjct  1010  YPKVEEGAPNGSPVIKVQATDEDKGVNGQVKYSIVQQP  1047

 Score = 44.3 bits (103),  Expect = 0.009, Method: Compositional matrix adjust.
 Identities = 48/199 (24%), Positives = 73/199 (37%), Gaps = 19/199 (10%)

Query  27    DYQATQKITYRISGVGIDQPPFGIFVVDKNTGDINITAIVDREETPSFLITCRALNAQGL  86
             D     K++Y I      Q  F I       G + I   +DRE TP   +   A++
Sbjct  1925  DQGGKSKVSYAIDRSSDRQRQFSI----NQEGTVTIQRNLDREVTPRHQVKILAIDDGIP  1980

Query  87    DVEKPLILTVKILDINDNPPVFSQQIFMGEIEENSASNSLVMILNATDADEPNHLNSKIA  146
                    LTV + DINDNPP F +  +   + E+     +V IL   D D
Sbjct  1981  PKTATATLTVIVQDINDNPPKFLKD-YRPVLPEHVPPRKVVEILATDDDDRSKSNGPPFQ  2039

Query  147   FKIVSQEPAGTPMFLLSRNTGEVRTLTN-----------SLDREQASSYRLVVSGADKDG  195
             F++   +P    +   S    + +   N           S DREQ   Y + +   D
Sbjct  2040  FRL---DPGADDIIRASFKVEQDQKGANGDGMAIVSSLRSFDREQQKEYLIPIVIKDHGN  2096

Query  196   EGLSTQCECNIKVKDVNDN  214
               ++      + + DVNDN
Sbjct  2097  PAMTGTSTLTVVIGDVNDN  2115

 Score = 40.8 bits (94),  Expect = 0.12, Method: Compositional matrix adjust.
 Identities = 54/235 (23%), Positives = 99/235 (42%), Gaps = 22/235 (9%)

Query  4    KFAKPCREGE--DNSKRNP-IAKITSDYQATQKITYRISGVGIDQPPFGIFVVDKNTGDI  60
            +F  P  E E  +N K++  I  + +   A ++I Y +   G      G F +   +G +
Sbjct  685  QFYMPSYEAEIPENQKKDSDIISVKAKSFADREIRYTLKAQGQGA---GTFNIGPTSGIV  741

Query  61   NITAIVDREE--TPSF--LITCRALNAQGLDVEKPLILTVKILDINDNPPVFSQQIFMGE  116
             +   +D E+   P    L+     ++ G      + LT+++ D+NDN P F    +
Sbjct  742  KLAKELDFEDLRQPHVYSLVVTATEDSGGFSTS--VELTIRVTDVNDNAPKFELPDYQAH  799

Query  117  -IEENSASNSLVMILNATDADEPNHLNSKIAFKIVSQEPAGTPMFLLSRNTGEVRTLTNS  175
             ++E+    + ++ + A DAD  +  N++I + +VS +      F +  N   V
Sbjct  800  NVDEDIPLGTSILKVKAMDAD--SGANAEIEY-LVSDD-----HFAVDSNGIIVNNKQLD  851

Query  176  LDREQASSYRLVVSGADKDGEGLSTQCECNIKVKDVNDNFPMFRDSQYSARIEEN  230
             D   A  Y  VV+  DK     +      +  K+ ND  P F    Y+  ++EN
Sbjct  852  ADNNNA-YYEFVVTAKDKGEPAKTGTATVRVYTKNKNDEEPKFSQQVYTPNVDEN  905

 Score = 39.3 bits (90),  Expect = 0.35, Method: Compositional matrix adjust.
 Identities = 89/389 (23%), Positives = 149/389 (38%), Gaps = 62/389 (16%)

Query  75   LITCRALNAQGLDVEKPLILTVKILDINDNPPVFSQQIF--MGEIEENSASNSLVMILNA  132
            +IT +  NA GL       + + + D+ND PP F  +       ++ N+  N+ V  L A
Sbjct  546  IITNQGHNA-GLKYTDNQRVIIHVKDVNDEPPYFINRPLPMQAVVQLNAPPNTPVFTLQA  604
```

```
Query  133  TDADEPNHLNSKIAFKIVSQEPAGTPMFLLSRNTGEVRTLTNSLDREQASSYRLVVSGAD  192
             D D      +  I + IV      G    F +    +G VRT     L  +      Y L V    D
Sbjct  605  RDPD----TDHNIHYFIVRDRTGG--RFEVDERSGVVRTRGTDL-FQLDMEYVLYVKAED  657

Query  193  KDGE----GLSTQCECNIKVKDVNDNFPMFRDSQYSARIEEN-ILSSELLRFQVTDLDEE  247
            ++G+         +  E  + +         P F     Y A I EN     S+++  +      +
Sbjct  658  QNGKVDDRRFQSTPEERLSIVG-GKRAPQFYMPSYEAEIPENQKKDSDIISVKA----KS  712

Query  248  YTDNWLAVYFFTSGNEGNWFEIQTDPRTNEGILKVVKALDYEQLQSVKLS----IAVKNK  303
            + D  +         G      F I          GI+K+ K LD+E L+    +       A ++
Sbjct  713  FADREIRYTLKAQGQGAGTFNIGP----TSGIVKLAKELDFEDLRQPHVYSLVVTATEDS  768

Query  304  AEFHQSVISRYRVQSTPVTIQVINVREGIAFRPASKTFTVQKGISSKKLVDYILGT----  359
              F  SV           +TI+V +V +         +  F +     +      D  LGT
Sbjct  769  GGFSTSV---------ELTIRVTDVNDN------APKFELPDYQAHNVDEDIPLGTSILK  813

Query  360  YQAIDEDTNKAASNVKYVMGRN----DGGYLMIDSKTAEIKFVKNMNRDSTFIVNKTITA  415
             +A+D D+  A + ++Y++  +     D    +++++K  +      N N     F+V
Sbjct  814  VKAMDADSG-ANAEIEYLVSDDHFAVDSNGIIVNNKQLD---ADNNNAYYEFVVTAKDKG  869

Query  416  EVLAIDEYTGKTSTGTVYVRVPDFNDNCP  444
            E          KT T TV V    + ND  P
Sbjct  870  E-------PAKTGTATVRVYTKNKNDEEP  891

>AALB20_033171.P66510 | transcript=AALB20_033171.R66510 | gene=AALB20_033171
| organism=Anopheles_albimanus_STECLA_2020 |
gene_product=neural-cadherin, transcript variant X38 |
transcript_product=neural-cadherin,
transcript variant X29 | location=CM024292:75037560-75208393(+)
| protein_length=3125 | sequence_SO=chromosome
| SO=protein_coding_gene | is_pseudo=false
Length=3125

 Score = 147 bits (370),  Expect = 3e-35, Method: Compositional matrix adjust.
 Identities = 128/446 (29%), Positives = 217/446 (49%), Gaps = 36/446 (8%)

Query  20    PIAKITS---DYQATQKITYRISGVGID--QPPFGIFVVDKNTGDINITAIVDREET---  71
             PI ++T+    D     Q I Y ++G GID    P     F +++ TG+I +    +DR++
Sbjct  1566  PIMQVTATDGDKDRPQNIVYFLTGQGIDPDNPANSKFDINRTTGEIFVLKPLDRDQPNGR  1625

Query  72    PSFLITCRALNAQGLDVEKPLILTVKILDINDNPPVFSQQIFMGEIEENSASNSLVMILN  131
             P +   T  A +  G  +      + V + DINDN P+F Q ++ G + EN  +  +VM +
Sbjct  1626  PQWRFTVFAQDEGGEGLVGYADVQVNLKDINDNAPIFPQGVYFGNVTENGTAGMVVMTMT  1685

Query  132   ATDADEPNH-LNSKIAFKI---VSQEPAGTPMFLLSRNTGEVRTLTNSLDREQASSYRLV  187
             A D D+PN    N+K+ + I    V +E  G+P+F +  +TG ++T     LDRE+    Y +
Sbjct  1686  AVDYDDPNEGTNAKLIYSIEKNVIEEETGSPIFEIEADTGVIKTAVCCLDRERTPDYSIQ  1745

Query  188   VSGADKDGEGLSTQCECNIKVKDVNDNFPMFRDSQYSARIEENILSS--ELLRFQVTDLD  245
             V      DG GL       +I+VKD+ND  P F    ++    ++E    ++  E+     VT  D
Sbjct  1746  VVAM--DGGGLKGTGTASIRVKDINDMPPQFTKDEWFTEVDETDGTNLPEMPILTVTVHD  1803

Query  246   EEYTDNWLAVYFFTSGNEGNWFEIQTDPRTNEGI--LKVVKALDYE---QLQSVKLSIAV  300
             E+ T+ +         SG    + F +     R N+G    LK+V+ LDYE    Q     +  I V
Sbjct  1804  EDETNKFQYKVIDNSGYGADKFTMV---RNNDGTGSLKIVQPLDYEDPLQSNGFRFRIQV  1860

Query  301   KNKAEFHQSVISRYRVQSTPVTIQVINVREGIAFRPASKTFTVQKGISSKKLVDYILGTY  360
              +K E + +    +Y V   + V +++ ++ +     +P   +    ++   +       V    L T+
Sbjct  1861  NDKGEDNDN--DKYHVAYSWVVVKLRDINDN---KPQFERPNIEVSVYENADVGKTLETF  1915

Query  361   QAIDEDTNKAASNVKYVMGR-NDGGYLMIDSKTAEIKFVKNMNRDSTFIVNKTITAEVLA  419
             +A D D      S V Y + R +D         ++    +    +N++R+     V        ++LA
Sbjct  1916  KATDPDQG-GKSKVSYAIDRSSDRQRQFSINQEGTVTIQRNLDRE----VTPRHQVKILA  1970

Query  420   IDE-YTGKTSTGTVYVRVPDFNDNCP  444
             ID+     KT+T T+ V V D NDN P
Sbjct  1971  IDDGIPPKTATATLTVIVQDINDNPP  1996
```

```
 Score = 79.3 bits (194),  Expect = 2e-13, Method: Compositional matrix adjust.
 Identities = 104/419 (25%), Positives = 173/419 (41%), Gaps = 51/419 (12%)

Query  50    IFVVDKNTGDINITAIVDREETPSFLITCRAL----------NAQGLDVEKPLILTVKIL  99
              F ++  +G+I    + DRE+  ++ +   A           N Q   V K   + + I
Sbjct  1381  FFEINLQSGEIFTKVVFDREKQGAYALEVEARDGAPSARPNSNQQPNSVTK--FIRIGIA  1438

Query  100   DINDNPPVFSQQIFMGEIEENSASNSLVMILNATDADEPNHLNSKIAFKIVSQEPAGTPM  159
             D NDNPP F + ++  E++EN      V+ + A D DE    +S+I ++I S    G
Sbjct  1439  DKNDNPPYFDKALYEAEVDENEDIQHTVLTVTAKDHDE----SSRIRYEITSGNIGGA--  1492

Query  160   FLLSRNTGEVRTLTNSLDREQASSYRLVVSGADKDGEGLSTQCECNIKVKDVNDNFPMFR  219
             F +   TG +  +  +LD E    Y L +  +D   E  +      I VKDVND  P F
Sbjct  1493  FAVKNMTGAI-YVAGALDYETRKRYHLTLVASDSLNENATI---IEIHVKDVNDMPPTFS  1548

Query  220   DSQYSARI--EENILSSELLRFQVTDLDEEYTDNWLAVYFFTSG--NEGNWFEIQTDPRT  275
                Y A    E+ +    +++   TD D++   N   VYF T    +  N    + D
Sbjct  1549  QQIYEATQDEEQTVTDRPIMQVTATDGDKDRPQN--IVYFLTGQGIDPDNPANSKFDINR  1606

Query  276   NEGILKVVKALDYEQLQS-VKLSIAVKNKAEFHQSVISRYRVQSTPVTIQVINVREGIAF  334
               G + V+K LD +Q      +    V  + E  + ++    VQ     + + ++ +
Sbjct  1607  TTGEIFVLKPLDRDQPNGRPQWRFTVFAQDEGGEGLVGYADVQ-----VNLKDINDNAPI  1661

Query  335   RPASKTFTVQKGISSKKLVDYILGTYQAIDEDTNKAASNVK--YVMGRN------DGGYL  386
              P    F     ++       ++ T  A+D D     +N K  Y + +N
Sbjct  1662  FPQGVYF---GNVTENGTAGMVVMTMTAVDYDDPNEGTNAKLIYSIEKNVIEEETGSPIF  1718

Query  387   MIDSKTAEIK-FVKNMNRDSTFIVNKTITAEVLAIDEYTGKTSTGTVYVRVPDFNDNCP  444
              I++ T  IK  V  ++R+ T       + +V+A+D   G   TGT  +RV D ND  P
Sbjct  1719  EIEADTGVIKTAVCCLDRERT----PDYSIQVVAMDG-GGLKGTGTASIRVKDINDMPP  1772

 Score = 77.4 bits (189),  Expect = 6e-13, Method: Compositional matrix adjust.
 Identities = 107/426 (25%), Positives = 179/426 (42%), Gaps = 51/426 (12%)

Query  53    VDKNTGDINITAIVDREETPSFLITCRALNAQGLDVEKPLILTVKILDINDNPPVFSQ-Q  111
             +D++  ++N+TA+ D         +C  +N          ++ V I D+NDN PVF
Sbjct  950   LDRDKYELNVTAMDDG--------SC-CVNGDATIHTSTAVVVVFITDVNDNKPVFKDCG  1000

Query  112   IFMGEIEENSASNSLVMILNATDADEPNHLNSKIAFKIVSQEPAGTPMFLLSRNTGEVRT  171
              +  ++EE + + S V+ + ATD D+   +N ++ + IV Q       F +   TGEV T
Sbjct  1001  TYYPKVEEGAPNGSPVIKVQATDEDK--GVNGQVKYSIVQQPNQKGTKFTVDEETGEVST  1058

Query  172   LTNSLDRE--QASSYRLVVSGADKDGEGLSTQCECNIKVKDVNDNFPMFRDSQYSARIEE  229
                  DRE        + V   D+    L   C   +++ DVNDN P+F   +Y   +++
Sbjct  1059  -NKVFDREGDDGKFVSVTVKATDQGEPSLEGVCSFTVEITDVNDNPPLFDRQKYVENVKQ  1117

Query  230   NI-LSSELLRFQVTDLDEEYTDNWLAVYFFT---SGNEGNWFEIQTDPRTNEGILKVVKA  285
             +  + + +LR  V+  DE+  +N   VY  +   S N+  +FEIQ  P +   +LK  K
Sbjct  1118  DASIGTNILR--VSASDEDADNNGAIVYTLSAPHSPNDLEYFEIQ--PESGWIVLK--KP  1171

Query  286   LD--YEQLQSVKLSIAVKNK------AEFHQSVISRYRVQSTPV----TIQVINVREGIA  333
             LD  Y + +  +L +   ++      A+    +    R    P+        I++RE +
Sbjct  1172  LDGCYSR-ERYRLKVYASDRGDPPSTADVDVELDVVDRNNKPPIWDRNIYGPIHIRENVT  1230

Query  334   FRPASKTFTVQKGISSKKLVDYILGTYQAIDEDTNKAASNVKYVMGRNDGGYLMIDSKTA  393
                         GI     V Y L         TNK   +  Y+  R D G        A
Sbjct  1231  VGTVVTAIKASSGIEGNPTVFYRL--MPGSTAQTNKL--HTFYLQQRPDNG-----DTWA  1281

Query  394   EIKFVKNMNRDSTFIVNKTITAEVLAIDEYTGKTSTGTVYVRVPDFNDNCPTAV-LEKDA  452
             +IK    ++ ++    N TI  E     +     S  TVY+++ D ND  P     E++
Sbjct  1282  DIKVNHPLDFETIKEYNLTIRVENNGAQQL---ASEATVYIQLEDVNDEIPLFTEREQET  1338

Query  453   VCSSSP  458
             V    P
Sbjct  1339  VLEGEP  1344

 Score = 63.2 bits (152),  Expect = 1e-08, Method: Compositional matrix adjust.
 Identities = 86/364 (24%), Positives = 156/364 (43%), Gaps = 38/364 (10%)
```

```
Query  51    FVVDKNTGDINITAIVDRE-ETPSFL-ITCRALNAQGLDVEKPLILTVKILDINDNPPVF  108
             F VD+ TG+++   + DRE +   F+ +T +A +     +E     TV+I D+NDNPP+F
Sbjct  1047  FTVDEETGEVSTNKVFDREGDDGKFVSVTVKATDQGEPSLEGVCSFTVEITDVNDNPPLF  1106

Query  109   SQQIFMGEIEENSASNSLVMILNATDADEPNHLNSKIAFKIVS-QEPAGTPMFLLSRNTG  167
              +Q ++  ++++++  + ++ ++A+D D  N  N  I + + +   P     F +   +G
Sbjct  1107  DRQKYVENVKQDASIGTNILRVSASDEDADN--NGAIVYTLSAPHSPNDLEYFEIQPESG  1164

Query  168   EV---RTLTNSLDREQASSYRLVVSGADKDGEGLSTQCECNIKVKDVNDNFPMFRDSQYS  224
              +   + L     RE+   YRL V  +D+ G+  ST            +N P   D
Sbjct  1165  WIVLKKPLDGCYSRER---YRLKVYASDR-GDPPSTADVDVELDVVDRNNKPPIWDRNIY  1220

Query  225   A--RIEENILSSELLRFQVTDLDEEYTDNWLAVYFFTSG-----NEGNWFEIQTDPRTNE  277
                 I EN+    ++             N    Y    G     N+ + F +Q  P   +
Sbjct  1221  GPIHIRENVTVGTVV--TAIKASSGIEGNPTVFYRLMPGSTAQTNKLHTFYLQQRPDNGD  1278

Query  278   --GILKVVKALDYEQLQSVKLSIAVKNKAEFHQSVISRYRVQSTPVTIQVINVREGIAFR  335
                 +KV   LD+E ++   L+I V+N         ++       V IQ+ +V + I
Sbjct  1279  TWADIKVNHPLDFETIKEYNLTIRVENNG-------AQQLASEATVYIQLEDVNDEIPLF  1331

Query  336   PASKTFTVQKGISSKKLVDYILGTYQAIDEDTNKAASNVKYVM---GRNDG-GYLMIDSK  391
                +  TV +G      V  +     AID+D     + V Y +    RN+G  +  I+ +
Sbjct  1332  TEREQETVLEGEPIGTKVTQV----NAIDKDGTFPNNQVYYYIVDSPRNEGKDFFEINLQ  1387

Query  392   TAEI  395
             + EI
Sbjct  1388  SGEI  1391

 Score = 55.8 bits (133),  Expect = 3e-06, Method: Compositional matrix adjust.
 Identities = 49/158 (31%), Positives = 78/158 (49%), Gaps = 24/158 (15%)

Query  102   NDNPPVFSQQIFMGEIEENSASNSLVMILNATDADEPNHLNSKIAFKIVSQEPAGTP--M  159
             ND  P FSQQ++   ++EN+  N+LV  + A+D D  N     + F  V     GT
Sbjct  880   NDEEPKFSQQVYTPNVDENAGPNTLVTTVVASDKDGDN-----VRFGFVG---GGTSSGQ  931

Query  160   FLLSRNTGEVRTLTN--SLDREQASSYRLVVSGADK-------DGEGLSTQCECNIKVKD  210
             F++   TG +R  +   SLDR++   Y L V+  D        D    ++     + + D
Sbjct  932   FVIEEITGVIRLHSKAISLDRDK---YELNVTAMDDGSCCVNGDATIHTSTAVVVVFITD  988

Query  211   VNDNFPMFRD-SQYSARIEENILS-SELLRFQVTDLDE  246
             VNDN P+F+D   Y  ++EE   + S +++ Q TD D+
Sbjct  989   VNDNKPVFKDCGTYYPKVEEGAPNGSPVIKVQATDEDK  1026

 Score = 53.5 bits (127),  Expect = 1e-05, Method: Compositional matrix adjust.
 Identities = 70/306 (23%), Positives = 126/306 (41%), Gaps = 30/306 (10%)

Query  20    PIAKIT-SDYQATQKITYRI---SGVGIDQPPFGIFVVDKNTGDINITAIVDRE---ETP  72
             PI  +T  D   T K  Y++   SG G D+  F +   +  TG + I   +D E   ++
Sbjct  1795  PILTVTVHDEDETNKFQYKVIDNSGYGADK--FTMVRNNDGTGSLKIVQPLDYEDPLQSN  1852

Query  73    SFLITCRALNAQGLDVEKPLI------LTVKILDINDNPPVFSQQIFMGEIEENSASNSL  126
              F    + +N +G D +          + VK+ DINDN P F +      + EN+
Sbjct  1853  GFRFRIQ-VNDKGEDNDNDKYHVAYSWVVVKLRDINDNKPQFERPNIEVSVYENADVGKT  1911

Query  127   VMILNATDADEPNHLNSKIAFKIVSQEPAGTPMFLLSRNTGEVRTLTNSLDREQASSYRL  186
             +    ATD D+     SK+++ I   + +       S N     T+  +LDRE    +++
Sbjct  1912  LETFKATDPDQGG--KSKVSYAI---DRSSDRQRQFSINQEGTVTIQRNLDREVTPRHQV  1966

Query  187   VVSGADKDGEGLSTQCECNIKVKDVNDNFPMFRDSQYSARIEENILSSELLRFQVTDLDE  246
              +   D      +      + V+D+NDN P F    Y   + E++   +++     TD D+
Sbjct  1967  KILAIDDGIPPKTATATLTVIVQDINDNPPKFL-KDYRPVLPEHVPPRKVVEILATDDDD  2025

Query  247   EYTDNWLAVYFFTSGNEGNW----FEIQTDPRTNEG----ILKVVKALDYEQLQSVKLSI  298
                 N     F       +     F+++ D +   G    I+  +++ D EQ +   + I
Sbjct  2026  RSKSNGPPFQFRLDPGADDIIRASFKVEQDQKGANGDGMAIVSSLRSFDREQQKEYLIPI  2085

Query  299   AVKNKA  304
              +K+
Sbjct  2086  VIKDHG  2091
```

```
 Score = 51.2 bits (121),  Expect = 6e-05, Method: Compositional matrix adjust.
 Identities = 96/398 (24%), Positives = 163/398 (41%), Gaps = 54/398 (14%)

Query  106   PVFSQQIFMGEIEENSASNSLVMILNATDADEPNHLNSKIAFKIVSQ-EPAGTPMFLLSR  164
             P F    +  EI EN   +S ++ + A       +  + +I + + +Q + AGT  F +
Sbjct  677   PQFYMPSYEAEIPENQKKDSDIISVKAK-----SFADREIRYTLKAQGQGAGT--FNIGP  729

Query  165   NTGEVRTLTNSLDRE---QASSYRLVVSGADKDGEGLSTQCECNIKVKDVNDNFPMFRDS  221
              +G V+ L   LD E   Q   Y LVV+ A +D  G ST  E  I+V DVNDN P F
Sbjct  730   TSGIVK-LAKELDFEDLRQPHVYSLVVT-ATEDSGGFSTSVELTIRVTDVNDNAPKFELP  787

Query  222   QYSAR-IEENI-LSSELLRFQVTDLDEEYTDNWLAVYFFTSGNEGNWFEIQTDPRTNEGI  279
              Y A  ++E+I + + +L+ +  D D          + +  S +    F + ++     GI
Sbjct  788   DYQAHNVDEDIQIGTSILKVKAMDADSGAN---AEIEYLVSDDH---FAVDSN-----GI  836

Query  280   LKVVKALDYEQLQSV-KLSIAVKNKAEFHQSVISRYRVQSTPVTIQVINVREGIAFRPAS  338
             +   K LD +   +  +  +  K+K E  ++  +  RV +            E   F
Sbjct  837   IVNNKQLDADNNNAYYEFVVTAKDKGEPAKTGTATVRVYTKNKN------DEEPKFSQQV  890

Query  339   KTFTVQKGISSKKLVDYILGTYQAIDEDTNKAASNVKY--VMGRNDGGYLMIDSKTAEIK  396
              T  V +      LV  ++ +          +K   NV++  V G    G  +I+  T  I+
Sbjct  891   YTPNVDENAGPNTLVTTVVAS--------DKDGDNVRFGFVGGGTSSGQFVIEEITGVIR  942

Query  397   FVK---NMNRDSTFI-VNKTITAEVLAIDEYTGKTSTGTVYVRVPDFNDN------CPTA  446
                  +++RD   + V              + T  TST  V V + D NDN      C T
Sbjct  943   LHSKAISLDRDKYELNVTAMDDGSCCVNGDATIHTSTAVVVVFITDVNDNKPVFKDCGTY  1002

Query  447   VLE-KDAVCSSSPSVVVSARTLNNRYTGPYTFALEDQP  483
               + ++   + SP + V A   +    G   +++  QP
Sbjct  1003  YPKVEEGAPNGSPVIKVQATDEDKGVNGQVKYSIVQQP  1040

 Score = 44.3 bits (103),  Expect = 0.009, Method: Compositional matrix adjust.
 Identities = 48/199 (24%), Positives = 73/199 (37%), Gaps = 19/199 (10%)

Query  27    DYQATQKITYRISGVGIDQPPFGIFVVDKNTGDINITAIVDREETPSFLITCRALNAQGL  86
             D     K++Y I      Q  F I       G + I   +DRE TP   +   A++
Sbjct  1921  DQGGKSKVSYAIDRSSDRQRQFSI----NQEGTVTIQRNLDREVTPRHQVKILAIDDGIP  1976

Query  87    DVEKPLILTVKILDINDNPPVFSQQIFMGEIEENSASNSLVMILNATDADEPNHLNSKIA  146
                    LTV + DINDNPP F +  +   + E+     +V IL   D D
Sbjct  1977  PKTATATLTVIVQDINDNPPKFLKD-YRPVLPEHVPPRKVVEILATDDDDRSKSNGPPFQ  2035

Query  147   FKIVSQEPAGTPMFLLSRNTGEVRTLTN-----------SLDREQASSYRLVVSGADKDG  195
             F++    +P    +   S    + +   N           S DREQ   Y + +   D
Sbjct  2036  FRL---DPGADDIIRASFKVEQDQKGANGDGMAIVSSLRSFDREQQKEYLIPIVIKDHGN  2092

Query  196   EGLSTQCECNIKVKDVNDN  214
               ++      + + DVNDN
Sbjct  2093  PAMTGTSTLTVVIGDVNDN  2111

 Score = 40.4 bits (93),  Expect = 0.13, Method: Compositional matrix adjust.
 Identities = 54/235 (23%), Positives = 99/235 (42%), Gaps = 22/235 (9%)

Query  4    KFAKPCREGE--DNSKRNP-IAKITSDYQATQKITYRISGVGIDQPPFGIFVVDKNTGDI  60
            +F  P  E E  +N K++  I  + +   A ++I Y +   G      G F +   +G +
Sbjct  678  QFYMPSYEAEIPENQKKDSDIISVKAKSFADREIRYTLKAQGQGA---GTFNIGPTSGIV  734

Query  61   NITAIVDREE--TPSF--LITCRALNAQGLDVEKPLILTVKILDINDNPPVFSQQIFMGE  116
             +   +D E+   P    L+      ++ G       + LT+++ D+NDN P F    +
Sbjct  735  KLAKELDFEDLRQPHVYSLVVTATEDSGGFSTS--VELTIRVTDVNDNAPKFELPDYQAH  792

Query  117  -IEENSASNSLVMILNATDADEPNHLNSKIAFKIVSQEPAGTPMFLLSRNTGEVRTLTNS  175
             ++E+     + ++ + A DAD   +  N++I + +VS +       F +  N   V
Sbjct  793  NVDEDIQIGTSILKVKAMDAD--SGANAEIEY-LVSDD-----HFAVDSNGIIVNNKQLD  844

Query  176  LDREQASSYRLVVSGADKDGEGLSTQCECNIKVKDVNDNFPMFRDSQYSARIEEN  230
             D   A  Y  VV+  DK      +       + K+ ND  P F    Y+  ++EN
```

```
Sbjct  845  ADNNNA-YYEFVVTAKDKGEPAKTGTATVRVYTKNKNDEEPKFSQQVYTPNVDEN  898

 Score = 39.3 bits (90),  Expect = 0.35, Method: Compositional matrix adjust.
 Identities = 55/225 (24%), Positives = 88/225 (39%), Gaps = 24/225 (11%)

Query  75   LITCRALNAQGLDVEKPLILTVKILDINDNPPVFSQQIF--MGEIEENSASNSLVMILNA  132
            +IT +  NA GL       + + + D+ND PP F  +       ++ N+  N+ V  L A
Sbjct  539  IITNQGHNA-GLKYTDNQRVIIHVKDVNDEPPYFINRPLPMQAVVQLNAPPNTPVFTLQA  597

Query  133  TDADEPNHLNSKIAFKIVSQEPAGTPMFLLSRNTGEVRTLTNSLDREQASSYRLVVSGAD  192
             D D     +  I + IV     G   F +   +G VRT    L  +    Y L V   D
Sbjct  598  RDPD----TDHNIHYFIVRDRTGG--RFEVDERSGVVRTRGTDL-FQLDMEYVLYVKAED  650

Query  193  KDGE----GLSTQCECNIKVKDVNDNFPMFRDSQYSARIEEN-ILSSELLRFQVTDLDEE  247
            ++G+       +  E  + +       P F    Y A I EN    S+++  +     +
Sbjct  651  QNGKVDDRRFQSTPEERLSIVG-GKRAPQFYMPSYEAEIPENQKKDSDIISVKA----KS  705

Query  248  YTDNWLAVYFFTSGNEGNWFEIQTDPRTNEGILKVVKALDYEQLQ  292
            + D  +       G     F I        GI+K+ K LD+E L+
Sbjct  706  FADREIRYTLKAQGQGAGTFNIG----PTSGIVKLAKELDFEDLR  746

>AMAC_003346.P3003 | transcript=AMAC_003346.R3003 | gene=AMAC_003346
| organism=Anopheles_maculipalpis_AmacGA1 | gene_product=neural-cadherin,
transcript variant X12 | transcript_product=neural-cadherin,
transcript variant X9 | location=AmacGA1_3RL:17033394-17218514(+)
| protein_length=3107 | sequence_SO=chromosome
| SO=protein_coding_gene | is_pseudo=false
Length=3107

 Score = 147 bits (370),  Expect = 3e-35, Method: Compositional matrix adjust.
 Identities = 128/446 (29%), Positives = 217/446 (49%), Gaps = 36/446 (8%)

Query  20    PIAKITS---DYQATQKITYRISGVGID--QPPFGIFVVDKNTGDINITAIVDREET---  71
             PI ++T+   D    Q I Y ++G GID   P    F +++ TG+I +   +DR++
Sbjct  1547  PIMQVTATDGDKDRPQNIVYFLTGQGIDPDNPANSKFDINRTTGEIFVLKPLDRDQPNGR  1606

Query  72    PSFLITCRALNAQGLDVEKPLILTVKILDINDNPPVFSQQIFMGEIEENSASNSLVMILN  131
             P +  T  A +  G  +     + V + DINDN P+F Q ++ G + EN  +  +VM +
Sbjct  1607  PQWRFTVFAQDEGGEGLVGYADVQVNLKDINDNAPIFPQGVYFGNVTENGTAGMVVMTMT  1666

Query  132   ATDADEPNH-LNSKIAFKI---VSQEPAGTPMFLLSRNTGEVRTLTNSLDREQASSYRLV  187
             A D D+PN   N+K+ + I   V +E  G+P+F +  +TG ++T    LDRE+   Y +
Sbjct  1667  AVDYDDPNEGTNAKLIYSIEKNVIEEETGSPIFEIEADTGVIKTAVCCLDRERTPDYSIQ  1726

Query  188   VSGADKDGEGLSTQCECNIKVKDVNDNFPMFRDSQYSARIEENILSS--ELLRFQVTDLD  245
             V     DG GL      +I+VKD+ND  P F   ++   ++E   ++  E+    VT  D
Sbjct  1727  VVAM--DGGGLKGTGTASIRVKDINDMPPQFTKDEWFTEVDETDGTNLPEMPILTVTVHD  1784

Query  246   EEYTDNWLAVYFFTSGNEGNWFEIQTDPRTNEGI--LKVVKALDYE---QLQSVKLSIAV  300
             E+ T+ +        SG    + F +    R N+G   LK+V+ LDYE   Q    +  I V
Sbjct  1785  EDETNKFQYKVIDNSGYGADKFTMV---RNNDGTGSLKIVQPLDYEDPLQSNGFRFRIQV  1841

Query  301   KNKAEFHQSVISRYRVQSTPVTIQVINVREGIAFRPASKTFTVQKGISSKKLVDYILGTY  360
              +K E + +   +Y V  + V +++ ++ +    +P  +   ++  +      V   L T+
Sbjct  1842  NDKGEDNDN--DKYHVAYSWVVVKLRDINDN---KPQFERPNIEVSVYENADVGKTLETF  1896

Query  361   QAIDEDTNKAASNVKYVMGR-NDGGYLMIDSKTAEIKFVKNMNRDSTFIVNKTITAEVLA  419
             +A D D     S V Y + R +D        ++   +   +N++R+    V      ++LA
Sbjct  1897  KATDPDQG-GKSKVSYAIDRSSDRQRQFSINQEGTVTIQRNLDRE----VTPRHQVKILA  1951

Query  420   IDE-YTGKTSTGTVYYVRVPDFNDNCP  444
             ID+    KT+T T+ V V D NDN P
Sbjct  1952  IDDGIPPKTATATLTVIVQDINDNPP  1977

 Score = 76.6 bits (187),  Expect = 1e-12, Method: Compositional matrix adjust.
 Identities = 103/418 (25%), Positives = 174/418 (42%), Gaps = 50/418 (12%)

Query  50    IFVVDKNTGDINITAIVDREETPSFLITCRAL----------NAQGLDVEKPLILTVKIL  99
```

```
              F ++  +G+I    + DRE+  ++ +   A           N Q   V K   + + I
Sbjct  1363  FFEINLQSGEIFTKVVFDREKQGAYALEVEARDGAPSARPNSNQQPNSVTK--FIRIGIA  1420

Query  100   DINDNPPVFSQQIFMGEIEENSASNSLVMILNATDADEPNHLNSKIAFKIVSQEPAGTPM  159
             D NDNPP F + ++  E++EN      V+ + A D DE    +S+I ++I S    G
Sbjct  1421  DKNDNPPYFDKALYEAEVDENEDIQHTVLTVTAKDHDE----SSRIRYEITSGNIGGA--  1474

Query  160   FLLSRNTGEVRTLTNSLDREQASSYRLVVSGADKDGEGLSTQCECNIKVKDVNDNFPMFR  219
             F +   TG + +  +LD E    Y L +  +D   E  +      I VKDVND  P F
Sbjct  1475  FAVKNMTGAI-YVAGALDYETRKRYHLTLVASDSLNENETIIV---INVKDVNDMPPSFP  1530

Query  220   DSQYSARIEENI-LSSELLRFQVTDLDEEYTDNWLAVYFFTSG--NEGNWFEIQTDPRTN  276
                Y   ++E + +   +++   TD D++   N   VYF T    +  N    + D
Sbjct  1531  QQVYERTMDEELAVPFPIMQVTATDGDKDRPQN--IVYFLTGQGIDPDNPANSKFDINRT  1588

Query  277   EGILKVVKALDYEQLQS-VKLSIAVKNKAEFHQSVISRYRVQSTPVTIQVINVREGIAFR  335
              G + V+K LD +Q     +    V  + E  + ++    VQ     + + ++ +
Sbjct  1589  TGEIFVLKPLDRDQPNGRPQWRFTVFAQDEGGEGLVGYADVQ-----VNLKDINDNAPIF  1643

Query  336   PASKTFTVQKGISSKKLVDYILGTYQAIDEDTNKAASNVK--YVMGRN------DGGYLM  387
             P    F     ++       ++ T  A+D D     +N K  Y + +N
Sbjct  1644  PQGVYF---GNVTENGTAGMVVMTMTAVDYDDPNEGTNAKLIYSIEKNVIEEETGSPIFE  1700

Query  388   IDSKTAEIK-FVKNMNRDSTFIVNKTITAEVLAIDEYTGKTSTGTVYVRVPDFNDNCP  444
             I++ T  IK  V  ++R+ T       + +V+A+D   G   TGT  +RV D ND  P
Sbjct  1701  IEADTGVIKTAVCCLDRERT----PDYSIQVVAMDG-GGLKGTGTASIRVKDINDMPP  1753

 Score = 76.3 bits (186),  Expect = 1e-12, Method: Compositional matrix adjust.
 Identities = 89/361 (25%), Positives = 167/361 (46%), Gaps = 36/361 (10%)

Query  51    FVVDKNTGDINITAIVDRE-ETPSFL-ITCRALNAQGLDVEKPLILTVKILDINDNPPVF  108
             F VD+ TG+++   + DRE +   F+ +T +A +     +E     TV+I D+NDNPP+F
Sbjct  1033  FTVDEETGEVSTNKVFDREGDDGKFVSVTVKATDQGEPSLEGVCSFTVEITDVNDNPPLF  1092

Query  109   SQQIFMGEIEENSASNSLVMILNATDADEPNHLNSKIAFKIVS-QEPAGTPMFLLSRNTG  167
              +Q ++  ++++++  + ++ ++A+D D  N  N  I + + +   P     F +   +G
Sbjct  1093  DRQKYVENVKQDASIGTNILRVSASDEDADN--NGAIVYTLSAPHSPNDLEYFEIQPESG  1150

Query  168   EVRTLTNSLDREQASSYRLVVSGADKDGEGLSTQCECNIKVKDVNDNFPMFRDSQYSA-R  226
              +  L   LDRE   +Y+L     DK    LS   E  I V D  +N P++ ++ Y
Sbjct  1151  WI-VLKKPLDRE---TYKLEAMAQDKGYPPLSRTVEVQIDVVDRANNPPVWDNTVYGPIY  1206

Query  227   IEENI-LSSELLRFQVTDLDEEYTDNWLAVYFFTSG-----NEGNWFEIQTDPRTNE--G  278
             ++EN+ +  +++  + +   E    N    Y    G     N+ + F +Q  P   +
Sbjct  1207  VKENMPVGGKVVSIKASSGIE---GNPTVFYRLMPGSTAQTNKLHTFYLQQRPDNGDTWA  1263

Query  279   ILKVVKALDYEQLQSVKLSIAVKNKAEFHQSVISRYRVQSTPVTIQVINVREGIAFRPAS  338
              +KV   LD+E ++   L++ V+N          ++       V IQ+ +V + I
Sbjct  1264  DIKVNHPLDFETIKEYNLTVRVENNG-------AQQLASEATVYIQLEDVNDEIPLFTER  1316

Query  339   KTFTVQKGISSKKLVDYILGTYQAIDEDTNKAASNVKYVM---GRNDG-GYLMIDSKTAE  394
             +  TV +G      V  +     AID+D     + V Y +    RN+G  +  I+ ++ E
Sbjct  1317  EQETVLEGEPIGTKVTQV----NAIDKDGTFPNNQVYYYIVDSPRNEGKDFFEINLQSGE  1372

Query  395   I  395
             I
Sbjct  1373  I  1373

 Score = 55.8 bits (133),  Expect = 3e-06, Method: Compositional matrix adjust.
 Identities = 49/158 (31%), Positives = 78/158 (49%), Gaps = 24/158 (15%)

Query  102   NDNPPVFSQQIFMGEIEENSASNSLVMILNATDADEPNHLNSKIAFKIVSQEPAGTP--M  159
             ND  P FSQQ++   ++EN+  N+LV  + A+D D  N      + F  V     GT
Sbjct  866   NDEEPKFSQQVYTPNVDENAGPNTLVTTVVASDKDGDN-----VRFGFVG---GGTSSGQ  917

Query  160   FLLSRNTGEVRTLTN--SLDREQASSYRLVVSGADK-------DGEGLSTQCECNIKVKD  210
             F++   TG +R  +   SLDR++   Y L V+  D         D    ++      + + D
Sbjct  918   FVIEEITGVIRLHSKAISLDRDK---YELNVTAMDDGSCCVNGDATIHTSTAVVVVFITD  974
```

```
Query  211   VNDNFPMFRD-SQYSARIEENILS-SELLRFQVTDLDE  246
             VNDN P+F+D   Y  ++EE   + S +++ Q TD D+
Sbjct  975   VNDNKPVFKDCGTYYPKVEEGAPNGSPVIKVQATDEDK  1012

 Score = 53.9 bits (128),  Expect = 1e-05, Method: Compositional matrix adjust.
 Identities = 70/306 (23%), Positives = 126/306 (41%), Gaps = 30/306 (10%)

Query  20    PIAKIT-SDYQATQKITYRI---SGVGIDQPPFGIFVVDKNTGDINITAIVDRE---ETP  72
             PI  +T  D   T K  Y++   SG G D+  F +   +  TG + I   +D E   ++
Sbjct  1776  PILTVTVHDEDETNKFQYKVIDNSGYGADK--FTMVRNNDGTGSLKIVQPLDYEDPLQSN  1833

Query  73    SFLITCRALNAQGLDVEKPLI------LTVKILDINDNPPVFSQQIFMGEIEENSASNSL  126
              F    + +N +G D +          + VK+ DINDN P F +      + EN+
Sbjct  1834  GFRFRIQ-VNDKGEDNDNDKYHVAYSWVVVKLRDINDNKPQFERPNIEVSVYENADVGKT  1892

Query  127   VMILNATDADEPNHLNSKIAFKIVSQEPAGTPMFLLSRNTGEVRTLTNSLDREQASSYRL  186
             +    ATD D+     SK+++ I   + +       S N     T+  +LDRE    +++
Sbjct  1893  LETFKATDPDQGG--KSKVSYAI---DRSSDRQRQFSINQEGTVTIQRNLDREVTPRHQV  1947

Query  187   VVSGADKDGEGLSTQCECNIKVKDVNDNFPMFRDSQYSARIEENILSSELLRFQVTDLDE  246
              +   D      +      + V+D+NDN P F    Y   + E++   +++    TD D+
Sbjct  1948  KILAIDDGIPPKTATATLTVIVQDINDNPPKFL-KDYRPVLPEHVPPRKVVEILATDDDD  2006

Query  247   EYTDNWLAVYFFTSGNEGNW----FEIQTDPRTNEG----ILKVVKALDYEQLQSVKLSI  298
                 N     F       +     F+++ D +   G    I+  +++ D EQ +   + I
Sbjct  2007  RSKSNGPPFQFRLDPGADDIIRASFKVEQDQKGANGDGMAIVSSLRSFDREQQKEYLIPI  2066

Query  299   AVKNKA  304
              +K+
Sbjct  2067  VIKDHG  2072

 Score = 52.4 bits (124),  Expect = 3e-05, Method: Compositional matrix adjust.
 Identities = 97/398 (24%), Positives = 163/398 (41%), Gaps = 54/398 (14%)

Query  106   PVFSQQIFMGEIEENSASNSLVMILNATDADEPNHLNSKIAFKIVSQ-EPAGTPMFLLSR  164
             P F    +  EI EN   +S ++ + A      +  + +I + + +Q + AGT  F +
Sbjct  663   PQFYMPSYEAEIPENQKKDSDIISVKAK-----SFADREIRYTLKAQGQGAGT--FNIGP  715

Query  165   NTGEVRTLTNSLDRE---QASSYRLVVSGADKDGEGLSTQCECNIKVKDVNDNFPMFRDS  221
              +G V+ L   LD E   Q   Y LVV+ A +D  G ST  E  I+V DVNDN P F
Sbjct  716   TSGIVK-LAKELDFEDLRQPHVYSLVVT-ATEDSGGFSTSVELTIRVTDVNDNAPKFELP  773

Query  222   QYSAR-IEENI-LSSELLRFQVTDLDEEYTDNWLAVYFFTSGNEGNWFEIQTDPRTNEGI  279
              Y A  ++E+I L + +L+ +  D D         + +  S +    F + ++     GI
Sbjct  774   DYQAHNVDEDIPLGTSILKVKAMDADSGAN---AEIEYLVSDDH---FAVDSN-----GI  822

Query  280   LKVVKALDYEQLQS-VKLSIAVKNKAEFHQSVISRYRVQSTPVTIQVINVREGIAFRPAS  338
             +   K LD +   +  +  +  K+K E  ++  +  RV +           E   F
Sbjct  823   IVNNKQLDADNNNAYYEFVVTAKDKGEPAKTGTATVRVYTKNKN------DEEPKFSQQV  876

Query  339   KTFTVQKGISSKKLVDYILGTYQAIDEDTNKAASNVK--YVMGRNDGGYLMIDSKTAEIK  396
              T  V +      LV  ++         ++K   NV+  +V G    G  +I+  T  I+
Sbjct  877   YTPNVDENAGPNTLVTTVVA--------SDKDGDNVRFGFVGGGTSSGQFVIEEITGVIR  928

Query  397   FVK---NMNRDSTFI-VNKTITAEVLAIDEYTGKTSTGTVYVRVPDFNDN------CPTA  446
                   +++RD   + V            + T  TST  V V + D NDN      C T
Sbjct  929   LHSKAISLDRDKYELNVTAMDDGSCCVNGDATIHTSTAVVVVFITDVNDNKPVFKDCGTY  988

Query  447   VLE-KDAVCSSSPSVVVSARTLNNRYTGPYTFALEDQP  483
               + ++   + SP + V A   +    G   +++  QP
Sbjct  989   YPKVEEGAPNGSPVIKVQATDEDKGVNGQVKYSIVQQP  1026

 Score = 44.3 bits (103),  Expect = 0.009, Method: Compositional matrix adjust.
 Identities = 48/199 (24%), Positives = 73/199 (37%), Gaps = 19/199 (10%)

Query  27    DYQATQKITYRISGVGIDQPPFGIFVVDKNTGDINITAIVDREETPSFLITCRALNAQGL  86
             D     K++Y I      Q  F I       G + I   +DRE TP   +   A++
Sbjct  1902  DQGGKSKVSYAIDRSSDRQRQFSI----NQEGTVTIQRNLDREVTPRHQVKILAIDDGIP  1957
```

```
Query  87    DVEKPLILTVKILDINDNPPVFSQQIFMGEIEENSASNSLVMILNATDADEPNHLNSKIA  146
                    LTV + DINDNPP F +  +   + E+     +V IL   D D
Sbjct  1958  PKTATATLTVIVQDINDNPPKFLKD-YRPVLPEHVPPRKVVEILATDDDDRSKSNGPPFQ  2016

Query  147   FKIVSQEPAGTPMFLLSRNTGEVRTLTN-----------SLDREQASSYRLVVSGADKDG  195
             F++   +P    +   S    + +   N           S DREQ   Y + +   D
Sbjct  2017  FRL---DPGADDIIRASFKVEQDQKGANGDGMAIVSSLRSFDREQQKEYLIPIVIKDHGN  2073

Query  196   EGLSTQCECNIKVKDVNDN  214
               ++      + + DVNDN
Sbjct  2074  PAMTGTSTLTVVIGDVNDN  2092

 Score = 40.8 bits (94),  Expect = 0.12, Method: Compositional matrix adjust.
 Identities = 54/235 (23%), Positives = 99/235 (42%), Gaps = 22/235 (9%)

Query  4    KFAKPCREGE--DNSKRNP-IAKITSDYQATQKITYRISGVGIDQPPFGIFVVDKNTGDI  60
            +F  P  E E  +N K++  I  + +   A ++I Y +   G      G F +   +G +
Sbjct  664  QFYMPSYEAEIPENQKKDSDIISVKAKSFADREIRYTLKAQGQGA---GTFNIGPTSGIV  720

Query  61   NITAIVDREE--TPSF--LITCRALNAQGLDVEKPLILTVKILDINDNPPVFSQQIFMGE  116
              +   +D E+   P    L+      ++ G      + LT+++ D+NDN P F    +
Sbjct  721  KLAKELDFEDLRQPHVYSLVVTATEDSGGFSTS--VELTIRVTDVNDNAPKFELPDYQAH  778

Query  117  -IEENSASNSLVMILNATDADEPNHLNSKIAFKIVSQEPAGTPMFLLSRNTGEVRTLTNS  175
              ++E+    + ++ + A DAD  +  N++I + +VS +      F +  N   V
Sbjct  779  NVDEDIPLGTSILKVKAMDAD--SGANAEIEY-LVSDD-----HFAVDSNGIIVNNKQLD  830

Query  176  LDREQASSYRLVVSGADKDGEGLSTQCECNIKVKDVNDNFPMFRDSQYSARIEEN  230
              D   A  Y  VV+  DK      +       +  K+ ND  P F    Y+  ++EN
Sbjct  831  ADNNNA-YYEFVVTAKDKGEPAKTGTATVRVYTKNKNDEEPKFSQQVYTPNVDEN  884

 Score = 39.3 bits (90),  Expect = 0.36, Method: Compositional matrix adjust.
 Identities = 89/389 (23%), Positives = 149/389 (38%), Gaps = 62/389 (16%)

Query  75   LITCRALNAQGLDVEKPLILTVKILDINDNPPVFSQQIF--MGEIEENSASNSLVMILNA  132
            +IT +  NA GL       + + + D+ND PP F  +        ++ N+  N+ V  L A
Sbjct  525  IITNQGHNA-GLKYTDNQRVIIHVKDVNDEPPYFINRPLPMQAVVQLNAPPNTPVFTLQA  583

Query  133  TDADEPNHLNSKIAFKIVSQEPAGTPMFLLSRNTGEVRTLTNSLDREQASSYRLVVSGAD  192
              D D     +  I + IV     G   F +   +G VRT    L  +    Y L V   D
Sbjct  584  RDPD----TDHNIHYFIVRDRTGG--RFEVDERSGVVRTRGTDL-FQLDMEYVLYVKAED  636

Query  193  KDGE----GLSTQCECNIKVKDVNDNFPMFRDSQYSARIEEN-ILSSELLRFQVTDLDEE  247
            ++G+        +  E  + +        P F    Y A I EN    S+++  +     +
Sbjct  637  QNGKVDDRRFQSTPEERLSIVG-GKRAPQFYMPSYEAEIPENQKKDSDIISVKA----KS  691

Query  248  YTDNWLAVYFFTSGNEGNWFEIQTDPRTNEGILKVVKALDYEQLQSVKLS----IAVKNK  303
            + D  +        G     F I         GI+K+ K LD+E L+   +      A ++
Sbjct  692  FADREIRYTLKAQGQGAGTFNIGP----TSGIVKLAKELDFEDLRQPHVYSLVVTATEDS  747

Query  304  AEFHQSVISRYRVQSTPVTIQVINVREGIAFRPASKTFTVQKGISSKKLVDYILGT----  359
              F  SV           +TI+V +V +       +  F +     +     D  LGT
Sbjct  748  GGFSTSV---------ELTIRVTDVNDN------APKFELPDYQAHNVDEDIPLGTSILK  792

Query  360  YQAIDEDTNKAASNVKYVMGRN----DGGYLMIDSKTAEIKFVKNMNRDSTFIVNKTITA  415
             +A+D D+  A + ++Y++  +    D    +++++K  +     N N    F+V
Sbjct  793  VKAMDADSG-ANAEIEYLVSDDHFAVDSNGIIVNNKQLD---ADNNNAYYEFVVTAKDKG  848

Query  416  EVLAIDEYTGKTSTGTVYYVRVPDFNDNCP  444
            E         KT T TV V   + ND  P
Sbjct  849  E-------PAKTGTATVRVYTKNKNDEEP  870
```

>ASTEI20_040830.P55068 | transcript=ASTEI20_040830.R55068 | gene=ASTEI20_040830 | organism=Anopheles_stephensi_UCISS2018 | gene_product=neural-cadherin, transcript variant X5 | transcript_product=neural-cadherin, transcript variant X14 | location=CM023249:57079885-57269506(-)

```
| protein_length=3108 | sequence_SO=chromosome
| SO=protein_coding_gene | is_pseudo=false
Length=3108

 Score = 147 bits (370),  Expect = 3e-35, Method: Compositional matrix adjust.
 Identities = 128/446 (29%), Positives = 217/446 (49%), Gaps = 36/446 (8%)

Query  20    PIAKITS---DYQATQKITYRISGVGID--QPPFGIFVVDKNTGDINITAIVDREET---  71
             PI ++T+   D    Q I Y ++G GID   P    F +++ TG+I +   +DR++
Sbjct  1548  PIMQVTATDGDKDRPQNIVYFLTGQGIDPDNPANSKFDINRTTGEIFVLKPLDRDQPNGR  1607

Query  72    PSFLITCRALNAQGLDVEKPLILTVKILDINDNPPVFSQQIFMGEIEENSASNSLVMILN  131
             P +  T  A +  G  +     + V + DINDN P+F Q ++ G + EN  +  +VM +
Sbjct  1608  PQWRFTVFAQDEGGEGLVGYADVQVNLKDINDNAPIFPQGVYFGNVTENGTAGMVVMTMT  1667

Query  132   ATDADEPNH-LNSKIAFKI---VSQEPAGTPMFLLSRNTGEVRTLTNSLDREQASSYRLV  187
             A D D+PN   N+K+ + I   V +E  G+P+F +  +TG ++T    LDRE+   Y +
Sbjct  1668  AVDYDDPNEGTNAKLIYSIEKNVIEEETGSPIFEIEADTGVIKTAVCCLDRERTPDYSIQ  1727

Query  188   VSGADKDGEGLSTQCECNIKVKDVNDNFPMFRDSQYSARIEENILSS--ELLRFQVTDLD  245
             V     DG GL      +I+VKD+ND  P F   ++   ++E   ++  E+    VT  D
Sbjct  1728  VVAM--DGGGLKGTGTASIRVKDINDMPPQFTKDEWFTEVDETDGTNLPEMPILTVTVHD  1785

Query  246   EEYTDNWLAVYFFTSGNEGNWFEIQTDPRTNEGI--LKVVKALDYE---QLQSVKLSIAV  300
             E+ T+ +       SG   + F +    R N+G   LK+V+ LDYE   Q    +  I V
Sbjct  1786  EDETNKFQYKVIDNSGYGADKFTMV---RNNDGTGSLKIVQPLDYEDPLQSNGFRFRIQV  1842

Query  301   KNKAEFHQSVISRYRVQSTPVTIQVINVREGIAFRPASKTFTVQKGISSKKLVDYILGTY  360
              +K E + +   +Y V  + V +++ ++ +    +P  +   ++  +     V   L T+
Sbjct  1843  NDKGEDNDN--DKYHVAYSWVVVKLRDINDN---KPQFERPNIEVSVYENADVGKTLETF  1897

Query  361   QAIDEDTNKAASNVKYVMGR-NDGGYLMIDSKTAEIKFVKNMNRDSTFIVNKTITAEVLA  419
             +A D D     S V Y + R +D       ++   +   +N++R+    V      ++LA
Sbjct  1898  KATDPDQG-GKSKVSYAIDRSSDRQRQFSINQEGTVTIQRNLDRE----VTPRHQVKILA  1952

Query  420   IDE-YTGKTSTGTVYVRVPDFNDNCP  444
             ID+    KT+T T+ V V D NDN P
Sbjct  1953  IDDGIPPKTATATLTVIVQDINDNPP  1978

 Score = 76.6 bits (187),  Expect = 1e-12, Method: Compositional matrix adjust.
 Identities = 103/418 (25%), Positives = 174/418 (42%), Gaps = 50/418 (12%)

Query  50    IFVVDKNTGDINITAIVDREETPSFLITCRAL----------NAQGLDVEKPLILTVKIL  99
              F ++  +G+I    + DRE+  ++ +   A          N Q   V K   + + I
Sbjct  1364  FFEINLQSGEIFTKVVFDREKQGAYALEVEARDGAPSARPNSNQQPNSVTK--FIRIGIA  1421

Query  100   DINDNPPVFSQQIFMGEIEENSASNSLVMILNATDADEPNHLNSKIAFKIVSQEPAGTPM  159
             D NDNPP F + ++  E++EN      V+ + A D DE    +S+I ++I S    G
Sbjct  1422  DKNDNPPYFDKALYEAEVDENEDIQHTVLTVTAKDHDE----SSRIRYEITSGNIGGA--  1475

Query  160   FLLSRNTGEVRTLTNSLDREQASSYRLVVSGADKDGEGLSTQCECNIKVKDVNDNFPMFR  219
             F +   TG +  +  +LD E    Y L +  +D   E  +      I VKDVND  P F
Sbjct  1476  FAVKNMTGAI-YVAGALDYETRKRYHLTLVASDSLNENETIIV---INVKDVNDMPPSFP  1531

Query  220   DSQYSARIEENI-LSSELLRFQVTDLDEEYTDNWLAVYFFTSG--NEGNWFEIQTDPRTN  276
                Y   ++E + +   +++   TD D++   N   VYF T    +  N    + D
Sbjct  1532  QQVYERTMDEELAVPFPIMQVTATDGDKDRPQN--IVYFLTGQGIDPDNPANSKFDINRT  1589

Query  277   EGILKVVKALDYEQLQS-VKLSIAVKNKAEFHQSVISRYRVQSTPVTIQVINVREGIAFR  335
              G + V+K LD +Q     +    V  + E  + ++    VQ     + + ++ +
Sbjct  1590  TGEIFVLKPLDRDQPNGRPQWRFTVFAQDEGGEGLVGYADVQ-----VNLKDINDNAPIF  1644

Query  336   PASKTFTVQKGISSKKLVDYILGTYQAIDEDTNKAASNVK--YVMGRN------DGGYLM  387
             P    F     ++       ++ T  A+D D     +N K  Y + +N
Sbjct  1645  PQGVYF---GNVTENGTAGMVVMTMTAVDYDDPNEGTNAKLIYSIEKNVIEEETGSPIFE  1701

Query  388   IDSKTAEIK-FVKNMNRDSTFIVNKTITAEVLAIDEYTGKTSTGTVYVRVPDFNDNCP  444
             I++ T  IK  V  ++R+ T       + +V+A+D   G   TGT  +RV D ND  P
Sbjct  1702  IEADTGVIKTAVCCLDRERT----PDYSIQVVAMDG-GGLKGTGTASIRVKDINDMPP  1754
```

```
 Score = 76.3 bits (186),  Expect = 1e-12, Method: Compositional matrix adjust.
 Identities = 89/361 (25%), Positives = 167/361 (46%), Gaps = 36/361 (10%)

Query  51    FVVDKNTGDINITAIVDRE-ETPSFL-ITCRALNAQGLDVEKPLILTVKILDINDNPPVF  108
             F VD+ TG+++   + DRE +   F+ +T +A +     +E     TV+I D+NDNPP+F
Sbjct  1034  FTVDEETGEVSTNKVFDREGDDGKFVSVTVKATDQGEPSLEGVCSFTVEITDVNDNPPLF  1093

Query  109   SQQIFMGEIEENSASNSLVMILNATDADEPNHLNSKIAFKIVS-QEPAGTPMFLLSRNTG  167
              +Q ++  ++++++  + ++ ++A+D D  N  N  I + + +   P     F +   +G
Sbjct  1094  DRQKYVENVKQDASIGTNILRVSASDEDADN--NGAIVYTLSAPHSPNDLEYFEIQPESG  1151

Query  168   EVRTLTNSLDREQASSYRLVVSGADKDGEGLSTQCECNIKVKDVNDNFPMFRDSQYSA-R  226
              +  L   LDRE   +Y+L     DK    LS   E  I V D  +N P++ ++ Y
Sbjct  1152  WI-VLKKPLDRE---TYKLEAMAQDKGYPPLSRTVEVQIDVVDRANNPPVWDNTVYGPIY  1207

Query  227   IEENI-LSSELLRFQVTDLDEEYTDNWLAVYFFTSG-----NEGNWFEIQTDPRTNE--G  278
             ++EN+ +  +++  + +   E    N    Y    G     N+ + F +Q  P   +
Sbjct  1208  VKENMPVGGKVVSIKASSGIE---GNPTVFYRLMPGSTAQTNKLHTFYLQQRPDNGDTWA  1264

Query  279   ILKVVKALDYEQLQSVKLSIAVKNKAEFHQSVISRYRVQSTPVTIQVINVREGIAFRPAS  338
              +KV   LD+E ++   L++ V+N         ++       V IQ+ +V + I
Sbjct  1265  DIKVNHPLDFETIKEYNLTVRVENNG-------AQQLASEATVYIQLEDVNDEIPLFTER  1317

Query  339   KTFTVQKGISSKKLVDYILGTYQAIDEDTNKAASNVKYVM---GRNDG-GYLMIDSKTAE  394
             +  TV +G      V  +     AID+D     + V Y +    RN+G  +  I+ ++ E
Sbjct  1318  EQETVLEGEPIGTKVTQV----NAIDKDGTFPNNQVYYYIVDSPRNEGKDFFEINLQSGE  1373

Query  395   I  395
             I
Sbjct  1374  I  1374

 Score = 55.8 bits (133),  Expect = 3e-06, Method: Compositional matrix adjust.
 Identities = 49/158 (31%), Positives = 78/158 (49%), Gaps = 24/158 (15%)

Query  102   NDNPPVFSQQIFMGEIEENSASNSLVMILNATDADEPNHLNSKIAFKIVSQEPAGTP--M  159
             ND  P FSQQ++   ++EN+  N+LV  + A+D D  N     + F  V     GT
Sbjct  867   NDEEPKFSQQVYTPNVDENAGPNTLVTTVVASDKDGDN-----VRFGFVG---GGTSSGQ  918

Query  160   FLLSRNTGEVRTLTN--SLDREQASSYRLVVSGADK-------DGEGLSTQCECNIKVKD  210
             F++   TG +R  +   SLDR++   Y L V+  D        D    ++     + + D
Sbjct  919   FVIEEITGVIRLHSKAISLDRDK---YELNVTAMDDGSCCVNGDATIHTSTAVVVVFITD  975

Query  211   VNDNFPMFRD-SQYSARIEENILS-SELLRFQVTDLDE  246
             VNDN P+F+D   Y  ++EE   + S +++ Q TD D+
Sbjct  976   VNDNKPVFKDCGTYYPKVEEGAPNGSPVIKVQATDEDK  1013

 Score = 53.5 bits (127),  Expect = 1e-05, Method: Compositional matrix adjust.
 Identities = 70/306 (23%), Positives = 126/306 (41%), Gaps = 30/306 (10%)

Query  20    PIAKIT-SDYQATQKITYRI---SGVGIDQPPFGIFVVDKNTGDINITAIVDRE---ETP  72
             PI  +T  D   T K  Y++   SG G D+  F +   +  TG + I   +D E   ++
Sbjct  1777  PILTVTVHDEDETNKFQYKVIDNSGYGADK--FTMVRNNDGTGSLKIVQPLDYEDPLQSN  1834

Query  73    SFLITCRALNAQGLDVEKPLI------LTVKILDINDNPPVFSQQIFMGEIEENSASNSL  126
              F    + +N +G D +          + VK+ DINDN P F +      + EN+
Sbjct  1835  GFRFRIQ-VNDKGEDNDNDKYHVAYSWVVVKLRDINDNKPQFERPNIEVSVYENADVGKT  1893

Query  127   VMILNATDADEPNHLNSKIAFKIVSQEPAGTPMFLLSRNTGEVRTLTNSLDREQASSYRL  186
             +    ATD D+     SK+++ I   + +       S N     T+  +LDRE    +++
Sbjct  1894  LETFKATDPDQGG--KSKVSYAI---DRSSDRQRQFSINQEGTVTIQRNLDREVTPRHQV  1948

Query  187   VVSGADKDGEGLSTQCECNIKVKDVNDNFPMFRDSQYSARIEENILSSELLRFQVTDLDE  246
              +   D      +      + V+D+NDN P F    Y   + E++   +++    TD D+
Sbjct  1949  KILAIDDGIPPKTATATLTVIVQDINDNPPKFL-KDYRPVLPEHVPPRKVVEILATDDDD  2007

Query  247   EYTDNWLAVYFFTSGNEGNW----FEIQTDPRTNEG----ILKVVKALDYEQLQSVKLSI  298
                 N     F       +     F+++ D +   G    I+  +++ D EQ +   + I
Sbjct  2008  RSKSNGPPFQFRLDPGADDIIRASFKVEQDQKGANGDGMAIVSSLRSFDREQQKEYLIPI  2067
```

```
Query  299   AVKNKA  304
              +K+
Sbjct  2068  VIKDHG  2073

 Score = 52.4 bits (124),  Expect = 3e-05, Method: Compositional matrix adjust.
 Identities = 97/398 (24%), Positives = 163/398 (41%), Gaps = 54/398 (14%)

Query  106   PVFSQQIFMGEIEENSASNSLVMILNATDADEPNHLNSKIAFKIVSQ-EPAGTPMFLLSR  164
             P F    +  EI EN   +S ++ + A      +  + +I + + +Q + AGT  F +
Sbjct  664   PQFYMPSYEAEIPENQKKDSDIISVKAK-----SFADREIRYTLKAQGQGAGT--FNIGP  716

Query  165   NTGEVRTLTNSLDRE---QASSYRLVVSGADKDGEGLSTQCECNIKVKDVNDNFPMFRDS  221
              +G V+ L   LD E   Q   Y LVV+ A +D  G ST  E  I+V DVNDN P F
Sbjct  717   TSGIVK-LAKELDFEDLRQPHVYSLVVT-ATEDSGGFSTSVELTIRVTDVNDNAPKFELP  774

Query  222   QYSAR-IEENI-LSSELLRFQVTDLDEEYTDNWLAVYFFTSGNEGNWFEIQTDPRTNEGI  279
              Y A  ++E+I L + +L+ +  D D          + +  S +    F + ++     GI
Sbjct  775   DYQAHNVDEDIPLGTSILKVKAMDADSGAN---AEIEYLVSDDH---FAVDSN-----GI  823

Query  280   LKVVKALDYEQLQS-VKLSIAVKNKAEFHQSVISRYRVQSTPVTIQVINVREGIAFRPAS  338
             +   K LD +   +  +  +  K+K E  ++  +  RV +            E   F
Sbjct  824   IVNNKQLDADNNNAYYEFVVTAKDKGEPAKTGTATVRVYTKNKN------DEEPKFSQQV  877

Query  339   KTFTVQKGISSKKLVDYILGTYQAIDEDTNKAASNVK--YVMGRNDGGYLMIDSKTAEIK  396
              T  V +      LV  ++          ++K   NV+  +V G    G  +I+  T  I+
Sbjct  878   YTPNVDENAGPNTLVTTVVA--------SDKDGDNVRFGFVGGGTSSGQFVIEEITGVIR  929

Query  397   FVK---NMNRDSTFI-VNKTITAEVLAIDEYTGKTSTGTVYVRVPDFNDN------CPTA  446
                   +++RD   + V             + T  TST  V V + D NDN      C T
Sbjct  930   LHSKAISLDRDKYELNVTAMDDGSCCVNGDATIHTSTAVVVVFITDVNDNKPVFKDCGTY  989

Query  447   VLE-KDAVCSSSPSVVVSARTLNNRYTGPYTFALEDQP  483
               + ++   + SP + V A   +    G   +++  QP
Sbjct  990   YPKVEEGAPNGSPVIKVQATDEDKGVNGQVKYSIVQQP  1027

 Score = 44.3 bits (103),  Expect = 0.009, Method: Compositional matrix adjust.
 Identities = 48/199 (24%), Positives = 73/199 (37%), Gaps = 19/199 (10%)

Query  27    DYQATQKITYRISGVGIDQPPFGIFVVDKNTGDINITAIVDREETPSFLITCRALNAQGL  86
             D     K++Y I      Q  F I       G + I   +DRE TP   +   A++
Sbjct  1903  DQGGKSKVSYAIDRSSDRQRQFSI----NQEGTVTIQRNLDREVTPRHQVKILAIDDGIP  1958

Query  87    DVEKPLILTVKILDINDNPPVFSQQIFMGEIEENSASNSLVMILNATDADEPNHLNSKIA  146
                     LTV + DINDNPP F +  +   + E+     +V IL   D D
Sbjct  1959  PKTATATLTVIVQDINDNPPKFLKD-YRPVLPEHVPPRKVVEILATDDDDRSKSNGPPFQ  2017

Query  147   FKIVSQEPAGTPMFLLSRNTGEVRTLTN-----------SLDREQASSYRLVVSGADKDG  195
             F++   +P    +   S    + +   N           S DREQ   Y + +   D
Sbjct  2018  FRL---DPGADDIIRASFKVEQDQKGANGDGMAIVSSLRSFDREQQKEYLIPIVIKDHGN  2074

Query  196   EGLSTQCECNIKVKDVNDN  214
              ++      + + DVNDN
Sbjct  2075  PAMTGTSTLTVVIGDVNDN  2093

 Score = 40.8 bits (94),  Expect = 0.13, Method: Compositional matrix adjust.
 Identities = 55/242 (23%), Positives = 102/242 (42%), Gaps = 22/242 (9%)

Query  4    KFAKPCREGE--DNSKRNP-IAKITSDYQATQKITYRISGVGIDQPPFGIFVVDKNTGDI  60
            +F  P  E E  +N K++  I  + +   A ++I Y +   G       G F +   +G +
Sbjct  665  QFYMPSYEAEIPENQKKDSDIISVKAKSFADREIRYTLKAQGQGA---GTFNIGPTSGIV  721

Query  61   NITAIVDREE--TPSF--LITCRALNAQGLDVEKPLILTVKILDINDNPPVFSQQIFMGE  116
              +   +D E+   P    L+      ++ G       + LT+++ D+NDN P F    +
Sbjct  722  KLAKELDFEDLRQPHVYSLVVTATEDSGGFSTS--VELTIRVTDVNDNAPKFELPDYQAH  779

Query  117  -IEENSASNSLVMILNATDADEPNHLNSKIAFKIVSQEPAGTPMFLLSRNTGEVRTLTNS  175
             ++E+    + ++ + A DAD   +  N++I + +VS +       F +  N   V
```

```
Sbjct  780  NVDEDIPLGTSILKVKAMDAD--SGANAEIEY-LVSDD-----HFAVDSNGIIVNNKQLD  831

Query  176  LDREQASSYRLVVSGADKDGEGLSTQCECNIKVKDVNDNFPMFRDSQYSARIEENILSSE  235
             D   A  Y  VV+  DK     +      +  K+ ND  P F    Y+  ++EN   +
Sbjct  832  ADNNNA-YYEFVVTAKDKGEPAKTGTATVRVYTKNKNDEEPKFSQQVYTPNVDENAGPNT  890

Query  236  LL  237
            L+
Sbjct  891  LV  892

 Score = 39.3 bits (90),  Expect = 0.36, Method: Compositional matrix adjust.
 Identities = 89/389 (23%), Positives = 149/389 (38%), Gaps = 62/389 (16%)

Query  75   LITCRALNAQGLDVEKPLILTVKILDINDNPPVFSQQIF--MGEIEENSASNSLVMILNA  132
            +IT +  NA GL       + + + D+ND PP F  +       ++ N+  N+ V  L A
Sbjct  526  IITNQGHNA-GLKYTDNQRVIIHVKDVNDEPPYFINRPLPMQAVVQLNAPPNTPVFTLQA  584

Query  133  TDADEPNHLNSKIAFKIVSQEPAGTPMFLLSRNTGEVRTLTNSLDREQASSYRLVVSGAD  192
             D D     +  I + IV     G   F +   +G VRT    L  +    Y L V   D
Sbjct  585  RDPD----TDHNIHYFIVRDRTGG--RFEVDERSGVVRTRGTDL-FQLDMEYVLYVKAED  637

Query  193  KDGE----GLSTQCECNIKVKDVNDNFPMFRDSQYSARIEEN-ILSSELLRFQVTDLDEE  247
            ++G+       +  E  + +       P F    Y A I EN    S+++  +     +
Sbjct  638  QNGKVDDRRFQSTPEERLSIVG-GKRAPQFYMPSYEAEIPENQKKDSDIISVKA----KS  692

Query  248  YTDNWLAVYFFTSGNEGNWFEIQTDPRTNEGILKVVKALDYEQLQSVKLS----IAVKNK  303
            + D  +       G     F I        GI+K+ K LD+E L+   +      A ++
Sbjct  693  FADREIRYTLKAQGQGAGTFNIGP----TSGIVKLAKELDFEDLRQPHVYSLVVTATEDS  748

Query  304  AEFHQSVISRYRVQSTPVTIQVINVREGIAFRPASKTFTVQKGISSKKLVDYILGT----  359
              F  SV          +TI+V +V +       +  F +    +     D  LGT
Sbjct  749  GGFSTSV---------ELTIRVTDVNDN------APKFELPDYQAHNVDEDIPLGTSILK  793

Query  360  YQAIDEDTNKAASNVKYVMGRN----DGGYLMIDSKTAEIKFVKNMNRDSTFIVNKTITA  415
             +A+D D+  A + ++Y++  +    D   +++++K  +     N N    F+V
Sbjct  794  VKAMDADSG-ANAEIEYLVSDDHFAVDSNGIIVNNKQLD---ADNNNAYYEFVVTAKDKG  849

Query  416  EVLAIDEYTGKTSTGTVYVRVPDFNDNCP  444
            E         KT T TV V   + ND  P
Sbjct  850  E-------PAKTGTATVRVYTKNKNDEEP  871

>AFUN2_006054.P9879 | transcript=AFUN2_006054.R9879 | gene=AFUN2_006054
| organism=Anopheles_funestus_AfunGA1 | gene_product=neural-cadherin,
transcript variant X17, neural-cadherin,
transcript variant X22, neural-cadherin, transcript variant
X12, neural-cadherin, transcript variant X1, neural-cadherin,
transcript variant X15, neural-cadherin, transcript variant
X2, neural-cadherin, transcript variant X23, neural-cadherin,
transcript variant X26, neural-cadherin, transcript variant
X24, neural-cadherin, transcript variant X7, neural-cadherin,
transcript variant X6, neural-cadherin, transcript variant
X18, neural-cadherin, transcript variant X3, neural-cadherin,
transcript variant X9, neural-cadherin, transcript variant
X14, neural-cadherin, transcript variant X4, neural-cadherin,
transcript variant X25, neural-cadherin, transcript
variant X21, neural-cadherin, transcript variant X20, neural-cadherin,
transcript variant X19, neural-cadherin, transcript
variant X13, neural-cadherin, transcript variant X11, neural-cadherin,
transcript variant X16, neural-cadherin, transcript
variant X5, neural-cadherin, transcript variant X10,
neural-cadherin, transcript variant X8 | transcript_product=neural-cadherin,
transcript variant X21 | location=AfunGA1_2RL:82372181-82559663(-)
| protein_length=3109 | sequence_SO=chromosome
| SO=protein_coding_gene | is_pseudo=false
Length=3109

 Score = 147 bits (370),  Expect = 3e-35, Method: Compositional matrix adjust.
 Identities = 128/446 (29%), Positives = 217/446 (49%), Gaps = 36/446 (8%)
```

```
Query  20    PIAKITS---DYQATQKITYRISGVGID--QPPFGIFVVDKNTGDINITAIVDREET---  71
             PI ++T+   D    Q I Y ++G GID   P    F +++ TG+I +   +DR++
Sbjct  1549  PIMQVTATDGDKDRPQNIVYFLTGQGIDPDNPANSKFDINRTTGEIFVLKPLDRDQPNGR  1608

Query  72    PSFLITCRALNAQGLDVEKPLILTVKILDINDNPPVFSQQIFMGEIEENSASNSLVMILN  131
             P +  T  A +  G  +     + V + DINDN P+F Q ++ G + EN  +  +VM +
Sbjct  1609  PQWRFTVFAQDEGGEGLVGYADVQVNLKDINDNAPIFPQGVYFGNVTENGTAGMVVMTMT  1668

Query  132   ATDADEPNH-LNSKIAFKI---VSQEPAGTPMFLLSRNTGEVRTLTNSLDREQASSYRLV  187
             A D D+PN   N+K+ + I   V +E  G+P+F +  +TG ++T    LDRE+   Y +
Sbjct  1669  AVDYDDPNEGTNAKLIYSIEKNVIEEETGSPIFEIEADTGVIKTAVCCLDRERTPDYSIQ  1728

Query  188   VSGADKDGEGLSTQCECNIKVKDVNDNFPMFRDSQYSARIEENILSS--ELLRFQVTDLD  245
             V     DG GL      +I+VKD+ND  P F   ++   ++E   ++  E+    VT  D
Sbjct  1729  VVAM--DGGGLKGTGTASIRVKDINDMPPQFTKDEWFTEVDETDGTNLPEMPILTVTVHD  1786

Query  246   EEYTDNWLAVYFFTSGNEGNWFEIQTDPRTNEGI--LKVVKALDYE---QLQSVKLSIAV  300
             E+ T+ +        SG   + F +    R N+G   LK+V+ LDYE   Q    +  I V
Sbjct  1787  EDETNKFQYKVIDNSGYGADKFTMV---RNNDGTGSLKIVQPLDYEDPLQSNGFRFRIQV  1843

Query  301   KNKAEFHQSVISRYRVQSTPVTIQVINVREGIAFRPASKTFTVQKGISSKKLVDYILGTY  360
              +K E + +   +Y V  + V +++ ++ +    +P  +   ++  +     V   L T+
Sbjct  1844  NDKGEDNDN--DKYHVAYSWVVVKLRDINDN---KPQFERPNIEVSVYENADVGKTLETF  1898

Query  361   QAIDEDTNKAASNVKYVMGR-NDGGYLMIDSKTAEIKFVKNMNRDSTFIVNKTITAEVLA  419
             +A D D     S V Y + R +D       ++   +   +N++R+    V      ++LA
Sbjct  1899  KATDPDQG-GKSKVSYAIDRSSDRQRQFSINQEGTVTIQRNLDRE----VTPRHQVKILA  1953

Query  420   IDE-YTGKTSTGTVYYVRVPDFNDNCP  444
             ID+    KT+T T+ V V D NDN P
Sbjct  1954  IDDGIPPKTATATLTVIVQDINDNPP  1979

 Score = 78.2 bits (191),  Expect = 3e-13, Method: Compositional matrix adjust.
 Identities = 106/424 (25%), Positives = 180/424 (42%), Gaps = 51/424 (12%)

Query  53    VDKNTGDINITAIVDREETPSFLITCRALNAQGLDVEKPLILTVKILDINDNPPVFSQ-Q  111
             +D++  ++N+TA+ D         +C  +N          ++ V I D+NDN PVF
Sbjct  938   LDRDKYELNVTAMDDG---------SC-CVNGDATIHTSTAVVVVFITDVNDNKPVFKDCG  988

Query  112   IFMGEIEENSASNSLVMILNATDADEPNHLNSKIAFKIVSQEPAGTPMFLLSRNTGEVRT  171
              +  ++EE + + S V+ + ATD D+   +N ++ + IV Q       F +   TGEV T
Sbjct  989   TYYPKVEEGAPNGSPVIKVQATDEDK--GVNGQVKYSIVQQPNQKGTKFTVDEETGEVST  1046

Query  172   LTNSLDRE--QASSYRLVVSGADKDGEGLSTQCECNIKVKDVNDNFPMFRDSQYSARIEE  229
                  DRE        + V   D+    L   C   +++ DVNDN P+F   +Y   +++
Sbjct  1047  -NKVFDREGDDGKFVSVTVKATDQGEPSLEGVCSFTVEITDVNDNPPLFDRQKYVENVKQ  1105

Query  230   NI-LSSELLRFQVTDLDEEYTDNWLAVYFFT---SGNEGNWFEIQTDPRTNEGILKVVKA  285
             +  + + +LR  V+  DE+  +N   VY  +   S N+  +FEIQ  P +   +LK  K
Sbjct  1106  DASIGTNILR--VSASDEDADNNGAIVYTLSAPHSPNDLEYFEIQ--PESGWIVLK--KP  1159

Query  286   LDYEQLQSVKLSIAVKNK------AEFHQSVISRYRVQSTPVTIQV----INVREGIAFR  335
             LD E+    +L +   ++      A+    +    R    P+  +     I++RE +
Sbjct  1160  LDRERY---RLKVYASDRGDPPSTADVDVELDVVDRNNKPPIWDRTIYGPIHIRENVTVG  1216

Query  336   PASKTFTVQKGISSKKLVDYILGTYQAIDEDTNKAASNVKYVMGRNDGGYLMIDSKTAEI  395
                 +     GI     V Y L         TNK   +  Y+  R D G        A+I
Sbjct  1217  TVVTSVKASSGIEGNPTVFYRL--MPGSTAQTNKL--HTFYLQQRPDNG-----DTWADI  1267

Query  396   KFVKNMNRDSTFIVNKTITAEVLAIDEYTGKTSTGTVYYVRVPDFNDNCPTAV-LEKDAVC  454
             K    ++ ++    N T+  E     +     S  TVY+++ D ND  P     E++ V
Sbjct  1268  KVNHPLDFETIKEYNLTVRVENNGAQQL---ASEATVYIQLEDVNDEIPLFTEREQETVL  1324

Query  455   SSSP  458
                P
Sbjct  1325  EGEP  1328

 Score = 76.3 bits (186),  Expect = 1e-12, Method: Compositional matrix adjust.
 Identities = 103/418 (25%), Positives = 174/418 (42%), Gaps = 50/418 (12%)
```

```
Query  50    IFVVDKNTGDINITAIVDREETPSFLITCRAL----------NAQGLDVEKPLILTVKIL  99
              F ++  +G+I    + DRE+  ++ +   A           N Q   V K   + + I
Sbjct  1365  FFEINLQSGEIFTKVVFDREKQGAYALEVEARDGAPSARPNSNQQPNSVTK--FIRIGIA  1422

Query  100   DINDNPPVFSQQIFMGEIEENSASNSLVMILNATDADEPNHLNSKIAFKIVSQEPAGTPM  159
             D NDNPP F + ++  E++EN      V+ + A D DE    +S+I ++I S    G
Sbjct  1423  DKNDNPPYFDKALYEAEVDENEDIQHTVLTVTAKDHDE----SSRIRYEITSGNIGGA--  1476

Query  160   FLLSRNTGEVRTLTNSLDREQASSYRLVVSGADKDGEGLSTQCECNIKVKDVNDNFPMFR  219
             F +   TG +  +  +LD E    Y L +  +D   E  +      I VKDVND  P F
Sbjct  1477  FAVKNMTGAI-YVAGALDYETRKRYHLTLVASDSLNENETIIV---INVKDVNDMPPSFP  1532

Query  220   DSQYSARIEENI-LSSELLRFQVTDLDEEYTDNWLAVYFFTSG--NEGNWFEIQTDPRTN  276
                Y   ++E + +   +++   TD D++   N   VYF T    +  N    + D
Sbjct  1533  QQVYERTMDEELAVPFPIMQVTATDGDKDRPQN--IVYFLTGQGIDPDNPANSKFDINRT  1590

Query  277   EGILKVVKALDYEQLQS-VKLSIAVKNKAEFHQSVISRYRVQSTPVTIQVINVREGIAFR  335
              G + V+K LD +Q     +    V  + E  + ++    VQ     + + ++ +
Sbjct  1591  TGEIFVLKPLDRDQPNGRPQWRFTVFAQDEGGEGLVGYADVQ-----VNLKDINDNAPIF  1645

Query  336   PASKTFTVQKGISSKKLVDYILGTYQAIDEDTNKAASNVK--YVMGRN------DGGYLM  387
             P    F     ++        ++ T  A+D D     +N K  Y + +N
Sbjct  1646  PQGVYF---GNVTENGTAGMVVMTMTAVDYDDPNEGTNAKLIYSIEKNVIEEETGSPIFE  1702

Query  388   IDSKTAEIK-FVKNMNRDSTFIVNKTITAEVLAIDEYTGKTSTGTVYVRVPDFNDNCP  444
             I++ T  IK  V  ++R+ T       + +V+A+D   G   TGT  +RV D ND  P
Sbjct  1703  IEADTGVIKTAVCCLDRERT----PDYSIQVVAMDG-GGLKGTGTASIRVKDINDMPP  1755

 Score = 65.1 bits (157),  Expect = 4e-09, Method: Compositional matrix adjust.
 Identities = 85/359 (24%), Positives = 153/359 (43%), Gaps = 32/359 (9%)

Query  51    FVVDKNTGDINITAIVDRE-ETPSFL-ITCRALNAQGLDVEKPLILTVKILDINDNPPVF  108
             F VD+ TG+++   + DRE +   F+ +T +A +     +E     TV+I D+NDNPP+F
Sbjct  1035  FTVDEETGEVSTNKVFDREGDDGKFVSVTVKATDQGEPSLEGVCSFTVEITDVNDNPPLF  1094

Query  109   SQQIFMGEIEENSASNSLVMILNATDADEPNHLNSKIAFKIVS-QEPAGTPMFLLSRNTG  167
              +Q ++  ++++++  + ++ ++A+D D  N  N  I + + +   P     F +   +G
Sbjct  1095  DRQKYVENVKQDASIGTNILRVSASDEDADN--NGAIVYTLSAPHSPNDLEYFEIQPESG  1152

Query  168   EVRTLTNSLDREQASSYRLVVSGADKDGEGLSTQCECNIKVKDVNDNFPMFRDSQYSARI  227
              +  L   LDRE+   YRL V  +D+ G+  ST            +N P   D      I
Sbjct  1153  WI-VLKKPLDRER---YRLKVYASDR-GDPPSTADVDVELDVVDRNNKPPIWDRTIYGPI  1207

Query  228   EENILSSELLRFQVTDLDEEYTDNWLAVYFFTSG-----NEGNWFEIQTDPRTNE--GIL  280
                   +                N    Y    G     N+ + F +Q  P   +    +
Sbjct  1208  HIRENVTVGTVVTSVKASSGIEGNPTVFYRLMPGSTAQTNKLHTFYLQQRPDNGDTWADI  1267

Query  281   KVVKALDYEQLQSVKLSIAVKNKAEFHQSVISRYRVQSTPVTIQVINVREGIAFRPASKT  340
             KV   LD+E ++   L++ V+N         ++       V IQ+ +V + I      +
Sbjct  1268  KVNHPLDFETIKEYNLTVRVENNG-------AQQLASEATVYIQLEDVNDEIPLFTEREQ  1320

Query  341   FTVQKGISSKKLVDYILGTYQAIDEDTNKAASNVKYVM---GRNDG-GYLMIDSKTAEI  395
              TV +G      V  +     AID+D     + V Y +    RN+G  +  I+ ++ EI
Sbjct  1321  ETVLEGEPIGTKVTQV----NAIDKDGTFPNNQVYYYIVDSPRNEGKDFFEINLQSGEI  1375

 Score = 55.5 bits (132),  Expect = 3e-06, Method: Compositional matrix adjust.
 Identities = 49/158 (31%), Positives = 78/158 (49%), Gaps = 24/158 (15%)

Query  102   NDNPPVFSQQIFMGEIEENSASNSLVMILNATDADEPNHLNSKIAFKIVSQEPAGTP--M  159
             ND  P FSQQ++   ++EN+  N+LV  + A+D D  N     + F  V     GT
Sbjct  868   NDEEPKFSQQVYTPNVDENAGPNTLVTTVVASDKDGDN-----VRFGFVG---GGTSSGQ  919

Query  160   FLLSRNTGEVRTLTN--SLDREQASSYRLVVSGADK-------DGEGLSTQCECNIKVKD  210
             F++   TG +R  +   SLDR++   Y L V+  D        D    ++     + + D
Sbjct  920   FVIEEITGVIRLHSKAISLDRDK---YELNVTAMDDGSCCVNGDATIHTSTAVVVVFITD  976

Query  211   VNDNFPMFRD-SQYSARIEENILS-SELLRFQVTDLDE  246
             VNDN P+F+D   Y  ++EE   + S +++ Q TD D+
```

```
Sbjct  977   VNDNKPVFKDCGTYYPKVEEGAPNGSPVIKVQATDEDK  1014

 Score = 53.5 bits (127),  Expect = 1e-05, Method: Compositional matrix adjust.
 Identities = 70/306 (23%), Positives = 126/306 (41%), Gaps = 30/306 (10%)

Query  20    PIAKIT-SDYQATQKITYRI---SGVGIDQPPFGIFVVDKNTGDINITAIVDRE---ETP  72
             PI  +T  D   T K  Y++   SG G D+  F +   +  TG + I   +D E   ++
Sbjct  1778  PILTVTVHDEDETNKFQYKVIDNSGYGADK--FTMVRNNDGTGSLKIVQPLDYEDPLQSN  1835

Query  73    SFLITCRALNAQGLDVEKPLI------LTVKILDINDNPPVFSQQIFMGEIEENSASNSL  126
              F    + +N +G D +          + VK+ DINDN P F +      + EN+
Sbjct  1836  GFRFRIQ-VNDKGEDNDNDKYHVAYSWVVVKLRDINDNKPQFERPNIEVSVYENADVGKT  1894

Query  127   VMILNATDADEPNHLNSKIAFKIVSQEPAGTPMFLLSRNTGEVRTLTNSLDREQASSYRL  186
             +    ATD D+     SK+++ I   + +       S N     T+  +LDRE    +++
Sbjct  1895  LETFKATDPDQGG--KSKVSYAI---DRSSDRQRQFSINQEGTVTIQRNLDREVTPRHQV  1949

Query  187   VVSGADKDGEGLSTQCECNIKVKDVNDNFPMFRDSQYSARIEENILSSELLRFQVTDLDE  246
              +   D      +      + V+D+NDN P F    Y   + E++   +++    TD D+
Sbjct  1950  KILAIDDGIPPKTATATLTVIVQDINDNPPKFL-KDYRPVLPEHVPPRKVVEILATDDDD  2008

Query  247   EYTDNWLAVYFFTSGNEGNW----FEIQTDPRTNEG----ILKVVKALDYEQLQSVKLSI  298
                 N     F       +     F+++ D +   G    I+  +++ D EQ +   + I
Sbjct  2009  RSKSNGPPFQFRLDPGADDIIRASFKVEQDQKGANGDGMAIVSSLRSFDREQQKEYLIPI  2068

Query  299   AVKNKA  304
              +K+
Sbjct  2069  VIKDHG  2074

 Score = 52.4 bits (124),  Expect = 3e-05, Method: Compositional matrix adjust.
 Identities = 97/398 (24%), Positives = 163/398 (41%), Gaps = 54/398 (14%)

Query  106   PVFSQQIFMGEIEENSASNSLVMILNATDADEPNHLNSKIAFKIVSQ-EPAGTPMFLLSR  164
             P F    +  EI EN   +S ++ + A      +    +I + + +Q + AGT  F +
Sbjct  665   PQFYMPSYEAEIPENQKKDSDIISVKAK-----SFADREIRYTLKAQGQGAGT--FNIGP  717

Query  165   NTGEVRTLTNSLDRE---QASSYRLVVSGADKDGEGLSTQCECNIKVKDVNDNFPMFRDS  221
              +G V+ L   LD E   Q   Y LVV+ A +D  G ST  E  I+V DVNDN P F
Sbjct  718   TSGIVK-LAKELDFEDLRQPHVYSLVVT-ATEDSGGFSTSVELTIRVTDVNDNAPKFELP  775

Query  222   QYSAR-IEENI-LSSELLRFQVTDLDEEYTDNWLAVYFFTSGNEGNWFEIQTDPRTNEGI  279
              Y A  ++E+I L + +L+ +  D D         + +  S +    F + ++     GI
Sbjct  776   DYQAHNVDEDIPLGTSILKVKAMDADSGAN---AEIEYLVSDDH---FAVDSN-----GI  824

Query  280   LKVVKALDYEQLQS-VKLSIAVKNKAEFHQSVISRYRVQSTPVTIQVINVREGIAFRPAS  338
             +   K LD +   +  +  +  K+K E  ++  +  RV +           E   F
Sbjct  825   IVNNKQLDADNNNAYYEFVVTAKDKGEPAKTGTATVRVYTKNKN------DEEPKFSQQV  878

Query  339   KTFTVQKGISSKKLVDYILGTYQAIDEDTNKAASNVK--YVMGRNDGGYLMIDSKTAEIK  396
              T  V +      LV  ++         ++K   NV+  +V G    G  +I+  T  I+
Sbjct  879   YTPNVDENAGPNTLVTTVVA--------SDKDGDNVRFGFVGGGTSSGQFVIEEITGVIR  930

Query  397   FVK---NMNRDSTFI-VNKTITAEVLAIDEYTGKTSTGTVYVRVPDFNDN------CPTA  446
                   +++RD   + V            + T  TST  V V + D NDN      C T
Sbjct  931   LHSKAISLDRDKYELNVTAMDDGSCCVNGDATIHTSTAVVVVFITDVNDNKPVFKDCGTY  990

Query  447   VLE-KDAVCSSSPSVVVSARTLNNRYTGPYTFALEDQP  483
               + ++   + SP + V A   +    G   +++  QP
Sbjct  991   YPKVEEGAPNGSPVIKVQATDEDKGVNGQVKYSIVQQP  1028

 Score = 44.3 bits (103),  Expect = 0.009, Method: Compositional matrix adjust.
 Identities = 48/199 (24%), Positives = 73/199 (37%), Gaps = 19/199 (10%)

Query  27    DYQATQKITYRISGVGIDQPPFGIFVVDKNTGDINITAIVDREETPSFLITCRALNAQGL  86
             D     K++Y I      Q  F I       G + I   +DRE TP   +   A++
Sbjct  1904  DQGGKSKVSYAIDRSSDRQRQFSI----NQEGTVTIQRNLDREVTPRHQVKILAIDDGIP  1959

Query  87    DVEKPLILTVKILDINDNPPVFSQQIFMGEIEENSASNSLVMILNATDADEPNHLNSKIA  146
```

```
                     LTV + DINDNPP F +  +   + E+     +V IL   D D
Sbjct  1960  PKTATATLTVIVQDINDNPPKFLKD-YRPVLPEHVPPRKVVEILATDDDDRSKSNGPPFQ  2018

Query  147   FKIVSQEPAGTPMFLLSRNTGEVRTLTN-----------SLDREQASSYRLVVSGADKDG  195
             F++   +P    +   S    + +   N           S DREQ   Y + +   D
Sbjct  2019  FRL---DPGADDIIRASFKVEQDQKGANGDGMAIVSSLRSFDREQQKEYLIPIVIKDHGN  2075

Query  196   EGLSTQCECNIKVKDVNDN  214
               ++      + + DVNDN
Sbjct  2076  PAMTGTSTLTVVIGDVNDN  2094

 Score = 40.4 bits (93),  Expect = 0.13, Method: Compositional matrix adjust.
 Identities = 55/242 (23%), Positives = 102/242 (42%), Gaps = 22/242 (9%)

Query  4    KFAKPCREGE--DNSKRNP-IAKITSDYQATQKITYRISGVGIDQPPFGIFVVDKNTGDI  60
            +F  P  E E  +N K++  I  + +   A ++I Y +   G      G F +   +G +
Sbjct  666  QFYMPSYEAEIPENQKKDSDIISVKAKSFADREIRYTLKAQGQGA---GTFNIGPTSGIV  722

Query  61   NITAIVDREE--TPSF--LITCRALNAQGLDVEKPLILTVKILDINDNPPVFSQQIFMGE  116
             +   +D E+   P    L+     ++ G      + LT+++ D+NDN P F    +
Sbjct  723  KLAKELDFEDLRQPHVYSLVVTATEDSGGFSTS--VELTIRVTDVNDNAPKFELPDYQAH  780

Query  117  -IEENSASNSLVMILNATDADEPNHLNSKIAFKIVSQEPAGTPMFLLSRNTGEVRTLTNS  175
             ++E+    + ++ + A DAD  +  N++I + +VS +      F +  N   V
Sbjct  781  NVDEDIPLGTSILKVKAMDAD--SGANAEIEY-LVSDD-----HFAVDSNGIIVNNKQLD  832

Query  176  LDREQASSYRLVVSGADKDGEGLSTQCECNIKVKDVNDNFPMFRDSQYSARIEENILSSE  235
             D   A  Y  VV+  DK     +      +  K+ ND  P F    Y+  ++EN   +
Sbjct  833  ADNNNA-YYEFVVTAKDKGEPAKTGTATVRVYTKNKNDEEPKFSQQVYTPNVDENAGPNT  891

Query  236  LL  237
            L+
Sbjct  892  LV  893

 Score = 39.3 bits (90),  Expect = 0.37, Method: Compositional matrix adjust.
 Identities = 89/389 (23%), Positives = 149/389 (38%), Gaps = 62/389 (16%)

Query  75   LITCRALNAQGLDVEKPLILTVKILDINDNPPVFSQQIF--MGEIEENSASNSLVMILNA  132
            +IT +  NA GL       + + + D+ND PP F  +       ++ N+  N+ V  L A
Sbjct  527  IITNQGHNA-GLKYTDNQRVIIHVKDVNDEPPYFINRPLPMQAVVQLNAPPNTPVFTLQA  585

Query  133  TDADEPNHLNSKIAFKIVSQEPAGTPMFLLSRNTGEVRTLTNSLDREQASSYRLVVSGAD  192
             D D     +  I + IV     G   F +   +G VRT    L  +    Y L V   D
Sbjct  586  RDPD----TDHNIHYFIVRDRTGG--RFEVDERSGVVRTRGTDL-FQLDMEYVLYVKAED  638

Query  193  KDGE----GLSTQCECNIKVKDVNDNFPMFRDSQYSARIEEN-ILSSELLRFQVTDLDEE  247
            ++G+       +  E  + +        P F    Y A I EN    S+++  +     +
Sbjct  639  QNGKVDDRRFQSTPEERLSIVG-GKRAPQFYMPSYEAEIPENQKKDSDIISVKA----KS  693

Query  248  YTDNWLAVYFFTSGNEGNWFEIQTDPRTNEGILKVVKALDYEQLQSVKLS----IAVKNK  303
            + D  +       G     F I         GI+K+ K LD+E L+   +      A ++
Sbjct  694  FADREIRYTLKAQGQGAGTFNIGP----TSGIVKLAKELDFEDLRQPHVYSLVVTATEDS  749

Query  304  AEFHQSVISRYRVQSTPVTIQVINVREGIAFRPASKTFTVQKGISSKKLVDYILGT----  359
              F  SV           +TI+V +V +       +  F +    +     D  LGT
Sbjct  750  GGFSTSV---------ELTIRVTDVNDN------APKFELPDYQAHNVDEDIPLGTSILK  794

Query  360  YQAIDEDTNKAASNVKYVMGRN----DGGYLMIDSKTAEIKFVKNMNRDSTFIVNKTITA  415
             +A+D D+  A + ++Y++  +    D    +++++K  +     N N    F+V
Sbjct  795  VKAMDADSG-ANAEIEYLVSDDHFAVDSNGIIVNNKQLD---ADNNNAYYEFVVTAKDKG  850

Query  416  EVLAIDEYTGKTSTGTVYVRVPDFNDNCP  444
            E          KT T TV V   + ND  P
Sbjct  851  E-------PAKTGTATVRVYTKNKNDEEP  872

>AFUN2_006054.P9862 | transcript=AFUN2_006054.R9862 | gene=AFUN2_006054
| organism=Anopheles_funestus_AfunGA1 | gene_product=neural-cadherin,
transcript variant X17, neural-cadherin,
```

transcript variant X22, neural-cadherin, transcript variant X12, neural-cadherin, transcript variant X1, neural-cadherin, transcript variant X15, neural-cadherin, transcript variant X2, neural-cadherin, transcript variant X23, neural-cadherin, transcript variant X26, neural-cadherin, transcript variant X24, neural-cadherin, transcript variant X7, neural-cadherin, transcript variant X6, neural-cadherin, transcript variant X18, neural-cadherin, transcript variant X3, neural-cadherin, transcript variant X9, neural-cadherin, transcript variant X14, neural-cadherin, transcript variant X4, neural-cadherin, transcript variant X25, neural-cadherin, transcript variant X21, neural-cadherin, transcript variant X20, neural-cadherin, transcript variant X19, neural-cadherin, transcript variant X13, neural-cadherin, transcript variant X11, neural-cadherin, transcript variant X16, neural-cadherin, transcript variant X5, neural-cadherin, transcript variant X10, neural-cadherin, transcript variant X8 | transcript_product=neural-cadherin, transcript variant X22 | location=AfunGA1_2RL:82372181-82559663(-) | protein_length=3109 | sequence_SO=chromosome | SO=protein_coding_gene | is_pseudo=false
Length=3109

```
 Score = 147 bits (370),  Expect = 3e-35, Method: Compositional matrix adjust.
 Identities = 128/446 (29%), Positives = 217/446 (49%), Gaps = 36/446 (8%)

Query  20    PIAKITS---DYQATQKITYRISGVGID--QPPFGIFVVDKNTGDINITAIVDREET---  71
             PI ++T+   D    Q I Y ++G GID   P    F +++ TG+I +   +DR++
Sbjct  1549  PIMQVTATDGDKDRPQNIVYFLTGQGIDPDNPANSKFDINRTTGEIFVLKPLDRDQPNGR  1608

Query  72    PSFLITCRALNAQGLDVEKPLILTVKILDINDNPPVFSQQIFMGEIEENSASNSLVMILN  131
             P +  T  A +  G  +     + V + DINDN P+F Q ++ G + EN  +  +VM +
Sbjct  1609  PQWRFTVFAQDEGGEGLVGYADVQVNLKDINDNAPIFPQGVYFGNVTENGTAGMVVMTMT  1668

Query  132   ATDADEPNH-LNSKIAFKI---VSQEPAGTPMFLLSRNTGEVRTLTNSLDREQASSYRLV  187
             A D D+PN   N+K+ + I   V +E  G+P+F +  +TG ++T    LDRE+   Y +
Sbjct  1669  AVDYDDPNEGTNAKLIYSIEKNVIEEETGSPIFEIEADTGVIKTAVCCLDRERTPDYSIQ  1728

Query  188   VSGADKDGEGLSTQCECNIKVKDVNDNFPMFRDSQYSARIEENILSS--ELLRFQVTDLD  245
             V     DG GL       +I+VKD+ND  P F   ++   ++E   ++  E+    VT  D
Sbjct  1729  VVAM--DGGGLKGTGTASIRVKDINDMPPQFTKDEWFTEVDETDGTNLPEMPILTVTVHD  1786

Query  246   EEYTDNWLAVYFFTSGNEGNWFEIQTDPRTNEGI--LKVVKALDYE---QLQSVKLSIAV  300
             E+ T+ +        SG    + F +    R N+G   LK+V+ LDYE   Q    +  I V
Sbjct  1787  EDETNKFQYKVIDNSGYGADKFTMV---RNNDGTGSLKIVQPLDYEDPLQSNGFRFRIQV  1843

Query  301   KNKAEFHQSVISRYRVQSTPVTIQVINVREGIAFRPASKTFTVQKGISSKKLVDYILGTY  360
              +K E + +   +Y V  + V +++ ++ +    +P  +   ++  +     V   L T+
Sbjct  1844  NDKGEDNDN--DKYHVAYSWVVVKLRDINDN---KPQFERPNIEVSVYENADVGKTLETF  1898

Query  361   QAIDEDTNKAASNVKYVMGR-NDGGYLMIDSKTAEIKFVKNMNRDSTFIVNKTITAEVLA  419
             +A D D     S V Y + R +D        ++   +   +N++R+    V       ++LA
Sbjct  1899  KATDPDQG-GKSKVSYAIDRSSDRQRQFSINQEGTVTIQRNLDRE----VTPRHQVKILA  1953

Query  420   IDE-YTGKTSTGTVYYVRVPDFNDNCP  444
             ID+    KT+T T+ V V D NDN P
Sbjct  1954  IDDGIPPKTATATLTVIVQDINDNPP  1979

 Score = 76.3 bits (186),  Expect = 1e-12, Method: Compositional matrix adjust.
 Identities = 103/418 (25%), Positives = 174/418 (42%), Gaps = 50/418 (12%)

Query  50    IFVVDKNTGDINITAIVDREETPSFLITCRAL----------NAQGLDVEKPLILTVKIL  99
              F ++  +G+I     + DRE+  ++ +   A             N Q   V K   + + I
Sbjct  1365  FFEINLQSGEIFTKVVFDREKQGAYALEVEARDGAPSARPNSNQQPNSVTK--FIRIGIA  1422

Query  100   DINDNPPVFSQQIFMGEIEENSASNSLVMILNATDADEPNHLNSKIAFKIVSQEPAGTPM  159
             D NDNPP F + ++  E++EN       V+ + A D DE    +S+I ++I S    G
Sbjct  1423  DKNDNPPYFDKALYEAEVDENEDIQHTVLTVTAKDHDE----SSRIRYEITSGNIGGA--  1476

Query  160   FLLSRNTGEVRTLTNSLDREQASSYRLVVSGADKDGEGLSTQCECNIKVKDVNDNFPMFR  219
             F +   TG +  +  +LD E    Y L +  +D   E  +       I VKDVND  P F
```

```
Sbjct  1477   FAVKNMTGAI-YVAGALDYETRKRYHLTLVASDSLNENETIIV---INVKDVNDMPPSFP  1532

Query  220    DSQYSARIEENI-LSSELLRFQVTDLDEEYTDNWLAVYFFTSG--NEGNWFEIQTDPRTN  276
                 Y   ++E + +   +++   TD D++   N   VYF T    +  N    + D
Sbjct  1533   QQVYERTMDEELAVPFPIMQVTATDGDKDRPQN--IVYFLTGQGIDPDNPANSKFDINRT  1590

Query  277    EGILKVVKALDYEQLQS-VKLSIAVKNKAEFHQSVISRYRVQSTPVTIQVINVREGIAFR  335
               G + V+K LD +Q     +    V  + E  + ++    VQ     + + ++ +
Sbjct  1591   TGEIFVLKPLDRDQPNGRPQWRFTVFAQDEGGEGLVGYADVQ-----VNLKDINDNAPIF  1645

Query  336    PASKTFTVQKGISSKKLVDYILGTYQAIDEDTNKAASNVK--YVMGRN------DGGYLM  387
              P    F     ++       ++ T  A+D D     +N K  Y + +N
Sbjct  1646   PQGVYF---GNVTENGTAGMVVMTMTAVDYDDPNEGTNAKLIYSIEKNVIEEETGSPIFE  1702

Query  388    IDSKTAEIK-FVKNMNRDSTFIVNKTITAEVLAIDEYTGKTSTGTVYVRVPDFNDNCP  444
              I++ T  IK  V  ++R+ T       + +V+A+D   G   TGT  +RV D ND  P
Sbjct  1703   IEADTGVIKTAVCCLDRERT----PDYSIQVVAMDG-GGLKGTGTASIRVKDINDMPP  1755

 Score = 75.9 bits (185),  Expect = 2e-12, Method: Compositional matrix adjust.
 Identities = 89/361 (25%), Positives = 167/361 (46%), Gaps = 36/361 (10%)

Query  51     FVVDKNTGDINITAIVDRE-ETPSFL-ITCRALNAQGLDVEKPLILTVKILDINDNPPVF  108
              F VD+ TG+++   + DRE +   F+ +T +A +     +E     TV+I D+NDNPP+F
Sbjct  1035   FTVDEETGEVSTNKVFDREGDDGKFVSVTVKATDQGEPSLEGVCSFTVEITDVNDNPPLF  1094

Query  109    SQQIFMGEIEENSASNSLVMILNATDADEPNHLNSKIAFKIVS-QEPAGTPMFLLSRNTG  167
               +Q ++  ++++++  + ++ ++A+D D  N  N  I + + +   P     F +   +G
Sbjct  1095   DRQKYVENVKQDASIGTNILRVSASDEDADN--NGAIVYTLSAPHSPNDLEYFEIQPESG  1152

Query  168    EVRTLTNSLDREQASSYRLVVSGADKDGEGLSTQCECNIKVKDVNDNFPMFRDSQYSA-R  226
               +  L   LDRE   +Y+L     DK    LS   E  I V D  +N P++ ++ Y
Sbjct  1153   WI-VLKKPLDRE---TYKLEAMAQDKGYPPLSRTVEVQIDVVDRANNPPVWDNTVYGPIY  1208

Query  227    IEENI-LSSELLRFQVTDLDEEYTDNWLAVYFFTSG-----NEGNWFEIQTDPRTNE--G  278
              ++EN+ +  +++  + +   E    N    Y    G     N+ + F +Q  P   +
Sbjct  1209   VKENMPVGGKVVSIKASSGIE---GNPTVFYRLMPGSTAQTNKLHTFYLQQRPDNGDTWA  1265

Query  279    ILKVVKALDYEQLQSVKLSIAVKNKAEFHQSVISRYRVQSTPVTIQVINVREGIAFRPAS  338
               +KV   LD+E ++   L++ V+N         ++       V IQ+ +V + I
Sbjct  1266   DIKVNHPLDFETIKEYNLTVRVENNG-------AQQLASEATVYIQLEDVNDEIPLFTER  1318

Query  339    KTFTVQKGISSKKLVDYILGTYQAIDEDTNKAASNVKYVM---GRNDG-GYLMIDSKTAE  394
              +  TV +G      V  +     AID+D     + V Y +    RN+G  +  I+ ++ E
Sbjct  1319   EQETVLEGEPIGTKVTQV----NAIDKDGTFPNNQVYYYIVDSPRNEGKDFFEINLQSGE  1374

Query  395    I  395
              I
Sbjct  1375   I  1375

 Score = 55.8 bits (133),  Expect = 3e-06, Method: Compositional matrix adjust.
 Identities = 49/158 (31%), Positives = 78/158 (49%), Gaps = 24/158 (15%)

Query  102    NDNPPVFSQQIFMGEIEENSASNSLVMILNATDADEPNHLNSKIAFKIVSQEPAGTP--M  159
              ND  P FSQQ++   ++EN+  N+LV  + A+D D  N     + F  V     GT
Sbjct  868    NDEEPKFSQQVYTPNVDENAGPNTLVTTVVASDKDGDN-----VRFGFVG---GGTSSGQ  919

Query  160    FLLSRNTGEVRTLTN--SLDREQASSYRLVVSGADK-------DGEGLSTQCECNIKVKD  210
              F++   TG +R  +   SLDR++   Y L V+  D        D    ++     + + D
Sbjct  920    FVIEEITGVIRLHSKAISLDRDK---YELNVTAMDDGSCCVNGDATIHTSTAVVVVFITD  976

Query  211    VNDNFPMFRD-SQYSARIEENILS-SELLRFQVTDLDE  246
              VNDN P+F+D   Y  ++EE   + S +++ Q TD D+
Sbjct  977    VNDNKPVFKDCGTYYPKVEEGAPNGSPVIKVQATDEDK  1014

 Score = 53.5 bits (127),  Expect = 1e-05, Method: Compositional matrix adjust.
 Identities = 70/306 (23%), Positives = 126/306 (41%), Gaps = 30/306 (10%)

Query  20     PIAKIT-SDYQATQKITYRI---SGVGIDQPPFGIFVVDKNTGDINITAIVDRE---ETP  72
```

```
             PI  +T  D   T K  Y++   SG G D+  F +   +  TG + I   +D E   ++
Sbjct  1778  PILTVTVHDEDETNKFQYKVIDNSGYGADK--FTMVRNNDGTGSLKIVQPLDYEDPLQSN  1835

Query  73    SFLITCRALNAQGLDVEKPLI------LTVKILDINDNPPVFSQQIFMGEIEENSASNSL  126
              F    + +N +G D +           + VK+ DINDN P F +      + EN+
Sbjct  1836  GFRFRIQ-VNDKGEDNDNDKYHVAYSWVVVKLRDINDNKPQFERPNIEVSVYENADVGKT  1894

Query  127   VMILNATDADEPNHLNSKIAFKIVSQEPAGTPMFLLSRNTGEVRTLTNSLDREQASSYRL  186
             +    ATD D+     SK+++ I    + +        S N     T+  +LDRE    +++
Sbjct  1895  LETFKATDPDQGG--KSKVSYAI---DRSSDRQRQFSINQEGTVTIQRNLDREVTPRHQV  1949

Query  187   VVSGADKDGEGLSTQCECNIKVKDVNDNFPMFRDSQYSARIEENILSSELLRFQVTDLDE  246
              +   D      +      + V+D+NDN P F    Y   + E++   +++    TD D+
Sbjct  1950  KILAIDDGIPPKTATATLTVIVQDINDNPPKFL-KDYRPVLPEHVPPRKVVEILATDDDD  2008

Query  247   EYTDNWLAVYFFTSGNEGNW----FEIQTDPRTNEG----ILKVVKALDYEQLQSVKLSI  298
                 N     F       +     F+++ D +   G    I+  +++ D EQ +   + I
Sbjct  2009  RSKSNGPPFQFRLDPGADDIIRASFKVEQDQKGANGDGMAIVSSLRSFDREQQKEYLIPI  2068

Query  299   AVKNKA  304
              +K+
Sbjct  2069  VIKDHG  2074

 Score = 52.4 bits (124),  Expect = 3e-05, Method: Compositional matrix adjust.
 Identities = 97/398 (24%), Positives = 163/398 (41%), Gaps = 54/398 (14%)

Query  106   PVFSQQIFMGEIEENSASNSLVMILNATDADEPNHLNSKIAFKIVSQ-EPAGTPMFLLSR  164
             P F    +  EI EN   +S ++ + A      +  + +I + + + +Q + AGT  F +
Sbjct  665   PQFYMPSYEAEIPENQKKDSDIISVKAK-----SFADREIRYTLKAQGQGAGT--FNIGP  717

Query  165   NTGEVRTLTNSLDRE---QASSYRLVVSGADKDGEGLSTQCECNIKVKDVNDNFPMFRDS  221
              +G V+ L   LD E   Q   Y LVV+ A +D  G ST  E  I+V DVNDN P F
Sbjct  718   TSGIVK-LAKELDFEDLRQPHVYSLVVT-ATEDSGGFSTSVELTIRVTDVNDNAPKFELP  775

Query  222   QYSAR-IEENI-LSSELLRFQVTDLDEEYTDNWLAVYFFTSGNEGNWFEIQTDPRTNEGI  279
              Y A  ++E+I L + +L+ +  D D          + +  S +    F + ++     GI
Sbjct  776   DYQAHNVDEDIPLGTSILKVKAMDADSGAN---AEIEYLVSDDH---FAVDSN-----GI  824

Query  280   LKVVKALDYEQLQS-VKLSIAVKNKAEFHQSVISRYRVQSTPVTIQVINVREGIAFRPAS  338
             +   K LD +   +  +  + K+K E  ++  +  RV +             E   F
Sbjct  825   IVNNKQLDADNNNAYYEFVVTAKDKGEPAKTGTATVRVYTKNKN------DEEPKFSQQV  878

Query  339   KTFTVQKGISSKKLVDYILGTYQAIDEDTNKAASNVK--YVMGRNDGGYLMIDSKTAEIK  396
              T V +      LV  ++          ++K   NV+  +V G    G  +I+  T  I+
Sbjct  879   YTPNVDENAGPNTLVTTVVA--------SDKDGDNVRFGFVGGGTSSGQFVIEEITGVIR  930

Query  397   FVK---NMNRDSTFI-VNKTITAEVLAIDEYTGKTSTGTVYVRVPDFNDN------CPTA  446
                  +++RD   + V              + T  TST  V V + D NDN      C T
Sbjct  931   LHSKAISLDRDKYELNVTAMDDGSCCVNGDATIHTSTAVVVVFITDVNDNKPVFKDCGTY  990

Query  447   VLE-KDAVCSSSPSVVVSARTLNNRYTGPYTFALEDQP  483
               + ++   + SP + V A   +    G   +++  QP
Sbjct  991   YPKVEEGAPNGSPVIKVQATDEDKGVNGQVKYSIVQQP  1028

 Score = 44.3 bits (103),  Expect = 0.009, Method: Compositional matrix adjust.
 Identities = 48/199 (24%), Positives = 73/199 (37%), Gaps = 19/199 (10%)

Query  27    DYQATQKITYRISGVGIDQPPFGIFVVDKNTGDINITAIVDREETPSFLITCRALNAQGL  86
             D     K++Y I      Q  F I        G + I   +DRE TP   +    A++
Sbjct  1904  DQGGKSKVSYAIDRSSDRQRQFSI----NQEGTVTIQRNLDREVTPRHQVKILAIDDGIP  1959

Query  87    DVEKPLILTVKILDINDNPPVFSQQIFMGEIEENSASNSLVMILNATDADEPNHLNSKIA  146
                    LTV + DINDNPP F +  +    + E+     +V IL   D D
Sbjct  1960  PKTATATLTVIVQDINDNPPKFLKD-YRPVLPEHVPPRKVVEILATDDDDRSKSNGPPFQ  2018

Query  147   FKIVSQEPAGTPMFLLSRNTGEVRTLTN-----------SLDREQASSYRLVVSGADKDG  195
             F++   +P     +   S     + +   N           S DREQ    Y + +   D
Sbjct  2019  FRL---DPGADDIIRASFKVEQDQKGANGDGMAIVSSLRSFDREQQKEYLIPIVIKDHGN  2075
```

```
Query  196   EGLSTQCECNIKVKDVNDN  214
               ++      + + DVNDN
Sbjct  2076  PAMTGTSTLTVVIGDVNDN  2094

 Score = 40.4 bits (93),  Expect = 0.14, Method: Compositional matrix adjust.
 Identities = 55/242 (23%), Positives = 102/242 (42%), Gaps = 22/242 (9%)

Query  4    KFAKPCREGE--DNSKRNP-IAKITSDYQATQKITYRISGVGIDQPPFGIFVVDKNTGDI  60
            +F  P  E E  +N K++  I  + +   A ++I Y +   G      G F +   +G +
Sbjct  666  QFYMPSYEAEIPENQKKDSDIISVKAKSFADREIRYTLKAQGQGA---GTFNIGPTSGIV  722

Query  61   NITAIVDREE--TPSF--LITCRALNAQGLDVEKPLILTVKILDINDNPPVFSQQIFMGE  116
             +   +D E+   P    L+     ++ G      + LT+++ D+NDN P F    +
Sbjct  723  KLAKELDFEDLRQPHVYSLVVTATEDSGGFSTS--VELTIRVTDVNDNAPKFELPDYQAH  780

Query  117  -IEENSASNSLVMILNATDADEPNHLNSKIAFKIVSQEPAGTPMFLLSRNTGEVRTLTNS  175
             ++E+    + ++ + A DAD  +  N++I + +VS +      F +  N   V
Sbjct  781  NVDEDIPLGTSILKVKAMDAD--SGANAEIEY-LVSDD-----HFAVDSNGIIVNNKQLD  832

Query  176  LDREQASSYRLVVSGADKDGEGLSTQCECNIKVKDVNDNFPMFRDSQYSARIEENILSSE  235
             D   A  Y  VV+  DK     +      +  K+ ND  P F    Y+  ++EN   +
Sbjct  833  ADNNNA-YYEFVVTAKDKGEPAKTGTATVRVYTKNKNDEEPKFSQQVYTPNVDENAGPNT  891

Query  236  LL  237
            L+
Sbjct  892  LV  893

 Score = 38.9 bits (89),  Expect = 0.40, Method: Compositional matrix adjust.
 Identities = 55/225 (24%), Positives = 88/225 (39%), Gaps = 24/225 (11%)

Query  75   LITCRALNAQGLDVEKPLILTVKILDINDNPPVFSQQIF--MGEIEENSASNSLVMILNA  132
            +IT +  NA GL       + + + D+ND PP F  +       ++ N+  N+ V  L A
Sbjct  527  IITNQGHNA-GLKYTDNQRVIIHVKDVNDEPPYFINRPLPMQAVVQLNAPPNTPVFTLQA  585

Query  133  TDADEPNHLNSKIAFKIVSQEPAGTPMFLLSRNTGEVRTLTNSLDREQASSYRLVVSGAD  192
             D D     +  I + IV     G   F +   +G VRT    L  +     Y L V   D
Sbjct  586  RDPD----TDHNIHYFIVRDRTGG--RFEVDERSGVVRTRGTDL-FQLDMEYVLYVKAED  638

Query  193  KDGE----GLSTQCECNIKVKDVNDNFPMFRDSQYSARIEEN-ILSSELLRFQVTDLDEE  247
            ++G+       +  E  + +       P F    Y A I EN    S+++  +     +
Sbjct  639  QNGKVDDRRFQSTPEERLSIVG-GKRAPQFYMPSYEAEIPENQKKDSDIISVKA----KS  693

Query  248  YTDNWLAVYFFTSGNEGNWFEIQTDPRTNEGILKVVKALDYEQLQ  292
            + D  +       G     F I        GI+K+ K LD+E L+
Sbjct  694  FADREIRYTLKAQGQGAGTFNIGP----TSGIVKLAKELDFEDLR  734

>AMEM21_003341.P6433 | transcript=AMEM21_003341.R6433 | gene=AMEM21_003341
| organism=Anopheles_merus_MAF_2021 | gene_product=neural-cadherin,
transcript variant X1 | transcript_product=neural-cadherin,
transcript variant X16 | location=CM029766:40289848-40498951(+)
| protein_length=3099 | sequence_SO=chromosome
| SO=protein_coding_gene | is_pseudo=false
Length=3099

 Score = 146 bits (369),  Expect = 4e-35, Method: Compositional matrix adjust.
 Identities = 128/446 (29%), Positives = 217/446 (49%), Gaps = 36/446 (8%)

Query  20    PIAKITS---DYQATQKITYRISGVGID--QPPFGIFVVDKNTGDINITAIVDREET---  71
             PI ++T+   D    Q I Y ++G GID   P    F +++ TG+I +   +DR++
Sbjct  1539  PIMQVTATDGDKDRPQNIVYFLTGQGIDPDNPANSKFDINRTTGEIFVLKPLDRDQPNGR  1598

Query  72    PSFLITCRALNAQGLDVEKPLILTVKILDINDNPPVFSQQIFMGEIEENSASNSLVMILN  131
             P +  T  A +  G  +     + V + DINDN P+F Q ++ G + EN  +  +VM +
Sbjct  1599  PQWRFTVFAQDEGGEGLVGYADVQVNLKDINDNAPIFPQGVYFGNVTENGTAGMVVMTMT  1658

Query  132   ATDADEPNH-LNSKIAFKI---VSQEPAGTPMFLLSRNTGEVRTLTNSLDREQASSYRLV  187
             A D D+PN   N+K+ + I   V +E  G+P+F +  +TG ++T    LDRE+   Y +
Sbjct  1659  AVDYDDPNEGTNAKLIYSIEKNVIEEETGSPIFEIEADTGVIKTAVCCLDRERTPDYSIQ  1718
```

```
Query  188   VSGADKDGEGLSTQCECNIKVKDVNDNFPMFRDSQYSARIEENILSS--ELLRFQVTDLD  245
             V     DG GL      +I+VKD+ND  P F   ++   ++E   ++  E+    VT  D
Sbjct  1719  VVAM--DGGGLKGTGTASIRVKDINDMPPQFTKDEWFTEVDETDGTNLPEMPILTVTVHD  1776

Query  246   EEYTDNWLAVYFFTSGNEGNWFEIQTDPRTNEGI--LKVVKALDYE---QLQSVKLSIAV  300
             E+ T+ +        SG    + F +    R N+G   LK+V+ LDYE   Q    +  I V
Sbjct  1777  EDETNKFQYKVIDNSGYGADKFTMV---RNNDGTGSLKIVQPLDYEDPLQSNGFRFRIQV  1833

Query  301   KNKAEFHQSVISRYRVQSTPVTIQVINVREGIAFRPASKTFTVQKGISSKKLVDYILGTY  360
              +K E + +    +Y V  + V +++ ++ +    +P  +   ++  +     V   L T+
Sbjct  1834  NDKGEDNDN--DKYHVAYSWVVVKLRDINDN---KPQFERPNIEVSVYENADVGKTLETF  1888

Query  361   QAIDEDTNKAASNVKYVMGR-NDGGYLMIDSKTAEIKFVKNMNRDSTFIVNKTITAEVLA  419
             +A D D     S V Y + R +D       ++   +   +N++R+    V       ++LA
Sbjct  1889  KATDPDQG-GKSKVSYAIDRSSDRQRQFSINQEGTVTIQRNLDRE----VTPRHQVKILA  1943

Query  420   IDE-YTGKTSTGTVYVRVPDFNDNCP  444
             ID+    KT+T T+ V V D NDN P
Sbjct  1944  IDDGIPPKTATATLTVIVQDINDNPP  1969

 Score = 75.9 bits (185),  Expect = 2e-12, Method: Compositional matrix adjust.
 Identities = 89/361 (25%), Positives = 167/361 (46%), Gaps = 36/361 (10%)

Query  51    FVVDKNTGDINITAIVDRE-ETPSFL-ITCRALNAQGLDVEKPLILTVKILDINDNPPVF  108
             F VD+ TG+++   + DRE +   F+ +T +A +     +E     TV+I D+NDNPP+F
Sbjct  1025  FTVDEETGEVSTNKVFDREGDDGKFVSVTVKATDQGEPSLEGVCSFTVEITDVNDNPPLF  1084

Query  109   SQQIFMGEIEENSASNSLVMILNATDADEPNHLNSKIAFKIVS-QEPAGTPMFLLSRNTG  167
              +Q ++  ++++++  + ++ ++A+D D  N  N  I + + +   P     F +   +G
Sbjct  1085  DRQKYVENVKQDASIGTNILRVSASDEDADN--NGAIVYTLSAPHSPNDLEYFEIQPESG  1142

Query  168   EVRTLTNSLDREQASSYRLVVSGADKDGEGLSTQCECNIKVKDVNDNFPMFRDSQYSA-R  226
              +  L   LDRE   +Y+L      DK     LS   E  I V D  +N P++ ++ Y
Sbjct  1143  WI-VLKKPLDRE---TYKLEAMAQDKGYPPLSRTVEVQIDVVDRANNPPVWDNTVYGPIY  1198

Query  227   IEENI-LSSELLRFQVTDLDEEYTDNWLAVYFFTSG-----NEGNWFEIQTDPRTNE--G  278
             ++EN+ +  +++  + +   E    N    Y    G     N+ + F +Q  P   +
Sbjct  1199  VKENMPVGGKVVSIKASSGIE---GNPTVFYRLMPGSTAQTNKLHTFYLQQRPDNGDTWA  1255

Query  279   ILKVVKALDYEQLQSVKLSIAVKNKAEFHQSVISRYRVQSTPVTIQVINVREGIAFRPAS  338
              +KV   LD+E ++   L++ V+N          ++        V IQ+ +V + I
Sbjct  1256  DIKVNHPLDFETIKEYNLTVRVENNG-------AQQLASEATVYIQLEDVNDEIPLFTER  1308

Query  339   KTFTVQKGISSKKLVDYILGTYQAIDEDTNKAASNVKYVM---GRNDG-GYLMIDSKTAE  394
             +  TV +G      V  +     AID+D     + V Y +    RN+G  +  I+ ++ E
Sbjct  1309  EQETVLEGEPIGTKVTQV----NAIDKDGTFPNNQVYYYIVDSPRNEGKDFFEINLQSGE  1364

Query  395   I  395
             I
Sbjct  1365  I  1365

 Score = 75.5 bits (184),  Expect = 2e-12, Method: Compositional matrix adjust.
 Identities = 103/418 (25%), Positives = 174/418 (42%), Gaps = 50/418 (12%)

Query  50    IFVVDKNTGDINITAIVDREETPSFLITCRAL----------NAQGLDVEKPLILTVKIL  99
              F ++  +G+I    + DRE+  ++ +   A           N Q   V K   + + I
Sbjct  1355  FFEINLQSGEIFTKVVFDREKQGAYALEVEARDGAPSARPNSNQQPNSVTK--FIRIGIA  1412

Query  100   DINDNPPVFSQQIFMGEIEENSASNSLVMILNATDADEPNHLNSKIAFKIVSQEPAGTPM  159
             D NDNPP F + ++  E++EN       V+ + A D DE    +S+I ++I S    G
Sbjct  1413  DKNDNPPYFDKALYEAEVDENEDIQHTVLTVTAKDHDE----SSRIRYEITSGNIGGA--  1466

Query  160   FLLSRNTGEVRTLTNSLDREQASSYRLVVSGADKDGEGLSTQCECNIKVKDVNDNFPMFR  219
             F +   TG +  +  +LD E    Y L +  +D    E  +       I VKDVND  P F
Sbjct  1467  FAVKNMTGAI-YVAGALDYETRKRYYLTLVASDSLNENETIIV---INVKDVNDMPPSFP  1522

Query  220   DSQYSARIEENI-LSSELLRFQVTDLDEEYTDNWLAVYFFTSG--NEGNWFEIQTDPRTN  276
                Y   ++E + +   +++   TD D++   N   VYF T     +  N     + D
```

```
Sbjct  1523  QQVYERTMDEELAVPFPIMQVTATDGDKDRPQN--IVYFLTGQGIDPDNPANSKFDINRT  1580

Query  277   EGILKVVKALDYEQLQS-VKLSIAVKNKAEFHQSVISRYRVQSTPVTIQVINVREGIAFR  335
              G + V+K LD +Q     +    V  + E  + ++    VQ     + + ++ +
Sbjct  1581  TGEIFVLKPLDRDQPNGRPQWRFTVFAQDEGGEGLVGYADVQ-----VNLKDINDNAPIF  1635

Query  336   PASKTFTVQKGISSKKLVDYILGTYQAIDEDTNKAASNVK--YVMGRN------DGGYLM  387
             P    F     ++        ++ T  A+D D     +N K  Y + +N
Sbjct  1636  PQGVYF---GNVTENGTAGMVVMTMTAVDYDDPNEGTNAKLIYSIEKNVIEEETGSPIFE  1692

Query  388   IDSKTAEIK-FVKNMNRDSTFIVNKTITAEVLAIDEYTGKTSTGTVYVRVPDFNDNCP  444
             I++ T  IK  V  ++R+ T        + +V+A+D   G   TGT  +RV D ND  P
Sbjct  1693  IEADTGVIKTAVCCLDRERT----PDYSIQVVAMDG-GGLKGTGTASIRVKDINDMPP  1745

 Score = 55.5 bits (132),  Expect = 3e-06, Method: Compositional matrix adjust.
 Identities = 49/158 (31%), Positives = 78/158 (49%), Gaps = 24/158 (15%)

Query  102   NDNPPVFSQQIFMGEIEENSASNSLVMILNATDADEPNHLNSKIAFKIVSQEPAGTP--M  159
             ND  P FSQQ++   ++EN+  N+LV  + A+D D  N     + F  V     GT
Sbjct  858   NDEEPKFSQQVYTPNVDENAGPNTLVTTVVASDKDGDN-----VRFGFVG---GGTSSGQ  909

Query  160   FLLSRNTGEVRTLTN--SLDREQASSYRLVVSGADK-------DGEGLSTQCECNIKVKD  210
             F++   TG +R  +   SLDR++   Y L V+  D         D    ++     + + D
Sbjct  910   FVIEEITGVIRLHSKAISLDRDK---YELNVTAMDDGSCCVNGDATIHTSTAVVVVFITD  966

Query  211   VNDNFPMFRD-SQYSARIEENILS-SELLRFQVTDLDE  246
             VNDN P+F+D   Y  ++EE   + S +++ Q TD D+
Sbjct  967   VNDNKPVFKDCGTYYPKVEEGAPNGSPVIKVQATDEDK  1004

 Score = 53.5 bits (127),  Expect = 1e-05, Method: Compositional matrix adjust.
 Identities = 70/306 (23%), Positives = 126/306 (41%), Gaps = 30/306 (10%)

Query  20    PIAKIT-SDYQATQKITYRI---SGVGIDQPPFGIFVVDKNTGDINITAIVDRE---ETP  72
             PI  +T  D   T K  Y++   SG G D+  F +   +  TG + I   +D E   ++
Sbjct  1768  PILTVTVHDEDETNKFQYKVIDNSGYGADK--FTMVRNNDGTGSLKIVQPLDYEDPLQSN  1825

Query  73    SFLITCRALNAQGLDVEKPLI------LTVKILDINDNPPVFSQQIFMGEIEENSASNSL  126
              F    + +N +G D +           + VK+ DINDN P F +      + EN+
Sbjct  1826  GFRFRIQ-VNDKGEDNDNDKYHVAYSWVVVKLRDINDNKPQFERPNIEVSVYENADVGKT  1884

Query  127   VMILNATDADEPNHLNSKIAFKIVSQEPAGTPMFLLSRNTGEVRTLTNSLDREQASSYRL  186
             +    ATD D+     SK+++ I   + +        S N     T+  +LDRE    +++
Sbjct  1885  LETFKATDPDQGG--KSKVSYAI---DRSSDRQRQFSINQEGTVTIQRNLDREVTPRHQV  1939

Query  187   VVSGADKDGEGLSTQCECNIKVKDVNDNFPMFRDSQYSARIEENILSSELLRFQVTDLDE  246
              +   D      +       + V+D+NDN P F    Y   + E++   +++    TD D+
Sbjct  1940  KILAIDDGIPPKTATATLTVIVQDINDNPPKFL-KDYRPVLPEHVPPRKVVEILATDDDD  1998

Query  247   EYTDNWLAVYFFTSGNEGNW----FEIQTDPRTNEG----ILKVVKALDYEQLQSVKLSI  298
                N     F       +     F+++ D +   G    I+  +++ D EQ +   + I
Sbjct  1999  RSKSNGPPFQFRLDPGADDIIRASFKVEQDQKGANGDGMAIVSSLRSFDREQQKEYLIPI  2058

Query  299   AVKNKA  304
              +K+
Sbjct  2059  VIKDHG  2064

 Score = 51.6 bits (122),  Expect = 5e-05, Method: Compositional matrix adjust.
 Identities = 97/398 (24%), Positives = 163/398 (41%), Gaps = 54/398 (14%)

Query  106   PVFSQQIFMGEIEENSASNSLVMILNATDADEPNHLNSKIAFKIVSQ-EPAGTPMFLLSR  164
             P F     +  EI EN   +S ++ + A       +  + +I + + +Q + AGT  F +
Sbjct  655   PQFYMPSYEAEIPENQKKDSDIISVKAK-----SFADREIRYTLKAQGQGAGT--FNIGP  707

Query  165   NTGEVRTLTNSLDRE---QASSYRLVVSGADKDGEGLSTQCECNIKVKDVNDNFPMFRDS  221
              +G V+ L   LD E   Q   Y LVV+ A +D  G ST  E  I+V DVNDN P F
Sbjct  708   TSGIVK-LAKELDFEDLRQPHVYSLVVT-ATEDSGGFSTSVELTIRVTDVNDNAPKFELP  765

Query  222   QYSAR-IEENI-LSSELLRFQVTDLDEEYTDNWLAVYFFTSGNEGNWFEIQTDPRTNEGI  279
```

```
               Y A  ++E+I L + +L+ +  D D         + +  S +    F + ++     GI
Sbjct  766   DYQAHNVDEDIPLGTSILKVKAMDSDSGAN---AEIEYLVSDDH---FAVDSN-----GI  814

Query  280   LKVVKALDYEQLQS-VKLSIAVKNKAEFHQSVISRYRVQSTPVTIQVINVREGIAFRPAS  338
             +   K LD +   +  +  +  K+K E  ++  +  RV +            E   F
Sbjct  815   IVNNKQLDADNNNAYYEFVVTAKDKGEPAKTGTATVRVYTKNKN------DEEPKFSQQV  868

Query  339   KTFTVQKGISSKKLVDYILGTYQAIDEDTNKAASNVK--YVMGRNDGGYLMIDSKTAEIK  396
              T  V +      LV  ++          ++K   NV+  +V G    G  +I+  T  I+
Sbjct  869   YTPNVDENAGPNTLVTTVVA--------SDKDGDNVRFGFVGGGTSSGQFVIEEITGVIR  920

Query  397   FVK---NMNRDSTFI-VNKTITAEVLAIDEYTGKTSTGTVYVRVPDFNDN------CPTA  446
                   +++RD   + V             + T  TST  V V + D NDN      C T
Sbjct  921   LHSKAISLDRDKYELNVTAMDDGSCCVNGDATIHTSTAVVVVFITDVNDNKPVFKDCGTY  980

Query  447   VLE-KDAVCSSSPSVVVSARTLNNRYTGPYTFALEDQP  483
               + ++   + SP + V A   +    G   +++  QP
Sbjct  981   YPKVEEGAPNGSPVIKVQATDEDKGVNGQVKYSIVQQP  1018

 Score = 44.3 bits (103),  Expect = 0.010, Method: Compositional matrix adjust.
 Identities = 48/199 (24%), Positives = 73/199 (37%), Gaps = 19/199 (10%)

Query  27    DYQATQKITYRISGVGIDQPPFGIFVVDKNTGDINITAIVDREETPSFLITCRALNAQGL  86
             D     K++Y I      Q  F I       G + I   +DRE TP   +   A++
Sbjct  1894  DQGGKSKVSYAIDRSSDRQRQFSI----NQEGTVTIQRNLDREVTPRHQVKILAIDDGIP  1949

Query  87    DVEKPLILTVKILDINDNPPVFSQQIFMGEIEENSASNSLVMILNATDADEPNHLNSKIA  146
                    LTV + DINDNPP F +  +   + E+     +V IL   D D
Sbjct  1950  PKTATATLTVIVQDINDNPPKFLKD-YRPVLPEHVPPRKVVEILATDDDDRSKSNGPPFQ  2008

Query  147   FKIVSQEPAGTPMFLLSRNTGEVRTLTN-----------SLDREQASSYRLVVSGADKDG  195
             F++   +P    +   S    + +   N           S DREQ   Y + +   D
Sbjct  2009  FRL---DPGADDIIRASFKVEQDQKGANGDGMAIVSSLRSFDREQQKEYLIPIVIKDHGN  2065

Query  196   EGLSTQCECNIKVKDVNDN  214
               ++       + + DVNDN
Sbjct  2066  PAMTGTSTLTVVIGDVNDN  2084

 Score = 39.3 bits (90),  Expect = 0.33, Method: Compositional matrix adjust.
 Identities = 54/242 (22%), Positives = 102/242 (42%), Gaps = 22/242 (9%)

Query  4    KFAKPCREGE--DNSKRNP-IAKITSDYQATQKITYRISGVGIDQPPFGIFVVDKNTGDI  60
            +F  P  E E  +N K++  I  + +   A ++I Y +   G      G F +   +G +
Sbjct  656  QFYMPSYEAEIPENQKKDSDIISVKAKSFADREIRYTLKAQGQGA---GTFNIGPTSGIV  712

Query  61   NITAIVDREE--TPSF--LITCRALNAQGLDVEKPLILTVKILDINDNPPVFSQQIFMGE  116
              +   +D E+   P    L+     ++ G       + LT+++ D+NDN P F    +
Sbjct  713  KLAKELDFEDLRQPHVYSLVVTATEDSGGFSTS--VELTIRVTDVNDNAPKFELPDYQAH  770

Query  117  -IEENSASNSLVMILNATDADEPNHLNSKIAFKIVSQEPAGTPMFLLSRNTGEVRTLTNS  175
             ++E+    + ++ +  A D+D  +  N++I + +VS +       F +  N   V
Sbjct  771  NVDEDIPLGTSILKVKAMDSD--SGANAEIEY-LVSDD-----HFAVDSNGIIVNNKQLD  822

Query  176  LDREQASSYRLVVSGADKDGEGLSTQCECNIKVKDVNDNFPMFRDSQYSARIEENILSSE  235
             D   A  Y  VV+  DK      +       +  K+ ND  P F    Y+  ++EN   +
Sbjct  823  ADNNNA-YYEFVVTAKDKGEPAKTGTATVRVYTKNKNDEEPKFSQQVYTPNVDENAGPNT  881

Query  236  LL  237
            L+
Sbjct  882  LV  883

 Score = 39.3 bits (90),  Expect = 0.35, Method: Compositional matrix adjust.
 Identities = 89/389 (23%), Positives = 149/389 (38%), Gaps = 62/389 (16%)

Query  75   LITCRALNAQGLDVEKPLILTVKILDINDNPPVFSQQIF--MGEIEENSASNSLVMILNA  132
            +IT +  NA GL       + + + D+ND PP F  +       ++ N+  N+ V  L A
Sbjct  517  IITNQGHNA-GLKYTDNQRVIIHVKDVNDEPPYFINRPLPMQAVVQLNAPPNTPVFTLQA  575
```

```
Query  133  TDADEPNHLNSKIAFKIVSQEPAGTPMFLLSRNTGEVRTLTNSLDREQASSYRLVVSGAD  192
             D D      + I + IV     G   F +   +G VRT    L  +    Y L V   D
Sbjct  576  RDPD----TDHNIHYFIVRDRTGG--RFEVDERSGVVRTRGTDL-FQLDMEYVLYVKAED  628

Query  193  KDGE----GLSTQCECNIKVKDVNDNFPMFRDSQYSARIEEN-ILSSELLRFQVTDLDEE  247
            ++G+       +  E  + +       P F    Y A I EN    S+++  +     +
Sbjct  629  QNGKVDDRRFQSTPEERLSIVG-GKRAPQFYMPSYEAEIPENQKKDSDIISVKA----KS  683

Query  248  YTDNWLAVYFFTSGNEGNWFEIQTDPRTNEGILKVVKALDYEQLQSVKLS----IAVKNK  303
            + D  +       G     F I        GI+K+ K LD+E L+   +      A ++
Sbjct  684  FADREIRYTLKAQGQGAGTFNIGP----TSGIVKLAKELDFEDLRQPHVYSLVVTATEDS  739

Query  304  AEFHQSVISRYRVQSTPVTIQVINVREGIAFRPASKTFTVQKGISSKKLVDYILGT----  359
              F  SV          +TI+V +V +       +  F +    +     D  LGT
Sbjct  740  GGFSTSV---------ELTIRVTDVNDN------APKFELPDYQAHNVDEDIPLGTSILK  784

Query  360  YQAIDEDTNKAASNVKYVMGRN----DGGYLMIDSKTAEIKFVKNMNRDSTFIVNKTITA  415
             +A+D D+  A + ++Y++  +    D   +++++K  +     N N    F+V
Sbjct  785  VKAMDSDSG-ANAEIEYLVSDDHFAVDSNGIIVNNKQLD---ADNNNAYYEFVVTAKDKG  840

Query  416  EVLAIDEYTGKTSTGTVYVRVPDFNDNCP  444
            E         KT T TV V   + ND  P
Sbjct  841  E-------PAKTGTATVRVYTKNKNDEEP  862

>AGAMI1_007196.P14110 | transcript=AGAMI1_007196.R14110 | gene=AGAMI1_007196
| organism=Anopheles_gambiae_Ifakara | gene_product=neural-cadherin
| transcript_product=neural-cadherin,
transcript variant X14 | location=AgamI1_3RL:41252608-41455387(+)
| protein_length=3099 | sequence_SO=chromosome | SO=protein_coding_gene
| is_pseudo=false
Length=3099

 Score = 146 bits (369),  Expect = 4e-35, Method: Compositional matrix adjust.
 Identities = 128/446 (29%), Positives = 217/446 (49%), Gaps = 36/446 (8%)

Query  20    PIAKITS---DYQATQKITYRISGVGID--QPPFGIFVVDKNTGDINITAIVDREET---  71
             PI ++T+   D    Q I Y ++G GID   P    F +++ TG+I +   +DR++
Sbjct  1539  PIMQVTATDGDKDRPQNIVYFLTGQGIDPDNPANSKFDINRTTGEIFVLKPLDRDQPNGR  1598

Query  72    PSFLITCRALNAQGLDVEKPLILTVKILDINDNPPVFSQQIFMGEIEENSASNSLVMILN  131
             P +  T  A +  G  +     + V + DINDN P+F Q ++ G + EN  +  +VM +
Sbjct  1599  PQWRFTVFAQDEGGEGLVGYADVQVNLKDINDNAPIFPQGVYFGNVTENGTAGMVVMTMT  1658

Query  132   ATDADEPNH-LNSKIAFKI---VSQEPAGTPMFLLSRNTGEVRTLTNSLDREQASSYRLV  187
             A D D+PN   N+K+ + I   V +E  G+P+F +  +TG ++T    LDRE+   Y +
Sbjct  1659  AVDYDDPNEGTNAKLIYSIEKNVIEEETGSPIFEIEADTGVIKTAVCCLDRERTPDYSIQ  1718

Query  188   VSGADKDGEGLSTQCECNIKVKDVNDNFPMFRDSQYSARIEENILSS--ELLRFQVTDLD  245
             V     DG GL      +I+VKD+ND  P F   ++   ++E   ++  E+    VT  D
Sbjct  1719  VVAM--DGGGLKGTGTASIRVKDINDMPPQFTKDEWFTEVDETDGTNLPEMPILTVTVHD  1776

Query  246   EEYTDNWLAVYFFTSGNEGNWFEIQTDPRTNEGI--LKVVKALDYE---QLQSVKLSIAV  300
             E+ T+ +       SG    + F +    R N+G   LK+V+ LDYE   Q    +  I V
Sbjct  1777  EDETNKFQYKVIDNSGYGADKFTMV---RNNDGTGSLKIVQPLDYEDPLQSNGFRFRIQV  1833

Query  301   KNKAEFHQSVISRYRVQSTPVTIQVINVREGIAFRPASKTFTVQKGISSKKLVDYILGTY  360
              +K E + +   +Y V  + V +++ ++ +    +P  +   ++  +     V   L T+
Sbjct  1834  NDKGEDNDN--DKYHVAYSWVVVKLRDINDN---KPQFERPNIEVSVYENADVGKTLETF  1888

Query  361   QAIDEDTNKAASNVKYVMGR-NDGGYLMIDSKTAEIKFVKNMNRDSTFIVNKTITAEVLA  419
             +A D D     S V Y + R +D       ++   +   +N++R+    V      ++LA
Sbjct  1889  KATDPDQG-GKSKVSYAIDRSSDRQRQFSINQEGTVTIQRNLDRE----VTPRHQVKILA  1943

Query  420   IDE-YTGKTSTGTVYVRVPDFNDNCP  444
             ID+    KT+T T+ V V D NDN P
Sbjct  1944  IDDGIPPKTATATLTVIVQDINDNPP  1969

 Score = 75.9 bits (185),  Expect = 2e-12, Method: Compositional matrix adjust.
 Identities = 89/361 (25%), Positives = 167/361 (46%), Gaps = 36/361 (10%)
```

```
Query  51    FVVDKNTGDINITAIVDRE-ETPSFL-ITCRALNAQGLDVEKPLILTVKILDINDNPPVF  108
             F VD+ TG+++   + DRE +   F+ +T +A +     +E     TV+I D+NDNPP+F
Sbjct  1025  FTVDEETGEVSTNKVFDREGDDGKFVSVTVKATDQGEPSLEGVCSFTVEITDVNDNPPLF  1084

Query  109   SQQIFMGEIEENSASNSLVMILNATDADEPNHLNSKIAFKIVS-QEPAGTPMFLLSRNTG  167
              +Q ++  ++++++  + ++ ++A+D D  N  N  I + + +   P     F +   +G
Sbjct  1085  DRQKYVENVKQDASIGTNILRVSASDEDADN--NGAIVYTLSAPHSPNDLEYFEIQPESG  1142

Query  168   EVRTLTNSLDREQASSYRLVVSGADKDGEGLSTQCECNIKVKDVNDNFPMFRDSQYSA-R  226
              +  L   LDRE   +Y+L      DK    LS   E  I V D  +N P++ ++ Y
Sbjct  1143  WI-VLKKPLDRE---TYKLEAMAQDKGYPPLSRTVEVQIDVVDRANNPPVWDNTVYGPIY  1198

Query  227   IEENI-LSSELLRFQVTDLDEEYTDNWLAVYFFTSG-----NEGNWFEIQTDPRTNE--G  278
             ++EN+ +  +++  + +   E    N    Y    G     N+ + F +Q  P   +
Sbjct  1199  VKENMPVGGKVVSIKASSGIE---GNPTVFYRLMPGSTAQTNKLHTFYLQQRPDNGDTWA  1255

Query  279   ILKVVKALDYEQLQSVKLSIAVKNKAEFHQSVISRYRVQSTPVTIQVINVREGIAFRPAS  338
              +KV   LD+E ++   L++ V+N          ++       V IQ+ +V + I
Sbjct  1256  DIKVNHPLDFETIKEYNLTVRVENNG-------AQQLASEATVYIQLEDVNDEIPLFTER  1308

Query  339   KTFTVQKGISSKKLVDYILGTYQAIDEDTNKAASNVKYVM---GRNDG-GYLMIDSKTAE  394
             +  TV +G      V  +     AID+D     + V Y +    RN+G  +  I+ ++ E
Sbjct  1309  EQETVLEGEPIGTKVTQV----NAIDKDGTFPNNQVYYYIVDSPRNEGKDFFEINLQSGE  1364

Query  395   I  395
             I
Sbjct  1365  I  1365

 Score = 75.5 bits (184),  Expect = 2e-12, Method: Compositional matrix adjust.
 Identities = 103/418 (25%), Positives = 174/418 (42%), Gaps = 50/418 (12%)

Query  50    IFVVDKNTGDINITAIVDREETPSFLITCRAL----------NAQGLDVEKPLILTVKIL  99
              F ++  +G+I    + DRE+  ++ +   A            N Q   V K   + + I
Sbjct  1355  FFEINLQSGEIFTKVVFDREKQGAYALEVEARDGAPSARPNSNQQPNSVTK--FIRIGIA  1412

Query  100   DINDNPPVFSQQIFMGEIEENSASNSLVMILNATDADEPNHLNSKIAFKIVSQEPAGTPM  159
             D NDNPP F + ++  E++EN      V+ + A D DE    +S+I ++I S    G
Sbjct  1413  DKNDNPPYFDKALYEAEVDENEDIQHTVLTVTAKDHDE----SSRIRYEITSGNIGGA--  1466

Query  160   FLLSRNTGEVRTLTNSLDREQASSYRLVVSGADKDGEGLSTQCECNIKVKDVNDNFPMFR  219
             F +   TG +  +  +LD E    Y L +  +D   E  +       I VKDVND  P F
Sbjct  1467  FAVKNMTGAI-YVAGALDYETRKRYYLTLVASDSLNENETIIV---INVKDVNDMPPSFP  1522

Query  220   DSQYSARIEENI-LSSELLRFQVTDLDEEYTDNWLAVYFFTSG--NEGNWFEIQTDPRTN  276
                Y   ++E + +   +++   TD D++   N   VYF T    +  N    + D
Sbjct  1523  QQVYERTMDEELAVPFPIMQVTATDGDKDRPQN--IVYFLTGQGIDPDNPANSKFDINRT  1580

Query  277   EGILKVVKALDYEQLQS-VKLSIAVKNKAEFHQSVISRYRVQSTPVTIQVINVREGIAFR  335
              G + V+K LD +Q      +    V  + E  + ++    VQ     + + ++ +
Sbjct  1581  TGEIFVLKPLDRDQPNGRPQWRFTVFAQDEGGEGLVGYADVQ-----VNLKDINDNAPIF  1635

Query  336   PASKTFTVQKGISSKKLVDYILGTYQAIDEDTNKAASNVK--YVMGRN------DGGYLM  387
             P    F     ++        ++ T  A+D D     +N K  Y + +N
Sbjct  1636  PQGVYF---GNVTENGTAGMVVMTMTAVDYDDPNEGTNAKLIYSIEKNVIEEETGSPIFE  1692

Query  388   IDSKTAEIK-FVKNMNRDSTFIVNKTITAEVLAIDEYTGKTSTGTVYVRVPDFNDNCP  444
             I++ T  IK  V  ++R+ T        + +V+A+D   G   TGT  +RV D ND  P
Sbjct  1693  IEADTGVIKTAVCCLDRERT----PDYSIQVVAMDG-GGLKGTGTASIRVKDINDMPP  1745

 Score = 55.5 bits (132),  Expect = 3e-06, Method: Compositional matrix adjust.
 Identities = 49/158 (31%), Positives = 78/158 (49%), Gaps = 24/158 (15%)

Query  102   NDNPPVFSQQIFMGEIEENSASNSLVMILNATDADEPNHLNSKIAFKIVSQEPAGTP--M  159
             ND  P FSQQ++   ++EN+  N+LV  + A+D D  N     + F  V     GT
Sbjct  858   NDEEPKFSQQVYTPNVDENAGPNTLVTTVVASDKDGDN-----VRFGFVG---GGTSSGQ  909

Query  160   FLLSRNTGEVRTLTN--SLDREQASSYRLVVSGADK-------DGEGLSTQCECNIKVKD  210
             F++   TG +R  +   SLDR++   Y L V+  D         D    ++     + + D
```

```
Sbjct  910   FVIEEITGVIRLHSKAISLDRDK---YELNVTAMDDGSCCVNGDATIHTSTAVVVVFITD  966

Query  211   VNDNFPMFRD-SQYSARIEENILS-SELLRFQVTDLDE  246
             VNDN P+F+D   Y  ++EE   + S +++ Q TD D+
Sbjct  967   VNDNKPVFKDCGTYYPKVEEGAPNGSPVIKVQATDEDK  1004

 Score = 53.5 bits (127),  Expect = 1e-05, Method: Compositional matrix adjust.
 Identities = 70/306 (23%), Positives = 126/306 (41%), Gaps = 30/306 (10%)

Query  20    PIAKIT-SDYQATQKITYRI---SGVGIDQPPFGIFVVDKNTGDINITAIVDRE---ETP  72
             PI  +T  D   T K  Y++   SG G D+  F +   +  TG + I   +D E   ++
Sbjct  1768  PILTVTVHDEDETNKFQYKVIDNSGYGADK--FTMVRNNDGTGSLKIVQPLDYEDPLQSN  1825

Query  73    SFLITCRALNAQGLDVEKPLI------LTVKILDINDNPPVFSQQIFMGEIEENSASNSL  126
              F    + +N +G D +          + VK+ DINDN P F +      + EN+
Sbjct  1826  GFRFRIQ-VNDKGEDNDNDKYHVAYSWVVVKLRDINDNKPQFERPNIEVSVYENADVGKT  1884

Query  127   VMILNATDADEPNHLNSKIAFKIVSQEPAGTPMFLLSRNTGEVRTLTNSLDREQASSYRL  186
             +    ATD D+     SK+++ I   + +       S N     T+  +LDRE    +++
Sbjct  1885  LETFKATDPDQGG--KSKVSYAI---DRSSDRQRQFSINQEGTVTIQRNLDREVTPRHQV  1939

Query  187   VVSGADKDGEGLSTQCECNIKVKDVNDNFPMFRDSQYSARIEENILSSELLRFQVTDLDE  246
              +   D      +      + V+D+NDN P F    Y   + E++   +++    TD D+
Sbjct  1940  KILAIDDGIPPKTATATLTVIVQDINDNPPKFL-KDYRPVLPEHVPPRKVVEILATDDDD  1998

Query  247   EYTDNWLAVYFFTSGNEGNW----FEIQTDPRTNEG----ILKVVKALDYEQLQSVKLSI  298
                 N     F       +     F+++ D +   G    I+  +++ D EQ +   + I
Sbjct  1999  RSKSNGPPFQFRLDPGADDIIRASFKVEQDQKGANGDGMAIVSSLRSFDREQQKEYLIPI  2058

Query  299   AVKNKA  304
              +K+
Sbjct  2059  VIKDHG  2064

 Score = 51.6 bits (122),  Expect = 5e-05, Method: Compositional matrix adjust.
 Identities = 97/398 (24%), Positives = 163/398 (41%), Gaps = 54/398 (14%)

Query  106   PVFSQQIFMGEIEENSASNSLVMILNATDADEPNHLNSKIAFKIVSQ-EPAGTPMFLLSR  164
             P F    +  EI EN   +S ++ + A      +  + +I + + +Q + AGT  F +
Sbjct  655   PQFYMPSYEAEIPENQKKDSDIISVKAK-----SFADREIRYTLKAQGQGAGT--FNIGP  707

Query  165   NTGEVRTLTNSLDRE---QASSYRLVVSGADKDGEGLSTQCECNIKVKDVNDNFPMFRDS  221
              +G V+ L   LD E   Q   Y LVV+ A +D  G ST  E  I+V DVNDN P F
Sbjct  708   TSGIVK-LAKELDFEDLRQPHVYSLVVT-ATEDSGGFSTSVELTIRVTDVNDNAPKFELP  765

Query  222   QYSAR-IEENI-LSSELLRFQVTDLDEEYTDNWLAVYFFTSGNEGNWFEIQTDPRTNEGI  279
              Y A  ++E+I L + +L+ +  D D         + +  S +    F + ++     GI
Sbjct  766   DYQAHNVDEDIPLGTSILKVKAMDSDSGAN---AEIEYLVSDDH---FAVDSN-----GI  814

Query  280   LKVVKALDYEQLQS-VKLSIAVKNKAEFHQSVISRYRVQSTPVTIQVINVREGIAFRPAS  338
             +   K LD +   +  +  +  K+K E  ++  +  RV +           E   F
Sbjct  815   IVNNKQLDADNNNAYYEFVVTAKDKGEPAKTGTATVRVYTKNKN------DEEPKFSQQV  868

Query  339   KTFTVQKGISSKKLVDYILGTYQAIDEDTNKAASNVK--YVMGRNDGGYLMIDSKTAEIK  396
              T  V +      LV  ++         ++K   NV+  +V G    G  +I+  T  I+
Sbjct  869   YTPNVDENAGPNTLVTTVVA--------SDKDGDNVRFGFVGGGTSSGQFVIEEITGVIR  920

Query  397   FVK---NMNRDSTFI-VNKTITAEVLAIDEYTGKTSTGTVYVRVPDFNDN------CPTA  446
                   +++RD   + V            + T  TST  V V + D NDN      C T
Sbjct  921   LHSKAISLDRDKYELNVTAMDDGSCCVNGDATIHTSTAVVVVFITDVNDNKPVFKDCGTY  980

Query  447   VLE-KDAVCSSSPSVVVSARTLNNRYTGPYTFALEDQP  483
               + ++   + SP + V A   +    G   +++  QP
Sbjct  981   YPKVEEGAPNGSPVIKVQATDEDKGVNGQVKYSIVQQP  1018

 Score = 44.3 bits (103),  Expect = 0.010, Method: Compositional matrix adjust.
 Identities = 48/199 (24%), Positives = 73/199 (37%), Gaps = 19/199 (10%)

Query  27    DYQATQKITYRISGVGIDQPPFGIFVVDKNTGDINITAIVDREETPSFLITCRALNAQGL  86
```

```
               D     K++Y I      Q  F I       G + I   +DRE TP   +   A++
Sbjct  1894  DQGGKSKVSYAIDRSSDRQRQFSI----NQEGTVTIQRNLDREVTPRHQVKILAIDDGIP  1949

Query  87    DVEKPLILTVKILDINDNPPVFSQQIFMGEIEENSASNSLVMILNATDADEPNHLNSKIA  146
                     LTV + DINDNPP F +  +   + E+     +V IL   D D
Sbjct  1950  PKTATATLTVIVQDINDNPPKFLKD-YRPVLPEHVPPRKVVEILATDDDDRSKSNGPPFQ  2008

Query  147   FKIVSQEPAGTPMFLLSRNTGEVRTLTN-----------SLDREQASSYRLVVSGADKDG  195
             F++   +P    +   S    + +   N           S DREQ   Y + +   D
Sbjct  2009  FRL---DPGADDIIRASFKVEQDQKGANGDGMAIVSSLRSFDREQQKEYLIPIVIKDHGN  2065

Query  196   EGLSTQCECNIKVKDVNDN  214
               ++      + + DVNDN
Sbjct  2066  PAMTGTSTLTVVIGDVNDN  2084

 Score = 39.3 bits (90),  Expect = 0.33, Method: Compositional matrix adjust.
 Identities = 54/242 (22%), Positives = 102/242 (42%), Gaps = 22/242 (9%)

Query  4    KFAKPCREGE--DNSKRNP-IAKITSDYQATQKITYRISGVGIDQPPFGIFVVDKNTGDI  60
            +F  P  E E  +N K++  I  + +   A ++I Y +   G      G F +   +G +
Sbjct  656  QFYMPSYEAEIPENQKKDSDIISVKAKSFADREIRYTLKAQGQGA---GTFNIGPTSGIV  712

Query  61   NITAIVDREE--TPSF--LITCRALNAQGLDVEKPLILTVKILDINDNPPVFSQQIFMGE  116
              +   +D E+   P    L+     ++ G      + LT+++ D+NDN P F    +
Sbjct  713  KLAKELDFEDLRQPHVYSLVVTATEDSGGFSTS--VELTIRVTDVNDNAPKFELPDYQAH  770

Query  117  -IEENSASNSLVMILNATDADEPNHLNSKIAFKIVSQEPAGTPMFLLSRNTGEVRTLTNS  175
             ++E+    + ++ + A D+D  +  N++I + +VS +      F +  N   V
Sbjct  771  NVDEDIPLGTSILKVKAMDSD--SGANAEIEY-LVSDD-----HFAVDSNGIIVNNKQLD  822

Query  176  LDREQASSYRLVVSGADKDGEGLSTQCECNIKVKDVNDNFPMFRDSQYSARIEENILSSE  235
             D   A  Y  VV+  DK      +       +  K+ ND  P F    Y+  ++EN   +
Sbjct  823  ADNNNA-YYEFVVTAKDKGEPAKTGTATVRVYTKNKNDEEPKFSQQVYTPNVDENAGPNT  881

Query  236  LL  237
            L+
Sbjct  882  LV  883

 Score = 39.3 bits (90),  Expect = 0.34, Method: Compositional matrix adjust.
 Identities = 89/389 (23%), Positives = 149/389 (38%), Gaps = 62/389 (16%)

Query  75   LITCRALNAQGLDVEKPLILTVKILDINDNPPVFSQQIF--MGEIEENSASNSLVMILNA  132
            +IT +  NA GL       + + + D+ND PP F  +        ++ N+  N+ V  L A
Sbjct  517  IITNQGHNA-GLKYTDNQRVIIHVKDVNDEPPYFINRPLPMQAVVQLNAPPNTPVFTLQA  575

Query  133  TDADEPNHLNSKIAFKIVSQEPAGTPMFLLSRNTGEVRTLTNSLDREQASSYRLVVSGAD  192
             D D     +  I + IV     G   F +   +G VRT    L  +    Y L V   D
Sbjct  576  RDPD----TDHNIHYFIVRDRTGG--RFEVDERSGVVRTRGTDL-FQLDMEYVLYVKAED  628

Query  193  KDGE----GLSTQCECNIKVKDVNDNFPMFRDSQYSARIEEN-ILSSELLRFQVTDLDEE  247
            ++G+        +  E  + +        P F    Y A I EN    S+++  +     +
Sbjct  629  QNGKVDDRRFQSTPEERLSIVG-GKRAPQFYMPSYEAEIPENQKKDSDIISVKA----KS  683

Query  248  YTDNWLAVYFFTSGNEGNWFEIQTDPRTNEGILKVVKALDYEQLQSVKLS----IAVKNK  303
            + D  +        G     F I          GI+K+ K LD+E L+   +      A ++
Sbjct  684  FADREIRYTLKAQGQGAGTFNIGP----TSGIVKLAKELDFEDLRQPHVYSLVVTATEDS  739

Query  304  AEFHQSVISRYRVQSTPVTIQVINVREGIAFRPASKTFTVQKGISSKKLVDYILGT----  359
              F  SV           +TI+V +V +        +  F +     +     D  LGT
Sbjct  740  GGFSTSV---------ELTIRVTDVNDN------APKFELPDYQAHNVDEDIPLGTSILK  784

Query  360  YQAIDEDTNKAASNVKYVMGRN----DGGYLMIDSKTAEIKFVKNMNRDSTFIVNKTITA  415
             +A+D D+  A + ++Y++  +    D    +++++K  +     N N    F+V
Sbjct  785  VKAMDSDSG-ANAEIEYLVSDDHFAVDSNGIIVNNKQLD---ADNNNAYYEFVVTAKDKG  840

Query  416  EVLAIDEYTGKTSTGTVYVRVPDFNDNCP  444
            E         KT T TV V   + ND  P
Sbjct  841  E-------PAKTGTATVRVYTKNKNDEEP  862
```

```
>ACMO_007424.P13509 | transcript=ACMO_007424.R13509 | gene=ACMO_007424
| organism=Anopheles_coluzzii_MOPTI | gene_product=neural-cadherin,
transcript variant X4 | transcript_product=neural-cadherin,
transcript variant X13 | location=CM029349:40739311-40945968(+)
| protein_length=3099 | sequence_SO=chromosome
| SO=protein_coding_gene | is_pseudo=false
Length=3099

 Score = 146 bits (369),  Expect = 4e-35, Method: Compositional matrix adjust.
 Identities = 128/446 (29%), Positives = 217/446 (49%), Gaps = 36/446 (8%)

Query  20    PIAKITS---DYQATQKITYRISGVGID--QPPFGIFVVDKNTGDINITAIVDREET---  71
             PI ++T+   D    Q I Y ++G GID   P    F +++ TG+I +   +DR++
Sbjct  1539  PIMQVTATDGDKDRPQNIVYFLTGQGIDPDNPANSKFDINRTTGEIFVLKPLDRDQPNGR  1598

Query  72    PSFLITCRALNAQGLDVEKPLILTVKILDINDNPPVFSQQIFMGEIEENSASNSLVMILN  131
             P +  T  A +  G  +     + V + DINDN P+F Q ++ G + EN  +  +VM +
Sbjct  1599  PQWRFTVFAQDEGGEGLVGYADVQVNLKDINDNAPIFPQGVYFGNVTENGTAGMVVMTMT  1658

Query  132   ATDADEPNH-LNSKIAFKI---VSQEPAGTPMFLLSRNTGEVRTLTNSLDREQASSYRLV  187
             A D D+PN   N+K+ + I   V +E  G+P+F +  +TG ++T    LDRE+   Y +
Sbjct  1659  AVDYDDPNEGTNAKLIYSIEKNVIEEETGSPIFEIEADTGVIKTAVCCLDRERTPDYSIQ  1718

Query  188   VSGADKDGEGLSTQCECNIKVKDVNDNFPMFRDSQYSARIEENILSS--ELLRFQVTDLD  245
             V     DG GL       +I+VKD+ND  P F   ++   ++E   ++  E+    VT  D
Sbjct  1719  VVAM--DGGGLKGTGTASIRVKDINDMPPQFTKDEWFTEVDETDGTNLPEMPILTVTVHD  1776

Query  246   EEYTDNWLAVYFFTSGNEGNWFEIQTDPRTNEGI--LKVVKALDYE---QLQSVKLSIAV  300
             E+ T+ +        SG    + F +    R N+G   LK+V+ LDYE   Q    +  I V
Sbjct  1777  EDETNKFQYKVIDNSGYGADKFTMV---RNNDGTGSLKIVQPLDYEDPLQSNGFRFRIQV  1833

Query  301   KNKAEFHQSVISRYRVQSTPVTIQVINVREGIAFRPASKTFTVQKGISSKKLVDYILGTY  360
              +K E + +   +Y V  + V +++ ++ +    +P  +   ++  +      V   L T+
Sbjct  1834  NDKGEDNDN--DKYHVAYSWVVVKLRDINDN---KPQFERPNIEVSVYENADVGKTLETF  1888

Query  361   QAIDEDTNKAASNVKYVMGR-NDGGYLMIDSKTAEIKFVKNMNRDSTFIVNKTITAEVLA  419
             +A D D     S V Y + R +D       ++   +   +N++R+    V       ++LA
Sbjct  1889  KATDPDQG-GKSKVSYAIDRSSDRQRQFSINQEGTVTIQRNLDRE----VTPRHQVKILA  1943

Query  420   IDE-YTGKTSTGTVYVRVPDFNDNCP  444
             ID+    KT+T T+ V V D NDN P
Sbjct  1944  IDDGIPPKTATATLTVIVQDINDNPP  1969

 Score = 75.9 bits (185),  Expect = 2e-12, Method: Compositional matrix adjust.
 Identities = 89/361 (25%), Positives = 167/361 (46%), Gaps = 36/361 (10%)

Query  51    FVVDKNTGDINITAIVDRE-ETPSFL-ITCRALNAQGLDVEKPLILTVKILDINDNPPVF  108
             F VD+ TG+++   + DRE +   F+ +T +A +      +E      TV+I D+NDNPP+F
Sbjct  1025  FTVDEETGEVSTNKVFDREGDDGKFVSVTVKATDQGEPSLEGVCSFTVEITDVNDNPPLF  1084

Query  109   SQQIFMGEIEENSASNSLVMILNATDADEPNHLNSKIAFKIVS-QEPAGTPMFLLSRNTG  167
              +Q ++  ++++++  + ++ ++A+D D  N  N  I + + +   P      F +   +G
Sbjct  1085  DRQKYVENVKQDASIGTNILRVSASDEDADN--NGAIVYTLSAPHSPNDLEYFEIQPESG  1142

Query  168   EVRTLTNSLDREQASSYRLVVSGADKDGEGLSTQCECNIKVKDVNDNFPMFRDSQYSA-R  226
              +  L   LDRE   +Y+L      DK     LS   E  I V D  +N P++ ++ Y
Sbjct  1143  WI-VLKKPLDRE---TYKLEAMAQDKGYPPLSRTVEVQIDVVDRANNPPVWDNTVYGPIY  1198

Query  227   IEENI-LSSELLRFQVTDLDEEYTDNWLAVYFFTSG-----NEGNWFEIQTDPRTNE--G  278
             ++EN+ +  +++  + +   E    N    Y    G     N+ + F +Q  P   +
Sbjct  1199  VKENMPVGGKVVSIKASSGIE---GNPTVFYRLMPGSTAQTNKLHTFYLQQRPDNGDTWA  1255

Query  279   ILKVVKALDYEQLQSVKLSIAVKNKAEFHQSVISRYRVQSTPVTIQVINVREGIAFRPAS  338
              +KV   LD+E ++   L++ V+N           ++        V IQ+ +V + I
Sbjct  1256  DIKVNHPLDFETIKEYNLTVRVENNG-------AQQLASEATVYIQLEDVNDEIPLFTER  1308

Query  339   KTFTVQKGISSKKLVDYILGTYQAIDEDTNKAASNVKYVM---GRNDG-GYLMIDSKTAE  394
             +  TV +G       V  +     AID+D     + V Y +    RN+G  +  I+ ++ E
Sbjct  1309  EQETVLEGEPIGTKVTQV----NAIDKDGTFPNNQVYYYIVDSPRNEGKDFFEINLQSGE  1364
```

```
Query  395   I  395
             I
Sbjct  1365  I  1365

 Score = 75.5 bits (184),  Expect = 2e-12, Method: Compositional matrix adjust.
 Identities = 103/418 (25%), Positives = 174/418 (42%), Gaps = 50/418 (12%)

Query  50    IFVVDKNTGDINITAIVDREETPSFLITCRAL----------NAQGLDVEKPLILTVKIL  99
              F ++  +G+I    + DRE+  ++ +   A            N Q   V K   + + I
Sbjct  1355  FFEINLQSGEIFTKVVFDREKQGAYALEVEARDGAPSARPNSNQQPNSVTK--FIRIGIA  1412

Query  100   DINDNPPVFSQQIFMGEIEENSASNSLVMILNATDADEPNHLNSKIAFKIVSQEPAGTPM  159
             D NDNPP F + ++  E++EN       V+ + A D DE    +S+I ++I S    G
Sbjct  1413  DKNDNPPYFDKALYEAEVDENEDIQHTVLTVTAKDHDE----SSRIRYEITSGNIGGA--  1466

Query  160   FLLSRNTGEVRTLTNSLDREQASSYRLVVSGADKDGEGLSTQCECNIKVKDVNDNFPMFR  219
             F +   TG +  +  +LD E    Y L +  +D   E  +       I VKDVND  P F
Sbjct  1467  FAVKNMTGAI-YVAGALDYETRKRYYLTLVASDSLNENETIIV---INVKDVNDMPPSFP  1522

Query  220   DSQYSARIEENI-LSSELLRFQVTDLDEEYTDNWLAVYFFTSG--NEGNWFEIQTDPRTN  276
                Y   ++E + +   +++   TD D++   N   VYF T    +  N    + D
Sbjct  1523  QQVYERTMDEELAVPFPIMQVTATDGDKDRPQN--IVYFLTGQGIDPDNPANSKFDINRT  1580

Query  277   EGILKVVKALDYEQLQS-VKLSIAVKNKAEFHQSVISRYRVQSTPVTIQVINVREGIAFR  335
              G + V+K LD +Q     +    V  + E  + ++    VQ     + + ++ +
Sbjct  1581  TGEIFVLKPLDRDQPNGRPQWRFTVFAQDEGGEGLVGYADVQ-----VNLKDINDNAPIF  1635

Query  336   PASKTFTVQKGISSKKLVDYILGTYQAIDEDTNKAASNVK--YVMGRN------DGGYLM  387
             P    F     ++        ++ T  A+D D     +N K  Y + +N
Sbjct  1636  PQGVYF---GNVTENGTAGMVVMTMTAVDYDDPNEGTNAKLIYSIEKNVIEEETGSPIFE  1692

Query  388   IDSKTAEIK-FVKNMNRDSTFIVNKTITAEVLAIDEYTGKTSTGTVYVRVPDFNDNCP  444
             I++ T  IK  V  ++R+ T        + +V+A+D   G   TGT  +RV D ND  P
Sbjct  1693  IEADTGVIKTAVCCLDRERT----PDYSIQVVAMDG-GGLKGTGTASIRVKDINDMPP  1745

 Score = 55.5 bits (132),  Expect = 3e-06, Method: Compositional matrix adjust.
 Identities = 49/158 (31%), Positives = 78/158 (49%), Gaps = 24/158 (15%)

Query  102   NDNPPVFSQQIFMGEIEENSASNSLVMILNATDADEPNHLNSKIAFKIVSQEPAGTP--M  159
             ND  P FSQQ++   ++EN+  N+LV  + A+D D  N     + F  V     GT
Sbjct  858   NDEEPKFSQQVYTPNVDENAGPNTLVTTVVASDKDGDN-----VRFGFVG---GGTSSGQ  909

Query  160   FLLSRNTGEVRTLTN--SLDREQASSYRLVVSGADK-------DGEGLSTQCECNIKVKD  210
             F++    TG +R  +   SLDR++    Y L V+  D         D    ++      + + D
Sbjct  910   FVIEEITGVIRLHSKAISLDRDK---YELNVTAMDDGSCCVNGDATIHTSTAVVVVFITD  966

Query  211   VNDNFPMFRD-SQYSARIEENILS-SELLRFQVTDLDE  246
             VNDN P+F+D   Y  ++EE   + S +++ Q TD D+
Sbjct  967   VNDNKPVFKDCGTYYPKVEEGAPNGSPVIKVQATDEDK  1004

 Score = 53.5 bits (127),  Expect = 1e-05, Method: Compositional matrix adjust.
 Identities = 70/306 (23%), Positives = 126/306 (41%), Gaps = 30/306 (10%)

Query  20    PIAKIT-SDYQATQKITYRI---SGVGIDQPPFGIFVVDKNTGDINITAIVDRE---ETP  72
             PI  +T  D   T K  Y++   SG G D+  F +   +  TG + I   +D E   ++
Sbjct  1768  PILTVTVHDEDETNKFQYKVIDNSGYGADK--FTMVRNNDGTGSLKIVQPLDYEDPLQSN  1825

Query  73    SFLITCRALNAQGLDVEKPLI------LTVKILDINDNPPVFSQQIFMGEIEENSASNSL  126
              F    + +N +G D +            + VK+ DINDN P F +       + EN+
Sbjct  1826  GFRFRIQ-VNDKGEDNDNDKYHVAYSWVVVKLRDINDNKPQFERPNIEVSVYENADVGKT  1884

Query  127   VMILNATDADEPNHLNSKIAFKIVSQEPAGTPMFLLSRNTGEVRTLTNSLDREQASSYRL  186
             +    ATD D+      SK+++ I   + +          S N      T+  +LDRE    +++
Sbjct  1885  LETFKATDPDQGG--KSKVSYAI---DRSSDRQRQFSINQEGTVTIQRNLDREVTPRHQV  1939

Query  187   VVSGADKDGEGLSTQCECNIKVKDVNDNFPMFRDSQYSARIEENILSSELLRFQVTDLDE  246
              +   D       +       + V+D+NDN P F    Y   + E++   +++    TD D+
```

```
Sbjct  1940  KILAIDDGIPPKTATATLTVIVQDINDNPPKFL-KDYRPVLPEHVPPRKVVEILATDDDD  1998

Query  247   EYTDNWLAVYFFTSGNEGNW----FEIQTDPRTNEG----ILKVVKALDYEQLQSVKLSI  298
                 N     F       +     F+++ D +   G    I+  +++ D EQ +   + I
Sbjct  1999  RSKSNGPPFQFRLDPGADDIIRASFKVEQDQKGANGDGMAIVSSLRSFDREQQKEYLIPI  2058

Query  299   AVKNKA  304
              +K+
Sbjct  2059  VIKDHG  2064

 Score = 51.6 bits (122),  Expect = 5e-05, Method: Compositional matrix adjust.
 Identities = 97/398 (24%), Positives = 163/398 (41%), Gaps = 54/398 (14%)

Query  106   PVFSQQIFMGEIEENSASNSLVMILNATDADEPNHLNSKIAFKIVSQ-EPAGTPMFLLSR  164
             P F    +  EI EN   +S ++ + A      +  + +I + + +Q + AGT  F +
Sbjct  655   PQFYMPSYEAEIPENQKKDSDIISVKAK-----SFADREIRYTLKAQGQGAGT--FNIGP  707

Query  165   NTGEVRTLTNSLDRE---QASSYRLVVSGADKDGEGLSTQCECNIKVKDVNDNFPMFRDS  221
              +G V+ L   LD E   Q   Y LVV+ A +D  G ST  E  I+V DVNDN P F
Sbjct  708   TSGIVK-LAKELDFEDLRQPHVYSLVVT-ATEDSGGFSTSVELTIRVTDVNDNAPKFELP  765

Query  222   QYSAR-IEENI-LSSELLRFQVTDLDEEYTDNWLAVYFFTSGNEGNWFEIQTDPRTNEGI  279
              Y A  ++E+I L + +L+ +  D D         + +  S +    F + ++     GI
Sbjct  766   DYQAHNVDEDIPLGTSILKVKAMDSDSGAN---AEIEYLVSDDH---FAVDSN-----GI  814

Query  280   LKVVKALDYEQLQS-VKLSIAVKNKAEFHQSVISRYRVQSTPVTIQVINVREGIAFRPAS  338
             +   K LD +   +  +  +  K+K E  ++  +  RV +           E   F
Sbjct  815   IVNNKQLDADNNNAYYEFVVTAKDKGEPAKTGTATVRVYTKNKN------DEEPKFSQQV  868

Query  339   KTFTVQKGISSKKLVDYILGTYQAIDEDTNKAASNVK--YVMGRNDGGYLMIDSKTAEIK  396
              T  V +      LV  ++         ++K   NV+  +V G    G  +I+  T  I+
Sbjct  869   YTPNVDENAGPNTLVTTVVA--------SDKDGDNVRFGFVGGGTSSGQFVIEEITGVIR  920

Query  397   FVK---NMNRDSTFI-VNKTITAEVLAIDEYTGKTSTGTVYVRVPDFNDN------CPTA  446
                   +++RD   + V            + T  TST  V V + D NDN      C T
Sbjct  921   LHSKAISLDRDKYELNVTAMDDGSCCVNGDATIHTSTAVVVVFITDVNDNKPVFKDCGTY  980

Query  447   VLE-KDAVCSSSPSVVVSARTLNNRYTGPYTFALEDQP  483
               + ++   + SP + V A   +    G   +++  QP
Sbjct  981   YPKVEEGAPNGSPVIKVQATDEDKGVNGQVKYSIVQQP  1018

 Score = 44.3 bits (103),  Expect = 0.010, Method: Compositional matrix adjust.
 Identities = 48/199 (24%), Positives = 73/199 (37%), Gaps = 19/199 (10%)

Query  27    DYQATQKITYRISGVGIDQPPFGIFVVDKNTGDINITAIVDREETPSFLITCRALNAQGL  86
             D     K++Y I      Q  F I       G + I   +DRE TP   +   A++
Sbjct  1894  DQGGKSKVSYAIDRSSDRQRQFSI----NQEGTVTIQRNLDREVTPRHQVKILAIDDGIP  1949

Query  87    DVEKPLILTVKILDINDNPPVFSQQIFMGEIEENSASNSLVMILNATDADEPNHLNSKIA  146
                    LTV + DINDNPP F +  +   + E+     +V IL   D D
Sbjct  1950  PKTATATLTVIVQDINDNPPKFLKD-YRPVLPEHVPPRKVVEILATDDDDRSKSNGPPFQ  2008

Query  147   FKIVSQEPAGTPMFLLSRNTGEVRTLTN-----------SLDREQASSYRLVVSGADKDG  195
             F++   +P    +   S    + +   N           S DREQ   Y + +   D
Sbjct  2009  FRL---DPGADDIIRASFKVEQDQKGANGDGMAIVSSLRSFDREQQKEYLIPIVIKDHGN  2065

Query  196   EGLSTQCECNIKVKDVNDN  214
               ++      + + DVNDN
Sbjct  2066  PAMTGTSTLTVVIGDVNDN  2084

 Score = 39.3 bits (90),  Expect = 0.33, Method: Compositional matrix adjust.
 Identities = 54/242 (22%), Positives = 102/242 (42%), Gaps = 22/242 (9%)

Query  4    KFAKPCREGE--DNSKRNP-IAKITSDYQATQKITYRISGVGIDQPPFGIFVVDKNTGDI  60
            +F  P  E E  +N K++  I  + +   A ++I Y +   G      G F +   +G +
Sbjct  656  QFYMPSYEAEIPENQKKDSDIISVKAKSFADREIRYTLKAQGQGA---GTFNIGPTSGIV  712

Query  61   NITAIVDREE--TPSF--LITCRALNAQGLDVEKPLILTVKILDINDNPPVFSQQIFMGE  116
```

```
               +   +D E+   P    L+     ++ G      + LT+++ D+NDN P F    +
Sbjct  713  KLAKELDFEDLRQPHVYSLVVTATEDSGGFSTS--VELTIRVTDVNDNAPKFELPDYQAH  770

Query  117  -IEENSASNSLVMILNATDADEPNHLNSKIAFKIVSQEPAGTPMFLLSRNTGEVRTLTNS  175
             ++E+    + ++ + A D+D  +  N++I + +VS +      F +  N   V
Sbjct  771  NVDEDIPLGTSILKVKAMDSD--SGANAEIEY-LVSDD-----HFAVDSNGIIVNNKQLD  822

Query  176  LDREQASSYRLVVSGADKDGEGLSTQCECNIKVKDVNDNFPMFRDSQYSARIEENILSSE  235
             D   A  Y  VV+  DK     +      +  K+ ND  P F    Y+  ++EN   +
Sbjct  823  ADNNNA-YYEFVVTAKDKGEPAKTGTATVRVYTKNKNDEEPKFSQQVYTPNVDENAGPNT  881

Query  236  LL  237
            L+
Sbjct  882  LV  883

 Score = 39.3 bits (90),  Expect = 0.35, Method: Compositional matrix adjust.
 Identities = 89/389 (23%), Positives = 149/389 (38%), Gaps = 62/389 (16%)

Query  75   LITCRALNAQGLDVEKPLILTVKILDINDNPPVFSQQIF--MGEIEENSASNSLVMILNA  132
            +IT +  NA GL       + + + D+ND PP F  +       ++ N+  N+ V  L A
Sbjct  517  IITNQGHNA-GLKYTDNQRVIIHVKDVNDEPPYFINRPLPMQAVVQLNAPPNTPVFTLQA  575

Query  133  TDADEPNHLNSKIAFKIVSQEPAGTPMFLLSRNTGEVRTLTNSLDREQASSYRLVVSGAD  192
             D D     +  I + IV     G   F +   +G VRT    L  +    Y L V   D
Sbjct  576  RDPD----TDHNIHYFIVRDRTGG--RFEVDERSGVVRTRGTDL-FQLDMEYVLYVKAED  628

Query  193  KDGE----GLSTQCECNIKVKDVNDNFPMFRDSQYSARIEEN-ILSSELLRFQVTDLDEE  247
            ++G+        +  E  + +        P F    Y A I EN    S+++  +     +
Sbjct  629  QNGKVDDRRFQSTPEERLSIVG-GKRAPQFYMPSYEAEIPENQKKDSDIISVKA----KS  683

Query  248  YTDNWLAVYFFTSGNEGNWFEIQTDPRTNEGILKVVKALDYEQLQSVKLS----IAVKNK  303
            + D  +       G     F I        GI+K+ K LD+E L+   +       A ++
Sbjct  684  FADREIRYTLKAQGQGAGTFNIGP----TSGIVKLAKELDFEDLRQPHVYSLVVTATEDS  739

Query  304  AEFHQSVISRYRVQSTPVTIQVINVREGIAFRPASKTFTVQKGISSKKLVDYILGT----  359
              F  SV           +TI+V +V +       +  F +    +     D  LGT
Sbjct  740  GGFSTSV---------ELTIRVTDVNDN------APKFELPDYQAHNVDEDIPLGTSILK  784

Query  360  YQAIDEDTNKAASNVKYVMGRN----DGGYLMIDSKTAEIKFVKNMNRDSTFIVNKTITA  415
             +A+D D+  A + ++Y++  +    D    +++++K  +     N N    F+V
Sbjct  785  VKAMDSDSG-ANAEIEYLVSDDHFAVDSNGIIVNNKQLD---ADNNNAYYEFVVTAKDKG  840

Query  416  EVLAIDEYTGKTSTGTVYVYRVPDFNDNCP  444
            E         KT T TV V   + ND  P
Sbjct  841  E-------PAKTGTATVRVYTKNKNDEEP  862

>ACON2_034794.P62457 | transcript=ACON2_034794.R62457 | gene=ACON2_034794
| organism=Anopheles_coluzzii_AcolN3 | gene_product=neural-cadherin,
transcript variant X15, neural-cadherin,
transcript variant X13, neural-cadherin, transcript variant
X8, neural-cadherin, transcript variant X1, neural-cadherin,
transcript variant X3, neural-cadherin, transcript variant
X10, neural-cadherin, transcript variant X16, neural-cadherin,
transcript variant X2, neural-cadherin, transcript variant
X12, neural-cadherin, transcript variant X4, neural-cadherin,
transcript variant X7, neural-cadherin, transcript variant
X9, neural-cadherin, transcript variant X5, neural-cadherin,
transcript variant X17, neural-cadherin, transcript variant
X14, neural-cadherin, transcript variant X11, neural-cadherin,
transcript variant X6 | transcript_product=neural-cadherin,
transcript variant X13 | location=AcolN3_3RL:40912217-41119468(+)
| protein_length=3099 | sequence_SO=chromosome
| SO=protein_coding_gene | is_pseudo=false
Length=3099

 Score = 146 bits (369),  Expect = 4e-35, Method: Compositional matrix adjust.
 Identities = 128/446 (29%), Positives = 217/446 (49%), Gaps = 36/446 (8%)

Query  20    PIAKITS---DYQATQKITYRISGVGID--QPPFGIFVVDKNTGDINITAIVDREET---  71
```

```
             PI ++T+   D    Q I Y ++G GID   P    F +++ TG+I +   +DR++
Sbjct  1539  PIMQVTATDGDKDRPQNIVYFLTGQGIDPDNPANSKFDINRTTGEIFVLKPLDRDQPNGR  1598

Query  72    PSFLITCRALNAQGLDVEKPLILTVKILDINDNPPVFSQQIFMGEIEENSASNSLVMILN  131
             P +  T  A +  G  +     + V + DINDN P+F Q ++ G + EN  +  +VM +
Sbjct  1599  PQWRFTVFAQDEGGEGLVGYADVQVNLKDINDNAPIFPQGVYFGNVTENGTAGMVVMTMT  1658

Query  132   ATDADEPNH-LNSKIAFKI---VSQEPAGTPMFLLSRNTGEVRTLTNSLDREQASSYRLV  187
             A D D+PN   N+K+ + I   V +E  G+P+F +  +TG ++T    LDRE+   Y +
Sbjct  1659  AVDYDDPNEGTNAKLIYSIEKNVIEEETGSPIFEIEADTGVIKTAVCCLDRERTPDYSIQ  1718

Query  188   VSGADKDGEGLSTQCECNIKVKDVNDNFPMFRDSQYSARIEENILSS--ELLRFQVTDLD  245
             V     DG GL       +I+VKD+ND  P F   ++   ++E   ++  E+    VT  D
Sbjct  1719  VVAM--DGGGLKGTGTASIRVKDINDMPPQFTKDEWFTEVDETDGTNLPEMPILTVTVHD  1776

Query  246   EEYTDNWLAVYFFTSGNEGNWFEIQTDPRTNEGI--LKVVKALDYE---QLQSVKLSIAV  300
             E+ T+ +        SG    + F +    R N+G   LK+V+ LDYE   Q    +  I V
Sbjct  1777  EDETNKFQYKVIDNSGYGADKFTMV---RNNDGTGSLKIVQPLDYEDPLQSNGFRFRIQV  1833

Query  301   KNKAEFHQSVISRYRVQSTPVTIQVINVREGIAFRPASKTFTVQKGISSKKLVDYILGTY  360
              +K E + +   +Y V  + V +++ ++ +    +P  +   ++  +     V   L T+
Sbjct  1834  NDKGEDNDN--DKYHVAYSWVVVKLRDINDN---KPQFERPNIEVSVYENADVGKTLETF  1888

Query  361   QAIDEDTNKAASNVKYVMGR-NDGGYLMIDSKTAEIKFVKNMNRDSTFIVNKTITAEVLA  419
             +A D D     S V Y + R +D        ++   +   +N++R+    V       ++LA
Sbjct  1889  KATDPDQG-GKSKVSYAIDRSSDRQRQFSINQEGTVTIQRNLDRE----VTPRHQVKILA  1943

Query  420   IDE-YTGKTSTGTVYVRVPDFNDNCP  444
             ID+    KT+T T+ V V D NDN P
Sbjct  1944  IDDGIPPKTATATLTVIVQDINDNPP  1969

 Score = 75.9 bits (185),  Expect = 2e-12, Method: Compositional matrix adjust.
 Identities = 89/361 (25%), Positives = 167/361 (46%), Gaps = 36/361 (10%)

Query  51    FVVDKNTGDINITAIVDRE-ETPSFL-ITCRALNAQGLDVEKPLILTVKILDINDNPPVF  108
             F VD+ TG+++   + DRE +   F+ +T +A +    +E      TV+I D+NDNPP+F
Sbjct  1025  FTVDEETGEVSTNKVFDREGDDGKFVSVTVKATDQGEPSLEGVCSFTVEITDVNDNPPLF  1084

Query  109   SQQIFMGEIEENSASNSLVMILNATDADEPNHLNSKIAFKIVS-QEPAGTPMFLLSRNTG  167
              +Q ++  ++++++  + ++ ++A+D D  N  N  I + + +   P     F +   +G
Sbjct  1085  DRQKYVENVKQDASIGTNILRVSASDEDADN--NGAIVYTLSAPHSPNDLEYFEIQPESG  1142

Query  168   EVRTLTNSLDREQASSYRLVVSGADKDGEGLSTQCECNIKVKDVNDNFPMFRDSQYSA-R  226
              +  L   LDRE   +Y+L      DK    LS   E  I V D  +N P++ ++ Y
Sbjct  1143  WI-VLKKPLDRE---TYKLEAMAQDKGYPPLSRTVEVQIDVVDRANNPPVWDNTVYGPIY  1198

Query  227   IEENI-LSSELLRFQVTDLDEEYTDNWLAVYFFTSG-----NEGNWFEIQTDPRTNE--G  278
             ++EN+ +  +++  + +   E    N    Y    G     N+ + F +Q  P   +
Sbjct  1199  VKENMPVGGKVVSIKASSGIE---GNPTVFYRLMPGSTAQTNKLHTFYLQQRPDNGDTWA  1255

Query  279   ILKVVKALDYEQLQSVKLSIAVKNKAEFHQSVISRYRVQSTPVTIQVINVREGIAFRPAS  338
              +KV   LD+E ++   L++ V+N          ++        V IQ+ +V + I
Sbjct  1256  DIKVNHPLDFETIKEYNLTVRVENNG-------AQQLASEATVYIQLEDVNDEIPLFTER  1308

Query  339   KTFTVQKGISSKKLVDYILGTYQAIDEDTNKAASNVKYVM---GRNDG-GYLMIDSKTAE  394
             +  TV +G      V  +     AID+D     + V Y +    RN+G  +  I+ ++ E
Sbjct  1309  EQETVLEGEPIGTKVTQV----NAIDKDGTFPNNQVYYYIVDSPRNEGKDFFEINLQSGE  1364

Query  395   I  395
             I
Sbjct  1365  I  1365

 Score = 75.5 bits (184),  Expect = 2e-12, Method: Compositional matrix adjust.
 Identities = 103/418 (25%), Positives = 174/418 (42%), Gaps = 50/418 (12%)

Query  50    IFVVDKNTGDINITAIVDREETPSFLITCRAL----------NAQGLDVEKPLILTVKIL  99
              F ++  +G+I    + DRE+  ++ +   A            N Q   V K   + + I
Sbjct  1355  FFEINLQSGEIFTKVVFDREKQGAYALEVEARDGAPSARPNSNQQPNSVTK--FIRIGIA  1412
```

```
Query  100   DINDNPPVFSQQIFMGEIEENSASNSLVMILNATDADEPNHLNSKIAFKIVSQEPAGTPM  159
             D NDNPP F + ++  E++EN      V+ + A D DE    +S+I ++I S    G
Sbjct  1413  DKNDNPPYFDKALYEAEVDENEDIQHTVLTVTAKDHDE----SSRIRYEITSGNIGGA--  1466

Query  160   FLLSRNTGEVRTLTNSLDREQASSYRLVVSGADKDGEGLSTQCECNIKVKDVNDNFPMFR  219
             F +   TG +  +  +LD E    Y L +  +D   E  +      I VKDVND  P F
Sbjct  1467  FAVKNMTGAI-YVAGALDYETRKRYYLTLVASDSLNENETIIV---INVKDVNDMPPSFP  1522

Query  220   DSQYSARIEENI-LSSELLRFQVTDLDEEYTDNWLAVYFFTSG--NEGNWFEIQTDPRTN  276
                Y   ++E + +   +++   TD D++   N   VYF T    +  N    + D
Sbjct  1523  QQVYERTMDEELAVPFPIMQVTATDGDKDRPQN--IVYFLTGQGIDPDNPANSKFDINRT  1580

Query  277   EGILKVVKALDYEQLQS-VKLSIAVKNKAEFHQSVISRYRVQSTPVTIQVINVREGIAFR  335
              G + V+K LD +Q     +    V  + E  + ++    VQ     + + ++ +
Sbjct  1581  TGEIFVLKPLDRDQPNGRPQWRFTVFAQDEGGEGLVGYADVQ-----VNLKDINDNAPIF  1635

Query  336   PASKTFTVQKGISSKKLVDYILGTYQAIDEDTNKAASNVK--YVMGRN------DGGYLM  387
             P    F     ++       ++ T  A+D D     +N K  Y + +N
Sbjct  1636  PQGVYF---GNVTENGTAGMVVMTMTAVDYDDPNEGTNAKLIYSIEKNVIEEETGSPIFE  1692

Query  388   IDSKTAEIK-FVKNMNRDSTFIVNKTITAEVLAIDEYTGKTSTGTVYVRVPDFNDNCP  444
             I++ T  IK  V  ++R+ T       + +V+A+D   G   TGT  +RV D ND  P
Sbjct  1693  IEADTGVIKTAVCCLDRERT----PDYSIQVVAMDG-GGLKGTGTASIRVKDINDMPP  1745

 Score = 55.5 bits (132),  Expect = 3e-06, Method: Compositional matrix adjust.
 Identities = 49/158 (31%), Positives = 78/158 (49%), Gaps = 24/158 (15%)

Query  102   NDNPPVFSQQIFMGEIEENSASNSLVMILNATDADEPNHLNSKIAFKIVSQEPAGTP--M  159
             ND  P FSQQ++   ++EN+  N+LV  + A+D D  N     + F  V     GT
Sbjct  858   NDEEPKFSQQVYTPNVDENAGPNTLVTTVVASDKDGDN-----VRFGFVG---GGTSSGQ  909

Query  160   FLLSRNTGEVRTLTN--SLDREQASSYRLVVSGADK-------DGEGLSTQCECNIKVKD  210
             F++   TG +R  +   SLDR++   Y L V+  D        D    ++     + + D
Sbjct  910   FVIEEITGVIRLHSKAISLDRDK---YELNVTAMDDGSCCVNGDATIHTSTAVVVVFITD  966

Query  211   VNDNFPMFRD-SQYSARIEENILS-SELLRFQVTDLDE  246
             VNDN P+F+D   Y  ++EE   + S +++ Q TD D+
Sbjct  967   VNDNKPVFKDCGTYYPKVEEGAPNGSPVIKVQATDEDK  1004

 Score = 53.5 bits (127),  Expect = 1e-05, Method: Compositional matrix adjust.
 Identities = 70/306 (23%), Positives = 126/306 (41%), Gaps = 30/306 (10%)

Query  20    PIAKIT-SDYQATQKITYRI---SGVGIDQPPFGIFVVDKNTGDINITAIVDRE---ETP  72
             PI  +T  D   T K  Y++   SG G D+  F +   +  TG + I   +D E   ++
Sbjct  1768  PILTVTVHDEDETNKFQYKVIDNSGYGADK--FTMVRNNDGTGSLKIVQPLDYEDPLQSN  1825

Query  73    SFLITCRALNAQGLDVEKPLI------LTVKILDINDNPPVFSQQIFMGEIEENSASNSL  126
              F    + +N +G D +          + VK+ DINDN P F +      + EN+
Sbjct  1826  GFRFRIQ-VNDKGEDNDNDKYHVAYSWVVVKLRDINDNKPQFERPNIEVSVYENADVGKT  1884

Query  127   VMILNATDADEPNHLNSKIAFKIVSQEPAGTPMFLLSRNTGEVRTLTNSLDREQASSYRL  186
             +    ATD D+     SK+++ I   + +       S N     T+  +LDRE    +++
Sbjct  1885  LETFKATDPDQGG--KSKVSYAI---DRSSDRQRQFSINQEGTVTIQRNLDREVTPRHQV  1939

Query  187   VVSGADKDGEGLSTQCECNIKVKDVNDNFPMFRDSQYSARIEENILSSELLRFQVTDLDE  246
              +   D      +      + V+D+NDN P F    Y   + E++   +++    TD D+
Sbjct  1940  KILAIDDGIPPKTATATLTVIVQDINDNPPKFL-KDYRPVLPEHVPPRKVVEILATDDDD  1998

Query  247   EYTDNWLAVYFFTSGNEGNW----FEIQTDPRTNEG----ILKVVKALDYEQLQSVKLSI  298
                 N     F       +     F+++ D +   G    I+  +++ D EQ +   + I
Sbjct  1999  RSKSNGPPFQFRLDPGADDIIRASFKVEQDQKGANGDGMAIVSSLRSFDREQQKEYLIPI  2058

Query  299   AVKNKA  304
              +K+
Sbjct  2059  VIKDHG  2064

 Score = 51.6 bits (122),  Expect = 5e-05, Method: Compositional matrix adjust.
 Identities = 97/398 (24%), Positives = 163/398 (41%), Gaps = 54/398 (14%)
```

```
Query  106   PVFSQQIFMGEIEENSASNSLVMILNATDADEPNHLNSKIAFKIVSQ-EPAGTPMFLLSR  164
             P F    +  EI EN   +S ++ + A      +  + +I + + +Q + AGT  F +
Sbjct  655   PQFYMPSYEAEIPENQKKDSDIISVKAK-----SFADREIRYTLKAQGQGAGT--FNIGP  707

Query  165   NTGEVRTLTNSLDRE---QASSYRLVVSGADKDGEGLSTQCECNIKVKDVNDNFPMFRDS  221
              +G V+ L   LD E   Q   Y LVV+ A +D  G ST  E  I+V DVNDN P F
Sbjct  708   TSGIVK-LAKELDFEDLRQPHVYSLVVT-ATEDSGGFSTSVELTIRVTDVNDNAPKFELP  765

Query  222   QYSAR-IEENI-LSSELLRFQVTDLDEEYTDNWLAVYFFTSGNEGNWFEIQTDPRTNEGI  279
              Y A  ++E+I L + +L+ +  D D         + +  S +    F + ++     GI
Sbjct  766   DYQAHNVDEDIPLGTSILKVKAMDSDSGAN---AEIEYLVSDDH---FAVDSN-----GI  814

Query  280   LKVVKALDYEQLQS-VKLSIAVKNKAEFHQSVISRYRVQSTPVTIQVINVREGIAFRPAS  338
             +   K LD +   +  +  +  K+K E  ++  +  RV +           E   F
Sbjct  815   IVNNKQLDADNNNAYYEFVVTAKDKGEPAKTGTATVRVYTKNKN------DEEPKFSQQV  868

Query  339   KTFTVQKGISSKKLVDYILGTYQAIDEDTNKAASNVK--YVMGRNDGGYLMIDSKTAEIK  396
              T  V +      LV  ++         ++K   NV+  +V G    G  +I+  T  I+
Sbjct  869   YTPNVDENAGPNTLVTTVVA--------SDKDGDNVRFGFVGGGTSSGQFVIEEITGVIR  920

Query  397   FVK---NMNRDSTFI-VNKTITAEVLAIDEYTGKTSTGTVYVRVPDFNDN------CPTA  446
                   +++RD   + V            + T  TST  V V + D NDN      C T
Sbjct  921   LHSKAISLDRDKYELNVTAMDDGSCCVNGDATIHTSTAVVVVFITDVNDNKPVFKDCGTY  980

Query  447   VLE-KDAVCSSSPSVVVSARTLNNRYTGPYTFALEDQP  483
               + ++   + SP + V A    +   G   +++  QP
Sbjct  981   YPKVEEGAPNGSPVIKVQATDEDKGVNGQVKYSIVQQP  1018

 Score = 44.3 bits (103),  Expect = 0.010, Method: Compositional matrix adjust.
 Identities = 48/199 (24%), Positives = 73/199 (37%), Gaps = 19/199 (10%)

Query  27    DYQATQKITYRISGVGIDQPPFGIFVVDKNTGDINITAIVDREETPSFLITCRALNAQGL  86
             D     K++Y I      Q  F I       G + I   +DRE TP   +   A++
Sbjct  1894  DQGGKSKVSYAIDRSSDRQRQFSI----NQEGTVTIQRNLDREVTPRHQVKILAIDDGIP  1949

Query  87    DVEKPLILTVKILDINDNPPVFSQQIFMGEIEENSASNSLVMILNATDADEPNHLNSKIA  146
                    LTV + DINDNPP F +  +   + E+     +V IL   D D
Sbjct  1950  PKTATATLTVIVQDINDNPPKFLKD-YRPVLPEHVPPRKVVEILATDDDDRSKSNGPPFQ  2008

Query  147   FKIVSQEPAGTPMFLLSRNTGEVRTLTN-----------SLDREQASSYRLVVSGADKDG  195
             F++   +P    +   S    + +   N           S DREQ   Y + +   D
Sbjct  2009  FRL---DPGADDIIRASFKVEQDQKGANGDGMAIVSSLRSFDREQQKEYLIPIVIKDHGN  2065

Query  196   EGLSTQCECNIKVKDVNDN  214
               ++      + + DVNDN
Sbjct  2066  PAMTGTSTLTVVIGDVNDN  2084

 Score = 39.3 bits (90),  Expect = 0.33, Method: Compositional matrix adjust.
 Identities = 54/242 (22%), Positives = 102/242 (42%), Gaps = 22/242 (9%)

Query  4    KFAKPCREGE--DNSKRNP-IAKITSDYQATQKITYRISGVGIDQPPFGIFVVDKNTGDI  60
            +F  P  E E  +N K++  I  + +   A ++I Y +   G      G F +   +G +
Sbjct  656  QFYMPSYEAEIPENQKKDSDIISVKAKSFADREIRYTLKAQGQGA---GTFNIGPTSGIV  712

Query  61   NITAIVDREE--TPSF--LITCRALNAQGLDVEKPLILTVKILDINDNPPVFSQQIFMGE  116
             +   +D E+   P    L+     ++ G      + LT+++ D+NDN P F    +
Sbjct  713  KLAKELDFEDLRQPHVYSLVVTATEDSGGFSTS--VELTIRVTDVNDNAPKFELPDYQAH  770

Query  117  -IEENSASNSLVMILNATDADEPNHLNSKIAFKIVSQEPAGTPMFLLSRNTGEVRTLTNS  175
             ++E+    + ++ + A D+D  +  N++I + +VS +      F +  N   V
Sbjct  771  NVDEDIPLGTSILKVKAMDSD--SGANAEIEY-LVSDD-----HFAVDSNGIIVNNKQLD  822

Query  176  LDREQASSYRLVVSGADKDGEGLSTQCECNIKVKDVNDNFPMFRDSQYSARIEENILSSE  235
             D   A  Y  VV+  DK     +      +  K+ ND  P F    Y+  ++EN   +
Sbjct  823  ADNNNA-YYEFVVTAKDKGEPAKTGTATVRVYTKNKNDEEPKFSQQVYTPNVDENAGPNT  881

Query  236  LL  237
            L+
```

```
Sbjct  882  LV  883

 Score = 39.3 bits (90),  Expect = 0.34, Method: Compositional matrix adjust.
 Identities = 89/389 (23%), Positives = 149/389 (38%), Gaps = 62/389 (16%)

Query  75   LITCRALNAQGLDVEKPLILTVKILDINDNPPVFSQQIF--MGEIEENSASNSLVMILNA  132
            +IT +  NA GL       + + + D+ND PP F  +       ++ N+  N+ V  L A
Sbjct  517  IITNQGHNA-GLKYTDNQRVIIHVKDVNDEPPYFINRPLPMQAVVQLNAPPNTPVFTLQA  575

Query  133  TDADEPNHLNSKIAFKIVSQEPAGTPMFLLSRNTGEVRTLTNSLDREQASSYRLVVSGAD  192
             D D     +  I + IV     G   F +   +G VRT    L  +    Y L V   D
Sbjct  576  RDPD----TDHNIHYFIVRDRTGG--RFEVDERSGVVRTRGTDL-FQLDMEYVLYVKAED  628

Query  193  KDGE----GLSTQCECNIKVKDVNDNFPMFRDSQYSARIEEN-ILSSELLRFQVTDLDEE  247
            ++G+        +  E  + +        P F    Y A I EN    S+++  +     +
Sbjct  629  QNGKVDDRRFQSTPEERLSIVG-GKRAPQFYMPSYEAEIPENQKKDSDIISVKA----KS  683

Query  248  YTDNWLAVYFFTSGNEGNWFEIQTDPRTNEGILKVVKALDYEQLQSVKLS----IAVKNK  303
            + D  +        G     F I         GI+K+ K LD+E L+   +       A ++
Sbjct  684  FADREIRYTLKAQGQGAGTFNIGP----TSGIVKLAKELDFEDLRQPHVYSLVVTATEDS  739

Query  304  AEFHQSVISRYRVQSTPVTIQVINVREGIAFRPASKTFTVQKGISSKKLVDYILGT----  359
              F  SV           +TI+V +V +        +  F +    +     D  LGT
Sbjct  740  GGFSTSV---------ELTIRVTDVNDN------APKFELPDYQAHNVDEDIPLGTSILK  784

Query  360  YQAIDEDTNKAASNVKYVMGRN----DGGYLMIDSKTAEIKFVKNMNRDSTFIVNKTITA  415
             +A+D D+  A + ++Y++  +    D   +++++K  +     N N    F+V
Sbjct  785  VKAMDSDSG-ANAEIEYLVSDDHFAVDSNGIIVNNKQLD---ADNNNAYYEFVVTAKDKG  840

Query  416  EVLAIDEYTGKTSTGTVYVRVPDFNDNCP  444
            E         KT T TV V   + ND  P
Sbjct  841  E-------PAKTGTATVRVYTKNKNDEEP  862

>AAEL000597-PB | transcript=AAEL000597-RB | gene=AAEL000597 |
organism=Aedes_aegypti_LVP_AGWG | gene_product=cadherin | transcript_product=cadherin
| location=AaegL5_2:408019463-408166166(-)
| protein_length=1717 | sequence_SO=chromosome | SO=protein_coding_gene
| is_pseudo=false
Length=1717

 Score = 146 bits (368),  Expect = 4e-35, Method: Compositional matrix adjust.
 Identities = 130/449 (29%), Positives = 216/449 (48%), Gaps = 47/449 (10%)

Query  21   IAKITSDYQATQKITYRISGVGID--QPPFGIFVVDKNTGDINITAIVDREET---PSFL  75
            +     D    Q I Y ++G GID   P    F +++ +GDI +   +DR++    P +
Sbjct  160  VTATDGDKDRPQNIVYFLTGQGIDPDNPANSKFDINRTSGDIFVLKPLDRDQPNGRPQWR  219

Query  76   ITCRALNAQGLDVEKPLILTVKILDINDNPPVFSQQIFMGEIEENSASNSLVMILNATDA  135
             T  A +  G  +     + V + DINDN P+F Q ++ G + EN  +  +VM + A D
Sbjct  220  FTVFAQDEGGEGLVGYADVQVNLKDINDNAPIFPQGVYFGNVTENGTAGMVVMTMTAVDY  279

Query  136  DEPNH-LNSKIAFKI---VSQEPAGTPMFLLSRNTGEVRTLTNSLDREQASSYRLVVSGA  191
            D+PN   N+K+ + I   V +E  G+P+F +  +TG ++T    LDRE+   Y + V
Sbjct  280  DDPNEGTNAKLIYSIEKNVIEEETGSPIFEIEADTGVIKTAVCCLDRERTPDYSIQVVAM  339

Query  192  DKDGEGLSTQCECNIKVKDVNDNFPMFRDSQYSARIEENILSS--ELLRFQVTDLDEEYT  249
              DG GL       +I+VKD+ND  P F    ++   ++E   ++  E+    VT  DE+ T
Sbjct  340  --DGGGLKGTGTASIRVKDINDMPPQFTKDEWFTEVDETDGTNLPEMPILTVTVHDEDET  397

Query  250  DNWLAVYFFTSGNEGNWFEIQTDPRTNEGI--LKVVKALDYE-QLQS--VKLSIAVKNKA  304
            + +        SG    + F +     R N+G   LK+V+ LDYE QLQS    +  I V +K
Sbjct  398  NKFQYKVIDNSGYGADKFTMV---RNNDGTGSLKIVQPLDYEDQLQSNGFRFRIQVNDKG  454

Query  305  EFHQSVISRYRVQSTPVTIQVINVREGIAFRPASKTFTVQKGISSKKLVDYILGTYQAID  364
            E + +    +Y V  + V +++ ++ ++ +     +P  +    ++  +       V   L T++A D
Sbjct  455  EDNDN--DKYHVAYSWVVVKLRDINDN---KPQFERPNIEVSVYENADVGKTLETFKATD  509

Query  365  EDTNKAASNVKYVMGRNDGGYLMIDSKTAEIKFVKNMNRDSTFIVNKTITAEV-------  417
             D     S V Y + R          S   + +F  ++N++ T  + + +  EV
```

```
Sbjct  510  PDQG-GRSKVSYAIDR---------SSDRQRQF--SINQEGTVTIQRQLDREVTPRHQVK  557

Query  418  -LAIDE-YTGKTSTGTVYVRVPDFNDNCP  444
             LAID+    KT+T T+ V V D NDN P
Sbjct  558  ILAIDDGIPPKTATATLTVIVQDINDNPP  586

 Score = 84.0 bits (206),  Expect = 5e-15, Method: Compositional matrix adjust.
 Identities = 98/367 (27%), Positives = 158/367 (43%), Gaps = 40/367 (11%)

Query  93   ILTVKILDINDNPPVFSQQIFMGEIEENSASNSLVMILNATDADEPNHLNSKIAFKIVSQ  152
             + + I D NDNPP F + ++  E++EN      V+ + A D DE    +S+I ++I S
Sbjct  21   FIRIGIADKNDNPPYFDKALYEAEVDENEDIQHTVLTVTAKDHDE----SSRIRYEITSG  76

Query  153  EPAGTPMFLLSRNTGEVRTLTNSLDREQASSYRLVVSGADKDGEGLSTQCECNIKVKDVN  212
               G   F +   TG +  +  +LD E    Y L ++ +D   E  +T      I VKDVN
Sbjct  77   NIGGA--FAVKNMTGAI-YVAGALDYETRKRYELRLAASDNLKENYTTVV---IHVKDVN  130

Query  213  DNFPMFRDSQYSARIEENI---LSSELLRFQVTDLDEEYTDNWLAVYFFTSG--NEGNWF  267
            DN P+F    Y A+I E      L   +L+   TD D++   N   VYF T    +  N
Sbjct  131  DNPPVFGRPTYQAQITEEDDRNLPKRILQVTATDGDKDRPQN--IVYFLTGQGIDPDNPA  188

Query  268  EIQTDPRTNEGILKVVKALDYEQLQS-VKLSIAVKNKAEFHQSVISRYRVQSTPVTIQVI  326
              + D     G + V+K LD +Q      +    V  + E  + ++    VQ     + +
Sbjct  189  NSKFDINRTSGDIFVLKPLDRDQPNGRPQWRFTVFAQDEGGEGLVGYADVQ-----VNLK  243

Query  327  NVREGIAFRPASKTFTVQKGISSKKLVDYILGTYQAIDEDTNKAASNVK--YVMGRN---  381
            ++ +      P    F      ++         ++ T  A+D D      +N K  Y + +N
Sbjct  244  DINDNAPIFPQGVYF---GNVTENGTAGMVVMTMTAVDYDDPNEGTNAKLIYSIEKNVIE  300

Query  382  ---DGGYLMIDSKTAEIK-FVKNMNRDSTFIVNKTITAEVLAIDEYTGKTSTGTVYVRVP  437
                         I++ T  IK  V  ++R+ T         + +V+A+D   G   TGT  +RV
Sbjct  301  EETGSPIFEIEADTGVIKTAVCCLDRERT----PDYSIQVVAMDG-GGLKGTGTASIRVK  355

Query  438  DFNDNCP  444
            D ND  P
Sbjct  356  DINDMPP  362

 Score = 53.5 bits (127),  Expect = 1e-05, Method: Compositional matrix adjust.
 Identities = 70/306 (23%), Positives = 125/306 (41%), Gaps = 30/306 (10%)

Query  20   PIAKIT-SDYQATQKITYRI---SGVGIDQPPFGIFVVDKNTGDINITAIVDRE---ETP  72
            PI  +T  D   T K  Y++   SG G D+  F +   +  TG + I   +D E   ++
Sbjct  385  PILTVTVHDEDETNKFQYKVIDNSGYGADK--FTMVRNNDGTGSLKIVQPLDYEDQLQSN  442

Query  73   SFLITCRALNAQGLDVEKPLI------LTVKILDINDNPPVFSQQIFMGEIEENSASNSL  126
             F    + +N +G D +           + VK+ DINDN P F +       + EN+
Sbjct  443  GFRFRIQ-VNDKGEDNDNDKYHVAYSWVVVKLRDINDNKPQFERPNIEVSVYENADVGKT  501

Query  127  VMILNATDADEPNHLNSKIAFKIVSQEPAGTPMFLLSRNTGEVRTLTNSLDREQASSYRL  186
            +    ATD D+      SK+++ I   + +        S N      T+   LDRE    +++
Sbjct  502  LETFKATDPDQGGR--SKVSYAI---DRSSDRQRQFSINQEGTVTIQRQLDREVTPRHQV  556

Query  187  VVSGADKDGEGLSTQCECNIKVKDVNDNFPMFRDSQYSARIEENILSSELLRFQVTDLDE  246
             +    D       +       + V+D+NDN P F     Y    + E++   +++     TD D+
Sbjct  557  KILAIDDGIPPKTATATLTVIVQDINDNPPKFL-KDYRPVLPEHVPPRKVVEILATDDDD  615

Query  247  EYTDNWLAVYFFTSGNEGNW----FEIQTDPRTNEG----ILKVVKALDYEQLQSVKLSI  298
                N     F        +     F+++ D +   G    I+  +++ D EQ +   + I
Sbjct  616  RSKSNGPPFQFRLDPGADDIIRASFKVEQDQKGANGDGMAIVSSLRSFDREQQKEYLIPI  675

Query  299  AVKNKA  304
             +K+
Sbjct  676  VIKDHG  681

 Score = 44.3 bits (103),  Expect = 0.009, Method: Compositional matrix adjust.
 Identities = 48/199 (24%), Positives = 73/199 (37%), Gaps = 19/199 (10%)

Query  27   DYQATQKITYRISGVGIDQPPFGIFVVDKNTGDINITAIVDREETPSFLITCRALNAQGL  86
```

```
               D     K++Y I      Q  F I       G + I   +DRE TP   +   A++
Sbjct  511  DQGGRSKVSYAIDRSSDRQRQFSI----NQEGTVTIQRQLDREVTPRHQVKILAIDDGIP  566

Query  87   DVEKPLILTVKILDINDNPPVFSQQIFMGEIEENSASNSLVMILNATDADEPNHLNSKIA  146
                    LTV + DINDNPP F +  +   + E+     +V IL   D D
Sbjct  567  PKTATATLTVIVQDINDNPPKFLKD-YRPVLPEHVPPRKVVEILATDDDDRSKSNGPPFQ  625

Query  147  FKIVSQEPAGTPMFLLSRNTGEVRTLTN-----------SLDREQASSYRLVVSGADKDG  195
            F++   +P    +   S    + +   N           S DREQ   Y + +   D
Sbjct  626  FRL---DPGADDIIRASFKVEQDQKGANGDGMAIVSSLRSFDREQQKEYLIPIVIKDHGN  682

Query  196  EGLSTQCECNIKVKDVNDN  214
              ++      + + DVNDN
Sbjct  683  PAMTGTSTLTVVIGDVNDN  701

>AALFPA_070705.P41263 | transcript=AALFPA_070705.R41263 | gene=AALFPA_070705
| organism=Aedes_albopictus_Foshan_FPA | gene_product=neural-cadherin
| transcript_product=neural-cadherin
| location=SWKY01000002:142685955-142823546(-) | protein_length=1717
| sequence_SO=supercontig | SO=protein_coding_gene
| is_pseudo=false
Length=1717

 Score = 146 bits (368),  Expect = 5e-35, Method: Compositional matrix adjust.
 Identities = 130/449 (29%), Positives = 216/449 (48%), Gaps = 47/449 (10%)

Query  21   IAKITSDYQATQKITYRISGVGID--QPPFGIFVVDKNTGDINITAIVDREET---PSFL  75
            +     D    Q I Y ++G GID   P    F +++ +GDI +   +DR++    P +
Sbjct  160  VTATDGDKDRPQNIVYFLTGQGIDPDNPANSKFDINRTSGDIFVLKPLDRDQPNGRPQWR  219

Query  76   ITCRALNAQGLDVEKPLILTVKILDINDNPPVFSQQIFMGEIEENSASNSLVMILNATDA  135
             T  A +  G  +     + V + DINDN P+F Q ++ G + EN  +  +VM + A D
Sbjct  220  FTVFAQDEGGEGLVGYADVQVNLKDINDNAPIFPQGVYFGNVTENGTAGMVVMTMTAVDY  279

Query  136  DEPNH-LNSKIAFKI---VSQEPAGTPMFLLSRNTGEVRTLTNSLDREQASSYRLVVSGA  191
            D+PN   N+K+ + I   V +E  G+P+F +  +TG ++T    LDRE+   Y + V
Sbjct  280  DDPNEGTNAKLIYSIEKNVIEEETGSPIFEIEADTGVIKTAVCCLDRERTPDYSIQVVAM  339

Query  192  DKDGEGLSTQCECNIKVKDVNDNFPMFRDSQYSARIEENILSS--ELLRFQVTDLDEEYT  249
              DG GL       +I+VKD+ND  P F   ++   ++E   ++  E+    VT  DE+ T
Sbjct  340  --DGGGLKGTGTASIRVKDINDMPPQFTKDEWFTEVDETDGTNLPEMPILTVTVHDEDET  397

Query  250  DNWLAVYFFTSGNEGNWFEIQTDPRTNEGI--LKVVKALDYE-QLQS--VKLSIAVKNKA  304
            + +         SG    + F +     R N+G   LK+V+ LDYE QLQS   +  I V +K
Sbjct  398  NKFQYKVIDNSGYGADKFTMV---RNNDGTGSLKIVQPLDYEDQLQSNGFRFRIQVNDKG  454

Query  305  EFHQSVISRYRVQSTPVTIQVINVREGIAFRPASKTFTVQKGISSKKLVDYILGTYQAID  364
            E + +   +Y V  + V +++ ++ +    +P  +   ++  +      V   L T++A D
Sbjct  455  EDNDN--DKYHVAYSWVVVKLRDINDN---KPQFERPNIEVSVYENADVGKTLETFKATD  509

Query  365  EDTNKAASNVKYVMGRNDGGYLMIDSKTAEIKFVKNMNRDSTFIVNKTITAEV-------  417
             D     S V Y + R          S   + +F  ++N++ T  + + +  EV
Sbjct  510  PDQG-GRSKVSYAIDR---------SSDRQRQF--SINQEGTVTIQRQLDREVTPRHQVK  557

Query  418  -LAIDE-YTGKTSTGTVYVRVPDFNDNCP  444
             LAID+    KT+T T+ V V D NDN P
Sbjct  558  ILAIDDGIPPKTATATLTVIVQDINDNPP  586

 Score = 83.6 bits (205),  Expect = 6e-15, Method: Compositional matrix adjust.
 Identities = 98/367 (27%), Positives = 158/367 (43%), Gaps = 40/367 (11%)

Query  93   ILTVKILDINDNPPVFSQQIFMGEIEENSASNSLVMILNATDADEPNHLNSKIAFKIVSQ  152
             + + I D NDNPP F + ++  E++EN      V+ + A D DE    +S+I ++I S
Sbjct  21   FIRIGIADKNDNPPYFDKALYEAEVDENEDIQHTVLTVTAKDHDE----SSRIRYEITSG  76

Query  153  EPAGTPMFLLSRNTGEVRTLTNSLDREQASSYRLVVSGADKDGEGLSTQCECNIKVKDVN  212
              G   F +   TG +  +  +LD E    Y L ++ +D   E  +T     I VKDVN
Sbjct  77   NIGGA--FAVKNMTGAI-YVAGALDYETRKRYELRLAASDNLKENYTTVV---IHVKDVN  130
```

```
Query  213  DNFPMFRDSQYSARIEENI---LSSELLRFQVTDLDEEYTDNWLAVYFFTSG--NEGNWF  267
            DN P+F    Y A+I E     L   +L+   TD D++   N   VYF T    +  N
Sbjct  131  DNPPVFGRPTYQAQITEEDDRNLPKRILQVTATDGDKDRPQN--IVYFLTGQGIDPDNPA  188

Query  268  EIQTDPRTNEGILKVVKALDYEQLQS-VKLSIAVKNKAEFHQSVISRYRVQSTPVTIQVI  326
              + D     G + V+K LD +Q     +    V  + E  + ++    VQ     + +
Sbjct  189  NSKFDINRTSGDIFVLKPLDRDQPNGRPQWRFTVFAQDEGGEGLVGYADVQ-----VNLK  243

Query  327  NVREGIAFRPASKTFTVQKGISSKKLVDYILGTYQAIDEDTNKAASNVK--YVMGRN---  381
            ++ +     P    F     ++       ++ T  A+D D     +N K  Y + +N
Sbjct  244  DINDNAPIFPQGVYF---GNVTENGTAGMVVMTMTAVDYDDPNEGTNAKLIYSIEKNVIE  300

Query  382  ---DGGYLMIDSKTAEIK-FVKNMNRDSTFIVNKTITAEVLAIDEYTGKTSTGTVYVRVP  437
                     I++ T  IK  V  ++R+ T       + +V+A+D   G   TGT  +RV
Sbjct  301  EETGSPIFEIEADTGVIKTAVCCLDRERT----PDYSIQVVAMDG-GGLKGTGTASIRVK  355

Query  438  DFNDNCP  444
            D ND  P
Sbjct  356  DINDMPP  362

 Score = 53.5 bits (127),  Expect = 1e-05, Method: Compositional matrix adjust.
 Identities = 70/306 (23%), Positives = 125/306 (41%), Gaps = 30/306 (10%)

Query  20   PIAKIT-SDYQATQKITYRI---SGVGIDQPPFGIFVVDKNTGDINITAIVDRE---ETP  72
            PI  +T  D   T K  Y++   SG G D+  F +   +  TG + I   +D E   ++
Sbjct  385  PILTVTVHDEDETNKFQYKVIDNSGYGADK--FTMVRNNDGTGSLKIVQPLDYEDQLQSN  442

Query  73   SFLITCRALNAQGLDVEKPLI------LTVKILDINDNPPVFSQQIFMGEIEENSASNSL  126
             F    + +N +G D +          + VK+ DINDN P F +      + EN+
Sbjct  443  GFRFRIQ-VNDKGEDNDNDKYHVAYSWVVVKLRDINDNKPQFERPNIEVSVYENADVGKT  501

Query  127  VMILNATDADEPNHLNSKIAFKIVSQEPAGTPMFLLSRNTGEVRTLTNSLDREQASSYRL  186
            +    ATD D+     SK+++ I   + +       S N     T+   LDRE    +++
Sbjct  502  LETFKATDPDQGGR--SKVSYAI---DRSSDRQRQFSINQEGTVTIQRQLDREVTPRHQV  556

Query  187  VVSGADKDGEGLSTQCECNIKVKDVNDNFPMFRDSQYSARIEENILSSELLRFQVTDLDE  246
             +   D      +      + V+D+NDN P F    Y   + E++   +++    TD D+
Sbjct  557  KILAIDDGIPPKTATATLTVIVQDINDNPPKFL-KDYRPVLPEHVPPRKVVEILATDDDD  615

Query  247  EYTDNWLAVYFFTSGNEGNW----FEIQTDPRTNEG----ILKVVKALDYEQLQSVKLSI  298
                N     F       +     F+++ D +   G    I+  +++ D EQ +   + I
Sbjct  616  RSKSNGPPFQFRLDPGADDIIRASFKVEQDQKGANGDGMAIVSSLRSFDREQQKEYLIPI  675

Query  299  AVKNKA  304
             +K+
Sbjct  676  VIKDHG  681

 Score = 44.3 bits (103),  Expect = 0.009, Method: Compositional matrix adjust.
 Identities = 48/199 (24%), Positives = 73/199 (37%), Gaps = 19/199 (10%)

Query  27   DYQATQKITYRISGVGIDQPPFGIFVVDKNTGDINITAIVDREETPSFLITCRALNAQGL  86
            D     K++Y I      Q  F I       G + I   +DRE TP   +   A++
Sbjct  511  DQGGRSKVSYAIDRSSDRQRQFSI----NQEGTVTIQRQLDREVTPRHQVKILAIDDGIP  566

Query  87   DVEKPLILTVKILDINDNPPVFSQQIFMGEIEENSASNSLVMILNATDADEPNHLNSKIA  146
                   LTV + DINDNPP F +  +   + E+     +V IL   D D
Sbjct  567  PKTATATLTVIVQDINDNPPKFLKD-YRPVLPEHVPPRKVVEILATDDDDRSKSNGPPFQ  625

Query  147  FKIVSQEPAGTPMFLLSRNTGEVRTLTN-----------SLDREQASSYRLVVSGADKDG  195
            F++   +P    +   S    + +   N           S DREQ   Y + +   D
Sbjct  626  FRL---DPGADDIIRASFKVEQDQKGANGDGMAIVSSLRSFDREQQKEYLIPIVIKDHGN  682

Query  196  EGLSTQCECNIKVKDVNDN  214
              ++      + + DVNDN
Sbjct  683  PAMTGTSTLTVVIGDVNDN  701

>AALC636_024560.P34471 | transcript=AALC636_024560.R34471 | gene=AALC636_024560
| organism=Aedes_albopictus_C6/36_cell_line
```

```
| gene_product=neural-cadherin | transcript_product=neural-cadherin
| location=MNAF02000948:246376-395582(+) | protein_length=1717
| sequence_SO=supercontig | SO=protein_coding_gene
| is_pseudo=false
Length=1717

 Score = 146 bits (368),  Expect = 5e-35, Method: Compositional matrix adjust.
 Identities = 130/449 (29%), Positives = 216/449 (48%), Gaps = 47/449 (10%)

Query  21   IAKITSDYQATQKITYRISGVGID--QPPFGIFVVDKNTGDINITAIVDREET---PSFL  75
            +     D    Q I Y ++G GID   P    F +++ +GDI +   +DR++    P +
Sbjct  160  VTATDGDKDRPQNIVYFLTGQGIDPDNPANSKFDINRTSGDIFVLKPLDRDQPNGRPQWR  219

Query  76   ITCRALNAQGLDVEKPLILTVKILDINDNPPVFSQQIFMGEIEENSASNSLVMILNATDA  135
             T  A +  G  +      + V + DINDN P+F Q ++ G + EN  +  +VM + A D
Sbjct  220  FTVFAQDEGGEGLVGYADVQVNLKDINDNAPIFPQGVYFGNVTENGTAGMVVMTMTAVDY  279

Query  136  DEPNH-LNSKIAFKI---VSQEPAGTPMFLLSRNTGEVRTLTNSLDREQASSYRLVVSGA  191
            D+PN   N+K+ + I   V +E  G+P+F +  +TG ++T    LDRE+   Y + V
Sbjct  280  DDPNEGTNAKLIYSIEKNVIEEETGSPIFEIEADTGVIKTAVCCLDRERTPDYSIQVVAM  339

Query  192  DKDGEGLSTQCECNIKVKDVNDNFPMFRDSQYSARIEENILSS--ELLRFQVTDLDEEYT  249
              DG GL       +I+VKD+ND  P F   ++   ++E   ++  E+    VT  DE+ T
Sbjct  340  --DGGGLKGTGTASIRVKDINDMPPQFTKDEWFTEVDETDGTNLPEMPILTVTVHDEDET  397

Query  250  DNWLAVYFFTSGNEGNWFEIQTDPRTNEGI--LKVVKALDYE-QLQS--VKLSIAVKNKA  304
            + +        SG    + F +    R N+G   LK+V+ LDYE QLQS   +  I V +K
Sbjct  398  NKFQYKVIDNSGYGADKFTMV---RNNDGTGSLKIVQPLDYEDQLQSNGFRFRIQVNDKG  454

Query  305  EFHQSVISRYRVQSTPVTIQVINVREGIAFRPASKTFTVQKGISSKKLVDYILGTYQAID  364
            E + +   +Y V  + V +++ ++ +    +P  +   ++  +      V   L T++A D
Sbjct  455  EDNDN--DKYHVAYSWVVVKLRDINDN---KPQFERPNIEVSVYENADVGKTLETFKATD  509

Query  365  EDTNKAASNVKYVMGRNDGGYLMIDSKTAEIKFVKNMNRDSTFIVNKTITAEV-------  417
             D     S V Y + R          S   + +F  ++N++ T  + + +  EV
Sbjct  510  PDQG-GRSKVSYAIDR---------SSDRQRQF--SINQEGTVTIQRQLDREVTPRHQVK  557

Query  418  -LAIDE-YTGKTSTGTVYVRVPDFNDNCP  444
             LAID+    KT+T T+ V V D NDN P
Sbjct  558  ILAIDDGIPPKTATATLTVIVQDINDNPP  586

 Score = 83.6 bits (205),  Expect = 6e-15, Method: Compositional matrix adjust.
 Identities = 98/367 (27%), Positives = 158/367 (43%), Gaps = 40/367 (11%)

Query  93   ILTVKILDINDNPPVFSQQIFMGEIEENSASNSLVMILNATDADEPNHLNSKIAFKIVSQ  152
             + + I D NDNPP F + ++  E++EN      V+ + A D DE    +S+I ++I S
Sbjct  21   FIRIGIADKNDNPPYFDKALYEAEVDENEDIQHTVLTVTAKDHDE----SSRIRYEITSG  76

Query  153  EPAGTPMFLLSRNTGEVRTLTNSLDREQASSYRLVVSGADKDGEGLSTQCECNIKVKDVN  212
               G   F +   TG +  +  +LD E    Y L ++ +D   E  +T      I VKDVN
Sbjct  77   NIGGA--FAVKNMTGAI-YVAGALDYETRKRYELRLAASDNLKENYTTVV---IHVKDVN  130

Query  213  DNFPMFRDSQYSARIEENI---LSSELLRFQVTDLDEEYTDNWLAVYFFTSG--NEGNWF  267
            DN P+F    Y A+I E     L   +L+   TD D++   N   VYF T    +  N
Sbjct  131  DNPPVFGRPTYQAQITEEDDRNLPKRILQVTATDGDKDRPQN--IVYFLTGQGIDPDNPA  188

Query  268  EIQTDPRTNEGILKVVKALDYEQLQS-VKLSIAVKNKAEFHQSVISRYRVQSTPVTIQVI  326
              + D     G + V+K LD +Q      +    V  + E  + ++    VQ     + +
Sbjct  189  NSKFDINRTSGDIFVLKPLDRDQPNGRPQWRFTVFAQDEGGEGLVGYADVQ-----VNLK  243

Query  327  NVREGIAFRPASKTFTVQKGISSKKLVDYILGTYQAIDEDTNKAASNVK--YVMGRN---  381
            ++ +      P    F     ++         ++ T  A+D D     +N K  Y + +N
Sbjct  244  DINDNAPIFPQGVYF---GNVTENGTAGMVVMTMTAVDYDDPNEGTNAKLIYSIEKNVIE  300

Query  382  ---DGGYLMIDSKTAEIK-FVKNMNRDSTFIVNKTITAEVLAIDEYTGKTSTGTVYVRVP  437
                     I++ T  IK  V  ++R+ T        + +V+A+D   G   TGT  +RV
Sbjct  301  EETGSPIFEIEADTGVIKTAVCCLDRERT----PDYSIQVVAMDG-GGLKGTGTASIRVK  355

Query  438  DFNDNCP  444
            D ND  P
```

```
Sbjct  356  DINDMPP  362

 Score = 53.5 bits (127),  Expect = 1e-05, Method: Compositional matrix adjust.
 Identities = 70/306 (23%), Positives = 125/306 (41%), Gaps = 30/306 (10%)

Query  20   PIAKIT-SDYQATQKITYRI---SGVGIDQPPFGIFVVDKNTGDINITAIVDRE---ETP  72
            PI  +T  D   T K  Y++   SG G D+  F +   +  TG + I   +D E   ++
Sbjct  385  PILTVTVHDEDETNKFQYKVIDNSGYGADK--FTMVRNNDGTGSLKIVQPLDYEDQLQSN  442

Query  73   SFLITCRALNAQGLDVEKPLI------LTVKILDINDNPPVFSQQIFMGEIEENSASNSL  126
             F    + +N +G D +           + VK+ DINDN P F +       + EN+
Sbjct  443  GFRFRIQ-VNDKGEDNDNDKYHVAYSWVVVKLRDINDNKPQFERPNIEVSVYENADVGKT  501

Query  127  VMILNATDADEPNHLNSKIAFKIVSQEPAGTPMFLLSRNTGEVRTLTNSLDREQASSYRL  186
            +    ATD D+      SK+++ I   + +        S N      T+   LDRE    +++
Sbjct  502  LETFKATDPDQGGR--SKVSYAI---DRSSDRQRQFSINQEGTVTIQRQLDREVTPRHQV  556

Query  187  VVSGADKDGEGLSTQCECNIKVKDVNDNFPMFRDSQYSARIEENILSSELLRFQVTDLDE  246
             +   D      +       + V+D+NDN P F    Y   + E++   +++     TD D+
Sbjct  557  KILAIDDGIPPKTATATLTVIVQDINDNPPKFL-KDYRPVLPEHVPPRKVVEILATDDDD  615

Query  247  EYTDNWLAVYFFTSGNEGNW----FEIQTDPRTNEG----ILKVVKALDYEQLQSVKLSI  298
               N     F       +     F+++ D +   G    I+  +++ D EQ +   + I
Sbjct  616  RSKSNGPPFQFRLDPGADDIIRASFKVEQDQKGANGDGMAIVSSLRSFDREQQKEYLIPI  675

Query  299  AVKNKA  304
             +K+
Sbjct  676  VIKDHG  681

 Score = 44.3 bits (103),  Expect = 0.009, Method: Compositional matrix adjust.
 Identities = 48/199 (24%), Positives = 73/199 (37%), Gaps = 19/199 (10%)

Query  27   DYQATQKITYRISGVGIDQPPFGIFVVDKNTGDINITAIVDREETPSFLITCRALNAQGL  86
            D     K++Y I      Q  F I        G + I   +DRE TP   +   A++
Sbjct  511  DQGGRSKVSYAIDRSSDRQRQFSI----NQEGTVTIQRQLDREVTPRHQVKILAIDDGIP  566

Query  87   DVEKPLILTVKILDINDNPPVFSQQIFMGEIEENSASNSLVMILNATDADEPNHLNSKIA  146
                   LTV + DINDNPP F +  +   + E+      +V IL   D D
Sbjct  567  PKTATATLTVIVQDINDNPPKFLKD-YRPVLPEHVPPRKVVEILATDDDDRSKSNGPPFQ  625

Query  147  FKIVSQEPAGTPMFLLSRNTGEVRTLTN-----------SLDREQASSYRLVVSGADKDG  195
            F++    +P    +   S    + +   N           S DREQ   Y + +   D
Sbjct  626  FRL---DPGADDIIRASFKVEQDQKGANGDGMAIVSSLRSFDREQQKEYLIPIVIKDHGN  682

Query  196  EGLSTQCECNIKVKDVNDN  214
             ++      + + DVNDN
Sbjct  683  PAMTGTSTLTVVIGDVNDN  701

>SCYAPA1_024866.P32618 | transcript=SCYAPA1_024866.R32618 | gene=SCYAPA1_024866
| organism=Sabethes_cyaneus_ScyaPA1 | gene_product=neural-cadherin
| transcript_product=neural-cadherin
| location=ScyaPA1_3:224633026-224769901(-) | protein_length=1717
| sequence_SO=chromosome | SO=protein_coding_gene |
is_pseudo=false
Length=1717

 Score = 145 bits (367),  Expect = 6e-35, Method: Compositional matrix adjust.
 Identities = 130/444 (29%), Positives = 215/444 (48%), Gaps = 47/444 (11%)

Query  26   SDYQATQKITYRISGVGID--QPPFGIFVVDKNTGDINITAIVDREET---PSFLITCRA  80
             D    Q I Y ++G GID    P     F +++ TG+I +    +DR++     P +  T  A
Sbjct  165  GDKDRPQNIVYFLTGQGIDPDNPANSKFDINRTTGEIFVLKPLDRDQPNGRPQWRFTVFA  224

Query  81   LNAQGLDVEKPLILTVKILDINDNPPVFSQQIFMGEIEENSASNSLVMILNATDADEPNH  140
             +  G  +      + V + DINDN P+F Q ++ G + EN  +  +VM + A D D+PN
Sbjct  225  QDEGGEGLVGYADVQVNLKDINDNAPIFPQGVYFGNVTENGTAGMVVMTMTAVDYDDPNE  284

Query  141  -LNSKIAFKI---VSQEPAGTPMFLLSRNTGEVRTLTNSLDREQASSYRLVVSGADKDGE  196
```

```
                 N+K+ + I   V +E  G+P+F +   +TG ++T    LDRE+   Y + V     DG
Sbjct  285  GTNAKLIYSIEKNVIEEETGSPIFEIEADTGVIKTAVCCLDRERTPDYSIQVVAM--DGG  342

Query  197  GLSTQCECNIKVKDVNDNFPMFRDSQYSARIEENILSS--ELLRFQVTDLDEEYTDNWLA  254
            GL       +I+VKD+ND  P F   ++   ++E   ++  E+    VT  DE+ T+ +
Sbjct  343  GLKGTGTASIRVKDINDMPPQFTKDEWFTEVDETDGTNLPEMPILTVTVHDEDETNKFQY  402

Query  255  VYFFTSGNEGNWFEIQTDPRTNEGI--LKVVKALDYE-QLQS--VKLSIAVKNKAEFHQS  309
                 SG    + F +    R N+G   LK+V+ LDYE QLQS   +  I V +K E + +
Sbjct  403  KVIDNSGYGADKFTMV---RNNDGTGSLKIVQPLDYEDQLQSNGFRFRIQVNDKGEDNDN  459

Query  310  VISRYRVQSTPVTIQVINVREGIAFRPASKTFTVQKGISSKKLVDYILGTYQAIDEDTNK  369
               +Y V  + V +++ ++ +    +P  +   ++  +     V   L T++A D D
Sbjct  460  --DKYHVAYSWVVVKLRDINDN---KPQFERPNIEVSVYENADVGKTLETFKATDPDQG-  513

Query  370  AASNVKYVMGRNDGGYLMIDSKTAEIKFVKNMNRDSTFIVNKTITAEV--------LAID  421
              S V Y + R          S   + +F  ++N++ T  + + +  EV         LAID
Sbjct  514  GKSKVSYAIDR---------SSDRQRQF--SINQEGTVTIQRQLDREVTPRHQVKILAID  562

Query  422  E-YTGKTSTGTVYVRVPDFNDNCP  444
            +    KT+T T+ V V D NDN P
Sbjct  563  DGIPPKTATATLTVIVQDINDNPP  586

 Score = 80.5 bits (197),  Expect = 6e-14, Method: Compositional matrix adjust.
 Identities = 96/367 (26%), Positives = 157/367 (43%), Gaps = 40/367 (11%)

Query  93   ILTVKILDINDNPPVFSQQIFMGEIEENSASNSLVMILNATDADEPNHLNSKIAFKIVSQ  152
              + + I D NDNPP F + ++  E++EN       V+ + A D DE    +S+I ++I S
Sbjct  21   FIRIGIADKNDNPPYFDKALYEAEVDENEDIQHTVLTVTAKDHDE----SSRIRYEITSG  76

Query  153  EPAGTPMFLLSRNTGEVRTLTNSLDREQASSYRLVVSGADKDGEGLSTQCECNIKVKDVN  212
               G   F +   TG +  +  +LD E    Y L ++ +D   E  +T      I VKDVN
Sbjct  77   NIGGA--FAVKNMTGAI-YVAGALDYETRKRYELRLAASDNLKENYTTVV---IHVKDVN  130

Query  213  DNFPMFRDSQYSARIEENI---LSSELLRFQVTDLDEEYTDNWLAVYFFTSG--NEGNWF  267
            DN P+F    Y  +I E     L   +L+   +D D++   N   VYF T    +  N
Sbjct  131  DNPPVFERPTYRTQITEEDDRNLPKRVLQVTASDGDKDRPQN--IVYFLTGQGIDPDNPA  188

Query  268  EIQTDPRTNEGILKVVKALDYEQLQS-VKLSIAVKNKAEFHQSVISRYRVQSTPVTIQVI  326
              + D     G + V+K LD +Q      +     V  + E  + ++    VQ     + +
Sbjct  189  NSKFDINRTTGEIFVLKPLDRDQPNGRPQWRFTVFAQDEGGEGLVGYADVQ-----VNLK  243

Query  327  NVREGIAFRPASKTFTVQKGISSKKLVDYILGTYQAIDEDTNKAASNVK--YVMGRN---  381
            ++ +      P    F      ++         ++ T  A+D D     +N K  Y + +N
Sbjct  244  DINDNAPIFPQGVYF---GNVTENGTAGMVVMTMTAVDYDDPNEGTNAKLIYSIEKNVIE  300

Query  382  ---DGGYLMIDSKTAEIK-FVKNMNRDSTFIVNKTITAEVLAIDEYTGKTSTGTVYVRVP  437
                      I++ T  IK  V  ++R+ T       + +V+A+D   G   TGT  +RV
Sbjct  301  EETGSPIFEIEADTGVIKTAVCCLDRERT----PDYSIQVVAMDG-GGLKGTGTASIRVK  355

Query  438  DFNDNCP  444
            D ND  P
Sbjct  356  DINDMPP  362

 Score = 53.5 bits (127),  Expect = 1e-05, Method: Compositional matrix adjust.
 Identities = 70/306 (23%), Positives = 125/306 (41%), Gaps = 30/306 (10%)

Query  20   PIAKIT-SDYQATQKITYRI---SGVGIDQPPFGIFVVDKNTGDINITAIVDRE---ETP  72
            PI  +T  D   T K  Y++    SG G D+  F +    +  TG + I   +D E   ++
Sbjct  385  PILTVTVHDEDETNKFQYKVIDNSGYGADK--FTMVRNNDGTGSLKIVQPLDYEDQLQSN  442

Query  73   SFLITCRALNAQGLDVEKPLI------LTVKILDINDNPPVFSQQIFMGEIEENSASNSL  126
              F     + +N +G D +            + VK+ DINDN P F +       + EN+
Sbjct  443  GFRFRIQ-VNDKGEDNDNDKYHVAYSWVVVKLRDINDNKPQFERPNIEVSVYENADVGKT  501

Query  127  VMILNATDADEPNHLNSKIAFKIVSQEPAGTPMFLLSRNTGEVRTLTNSLDREQASSYRL  186
            +    ATD D+      SK+++ I    + +        S N     T+   LDRE    +++
Sbjct  502  LETFKATDPDQGG--KSKVSYAI---DRSSDRQRQFSINQEGTVTIQRQLDREVTPRHQV  556
```

```
Query  187  VVSGADKDGEGLSTQCECNIKVKDVNDNFPMFRDSQYSARIEENILSSELLRFQVTDLDE  246
              +   D       +       + V+D+NDN P F    Y   + E++   +++    TD D+
Sbjct  557  KILAIDDGIPPKTATATLTVIVQDINDNPPKFL-KDYRPVLPEHVPPRKVVEILATDDDD  615

Query  247  EYTDNWLAVYFFTSGNEGNW----FEIQTDPRTNEG----ILKVVKALDYEQLQSVKLSI  298
                N     F       +     F+++ D +   G    I+  +++ D EQ +    + I
Sbjct  616  RSKSNGPPFQFRLDPGADDIIRASFKVEQDQKGANGDGMAIVSSLRSFDREQQKEYLIPI  675

Query  299  AVKNKA  304
             +K+
Sbjct  676  VIKDHG  681

 Score = 44.7 bits (104),  Expect = 0.008, Method: Compositional matrix adjust.
 Identities = 48/199 (24%), Positives = 73/199 (37%), Gaps = 19/199 (10%)

Query  27   DYQATQKITYRISGVGIDQPPFGIFVVDKNTGDINITAIVDREETPSFLITCRALNAQGL  86
            D     K++Y I      Q  F I        G + I   +DRE TP    +    A++
Sbjct  511  DQGGKSKVSYAIDRSSDRQRQFSI----NQEGTVTIQRQLDREVTPRHQVKILAIDDGIP  566

Query  87   DVEKPLILTVKILDINDNPPVFSQQIFMGEIEENSASNSLVMILNATDADEPNHLNSKIA  146
                     LTV + DINDNPP F +   +   + E+     +V IL   D D
Sbjct  567  PKTATATLTVIVQDINDNPPKFLKD-YRPVLPEHVPPRKVVEILATDDDDRSKSNGPPFQ  625

Query  147  FKIVSQEPAGTPMFLLSRNTGEVRTLTN-----------SLDREQASSYRLVVSGADKDG  195
            F++    +P     +   S     + +    N            S DREQ    Y + +    D
Sbjct  626  FRL---DPGADDIIRASFKVEQDQKGANGDGMAIVSSLRSFDREQQKEYLIPIVIKDHGN  682

Query  196  EGLSTQCECNIKVKDVNDN  214
              ++        + + DVNDN
Sbjct  683  PAMTGTSTLTVVIGDVNDN  701

>CQUJHB012679.P19663 | transcript=CQUJHB012679.R19663 | gene=CQUJHB012679
| organism=Culex_quinquefasciatus_JHB_2020 | gene_product=neural-cadherin
| transcript_product=neural-cadherin
| location=CM027411.1:213863109-213962495(+) | protein_length=1717
| sequence_SO=chromosome | SO=protein_coding_gene
| is_pseudo=false
Length=1717

 Score = 145 bits (366),  Expect = 7e-35, Method: Compositional matrix adjust.
 Identities = 130/449 (29%), Positives = 216/449 (48%), Gaps = 47/449 (10%)

Query  21   IAKITSDYQATQKITYRISGVGID--QPPFGIFVVDKNTGDINITAIVDREET---PSFL  75
            +      D     Q I Y ++G GID    P     F +++ TG+I +    +DR++     P +
Sbjct  160  VTATDGDKDRPQNIVYFLTGQGIDPDNPANSKFDINRTTGEIFVLKPLDRDQPNGRPQWR  219

Query  76   ITCRALNAQGLDVEKPLILTVKILDINDNPPVFSQQIFMGEIEENSASNSLVMILNATDA  135
             T   A +   G   +      + V + DINDN P+F Q ++ G + EN   +  +VM + A D
Sbjct  220  FTVFAQDEGGEGLVGYADVQVNLKDINDNAPIFPQGVYFGNVTENGTAGMVVMTMTAVDY  279

Query  136  DEPNH-LNSKIAFKI---VSQEPAGTPMFLLSRNTGEVRTLTNSLDREQASSYRLVVSGA  191
            D+PN   N+K+ + I   V +E  G+P+F +   +TG ++T    LDRE+    Y + V
Sbjct  280  DDPNEGTNAKLIYSIEKNVIEEETGSPIFEIEADTGVIKTAVCCLDRERTPDYSIQVVAM  339

Query  192  DKDGEGLSTQCECNIKVKDVNDNFPMFRDSQYSARIEENILSS--ELLRFQVTDLDEEYT  249
              DG GL       +I+VKD+ND  P F   ++   ++E   ++  E+    VT  DE+ T
Sbjct  340  --DGGGLKGTGTASIRVKDINDMPPQFTKDEWFTEVDETDGTNLPEMPILTVTVHDEDET  397

Query  250  DNWLAVYFFTSGNEGNWFEIQTDPRTNEGI--LKVVKALDYE-QLQS--VKLSIAVKNKA  304
            + +         SG    + F +     R N+G   LK+V+ LDYE QLQS    +  I V +K
Sbjct  398  NKFQYKVIDNSGYGADKFTMV---RNNDGTGSLKIVQPLDYEDQLQSNGFRFRIQVNDKG  454

Query  305  EFHQSVISRYRVQSTPVTIQVINVREGIAFRPASKTFTVQKGISSKKLVDYILGTYQAID  364
            E + +    +Y V   + V +++ ++ +     +P   +    ++   +       V   L T++A D
Sbjct  455  EDNDN--DKYHVAYSWVVVKLRDINDN---KPQFERPNIEVSVYENADVGKTLETFKATD  509

Query  365  EDTNKAASNVKYVMGRNDGGYLMIDSKTAEIKFVVKNMNRDSTFIVNKTITAEV-------  417
             D     S V Y + R           S    + +F   ++N++  T   + + +  EV
Sbjct  510  PDQG-GKSKVSYAIDR---------SSDRQRQF--SINQEGTVTIQRQLDREVTPRHQVK  557
```

```
Query  418  -LAIDE-YTGKTSTGTVYYVRVPDFNDNCP  444
             LAID+    KT+T T+ V V D NDN P
Sbjct  558  ILAIDDGIPPKTATATLTVIVQDINDNPP  586

 Score = 81.3 bits (199),  Expect = 3e-14, Method: Compositional matrix adjust.
 Identities = 97/367 (26%), Positives = 157/367 (43%), Gaps = 40/367 (11%)

Query  93   ILTVKILDINDNPPVFSQQIFMGEIEENSASNSLVMILNATDADEPNHLNSKIAFKIVSQ  152
             + + I D NDNPP F + ++  E++EN      V+ + A D DE    +S+I ++I S
Sbjct  21   FIRIGIADKNDNPPYFDKALYEAEVDENEDIQHTVLTVTAKDHDE----SSRIRYEITSG  76

Query  153  EPAGTPMFLLSRNTGEVRTLTNSLDREQASSYRLVVSGADKDGEGLSTQCECNIKVKDVN  212
               G   F +   TG +  +  +LD E    Y L ++ +D   E  +T     I VKDVN
Sbjct  77   NIGGA--FAVKNMTGAI-YVAGALDYETRKRYELRLAASDNLKENYTTVV---IHVKDVN  130

Query  213  DNFPMFRDSQYSARIEENI---LSSELLRFQVTDLDEEYTDNWLAVYFFTSG--NEGNWF  267
            DN P+F    Y  +I E     L   +L+   TD D++   N   VYF T    +  N
Sbjct  131  DNPPVFERPTYRTQITEEDDRNLPKRVLQVTATDGDKDRPQN--IVYFLTGQGIDPDNPA  188

Query  268  EIQTDPRTNEGILKVVKALDYEQLQS-VKLSIAVKNKAEFHQSVISRYRVQSTPVTIQVI  326
              + D     G + V+K LD +Q     +    V  + E  + ++    VQ     + +
Sbjct  189  NSKFDINRTTGEIFVLKPLDRDQPNGRPQWRFTVFAQDEGGEGLVGYADVQ-----VNLK  243

Query  327  NVREGIAFRPASKTFTVQKGISSKKLVDYILGTYQAIDEDTNKAASNVK--YVMGRN---  381
            ++ +     P    F     ++       ++ T  A+D D     +N K  Y + +N
Sbjct  244  DINDNAPIFPQGVYF---GNVTENGTAGMVVMTMTAVDYDDPNEGTNAKLIYSIEKNVIE  300

Query  382  ---DGGYLMIDSKTAEIK-FVKNMNRDSTFIVNKTITAEVLAIDEYTGKTSTGTVYYVRVP  437
                     I++ T  IK  V  ++R+ T       + +V+A+D   G   TGT  +RV
Sbjct  301  EETGSPIFEIEADTGVIKTAVCCLDRERT----PDYSIQVVAMDG-GGLKGTGTASIRVK  355

Query  438  DFNDNCP  444
            D ND  P
Sbjct  356  DINDMPP  362

 Score = 53.1 bits (126),  Expect = 2e-05, Method: Compositional matrix adjust.
 Identities = 70/306 (23%), Positives = 125/306 (41%), Gaps = 30/306 (10%)

Query  20   PIAKIT-SDYQATQKITYRI---SGVGIDQPPFGIFVVDKNTGDINITAIVDRE---ETP  72
            PI  +T  D   T K  Y++   SG G D+  F +   +  TG + I   +D E   ++
Sbjct  385  PILTVTVHDEDETNKFQYKVIDNSGYGADK--FTMVRNNDGTGSLKIVQPLDYEDQLQSN  442

Query  73   SFLITCRALNAQGLDVEKPLI------LTVKILDINDNPPVFSQQIFMGEIEENSASNSL  126
             F    + +N +G D +          + VK+ DINDN P F +      + EN+
Sbjct  443  GFRFRIQ-VNDKGEDNDNDKYHVAYSWVVVKLRDINDNKPQFERPNIEVSVYENADVGKT  501

Query  127  VMILNATDADEPNHLNSKIAFKIVSQEPAGTPMFLLSRNTGEVRTLTNSLDREQASSYRL  186
            +    ATD D+     SK+++ I   + +       S N     T+   LDRE    +++
Sbjct  502  LETFKATDPDQGG--KSKVSYAI---DRSSDRQRQFSINQEGTVTIQRQLDREVTPRHQV  556

Query  187  VVSGADKDGEGLSTQCECNIKVKDVNDNFPMFRDSQYSARIEENILSSELLRFQVTDLDE  246
             +   D      +      + V+D+NDN P F    Y   + E++   +++    TD D+
Sbjct  557  KILAIDDGIPPKTATATLTVIVQDINDNPPKFL-KDYRPVLPEHVPPRKVVEILATDDDD  615

Query  247  EYTDNWLAVYFFTSGNEGNW----FEIQTDPRTNEG----ILKVVKALDYEQLQSVKLSI  298
                N     F       +     F+++ D +   G    I+  +++ D EQ +   + I
Sbjct  616  RSKSNGPPFQFRLDPGADDIIRASFKVEQDQKGANGDGMAIVSSLRSFDREQQKEYLIPI  675

Query  299  AVKNKA  304
             +K+
Sbjct  676  VIKDHG  681

 Score = 44.3 bits (103),  Expect = 0.009, Method: Compositional matrix adjust.
 Identities = 48/199 (24%), Positives = 73/199 (37%), Gaps = 19/199 (10%)

Query  27   DYQATQKITYRISGVGIDQPPFGIFVVDKNTGDINITAIVDREETPSFLITCRALNAQGL  86
            D     K++Y I      Q  F I       G + I   +DRE TP   +   A++
```

```
Sbjct  511  DQGGKSKVSYAIDRSSDRQRQFSI----NQEGTVTIQRQLDREVTPRHQVKILAIDDGIP  566

Query  87   DVEKPLILTVKILDINDNPPVFSQQIFMGEIEENSASNSLVMILNATDADEPNHLNSKIA  146
                   LTV + DINDNPP F +  +   + E+     +V IL   D D
Sbjct  567  PKTATATLTVIVQDINDNPPKFLKD-YRPVLPEHVPPRKVVEILATDDDDRSKSNGPPFQ  625

Query  147  FKIVSQEPAGTPMFLLSRNTGEVRTLTN-----------SLDREQASSYRLVVSGADKDG  195
            F++   +P    +   S    + +   N           S DREQ   Y + +   D
Sbjct  626  FRL---DPGADDIIRASFKVEQDQKGANGDGMAIVSSLRSFDREQQKEYLIPIVIKDHGN  682

Query  196  EGLSTQCECNIKVKDVNDN  214
              ++      + + DVNDN
Sbjct  683  PAMTGTSTLTVVIGDVNDN  701

>ASTEI02004-PA | transcript=ASTEI02004-RA | gene=ASTEI02004 |
organism=Anopheles_stephensi_Indian | gene_product=unspecified
product | transcript_product=unspecified product |
location=KE388896:2997203-3024673(+)
| protein_length=1652 | sequence_SO=supercontig
| SO=protein_coding_gene | is_pseudo=false
Length=1652

 Score = 145 bits (365),  Expect = 1e-34, Method: Compositional matrix adjust.
 Identities = 125/442 (28%), Positives = 212/442 (48%), Gaps = 33/442 (7%)

Query  21   IAKITSDYQATQKITYRISGVGID--QPPFGIFVVDKNTGDINITAIVDREET---PSFL  75
            +     D    Q I Y ++G GID   P    F +++ TG+I +   +DR++    P +
Sbjct  23   VTATDGDKDRPQNIVYFLTGQGIDPDNPANSKFDINRTTGEIFVLKPLDRDQPNGRPQWR  82

Query  76   ITCRALNAQGLDVEKPLILTVKILDINDNPPVFSQQIFMGEIEENSASNSLVMILNATDA  135
             T  A +  G  +     + V + DINDN P+F Q ++ G + EN  +  +VM + A D
Sbjct  83   FTVFAQDEGGEGLVGYADVQVNLKDINDNAPIFPQGVYFGNVTENGTAGMVVMTMTAVDY  142

Query  136  DEPNH-LNSKIAFKI---VSQEPAGTPMFLLSRNTGEVRTLTNSLDREQASSYRLVVSGA  191
            D+PN   N+K+ + I   V +E  G+P+F +  +TG ++T    LDRE+   Y + V
Sbjct  143  DDPNEGTNAKLIYSIEKNVIEEETGSPIFEIEADTGVIKTAVCCLDRERTPDYSIQVVAM  202

Query  192  DKDGEGLSTQCECNIKVKDVNDNFPMFRDSQYSARIEENILSS--ELLRFQVTDLDEEYT  249
              DG GL      +I+VKD+ND  P F    ++   ++E   ++  E+    VT  DE+ T
Sbjct  203  --DGGGLKGTGTASIRVKDINDMPPQFTKDEWFTEVDETDGTNLPEMPILTVTVHDEDET  260

Query  250  DNWLAVYFFTSGNEGNWFEIQTDPRTNEGI--LKVVKALDYE---QLQSVKLSIAVKNKA  304
            + +       SG   + F +    R N+G   LK+V+ LDYE   Q    +  I V +K
Sbjct  261  NKFQYKVIDNSGYGADKFTMV---RNNDGTGSLKIVQPLDYEDPLQSNGFRFRIQVNDKG  317

Query  305  EFHQSVISRYRVQSTPVTIQVINVREGIAFRPASKTFTVQKGISSKKLVDYILGTYQAID  364
            E + +   +Y V  + V +++ ++ +    +P  +   ++  +     V   L T++A D
Sbjct  318  EDNDN--DKYHVAYSWVVVKLRDINDN---KPQFERPNIEVSVYENADVGKTLETFKATD  372

Query  365  EDTNKAASNVKYVMGR-NDGGYLMIDSKTAEIKFVKNMNRDSTFIVNKTITAEVLAIDE-  422
             D     S V Y + R +D       ++   +   +N++R+    V      ++LAID+
Sbjct  373  PDQG-GKSKVSYAIDRSSDRQRQFSINQEGTVTIQRNLDRE----VTPRHQVKILAIDDG  427

Query  423  YTGKTSTGTVYVRVPDFNDNCP  444
               KT+T T+ V V D NDN P
Sbjct  428  IPPKTATATLTVIVQDINDNPP  449

 Score = 53.5 bits (127),  Expect = 1e-05, Method: Compositional matrix adjust.
 Identities = 70/306 (23%), Positives = 126/306 (41%), Gaps = 30/306 (10%)

Query  20   PIAKIT-SDYQATQKITYRI---SGVGIDQPPFGIFVVDKNTGDINITAIVDRE---ETP  72
            PI  +T  D   T K  Y++   SG G D+  F +   +  TG + I   +D E   ++
Sbjct  248  PILTVTVHDEDETNKFQYKVIDNSGYGADK--FTMVRNNDGTGSLKIVQPLDYEDPLQSN  305

Query  73   SFLITCRALNAQGLDVEKPLI------LTVKILDINDNPPVFSQQIFMGEIEENSASNSL  126
             F    + +N +G D +          + VK+ DINDN P F +      + EN+
Sbjct  306  GFRFRIQ-VNDKGEDNDNDKYHVAYSWVVVKLRDINDNKPQFERPNIEVSVYENADVGKT  364

Query  127  VMILNATDADEPNHLNSKIAFKIVSQEPAGTPMFLLSRNTGEVRTLTNSLDREQASSYRL  186
```

```
            +    ATD D+     SK+++ I   + +       S N     T+  +LDRE    +++
Sbjct  365  LETFKATDPDQGG--KSKVSYAI---DRSSDRQRQFSINQEGTVTIQRNLDREVTPRHQV  419

Query  187  VVSGADKDGEGLSTQCECNIKVKDVNDNFPMFRDSQYSARIEENILSSELLRFQVTDLDE  246
             +   D      +      + V+D+NDN P F    Y   + E++   +++    TD D+
Sbjct  420  KILAIDDGIPPKTATATLTVIVQDINDNPPKFL-KDYRPVLPEHVPPRKVVEILATDDDD  478

Query  247  EYTDNWLAVYFFTSGNEGNW----FEIQTDPRTNEG----ILKVVKALDYEQLQSVKLSI  298
                N     F       +     F+++ D +   G    I+  +++ D EQ +   + I
Sbjct  479  RSKSNGPPFQFRLDPGADDIIRASFKVEQDQKGANGDGMAIVSSLRSFDREQQKEYLIPI  538

Query  299  AVKNKA  304
             +K+
Sbjct  539  VIKDHG  544

 Score = 43.9 bits (102),  Expect = 0.012, Method: Compositional matrix adjust.
 Identities = 48/199 (24%), Positives = 73/199 (37%), Gaps = 19/199 (10%)

Query  27   DYQATQKITYRISGVGIDQPPFGIFVVDKNTGDINITAIVDREETPSFLITCRALNAQGL  86
            D     K++Y I      Q  F I       G + I   +DRE TP   +   A++
Sbjct  374  DQGGKSKVSYAIDRSSDRQRQFSI----NQEGTVTIQRNLDREVTPRHQVKILAIDDGIP  429

Query  87   DVEKPLILTVKILDINDNPPVFSQQIFMGEIEENSASNSLVMILNATDADEPNHLNSKIA  146
                   LTV + DINDNPP F +  +   + E+     +V IL   D D
Sbjct  430  PKTATATLTVIVQDINDNPPKFLKD-YRPVLPEHVPPRKVVEILATDDDDRSKSNGPPFQ  488

Query  147  FKIVSQEPAGTPMFLLSRNTGEVRTLTN-----------SLDREQASSYRLVVSGADKDG  195
            F++   +P    +   S    + +   N           S DREQ   Y + +   D
Sbjct  489  FRL---DPGADDIIRASFKVEQDQKGANGDGMAIVSSLRSFDREQQKEYLIPIVIKDHGN  545

Query  196  EGLSTQCECNIKVKDVNDN  214
              ++      + + DVNDN
Sbjct  546  PAMTGTSTLTVVIGDVNDN  564

>ADAR2_001930.P21641 | transcript=ADAR2_001930.R21641 | gene=ADAR2_001930
| organism=Anopheles_darlingi_AdarGF1 | gene_product=neural-cadherin,
transcript variant X1 | transcript_product=neural-cadherin,
transcript variant X1 | location=AdarGF1_2RL:75104590-75171550(+)
| protein_length=1716 | sequence_SO=chromosome
| SO=protein_coding_gene | is_pseudo=false
Length=1716

 Score = 144 bits (364),  Expect = 1e-34, Method: Compositional matrix adjust.
 Identities = 125/442 (28%), Positives = 212/442 (48%), Gaps = 33/442 (7%)

Query  21   IAKITSDYQATQKITYRISGVGID--QPPFGIFVVDKNTGDINITAIVDREET---PSFL  75
            +     D    Q I Y ++G GID   P    F +++ TG+I +   +DR++    P +
Sbjct  160  VTATDGDKDRPQNIVYFLTGQGIDPDNPANSKFDINRTTGEIFVLKPLDRDQPNGRPQWR  219

Query  76   ITCRALNAQGLDVEKPLILTVKILDINDNPPVFSQQIFMGEIEENSASNSLVMILNATDA  135
             T  A +  G  +     + V + DINDN P+F Q ++ G + EN  +  +VM + A D
Sbjct  220  FTVFAQDEGGEGLVGYADVQVNLKDINDNAPIFPQGVYFGNVTENGTAGMVVMTMTAVDY  279

Query  136  DEPNH-LNSKIAFKI---VSQEPAGTPMFLLSRNTGEVRTLTNSLDREQASSYRLVVSGA  191
            D+PN   N+K+ + I   V +E  G+P+F +  +TG ++T    LDRE+   Y + V
Sbjct  280  DDPNEGTNAKLIYSIEKNVIEEETGSPIFEIEADTGVIKTAVCCLDRERTPDYSIQVVAM  339

Query  192  DKDGEGLSTQCECNIKVKDVNDNFPMFRDSQYSARIEENILSS--ELLRFQVTDLDEEYT  249
              DG GL      +I+VKD+ND  P F   ++   ++E   ++  E+    VT  DE+ T
Sbjct  340  --DGGGLKGTGTASIRVKDINDMPPQFTKDEWFTEVDETDGTNLPEMPILTVTVHDEDET  397

Query  250  DNWLAVYFFTSGNEGNWFEIQTDPRTNEGI--LKVVKALDYE---QLQSVKLSIAVKNKA  304
            + +       SG   + F +    R N+G   LK+V+ LDYE   Q    +  I V +K
Sbjct  398  NKFQYKVIDNSGYGADKFTMV---RNNDGTGSLKIVQPLDYEDPLQSNGFRFRIQVNDKG  454

Query  305  EFHQSVISRYRVQSTPVTIQVINVREGIAFRPASKTFTVQKGISSKKLVDYILGTYQAID  364
            E + +   +Y V  + V +++ ++ +    +P  +   ++  +     V   L T++A D
Sbjct  455  EDNDN--DKYHVAYSWVVVKLRDINDN---KPQFERPNIEVSVYENADVGKTLETFKATD  509
```

```
Query  365  EDTNKAASNVKYVMGR-NDGGYLMIDSKTAEIKFVKNMNRDSTFIVNKTITAEVLAIDE-  422
             D     S V Y + R +D       ++  +   +N++R+    V      ++LAID+
Sbjct  510  PDQG-GKSKVSYAIDRSSDRQRQFSINQEGTVTIQRNLDRE----VTPRHQVKILAIDDG  564

Query  423  YTGKTSTGTVYYRVPDFNDNCP  444
               KT+T T+ V V D NDN P
Sbjct  565  IPPKTATATLTVIVQDINDNPP  586

 Score = 81.6 bits (200),  Expect = 2e-14, Method: Compositional matrix adjust.
 Identities = 97/367 (26%), Positives = 157/367 (43%), Gaps = 40/367 (11%)

Query  93   ILTVKILDINDNPPVFSQQIFMGEIEENSASNSLVMILNATDADEPNHLNSKIAFKIVSQ  152
             + + I D NDNPP F + ++  E++EN      V+ + A D DE    +S+I ++I S
Sbjct  21   FIRIGIADKNDNPPYFDKALYEAEVDENEDIQHTVLTVTAKDHDE----SSRIRYEITSG  76

Query  153  EPAGTPMFLLSRNTGEVRTLTNSLDREQASSYRLVVSGADKDGEGLSTQCECNIKVKDVN  212
               G     F + TG +  +  +LD E    Y L ++ +D   E  +T      I VKDVN
Sbjct  77   NIGGA--FAVKNMTGAI-YVAGALDYETRKRYELRLAASDNLKENYTTVV---IHVKDVN  130

Query  213  DNFPMFRDSQYSARIEENI---LSSELLRFQVTDLDEEYTDNWLAVYFFTSG--NEGNWF  267
            DN P+F    Y  +I E     L   +L+   TD D++   N   VYF T    +  N
Sbjct  131  DNPPVFERPTYRTQITEEDDRNLPKRVLQVTATDGDKDRPQN--IVYFLTGQGIDPDNPA  188

Query  268  EIQTDPRTNEGILKVVKALDYEQLQS-VKLSIAVKNKAEFHQSVISRYRVQSTPVTIQVI  326
             + D     G + V+K LD +Q      +    V  + E  + ++    VQ     + +
Sbjct  189  NSKFDINRTTGEIFVLKPLDRDQPNGRPQWRFTVFAQDEGGEGLVGYADVQ-----VNLK  243

Query  327  NVREGIAFRPASKTFTVQKGISSKKLVDYILGTYQAIDEDTNKAASNVK--YVMGRN---  381
            ++ +     P    F     ++       ++ T  A+D D     +N K  Y + +N
Sbjct  244  DINDNAPIFPQGVYF---GNVTENGTAGMVVMTMTAVDYDDPNEGTNAKLIYSIEKNVIE  300

Query  382  ---DGGYLMIDSKTAEIK-FVKNMNRDSTFIVNKTITAEVLAIDEYTGKTSTGTVYVRVP  437
                     I++ T  IK  V  ++R+ T       + +V+A+D   G   TGT  +RV
Sbjct  301  EETGSPIFEIEADTGVIKTAVCCLDRERT----PDYSIQVVAMDG-GGLKGTGTASIRVK  355

Query  438  DFNDNCP  444
            D ND  P
Sbjct  356  DINDMPP  362

 Score = 53.9 bits (128),  Expect = 1e-05, Method: Compositional matrix adjust.
 Identities = 70/306 (23%), Positives = 126/306 (41%), Gaps = 30/306 (10%)

Query  20   PIAKIT-SDYQATQKITYRI---SGVGIDQPPFGIFVVDKNTGDINITAIVDRE---ETP  72
            PI  +T  D   T K  Y++   SG G D+  F +   +  TG + I   +D E   ++
Sbjct  385  PILTVTVHDEDETNKFQYKVIDNSGYGADK--FTMVRNNDGTGSLKIVQPLDYEDPLQSN  442

Query  73   SFLITCRALNAQGLDVEKPLI------LTVKILDINDNPPVFSQQIFMGEIEENSASNSL  126
             F    + +N +G D +           + VK+ DINDN P F +       + EN+
Sbjct  443  GFRFRIQ-VNDKGEDNDNDKYHVAYSWVVVKLRDINDNKPQFERPNIEVSVYENADVGKT  501

Query  127  VMILNATDADEPNHLNSKIAFKIVSQEPAGTPMFLLSRNTGEVRTLTNSLDREQASSYRL  186
            +    ATD D+      SK+++ I   + +         S N      T+  +LDRE    +++
Sbjct  502  LETFKATDPDQGG--KSKVSYAI---DRSSDRQRQFSINQEGTVTIQRNLDREVTPRHQV  556

Query  187  VVSGADKDGEGLSTQCECNIKVKDVNDNFPMFRDSQYSARIEENILSSELLRFQVTDLDE  246
             +   D       +       + V+D+NDN P F    Y   + E++   +++     TD D+
Sbjct  557  KILAIDDGIPPKTATATLTVIVQDINDNPPKFL-KDYRPVLPEHVPPRKVVEILATDDDD  615

Query  247  EYTDNWLAVYFFTSGNEGNW----FEIQTDPRTNEG----ILKVVKALDYEQLQSVKLSI  298
               N      F        +     F+++ D +   G    I+  +++ D EQ +   + I
Sbjct  616  RSKSNGPPFQFRLDPGADDIIRASFKVEQDQKGANGDGMAIVSSLRSFDREQQKEYLIPI  675

Query  299  AVKNKA  304
             +K+
Sbjct  676  VIKDHG  681

 Score = 44.3 bits (103),  Expect = 0.010, Method: Compositional matrix adjust.
 Identities = 48/199 (24%), Positives = 73/199 (37%), Gaps = 19/199 (10%)
```

```
Query  27   DYQATQKITYRISGVGIDQPPFGIFVVDKNTGDINITAIVDREETPSFLITCRALNAQGL  86
            D     K++Y I      Q  F I       G + I   +DRE TP   +   A++
Sbjct  511  DQGGKSKVSYAIDRSSDRQRQFSI----NQEGTVTIQRNLDREVTPRHQVKILAIDDGIP  566

Query  87   DVEKPLILTVKILDINDNPPVFSQQIFMGEIEENSASNSLVMILNATDADEPNHLNSKIA  146
                   LTV + DINDNPP F +  +   + E+     +V IL   D D
Sbjct  567  PKTATATLTVIVQDINDNPPKFLKD-YRPVLPEHVPPRKVVEILATDDDDRSKSNGPPFQ  625

Query  147  FKIVSQEPAGTPMFLLSRNTGEVRTLTN-----------SLDREQASSYRLVVSGADKDG  195
            F++   +P    +   S    + +   N           S DREQ   Y + +   D
Sbjct  626  FRL---DPGADDIIRASFKVEQDQKGANGDGMAIVSSLRSFDREQQKEYLIPIVIKDHGN  682

Query  196  EGLSTQCECNIKVKDVNDN  214
              ++      + + DVNDN
Sbjct  683  PAMTGTSTLTVVIGDVNDN  701

>AALB20_033171.P66492 | transcript=AALB20_033171.R66492 | gene=AALB20_033171
| organism=Anopheles_albimanus_STECLA_2020 |
gene_product=neural-cadherin, transcript variant X38 |
transcript_product=neural-cadherin,
transcript variant X39 | location=CM024292:75144349-75208393(+)
| protein_length=1716 | sequence_SO=chromosome
| SO=protein_coding_gene | is_pseudo=false
Length=1716

 Score = 144 bits (364),  Expect = 1e-34, Method: Compositional matrix adjust.
 Identities = 125/442 (28%), Positives = 212/442 (48%), Gaps = 33/442 (7%)

Query  21   IAKITSDYQATQKITYRISGVGID--QPPFGIFVVDKNTGDINITAIVDREET---PSFL  75
            +     D    Q I Y ++G GID   P    F +++ TG+I +   +DR++    P +
Sbjct  160  VTATDGDKDRPQNIVYFLTGQGIDPDNPANSKFDINRTTGEIFVLKPLDRDQPNGRPQWR  219

Query  76   ITCRALNAQGLDVEKPLILTVKILDINDNPPVFSQQIFMGEIEENSASNSLVMILNATDA  135
             T  A +  G  +     + V + DINDN P+F Q ++ G + EN  +  +VM + A D
Sbjct  220  FTVFAQDEGGEGLVGYADVQVNLKDINDNAPIFPQGVYFGNVTENGTAGMVVMTMTAVDY  279

Query  136  DEPNH-LNSKIAFKI---VSQEPAGTPMFLLSRNTGEVRTLTNSLDREQASSYRLVVSGA  191
            D+PN   N+K+ + I   V +E  G+P+F +  +TG ++T    LDRE+   Y + V
Sbjct  280  DDPNEGTNAKLIYSIEKNVIEEETGSPIFEIEADTGVIKTAVCCLDRERTPDYSIQVVAM  339

Query  192  DKDGEGLSTQCECNIKVKDVNDNFPMFRDSQYSARIEENILSS--ELLRFQVTDLDEEYT  249
              DG GL      +I+VKD+ND  P F   ++   ++E   ++  E+    VT  DE+ T
Sbjct  340  --DGGGLKGTGTASIRVKDINDMPPQFTKDEWFTEVDETDGTNLPEMPILTVTVHDEDET  397

Query  250  DNWLAVYFFTSGNEGNWFEIQTDPRTNEGI--LKVVKALDYE---QLQSVKLSIAVKNKA  304
            + +       SG   + F +    R N+G   LK+V+ LDYE   Q    +  I V +K
Sbjct  398  NKFQYKVIDNSGYGADKFTMV---RNNDGTGSLKIVQPLDYEDPLQSNGFRFRIQVNDKG  454

Query  305  EFHQSVISRYRVQSTPVTIQVINVREGIAFRPASKTFTVQKGISSKKLVDYILGTYQAID  364
            E + +   +Y V  + V +++ ++ +    +P  +   ++  +     V   L T++A D
Sbjct  455  EDNDN--DKYHVAYSWVVVKLRDINDN---KPQFERPNIEVSVYENADVGKTLETFKATD  509

Query  365  EDTNKAASNVKYVMGR-NDGGYLMIDSKTAEIKFVKNMNRDSTFIVNKTITAEVLAIDE-  422
             D     S V Y + R +D       ++   +   +N++R+    V      ++LAID+
Sbjct  510  PDQG-GKSKVSYAIDRSSDRQRQFSINQEGTVTIQRNLDRE----VTPRHQVKILAIDDG  564

Query  423  YTGKTSTGTVYVRVPDFNDNCP  444
               KT+T T+ V V D NDN P
Sbjct  565  IPPKTATATLTVIVQDINDNPP  586

 Score = 81.6 bits (200),  Expect = 2e-14, Method: Compositional matrix adjust.
 Identities = 97/367 (26%), Positives = 157/367 (43%), Gaps = 40/367 (11%)

Query  93   ILTVKILDINDNPPVFSQQIFMGEIEENSASNSLVMILNATDADEPNHLNSKIAFKIVSQ  152
             + + I D NDNPP F + ++  E++EN      V+ + A D DE    +S+I ++I S
Sbjct  21   FIRIGIADKNDNPPYFDKALYEAEVDENEDIQHTVLTVTAKDHDE----SSRIRYEITSG  76

Query  153  EPAGTPMFLLSRNTGEVRTLTNSLDREQASSYRLVVSGADKDGEGLSTQCECNIKVKDVN  212
```

```
               G   F +   TG +  +  +LD E    Y L ++ +D   E  +T     I VKDVN
Sbjct  77   NIGGA--FAVKNMTGAI-YVAGALDYETRKRYELRLAASDNLKENYTTVV---IHVKDVN  130

Query  213  DNFPMFRDSQYSARIEENI---LSSELLRFQVTDLDEEYTDNWLAVYFFTSG--NEGNWF  267
            DN P+F    Y  +I E     L   +L+   TD D++   N   VYF T    +  N
Sbjct  131  DNPPVFERPTYRTQITEEDDRNLPKRVLQVTATDGDKDRPQN--IVYFLTGQGIDPDNPA  188

Query  268  EIQTDPRTNEGILKVVKALDYEQLQS-VKLSIAVKNKAEFHQSVISRYRVQSTPVTIQVI  326
              + D     G + V+K LD +Q     +    V  + E  + ++    VQ     + +
Sbjct  189  NSKFDINRTTGEIFVLKPLDRDQPNGRPQWRFTVFAQDEGGEGLVGYADVQ-----VNLK  243

Query  327  NVREGIAFRPASKTFTVQKGISSKKLVDYILGTYQAIDEDTNKAASNVK--YVMGRN---  381
            ++ +     P    F     ++       ++ T  A+D D     +N K  Y + +N
Sbjct  244  DINDNAPIFPQGVYF---GNVTENGTAGMVVMTMTAVDYDDPNEGTNAKLIYSIEKNVIE  300

Query  382  ---DGGYLMIDSKTAEIK-FVKNMNRDSTFIVNKTITAEVLAIDEYTGKTSTGTVYVRVP  437
                     I++ T  IK  V  ++R+ T       + +V+A+D   G   TGT  +RV
Sbjct  301  EETGSPIFEIEADTGVIKTAVCCLDRERT----PDYSIQVVAMDG-GGLKGTGTASIRVK  355

Query  438  DFNDNCP  444
            D ND  P
Sbjct  356  DINDMPP  362

 Score = 53.5 bits (127),  Expect = 1e-05, Method: Compositional matrix adjust.
 Identities = 70/306 (23%), Positives = 126/306 (41%), Gaps = 30/306 (10%)

Query  20   PIAKIT-SDYQATQKITYRI---SGVGIDQPPFGIFVVDKNTGDINITAIVDRE---ETP  72
            PI  +T  D   T K  Y++   SG G D+  F +   +  TG + I   +D E   ++
Sbjct  385  PILTVTVHDEDETNKFQYKVIDNSGYGADK--FTMVRNNDGTGSLKIVQPLDYEDPLQSN  442

Query  73   SFLITCRALNAQGLDVEKPLI------LTVKILDINDNPPVFSQQIFMGEIEENSASNSL  126
             F    + +N +G D +           + VK+ DINDN P F +      + EN+
Sbjct  443  GFRFRIQ-VNDKGEDNDNDKYHVAYSWVVVKLRDINDNKPQFERPNIEVSVYENADVGKT  501

Query  127  VMILNATDADEPNHLNSKIAFKIVSQEPAGTPMFLLSRNTGEVRTLTNSLDREQASSYRL  186
            +    ATD D+     SK+++ I   + +       S N     T+  +LDRE    +++
Sbjct  502  LETFKATDPDQGG--KSKVSYAI---DRSSDRQRQFSINQEGTVTIQRNLDREVTPRHQV  556

Query  187  VVSGADKDGEGLSTQCECNIKVKDVNDNFPMFRDSQYSARIEENILSSELLRFQVTDLDE  246
             +   D      +      + V+D+NDN P F    Y   + E++   +++    TD D+
Sbjct  557  KILAIDDGIPPKTATATLTVIVQDINDNPPKFL-KDYRPVLPEHVPPRKVVEILATDDDD  615

Query  247  EYTDNWLAVYFFTSGNEGNW----FEIQTDPRTNEG----ILKVVKALDYEQLQSVKLSI  298
                N     F       +     F+++ D +   G    I+  +++ D EQ +   + I
Sbjct  616  RSKSNGPPFQFRLDPGADDIIRASFKVEQDQKGANGDGMAIVSSLRSFDREQQKEYLIPI  675

Query  299  AVKNKA  304
             +K+
Sbjct  676  VIKDHG  681

 Score = 44.3 bits (103),  Expect = 0.010, Method: Compositional matrix adjust.
 Identities = 48/199 (24%), Positives = 73/199 (37%), Gaps = 19/199 (10%)

Query  27   DYQATQKITYRISGVGIDQPPFGIFVVDKNTGDINITAIVDREETPSFLITCRALNAQGL  86
            D     K++Y I      Q  F I       G + I   +DRE TP   +   A++
Sbjct  511  DQGGKSKVSYAIDRSSDRQRQFSI----NQEGTVTIQRNLDREVTPRHQVKILAIDDGIP  566

Query  87   DVEKPLILTVKILDINDNPPVFSQQIFMGEIEENSASNSLVMILNATDADEPNHLNSKIA  146
                   LTV + DINDNPP F +  +   + E+     +V IL   D D
Sbjct  567  PKTATATLTVIVQDINDNPPKFLKD-YRPVLPEHVPPRKVVEILATDDDDRSKSNGPPFQ  625

Query  147  FKIVSQEPAGTPMFLLSRNTGEVRTLTN-----------SLDREQASSYRLVVSGADKDG  195
            F++   +P    +   S    + +   N           S DREQ   Y + +   D
Sbjct  626  FRL---DPGADDIIRASFKVEQDQKGANGDGMAIVSSLRSFDREQQKEYLIPIVIKDHGN  682

Query  196  EGLSTQCECNIKVKDVNDN  214
              ++      + + DVNDN
Sbjct  683  PAMTGTSTLTVVIGDVNDN  701
```

```
>AFAF019387-PA | transcript=AFAF019387-RA | gene=AFAF019387 |
organism=Anopheles_farauti_FAR1 | gene_product=unspecified product
| transcript_product=unspecified product | location=KI915042:9063109-9086764(+)
| protein_length=1349 | sequence_SO=supercontig
| SO=protein_coding_gene | is_pseudo=false
Length=1349

 Score = 144 bits (364),  Expect = 1e-34, Method: Compositional matrix adjust.
 Identities = 126/442 (29%), Positives = 212/442 (48%), Gaps = 33/442 (7%)

Query  21   IAKITSDYQATQKITYRISGVGID--QPPFGIFVVDKNTGDINITAIVDREE---TPSFL  75
            +     D    Q I Y ++G GID   P    F +++ TG+I +   +DR++    P +
Sbjct  15   VTATDGDKDRPQNIVYFLTGQGIDPDNPANSKFDINRTTGEIFVLKPLDRDQPNGRPQWR  74

Query  76   ITCRALNAQGLDVEKPLILTVKILDINDNPPVFSQQIFMGEIEENSASNSLVMILNATDA  135
             T  A +  G  +     + V + DINDN P+F Q ++ G + EN  +  +VM + A D
Sbjct  75   FTVFAQDEGGEGLVGYADVQVNLKDINDNAPIFPQGVYFGNVTENGTAGMVVMTMTAVDY  134

Query  136  DEPNH-LNSKIAFKI---VSQEPAGTPMFLLSRNTGEVRTLTNSLDREQASSYRLVVSGA  191
            D+PN   N+K+ + I   V +E  G+P+F +  +TG ++T    LDRE+   Y + V
Sbjct  135  DDPNEGTNAKLIYSIEKNVIEEETGSPIFEIEADTGVIKTAVCCLDRERTPDYSIQVVAM  194

Query  192  DKDGEGLSTQCECNIKVKDVNDNFPMFRDSQYSARIEENILSS--ELLRFQVTDLDEEYT  249
              DG GL       +I+VKD+ND  P F   ++   ++E   ++  E+    VT  DE+ T
Sbjct  195  --DGGGLKGTGTASIRVKDINDMPPQFTKDEWFTEVDETDGTNLPEMPILTVTVHDEDET  252

Query  250  DNWLAVYFFTSGNEGNWFEIQTDPRTNEGI--LKVVKALDYE---QLQSVKLSIAVKNKA  304
            + +        SG   + F   T  R N+G   LK+V+ LDYE   Q    +  I V +K
Sbjct  253  NKFQYKVIDNSGYGADKF---TMVRNNDGTGSLKIVQPLDYEDPLQSNGFRFRIQVNDKG  309

Query  305  EFHQSVISRYRVQSTPVTIQVINVREGIAFRPASKTFTVQKGISSKKLVDYILGTYQAID  364
            E + +    +Y V  + V +++ ++ +    +P  +   ++  +      V   L T++A D
Sbjct  310  EDNDN--DKYHVAYSWVVVKLRDINDN---KPQFERPNIEVSVYENADVGKTLETFKATD  364

Query  365  EDTNKAASNVKYVMGR-NDGGYLMIDSKTAEIKFVKNMNRDSTFIVNKTITAEVLAIDE-  422
             D     S V Y + R +D        ++   +   +N++R+    V       ++LAID+
Sbjct  365  PDQG-GKSKVSYAIDRSSDRQRQFSINQEGTVTIQRNLDRE----VTPRHQVKILAIDDG  419

Query  423  YTGKTSTGTVYYVRVPDFNDNCP  444
               KT+T T+ V V D NDN P
Sbjct  420  IPPKTATATLTVIVQDINDNPP  441

 Score = 53.1 bits (126),  Expect = 2e-05, Method: Compositional matrix adjust.
 Identities = 70/306 (23%), Positives = 126/306 (41%), Gaps = 30/306 (10%)

Query  20   PIAKIT-SDYQATQKITYRI---SGVGIDQPPFGIFVVDKNTGDINITAIVDRE---ETP  72
            PI  +T  D   T K  Y++   SG G D+  F +   +  TG + I   +D E   ++
Sbjct  240  PILTVTVHDEDETNKFQYKVIDNSGYGADK--FTMVRNNDGTGSLKIVQPLDYEDPLQSN  297

Query  73   SFLITCRALNAQGLDVEKPLI------LTVKILDINDNPPVFSQQIFMGEIEENSASNSL  126
             F    + +N +G D +           + VK+ DINDN P F +       + EN+
Sbjct  298  GFRFRIQ-VNDKGEDNDNDKYHVAYSWVVVKLRDINDNKPQFERPNIEVSVYENADVGKT  356

Query  127  VMILNATDADEPNHLNSKIAFKIVSQEPAGTPMFLLSRNTGEVRTLTNSLDREQASSYRL  186
            +    ATD D+      SK+++ I   + +        S N     T+  +LDRE    +++
Sbjct  357  LETFKATDPDQGG--KSKVSYAI---DRSSDRQRQFSINQEGTVTIQRNLDREVTPRHQV  411

Query  187  VVSGADKDGEGLSTQCECNIKVKDVNDNFPMFRDSQYSARIEENILSSELLRFQVTDLDE  246
             +   D      +       + V+D+NDN P F     Y   + E++   +++     TD D+
Sbjct  412  KILAIDDGIPPKTATATLTVIVQDINDNPPKFL-KDYRPVLPEHVPPRKVVEILATDDDD  470

Query  247  EYTDNWLAVYFFTSGNEGNW----FEIQTDPRTNEG----ILKVVKALDYEQLQSVKLSI  298
                N     F       +     F+++ D +   G    I+  +++ D EQ +    + I
Sbjct  471  RSKSNGPPFQFRLDPGADDIIRASFKVEQDQKGANGDGMAIVSSLRSFDREQQKEYLIPI  530

Query  299  AVKNKA  304
             +K+
Sbjct  531  VIKDHG  536
```

```
 Score = 43.9 bits (102),  Expect = 0.011, Method: Compositional matrix adjust.
 Identities = 48/199 (24%), Positives = 73/199 (37%), Gaps = 19/199 (10%)

Query  27   DYQATQKITYRISGVGIDQPPFGIFVVDKNTGDINITAIVDREETPSFLITCRALNAQGL  86
            D     K++Y I      Q  F I       G + I   +DRE TP   +   A++
Sbjct  366  DQGGKSKVSYAIDRSSDRQRQFSI----NQEGTVTIQRNLDREVTPRHQVKILAIDDGIP  421

Query  87   DVEKPLILTVKILDINDNPPVFSQQIFMGEIEENSASNSLVMILNATDADEPNHLNSKIA  146
                   LTV + DINDNPP F +  +   + E+     +V IL   D D
Sbjct  422  PKTATATLTVIVQDINDNPPKFLKD-YRPVLPEHVPPRKVVEILATDDDDRSKSNGPPFQ  480

Query  147  FKIVSQEPAGTPMFLLSRNTGEVRTLTN-----------SLDREQASSYRLVVSGADKDG  195
            F++   +P    +   S    + +   N           S DREQ   Y + +   D
Sbjct  481  FRL---DPGADDIIRASFKVEQDQKGANGDGMAIVSSLRSFDREQQKEYLIPIVIKDHGN  537

Query  196  EGLSTQCECNIKVKDVNDN  214
              ++      + + DVNDN
Sbjct  538  PAMTGTSTLTVVIGDVNDN  556

>AARA017833-PA | transcript=AARA017833-RA | gene=AARA017833 |
organism=Anopheles_arabiensis_Dongola | gene_product=unspecified
product | transcript_product=unspecified product |
location=KB705006:7400057-7608391(-)
| protein_length=3101 | sequence_SO=supercontig
| SO=protein_coding_gene | is_pseudo=false
Length=3101

 Score = 145 bits (365),  Expect = 1e-34, Method: Compositional matrix adjust.
 Identities = 125/437 (29%), Positives = 211/437 (48%), Gaps = 33/437 (8%)

Query  26    SDYQATQKITYRISGVGID--QPPFGIFVVDKNTGDINITAIVDREET---PSFLITCRA  80
              D    Q I Y ++G GID   P    F +++ TG+I +   +DR++    P +  T  A
Sbjct  1550  GDKDRPQNIVYFLTGQGIDPDNPANSKFDINRTTGEIFVLKPLDRDQPNGRPQWRFTVFA  1609

Query  81    LNAQGLDVEKPLILTVKILDINDNPPVFSQQIFMGEIEENSASNSLVMILNATDADEPNH  140
              +  G  +     + V + DINDN P+F Q ++ G + EN  +  +VM + A D D+PN
Sbjct  1610  QDEGGEGLVGYADVQVNLKDINDNAPIFPQGVYFGNVTENGTAGMVVMTMTAVDYDDPNE  1669

Query  141   -LNSKIAFKI---VSQEPAGTPMFLLSRNTGEVRTLTNSLDREQASSYRLVVSGADKDGE  196
               N+K+ + I   V +E  G+P+F +  +TG ++T    LDRE+   Y + V     DG
Sbjct  1670  GTNAKLIYSIEKNVIEEETGSPIFEIEADTGVIKTAVCCLDRERTPDYSIQVVAM--DGG  1727

Query  197   GLSTQCECNIKVKDVNDNFPMFRDSQYSARIEENILSS--ELLRFQVTDLDEEYTDNWLA  254
             GL      +I+VKD+ND  P F   ++   ++E   ++  E+    VT  DE+ T+ +
Sbjct  1728  GLKGTGTASIRVKDINDMPPQFTKDEWFTEVDETDGTNLPEMPILTVTVHDEDETNKFQY  1787

Query  255   VYFFTSGNEGNWFEIQTDPRTNEGI--LKVVKALDYE---QLQSVKLSIAVKNKAEFHQS  309
                  SG   + F +    R N+G   LK+V+ LDYE   Q    +  I V +K E + +
Sbjct  1788  KVIDNSGYGADKFTMV---RNNDGTGSLKIVQPLDYEDPLQSNGFRFRIQVNDKGEDNDN  1844

Query  310   VISRYRVQSTPVTIQVINVREGIAFRPASKTFTVQKGISSKKLVDYILGTYQAIDEDTNK  369
                +Y V  + V +++ ++ +    +P  +   ++  +     V   L T++A D D
Sbjct  1845  --DKYHVAYSWVVVKLRDINDN---KPQFERPNIEVSVYENADVGKTLETFKATDPDQG-  1898

Query  370   AASNVKYVMGR-NDGGYLMIDSKTAEIKFVKNMNRDSTFIVNKTITAEVLAIDE-YTGKT  427
               S V Y + R +D       ++   +   +N++R+    V      ++LAID+    KT
Sbjct  1899  GKSKVSYAIDRSSDRQRQFSINQEGTVTIQRNLDRE----VTPRHQVKILAIDDGIPPKT  1954

Query  428   STGTVYVRVPDFNDNCP  444
             +T T+ V V D NDN P
Sbjct  1955  ATATLTVIVQDINDNPP  1971

 Score = 85.9 bits (211),  Expect = 1e-15, Method: Compositional matrix adjust.
 Identities = 108/420 (26%), Positives = 177/420 (42%), Gaps = 52/420 (12%)

Query  50    IFVVDKNTGDINITAIVDREETPSFLITCRAL----------NAQGLDVEKPLILTVKIL  99
              F ++  +G+I    + DRE+  ++ +   A           N Q   V K   + + I
Sbjct  1355  FFEINLQSGEIFTKVVFDREKQGAYALEVEARDGAPSARPNSNQQPNSVTK--FIRIGIA  1412
```

```
Query  100    DINDNPPVFSQQIFMGEIEENSASNSLVMILNATDADEPNHLNSKIAFKIVSQEPAGTPM  159
              D NDNPP F + ++  E++EN      V+ + A D DE    +S+I ++I S    G
Sbjct  1413  DKNDNPPYFDKALYEAEVDENEDIQHTVLTVTAKDHDE----SSRIRYEITSGNIGGA--  1466

Query  160    FLLSRNTGEVRTLTNSLDREQASSYRLVVSGADKDGEGLSTQCECNIKVKDVNDNFPMFR  219
              F +   TG +  +  +LD E    Y L ++ +D   E  +T      I VKDVNDN P+F
Sbjct  1467  FAVKNMTGAI-YVAGALDYETRKRYELRLAASDNLKENYTTVV---IHVKDVNDNPPVFE  1522

Query  220    DSQYSARIEENI---LSSELLRFQVTDLDEEYTDNWLAVYFFTSG--NEGNWFEIQTDPR  274
                Y  +I E     L   +L+   TD D++   N   VYF T    +  N    + D
Sbjct  1523  RPTYRTQITEEDDRNLPKRVLQVTATDGDKDRPQN--IVYFLTGQGIDPDNPANSKFDIN  1580

Query  275    TNEGILKVVKALDYEQLQS-VKLSIAVKNKAEFHQSVISRYRVQSTPVTIQVINVREGIA  333
                 G + V+K LD +Q     +    V  + E  + ++    VQ     + + ++ +
Sbjct  1581  RTTGEIFVLKPLDRDQPNGRPQWRFTVFAQDEGGEGLVGYADVQ-----VNLKDINDNAP  1635

Query  334    FRPASKTFTVQKGISSKKLVDYILGTYQAIDEDTNKAASNVK--YVMGRN------DGGY  385
                P    F     ++        ++ T  A+D D     +N K  Y + +N
Sbjct  1636  IFPQGVYF---GNVTENGTAGMVVMTMTAVDYDDPNEGTNAKLIYSIEKNVIEEETGSPI  1692

Query  386    LMIDSKTAEIK-FVKNMNRDSTFIVNKTITAEVLAIDEYTGKTSTGTVYVRVPDFNDNCP  444
                I++ T  IK  V  ++R+ T        + +V+A+D   G   TGT  +RV D ND  P
Sbjct  1693  FEIEADTGVIKTAVCCLDRERT----PDYSIQVVAMDG-GGLKGTGTASIRVKDINDMPP  1747

 Score = 79.0 bits (193),  Expect = 2e-13, Method: Compositional matrix adjust.
 Identities = 106/424 (25%), Positives = 180/424 (42%), Gaps = 51/424 (12%)

Query  53     VDKNTGDINITAIVDREETPSFLITCRALNAQGLDVEKPLILTVKILDINDNPPVFSQ-Q  111
              +D++  ++N+TA+ D          +C  +N           ++ V I D+NDN PVF
Sbjct  928   LDRDKYELNVTAMDDG--------SC-CVNGDATIHTSTAVVVVFITDVNDNKPVFKDCG  978

Query  112    IFMGEIEENSASNSLVMILNATDADEPNHLNSKIAFKIVSQEPAGTPMFLLSRNTGEVRT  171
               +  ++EE + + S V+ + ATD D+   +N ++ + IV Q        F +   TGEV T
Sbjct  979   TYYPKVEEGAPNGSPVIKVQATDEDK--GVNGQVKYSIVQQPNQKGTKFTVDEETGEVST  1036

Query  172    LTNSLDRE--QASSYRLVVSGADKDGEGLSTQCECNIKVKDVNDNFPMFRDSQYSARIEE  229
                   DRE         + V   D+    L   C    +++ DVNDN P+F   +Y   +++
Sbjct  1037  -NKVFDREGDDGKFVSVTVKATDQGEPSLEGVCSFTVEITDVNDNPPLFDRQKYVENVKQ  1095

Query  230    NI-LSSELLRFQVTDLDEEYTDNWLAVYFFT---SGNEGNWFEIQTDPRTNEGILKVVKA  285
              +  + + +LR  V+  DE+  +N   VY  +   S N+  +FEIQ  P +   +LK  K
Sbjct  1096  DASIGTNILR--VSASDEDADNNGAIVYTLSAPHSPNDLEYFEIQ--PESGWIVLK--KP  1149

Query  286    LDYEQLQSVKLSIAVKNK------AEFHQSVISRYRVQSTPVTIQV----INVREGIAFR  335
              LD E+    +L +   ++      A+    +    R    P+  +      I++RE +
Sbjct  1150  LDRERY---RLKVYASDRGDPPSTADVDVELDVVDRNNKPPIWDRTIYGPIHIRENVTVG  1206

Query  336    PASKTFTVQKGISSKKLVDYILGTYQAIDEDTNKAASNVKYVMGRNDGGYLMIDSKTAEI  395
                  +     GI     V Y L          TNK   +  Y+  R D G         A+I
Sbjct  1207  TVVTSVKASSGIEGNPTVFYRL--MPGSTAQTNKL--HTFYLQQRPDNG-----DTWADI  1257

Query  396    KFVKNMNRDSTFIVNKTITAEVLAIDEYTGKTSTGTVYVRVPDFNDNCPTAV-LEKDAVC  454
              K    ++ ++     N T+  E     +      S  TVY+++ D ND  P      E++ V
Sbjct  1258  KVNHPLDFETIKEYNLTVRVENNGAQQL---ASEATVYIQLEDVNDEIPLFTEREQETVL  1314

Query  455    SSSP  458
                P
Sbjct  1315  EGEP  1318

 Score = 65.5 bits (158),  Expect = 3e-09, Method: Compositional matrix adjust.
 Identities = 85/359 (24%), Positives = 153/359 (43%), Gaps = 32/359 (9%)

Query  51     FVVDKNTGDINITAIVDRE-ETPSFL-ITCRALNAQGLDVEKPLILTVKILDINDNPPVF  108
              F VD+ TG+++    + DRE +   F+ +T +A +      +E      TV+I D+NDNPP+F
Sbjct  1025  FTVDEETGEVSTNKVFDREGDDGKFVSVTVKATDQGEPSLEGVCSFTVEITDVNDNPPLF  1084

Query  109    SQQIFMGEIEENSASNSLVMILNATDADEPNHLNSKIAFKIVS-QEPAGTPMFLLSRNTG  167
               +Q ++  ++++++  + ++ ++A+D D  N  N  I + + +   P     F +   +G
```

```
Sbjct  1085  DRQKYVENVKQDASIGTNILRVSASDEDADN--NGAIVYTLSAPHSPNDLEYFEIQPESG  1142

Query  168   EVRTLTNSLDREQASSYRLVVSGADKDGEGLSTQCECNIKVKDVNDNFPMFRDSQYSARI  227
              +  L   LDRE+   YRL V  +D+ G+  ST            +N P   D      I
Sbjct  1143  WI-VLKKPLDRER---YRLKVYASDR-GDPPSTADVDVELDVVDRNNKPPIWDRTIYGPI  1197

Query  228   EENILSSELLRFQVTDLDEEYTDNWLAVYFFTSG-----NEGNWFEIQTDPRTNE--GIL  280
                    +                N    Y    G     N+ + F +Q  P   +    +
Sbjct  1198  HIRENVTVGTVVTSVKASSGIEGNPTVFYRLMPGSTAQTNKLHTFYLQQRPDNGDTWADI  1257

Query  281   KVVKALDYEQLQSVKLSIAVKNKAEFHQSVISRYRVQSTPVTIQVINVREGIAFRPASKT  340
             KV   LD+E ++   L++ V+N         ++       V IQ+ +V + I      +
Sbjct  1258  KVNHPLDFETIKEYNLTVRVENNG-------AQQLASEATVYIQLEDVNDEIPLFTEREQ  1310

Query  341   FTVQKGISSKKLVDYILGTYQAIDEDTNKAASNVKYVM---GRNDG-GYLMIDSKTAEI  395
              TV +G      V  +     AID+D     + V Y +    RN+G  +  I+ ++ EI
Sbjct  1311  ETVLEGEPIGTKVTQV----NAIDKDGTFPNNQVYYYIVDSPRNEGKDFFEINLQSGEI  1365

 Score = 55.8 bits (133),  Expect = 3e-06, Method: Compositional matrix adjust.
 Identities = 49/158 (31%), Positives = 78/158 (49%), Gaps = 24/158 (15%)

Query  102   NDNPPVFSQQIFMGEIEENSASNSLVMILNATDADEPNHLNSKIAFKIVSQEPAGTP--M  159
             ND  P FSQQ++   ++EN+  N+LV  + A+D D  N     + F  V     GT
Sbjct  858   NDEEPKFSQQVYTPNVDENAGPNTLVTTVVASDKDGDN-----VRFGFVG---GGTSSGQ  909

Query  160   FLLSRNTGEVRTLTN--SLDREQASSYRLVVSGADK-------DGEGLSTQCECNIKVKD  210
             F++   TG +R  +   SLDR++   Y L V+  D        D    ++     + + D
Sbjct  910   FVIEEITGVIRLHSKAISLDRDK---YELNVTAMDDGSCCVNGDATIHTSTAVVVVFITD  966

Query  211   VNDNFPMFRD-SQYSARIEENILS-SELLRFQVTDLDE  246
             VNDN P+F+D   Y  ++EE   + S +++ Q TD D+
Sbjct  967   VNDNKPVFKDCGTYYPKVEEGAPNGSPVIKVQATDEDK  1004

 Score = 53.9 bits (128),  Expect = 1e-05, Method: Compositional matrix adjust.
 Identities = 70/306 (23%), Positives = 126/306 (41%), Gaps = 30/306 (10%)

Query  20    PIAKIT-SDYQATQKITYRI---SGVGIDQPPFGIFVVDKNTGDINITAIVDRE---ETP  72
             PI  +T  D   T K  Y++   SG G D+  F +   +  TG + I   +D E   ++
Sbjct  1770  PILTVTVHDEDETNKFQYKVIDNSGYGADK--FTMVRNNDGTGSLKIVQPLDYEDPLQSN  1827

Query  73    SFLITCRALNAQGLDVEKPLI------LTVKILDINDNPPVFSQQIFMGEIEENSASNSL  126
              F    + +N +G D +          + VK+ DINDN P F +      + EN+
Sbjct  1828  GFRFRIQ-VNDKGEDNDNDKYHVAYSWVVVKLRDINDNKPQFERPNIEVSVYENADVGKT  1886

Query  127   VMILNATDADEPNHLNSKIAFKIVSQEPAGTPMFLLSRNTGEVRTLTNSLDREQASSYRL  186
             +    ATD D+     SK+++ I   + +       S N     T+  +LDRE    +++
Sbjct  1887  LETFKATDPDQGG--KSKVSYAI---DRSSDRQRQFSINQEGTVTIQRNLDREVTPRHQV  1941

Query  187   VVSGADKDGEGLSTQCECNIKVKDVNDNFPMFRDSQYSARIEENILSSELLRFQVTDLDE  246
              +   D      +      + V+D+NDN P F    Y   + E++   +++    TD D+
Sbjct  1942  KILAIDDGIPPKTATATLTVIVQDINDNPPKFL-KDYRPVLPEHVPPRKVVEILATDDDD  2000

Query  247   EYTDNWLAVYFFTSGNEGNW----FEIQTDPRTNEG----ILKVVKALDYEQLQSVKLSI  298
                 N     F       +     F+++ D +   G    I+  +++ D EQ +   + I
Sbjct  2001  RSKSNGPPFQFRLDPGADDIIRASFKVEQDQKGANGDGMAIVSSLRSFDREQQKEYLIPI  2060

Query  299   AVKNKA  304
              +K+
Sbjct  2061  VIKDHG  2066

 Score = 52.4 bits (124),  Expect = 4e-05, Method: Compositional matrix adjust.
 Identities = 97/398 (24%), Positives = 163/398 (41%), Gaps = 54/398 (14%)

Query  106   PVFSQQIFMGEIEENSASNSLVMILNATDADEPNHLNSKIAFKIVSQ-EPAGTPMFLLSR  164
             P F    +  EI EN   +S ++ + A      +  + +I + + +Q + AGT  F +
Sbjct  655   PQFYMPSYEAEIPENQKKDSDIISVKAK-----SFADREIRYTLKAQGQGAGT--FNIGP  707

Query  165   NTGEVRTLTNSLDRE---QASSYRLVVSGADKDGEGLSTQCECNIKVKDVNDNFPMFRDS  221
```

```
              +G V+ L   LD E   Q   Y LVV+ A +D  G ST  E  I+V DVNDN P F
Sbjct  708   TSGIVK-LAKELDFEDLRQPHVYSLVVT-ATEDSGGFSTSVELTIRVTDVNDNAPKFELP  765

Query  222   QYSAR-IEENI-LSSELLRFQVTDLDEEYTDNWLAVYFFTSGNEGNWFEIQTDPRTNEGI  279
              Y A  ++E+I L + +L+ +  D D         + +  S +    F + ++     GI
Sbjct  766   DYQAHNVDEDIPLGTSILKVKAMDSDSGAN---AEIEYLVSDDH---FAVDSN-----GI  814

Query  280   LKVVKALDYEQLQS-VKLSIAVKNKAEFHQSVISRYRVQSTPVTIQVINVREGIAFRPAS  338
             +   K LD +   +  +  +  K+K E  ++  +  RV +           E   F
Sbjct  815   IVNNKQLDADNNNAYYEFVVTAKDKGEPAKTGTATVRVYTKNKN------DEEPKFSQQV  868

Query  339   KTFTVQKGISSKKLVDYILGTYQAIDEDTNKAASNVK--YVMGRNDGGYLMIDSKTAEIK  396
              T  V +      LV  ++         ++K   NV+  +V G    G  +I+  T  I+
Sbjct  869   YTPNVDENAGPNTLVTTVVA--------SDKDGDNVRFGFVGGGTSSGQFVIEEITGVIR  920

Query  397   FVK---NMNRDSTFI-VNKTITAEVLAIDEYTGKTSTGTVYVRVPDFNDN------CPTA  446
                   +++RD   + V            + T  TST  V V + D NDN      C T
Sbjct  921   LHSKAISLDRDKYELNVTAMDDGSCCVNGDATIHTSTAVVVVFITDVNDNKPVFKDCGTY  980

Query  447   VLE-KDAVCSSSPSVVVSARTLNNRYTGPYTFALEDQP  483
               + ++   + SP + V A   +    G   +++  QP
Sbjct  981   YPKVEEGAPNGSPVIKVQATDEDKGVNGQVKYSIVQQP  1018

 Score = 44.7 bits (104),  Expect = 0.008, Method: Compositional matrix adjust.
 Identities = 48/199 (24%), Positives = 73/199 (37%), Gaps = 19/199 (10%)

Query  27    DYQATQKITYRISGVGIDQPPFGIFVVDKNTGDINITAIVDREETPSFLITCRALNAQGL  86
             D     K++Y I      Q  F I       G + I   +DRE TP   +   A++
Sbjct  1896  DQGGKSKVSYAIDRSSDRQRQFSI----NQEGTVTIQRNLDREVTPRHQVKILAIDDGIP  1951

Query  87    DVEKPLILTVKILDINDNPPVFSQQIFMGEIEENSASNSLVMILNATDADEPNHLNSKIA  146
                    LTV + DINDNPP F +  +   + E+     +V IL   D D
Sbjct  1952  PKTATATLTVIVQDINDNPPKFLKD-YRPVLPEHVPPRKVVEILATDDDDRSKSNGPPFQ  2010

Query  147   FKIVSQEPAGTPMFLLSRNTGEVRTLTN-----------SLDREQASSYRLVVSGADKDG  195
             F++   +P    +   S    + +   N           S DREQ   Y + +   D
Sbjct  2011  FRL---DPGADDIIRASFKVEQDQKGANGDGMAIVSSLRSFDREQQKEYLIPIVIKDHGN  2067

Query  196   EGLSTQCECNIKVKDVNDN  214
               ++      + + DVNDN
Sbjct  2068  PAMTGTSTLTVVIGDVNDN  2086

 Score = 40.0 bits (92),  Expect = 0.21, Method: Compositional matrix adjust.
 Identities = 89/389 (23%), Positives = 149/389 (38%), Gaps = 62/389 (16%)

Query  75   LITCRALNAQGLDVEKPLILTVKILDINDNPPVFSQQIF--MGEIEENSASNSLVMILNA  132
            +IT +  NA GL       + + + D+ND PP F  +       ++ N+  N+ V  L A
Sbjct  517  IITNQGHNA-GLKYTDNQRVIIHVKDVNDEPPYFINRPLPMQAVVQLNAPPNTPVFTLQA  575

Query  133  TDADEPNHLNSKIAFKIVSQEPAGTPMFLLSRNTGEVRTLTNSLDREQASSYRLVVSGAD  192
             D D     +  I + IV     G   F +   +G VRT    L  +    Y L V   D
Sbjct  576  RDPD----TDHNIHYFIVRDRTGG--RFEVDERSGVVRTRGTDL-FQLDMEYVLYVKAED  628

Query  193  KDGE----GLSTQCECNIKVKDVNDNFPMFRDSQYSARIEEN-ILSSELLRFQVTDLDEE  247
            ++G+       +  E  + +       P F    Y A I EN    S+++  +     +
Sbjct  629  QNGKVDDRRFQSTPEERLSIVG-GKRAPQFYMPSYEAEIPENQKKDSDIISVKA----KS  683

Query  248  YTDNWLAVYFFTSGNEGNWFEIQTDPRTNEGILKVVKALDYEQLQSVKLS----IAVKNK  303
            + D  +       G     F I        GI+K+ K LD+E L+   +      A ++
Sbjct  684  FADREIRYTLKAQGQGAGTFNIGP----TSGIVKLAKELDFEDLRQPHVYSLVVTATEDS  739

Query  304  AEFHQSVISRYRVQSTPVTIQVINVREGIAFRPASKTFTVQKGISSKKLVDYILGT----  359
              F  SV          +TI+V +V +       +  F +    +     D  LGT
Sbjct  740  GGFSTSV---------ELTIRVTDVNDN------APKFELPDYQAHNVDEDIPLGTSILK  784

Query  360  YQAIDEDTNKAASNVKYVMGRN----DGGYLMIDSKTAEIKFVKNMNRDSTFIVNKTITA  415
             +A+D D+  A + ++Y++  +    D   +++++K  +     N N    F+V
Sbjct  785  VKAMDSDSG-ANAEIEYLVSDDHFAVDSNGIIVNNKQLD---ADNNNAYYEFVVTAKDKG  840
```

```
Query  416  EVLAIDEYTGKTSTGTVYVRVPDFNDNCP  444
            E         KT T TV V   + ND  P
Sbjct  841  E-------PAKTGTATVRVYTKNKNDEEP  862

 Score = 39.7 bits (91),  Expect = 0.26, Method: Compositional matrix adjust.
 Identities = 54/242 (22%), Positives = 102/242 (42%), Gaps = 22/242 (9%)

Query  4    KFAKPCREGE--DNSKRNP-IAKITSDYQATQKITYRISGVGIDQPPFGIFVVDKNTGDI  60
            +F  P  E E  +N K++  I  + +   A ++I Y +   G      G F +   +G +
Sbjct  656  QFYMPSYEAEIPENQKKDSDIISVKAKSFADREIRYTLKAQGQGA---GTFNIGPTSGIV  712

Query  61   NITAIVDREE--TPSF--LITCRALNAQGLDVEKPLILTVKILDINDNPPVFSQQIFMGE  116
             +   +D E+   P    L+     ++ G      + LT+++ D+NDN P F    +
Sbjct  713  KLAKELDFEDLRQPHVYSLVVTATEDSGGFSTS--VELTIRVTDVNDNAPKFELPDYQAH  770

Query  117  -IEENSASNSLVMILNATDADEPNHLNSKIAFKIVSQEPAGTPMFLLSRNTGEVRTLTNS  175
             ++E+    + ++ + A D+D  +  N++I + +VS +      F +  N   V
Sbjct  771  NVDEDIPLGTSILKVKAMDSD--SGANAEIEY-LVSDD-----HFAVDSNGIIVNNKQLD  822

Query  176  LDREQASSYRLVVSGADKDGEGLSTQCECNIKVKDVNDNFPMFRDSQYSARIEENILSSE  235
             D   A  Y  VV+  DK      +      +  K+ ND  P F    Y+  ++EN   +
Sbjct  823  ADNNNA-YYEFVVTAKDKGEPAKTGTATVRVYTKNKNDEEPKFSQQVYTPNVDENAGPNT  881

Query  236  LL  237
            L+
Sbjct  882  LV  883

>AARA21_009090.P17039 | transcript=AARA21_009090.R17039 | gene=AARA21_009090
| organism=Anopheles_arabiensis_DONGOLA_2021
| gene_product=neural-cadherin, transcript variant X5 |
transcript_product=neural-cadherin,
transcript variant X10 | location=CM029353:40466514-40666848(+)
| protein_length=3101 |
sequence_SO=chromosome | SO=protein_coding_gene | is_pseudo=false
Length=3101

 Score = 145 bits (365),  Expect = 1e-34, Method: Compositional matrix adjust.
 Identities = 125/437 (29%), Positives = 211/437 (48%), Gaps = 33/437 (8%)

Query  26    SDYQATQKITYRISGVGID--QPPFGIFVVDKNTGDINITAIVDREET---PSFLITCRA  80
              D    Q I Y ++G GID   P    F +++ TG+I +   +DR++    P +  T  A
Sbjct  1550  GDKDRPQNIVYFLTGQGIDPDNPANSKFDINRTTGEIFVLKPLDRDQPNGRPQWRFTVFA  1609

Query  81    LNAQGLDVEKPLILTVKILDINDNPPVFSQQIFMGEIEENSASNSLVMILNATDADEPNH  140
              +  G  +      + V + DINDN P+F Q ++ G + EN  +  +VM + A D D+PN
Sbjct  1610  QDEGGEGLVGYADVQVNLKDINDNAPIFPQGVYFGNVTENGTAGMVVMTMTAVDYDDPNE  1669

Query  141   -LNSKIAFKI---VSQEPAGTPMFLLSRNTGEVRTLTNSLDREQASSYRLVVSGADKDGE  196
               N+K+ + I   V +E  G+P+F +  +TG ++T    LDRE+   Y + V     DG
Sbjct  1670  GTNAKLIYSIEKNVIEEETGSPIFEIEADTGVIKTAVCCLDRERTPDYSIQVVAM--DGG  1727

Query  197   GLSTQCECNIKVKDVNDNFPMFRDSQYSARIEENILSS--ELLRFQVTDLDEEYTDNWLA  254
             GL      +I+VKD+ND  P F  ++   ++E   ++  E+    VT  DE+ T+ +
Sbjct  1728  GLKGTGTASIRVKDINDMPPQFTKDEWFTEVDETDGTNLPEMPILTVTVHDEDETNKFQY  1787

Query  255   VYFFTSGNEGNWFEIQTDPRTNEGI--LKVVKALDYE---QLQSVKLSIAVKNKAEFHQS  309
                  SG   + F +    R N+G   LK+V+ LDYE   Q     +  I V +K E + +
Sbjct  1788  KVIDNSGYGADKFTMV---RNNDGTGSLKIVQPLDYEDPLQSNGFRFRIQVNDKGEDNDN  1844

Query  310   VISRYRVQSTPVTIQVINVREGIAFRPASKTFTVQKGISSKKLVDYILGTYQAIDEDTNK  369
                +Y V  + V +++ ++ +    +P  +   ++  +     V   L T++A D D
Sbjct  1845  --DKYHVAYSWVVVKLRDINDN---KPQFERPNIEVSVYENADVGKTLETFKATDPDQG-  1898

Query  370   AASNVKYVMGR-NDGGYLMIDSKTAEIKFVKNMNRDSTFIVNKTITAEVLAIDE-YTGKT  427
               S V Y + R +D       ++   +   +N++R+    V      ++LAID+    KT
Sbjct  1899  GKSKVSYAIDRSSDRQRQFSINQEGTVTIQRNLDRE----VTPRHQVKILAIDDGIPPKT  1954

Query  428   STGTVYVRVPDFNDNCP  444
             +T T+ V V D NDN P
```

```
Sbjct  1955  ATATLTVIVQDINDNPP  1971

 Score = 85.9 bits (211),  Expect = 1e-15, Method: Compositional matrix adjust.
 Identities = 108/420 (26%), Positives = 177/420 (42%), Gaps = 52/420 (12%)

Query  50    IFVVDKNTGDINITAIVDREETPSFLITCRAL----------NAQGLDVEKPLILTVKIL  99
              F ++  +G+I    + DRE+  ++ +   A           N Q   V K   + + I
Sbjct  1355  FFEINLQSGEIFTKVVFDREKQGAYALEVEARDGAPSARPNSNQQPNSVTK--FIRIGIA  1412

Query  100   DINDNPPVFSQQIFMGEIEENSASNSLVMILNATDADEPNHLNSKIAFKIVSQEPAGTPM  159
             D NDNPP F + ++  E++EN      V+ + A D DE    +S+I ++I S    G
Sbjct  1413  DKNDNPPYFDKALYEAEVDENEDIQHTVLTVTAKDHDE----SSRIRYEITSGNIGGA--  1466

Query  160   FLLSRNTGEVRTLTNSLDREQASSYRLVVSGADKDGEGLSTQCECNIKVKDVNDNFPMFR  219
             F +   TG +  +  +LD E    Y L ++ +D   E  +T      I VKDVNDN P+F
Sbjct  1467  FAVKNMTGAI-YVAGALDYETRKRYELRLAASDNLKENYTTVV---IHVKDVNDNPPVFE  1522

Query  220   DSQYSARIEENI---LSSELLRFQVTDLDEEYTDNWLAVYFFTSG--NEGNWFEIQTDPR  274
               Y  +I E     L   +L+   TD D++   N   VYF T    +  N    + D
Sbjct  1523  RPTYRTQITEEDDRNLPKRVLQVTATDGDKDRPQN--IVYFLTGQGIDPDNPANSKFDIN  1580

Query  275   TNEGILKVVKALDYEQLQS-VKLSIAVKNKAEFHQSVISRYRVQSTPVTIQVINVREGIA  333
                G + V+K LD +Q      +    V  + E  + ++    VQ      + + ++ +
Sbjct  1581  RTTGEIFVLKPLDRDQPNGRPQWRFTVFAQDEGGEGLVGYADVQ-----VNLKDINDNAP  1635

Query  334   FRPASKTFTVQKGISSKKLVDYILGTYQAIDEDTNKAASNVK--YVMGRN------DGGY  385
              P    F     ++        ++ T  A+D D     +N K  Y + +N
Sbjct  1636  IFPQGVYF---GNVTENGTAGMVVMTMTAVDYDDPNEGTNAKLIYSIEKNVIEEETGSPI  1692

Query  386   LMIDSKTAEIK-FVKNMNRDSTFIVNKTITAEVLAIDEYTGKTSTGTVYVRVPDFNDNCP  444
               I++ T  IK  V  ++R+ T        + +V+A+D   G   TGT  +RV D ND  P
Sbjct  1693  FEIEADTGVIKTAVCCLDRERT----PDYSIQVVAMDG-GGLKGTGTASIRVKDINDMPP  1747

 Score = 79.0 bits (193),  Expect = 2e-13, Method: Compositional matrix adjust.
 Identities = 106/424 (25%), Positives = 180/424 (42%), Gaps = 51/424 (12%)

Query  53    VDKNTGDINITAIVDREETPSFLITCRALNAQGLDVEKPLILTVKILDINDNPPVFSQ-Q  111
             +D++  ++N+TA+ D          +C  +N           ++ V I D+NDN PVF
Sbjct  928   LDRDKYELNVTAMDDG--------SC-CVNGDATIHTSTAVVVVFITDVNDNKPVFKDCG  978

Query  112   IFMGEIEENSASNSLVMILNATDADEPNHLNSKIAFKIVSQEPAGTPMFLLSRNTGEVRT  171
              +  ++EE + + S V+ + ATD D+    +N ++ + IV Q        F +   TGEV T
Sbjct  979   TYYPKVEEGAPNGSPVIKVQATDEDK--GVNGQVKYSIVQQPNQKGTKFTVDEETGEVST  1036

Query  172   LTNSLDRE--QASSYRLVVSGADKDGEGLSTQCECNIKVKDVNDNFPMFRDSQYSARIEE  229
                  DRE         + V   D+    L   C   +++ DVNDN P+F   +Y   +++
Sbjct  1037  -NKVFDREGDDGKFVSVTVKATDQGEPSLEGVCSFTVEITDVNDNPPLFDRQKYVENVKQ  1095

Query  230   NI-LSSELLRFQVTDLDEEYTDNWLAVYFFT---SGNEGNWFEIQTDPRTNEGILKVVKA  285
             +  + + +LR  V+  DE+  +N   VY  +   S N+  +FEIQ  P +   +LK  K
Sbjct  1096  DASIGTNILR--VSASDEDADNNGAIVYTLSAPHSPNDLEYFEIQ--PESGWIVLK--KP  1149

Query  286   LDYEQLQSVKLSIAVKNK------AEFHQSVISRYRVQSTPVTIQV----INVREGIAFR  335
             LD E+     +L +    ++       A+     +     R     P+  +      I++RE +
Sbjct  1150  LDRERY---RLKVYASDRGDPPSTADVDVELDVVDRNNKPPIWDRTIYGPIHIRENVTVG  1206

Query  336   PASKTFTVQKGISSKKLVDYILGTYQAIDEDTNKAASNVKYVMGRNDGGYLMIDSKTAEI  395
                +      GI      V Y L           TNK   +  Y+  R D G         A+I
Sbjct  1207  TVVTSVKASSGIEGNPTVFYRL--MPGSTAQTNKL--HTFYLQQRPDNG-----DTWADI  1257

Query  396   KFVKNMNRDSTFIVNKTITAEVLAIDEYTGKTSTGTVYVRVPDFNDNCPTAV-LEKDAVC  454
             K     ++ ++      N T+  E       +      S  TVY+++ D ND  P      E++ V
Sbjct  1258  KVNHPLDFETIKEYNLTVRVENNGAQQL---ASEATVYIQLEDVNDEIPLFTEREQETVL  1314

Query  455   SSSP  458
               P
Sbjct  1315  EGEP  1318
```

```
 Score = 65.5 bits (158),  Expect = 3e-09, Method: Compositional matrix adjust.
 Identities = 85/359 (24%), Positives = 153/359 (43%), Gaps = 32/359 (9%)

Query  51    FVVDKNTGDINITAIVDRE-ETPSFL-ITCRALNAQGLDVEKPLILTVKILDINDNPPVF  108
             F VD+ TG+++   + DRE +   F+ +T +A +     +E     TV+I D+NDNPP+F
Sbjct  1025  FTVDEETGEVSTNKVFDREGDDGKFVSVTVKATDQGEPSLEGVCSFTVEITDVNDNPPLF  1084

Query  109   SQQIFMGEIEENSASNSLVMILNATDADEPNHLNSKIAFKIVS-QEPAGTPMFLLSRNTG  167
              +Q ++  ++++++  + ++ ++A+D D  N  N  I + + +   P     F +   +G
Sbjct  1085  DRQKYVENVKQDASIGTNILRVSASDEDADN--NGAIVYTLSAPHSPNDLEYFEIQPESG  1142

Query  168   EVRTLTNSLDREQASSYRLVVSGADKDGEGLSTQCECNIKVKDVNDNFPMFRDSQYSARI  227
              +  L   LDRE+   YRL V  +D+ G+  ST            +N P   D      I
Sbjct  1143  WI-VLKKPLDRER---YRLKVYASDR-GDPPSTADVDVELDVVDRNNKPPIWDRTIYGPI  1197

Query  228   EENILSSELLRFQVTDLDEEYTDNWLAVYFFTSG-----NEGNWFEIQTDPRTNE--GIL  280
                   +                N    Y    G     N+ + F +Q  P   +    +
Sbjct  1198  HIRENVTVGTVVTSVKASSGIEGNPTVFYRLMPGSTAQTNKLHTFYLQQRPDNGDTWADI  1257

Query  281   KVVKALDYEQLQSVKLSIAVKNKAEFHQSVISRYRVQSTPVTIQVINVREGIAFRPASKT  340
             KV   LD+E ++   L++ V+N         ++       V IQ+ +V + I      +
Sbjct  1258  KVNHPLDFETIKEYNLTVRVENNG-------AQQLASEATVYIQLEDVNDEIPLFTEREQ  1310

Query  341   FTVQKGISSKKLVDYILGTYQAIDEDTNKAASNVKYVM---GRNDG-GYLMIDSKTAEI  395
              TV +G      V  +     AID+D     + V Y +    RN+G  +  I+ ++ EI
Sbjct  1311  ETVLEGEPIGTKVTQV----NAIDKDGTFPNNQVYYYIVDSPRNEGKDFFEINLQSGEI  1365

 Score = 55.8 bits (133),  Expect = 3e-06, Method: Compositional matrix adjust.
 Identities = 49/158 (31%), Positives = 78/158 (49%), Gaps = 24/158 (15%)

Query  102   NDNPPVFSQQIFMGEIEENSASNSLVMILNATDADEPNHLNSKIAFKIVSQEPAGTP--M  159
             ND  P FSQQ++   ++EN+  N+LV  + A+D D  N     + F  V     GT
Sbjct  858   NDEEPKFSQQVYTPNVDENAGPNTLVTTVVASDKDGDN-----VRFGFVG---GGTSSGQ  909

Query  160   FLLSRNTGEVRTLTN--SLDREQASSYRLVVSGADK-------DGEGLSTQCECNIKVKD  210
             F++   TG +R  +   SLDR++   Y L V+  D        D    ++     + + D
Sbjct  910   FVIEEITGVIRLHSKAISLDRDK---YELNVTAMDDGSCCVNGDATIHTSTAVVVVFITD  966

Query  211   VNDNFPMFRD-SQYSARIEENILS-SELLRFQVTDLDE  246
             VNDN P+F+D   Y  ++EE   + S +++ Q TD D+
Sbjct  967   VNDNKPVFKDCGTYYPKVEEGAPNGSPVIKVQATDEDK  1004

 Score = 53.9 bits (128),  Expect = 1e-05, Method: Compositional matrix adjust.
 Identities = 70/306 (23%), Positives = 126/306 (41%), Gaps = 30/306 (10%)

Query  20    PIAKIT-SDYQATQKITYRI---SGVGIDQPPFGIFVVDKNTGDINITAIVDRE---ETP  72
             PI  +T  D   T K  Y++   SG G D+  F +   +  TG + I   +D E   ++
Sbjct  1770  PILTVTVHDEDETNKFQYKVIDNSGYGADK--FTMVRNNDGTGSLKIVQPLDYEDPLQSN  1827

Query  73    SFLITCRALNAQGLDVEKPLI------LTVKILDINDNPPVFSQQIFMGEIEENSASNSL  126
              F    + +N +G D +          + VK+ DINDN P F +      + EN+
Sbjct  1828  GFRFRIQ-VNDKGEDNDNDKYHVAYSWVVVKLRDINDNKPQFERPNIEVSVYENADVGKT  1886

Query  127   VMILNATDADEPNHLNSKIAFKIVSQEPAGTPMFLLSRNTGEVRTLTNSLDREQASSYRL  186
             +    ATD D+     SK+++ I   + +       S N     T+  +LDRE    +++
Sbjct  1887  LETFKATDPDQGG--KSKVSYAI---DRSSDRQRQFSINQEGTVTIQRNLDREVTPRHQV  1941

Query  187   VVSGADKDGEGLSTQCECNIKVKDVNDNFPMFRDSQYSARIEENILSSELLRFQVTDLDE  246
              +   D      +      + V+D+NDN P F    Y   + E++   +++    TD D+
Sbjct  1942  KILAIDDGIPPKTATATLTVIVQDINDNPPKFL-KDYRPVLPEHVPPRKVVEILATDDDD  2000

Query  247   EYTDNWLAVYFFTSGNEGNW----FEIQTDPRTNEG----ILKVVKALDYEQLQSVKLSI  298
                 N     F       +     F+++ D +   G    I+  +++ D EQ +   + I
Sbjct  2001  RSKSNGPPFQFRLDPGADDIIRASFKVEQDQKGANGDGMAIVSSLRSFDREQQKEYLIPI  2060

Query  299   AVKNKA  304
              +K+
Sbjct  2061  VIKDHG  2066
```

```
 Score = 52.4 bits (124),  Expect = 4e-05, Method: Compositional matrix adjust.
 Identities = 97/398 (24%), Positives = 163/398 (41%), Gaps = 54/398 (14%)

Query  106   PVFSQQIFMGEIEENSASNSLVMILNATDADEPNHLNSKIAFKIVSQ-EPAGTPMFLLSR  164
             P F    +  EI EN   +S ++ + A      +  + +I + + +Q + AGT  F +
Sbjct  655   PQFYMPSYEAEIPENQKKDSDIISVKAK-----SFADREIRYTLKAQGQGAGT--FNIGP  707

Query  165   NTGEVRTLTNSLDRE---QASSYRLVVSGADKDGEGLSTQCECNIKVKDVNDNFPMFRDS  221
              +G V+ L   LD E   Q   Y LVV+ A +D  G ST  E  I+V DVNDN P F
Sbjct  708   TSGIVK-LAKELDFEDLRQPHVYSLVVT-ATEDSGGFSTSVELTIRVTDVNDNAPKFELP  765

Query  222   QYSAR-IEENI-LSSELLRFQVTDLDEEYTDNWLAVYFFTSGNEGNWFEIQTDPRTNEGI  279
              Y A  ++E+I L + +L+ +  D D         + +  S +    F + ++     GI
Sbjct  766   DYQAHNVDEDIPLGTSILKVKAMDSDSGAN---AEIEYLVSDDH---FAVDSN-----GI  814

Query  280   LKVVKALDYEQLQS-VKLSIAVKNKAEFHQSVISRYRVQSTPVTIQVINVREGIAFRPAS  338
             +   K LD +   +  +  +  K+K E  ++  +  RV +           E   F
Sbjct  815   IVNNKQLDADNNNAYYEFVVTAKDKGEPAKTGTATVRVYTKNKN------DEEPKFSQQV  868

Query  339   KTFTVQKGISSKKLVDYILGTYQAIDEDTNKAASNVK--YVMGRNDGGYLMIDSKTAEIK  396
              T  V +      LV  ++         ++K   NV+  +V G    G  +I+  T  I+
Sbjct  869   YTPNVDENAGPNTLVTTVVA--------SDKDGDNVRFGFVGGGTSSGQFVIEEITGVIR  920

Query  397   FVK---NMNRDSTFI-VNKTITAEVLAIDEYTGKTSTGTVYVRVPDFNDN------CPTA  446
                   +++RD   + V            + T  TST  V V + D NDN      C T
Sbjct  921   LHSKAISLDRDKYELNVTAMDDGSCCVNGDATIHTSTAVVVVFITDVNDNKPVFKDCGTY  980

Query  447   VLE-KDAVCSSSPSVVVSARTLNNRYTGPYTFALEDQP  483
               + ++   + SP + V A   +    G   +++  QP
Sbjct  981   YPKVEEGAPNGSPVIKVQATDEDKGVNGQVKYSIVQQP  1018

 Score = 44.7 bits (104),  Expect = 0.008, Method: Compositional matrix adjust.
 Identities = 48/199 (24%), Positives = 73/199 (37%), Gaps = 19/199 (10%)

Query  27    DYQATQKITYRISGVGIDQPPFGIFVVDKNTGDINITAIVDREETPSFLITCRALNAQGL  86
             D     K++Y I      Q  F I       G + I   +DRE TP   +   A++
Sbjct  1896  DQGGKSKVSYAIDRSSDRQRQFSI----NQEGTVTIQRNLDREVTPRHQVKILAIDDGIP  1951

Query  87    DVEKPLILTVKILDINDNPPVFSQQIFMGEIEENSASNSLVMILNATDADEPNHLNSKIA  146
                    LTV + DINDNPP F +  +   + E+     +V IL   D D
Sbjct  1952  PKTATATLTVIVQDINDNPPKFLKD-YRPVLPEHVPPRKVVEILATDDDDRSKSNGPPFQ  2010

Query  147   FKIVSQEPAGTPMFLLSRNTGEVRTLTN-----------SLDREQASSYRLVVSGADKDG  195
             F++   +P    +   S    + +   N           S DREQ   Y + +   D
Sbjct  2011  FRL---DPGADDIIRASFKVEQDQKGANGDGMAIVSSLRSFDREQQKEYLIPIVIKDHGN  2067

Query  196   EGLSTQCECNIKVKDVNDN  214
               ++      + + DVNDN
Sbjct  2068  PAMTGTSTLTVVIGDVNDN  2086

 Score = 40.0 bits (92),  Expect = 0.21, Method: Compositional matrix adjust.
 Identities = 89/389 (23%), Positives = 149/389 (38%), Gaps = 62/389 (16%)

Query  75   LITCRALNAQGLDVEKPLILTVKILDINDNPPVFSQQIF--MGEIEENSASNSLVMILNA  132
            +IT +  NA GL       + + + D+ND PP F  +       ++ N+  N+ V  L A
Sbjct  517  IITNQGHNA-GLKYTDNQRVIIHVKDVNDEPPYFINRPLPMQAVVQLNAPPNTPVFTLQA  575

Query  133  TDADEPNHLNSKIAFKIVSQEPAGTPMFLLSRNTGEVRTLTNSLDREQASSYRLVVSGAD  192
             D D     +  I + IV     G   F +   +G VRT    L  +    Y L V   D
Sbjct  576  RDPD----TDHNIHYFIVRDRTGG--RFEVDERSGVVRTRGTDL-FQLDMEYVLYVKAED  628

Query  193  KDGE----GLSTQCECNIKVKDVNDNFPMFRDSQYSARIEEN-ILSSELLRFQVTDLDEE  247
            ++G+       +  E  + +       P F    Y A I EN    S+++  +      +
Sbjct  629  QNGKVDDRRFQSTPEERLSIVG-GKRAPQFYMPSYEAEIPENQKKDSDIISVKA----KS  683

Query  248  YTDNWLAVYFFTSGNEGNWFEIQTDPRTNEGILKVVKALDYEQLQSVKLS----IAVKNK  303
            + D  +       G     F I        GI+K+ K LD+E L+   +      A ++
Sbjct  684  FADREIRYTLKAQGQGAGTFNIGP----TSGIVKLAKELDFEDLRQPHVYSLVVTATEDS  739
```

```
Query  304  AEFHQSVISRYRVQSTPVTIQVINVREGIAFRPASKTFTVQKGISSKKLVDYILGT----  359
              F  SV          +TI+V +V +       +  F +    +     D  LGT
Sbjct  740  GGFSTSV---------ELTIRVTDVNDN------APKFELPDYQAHNVDEDIPLGTSILK  784

Query  360  YQAIDEDTNKAASNVKYVMGRN----DGGYLMIDSKTAEIKFVKNMNRDSTFIVNKTITA  415
             +A+D D+  A + ++Y++  +    D   +++++K  +     N N    F+V
Sbjct  785  VKAMDSDSG-ANAEIEYLVSDDHFAVDSNGIIVNNKQLD---ADNNNAYYEFVVTAKDKG  840

Query  416  EVLAIDEYTGKTSTGTVYVRVPDFNDNCP  444
            E         KT T TV V   + ND  P
Sbjct  841  E-------PAKTGTATVRVYTKNKNDEEP  862

 Score = 39.7 bits (91),  Expect = 0.26, Method: Compositional matrix adjust.
 Identities = 54/242 (22%), Positives = 102/242 (42%), Gaps = 22/242 (9%)

Query  4    KFAKPCREGE--DNSKRNP-IAKITSDYQATQKITYRISGVGIDQPPFGIFVVDKNTGDI  60
            +F  P  E E  +N K++  I  + +   A ++I Y +   G      G F +   +G +
Sbjct  656  QFYMPSYEAEIPENQKKDSDIISVKAKSFADREIRYTLKAQGQGA---GTFNIGPTSGIV  712

Query  61   NITAIVDREE--TPSF--LITCRALNAQGLDVEKPLILTVKILDINDNPPVFSQQIFMGE  116
             +   +D E+   P    L+     ++ G      + LT+++ D+NDN P F    +
Sbjct  713  KLAKELDFEDLRQPHVYSLVVTATEDSGGFSTS--VELTIRVTDVNDNAPKFELPDYQAH  770

Query  117  -IEENSASNSLVMILNATDADEPNHLNSKIAFKIVSQEPAGTPMFLLSRNTGEVRTLTNS  175
             ++E+    + ++ + A D+D  +  N++I + +VS +      F +  N   V
Sbjct  771  NVDEDIPLGTSILKVKAMDSD--SGANAEIEY-LVSDD-----HFAVDSNGIIVNNKQLD  822

Query  176  LDREQASSYRLVVSGADKDGEGLSTQCECNIKVKDVNDNFPMFRDSQYSARIEENILSSE  235
             D   A  Y  VV+  DK     +      +  K+ ND  P F    Y+  ++EN   +
Sbjct  823  ADNNNA-YYEFVVTAKDKGEPAKTGTATVRVYTKNKNDEEPKFSQQVYTPNVDENAGPNT  881

Query  236  LL  237
            L+
Sbjct  882  LV  883

>ACMO_007424.P13503 | transcript=ACMO_007424.R13503 | gene=ACMO_007424
| organism=Anopheles_coluzzii_MOPTI | gene_product=neural-cadherin,
transcript variant X4 | transcript_product=neural-cadherin,
transcript variant X6 | location=CM029349:40739311-40945968(+)
| protein_length=3104 | sequence_SO=chromosome
| SO=protein_coding_gene | is_pseudo=false
Length=3104

 Score = 145 bits (365),  Expect = 1e-34, Method: Compositional matrix adjust.
 Identities = 125/437 (29%), Positives = 211/437 (48%), Gaps = 33/437 (8%)

Query  26    SDYQATQKITYRISGVGID--QPPFGIFVVDKNTGDINITAIVDREET---PSFLITCRA  80
              D    Q I Y ++G GID   P    F +++ TG+I +   +DR++    P +  T  A
Sbjct  1554  GDKDRPQNIVYFLTGQGIDPDNPANSKFDINRTTGEIFVLKPLDRDQPNGRPQWRFTVFA  1613

Query  81    LNAQGLDVEKPLILTVKILDINDNPPVFSQQIFMGEIEENSASNSLVMILNATDADEPNH  140
              +  G  +     + V + DINDN P+F Q ++ G + EN  +  +VM + A D D+PN
Sbjct  1614  QDEGGEGLVGYADVQVNLKDINDNAPIFPQGVYFGNVTENGTAGMVVMTMTAVDYDDPNE  1673

Query  141   -LNSKIAFKI---VSQEPAGTPMFLLSRNTGEVRTLTNSLDREQASSYRLVVSGADKDGE  196
               N+K+ + I   V +E  G+P+F +  +TG ++T    LDRE+   Y + V     DG
Sbjct  1674  GTNAKLIYSIEKNVIEEETGSPIFEIEADTGVIKTAVCCLDRERTPDYSIQVVAM--DGG  1731

Query  197   GLSTQCECNIKVKDVNDNFPMFRDSQYSARIEENILSS--ELLRFQVTDLDEEYTDNWLA  254
             GL      +I+VKD+ND  P F   ++   ++E   ++  E+    VT  DE+ T+ +
Sbjct  1732  GLKGTGTASIRVKDINDMPPQFTKDEWFTEVDETDGTNLPEMPILTVTVHDEDETNKFQY  1791

Query  255   VYFFTSGNEGNWFEIQTDPRTNEGI--LKVVKALDYE---QLQSVKLSIAVKNKAEFHQS  309
                  SG   +  F +    R N+G   LK+V+ LDYE   Q    +  I V +K E + +
Sbjct  1792  KVIDNSGYGADKFTMV---RNNDGTGSLKIVQPLDYEDPLQSNGFRFRIQVNDKGEDNDN  1848

Query  310   VISRYRVQSTPVTIQVINVREGIAFRPASKTFTVQKGISSKKLVDYILGTYQAIDEDTNK  369
                +Y V  + V +++ ++ +    +P  +   ++  +      V   L T++A D D
```

```
Sbjct  1849  --DKYHVAYSWVVVKLRDINDN---KPQFERPNIEVSVYENADVGKTLETFKATDPDQG-  1902

Query  370   AASNVKYVMGR-NDGGYLMIDSKTAEIKFVKNMNRDSTFIVNKTITAEVLAIDE-YTGKT  427
               S V Y + R +D       ++   +   +N++R+    V      ++LAID+    KT
Sbjct  1903  GKSKVSYAIDRSSDRQRQFSINQEGTVTIQRNLDRE----VTPRHQVKILAIDDGIPPKT  1958

Query  428   STGTVYVRVPDFNDNCP  444
             +T T+ V V  D NDN P
Sbjct  1959  ATATLTVIVQDINDNPP  1975

 Score = 86.3 bits (212),  Expect = 1e-15, Method: Compositional matrix adjust.
 Identities = 108/420 (26%), Positives = 177/420 (42%), Gaps = 52/420 (12%)

Query  50    IFVVDKNTGDINITAIVDREETPSFLITCRAL----------NAQGLDVEKPLILTVKIL  99
              F ++  +G+I    + DRE+  ++ +   A           N Q   V K   + + I
Sbjct  1359  FFEINLQSGEIFTKVVFDREKQGAYALEVEARDGAPSARPNSNQQPNSVTK--FIRIGIA  1416

Query  100   DINDNPPVFSQQIFMGEIEENSASNSLVMILNATDADEPNHLNSKIAFKIVSQEPAGTPM  159
             D NDNPP F + ++  E++EN      V+ + A D DE    +S+I ++I S    G
Sbjct  1417  DKNDNPPYFDKALYEAEVDENEDIQHTVLTVTAKDHDE----SSRIRYEITSGNIGGA--  1470

Query  160   FLLSRNTGEVRTLTNSLDREQASSYRLVVSGADKDGEGLSTQCECNIKVKDVNDNFPMFR  219
             F +   TG +  +  +LD E    Y L ++ +D   E  +T      I VKDVNDN P+F
Sbjct  1471  FAVKNMTGAI-YVAGALDYETRKRYELRLAASDNLKENYTTVV---IHVKDVNDNPPVFE  1526

Query  220   DSQYSARIEENI---LSSELLRFQVTDLDEEYTDNWLAVYFFTSG--NEGNWFEIQTDPR  274
                Y  +I E     L   +L+   TD D++   N   VYF T    +  N    + D
Sbjct  1527  RPTYRTQITEEDDRNLPKRVLQVTATDGDKDRPQN--IVYFLTGQGIDPDNPANSKFDIN  1584

Query  275   TNEGILKVVKALDYEQLQS-VKLSIAVKNKAEFHQSVISRYRVQSTPVTIQVINVREGIA  333
                G + V+K LD +Q      +    V  + E  + ++    VQ     + + ++ +
Sbjct  1585  RTTGEIFVLKPLDRDQPNGRPQWRFTVFAQDEGGEGLVGYADVQ-----VNLKDINDNAP  1639

Query  334   FRPASKTFTVQKGISSKKLVDYILGTYQAIDEDTNKAASNVK--YVMGRN------DGGY  385
               P    F     ++        ++ T  A+D D     +N K  Y + +N
Sbjct  1640  IFPQGVYF---GNVTENGTAGMVVMTMTAVDYDDPNEGTNAKLIYSIEKNVIEEETGSPI  1696

Query  386   LMIDSKTAEIK-FVKNMNRDSTFIVNKTITAEVLAIDEYTGKTSTGTVYVRVPDFNDNCP  444
               I++ T  IK  V  ++R+ T        + +V+A+D   G   TGT  +RV D ND  P
Sbjct  1697  FEIEADTGVIKTAVCCLDRERT----PDYSIQVVAMDG-GGLKGTGTASIRVKDINDMPP  1751

 Score = 76.3 bits (186),  Expect = 1e-12, Method: Compositional matrix adjust.
 Identities = 106/426 (25%), Positives = 181/426 (42%), Gaps = 51/426 (12%)

Query  53    VDKNTGDINITAIVDREETPSFLITCRALNAQGLDVEKPLILTVKILDINDNPPVFSQ-Q  111
             +D++  ++N+TA+ D          +C  +N           ++ V I D+NDN PVF
Sbjct  928   LDRDKYELNVTAMDDG--------SC-CVNGDATIHTSTAVVVVFITDVNDNKPVFKDCG  978

Query  112   IFMGEIEENSASNSLVMILNATDADEPNHLNSKIAFKIVSQEPAGTPMFLLSRNTGEVRT  171
              +  ++EE + + S V+ + ATD D+    +N ++ + IV Q        F +   TGEV T
Sbjct  979   TYYPKVEEGAPNGSPVIKVQATDEDK--GVNGQVKYSIVQQPNQKGTKFTVDEETGEVST  1036

Query  172   LTNSLDRE--QASSYRLVVSGADKDGEGLSTQCECNIKVKDVNDNFPMFRDSQYSARIEE  229
                  DRE         + V   D+     L   C    +++ DVNDN P+F    +Y   +++
Sbjct  1037  -NKVFDREGDDGKFVSVTVKATDQGEPSLEGVCSFTVEITDVNDNPPLFDRQKYVENVKQ  1095

Query  230   NI-LSSELLRFQVTDLDEEYTDNWLAVYFFT---SGNEGNWFEIQTDPRTNEGILKVVKA  285
             +  + + +LR  V+  DE+  +N   VY  +   S N+  +FEIQ  P +   +LK  K
Sbjct  1096  DASIGTNILR--VSASDEDADNNGAIVYTLSAPHSPNDLEYFEIQ--PESGWIVLK--KP  1149

Query  286   LD--YEQLQSVKLSIAVKNK------AEFHQSVISRYRVQSTPVTIQV----INVREGIA  333
             LD  Y + +  +L +   ++      A+     +   R    P+  +     I++RE +
Sbjct  1150  LDGCYSR-ERYRLKVYASDRGDPPSTADVDVELDVVDRNNKPPIWDRTIYGPIHIRENVT  1208

Query  334   FRPASKTFTVQKGISSKKLVDYILGTYQAIDEDTNKAASNVKYVMGRNDGGYLMIDSKTA  393
                   +      GI      V Y L           TNK   +  Y+  R D G         A
Sbjct  1209  VGTVVTSVKASSGIEGNPTVFYRL--MPGSTAQTNKL--HTFYLQQRPDNG-----DTWA  1259

Query  394   EIKFVKNMNRDSTFIVNKTITAEVLAIDEYTGKTSTGTVYVRVPDFNDNCPTAV-LEKDA  452
```

```
             +IK    ++ ++    N T+  E     +     S  TVY+++ D ND  P     E++
Sbjct  1260  DIKVNHPLDFETIKEYNLTVRVENNGAQQL---ASEATVYIQLEDVNDEIPLFTEREQET  1316

Query  453   VCSSSP  458
             V    P
Sbjct  1317  VLEGEP  1322

 Score = 60.8 bits (146),  Expect = 7e-08, Method: Compositional matrix adjust.
 Identities = 83/362 (23%), Positives = 152/362 (42%), Gaps = 34/362 (9%)

Query  51    FVVDKNTGDINITAIVDRE-ETPSFL-ITCRALNAQGLDVEKPLILTVKILDINDNPPVF  108
             F VD+ TG+++   + DRE +   F+ +T +A +     +E     TV+I D+NDNPP+F
Sbjct  1025  FTVDEETGEVSTNKVFDREGDDGKFVSVTVKATDQGEPSLEGVCSFTVEITDVNDNPPLF  1084

Query  109   SQQIFMGEIEENSASNSLVMILNATDADEPNHLNSKIAFKIVS-QEPAGTPMFLLSRNTG  167
              +Q ++  ++++++  + ++ ++A+D D  N  N  I + + +   P     F +   +G
Sbjct  1085  DRQKYVENVKQDASIGTNILRVSASDEDADN--NGAIVYTLSAPHSPNDLEYFEIQPESG  1142

Query  168   EV---RTLTNSLDREQASSYRLVVSGADKDGEGLSTQCECNIKVKDVNDNFPMFRDSQYS  224
              +   + L     RE+   YRL V  +D+ G+  ST            +N P   D
Sbjct  1143  WIVLKKPLDGCYSRER---YRLKVYASDR-GDPPSTADVDVELDVVDRNNKPPIWDRTIY  1198

Query  225   ARIEENILSSELLRFQVTDLDEEYTDNWLAVYFFTSG-----NEGNWFEIQTDPRTNE--  277
               I      +                N    Y    G     N+ + F +Q  P   +
Sbjct  1199  GPIHIRENVTVGTVVTSVKASSGIEGNPTVFYRLMPGSTAQTNKLHTFYLQQRPDNGDTW  1258

Query  278   GILKVVKALDYEQLQSVKLSIAVKNKAEFHQSVISRYRVQSTPVTIQVINVREGIAFRPA  337
               +KV   LD+E ++   L++ V+N         ++       V IQ+ +V + I
Sbjct  1259  ADIKVNHPLDFETIKEYNLTVRVENNG-------AQQLASEATVYIQLEDVNDEIPLFTE  1311

Query  338   SKTFTVQKGISSKKLVDYILGTYQAIDEDTNKAASNVKYVM---GRNDG-GYLMIDSKTA  393
              +  TV +G      V  +     AID+D     + V Y +    RN+G  +  I+ ++
Sbjct  1312  REQETVLEGEPIGTKVTQV----NAIDKDGTFPNNQVYYYIVDSPRNEGKDFFEINLQSG  1367

Query  394   EI  395
             EI
Sbjct  1368  EI  1369

 Score = 55.8 bits (133),  Expect = 2e-06, Method: Compositional matrix adjust.
 Identities = 49/158 (31%), Positives = 78/158 (49%), Gaps = 24/158 (15%)

Query  102   NDNPPVFSQQIFMGEIEENSASNSLVMILNATDADEPNHLNSKIAFKIVSQEPAGTP--M  159
             ND  P FSQQ++   ++EN+  N+LV  + A+D D  N     + F  V     GT
Sbjct  858   NDEEPKFSQQVYTPNVDENAGPNTLVTTVVASDKDGDN-----VRFGFVG---GGTSSGQ  909

Query  160   FLLSRNTGEVRTLTN--SLDREQASSYRLVVSGADK-------DGEGLSTQCECNIKVKD  210
             F++   TG +R  +   SLDR++   Y L V+  D        D    ++     + + D
Sbjct  910   FVIEEITGVIRLHSKAISLDRDK---YELNVTAMDDGSCCVNGDATIHTSTAVVVVFITD  966

Query  211   VNDNFPMFRD-SQYSARIEENILS-SELLRFQVTDLDE  246
             VNDN P+F+D   Y  ++EE   + S +++ Q TD D+
Sbjct  967   VNDNKPVFKDCGTYYPKVEEGAPNGSPVIKVQATDEDK  1004

 Score = 53.9 bits (128),  Expect = 1e-05, Method: Compositional matrix adjust.
 Identities = 70/306 (23%), Positives = 126/306 (41%), Gaps = 30/306 (10%)

Query  20    PIAKIT-SDYQATQKITYRI---SGVGIDQPPFGIFVVDKNTGDINITAIVDRE---ETP  72
             PI  +T  D   T K  Y++   SG G D+  F +   +  TG + I   +D E   ++
Sbjct  1774  PILTVTVHDEDETNKFQYKVIDNSGYGADK--FTMVRNNDGTGSLKIVQPLDYEDPLQSN  1831

Query  73    SFLITCRALNAQGLDVEKPLI------LTVKILDINDNPPVFSQQIFMGEIEENSASNSL  126
              F    + +N +G D +          + VK+ DINDN P F +      + EN+
Sbjct  1832  GFRFRIQ-VNDKGEDNDNDKYHVAYSWVVVKLRDINDNKPQFERPNIEVSVYENADVGKT  1890

Query  127   VMILNATDADEPNHLNSKIAFKIVSQEPAGTPMFLLSRNTGEVRTLTNSLDREQASSYRL  186
             +    ATD D+     SK+++ I   + +       S N     T+  +LDRE    +++
Sbjct  1891  LETFKATDPDQGG--KSKVSYAI---DRSSDRQRQFSINQEGTVTIQRNLDREVTPRHQV  1945
```

```
Query  187   VVSGADKDGEGLSTQCECNIKVKDVNDNFPMFRDSQYSARIEENILSSELLRFQVTDLDE  246
              +   D      +      + V+D+NDN P F    Y   + E++   +++    TD D+
Sbjct  1946  KILAIDDGIPPKTATATLTVIVQDINDNPPKFL-KDYRPVLPEHVPPRKVVEILATDDDD  2004

Query  247   EYTDNWLAVYFFTSGNEGNW----FEIQTDPRTNEG----ILKVVKALDYEQLQSVKLSI  298
                  N     F       +     F+++ D +   G    I+  +++ D EQ +   + I
Sbjct  2005  RSKSNGPPFQFRLDPGADDIIRASFKVEQDQKGANGDGMAIVSSLRSFDREQQKEYLIPI  2064

Query  299   AVKNKA  304
              +K+
Sbjct  2065  VIKDHG  2070

 Score = 52.4 bits (124),  Expect = 3e-05, Method: Compositional matrix adjust.
 Identities = 97/398 (24%), Positives = 163/398 (41%), Gaps = 54/398 (14%)

Query  106   PVFSQQIFMGEIEENSASNSLVMILNATDADEPNHLNSKIAFKIVSQ-EPAGTPMFLLSR  164
             P F    +  EI EN   +S ++ + A       +  + +I + + +Q + AGT  F +
Sbjct  655   PQFYMPSYEAEIPENQKKDSDIISVKAK-----SFADREIRYTLKAQGQGAGT--FNIGP  707

Query  165   NTGEVRTLTNSLDRE---QASSYRLVVSGADKDGEGLSTQCECNIKVKDVNDNFPMFRDS  221
              +G V+ L   LD E   Q   Y LVV+ A +D  G ST  E  I+V DVNDN P F
Sbjct  708   TSGIVK-LAKELDFEDLRQPHVYSLVVT-ATEDSGGFSTSVELTIRVTDVNDNAPKFELP  765

Query  222   QYSAR-IEENI-LSSELLRFQVTDLDEEYTDNWLAVYFFTSGNEGNWFEIQTDPRTNEGI  279
              Y A  ++E+I L + +L+ +  D D          + +  S +    F + ++     GI
Sbjct  766   DYQAHNVDEDIPLGTSILKVKAMDSDSGAN---AEIEYLVSDDH---FAVDSN-----GI  814

Query  280   LKVVKALDYEQLQS-VKLSIAVKNKAEFHQSVISRYRVQSTPVTIQVINVREGIAFRPAS  338
             +   K LD +   +  +  +  K+K E  ++  +  RV +             E   F
Sbjct  815   IVNNKQLDADNNNAYYEFVVTAKDKGEPAKTGTATVRVYTKNKN------DEEPKFSQQV  868

Query  339   KTFTVQKGISSKKLVDYILGTYQAIDEDTNKAASNVK--YVMGRNDGGYLMIDSKTAEIK  396
              T  V +       LV  ++          ++K   NV+  +V G    G  +I+  T  I+
Sbjct  869   YTPNVDENAGPNTLVTTVVA--------SDKDGDNVRFGFVGGGTSSGQFVIEEITGVIR  920

Query  397   FVK---NMNRDSTFI-VNKTITAEVLAIDEYTGKTSTGTVYVRVPDFNDN------CPTA  446
                   +++RD   + V              + T  TST  V V + D NDN      C T
Sbjct  921   LHSKAISLDRDKYELNVTAMDDGSCCVNGDATIHTSTAVVVVFITDVNDNKPVFKDCGTY  980

Query  447   VLE-KDAVCSSSPSVVVSARTLNNRYTGPYTFALEDQP  483
               + ++   + SP + V A   +    G   +++  QP
Sbjct  981   YPKVEEGAPNGSPVIKVQATDEDKGVNGQVKYSIVQQP  1018

 Score = 44.7 bits (104),  Expect = 0.007, Method: Compositional matrix adjust.
 Identities = 48/199 (24%), Positives = 73/199 (37%), Gaps = 19/199 (10%)

Query  27    DYQATQKITYRISGVGIDQPPFGIFVVDKNTGDINITAIVDREETPSFLITCRALNAQGL  86
             D     K++Y I       Q  F I        G + I   +DRE TP   +   A++
Sbjct  1900  DQGGKSKVSYAIDRSSDRQRQFSI----NQEGTVTIQRNLDREVTPRHQVKILAIDDGIP  1955

Query  87    DVEKPLILTVKILDINDNPPVFSQQIFMGEIEENSASNSLVMILNATDADEPNHLNSKIA  146
                     LTV + DINDNPP F +  +    + E+      +V IL   D D
Sbjct  1956  PKTATATLTVIVQDINDNPPKFLKD-YRPVLPEHVPPRKVVEILATDDDDRSKSNGPPFQ  2014

Query  147   FKIVSQEPAGTPMFLLSRNTGEVRTLTN-----------SLDREQASSYRLVVSGADKDG  195
             F++   +P     +   S     + +   N             S DREQ    Y + +   D
Sbjct  2015  FRL---DPGADDIIRASFKVEQDQKGANGDGMAIVSSLRSFDREQQKEYLIPIVIKDHGN  2071

Query  196   EGLSTQCECNIKVKDVNDN  214
               ++       + + DVNDN
Sbjct  2072  PAMTGTSTLTVVIGDVNDN  2090

 Score = 40.0 bits (92),  Expect = 0.19, Method: Compositional matrix adjust.
 Identities = 89/389 (23%), Positives = 149/389 (38%), Gaps = 62/389 (16%)

Query  75   LITCRALNAQGLDVEKPLILTVKILDINDNPPVFSQQIF--MGEIEENSASNSLVMILNA  132
            +IT +   NA GL        + + + D+ND PP F  +        ++ N+  N+ V  L A
Sbjct  517  IITNQGHNA-GLKYTDNQRVIIHVKDVNDEPPYFINRPLPMQAVVQLNAPPNTPVFTLQA  575
```

```
Query  133  TDADEPNHLNSKIAFKIVSQEPAGTPMFLLSRNTGEVRTLTNSLDREQASSYRLVVSGAD  192
             D D     +  I + IV     G   F +   +G VRT    L  +    Y L V   D
Sbjct  576  RDPD----TDHNIHYFIVRDRTGG--RFEVDERSGVVRTRGTDL-FQLDMEYVLYVKAED  628

Query  193  KDGE----GLSTQCECNIKVKDVNDNFPMFRDSQYSARIEEN-ILSSELLRFQVTDLDEE  247
            ++G+        +  E  + +       P F    Y A I EN    S+++  +     +
Sbjct  629  QNGKVDDRRFQSTPEERLSIVG-GKRAPQFYMPSYEAEIPENQKKDSDIISVKA----KS  683

Query  248  YTDNWLAVYFFTSGNEGNWFEIQTDPRTNEGILKVVKALDYEQLQSVKLS----IAVKNK  303
            + D  +        G     F I         GI+K+ K LD+E L+   +       A ++
Sbjct  684  FADREIRYTLKAQGQGAGTFNIGP----TSGIVKLAKELDFEDLRQPHVYSLVVTATEDS  739

Query  304  AEFHQSVISRYRVQSTPVTIQVINVREGIAFRPASKTFTVQKGISSKKLVDYILGT----  359
              F  SV           +TI+V +V +       +  F +    +     D  LGT
Sbjct  740  GGFSTSV---------ELTIRVTDVNDN------APKFELPDYQAHNVDEDIPLGTSILK  784

Query  360  YQAIDEDTNKAASNVKYVMGRN----DGGYLMIDSKTAEIKFVKNMNRDSTFIVNKTITA  415
             +A+D D+  A + ++Y++  +    D   +++++K  +     N N    F+V
Sbjct  785  VKAMDSDSG-ANAEIEYLVSDDHFAVDSNGIIVNNKQLD---ADNNNAYYEFVVTAKDKG  840

Query  416  EVLAIDEYTGKTSTGTVYVRVPDFNDNCP  444
            E         KT T TV V   + ND  P
Sbjct  841  E-------PAKTGTATVRVYTKNKNDEEP  862

 Score = 39.7 bits (91),  Expect = 0.24, Method: Compositional matrix adjust.
 Identities = 54/242 (22%), Positives = 102/242 (42%), Gaps = 22/242 (9%)

Query  4    KFAKPCREGE--DNSKRNP-IAKITSDYQATQKITYRISGVGIDQPPFGIFVVDKNTGDI  60
            +F  P  E E  +N K++  I  + +   A ++I Y +   G      G F +   +G +
Sbjct  656  QFYMPSYEAEIPENQKKDSDIISVKAKSFADREIRYTLKAQGQGA---GTFNIGPTSGIV  712

Query  61   NITAIVDREE--TPSF--LITCRALNAQGLDVEKPLILTVKILDINDNPPVFSQQIFMGE  116
             +   +D E+   P    L+     ++ G       + LT+++ D+NDN P F    +
Sbjct  713  KLAKELDFEDLRQPHVYSLVVTATEDSGGFSTS--VELTIRVTDVNDNAPKFELPDYQAH  770

Query  117  -IEENSASNSLVMILNATDADEPNHLNSKIAFKIVSQEPAGTPMFLLSRNTGEVRTLTNS  175
             ++E+    + ++ + A D+D  +  N++I + +VS +       F +  N   V
Sbjct  771  NVDEDIPLGTSILKVKAMDSD--SGANAEIEY-LVSDD-----HFAVDSNGIIVNNKQLD  822

Query  176  LDREQASSYRLVVSGADKDGEGLSTQCECNIKVKDVNDNFPMFRDSQYSARIEENILSSE  235
             D   A  Y  VV+  DK      +      +  K+ ND  P F    Y+  ++EN   +
Sbjct  823  ADNNNA-YYEFVVTAKDKGEPAKTGTATVRVYTKNKNDEEPKFSQQVYTPNVDENAGPNT  881

Query  236  LL  237
            L+
Sbjct  882  LV  883

>ADAR2_001930.P21643 | transcript=ADAR2_001930.R21643 | gene=ADAR2_001930
| organism=Anopheles_darlingi_AdarGF1 | gene_product=neural-cadherin,
transcript variant X6 | transcript_product=neural-cadherin,
transcript variant X6 | location=AdarGF1_2RL:74991964-75171550(+)
| protein_length=3130 | sequence_SO=chromosome
| SO=protein_coding_gene | is_pseudo=false
Length=3130

 Score = 145 bits (365),  Expect = 1e-34, Method: Compositional matrix adjust.
 Identities = 125/437 (29%), Positives = 211/437 (48%), Gaps = 33/437 (8%)

Query  26    SDYQATQKITYRISGVGID--QPPFGIFVVDKNTGDINITAIVDREET---PSFLITCRA  80
              D    Q I Y ++G GID    P     F +++ TG+I +    +DR++     P +  T  A
Sbjct  1579  GDKDRPQNIVYFLTGQGIDPDNPANSKFDINRTTGEIFVLKPLDRDQPNGRPQWRFTVFA  1638

Query  81    LNAQGLDVEKPLILTVKILDINDNPPVFSQQIFMGEIEENSASNSLVMILNATDADEPNH  140
               +  G  +      + V + DINDN P+F Q ++ G + EN  +  +VM + A D D+PN
Sbjct  1639  QDEGGEGLVGYADVQVNLKDINDNAPIFPQGVYFGNVTENGTAGMVVMTMTAVDYDDPNE  1698

Query  141   -LNSKIAFKI---VSQEPAGTPMFLLSRNTGEVRTLTNSLDREQASSYRLVVSGADKDGE  196
                N+K+ + I   V +E  G+P+F +  +TG ++T    LDRE+   Y + V      DG
```

```
Sbjct  1699  GTNAKLIYSIEKNVIEEETGSPIFEIEADTGVIKTAVCCLDRERTPDYSIQVVAM--DGG  1756

Query  197   GLSTQCECNIKVKDVNDNFPMFRDSQYSARIEENILSS--ELLRFQVTDLDEEYTDNWLA  254
             GL      +I+VKD+ND  P F   ++   ++E   ++  E+    VT  DE+ T+ +
Sbjct  1757  GLKGTGTASIRVKDINDMPPQFTKDEWFTEVDETDGTNLPEMPILTVTVHDEDETNKFQY  1816

Query  255   VYFFTSGNEGNWFEIQTDPRTNEGI--LKVVKALDYE---QLQSVKLSIAVKNKAEFHQS  309
                  SG   + F +    R N+G   LK+V+ LDYE   Q    +  I V +K E + +
Sbjct  1817  KVIDNSGYGADKFTMV---RNNDGTGSLKIVQPLDYEDPLQSNGFRFRIQVNDKGEDNDN  1873

Query  310   VISRYRVQSTPVTIQVINVREGIAFRPASKTFTVQKGISSKKLVDYILGTYQAIDEDTNK  369
                +Y V  + V +++ ++ +    +P  +   ++  +     V   L T++A D D
Sbjct  1874  --DKYHVAYSWVVVKLRDINDN---KPQFERPNIEVSVYENADVGKTLETFKATDPDQG-  1927

Query  370   AASNVKYVMGR-NDGGYLMIDSKTAEIKFVKNMNRDSTFIVNKTITAEVLAIDE-YTGKT  427
               S V Y + R +D       ++   +   +N++R+    V      ++LAID+    KT
Sbjct  1928  GKSKVSYAIDRSSDRQRQFSINQEGTVTIQRNLDRE----VTPRHQVKILAIDDGIPPKT  1983

Query  428   STGTVYVRVPDFNDNCP  444
             +T T+ V V D NDN P
Sbjct  1984  ATATLTVIVQDINDNPP  2000

 Score = 85.9 bits (211),  Expect = 1e-15, Method: Compositional matrix adjust.
 Identities = 108/420 (26%), Positives = 177/420 (42%), Gaps = 52/420 (12%)

Query  50    IFVVDKNTGDINITAIVDREETPSFLITCRAL----------NAQGLDVEKPLILTVKIL  99
              F ++  +G+I    + DRE+  ++ +   A           N Q   V K   + + I
Sbjct  1384  FFEINLQSGEIFTKVVFDREKQGAYALEVEARDGAPSARPNSNQQPNSVTK--FIRIGIA  1441

Query  100   DINDNPPVFSQQIFMGEIEENSASNSLVMILNATDADEPNHLNSKIAFKIVSQEPAGTPM  159
             D NDNPP F + ++  E++EN      V+ + A D DE    +S+I ++I S    G
Sbjct  1442  DKNDNPPYFDKALYEAEVDENEDIQHTVLTVTAKDHDE----SSRIRYEITSGNIGGA--  1495

Query  160   FLLSRNTGEVRTLTNSLDREQASSYRLVVSGADKDGEGLSTQCECNIKVKDVNDNFPMFR  219
             F +   TG +  +  +LD E    Y L ++ +D   E  +T     I VKDVNDN P+F
Sbjct  1496  FAVKNMTGAI-YVAGALDYETRKRYELRLAASDNLKENYTTVV---IHVKDVNDNPPVFE  1551

Query  220   DSQYSARIEENI---LSSELLRFQVTDLDEEYTDNWLAVYFFTSG--NEGNWFEIQTDPR  274
                Y  +I E     L   +L+   TD D++   N   VYF T    +  N    + D
Sbjct  1552  RPTYRTQITEEDDRNLPKRVLQVTATDGDKDRPQN--IVYFLTGQGIDPDNPANSKFDIN  1609

Query  275   TNEGILKVVKALDYEQLQS-VKLSIAVKNKAEFHQSVISRYRVQSTPVTIQVINVREGIA  333
                G + V+K LD +Q     +    V  + E  + ++    VQ     + + ++ +
Sbjct  1610  RTTGEIFVLKPLDRDQPNGRPQWRFTVFAQDEGGEGLVGYADVQ-----VNLKDINDNAP  1664

Query  334   FRPASKTFTVQKGISSKKLVDYILGTYQAIDEDTNKAASNVK--YVMGRN------DGGY  385
               P    F     ++        ++ T  A+D D     +N K  Y + +N
Sbjct  1665  IFPQGVYF---GNVTENGTAGMVVMTMTAVDYDDPNEGTNAKLIYSIEKNVIEEETGSPI  1721

Query  386   LMIDSKTAEIK-FVKNMNRDSTFIVNKTITAEVLAIDEYTGKTSTGTVYVRVPDFNDNCP  444
               I++ T  IK  V  ++R+ T       + +V+A+D   G   TGT  +RV D ND  P
Sbjct  1722  FEIEADTGVIKTAVCCLDRERT----PDYSIQVVAMDG-GGLKGTGTASIRVKDINDMPP  1776

 Score = 80.5 bits (197),  Expect = 6e-14, Method: Compositional matrix adjust.
 Identities = 107/424 (25%), Positives = 178/424 (42%), Gaps = 51/424 (12%)

Query  53    VDKNTGDINITAIVDREETPSFLITCRALNAQGLDVEKPLILTVKILDINDNPPVFSQ-Q  111
             +D++  ++N+TA+ D          +C  +N          ++ V I D+NDN PVF
Sbjct  957   LDRDKYELNVTAMDDG--------SC-CVNGDATIHTSTAVVVVFITDVNDNKPVFKDCG  1007

Query  112   IFMGEIEENSASNSLVMILNATDADEPNHLNSKIAFKIVSQEPAGTPMFLLSRNTGEVRT  171
              +  ++EE + + S V+ + ATD D+   +N ++ + IV Q       F +   TGEV T
Sbjct  1008  TYYPKVEEGAPNGSPVIKVQATDEDK--GVNGQVKYSIVQQPNQKGTKFTVDEETGEVST  1065

Query  172   LTNSLDRE--QASSYRLVVSGADKDGEGLSTQCECNIKVKDVNDNFPMFRDSQYSARIEE  229
                  DRE        + V   D+    L   C   +++ DVNDN P+F   +Y   +++
Sbjct  1066  -NKVFDREGDDGKFVSVTVKATDQGEPSLEGVCSFTVEITDVNDNPPLFDRQKYVENVKQ  1124

Query  230   NI-LSSELLRFQVTDLDEEYTDNWLAVYFFT---SGNEGNWFEIQTDPRTNEGILKVVKA  285
```

```
             +  + + +LR  V+  DE+  +N   VY  +   S N+  +FEIQ  P +   +LK  K
Sbjct  1125  DASIGTNILR--VSASDEDADNNGAIVYTLSAPHSPNDLEYFEIQ--PESGWIVLK--KP  1178

Query  286   LDYEQLQSVKLSIAVKNK------AEFHQSVISRYRVQSTPV----TIQVINVREGIAFR  335
             LD E+    +L +   ++      A+    +    R    P+        I++RE +
Sbjct  1179  LDRERY---RLKVYASDRGDPPSTADVDVELDVVDRNNKPPIWDRNIYGPIHIRENVTVG  1235

Query  336   PASKTFTVQKGISSKKLVDYILGTYQAIDEDTNKAASNVKYVMGRNDGGYLMIDSKTAEI  395
                       GI     V Y L          TNK   +  Y+  R D G         A+I
Sbjct  1236  TVVTAIKASSGIEGNPTVFYRL--MPGSTAQTNKL--HTFYLQQRPDNG-----DTWADI  1286

Query  396   KFVKNMNRDSTFIVNKTITAEVLAIDEYTGKTSTGTVYVRVPDFNDNCPTAV-LEKDAVC  454
             K    ++ ++    N TI  E     +     S  TVY+++ D ND  P     E++ V
Sbjct  1287  KVNHPLDFETIKEYNLTIRVENNGAQQL---ASEATVYIQLEDVNDEIPLFTEREQETVL  1343

Query  455   SSSP  458
                P
Sbjct  1344  EGEP  1347

 Score = 68.6 bits (166),  Expect = 4e-10, Method: Compositional matrix adjust.
 Identities = 88/361 (24%), Positives = 157/361 (43%), Gaps = 36/361 (10%)

Query  51    FVVDKNTGDINITAIVDRE-ETPSFL-ITCRALNAQGLDVEKPLILTVKILDINDNPPVF  108
             F VD+ TG+++   + DRE +   F+ +T +A +     +E     TV+I D+NDNPP+F
Sbjct  1054  FTVDEETGEVSTNKVFDREGDDGKFVSVTVKATDQGEPSLEGVCSFTVEITDVNDNPPLF  1113

Query  109   SQQIFMGEIEENSASNSLVMILNATDADEPNHLNSKIAFKIVS-QEPAGTPMFLLSRNTG  167
              +Q ++  ++++++  + ++ ++A+D D  N  N  I + + +   P     F +   +G
Sbjct  1114  DRQKYVENVKQDASIGTNILRVSASDEDADN--NGAIVYTLSAPHSPNDLEYFEIQPESG  1171

Query  168   EVRTLTNSLDREQASSYRLVVSGADKDGEGLSTQCECNIKVKDVNDNFPMFRDSQYSA--  225
              +  L   LDRE+   YRL V  +D+ G+  ST            +N P   D
Sbjct  1172  WI-VLKKPLDRER---YRLKVYASDR-GDPPSTADVDVELDVVDRNNKPPIWDRNIYGPI  1226

Query  226   RIEENILSSELLRFQVTDLDEEYTDNWLAVYFFTSG-----NEGNWFEIQTDPRTNE--G  278
              I EN+    ++               N    Y    G     N+ + F +Q  P   +
Sbjct  1227  HIRENVTVGTVV--TAIKASSGIEGNPTVFYRLMPGSTAQTNKLHTFYLQQRPDNGDTWA  1284

Query  279   ILKVVKALDYEQLQSVKLSIAVKNKAEFHQSVISRYRVQSTPVTIQVINVREGIAFRPAS  338
              +KV   LD+E ++   L+I V+N          ++        V IQ+ +V + I
Sbjct  1285  DIKVNHPLDFETIKEYNLTIRVENNG-------AQQLASEATVYIQLEDVNDEIPLFTER  1337

Query  339   KTFTVQKGISSKKLVDYILGTYQAIDEDTNKAASNVKYVM---GRNDG-GYLMIDSKTAE  394
             +  TV +G      V  +     AID+D     + V Y +    RN+G  +  I+ ++ E
Sbjct  1338  EQETVLEGEPIGTKVTQV----NAIDKDGTFPNNQVYYYIVDSPRNEGKDFFEINLQSGE  1393

Query  395   I  395
             I
Sbjct  1394  I  1394

 Score = 55.8 bits (133),  Expect = 3e-06, Method: Compositional matrix adjust.
 Identities = 49/158 (31%), Positives = 78/158 (49%), Gaps = 24/158 (15%)

Query  102   NDNPPVFSQQIFMGEIEENSASNSLVMILNATDADEPNHLNSKIAFKIVSQEPAGTP--M  159
             ND  P FSQQ++   ++EN+  N+LV  + A+D D  N      + F  V     GT
Sbjct  887   NDEEPKFSQQVYTPNVDENAGPNTLVTTVVASDKDGDN-----VRFGFVG---GGTSSGQ  938

Query  160   FLLSRNTGEVRTLTN--SLDREQASSYRLVVSGADK-------DGEGLSTQCECNIKVKD  210
             F++   TG +R  +   SLDR+   Y L V+  D         D    ++     + + D
Sbjct  939   FVIEEITGVIRLHSKAISLDRDK---YELNVTAMDDGSCCVNGDATIHTSTAVVVVFITD  995

Query  211   VNDNFPMFRD-SQYSARIEENILS-SELLRFQVTDLDE  246
             VNDN P+F+D   Y  ++EE   + S +++ Q TD D+
Sbjct  996   VNDNKPVFKDCGTYYPKVEEGAPNGSPVIKVQATDEDK  1033

 Score = 53.9 bits (128),  Expect = 1e-05, Method: Compositional matrix adjust.
 Identities = 70/306 (23%), Positives = 126/306 (41%), Gaps = 30/306 (10%)
```

```
Query  20    PIAKIT-SDYQATQKITYRI---SGVGIDQPPFGIFVVDKNTGDINITAIVDRE---ETP  72
             PI  +T  D   T K  Y++   SG G D+  F +   +  TG + I   +D E   ++
Sbjct  1799  PILTVTVHDEDETNKFQYKVIDNSGYGADK--FTMVRNNDGTGSLKIVQPLDYEDPLQSN  1856

Query  73    SFLITCRALNAQGLDVEKPLI------LTVKILDINDNPPVFSQQIFMGEIEENSASNSL  126
              F    + +N +G D +          + VK+ DINDN P F +      + EN+
Sbjct  1857  GFRFRIQ-VNDKGEDNDNDKYHVAYSWVVVKLRDINDNKPQFERPNIEVSVYENADVGKT  1915

Query  127   VMILNATDADEPNHLNSKIAFKIVSQEPAGTPMFLLSRNTGEVRTLTNSLDREQASSYRL  186
             +    ATD D+     SK+++ I   + +       S N     T+  +LDRE    +++
Sbjct  1916  LETFKATDPDQGG--KSKVSYAI---DRSSDRQRQFSINQEGTVTIQRNLDREVTPRHQV  1970

Query  187   VVSGADKDGEGLSTQCECNIKVKDVNDNFPMFRDSQYSARIEENILSSELLRFQVTDLDE  246
              +   D      +      + V+D+NDN P F    Y   + E++   +++    TD D+
Sbjct  1971  KILAIDDGIPPKTATATLTVIVQDINDNPPKFL-KDYRPVLPEHVPPRKVVEILATDDDD  2029

Query  247   EYTDNWLAVYFFTSGNEGNW----FEIQTDPRTNEG----ILKVVKALDYEQLQSVKLSI  298
                 N     F       +     F+++ D +   G    I+  +++ D EQ +   + I
Sbjct  2030  RSKSNGPPFQFRLDPGADDIIRASFKVEQDQKGANGDGMAIVSSLRSFDREQQKEYLIPI  2089

Query  299   AVKNKA  304
              +K+
Sbjct  2090  VIKDHG  2095

 Score = 52.8 bits (125),  Expect = 2e-05, Method: Compositional matrix adjust.
 Identities = 97/398 (24%), Positives = 163/398 (41%), Gaps = 54/398 (14%)

Query  106   PVFSQQIFMGEIEENSASNSLVMILNATDADEPNHLNSKIAFKIVSQ-EPAGTPMFLLSR  164
             P F    +  EI EN   +S ++ + A      +  + +I + + +Q + AGT  F +
Sbjct  684   PQFYMPSYEAEIPENQKKDSDIISVKAK-----SFADREIRYTLKAQGQGAGT--FNIGP  736

Query  165   NTGEVRTLTNSLDRE---QASSYRLVVSGADKDGEGLSTQCECNIKVKDVNDNFPMFRDS  221
              +G V+ L   LD E   Q   Y LVV+ A +D  G ST  E  I+V DVNDN P F
Sbjct  737   TSGIVK-LAKELDFEDLRQPHVYSLVVT-ATEDSGGFSTSVELTIRVTDVNDNAPKFELP  794

Query  222   QYSAR-IEENI-LSSELLRFQVTDLDEEYTDNWLAVYFFTSGNEGNWFEIQTDPRTNEGI  279
              Y A  ++E+I L + +L+ +  D D         + +  S +    F + ++     GI
Sbjct  795   DYQAHNVDEDIPLGTSILKVKAMDADSGAN---AEIEYLVSDDH---FAVDSN-----GI  843

Query  280   LKVVKALDYEQLQS-VKLSIAVKNKAEFHQSVISRYRVQSTPVTIQVINVREGIAFRPAS  338
             +   K LD +   +  +  +  K+K E  ++  +  RV +           E   F
Sbjct  844   IVNNKQLDADNNNAYYEFVVTAKDKGEPAKTGTATVRVYTKNKN------DEEPKFSQQV  897

Query  339   KTFTVQKGISSKKLVDYILGTYQAIDEDTNKAASNVK--YVMGRNDGGYLMIDSKTAEIK  396
              T  V +      LV  ++         ++K   NV+  +V G    G  +I+  T  I+
Sbjct  898   YTPNVDENAGPNTLVTTVVA--------SDKDGDNVRFGFVGGGTSSGQFVIEEITGVIR  949

Query  397   FVK---NMNRDSTFI-VNKTITAEVLAIDEYTGKTSTGTVYVRVPDFNDN------CPTA  446
                   +++RD   + V            + T  TST  V V + D NDN      C T
Sbjct  950   LHSKAISLDRDKYELNVTAMDDGSCCVNGDATIHTSTAVVVVFITDVNDNKPVFKDCGTY  1009

Query  447   VLE-KDAVCSSSPSVVVSARTLNNRYTGPYTFALEDQP  483
               + ++   + SP + V A   +    G   +++  QP
Sbjct  1010  YPKVEEGAPNGSPVIKVQATDEDKGVNGQVKYSIVQQP  1047

 Score = 44.7 bits (104),  Expect = 0.008, Method: Compositional matrix adjust.
 Identities = 48/199 (24%), Positives = 73/199 (37%), Gaps = 19/199 (10%)

Query  27    DYQATQKITYRISGVGIDQPPFGIFVVDKNTGDINITAIVDREETPSFLITCRALNAQGL  86
             D     K++Y I      Q  F I       G + I   +DRE TP   +   A++
Sbjct  1925  DQGGKSKVSYAIDRSSDRQRQFSI----NQEGTVTIQRNLDREVTPRHQVKILAIDDGIP  1980

Query  87    DVEKPLILTVKILDINDNPPVFSQQIFMGEIEENSASNSLVMILNATDADEPNHLNSKIA  146
                    LTV + DINDNPP F +  +   + E+     +V IL   D D
Sbjct  1981  PKTATATLTVIVQDINDNPPKFLKD-YRPVLPEHVPPRKVVEILATDDDDRSKSNGPPFQ  2039

Query  147   FKIVSQEPAGTPMFLLSRNTGEVRTLTN-----------SLDREQASSYRLVVSGADKDG  195
             F++   +P    +   S    + +   N           S DREQ   Y + +   D
Sbjct  2040  FRL---DPGADDIIRASFKVEQDQKGANGDGMAIVSSLRSFDREQQKEYLIPIVIKDHGN  2096
```

```
Query  196   EGLSTQCECNIKVKDVNDN  214
               ++      + + DVNDN
Sbjct  2097  PAMTGTSTLTVVIGDVNDN  2115

 Score = 40.8 bits (94),  Expect = 0.11, Method: Compositional matrix adjust.
 Identities = 55/242 (23%), Positives = 102/242 (42%), Gaps = 22/242 (9%)

Query  4    KFAKPCREGE--DNSKRNP-IAKITSDYQATQKITYRISGVGIDQPPFGIFVVDKNTGDI  60
            +F  P  E E  +N K++  I  + +   A ++I Y +   G      G F +   +G +
Sbjct  685  QFYMPSYEAEIPENQKKDSDIISVKAKSFADREIRYTLKAQGQGA---GTFNIGPTSGIV  741

Query  61   NITAIVDREE--TPSF--LITCRALNAQGLDVEKPLILTVKILDINDNPPVFSQQIFMGE  116
             +   +D E+   P    L+     ++ G      + LT+++ D+NDN P F    +
Sbjct  742  KLAKELDFEDLRQPHVYSLVVTATEDSGGFSTS--VELTIRVTDVNDNAPKFELPDYQAH  799

Query  117  -IEENSASNSLVMILNATDADEPNHLNSKIAFKIVSQEPAGTPMFLLSRNTGEVRTLTNS  175
             ++E+    + ++ + A DAD  +  N++I + +VS +      F +  N   V
Sbjct  800  NVDEDIPLGTSILKVKAMDAD--SGANAEIEY-LVSDD-----HFAVDSNGIIVNNKQLD  851

Query  176  LDREQASSYRLVVSGADKDGEGLSTQCECNIKVKDVNDNFPMFRDSQYSARIEENILSSE  235
             D   A  Y  VV+  DK     +      +  K+ ND  P F    Y+  ++EN   +
Sbjct  852  ADNNNA-YYEFVVTAKDKGEPAKTGTATVRVYTKNKNDEEPKFSQQVYTPNVDENAGPNT  910

Query  236  LL  237
            L+
Sbjct  911  LV  912

 Score = 39.7 bits (91),  Expect = 0.24, Method: Compositional matrix adjust.
 Identities = 89/389 (23%), Positives = 149/389 (38%), Gaps = 62/389 (16%)

Query  75   LITCRALNAQGLDVEKPLILTVKILDINDNPPVFSQQIF--MGEIEENSASNSLVMILNA  132
            +IT +  NA GL       + + + D+ND PP F  +       ++ N+  N+ V  L A
Sbjct  546  IITNQGHNA-GLKYTDNQRVIIHVKDVNDEPPYFINRPLPMQAVVQLNAPPNTPVFTLQA  604

Query  133  TDADEPNHLNSKIAFKIVSQEPAGTPMFLLSRNTGEVRTLTNSLDREQASSYRLVVSGAD  192
             D D     +  I + IV     G   F +   +G VRT    L  +    Y L V   D
Sbjct  605  RDPD----TDHNIHYFIVRDRTGG--RFEVDERSGVVRTRGTDL-FQLDMEYVLYVKAED  657

Query  193  KDGE----GLSTQCECNIKVKDVNDNFPMFRDSQYSARIEEN-ILSSELLRFQVTDLDEE  247
            ++G+       +  E  + +       P F    Y A I EN    S+++  +      +
Sbjct  658  QNGKVDDRRFQSTPEERLSIVG-GKRAPQFYMPSYEAEIPENQKKDSDIISVKA----KS  712

Query  248  YTDNWLAVYFFTSGNEGNWFEIQTDPRTNEGILKVVKALDYEQLQSVKLS----IAVKNK  303
            + D  +       G     F I        GI+K+ K LD+E L+   +      A ++
Sbjct  713  FADREIRYTLKAQGQGAGTFNIGP----TSGIVKLAKELDFEDLRQPHVYSLVVTATEDS  768

Query  304  AEFHQSVISRYRVQSTPVTIQVINVREGIAFRPASKTFTVQKGISSKKLVDYILGT----  359
              F  SV          +TI+V +V +       +  F +    +     D  LGT
Sbjct  769  GGFSTSV---------ELTIRVTDVNDN------APKFELPDYQAHNVDEDIPLGTSILK  813

Query  360  YQAIDEDTNKAASNVKYVMGRN----DGGYLMIDSKTAEIKFVKNMNRDSTFIVNKTITA  415
             +A+D D+  A + ++Y++  +    D   +++++K  +     N N    F+V
Sbjct  814  VKAMDADSG-ANAEIEYLVSDDHFAVDSNGIIVNNKQLD---ADNNNAYYEFVVTAKDKG  869

Query  416  EVLAIDEYTGKTSTGTVYVRVPDFNDNCP  444
            E         KT T TV V   + ND  P
Sbjct  870  E-------PAKTGTATVRVYTKNKNDEEP  891

>ANILCM1_038625.P14110 | transcript=ANILCM1_038625.R14110 | gene=ANILCM1_038625
| organism=Anopheles_nili_AnilCM1 | gene_product=neural-cadherin,
transcript variant X2 | transcript_product=neural-cadherin,
transcript variant X3 | location=AnilCM1_3RL:7464564-7527461(+)
| protein_length=1716 | sequence_SO=chromosome
| SO=protein_coding_gene | is_pseudo=false
Length=1716

 Score = 144 bits (364),  Expect = 1e-34, Method: Compositional matrix adjust.
```

```
 Identities = 125/442 (28%), Positives = 212/442 (48%), Gaps = 33/442 (7%)

Query  21   IAKITSDYQATQKITYRISGVGID--QPPFGIFVVDKNTGDINITAIVDREET---PSFL  75
            +     D    Q I Y ++G GID   P    F +++ TG+I +   +DR++    P +
Sbjct  160  VTATDGDKDRPQNIVYFLTGQGIDPDNPANSKFDINRTTGEIFVLKPLDRDQPNGRPQWR  219

Query  76   ITCRALNAQGLDVEKPLILTVKILDINDNPPVFSQQIFMGEIEENSASNSLVMILNATDA  135
             T  A +  G  +     + V + DINDN P+F Q ++ G + EN  +  +VM + A D
Sbjct  220  FTVFAQDEGGEGLVGYADVQVNLKDINDNAPIFPQGVYFGNVTENGTAGMVVMTMTAVDY  279

Query  136  DEPNH-LNSKIAFKI---VSQEPAGTPMFLLSRNTGEVRTLTNSLDREQASSYRLVVSGA  191
            D+PN   N+K+ + I   V +E  G+P+F +  +TG ++T    LDRE+   Y + V
Sbjct  280  DDPNEGTNAKLIYSIEKNVIEEETGSPIFEIEADTGVIKTAVCCLDRERTPDYSIQVVAM  339

Query  192  DKDGEGLSTQCECNIKVKDVNDNFPMFRDSQYSARIEENILSS--ELLRFQVTDLDEEYT  249
              DG GL      +I+VKD+ND  P F   ++   ++E   ++  E+    VT  DE+ T
Sbjct  340  --DGGGLKGTGTASIRVKDINDMPPQFTKDEWFTEVDETDGTNLPEMPILTVTVHDEDET  397

Query  250  DNWLAVYFFTSGNEGNWFEIQTDPRTNEGI--LKVVKALDYE---QLQSVKLSIAVKNKA  304
            + +       SG   + F +    R N+G   LK+V+ LDYE   Q    +  I V +K
Sbjct  398  NKFQYKVIDNSGYGADKFTMV---RNNDGTGSLKIVQPLDYEDPLQSNGFRFRIQVNDKG  454

Query  305  EFHQSVISRYRVQSTPVTIQVINVREGIAFRPASKTFTVQKGISSKKLVDYILGTYQAID  364
            E + +   +Y V  + V +++ ++ +    +P  +   ++  +     V   L T++A D
Sbjct  455  EDNDN--DKYHVAYSWVVVKLRDINDN---KPQFERPNIEVSVYENADVGKTLETFKATD  509

Query  365  EDTNKAASNVKYVMGR-NDGGYLMIDSKTAEIKFVKNMNRDSTFIVNKTITAEVLAIDE-  422
             D     S V Y + R +D       ++   +   +N++R+    V      ++LAID+
Sbjct  510  PDQG-GKSKVSYAIDRSSDRQRQFSINQEGTVTIQRNLDRE----VTPRHQVKILAIDDG  564

Query  423  YTGKTSTGTVYYVRVPDFNDNCP  444
               KT+T T+ V V D NDN P
Sbjct  565  IPPKTATATLTVIVQDINDNPP  586

 Score = 81.6 bits (200),  Expect = 3e-14, Method: Compositional matrix adjust.
 Identities = 97/367 (26%), Positives = 157/367 (43%), Gaps = 40/367 (11%)

Query  93   ILTVKILDINDNPPVFSQQIFMGEIEENSASNSLVMILNATDADEPNHLNSKIAFKIVSQ  152
             + + I D NDNPP F + ++  E++EN      V+ + A D DE    +S+I ++I S
Sbjct  21   FIRIGIADKNDNPPYFDKALYEAEVDENEDIQHTVLTVTAKDHDE----SSRIRYEITSG  76

Query  153  EPAGTPMFLLSRNTGEVRTLTNSLDREQASSYRLVVSGADKDGEGLSTQCECNIKVKDVN  212
               G   F +   TG +  +  +LD E    Y L ++ +D   E  +T     I VKDVN
Sbjct  77   NIGGA--FAVKNMTGAI-YVAGALDYETRKRYELRLAASDNLKENYTTVV---IHVKDVN  130

Query  213  DNFPMFRDSQYSARIEENI---LSSELLRFQVTDLDEEYTDNWLAVYFFTSG--NEGNWF  267
            DN P+F    Y  +I E     L   +L+   TD D++   N   VYF T    +  N
Sbjct  131  DNPPVFERPTYRTQITEEDDRNLPKRVLQVTATDGDKDRPQN--IVYFLTGQGIDPDNPA  188

Query  268  EIQTDPRTNEGILKVVKALDYEQLQS-VKLSIAVKNKAEFHQSVISRYRVQSTPVTIQVI  326
              + D     G + V+K LD +Q     +    V  + E  + ++    VQ     + +
Sbjct  189  NSKFDINRTTGEIFVLKPLDRDQPNGRPQWRFTVFAQDEGGEGLVGYADVQ-----VNLK  243

Query  327  NVREGIAFRPASKTFTVQKGISSKKLVDYILGTYQAIDEDTNKAASNVK--YVMGRN---  381
            ++ +     P    F     ++        ++ T  A+D D     +N K  Y + +N
Sbjct  244  DINDNAPIFPQGVYF---GNVTENGTAGMVVMTMTAVDYDDPNEGTNAKLIYSIEKNVIE  300

Query  382  ---DGGYLMIDSKTAEIK-FVKNMNRDSTFIVNKTITAEVLAIDEYTGKTSTGTVYYVRVP  437
                     I++ T  IK  V  ++R+ T       + +V+A+D   G   TGT  +RV
Sbjct  301  EETGSPIFEIEADTGVIKTAVCCLDRERT----PDYSIQVVAMDG-GGLKGTGTASIRVK  355

Query  438  DFNDNCP  444
            D ND  P
Sbjct  356  DINDMPP  362

 Score = 53.5 bits (127),  Expect = 1e-05, Method: Compositional matrix adjust.
 Identities = 70/306 (23%), Positives = 126/306 (41%), Gaps = 30/306 (10%)

Query  20   PIAKIT-SDYQATQKITYRI---SGVGIDQPPFGIFVVDKNTGDINITAIVDRE---ETP  72
```

```
            PI  +T  D   T K  Y++   SG G D+  F +   +  TG + I   +D E   ++
Sbjct  385  PILTVTVHDEDETNKFQYKVIDNSGYGADK--FTMVRNNDGTGSLKIVQPLDYEDPLQSN  442

Query  73   SFLITCRALNAQGLDVEKPLI------LTVKILDINDNPPVFSQQIFMGEIEENSASNSL  126
             F    + +N +G D +           + VK+ DINDN P F +      + EN+
Sbjct  443  GFRFRIQ-VNDKGEDNDNDKYHVAYSWVVVKLRDINDNKPQFERPNIEVSVYENADVGKT  501

Query  127  VMILNATDADEPNHLNSKIAFKIVSQEPAGTPMFLLSRNTGEVRTLTNSLDREQASSYRL  186
            +    ATD D+     SK+++ I   + +       S N     T+  +LDRE    +++
Sbjct  502  LETFKATDPDQGG--KSKVSYAI---DRSSDRQRQFSINQEGTVTIQRNLDREVTPRHQV  556

Query  187  VVSGADKDGEGLSTQCECNIKVKDVNDNFPMFRDSQYSARIEENILSSELLRFQVTDLDE  246
             +   D      +      + V+D+NDN P F    Y   + E++   +++    TD D+
Sbjct  557  KILAIDDGIPPKTATATLTVIVQDINDNPPKFL-KDYRPVLPEHVPPRKVVEILATDDDD  615

Query  247  EYTDNWLAVYFFTSGNEGNW----FEIQTDPRTNEG----ILKVVKALDYEQLQSVKLSI  298
                N     F       +     F+++ D +   G    I+  +++ D EQ +   + I
Sbjct  616  RSKSNGPPFQFRLDPGADDIIRASFKVEQDQKGANGDGMAIVSSLRSFDREQQKEYLIPI  675

Query  299  AVKNKA  304
             +K+
Sbjct  676  VIKDHG  681

 Score = 44.3 bits (103),  Expect = 0.010, Method: Compositional matrix adjust.
 Identities = 48/199 (24%), Positives = 73/199 (37%), Gaps = 19/199 (10%)

Query  27   DYQATQKITYRISGVGIDQPPFGIFVVDKNTGDINITAIVDREETPSFLITCRALNAQGL  86
            D     K++Y I      Q  F I       G + I   +DRE TP   +    A++
Sbjct  511  DQGGKSKVSYAIDRSSDRQRQFSI----NQEGTVTIQRNLDREVTPRHQVKILAIDDGIP  566

Query  87   DVEKPLILTVKILDINDNPPVFSQQIFMGEIEENSASNSLVMILNATDADEPNHLNSKIA  146
                   LTV + DINDNPP F +  +   + E+     +V IL   D D
Sbjct  567  PKTATATLTVIVQDINDNPPKFLKD-YRPVLPEHVPPRKVVEILATDDDDRSKSNGPPFQ  625

Query  147  FKIVSQEPAGTPMFLLSRNTGEVRTLTN-----------SLDREQASSYRLVVSGADKDG  195
            F++   +P    +   S    + +   N           S DREQ   Y + +   D
Sbjct  626  FRL---DPGADDIIRASFKVEQDQKGANGDGMAIVSSLRSFDREQQKEYLIPIVIKDHGN  682

Query  196  EGLSTQCECNIKVKDVNDN  214
              ++      + + DVNDN
Sbjct  683  PAMTGTSTLTVVIGDVNDN  701

>AARA21_009090.P17044 | transcript=AARA21_009090.R17044 | gene=AARA21_009090
| organism=Anopheles_arabiensis_DONGOLA_2021
| gene_product=neural-cadherin, transcript variant X5 |
transcript_product=neural-cadherin,
transcript variant X5 | location=CM029353:40466514-40666848(+)
| protein_length=3105 | sequence_SO=chromosome
| SO=protein_coding_gene | is_pseudo=false
Length=3105

 Score = 144 bits (364),  Expect = 1e-34, Method: Compositional matrix adjust.
 Identities = 125/437 (29%), Positives = 211/437 (48%), Gaps = 33/437 (8%)

Query  26    SDYQATQKITYRISGVGID--QPPFGIFVVDKNTGDINITAIVDREET---PSFLITCRA  80
              D    Q I Y ++G GID   P    F +++ TG+I +   +DR++    P +  T  A
Sbjct  1554  GDKDRPQNIVYFLTGQGIDPDNPANSKFDINRTTGEIFVLKPLDRDQPNGRPQWRFTVFA  1613

Query  81    LNAQGLDVEKPLILTVKILDINDNPPVFSQQIFMGEIEENSASNSLVMILNATDADEPNH  140
              +  G  +     + V + DINDN P+F Q ++ G + EN  +  +VM + A D D+PN
Sbjct  1614  QDEGGEGLVGYADVQVNLKDINDNAPIFPQGVYFGNVTENGTAGMVVMTMTAVDYDDPNE  1673

Query  141   -LNSKIAFKI---VSQEPAGTPMFLLSRNTGEVRTLTNSLDREQASSYRLVVSGADKDGE  196
               N+K+ + I   V +E  G+P+F +  +TG ++T    LDRE+   Y + V     DG
Sbjct  1674  GTNAKLIYSIEKNVIEEETGSPIFEIEADTGVIKTAVCCLDRERTPDYSIQVVAM--DGG  1731

Query  197   GLSTQCECNIKVKDVNDNFPMFRDSQYSARIEENILSS--ELLRFQVTDLDEEYTDNWLA  254
             GL      +I+VKD+ND  P F   ++   ++E   ++  E+    VT  DE+ T+ +
Sbjct  1732  GLKGTGTASIRVKDINDMPPQFTKDEWFTEVDETDGTNLPEMPILTVTVHDEDETNKFQY  1791
```

```
Query  255   VYFFTSGNEGNWFEIQTDPRTNEGI--LKVVKALDYE---QLQSVKLSIAVKNKAEFHQS  309
                  SG   + F +    R N+G   LK+V+ LDYE   Q    +  I V +K E + +
Sbjct  1792  KVIDNSGYGADKFTMV---RNNDGTGSLKIVQPLDYEDPLQSNGFRFRIQVNDKGEDNDN  1848

Query  310   VISRYRVQSTPVTIQVINVREGIAFRPASKTFTVQKGISSKKLVDYILGTYQAIDEDTNK  369
                +Y V  + V +++ ++ +    +P  +   ++  +     V   L T++A D D
Sbjct  1849  --DKYHVAYSWVVVKLRDINDN---KPQFERPNIEVSVYENADVGKTLETFKATDPDQG-  1902

Query  370   AASNVKYVMGR-NDGGYLMIDSKTAEIKFVKNMNRDSTFIVNKTITAEVLAIDE-YTGKT  427
               S V Y + R +D       ++   +   +N++R+    V      ++LAID+    KT
Sbjct  1903  GKSKVSYAIDRSSDRQRQFSINQEGTVTIQRNLDRE----VTPRHQVKILAIDDGIPPKT  1958

Query  428   STGTVYVRVPDFNDNCP  444
             +T T+ V V D NDN P
Sbjct  1959  ATATLTVIVQDINDNPP  1975

 Score = 85.9 bits (211),  Expect = 1e-15, Method: Compositional matrix adjust.
 Identities = 108/420 (26%), Positives = 177/420 (42%), Gaps = 52/420 (12%)

Query  50    IFVVDKNTGDINITAIVDREETPSFLITCRAL----------NAQGLDVEKPLILTVKIL  99
              F ++  +G+I    + DRE+  ++ +   A           N Q   V K   + + I
Sbjct  1359  FFEINLQSGEIFTKVVFDREKQGAYALEVEARDGAPSARPNSNQQPNSVTK--FIRIGIA  1416

Query  100   DINDNPPVFSQQIFMGEIEENSASNSLVMILNATDADEPNHLNSKIAFKIVSQEPAGTPM  159
             D NDNPP F + ++  E++EN      V+ + A D DE    +S+I ++I S    G
Sbjct  1417  DKNDNPPYFDKALYEAEVDENEDIQHTVLTVTAKDHDE----SSRIRYEITSGNIGGA--  1470

Query  160   FLLSRNTGEVRTLTNSLDREQASSYRLVVSGADKDGEGLSTQCECNIKVKDVNDNFPMFR  219
             F +   TG +  +  +LD E    Y L ++ +D   E  +T     I VKDVNDN P+F
Sbjct  1471  FAVKNMTGAI-YVAGALDYETRKRYELRLAASDNLKENYTTVV---IHVKDVNDNPPVFE  1526

Query  220   DSQYSARIEENI---LSSELLRFQVTDLDEEYTDNWLAVYFFTSG--NEGNWFEIQTDPR  274
                Y  +I E     L   +L+   TD D++   N   VYF T    +  N    + D
Sbjct  1527  RPTYRTQITEEDDRNLPKRVLQVTATDGDKDRPQN--IVYFLTGQGIDPDNPANSKFDIN  1584

Query  275   TNEGILKVVKALDYEQLQS-VKLSIAVKNKAEFHQSVISRYRVQSTPVTIQVINVREGIA  333
                G + V+K LD +Q     +    V  + E  + ++    VQ     + + ++ +
Sbjct  1585  RTTGEIFVLKPLDRDQPNGRPQWRFTVFAQDEGGEGLVGYADVQ-----VNLKDINDNAP  1639

Query  334   FRPASKTFTVQKGISSKKLVDYILGTYQAIDEDTNKAASNVK--YVMGRN------DGGY  385
               P    F     ++       ++ T  A+D D     +N K  Y + +N
Sbjct  1640  IFPQGVYF---GNVTENGTAGMVVMTMTAVDYDDPNEGTNAKLIYSIEKNVIEEETGSPI  1696

Query  386   LMIDSKTAEIK-FVKNMNRDSTFIVNKTITAEVLAIDEYTGKTSTGTVYVRVPDFNDNCP  444
               I++ T  IK  V  ++R+ T       + +V+A+D   G   TGT  +RV D ND  P
Sbjct  1697  FEIEADTGVIKTAVCCLDRERT----PDYSIQVVAMDG-GGLKGTGTASIRVKDINDMPP  1751

 Score = 76.3 bits (186),  Expect = 1e-12, Method: Compositional matrix adjust.
 Identities = 106/426 (25%), Positives = 181/426 (42%), Gaps = 51/426 (12%)

Query  53    VDKNTGDINITAIVDREETPSFLITCRALNAQGLDVEKPLILTVKILDINDNPPVFSQ-Q  111
             +D++  ++N+TA+ D         +C  +N          ++ V I D+NDN PVF
Sbjct  928   LDRDKYELNVTAMDDG--------SC-CVNGDATIHTSTAVVVVFITDVNDNKPVFKDCG  978

Query  112   IFMGEIEENSASNSLVMILNATDADEPNHLNSKIAFKIVSQEPAGTPMFLLSRNTGEVRT  171
              +  ++EE + + S V+ + ATD D+   +N ++ + IV Q       F +   TGEV T
Sbjct  979   TYYPKVEEGAPNGSPVIKVQATDEDK--GVNGQVKYSIVQQPNQKGTKFTVDEETGEVST  1036

Query  172   LTNSLDRE--QASSYRLVVSGADKDGEGLSTQCECNIKVKDVNDNFPMFRDSQYSARIEE  229
                  DRE        + V   D+    L   C   +++ DVNDN P+F   +Y   +++
Sbjct  1037  -NKVFDREGDDGKFVSVTVKATDQGEPSLEGVCSFTVEITDVNDNPPLFDRQKYVENVKQ  1095

Query  230   NI-LSSELLRFQVTDLDEEYTDNWLAVYFFT---SGNEGNWFEIQTDPRTNEGILKVVKA  285
             +  + + +LR  V+  DE+  +N   VY  +   S N+  +FEIQ  P +   +LK  K
Sbjct  1096  DASIGTNILR--VSASDEDADNNGAIVYTLSAPHSPNDLEYFEIQ--PESGWIVLK--KP  1149

Query  286   LD--YEQLQSVKLSIAVKNK------AEFHQSVISRYRVQSTPVTIQV----INVREGIA  333
             LD  Y + +  +L +   ++      A+    +    R    P+  +     I++RE +
```

```
Sbjct  1150  LDGCYSR-ERYRLKVYASDRGDPPSTADVDELDVVDRNNKPPIWDRTIYGPIHIRENVT  1208

Query  334   FRPASKTFTVQKGISSKKLVDYILGTYQAIDEDTNKAASNVKYVMGRNDGGYLMIDSKTA  393
                   +     GI     V Y L         TNK   +  Y+  R D G        A
Sbjct  1209  VGTVVTSVKASSGIEGNPTVFYRL--MPGSTAQTNKL--HTFYLQQRPDNG-----DTWA  1259

Query  394   EIKFVKNMNRDSTFIVNKTITAEVLAIDEYTGKTSTGTVYYRVPDFNDNCPTAV-LEKDA  452
             +IK    ++ ++    N T+  E     +     S  TVY+++ D ND  P     E++
Sbjct  1260  DIKVNHPLDFETIKEYNLTVRVENNGAQQL---ASEATVYIQLEDVNDEIPLFTEREQET  1316

Query  453   VCSSSP  458
             V    P
Sbjct  1317  VLEGEP  1322

 Score = 60.8 bits (146),  Expect = 9e-08, Method: Compositional matrix adjust.
 Identities = 83/362 (23%), Positives = 152/362 (42%), Gaps = 34/362 (9%)

Query  51    FVVDKNTGDINITAIVDRE-ETPSFL-ITCRALNAQGLDVEKPLILTVKILDINDNPPVF  108
             F VD+ TG+++   + DRE +   F+ +T +A +     +E     TV+I D+NDNPP+F
Sbjct  1025  FTVDEETGEVSTNKVFDREGDDGKFVSVTVKATDQGEPSLEGVCSFTVEITDVNDNPPLF  1084

Query  109   SQQIFMGEIEENSASNSLVMILNATDADEPNHLNSKIAFKIVS-QEPAGTPMFLLSRNTG  167
              +Q ++  ++++++  + ++ ++A+D D  N  N  I + + +   P     F +   +G
Sbjct  1085  DRQKYVENVKQDASIGTNILRVSASDEDADN--NGAIVYTLSAPHSPNDLEYFEIQPESG  1142

Query  168   EV---RTLTNSLDREQASSYRLVVSGADKDGEGLSTQCECNIKVKDVNDNFPMFRDSQYS  224
              +   + L     RE+   YRL V  +D+ G+  ST            +N P   D
Sbjct  1143  WIVLKKPLDGCYSRER---YRLKVYASDR-GDPPSTADVDVELDVVDRNNKPPIWDRTIY  1198

Query  225   ARIEENILSSELLRFQVTDLDEEYTDNWLAVYFFTSG-----NEGNWFEIQTDPRTNE--  277
               I      +                N    Y    G     N+ + F +Q  P   +
Sbjct  1199  GPIHIRENVTVGTVVTSVKASSGIEGNPTVFYRLMPGSTAQTNKLHTFYLQQRPDNGDTW  1258

Query  278   GILKVVKALDYEQLQSVKLSIAVKNKAEFHQSVISRYRVQSTPVTIQVINVREGIAFRPA  337
               +KV   LD+E ++   L++ V+N         ++       V IQ+ +V + I
Sbjct  1259  ADIKVNHPLDFETIKEYNLTVRVENNG-------AQQLASEATVYIQLEDVNDEIPLFTE  1311

Query  338   SKTFTVQKGISSKKLVDYILGTYQAIDEDTNKAASNVKYVM---GRNDG-GYLMIDSKTA  393
              +  TV +G      V  +     AID+D     + V Y +    RN+G  +  I+ ++
Sbjct  1312  REQETVLEGEPIGTKVTQV----NAIDKDGTFPNNQVYYYIVDSPRNEGKDFFEINLQSG  1367

Query  394   EI  395
             EI
Sbjct  1368  EI  1369

 Score = 55.8 bits (133),  Expect = 3e-06, Method: Compositional matrix adjust.
 Identities = 49/158 (31%), Positives = 78/158 (49%), Gaps = 24/158 (15%)

Query  102   NDNPPVFSQQIFMGEIEENSASNSLVMILNATDADEPNHLNSKIAFKIVSQEPAGTP--M  159
             ND  P FSQQ++   ++EN+  N+LV  + A+D D  N     + F  V     GT
Sbjct  858   NDEEPKFSQQVYTPNVDENAGPNTLVTTVVASDKDGDN-----VRFGFVG---GGTSSGQ  909

Query  160   FLLSRNTGEVRTLTN--SLDREQASSYRLVVSGADK-------DGEGLSTQCECNIKVKD  210
             F++   TG +R  +   SLDR++   Y L V+  D        D    ++     + + D
Sbjct  910   FVIEEITGVIRLHSKAISLDRDK---YELNVTAMDDGSCCVNGDATIHTSTAVVVVFITD  966

Query  211   VNDNFPMFRD-SQYSARIEENILS-SELLRFQVTDLDE  246
             VNDN P+F+D   Y  ++EE   + S +++ Q TD D+
Sbjct  967   VNDNKPVFKDCGTYYPKVEEGAPNGSPVIKVQATDEDK  1004

 Score = 53.9 bits (128),  Expect = 1e-05, Method: Compositional matrix adjust.
 Identities = 70/306 (23%), Positives = 126/306 (41%), Gaps = 30/306 (10%)

Query  20    PIAKIT-SDYQATQKITYRI---SGVGIDQPPFGIFVVDKNTGDINITAIVDRE---ETP  72
             PI  +T  D   T K  Y++   SG G D+  F +   +  TG + I   +D E   ++
Sbjct  1774  PILTVTVHDEDETNKFQYKVIDNSGYGADK--FTMVRNNDGTGSLKIVQPLDYEDPLQSN  1831

Query  73    SFLITCRALNAQGLDVEKPLI------LTVKILDINDNPPVFSQQIFMGEIEENSASNSL  126
```

```
              F    + +N +G D +          + VK+ DIDN  P F +       + EN+
Sbjct  1832  GFRFRIQ-VNDKGEDNDNDKYHVAYSWVVVKLRDINDNKPQFERPNIEVSVYENADVGKT  1890

Query  127   VMILNATDADEPNHLNSKIAFKIVSQEPAGTPMFLLSRNTGEVRTLTNSLDREQASSYRL  186
             +    ATD D+     SK+++ I    + +        S N      T+  +LDRE    +++
Sbjct  1891  LETFKATDPDQGG--KSKVSYAI---DRSSDRQRQFSINQEGTVTIQRNLDREVTPRHQV  1945

Query  187   VVSGADKDGEGLSTQCECNIKVKDVNDNFPMFRDSQYSARIEENILSSELLRFQVTDLDE  246
              +   D      +       + V+D+NDN P F    Y    + E++    +++     TD D+
Sbjct  1946  KILAIDDGIPPKTATATLTVIVQDINDNPPKFL-KDYRPVLPEHVPPRKVVEILATDDDD  2004

Query  247   EYTDNWLAVYFFTSGNEGNW----FEIQTDPRTNEG----ILKVVKALDYEQLQSVKLSI  298
                 N     F       +     F+++ D +    G     I+  +++ D EQ +    + I
Sbjct  2005  RSKSNGPPFQFRLDPGADDIIRASFKVEQDQKGANGDGMAIVSSLRSFDREQQKEYLIPI  2064

Query  299   AVKNKA  304
              +K+
Sbjct  2065  VIKDHG  2070

 Score = 52.4 bits (124),  Expect = 4e-05, Method: Compositional matrix adjust.
 Identities = 97/398 (24%), Positives = 163/398 (41%), Gaps = 54/398 (14%)

Query  106   PVFSQQIFMGEIEENSASNSLVMILNATDADEPNHLNSKIAFKIVSQ-EPAGTPMFLLSR  164
             P F    +  EI EN   +S ++ + A       +  + +I + + +Q + AGT  F +
Sbjct  655   PQFYMPSYEAEIPENQKKDSDIISVKAK-----SFADREIRYTLKAQGQGAGT--FNIGP  707

Query  165   NTGEVRTLTNSLDRE---QASSYRLVVSGADKDGEGLSTQCECNIKVKDVNDNFPMFRDS  221
              +G V+ L   LD E   Q   Y LVV+ A +D  G ST  E  I+V DVNDN P F
Sbjct  708   TSGIVK-LAKELDFEDLRQPHVYSLVVT-ATEDSGGFSTSVELTIRVTDVNDNAPKFELP  765

Query  222   QYSAR-IEENI-LSSELLRFQVTDLDEEYTDNWLAVYFFTSGNEGNWFEIQTDPRTNEGI  279
              Y A  ++E+I L + +L+ +  D D          + +  S +    F + ++     GI
Sbjct  766   DYQAHNVDEDIPLGTSILKVKAMDSDSGAN---AEIEYLVSDDH---FAVDSN-----GI  814

Query  280   LKVVKALDYEQLQS-VKLSIAVKNKAEFHQSVISRYRVQSTPVTIQVINVREGIAFRPAS  338
             +   K LD +   +  +  +  K+K E  ++  +  RV +             E  F
Sbjct  815   IVNNKQLDADNNNAYYEFVVTAKDKGEPAKTGTATVRVYTKNKN------DEEPKFSQQV  868

Query  339   KTFTVQKGISSKKLVDYILGTYQAIDEDTNKAASNVK--YVMGRNDGGYLMIDSKTAEIK  396
              T  V +       LV  ++          ++K   NV+  +V G    G  +I+  T  I+
Sbjct  869   YTPNVDENAGPNTLVTTVVA--------SDKDGDNVRFGFVGGGTSSGQFVIEEITGVIR  920

Query  397   FVK---NMNRDSTFI-VNKTITAEVLAIDEYTGKTSTGTVYVRVPDFNDN------CPTA  446
                   +++RD   + V              + T  TST  V V + D NDN      C T
Sbjct  921   LHSKAISLDRDKYELNVTAMDDGSCCVNGDATIHTSTAVVVVFITDVNDNKPVFKDCGTY  980

Query  447   VLE-KDAVCSSSPSVVVSARTLNNRYTGPYTFALEDQP  483
               + ++    + SP + V A    +    G    +++  QP
Sbjct  981   YPKVEEGAPNGSPVIKVQATDEDKGVNGQVKYSIVQQP  1018

 Score = 44.7 bits (104),  Expect = 0.008, Method: Compositional matrix adjust.
 Identities = 48/199 (24%), Positives = 73/199 (37%), Gaps = 19/199 (10%)

Query  27    DYQATQKITYRISGVGIDQPPFGIFVVDKNTGDINITAIVDREETPSFLITCRALNAQGL  86
             D      K++Y I       Q  F I        G + I   +DRE TP    +    A++
Sbjct  1900  DQGGKSKVSYAIDRSSDRQRQFSI----NQEGTVTIQRNLDREVTPRHQVKILAIDDGIP  1955

Query  87    DVEKPLILTVKILDINDNPPVFSQQIFMGEIEENSASNSLVMILNATDADEPNHLNSKIA  146
                    LTV + DINDNPP F +  +    + E+      +V IL    D D
Sbjct  1956  PKTATATLTVIVQDINDNPPKFLKD-YRPVLPEHVPPRKVVEILATDDDDRSKSNGPPFQ  2014

Query  147   FKIVSQEPAGTPMFLLSRNTGEVRTLTN-----------SLDREQASSYRLVVSGADKDG  195
             F++    +P     +    S     + +    N                S DREQ    Y + +    D
Sbjct  2015  FRL---DPGADDIIRASFKVEQDQKGANGDGMAIVSSLRSFDREQQKEYLIPIVIKDHGN  2071

Query  196   EGLSTQCECNIKVKDVNDN  214
               ++        + + DVNDN
Sbjct  2072  PAMTGTSTLTVVIGDVNDN  2090
```

```
 Score = 40.0 bits (92),  Expect = 0.21, Method: Compositional matrix adjust.
 Identities = 89/389 (23%), Positives = 149/389 (38%), Gaps = 62/389 (16%)

Query  75   LITCRALNAQGLDVEKPLILTVKILDINDNPPVFSQQIF--MGEIEENSASNSLVMILNA  132
            +IT +  NA GL       + + + D+ND PP F  +       ++ N+  N+ V  L A
Sbjct  517  IITNQGHNA-GLKYTDNQRVIIHVKDVNDEPPYFINRPLPMQAVVQLNAPPNTPVFTLQA  575

Query  133  TDADEPNHLNSKIAFKIVSQEPAGTPMFLLSRNTGEVRTLTNSLDREQASSYRLVVSGAD  192
             D D     +  I + IV     G   F +   +G VRT    L  +    Y L V   D
Sbjct  576  RDPD----TDHNIHYFIVRDRTGG--RFEVDERSGVVRTRGTDL-FQLDMEYVLYVKAED  628

Query  193  KDGE----GLSTQCECNIKVKDVNDNFPMFRDSQYSARIEEN-ILSSELLRFQVTDLDEE  247
            ++G+        +  E  + +       P F    Y A I EN    S+++  +     +
Sbjct  629  QNGKVDDRRFQSTPEERLSIVG-GKRAPQFYMPSYEAEIPENQKKDSDIISVKA----KS  683

Query  248  YTDNWLAVYFFTSGNEGNWFEIQTDPRTNEGILKVVKALDYEQLQSVKLS----IAVKNK  303
            + D  +       G     F I         GI+K+ K LD+E L+   +      A ++
Sbjct  684  FADREIRYTLKAQGQGAGTFNIGP----TSGIVKLAKELDFEDLRQPHVYSLVVTATEDS  739

Query  304  AEFHQSVISRYRVQSTPVTIQVINVREGIAFRPASKTFTVQKGISSKKLVDYILGT----  359
              F  SV          +TI+V +V +       +  F +    +     D  LGT
Sbjct  740  GGFSTSV---------ELTIRVTDVNDN------APKFELPDYQAHNVDEDIPLGTSILK  784

Query  360  YQAIDEDTNKAASNVKYVMGRN----DGGYLMIDSKTAEIKFVKNMNRDSTFIVNKTITA  415
             +A+D D+  A + ++Y++  +    D   +++++K  +     N N    F+V
Sbjct  785  VKAMDSDSG-ANAEIEYLVSDDHFAVDSNGIIVNNKQLD---ADNNNAYYEFVVTAKDKG  840

Query  416  EVLAIDEYTGKTSTGTVYVRVPDFNDNCP  444
            E         KT T TV V   + ND  P
Sbjct  841  E-------PAKTGTATVRVYTKNKNDEEP  862

 Score = 39.7 bits (91),  Expect = 0.25, Method: Compositional matrix adjust.
 Identities = 54/242 (22%), Positives = 102/242 (42%), Gaps = 22/242 (9%)

Query  4    KFAKPCREGE--DNSKRNP-IAKITSDYQATQKITYRISGVGIDQPPFGIFVVDKNTGDI  60
            +F  P  E E  +N K++  I  + +  A ++I Y +   G      G F +   +G +
Sbjct  656  QFYMPSYEAEIPENQKKDSDIISVKAKSFADREIRYTLKAQGQGA---GTFNIGPTSGIV  712

Query  61   NITAIVDREE--TPSF--LITCRALNAQGLDVEKPLILTVKILDINDNPPVFSQQIFMGE  116
             +   +D E+   P    L+     ++ G       + LT+++ D+NDN P F    +
Sbjct  713  KLAKELDFEDLRQPHVYSLVVTATEDSGGFSTS--VELTIRVTDVNDNAPKFELPDYQAH  770

Query  117  -IEENSASNSLVMILNATDADEPNHLNSKIAFKIVSQEPAGTPMFLLSRNTGEVRTLTNS  175
             ++E+    + ++ + A D+D  +  N++I + +VS +      F +  N   V
Sbjct  771  NVDEDIPLGTSILKVKAMDSD--SGANAEIEY-LVSDD-----HFAVDSNGIIVNNKQLD  822

Query  176  LDREQASSYRLVVSGADKDGEGLSTQCECNIKVKDVNDNFPMFRDSQYSARIEENILSSE  235
             D   A  Y  VV+  DK     +      +  K+ ND  P F    Y+  ++EN   +
Sbjct  823  ADNNNA-YYEFVVTAKDKGEPAKTGTATVRVYTKNKNDEEPKFSQQVYTPNVDENAGPNT  881

Query  236  LL  237
            L+
Sbjct  882  LV  883

>AARA21_009090.P17042 | transcript=AARA21_009090.R17042 | gene=AARA21_009090
| organism=Anopheles_arabiensis_DONGOLA_2021
| gene_product=neural-cadherin, transcript variant X5 |
transcript_product=neural-cadherin,
transcript variant X2 | location=CM029353:40466514-40666848(+)
| protein_length=3105 | sequence_SO=chromosome
| SO=protein_coding_gene | is_pseudo=false
Length=3105

 Score = 144 bits (364),  Expect = 1e-34, Method: Compositional matrix adjust.
 Identities = 125/437 (29%), Positives = 211/437 (48%), Gaps = 33/437 (8%)

Query  26    SDYQATQKITYRISGVGID--QPPFGIFVVDKNTGDINITAIVDREET---PSFLITCRA  80
              D    Q I Y ++G GID   P    F +++ TG+I +   +DR++     P +  T  A
```

```
Sbjct  1554  GDKDRPQNIVYFLTGQGIDPDNPANSKFDINRTTGEIFVLKPLDRDQPNGRPQWRFTVFA  1613

Query  81    LNAQGLDVEKPLILTVKILDINDNPPVFSQQIFMGEIEENSASNSLVMILNATDADEPNH  140
              +  G  +     + V + DINDN P+F Q ++ G + EN  +  +VM + A D D+PN
Sbjct  1614  QDEGGEGLVGYADVQVNLKDINDNAPIFPQGVYFGNVTENGTAGMVVMTMTAVDYDDPNE  1673

Query  141   -LNSKIAFKI---VSQEPAGTPMFLLSRNTGEVRTLTNSLDREQASSYRLVVSGADKDGE  196
               N+K+ + I   V +E  G+P+F +  +TG ++T    LDRE+   Y + V     DG
Sbjct  1674  GTNAKLIYSIEKNVIEEETGSPIFEIEADTGVIKTAVCCLDERTPDYSIQVVAM--DGG  1731

Query  197   GLSTQCECNIKVKDVNDNFPMFRDSQYSARIEENILSS--ELLRFQVTDLDEEYTDNWLA  254
             GL      +I+VKD+ND  P F   ++   ++E   ++  E+    VT  DE+ T+ +
Sbjct  1732  GLKGTGTASIRVKDINDMPPQFTKDEWFTEVDETDGTNLPEMPILTVTVHDEDETNKFQY  1791

Query  255   VYFFTSGNEGNWFEIQTDPRTNEGI--LKVVKALDYE---QLQSVKLSIAVKNKAEFHQS  309
                  SG   + F +    R N+G   LK+V+ LDYE   Q    +  I V +K E + +
Sbjct  1792  KVIDNSGYGADKFTMV---RNNDGTGSLKIVQPLDYEDPLQSNGFRFRIQVNDKGEDNDN  1848

Query  310   VISRYRVQSTPVTIQVINVREGIAFRPASKTFTVQKGISSKKLVDYILGTYQAIDEDTNK  369
                +Y V  + V +++ ++ +    +P  +   ++  +     V   L T++A D D
Sbjct  1849  --DKYHVAYSWVVVKLRDINDN---KPQFERPNIEVSVYENADVGKTLETFKATDPDQG-  1902

Query  370   AASNVKYVMGR-NDGGYLMIDSKTAEIKFVKNMNRDSTFIVNKTITAEVLAIDE-YTGKT  427
               S V Y + R +D       ++   +   +N++R+    V      ++LAID+    KT
Sbjct  1903  GKSKVSYAIDRSSDRQRQFSINQEGTVTIQRNLDRE----VTPRHQVKILAIDDGIPPKT  1958

Query  428   STGTVYVRVPDFNDNCP  444
             +T T+ V V D NDN P
Sbjct  1959  ATATLTVIVQDINDNPP  1975

 Score = 85.9 bits (211),  Expect = 1e-15, Method: Compositional matrix adjust.
 Identities = 108/420 (26%), Positives = 177/420 (42%), Gaps = 52/420 (12%)

Query  50    IFVVDKNTGDINITAIVDREETPSFLITCRAL----------NAQGLDVEKPLILTVKIL  99
              F ++  +G+I    + DRE+  ++ +   A           N Q   V K   + + I
Sbjct  1359  FFEINLQSGEIFTKVVFDREKQGAYALEVEARDGAPSARPNSNQQPNSVTK--FIRIGIA  1416

Query  100   DINDNPPVFSQQIFMGEIEENSASNSLVMILNATDADEPNHLNSKIAFKIVSQEPAGTPM  159
             D NDNPP F + ++  E++EN      V+ + A D DE    +S+I ++I S    G
Sbjct  1417  DKNDNPPYFDKALYEAEVDENEDIQHTVLTVTAKDHDE----SSRIRYEITSGNIGGA--  1470

Query  160   FLLSRNTGEVRTLTNSLDREQASSYRLVVSGADKDGEGLSTQCECNIKVKDVNDNFPMFR  219
             F +   TG +  +  +LD E    Y L ++ +D   E  +T     I VKDVNDN P+F
Sbjct  1471  FAVKNMTGAI-YVAGALDYETRKRYELRLAASDNLKENYTTVV---IHVKDVNDNPPVFE  1526

Query  220   DSQYSARIEENI---LSSELLRFQVTDLDEEYTDNWLAVYFFTSG--NEGNWFEIQTDPR  274
                Y  +I E     L   +L+   TD D++   N   VYF T    +  N    + D
Sbjct  1527  RPTYRTQITEEDDRNLPKRVLQVTATDGDKDRPQN--IVYFLTGQGIDPDNPANSKFDIN  1584

Query  275   TNEGILKVVKALDYEQLQS-VKLSIAVKNKAEFHQSVISRYRVQSTPVTIQVINVREGIA  333
                G + V+K LD +Q     +    V  + E  + ++    VQ     + + ++ +
Sbjct  1585  RTTGEIFVLKPLDRDQPNGRPQWRFTVFAQDEGGEGLVGYADVQ-----VNLKDINDNAP  1639

Query  334   FRPASKTFTVQKGISSKKLVDYILGTYQAIDEDTNKAASNVK--YVMGRN------DGGY  385
               P    F     ++       ++ T  A+D D     +N K  Y + +N
Sbjct  1640  IFPQGVYF---GNVTENGTAGMVVMTMTAVDYDDPNEGTNAKLIYSIEKNVIEEETGSPI  1696

Query  386   LMIDSKTAEIK-FVKNMNRDSTFIVNKTITAEVLAIDEYTGKTSTGTVYVRVPDFNDNCP  444
               I++ T  IK  V  ++R+ T       + +V+A+D   G   TGT  +RV D ND  P
Sbjct  1697  FEIEADTGVIKTAVCCLDERT----PDYSIQVVAMDG-GGLKGTGTASIRVKDINDMPP  1751

 Score = 76.3 bits (186),  Expect = 1e-12, Method: Compositional matrix adjust.
 Identities = 106/426 (25%), Positives = 181/426 (42%), Gaps = 51/426 (12%)

Query  53    VDKNTGDINITAIVDREETPSFLITCRALNAQGLDVEKPLILTVKILDINDNPPVFSQ-Q  111
             +D++  ++N+TA+ D         +C  +N          ++ V I D+NDN PVF
Sbjct  928   LDRDKYELNVTAMDDG--------SC-CVNGDATIHTSTAVVVVFITDVNDNKPVFKDCG  978

Query  112   IFMGEIEENSASNSLVMILNATDADEPNHLNSKIAFKIVSQEPAGTPMFLLSRNTGEVRT  171
```

```
              +  ++EE + + S V+ + ATD D+   +N ++ + IV Q       F +   TGEV T
Sbjct  979   TYYPKVEEGAPNGSPVIKVQATDEDK--GVNGQVKYSIVQQPNQKGTKFTVDEETGEVST  1036

Query  172   LTNSLDRE--QASSYRLVVSGADKDGEGLSTQCECNIKVKDVNDNFPMFRDSQYSARIEE  229
                  DRE        + V   D+    L   C   +++ DVNDN P+F   +Y    +++
Sbjct  1037  -NKVFDREGDDGKFVSVTVKATDQGEPSLEGVCSFTVEITDVNDNPPLFDRQKYVENVKQ  1095

Query  230   NI-LSSELLRFQVTDLDEEYTDNWLAVYFFT---SGNEGNWFEIQTDPRTNEGILKVVKA  285
             +  + + +LR  V+  DE+  +N   VY  +   S N+  +FEIQ  P +   +LK  K
Sbjct  1096  DASIGTNILR--VSASDEDADNNGAIVYTLSAPHSPNDLEYFEIQ--PESGWIVLK--KP  1149

Query  286   LD--YEQLQSVKLSIAVKNK------AEFHQSVISRYRVQSTPVTIQV----INVREGIA  333
             LD  Y + +  +L +   ++      A+    +    R    P+  +     I++RE +
Sbjct  1150  LDGCYSR-ERYRLKVYASDRGDPPSTADVDVELDVVDRNNKPPIWDRTIYGPIHIRENVT  1208

Query  334   FRPASKTFTVQKGISSKKLVDYILGTYQAIDEDTNKAASNVKYVMGRNDGGYLMIDSKTA  393
                   +     GI     V Y L         TNK   +  Y+  R D G        A
Sbjct  1209  VGTVVTSVKASSGIEGNPTVFYRL--MPGSTAQTNKL--HTFYLQQRPDNG-----DTWA  1259

Query  394   EIKFVKNMNRDSTFIVNKTITAEVLAIDEYTGKTSTGTVYVRVPDFNDNCPTAV-LEKDA  452
             +IK    ++ ++    N T+  E     +     S  TVY+++ D ND  P     E++
Sbjct  1260  DIKVNHPLDFETIKEYNLTVRVENNGAQQL---ASEATVYIQLEDVNDEIPLFTEREQET  1316

Query  453   VCSSSP  458
             V    P
Sbjct  1317  VLEGEP  1322

 Score = 60.8 bits (146),  Expect = 9e-08, Method: Compositional matrix adjust.
 Identities = 83/362 (23%), Positives = 152/362 (42%), Gaps = 34/362 (9%)

Query  51    FVVDKNTGDINITAIVDRE-ETPSFL-ITCRALNAQGLDVEKPLILTVKILDINDNPPVF  108
             F VD+ TG+++   + DRE +   F+ +T +A +     +E     TV+I D+NDNPP+F
Sbjct  1025  FTVDEETGEVSTNKVFDREGDDGKFVSVTVKATDQGEPSLEGVCSFTVEITDVNDNPPLF  1084

Query  109   SQQIFMGEIEENSASNSLVMILNATDADEPNHLNSKIAFKIVS-QEPAGTPMFLLSRNTG  167
              +Q ++  ++++++  + ++ ++A+D D  N  N  I + + +   P     F +   +G
Sbjct  1085  DRQKYVENVKQDASIGTNILRVSASDEDADN--NGAIVYTLSAPHSPNDLEYFEIQPESG  1142

Query  168   EV---RTLTNSLDREQASSYRLVVSGADKDGEGLSTQCECNIKVKDVNDNFPMFRDSQYS  224
              +   + L     RE+   YRL V  +D+ G+  ST            +N P   D
Sbjct  1143  WIVLKKPLDGCYSRER---YRLKVYASDR-GDPPSTADVDVELDVVDRNNKPPIWDRTIY  1198

Query  225   ARIEENILSSELLRFQVTDLDEEYTDNWLAVYFFTSG-----NEGNWFEIQTDPRTNE--  277
               I      +                N    Y    G     N+ + F +Q  P   +
Sbjct  1199  GPIHIRENVTVGTVVTSVKASSGIEGNPTVFYRLMPGSTAQTNKLHTFYLQQRPDNGDTW  1258

Query  278   GILKVVKALDYEQLQSVKLSIAVKNKAEFHQSVISRYRVQSTPVTIQVINVREGIAFRPA  337
               +KV   LD+E ++   L++ V+N         ++       V IQ+ +V + I
Sbjct  1259  ADIKVNHPLDFETIKEYNLTVRVENNG-------AQQLASEATVYIQLEDVNDEIPLFTE  1311

Query  338   SKTFTVQKGISSKKLVDYILGTYQAIDEDTNKAASNVKYVM---GRNDG-GYLMIDSKTA  393
              +  TV +G      V  +     AID+D     + V Y +    RN+G  +  I+ ++
Sbjct  1312  REQETVLEGEPIGTKVTQV----NAIDKDGTFPNNQVYYYIVDSPRNEGKDFFEINLQSG  1367

Query  394   EI  395
             EI
Sbjct  1368  EI  1369

 Score = 55.8 bits (133),  Expect = 3e-06, Method: Compositional matrix adjust.
 Identities = 49/158 (31%), Positives = 78/158 (49%), Gaps = 24/158 (15%)

Query  102   NDNPPVFSQQIFMGEIEENSASNSLVMILNATDADEPNHLNSKIAFKIVSQEPAGTP--M  159
             ND  P FSQQ++   ++EN+  N+LV  + A+D D  N     + F  V     GT
Sbjct  858   NDEEPKFSQQVYTPNVDENAGPNTLVTTVVASDKDGDN-----VRFGFVG---GGTSSGQ  909

Query  160   FLLSRNTGEVRTLTN--SLDREQASSYRLVVSGADK-------DGEGLSTQCECNIKVKD  210
             F++   TG +R  +   SLDR++   Y L V+  D        D    ++     + + D
Sbjct  910   FVIEEITGVIRLHSKAISLDRDK---YELNVTAMDDGSCCVNGDATIHTSTAVVVVFITD  966
```

```
Query  211   VNDNFPMFRD-SQYSARIEENILS-SELLRFQVTDLDE  246
             VNDN P+F+D   Y  ++EE   + S +++ Q TD D+
Sbjct  967   VNDNKPVFKDCGTYYPKVEEGAPNGSPVIKVQATDEDK  1004

 Score = 53.9 bits (128),  Expect = 1e-05, Method: Compositional matrix adjust.
 Identities = 70/306 (23%), Positives = 126/306 (41%), Gaps = 30/306 (10%)

Query  20    PIAKIT-SDYQATQKITYRI---SGVGIDQPPFGIFVVDKNTGDINITAIVDRE---ETP  72
             PI  +T  D   T K  Y++   SG G D+  F +   +  TG + I   +D E   ++
Sbjct  1774  PILTVTVHDEDETNKFQYKVIDNSGYGADK--FTMVRNNDGTGSLKIVQPLDYEDPLQSN  1831

Query  73    SFLITCRALNAQGLDVEKPLI------LTVKILDINDNPPVFSQQIFMGEIEENSASNSL  126
              F    + +N +G D +          + VK+ DINDN P F +      + EN+
Sbjct  1832  GFRFRIQ-VNDKGEDNDNDKYHVAYSWVVVKLRDINDNKPQFERPNIEVSVYENADVGKT  1890

Query  127   VMILNATDADEPNHLNSKIAFKIVSQEPAGTPMFLLSRNTGEVRTLTNSLDREQASSYRL  186
             +    ATD D+      SK+++ I   + +       S N     T+  +LDRE    +++
Sbjct  1891  LETFKATDPDQGG--KSKVSYAI---DRSSDRQRQFSINQEGTVTIQRNLDREVTPRHQV  1945

Query  187   VVSGADKDGEGLSTQCECNIKVKDVNDNFPMFRDSQYSARIEENILSSELLRFQVTDLDE  246
              +   D      +      + V+D+NDN P F    Y   + E++   +++    TD D+
Sbjct  1946  KILAIDDGIPPKTATATLTVIVQDINDNPPKFL-KDYRPVLPEHVPPRKVVEILATDDDD  2004

Query  247   EYTDNWLAVYFFTSGNEGNW----FEIQTDPRTNEG----ILKVVKALDYEQLQSVKLSI  298
                 N     F       +     F+++ D +   G    I+  +++ D EQ +   + I
Sbjct  2005  RSKSNGPPFQFRLDPGADDIIRASFKVEQDQKGANGDGMAIVSSLRSFDREQQKEYLIPI  2064

Query  299   AVKNKA  304
              +K+
Sbjct  2065  VIKDHG  2070

 Score = 52.4 bits (124),  Expect = 4e-05, Method: Compositional matrix adjust.
 Identities = 97/398 (24%), Positives = 163/398 (41%), Gaps = 54/398 (14%)

Query  106   PVFSQQIFMGEIEENSASNSLVMILNATDADEPNHLNSKIAFKIVSQ-EPAGTPMFLLSR  164
             P F    +  EI EN   +S ++ + A      +  + +I + + +Q + AGT  F +
Sbjct  655   PQFYMPSYEAEIPENQKKDSDIISVKAK-----SFADREIRYTLKAQGQGAGT--FNIGP  707

Query  165   NTGEVRTLTNSLDRE---QASSYRLVVSGADKDGEGLSTQCECNIKVKDVNDNFPMFRDS  221
              +G V+ L   LD E   Q   Y LVV+ A +D  G ST  E  I+V DVNDN P F
Sbjct  708   TSGIVK-LAKELDFEDLRQPHVYSLVVT-ATEDSGGFSTSVELTIRVTDVNDNAPKFELP  765

Query  222   QYSAR-IEENI-LSSELLRFQVTDLDEEYTDNWLAVYFFTSGNEGNWFEIQTDPRTNEGI  279
              Y A  ++E+I L + +L+ +  D D         + +  S +    F + ++     GI
Sbjct  766   DYQAHNVDEDIPLGTSILKVKAMDSDSGAN---AEIEYLVSDDH---FAVDSN-----GI  814

Query  280   LKVVKALDYEQLQS-VKLSIAVKNKAEFHQSVISRYRVQSTPVTIQVINVREGIAFRPAS  338
             +   K LD +   +  +  +  K+K E  ++  +  RV +           E   F
Sbjct  815   IVNNKQLDADNNNAYYEFVVTAKDKGEPAKTGTATVRVYTKNKN------DEEPKFSQQV  868

Query  339   KTFTVQKGISSKKLVDYILGTYQAIDEDTNKAASNVK--YVMGRNDGGYLMIDSKTAEIK  396
              T  V +      LV  ++         ++K   NV+  +V G    G  +I+  T  I+
Sbjct  869   YTPNVDENAGPNTLVTTVVA--------SDKDGDNVRFGFVGGGTSSGQFVIEEITGVIR  920

Query  397   FVK---NMNRDSTFI-VNKTITAEVLAIDEYTGKTSTGTVYVRVPDFNDN------CPTA  446
                   +++RD   + V             + T  TST  V V + D NDN      C T
Sbjct  921   LHSKAISLDRDKYELNVTAMDDGSCCVNGDATIHTSTAVVVVFITDVNDNKPVFKDCGTY  980

Query  447   VLE-KDAVCSSSPSVVVSARTLNNRYTGPYTFALEDQP  483
               + ++   + SP + V A   +    G   +++  QP
Sbjct  981   YPKVEEGAPNGSPVIKVQATDEDKGVNGQVKYSIVQQP  1018

 Score = 44.7 bits (104),  Expect = 0.008, Method: Compositional matrix adjust.
 Identities = 48/199 (24%), Positives = 73/199 (37%), Gaps = 19/199 (10%)

Query  27    DYQATQKITYRISGVGIDQPPFGIFVVDKNTGDINITAIVDREETPSFLITCRALNAQGL  86
             D     K++Y I      Q  F I       G + I   +DRE TP   +   A++
Sbjct  1900  DQGGKSKVSYAIDRSSDRQRQFSI----NQEGTVTIQRNLDREVTPRHQVKILAIDDGIP  1955
```

```
Query  87    DVEKPLILTVKILDINDNPPVFSQQIFMGEIEENSASNSLVMILNATDADEPNHLNSKIA  146
                    LTV + DINDNPP F +  +   + E+     +V IL   D D
Sbjct  1956  PKTATATLTVIVQDINDNPPKFLKD-YRPVLPEHVPPRKVVEILATDDDDRSKSNGPPFQ  2014

Query  147   FKIVSQEPAGTPMFLLSRNTGEVRTLTN-----------SLDREQASSYRLVVSGADKDG  195
             F++   +P    +   S    + +   N           S DREQ   Y + +   D
Sbjct  2015  FRL---DPGADDIIRASFKVEQDQKGANGDGMAIVSSLRSFDREQQKEYLIPIVIKDHGN  2071

Query  196   EGLSTQCECNIKVKDVNDN  214
               ++      + + DVNDN
Sbjct  2072  PAMTGTSTLTVVIGDVNDN  2090

 Score = 40.0 bits (92),  Expect = 0.21, Method: Compositional matrix adjust.
 Identities = 89/389 (23%), Positives = 149/389 (38%), Gaps = 62/389 (16%)

Query  75   LITCRALNAQGLDVEKPLILTVKILDINDNPPVFSQQIF--MGEIEENSASNSLVMILNA  132
            +IT +  NA GL       + + + D+ND PP F  +       ++ N+  N+ V  L A
Sbjct  517  IITNQGHNA-GLKYTDNQRVIIHVKDVNDEPPYFINRPLPMQAVVQLNAPPNTPVFTLQA  575

Query  133  TDADEPNHLNSKIAFKIVSQEPAGTPMFLLSRNTGEVRTLTNSLDREQASSYRLVVSGAD  192
             D D     +  I + IV     G   F +   +G VRT    L  +    Y L V   D
Sbjct  576  RDPD----TDHNIHYFIVRDRTGG--RFEVDERSGVVRTRGTDL-FQLDMEYVLYVKAED  628

Query  193  KDGE----GLSTQCECNIKVKDVNDNFPMFRDSQYSARIEEN-ILSSELLRFQVTDLDEE  247
            ++G+       +  E  + +       P F    Y A I EN    S+++  +     +
Sbjct  629  QNGKVDDRRFQSTPEERLSIVG-GKRAPQFYMPSYEAEIPENQKKDSDIISVKA----KS  683

Query  248  YTDNWLAVYFFTSGNEGNWFEIQTDPRTNEGILKVVKALDYEQLQSVKLS----IAVKNK  303
            + D  +       G     F I        GI+K+ K LD+E L+   +      A ++
Sbjct  684  FADREIRYTLKAQGQGAGTFNIGP----TSGIVKLAKELDFEDLRQPHVYSLVVTATEDS  739

Query  304  AEFHQSVISRYRVQSTPVTIQVINVREGIAFRPASKTFTVQKGISSKKLVDYILGT----  359
              F  SV          +TI+V +V +       +  F +    +     D  LGT
Sbjct  740  GGFSTSV---------ELTIRVTDVNDN------APKFELPDYQAHNVDEDIPLGTSILK  784

Query  360  YQAIDEDTNKAASNVKYVMGRN----DGGYLMIDSKTAEIKFVKNMNRDSTFIVNKTITA  415
             +A+D D+  A + ++Y++  +    D   +++++K  +     N N    F+V
Sbjct  785  VKAMDSDSG-ANAEIEYLVSDDHFAVDSNGIIVNNKQLD---ADNNNAYYEFVVTAKDKG  840

Query  416  EVLAIDEYTGKTSTGTVYVRVPDFNDNCP  444
            E         KT T TV V   + ND  P
Sbjct  841  E-------PAKTGTATVRVYTKNKNDEEP  862

 Score = 39.7 bits (91),  Expect = 0.25, Method: Compositional matrix adjust.
 Identities = 54/242 (22%), Positives = 102/242 (42%), Gaps = 22/242 (9%)

Query  4    KFAKPCREGE--DNSKRNP-IAKITSDYQATQKITYRISGVGIDQPPFGIFVVDKNTGDI  60
            +F  P  E E  +N K++  I  + +   A ++I Y +   G      G F +   +G +
Sbjct  656  QFYMPSYEAEIPENQKKDSDIISVKAKSFADREIRYTLKAQGQGA---GTFNIGPTSGIV  712

Query  61   NITAIVDREE--TPSF--LITCRALNAQGLDVEKPLILTVKILDINDNPPVFSQQIFMGE  116
             +   +D E+   P    L+     ++ G      + LT+++ D+NDN P F    +
Sbjct  713  KLAKELDFEDLRQPHVYSLVVTATEDSGGFSTS--VELTIRVTDVNDNAPKFELPDYQAH  770

Query  117  -IEENSASNSLVMILNATDADEPNHLNSKIAFKIVSQEPAGTPMFLLSRNTGEVRTLTNS  175
             ++E+    + ++ + A D+D  +  N++I + +VS +      F +  N   V
Sbjct  771  NVDEDIPLGTSILKVKAMDSD--SGANAEIEY-LVSDD-----HFAVDSNGIIVNNKQLD  822

Query  176  LDREQASSYRLVVSGADKDGEGLSTQCECNIKVKDVNDNFPMFRDSQYSARIEENILSSE  235
             D   A  Y  VV+  DK     +      +  K+ ND  P F    Y+  ++EN   +
Sbjct  823  ADNNNA-YYEFVVTAKDKGEPAKTGTATVRVYTKNKNDEEPKFSQQVYTPNVDENAGPNT  881

Query  236  LL  237
            L+
Sbjct  882  LV  883

>AARA21_009090.P17038 | transcript=AARA21_009090.R17038 | gene=AARA21_009090
```

```
| organism=Anopheles_arabiensis_DONGOLA_2021
| gene_product=neural-cadherin, transcript variant X5 |
transcript_product=neural-cadherin,
transcript variant X4 | location=CM029353:40466514-40666848(+)
| protein_length=3105 | sequence_SO=chromosome
| SO=protein_coding_gene | is_pseudo=false
Length=3105

 Score = 144 bits (364),  Expect = 1e-34, Method: Compositional matrix adjust.
 Identities = 125/437 (29%), Positives = 211/437 (48%), Gaps = 33/437 (8%)

Query  26    SDYQATQKITYRISGVGID--QPPFGIFVVDKNTGDINITAIVDREET---PSFLITCRA  80
              D    Q I Y ++G GID   P    F +++ TG+I +   +DR++    P +  T  A
Sbjct  1554  GDKDRPQNIVYFLTGQGIDPDNPANSKFDINRTTGEIFVLKPLDRDQPNGRPQWRFTVFA  1613

Query  81    LNAQGLDVEKPLILTVKILDINDNPPVFSQQIFMGEIEENSASNSLVMILNATDADEPNH  140
              +  G  +     + V + DINDN P+F Q ++ G + EN  +  +VM + A D D+PN
Sbjct  1614  QDEGGEGLVGYADVQVNLKDINDNAPIFPQGVYFGNVTENGTAGMVVMTMTAVDYDDPNE  1673

Query  141   -LNSKIAFKI---VSQEPAGTPMFLLSRNTGEVRTLTNSLDREQASSYRLVVSGADKDGE  196
                N+K+ + I   V +E  G+P+F +  +TG ++T    LDRE+   Y + V     DG
Sbjct  1674  GTNAKLIYSIEKNVIEEETGSPIFEIEADTGVIKTAVCCLDRERTPDYSIQVVAM--DGG  1731

Query  197   GLSTQCECNIKVKDVNDNFPMFRDSQYSARIEENILSS--ELLRFQVTDLDEEYTDNWLA  254
             GL      +I+VKD+ND  P F   ++   ++E   ++  E+    VT  DE+ T+ +
Sbjct  1732  GLKGTGTASIRVKDINDMPPQFTKDEWFTEVDETDGTNLPEMPILTVTVHDEDETNKFQY  1791

Query  255   VYFFTSGNEGNWFEIQTDPRTNEGI--LKVVKALDYE---QLQSVKLSIAVKNKAEFHQS  309
                  SG   + F +    R N+G   LK+V+ LDYE   Q    +  I V +K E + +
Sbjct  1792  KVIDNSGYGADKFTMV---RNNDGTGSLKIVQPLDYEDPLQSNGFRFRIQVNDKGEDNDN  1848

Query  310   VISRYRVQSTPVTIQVINVREGIAFRPASKTFTVQKGISSKKLVDYILGTYQAIDEDTNK  369
                +Y V  + V +++ ++ +    +P  +   ++  +     V   L T++A D D
Sbjct  1849  --DKYHVAYSWVVVKLRDINDN---KPQFERPNIEVSVYENADVGKTLETFKATDPDQG-  1902

Query  370   AASNVKYVMGR-NDGGYLMIDSKTAEIKFVKNMNRDSTFIVNKTITAEVLAIDE-YTGKT  427
               S V Y + R +D       ++   +   +N++R+    V      ++LAID+    KT
Sbjct  1903  GKSKVSYAIDRSSDRQRQFSINQEGTVTIQRNLDRE----VTPRHQVKILAIDDGIPPKT  1958

Query  428   STGTVYVRVPDFNDNCP  444
             +T T+ V V D NDN P
Sbjct  1959  ATATLTVIVQDINDNPP  1975

 Score = 85.9 bits (211),  Expect = 1e-15, Method: Compositional matrix adjust.
 Identities = 108/420 (26%), Positives = 177/420 (42%), Gaps = 52/420 (12%)

Query  50    IFVVDKNTGDINITAIVDREETPSFLITCRAL----------NAQGLDVEKPLILTVKIL  99
              F ++  +G+I    + DRE+  ++ +   A           N Q   V K   + + I
Sbjct  1359  FFEINLQSGEIFTKVVFDREKQGAYALEVEARDGAPSARPNSNQQPNSVTK--FIRIGIA  1416

Query  100   DINDNPPVFSQQIFMGEIEENSASNSLVMILNATDADEPNHLNSKIAFKIVSQEPAGTPM  159
             D NDNPP F + ++  E++EN      V+ + A D DE    +S+I ++I S    G
Sbjct  1417  DKNDNPPYFDKALYEAEVDENEDIQHTVLTVTAKDHDE----SSRIRYEITSGNIGGA--  1470

Query  160   FLLSRNTGEVRTLTNSLDREQASSYRLVVSGADKDGEGLSTQCECNIKVKDVNDNFPMFR  219
             F +   TG +  +  +LD E    Y L ++ +D   E  +T     I VKDVNDN P+F
Sbjct  1471  FAVKNMTGAI-YVAGALDYETRKRYELRLAASDNLKENYTTVV---IHVKDVNDNPPVFE  1526

Query  220   DSQYSARIEENI---LSSELLRFQVTDLDEEYTDNWLAVYFFTSG--NEGNWFEIQTDPR  274
                Y  +I E     L   +L+   TD D++   N   VYF T    +  N    + D
Sbjct  1527  RPTYRTQITEEDDRNLPKRVLQVTATDGDKDRPQN--IVYFLTGQGIDPDNPANSKFDIN  1584

Query  275   TNEGILKVVKALDYEQLQS-VKLSIAVKNKAEFHQSVISRYRVQSTPVTIQVINVREGIA  333
                G + V+K LD +Q     +    V  + E  + ++    VQ     + + ++ +
Sbjct  1585  RTTGEIFVLKPLDRDQPNGRPQWRFTVFAQDEGGEGLVGYADVQ-----VNLKDINDNAP  1639

Query  334   FRPASKTFTVQKGISSKKLVDYILGTYQAIDEDTNKAASNVK--YVMGRN------DGGY  385
               P    F     ++        ++ T  A+D D     +N K  Y + +N
Sbjct  1640  IFPQGVYF---GNVTENGTAGMVVMTMTAVDYDDPNEGTNAKLIYSIEKNVIEEETGSPI  1696
```

```
Query  386   LMIDSKTAEIK-FVKNMNRDSTFIVNKTITAEVLAIDEYTGKTSTGTVYYVRVPDFNDNCP  444
               I++ T  IK  V  ++R+ T        + +V+A+D   G   TGT  +RV D ND  P
Sbjct  1697  FEIEADTGVIKTAVCCLDRERT----PDYSIQVVAMDG-GGLKGTGTASIRVKDINDMPP  1751

 Score = 76.3 bits (186),  Expect = 1e-12, Method: Compositional matrix adjust.
 Identities = 106/426 (25%), Positives = 181/426 (42%), Gaps = 51/426 (12%)

Query  53    VDKNTGDINITAIVDREETPSFLITCRALNAQGLDVEKPLILTVKILDINDNPPVFSQ-Q  111
             +D++  ++N+TA+ D          +C  +N           ++ V I D+NDN PVF
Sbjct  928   LDRDKYELNVTAMDDG--------SC-CVNGDATIHTSTAVVVVFITDVNDNKPVFKDCG  978

Query  112   IFMGEIEENSASNSLVMILNATDADEPNHLNSKIAFKIVSQEPAGTPMFLLSRNTGEVRT  171
              +  ++EE + + S V+ + ATD D+    +N ++ + IV Q        F +    TGEV T
Sbjct  979   TYYPKVEEGAPNGSPVIKVQATDEDK--GVNGQVKYSIVQQPNQKGTKFTVDEETGEVST  1036

Query  172   LTNSLDRE--QASSYRLVVSGADKDGEGLSTQCECNIKVKDVNDNFPMFRDSQYSARIEE  229
                  DRE         + V   D+     L   C     +++ DVNDN P+F    +Y   +++
Sbjct  1037  -NKVFDREGDDGKFVSVTVKATDQGEPSLEGVCSFTVEITDVNDNPPLFDRQKYVENVKQ  1095

Query  230   NI-LSSELLRFQVTDLDEEYTDNWLAVYFFT---SGNEGNWFEIQTDPRTNEGILKVVKA  285
             +  + + +LR  V+  DE+  +N   VY  +   S N+  +FEIQ  P +    +LK  K
Sbjct  1096  DASIGTNILR--VSASDEDADNNGAIVYTLSAPHSPNDLEYFEIQ--PESGWIVLK--KP  1149

Query  286   LD--YEQLQSVKLSIAVKNK------AEFHQSVISRYRVQSTPVTIQV----INVREGIA  333
             LD  Y + +  +L +   ++       A+     +     R     P+   +      I++RE +
Sbjct  1150  LDGCYSR-ERYRLKVYASDRGDPPSTADVDVELDVVDRNNKPPIWDRTIYGPIHIRENVT  1208

Query  334   FRPASKTFTVQKGISSKKLVDYILGTYQAIDEDTNKAASNVKYVMGRNDGGYLMIDSKTA  393
                   +     GI      V Y L           TNK    +  Y+  R D G         A
Sbjct  1209  VGTVVTSVKASSGIEGNPTVFYRL--MPGSTAQTNKL--HTFYLQQRPDNG-----DTWA  1259

Query  394   EIKFVKNMNRDSTFIVNKTITAEVLAIDEYTGKTSTGTVYYVRVPDFNDNCPTAV-LEKDA  452
             +IK    ++ ++    N T+  E      +      S  TVY+++ D ND  P      E++
Sbjct  1260  DIKVNHPLDFETIKEYNLTVRVENNGAQQL---ASEATVYIQLEDVNDEIPLFTEREQET  1316

Query  453   VCSSSP  458
             V   P
Sbjct  1317  VLEGEP  1322

 Score = 60.8 bits (146),  Expect = 9e-08, Method: Compositional matrix adjust.
 Identities = 83/362 (23%), Positives = 152/362 (42%), Gaps = 34/362 (9%)

Query  51    FVVDKNTGDINITAIVDRE-ETPSFL-ITCRALNAQGLDVEKPLILTVKILDINDNPPVF  108
             F VD+ TG+++   + DRE +   F+ +T +A +      +E      TV+I D+NDNPP+F
Sbjct  1025  FTVDEETGEVSTNKVFDREGDDGKFVSVTVKATDQGEPSLEGVCSFTVEITDVNDNPPLF  1084

Query  109   SQQIFMGEIEENSASNSLVMILNATDADEPNHLNSKIAFKIVS-QEPAGTPMFLLSRNTG  167
              +Q ++   ++++++   + ++ ++A+D D   N  N   I + + +   P      F +   +G
Sbjct  1085  DRQKYVENVKQDASIGTNILRVSASDEDADN--NGAIVYTLSAPHSPNDLEYFEIQPESG  1142

Query  168   EV---RTLTNSLDREQASSYRLVVSGADKDGEGLSTQCECNIKVKDVNDNFPMFRDSQYS  224
              +    + L      RE+   YRL V  +D+ G+  ST              +N P   D
Sbjct  1143  WIVLKKPLDGCYSRER---YRLKVYASDR-GDPPSTADVDVELDVVDRNNKPPIWDRTIY  1198

Query  225   ARIEENILSSELLRFQVTDLDEEYTDNWLAVYFFTSG-----NEGNWFEIQTDPRTNE--  277
               I       +                  N    Y    G      N+ + F +Q  P   +
Sbjct  1199  GPIHIRENVTVGTVVTSVKASSGIEGNPTVFYRLMPGSTAQTNKLHTFYLQQRPDNGDTW  1258

Query  278   GILKVVKALDYEQLQSVKLSIAVKNKAEFHQSVISRYRVQSTPVTIQVINVREGIAFRPA  337
               +KV   LD+E ++   L++ V+N         ++        V IQ+ +V + I
Sbjct  1259  ADIKVNHPLDFETIKEYNLTVRVENNG-------AQQLASEATVYIQLEDVNDEIPLFTE  1311

Query  338   SKTFTVQKGISSKKLVDYILGTYQAIDEDTNKAASNVKYVM---GRNDG-GYLMIDSKTA  393
              +  TV +G      V  +     AID+D      + V Y +     RN+G  +  I+ ++
Sbjct  1312  REQETVLEGEPIGTKVTQV----NAIDKDGTFPNNQVYYYIVDSPRNEGKDFFEINLQSG  1367

Query  394   EI  395
             EI
Sbjct  1368  EI  1369
```

```
 Score = 55.8 bits (133),  Expect = 3e-06, Method: Compositional matrix adjust.
 Identities = 49/158 (31%), Positives = 78/158 (49%), Gaps = 24/158 (15%)

Query  102   NDNPPVFSQQIFMGEIEENSASNSLVMILNATDADEPNHLNSKIAFKIVSQEPAGTP--M  159
             ND  P FSQQ++   ++EN+  N+LV  + A+D D  N     + F  V     GT
Sbjct  858   NDEEPKFSQQVYTPNVDENAGPNTLVTTVVASDKDGDN-----VRFGFVG---GGTSSGQ  909

Query  160   FLLSRNTGEVRTLTN--SLDREQASSYRLVVSGADK-------DGEGLSTQCECNIKVKD  210
             F++   TG +R  +   SLDR++   Y L V+  D        D    ++     + + D
Sbjct  910   FVIEEITGVIRLHSKAISLDRDK---YELNVTAMDDGSCCVNGDATIHTSTAVVVVFITD  966

Query  211   VNDNFPMFRD-SQYSARIEENILS-SELLRFQVTDLDE  246
             VNDN P+F+D   Y  ++EE   + S +++ Q TD D+
Sbjct  967   VNDNKPVFKDCGTYYPKVEEGAPNGSPVIKVQATDEDK  1004

 Score = 53.9 bits (128),  Expect = 1e-05, Method: Compositional matrix adjust.
 Identities = 70/306 (23%), Positives = 126/306 (41%), Gaps = 30/306 (10%)

Query  20    PIAKIT-SDYQATQKITYRI---SGVGIDQPPFGIFVVDKNTGDINITAIVDRE---ETP  72
             PI  +T  D   T K  Y++   SG G D+  F +   +  TG + I   +D E   ++
Sbjct  1774  PILTVTVHDEDETNKFQYKVIDNSGYGADK--FTMVRNNDGTGSLKIVQPLDYEDPLQSN  1831

Query  73    SFLITCRALNAQGLDVEKPLI------LTVKILDINDNPPVFSQQIFMGEIEENSASNSL  126
              F    + +N +G D +          + VK+ DINDN P F +      + EN+
Sbjct  1832  GFRFRIQ-VNDKGEDNDNDKYHVAYSWVVVKLRDINDNKPQFERPNIEVSVYENADVGKT  1890

Query  127   VMILNATDADEPNHLNSKIAFKIVSQEPAGTPMFLLSRNTGEVRTLTNSLDREQASSYRL  186
             +    ATD D+     SK+++ I   + +       S N     T+  +LDRE    +++
Sbjct  1891  LETFKATDPDQGG--KSKVSYAI---DRSSDRQRQFSINQEGTVTIQRNLDREVTPRHQV  1945

Query  187   VVSGADKDGEGLSTQCECNIKVKDVNDNFPMFRDSQYSARIEENILSSELLRFQVTDLDE  246
              +   D      +      + V+D+NDN P F    Y   + E++   +++    TD D+
Sbjct  1946  KILAIDDGIPPKTATATLTVIVQDINDNPPKFL-KDYRPVLPEHVPPRKVVEILATDDDD  2004

Query  247   EYTDNWLAVYFFTSGNEGNW----FEIQTDPRTNEG----ILKVVKALDYEQLQSVKLSI  298
                 N     F       +     F+++ D +   G    I+  +++ D EQ +   + I
Sbjct  2005  RSKSNGPPFQFRLDPGADDIIRASFKVEQDQKGANGDGMAIVSSLRSFDREQQKEYLIPI  2064

Query  299   AVKNKA  304
              +K+
Sbjct  2065  VIKDHG  2070

 Score = 52.4 bits (124),  Expect = 4e-05, Method: Compositional matrix adjust.
 Identities = 97/398 (24%), Positives = 163/398 (41%), Gaps = 54/398 (14%)

Query  106   PVFSQQIFMGEIEENSASNSLVMILNATDADEPNHLNSKIAFKIVSQ-EPAGTPMFLLSR  164
             P F    +  EI EN   +S ++ + A      +  + +I + + +Q + AGT  F +
Sbjct  655   PQFYMPSYEAEIPENQKKDSDIISVKAK-----SFADREIRYTLKAQGQGAGT--FNIGP  707

Query  165   NTGEVRTLTNSLDRE---QASSYRLVVSGADKDGEGLSTQCECNIKVKDVNDNFPMFRDS  221
              +G V+ L   LD E   Q   Y LVV+ A +D  G ST  E  I+V DVNDN P F
Sbjct  708   TSGIVK-LAKELDFEDLRQPHVYSLVVT-ATEDSGGFSTSVELTIRVTDVNDNAPKFELP  765

Query  222   QYSAR-IEENI-LSSELLRFQVTDLDEEYTDNWLAVYFFTSGNEGNWFEIQTDPRTNEGI  279
              Y A  ++E+I L + +L+ +  D D         + +  S +    F + ++     GI
Sbjct  766   DYQAHNVDEDIPLGTSILKVKAMDSDSGAN---AEIEYLVSDDH---FAVDSN-----GI  814

Query  280   LKVVKALDYEQLQS-VKLSIAVKNKAEFHQSVISRYRVQSTPVTIQVINVREGIAFRPAS  338
             +   K LD +   +  +  +  K+K E  ++  +  RV +           E   F
Sbjct  815   IVNNKQLDADNNNAYYEFVVTAKDKGEPAKTGTATVRVYTKNKN------DEEPKFSQQV  868

Query  339   KTFTVQKGISSKKLVDYILGTYQAIDEDTNKAASNVK--YVMGRNDGGYLMIDSKTAEIK  396
              T  V +      LV  ++         ++K   NV+  +V G    G  +I+  T  I+
Sbjct  869   YTPNVDENAGPNTLVTTVVA--------SDKDGDNVRFGFVGGGTSSGQFVIEEITGVIR  920

Query  397   FVK---NMNRDSTFI-VNKTITAEVLAIDEYTGKTSTGTVYVRVPDFNDN------CPTA  446
                   +++RD   + V            + T  TST  V V + D NDN      C T
```

```
Sbjct  921   LHSKAISLDRDKYELNVTAMDDGSCCVNGDATIHTSTAVVVVFITDVNDNKPVFKDCGTY  980

Query  447   VLE-KDAVCSSSPSVVVSARTLNNRYTGPYTFALEDQP  483
               + ++   + SP + V A   +    G   +++  QP
Sbjct  981   YPKVEEGAPNGSPVIKVQATDEDKGVNGQVKYSIVQQP  1018

 Score = 44.7 bits (104),  Expect = 0.008, Method: Compositional matrix adjust.
 Identities = 48/199 (24%), Positives = 73/199 (37%), Gaps = 19/199 (10%)

Query  27    DYQATQKITYRISGVGIDQPPFGIFVVDKNTGDINITAIVDREETPSFLITCRALNAQGL  86
             D     K++Y I      Q  F I       G + I   +DRE TP   +   A++
Sbjct  1900  DQGGKSKVSYAIDRSSDRQRQFSI----NQEGTVTIQRNLDREVTPRHQVKILAIDDGIP  1955

Query  87    DVEKPLILTVKILDINDNPPVFSQQIFMGEIEENSASNSLVMILNATDADEPNHLNSKIA  146
                    LTV + DINDNPP F +  +   + E+     +V IL   D D
Sbjct  1956  PKTATATLTVIVQDINDNPPKFLKD-YRPVLPEHVPPRKVVEILATDDDDRSKSNGPPFQ  2014

Query  147   FKIVSQEPAGTPMFLLSRNTGEVRTLTN-----------SLDREQASSYRLVVSGADKDG  195
             F++   +P    +   S    + +   N           S DREQ   Y + +   D
Sbjct  2015  FRL---DPGADDIIRASFKVEQDQKGANGDGMAIVSSLRSFDREQQKEYLIPIVIKDHGN  2071

Query  196   EGLSTQCECNIKVKDVNDN  214
               ++      + + DVNDN
Sbjct  2072  PAMTGTSTLTVVIGDVNDN  2090

 Score = 40.0 bits (92),  Expect = 0.21, Method: Compositional matrix adjust.
 Identities = 89/389 (23%), Positives = 149/389 (38%), Gaps = 62/389 (16%)

Query  75   LITCRALNAQGLDVEKPLILTVKILDINDNPPVFSQQIF--MGEIEENSASNSLVMILNA  132
            +IT +  NA GL       + + + D+ND PP F  +       ++ N+  N+ V  L A
Sbjct  517  IITNQGHNA-GLKYTDNQRVIIHVKDVNDEPPYFINRPLPMQAVVQLNAPPNTPVFTLQA  575

Query  133  TDADEPNHLNSKIAFKIVSQEPAGTPMFLLSRNTGEVRTLTNSLDREQASSYRLVVSGAD  192
             D D     +  I + IV     G   F +   +G VRT    L  +    Y L V   D
Sbjct  576  RDPD----TDHNIHYFIVRDRTGG--RFEVDERSGVVRTRGTDL-FQLDMEYVLYVKAED  628

Query  193  KDGE----GLSTQCECNIKVKDVNDNFPMFRDSQYSARIEEN-ILSSELLRFQVTDLDEE  247
            ++G+       +  E  + +       P F    Y A I EN    S+++  +     +
Sbjct  629  QNGKVDDRRFQSTPEERLSIVG-GKRAPQFYMPSYEAEIPENQKKDSDIISVKA----KS  683

Query  248  YTDNWLAVYFFTSGNEGNWFEIQTDPRTNEGILKVVKALDYEQLQSVKLS----IAVKNK  303
            + D  +       G     F I        GI+K+ K LD+E L+   +      A ++
Sbjct  684  FADREIRYTLKAQGQGAGTFNIGP----TSGIVKLAKELDFEDLRQPHVYSLVVTATEDS  739

Query  304  AEFHQSVISRYRVQSTPVTIQVINVREGIAFRPASKTFTVQKGISSKKLVDYILGT----  359
              F  SV          +TI+V +V +       +  F +    +     D  LGT
Sbjct  740  GGFSTSV---------ELTIRVTDVNDN------APKFELPDYQAHNVDEDIPLGTSILK  784

Query  360  YQAIDEDTNKAASNVKYVMGRN----DGGYLMIDSKTAEIKFVKNMNRDSTFIVNKTITA  415
             +A+D D+  A + ++Y++  +    D   +++++K  +     N N    F+V
Sbjct  785  VKAMDSDSG-ANAEIEYLVSDDHFAVDSNGIIVNNKQLD---ADNNNAYYEFVVTAKDKG  840

Query  416  EVLAIDEYTGKTSTGTVYYVRVPDFNDNCP  444
            E         KT T TV V   + ND  P
Sbjct  841  E-------PAKTGTATVRVYTKNKNDEEP  862

 Score = 39.7 bits (91),  Expect = 0.25, Method: Compositional matrix adjust.
 Identities = 54/242 (22%), Positives = 102/242 (42%), Gaps = 22/242 (9%)

Query  4    KFAKPCREGE--DNSKRNP-IAKITSDYQATQKITYRISGVGIDQPPFGIFVVDKNTGDI  60
            +F  P  E E  +N K++  I  + +   A ++I Y +   G      G F +   +G +
Sbjct  656  QFYMPSYEAEIPENQKKDSDIISVKAKSFADREIRYTLKAQGQGA---GTFNIGPTSGIV  712

Query  61   NITAIVDREE--TPSF--LITCRALNAQGLDVEKPLILTVKILDINDNPPVFSQQIFMGE  116
             +   +D E+   P    L+     ++ G      + LT+++ D+NDN P F    +
Sbjct  713  KLAKELDFEDLRQPHVYSLVVTATEDSGGFSTS--VELTIRVTDVNDNAPKFELPDYQAH  770

Query  117  -IEENSASNSLVMILNATDADEPNHLNSKIAFKIVSQEPAGTPMFLLSRNTGEVRTLTNS  175
```

```
             ++E+    + ++ + A D+D  +  N++I + +VS +      F +  N   V
Sbjct  771  NVDEDIPLGTSILKVKAMDSD--SGANAEIEY-LVSDD-----HFAVDSNGIIVNNKQLD  822

Query  176  LDREQASSYRLVVSGADKDGEGLSTQCECNIKVKDVNDNFPMFRDSQYSARIEENILSSE  235
             D   A  Y  VV+  DK     +      +  K+ ND  P F    Y+  ++EN   +
Sbjct  823  ADNNNA-YYEFVVTAKDKGEPAKTGTATVRVYTKNKNDEEPKFSQQVYTPNVDENAGPNT  881

Query  236  LL  237
            L+
Sbjct  882  LV  883

>AARA21_009090.P17037 | transcript=AARA21_009090.R17037 | gene=AARA21_009090
| organism=Anopheles_arabiensis_DONGOLA_2021
| gene_product=neural-cadherin, transcript variant X5 |
transcript_product=neural-cadherin,
transcript variant X3 | location=CM029353:40466514-40666848(+)
| protein_length=3105 | sequence_SO=chromosome
| SO=protein_coding_gene | is_pseudo=false
Length=3105

 Score = 144 bits (364),  Expect = 1e-34, Method: Compositional matrix adjust.
 Identities = 125/437 (29%), Positives = 211/437 (48%), Gaps = 33/437 (8%)

Query  26    SDYQATQKITYRISGVGID--QPPFGIFVVDKNTGDINITAIVDREET---PSFLITCRA  80
              D    Q I Y ++G GID   P    F +++ TG+I +   +DR++    P +  T  A
Sbjct  1554  GDKDRPQNIVYFLTGQGIDPDNPANSKFDINRTTGEIFVLKPLDRDQPNGRPQWRFTVFA  1613

Query  81    LNAQGLDVEKPLILTVKILDINDNPPVFSQQIFMGEIEENSASNSLVMILNATDADEPNH  140
              +  G  +     + V + DINDN P+F Q ++ G + EN  +  +VM + A D D+PN
Sbjct  1614  QDEGGEGLVGYADVQVNLKDINDNAPIFPQGVYFGNVTENGTAGMVVMTMTAVDYDDPNE  1673

Query  141   -LNSKIAFKI---VSQEPAGTPMFLLSRNTGEVRTLTNSLDREQASSYRLVVSGADKDGE  196
                N+K+ + I   V +E  G+P+F +  +TG ++T    LDRE+   Y + V     DG
Sbjct  1674  GTNAKLIYSIEKNVIEEETGSPIFEIEADTGVIKTAVCCLDRERTPDYSIQVVAM--DGG  1731

Query  197   GLSTQCECNIKVKDVNDNFPMFRDSQYSARIEENILSS--ELLRFQVTDLDEEYTDNWLA  254
              GL      +I+VKD+ND  P F   ++   ++E   ++  E+    VT  DE+ T+ +
Sbjct  1732  GLKGTGTASIRVKDINDMPPQFTKDEWFTEVDETDGTNLPEMPILTVTVHDEDETNKFQY  1791

Query  255   VYFFTSGNEGNWFEIQTDPRTNEGI--LKVVKALDYE---QLQSVKLSIAVKNKAEFHQS  309
                   SG   + F +    R N+G   LK+V+ LDYE   Q    +  I V +K E + +
Sbjct  1792  KVIDNSGYGADKFTMV---RNNDGTGSLKIVQPLDYEDPLQSNGFRFRIQVNDKGEDNDN  1848

Query  310   VISRYRVQSTPVTIQVINVREGIAFRPASKTFTVQKGISSKKLVDYILGTYQAIDEDTNK  369
                 +Y V  + V +++ ++ +    +P  +   ++  +     V   L T++A D D
Sbjct  1849  --DKYHVAYSWVVVKLRDINDN---KPQFERPNIEVSVYENADVGKTLETFKATDPDQG-  1902

Query  370   AASNVKYVMGR-NDGGYLMIDSKTAEIKFVKNMNRDSTFIVNKTITAEVLAIDE-YTGKT  427
                S V Y + R +D       ++   +   +N++R+    V      ++LAID+    KT
Sbjct  1903  GKSKVSYAIDRSSDRQRQFSINQEGTVTIQRNLDRE----VTPRHQVKILAIDDGIPPKT  1958

Query  428   STGTVYVRVPDFNDNCP  444
              +T T+ V V D NDN P
Sbjct  1959  ATATLTVIVQDINDNPP  1975

 Score = 85.9 bits (211),  Expect = 1e-15, Method: Compositional matrix adjust.
 Identities = 108/420 (26%), Positives = 177/420 (42%), Gaps = 52/420 (12%)

Query  50    IFVVDKNTGDINITAIVDREETPSFLITCRAL----------NAQGLDVEKPLILTVKIL  99
               F ++  +G+I    + DRE+  ++ +   A           N Q   V K   + + I
Sbjct  1359  FFEINLQSGEIFTKVVFDREKQGAYALEVEARDGAPSARPNSNQQPNSVTK--FIRIGIA  1416

Query  100   DINDNPPVFSQQIFMGEIEENSASNSLVMILNATDADEPNHLNSKIAFKIVSQEPAGTPM  159
              D NDNPP F + ++  E++EN      V+ + A D DE    +S+I ++I S    G
Sbjct  1417  DKNDNPPYFDKALYEAEVDENEDIQHTVLTVTAKDHDE----SSRIRYEITSGNIGGA--  1470

Query  160   FLLSRNTGEVRTLTNSLDREQASSYRLVVSGADKDGEGLSTQCECNIKVKDVNDNFPMFR  219
              F +   TG +  +  +LD E    Y L ++ +D   E  +T     I VKDVNDN P+F
Sbjct  1471  FAVKNMTGAI-YVAGALDYETRKRYELRLAASDNLKENYTTVV---IHVKDVNDNPPVFE  1526
```

```
Query  220   DSQYSARIEENI---LSSELLRFQVTDLDEEYTDNWLAVYFFTSG--NEGNWFEIQTDPR  274
                Y  +I E     L   +L+   TD D++    N   VYF T    +  N    + D
Sbjct  1527  RPTYRTQITEEDDRNLPKRVLQVTATDGDKDRPQN--IVYFLTGQGIDPDNPANSKFDIN  1584

Query  275   TNEGILKVVKALDYEQLQS-VKLSIAVKNKAEFHQSVISRYRVQSTPVTIQVINVREGIA  333
                G + V+K LD +Q      +    V  + E  + ++    VQ     + + ++ +
Sbjct  1585  RTTGEIFVLKPLDRDQPNGRPQWRFTVFAQDEGGEGLVGYADVQ-----VNLKDINDNAP  1639

Query  334   FRPASKTFTVQKGISSKKLVDYILGTYQAIDEDTNKAASNVK--YVMGRN------DGGY  385
              P    F     ++        ++ T  A+D D     +N K  Y + +N
Sbjct  1640  IFPQGVYF---GNVTENGTAGMVVMTMTAVDYDDPNEGTNAKLIYSIEKNVIEEETGSPI  1696

Query  386   LMIDSKTAEIK-FVKNMNRDSTFIVNKTITAEVLAIDEYTGKTSTGTVYVRVPDFNDNCP  444
                I++ T  IK  V  ++R+ T        + +V+A+D   G   TGT  +RV D ND  P
Sbjct  1697  FEIEADTGVIKTAVCCLDRERT----PDYSIQVVAMDG-GGLKGTGTASIRVKDINDMPP  1751

 Score = 76.3 bits (186),  Expect = 1e-12, Method: Compositional matrix adjust.
 Identities = 106/426 (25%), Positives = 181/426 (42%), Gaps = 51/426 (12%)

Query  53    VDKNTGDINITAIVDREETPSFLITCRALNAQGLDVEKPLILTVKILDINDNPPVFSQ-Q  111
             +D++  ++N+TA+ D          +C   +N           ++ V I D+NDN PVF
Sbjct  928   LDRDKYELNVTAMDDG--------SC-CVNGDATIHTSTAVVVVFITDVNDNKPVFKDCG  978

Query  112   IFMGEIEENSASNSLVMILNATDADEPNHLNSKIAFKIVSQEPAGTPMFLLSRNTGEVRT  171
              +  ++EE + + S V+ + ATD D+    +N ++ + IV Q        F +   TGEV T
Sbjct  979   TYYPKVEEGAPNGSPVIKVQATDEDK--GVNGQVKYSIVQQPNQKGTKFTVDEETGEVST  1036

Query  172   LTNSLDRE--QASSYRLVVSGADKDGEGLSTQCECNIKVKDVNDNFPMFRDSQYSARIEE  229
                  DRE         + V   D+     L   C    +++ DVNDN P+F    +Y   +++
Sbjct  1037  -NKVFDREGDDGKFVSVTVKATDQGEPSLEGVCSFTVEITDVNDNPPLFDRQKYVENVKQ  1095

Query  230   NI-LSSELLRFQVTDLDEEYTDNWLAVYFFT---SGNEGNWFEIQTDPRTNEGILKVVKA  285
             +   + + +LR  V+   DE+   +N    VY  +    S N+   +FEIQ   P +    +LK  K
Sbjct  1096  DASIGTNILR--VSASDEDADNNGAIVYTLSAPHSPNDLEYFEIQ--PESGWIVLK--KP  1149

Query  286   LD--YEQLQSVKLSIAVKNK------AEFHQSVISRYRVQSTPVTIQV----INVREGIA  333
             LD  Y + +  +L +   ++      A+     +     R     P+   +     I++RE +
Sbjct  1150  LDGCYSR-ERYRLKVYASDRGDPPSTADVDVELDVVDRNNKPPIWDRTIYGPIHIRENVT  1208

Query  334   FRPASKTFTVQKGISSKKLVDYILGTYQAIDEDTNKAASNVKYVMGRNDGGYLMIDSKTA  393
                   +     GI     V Y L          TNK    +  Y+  R D G        A
Sbjct  1209  VGTVVTSVKASSGIEGNPTVFYRL--MPGSTAQTNKL--HTFYLQQRPDNG-----DTWA  1259

Query  394   EIKFVKNMNRDSTFIVNKTITAEVLAIDEYTGKTSTGTVYVRVPDFNDNCPTAV-LEKDA  452
             +IK    ++ ++    N T+  E       +      S  TVY+++ D ND  P     E++
Sbjct  1260  DIKVNHPLDFETIKEYNLTVRVENNGAQQL---ASEATVYIQLEDVNDEIPLFTEREQET  1316

Query  453   VCSSSP  458
             V    P
Sbjct  1317  VLEGEP  1322

 Score = 60.8 bits (146),  Expect = 9e-08, Method: Compositional matrix adjust.
 Identities = 83/362 (23%), Positives = 152/362 (42%), Gaps = 34/362 (9%)

Query  51    FVVDKNTGDINITAIVDRE-ETPSFL-ITCRALNAQGLDVEKPLILTVKILDINDNPPVF  108
             F VD+ TG+++    + DRE +    F+ +T +A +      +E      TV+I D+NDNPP+F
Sbjct  1025  FTVDEETGEVSTNKVFDREGDDGKFVSVTVKATDQGEPSLEGVCSFTVEITDVNDNPPLF  1084

Query  109   SQQIFMGEIEENSASNSLVMILNATDADEPNHLNSKIAFKIVS-QEPAGTPMFLLSRNTG  167
              +Q ++   ++++++  + ++ ++A+D D  N  N  I + + + +   P     F +   +G
Sbjct  1085  DRQKYVENVKQDASIGTNILRVSASDEDADN--NGAIVYTLSAPHSPNDLEYFEIQPESG  1142

Query  168   EV---RTLTNSLDREQASSYRLVVSGADKDGEGLSTQCECNIKVKDVNDNFPMFRDSQYS  224
              +    + L      RE+    YRL V  +D+ G+  ST            +N P   D
Sbjct  1143  WIVLKKPLDGCYSRER---YRLKVYASDR-GDPPSTADVDVELDVVDRNNKPPIWDRTIY  1198

Query  225   ARIEENILSSELLRFQVTDLDEEYTDNWLAVYFFTSG-----NEGNWFEIQTDPRTNE--  277
               I       +                 N    Y    G     N+ + F +Q  P   +
```

```
Sbjct  1199  GPIHIRENVTVGTVVTSVKASSGIEGNPTVFYRLMPGSTAQTNKLHTFYLQQRPDNGDTW  1258

Query  278   GILKVVKALDYEQLQSVKLSIAVKNKAEFHQSVISRYRVQSTPVTIQVINVREGIAFRPA  337
               +KV   LD+E ++   L++ V+N         ++       V IQ+ +V + I
Sbjct  1259  ADIKVNHPLDFETIKEYNLTVRVENNG-------AQQLASEATVYIQLEDVNDEIPLFTE  1311

Query  338   SKTFTVQKGISSKKLVDYILGTYQAIDEDTNKAASNVKYVM---GRNDG-GYLMIDSKTA  393
              +  TV +G      V  +     AID+D     + V Y +    RN+G  +  I+ ++
Sbjct  1312  REQETVLEGEPIGTKVTQV----NAIDKDGTFPNNQVYYYIVDSPRNEGKDFFEINLQSG  1367

Query  394   EI  395
             EI
Sbjct  1368  EI  1369

 Score = 55.8 bits (133),  Expect = 3e-06, Method: Compositional matrix adjust.
 Identities = 49/158 (31%), Positives = 78/158 (49%), Gaps = 24/158 (15%)

Query  102   NDNPPVFSQQIFMGEIEENSASNSLVMILNATDADEPNHLNSKIAFKIVSQEPAGTP--M  159
             ND  P FSQQ++   ++EN+  N+LV  + A+D D  N     + F  V     GT
Sbjct  858   NDEEPKFSQQVYTPNVDENAGPNTLVTTVVASDKDGDN-----VRFGFVG---GGTSSGQ  909

Query  160   FLLSRNTGEVRTLTN--SLDREQASSYRLVVSGADK-------DGEGLSTQCECNIKVKD  210
             F++   TG +R  +   SLDR++   Y L V+  D        D    ++     + + D
Sbjct  910   FVIEEITGVIRLHSKAISLDRDK---YELNVTAMDDGSCCVNGDATIHTSTAVVVVFITD  966

Query  211   VNDNFPMFRD-SQYSARIEENILS-SELLRFQVTDLDE  246
             VNDN P+F+D   Y  ++EE   + S +++ Q TD D+
Sbjct  967   VNDNKPVFKDCGTYYPKVEEGAPNGSPVIKVQATDEDK  1004

 Score = 53.9 bits (128),  Expect = 1e-05, Method: Compositional matrix adjust.
 Identities = 70/306 (23%), Positives = 126/306 (41%), Gaps = 30/306 (10%)

Query  20    PIAKIT-SDYQATQKITYRI---SGVGIDQPPFGIFVVDKNTGDINITAIVDRE---ETP  72
             PI  +T  D   T K  Y++   SG G D+  F +   +  TG + I   +D E   ++
Sbjct  1774  PILTVTVHDEDETNKFQYKVIDNSGYGADK--FTMVRNNDGTGSLKIVQPLDYEDPLQSN  1831

Query  73    SFLITCRALNAQGLDVEKPLI------LTVKILDINDNPPVFSQQIFMGEIEENSASNSL  126
              F    + +N +G D +          + VK+ DINDN P F +      + EN+
Sbjct  1832  GFRFRIQ-VNDKGEDNDNDKYHVAYSWVVVKLRDINDNKPQFERPNIEVSVYENADVGKT  1890

Query  127   VMILNATDADEPNHLNSKIAFKIVSQEPAGTPMFLLSRNTGEVRTLTNSLDREQASSYRL  186
             +    ATD D+     SK+++ I   + +       S N     T+  +LDRE    +++
Sbjct  1891  LETFKATDPDQGG--KSKVSYAI---DRSSDRQRQFSINQEGTVTIQRNLDREVTPRHQV  1945

Query  187   VVSGADKDGEGLSTQCECNIKVKDVNDNFPMFRDSQYSARIEENILSSELLRFQVTDLDE  246
              +   D      +      + V+D+NDN P F    Y   + E++   +++    TD D+
Sbjct  1946  KILAIDDGIPPKTATATLTVIVQDINDNPPKFL-KDYRPVLPEHVPPRKVVEILATDDDD  2004

Query  247   EYTDNWLAVYFFTSGNEGNW----FEIQTDPRTNEG----ILKVVKALDYEQLQSVKLSI  298
                 N     F       +     F+++ D +   G    I+  +++ D EQ +   + I
Sbjct  2005  RSKSNGPPFQFRLDPGADDIIRASFKVEQDQKGANGDGMAIVSSLRSFDREQQKEYLIPI  2064

Query  299   AVKNKA  304
              +K+
Sbjct  2065  VIKDHG  2070

 Score = 52.4 bits (124),  Expect = 4e-05, Method: Compositional matrix adjust.
 Identities = 97/398 (24%), Positives = 163/398 (41%), Gaps = 54/398 (14%)

Query  106   PVFSQQIFMGEIEENSASNSLVMILNATDADEPNHLNSKIAFKIVSQ-EPAGTPMFLLSR  164
             P F    +  EI EN   +S ++ + A      +  + +I + + +Q + AGT  F +
Sbjct  655   PQFYMPSYEAEIPENQKKDSDIISVKAK-----SFADREIRYTLKAQGQGAGT--FNIGP  707

Query  165   NTGEVRTLTNSLDRE---QASSYRLVVSGADKDGEGLSTQCECNIKVKDVNDNFPMFRDS  221
              +G V+ L   LD E   Q   Y LVV+ A +D  G ST  E  I+V DVNDN P F
Sbjct  708   TSGIVK-LAKELDFEDLRQPHVYSLVVT-ATEDSGGFSTSVELTIRVTDVNDNAPKFELP  765

Query  222   QYSAR-IEENI-LSSELLRFQVTDLDEEYTDNWLAVYFFTSGNEGNWFEIQTDPRTNEGI  279
```

```
                 Y A  ++E+I L + +L+ +  D D         + +  S +    F + ++     GI
Sbjct  766   DYQAHNVDEDIPLGTSILKVKAMDSDSGAN---AEIEYLVSDDH---FAVDSN-----GI  814

Query  280   LKVVKALDYEQLQS-VKLSIAVKNKAEFHQSVISRYRVQSTPVTIQVINVREGIAFRPAS  338
             +   K LD +    +  +  +  K+K E  ++  +  RV +            E   F
Sbjct  815   IVNNKQLDADNNNAYYEFVVTAKDKGEPAKTGTATVRVYTKNKN------DEEPKFSQQV  868

Query  339   KTFTVQKGISSKKLVDYILGTYQAIDEDTNKAASNVK--YVMGRNDGGYLMIDSKTAEIK  396
              T  V +      LV  ++          ++K   NV+  +V G    G  +I+  T  I+
Sbjct  869   YTPNVDENAGPNTLVTTVVA--------SDKDGDNVRFGFVGGGTSSGQFVIEEITGVIR  920

Query  397   FVK---NMNRDSTFI-VNKTITAEVLAIDEYTGKTSTGTVYVRVPDFNDN------CPTA  446
                   +++RD   + V             + T  TST  V V + D NDN      C T
Sbjct  921   LHSKAISLDRDKYELNVTAMDDGSCCVNGDATIHTSTAVVVVFITDVNDNKPVFKDCGTY  980

Query  447   VLE-KDAVCSSSPSVVVSARTLNNRYTGPYTFALEDQP  483
               + ++   + SP + V A   +    G   +++  QP
Sbjct  981   YPKVEEGAPNGSPVIKVQATDEDKGVNGQVKYSIVQQP  1018

 Score = 44.7 bits (104),  Expect = 0.008, Method: Compositional matrix adjust.
 Identities = 48/199 (24%), Positives = 73/199 (37%), Gaps = 19/199 (10%)

Query  27    DYQATQKITYRISGVGIDQPPFGIFVVDKNTGDINITAIVDREETPSFLITCRALNAQGL  86
             D     K++Y I      Q  F I       G + I   +DRE TP   +   A++
Sbjct  1900  DQGGKSKVSYAIDRSSDRQRQFSI----NQEGTVTIQRNLDREVTPRHQVKILAIDDGIP  1955

Query  87    DVEKPLILTVKILDINDNPPVFSQQIFMGEIEENSASNSLVMILNATDADEPNHLNSKIA  146
                    LTV + DINDNPP F +   +   + E+     +V IL   D D
Sbjct  1956  PKTATATLTVIVQDINDNPPKFLKD-YRPVLPEHVPPRKVVEILATDDDDRSKSNGPPFQ  2014

Query  147   FKIVSQEPAGTPMFLLSRNTGEVRTLTN-----------SLDREQASSYRLVVSGADKDG  195
             F++   +P    +   S    + +   N            S DREQ   Y + +   D
Sbjct  2015  FRL---DPGADDIIRASFKVEQDQKGANGDGMAIVSSLRSFDREQQKEYLIPIVIKDHGN  2071

Query  196   EGLSTQCECNIKVKDVNDN  214
               ++      + + DVNDN
Sbjct  2072  PAMTGTSTLTVVIGDVNDN  2090

 Score = 40.0 bits (92),  Expect = 0.21, Method: Compositional matrix adjust.
 Identities = 89/389 (23%), Positives = 149/389 (38%), Gaps = 62/389 (16%)

Query  75   LITCRALNAQGLDVEKPLILTVKILDINDNPPVFSQQIF--MGEIEENSASNSLVMILNA  132
            +IT +  NA GL       + + + D+ND PP F  +       ++ N+  N+ V  L A
Sbjct  517  IITNQGHNA-GLKYTDNQRVIIHVKDVNDEPPYFINRPLPMQAVVQLNAPPNTPVFTLQA  575

Query  133  TDADEPNHLNSKIAFKIVSQEPAGTPMFLLSRNTGEVRTLTNSLDREQASSYRLVVSGAD  192
             D D     +  I + IV     G   F +   +G VRT    L  +     Y L V   D
Sbjct  576  RDPD----TDHNIHYFIVRDRTGG--RFEVDERSGVVRTRGTDL-FQLDMEYVLYVKAED  628

Query  193  KDGE----GLSTQCECNIKVKDVNDNFPMFRDSQYSARIEEN-ILSSELLRFQVTDLDEE  247
            ++G+        +  E  + +        P F    Y A I EN    S+++  +     +
Sbjct  629  QNGKVDDRRFQSTPEERLSIVG-GKRAPQFYMPSYEAEIPENQKKDSDIISVKA----KS  683

Query  248  YTDNWLAVYFFTSGNEGNWFEIQTDPRTNEGILKVVKALDYEQLQSVKLS----IAVKNK  303
            + D  +        G     F I          GI+K+ K LD+E L+   +       A ++
Sbjct  684  FADREIRYTLKAQGQGAGTFNIGP----TSGIVKLAKELDFEDLRQPHVYSLVVTATEDS  739

Query  304  AEFHQSVISRYRVQSTPVTIQVINVREGIAFRPASKTFTVQKGISSKKLVDYILGT----  359
              F  SV           +TI+V +V +        +  F +    +     D  LGT
Sbjct  740  GGFSTSV---------ELTIRVTDVNDN------APKFELPDYQAHNVDEDIPLGTSILK  784

Query  360  YQAIDEDTNKAASNVKYVMGRN----DGGYLMIDSKTAEIKFVKNMNRDSTFIVNKTITA  415
             +A+D D+  A + ++Y++  +     D    +++++K  +     N N    F+V
Sbjct  785  VKAMDSDSG-ANAEIEYLVSDDHFAVDSNGIIVNNKQLD---ADNNNAYYEFVVTAKDKG  840

Query  416  EVLAIDEYTGKTSTGTVYVRVPDFNDNCP  444
            E          KT T TV V   + ND  P
Sbjct  841  E-------PAKTGTATVRVYTKNKNDEEP  862
```

```
 Score = 39.7 bits (91),  Expect = 0.25, Method: Compositional matrix adjust.
 Identities = 54/242 (22%), Positives = 102/242 (42%), Gaps = 22/242 (9%)

Query  4    KFAKPCREGE--DNSKRNP-IAKITSDYQATQKITYRISGVGIDQPPFGIFVVDKNTGDI  60
            +F  P  E E  +N K++  I  + +   A ++I Y +   G      G F +   +G +
Sbjct  656  QFYMPSYEAEIPENQKKDSDIISVKAKSFADREIRYTLKAQGQGA---GTFNIGPTSGIV  712

Query  61   NITAIVDREE--TPSF--LITCRALNAQGLDVEKPLILTVKILDINDNPPVFSQQIFMGE  116
             +   +D E+    P    L+      ++ G       + LT+++ D+NDN P F    +
Sbjct  713  KLAKELDFEDLRQPHVYSLVVTATEDSGGFSTS--VELTIRVTDVNDNAPKFELPDYQAH  770

Query  117  -IEENSASNSLVMILNATDADEPNHLNSKIAFKIVSQEPAGTPMFLLSRNTGEVRTLTNS  175
             ++E+    + ++ + A D+D  +  N++I + +VS +      F +  N   V
Sbjct  771  NVDEDIPLGTSILKVKAMDSD--SGANAEIEY-LVSDD-----HFAVDSNGIIVNNKQLD  822

Query  176  LDREQASSYRLVVSGADKDGEGLSTQCECNIKVKDVNDNFPMFRDSQYSARIEENILSSE  235
             D   A  Y  VV+  DK      +       +  K+ ND  P F    Y+  ++EN   +
Sbjct  823  ADNNNA-YYEFVVTAKDKGEPAKTGTATVRVYTKNKNDEEPKFSQQVYTPNVDENAGPNT  881

Query  236  LL  237
            L+
Sbjct  882  LV  883

>AARA21_009090.P17033 | transcript=AARA21_009090.R17033 | gene=AARA21_009090
| organism=Anopheles_arabiensis_DONGOLA_2021
| gene_product=neural-cadherin, transcript variant X5 |
transcript_product=neural-cadherin,
transcript variant X1 | location=CM029353:40466514-40666848(+)
| protein_length=3105 | sequence_SO=chromosome
| SO=protein_coding_gene | is_pseudo=false
Length=3105

 Score = 144 bits (364),  Expect = 1e-34, Method: Compositional matrix adjust.
 Identities = 125/437 (29%), Positives = 211/437 (48%), Gaps = 33/437 (8%)

Query  26    SDYQATQKITYRISGVGID--QPPFGIFVVDKNTGDINITAIVDREET---PSFLITCRA  80
              D    Q I Y ++G GID   P    F +++ TG+I +   +DR++    P +  T  A
Sbjct  1554  GDKDRPQNIVYFLTGQGIDPDNPANSKFDINRTTGEIFVLKPLDRDQPNGRPQWRFTVFA  1613

Query  81    LNAQGLDVEKPLILTVKILDINDNPPVFSQQIFMGEIEENSASNSLVMILNATDADEPNH  140
              +  G  +     + V + DINDN P+F Q ++ G + EN  +  +VM + A D D+PN
Sbjct  1614  QDEGGEGLVGYADVQVNLKDINDNAPIFPQGVYFGNVTENGTAGMVVMTMTAVDYDDPNE  1673

Query  141   -LNSKIAFKI---VSQEPAGTPMFLLSRNTGEVRTLTNSLDREQASSYRLVVSGADKDGE  196
                N+K+ + I   V +E  G+P+F +  +TG ++T    LDRE+   Y + V     DG
Sbjct  1674  GTNAKLIYSIEKNVIEEETGSPIFEIEADTGVIKTAVCCLDRERTPDYSIQVVAM--DGG  1731

Query  197   GLSTQCECNIKVKDVNDNFPMFRDSQYSARIEENILSS--ELLRFQVTDLDEEYTDNWLA  254
             GL      +I+VKD+ND  P F   ++   ++E   ++  E+    VT  DE+ T+ +
Sbjct  1732  GLKGTGTASIRVKDINDMPPQFTKDEWFTEVDETDGTNLPEMPILTVTVHDEDETNKFQY  1791

Query  255   VYFFTSGNEGNWFEIQTDPRTNEGI--LKVVKALDYE---QLQSVKLSIAVKNKAEFHQS  309
                  SG   + F +    R N+G   LK+V+ LDYE   Q    +  I V +K E + +
Sbjct  1792  KVIDNSGYGADKFTMV---RNNDGTGSLKIVQPLDYEDPLQSNGFRFRIQVNDKGEDNDN  1848

Query  310   VISRYRVQSTPVTIQVINVREGIAFRPASKTFTVQKGISSKKLVDYILGTYQAIDEDTNK  369
                +Y V  + V +++ ++ +    +P  +   ++  +     V   L T++A D D
Sbjct  1849  --DKYHVAYSWVVVKLRDINDN---KPQFERPNIEVSVYENADVGKTLETFKATDPDQG-  1902

Query  370   AASNVKYVMGR-NDGGYLMIDSKTAEIKFVKNMNRDSTFIVNKTITAEVLAIDE-YTGKT  427
               S V Y + R +D        ++   +   +N++R+    V       ++LAID+    KT
Sbjct  1903  GKSKVSYAIDRSSDRQRQFSINQEGTVTIQRNLDRE----VTPRHQVKILAIDDGIPPKT  1958

Query  428   STGTVYYVRVPDFNDNCP  444
             +T T+ V V D NDN P
Sbjct  1959  ATATLTVIVQDINDNPP  1975

 Score = 85.9 bits (211),  Expect = 1e-15, Method: Compositional matrix adjust.
```

```
 Identities = 108/420 (26%), Positives = 177/420 (42%), Gaps = 52/420 (12%)

Query  50    IFVVDKNTGDINITAIVDREETPSFLITCRAL----------NAQGLDVEKPLILTVKIL  99
              F ++  +G+I    + DRE+  ++ +   A           N Q   V K   + + I
Sbjct  1359  FFEINLQSGEIFTKVVFDREKQGAYALEVEARDGAPSARPNSNQQPNSVTK--FIRIGIA  1416

Query  100   DINDNPPVFSQQIFMGEIEENSASNSLVMILNATDADEPNHLNSKIAFKIVSQEPAGTPM  159
             D NDNPP F + ++  E++EN      V+ + A D DE    +S+I ++I S    G
Sbjct  1417  DKNDNPPYFDKALYEAEVDENEDIQHTVLTVTAKDHDE----SSRIRYEITSGNIGGA--  1470

Query  160   FLLSRNTGEVRTLTNSLDREQASSYRLVVSGADKDGEGLSTQCECNIKVKDVNDNFPMFR  219
             F +   TG +  +  +LD E    Y L ++ +D   E  +T     I VKDVNDN P+F
Sbjct  1471  FAVKNMTGAI-YVAGALDYETRKRYELRLAASDNLKENYTTVV---IHVKDVNDNPPVFE  1526

Query  220   DSQYSARIEENI---LSSELLRFQVTDLDEEYTDNWLAVYFFTSG--NEGNWFEIQTDPR  274
                Y  +I E     L   +L+   TD D++    N   VYF T    +  N    + D
Sbjct  1527  RPTYRTQITEEDDRNLPKRVLQVTATDGDKDRPQN--IVYFLTGQGIDPDNPANSKFDIN  1584

Query  275   TNEGILKVVKALDYEQLQS-VKLSIAVKNKAEFHQSVISRYRVQSTPVTIQVINVREGIA  333
                G + V+K LD +Q     +    V  + E  + ++    VQ     + + ++ +
Sbjct  1585  RTTGEIFVLKPLDRDQPNGRPQWRFTVFAQDEGGEGLVGYADVQ-----VNLKDINDNAP  1639

Query  334   FRPASKTFTVQKGISSKKLVDYILGTYQAIDEDTNKAASNVK--YVMGRN------DGGY  385
               P    F     ++        ++ T  A+D D     +N K  Y + +N
Sbjct  1640  IFPQGVYF---GNVTENGTAGMVVMTMTAVDYDDPNEGTNAKLIYSIEKNVIEEETGSPI  1696

Query  386   LMIDSKTAEIK-FVKNMNRDSTFIVNKTITAEVLAIDEYTGKTSTGTVYVRVPDFNDNCP  444
               I++ T  IK  V  ++R+ T       + +V+A+D   G   TGT  +RV D ND  P
Sbjct  1697  FEIEADTGVIKTAVCCLDRERT----PDYSIQVVAMDG-GGLKGTGTASIRVKDINDMPP  1751

 Score = 76.3 bits (186),  Expect = 1e-12, Method: Compositional matrix adjust.
 Identities = 106/426 (25%), Positives = 181/426 (42%), Gaps = 51/426 (12%)

Query  53    VDKNTGDINITAIVDREETPSFLITCRALNAQGLDVEKPLILTVKILDINDNPPVFSQ-Q  111
             +D++  ++N+TA+ D         +C   +N           ++ V I D+NDN PVF
Sbjct  928   LDRDKYELNVTAMDDG--------SC-CVNGDATIHTSTAVVVVFITDVNDNKPVFKDCG  978

Query  112   IFMGEIEENSASNSLVMILNATDADEPNHLNSKIAFKIVSQEPAGTPMFLLSRNTGEVRT  171
              +  ++EE + + S V+ + ATD D+   +N ++ + IV Q       F +   TGEV T
Sbjct  979   TYYPKVEEGAPNGSPVIKVQATDEDK--GVNGQVKYSIVQQPNQKGTKFTVDEETGEVST  1036

Query  172   LTNSLDRE--QASSYRLVVSGADKDGEGLSTQCECNIKVKDVNDNFPMFRDSQYSARIEE  229
                  DRE        + V   D+     L   C   +++ DVNDN P+F   +Y   +++
Sbjct  1037  -NKVFDREGDDGKFVSVTVKATDQGEPSLEGVCSFTVEITDVNDNPPLFDRQKYVENVKQ  1095

Query  230   NI-LSSELLRFQVTDLDEEYTDNWLAVYFFT---SGNEGNWFEIQTDPRTNEGILKVVKA  285
             +  + + +LR  V+  DE+  +N   VY  +   S N+  +FEIQ  P +   +LK  K
Sbjct  1096  DASIGTNILR--VSASDEDADNNGAIVYTLSAPHSPNDLEYFEIQ--PESGWIVLK--KP  1149

Query  286   LD--YEQLQSVKLSIAVKNK------AEFHQSVISRYRVQSTPVTIQV----INVREGIA  333
             LD  Y + +  +L +   ++      A+    +    R    P+  +     I++RE +
Sbjct  1150  LDGCYSR-ERYRLKVYASDRGDPPSTADVDVELDVVDRNNKPPIWDRTIYGPIHIRENVT  1208

Query  334   FRPASKTFTVQKGISSKKLVDYILGTYQAIDEDTNKAASNVKYVMGRNDGGYLMIDSKTA  393
                   +     GI     V Y L         TNK   +  Y+  R D G        A
Sbjct  1209  VGTVVTSVKASSGIEGNPTVFYRL--MPGSTAQTNKL--HTFYLQQRPDNG-----DTWA  1259

Query  394   EIKFVKNMNRDSTFIVNKTITAEVLAIDEYTGKTSTGTVYVRVPDFNDNCPTAV-LEKDA  452
             +IK    ++ ++    N T+  E     +     S  TVY+++ D ND  P     E++
Sbjct  1260  DIKVNHPLDFETIKEYNLTVRVENNGAQQL---ASEATVYIQLEDVNDEIPLFTEREQET  1316

Query  453   VCSSSP  458
             V    P
Sbjct  1317  VLEGEP  1322

 Score = 60.8 bits (146),  Expect = 9e-08, Method: Compositional matrix adjust.
 Identities = 83/362 (23%), Positives = 152/362 (42%), Gaps = 34/362 (9%)

Query  51    FVVDKNTGDINITAIVDRE-ETPSFL-ITCRALNAQGLDVEKPLILTVKILDINDNPPVF  108
```

```
             F VD+ TG+++   + DRE +   F+ +T +A +     +E     TV+I D+NDNPP+F
Sbjct  1025  FTVDEETGEVSTNKVFDREGDDGKFVSVTVKATDQGEPSLEGVCSFTVEITDVNDNPPLF  1084

Query  109   SQQIFMGEIEENSASNSLVMILNATDADEPNHLNSKIAFKIVS-QEPAGTPMFLLSRNTG  167
              +Q ++  ++++++  + ++ ++A+D D  N  N  I + + +   P     F +   +G
Sbjct  1085  DRQKYVENVKQDASIGTNILRVSASDEDADN--NGAIVYTLSAPHSPNDLEYFEIQPESG  1142

Query  168   EV---RTLTNSLDREQASSYRLVVSGADKDGEGLSTQCECNIKVKDVNDNFPMFRDSQYS  224
              +   + L     RE+   YRL V  +D+ G+  ST            +N P   D
Sbjct  1143  WIVLKKPLDGCYSRER---YRLKVYASDR-GDPPSTADVDVELDVVDRNNKPPIWDRTIY  1198

Query  225   ARIEENILSSELLRFQVTDLDEEYTDNWLAVYFFTSG-----NEGNWFEIQTDPRTNE--  277
               I      +                N    Y    G     N+ + F +Q  P   +
Sbjct  1199  GPIHIRENVTVGTVVTSVKASSGIEGNPTVFYRLMPGSTAQTNKLHTFYLQQRPDNGDTW  1258

Query  278   GILKVVKALDYEQLQSVKLSIAVKNKAEFHQSVISRYRVQSTPVTIQVINVREGIAFRPA  337
               +KV   LD+E ++   L++ V+N         ++       V IQ+ +V + I
Sbjct  1259  ADIKVNHPLDFETIKEYNLTVRVENNG-------AQQLASEATVYIQLEDVNDEIPLFTE  1311

Query  338   SKTFTVQKGISSKKLVDYILGTYQAIDEDTNKAASNVKYVM---GRNDG-GYLMIDSKTA  393
              +  TV +G      V  +     AID+D     + V Y +    RN+G  +  I+ ++
Sbjct  1312  REQETVLEGEPIGTKVTQV----NAIDKDGTFPNNQVYYYIVDSPRNEGKDFFEINLQSG  1367

Query  394   EI  395
             EI
Sbjct  1368  EI  1369

 Score = 55.8 bits (133),  Expect = 3e-06, Method: Compositional matrix adjust.
 Identities = 49/158 (31%), Positives = 78/158 (49%), Gaps = 24/158 (15%)

Query  102   NDNPPVFSQQIFMGEIEENSASNSLVMILNATDADEPNHLNSKIAFKIVSQEPAGTP--M  159
             ND  P FSQQ++   ++EN+  N+LV  + A+D D  N     + F  V     GT
Sbjct  858   NDEEPKFSQQVYTPNVDENAGPNTLVTTVVASDKDGDN-----VRFGFVG---GGTSSGQ  909

Query  160   FLLSRNTGEVRTLTN--SLDREQASSYRLVVSGADK-------DGEGLSTQCECNIKVKD  210
             F++   TG +R  +   SLDR++   Y L V+  D        D    ++     + + D
Sbjct  910   FVIEEITGVIRLHSKAISLDRDK---YELNVTAMDDGSCCVNGDATIHTSTAVVVVFITD  966

Query  211   VNDNFPMFRD-SQYSARIEENILS-SELLRFQVTDLDE  246
             VNDN P+F+D   Y  ++EE   + S +++ Q TD D+
Sbjct  967   VNDNKPVFKDCGTYYPKVEEGAPNGSPVIKVQATDEDK  1004

 Score = 53.9 bits (128),  Expect = 1e-05, Method: Compositional matrix adjust.
 Identities = 70/306 (23%), Positives = 126/306 (41%), Gaps = 30/306 (10%)

Query  20    PIAKIT-SDYQATQKITYRI---SGVGIDQPPFGIFVVDKNTGDINITAIVDRE---ETP  72
             PI  +T  D   T K  Y++   SG G D+  F +   +  TG + I   +D E   ++
Sbjct  1774  PILTVTVHDEDETNKFQYKVIDNSGYGADK--FTMVRNNDGTGSLKIVQPLDYEDPLQSN  1831

Query  73    SFLITCRALNAQGLDVEKPLI------LTVKILDINDNPPVFSQQIFMGEIEENSASNSL  126
              F    + +N +G D +          + VK+ DINDN P F +      + EN+
Sbjct  1832  GFRFRIQ-VNDKGEDNDNDKYHVAYSWVVVKLRDINDNKPQFERPNIEVSVYENADVGKT  1890

Query  127   VMILNATDADEPNHLNSKIAFKIVSQEPAGTPMFLLSRNTGEVRTLTNSLDREQASSYRL  186
             +    ATD D+     SK+++ I   + +       S N     T+  +LDRE    +++
Sbjct  1891  LETFKATDPDQGG--KSKVSYAI---DRSSDRQRQFSINQEGTVTIQRNLDREVTPRHQV  1945

Query  187   VVSGADKDGEGLSTQCECNIKVKDVNDNFPMFRDSQYSARIEENILSSELLRFQVTDLDE  246
              +   D      +      + V+D+NDN P F    Y   + E++   +++    TD D+
Sbjct  1946  KILAIDDGIPPKTATATLTVIVQDINDNPPKFL-KDYRPVLPEHVPPRKVVEILATDDDD  2004

Query  247   EYTDNWLAVYFFTSGNEGNW----FEIQTDPRTNEG----ILKVVKALDYEQLQSVKLSI  298
                 N     F       +     F+++ D +   G    I+  +++ D EQ +   + I
Sbjct  2005  RSKSNGPPFQFRLDPGADDIIRASFKVEQDQKGANGDGMAIVSSLRSFDREQQKEYLIPI  2064

Query  299   AVKNKA  304
              +K+
Sbjct  2065  VIKDHG  2070
```

```
 Score = 52.4 bits (124),  Expect = 4e-05, Method: Compositional matrix adjust.
 Identities = 97/398 (24%), Positives = 163/398 (41%), Gaps = 54/398 (14%)

Query  106   PVFSQQIFMGEIEENSASNSLVMILNATDADEPNHLNSKIAFKIVSQ-EPAGTPMFLLSR  164
             P F    +  EI EN   +S ++ + A       +  + +I + + +Q + AGT  F +
Sbjct  655   PQFYMPSYEAEIPENQKKDSDIISVKAK-----SFADREIRYTLKAQGQGAGT--FNIGP  707

Query  165   NTGEVRTLTNSLDRE---QASSYRLVVSGADKDGEGLSTQCECNIKVKDVNDNFPMFRDS  221
              +G V+ L   LD E   Q   Y LVV+ A +D  G ST  E  I+V DVNDN P F
Sbjct  708   TSGIVK-LAKELDFEDLRQPHVYSLVVT-ATEDSGGFSTSVELTIRVTDVNDNAPKFELP  765

Query  222   QYSAR-IEENI-LSSELLRFQVTDLDEEYTDNWLAVYFFTSGNEGNWFEIQTDPRTNEGI  279
              Y A  ++E+I L + +L+ +  D D          + +  S +    F + ++     GI
Sbjct  766   DYQAHNVDEDIPLGTSILKVKAMDSDSGAN---AEIEYLVSDDH---FAVDSN-----GI  814

Query  280   LKVVKALDYEQLQS-VKLSIAVKNKAEFHQSVISRYRVQSTPVTIQVINVREGIAFRPAS  338
             +   K LD +   +  +  +  K+K E  ++  +  RV +             E   F
Sbjct  815   IVNNKQLDADNNNAYYEFVVTAKDKGEPAKTGTATVRVYTKNKN------DEEPKFSQQV  868

Query  339   KTFTVQKGISSKKLVDYILGTYQAIDEDTNKAASNVK--YVMGRNDGGYLMIDSKTAEIK  396
              T  V +      LV  ++          ++K   NV+  +V G    G  +I+  T  I+
Sbjct  869   YTPNVDENAGPNTLVTTVVA--------SDKDGDNVRFGFVGGGTSSGQFVIEEITGVIR  920

Query  397   FVK---NMNRDSTFI-VNKTITAEVLAIDEYTGKTSTGTVYVRVPDFNDN------CPTA  446
                   +++RD   + V              + T  TST  V V + D NDN      C T
Sbjct  921   LHSKAISLDRDKYELNVTAMDDGSCCVNGDATIHTSTAVVVVFITDVNDNKPVFKDCGTY  980

Query  447   VLE-KDAVCSSSPSVVVSARTLNNRYTGPYTFALEDQP  483
               + ++   + SP + V A   +    G   +++  QP
Sbjct  981   YPKVEEGAPNGSPVIKVQATDEDKGVNGQVKYSIVQQP  1018

 Score = 44.7 bits (104),  Expect = 0.008, Method: Compositional matrix adjust.
 Identities = 48/199 (24%), Positives = 73/199 (37%), Gaps = 19/199 (10%)

Query  27    DYQATQKITYRISGVGIDQPPFGIFVVDKNTGDINITAIVDREETPSFLITCRALNAQGL  86
             D     K++Y I      Q  F I        G + I   +DRE TP   +   A++
Sbjct  1900  DQGGKSKVSYAIDRSSDRQRQFSI----NQEGTVTIQRNLDREVTPRHQVKILAIDDGIP  1955

Query  87    DVEKPLILTVKILDINDNPPVFSQQIFMGEIEENSASNSLVMILNATDADEPNHLNSKIA  146
                    LTV + DINDNPP F +  +    + E+     +V IL   D D
Sbjct  1956  PKTATATLTVIVQDINDNPPKFLKD-YRPVLPEHVPPRKVVEILATDDDDRSKSNGPPFQ  2014

Query  147   FKIVSQEPAGTPMFLLSRNTGEVRTLTN-----------SLDREQASSYRLVVSGADKDG  195
             F++   +P     +   S    + +   N           S DREQ   Y + +   D
Sbjct  2015  FRL---DPGADDIIRASFKVEQDQKGANGDGMAIVSSLRSFDREQQKEYLIPIVIKDHGN  2071

Query  196   EGLSTQCECNIKVKDVNDN  214
               ++      + + DVNDN
Sbjct  2072  PAMTGTSTLTVVIGDVNDN  2090

 Score = 40.0 bits (92),  Expect = 0.21, Method: Compositional matrix adjust.
 Identities = 89/389 (23%), Positives = 149/389 (38%), Gaps = 62/389 (16%)

Query  75   LITCRALNAQGLDVEKPLILTVKILDINDNPPVFSQQIF--MGEIEENSASNSLVMILNA  132
            +IT +  NA GL       + + + D+ND PP F  +       ++ N+  N+ V  L A
Sbjct  517  IITNQGHNA-GLKYTDNQRVIIHVKDVNDEPPYFINRPLPMQAVVQLNAPPNTPVFTLQA  575

Query  133  TDADEPNHLNSKIAFKIVSQEPAGTPMFLLSRNTGEVRTLTNSLDREQASSYRLVVSGAD  192
             D D     +  I + IV     G   F +   +G VRT    L  +     Y L V  D
Sbjct  576  RDPD----TDHNIHYFIVRDRTGG--RFEVDERSGVVRTRGTDL-FQLDMEYVLYVKAED  628

Query  193  KDGE----GLSTQCECNIKVKDVNDNFPMFRDSQYSARIEEN-ILSSELLRFQVTDLDEE  247
            ++G+        +  E  + +        P F    Y A I EN    S+++  +      +
Sbjct  629  QNGKVDDRRFQSTPEERLSIVG-GKRAPQFYMPSYEAEIPENQKKDSDIISVKA----KS  683

Query  248  YTDNWLAVYFFTSGNEGNWFEIQTDPRTNEGILKVVKALDYEQLQSVKLS----IAVKNK  303
            + D  +        G     F I         GI+K+ K LD+E L+   +      A ++
Sbjct  684  FADREIRYTLKAQGQGAGTFNIGP----TSGIVKLAKELDFEDLRQPHVYSLVVTATEDS  739
```

```
Query  304  AEFHQSVISRYRVQSTPVTIQVINVREGIAFRPASKTFTVQKGISSKKLVDYILGT----  359
              F  SV           +TI+V +V +        +  F +     +     D  LGT
Sbjct  740  GGFSTSV---------ELTIRVTDVNDN------APKFELPDYQAHNVDEDIPLGTSILK  784

Query  360  YQAIDEDTNKAASNVKYVMGRN----DGGYLMIDSKTAEIKFVKNMNRDSTFIVNKTITA  415
             +A+D D+  A + ++Y++  +    D   +++++K  +     N N    F+V
Sbjct  785  VKAMDSDSG-ANAEIEYLVSDDHFAVDSNGIIVNNKQLD---ADNNNAYYEFVVTAKDKG  840

Query  416  EVLAIDEYTGKTSTGTVYVRVPDFNDNCP  444
            E          KT T TV V   + ND  P
Sbjct  841  E-------PAKTGTATVRVYTKNKNDEEP  862

 Score = 39.7 bits (91),  Expect = 0.25, Method: Compositional matrix adjust.
 Identities = 54/242 (22%), Positives = 102/242 (42%), Gaps = 22/242 (9%)

Query  4    KFAKPCREGE--DNSKRNP-IAKITSDYQATQKITYRISGVGIDQPPFGIFVVDKNTGDI  60
            +F  P  E E  +N K++  I  + +   A ++I Y +   G       G F +   +G +
Sbjct  656  QFYMPSYEAEIPENQKKDSDIISVKAKSFADREIRYTLKAQGQGA---GTFNIGPTSGIV  712

Query  61   NITAIVDREE--TPSF--LITCRALNAQGLDVEKPLILTVKILDINDNPPVFSQQIFMGE  116
             +   +D E+   P    L+      ++ G       + LT+++ D+NDN P F    +
Sbjct  713  KLAKELDFEDLRQPHVYSLVVTATEDSGGFSTS--VELTIRVTDVNDNAPKFELPDYQAH  770

Query  117  -IEENSASNSLVMILNATDADEPNHLNSKIAFKIVSQEPAGTPMFLLSRNTGEVRTLTNS  175
             ++E+    + ++ + A D+D  +  N++I + +VS +       F +  N   V
Sbjct  771  NVDEDIPLGTSILKVKAMDSD--SGANAEIEY-LVSDD-----HFAVDSNGIIVNNKQLD  822

Query  176  LDREQASSYRLVVSGADKDGEGLSTQCECNIKVKDVNDNFPMFRDSQYSARIEENILSSE  235
             D   A  Y  VV+  DK      +       +  K+ ND  P F     Y+  ++EN   +
Sbjct  823  ADNNNA-YYEFVVTAKDKGEPAKTGTATVRVYTKNKNDEEPKFSQQVYTPNVDENAGPNT  881

Query  236  LL  237
            L+
Sbjct  882  LV  883

>AALB20_033171.P66528 | transcript=AALB20_033171.R66528 | gene=AALB20_033171
| organism=Anopheles_albimanus_STECLA_2020 |
gene_product=neural-cadherin, transcript variant X38 |
transcript_product=neural-cadherin,
transcript variant X19 | location=CM024292:75037560-75208393(+)
| protein_length=3131 | sequence_SO=chromosome
| SO=protein_coding_gene | is_pseudo=false
Length=3131

 Score = 144 bits (364),  Expect = 1e-34, Method: Compositional matrix adjust.
 Identities = 125/437 (29%), Positives = 211/437 (48%), Gaps = 33/437 (8%)

Query  26    SDYQATQKITYRISGVGID--QPPFGIFVVDKNTGDINITAIVDREET---PSFLITCRA  80
               D    Q I Y ++G GID    P    F +++ TG+I +    +DR++     P +  T  A
Sbjct  1580  GDKDRPQNIVYFLTGQGIDPDNPANSKFDINRTTGEIFVLKPLDRDQPNGRPQWRFTVFA  1639

Query  81    LNAQGLDVEKPLILTVKILDINDNPPVFSQQIFMGEIEENSASNSLVMILNATDADEPNH  140
               +  G  +      + V + DINDN P+F Q ++ G + EN  +  +VM + A D D+PN
Sbjct  1640  QDEGGEGLVGYADVQVNLKDINDNAPIFPQGVYFGNVTENGTAGMVVMTMTAVDYDDPNE  1699

Query  141   -LNSKIAFKI---VSQEPAGTPMFLLSRNTGEVRTLTNSLDREQASSYRLVVSGADKDGE  196
                 N+K+ + I   V +E  G+P+F +  +TG ++T     LDRE+   Y + V      DG
Sbjct  1700  GTNAKLIYSIEKNVIEEETGSPIFEIEADTGVIKTAVCCLDRERTPDYSIQVVAM--DGG  1757

Query  197   GLSTQCECNIKVKDVNDNFPMFRDSQYSARIEENILSS--ELLRFQVTDLDEEYTDNWLA  254
             GL       +I+VKD+ND  P F   ++    ++E   ++  E+    VT  DE+ T+ +
Sbjct  1758  GLKGTGTASIRVKDINDMPPQFTKDEWFTEVDETDGTNLPEMPILTVTVHDEDETNKFQY  1817

Query  255   VYFFTSGNEGNWFEIQTDPRTNEGI--LKVVKALDYE---QLQSVKLSIAVKNKAEFHQS  309
                    SG    + F +     R N+G   LK+V+ LDYE   Q     +  I V +K E + +
Sbjct  1818  KVIDNSGYGADKFTMV---RNNDGTGSLKIVQPLDYEDPLQSNGFRFRIQVNDKGEDNDN  1874

Query  310   VISRYRVQSTPVTIQVINVREGIAFRPASKTFTVQKGISSKKLVDYILGTYQAIDEDTNK  369
```

```
                +Y V  + V +++ ++ +    +P  +   ++  +     V   L T++A D D
Sbjct  1875  --DKYHVAYSWVVVKLRDINDN---KPQFERPNIEVSVYENADVGKTLETFKATDPDQG-  1928

Query  370   AASNVKYVMGR-NDGGYLMIDSKTAEIKFVKNMNRDSTFIVNKTITAEVLAIDE-YTGKT  427
               S V Y + R +D       ++   +   +N++R+    V      ++LAID+    KT
Sbjct  1929  GKSKVSYAIDRSSDRQRQFSINQEGTVTIQRNLDRE----VTPRHQVKILAIDDGIPPKT  1984

Query  428   STGTVYVRVPDFNDNCP  444
             +T T+ V V D NDN P
Sbjct  1985  ATATLTVIVQDINDNPP  2001

 Score = 85.9 bits (211),  Expect = 1e-15, Method: Compositional matrix adjust.
 Identities = 108/420 (26%), Positives = 177/420 (42%), Gaps = 52/420 (12%)

Query  50    IFVVDKNTGDINITAIVDREETPSFLITCRAL----------NAQGLDVEKPLILTVKIL  99
              F ++  +G+I    + DRE+  ++ + A           N Q   V K   + + I
Sbjct  1385  FFEINLQSGEIFTKVVFDREKQGAYALEVEARDGAPSARPNSNQQPNSVTK--FIRIGIA  1442

Query  100   DINDNPPVFSQQIFMGEIEENSASNSLVMILNATDADEPNHLNSKIAFKIVSQEPAGTPM  159
             D NDNPP F + ++  E++EN      V+ + A D DE    +S+I ++I S    G
Sbjct  1443  DKNDNPPYFDKALYEAEVDENEDIQHTVLTVTAKDHDE----SSRIRYEITSGNIGGA--  1496

Query  160   FLLSRNTGEVRTLTNSLDREQASSYRLVVSGADKDGEGLSTQCECNIKVKDVNDNFPMFR  219
             F +   TG +  +  +LD E    Y L ++ +D   E  +T     I VKDVNDN P+F
Sbjct  1497  FAVKNMTGAI-YVAGALDYETRKRYELRLAASDNLKENYTTVV---IHVKDVNDNPPVFE  1552

Query  220   DSQYSARIEENI---LSSELLRFQVTDLDEEYTDNWLAVYFFTSG--NEGNWFEIQTDPR  274
                Y  +I E     L   +L+   TD D++   N   VYF T    +  N    + D
Sbjct  1553  RPTYRTQITEEDDRNLPKRVLQVTATDGDKDRPQN--IVYFLTGQGIDPDNPANSKFDIN  1610

Query  275   TNEGILKVVKALDYEQLQS-VKLSIAVKNKAEFHQSVISRYRVQSTPVTIQVINVREGIA  333
                G + V+K LD +Q     +    V  + E  + ++    VQ     + + ++ +
Sbjct  1611  RTTGEIFVLKPLDRDQPNGRPQWRFTVFAQDEGGEGLVGYADVQ-----VNLKDINDNAP  1665

Query  334   FRPASKTFTVQKGISSKKLVDYILGTYQAIDEDTNKAASNVK--YVMGRN------DGGY  385
               P    F     ++       ++ T  A+D D     +N K  Y + +N
Sbjct  1666  IFPQGVYF---GNVTENGTAGMVVMTMTAVDYDDPNEGTNAKLIYSIEKNVIEEETGSPI  1722

Query  386   LMIDSKTAEIK-FVKNMNRDSTFIVNKTITAEVLAIDEYTGKTSTGTVYVRVPDFNDNCP  444
               I++ T  IK  V  ++R+ T       + +V+A+D   G   TGT  +RV D ND  P
Sbjct  1723  FEIEADTGVIKTAVCCLDRERT----PDYSIQVVAMDG-GGLKGTGTASIRVKDINDMPP  1777

 Score = 75.9 bits (185),  Expect = 2e-12, Method: Compositional matrix adjust.
 Identities = 108/429 (25%), Positives = 178/429 (41%), Gaps = 53/429 (12%)

Query  53    VDKNTGDINITAIVDREETPSFLITCRALNAQGLDVEKPLILTVKILDINDNPPVFSQ-Q  111
             +D++  ++N+TA+ D         +C  +N          ++ V I D+NDN PVF
Sbjct  950   LDRDKYELNVTAMDDG--------SC-CVNGDATIHTSTAVVVVFITDVNDNKPVFKDCG  1000

Query  112   IFMGEIEENSASNSLVMILNATDADEPNHLNSKIAFKIVSQEPAGTPMFLLSRNTGEVRT  171
              +  ++EE + + S V+ + ATD D+   +N ++ + IV Q       F +   TGEV T
Sbjct  1001  TYYPKVEEGAPNGSPVIKVQATDEDK--GVNGQVKYSIVQQPNQKGTKFTVDEETGEVST  1058

Query  172   LTNSLDRE--QASSYRLVVSGADKDGEGLSTQCECNIKVKDVNDNFPMFRDSQYSARIEE  229
                  DRE        + V   D+     L   C   +++ DVNDN P+F   +Y   +++
Sbjct  1059  -NKVFDREGDDGKFVSVTVKATDQGEPSLEGVCSFTVEITDVNDNPPLFDRQKYVENVKQ  1117

Query  230   NI-LSSELLRFQVTDLDEEYTDNWLAVYFFT---SGNEGNWFEIQTDPRTNEGILKVVKA  285
             +  + + +LR  V+  DE+  +N   VY  +   S N+  +FEIQ  P +   +LK  K
Sbjct  1118  DASIGTNILR--VSASDEDADNNGAIVYTLSAPHSPNDLEYFEIQ--PESGWIVLK--KP  1171

Query  286   LD-YEQLQSVKLSIAVK----------NKAEFHQSVISRYRVQSTPV----TIQVINVRE  330
             LD    + SV+    +K          + A+    +    R    P+        I++RE
Sbjct  1172  LDGCYSVSSVRERYRLKVYASDRGDPPSTADVDVELDVVDRNNKPPIWDRNIYGPIHIRE  1231

Query  331   GIAFRPASKTFTVQKGISSKKLVDYILGTYQAIDEDTNKAASNVKYVMGRNDGGYLMIDS  390
              +             GI     V Y L         TNK   +  Y+  R D G
Sbjct  1232  NVTVGTVVTAIKASSGIEGNPTVFYRL--MPGSTAQTNKL--HTFYLQQRPDNG-----D  1282
```

```
Query  391   KTAEIKFVKNMNRDSTFIVNKTITAEVLAIDEYTGKTSTGTVYYVRVPDFNDNCPTAV-LE  449
               A+IK    ++ ++    N TI  E     +     S  TVY+++ D ND  P     E
Sbjct  1283  TWADIKVNHPLDFETIKEYNLTIRVENNGAQQL---ASEATVYIQLEDVNDEIPLFTERE  1339

Query  450   KDAVCSSSP  458
             ++ V    P
Sbjct  1340  QETVLEGEP  1348

 Score = 61.2 bits (147),  Expect = 5e-08, Method: Compositional matrix adjust.
 Identities = 87/366 (24%), Positives = 156/366 (43%), Gaps = 38/366 (10%)

Query  51    FVVDKNTGDINITAIVDRE-ETPSFL-ITCRALNAQGLDVEKPLILTVKILDINDNPPVF  108
             F VD+ TG+++   + DRE +   F+ +T +A +     +E     TV+I D+NDNPP+F
Sbjct  1047  FTVDEETGEVSTNKVFDREGDDGKFVSVTVKATDQGEPSLEGVCSFTVEITDVNDNPPLF  1106

Query  109   SQQIFMGEIEENSASNSLVMILNATDADEPNHLNSKIAFKIVS-QEPAGTPMFLLSRNTG  167
              +Q ++  ++++++  + ++ ++A+D D  N  N  I + + +   P     F +   +G
Sbjct  1107  DRQKYVENVKQDASIGTNILRVSASDEDADN--NGAIVYTLSAPHSPNDLEYFEIQPESG  1164

Query  168   EVRTLTNSLDREQASS-----YRLVVSGADKDGEGLSTQCECNIKVKDVNDNFPMFRDSQ  222
              +  L   LD   + S     YRL V  +D+ G+  ST            +N P   D
Sbjct  1165  WI-VLKKPLDGCYSVSSVRERYRLKVYASDR-GDPPSTADVDVELDVVDRNNKPPIWDRN  1222

Query  223   YSA--RIEENILSSELLRFQVTDLDEEYTDNWLAVYFFTSG-----NEGNWFEIQTDPRT  275
                   I EN+    ++             N    Y    G     N+ + F +Q  P
Sbjct  1223  IYGPIHIRENVTVGTVV--TAIKASSGIEGNPTVFYRLMPGSTAQTNKLHTFYLQQRPDN  1280

Query  276   NE--GILKVVKALDYEQLQSVKLSIAVKNKAEFHQSVISRYRVQSTPVTIQVINVREGIA  333
              +    +KV   LD+E ++   L+I V+N         ++        V IQ+ +V + I
Sbjct  1281  GDTWADIKVNHPLDFETIKEYNLTIRVENNG-------AQQLASEATVYIQLEDVNDEIP  1333

Query  334   FRPASKTFTVQKGISSKKLVDYILGTYQAIDEDTNKAASNVKYVM---GRNDG-GYLMID  389
                  +  TV +G      V  +     AID+D     + V Y +    RN+G  +  I+
Sbjct  1334  LFTEREQETVLEGEPIGTKVTQV----NAIDKDGTFPNNQVYYYIVDSPRNEGKDFFEIN  1389

Query  390   SKTAEI  395
              ++ EI
Sbjct  1390  LQSGEI  1395

 Score = 55.8 bits (133),  Expect = 3e-06, Method: Compositional matrix adjust.
 Identities = 49/158 (31%), Positives = 78/158 (49%), Gaps = 24/158 (15%)

Query  102   NDNPPVFSQQIFMGEIEENSASNSLVMILNATDADEPNHLNSKIAFKIVSQEPAGTP--M  159
             ND  P FSQQ++   ++EN+  N+LV  + A+D D  N     + F  V     GT
Sbjct  880   NDEEPKFSQQVYTPNVDENAGPNTLVTTVVASDKDGDN-----VRFGFVG---GGTSSGQ  931

Query  160   FLLSRNTGEVRTLTN--SLDREQASSYRLVVSGADK-------DGEGLSTQCECNIKVKD  210
             F++   TG +R  +   SLDR++   Y L V+  D        D    ++     + + D
Sbjct  932   FVIEEITGVIRLHSKAISLDRDK---YELNVTAMDDGSCCVNGDATIHTSTAVVVVFITD  988

Query  211   VNDNFPMFRD-SQYSARIEENILS-SELLRFQVTDLDE  246
             VNDN P+F+D   Y  ++EE   + S +++ Q TD D+
Sbjct  989   VNDNKPVFKDCGTYYPKVEEGAPNGSPVIKVQATDEDK  1026

 Score = 53.9 bits (128),  Expect = 1e-05, Method: Compositional matrix adjust.
 Identities = 70/306 (23%), Positives = 126/306 (41%), Gaps = 30/306 (10%)

Query  20    PIAKIT-SDYQATQKITYRI---SGVGIDQPPFGIFVVDKNTGDINITAIVDRE---ETP  72
             PI  +T  D   T K  Y++   SG G D+  F +   +  TG + I   +D E   ++
Sbjct  1800  PILTVTVHDEDETNKFQYKVIDNSGYGADK--FTMVRNNDGTGSLKIVQPLDYEDPLQSN  1857

Query  73    SFLITCRALNAQGLDVEKPLI------LTVKILDINDNPPVFSQQIFMGEIEENSASNSL  126
              F    + +N +G D +          + VK+ DINDN P F +      + EN+
Sbjct  1858  GFRFRIQ-VNDKGEDNDNDKYHVAYSWVVVKLRDINDNKPQFERPNIEVSVYENADVGKT  1916

Query  127   VMILNATDADEPNHLNSKIAFKIVSQEPAGTPMFLLSRNTGEVRTLTNSLDREQASSYRL  186
             +    ATD D+     SK+++ I   + +       S N     T+  +LDRE    +++
Sbjct  1917  LETFKATDPDQGG--KSKVSYAI---DRSSDRQRQFSINQEGTVTIQRNLDREVTPRHQV  1971
```

```
Query  187   VVSGADKDGEGLSTQCECNIKVKDVNDNFPMFRDSQYSARIEENILSSELLRFQVTDLDE  246
              +    D       +       + V+D+NDN  P F     Y    + E++    +++     TD D+
Sbjct  1972  KILAIDDGIPPKTATATLTVIVQDINDNPPKFL-KDYRPVLPEHVPPRKVVEILATDDDD  2030

Query  247   EYTDNWLAVYFFTSGNEGNW----FEIQTDPRTNEG----ILKVVKALDYEQLQSVKLSI  298
                 N     F        +     F+++ D +   G     I+  +++ D EQ +    + I
Sbjct  2031  RSKSNGPPFQFRLDPGADDIIRASFKVEQDQKGANGDGMAIVSSLRSFDREQQKEYLIPI  2090

Query  299   AVKNKA  304
              +K+
Sbjct  2091  VIKDHG  2096

 Score = 51.6 bits (122),  Expect = 5e-05, Method: Compositional matrix adjust.
 Identities = 96/398 (24%), Positives = 163/398 (41%), Gaps = 54/398 (14%)

Query  106   PVFSQQIFMGEIEENSASNSLVMILNATDADEPNHLNSKIAFKIVSQ-EPAGTPMFLLSR  164
             P F    +  EI EN   +S ++ + A       +  + +I + + +Q + AGT  F +
Sbjct  677   PQFYMPSYEAEIPENQKKDSDIISVKAK-----SFADREIRYTLKAQGQGAGT--FNIGP  729

Query  165   NTGEVRTLTNSLDRE---QASSYRLVVSGADKDGEGLSTQCECNIKVKDVNDNFPMFRDS  221
              +G V+ L   LD E   Q   Y LVV+ A +D  G ST  E  I+V DVNDN P F
Sbjct  730   TSGIVK-LAKELDFEDLRQPHVYSLVVT-ATEDSGGFSTSVELTIRVTDVNDNAPKFELP  787

Query  222   QYSAR-IEENI-LSSELLRFQVTDLDEEYTDNWLAVYFFTSGNEGNWFEIQTDPRTNEGI  279
              Y A  ++E+I + + +L+ +  D D           + +  S +    F + ++     GI
Sbjct  788   DYQAHNVDEDIQIGTSILKVKAMDADSGAN---AEIEYLVSDDH---FAVDSN-----GI  836

Query  280   LKVVKALDYEQLQSV-KLSIAVKNKAEFHQSVISRYRVQSTPVTIQVINVREGIAFRPAS  338
             +   K LD +   +  +  +  K+K E  ++  +  RV +            E   F
Sbjct  837   IVNNKQLDADNNNAYYEFVVTAKDKGEPAKTGTATVRVYTKNKN------DEEPKFSQQV  890

Query  339   KTFTVQKGISSKKLVDYILGTYQAIDEDTNKAASNVKY--VMGRNDGGYLMIDSKTAEIK  396
              T  V +       LV  ++ +         +K   NV++  V G     G  +I+  T  I+
Sbjct  891   YTPNVDENAGPNTLVTTVVAS--------DKDGDNVRFGFVGGGTSSGQFVIEEITGVIR  942

Query  397   FVK---NMNRDSTFI-VNKTITAEVLAIDEYTGKTSTGTVYVRVPDFNDN------CPTA  446
                  +++RD   + V              + T  TST  V V + D NDN       C T
Sbjct  943   LHSKAISLDRDKYELNVTAMDDGSCCVNGDATIHTSTAVVVVFITDVNDNKPVFKDCGTY  1002

Query  447   VLE-KDAVCSSSPSVVVSARTLNNRYTGPYTFALEDQP  483
               + ++    + SP + V A    +    G   +++   QP
Sbjct  1003  YPKVEEGAPNGSPVIKVQATDEDKGVNGQVKYSIVQQP  1040

 Score = 44.7 bits (104),  Expect = 0.008, Method: Compositional matrix adjust.
 Identities = 48/199 (24%), Positives = 73/199 (37%), Gaps = 19/199 (10%)

Query  27    DYQATQKITYRISGVGIDQPPFGIFVVDKNTGDINITAIVDREETPSFLITCRALNAQGL  86
             D     K++Y I       Q  F I        G + I   +DRE TP   +    A++
Sbjct  1926  DQGGKSKVSYAIDRSSDRQRQFSI----NQEGTVTIQRNLDREVTPRHQVKILAIDDGIP  1981

Query  87    DVEKPLILTVKILDINDNPPVFSQQIFMGEIEENSASNSLVMILNATDADEPNHLNSKIA  146
                   LTV + DINDNPP F +  +   + E+     +V IL   D D
Sbjct  1982  PKTATATLTVIVQDINDNPPKFLKD-YRPVLPEHVPPRKVVEILATDDDDRSKSNGPPFQ  2040

Query  147   FKIVSQEPAGTPMFLLSRNTGEVRTLTN-----------SLDREQASSYRLVVSGADKDG  195
             F++    +P     +    S     + +    N            S DREQ    Y + +    D
Sbjct  2041  FRL---DPGADDIIRASFKVEQDQKGANGDGMAIVSSLRSFDREQQKEYLIPIVIKDHGN  2097

Query  196   EGLSTQCECNIKVKDVNDN  214
              ++       + + DVNDN
Sbjct  2098  PAMTGTSTLTVVIGDVNDN  2116

 Score = 40.8 bits (94),  Expect = 0.11, Method: Compositional matrix adjust.
 Identities = 55/242 (23%), Positives = 102/242 (42%), Gaps = 22/242 (9%)

Query  4     KFAKPCREGE--DNSKRNP-IAKITSDYQATQKITYRISGVGIDQPPFGIFVVDKNTGDI  60
             +F  P  E E  +N K++  I  + +   A ++I Y +   G       G F +   +G +
```

```
Sbjct  678  QFYMPSYEAEIPENQKKDSDIISVKAKSFADREIRYTLKAQGQGA---GTFNIGPTSGIV  734

Query  61   NITAIVDREE--TPSF--LITCRALNAQGLDVEKPLILTVKILDINDNPPVFSQQIFMGE  116
             +   +D E+   P    L+     ++ G      + LT+++ D+NDN P F    +
Sbjct  735  KLAKELDFEDLRQPHVYSLVVTATEDSGGFSTS--VELTIRVTDVNDNAPKFELPDYQAH  792

Query  117  -IEENSASNSLVMILNATDADEPNHLNSKIAFKIVSQEPAGTPMFLLSRNTGEVRTLTNS  175
             ++E+    + ++ + A DAD  +  N++I + +VS +      F +  N   V
Sbjct  793  NVDEDIQIGTSILKVKAMDAD--SGANAEIEY-LVSDD-----HFAVDSNGIIVNNKQLD  844

Query  176  LDREQASSYRLVVSGADKDGEGLSTQCECNIKVKDVNDNFPMFRDSQYSARIEENILSSE  235
             D   A  Y  VV+  DK     +      +  K+ ND  P F    Y+  ++EN   +
Sbjct  845  ADNNNA-YYEFVVTAKDKGEPAKTGTATVRVYTKNKNDEEPKFSQQVYTPNVDENAGPNT  903

Query  236  LL  237
            L+
Sbjct  904  LV  905

 Score = 39.3 bits (90),  Expect = 0.30, Method: Compositional matrix adjust.
 Identities = 55/225 (24%), Positives = 88/225 (39%), Gaps = 24/225 (11%)

Query  75   LITCRALNAQGLDVEKPLILTVKILDINDNPPVFSQQIF--MGEIEENSASNSLVMILNA  132
            +IT +  NA GL       + + + D+ND PP F  +       ++ N+  N+ V  L A
Sbjct  539  IITNQGHNA-GLKYTDNQRVIIHVKDVNDEPPYFINRPLPMQAVVQLNAPPNTPVFTLQA  597

Query  133  TDADEPNHLNSKIAFKIVSQEPAGTPMFLLSRNTGEVRTLTNSLDREQASSYRLVVSGAD  192
             D D     +  I + IV     G   F +   +G VRT    L  +    Y L V   D
Sbjct  598  RDPD----TDHNIHYFIVRDRTGG--RFEVDERSGVVRTRGTDL-FQLDMEYVLYVKAED  650

Query  193  KDGE----GLSTQCECNIKVKDVNDNFPMFRDSQYSARIEEN-ILSSELLRFQVTDLDEE  247
            ++G+       +  E  + +       P F    Y A I EN    S+++  +     +
Sbjct  651  QNGKVDDRRFQSTPEERLSIVG-GKRAPQFYMPSYEAEIPENQKKDSDIISVKA----KS  705

Query  248  YTDNWLAVYFFTSGNEGNWFEIQTDPRTNEGILKVVKALDYEQLQ  292
            + D  +       G     F I        GI+K+ K LD+E L+
Sbjct  706  FADREIRYTLKAQGQGAGTFNIG----PTSGIVKLAKELDFEDLR  746

>AALB20_033171.P66526 | transcript=AALB20_033171.R66526 | gene=AALB20_033171
| organism=Anopheles_albimanus_STECLA_2020 |
gene_product=neural-cadherin, transcript variant X38 |
transcript_product=neural-cadherin,
transcript variant X3 | location=CM024292:75037560-75208393(+)
| protein_length=3131 | sequence_SO=chromosome
| SO=protein_coding_gene | is_pseudo=false
Length=3131

 Score = 144 bits (364),  Expect = 1e-34, Method: Compositional matrix adjust.
 Identities = 125/437 (29%), Positives = 211/437 (48%), Gaps = 33/437 (8%)

Query  26    SDYQATQKITYRISGVGID--QPPFGIFVVDKNTGDINITAIVDREET---PSFLITCRA  80
              D    Q I Y ++G GID   P    F +++ TG+I +   +DR++    P +  T  A
Sbjct  1580  GDKDRPQNIVYFLTGQGIDPDNPANSKFDINRTTGEIFVLKPLDRDQPNGRPQWRFTVFA  1639

Query  81    LNAQGLDVEKPLILTVKILDINDNPPVFSQQIFMGEIEENSASNSLVMILNATDADEPNH  140
              +  G  +     + V + DINDN P+F Q ++ G + EN  +  +VM + A D D+PN
Sbjct  1640  QDEGGEGLVGYADVQVNLKDINDNAPIFPQGVYFGNVTENGTAGMVVMTMTAVDYDDPNE  1699

Query  141   -LNSKIAFKI---VSQEPAGTPMFLLSRNTGEVRTLTNSLDREQASSYRLVVSGADKDGE  196
                N+K+ + I   V +E  G+P+F +  +TG ++T    LDRE+   Y + V     DG
Sbjct  1700  GTNAKLIYSIEKNVIEEETGSPIFEIEADTGVIKTAVCCLDRERTPDYSIQVVAM--DGG  1757

Query  197   GLSTQCECNIKVKDVNDNFPMFRDSQYSARIEENILSS--ELLRFQVTDLDEEYTDNWLA  254
              GL      +I+VKD+ND  P F   ++   ++E   ++  E+    VT  DE+ T+ +
Sbjct  1758  GLKGTGTASIRVKDINDMPPQFTKDEWFTEVDETDGTNLPEMPILTVTVHDEDETNKFQY  1817

Query  255   VYFFTSGNEGNWFEIQTDPRTNEGI--LKVVKALDYE---QLQSVKLSIAVKNKAEFHQS  309
                   SG    + F +   R N+G   LK+V+ LDYE   Q    +  I V +K E + +
Sbjct  1818  KVIDNSGYGADKFTMV---RNNDGTGSLKIVQPLDYEDPLQSNGFRFIQVNDKGEDNDN  1874
```

```
Query  310   VISRYRVQSTPVTIQVINVREGIAFRPASKTFTVQKGISSKKLVDYILGTYQAIDEDTNK  369
                +Y V  + V +++ ++ +    +P  +   ++  +     V   L T++A D D
Sbjct  1875  --DKYHVAYSWVVVKLRDINDN---KPQFERPNIEVSVYENADVGKTLETFKATDPDQG-  1928

Query  370   AASNVKYVMGR-NDGGYLMIDSKTAEIKFVKNMNRDSTFIVNKTITAEVLAIDE-YTGKT  427
               S V Y + R +D       ++   +   +N++R+    V      ++LAID+    KT
Sbjct  1929  GKSKVSYAIDRSSDRQRQFSINQEGTVTIQRNLDRE----VTPRHQVKILAIDDGIPPKT  1984

Query  428   STGTVYVRVPDFNDNCP  444
             +T T+ V V D NDN P
Sbjct  1985  ATATLTVIVQDINDNPP  2001

 Score = 85.9 bits (211),  Expect = 1e-15, Method: Compositional matrix adjust.
 Identities = 108/420 (26%), Positives = 177/420 (42%), Gaps = 52/420 (12%)

Query  50    IFVVDKNTGDINITAIVDREETPSFLITCRAL----------NAQGLDVEKPLILTVKIL  99
              F ++  +G+I    + DRE+  ++ +   A           N Q   V K   + + I
Sbjct  1385  FFEINLQSGEIFTKVVFDREKQGAYALEVEARDGAPSARPNSNQQPNSVTK--FIRIGIA  1442

Query  100   DINDNPPVFSQQIFMGEIEENSASNSLVMILNATDADEPNHLNSKIAFKIVSQEPAGTPM  159
             D NDNPP F + ++  E++EN      V+ + A D DE    +S+I ++I S    G
Sbjct  1443  DKNDNPPYFDKALYEAEVDENEDIQHTVLTVTAKDHDE----SSRIRYEITSGNIGGA--  1496

Query  160   FLLSRNTGEVRTLTNSLDREQASSYRLVVSGADKDGEGLSTQCECNIKVKDVNDNFPMFR  219
             F +   TG +  +  +LD E    Y L ++ +D   E  +T     I VKDVNDN P+F
Sbjct  1497  FAVKNMTGAI-YVAGALDYETRKRYELRLAASDNLKENYTTVV---IHVKDVNDNPPVFE  1552

Query  220   DSQYSARIEENI---LSSELLRFQVTDLDEEYTDNWLAVYFFTSG--NEGNWFEIQTDPR  274
                Y  +I E     L   +L+   TD D++   N   VYF T    +  N    + D
Sbjct  1553  RPTYRTQITEEDDRNLPKRVLQVTATDGDKDRPQN--IVYFLTGQGIDPDNPANSKFDIN  1610

Query  275   TNEGILKVVKALDYEQLQS-VKLSIAVKNKAEFHQSVISRYRVQSTPVTIQVINVREGIA  333
                G + V+K LD +Q     +    V  + E  + ++    VQ     + + ++ +
Sbjct  1611  RTTGEIFVLKPLDRDQPNGRPQWRFTVFAQDEGGEGLVGYADVQ-----VNLKDINDNAP  1665

Query  334   FRPASKTFTVQKGISSKKLVDYILGTYQAIDEDTNKAASNVK--YVMGRN------DGGY  385
               P    F     ++       ++ T  A+D D     +N K  Y + +N
Sbjct  1666  IFPQGVYF---GNVTENGTAGMVVMTMTAVDYDDPNEGTNAKLIYSIEKNVIEEETGSPI  1722

Query  386   LMIDSKTAEIK-FVKNMNRDSTFIVNKTITAEVLAIDEYTGKTSTGTVYVRVPDFNDNCP  444
               I++ T  IK  V  ++R+ T       + +V+A+D   G   TGT  +RV D ND  P
Sbjct  1723  FEIEADTGVIKTAVCCLDRERT----PDYSIQVVAMDG-GGLKGTGTASIRVKDINDMPP  1777

 Score = 75.9 bits (185),  Expect = 2e-12, Method: Compositional matrix adjust.
 Identities = 108/429 (25%), Positives = 178/429 (41%), Gaps = 53/429 (12%)

Query  53    VDKNTGDINITAIVDREETPSFLITCRALNAQGLDVEKPLILTVKILDINDNPPVFSQ-Q  111
             +D++  ++N+TA+ D         +C  +N          ++ V I D+NDN PVF
Sbjct  950   LDRDKYELNVTAMDDG--------SC-CVNGDATIHTSTAVVVVFITDVNDNKPVFKDCG  1000

Query  112   IFMGEIEENSASNSLVMILNATDADEPNHLNSKIAFKIVSQEPAGTPMFLLSRNTGEVRT  171
              +  ++EE + + S V+ + ATD D+   +N ++ + IV Q       F +   TGEV T
Sbjct  1001  TYYPKVEEGAPNGSPVIKVQATDEDK--GVNGQVKYSIVQQPNQKGTKFTVDEETGEVST  1058

Query  172   LTNSLDRE--QASSYRLVVSGADKDGEGLSTQCECNIKVKDVNDNFPMFRDSQYSARIEE  229
                  DRE        + V   D+    L   C   +++ DVNDN P+F   +Y   +++
Sbjct  1059  -NKVFDREGDDGKFVSVTVKATDQGEPSLEGVCSFTVEITDVNDNPPLFDRQKYVENVKQ  1117

Query  230   NI-LSSELLRFQVTDLDEEYTDNWLAVYFFT---SGNEGNWFEIQTDPRTNEGILKVVKA  285
             +  + + +LR  V+  DE+  +N   VY  +   S N+  +FEIQ  P +   +LK  K
Sbjct  1118  DASIGTNILR--VSASDEDADNNGAIVYTLSAPHSPNDLEYFEIQ--PESGWIVLK--KP  1171

Query  286   LD-YEQLQSVKLSIAVK----------NKAEFHQSVISRYRVQSTPV----TIQVINVRE  330
             LD    + SV+    +K          + A+    +    R    P+        I++RE
Sbjct  1172  LDGCYSVSSVRERYRLKVYASDRGDPPSTADVDVELDVVDRNNKPPIWDRNIYGPIHIRE  1231

Query  331   GIAFRPASKTFTVQKGISSKKLVDYILGTYQAIDEDTNKAASNVKYVMGRNDGGYLMIDS  390
              +             GI     V Y L         TNK   +  Y+  R D G
Sbjct  1232  NVTVGTVVTAIKASSGIEGNPTVFYRL--MPGSTAQTNKL--HTFYLQQRPDNG-----D  1282
```

```
Query  391   KTAEIKFVKNMNRDSTFIVNKTITAEVLAIDEYTGKTSTGTVYVRVPDFNDNCPTAV-LE  449
               A+IK    ++ ++    N TI  E     +     S  TVY+++ D ND  P     E
Sbjct  1283  TWADIKVNHPLDFETIKEYNLTIRVENNGAQQL---ASEATVYIQLEDVNDEIPLFTERE  1339

Query  450   KDAVCSSSP  458
             ++ V    P
Sbjct  1340  QETVLEGEP  1348

 Score = 61.2 bits (147),  Expect = 5e-08, Method: Compositional matrix adjust.
 Identities = 87/366 (24%), Positives = 156/366 (43%), Gaps = 38/366 (10%)

Query  51    FVVDKNTGDINITAIVDRE-ETPSFL-ITCRALNAQGLDVEKPLILTVKILDINDNPPVF  108
             F VD+ TG+++   + DRE +   F+ +T +A +     +E     TV+I D+NDNPP+F
Sbjct  1047  FTVDEETGEVSTNKVFDREGDDGKFVSVTVKATDQGEPSLEGVCSFTVEITDVNDNPPLF  1106

Query  109   SQQIFMGEIEENSASNSLVMILNATDADEPNHLNSKIAFKIVS-QEPAGTPMFLLSRNTG  167
              +Q ++  ++++++  + ++ ++A+D D  N  N  I + + +   P     F +   +G
Sbjct  1107  DRQKYVENVKQDASIGTNILRVSASDEDADN--NGAIVYTLSAPHSPNDLEYFEIQPESG  1164

Query  168   EVRTLTNSLDREQASS-----YRLVVSGADKDGEGLSTQCECNIKVKDVNDNFPMFRDSQ  222
              +  L   LD   + S     YRL V  +D+ G+  ST            +N P   D
Sbjct  1165  WI-VLKKPLDGCYSVSSVRERYRLKVYASDR-GDPPSTADVDVELDVVDRNNKPPIWDRN  1222

Query  223   YSA--RIEENILSSELLRFQVTDLDEEYTDNWLAVYFFTSG-----NEGNWFEIQTDPRT  275
                   I EN+    ++             N    Y    G     N+ + F +Q  P
Sbjct  1223  IYGPIHIRENVTVGTVV--TAIKASSGIEGNPTVFYRLMPGSTAQTNKLHTFYLQQRPDN  1280

Query  276   NE--GILKVVKALDYEQLQSVKLSIAVKNKAEFHQSVISRYRVQSTPVTIQVINVREGIA  333
              +    +KV   LD+E ++   L+I V+N         ++       V IQ+ +V + I
Sbjct  1281  GDTWADIKVNHPLDFETIKEYNLTIRVENNG-------AQQLASEATVYIQLEDVNDEIP  1333

Query  334   FRPASKTFTVQKGISSKKLVDYILGTYQAIDEDTNKAASNVKYVM---GRNDG-GYLMID  389
                  +  TV +G      V  +     AID+D     + V Y +    RN+G  +  I+
Sbjct  1334  LFTEREQETVLEGEPIGTKVTQV----NAIDKDGTFPNNQVYYYIVDSPRNEGKDFFEIN  1389

Query  390   SKTAEI  395
              ++ EI
Sbjct  1390  LQSGEI  1395

 Score = 55.8 bits (133),  Expect = 3e-06, Method: Compositional matrix adjust.
 Identities = 49/158 (31%), Positives = 78/158 (49%), Gaps = 24/158 (15%)

Query  102   NDNPPVFSQQIFMGEIEENSASNSLVMILNATDADEPNHLNSKIAFKIVSQEPAGTP--M  159
             ND  P FSQQ++   ++EN+  N+LV  + A+D D  N     + F  V      GT
Sbjct  880   NDEEPKFSQQVYTPNVDENAGPNTLVTTVVASDKDGDN-----VRFGFVG---GGTSSGQ  931

Query  160   FLLSRNTGEVRTLTN--SLDREQASSYRLVVSGADK-------DGEGLSTQCECNIKVKD  210
             F++   TG +R  +   SLDR++   Y L V+  D        D    ++     + + D
Sbjct  932   FVIEEITGVIRLHSKAISLDRDK---YELNVTAMDDGSCCVNGDATIHTSTAVVVVFITD  988

Query  211   VNDNFPMFRD-SQYSARIEENILS-SELLRFQVTDLDE  246
             VNDN P+F+D   Y  ++EE   + S +++ Q TD D+
Sbjct  989   VNDNKPVFKDCGTYYPKVEEGAPNGSPVIKVQATDEDK  1026

 Score = 53.9 bits (128),  Expect = 1e-05, Method: Compositional matrix adjust.
 Identities = 70/306 (23%), Positives = 126/306 (41%), Gaps = 30/306 (10%)

Query  20    PIAKIT-SDYQATQKITYRI---SGVGIDQPPFGIFVVDKNTGDINITAIVDRE---ETP  72
             PI  +T  D   T K  Y++   SG G D+  F +   +  TG + I   +D E   ++
Sbjct  1800  PILTVTVHDEDETNKFQYKVIDNSGYGADK--FTMVRNNDGTGSLKIVQPLDYEDPLQSN  1857

Query  73    SFLITCRALNAQGLDVEKPLI------LTVKILDINDNPPVFSQQIFMGEIEENSASNSL  126
              F    + +N +G D +          + VK+ DINDN P F +      + EN+
Sbjct  1858  GFRFRIQ-VNDKGEDNDNDKYHVAYSWVVVKLRDINDNKPQFERPNIEVSVYENADVGKT  1916

Query  127   VMILNATDADEPNHLNSKIAFKIVSQEPAGTPMFLLSRNTGEVRTLTNSLDREQASSYRL  186
             +    ATD D+     SK+++ I   + +       S N     T+  +LDRE    +++
```

```
Sbjct  1917  LETFKATDPDQGG--KSKVSYAI---DRSSDRQRQFSINQEGTVTIQRNLDREVTPRHQV  1971

Query  187   VVSGADKDGEGLSTQCECNIKVKDVNDNFPMFRDSQYSARIEENILSSELLRFQVTDLDE  246
              +   D      +      + V+D+NDN P F    Y   + E++   +++    TD D+
Sbjct  1972  KILAIDDGIPPKTATATLTVIVQDINDNPPKFL-KDYRPVLPEHVPPRKVVEILATDDDD  2030

Query  247   EYTDNWLAVYFFTSGNEGNW----FEIQTDPRTNEG----ILKVVKALDYEQLQSVKLSI  298
                  N     F       +     F+++ D +   G    I+  +++ D EQ +   + I
Sbjct  2031  RSKSNGPPFQFRLDPGADDIIRASFKVEQDQKGANGDGMAIVSSLRSFDREQQKEYLIPI  2090

Query  299   AVKNKA  304
              +K+
Sbjct  2091  VIKDHG  2096

 Score = 51.6 bits (122),  Expect = 5e-05, Method: Compositional matrix adjust.
 Identities = 96/398 (24%), Positives = 163/398 (41%), Gaps = 54/398 (14%)

Query  106   PVFSQQIFMGEIEENSASNSLVMILNATDADEPNHLNSKIAFKIVSQ-EPAGTPMFLLSR  164
             P F    +  EI EN   +S ++ + A       +  + +I + + +Q + AGT  F +
Sbjct  677   PQFYMPSYEAEIPENQKKDSDIISVKAK-----SFADREIRYTLKAQGQGAGT--FNIGP  729

Query  165   NTGEVRTLTNSLDRE---QASSYRLVVSGADKDGEGLSTQCECNIKVKDVNDNFPMFRDS  221
              +G V+ L   LD E   Q   Y LVV+ A +D  G ST  E  I+V DVNDN P F
Sbjct  730   TSGIVK-LAKELDFEDLRQPHVYSLVVT-ATEDSGGFSTSVELTIRVTDVNDNAPKFELP  787

Query  222   QYSAR-IEENI-LSSELLRFQVTDLDEEYTDNWLAVYFFTSGNEGNWFEIQTDPRTNEGI  279
              Y A  ++E+I + + +L+ +  D D          + +  S +    F + ++     GI
Sbjct  788   DYQAHNVDEDIQIGTSILKVKAMDADSGAN---AEIEYLVSDDH---FAVDSN-----GI  836

Query  280   LKVVKALDYEQLQSV-KLSIAVKNKAEFHQSVISRYRVQSTPVTIQVINVREGIAFRPAS  338
             +   K LD +   +  +  K+K E  ++  +  RV +            E  F
Sbjct  837   IVNNKQLDADNNNAYYEFVVTAKDKGEPAKTGTATVRVYTKNKN------DEEPKFSQQV  890

Query  339   KTFTVQKGISSKKLVDYILGTYQAIDEDTNKAASNVKY--VMGRNDGGYLMIDSKTAEIK  396
              T  V +      LV  ++ +         +K   NV++  V G    G  +I+  T  I+
Sbjct  891   YTPNVDENAGPNTLVTTVVAS--------DKDGDNVRFGFVGGGTSSGQFVIEEITGVIR  942

Query  397   FVK---NMNRDSTFI-VNKTITAEVLAIDEYTGKTSTGTVYVRVPDFNDN------CPTA  446
                   +++RD   + V              + T  TST  V V + D NDN      C T
Sbjct  943   LHSKAISLDRDKYELNVTAMDDGSCCVNGDATIHTSTAVVVVFITDVNDNKPVFKDCGTY  1002

Query  447   VLE-KDAVCSSSPSVVVSARTLNNRYTGPYTFALEDQP  483
               + ++    + SP + V A    +    G   +++  QP
Sbjct  1003  YPKVEEGAPNGSPVIKVQATDEDKGVNGQVKYSIVQQP  1040

 Score = 44.7 bits (104),  Expect = 0.008, Method: Compositional matrix adjust.
 Identities = 48/199 (24%), Positives = 73/199 (37%), Gaps = 19/199 (10%)

Query  27    DYQATQKITYRISGVGIDQPPFGIFVVDKNTGDINITAIVDREETPSFLITCRALNAQGL  86
             D     K++Y I      Q  F I        G + I   +DRE TP   +    A++
Sbjct  1926  DQGGKSKVSYAIDRSSDRQRQFSI----NQEGTVTIQRNLDREVTPRHQVKILAIDDGIP  1981

Query  87    DVEKPLILTVKILDINDNPPVFSQQIFMGEIEENSASNSLVMILNATDADEPNHLNSKIA  146
                      LTV + DINDNPP F +  +    + E+     +V IL   D D
Sbjct  1982  PKTATATLTVIVQDINDNPPKFLKD-YRPVLPEHVPPRKVVEILATDDDDRSKSNGPPFQ  2040

Query  147   FKIVSQEPAGTPMFLLSRNTGEVRTLTN-----------SLDREQASSYRLVVSGADKDG  195
             F++    +P    +   S    + +   N           S DREQ    Y + +   D
Sbjct  2041  FRL---DPGADDIIRASFKVEQDQKGANGDGMAIVSSLRSFDREQQKEYLIPIVIKDHGN  2097

Query  196   EGLSTQCECNIKVKDVNDN  214
              ++       + + DVNDN
Sbjct  2098  PAMTGTSTLTVVIGDVNDN  2116

 Score = 40.8 bits (94),  Expect = 0.11, Method: Compositional matrix adjust.
 Identities = 55/242 (23%), Positives = 102/242 (42%), Gaps = 22/242 (9%)

Query  4    KFAKPCREGE--DNSKRNP-IAKITSDYQATQKITYRISGVGIDQPPFGIFVVDKNTGDI  60
```

```
            +F  P  E E  +N K++  I  + +   A ++I Y +   G      G F +   +G +
Sbjct  678  QFYMPSYEAEIPENQKKDSDIISVKAKSFADREIRYTLKAQGQGA---GTFNIGPTSGIV  734

Query  61   NITAIVDREE--TPSF--LITCRALNAQGLDVEKPLILTVKILDINDNPPVFSQQIFMGE  116
             +   +D E+   P    L+     ++ G      + LT+++ D+NDN P F    +
Sbjct  735  KLAKELDFEDLRQPHVYSLVVTATEDSGGFSTS--VELTIRVTDVNDNAPKFELPDYQAH  792

Query  117  -IEENSASNSLVMILNATDADEPNHLNSKIAFKIVSQEPAGTPMFLLSRNTGEVRTLTNS  175
             ++E+    + ++ + A DAD  +  N++I + +VS +      F +  N   V
Sbjct  793  NVDEDIQIGTSILKVKAMDAD--SGANAEIEY-LVSDD-----HFAVDSNGIIVNNKQLD  844

Query  176  LDREQASSYRLVVSGADKDGEGLSTQCECNIKVKDVNDNFPMFRDSQYSARIEENILSSE  235
             D   A  Y  VV+  DK      +       +  K+ ND  P F    Y+  ++EN   +
Sbjct  845  ADNNNA-YYEFVVTAKDKGEPAKTGTATVRVYTKNKNDEEPKFSQQVYTPNVDENAGPNT  903

Query  236  LL  237
            L+
Sbjct  904  LV  905

 Score = 39.3 bits (90),  Expect = 0.30, Method: Compositional matrix adjust.
 Identities = 55/225 (24%), Positives = 88/225 (39%), Gaps = 24/225 (11%)

Query  75   LITCRALNAQGLDVEKPLILTVKILDINDNPPVFSQQIF--MGEIEENSASNSLVMILNA  132
            +IT +  NA GL       + + + D+ND PP F  +       ++ N+  N+ V  L A
Sbjct  539  IITNQGHNA-GLKYTDNQRVIIHVKDVNDEPPYFINRPLPMQAVVQLNAPPNTPVFTLQA  597

Query  133  TDADEPNHLNSKIAFKIVSQEPAGTPMFLLSRNTGEVRTLTNSLDREQASSYRLVVSGAD  192
             D D     +  I + IV     G   F +   +G VRT    L  +    Y L V   D
Sbjct  598  RDPD----TDHNIHYFIVRDRTGG--RFEVDERSGVVRTRGTDL-FQLDMEYVLYVKAED  650

Query  193  KDGE----GLSTQCECNIKVKDVNDNFPMFRDSQYSARIEEN-ILSSELLRFQVTDLDEE  247
            ++G+        +  E  + +        P F    Y A I EN    S+++  +     +
Sbjct  651  QNGKVDDRRFQSTPEERLSIVG-GKRAPQFYMPSYEAEIPENQKKDSDIISVKA----KS  705

Query  248  YTDNWLAVYFFTSGNEGNWFEIQTDPRTNEGILKVVKALDYEQLQ  292
            + D  +        G     F I         GI+K+ K LD+E L+
Sbjct  706  FADREIRYTLKAQGQGAGTFNIG----PTSGIVKLAKELDFEDLR  746

>AALB20_033171.P66524 | transcript=AALB20_033171.R66524 | gene=AALB20_033171
| organism=Anopheles_albimanus_STECLA_2020 |
gene_product=neural-cadherin, transcript variant X38 |
transcript_product=neural-cadherin,
transcript variant X15 | location=CM024292:75037560-75208393(+)
| protein_length=3131 | sequence_SO=chromosome
| SO=protein_coding_gene | is_pseudo=false
Length=3131

 Score = 144 bits (364),  Expect = 1e-34, Method: Compositional matrix adjust.
 Identities = 125/437 (29%), Positives = 211/437 (48%), Gaps = 33/437 (8%)

Query  26    SDYQATQKITYRISGVGID--QPPFGIFVVDKNTGDINITAIVDREET---PSFLITCRA  80
              D    Q I Y ++G GID   P    F +++ TG+I +   +DR++    P +  T  A
Sbjct  1580  GDKDRPQNIVYFLTGQGIDPDNPANSKFDINRTTGEIFVLKPLDRDQPNGRPQWRFTVFA  1639

Query  81    LNAQGLDVEKPLILTVKILDINDNPPVFSQQIFMGEIEENSASNSLVMILNATDADEPNH  140
               +  G  +      + V + DINDN P+F Q ++ G + EN  +  +VM + A D D+PN
Sbjct  1640  QDEGGEGLVGYADVQVNLKDINDNAPIFPQGVYFGNVTENGTAGMVVMTMTAVDYDDPNE  1699

Query  141   -LNSKIAFKI---VSQEPAGTPMFLLSRNTGEVRTLTNSLDREQASSYRLVVSGADKDGE  196
                N+K+ + I   V +E  G+P+F +   +TG ++T    LDRE+   Y + V     DG
Sbjct  1700  GTNAKLIYSIEKNVIEEETGSPIFEIEADTGVIKTAVCCLDRERTPDYSIQVVAM--DGG  1757

Query  197   GLSTQCECNIKVKDVNDNFPMFRDSQYSARIEENILSS--ELLRFQVTDLDEEYTDNWLA  254
              GL       +I+VKD+ND  P F   ++   ++E   ++  E+    VT  DE+ T+ +
Sbjct  1758  GLKGTGTASIRVKDINDMPPQFTKDEWFTEVDETDGTNLPEMPILTVTVHDEDETNKFQY  1817

Query  255   VYFFTSGNEGNWFEIQTDPRTNEGI--LKVVKALDYE---QLQSVKLSIAVKNKAEFHQS  309
                   SG    + F +    R N+G   LK+V+ LDYE   Q     +  I V +K E + +
Sbjct  1818  KVIDNSGYGADKFTMV---RNNDGTGSLKIVQPLDYEDPLQSNGFRFRIQVNDKGEDNDN  1874
```

```
Query  310   VISRYRVQSTPVTIQVINVREGIAFRPASKTFTVQKGISSKKLVDYILGTYQAIDEDTNK  369
                +Y V  + V +++ ++ +    +P  +   ++  +     V   L T++A D D
Sbjct  1875  --DKYHVAYSWVVVKLRDINDN---KPQFERPNIEVSVYENADVGKTLETFKATDPDQG-  1928

Query  370   AASNVKYVMGR-NDGGYLMIDSKTAEIKFVKNMNRDSTFIVNKTITAEVLAIDE-YTGKT  427
               S V Y + R +D       ++   +   +N++R+    V      ++LAID+    KT
Sbjct  1929  GKSKVSYAIDRSSDRQRQFSINQEGTVTIQRNLDRE----VTPRHQVKILAIDDGIPPKT  1984

Query  428   STGTVYVRVPDFNDNCP  444
             +T T+ V V D NDN P
Sbjct  1985  ATATLTVIVQDINDNPP  2001

 Score = 85.9 bits (211),  Expect = 1e-15, Method: Compositional matrix adjust.
 Identities = 108/420 (26%), Positives = 177/420 (42%), Gaps = 52/420 (12%)

Query  50    IFVVDKNTGDINITAIVDREETPSFLITCRAL----------NAQGLDVEKPLILTVKIL  99
              F ++  +G+I    + DRE+  ++ +   A           N Q   V K   + + I
Sbjct  1385  FFEINLQSGEIFTKVVFDREKQGAYALEVEARDGAPSARPNSNQQPNSVTK--FIRIGIA  1442

Query  100   DINDNPPVFSQQIFMGEIEENSASNSLVMILNATDADEPNHLNSKIAFKIVSQEPAGTPM  159
             D NDNPP F + ++  E++EN      V+ + A D DE    +S+I ++I S    G
Sbjct  1443  DKNDNPPYFDKALYEAEVDENEDIQHTVLTVTAKDHDE----SSRIRYEITSGNIGGA--  1496

Query  160   FLLSRNTGEVRTLTNSLDREQASSYRLVVSGADKDGEGLSTQCECNIKVKDVNDNFPMFR  219
             F +   TG +  +  +LD E    Y L ++ +D   E  +T     I VKDVNDN P+F
Sbjct  1497  FAVKNMTGAI-YVAGALDYETRKRYELRLAASDNLKENYTTVV---IHVKDVNDNPPVFE  1552

Query  220   DSQYSARIEENI---LSSELLRFQVTDLDEEYTDNWLAVYFFTSG--NEGNWFEIQTDPR  274
                Y  +I E     L   +L+   TD D++   N   VYF T    +  N    + D
Sbjct  1553  RPTYRTQITEEDDRNLPKRVLQVTATDGDKDRPQN--IVYFLTGQGIDPDNPANSKFDIN  1610

Query  275   TNEGILKVVKALDYEQLQS-VKLSIAVKNKAEFHQSVISRYRVQSTPVTIQVINVREGIA  333
                G + V+K LD +Q     +    V  + E  + ++    VQ     + + ++ +
Sbjct  1611  RTTGEIFVLKPLDRDQPNGRPQWRFTVFAQDEGGEGLVGYADVQ-----VNLKDINDNAP  1665

Query  334   FRPASKTFTVQKGISSKKLVDYILGTYQAIDEDTNKAASNVK--YVMGRN------DGGY  385
               P    F     ++       ++ T  A+D D     +N K  Y + +N
Sbjct  1666  IFPQGVYF---GNVTENGTAGMVVMTMTAVDYDDPNEGTNAKLIYSIEKNVIEEETGSPI  1722

Query  386   LMIDSKTAEIK-FVKNMNRDSTFIVNKTITAEVLAIDEYTGKTSTGTVYVRVPDFNDNCP  444
               I++ T  IK  V  ++R+ T       + +V+A+D   G   TGT  +RV D ND  P
Sbjct  1723  FEIEADTGVIKTAVCCLDRERT----PDYSIQVVAMDG-GGLKGTGTASIRVKDINDMPP  1777

 Score = 75.9 bits (185),  Expect = 2e-12, Method: Compositional matrix adjust.
 Identities = 108/429 (25%), Positives = 178/429 (41%), Gaps = 53/429 (12%)

Query  53    VDKNTGDINITAIVDREETPSFLITCRALNAQGLDVEKPLILTVKILDINDNPPVFSQ-Q  111
             +D++  ++N+TA+ D         +C  +N          ++ V I D+NDN PVF
Sbjct  950   LDRDKYELNVTAMDDG--------SC-CVNGDATIHTSTAVVVVFITDVNDNKPVFKDCG  1000

Query  112   IFMGEIEENSASNSLVMILNATDADEPNHLNSKIAFKIVSQEPAGTPMFLLSRNTGEVRT  171
              +  ++EE + + S V+ + ATD D+   +N ++ + IV Q       F +   TGEV T
Sbjct  1001  TYYPKVEEGAPNGSPVIKVQATDEDK--GVNGQVKYSIVQQPNQKGTKFTVDEETGEVST  1058

Query  172   LTNSLDRE--QASSYRLVVSGADKDGEGLSTQCECNIKVKDVNDNFPMFRDSQYSARIEE  229
                  DRE        + V   D+    L   C   +++ DVNDN P+F   +Y   +++
Sbjct  1059  -NKVFDREGDDGKFVSVTVKATDQGEPSLEGVCSFTVEITDVNDNPPLFDRQKYVENVKQ  1117

Query  230   NI-LSSELLRFQVTDLDEEYTDNWLAVYFFT---SGNEGNWFEIQTDPRTNEGILKVVKA  285
             +  + + +LR  V+  DE+  +N   VY  +   S N+  +FEIQ  P +   +LK  K
Sbjct  1118  DASIGTNILR--VSASDEDADNNGAIVYTLSAPHSPNDLEYFEIQ--PESGWIVLK--KP  1171

Query  286   LD-YEQLQSVKLSIAVK----------NKAEFHQSVISRYRVQSTPV----TIQVINVRE  330
             LD    + SV+    +K          + A+    +    R    P+        I++RE
Sbjct  1172  LDGCYSVSSVRERYRLKVYASDRGDPPSTADVDVELDVVDRNNKPPIWDRNIYGPIHIRE  1231

Query  331   GIAFRPASKTFTVQKGISSKKLVDYILGTYQAIDEDTNKAASNVKYVMGRNDGGYLMIDS  390
              +             GI     V Y L         TNK   +  Y+  R D G
```

```
Sbjct  1232  NVTVGTVVTAIKASSGIEGNPTVFYRL--MPGSTAQTNKL--HTFYLQQRPDNG-----D  1282

Query  391   KTAEIKFVKNMNRDSTFIVNKTITAEVLAIDEYTGKTSTGTVYVRVPDFNDNCPTAV-LE  449
               A+IK    ++ ++    N TI   E     +      S  TVY+++ D ND  P     E
Sbjct  1283  TWADIKVNHPLDFETIKEYNLTIRVENNGAQQL---ASEATVYIQLEDVNDEIPLFTERE  1339

Query  450   KDAVCSSSP  458
             ++ V    P
Sbjct  1340  QETVLEGEP  1348

 Score = 61.2 bits (147),  Expect = 5e-08, Method: Compositional matrix adjust.
 Identities = 87/366 (24%), Positives = 156/366 (43%), Gaps = 38/366 (10%)

Query  51    FVVDKNTGDINITAIVDRE-ETPSFL-ITCRALNAQGLDVEKPLILTVKILDINDNPPVF  108
             F VD+ TG+++    + DRE +    F+ +T +A +      +E      TV+I D+NDNPP+F
Sbjct  1047  FTVDEETGEVSTNKVFDREGDDGKFVSVTVKATDQGEPSLEGVCSFTVEITDVNDNPPLF  1106

Query  109   SQQIFMGEIEENSASNSLVMILNATDADEPNHLNSKIAFKIVS-QEPAGTPMFLLSRNTG  167
              +Q ++   ++++++   + ++ ++A+D D  N  N  I + + +    P     F +   +G
Sbjct  1107  DRQKYVENVKQDASIGTNILRVSASDEDADN--NGAIVYTLSAPHSPNDLEYFEIQPESG  1164

Query  168   EVRTLTNSLDREQASS-----YRLVVSGADKDGEGLSTQCECNIKVKDVNDNFPMFRDSQ  222
              +  L   LD   + S      YRL V  +D+ G+  ST              +N P   D
Sbjct  1165  WI-VLKKPLDGCYSVSSVRERYRLKVYASDR-GDPPSTADVDVELDVVDRNNKPPIWDRN  1222

Query  223   YSA--RIEENILSSELLRFQVTDLDEEYTDNWLAVYFFTSG-----NEGNWFEIQTDPRT  275
                   I EN+    ++                N    Y    G     N+ + F +Q  P
Sbjct  1223  IYGPIHIRENVTVGTVV--TAIKASSGIEGNPTVFYRLMPGSTAQTNKLHTFYLQQRPDN  1280

Query  276   NE--GILKVVKALDYEQLQSVKLSIAVKNKAEFHQSVISRYRVQSTPVTIQVINVREGIA  333
              +     +KV   LD+E ++   L+I V+N           ++        V IQ+ +V + I
Sbjct  1281  GDTWADIKVNHPLDFETIKEYNLTIRVENNG-------AQQLASEATVYIQLEDVNDEIP  1333

Query  334   FRPASKTFTVQKGISSKKLVDYILGTYQAIDEDTNKAASNVKYVM---GRNDG-GYLMID  389
                  +  TV +G      V  +     AID+D     + V Y +    RN+G  +  I+
Sbjct  1334  LFTEREQETVLEGEPIGTKVTQV----NAIDKDGTFPNNQVYYYIVDSPRNEGKDFFEIN  1389

Query  390   SKTAEI  395
              ++ EI
Sbjct  1390  LQSGEI  1395

 Score = 55.8 bits (133),  Expect = 3e-06, Method: Compositional matrix adjust.
 Identities = 49/158 (31%), Positives = 78/158 (49%), Gaps = 24/158 (15%)

Query  102   NDNPPVFSQQIFMGEIEENSASNSLVMILNATDADEPNHLNSKIAFKIVSQEPAGTP--M  159
             ND  P FSQQ++   ++EN+  N+LV  + A+D D  N     + F  V     GT
Sbjct  880   NDEEPKFSQQVYTPNVDENAGPNTLVTTVVASDKDGDN-----VRFGFVG---GGTSSGQ  931

Query  160   FLLSRNTGEVRTLTN--SLDREQASSYRLVVSGADK-------DGEGLSTQCECNIKVKD  210
             F++   TG +R  +   SLDR++    Y L V+  D         D    ++      + + D
Sbjct  932   FVIEEITGVIRLHSKAISLDRDK---YELNVTAMDDGSCCVNGDATIHTSTAVVVVFITD  988

Query  211   VNDNFPMFRD-SQYSARIEENILS-SELLRFQVTDLDE  246
             VNDN P+F+D   Y  ++EE   + S +++ Q TD D+
Sbjct  989   VNDNKPVFKDCGTYYPKVEEGAPNGSPVIKVQATDEDK  1026

 Score = 53.9 bits (128),  Expect = 1e-05, Method: Compositional matrix adjust.
 Identities = 70/306 (23%), Positives = 126/306 (41%), Gaps = 30/306 (10%)

Query  20    PIAKIT-SDYQATQKITYRI---SGVGIDQPPFGIFVVDKNTGDINITAIVDRE---ETP  72
             PI  +T  D   T K  Y++   SG G D+  F +   +  TG + I   +D E   ++
Sbjct  1800  PILTVTVHDEDETNKFQYKVIDNSGYGADK--FTMVRNNDGTGSLKIVQPLDYEDPLQSN  1857

Query  73    SFLITCRALNAQGLDVEKPLI------LTVKILDINDNPPVFSQQIFMGEIEENSASNSL  126
              F    + +N +G D +           + VK+ DINDN P F +       + EN+
Sbjct  1858  GFRFRIQ-VNDKGEDNDNDKYHVAYSWVVVKLRDINDNKPQFERPNIEVSVYENADVGKT  1916

Query  127   VMILNATDADEPNHLNSKIAFKIVSQEPAGTPMFLLSRNTGEVRTLTNSLDREQASSYRL  186
```

```
             +    ATD D+     SK+++ I   + +       S N     T+  +LDRE    +++
Sbjct  1917  LETFKATDPDQGG--KSKVSYAI---DRSSDRQRQFSINQEGTVTIQRNLDREVTPRHQV  1971

Query  187   VVSGADKDGEGLSTQCECNIKVKDVNDNFPMFRDSQYSARIEENILSSELLRFQVTDLDE  246
              +   D      +      + V+D+NDN P F    Y   + E++   +++    TD D+
Sbjct  1972  KILAIDDGIPPKTATATLTVIVQDINDNPPKFL-KDYRPVLPEHVPPRKVVEILATDDDD  2030

Query  247   EYTDNWLAVYFFTSGNEGNW----FEIQTDPRTNEG----ILKVVKALDYEQLQSVKLSI  298
                 N     F       +     F+++ D +   G    I+  +++ D EQ +   + I
Sbjct  2031  RSKSNGPPFQFRLDPGADDIIRASFKVEQDQKGANGDGMAIVSSLRSFDREQQKEYLIPI  2090

Query  299   AVKNKA  304
              +K+
Sbjct  2091  VIKDHG  2096

 Score = 51.6 bits (122),  Expect = 5e-05, Method: Compositional matrix adjust.
 Identities = 96/398 (24%), Positives = 163/398 (41%), Gaps = 54/398 (14%)

Query  106   PVFSQQIFMGEIEENSASNSLVMILNATDADEPNHLNSKIAFKIVSQ-EPAGTPMFLLSR  164
             P F    +  EI EN   +S ++ + A      +  + +I + + +Q + AGT  F +
Sbjct  677   PQFYMPSYEAEIPENQKKDSDIISVKAK-----SFADREIRYTLKAQGQGAGT--FNIGP  729

Query  165   NTGEVRTLTNSLDRE---QASSYRLVVSGADKDGEGLSTQCECNIKVKDVNDNFPMFRDS  221
              +G V+ L   LD E   Q   Y LVV+ A +D  G ST  E  I+V DVNDN P F
Sbjct  730   TSGIVK-LAKELDFEDLRQPHVYSLVVT-ATEDSGGFSTSVELTIRVTDVNDNAPKFELP  787

Query  222   QYSAR-IEENI-LSSELLRFQVTDLDEEYTDNWLAVYFFTSGNEGNWFEIQTDPRTNEGI  279
              Y A  ++E+I + + +L+ +  D D         + +  S +    F + ++     GI
Sbjct  788   DYQAHNVDEDIQIGTSILKVKAMDADSGAN---AEIEYLVSDDH---FAVDSN-----GI  836

Query  280   LKVVKALDYEQLQSV-KLSIAVKNKAEFHQSVISRYRVQSTPVTIQVINVREGIAFRPAS  338
             +   K LD +   +  +  +  K+K E  ++  +  RV +           E   F
Sbjct  837   IVNNKQLDADNNNAYYEFVVTAKDKGEPAKTGTATVRVYTKNKN------DEEPKFSQQV  890

Query  339   KTFTVQKGISSKKLVDYILGTYQAIDEDTNKAASNVKY--VMGRNDGGYLMIDSKTAEIK  396
              T  V +      LV  ++ +        +K   NV++  V G    G  +I+  T  I+
Sbjct  891   YTPNVDENAGPNTLVTTVVAS--------DKDGDNVRFGFVGGGTSSGQFVIEEITGVIR  942

Query  397   FVK---NMNRDSTFI-VNKTITAEVLAIDEYTGKTSTGTVYVRVPDFNDN------CPTA  446
                   +++RD   + V            + T  TST  V V + D NDN      C T
Sbjct  943   LHSKAISLDRDKYELNVTAMDDGSCCVNGDATIHTSTAVVVVFITDVNDNKPVFKDCGTY  1002

Query  447   VLE-KDAVCSSSPSVVVSARTLNNRYTGPYTFALEDQP  483
               + ++   + SP + V A   +    G   +++  QP
Sbjct  1003  YPKVEEGAPNGSPVIKVQATDEDKGVNGQVKYSIVQQP  1040

 Score = 44.7 bits (104),  Expect = 0.008, Method: Compositional matrix adjust.
 Identities = 48/199 (24%), Positives = 73/199 (37%), Gaps = 19/199 (10%)

Query  27    DYQATQKITYRISGVGIDQPPFGIFVVDKNTGDINITAIVDREETPSFLITCRALNAQGL  86
             D     K++Y I      Q  F I       G + I   +DRE TP   +   A++
Sbjct  1926  DQGGKSKVSYAIDRSSDRQRQFSI----NQEGTVTIQRNLDREVTPRHQVKILAIDDGIP  1981

Query  87    DVEKPLILTVKILDINDNPPVFSQQIFMGEIEENSASNSLVMILNATDADEPNHLNSKIA  146
                    LTV + DINDNPP F +  +   + E+     +V IL   D D
Sbjct  1982  PKTATATLTVIVQDINDNPPKFLKD-YRPVLPEHVPPRKVVEILATDDDDRSKSNGPPFQ  2040

Query  147   FKIVSQEPAGTPMFLLSRNTGEVRTLTN-----------SLDREQASSYRLVVSGADKDG  195
             F++   +P    +   S    + +   N           S DREQ   Y + +   D
Sbjct  2041  FRL---DPGADDIIRASFKVEQDQKGANGDGMAIVSSLRSFDREQQKEYLIPIVIKDHGN  2097

Query  196   EGLSTQCECNIKVKDVNDN  214
               ++      + + DVNDN
Sbjct  2098  PAMTGTSTLTVVIGDVNDN  2116

 Score = 40.8 bits (94),  Expect = 0.11, Method: Compositional matrix adjust.
 Identities = 55/242 (23%), Positives = 102/242 (42%), Gaps = 22/242 (9%)
```

```
Query  4    KFAKPCREGE--DNSKRNP-IAKITSDYQATQKITYRISGVGIDQPPFGIFVVDKNTGDI  60
            +F  P  E E  +N K++  I  + +   A ++I Y +   G      G F +   +G +
Sbjct  678  QFYMPSYEAEIPENQKKDSDIISVKAKSFADREIRYTLKAQGQGA---GTFNIGPTSGIV  734

Query  61   NITAIVDREE--TPSF--LITCRALNAQGLDVEKPLILTVKILDINDNPPVFSQQIFMGE  116
             +   +D E+   P    L+     ++ G       + LT+++ D+NDN P F    +
Sbjct  735  KLAKELDFEDLRQPHVYSLVVTATEDSGGFSTS--VELTIRVTDVNDNAPKFELPDYQAH  792

Query  117  -IEENSASNSLVMILNATDADEPNHLNSKIAFKIVSQEPAGTPMFLLSRNTGEVRTLTNS  175
             ++E+    + ++ + A DAD  +  N++I + +VS +      F +  N   V
Sbjct  793  NVDEDIQIGTSILKVKAMDAD--SGANAEIEY-LVSDD-----HFAVDSNGIIVNNKQLD  844

Query  176  LDREQASSYRLVVSGADKDGEGLSTQCECNIKVKDVNDNFPMFRDSQYSARIEENILSSE  235
             D   A  Y  VV+  DK      +       +  K+ ND  P F    Y+  ++EN   +
Sbjct  845  ADNNNA-YYEFVVTAKDKGEPAKTGTATVRVYTKNKNDEEPKFSQQVYTPNVDENAGPNT  903

Query  236  LL  237
            L+
Sbjct  904  LV  905

 Score = 39.3 bits (90),  Expect = 0.30, Method: Compositional matrix adjust.
 Identities = 55/225 (24%), Positives = 88/225 (39%), Gaps = 24/225 (11%)

Query  75   LITCRALNAQGLDVEKPLILTVKILDINDNPPVFSQQIF--MGEIEENSASNSLVMILNA  132
            +IT +  NA GL       + + + D+ND PP F  +       ++ N+  N+ V  L A
Sbjct  539  IITNQGHNA-GLKYTDNQRVIIHVKDVNDEPPYFINRPLPMQAVVQLNAPPNTPVFTLQA  597

Query  133  TDADEPNHLNSKIAFKIVSQEPAGTPMFLLSRNTGEVRTLTNSLDREQASSYRLVVSGAD  192
             D D     +  I + IV     G   F +   +G VRT    L  +    Y L V   D
Sbjct  598  RDPD----TDHNIHYFIVRDRTGG--RFEVDERSGVVRTRGTDL-FQLDMEYVLYVKAED  650

Query  193  KDGE----GLSTQCECNIKVKDVNDNFPMFRDSQYSARIEEN-ILSSELLRFQVTDLDEE  247
            ++G+        +  E  + +        P F    Y A I EN    S+++  +     +
Sbjct  651  QNGKVDDRRFQSTPEERLSIVG-GKRAPQFYMPSYEAEIPENQKKDSDIISVKA----KS  705

Query  248  YTDNWLAVYFFTSGNEGNWFEIQTDPRTNEGILKVVKALDYEQLQ  292
            + D  +       G     F I         GI+K+ K LD+E L+
Sbjct  706  FADREIRYTLKAQGQGAGTFNIG----PTSGIVKLAKELDFEDLR  746

>AALB20_033171.P66522 | transcript=AALB20_033171.R66522 | gene=AALB20_033171
| organism=Anopheles_albimanus_STECLA_2020 |
gene_product=neural-cadherin, transcript variant X38 |
transcript_product=neural-cadherin,
transcript variant X14 | location=CM024292:75037560-75208393(+)
| protein_length=3131 | sequence_SO=chromosome
| SO=protein_coding_gene | is_pseudo=false
Length=3131

 Score = 144 bits (364),  Expect = 1e-34, Method: Compositional matrix adjust.
 Identities = 125/437 (29%), Positives = 211/437 (48%), Gaps = 33/437 (8%)

Query  26    SDYQATQKITYRISGVGID--QPPFGIFVVDKNTGDINITAIVDREET---PSFLITCRA  80
               D    Q I Y ++G GID   P    F +++ TG+I +   +DR++    P +  T  A
Sbjct  1580  GDKDRPQNIVYFLTGQGIDPDNPANSKFDINRTTGEIFVLKPLDRDQPNGRPQWRFTVFA  1639

Query  81    LNAQGLDVEKPLILTVKILDINDNPPVFSQQIFMGEIEENSASNSLVMILNATDADEPNH  140
               +  G  +      + V + DINDN P+F Q ++ G + EN  +  +VM + A D D+PN
Sbjct  1640  QDEGGEGLVGYADVQVNLKDINDNAPIFPQGVYFGNVTENGTAGMVVMTMTAVDYDDPNE  1699

Query  141   -LNSKIAFKI---VSQEPAGTPMFLLSRNTGEVRTLTNSLDREQASSYRLVVSGADKDGE  196
                N+K+ + I   V +E  G+P+F +   +TG ++T    LDRE+   Y + V      DG
Sbjct  1700  GTNAKLIYSIEKNVIEEETGSPIFEIEADTGVIKTAVCCLDRERTPDYSIQVVAM--DGG  1757

Query  197   GLSTQCECNIKVKDVNDNFPMFRDSQYSARIEENILSS--ELLRFQVTDLDEEYTDNWLA  254
               GL       +I+VKD+ND  P F   ++    ++E   ++  E+    VT  DE+ T+ +
Sbjct  1758  GLKGTGTASIRVKDINDMPPQFTKDEWFTEVDETDGTNLPEMPILTVTVHDEDETNKFQY  1817

Query  255   VYFFTSGNEGNWFEIQTDPRTNEGI--LKVVKALDYE---QLQSVKLSIAVKNKAEFHQS  309
                    SG   + F +    R N+G   LK+V+ LDYE   Q    +  I V +K E + +
```

```
Sbjct  1818  KVIDNSGYGADKFTMV---RNNDGTGSLKIVQPLDYEDPLQSNGFRFRIQVNDKGEDNDN  1874

Query  310   VISRYRVQSTPVTIQVINVREGIAFRPASKTFTVQKGISSKKLVDYILGTYQAIDEDTNK  369
                +Y V  + V +++ ++ +    +P  +   ++  +     V   L T++A D D
Sbjct  1875  --DKYHVAYSWVVVKLRDINDN---KPQFERPNIEVSVYENADVGKTLETFKATDPDQG-  1928

Query  370   AASNVKYVMGR-NDGGYLMIDSKTAEIKFVKNMNRDSTFIVNKTITAEVLAIDE-YTGKT  427
               S V Y + R +D       ++    +   +N++R+    V       ++LAID+    KT
Sbjct  1929  GKSKVSYAIDRSSDRQRQFSINQEGTVTIQRNLDRE----VTPRHQVKILAIDDGIPPKT  1984

Query  428   STGTVYVRVPDFNDNCP  444
             +T T+ V V D NDN P
Sbjct  1985  ATATLTVIVQDINDNPP  2001

 Score = 85.9 bits (211),  Expect = 1e-15, Method: Compositional matrix adjust.
 Identities = 108/420 (26%), Positives = 177/420 (42%), Gaps = 52/420 (12%)

Query  50    IFVVDKNTGDINITAIVDREETPSFLITCRAL----------NAQGLDVEKPLILTVKIL  99
              F ++  +G+I    + DRE+  ++ +   A            N Q   V K   + + I
Sbjct  1385  FFEINLQSGEIFTKVVFDREKQGAYALEVEARDGAPSARPNSNQQPNSVTK--FIRIGIA  1442

Query  100   DINDNPPVFSQQIFMGEIEENSASNSLVMILNATDADEPNHLNSKIAFKIVSQEPAGTPM  159
             D NDNPP F + ++  E++EN       V+ + A D DE    +S+I ++I S    G
Sbjct  1443  DKNDNPPYFDKALYEAEVDENEDIQHTVLTVTAKDHDE----SSRIRYEITSGNIGGA--  1496

Query  160   FLLSRNTGEVRTLTNSLDREQASSYRLVVSGADKDGEGLSTQCECNIKVKDVNDNFPMFR  219
             F +   TG +  +  +LD E    Y L ++ +D   E  +T     I VKDVNDN P+F
Sbjct  1497  FAVKNMTGAI-YVAGALDYETRKRYELRLAASDNLKENYTTVV---IHVKDVNDNPPVFE  1552

Query  220   DSQYSARIEENI---LSSELLRFQVTDLDEEYTDNWLAVYFFTSG--NEGNWFEIQTDPR  274
                Y  +I E      L   +L+   TD D++   N   VYF T    +  N    + D
Sbjct  1553  RPTYRTQITEEDDRNLPKRVLQVTATDGDKDRPQN--IVYFLTGQGIDPDNPANSKFDIN  1610

Query  275   TNEGILKVVKALDYEQLQS-VKLSIAVKNKAEFHQSVISRYRVQSTPVTIQVINVREGIA  333
                G + V+K LD +Q      +    V  + E  + ++    VQ     + + ++ +
Sbjct  1611  RTTGEIFVLKPLDRDQPNGRPQWRFTVFAQDEGGEGLVGYADVQ-----VNLKDINDNAP  1665

Query  334   FRPASKTFTVQKGISSKKLVDYILGTYQAIDEDTNKAASNVK--YVMGRN------DGGY  385
               P    F     ++        ++ T  A+D D     +N K  Y + +N
Sbjct  1666  IFPQGVYF---GNVTENGTAGMVVMTMTAVDYDDPNEGTNAKLIYSIEKNVIEEETGSPI  1722

Query  386   LMIDSKTAEIK-FVKNMNRDSTFIVNKTITAEVLAIDEYTGKTSTGTVYVRVPDFNDNCP  444
               I++ T  IK  V  ++R+ T        + +V+A+D   G   TGT  +RV D ND  P
Sbjct  1723  FEIEADTGVIKTAVCCLDRERT----PDYSIQVVAMDG-GGLKGTGTASIRVKDINDMPP  1777

 Score = 75.9 bits (185),  Expect = 2e-12, Method: Compositional matrix adjust.
 Identities = 108/429 (25%), Positives = 178/429 (41%), Gaps = 53/429 (12%)

Query  53    VDKNTGDINITAIVDREETPSFLITCRALNAQGLDVEKPLILTVKILDINDNPPVFSQ-Q  111
             +D++  ++N+TA+ D          +C  +N           ++ V I D+NDN PVF
Sbjct  950   LDRDKYELNVTAMDDG--------SC-CVNGDATIHTSTAVVVVFITDVNDNKPVFKDCG  1000

Query  112   IFMGEIEENSASNSLVMILNATDADEPNHLNSKIAFKIVSQEPAGTPMFLLSRNTGEVRT  171
              +  ++EE + + S V+ + ATD D+   +N ++ + IV Q        F +   TGEV T
Sbjct  1001  TYYPKVEEGAPNGSPVIKVQATDEDK--GVNGQVKYSIVQQPNQKGTKFTVDEETGEVST  1058

Query  172   LTNSLDRE--QASSYRLVVSGADKDGEGLSTQCECNIKVKDVNDNFPMFRDSQYSARIEE  229
                  DRE         + V   D+     L   C    +++ DVNDN P+F   +Y   +++
Sbjct  1059  -NKVFDREGDDGKFVSVTVKATDQGEPSLEGVCSFTVEITDVNDNPPLFDRQKYVENVKQ  1117

Query  230   NI-LSSELLRFQVTDLDEEYTDNWLAVYFFT---SGNEGNWFEIQTDPRTNEGILKVVKA  285
             +  + + +LR  V+  DE+  +N   VY  +   S N+  +FEIQ  P +   +LK  K
Sbjct  1118  DASIGTNILR--VSASDEDADNNGAIVYTLSAPHSPNDLEYFEIQ--PESGWIVLK--KP  1171

Query  286   LD-YEQLQSVKLSIAVK----------NKAEFHQSVISRYRVQSTPV----TIQVINVRE  330
             LD     + SV+    +K            + A+    +    R    P+         I++RE
Sbjct  1172  LDGCYSVSSVRERYRLKVYASDRGDPPSTADVDVELDVVDRNNKPPIWDRNIYGPIHIRE  1231

Query  331   GIAFRPASKTFTVQKGISSKKLVDYILGTYQAIDEDTNKAASNVKYVMGRNDGGYLMIDS  390
```

```
                +             GI     V Y L         TNK   +  Y+  R D G
Sbjct  1232  NVTVGTVVTAIKASSGIEGNPTVFYRL--MPGSTAQTNKL--HTFYLQQRPDNG-----D  1282

Query  391   KTAEIKFVKNMNRDSTFIVNKTITAEVLAIDEYTGKTSTGTVYVRVPDFNDNCPTAV-LE  449
               A+IK    ++ ++    N TI  E     +     S  TVY+++ D ND  P     E
Sbjct  1283  TWADIKVNHPLDFETIKEYNLTIRVENNGAQQL---ASEATVYIQLEDVNDEIPLFTERE  1339

Query  450   KDAVCSSSP  458
             ++ V    P
Sbjct  1340  QETVLEGEP  1348

 Score = 61.2 bits (147),  Expect = 5e-08, Method: Compositional matrix adjust.
 Identities = 87/366 (24%), Positives = 156/366 (43%), Gaps = 38/366 (10%)

Query  51    FVVDKNTGDINITAIVDRE-ETPSFL-ITCRALNAQGLDVEKPLILTVKILDINDNPPVF  108
             F VD+ TG+++   + DRE +   F+ +T +A +     +E      TV+I D+NDNPP+F
Sbjct  1047  FTVDEETGEVSTNKVFDREGDDGKFVSVTVKATDQGEPSLEGVCSFTVEITDVNDNPPLF  1106

Query  109   SQQIFMGEIEENSASNSLVMILNATDADEPNHLNSKIAFKIVS-QEPAGTPMFLLSRNTG  167
              +Q ++ ++++++   + ++ ++A+D D  N  N  I + + +   P     F +   +G
Sbjct  1107  DRQKYVENVKQDASIGTNILRVSASDEDADN--NGAIVYTLSAPHSPNDLEYFEIQPESG  1164

Query  168   EVRTLTNSLDREQASS-----YRLVVSGADKDGEGLSTQCECNIKVKDVNDNFPMFRDSQ  222
              +  L   LD   + S     YRL V  +D+ G+  ST            +N P   D
Sbjct  1165  WI-VLKKPLDGCYSVSSVRERYRLKVYASDR-GDPPSTADVDVELDVVDRNNKPPIWDRN  1222

Query  223   YSA--RIEENILSSELLRFQVTDLDEEYTDNWLAVYFFTSG-----NEGNWFEIQTDPRT  275
                   I EN+    ++             N    Y    G     N+ + F +Q  P
Sbjct  1223  IYGPIHIRENVTVGTVV--TAIKASSGIEGNPTVFYRLMPGSTAQTNKLHTFYLQQRPDN  1280

Query  276   NE--GILKVVKALDYEQLQSVKLSIAVKNKAEFHQSVISRYRVQSTPVTIQVINVREGIA  333
              +    +KV   LD+E ++   L+I V+N         ++       V IQ+ +V + I
Sbjct  1281  GDTWADIKVNHPLDFETIKEYNLTIRVENNG-------AQQLASEATVYIQLEDVNDEIP  1333

Query  334   FRPASKTFTVQKGISSKKLVDYILGTYQAIDEDTNKAASNVKYVM---GRNDG-GYLMID  389
                  +  TV +G      V  +     AID+D     + V Y +    RN+G  +  I+
Sbjct  1334  LFTEREQETVLEGEPIGTKVTQV----NAIDKDGTFPNNQVYYYIVDSPRNEGKDFFEIN  1389

Query  390   SKTAEI  395
              ++ EI
Sbjct  1390  LQSGEI  1395

 Score = 55.8 bits (133),  Expect = 3e-06, Method: Compositional matrix adjust.
 Identities = 49/158 (31%), Positives = 78/158 (49%), Gaps = 24/158 (15%)

Query  102   NDNPPVFSQQIFMGEIEENSASNSLVMILNATDADEPNHLNSKIAFKIVSQEPAGTP--M  159
             ND  P FSQQ++   ++EN+  N+LV  + A+D D  N     + F  V     GT
Sbjct  880   NDEEPKFSQQVYTPNVDENAGPNTLVTTVVASDKDGDN-----VRFGFVG---GGTSSGQ  931

Query  160   FLLSRNTGEVRTLTN--SLDREQASSYRLVVSGADK-------DGEGLSTQCECNIKVKD  210
             F++   TG +R  +   SLDR++   Y L V+  D        D    ++     + + D
Sbjct  932   FVIEEITGVIRLHSKAISLDRDK---YELNVTAMDDGSCCVNGDATIHTSTAVVVVFITD  988

Query  211   VNDNFPMFRD-SQYSARIEENILS-SELLRFQVTDLDE  246
             VNDN P+F+D   Y  ++EE   + S +++ Q TD D+
Sbjct  989   VNDNKPVFKDCGTYYPKVEEGAPNGSPVIKVQATDEDK  1026

 Score = 53.9 bits (128),  Expect = 1e-05, Method: Compositional matrix adjust.
 Identities = 70/306 (23%), Positives = 126/306 (41%), Gaps = 30/306 (10%)

Query  20    PIAKIT-SDYQATQKITYRI---SGVGIDQPPFGIFVVDKNTGDINITAIVDRE---ETP  72
             PI  +T  D   T K  Y++   SG G D+  F +   +  TG + I   +D E   ++
Sbjct  1800  PILTVTVHDEDETNKFQYKVIDNSGYGADK--FTMVRNNDGTGSLKIVQPLDYEDPLQSN  1857

Query  73    SFLITCRALNAQGLDVEKPLI------LTVKILDINDNPPVFSQQIFMGEIEENSASNSL  126
              F    + +N +G D +          + VK+ DINDN P F +      + EN+
Sbjct  1858  GFRFRIQ-VNDKGEDNDNDKYHVAYSWVVVKLRDINDNKPQFERPNIEVSVYENADVGKT  1916
```

```
Query  127   VMILNATDADEPNHLNSKIAFKIVSQEPAGTPMFLLSRNTGEVRTLTNSLDREQASSYRL  186
             +    ATD D+     SK+++ I   + +       S N     T+  +LDRE    +++
Sbjct  1917  LETFKATDPDQGG--KSKVSYAI---DRSSDRQRQFSINQEGTVTIQRNLDREVTPRHQV  1971

Query  187   VVSGADKDGEGLSTQCECNIKVKDVNDNFPMFRDSQYSARIEENILSSELLRFQVTDLDE  246
              +   D      +      + V+D+NDN P F    Y   + E++   +++    TD D+
Sbjct  1972  KILAIDDGIPPKTATATLTVIVQDINDNPPKFL-KDYRPVLPEHVPPRKVVEILATDDDD  2030

Query  247   EYTDNWLAVYFFTSGNEGNW----FEIQTDPRTNEG----ILKVVKALDYEQLQSVKLSI  298
                 N     F       +     F+++ D +   G    I+  +++ D EQ +   + I
Sbjct  2031  RSKSNGPPFQFRLDPGADDIIRASFKVEQDQKGANGDGMAIVSSLRSFDREQQKEYLIPI  2090

Query  299   AVKNKA  304
              +K+
Sbjct  2091  VIKDHG  2096

 Score = 51.6 bits (122),  Expect = 5e-05, Method: Compositional matrix adjust.
 Identities = 96/398 (24%), Positives = 163/398 (41%), Gaps = 54/398 (14%)

Query  106   PVFSQQIFMGEIEENSASNSLVMILNATDADEPNHLNSKIAFKIVSQ-EPAGTPMFLLSR  164
             P F    +  EI EN   +S ++ + A      +  + +I + + +Q + AGT  F +
Sbjct  677   PQFYMPSYEAEIPENQKKDSDIISVKAK-----SFADREIRYTLKAQGQGAGT--FNIGP  729

Query  165   NTGEVRTLTNSLDRE---QASSYRLVVSGADKDGEGLSTQCECNIKVKDVNDNFPMFRDS  221
              +G V+ L   LD E   Q   Y LVV+ A +D  G ST  E  I+V DVNDN P F
Sbjct  730   TSGIVK-LAKELDFEDLRQPHVYSLVVT-ATEDSGGFSTSVELTIRVTDVNDNAPKFELP  787

Query  222   QYSAR-IEENI-LSSELLRFQVTDLDEEYTDNWLAVYFFTSGNEGNWFEIQTDPRTNEGI  279
              Y A  ++E+I + + +L+ +  D D          + +  S +    F + ++     GI
Sbjct  788   DYQAHNVDEDIQIGTSILKVKAMDADSGAN---AEIEYLVSDDH---FAVDSN-----GI  836

Query  280   LKVVKALDYEQLQSV-KLSIAVKNKAEFHQSVISRYRVQSTPVTIQVINVREGIAFRPAS  338
             +   K LD +   +  +  +  K+K E  ++  +  RV +             E   F
Sbjct  837   IVNNKQLDADNNNAYYEFVVTAKDKGEPAKTGTATVRVYTKNKN------DEEPKFSQQV  890

Query  339   KTFTVQKGISSKKLVDYILGTYQAIDEDTNKAASNVKY--VMGRNDGGYLMIDSKTAEIK  396
              T  V +      LV  ++ +        +K   NV++  V G    G +I+  T  I+
Sbjct  891   YTPNVDENAGPNTLVTTVVAS--------DKDGDNVRFGFVGGGTSSGQFVIEEITGVIR  942

Query  397   FVK---NMNRDSTFI-VNKTITAEVLAIDEYTGKTSTGTVYVRVPDFNDN------CPTA  446
                  +++RD   + V             + T  TST  V V + D NDN      C T
Sbjct  943   LHSKAISLDRDKYELNVTAMDDGSCCVNGDATIHTSTAVVVVFITDVNDNKPVFKDCGTY  1002

Query  447   VLE-KDAVCSSSPSVVVSARTLNNRYTGPYTFALEDQP  483
               + ++   + SP + V A    +    G   +++  QP
Sbjct  1003  YPKVEEGAPNGSPVIKVQATDEDKGVNGQVKYSIVQQP  1040

 Score = 44.7 bits (104),  Expect = 0.008, Method: Compositional matrix adjust.
 Identities = 48/199 (24%), Positives = 73/199 (37%), Gaps = 19/199 (10%)

Query  27    DYQATQKITYRISGVGIDQPPFGIFVVDKNTGDINITAIVDREETPSFLITCRALNAQGL  86
             D     K++Y I      Q  F I        G + I   +DRE TP   +    A++
Sbjct  1926  DQGGKSKVSYAIDRSSDRQRQFSI----NQEGTVTIQRNLDREVTPRHQVKILAIDDGIP  1981

Query  87    DVEKPLILTVKILDINDNPPVFSQQIFMGEIEENSASNSLVMILNATDADEPNHLNSKIA  146
                    LTV + DINDNPP F +  +    + E+      +V IL   D D
Sbjct  1982  PKTATATLTVIVQDINDNPPKFLKD-YRPVLPEHVPPRKVVEILATDDDDRSKSNGPPFQ  2040

Query  147   FKIVSQEPAGTPMFLLSRNTGEVRTLTN-----------SLDREQASSYRLVVSGADKDG  195
             F++   +P    +   S    + +   N           S DREQ    Y + +   D
Sbjct  2041  FRL---DPGADDIIRASFKVEQDQKGANGDGMAIVSSLRSFDREQQKEYLIPIVIKDHGN  2097

Query  196   EGLSTQCECNIKVKDVNDN  214
               ++      + + DVNDN
Sbjct  2098  PAMTGTSTLTVVIGDVNDN  2116

 Score = 40.8 bits (94),  Expect = 0.11, Method: Compositional matrix adjust.
 Identities = 55/242 (23%), Positives = 102/242 (42%), Gaps = 22/242 (9%)
```

```
Query  4    KFAKPCREGE--DNSKRNP-IAKITSDYQATQKITYRISGVGIDQPPFGIFVVDKNTGDI  60
            +F  P  E E  +N K++  I  + +   A ++I Y +   G      G F +   +G +
Sbjct  678  QFYMPSYEAEIPENQKKDSDIISVKAKSFADREIRYTLKAQGQGA---GTFNIGPTSGIV  734

Query  61   NITAIVDREE--TPSF--LITCRALNAQGLDVEKPLILTVKILDINDNPPVFSQQIFMGE  116
             +   +D E+   P    L+     ++ G       + LT+++ D+NDN P F    +
Sbjct  735  KLAKELDFEDLRQPHVYSLVVTATEDSGGFSTS--VELTIRVTDVNDNAPKFELPDYQAH  792

Query  117  -IEENSASNSLVMILNATDADEPNHLNSKIAFKIVSQEPAGTPMFLLSRNTGEVRTLTNS  175
             ++E+    + ++ + A DAD  +  N++I + +VS +      F +  N   V
Sbjct  793  NVDEDIQIGTSILKVKAMDAD--SGANAEIEY-LVSDD-----HFAVDSNGIIVNNKQLD  844

Query  176  LDREQASSYRLVVSGADKDGEGLSTQCECNIKVKDVNDNFPMFRDSQYSARIEENILSSE  235
             D   A  Y  VV+  DK      +       +  K+ ND  P F    Y+  ++EN   +
Sbjct  845  ADNNNA-YYEFVVTAKDKGEPAKTGTATVRVYTKNKNDEEPKFSQQVYTPNVDENAGPNT  903

Query  236  LL  237
            L+
Sbjct  904  LV  905

 Score = 39.3 bits (90),  Expect = 0.30, Method: Compositional matrix adjust.
 Identities = 55/225 (24%), Positives = 88/225 (39%), Gaps = 24/225 (11%)

Query  75   LITCRALNAQGLDVEKPLILTVKILDINDNPPVFSQQIF--MGEIEENSASNSLVMILNA  132
            +IT +   NA GL       + + + D+ND PP F  +        ++ N+  N+ V  L A
Sbjct  539  IITNQGHNA-GLKYTDNQRVIIHVKDVNDEPPYFINRPLPMQAVVQLNAPPNTPVFTLQA  597

Query  133  TDADEPNHLNSKIAFKIVSQEPAGTPMFLLSRNTGEVRTLTNSLDREQASSYRLVVSGAD  192
             D D     +  I + IV     G   F +   +G VRT    L  +    Y L V   D
Sbjct  598  RDPD----TDHNIHYFIVRDRTGG--RFEVDERSGVVRTRGTDL-FQLDMEYVLYVKAED  650

Query  193  KDGE----GLSTQCECNIKVKDVNDNFPMFRDSQYSARIEEN-ILSSELLRFQVTDLDEE  247
            ++G+        +  E  + +        P F    Y A I EN    S+++  +     +
Sbjct  651  QNGKVDDRRFQSTPEERLSIVG-GKRAPQFYMPSYEAEIPENQKKDSDIISVKA----KS  705

Query  248  YTDNWLAVYFFTSGNEGNWFEIQTDPRTNEGILKVVKALDYEQLQ  292
            + D  +       G     F I        GI+K+ K LD+E L+
Sbjct  706  FADREIRYTLKAQGQGAGTFNIG----PTSGIVKLAKELDFEDLR  746

>AALB20_033171.P66521 | transcript=AALB20_033171.R66521 | gene=AALB20_033171
| organism=Anopheles_albimanus_STECLA_2020 |
gene_product=neural-cadherin, transcript variant X38 |
transcript_product=neural-cadherin,
transcript variant X17 | location=CM024292:75037560-75208393(+)
| protein_length=3131 | sequence_SO=chromosome
| SO=protein_coding_gene | is_pseudo=false
Length=3131

 Score = 144 bits (364),  Expect = 1e-34, Method: Compositional matrix adjust.
 Identities = 125/437 (29%), Positives = 211/437 (48%), Gaps = 33/437 (8%)

Query  26    SDYQATQKITYRISGVGID--QPPFGIFVVDKNTGDINITAIVDREET---PSFLITCRA  80
               D    Q I Y ++G GID   P    F +++ TG+I +   +DR++    P +  T  A
Sbjct  1580  GDKDRPQNIVYFLTGQGIDPDNPANSKFDINRTTGEIFVLKPLDRDQPNGRPQWRFTVFA  1639

Query  81    LNAQGLDVEKPLILTVKILDINDNPPVFSQQIFMGEIEENSASNSLVMILNATDADEPNH  140
               +  G  +      + V + DINDN P+F Q ++ G + EN  +  +VM + A D D+PN
Sbjct  1640  QDEGGEGLVGYADVQVNLKDINDNAPIFPQGVYFGNVTENGTAGMVVMTMTAVDYDDPNE  1699

Query  141   -LNSKIAFKI---VSQEPAGTPMFLLSRNTGEVRTLTNSLDREQASSYRLVVSGADKDGE  196
                 N+K+ + I    V +E  G+P+F +   +TG ++T     LDRE+    Y + V      DG
Sbjct  1700  GTNAKLIYSIEKNVIEEETGSPIFEIEADTGVIKTAVCCLDRERTPDYSIQVVAM--DGG  1757

Query  197   GLSTQCECNIKVKDVNDNFPMFRDSQYSARIEENILSS--ELLRFQVTDLDEEYTDNWLA  254
               GL       +I+VKD+ND  P F   ++   ++E   ++  E+    VT  DE+ T+ +
Sbjct  1758  GLKGTGTASIRVKDINDMPPQFTKDEWFTEVDETDGTNLPEMPILTVTVHDEDETNKFQY  1817

Query  255   VYFFTSGNEGNWFEIQTDPRTNEGI--LKVVKALDYE---QLQSVKLSIAVKNKAEFHQS  309
```

```
                  SG   + F +    R N+G   LK+V+ LDYE   Q    +  I V +K E + +
Sbjct  1818  KVIDNSGYGADKFTMV---RNNDGTGSLKIVQPLDYEDPLQSNGFRFRIQVNDKGEDNDN  1874

Query  310   VISRYRVQSTPVTIQVINVREGIAFRPASKTFTVQKGISSKKLVDYILGTYQAIDEDTNK  369
                +Y V  + V +++ ++ +    +P  +   ++  +     V   L T++A D D
Sbjct  1875  --DKYHVAYSWVVVKLRDINDN---KPQFERPNIEVSVYENADVGKTLETFKATDPDQG-  1928

Query  370   AASNVKYVMGR-NDGGYLMIDSKTAEIKFVKNMNRDSTFIVNKTITAEVLAIDE-YTGKT  427
               S V Y + R +D       ++   +   +N++R+    V      ++LAID+    KT
Sbjct  1929  GKSKVSYAIDRSSDRQRQFSINQEGTVTIQRNLDRE----VTPRHQVKILAIDDGIPPKT  1984

Query  428   STGTVYVRVPDFNDNCP  444
             +T T+ V V D NDN P
Sbjct  1985  ATATLTVIVQDINDNPP  2001

 Score = 85.9 bits (211),  Expect = 1e-15, Method: Compositional matrix adjust.
 Identities = 108/420 (26%), Positives = 177/420 (42%), Gaps = 52/420 (12%)

Query  50    IFVVDKNTGDINITAIVDREETPSFLITCRAL----------NAQGLDVEKPLILTVKIL  99
              F ++  +G+I    + DRE+  ++ +   A           N Q   V K   + + I
Sbjct  1385  FFEINLQSGEIFTKVVFDREKQGAYALEVEARDGAPSARPNSNQQPNSVTK--FIRIGIA  1442

Query  100   DINDNPPVFSQQIFMGEIEENSASNSLVMILNATDADEPNHLNSKIAFKIVSQEPAGTPM  159
             D NDNPP F + ++  E++EN      V+ + A D DE    +S+I ++I S    G
Sbjct  1443  DKNDNPPYFDKALYEAEVDENEDIQHTVLTVTAKDHDE----SSRIRYEITSGNIGGA--  1496

Query  160   FLLSRNTGEVRTLTNSLDREQASSYRLVVSGADKDGEGLSTQCECNIKVKDVNDNFPMFR  219
             F +   TG +  +  +LD E    Y L ++ +D   E  +T     I VKDVNDN P+F
Sbjct  1497  FAVKNMTGAI-YVAGALDYETRKRYELRLAASDNLKENYTTVV---IHVKDVNDNPPVFE  1552

Query  220   DSQYSARIEENI---LSSELLRFQVTDLDEEYTDNWLAVYFFTSG--NEGNWFEIQTDPR  274
                Y  +I E     L   +L+   TD D++   N   VYF T    +  N    + D
Sbjct  1553  RPTYRTQITEEDDRNLPKRVLQVTATDGDKDRPQN--IVYFLTGQGIDPDNPANSKFDIN  1610

Query  275   TNEGILKVVKALDYEQLQS-VKLSIAVKNKAEFHQSVISRYRVQSTPVTIQVINVREGIA  333
                G + V+K LD +Q     +    V  + E  + ++    VQ     + + ++ +
Sbjct  1611  RTTGEIFVLKPLDRDQPNGRPQWRFTVFAQDEGGEGLVGYADVQ-----VNLKDINDNAP  1665

Query  334   FRPASKTFTVQKGISSKKLVDYILGTYQAIDEDTNKAASNVK--YVMGRN------DGGY  385
               P    F     ++       ++ T  A+D D     +N K  Y + +N
Sbjct  1666  IFPQGVYF---GNVTENGTAGMVVMTMTAVDYDDPNEGTNAKLIYSIEKNVIEEETGSPI  1722

Query  386   LMIDSKTAEIK-FVKNMNRDSTFIVNKTITAEVLAIDEYTGKTSTGTVYVRVPDFNDNCP  444
               I++ T  IK  V  ++R+ T       + +V+A+D   G   TGT  +RV D ND  P
Sbjct  1723  FEIEADTGVIKTAVCCLDRERT----PDYSIQVVAMDG-GGLKGTGTASIRVKDINDMPP  1777

 Score = 75.9 bits (185),  Expect = 2e-12, Method: Compositional matrix adjust.
 Identities = 108/429 (25%), Positives = 178/429 (41%), Gaps = 53/429 (12%)

Query  53    VDKNTGDINITAIVDREETPSFLITCRALNAQGLDVEKPLILTVKILDINDNPPVFSQ-Q  111
             +D++  ++N+TA+ D         +C  +N          ++ V I D+NDN PVF
Sbjct  950   LDRDKYELNVTAMDDG--------SC-CVNGDATIHTSTAVVVVFITDVNDNKPVFKDCG  1000

Query  112   IFMGEIEENSASNSLVMILNATDADEPNHLNSKIAFKIVSQEPAGTPMFLLSRNTGEVRT  171
              +  ++EE + + S V+ + ATD D+   +N ++ + IV Q       F +   TGEV T
Sbjct  1001  TYYPKVEEGAPNGSPVIKVQATDEDK--GVNGQVKYSIVQQPNQKGTKFTVDEETGEVST  1058

Query  172   LTNSLDRE--QASSYRLVVSGADKDGEGLSTQCECNIKVKDVNDNFPMFRDSQYSARIEE  229
                  DRE        + V   D+    L   C   +++ DVNDN P+F   +Y   +++
Sbjct  1059  -NKVFDREGDDGKFVSVTVKATDQGEPSLEGVCSFTVEITDVNDNPPLFDRQKYVENVKQ  1117

Query  230   NI-LSSELLRFQVTDLDEEYTDNWLAVYFFT---SGNEGNWFEIQTDPRTNEGILKVVKA  285
             +  + + +LR  V+  DE+  +N   VY +    S N+  +FEIQ  P +   +LK  K
Sbjct  1118  DASIGTNILR--VSASDEDADNNGAIVYTLSAPHSPNDLEYFEIQ--PESGWIVLK--KP  1171

Query  286   LD-YEQLQSVKLSIAVK----------NKAEFHQSVISRYRVQSTPV----TIQVINVRE  330
             LD    + SV+    +K          + A+    +    R    P+        I++RE
Sbjct  1172  LDGCYSVSSVRERYRLKVYASDRGDPPSTADVDVELDVVDRNNKPPIWDRNIYGPIHIRE  1231
```

```
Query  331    GIAFRPASKTFTVQKGISSKKLVDYILGTYQAIDEDTNKAASNVKYVMGRNDGGYLMIDS  390
               +             GI     V Y L         TNK   +  Y+  R D G
Sbjct  1232   NVTVGTVVTAIKASSGIEGNPTVFYRL--MPGSTAQTNKL--HTFYLQQRPDNG-----D  1282

Query  391    KTAEIKFVKNMNRDSTFIVNKTITAEVLAIDEYTGKTSTGTVYVRVPDFNDNCPTAV-LE  449
                A+IK    ++ ++    N TI  E     +     S  TVY+++ D ND  P     E
Sbjct  1283   TWADIKVNHPLDFETIKEYNLTIRVENNGAQQL---ASEATVYIQLEDVNDEIPLFTERE  1339

Query  450    KDAVCSSSP  458
              ++ V    P
Sbjct  1340   QETVLEGEP  1348

 Score = 61.2 bits (147),  Expect = 5e-08, Method: Compositional matrix adjust.
 Identities = 87/366 (24%), Positives = 156/366 (43%), Gaps = 38/366 (10%)

Query  51     FVVDKNTGDINITAIVDRE-ETPSFL-ITCRALNAQGLDVEKPLILTVKILDINDNPPVF  108
              F VD+ TG+++   + DRE +   F+ +T +A +     +E     TV+I D+NDNPP+F
Sbjct  1047   FTVDEETGEVSTNKVFDREGDDGKFVSVTVKATDQGEPSLEGVCSFTVEITDVNDNPPLF  1106

Query  109    SQQIFMGEIEENSASNSLVMILNATDADEPNHLNSKIAFKIVS-QEPAGTPMFLLSRNTG  167
               +Q ++  ++++++  + ++ ++A+D D  N  N  I + + +   P     F +   +G
Sbjct  1107   DRQKYVENVKQDASIGTNILRVSASDEDADN--NGAIVYTLSAPHSPNDLEYFEIQPESG  1164

Query  168    EVRTLTNSLDREQASS-----YRLVVSGADKDGEGLSTQCECNIKVKDVNDNFPMFRDSQ  222
               +  L   LD   + S     YRL V  +D+ G+  ST            +N P   D
Sbjct  1165   WI-VLKKPLDGCYSVSSVRERYRLKVYASDR-GDPPSTADVDVELDVVDRNNKPPIWDRN  1222

Query  223    YSA--RIEENILSSELLRFQVTDLDEEYTDNWLAVYFFTSG-----NEGNWFEIQTDPRT  275
                    I EN+    ++             N    Y    G     N+ + F +Q  P
Sbjct  1223   IYGPIHIRENVTVGTVV--TAIKASSGIEGNPTVFYRLMPGSTAQTNKLHTFYLQQRPDN  1280

Query  276    NE--GILKVVKALDYEQLQSVKLSIAVKNKAEFHQSVISRYRVQSTPVTIQVINVREGIA  333
               +    +KV   LD+E ++   L+I V+N         ++       V IQ+ +V + I
Sbjct  1281   GDTWADIKVNHPLDFETIKEYNLTIRVENNG-------AQQLASEATVYIQLEDVNDEIP  1333

Query  334    FRPASKTFTVQKGISSKKLVDYILGTYQAIDEDTNKAASNVKYVM---GRNDG-GYLMID  389
                   +  TV +G      V  +     AID+D     + V Y +    RN+G  +  I+
Sbjct  1334   LFTEREQETVLEGEPIGTKVTQV----NAIDKDGTFPNNQVYYYIVDSPRNEGKDFFEIN  1389

Query  390    SKTAEI  395
               ++ EI
Sbjct  1390   LQSGEI  1395

 Score = 55.8 bits (133),  Expect = 3e-06, Method: Compositional matrix adjust.
 Identities = 49/158 (31%), Positives = 78/158 (49%), Gaps = 24/158 (15%)

Query  102    NDNPPVFSQQIFMGEIEENSASNSLVMILNATDADEPNHLNSKIAFKIVSQEPAGTP--M  159
              ND  P FSQQ++   ++EN+  N+LV  + A+D D  N     + F  V     GT
Sbjct  880    NDEEPKFSQQVYTPNVDENAGPNTLVTTVVASDKDGDN-----VRFGFVG---GGTSSGQ  931

Query  160    FLLSRNTGEVRTLTN--SLDREQASSYRLVVSGADK-------DGEGLSTQCECNIKVKD  210
              F++   TG +R  +   SLDR++   Y L V+  D        D    ++     + + D
Sbjct  932    FVIEEITGVIRLHSKAISLDRDK---YELNVTAMDDGSCCVNGDATIHTSTAVVVVFITD  988

Query  211    VNDNFPMFRD-SQYSARIEENILS-SELLRFQVTDLDE  246
              VNDN P+F+D   Y  ++EE   + S +++ Q TD D+
Sbjct  989    VNDNKPVFKDCGTYYPKVEEGAPNGSPVIKVQATDEDK  1026

 Score = 53.9 bits (128),  Expect = 1e-05, Method: Compositional matrix adjust.
 Identities = 70/306 (23%), Positives = 126/306 (41%), Gaps = 30/306 (10%)

Query  20     PIAKIT-SDYQATQKITYRI---SGVGIDQPPFGIFVVDKNTGDINITAIVDRE---ETP  72
              PI  +T  D   T K  Y++   SG G D+  F +   +  TG + I   +D E   ++
Sbjct  1800   PILTVTVHDEDETNKFQYKVIDNSGYGADK--FTMVRNNDGTGSLKIVQPLDYEDPLQSN  1857

Query  73     SFLITCRALNAQGLDVEKPLI------LTVKILDINDNPPVFSQQIFMGEIEENSASNSL  126
               F    + +N +G D +          + VK+ DINDN P F +      + EN+
Sbjct  1858   GFRFRIQ-VNDKGEDNDNDKYHVAYSWVVVKLRDINDNKPQFERPNIEVSVYENADVGKT  1916
```

```
Query  127   VMILNATDADEPNHLNSKIAFKIVSQEPAGTPMFLLSRNTGEVRTLTNSLDREQASSYRL  186
             +    ATD D+     SK+++ I   + +       S N     T+  +LDRE    +++
Sbjct  1917  LETFKATDPDQGG--KSKVSYAI---DRSSDRQRQFSINQEGTVTIQRNLDREVTPRHQV  1971

Query  187   VVSGADKDGEGLSTQCECNIKVKDVNDNFPMFRDSQYSARIEENILSSELLRFQVTDLDE  246
              +   D      +      + V+D+NDN P F    Y   + E++   +++    TD D+
Sbjct  1972  KILAIDDGIPPKTATATLTVIVQDINDNPPKFL-KDYRPVLPEHVPPRKVVEILATDDDD  2030

Query  247   EYTDNWLAVYFFTSGNEGNW----FEIQTDPRTNEG----ILKVVKALDYEQLQSVKLSI  298
                 N     F       +     F+++ D +   G    I+  +++ D EQ +   + I
Sbjct  2031  RSKSNGPPFQFRLDPGADDIIRASFKVEQDQKGANGDGMAIVSSLRSFDREQQKEYLIPI  2090

Query  299   AVKNKA  304
              +K+
Sbjct  2091  VIKDHG  2096

 Score = 51.6 bits (122),  Expect = 5e-05, Method: Compositional matrix adjust.
 Identities = 96/398 (24%), Positives = 163/398 (41%), Gaps = 54/398 (14%)

Query  106   PVFSQQIFMGEIEENSASNSLVMILNATDADEPNHLNSKIAFKIVSQ-EPAGTPMFLLSR  164
             P F    +  EI EN   +S ++ + A      +  + +I + + +Q + AGT  F +
Sbjct  677   PQFYMPSYEAEIPENQKKDSDIISVKAK-----SFADREIRYTLKAQGQGAGT--FNIGP  729

Query  165   NTGEVRTLTNSLDRE---QASSYRLVVSGADKDGEGLSTQCECNIKVKDVNDNFPMFRDS  221
              +G V+ L   LD E   Q   Y LVV+ A +D  G ST  E  I+V DVNDN P F
Sbjct  730   TSGIVK-LAKELDFEDLRQPHVYSLVVT-ATEDSGGFSTSVELTIRVTDVNDNAPKFELP  787

Query  222   QYSAR-IEENI-LSSELLRFQVTDLDEEYTDNWLAVYFFTSGNEGNWFEIQTDPRTNEGI  279
              Y A  ++E+I + + +L+ +  D D         + +  S +    F + ++     GI
Sbjct  788   DYQAHNVDEDIQIGTSILKVKAMDADSGAN---AEIEYLVSDDH---FAVDSN-----GI  836

Query  280   LKVVKALDYEQLQSV-KLSIAVKNKAEFHQSVISRYRVQSTPVTIQVINVREGIAFRPAS  338
             +   K LD +   +  +  +  K+K E  ++  +  RV +           E   F
Sbjct  837   IVNNKQLDADNNNAYYEFVVTAKDKGEPAKTGTATVRVYTKNKN------DEEPKFSQQV  890

Query  339   KTFTVQKGISSKKLVDYILGTYQAIDEDTNKAASNVKY--VMGRNDGGYLMIDSKTAEIK  396
              T  V +      LV  ++ +        +K   NV++  V G    G  +I+  T  I+
Sbjct  891   YTPNVDENAGPNTLVTTVVAS--------DKDGDNVRFGFVGGGTSSGQFVIEEITGVIR  942

Query  397   FVK---NMNRDSTFI-VNKTITAEVLAIDEYTGKTSTGTVYVRVPDFNDN------CPTA  446
                   +++RD   + V            + T  TST  V V + D NDN      C T
Sbjct  943   LHSKAISLDRDKYELNVTAMDDGSCCVNGDATIHTSTAVVVVFITDVNDNKPVFKDCGTY  1002

Query  447   VLE-KDAVCSSSPSVVVSARTLNNRYTGPYTFALEDQP  483
               + ++   + SP + V A   +    G   +++  QP
Sbjct  1003  YPKVEEGAPNGSPVIKVQATDEDKGVNGQVKYSIVQQP  1040

 Score = 44.7 bits (104),  Expect = 0.008, Method: Compositional matrix adjust.
 Identities = 48/199 (24%), Positives = 73/199 (37%), Gaps = 19/199 (10%)

Query  27    DYQATQKITYRISGVGIDQPPFGIFVVDKNTGDINITAIVDREETPSFLITCRALNAQGL  86
             D     K++Y I      Q  F I       G + I   +DRE TP   +   A++
Sbjct  1926  DQGGKSKVSYAIDRSSDRQRQFSI----NQEGTVTIQRNLDREVTPRHQVKILAIDDGIP  1981

Query  87    DVEKPLILTVKILDINDNPPVFSQQIFMGEIEENSASNSLVMILNATDADEPNHLNSKIA  146
                    LTV + DINDNPP F +  +   + E+     +V IL   D D
Sbjct  1982  PKTATATLTVIVQDINDNPPKFLKD-YRPVLPEHVPPRKVVEILATDDDDRSKSNGPPFQ  2040

Query  147   FKIVSQEPAGTPMFLLSRNTGEVRTLTN-----------SLDREQASSYRLVVSGADKDG  195
             F++   +P    +   S    + +   N           S DREQ   Y + +   D
Sbjct  2041  FRL---DPGADDIIRASFKVEQDQKGANGDGMAIVSSLRSFDREQQKEYLIPIVIKDHGN  2097

Query  196   EGLSTQCECNIKVKDVNDN  214
               ++      + + DVNDN
Sbjct  2098  PAMTGTSTLTVVIGDVNDN  2116

 Score = 40.8 bits (94),  Expect = 0.11, Method: Compositional matrix adjust.
```

```
 Identities = 55/242 (23%), Positives = 102/242 (42%), Gaps = 22/242 (9%)

Query  4    KFAKPCREGE--DNSKRNP-IAKITSDYQATQKITYRISGVGIDQPPFGIFVVDKNTGDI  60
            +F  P  E E  +N K++  I  + +   A ++I Y +   G      G F +   +G +
Sbjct  678  QFYMPSYEAEIPENQKKDSDIISVKAKSFADREIRYTLKAQGQGA---GTFNIGPTSGIV  734

Query  61   NITAIVDREE--TPSF--LITCRALNAQGLDVEKPLILTVKILDINDNPPVFSQQIFMGE  116
             +   +D E+    P    L+      ++ G       + LT+++ D+NDN P F    +
Sbjct  735  KLAKELDFEDLRQPHVYSLVVTATEDSGGFSTS--VELTIRVTDVNDNAPKFELPDYQAH  792

Query  117  -IEENSASNSLVMILNATDADEPNHLNSKIAFKIVSQEPAGTPMFLLSRNTGEVRTLTNS  175
             ++E+    + ++ + A DAD  +  N++I + +VS +      F +  N   V
Sbjct  793  NVDEDIQIGTSILKVKAMDAD--SGANAEIEY-LVSDD-----HFAVDSNGIIVNNKQLD  844

Query  176  LDREQASSYRLVVSGADKDGEGLSTQCECNIKVKDVNDNFPMFRDSQYSARIEENILSSE  235
             D   A  Y  VV+  DK      +       +  K+ ND  P F    Y+  ++EN   +
Sbjct  845  ADNNNA-YYEFVVTAKDKGEPAKTGTATVRVYTKNKNDEEPKFSQQVYTPNVDENAGPNT  903

Query  236  LL  237
            L+
Sbjct  904  LV  905

 Score = 39.3 bits (90),  Expect = 0.30, Method: Compositional matrix adjust.
 Identities = 55/225 (24%), Positives = 88/225 (39%), Gaps = 24/225 (11%)

Query  75   LITCRALNAQGLDVEKPLILTVKILDINDNPPVFSQQIF--MGEIEENSASNSLVMILNA  132
            +IT +  NA GL       + + + D+ND PP F  +        ++ N+  N+ V  L A
Sbjct  539  IITNQGHNA-GLKYTDNQRVIIHVKDVNDEPPYFINRPLPMQAVVQLNAPPNTPVFTLQA  597

Query  133  TDADEPNHLNSKIAFKIVSQEPAGTPMFLLSRNTGEVRTLTNSLDREQASSYRLVVSGAD  192
             D D     +  I + IV     G   F +   +G VRT    L  +    Y L V   D
Sbjct  598  RDPD----TDHNIHYFIVRDRTGG--RFEVDERSGVVRTRGTDL-FQLDMEYVLYVKAED  650

Query  193  KDGE----GLSTQCECNIKVKDVNDNFPMFRDSQYSARIEEN-ILSSELLRFQVTDLDEE  247
            ++G+        +  E  + +        P F    Y A I EN    S+++  +     +
Sbjct  651  QNGKVDDRRFQSTPEERLSIVG-GKRAPQFYMPSYEAEIPENQKKDSDIISVKA----KS  705

Query  248  YTDNWLAVYFFTSGNEGNWFEIQTDPRTNEGILKVVKALDYEQLQ  292
            + D  +       G     F I        GI+K+ K LD+E L+
Sbjct  706  FADREIRYTLKAQGQGAGTFNIG----PTSGIVKLAKELDFEDLR  746

>AALB20_033171.P66520 | transcript=AALB20_033171.R66520 | gene=AALB20_033171
| organism=Anopheles_albimanus_STECLA_2020 |
gene_product=neural-cadherin, transcript variant X38 |
transcript_product=neural-cadherin,
transcript variant X13 | location=CM024292:75037560-75208393(+)
| protein_length=3131 | sequence_SO=chromosome
| SO=protein_coding_gene | is_pseudo=false
Length=3131

 Score = 144 bits (364),  Expect = 1e-34, Method: Compositional matrix adjust.
 Identities = 125/437 (29%), Positives = 211/437 (48%), Gaps = 33/437 (8%)

Query  26    SDYQATQKITYRISGVGID--QPPFGIFVVDKNTGDINITAIVDREET---PSFLITCRA  80
               D    Q I Y ++G GID   P    F +++ TG+I +   +DR++    P +  T  A
Sbjct  1580  GDKDRPQNIVYFLTGQGIDPDNPANSKFDINRTTGEIFVLKPLDRDQPNGRPQWRFTVFA  1639

Query  81    LNAQGLDVEKPLILTVKILDINDNPPVFSQQIFMGEIEENSASNSLVMILNATDADEPNH  140
               +  G  +     + V + DINDN P+F Q ++ G + EN  +  +VM + A D D+PN
Sbjct  1640  QDEGGEGLVGYADVQVNLKDINDNAPIFPQGVYFGNVTENGTAGMVVMTMTAVDYDDPNE  1699

Query  141   -LNSKIAFKI---VSQEPAGTPMFLLSRNTGEVRTLTNSLDREQASSYRLVVSGADKDGE  196
                 N+K+ + I   V +E  G+P+F +  +TG ++T    LDRE+   Y + V     DG
Sbjct  1700  GTNAKLIYSIEKNVIEEETGSPIFEIEADTGVIKTAVCCLDRERTPDYSIQVVAM--DGG  1757

Query  197   GLSTQCECNIKVKDVNDNFPMFRDSQYSARIEENILSS--ELLRFQVTDLDEEYTDNWLA  254
             GL       +I+VKD+ND  P F   ++   ++E   ++  E+    VT  DE+ T+ +
Sbjct  1758  GLKGTGTASIRVKDINDMPPQFTKDEWFTEVDETDGTNLPEMPILTVTVHDEDETNKFQY  1817
```

```
Query  255   VYFFTSGNEGNWFEIQTDPRTNEGI--LKVVKALDYE---QLQSVKLSIAVKNKAEFHQS  309
                  SG    + F +    R N+G   LK+V+ LDYE   Q    +  I V +K E + +
Sbjct  1818  KVIDNSGYGADKFTMV---RNNDGTGSLKIVQPLDYEDPLQSNGFRFIQVNDKGEDNDN  1874

Query  310   VISRYRVQSTPVTIQVINVREGIAFRPASKTFTVQKGISSKKLVDYILGTYQAIDEDTNK  369
                +Y V  + V +++ ++ +     +P  +   ++  +      V   L T++A D D
Sbjct  1875  --DKYHVAYSWVVVKLRDINDN---KPQFERPNIEVSVYENADVGKTLETFKATDPDQG-  1928

Query  370   AASNVKYVMGR-NDGGYLMIDSKTAEIKFVKNMNRDSTFIVNKTITAEVLAIDE-YTGKT  427
               S V Y + R +D       ++    +   +N++R+    V       ++LAID+    KT
Sbjct  1929  GKSKVSYAIDRSSDRQRQFSINQEGTVTIQRNLDRE----VTPRHQVKILAIDDGIPPKT  1984

Query  428   STGTVYVRVPDFNDNCP  444
              +T T+ V V D NDN P
Sbjct  1985  ATATLTVIVQDINDNPP  2001

 Score = 85.9 bits (211),  Expect = 1e-15, Method: Compositional matrix adjust.
 Identities = 108/420 (26%), Positives = 177/420 (42%), Gaps = 52/420 (12%)

Query  50    IFVVDKNTGDINITAIVDREETPSFLITCRAL----------NAQGLDVEKPLILTVKIL  99
              F ++  +G+I    + DRE+  ++ +   A            N Q   V K   + + I
Sbjct  1385  FFEINLQSGEIFTKVVFDREKQGAYALEVEARDGAPSARPNSNQQPNSVTK--FIRIGIA  1442

Query  100   DINDNPPVFSQQIFMGEIEENSASNSLVMILNATDADEPNHLNSKIAFKIVSQEPAGTPM  159
             D NDNPP F + ++  E++EN       V+ + A D DE    +S+I ++I S    G
Sbjct  1443  DKNDNPPYFDKALYEAEVDENEDIQHTVLTVTAKDHDE----SSRIRYEITSGNIGGA--  1496

Query  160   FLLSRNTGEVRTLTNSLDREQASSYRLVVSGADKDGEGLSTQCECNIKVKDVNDNFPMFR  219
             F +   TG +  +  +LD E    Y L ++ +D   E  +T      I VKDVNDN P+F
Sbjct  1497  FAVKNMTGAI-YVAGALDYETRKRYELRLAASDNLKENYTTVV---IHVKDVNDNPPVFE  1552

Query  220   DSQYSARIEENI---LSSELLRFQVTDLDEEYTDNWLAVYFFTSG--NEGNWFEIQTDPR  274
               Y  +I E     L   +L+   TD D++   N   VYF T    +  N    + D
Sbjct  1553  RPTYRTQITEEDDRNLPKRVLQVTATDGDKDRPQN--IVYFLTGQGIDPDNPANSKFDIN  1610

Query  275   TNEGILKVVKALDYEQLQS-VKLSIAVKNKAEFHQSVISRYRVQSTPVTIQVINVREGIA  333
               G + V+K LD +Q     +    V  + E  + ++   VQ     + + ++ +
Sbjct  1611  RTTGEIFVLKPLDRDQPNGRPQWRFTVFAQDEGGEGLVGYADVQ-----VNLKDINDNAP  1665

Query  334   FRPASKTFTVQKGISSKKLVDYILGTYQAIDEDTNKAASNVK--YVMGRN------DGGY  385
               P    F     ++       ++ T  A+D D     +N K  Y + +N
Sbjct  1666  IFPQGVYF---GNVTENGTAGMVVMTMTAVDYDDPNEGTNAKLIYSIEKNVIEEETGSPI  1722

Query  386   LMIDSKTAEIK-FVKNMNRDSTFIVNKTITAEVLAIDEYTGKTSTGTVYVRVPDFNDNCP  444
              I++ T  IK  V  ++R+ T        + +V+A+D   G   TGT  +RV D ND  P
Sbjct  1723  FEIEADTGVIKTAVCCLDRERT----PDYSIQVVAMDG-GGLKGTGTASIRVKDINDMPP  1777

 Score = 75.9 bits (185),  Expect = 2e-12, Method: Compositional matrix adjust.
 Identities = 108/429 (25%), Positives = 178/429 (41%), Gaps = 53/429 (12%)

Query  53    VDKNTGDINITAIVDREETPSFLITCRALNAQGLDVEKPLILTVKILDINDNPPVFSQ-Q  111
             +D++  ++N+TA+ D          +C  +N          ++ V I D+NDN PVF
Sbjct  950   LDRDKYELNVTAMDDG--------SC-CVNGDATIHTSTAVVVVFITDVNDNKPVFKDCG  1000

Query  112   IFMGEIEENSASNSLVMILNATDADEPNHLNSKIAFKIVSQEPAGTPMFLLSRNTGEVRT  171
              +  ++EE + + S V+ + ATD D+    +N ++ + IV Q        F +   TGEV T
Sbjct  1001  TYYPKVEEGAPNGSPVIKVQATDEDK--GVNGQVKYSIVQQPNQKGTKFTVDEETGEVST  1058

Query  172   LTNSLDRE--QASSYRLVVSGADKDGEGLSTQCECNIKVKDVNDNFPMFRDSQYSARIEE  229
                  DRE          + V   D+     L   C    +++ DVNDN P+F    +Y   +++
Sbjct  1059  -NKVFDREGDDGKFVSVTVKATDQGEPSLEGVCSFTVEITDVNDNPPLFDRQKYVENVKQ  1117

Query  230   NI-LSSELLRFQVTDLDEEYTDNWLAVYFFT---SGNEGNWFEIQTDPRTNEGILKVVKA  285
             +  + + +LR  V+  DE+  +N    VY  +   S N+  +FEIQ  P +   +LK  K
Sbjct  1118  DASIGTNILR--VSASDEDADNNGAIVYTLSAPHSPNDLEYFEIQ--PESGWIVLK--KP  1171

Query  286   LD-YEQLQSVKLSIAVK----------NKAEFHQSVISRYRVQSTPV----TIQVINVRE  330
             LD     + SV+    +K             + A+    +     R    P+          I++RE
Sbjct  1172  LDGCYSVSSVRERYRLKVYASDRGDPPSTADVDVELDVVDRNNKPPIWDRNIYGPIHIRE  1231
```

```
Query  331   GIAFRPASKTFTVQKGISSKKLVDYILGTYQAIDEDTNKAASNVKYVMGRNDGGYLMIDS  390
              +             GI     V Y L         TNK   +  Y+  R D G
Sbjct  1232  NVTVGTVVTAIKASSGIEGNPTVFYRL--MPGSTAQTNKL--HTFYLQQRPDNG-----D  1282

Query  391   KTAEIKFVKNMNRDSTFIVNKTITAEVLAIDEYTGKTSTGTVYYVRVPDFNDNCPTAV-LE  449
               A+IK    ++ ++    N TI  E     +     S  TVY+++ D ND  P     E
Sbjct  1283  TWADIKVNHPLDFETIKEYNLTIRVENNGAQQL---ASEATVYIQLEDVNDEIPLFTERE  1339

Query  450   KDAVCSSSP  458
             ++ V    P
Sbjct  1340  QETVLEGEP  1348

 Score = 61.2 bits (147),  Expect = 5e-08, Method: Compositional matrix adjust.
 Identities = 87/366 (24%), Positives = 156/366 (43%), Gaps = 38/366 (10%)

Query  51    FVVDKNTGDINITAIVDRE-ETPSFL-ITCRALNAQGLDVEKPLILTVKILDINDNPPVF  108
             F VD+ TG+++   + DRE +   F+ +T +A +     +E     TV+I D+NDNPP+F
Sbjct  1047  FTVDEETGEVSTNKVFDREGDDGKFVSVTVKATDQGEPSLEGVCSFTVEITDVNDNPPLF  1106

Query  109   SQQIFMGEIEENSASNSLVMILNATDADEPNHLNSKIAFKIVS-QEPAGTPMFLLSRNTG  167
              +Q ++  ++++++  + ++ ++A+D D  N  N  I + + +   P     F +   +G
Sbjct  1107  DRQKYVENVKQDASIGTNILRVSASDEDADN--NGAIVYTLSAPHSPNDLEYFEIQPESG  1164

Query  168   EVRTLTNSLDREQASS-----YRLVVSGADKDGEGLSTQCECNIKVKDVNDNFPMFRDSQ  222
              +  L   LD   + S     YRL V  +D+ G+  ST            +N P   D
Sbjct  1165  WI-VLKKPLDGCYSVSSVRERYRLKVYASDR-GDPPSTADVDVELDVVDRNNKPPIWDRN  1222

Query  223   YSA--RIEENILSSELLRFQVTDLDEEYTDNWLAVYFFTSG-----NEGNWFEIQTDPRT  275
                    I EN+    ++              N    Y    G     N+ + F +Q  P
Sbjct  1223  IYGPIHIRENVTVGTVV--TAIKASSGIEGNPTVFYRLMPGSTAQTNKLHTFYLQQRPDN  1280

Query  276   NE--GILKVVKALDYEQLQSVKLSIAVKNKAEFHQSVISRYRVQSTPVTIQVINVREGIA  333
              +    +KV   LD+E ++   L+I V+N          ++       V IQ+ +V + I
Sbjct  1281  GDTWADIKVNHPLDFETIKEYNLTIRVENNG-------AQQLASEATVYIQLEDVNDEIP  1333

Query  334   FRPASKTFTVQKGISSKKLVDYILGTYQAIDEDTNKAASNVKYVM---GRNDG-GYLMID  389
                  +  TV +G      V  +     AID+D     + V Y +    RN+G  +  I+
Sbjct  1334  LFTEREQETVLEGEPIGTKVTQV----NAIDKDGTFPNNQVYYYIVDSPRNEGKDFFEIN  1389

Query  390   SKTAEI  395
              ++ EI
Sbjct  1390  LQSGEI  1395

 Score = 55.8 bits (133),  Expect = 3e-06, Method: Compositional matrix adjust.
 Identities = 49/158 (31%), Positives = 78/158 (49%), Gaps = 24/158 (15%)

Query  102   NDNPPVFSQQIFMGEIEENSASNSLVMILNATDADEPNHLNSKIAFKIVSQEPAGTP--M  159
             ND  P FSQQ++   ++EN+  N+LV  + A+D D  N     + F  V     GT
Sbjct  880   NDEEPKFSQQVYTPNVDENAGPNTLVTTVVASDKDGDN-----VRFGFVG---GGTSSGQ  931

Query  160   FLLSRNTGEVRTLTN--SLDREQASSYRLVVSGADK-------DGEGLSTQCECNIKVKD  210
             F++   TG +R  +   SLDR++   Y L V+  D        D    ++      + + D
Sbjct  932   FVIEEITGVIRLHSKAISLDRDK---YELNVTAMDDGSCCVNGDATIHTSTAVVVVFITD  988

Query  211   VNDNFPMFRD-SQYSARIEENILS-SELLRFQVTDLDE  246
             VNDN P+F+D   Y  ++EE   + S +++ Q TD D+
Sbjct  989   VNDNKPVFKDCGTYYPKVEEGAPNGSPVIKVQATDEDK  1026

 Score = 53.9 bits (128),  Expect = 1e-05, Method: Compositional matrix adjust.
 Identities = 70/306 (23%), Positives = 126/306 (41%), Gaps = 30/306 (10%)

Query  20    PIAKIT-SDYQATQKITYRI---SGVGIDQPPFGIFVVDKNTGDINITAIVDRE---ETP  72
             PI  +T  D   T K  Y++   SG G D+  F +   +  TG + I   +D E   ++
Sbjct  1800  PILTVTVHDEDETNKFQYKVIDNSGYGADK--FTMVRNNDGTGSLKIVQPLDYEDPLQSN  1857

Query  73    SFLITCRALNAQGLDVEKPLI------LTVKILDINDNPPVFSQQIFMGEIEENSASNSL  126
              F    + +N +G D +          + VK+ DINDN P F +      + EN+
```

```
Sbjct  1858  GFRFRIQ-VNDKGEDNDNDKYHVAYSWVVVKLRDINDNKPQFERPNIEVSVYENADVGKT  1916

Query  127   VMILNATDADEPNHLNSKIAFKIVSQEPAGTPMFLLSRNTGEVRTLTNSLDREQASSYRL  186
             +    ATD D+      SK+++ I    + +        S N      T+   +LDRE     +++
Sbjct  1917  LETFKATDPDQGG--KSKVSYAI---DRSSDRQRQFSINQEGTVTIQRNLDREVTPRHQV  1971

Query  187   VVSGADKDGEGLSTQCECNIKVKDVNDNFPMFRDSQYSARIEENILSSELLRFQVTDLDE  246
              +   D        +       + V+D+NDN P F     Y    + E++    +++     TD D+
Sbjct  1972  KILAIDDGIPPKTATATLTVIVQDINDNPPKFL-KDYRPVLPEHVPPRKVVEILATDDDD  2030

Query  247   EYTDNWLAVYFFTSGNEGNW----FEIQTDPRTNEG----ILKVVKALDYEQLQSVKLSI  298
                 N     F        +      F+++ D +    G     I+  +++ D EQ +    + I
Sbjct  2031  RSKSNGPPFQFRLDPGADDIIRASFKVEQDQKGANGDGMAIVSSLRSFDREQQKEYLIPI  2090

Query  299   AVKNKA  304
              +K+
Sbjct  2091  VIKDHG  2096

 Score = 51.6 bits (122),  Expect = 5e-05, Method: Compositional matrix adjust.
 Identities = 96/398 (24%), Positives = 163/398 (41%), Gaps = 54/398 (14%)

Query  106   PVFSQQIFMGEIEENSASNSLVMILNATDADEPNHLNSKIAFKIVSQ-EPAGTPMFLLSR  164
             P F    +  EI EN   +S ++ + A       +  + +I + + +Q + AGT  F +
Sbjct  677   PQFYMPSYEAEIPENQKKDSDIISVKAK-----SFADREIRYTLKAQGQGAGT--FNIGP  729

Query  165   NTGEVRTLTNSLDRE---QASSYRLVVSGADKDGEGLSTQCECNIKVKDVNDNFPMFRDS  221
              +G V+ L   LD E   Q   Y LVV+ A +D  G ST  E  I+V DVNDN P F
Sbjct  730   TSGIVK-LAKELDFEDLRQPHVYSLVVT-ATEDSGGFSTSVELTIRVTDVNDNAPKFELP  787

Query  222   QYSAR-IEENI-LSSELLRFQVTDLDEEYTDNWLAVYFFTSGNEGNWFEIQTDPRTNEGI  279
              Y A  ++E+I + + +L+ +  D D          + +  S +    F + ++     GI
Sbjct  788   DYQAHNVDEDIQIGTSILKVKAMDADSGAN---AEIEYLVSDDH---FAVDSN-----GI  836

Query  280   LKVVKALDYEQLQSV-KLSIAVKNKAEFHQSVISRYRVQSTPVTIQVINVREGIAFRPAS  338
             +   K LD +   +  +  +  K+K E  ++  +  RV +            E   F
Sbjct  837   IVNNKQLDADNNNAYYEFVVTAKDKGEPAKTGTATVRVYTKNKN------DEEPKFSQQV  890

Query  339   KTFTVQKGISSKKLVDYILGTYQAIDEDTNKAASNVKY--VMGRNDGGYLMIDSKTAEIK  396
              T  V +       LV  ++ +          +K   NV++  V G    G  +I+  T  I+
Sbjct  891   YTPNVDENAGPNTLVTTVVAS--------DKDGDNVRFGFVGGGTSSGQFVIEEITGVIR  942

Query  397   FVK---NMNRDSTFI-VNKTITAEVLAIDEYTGKTSTGTVYVRVPDFNDN------CPTA  446
                   +++RD   + V              + T  TST  V V + D NDN      C T
Sbjct  943   LHSKAISLDRDKYELNVTAMDDGSCCVNGDATIHTSTAVVVVFITDVNDNKPVFKDCGTY  1002

Query  447   VLE-KDAVCSSSPSVVVSARTLNNRYTGPYTFALEDQP  483
               + ++    + SP + V A    +    G    +++   QP
Sbjct  1003  YPKVEEGAPNGSPVIKVQATDEDKGVNGQVKYSIVQQP  1040

 Score = 44.7 bits (104),  Expect = 0.008, Method: Compositional matrix adjust.
 Identities = 48/199 (24%), Positives = 73/199 (37%), Gaps = 19/199 (10%)

Query  27    DYQATQKITYRISGVGIDQPPFGIFVVDKNTGDINITAIVDREETPSFLITCRALNAQGL  86
             D     K++Y I       Q  F I        G + I   +DRE TP    +   A++
Sbjct  1926  DQGGKSKVSYAIDRSSDRQRQFSI----NQEGTVTIQRNLDREVTPRHQVKILAIDDGIP  1981

Query  87    DVEKPLILTVKILDINDNPPVFSQQIFMGEIEENSASNSLVMILNATDADEPNHLNSKIA  146
                    LTV + DINDNPP F +  +    + E+      +V IL    D D
Sbjct  1982  PKTATATLTVIVQDINDNPPKFLKD-YRPVLPEHVPPRKVVEILATDDDDRSKSNGPPFQ  2040

Query  147   FKIVSQEPAGTPMFLLSRNTGEVRTLTN-----------SLDREQASSYRLVVSGADKDG  195
             F++    +P     +   S     + +    N             S DREQ    Y + +     D
Sbjct  2041  FRL---DPGADDIIRASFKVEQDQKGANGDGMAIVSSLRSFDREQQKEYLIPIVIKDHGN  2097

Query  196   EGLSTQCECNIKVKDVNDN  214
               ++       + + DVNDN
Sbjct  2098  PAMTGTSTLTVVIGDVNDN  2116
```

```
 Score = 40.8 bits (94),  Expect = 0.11, Method: Compositional matrix adjust.
 Identities = 55/242 (23%), Positives = 102/242 (42%), Gaps = 22/242 (9%)

Query  4    KFAKPCREGE--DNSKRNP-IAKITSDYQATQKITYRISGVGIDQPPFGIFVVDKNTGDI  60
            +F  P  E E  +N K++  I  + +   A ++I Y +   G      G F +   +G +
Sbjct  678  QFYMPSYEAEIPENQKKDSDIISVKAKSFADREIRYTLKAQGQGA---GTFNIGPTSGIV  734

Query  61   NITAIVDREE--TPSF--LITCRALNAQGLDVEKPLILTVKILDINDNPPVFSQQIFMGE  116
             +   +D E+   P    L+     ++ G       + LT+++ D+NDN P F    +
Sbjct  735  KLAKELDFEDLRQPHVYSLVVTATEDSGGFSTS--VELTIRVTDVNDNAPKFELPDYQAH  792

Query  117  -IEENSASNSLVMILNATDADEPNHLNSKIAFKIVSQEPAGTPMFLLSRNTGEVRTLTNS  175
             ++E+    + ++ + A DAD  +  N++I + +VS +      F +  N   V
Sbjct  793  NVDEDIQIGTSILKVKAMDAD--SGANAEIEY-LVSDD-----HFAVDSNGIIVNNKQLD  844

Query  176  LDREQASSYRLVVSGADKDGEGLSTQCECNIKVKDVNDNFPMFRDSQYSARIEENILSSE  235
             D   A  Y  VV+  DK     +      +  K+ ND  P F    Y+  ++EN   +
Sbjct  845  ADNNNA-YYEFVVTAKDKGEPAKTGTATVRVYTKNKNDEEPKFSQQVYTPNVDENAGPNT  903

Query  236  LL  237
            L+
Sbjct  904  LV  905

 Score = 39.3 bits (90),  Expect = 0.30, Method: Compositional matrix adjust.
 Identities = 55/225 (24%), Positives = 88/225 (39%), Gaps = 24/225 (11%)

Query  75   LITCRALNAQGLDVEKPLILTVKILDINDNPPVFSQQIF--MGEIEENSASNSLVMILNA  132
            +IT +  NA GL       + + + D+ND PP F  +       ++ N+  N+ V  L A
Sbjct  539  IITNQGHNA-GLKYTDNQRVIIHVKDVNDEPPYFINRPLPMQAVVQLNAPPNTPVFTLQA  597

Query  133  TDADEPNHLNSKIAFKIVSQEPAGTPMFLLSRNTGEVRTLTNSLDREQASSYRLVVSGAD  192
             D D     +  I + IV     G   F +   +G VRT    L  +    Y L V   D
Sbjct  598  RDPD----TDHNIHYFIVRDRTGG--RFEVDERSGVVRTRGTDL-FQLDMEYVLYVKAED  650

Query  193  KDGE----GLSTQCECNIKVKDVNDNFPMFRDSQYSARIEEN-ILSSELLRFQVTDLDEE  247
            ++G+        +  E  + +        P F    Y A I EN    S+++  +     +
Sbjct  651  QNGKVDDRRFQSTPEERLSIVG-GKRAPQFYMPSYEAEIPENQKKDSDIISVKA----KS  705

Query  248  YTDNWLAVYFFTSGNEGNWFEIQTDPRTNEGILKVVKALDYEQLQ  292
            + D  +       G     F I         GI+K+ K LD+E L+
Sbjct  706  FADREIRYTLKAQGQGAGTFNIG----PTSGIVKLAKELDFEDLR  746

>AALB20_033171.P66518 | transcript=AALB20_033171.R66518 | gene=AALB20_033171
| organism=Anopheles_albimanus_STECLA_2020 |
gene_product=neural-cadherin, transcript variant X38 |
transcript_product=neural-cadherin,
transcript variant X21 | location=CM024292:75037560-75208393(+)
| protein_length=3131 | sequence_SO=chromosome
| SO=protein_coding_gene | is_pseudo=false
Length=3131

 Score = 144 bits (364),  Expect = 1e-34, Method: Compositional matrix adjust.
 Identities = 125/437 (29%), Positives = 211/437 (48%), Gaps = 33/437 (8%)

Query  26    SDYQATQKITYRISGVGID--QPPFGIFVVDKNTGDINITAIVDREET---PSFLITCRA  80
              D    Q I Y ++G GID   P    F +++ TG+I +    +DR++     P +  T  A
Sbjct  1580  GDKDRPQNIVYFLTGQGIDPDNPANSKFDINRTTGEIFVLKPLDRDQPNGRPQWRFTVFA  1639

Query  81    LNAQGLDVEKPLILTVKILDINDNPPVFSQQIFMGEIEENSASNSLVMILNATDADEPNH  140
              +  G  +      + V + DINDN P+F Q ++ G + EN  +  +VM + A D D+PN
Sbjct  1640  QDEGGEGLVGYADVQVNLKDINDNAPIFPQGVYFGNVTENGTAGMVVMTMTAVDYDDPNE  1699

Query  141   -LNSKIAFKI---VSQEPAGTPMFLLSRNTGEVRTLTNSLDREQASSYRLVVSGADKDGE  196
                N+K+ + I    V +E  G+P+F +  +TG ++T    LDRE+   Y + V      DG
Sbjct  1700  GTNAKLIYSIEKNVIEEETGSPIFEIEADTGVIKTAVCCLDRERTPDYSIQVVAM--DGG  1757

Query  197   GLSTQCECNIKVKDVNDNFPMFRDSQYSARIEENILSS--ELLRFQVTDLDEEYTDNWLA  254
            GL        +I+VKD+ND  P F   ++   ++E   ++  E+    VT  DE+ T+ +
Sbjct  1758  GLKGTGTASIRVKDINDMPPQFTKDEWFTEVDETDGTNLPEMPILTVTVHDEDETNKFQY  1817
```

```
Query  255   VYFFTSGNEGNWFEIQTDPRTNEGI--LKVVKALDYE---QLQSVKLSIAVKNKAEFHQS  309
                  SG   + F +    R N+G   LK+V+ LDYE   Q    +  I V +K E + +
Sbjct  1818  KVIDNSGYGADKFTMV---RNNDGTGSLKIVQPLDYEDPLQSNGFRFRIQVNDKGEDNDN  1874

Query  310   VISRYRVQSTPVTIQVINVREGIAFRPASKTFTVQKGISSKKLVDYILGTYQAIDEDTNK  369
                +Y V  + V +++ ++ +    +P  +   ++  +     V   L T++A D D
Sbjct  1875  --DKYHVAYSWVVVKLRDINDN---KPQFERPNIEVSVYENADVGKTLETFKATDPDQG-  1928

Query  370   AASNVKYVMGR-NDGGYLMIDSKTAEIKFVKNMNRDSTFIVNKTITAEVLAIDE-YTGKT  427
               S V Y + R +D       ++   +   +N++R+    V      ++LAID+    KT
Sbjct  1929  GKSKVSYAIDRSSDRQRQFSINQEGTVTIQRNLDRE----VTPRHQVKILAIDDGIPPKT  1984

Query  428   STGTVYVRVPDFNDNCP  444
             +T T+ V V D NDN P
Sbjct  1985  ATATLTVIVQDINDNPP  2001

 Score = 85.9 bits (211),  Expect = 1e-15, Method: Compositional matrix adjust.
 Identities = 108/420 (26%), Positives = 177/420 (42%), Gaps = 52/420 (12%)

Query  50    IFVVDKNTGDINITAIVDREETPSFLITCRAL----------NAQGLDVEKPLILTVKIL  99
              F ++  +G+I    + DRE+  ++ +   A           N Q   V K   + + I
Sbjct  1385  FFEINLQSGEIFTKVVFDREKQGAYALEVEARDGAPSARPNSNQQPNSVTK--FIRIGIA  1442

Query  100   DINDNPPVFSQQIFMGEIEENSASNSLVMILNATDADEPNHLNSKIAFKIVSQEPAGTPM  159
             D NDNPP F + ++  E++EN      V+ + A D DE    +S+I ++I S    G
Sbjct  1443  DKNDNPPYFDKALYEAEVDENEDIQHTVLTVTAKDHDE----SSRIRYEITSGNIGGA--  1496

Query  160   FLLSRNTGEVRTLTNSLDREQASSYRLVVSGADKDGEGLSTQCECNIKVKDVNDNFPMFR  219
             F +   TG +  +  +LD E    Y L ++ +D   E  +T     I VKDVNDN P+F
Sbjct  1497  FAVKNMTGAI-YVAGALDYETRKRYELRLAASDNLKENYTTVV---IHVKDVNDNPPVFE  1552

Query  220   DSQYSARIEENI---LSSELLRFQVTDLDEEYTDNWLAVYFFTSG--NEGNWFEIQTDPR  274
                Y  +I E     L   +L+   TD D++   N   VYF T    +  N    + D
Sbjct  1553  RPTYRTQITEEDDRNLPKRVLQVTATDGDKDRPQN--IVYFLTGQGIDPDNPANSKFDIN  1610

Query  275   TNEGILKVVKALDYEQLQS-VKLSIAVKNKAEFHQSVISRYRVQSTPVTIQVINVREGIA  333
                G + V+K LD +Q     +    V  + E  + ++    VQ     + + ++ +
Sbjct  1611  RTTGEIFVLKPLDRDQPNGRPQWRFTVFAQDEGGEGLVGYADVQ-----VNLKDINDNAP  1665

Query  334   FRPASKTFTVQKGISSKKLVDYILGTYQAIDEDTNKAASNVK--YVMGRN------DGGY  385
               P    F     ++       ++ T  A+D D     +N K  Y + +N
Sbjct  1666  IFPQGVYF---GNVTENGTAGMVVMTMTAVDYDDPNEGTNAKLIYSIEKNVIEEETGSPI  1722

Query  386   LMIDSKTAEIK-FVKNMNRDSTFIVNKTITAEVLAIDEYTGKTSTGTVYVRVPDFNDNCP  444
               I++ T  IK  V  ++R+ T       + +V+A+D   G   TGT  +RV D ND  P
Sbjct  1723  FEIEADTGVIKTAVCCLDRERT----PDYSIQVVAMDG-GGLKGTGTASIRVKDINDMPP  1777

 Score = 75.9 bits (185),  Expect = 2e-12, Method: Compositional matrix adjust.
 Identities = 108/429 (25%), Positives = 178/429 (41%), Gaps = 53/429 (12%)

Query  53    VDKNTGDINITAIVDREETPSFLITCRALNAQGLDVEKPLILTVKILDINDNPPVFSQ-Q  111
             +D++  ++N+TA+ D         +C  +N          ++ V I D+NDN PVF
Sbjct  950   LDRDKYELNVTAMDDG--------SC-CVNGDATIHTSTAVVVVFITDVNDNKPVFKDCG  1000

Query  112   IFMGEIEENSASNSLVMILNATDADEPNHLNSKIAFKIVSQEPAGTPMFLLSRNTGEVRT  171
              +  ++EE + + S V+ + ATD D+   +N ++ + IV Q       F +   TGEV T
Sbjct  1001  TYYPKVEEGAPNGSPVIKVQATDEDK--GVNGQVKYSIVQQPNQKGTKFTVDEETGEVST  1058

Query  172   LTNSLDRE--QASSYRLVVSGADKDGEGLSTQCECNIKVKDVNDNFPMFRDSQYSARIEE  229
                  DRE        + V   D+    L   C   +++ DVNDN P+F   +Y   +++
Sbjct  1059  -NKVFDREGDDGKFVSVTVKATDQGEPSLEGVCSFTVEITDVNDNPPLFDRQKYVENVKQ  1117

Query  230   NI-LSSELLRFQVTDLDEEYTDNWLAVYFFT---SGNEGNWFEIQTDPRTNEGILKVVKA  285
             +  + + +LR  V+  DE+  +N   VY  +   S N+  +FEIQ  P +   +LK  K
Sbjct  1118  DASIGTNILR--VSASDEDADNNGAIVYTLSAPHSPNDLEYFEIQ--PESGWIVLK--KP  1171

Query  286   LD-YEQLQSVKLSIAVK----------NKAEFHQSVISRYRVQSTPV----TIQVINVRE  330
             LD    + SV+    +K          + A+    +    R    P+        I++RE
```

```
Sbjct  1172  LDGCYSVSSVRERYRLKVYASDRGDPPSTADVDVELDVVDRNNKPPIWDRNIYGPIHIRE  1231

Query  331   GIAFRPASKTFTVQKGISSKKLVDYILGTYQAIDEDTNKAASNVKYVMGRNDGGYLMIDS  390
              +             GI     V Y L         TNK   +  Y+  R D G
Sbjct  1232  NVTVGTVVTAIKASSGIEGNPTVFYRL--MPGSTAQTNKL--HTFYLQQRPDNG-----D  1282

Query  391   KTAEIKFVKNMNRDSTFIVNKTITAEVLAIDEYTGKTSTGTVYVRVPDFNDNCPTAV-LE  449
               A+IK    ++ ++    N TI  E     +     S  TVY+++ D ND   P     E
Sbjct  1283  TWADIKVNHPLDFETIKEYNLTIRVENNGAQQL---ASEATVYIQLEDVNDEIPLFTERE  1339

Query  450   KDAVCSSSP  458
             ++ V    P
Sbjct  1340  QETVLEGEP  1348

 Score = 61.2 bits (147),  Expect = 5e-08, Method: Compositional matrix adjust.
 Identities = 87/366 (24%), Positives = 156/366 (43%), Gaps = 38/366 (10%)

Query  51    FVVDKNTGDINITAIVDRE-ETPSFL-ITCRALNAQGLDVEKPLILTVKILDINDNPPVF  108
             F VD+ TG+++   + DRE +   F+ +T +A +     +E     TV+I D+NDNPP+F
Sbjct  1047  FTVDEETGEVSTNKVFDREGDDGKFVSVTVKATDQGEPSLEGVCSFTVEITDVNDNPPLF  1106

Query  109   SQQIFMGEIEENSASNSLVMILNATDADEPNHLNSKIAFKIVS-QEPAGTPMFLLSRNTG  167
              +Q ++  ++++++  + ++ ++A+D D  N  N  I + + +   P     F +   +G
Sbjct  1107  DRQKYVENVKQDASIGTNILRVSASDEDADN--NGAIVYTLSAPHSPNDLEYFEIQPESG  1164

Query  168   EVRTLTNSLDREQASS-----YRLVVSGADKDGEGLSTQCECNIKVKDVNDNFPMFRDSQ  222
              +  L   LD   + S     YRL V  +D+ G+  ST            +N P   D
Sbjct  1165  WI-VLKKPLDGCYSVSSVRERYRLKVYASDR-GDPPSTADVDVELDVVDRNNKPPIWDRN  1222

Query  223   YSA--RIEENILSSELLRFQVTDLDEEYTDNWLAVYFFTSG-----NEGNWFEIQTDPRT  275
                   I EN+    ++             N    Y    G     N+ + F +Q  P
Sbjct  1223  IYGPIHIRENVTVGTVV--TAIKASSGIEGNPTVFYRLMPGSTAQTNKLHTFYLQQRPDN  1280

Query  276   NE--GILKVVKALDYEQLQSVKLSIAVKNKAEFHQSVISRYRVQSTPVTIQVINVREGIA  333
              +    +KV   LD+E ++   L+I V+N         ++       V IQ+ +V + I
Sbjct  1281  GDTWADIKVNHPLDFETIKEYNLTIRVENNG-------AQQLASEATVYIQLEDVNDEIP  1333

Query  334   FRPASKTFTVQKGISSKKLVDYILGTYQAIDEDTNKAASNVKYVM---GRNDG-GYLMID  389
                  +  TV +G      V  +     AID+D     + V Y +    RN+G  +  I+
Sbjct  1334  LFTEREQETVLEGEPIGTKVTQV----NAIDKDGTFPNNQVYYYIVDSPRNEGKDFFEIN  1389

Query  390   SKTAEI  395
              ++ EI
Sbjct  1390  LQSGEI  1395

 Score = 55.8 bits (133),  Expect = 3e-06, Method: Compositional matrix adjust.
 Identities = 49/158 (31%), Positives = 78/158 (49%), Gaps = 24/158 (15%)

Query  102   NDNPPVFSQQIFMGEIEENSASNSLVMILNATDADEPNHLNSKIAFKIVSQEPAGTP--M  159
             ND  P FSQQ++   ++EN+  N+LV  + A+D D  N     + F  V     GT
Sbjct  880   NDEEPKFSQQVYTPNVDENAGPNTLVTTVVASDKDGDN-----VRFGFVG---GGTSSGQ  931

Query  160   FLLSRNTGEVRTLTN--SLDREQASSYRLVVSGADK-------DGEGLSTQCECNIKVKD  210
             F++   TG +R  +   SLDR++   Y L V+  D        D    ++     + + D
Sbjct  932   FVIEEITGVIRLHSKAISLDRDK---YELNVTAMDDGSCCVNGDATIHTSTAVVVVFITD  988

Query  211   VNDNFPMFRD-SQYSARIEENILS-SELLRFQVTDLDE  246
             VNDN P+F+D   Y  ++EE   + S +++ Q TD D+
Sbjct  989   VNDNKPVFKDCGTYYPKVEEGAPNGSPVIKVQATDEDK  1026

 Score = 53.9 bits (128),  Expect = 1e-05, Method: Compositional matrix adjust.
 Identities = 70/306 (23%), Positives = 126/306 (41%), Gaps = 30/306 (10%)

Query  20    PIAKIT-SDYQATQKITYRI---SGVGIDQPPFGIFVVDKNTGDINITAIVDRE---ETP  72
             PI  +T  D   T K  Y++   SG G D+  F +   +  TG + I   +D E   ++
Sbjct  1800  PILTVTVHDEDETNKFQYKVIDNSGYGADK--FTMVRNNDGTGSLKIVQPLDYEDPLQSN  1857

Query  73    SFLITCRALNAQGLDVEKPLI------LTVKILDINDNPPVFSQQIFMGEIEENSASNSL  126
```

```
              F    + +N +G D +          + VK+ DINDN P F +      + EN+
Sbjct  1858  GFRFRIQ-VNDKGEDNDNDKYHVAYSWVVVKLRDINDNKPQFERPNIEVSVYENADVGKT  1916

Query  127   VMILNATDADEPNHLNSKIAFKIVSQEPAGTPMFLLSRNTGEVRTLTNSLDREQASSYRL  186
             +    ATD D+     SK+++ I   + +       S N     T+  +LDRE    +++
Sbjct  1917  LETFKATDPDQGG--KSKVSYAI---DRSSDRQRQFSINQEGTVTIQRNLDREVTPRHQV  1971

Query  187   VVSGADKDGEGLSTQCECNIKVKDVNDNFPMFRDSQYSARIEENILSSELLRFQVTDLDE  246
              +   D      +      + V+D+NDN P F    Y   + E++   +++    TD D+
Sbjct  1972  KILAIDDGIPPKTATATLTVIVQDINDNPPKFL-KDYRPVLPEHVPPRKVVEILATDDDD  2030

Query  247   EYTDNWLAVYFFTSGNEGNW----FEIQTDPRTNEG----ILKVVKALDYEQLQSVKLSI  298
                 N     F      +      F+++ D +   G    I+  +++ D EQ +   + I
Sbjct  2031  RSKSNGPPFQFRLDPGADDIIRASFKVEQDQKGANGDGMAIVSSLRSFDREQQKEYLIPI  2090

Query  299   AVKNKA  304
              +K+
Sbjct  2091  VIKDHG  2096

 Score = 51.6 bits (122),  Expect = 5e-05, Method: Compositional matrix adjust.
 Identities = 96/398 (24%), Positives = 163/398 (41%), Gaps = 54/398 (14%)

Query  106   PVFSQQIFMGEIEENSASNSLVMILNATDADEPNHLNSKIAFKIVSQ-EPAGTPMFLLSR  164
             P F    +  EI EN   +S ++ + A      +  + +I + + +Q + AGT  F +
Sbjct  677   PQFYMPSYEAEIPENQKKDSDIISVKAK-----SFADREIRYTLKAQGQGAGT--FNIGP  729

Query  165   NTGEVRTLTNSLDRE---QASSYRLVVSGADKDGEGLSTQCECNIKVKDVNDNFPMFRDS  221
              +G V+ L   LD E   Q   Y LVV+ A +D  G ST  E  I+V DVNDN P F
Sbjct  730   TSGIVK-LAKELDFEDLRQPHVYSLVVT-ATEDSGGFSTSVELTIRVTDVNDNAPKFELP  787

Query  222   QYSAR-IEENI-LSSELLRFQVTDLDEEYTDNWLAVYFFTSGNEGNWFEIQTDPRTNEGI  279
              Y A  ++E+I + + +L+ +  D D         + +  S +    F + ++     GI
Sbjct  788   DYQAHNVDEDIQIGTSILKVKAMDADSGAN---AEIEYLVSDDH---FAVDSN-----GI  836

Query  280   LKVVKALDYEQLQSV-KLSIAVKNKAEFHQSVISRYRVQSTPVTIQVINVREGIAFRPAS  338
             +   K LD +   +  +  +  K+K E  ++  +  RV +           E   F
Sbjct  837   IVNNKQLDADNNNAYYEFVVTAKDKGEPAKTGTATVRVYTKNKN------DEEPKFSQQV  890

Query  339   KTFTVQKGISSKKLVDYILGTYQAIDEDTNKAASNVKY--VMGRNDGGYLMIDSKTAEIK  396
              T  V +      LV  ++ +        +K   NV++  V G    G  +I+  T  I+
Sbjct  891   YTPNVDENAGPNTLVTTVVAS--------DKDGDNVRFGFVGGGTSSGQFVIEEITGVIR  942

Query  397   FVK---NMNRDSTFI-VNKTITAEVLAIDEYTGKTSTGTVYVRVPDFNDN------CPTA  446
                   +++RD   + V            + T  TST  V V + D NDN      C T
Sbjct  943   LHSKAISLDRDKYELNVTAMDDGSCCVNGDATIHTSTAVVVVFITDVNDNKPVFKDCGTY  1002

Query  447   VLE-KDAVCSSSPSVVVSARTLNNRYTGPYTFALEDQP  483
               + ++   + SP + V A   +    G   +++  QP
Sbjct  1003  YPKVEEGAPNGSPVIKVQATDEDKGVNGQVKYSIVQQP  1040

 Score = 44.7 bits (104),  Expect = 0.008, Method: Compositional matrix adjust.
 Identities = 48/199 (24%), Positives = 73/199 (37%), Gaps = 19/199 (10%)

Query  27    DYQATQKITYRISGVGIDQPPFGIFVVDKNTGDINITAIVDREETPSFLITCRALNAQGL  86
             D     K++Y I      Q  F I       G + I   +DRE TP   +   A++
Sbjct  1926  DQGGKSKVSYAIDRSSDRQRQFSI----NQEGTVTIQRNLDREVTPRHQVKILAIDDGIP  1981

Query  87    DVEKPLILTVKILDINDNPPVFSQQIFMGEIEENSASNSLVMILNATDADEPNHLNSKIA  146
                    LTV + DINDNPP F +  +   + E+     +V IL   D D
Sbjct  1982  PKTATATLTVIVQDINDNPPKFLKD-YRPVLPEHVPPRKVVEILATDDDDRSKSNGPPFQ  2040

Query  147   FKIVSQEPAGTPMFLLSRNTGEVRTLTN-----------SLDREQASSYRLVVSGADKDG  195
             F++   +P    +   S    + +   N           S DREQ   Y + +   D
Sbjct  2041  FRL---DPGADDIIRASFKVEQDQKGANGDGMAIVSSLRSFDREQQKEYLIPIVIKDHGN  2097

Query  196   EGLSTQCECNIKVKDVNDN  214
               ++      + + DVNDN
Sbjct  2098  PAMTGTSTLTVVIGDVNDN  2116
```

```
 Score = 40.8 bits (94),  Expect = 0.11, Method: Compositional matrix adjust.
 Identities = 55/242 (23%), Positives = 102/242 (42%), Gaps = 22/242 (9%)

Query  4    KFAKPCREGE--DNSKRNP-IAKITSDYQATQKITYRISGVGIDQPPFGIFVVDKNTGDI  60
            +F  P  E E  +N K++  I  + +   A ++I Y +   G      G F +   +G +
Sbjct  678  QFYMPSYEAEIPENQKKDSDIISVKAKSFADREIRYTLKAQGQGA---GTFNIGPTSGIV  734

Query  61   NITAIVDREE--TPSF--LITCRALNAQGLDVEKPLILTVKILDINDNPPVFSQQIFMGE  116
             +   +D E+   P    L+     ++ G      + LT+++ D+NDN P F    +
Sbjct  735  KLAKELDFEDLRQPHVYSLVVTATEDSGGFSTS--VELTIRVTDVNDNAPKFELPDYQAH  792

Query  117  -IEENSASNSLVMILNATDADEPNHLNSKIAFKIVSQEPAGTPMFLLSRNTGEVRTLTNS  175
             ++E+    + ++ + A DAD  +  N++I + +VS +      F +  N   V
Sbjct  793  NVDEDIQIGTSILKVKAMDAD--SGANAEIEY-LVSDD-----HFAVDSNGIIVNNKQLD  844

Query  176  LDREQASSYRLVVSGADKDGEGLSTQCECNIKVKDVNDNFPMFRDSQYSARIEENILSSE  235
             D   A  Y  VV+  DK     +      +  K+ ND  P F    Y+  ++EN   +
Sbjct  845  ADNNNA-YYEFVVTAKDKGEPAKTGTATVRVYTKNKNDEEPKFSQQVYTPNVDENAGPNT  903

Query  236  LL  237
            L+
Sbjct  904  LV  905

 Score = 39.3 bits (90),  Expect = 0.30, Method: Compositional matrix adjust.
 Identities = 55/225 (24%), Positives = 88/225 (39%), Gaps = 24/225 (11%)

Query  75   LITCRALNAQGLDVEKPLILTVKILDINDNPPVFSQQIF--MGEIEENSASNSLVMILNA  132
            +IT +  NA GL       + + + D+ND PP F  +        ++ N+  N+ V  L A
Sbjct  539  IITNQGHNA-GLKYTDNQRVIIHVKDVNDEPPYFINRPLPMQAVVQLNAPPNTPVFTLQA  597

Query  133  TDADEPNHLNSKIAFKIVSQEPAGTPMFLLSRNTGEVRTLTNSLDREQASSYRLVVSGAD  192
             D D     +  I + IV     G   F +   +G VRT    L  +    Y L V   D
Sbjct  598  RDPD----TDHNIHYFIVRDRTGG--RFEVDERSGVVRTRGTDL-FQLDMEYVLYVKAED  650

Query  193  KDGE----GLSTQCECNIKVKDVNDNFPMFRDSQYSARIEEN-ILSSELLRFQVTDLDEE  247
            ++G+        +  E  + +       P F    Y A I EN    S+++  +     +
Sbjct  651  QNGKVDDRRFQSTPEERLSIVG-GKRAPQFYMPSYEAEIPENQKKDSDIISVKA----KS  705

Query  248  YTDNWLAVYFFTSGNEGNWFEIQTDPRTNEGILKVVKALDYEQLQ  292
            + D  +       G     F I        GI+K+ K LD+E L+
Sbjct  706  FADREIRYTLKAQGQGAGTFNIG----PTSGIVKLAKELDFEDLR  746

>AALB20_033171.P66515 | transcript=AALB20_033171.R66515 | gene=AALB20_033171
| organism=Anopheles_albimanus_STECLA_2020 |
gene_product=neural-cadherin, transcript variant X38 |
transcript_product=neural-cadherin,
transcript variant X4 | location=CM024292:75037560-75208393(+)
| protein_length=3131 | sequence_SO=chromosome
| SO=protein_coding_gene | is_pseudo=false
Length=3131

 Score = 144 bits (364),  Expect = 1e-34, Method: Compositional matrix adjust.
 Identities = 125/437 (29%), Positives = 211/437 (48%), Gaps = 33/437 (8%)

Query  26    SDYQATQKITYRISGVGID--QPPFGIFVVDKNTGDINITAIVDREET---PSFLITCRA  80
              D    Q I Y ++G GID   P    F +++ TG+I +   +DR++     P +  T  A
Sbjct  1580  GDKDRPQNIVYFLTGQGIDPDNPANSKFDINRTTGEIFVLKPLDRDQPNGRPQWRFTVFA  1639

Query  81    LNAQGLDVEKPLILTVKILDINDNPPVFSQQIFMGEIEENSASNSLVMILNATDADEPNH  140
              +  G  +      + V + DINDN P+F Q ++ G + EN  +  +VM + A D D+PN
Sbjct  1640  QDEGGEGLVGYADVQVNLKDINDNAPIFPQGVYFGNVTENGTAGMVVMTMTAVDYDDPNE  1699

Query  141   -LNSKIAFKI---VSQEPAGTPMFLLSRNTGEVRTLTNSLDREQASSYRLVVSGADKDGE  196
                N+K+ + I   V +E  G+P+F +  +TG ++T    LDRE+   Y + V      DG
Sbjct  1700  GTNAKLIYSIEKNVIEEETGSPIFEIEADTGVIKTAVCCLDRERTPDYSIQVVAM--DGG  1757

Query  197   GLSTQCECNIKVKDVNDNFPMFRDSQYSARIEENILSS--ELLRFQVTDLDEEYTDNWLA  254
              GL       +I+VKD+ND  P F   ++    ++E   ++  E+     VT  DE+ T+ +
```

```
Sbjct  1758  GLKGTGTASIRVKDINDMPPQFTKDEWFTEVDETDGTNLPEMPILTVTVHDEDETNKFQY  1817

Query  255   VYFFTSGNEGNWFEIQTDPRTNEGI--LKVVKALDYE---QLQSVKLSIAVKNKAEFHQS  309
                  SG   + F +    R N+G   LK+V+ LDYE   Q    +  I V +K E + +
Sbjct  1818  KVIDNSGYGADKFTMV---RNNDGTGSLKIVQPLDYEDPLQSNGFRFRIQVNDKGEDNDN  1874

Query  310   VISRYRVQSTPVTIQVINVREGIAFRPASKTFTVQKGISSKKLVDYILGTYQAIDEDTNK  369
                +Y V  + V +++ ++ +    +P  +   ++  +     V   L T++A D D
Sbjct  1875  --DKYHVAYSWVVVKLRDINDN---KPQFERPNIEVSVYENADVGKTLETFKATDPDQG-  1928

Query  370   AASNVKYVMGR-NDGGYLMIDSKTAEIKFVKNMNRDSTFIVNKTITAEVLAIDE-YTGKT  427
               S V Y + R +D       ++   +   +N++R+    V      ++LAID+    KT
Sbjct  1929  GKSKVSYAIDRSSDRQRQFSINQEGTVTIQRNLDRE----VTPRHQVKILAIDDGIPPKT  1984

Query  428   STGTVYVRVPDFNDNCP  444
              +T T+ V V D NDN P
Sbjct  1985  ATATLTVIVQDINDNPP  2001

 Score = 85.9 bits (211),  Expect = 1e-15, Method: Compositional matrix adjust.
 Identities = 108/420 (26%), Positives = 177/420 (42%), Gaps = 52/420 (12%)

Query  50    IFVVDKNTGDINITAIVDREETPSFLITCRAL----------NAQGLDVEKPLILTVKIL  99
              F ++  +G+I    + DRE+  ++ +   A           N Q   V K   + + I
Sbjct  1385  FFEINLQSGEIFTKVVFDREKQGAYALEVEARDGAPSARPNSNQQPNSVTK--FIRIGIA  1442

Query  100   DINDNPPVFSQQIFMGEIEENSASNSLVMILNATDADEPNHLNSKIAFKIVSQEPAGTPM  159
              D NDNPP F + ++  E++EN      V+ + A D DE    +S+I ++I S    G
Sbjct  1443  DKNDNPPYFDKALYEAEVDENEDIQHTVLTVTAKDHDE----SSRIRYEITSGNIGGA--  1496

Query  160   FLLSRNTGEVRTLTNSLDREQASSYRLVVSGADKDGEGLSTQCECNIKVKDVNDNFPMFR  219
              F +   TG +  +  +LD E    Y L ++ +D   E  +T     I VKDVNDN P+F
Sbjct  1497  FAVKNMTGAI-YVAGALDYETRKRYELRLAASDNLKENYTTVV---IHVKDVNDNPPVFE  1552

Query  220   DSQYSARIEENI---LSSELLRFQVTDLDEEYTDNWLAVYFFTSG--NEGNWFEIQTDPR  274
                 Y  +I E     L   +L+   TD D++   N   VYF T    +  N    + D
Sbjct  1553  RPTYRTQITEEDDRNLPKRVLQVTATDGDKDRPQN--IVYFLTGQGIDPDNPANSKFDIN  1610

Query  275   TNEGILKVVKALDYEQLQS-VKLSIAVKNKAEFHQSVISRYRVQSTPVTIQVINVREGIA  333
                 G + V+K LD +Q     +    V  + E  + ++    VQ     + + ++ +
Sbjct  1611  RTTGEIFVLKPLDRDQPNGRPQWRFTVFAQDEGGEGLVGYADVQ-----VNLKDINDNAP  1665

Query  334   FRPASKTFTVQKGISSKKLVDYILGTYQAIDEDTNKAASNVK--YVMGRN------DGGY  385
                P    F     ++       ++ T  A+D D     +N K  Y + +N
Sbjct  1666  IFPQGVYF---GNVTENGTAGMVVMTMTAVDYDDPNEGTNAKLIYSIEKNVIEEETGSPI  1722

Query  386   LMIDSKTAEIK-FVKNMNRDSTFIVNKTITAEVLAIDEYTGKTSTGTVYVRVPDFNDNCP  444
                I++ T  IK  V  ++R+ T       + +V+A+D   G   TGT  +RV D ND  P
Sbjct  1723  FEIEADTGVIKTAVCCLDRERT----PDYSIQVVAMDG-GGLKGTGTASIRVKDINDMPP  1777

 Score = 75.9 bits (185),  Expect = 2e-12, Method: Compositional matrix adjust.
 Identities = 108/429 (25%), Positives = 178/429 (41%), Gaps = 53/429 (12%)

Query  53    VDKNTGDINITAIVDREETPSFLITCRALNAQGLDVEKPLILTVKILDINDNPPVFSQ-Q  111
             +D++  ++N+TA+ D         +C  +N          ++ V I D+NDN PVF
Sbjct  950   LDRDKYELNVTAMDDG--------SC-CVNGDATIHTSTAVVVVFITDVNDNKPVFKDCG  1000

Query  112   IFMGEIEENSASNSLVMILNATDADEPNHLNSKIAFKIVSQEPAGTPMFLLSRNTGEVRT  171
              +  ++EE + + S V+ + ATD D+   +N ++ + IV Q       F +   TGEV T
Sbjct  1001  TYYPKVEEGAPNGSPVIKVQATDEDK--GVNGQVKYSIVQQPNQKGTKFTVDEETGEVST  1058

Query  172   LTNSLDRE--QASSYRLVVSGADKDGEGLSTQCECNIKVKDVNDNFPMFRDSQYSARIEE  229
                  DRE        + V   D+    L   C   +++ DVNDN P+F   +Y   +++
Sbjct  1059  -NKVFDREGDDGKFVSVTVKATDQGEPSLEGVCSFTVEITDVNDNPPLFDRQKYVENVKQ  1117

Query  230   NI-LSSELLRFQVTDLDEEYTDNWLAVYFFT---SGNEGNWFEIQTDPRTNEGILKVVKA  285
             +  + + +LR  V+  DE+  +N   VY +    S N+  +FEIQ  P +   +LK  K
Sbjct  1118  DASIGTNILR--VSASDEDADNNGAIVYTLSAPHSPNDLEYFEIQ--PESGWIVLK--KP  1171

Query  286   LD-YEQLQSVKLSIAVK----------NKAEFHQSVISRYRVQSTPV----TIQVINVRE  330
```

```
             LD    + SV+    +K           + A+    +    R    P+         I++RE
Sbjct  1172  LDGCYSVSSVRERYRLKVYASDRGDPPSTADVDVELDVVDRNNKPPIWDRNIYGPIHIRE  1231

Query  331   GIAFRPASKTFTVQKGISSKKLVDYILGTYQAIDEDTNKAASNVKYVMGRNDGGYLMIDS  390
              +              GI     V Y L          TNK    +  Y+  R D G
Sbjct  1232  NVTVGTVVTAIKASSGIEGNPTVFYRL--MPGSTAQTNKL--HTFYLQQRPDNG-----D  1282

Query  391   KTAEIKFVKNMNRDSTFIVNKTITAEVLAIDEYTGKTSTGTVYVRVPDFNDNCPTAV-LE  449
               A+IK    ++ ++    N TI  E     +     S  TVY+++ D ND  P     E
Sbjct  1283  TWADIKVNHPLDFETIKEYNLTIRVENNGAQQL---ASEATVYIQLEDVNDEIPLFTERE  1339

Query  450   KDAVCSSSP  458
             ++ V    P
Sbjct  1340  QETVLEGEP  1348

 Score = 61.2 bits (147),  Expect = 5e-08, Method: Compositional matrix adjust.
 Identities = 87/366 (24%), Positives = 156/366 (43%), Gaps = 38/366 (10%)

Query  51    FVVDKNTGDINITAIVDRE-ETPSFL-ITCRALNAQGLDVEKPLILTVKILDINDNPPVF  108
             F VD+ TG+++    + DRE +   F+ +T +A +      +E      TV+I D+NDNPP+F
Sbjct  1047  FTVDEETGEVSTNKVFDREGDDGKFVSVTVKATDQGEPSLEGVCSFTVEITDVNDNPPLF  1106

Query  109   SQQIFMGEIEENSASNSLVMILNATDADEPNHLNSKIAFKIVS-QEPAGTPMFLLSRNTG  167
              +Q ++  ++++++  + ++ ++A+D D  N  N  I + + +   P     F +   +G
Sbjct  1107  DRQKYVENVKQDASIGTNILRVSASDEDADN--NGAIVYTLSAPHSPNDLEYFEIQPESG  1164

Query  168   EVRTLTNSLDREQASS-----YRLVVSGADKDGEGLSTQCECNIKVKDVNDNFPMFRDSQ  222
              +  L   LD   + S      YRL V  +D+ G+  ST             +N P   D
Sbjct  1165  WI-VLKKPLDGCYSVSSVRERYRLKVYASDR-GDPPSTADVDVELDVVDRNNKPPIWDRN  1222

Query  223   YSA--RIEENILSSELLRFQVTDLDEEYTDNWLAVYFFTSG-----NEGNWFEIQTDPRT  275
                   I EN+    ++               N    Y    G     N+ + F +Q  P
Sbjct  1223  IYGPIHIRENVTVGTVV--TAIKASSGIEGNPTVFYRLMPGSTAQTNKLHTFYLQQRPDN  1280

Query  276   NE--GILKVVKALDYEQLQSVKLSIAVKNKAEFHQSVISRYRVQSTPVTIQVINVREGIA  333
              +    +KV   LD+E ++   L+I V+N          ++        V IQ+ +V + I
Sbjct  1281  GDTWADIKVNHPLDFETIKEYNLTIRVENNG-------AQQLASEATVYIQLEDVNDEIP  1333

Query  334   FRPASKTFTVQKGISSKKLVDYILGTYQAIDEDTNKAASNVKYVM---GRNDG-GYLMID  389
                  +  TV +G       V  +     AID+D     + V Y +    RN+G  +  I+
Sbjct  1334  LFTEREQETVLEGEPIGTKVTQV----NAIDKDGTFPNNQVYYYIVDSPRNEGKDFFEIN  1389

Query  390   SKTAEI  395
              ++ EI
Sbjct  1390  LQSGEI  1395

 Score = 55.8 bits (133),  Expect = 3e-06, Method: Compositional matrix adjust.
 Identities = 49/158 (31%), Positives = 78/158 (49%), Gaps = 24/158 (15%)

Query  102   NDNPPVFSQQIFMGEIEENSASNSLVMILNATDADEPNHLNSKIAFKIVSQEPAGTP--M  159
             ND  P FSQQ++   ++EN+  N+LV  + A+D D  N     + F  V     GT
Sbjct  880   NDEEPKFSQQVYTPNVDENAGPNTLVTTVVASDKDGDN-----VRFGFVG---GGTSSGQ  931

Query  160   FLLSRNTGEVRTLTN--SLDREQASSYRLVVSGADK-------DGEGLSTQCECNIKVKD  210
             F++   TG +R  +   SLDR++   Y L V+  D         D    ++     + + D
Sbjct  932   FVIEEITGVIRLHSKAISLDRDK---YELNVTAMDDGSCCVNGDATIHTSTAVVVVFITD  988

Query  211   VNDNFPMFRD-SQYSARIEENILS-SELLRFQVTDLDE  246
             VNDN P+F+D   Y  ++EE   + S +++ Q TD D+
Sbjct  989   VNDNKPVFKDCGTYYPKVEEGAPNGSPVIKVQATDEDK  1026

 Score = 53.9 bits (128),  Expect = 1e-05, Method: Compositional matrix adjust.
 Identities = 70/306 (23%), Positives = 126/306 (41%), Gaps = 30/306 (10%)

Query  20    PIAKIT-SDYQATQKITYRI---SGVGIDQPPFGIFVVDKNTGDINITAIVDRE---ETP  72
             PI  +T  D   T K  Y++   SG G D+  F +    +  TG + I    +D E   ++
Sbjct  1800  PILTVTVHDEDETNKFQYKVIDNSGYGADK--FTMVRNNDGTGSLKIVQPLDYEDPLQSN  1857
```

```
Query  73    SFLITCRALNAQGLDVEKPLI------LTVKILDINDNPPVFSQQIFMGEIEENSASNSL  126
              F    + +N +G D +          + VK+ DINDN P F +      + EN+
Sbjct  1858  GFRFRIQ-VNDKGEDNDNDKYHVAYSWVVVKLRDINDNKPQFERPNIEVSVYENADVGKT  1916

Query  127   VMILNATDADEPNHLNSKIAFKIVSQEPAGTPMFLLSRNTGEVRTLTNSLDREQASSYRL  186
             +    ATD D+     SK+++ I   + +       S N     T+  +LDRE    +++
Sbjct  1917  LETFKATDPDQGG--KSKVSYAI---DRSSDRQRQFSINQEGTVTIQRNLDREVTPRHQV  1971

Query  187   VVSGADKDGEGLSTQCECNIKVKDVNDNFPMFRDSQYSARIEENILSSELLRFQVTDLDE  246
              +   D      +      + V+D+NDN P F    Y   + E++   +++    TD D+
Sbjct  1972  KILAIDDGIPPKTATATLTVIVQDINDNPPKFL-KDYRPVLPEHVPPRKVVEILATDDDD  2030

Query  247   EYTDNWLAVYFFTSGNEGNW----FEIQTDPRTNEG----ILKVVKALDYEQLQSVKLSI  298
                 N     F       +     F+++ D +   G    I+  +++ D EQ +   + I
Sbjct  2031  RSKSNGPPFQFRLDPGADDIIRASFKVEQDQKGANGDGMAIVSSLRSFDREQQKEYLIPI  2090

Query  299   AVKNKA  304
              +K+
Sbjct  2091  VIKDHG  2096

 Score = 51.6 bits (122),  Expect = 5e-05, Method: Compositional matrix adjust.
 Identities = 96/398 (24%), Positives = 163/398 (41%), Gaps = 54/398 (14%)

Query  106   PVFSQQIFMGEIEENSASNSLVMILNATDADEPNHLNSKIAFKIVSQ-EPAGTPMFLLSR  164
             P F    +  EI EN   +S ++ + A      +  + +I + + +Q + AGT  F +
Sbjct  677   PQFYMPSYEAEIPENQKKDSDIISVKAK-----SFADREIRYTLKAQGQGAGT--FNIGP  729

Query  165   NTGEVRTLTNSLDRE---QASSYRLVVSGADKDGEGLSTQCECNIKVKDVNDNFPMFRDS  221
              +G V+ L   LD E   Q   Y LVV+ A +D  G ST  E  I+V DVNDN P F
Sbjct  730   TSGIVK-LAKELDFEDLRQPHVYSLVVT-ATEDSGGFSTSVELTIRVTDVNDNAPKFELP  787

Query  222   QYSAR-IEENI-LSSELLRFQVTDLDEEYTDNWLAVYFFTSGNEGNWFEIQTDPRTNEGI  279
              Y A  ++E+I + + +L+ +  D D         + +  S +    F + ++     GI
Sbjct  788   DYQAHNVDEDIQIGTSILKVKAMDADSGAN---AEIEYLVSDDH---FAVDSN-----GI  836

Query  280   LKVVKALDYEQLQSV-KLSIAVKNKAEFHQSVISRYRVQSTPVTIQVINVREGIAFRPAS  338
             +   K LD +  +   +  +  K+K E  ++  +  RV +           E   F
Sbjct  837   IVNNKQLDADNNNAYYEFVVTAKDKGEPAKTGTATVRVYTKNKN------DEEPKFSQQV  890

Query  339   KTFTVQKGISSKKLVDYILGTYQAIDEDTNKAASNVKY--VMGRNDGGYLMIDSKTAEIK  396
              T  V +      LV  ++ +        +K   NV++  V G    G  +I+  T  I+
Sbjct  891   YTPNVDENAGPNTLVTTVVAS--------DKDGDNVRFGFVGGGTSSGQFVIEEITGVIR  942

Query  397   FVK---NMNRDSTFI-VNKTITAEVLAIDEYTGKTSTGTVYVRVPDFNDN------CPTA  446
                   +++RD   + V            + T  TST  V V + D NDN      C T
Sbjct  943   LHSKAISLDRDKYELNVTAMDDGSCCVNGDATIHTSTAVVVVFITDVNDNKPVFKDCGTY  1002

Query  447   VLE-KDAVCSSSPSVVVSARTLNNRYTGPYTFALEDQP  483
               + ++   + SP + V A   +    G   +++  QP
Sbjct  1003  YPKVEEGAPNGSPVIKVQATDEDKGVNGQVKYSIVQQP  1040

 Score = 44.7 bits (104),  Expect = 0.008, Method: Compositional matrix adjust.
 Identities = 48/199 (24%), Positives = 73/199 (37%), Gaps = 19/199 (10%)

Query  27    DYQATQKITYRISGVGIDQPPFGIFVVDKNTGDINITAIVDREETPSFLITCRALNAQGL  86
             D     K++Y I      Q  F I       G + I   +DRE TP   +   A++
Sbjct  1926  DQGGKSKVSYAIDRSSDRQRQFSI----NQEGTVTIQRNLDREVTPRHQVKILAIDDGIP  1981

Query  87    DVEKPLILTVKILDINDNPPVFSQQIFMGEIEENSASNSLVMILNATDADEPNHLNSKIA  146
                    LTV + DINDNPP F +  +   + E+     +V IL   D D
Sbjct  1982  PKTATATLTVIVQDINDNPPKFLKD-YRPVLPEHVPPRKVVEILATDDDDRSKSNGPPFQ  2040

Query  147   FKIVSQEPAGTPMFLLSRNTGEVRTLTN-----------SLDREQASSYRLVVSGADKDG  195
             F++   +P    +   S    + +   N           S DREQ   Y + +   D
Sbjct  2041  FRL---DPGADDIIRASFKVEQDQKGANGDGMAIVSSLRSFDREQQKEYLIPIVIKDHGN  2097

Query  196   EGLSTQCECNIKVKDVNDN  214
               ++      + + DVNDN
Sbjct  2098  PAMTGTSTLTVVIGDVNDN  2116
```

```
 Score = 40.8 bits (94),  Expect = 0.11, Method: Compositional matrix adjust.
 Identities = 55/242 (23%), Positives = 102/242 (42%), Gaps = 22/242 (9%)

Query  4    KFAKPCREGE--DNSKRNP-IAKITSDYQATQKITYRISGVGIDQPPFGIFVVDKNTGDI  60
            +F  P  E E  +N K++  I  + +   A ++I Y +   G      G F +   +G +
Sbjct  678  QFYMPSYEAEIPENQKKDSDIISVKAKSFADREIRYTLKAQGQGA---GTFNIGPTSGIV  734

Query  61   NITAIVDREE--TPSF--LITCRALNAQGLDVEKPLILTVKILDINDNPPVFSQQIFMGE  116
             +   +D E+   P    L+     ++ G      + LT+++ D+NDN P F    +
Sbjct  735  KLAKELDFEDLRQPHVYSLVVTATEDSGGFSTS--VELTIRVTDVNDNAPKFELPDYQAH  792

Query  117  -IEENSASNSLVMILNATDADEPNHLNSKIAFKIVSQEPAGTPMFLLSRNTGEVRTLTNS  175
             ++E+    + ++ + A DAD  +  N++I + +VS +      F +  N   V
Sbjct  793  NVDEDIQIGTSILKVKAMDAD--SGANAEIEY-LVSDD-----HFAVDSNGIIVNNKQLD  844

Query  176  LDREQASSYRLVVSGADKDGEGLSTQCECNIKVKDVNDNFPMFRDSQYSARIEENILSSE  235
             D   A  Y  VV+  DK     +      +  K+ ND  P F    Y+  ++EN   +
Sbjct  845  ADNNNA-YYEFVVTAKDKGEPAKTGTATVRVYTKNKNDEEPKFSQQVYTPNVDENAGPNT  903

Query  236  LL  237
            L+
Sbjct  904  LV  905

 Score = 39.3 bits (90),  Expect = 0.30, Method: Compositional matrix adjust.
 Identities = 55/225 (24%), Positives = 88/225 (39%), Gaps = 24/225 (11%)

Query  75   LITCRALNAQGLDVEKPLILTVKILDINDNPPVFSQQIF--MGEIEENSASNSLVMILNA  132
            +IT +  NA GL       + + + D+ND PP F  +       ++ N+  N+ V  L A
Sbjct  539  IITNQGHNA-GLKYTDNQRVIIHVKDVNDEPPYFINRPLPMQAVVQLNAPPNTPVFTLQA  597

Query  133  TDADEPNHLNSKIAFKIVSQEPAGTPMFLLSRNTGEVRTLTNSLDREQASSYRLVVSGAD  192
             D D     +  I + IV     G   F +   +G VRT    L  +    Y L V   D
Sbjct  598  RDPD----TDHNIHYFIVRDRTGG--RFEVDERSGVVRTRGTDL-FQLDMEYVLYVKAED  650

Query  193  KDGE----GLSTQCECNIKVKDVNDNFPMFRDSQYSARIEEN-ILSSELLRFQVTDLDEE  247
            ++G+       +  E  + +       P F    Y A I EN    S+++  +     +
Sbjct  651  QNGKVDDRRFQSTPEERLSIVG-GKRAPQFYMPSYEAEIPENQKKDSDIISVKA----KS  705

Query  248  YTDNWLAVYFFTSGNEGNWFEIQTDPRTNEGILKVVKALDYEQLQ  292
            + D  +       G     F I        GI+K+ K LD+E L+
Sbjct  706  FADREIRYTLKAQGQGAGTFNIG----PTSGIVKLAKELDFEDLR  746

>AALB20_033171.P66514 | transcript=AALB20_033171.R66514 | gene=AALB20_033171
| organism=Anopheles_albimanus_STECLA_2020 |
gene_product=neural-cadherin, transcript variant X38 |
transcript_product=neural-cadherin,
transcript variant X20 | location=CM024292:75037560-75208393(+)
| protein_length=3131 | sequence_SO=chromosome
| SO=protein_coding_gene | is_pseudo=false
Length=3131

 Score = 144 bits (364),  Expect = 1e-34, Method: Compositional matrix adjust.
 Identities = 125/437 (29%), Positives = 211/437 (48%), Gaps = 33/437 (8%)

Query  26    SDYQATQKITYRISGVGID--QPPFGIFVVDKNTGDINITAIVDREET---PSFLITCRA  80
              D    Q I Y ++G GID   P    F +++ TG+I +   +DR++    P +  T  A
Sbjct  1580  GDKDRPQNIVYFLTGQGIDPDNPANSKFDINRTTGEIFVLKPLDRDQPNGRPQWRFTVFA  1639

Query  81    LNAQGLDVEKPLILTVKILDINDNPPVFSQQIFMGEIEENSASNSLVMILNATDADEPNH  140
              +  G  +     + V + DINDN P+F Q ++ G + EN  +  +VM + A D D+PN
Sbjct  1640  QDEGGEGLVGYADVQVNLKDINDNAPIFPQGVYFGNVTENGTAGMVVMTMTAVDYDDPNE  1699

Query  141   -LNSKIAFKI---VSQEPAGTPMFLLSRNTGEVRTLTNSLDREQASSYRLVVSGADKDGE  196
               N+K+ + I   V +E  G+P+F +  +TG ++T    LDRE+   Y + V      DG
Sbjct  1700  GTNAKLIYSIEKNVIEEETGSPIFEIEADTGVIKTAVCCLDRERTPDYSIQVVAM--DGG  1757

Query  197   GLSTQCECNIKVKDVNDNFPMFRDSQYSARIEENILSS--ELLRFQVTDLDEEYTDNWLA  254
```

```
             GL      +I+VKD+ND  P F   ++   ++E   ++  E+    VT  DE+ T+ +
Sbjct  1758  GLKGTGTASIRVKDINDMPPQFTKDEWFTEVDETDGTNLPEMPILTVTVHDEDETNKFQY  1817

Query  255   VYFFTSGNEGNWFEIQTDPRTNEGI--LKVVKALDYE---QLQSVKLSIAVKNKAEFHQS  309
                  SG   + F +    R N+G   LK+V+ LDYE   Q    +  I V +K E + +
Sbjct  1818  KVIDNSGYGADKFTMV---RNNDGTGSLKIVQPLDYEDPLQSNGFRFRIQVNDKGEDNDN  1874

Query  310   VISRYRVQSTPVTIQVINVREGIAFRPASKTFTVQKGISSKKLVDYILGTYQAIDEDTNK  369
                +Y V  + V +++ ++ +    +P  +   ++  +     V   L T++A D D
Sbjct  1875  --DKYHVAYSWVVVKLRDINDN---KPQFERPNIEVSVYENADVGKTLETFKATDPDQG-  1928

Query  370   AASNVKYVMGR-NDGGYLMIDSKTAEIKFVKNMNRDSTFIVNKTITAEVLAIDE-YTGKT  427
               S V Y + R +D       ++   +   +N++R+    V      ++LAID+    KT
Sbjct  1929  GKSKVSYAIDRSSDRQRQFSINQEGTVTIQRNLDRE----VTPRHQVKILAIDDGIPPKT  1984

Query  428   STGTVYVRVPDFNDNCP  444
             +T T+ V V D NDN P
Sbjct  1985  ATATLTVIVQDINDNPP  2001

 Score = 85.9 bits (211),  Expect = 1e-15, Method: Compositional matrix adjust.
 Identities = 108/420 (26%), Positives = 177/420 (42%), Gaps = 52/420 (12%)

Query  50    IFVVDKNTGDINITAIVDREETPSFLITCRAL----------NAQGLDVEKPLILTVKIL  99
              F ++  +G+I    + DRE+  ++ +   A           N Q   V K   + + I
Sbjct  1385  FFEINLQSGEIFTKVVFDREKQGAYALEVEARDGAPSARPNSNQQPNSVTK--FIRIGIA  1442

Query  100   DINDNPPVFSQQIFMGEIEENSASNSLVMILNATDADEPNHLNSKIAFKIVSQEPAGTPM  159
             D NDNPP F + ++  E++EN      V+ + A D DE    +S+I ++I S    G
Sbjct  1443  DKNDNPPYFDKALYEAEVDENEDIQHTVLTVTAKDHDE----SSRIRYEITSGNIGGA--  1496

Query  160   FLLSRNTGEVRTLTNSLDREQASSYRLVVSGADKDGEGLSTQCECNIKVKDVNDNFPMFR  219
             F +   TG +  +  +LD E    Y L ++ +D   E  +T     I VKDVNDN P+F
Sbjct  1497  FAVKNMTGAI-YVAGALDYETRKRYELRLAASDNLKENYTTVV---IHVKDVNDNPPVFE  1552

Query  220   DSQYSARIEENI---LSSELLRFQVTDLDEEYTDNWLAVYFFTSG--NEGNWFEIQTDPR  274
                Y  +I E     L   +L+   TD D++   N   VYF T    +  N    + D
Sbjct  1553  RPTYRTQITEEDDRNLPKRVLQVTATDGDKDRPQN--IVYFLTGQGIDPDNPANSKFDIN  1610

Query  275   TNEGILKVVKALDYEQLQS-VKLSIAVKNKAEFHQSVISRYRVQSTPVTIQVINVREGIA  333
                G + V+K LD +Q     +    V  + E  + ++    VQ     + + ++ +
Sbjct  1611  RTTGEIFVLKPLDRDQPNGRPQWRFTVFAQDEGGEGLVGYADVQ-----VNLKDINDNAP  1665

Query  334   FRPASKTFTVQKGISSKKLVDYILGTYQAIDEDTNKAASNVK--YVMGRN------DGGY  385
               P    F     ++       ++ T  A+D D     +N K  Y + +N
Sbjct  1666  IFPQGVYF---GNVTENGTAGMVVMTMTAVDYDDPNEGTNAKLIYSIEKNVIEEETGSPI  1722

Query  386   LMIDSKTAEIK-FVKNMNRDSTFIVNKTITAEVLAIDEYTGKTSTGTVYVRVPDFNDNCP  444
               I++ T  IK  V  ++R+ T       + +V+A+D   G   TGT  +RV D ND  P
Sbjct  1723  FEIEADTGVIKTAVCCLDRERT----PDYSIQVVAMDG-GGLKGTGTASIRVKDINDMPP  1777

 Score = 75.9 bits (185),  Expect = 2e-12, Method: Compositional matrix adjust.
 Identities = 108/429 (25%), Positives = 178/429 (41%), Gaps = 53/429 (12%)

Query  53    VDKNTGDINITAIVDREETPSFLITCRALNAQGLDVEKPLILTVKILDINDNPPVFSQ-Q  111
             +D++  ++N+TA+ D         +C  +N          ++ V I D+NDN PVF
Sbjct  950   LDRDKYELNVTAMDDG--------SC-CVNGDATIHTSTAVVVVFITDVNDNKPVFKDCG  1000

Query  112   IFMGEIEENSASNSLVMILNATDADEPNHLNSKIAFKIVSQEPAGTPMFLLSRNTGEVRT  171
              +  ++EE + + S V+ + ATD D+   +N ++ + IV Q       F +   TGEV T
Sbjct  1001  TYYPKVEEGAPNGSPVIKVQATDEDK--GVNGQVKYSIVQQPNQKGTKFTVDEETGEVST  1058

Query  172   LTNSLDRE--QASSYRLVVSGADKDGEGLSTQCECNIKVKDVNDNFPMFRDSQYSARIEE  229
                  DRE        + V   D+    L   C   +++ DVNDN P+F   +Y   +++
Sbjct  1059  -NKVFDREGDDGKFVSVTVKATDQGEPSLEGVCSFTVEITDVNDNPPLFDRQKYVENVKQ  1117

Query  230   NI-LSSELLRFQVTDLDEEYTDNWLAVYFFT---SGNEGNWFEIQTDPRTNEGILKVVKA  285
             +  + + +LR  V+  DE+  +N   VY  +   S N+  +FEIQ  P +   +LK  K
Sbjct  1118  DASIGTNILR--VSASDEDADNNGAIVYTLSAPHSPNDLEYFEIQ--PESGWIVLK--KP  1171
```

```
Query  286   LD-YEQLQSVKLSIAVK----------NKAEFHQSVISRYRVQSTPV----TIQVINVRE  330
             LD    + SV+    +K          + A+    +    R    P+        I++RE
Sbjct  1172  LDGCYSVSSVRERYRLKVYASDRGDPPSTADVDVELDVVDRNNKPPIWDRNIYGPIHIRE  1231

Query  331   GIAFRPASKTFTVQKGISSKKLVDYILGTYQAIDEDTNKAASNVKYVMGRNDGGYLMIDS  390
              +             GI     V Y L         TNK   +  Y+  R D G
Sbjct  1232  NVTVGTVVTAIKASSGIEGNPTVFYRL--MPGSTAQTNKL--HTFYLQQRPDNG-----D  1282

Query  391   KTAEIKFVKNMNRDSTFIVNKTITAEVLAIDEYTGKTSTGTVYVRVPDFNDNCPTAV-LE  449
               A+IK    ++ ++    N TI  E     +     S  TVY+++ D ND  P     E
Sbjct  1283  TWADIKVNHPLDFETIKEYNLTIRVENNGAQQL---ASEATVYIQLEDVNDEIPLFTERE  1339

Query  450   KDAVCSSSP  458
             ++ V    P
Sbjct  1340  QETVLEGEP  1348

 Score = 61.2 bits (147),  Expect = 5e-08, Method: Compositional matrix adjust.
 Identities = 87/366 (24%), Positives = 156/366 (43%), Gaps = 38/366 (10%)

Query  51    FVVDKNTGDINITAIVDRE-ETPSFL-ITCRALNAQGLDVEKPLILTVKILDINDNPPVF  108
             F VD+ TG+++   + DRE +   F+ +T +A +     +E     TV+I D+NDNPP+F
Sbjct  1047  FTVDEETGEVSTNKVFDREGDDGKFVSVTVKATDQGEPSLEGVCSFTVEITDVNDNPPLF  1106

Query  109   SQQIFMGEIEENSASNSLVMILNATDADEPNHLNSKIAFKIVS-QEPAGTPMFLLSRNTG  167
              +Q ++  ++++++  + ++ ++A+D D  N  N  I + + +   P     F +   +G
Sbjct  1107  DRQKYVENVKQDASIGTNILRVSASDEDADN--NGAIVYTLSAPHSPNDLEYFEIQPESG  1164

Query  168   EVRTLTNSLDREQASS-----YRLVVSGADKDGEGLSTQCECNIKVKDVNDNFPMFRDSQ  222
              +  L   LD   + S     YRL V  +D+ G+  ST            +N P   D
Sbjct  1165  WI-VLKKPLDGCYSVSSVRERYRLKVYASDR-GDPPSTADVDVELDVVDRNNKPPIWDRN  1222

Query  223   YSA--RIEENILSSELLRFQVTDLDEEYTDNWLAVYFFTSG-----NEGNWFEIQTDPRT  275
                   I EN+    ++             N    Y    G     N+ + F +Q  P
Sbjct  1223  IYGPIHIRENVTVGTVV--TAIKASSGIEGNPTVFYRLMPGSTAQTNKLHTFYLQQRPDN  1280

Query  276   NE--GILKVVKALDYEQLQSVKLSIAVKNKAEFHQSVISRYRVQSTPVTIQVINVREGIA  333
              +    +KV   LD+E ++   L+I V+N         ++       V IQ+ +V + I
Sbjct  1281  GDTWADIKVNHPLDFETIKEYNLTIRVENNG-------AQQLASEATVYIQLEDVNDEIP  1333

Query  334   FRPASKTFTVQKGISSKKLVDYILGTYQAIDEDTNKAASNVKYVM---GRNDG-GYLMID  389
                  +  TV +G      V  +     AID+D     + V Y +    RN+G  +  I+
Sbjct  1334  LFTEREQETVLEGEPIGTKVTQV----NAIDKDGTFPNNQVYYYIVDSPRNEGKDFFEIN  1389

Query  390   SKTAEI  395
              ++ EI
Sbjct  1390  LQSGEI  1395

 Score = 55.8 bits (133),  Expect = 3e-06, Method: Compositional matrix adjust.
 Identities = 49/158 (31%), Positives = 78/158 (49%), Gaps = 24/158 (15%)

Query  102   NDNPPVFSQQIFMGEIEENSASNSLVMILNATDADEPNHLNSKIAFKIVSQEPAGTP--M  159
             ND  P FSQQ++   ++EN+  N+LV  + A+D D  N     + F  V     GT
Sbjct  880   NDEEPKFSQQVYTPNVDENAGPNTLVTTVVASDKDGDN-----VRFGFVG---GGTSSGQ  931

Query  160   FLLSRNTGEVRTLTN--SLDREQASSYRLVVSGADK-------DGEGLSTQCECNIKVKD  210
             F++   TG +R  +   SLDR++   Y L V+  D        D    ++     + + D
Sbjct  932   FVIEEITGVIRLHSKAISLDRDK---YELNVTAMDDGSCCVNGDATIHTSTAVVVVFITD  988

Query  211   VNDNFPMFRD-SQYSARIEENILS-SELLRFQVTDLDE  246
             VNDN P+F+D   Y  ++EE   + S +++ Q TD D+
Sbjct  989   VNDNKPVFKDCGTYYPKVEEGAPNGSPVIKVQATDEDK  1026

 Score = 53.9 bits (128),  Expect = 1e-05, Method: Compositional matrix adjust.
 Identities = 70/306 (23%), Positives = 126/306 (41%), Gaps = 30/306 (10%)

Query  20    PIAKIT-SDYQATQKITYRI---SGVGIDQPPFGIFVVDKNTGDINITAIVDRE---ETP  72
             PI  +T  D   T K  Y++   SG G D+  F +   +  TG + I   +D E   ++
Sbjct  1800  PILTVTVHDEDETNKFQYKVIDNSGYGADK--FTMVRNNDGTGSLKIVQPLDYEDPLQSN  1857
```

```
Query  73    SFLITCRALNAQGLDVEKPLI------LTVKILDINDNPPVFSQQIFMGEIEENSASNSL  126
              F    + +N +G D +          + VK+ DINDN P F +      + EN+
Sbjct  1858  GFRFRIQ-VNDKGEDNDNDKYHVAYSWVVVKLRDINDNKPQFERPNIEVSVYENADVGKT  1916

Query  127   VMILNATDADEPNHLNSKIAFKIVSQEPAGTPMFLLSRNTGEVRTLTNSLDREQASSYRL  186
             +    ATD D+     SK+++ I   + +       S N     T+  +LDRE    +++
Sbjct  1917  LETFKATDPDQGG--KSKVSYAI---DRSSDRQRQFSINQEGTVTIQRNLDREVTPRHQV  1971

Query  187   VVSGADKDGEGLSTQCECNIKVKDVNDNFPMFRDSQYSARIEENILSSELLRFQVTDLDE  246
              +   D      +      + V+D+NDN P F    Y   + E++   +++    TD D+
Sbjct  1972  KILAIDDGIPPKTATATLTVIVQDINDNPPKFL-KDYRPVLPEHVPPRKVVEILATDDDD  2030

Query  247   EYTDNWLAVYFFTSGNEGNW----FEIQTDPRTNEG----ILKVVKALDYEQLQSVKLSI  298
                 N     F       +     F+++ D +   G    I+  +++ D EQ +   + I
Sbjct  2031  RSKSNGPPFQFRLDPGADDIIRASFKVEQDQKGANGDGMAIVSSLRSFDREQQKEYLIPI  2090

Query  299   AVKNKA  304
              +K+
Sbjct  2091  VIKDHG  2096

 Score = 51.6 bits (122),  Expect = 5e-05, Method: Compositional matrix adjust.
 Identities = 96/398 (24%), Positives = 163/398 (41%), Gaps = 54/398 (14%)

Query  106   PVFSQQIFMGEIEENSASNSLVMILNATDADEPNHLNSKIAFKIVSQ-EPAGTPMFLLSR  164
             P F    +  EI EN   +S ++ + A      +  + +I + + +Q + AGT  F +
Sbjct  677   PQFYMPSYEAEIPENQKKDSDIISVKAK-----SFADREIRYTLKAQGQGAGT--FNIGP  729

Query  165   NTGEVRTLTNSLDRE---QASSYRLVVSGADKDGEGLSTQCECNIKVKDVNDNFPMFRDS  221
              +G V+ L   LD E   Q   Y LVV+ A +D  G ST  E  I+V DVNDN P F
Sbjct  730   TSGIVK-LAKELDFEDLRQPHVYSLVVT-ATEDSGGFSTSVELTIRVTDVNDNAPKFELP  787

Query  222   QYSAR-IEENI-LSSELLRFQVTDLDEEYTDNWLAVYFFTSGNEGNWFEIQTDPRTNEGI  279
              Y A  ++E+I + + +L+ +  D D         + +  S +    F + ++     GI
Sbjct  788   DYQAHNVDEDIQIGTSILKVKAMDADSGAN---AEIEYLVSDDH---FAVDSN-----GI  836

Query  280   LKVVKALDYEQLQSV-KLSIAVKNKAEFHQSVISRYRVQSTPVTIQVINVREGIAFRPAS  338
             +   K LD +   +  +  +  K+K E  ++  +  RV +           E   F
Sbjct  837   IVNNKQLDADNNNAYYEFVVTAKDKGEPAKTGTATVRVYTKNKN------DEEPKFSQQV  890

Query  339   KTFTVQKGISSKKLVDYILGTYQAIDEDTNKAASNVKY--VMGRNDGGYLMIDSKTAEIK  396
              T  V +      LV  ++ +        +K   NV++  V G    G  +I+  T  I+
Sbjct  891   YTPNVDENAGPNTLVTTVVAS--------DKDGDNVRFGFVGGGTSSGQFVIEEITGVIR  942

Query  397   FVK---NMNRDSTFI-VNKTITAEVLAIDEYTGKTSTGTVYYVRVPDFNDN------CPTA  446
                   +++RD   + V            + T  TST  V V + D NDN      C T
Sbjct  943   LHSKAISLDRDKYELNVTAMDDGSCCVNGDATIHTSTAVVVVFITDVNDNKPVFKDCGTY  1002

Query  447   VLE-KDAVCSSSPSVVVSARTLNNRYTGPYTFALEDQP  483
               + ++   + SP + V A   +    G   +++  QP
Sbjct  1003  YPKVEEGAPNGSPVIKVQATDEDKGVNGQVKYSIVQQP  1040

 Score = 44.7 bits (104),  Expect = 0.008, Method: Compositional matrix adjust.
 Identities = 48/199 (24%), Positives = 73/199 (37%), Gaps = 19/199 (10%)

Query  27    DYQATQKITYRISGVGIDQPPFGIFVVDKNTGDINITAIVDREETPSFLITCRALNAQGL  86
             D     K++Y I      Q  F I       G + I   +DRE TP   +   A++
Sbjct  1926  DQGGKSKVSYAIDRSSDRQRQFSI----NQEGTVTIQRNLDREVTPRHQVKILAIDDGIP  1981

Query  87    DVEKPLILTVKILDINDNPPVFSQQIFMGEIEENSASNSLVMILNATDADEPNHLNSKIA  146
                    LTV + DINDNPP F +  +   + E+     +V IL   D D
Sbjct  1982  PKTATATLTVIVQDINDNPPKFLKD-YRPVLPEHVPPRKVVEILATDDDDRSKSNGPPFQ  2040

Query  147   FKIVSQEPAGTPMFLLSRNTGEVRTLTN-----------SLDREQASSYRLVVSGADKDG  195
             F++   +P    +   S    + +   N           S DREQ   Y + +   D
Sbjct  2041  FRL---DPGADDIIRASFKVEQDQKGANGDGMAIVSSLRSFDREQQKEYLIPIVIKDHGN  2097

Query  196   EGLSTQCECNIKVKDVNDN  214
               ++      + + DVNDN
```

```
Sbjct  2098  PAMTGTSTLTVVIGDVNDN  2116

 Score = 40.8 bits (94),  Expect = 0.11, Method: Compositional matrix adjust.
 Identities = 55/242 (23%), Positives = 102/242 (42%), Gaps = 22/242 (9%)

Query  4    KFAKPCREGE--DNSKRNP-IAKITSDYQATQKITYRISGVGIDQPPFGIFVVDKNTGDI  60
            +F  P  E E  +N K++  I  + +   A ++I Y +   G      G F +   +G +
Sbjct  678  QFYMPSYEAEIPENQKKDSDIISVKAKSFADREIRYTLKAQGQGA---GTFNIGPTSGIV  734

Query  61   NITAIVDREE--TPSF--LITCRALNAQGLDVEKPLILTVKILDINDNPPVFSQQIFMGE  116
             +   +D E+   P    L+     ++ G      + LT+++ D+NDN P F    +
Sbjct  735  KLAKELDFEDLRQPHVYSLVVTATEDSGGFSTS--VELTIRVTDVNDNAPKFELPDYQAH  792

Query  117  -IEENSASNSLVMILNATDADEPNHLNSKIAFKIVSQEPAGTPMFLLSRNTGEVRTLTNS  175
             ++E+    + ++ + A DAD  +  N++I + +VS +      F +  N   V
Sbjct  793  NVDEDIQIGTSILKVKAMDAD--SGANAEIEY-LVSDD-----HFAVDSNGIIVNNKQLD  844

Query  176  LDREQASSYRLVVSGADKDGEGLSTQCECNIKVKDVNDNFPMFRDSQYSARIEENILSSE  235
             D   A  Y  VV+  DK     +      +  K+ ND  P F    Y+  ++EN   +
Sbjct  845  ADNNNA-YYEFVVTAKDKGEPAKTGTATVRVYTKNKNDEEPKFSQQVYTPNVDENAGPNT  903

Query  236  LL  237
            L+
Sbjct  904  LV  905

 Score = 39.3 bits (90),  Expect = 0.30, Method: Compositional matrix adjust.
 Identities = 55/225 (24%), Positives = 88/225 (39%), Gaps = 24/225 (11%)

Query  75   LITCRALNAQGLDVEKPLILTVKILDINDNPPVFSQQIF--MGEIEENSASNSLVMILNA  132
            +IT +  NA GL       + + + D+ND PP F  +       ++ N+  N+ V  L A
Sbjct  539  IITNQGHNA-GLKYTDNQRVIIHVKDVNDEPPYFINRPLPMQAVVQLNAPPNTPVFTLQA  597

Query  133  TDADEPNHLNSKIAFKIVSQEPAGTPMFLLSRNTGEVRTLTNSLDREQASSYRLVVSGAD  192
             D D     +  I + IV     G   F +   +G VRT    L  +    Y L V   D
Sbjct  598  RDPD----TDHNIHYFIVRDRTGG--RFEVDERSGVVRTRGTDL-FQLDMEYVLYVKAED  650

Query  193  KDGE----GLSTQCECNIKVKDVNDNFPMFRDSQYSARIEEN-ILSSELLRFQVTDLDEE  247
            ++G+       +  E  + +       P F    Y A I EN    S+++  +     +
Sbjct  651  QNGKVDDRRFQSTPEERLSIVG-GKRAPQFYMPSYEAEIPENQKKDSDIISVKA----KS  705

Query  248  YTDNWLAVYFFTSGNEGNWFEIQTDPRTNEGILKVVKALDYEQLQ  292
            + D  +       G     F I        GI+K+ K LD+E L+
Sbjct  706  FADREIRYTLKAQGQGAGTFNIG----PTSGIVKLAKELDFEDLR  746

>AALB20_033171.P66509 | transcript=AALB20_033171.R66509 | gene=AALB20_033171
| organism=Anopheles_albimanus_STECLA_2020 |
gene_product=neural-cadherin, transcript variant X38 |
transcript_product=neural-cadherin,
transcript variant X18 | location=CM024292:75037560-75208393(+)
| protein_length=3131 | sequence_SO=chromosome
| SO=protein_coding_gene | is_pseudo=false
Length=3131

 Score = 144 bits (364),  Expect = 1e-34, Method: Compositional matrix adjust.
 Identities = 125/437 (29%), Positives = 211/437 (48%), Gaps = 33/437 (8%)

Query  26    SDYQATQKITYRISGVGID--QPPFGIFVVDKNTGDINITAIVDREET---PSFLITCRA  80
              D    Q I Y ++G GID   P    F +++ TG+I +   +DR++    P +  T  A
Sbjct  1580  GDKDRPQNIVYFLTGQGIDPDNPANSKFDINRTTGEIFVLKPLDRDQPNGRPQWRFTVFA  1639

Query  81    LNAQGLDVEKPLILTVKILDINDNPPVFSQQIFMGEIEENSASNSLVMILNATDADEPNH  140
              +  G  +     + V + DINDN P+F Q ++ G + EN  +  +VM + A D D+PN
Sbjct  1640  QDEGGEGLVGYADVQVNLKDINDNAPIFPQGVYFGNVTENGTAGMVVMTMTAVDYDDPNE  1699

Query  141   -LNSKIAFKI---VSQEPAGTPMFLLSRNTGEVRTLTNSLDREQASSYRLVVSGADKDGE  196
               N+K+ + I   V +E  G+P+F +  +TG ++T    LDRE+   Y + V     DG
Sbjct  1700  GTNAKLIYSIEKNVIEEETGSPIFEIEADTGVIKTAVCCLDRERTPDYSIQVVAM--DGG  1757
```

```
Query  197   GLSTQCECNIKVKDVNDNFPMFRDSQYSARIEENILSS--ELLRFQVTDLDEEYTDNWLA  254
             GL      +I+VKD+ND  P F   ++   ++E   ++  E+    VT  DE+ T+ +
Sbjct  1758  GLKGTGTASIRVKDINDMPPQFTKDEWFTEVDETDGTNLPEMPILTVTVHDEDETNKFQY  1817

Query  255   VYFFTSGNEGNWFEIQTDPRTNEGI--LKVVKALDYE---QLQSVKLSIAVKNKAEFHQS  309
                  SG   + F +    R N+G   LK+V+ LDYE   Q    +  I V +K E + +
Sbjct  1818  KVIDNSGYGADKFTMV---RNNDGTGSLKIVQPLDYEDPLQSNGFRFRIQVNDKGEDNDN  1874

Query  310   VISRYRVQSTPVTIQVINVREGIAFRPASKTFTVQKGISSKKLVDYILGTYQAIDEDTNK  369
                +Y V  + V +++ ++ +    +P  +   ++  +     V   L T++A D D
Sbjct  1875  --DKYHVAYSWVVVKLRDINDN---KPQFERPNIEVSVYENADVGKTLETFKATDPDQG-  1928

Query  370   AASNVKYVMGR-NDGGYLMIDSKTAEIKFVKNMNRDSTFIVNKTITAEVLAIDE-YTGKT  427
               S V Y + R +D       ++   +   +N++R+    V       ++LAID+    KT
Sbjct  1929  GKSKVSYAIDRSSDRQRQFSINQEGTVTIQRNLDRE----VTPRHQVKILAIDDGIPPKT  1984

Query  428   STGTVYVRVPDFNDNCP  444
             +T T+ V V D NDN P
Sbjct  1985  ATATLTVIVQDINDNPP  2001

 Score = 85.9 bits (211),  Expect = 1e-15, Method: Compositional matrix adjust.
 Identities = 108/420 (26%), Positives = 177/420 (42%), Gaps = 52/420 (12%)

Query  50    IFVVDKNTGDINITAIVDREETPSFLITCRAL----------NAQGLDVEKPLILTVKIL  99
              F ++  +G+I    + DRE+  ++ +   A            N Q   V K   + + I
Sbjct  1385  FFEINLQSGEIFTKVVFDREKQGAYALEVEARDGAPSARPNSNQQPNSVTK--FIRIGIA  1442

Query  100   DINDNPPVFSQQIFMGEIEENSASNSLVMILNATDADEPNHLNSKIAFKIVSQEPAGTPM  159
             D NDNPP F + ++  E++EN      V+ + A D DE    +S+I ++I S    G
Sbjct  1443  DKNDNPPYFDKALYEAEVDENEDIQHTVLTVTAKDHDE----SSRIRYEITSGNIGGA--  1496

Query  160   FLLSRNTGEVRTLTNSLDREQASSYRLVVSGADKDGEGLSTQCECNIKVKDVNDNFPMFR  219
             F +   TG +  +  +LD E    Y L ++ +D   E  +T      I VKDVNDN P+F
Sbjct  1497  FAVKNMTGAI-YVAGALDYETRKRYELRLAASDNLKENYTTVV---IHVKDVNDNPPVFE  1552

Query  220   DSQYSARIEENI---LSSELLRFQVTDLDEEYTDNWLAVYFFTSG--NEGNWFEIQTDPR  274
                Y  +I E     L   +L+   TD D++    N   VYF T    +  N    + D
Sbjct  1553  RPTYRTQITEEDDRNLPKRVLQVTATDGDKDRPQN--IVYFLTGQGIDPDNPANSKFDIN  1610

Query  275   TNEGILKVVKALDYEQLQS-VKLSIAVKNKAEFHQSVISRYRVQSTPVTIQVINVREGIA  333
                G + V+K LD +Q     +    V  + E  + ++    VQ     + + ++ +
Sbjct  1611  RTTGEIFVLKPLDRDQPNGRPQWRFTVFAQDEGGEGLVGYADVQ-----VNLKDINDNAP  1665

Query  334   FRPASKTFTVQKGISSKKLVDYILGTYQAIDEDTNKAASNVK--YVMGRN------DGGY  385
               P    F     ++       ++ T  A+D D     +N K  Y + +N
Sbjct  1666  IFPQGVYF---GNVTENGTAGMVVMTMTAVDYDDPNEGTNAKLIYSIEKNVIEEETGSPI  1722

Query  386   LMIDSKTAEIK-FVKNMNRDSTFIVNKTITAEVLAIDEYTGKTSTGTVYVRVPDFNDNCP  444
               I++ T  IK  V  ++R+ T        + +V+A+D   G   TGT  +RV D ND  P
Sbjct  1723  FEIEADTGVIKTAVCCLDRERT----PDYSIQVVAMDG-GGLKGTGTASIRVKDINDMPP  1777

 Score = 75.9 bits (185),  Expect = 2e-12, Method: Compositional matrix adjust.
 Identities = 108/429 (25%), Positives = 178/429 (41%), Gaps = 53/429 (12%)

Query  53    VDKNTGDINITAIVDREETPSFLITCRALNAQGLDVEKPLILTVKILDINDNPPVFSQ-Q  111
             +D++  ++N+TA+ D          +C  +N           ++ V I D+NDN PVF
Sbjct  950   LDRDKYELNVTAMDDG--------SC-CVNGDATIHTSTAVVVVFITDVNDNKPVFKDCG  1000

Query  112   IFMGEIEENSASNSLVMILNATDADEPNHLNSKIAFKIVSQEPAGTPMFLLSRNTGEVRT  171
              +  ++EE + + S V+ + ATD D+   +N ++ + IV Q        F +   TGEV T
Sbjct  1001  TYYPKVEEGAPNGSPVIKVQATDEDK--GVNGQVKYSIVQQPNQKGTKFTVDEETGEVST  1058

Query  172   LTNSLDRE--QASSYRLVVSGADKDGEGLSTQCECNIKVKDVNDNFPMFRDSQYSARIEE  229
                  DRE         + V   D+     L   C    +++ DVNDN P+F    +Y   +++
Sbjct  1059  -NKVFDREGDDGKFVSVTVKATDQGEPSLEGVCSFTVEITDVNDNPPLFDRQKYVENVKQ  1117

Query  230   NI-LSSELLRFQVTDLDEEYTDNWLAVYFFT---SGNEGNWFEIQTDPRTNEGILKVVKA  285
             +  + + +LR  V+  DE+  +N   VY  +   S N+  +FEIQ  P +   +LK  K
Sbjct  1118  DASIGTNILR--VSASDEDADNNGAIVYTLSAPHSPNDLEYFEIQ--PESGWIVLK--KP  1171
```

```
Query  286   LD-YEQLQSVKLSIAVK----------NKAEFHQSVISRYRVQSTPV----TIQVINVRE  330
             LD    + SV+    +K          + A+    +    R    P+        I++RE
Sbjct  1172  LDGCYSVSSVRERYRLKVYASDRGDPPSTADVDVELDVVDRNNKPPIWDRNIYGPIHIRE  1231

Query  331   GIAFRPASKTFTVQKGISSKKLVDYILGTYQAIDEDTNKAASNVKYVMGRNDGGYLMIDS  390
              +             GI     V Y L         TNK    + Y+  R D G
Sbjct  1232  NVTVGTVVTAIKASSGIEGNPTVFYRL--MPGSTAQTNKL--HTFYLQQRPDNG-----D  1282

Query  391   KTAEIKFVKNMNRDSTFIVNKTITAEVLAIDEYTGKTSTGTVYVRVPDFNDNCPTAV-LE  449
               A+IK    ++ ++    N TI  E     +     S  TVY+++ D ND  P     E
Sbjct  1283  TWADIKVNHPLDFETIKEYNLTIRVENNGAQQL---ASEATVYIQLEDVNDEIPLFTERE  1339

Query  450   KDAVCSSSP  458
             ++ V    P
Sbjct  1340  QETVLEGEP  1348

 Score = 61.2 bits (147),  Expect = 5e-08, Method: Compositional matrix adjust.
 Identities = 87/366 (24%), Positives = 156/366 (43%), Gaps = 38/366 (10%)

Query  51    FVVDKNTGDINITAIVDRE-ETPSFL-ITCRALNAQGLDVEKPLILTVKILDINDNPPVF  108
             F VD+ TG+++   + DRE +   F+ +T +A +     +E     TV+I D+NDNPP+F
Sbjct  1047  FTVDEETGEVSTNKVFDREGDDGKFVSVTVKATDQGEPSLEGVCSFTVEITDVNDNPPLF  1106

Query  109   SQQIFMGEIEENSASNSLVMILNATDADEPNHLNSKIAFKIVS-QEPAGTPMFLLSRNTG  167
              +Q ++  ++++++  + ++ ++A+D D  N  N  I + + +   P     F +   +G
Sbjct  1107  DRQKYVENVKQDASIGTNILRVSASDEDADN--NGAIVYTLSAPHSPNDLEYFEIQPESG  1164

Query  168   EVRTLTNSLDREQASS-----YRLVVSGADKDGEGLSTQCECNIKVKDVNDNFPMFRDSQ  222
              +  L   LD   + S     YRL V  +D+ G+  ST            +N P   D
Sbjct  1165  WI-VLKKPLDGCYSVSSVRERYRLKVYASDR-GDPPSTADVDVELDVVDRNNKPPIWDRN  1222

Query  223   YSA--RIEENILSSELLRFQVTDLDEEYTDNWLAVYFFTSG-----NEGNWFEIQTDPRT  275
                   I EN+    ++             N    Y    G     N+ + F +Q  P
Sbjct  1223  IYGPIHIRENVTVGTVV--TAIKASSGIEGNPTVFYRLMPGSTAQTNKLHTFYLQQRPDN  1280

Query  276   NE--GILKVVKALDYEQLQSVKLSIAVKNKAEFHQSVISRYRVQSTPVTIQVINVREGIA  333
              +    +KV   LD+E ++   L+I V+N         ++       V IQ+ +V + I
Sbjct  1281  GDTWADIKVNHPLDFETIKEYNLTIRVENNG-------AQQLASEATVYIQLEDVNDEIP  1333

Query  334   FRPASKTFTVQKGISSKKLVDYILGTYQAIDEDTNKAASNVKYVM---GRNDG-GYLMID  389
                  +  TV +G      V  +     AID+D     + V Y +    RN+G  +  I+
Sbjct  1334  LFTEREQETVLEGEPIGTKVTQV----NAIDKDGTFPNNQVYYYIVDSPRNEGKDFFEIN  1389

Query  390   SKTAEI  395
              ++ EI
Sbjct  1390  LQSGEI  1395

 Score = 55.8 bits (133),  Expect = 3e-06, Method: Compositional matrix adjust.
 Identities = 49/158 (31%), Positives = 78/158 (49%), Gaps = 24/158 (15%)

Query  102   NDNPPVFSQQIFMGEIEENSASNSLVMILNATDADEPNHLNSKIAFKIVSQEPAGTP--M  159
             ND  P FSQQ++   ++EN+  N+LV  + A+D D  N     + F  V     GT
Sbjct  880   NDEEPKFSQQVYTPNVDENAGPNTLVTTVVASDKDGDN-----VRFGFVG---GGTSSGQ  931

Query  160   FLLSRNTGEVRTLTN--SLDREQASSYRLVVSGADK-------DGEGLSTQCECNIKVKD  210
             F++   TG +R  +   SLDR++    Y L V+  D        D    ++     + + D
Sbjct  932   FVIEEITGVIRLHSKAISLDRDK---YELNVTAMDDGSCCVNGDATIHTSTAVVVVFITD  988

Query  211   VNDNFPMFRD-SQYSARIEENILS-SELLRFQVTDLDE  246
             VNDN P+F+D   Y  ++EE   + S +++ Q TD D+
Sbjct  989   VNDNKPVFKDCGTYYPKVEEGAPNGSPVIKVQATDEDK  1026

 Score = 53.9 bits (128),  Expect = 1e-05, Method: Compositional matrix adjust.
 Identities = 70/306 (23%), Positives = 126/306 (41%), Gaps = 30/306 (10%)

Query  20    PIAKIT-SDYQATQKITYRI---SGVGIDQPPFGIFVVDKNTGDINITAIVDRE---ETP  72
             PI  +T  D   T K  Y++   SG G D+  F +   +  TG + I   +D E   ++
```

```
Sbjct  1800  PILTVTVHDEDETNKFQYKVIDNSGYGADK--FTMVRNNDGTGSLKIVQPLDYEDPLQSN  1857

Query  73    SFLITCRALNAQGLDVEKPLI------LTVKILDINDNPPVFSQQIFMGEIEENSASNSL  126
              F    + +N +G D +          + VK+ DINDN P F +      + EN+
Sbjct  1858  GFRFRIQ-VNDKGEDNDNDKYHVAYSWVVVKLRDINDNKPQFERPNIEVSVYENADVGKT  1916

Query  127   VMILNATDADEPNHLNSKIAFKIVSQEPAGTPMFLLSRNTGEVRTLTNSLDREQASSYRL  186
             +    ATD D+     SK+++ I   + +       S N     T+  +LDRE    +++
Sbjct  1917  LETFKATDPDQGG--KSKVSYAI---DRSSDRQRQFSINQEGTVTIQRNLDREVTPRHQV  1971

Query  187   VVSGADKDGEGLSTQCECNIKVKDVNDNFPMFRDSQYSARIEENILSSELLRFQVTDLDE  246
              +   D      +      + V+D+NDN P F    Y   + E++   +++    TD D+
Sbjct  1972  KILAIDDGIPPKTATATLTVIVQDINDNPPKFL-KDYRPVLPEHVPPRKVVEILATDDDD  2030

Query  247   EYTDNWLAVYFFTSGNEGNW----FEIQTDPRTNEG----ILKVVKALDYEQLQSVKLSI  298
                 N     F       +     F+++ D +   G    I+  +++ D EQ +   + I
Sbjct  2031  RSKSNGPPFQFRLDPGADDIIRASFKVEQDQKGANGDGMAIVSSLRSFDREQQKEYLIPI  2090

Query  299   AVKNKA  304
              +K+
Sbjct  2091  VIKDHG  2096

 Score = 51.6 bits (122),  Expect = 5e-05, Method: Compositional matrix adjust.
 Identities = 96/398 (24%), Positives = 163/398 (41%), Gaps = 54/398 (14%)

Query  106   PVFSQQIFMGEIEENSASNSLVMILNATDADEPNHLNSKIAFKIVSQ-EPAGTPMFLLSR  164
             P F    +  EI EN   +S ++ + A      +  + +I + + +Q + AGT  F +
Sbjct  677   PQFYMPSYEAEIPENQKKDSDIISVKAK-----SFADREIRYTLKAQGQGAGT--FNIGP  729

Query  165   NTGEVRTLTNSLDRE---QASSYRLVVSGADKDGEGLSTQCECNIKVKDVNDNFPMFRDS  221
              +G V+ L   LD E   Q   Y LVV+ A +D  G ST  E  I+V DVNDN P F
Sbjct  730   TSGIVK-LAKELDFEDLRQPHVYSLVVT-ATEDSGGFSTSVELTIRVTDVNDNAPKFELP  787

Query  222   QYSAR-IEENI-LSSELLRFQVTDLDEEYTDNWLAVYFFTSGNEGNWFEIQTDPRTNEGI  279
              Y A  ++E+I + + +L+ +  D D         + +  S +    F + ++     GI
Sbjct  788   DYQAHNVDEDIQIGTSILKVKAMDADSGAN---AEIEYLVSDDH---FAVDSN-----GI  836

Query  280   LKVVKALDYEQLQSV-KLSIAVKNKAEFHQSVISRYRVQSTPVTIQVINVREGIAFRPAS  338
             +   K LD +   +  +  +  K+K E  ++  +  RV +           E   F
Sbjct  837   IVNNKQLDADNNNAYYEFVVTAKDKGEPAKTGTATVRVYTKNKN------DEEPKFSQQV  890

Query  339   KTFTVQKGISSKKLVDYILGTYQAIDEDTNKAASNVKY--VMGRNDGGYLMIDSKTAEIK  396
              T  V +      LV  ++ +        +K   NV++  V G    G  +I+  T  I+
Sbjct  891   YTPNVDENAGPNTLVTTVVAS--------DKDGDNVRFGFVGGGTSSGQFVIEEITGVIR  942

Query  397   FVK---NMNRDSTFI-VNKTITAEVLAIDEYTGKTSTGTVYVRVPDFNDN------CPTA  446
                   +++RD   + V            + T  TST  V V + D NDN      C T
Sbjct  943   LHSKAISLDRDKYELNVTAMDDGSCCVNGDATIHTSTAVVVVFITDVNDNKPVFKDCGTY  1002

Query  447   VLE-KDAVCSSSPSVVVSARTLNNRYTGPYTFALEDQP  483
               + ++   + SP + V A   +    G   +++  QP
Sbjct  1003  YPKVEEGAPNGSPVIKVQATDEDKGVNGQVKYSIVQQP  1040

 Score = 44.7 bits (104),  Expect = 0.008, Method: Compositional matrix adjust.
 Identities = 48/199 (24%), Positives = 73/199 (37%), Gaps = 19/199 (10%)

Query  27    DYQATQKITYRISGVGIDQPPFGIFVVDKNTGDINITAIVDREETPSFLITCRALNAQGL  86
             D     K++Y I      Q  F I       G + I   +DRE TP   +   A++
Sbjct  1926  DQGGKSKVSYAIDRSSDRQRQFSI----NQEGTVTIQRNLDREVTPRHQVKILAIDDGIP  1981

Query  87    DVEKPLILTVKILDINDNPPVFSQQIFMGEIEENSASNSLVMILNATDADEPNHLNSKIA  146
                    LTV + DINDNPP F +  +   + E+     +V IL   D D
Sbjct  1982  PKTATATLTVIVQDINDNPPKFLKD-YRPVLPEHVPPRKVVEILATDDDDRSKSNGPPFQ  2040

Query  147   FKIVSQEPAGTPMFLLSRNTGEVRTLTN-----------SLDREQASSYRLVVSGADKDG  195
             F++   +P    +   S    + +   N           S DREQ   Y + +   D
Sbjct  2041  FRL---DPGADDIIRASFKVEQDQKGANGDGMAIVSSLRSFDREQQKEYLIPIVIKDHGN  2097

Query  196   EGLSTQCECNIKVKDVNDN  214
```

```
                  ++      + + DVNDN
Sbjct  2098  PAMTGTSTLTVVIGDVNDN  2116

 Score = 40.8 bits (94),  Expect = 0.11, Method: Compositional matrix adjust.
 Identities = 55/242 (23%), Positives = 102/242 (42%), Gaps = 22/242 (9%)

Query  4    KFAKPCREGE--DNSKRNP-IAKITSDYQATQKITYRISGVGIDQPPFGIFVVDKNTGDI  60
            +F  P  E E  +N K++  I  + +   A ++I Y +   G      G F +   +G +
Sbjct  678  QFYMPSYEAEIPENQKKDSDIISVKAKSFADREIRYTLKAQGQGA---GTFNIGPTSGIV  734

Query  61   NITAIVDREE--TPSF--LITCRALNAQGLDVEKPLILTVKILDINDNPPVFSQQIFMGE  116
             +   +D E+   P    L+     ++ G      + LT+++ D+NDN P F    +
Sbjct  735  KLAKELDFEDLRQPHVYSLVVTATEDSGGFSTS--VELTIRVTDVNDNAPKFELPDYQAH  792

Query  117  -IEENSASNSLVMILNATDADEPNHLNSKIAFKIVSQEPAGTPMFLLSRNTGEVRTLTNS  175
             ++E+    + ++ + A DAD  +  N++I + +VS +      F +  N   V
Sbjct  793  NVDEDIQIGTSILKVKAMDAD--SGANAEIEY-LVSDD-----HFAVDSNGIIVNNKQLD  844

Query  176  LDREQASSYRLVVSGADKDGEGLSTQCECNIKVKDVNDNFPMFRDSQYSARIEENILSSE  235
             D  A  Y  VV+  DK     +      +  K+ ND  P F    Y+  ++EN   +
Sbjct  845  ADNNNA-YYEFVVTAKDKGEPAKTGTATVRVYTKNKNDEEPKFSQQVYTPNVDENAGPNT  903

Query  236  LL  237
            L+
Sbjct  904  LV  905

 Score = 39.3 bits (90),  Expect = 0.30, Method: Compositional matrix adjust.
 Identities = 55/225 (24%), Positives = 88/225 (39%), Gaps = 24/225 (11%)

Query  75   LITCRALNAQGLDVEKPLILTVKILDINDNPPVFSQQIF--MGEIEENSASNSLVMILNA  132
            +IT +  NA GL       + + + D+ND PP F  +       ++ N+  N+ V  L A
Sbjct  539  IITNQGHNA-GLKYTDNQRVIIHVKDVNDEPPYFINRPLPMQAVVQLNAPPNTPVFTLQA  597

Query  133  TDADEPNHLNSKIAFKIVSQEPAGTPMFLLSRNTGEVRTLTNSLDREQASSYRLVVSGAD  192
             D D     +  I + IV     G   F +   +G VRT    L  +    Y L V   D
Sbjct  598  RDPD----TDHNIHYFIVRDRTGG--RFEVDERSGVVRTRGTDL-FQLDMEYVLYVKAED  650

Query  193  KDGE----GLSTQCECNIKVKDVNDNFPMFRDSQYSARIEEN-ILSSELLRFQVTDLDEE  247
            ++G+        +  E  + +       P F    Y A I EN    S+++  +     +
Sbjct  651  QNGKVDDRRFQSTPEERLSIVG-GKRAPQFYMPSYEAEIPENQKKDSDIISVKA----KS  705

Query  248  YTDNWLAVYFFTSGNEGNWFEIQTDPRTNEGILKVVKALDYEQLQ  292
            + D  +       G     F I         GI+K+ K LD+E L+
Sbjct  706  FADREIRYTLKAQGQGAGTFNIG----PTSGIVKLAKELDFEDLR  746

>AALB20_033171.P66508 | transcript=AALB20_033171.R66508 | gene=AALB20_033171
| organism=Anopheles_albimanus_STECLA_2020 |
gene_product=neural-cadherin, transcript variant X38 |
transcript_product=neural-cadherin,
transcript variant X11 | location=CM024292:75037560-75208393(+)
| protein_length=3131 | sequence_SO=chromosome
| SO=protein_coding_gene | is_pseudo=false
Length=3131

 Score = 144 bits (364),  Expect = 1e-34, Method: Compositional matrix adjust.
 Identities = 125/437 (29%), Positives = 211/437 (48%), Gaps = 33/437 (8%)

Query  26    SDYQATQKITYRISGVGID--QPPFGIFVVDKNTGDINITAIVDREET---PSFLITCRA  80
               D    Q I Y ++G GID   P    F +++ TG+I +   +DR++    P +  T  A
Sbjct  1580  GDKDRPQNIVYFLTGQGIDPDNPANSKFDINRTTGEIFVLKPLDRDQPNGRPQWRFTVFA  1639

Query  81    LNAQGLDVEKPLILTVKILDINDNPPVFSQQIFMGEIEENSASNSLVMILNATDADEPNH  140
               +  G  +      + V + DINDN P+F Q ++ G + EN  +  +VM + A D D+PN
Sbjct  1640  QDEGGEGLVGYADVQVNLKDINDNAPIFPQGVYFGNVTENGTAGMVVMTMTAVDYDDPNE  1699

Query  141   -LNSKIAFKI---VSQEPAGTPMFLLSRNTGEVRTLTNSLDREQASSYRLVVSGADKDGE  196
                N+K+ + I   V +E  G+P+F +  +TG ++T    LDRE+   Y + V      DG
Sbjct  1700  GTNAKLIYSIEKNVIEEETGSPIFEIEADTGVIKTAVCCLDRERTPDYSIQVVAM--DGG  1757
```

```
Query  197   GLSTQCECNIKVKDVNDNFPMFRDSQYSARIEENILSS--ELLRFQVTDLDEEYTDNWLA  254
             GL      +I+VKD+ND  P F   ++    ++E   ++  E+    VT  DE+ T+ +
Sbjct  1758  GLKGTGTASIRVKDINDMPPQFTKDEWFTEVDETDGTNLPEMPILTVTVHDEDETNKFQY  1817

Query  255   VYFFTSGNEGNWFEIQTDPRTNEGI--LKVVKALDYE---QLQSVKLSIAVKNKAEFHQS  309
                  SG   + F +    R N+G   LK+V+ LDYE   Q    +  I V +K E + +
Sbjct  1818  KVIDNSGYGADKFTMV---RNNDGTGSLKIVQPLDYEDPLQSNGFRFRIQVNDKGEDNDN  1874

Query  310   VISRYRVQSTPVTIQVINVREGIAFRPASKTFTVQKGISSKKLVDYILGTYQAIDEDTNK  369
                +Y V  + V +++ ++ +    +P  +   ++  +     V   L T++A D D
Sbjct  1875  --DKYHVAYSWVVVKLRDINDN---KPQFERPNIEVSVYENADVGKTLETFKATDPDQG-  1928

Query  370   AASNVKYVMGR-NDGGYLMIDSKTAEIKFVKNMNRDSTFIVNKTITAEVLAIDE-YTGKT  427
               S V Y + R +D       ++    +   +N++R+    V       ++LAID+    KT
Sbjct  1929  GKSKVSYAIDRSSDRQRQFSINQEGTVTIQRNLDRE----VTPRHQVKILAIDDGIPPKT  1984

Query  428   STGTVYVRVPDFNDNCP  444
             +T T+ V V D NDN P
Sbjct  1985  ATATLTVIVQDINDNPP  2001

 Score = 85.9 bits (211),  Expect = 1e-15, Method: Compositional matrix adjust.
 Identities = 108/420 (26%), Positives = 177/420 (42%), Gaps = 52/420 (12%)

Query  50    IFVVDKNTGDINITAIVDREETPSFLITCRAL----------NAQGLDVEKPLILTVKIL  99
              F ++  +G+I    + DRE+  ++ +   A            N Q   V K   + + I
Sbjct  1385  FFEINLQSGEIFTKVVFDREKQGAYALEVEARDGAPSARPNSNQQPNSVTK--FIRIGIA  1442

Query  100   DINDNPPVFSQQIFMGEIEENSASNSLVMILNATDADEPNHLNSKIAFKIVSQEPAGTPM  159
             D NDNPP F + ++  E++EN      V+ + A D DE    +S+I ++I S    G
Sbjct  1443  DKNDNPPYFDKALYEAEVDENEDIQHTVLTVTAKDHDE----SSRIRYEITSGNIGGA--  1496

Query  160   FLLSRNTGEVRTLTNSLDREQASSYRLVVSGADKDGEGLSTQCECNIKVKDVNDNFPMFR  219
             F +   TG +  +  +LD E    Y L ++ +D   E  +T      I VKDVNDN P+F
Sbjct  1497  FAVKNMTGAI-YVAGALDYETRKRYELRLAASDNLKENYTTVV---IHVKDVNDNPPVFE  1552

Query  220   DSQYSARIEENI---LSSELLRFQVTDLDEEYTDNWLAVYFFTSG--NEGNWFEIQTDPR  274
                Y  +I E     L   +L+   TD D++   N   VYF T    +  N    + D
Sbjct  1553  RPTYRTQITEEDDRNLPKRVLQVTATDGDKDRPQN--IVYFLTGQGIDPDNPANSKFDIN  1610

Query  275   TNEGILKVVKALDYEQLQS-VKLSIAVKNKAEFHQSVISRYRVQSTPVTIQVINVREGIA  333
                G + V+K LD +Q     +    V  + E  + ++    VQ     + + ++ +
Sbjct  1611  RTTGEIFVLKPLDRDQPNGRPQWRFTVFAQDEGGEGLVGYADVQ-----VNLKDINDNAP  1665

Query  334   FRPASKTFTVQKGISSKKLVDYILGTYQAIDEDTNKAASNVK--YVMGRN------DGGY  385
               P    F     ++        ++ T  A+D D     +N K  Y + +N
Sbjct  1666  IFPQGVYF---GNVTENGTAGMVVMTMTAVDYDDPNEGTNAKLIYSIEKNVIEEETGSPI  1722

Query  386   LMIDSKTAEIK-FVKNMNRDSTFIVNKTITAEVLAIDEYTGKTSTGTVYVRVPDFNDNCP  444
               I++ T  IK  V  ++R+ T        + +V+A+D   G   TGT  +RV D ND  P
Sbjct  1723  FEIEADTGVIKTAVCCLDRERT----PDYSIQVVAMDG-GGLKGTGTASIRVKDINDMPP  1777

 Score = 75.9 bits (185),  Expect = 2e-12, Method: Compositional matrix adjust.
 Identities = 108/429 (25%), Positives = 178/429 (41%), Gaps = 53/429 (12%)

Query  53    VDKNTGDINITAIVDREETPSFLITCRALNAQGLDVEKPLILTVKILDINDNPPVFSQ-Q  111
             +D++  ++N+TA+ D          +C  +N           ++ V I D+NDN PVF
Sbjct  950   LDRDKYELNVTAMDDG--------SC-CVNGDATIHTSTAVVVVFITDVNDNKPVFKDCG  1000

Query  112   IFMGEIEENSASNSLVMILNATDADEPNHLNSKIAFKIVSQEPAGTPMFLLSRNTGEVRT  171
              +  ++EE + + S V+ + ATD D+   +N ++ + IV Q         F +   TGEV T
Sbjct  1001  TYYPKVEEGAPNGSPVIKVQATDEDK--GVNGQVKYSIVQQPNQKGTKFTVDEETGEVST  1058

Query  172   LTNSLDRE--QASSYRLVVSGADKDGEGLSTQCECNIKVKDVNDNFPMFRDSQYSARIEE  229
                  DRE          + V   D+     L   C    +++ DVNDN P+F    +Y   +++
Sbjct  1059  -NKVFDREGDDGKFVSVTVKATDQGEPSLEGVCSFTVEITDVNDNPPLFDRQKYVENVKQ  1117

Query  230   NI-LSSELLRFQVTDLDEEYTDNWLAVYFFT---SGNEGNWFEIQTDPRTNEGILKVVKA  285
             +  + + +LR  V+  DE+  +N    VY  +    S N+  +FEIQ  P +    +LK  K
```

```
Sbjct  1118  DASIGTNILR--VSASDEDADNNGAIVYTLSAPHSPNDLEYFEIQ--PESGWIVLK--KP  1171

Query  286   LD-YEQLQSVKLSIAVK----------NKAEFHQSVISRYRVQSTPV----TIQVINVRE  330
             LD    + SV+    +K          + A+    +    R    P+        I++RE
Sbjct  1172  LDGCYSVSSVRERYRLKVYASDRGDPPSTADVDVELDVVDRNNKPPIWDRNIYGPIHIRE  1231

Query  331   GIAFRPASKTFTVQKGISSKKLVDYILGTYQAIDEDTNKAASNVKYVMGRNDGGYLMIDS  390
              +             GI     V Y L         TNK   +  Y+  R D G
Sbjct  1232  NVTVGTVVTAIKASSGIEGNPTVFYRL--MPGSTAQTNKL--HTFYLQQRPDNG-----D  1282

Query  391   KTAEIKFVKNMNRDSTFIVNKTITAEVLAIDEYTGKTSTGTVYVRVPDFNDNCPTAV-LE  449
               A+IK    ++ ++    N TI  E     +     S  TVY+++ D ND  P     E
Sbjct  1283  TWADIKVNHPLDFETIKEYNLTIRVENNGAQQL---ASEATVYIQLEDVNDEIPLFTERE  1339

Query  450   KDAVCSSSP  458
             ++ V    P
Sbjct  1340  QETVLEGEP  1348

 Score = 61.2 bits (147),  Expect = 5e-08, Method: Compositional matrix adjust.
 Identities = 87/366 (24%), Positives = 156/366 (43%), Gaps = 38/366 (10%)

Query  51    FVVDKNTGDINITAIVDRE-ETPSFL-ITCRALNAQGLDVEKPLILTVKILDINDNPPVF  108
             F VD+ TG+++   + DRE +   F+ +T +A +     +E     TV+I D+NDNPP+F
Sbjct  1047  FTVDEETGEVSTNKVFDREGDDGKFVSVTVKATDQGEPSLEGVCSFTVEITDVNDNPPLF  1106

Query  109   SQQIFMGEIEENSASNSLVMILNATDADEPNHLNSKIAFKIVS-QEPAGTPMFLLSRNTG  167
              +Q ++  ++++++  + ++ ++A+D D  N  N  I + + +   P     F +   +G
Sbjct  1107  DRQKYVENVKQDASIGTNILRVSASDEDADN--NGAIVYTLSAPHSPNDLEYFEIQPESG  1164

Query  168   EVRTLTNSLDREQASS-----YRLVVSGADKDGEGLSTQCECNIKVKDVNDNFPMFRDSQ  222
              +  L   LD   + S     YRL V  +D+ G+  ST            +N P   D
Sbjct  1165  WI-VLKKPLDGCYSVSSVRERYRLKVYASDR-GDPPSTADVDVELDVVDRNNKPPIWDRN  1222

Query  223   YSA--RIEENILSSELLRFQVTDLDEEYTDNWLAVYFFTSG-----NEGNWFEIQTDPRT  275
                   I EN+    ++             N    Y    G     N+ + F +Q  P
Sbjct  1223  IYGPIHIRENVTVGTVV--TAIKASSGIEGNPTVFYRLMPGSTAQTNKLHTFYLQQRPDN  1280

Query  276   NE--GILKVVKALDYEQLQSVKLSIAVKNKAEFHQSVISRYRVQSTPVTIQVINVREGIA  333
              +    +KV   LD+E ++   L+I V+N         ++       V IQ+ +V + I
Sbjct  1281  GDTWADIKVNHPLDFETIKEYNLTIRVENNG-------AQQLASEATVYIQLEDVNDEIP  1333

Query  334   FRPASKTFTVQKGISSKKLVDYILGTYQAIDEDTNKAASNVKYVM---GRNDG-GYLMID  389
                  +  TV +G      V  +     AID+D     + V Y +    RN+G  +  I+
Sbjct  1334  LFTEREQETVLEGEPIGTKVTQV----NAIDKDGTFPNNQVYYYIVDSPRNEGKDFFEIN  1389

Query  390   SKTAEI  395
              ++ EI
Sbjct  1390  LQSGEI  1395

 Score = 55.8 bits (133),  Expect = 3e-06, Method: Compositional matrix adjust.
 Identities = 49/158 (31%), Positives = 78/158 (49%), Gaps = 24/158 (15%)

Query  102   NDNPPVFSQQIFMGEIEENSASNSLVMILNATDADEPNHLNSKIAFKIVSQEPAGTP--M  159
             ND  P FSQQ++   ++EN+  N+LV  + A+D D  N     + F  V     GT
Sbjct  880   NDEEPKFSQQVYTPNVDENAGPNTLVTTVVASDKDGDN-----VRFGFVG---GGTSSGQ  931

Query  160   FLLSRNTGEVRTLTN--SLDREQASSYRLVVSGADK-------DGEGLSTQCECNIKVKD  210
             F++   TG +R  +   SLDR++   Y L V+  D        D    ++     + + D
Sbjct  932   FVIEEITGVIRLHSKAISLDRDK---YELNVTAMDDGSCCVNGDATIHTSTAVVVVFITD  988

Query  211   VNDNFPMFRD-SQYSARIEENILS-SELLRFQVTDLDE  246
             VNDN P+F+D   Y  ++EE   + S +++ Q TD D+
Sbjct  989   VNDNKPVFKDCGTYYPKVEEGAPNGSPVIKVQATDEDK  1026

 Score = 53.9 bits (128),  Expect = 1e-05, Method: Compositional matrix adjust.
 Identities = 70/306 (23%), Positives = 126/306 (41%), Gaps = 30/306 (10%)

Query  20    PIAKIT-SDYQATQKITYRI---SGVGIDQPPFGIFVVDKNTGDINITAIVDRE---ETP  72
```

```
             PI  +T  D   T K  Y++   SG G D+  F +   +  TG + I   +D E   ++
Sbjct  1800  PILTVTVHDEDETNKFQYKVIDNSGYGADK--FTMVRNNDGTGSLKIVQPLDYEDPLQSN  1857

Query  73    SFLITCRALNAQGLDVEKPLI------LTVKILDINDNPPVFSQQIFMGEIEENSASNSL  126
              F    + +N +G D +          + VK+ DINDN P F +      + EN+
Sbjct  1858  GFRFRIQ-VNDKGEDNDNDKYHVAYSWVVVKLRDINDNKPQFERPNIEVSVYENADVGKT  1916

Query  127   VMILNATDADEPNHLNSKIAFKIVSQEPAGTPMFLLSRNTGEVRTLTNSLDREQASSYRL  186
             +    ATD D+     SK+++ I    + +      S N     T+  +LDRE    +++
Sbjct  1917  LETFKATDPDQGG--KSKVSYAI---DRSSDRQRQFSINQEGTVTIQRNLDREVTPRHQV  1971

Query  187   VVSGADKDGEGLSTQCECNIKVKDVNDNFPMFRDSQYSARIEENILSSELLRFQVTDLDE  246
              +   D      +      + V+D+NDN P F    Y   + E++   +++    TD D+
Sbjct  1972  KILAIDDGIPPKTATATLTVIVQDINDNPPKFL-KDYRPVLPEHVPPRKVVEILATDDDD  2030

Query  247   EYTDNWLAVYFFTSGNEGNW----FEIQTDPRTNEG----ILKVVKALDYEQLQSVKLSI  298
                 N     F       +     F+++ D +   G    I+  +++ D EQ +   + I
Sbjct  2031  RSKSNGPPFQFRLDPGADDIIRASFKVEQDQKGANGDGMAIVSSLRSFDREQQKEYLIPI  2090

Query  299   AVKNKA  304
              +K+
Sbjct  2091  VIKDHG  2096

 Score = 51.6 bits (122),  Expect = 5e-05, Method: Compositional matrix adjust.
 Identities = 96/398 (24%), Positives = 163/398 (41%), Gaps = 54/398 (14%)

Query  106   PVFSQQIFMGEIEENSASNSLVMILNATDADEPNHLNSKIAFKIVSQ-EPAGTPMFLLSR  164
             P F   +   EI EN   +S ++ + A      +  + +I + + +Q + AGT  F +
Sbjct  677   PQFYMPSYEAEIPENQKKDSDIISVKAK-----SFADREIRYTLKAQGQGAGT--FNIGP  729

Query  165   NTGEVRTLTNSLDRE---QASSYRLVVSGADKDGEGLSTQCECNIKVKDVNDNFPMFRDS  221
              +G V+ L   LD E   Q   Y LVV+ A +D  G ST  E  I+V DVNDN P F
Sbjct  730   TSGIVK-LAKELDFEDLRQPHVYSLVVT-ATEDSGGFSTSVELTIRVTDVNDNAPKFELP  787

Query  222   QYSAR-IEENI-LSSELLRFQVTDLDEEYTDNWLAVYFFTSGNEGNWFEIQTDPRTNEGI  279
              Y A  ++E+I + + +L+ +  D D         + +  S +    F + ++     GI
Sbjct  788   DYQAHNVDEDIQIGTSILKVKAMDADSGAN---AEIEYLVSDDH---FAVDSN-----GI  836

Query  280   LKVVKALDYEQLQSV-KLSIAVKNKAEFHQSVISRYRVQSTPVTIQVINVREGIAFRPAS  338
             +   K LD +   +  +  +  K+K E  ++  +  RV +           E   F
Sbjct  837   IVNNKQLDADNNNAYYEFVVTAKDKGEPAKTGTATVRVYTKNKN------DEEPKFSQQV  890

Query  339   KTFTVQKGISSKKLVDYILGTYQAIDEDTNKAASNVKY--VMGRNDGGYLMIDSKTAEIK  396
              T  V +      LV  ++ +        +K   NV++  V G    G  +I+  T  I+
Sbjct  891   YTPNVDENAGPNTLVTTVVAS--------DKDGDNVRFGFVGGGTSSGQFVIEEITGVIR  942

Query  397   FVK---NMNRDSTFI-VNKTITAEVLAIDEYTGKTSTGTVYVRVPDFNDN------CPTA  446
                   +++RD   + V            + T  TST  V V + D NDN      C T
Sbjct  943   LHSKAISLDRDKYELNVTAMDDGSCCVNGDATIHTSTAVVVVFITDVNDNKPVFKDCGTY  1002

Query  447   VLE-KDAVCSSSPSVVVSARTLNNRYTGPYTFALEDQP  483
               + ++   + SP + V A   +    G   +++  QP
Sbjct  1003  YPKVEEGAPNGSPVIKVQATDEDKGVNGQVKYSIVQQP  1040

 Score = 44.7 bits (104),  Expect = 0.008, Method: Compositional matrix adjust.
 Identities = 48/199 (24%), Positives = 73/199 (37%), Gaps = 19/199 (10%)

Query  27    DYQATQKITYRISGVGIDQPPFGIFVVDKNTGDINITAIVDREETPSFLITCRALNAQGL  86
             D     K++Y I      Q  F I       G + I   +DRE TP   +   A++
Sbjct  1926  DQGGKSKVSYAIDRSSDRQRQFSI----NQEGTVTIQRNLDREVTPRHQVKILAIDDGIP  1981

Query  87    DVEKPLILTVKILDINDNPPVFSQQIFMGEIEENSASNSLVMILNATDADEPNHLNSKIA  146
                    LTV + DINDNPP F +  +   + E+     +V IL   D D
Sbjct  1982  PKTATATLTVIVQDINDNPPKFLKD-YRPVLPEHVPPRKVVEILATDDDDRSKSNGPPFQ  2040

Query  147   FKIVSQEPAGTPMFLLSRNTGEVRTLTN-----------SLDREQASSYRLVVSGADKDG  195
             F++   +P    +   S    + +   N           S DREQ   Y + +   D
Sbjct  2041  FRL---DPGADDIIRASFKVEQDQKGANGDGMAIVSSLRSFDREQQKEYLIPIVIKDHGN  2097
```

```
Query  196   EGLSTQCECNIKVKDVNDN  214
               ++       + + DVNDN
Sbjct  2098  PAMTGTSTLTVVIGDVNDN  2116

 Score = 40.8 bits (94),  Expect = 0.11, Method: Compositional matrix adjust.
 Identities = 55/242 (23%), Positives = 102/242 (42%), Gaps = 22/242 (9%)

Query  4     KFAKPCREGE--DNSKRNP-IAKITSDYQATQKITYRISGVGIDQPPFGIFVVDKNTGDI  60
             +F  P  E E  +N K++  I  + +   A ++I Y +   G      G F +   +G +
Sbjct  678   QFYMPSYEAEIPENQKKDSDIISVKAKSFADREIRYTLKAQGQGA---GTFNIGPTSGIV  734

Query  61    NITAIVDREE--TPSF--LITCRALNAQGLDVEKPLILTVKILDINDNPPVFSQQIFMGE  116
              +   +D E+   P    L+     ++ G       + LT+++ D+NDN P F    +
Sbjct  735   KLAKELDFEDLRQPHVYSLVVTATEDSGGFSTS--VELTIRVTDVNDNAPKFELPDYQAH  792

Query  117   -IEENSASNSLVMILNATDADEPNHLNSKIAFKIVSQEPAGTPMFLLSRNTGEVRTLTNS  175
              ++E+    + ++ + A DAD  +  N++I + +VS +      F +  N   V
Sbjct  793   NVDEDIQIGTSILKVKAMDAD--SGANAEIEY-LVSDD-----HFAVDSNGIIVNNKQLD  844

Query  176   LDREQASSYRLVVSGADKDGEGLSTQCECNIKVKDVNDNFPMFRDSQYSARIEENILSSE  235
              D   A  Y  VV+  DK      +       +  K+ ND  P F    Y+  ++EN   +
Sbjct  845   ADNNNA-YYEFVVTAKDKGEPAKTGTATVRVYTKNKNDEEPKFSQQVYTPNVDENAGPNT  903

Query  236   LL  237
             L+
Sbjct  904   LV  905

 Score = 39.3 bits (90),  Expect = 0.30, Method: Compositional matrix adjust.
 Identities = 55/225 (24%), Positives = 88/225 (39%), Gaps = 24/225 (11%)

Query  75    LITCRALNAQGLDVEKPLILTVKILDINDNPPVFSQQIF--MGEIEENSASNSLVMILNA  132
             +IT +  NA GL        + + + D+ND PP F  +        ++ N+  N+ V  L A
Sbjct  539   IITNQGHNA-GLKYTDNQRVIIHVKDVNDEPPYFINRPLPMQAVVQLNAPPNTPVFTLQA  597

Query  133   TDADEPNHLNSKIAFKIVSQEPAGTPMFLLSRNTGEVRTLTNSLDREQASSYRLVVSGAD  192
              D D     +  I + IV     G   F +    +G VRT    L  +    Y L V   D
Sbjct  598   RDPD----TDHNIHYFIVRDRTGG--RFEVDERSGVVRTRGTDL-FQLDMEYVLYVKAED  650

Query  193   KDGE----GLSTQCECNIKVKDVNDNFPMFRDSQYSARIEEN-ILSSELLRFQVTDLDEE  247
             ++G+        +  E  + +        P F    Y A I EN    S+++  +     +
Sbjct  651   QNGKVDDRRFQSTPEERLSIVG-GKRAPQFYMPSYEAEIPENQKKDSDIISVKA----KS  705

Query  248   YTDNWLAVYFFTSGNEGNWFEIQTDPRTNEGILKVVKALDYEQLQ  292
             + D  +        G     F I        GI+K+ K LD+E L+
Sbjct  706   FADREIRYTLKAQGQGAGTFNIG----PTSGIVKLAKELDFEDLR  746

>AALB20_033171.P66507 | transcript=AALB20_033171.R66507 | gene=AALB20_033171
| organism=Anopheles_albimanus_STECLA_2020 |
gene_product=neural-cadherin, transcript variant X38 |
transcript_product=neural-cadherin,
transcript variant X9 | location=CM024292:75037560-75208393(+)
| protein_length=3131 | sequence_SO=chromosome
| SO=protein_coding_gene | is_pseudo=false
Length=3131

 Score = 144 bits (364),  Expect = 1e-34, Method: Compositional matrix adjust.
 Identities = 125/437 (29%), Positives = 211/437 (48%), Gaps = 33/437 (8%)

Query  26     SDYQATQKITYRISGVGID--QPPFGIFVVDKNTGDINITAIVDREET---PSFLITCRA  80
               D    Q I Y ++G GID    P    F +++ TG+I +    +DR++     P +  T  A
Sbjct  1580   GDKDRPQNIVYFLTGQGIDPDNPANSKFDINRTTGEIFVLKPLDRDQPNGRPQWRFTVFA  1639

Query  81     LNAQGLDVEKPLILTVKILDINDNPPVFSQQIFMGEIEENSASNSLVMILNATDADEPNH  140
                +  G  +      + V + DINDN P+F Q ++ G + EN  +  +VM + A D D+PN
Sbjct  1640   QDEGGEGLVGYADVQVNLKDINDNAPIFPQGVYFGNVTENGTAGMVVMTMTAVDYDDPNE  1699

Query  141    -LNSKIAFKI---VSQEPAGTPMFLLSRNTGEVRTLTNSLDREQASSYRLVVSGADKDGE  196
                 N+K+ + I    V +E  G+P+F +  +TG ++T     LDRE+    Y + V      DG
```

```
Sbjct  1700  GTNAKLIYSIEKNVIEEETGSPIFEIEADTGVIKTAVCCLDRERTPDYSIQVVAM--DGG  1757

Query  197   GLSTQCECNIKVKDVNDNFPMFRDSQYSARIEENILSS--ELLRFQVTDLDEEYTDNWLA  254
             GL      +I+VKD+ND  P F   ++   ++E   ++  E+    VT  DE+ T+   +
Sbjct  1758  GLKGTGTASIRVKDINDMPPQFTKDEWFTEVDETDGTNLPEMPILTVTVHDEDETNKFQY  1817

Query  255   VYFFTSGNEGNWFEIQTDPRTNEGI--LKVVKALDYE---QLQSVKLSIAVKNKAEFHQS  309
                  SG   + F +    R N+G   LK+V+ LDYE   Q    +  I V +K E + +
Sbjct  1818  KVIDNSGYGADKFTMV---RNNDGTGSLKIVQPLDYEDPLQSNGFRFRIQVNDKGEDNDN  1874

Query  310   VISRYRVQSTPVTIQVINVREGIAFRPASKTFTVQKGISSKKLVDYILGTYQAIDEDTNK  369
                +Y V  + V +++ ++ +    +P  +   ++  +     V   L T++A D D
Sbjct  1875  --DKYHVAYSWVVVKLRDINDN---KPQFERPNIEVSVYENADVGKTLETFKATDPDQG-  1928

Query  370   AASNVKYVMGR-NDGGYLMIDSKTAEIKFVKNMNRDSTFIVNKTITAEVLAIDE-YTGKT  427
               S V Y + R +D       ++   +   +N++R+    V      ++LAID+    KT
Sbjct  1929  GKSKVSYAIDRSSDRQRQFSINQEGTVTIQRNLDRE----VTPRHQVKILAIDDGIPPKT  1984

Query  428   STGTVYVRVPDFNDNCP  444
             +T T+ V V D NDN P
Sbjct  1985  ATATLTVIVQDINDNPP  2001

 Score = 85.9 bits (211),  Expect = 1e-15, Method: Compositional matrix adjust.
 Identities = 108/420 (26%), Positives = 177/420 (42%), Gaps = 52/420 (12%)

Query  50    IFVVDKNTGDINITAIVDREETPSFLITCRAL----------NAQGLDVEKPLILTVKIL  99
              F ++  +G+I    + DRE+  ++ +   A           N Q   V K   + + I
Sbjct  1385  FFEINLQSGEIFTKVVFDREKQGAYALEVEARDGAPSARPNSNQQPNSVTK--FIRIGIA  1442

Query  100   DINDNPPVFSQQIFMGEIEENSASNSLVMILNATDADEPNHLNSKIAFKIVSQEPAGTPM  159
             D NDNPP F + ++  E++EN      V+ + A D DE    +S+I ++I S    G
Sbjct  1443  DKNDNPPYFDKALYEAEVDENEDIQHTVLTVTAKDHDE----SSRIRYEITSGNIGGA--  1496

Query  160   FLLSRNTGEVRTLTNSLDREQASSYRLVVSGADKDGEGLSTQCECNIKVKDVNDNFPMFR  219
             F +   TG +  +  +LD E    Y L ++ +D   E  +T     I VKDVNDN P+F
Sbjct  1497  FAVKNMTGAI-YVAGALDYETRKRYELRLAASDNLKENYTTVV---IHVKDVNDNPPVFE  1552

Query  220   DSQYSARIEENI---LSSELLRFQVTDLDEEYTDNWLAVYFFTSG--NEGNWFEIQTDPR  274
                Y  +I E     L   +L+   TD D++   N   VYF T    +  N    + D
Sbjct  1553  RPTYRTQITEEDDRNLPKRVLQVTATDGDKDRPQN--IVYFLTGQGIDPDNPANSKFDIN  1610

Query  275   TNEGILKVVKALDYEQLQS-VKLSIAVKNKAEFHQSVISRYRVQSTPVTIQVINVREGIA  333
                G + V+K LD +Q     +    V  + E  + ++    VQ     + + ++ +
Sbjct  1611  RTTGEIFVLKPLDRDQPNGRPQWRFTVFAQDEGGEGLVGYADVQ-----VNLKDINDNAP  1665

Query  334   FRPASKTFTVQKGISSKKLVDYILGTYQAIDEDTNKAASNVK--YVMGRN------DGGY  385
              P    F     ++       ++ T  A+D D     +N K  Y + +N
Sbjct  1666  IFPQGVYF---GNVTENGTAGMVVMTMTAVDYDDPNEGTNAKLIYSIEKNVIEEETGSPI  1722

Query  386   LMIDSKTAEIK-FVKNMNRDSTFIVNKTITAEVLAIDEYTGKTSTGTVYVRVPDFNDNCP  444
               I++ T  IK  V  ++R+ T       + +V+A+D   G   TGT  +RV D ND  P
Sbjct  1723  FEIEADTGVIKTAVCCLDRERT----PDYSIQVVAMDG-GGLKGTGTASIRVKDINDMPP  1777

 Score = 75.9 bits (185),  Expect = 2e-12, Method: Compositional matrix adjust.
 Identities = 108/429 (25%), Positives = 178/429 (41%), Gaps = 53/429 (12%)

Query  53    VDKNTGDINITAIVDREETPSFLITCRALNAQGLDVEKPLILTVKILDINDNPPVFSQ-Q  111
             +D++  ++N+TA+ D         +C  +N           ++ V I D+NDN PVF
Sbjct  950   LDRDKYELNVTAMDDG--------SC-CVNGDATIHTSTAVVVVFITDVNDNKPVFKDCG  1000

Query  112   IFMGEIEENSASNSLVMILNATDADEPNHLNSKIAFKIVSQEPAGTPMFLLSRNTGEVRT  171
              +  ++EE + + S V+ + ATD D+   +N ++ + IV Q       F +   TGEV T
Sbjct  1001  TYYPKVEEGAPNGSPVIKVQATDEDK--GVNGQVKYSIVQQPNQKGTKFTVDEETGEVST  1058

Query  172   LTNSLDRE--QASSYRLVVSGADKDGEGLSTQCECNIKVKDVNDNFPMFRDSQYSARIEE  229
                  DRE        + V   D+    L   C   +++ DVNDN P+F   +Y   +++
Sbjct  1059  -NKVFDREGDDGKFVSVTVKATDQGEPSLEGVCSFTVEITDVNDNPPLFDRQKYVENVKQ  1117

Query  230   NI-LSSELLRFQVTDLDEEYTDNWLAVYFFT---SGNEGNWFEIQTDPRTNEGILKVVKA  285
```

```
              +  + + +LR  V+  DE+  +N   VY  +   S N+  +FEIQ  P +   +LK  K
Sbjct  1118  DASIGTNILR--VSASDEDADNNGAIVYTLSAPHSPNDLEYFEIQ--PESGWIVLK--KP  1171

Query  286   LD-YEQLQSVKLSIAVK----------NKAEFHQSVISRYRVQSTPV----TIQVINVRE  330
             LD    + SV+    +K          + A+    +    R    P+        I++RE
Sbjct  1172  LDGCYSVSSVRERYRLKVYASDRGDPPSTADVDVELDVVDRNNKPPIWDRNIYGPIHIRE  1231

Query  331   GIAFRPASKTFTVQKGISSKKLVDYILGTYQAIDEDTNKAASNVKYVMGRNDGGYLMIDS  390
              +             GI     V Y L         TNK   +  Y+  R D G
Sbjct  1232  NVTVGTVVTAIKASSGIEGNPTVFYRL--MPGSTAQTNKL--HTFYLQQRPDNG-----D  1282

Query  391   KTAEIKFVKNMNRDSTFIVNKTITAEVLAIDEYTGKTSTGTVYVRVPDFNDNCPTAV-LE  449
               A+IK    ++ ++    N TI  E     +     S  TVY+++ D ND  P     E
Sbjct  1283  TWADIKVNHPLDFETIKEYNLTIRVENNGAQQL---ASEATVYIQLEDVNDEIPLFTERE  1339

Query  450   KDAVCSSSP  458
             ++ V    P
Sbjct  1340  QETVLEGEP  1348

 Score = 61.2 bits (147),  Expect = 5e-08, Method: Compositional matrix adjust.
 Identities = 87/366 (24%), Positives = 156/366 (43%), Gaps = 38/366 (10%)

Query  51    FVVDKNTGDINITAIVDRE-ETPSFL-ITCRALNAQGLDVEKPLILTVKILDINDNPPVF  108
             F VD+ TG+++   + DRE +   F+ +T +A +     +E     TV+I D+NDNPP+F
Sbjct  1047  FTVDEETGEVSTNKVFDREGDDGKFVSVTVKATDQGEPSLEGVCSFTVEITDVNDNPPLF  1106

Query  109   SQQIFMGEIEENSASNSLVMILNATDADEPNHLNSKIAFKIVS-QEPAGTPMFLLSRNTG  167
              +Q ++  ++++++  + ++ ++A+D D  N  N  I + + +   P     F +   +G
Sbjct  1107  DRQKYVENVKQDASIGTNILRVSASDEDADN--NGAIVYTLSAPHSPNDLEYFEIQPESG  1164

Query  168   EVRTLTNSLDREQASS-----YRLVVSGADKDGEGLSTQCECNIKVKDVNDNFPMFRDSQ  222
              +  L   LD   + S     YRL V  +D+ G+  ST             +N P   D
Sbjct  1165  WI-VLKKPLDGCYSVSSVRERYRLKVYASDR-GDPPSTADVDVELDVVDRNNKPPIWDRN  1222

Query  223   YSA--RIEENILSSELLRFQVTDLDEEYTDNWLAVYFFTSG-----NEGNWFEIQTDPRT  275
                   I EN+    ++             N    Y    G     N+ + F +Q  P
Sbjct  1223  IYGPIHIRENVTVGTVV--TAIKASSGIEGNPTVFYRLMPGSTAQTNKLHTFYLQQRPDN  1280

Query  276   NE--GILKVVKALDYEQLQSVKLSIAVKNKAEFHQSVISRYRVQSTPVTIQVINVREGIA  333
              +    +KV   LD+E ++   L+I V+N          ++        V IQ+ +V + I
Sbjct  1281  GDTWADIKVNHPLDFETIKEYNLTIRVENNG-------AQQLASEATVYIQLEDVNDEIP  1333

Query  334   FRPASKTFTVQKGISSKKLVDYILGTYQAIDEDTNKAASNVKYVM---GRNDG-GYLMID  389
                  +  TV +G       V  +     AID+D     + V Y +    RN+G  +  I+
Sbjct  1334  LFTEREQETVLEGEPIGTKVTQV----NAIDKDGTFPNNQVYYYIVDSPRNEGKDFFEIN  1389

Query  390   SKTAEI  395
              ++ EI
Sbjct  1390  LQSGEI  1395

 Score = 55.8 bits (133),  Expect = 3e-06, Method: Compositional matrix adjust.
 Identities = 49/158 (31%), Positives = 78/158 (49%), Gaps = 24/158 (15%)

Query  102   NDNPPVFSQQIFMGEIEENSASNSLVMILNATDADEPNHLNSKIAFKIVSQEPAGTP--M  159
             ND  P FSQQ++   ++EN+  N+LV  + A+D D  N     + F  V     GT
Sbjct  880   NDEEPKFSQQVYTPNVDENAGPNTLVTTVVASDKDGDN-----VRFGFVG---GGTSSGQ  931

Query  160   FLLSRNTGEVRTLTN--SLDREQASSYRLVVSGADK-------DGEGLSTQCECNIKVKD  210
             F++   TG +R  +   SLDR++   Y L V+  D        D    ++     + + D
Sbjct  932   FVIEEITGVIRLHSKAISLDRDK---YELNVTAMDDGSCCVNGDATIHTSTAVVVVFITD  988

Query  211   VNDNFPMFRD-SQYSARIEENILS-SELLRFQVTDLDE  246
             VNDN P+F+D   Y  ++EE   + S +++ Q TD D+
Sbjct  989   VNDNKPVFKDCGTYYPKVEEGAPNGSPVIKVQATDEDK  1026

 Score = 53.9 bits (128),  Expect = 1e-05, Method: Compositional matrix adjust.
 Identities = 70/306 (23%), Positives = 126/306 (41%), Gaps = 30/306 (10%)
```

```
Query  20    PIAKIT-SDYQATQKITYRI---SGVGIDQPPFGIFVVDKNTGDINITAIVDRE---ETP  72
             PI  +T  D   T K  Y++   SG G D+  F +   +  TG + I   +D E   ++
Sbjct  1800  PILTVTVHDEDETNKFQYKVIDNSGYGADK--FTMVRNNDGTGSLKIVQPLDYEDPLQSN  1857

Query  73    SFLITCRALNAQGLDVEKPLI------LTVKILDINDNPPVFSQQIFMGEIEENSASNSL  126
              F    + +N +G D +          + VK+ DINDN P F +       + EN+
Sbjct  1858  GFRFRIQ-VNDKGEDNDNDKYHVAYSWVVVKLRDINDNKPQFERPNIEVSVYENADVGKT  1916

Query  127   VMILNATDADEPNHLNSKIAFKIVSQEPAGTPMFLLSRNTGEVRTLTNSLDREQASSYRL  186
             +    ATD D+     SK+++ I   + +       S N     T+  +LDRE    +++
Sbjct  1917  LETFKATDPDQGG--KSKVSYAI---DRSSDRQRQFSINQEGTVTIQRNLDREVTPRHQV  1971

Query  187   VVSGADKDGEGLSTQCECNIKVKDVNDNFPMFRDSQYSARIEENILSSELLRFQVTDLDE  246
              +   D      +      + V+D+NDN P F    Y   + E++   +++    TD D+
Sbjct  1972  KILAIDDGIPPKTATATLTVIVQDINDNPPKFL-KDYRPVLPEHVPPRKVVEILATDDDD  2030

Query  247   EYTDNWLAVYFFTSGNEGNW----FEIQTDPRTNEG----ILKVVKALDYEQLQSVKLSI  298
                 N     F       +     F+++ D +   G    I+  +++ D EQ +   + I
Sbjct  2031  RSKSNGPPFQFRLDPGADDIIRASFKVEQDQKGANGDGMAIVSSLRSFDREQQKEYLIPI  2090

Query  299   AVKNKA  304
              +K+
Sbjct  2091  VIKDHG  2096

 Score = 51.6 bits (122),  Expect = 5e-05, Method: Compositional matrix adjust.
 Identities = 96/398 (24%), Positives = 163/398 (41%), Gaps = 54/398 (14%)

Query  106   PVFSQQIFMGEIEENSASNSLVMILNATDADEPNHLNSKIAFKIVSQ-EPAGTPMFLLSR  164
             P F    +  EI EN   +S ++ + A      +  + +I + + +Q + AGT  F +
Sbjct  677   PQFYMPSYEAEIPENQKKDSDIISVKAK-----SFADREIRYTLKAQGQGAGT--FNIGP  729

Query  165   NTGEVRTLTNSLDRE---QASSYRLVVSGADKDGEGLSTQCECNIKVKDVNDNFPMFRDS  221
              +G V+ L   LD E   Q   Y LVV+ A +D  G ST  E  I+V DVNDN P F
Sbjct  730   TSGIVK-LAKELDFEDLRQPHVYSLVVT-ATEDSGGFSTSVELTIRVTDVNDNAPKFELP  787

Query  222   QYSAR-IEENI-LSSELLRFQVTDLDEEYTDNWLAVYFFTSGNEGNWFEIQTDPRTNEGI  279
              Y A  ++E+I + + +L+ + D D         + +  S +    F + ++     GI
Sbjct  788   DYQAHNVDEDIQIGTSILKVKAMDADSGAN---AEIEYLVSDDH---FAVDSN-----GI  836

Query  280   LKVVKALDYEQLQSV-KLSIAVKNKAEFHQSVISRYRVQSTPVTIQVINVREGIAFRPAS  338
             +   K LD +   +  +  +  K+K E  ++  +  RV +           E   F
Sbjct  837   IVNNKQLDADNNNAYYEFVVTAKDKGEPAKTGTATVRVYTKNKN------DEEPKFSQQV  890

Query  339   KTFTVQKGISSKKLVDYILGTYQAIDEDTNKAASNVKY--VMGRNDGGYLMIDSKTAEIK  396
              T  V +      LV  ++ +        +K   NV++  V G    G  +I+  T  I+
Sbjct  891   YTPNVDENAGPNTLVTTVVAS--------DKDGDNVRFGFVGGGTSSGQFVIEEITGVIR  942

Query  397   FVK---NMNRDSTFI-VNKTITAEVLAIDEYTGKTSTGTVYVRVPDFNDN------CPTA  446
                   +++RD   + V            + T  TST  V V + D NDN      C T
Sbjct  943   LHSKAISLDRDKYELNVTAMDDGSCCVNGDATIHTSTAVVVVFITDVNDNKPVFKDCGTY  1002

Query  447   VLE-KDAVCSSSPSVVVSARTLNNRYTGPYTFALEDQP  483
               + ++   + SP + V A   +    G   +++  QP
Sbjct  1003  YPKVEEGAPNGSPVIKVQATDEDKGVNGQVKYSIVQQP  1040

 Score = 44.7 bits (104),  Expect = 0.008, Method: Compositional matrix adjust.
 Identities = 48/199 (24%), Positives = 73/199 (37%), Gaps = 19/199 (10%)

Query  27    DYQATQKITYRISGVGIDQPPFGIFVVDKNTGDINITAIVDREETPSFLITCRALNAQGL  86
             D     K++Y I      Q  F I       G + I   +DRE TP   +   A++
Sbjct  1926  DQGGKSKVSYAIDRSSDRQRQFSI----NQEGTVTIQRNLDREVTPRHQVKILAIDDGIP  1981

Query  87    DVEKPLILTVKILDINDNPPVFSQQIFMGEIEENSASNSLVMILNATDADEPNHLNSKIA  146
                    LTV + DINDNPP F +  +   + E+     +V IL   D D
Sbjct  1982  PKTATATLTVIVQDINDNPPKFLKD-YRPVLPEHVPPRKVVEILATDDDDRSKSNGPPFQ  2040

Query  147   FKIVSQEPAGTPMFLLSRNTGEVRTLTN-----------SLDREQASSYRLVVSGADKDG  195
             F++   +P    +   S    + +   N           S DREQ   Y + +   D
Sbjct  2041  FRL---DPGADDIIRASFKVEQDQKGANGDGMAIVSSLRSFDREQQKEYLIPIVIKDHGN  2097
```

```

Query  196   EGLSTQCECNIKVKDVNDN  214
               ++      + + DVNDN
Sbjct  2098  PAMTGTSTLTVVIGDVNDN  2116

 Score = 40.8 bits (94),  Expect = 0.11, Method: Compositional matrix adjust.
 Identities = 55/242 (23%), Positives = 102/242 (42%), Gaps = 22/242 (9%)

Query  4    KFAKPCREGE--DNSKRNP-IAKITSDYQATQKITYRISGVGIDQPPFGIFVVDKNTGDI  60
            +F  P  E E  +N K++  I  + +   A ++I Y +   G      G F +   +G +
Sbjct  678  QFYMPSYEAEIPENQKKDSDIISVKAKSFADREIRYTLKAQGQGA---GTFNIGPTSGIV  734

Query  61   NITAIVDREE--TPSF--LITCRALNAQGLDVEKPLILTVKILDINDNPPVFSQQIFMGE  116
             +   +D E+   P    L+     ++ G      + LT+++ D+NDN P F    +
Sbjct  735  KLAKELDFEDLRQPHVYSLVVTATEDSGGFSTS--VELTIRVTDVNDNAPKFELPDYQAH  792

Query  117  -IEENSASNSLVMILNATDADEPNHLNSKIAFKIVSQEPAGTPMFLLSRNTGEVRTLTNS  175
             ++E+    + ++ + A DAD  +  N++I + +VS +      F +  N   V
Sbjct  793  NVDEDIQIGTSILKVKAMDAD--SGANAEIEY-LVSDD-----HFAVDSNGIIVNNKQLD  844

Query  176  LDREQASSYRLVVSGADKDGEGLSTQCECNIKVKDVNDNFPMFRDSQYSARIEENILSSE  235
             D   A  Y  VV+  DK     +      +  K+ ND  P F    Y+  ++EN   +
Sbjct  845  ADNNNA-YYEFVVTAKDKGEPAKTGTATVRVYTKNKNDEEPKFSQQVYTPNVDENAGPNT  903

Query  236  LL  237
            L+
Sbjct  904  LV  905

 Score = 39.3 bits (90),  Expect = 0.30, Method: Compositional matrix adjust.
 Identities = 55/225 (24%), Positives = 88/225 (39%), Gaps = 24/225 (11%)

Query  75   LITCRALNAQGLDVEKPLILTVKILDINDNPPVFSQQIF--MGEIEENSASNSLVMILNA  132
            +IT +  NA GL       + + + D+ND PP F  +       ++ N+  N+ V  L A
Sbjct  539  IITNQGHNA-GLKYTDNQRVIIHVKDVNDEPPYFINRPLPMQAVVQLNAPPNTPVFTLQA  597

Query  133  TDADEPNHLNSKIAFKIVSQEPAGTPMFLLSRNTGEVRTLTNSLDREQASSYRLVVSGAD  192
             D D     +  I + IV     G   F +   +G VRT    L  +    Y L V   D
Sbjct  598  RDPD----TDHNIHYFIVRDRTGG--RFEVDERSGVVRTRGTDL-FQLDMEYVLYVKAED  650

Query  193  KDGE----GLSTQCECNIKVKDVNDNFPMFRDSQYSARIEEN-ILSSELLRFQVTDLDEE  247
            ++G+       +  E  + +       P F    Y A I EN    S+++  +     +
Sbjct  651  QNGKVDDRRFQSTPEERLSIVG-GKRAPQFYMPSYEAEIPENQKKDSDIISVKA----KS  705

Query  248  YTDNWLAVYFFTSGNEGNWFEIQTDPRTNEGILKVVKALDYEQLQ  292
            + D  +       G     F I        GI+K+ K LD+E L+
Sbjct  706  FADREIRYTLKAQGQGAGTFNIG----PTSGIVKLAKELDFEDLR  746

>AALB20_033171.P66505 | transcript=AALB20_033171.R66505 | gene=AALB20_033171
| organism=Anopheles_albimanus_STECLA_2020 |
gene_product=neural-cadherin, transcript variant X38 |
transcript_product=neural-cadherin,
transcript variant X2 | location=CM024292:75037560-75208393(+)
| protein_length=3131 | sequence_SO=chromosome
| SO=protein_coding_gene | is_pseudo=false
Length=3131

 Score = 144 bits (364),  Expect = 1e-34, Method: Compositional matrix adjust.
 Identities = 125/437 (29%), Positives = 211/437 (48%), Gaps = 33/437 (8%)

Query  26    SDYQATQKITYRISGVGID--QPPFGIFVVDKNTGDINITAIVDREET---PSFLITCRA  80
              D    Q I Y ++G GID   P    F +++ TG+I +   +DR++    P +  T  A
Sbjct  1580  GDKDRPQNIVYFLTGQGIDPDNPANSKFDINRTTGEIFVLKPLDRDQPNGRPQWRFTVFA  1639

Query  81    LNAQGLDVEKPLILTVKILDINDNPPVFSQQIFMGEIEENSASNSLVMILNATDADEPNH  140
              +  G  +     + V + DINDN P+F Q ++ G + EN  +  +VM + A D D+PN
Sbjct  1640  QDEGGEGLVGYADVQVNLKDINDNAPIFPQGVYFGNVTENGTAGMVVMTMTAVDYDDPNE  1699

Query  141   -LNSKIAFKI---VSQEPAGTPMFLLSRNTGEVRTLTNSLDREQASSYRLVVSGADKDGE  196
```

```
                 N+K+ + I   V +E  G+P+F +  +TG ++T    LDRE+   Y + V     DG
Sbjct  1700  GTNAKLIYSIEKNVIEEETGSPIFEIEADTGVIKTAVCCLDRERTPDYSIQVVAM--DGG  1757

Query  197   GLSTQCECNIKVKDVNDNFPMFRDSQYSARIEENILSS--ELLRFQVTDLDEEYTDNWLA  254
             GL      +I+VKD+ND  P F   ++   ++E   ++  E+    VT  DE+ T+ +
Sbjct  1758  GLKGTGTASIRVKDINDMPPQFTKDEWFTEVDETDGTNLPEMPILTVTVHDEDETNKFQY  1817

Query  255   VYFFTSGNEGNWFEIQTDPRTNEGI--LKVVKALDYE---QLQSVKLSIAVKNKAEFHQS  309
                  SG   + F +    R N+G   LK+V+ LDYE   Q    +  I V +K E + +
Sbjct  1818  KVIDNSGYGADKFTMV---RNNDGTGSLKIVQPLDYEDPLQSNGFRFRIQVNDKGEDNDN  1874

Query  310   VISRYRVQSTPVTIQVINVREGIAFRPASKTFTVQKGISSKKLVDYILGTYQAIDEDTNK  369
                +Y V  + V +++ ++ +    +P  +   ++  +     V   L T++A D D
Sbjct  1875  --DKYHVAYSWVVVKLRDINDN---KPQFERPNIEVSVYENADVGKTLETFKATDPDQG-  1928

Query  370   AASNVKYVMGR-NDGGYLMIDSKTAEIKFVKNMNRDSTFIVNKTITAEVLAIDE-YTGKT  427
               S V Y + R +D       ++   +   +N++R+    V      ++LAID+    KT
Sbjct  1929  GKSKVSYAIDRSSDRQRQFSINQEGTVTIQRNLDRE----VTPRHQVKILAIDDGIPPKT  1984

Query  428   STGTVYVRVPDFNDNCP  444
             +T T+ V V D NDN P
Sbjct  1985  ATATLTVIVQDINDNPP  2001

 Score = 85.9 bits (211),  Expect = 1e-15, Method: Compositional matrix adjust.
 Identities = 108/420 (26%), Positives = 177/420 (42%), Gaps = 52/420 (12%)

Query  50    IFVVDKNTGDINITAIVDREETPSFLITCRAL----------NAQGLDVEKPLILTVKIL  99
              F ++  +G+I    + DRE+  ++ +   A           N Q   V K   + + I
Sbjct  1385  FFEINLQSGEIFTKVVFDREKQGAYALEVEARDGAPSARPNSNQQPNSVTK--FIRIGIA  1442

Query  100   DINDNPPVFSQQIFMGEIEENSASNSLVMILNATDADEPNHLNSKIAFKIVSQEPAGTPM  159
             D NDNPP F + ++  E++EN      V+ + A D DE    +S+I ++I S    G
Sbjct  1443  DKNDNPPYFDKALYEAEVDENEDIQHTVLTVTAKDHDE----SSRIRYEITSGNIGGA--  1496

Query  160   FLLSRNTGEVRTLTNSLDREQASSYRLVVSGADKDGEGLSTQCECNIKVKDVNDNFPMFR  219
             F +   TG +  +  +LD E    Y L ++ +D   E  +T     I VKDVNDN P+F
Sbjct  1497  FAVKNMTGAI-YVAGALDYETRKRYELRLAASDNLKENYTTVV---IHVKDVNDNPPVFE  1552

Query  220   DSQYSARIEENI---LSSELLRFQVTDLDEEYTDNWLAVYFFTSG--NEGNWFEIQTDPR  274
                Y  +I E     L   +L+   TD D++   N   VYF T    +  N    + D
Sbjct  1553  RPTYRTQITEEDDRNLPKRVLQVTATDGDKDRPQN--IVYFLTGQGIDPDNPANSKFDIN  1610

Query  275   TNEGILKVVKALDYEQLQS-VKLSIAVKNKAEFHQSVISRYRVQSTPVTIQVINVREGIA  333
                G + V+K LD +Q     +    V  + E  + ++    VQ     + + ++ +
Sbjct  1611  RTTGEIFVLKPLDRDQPNGRPQWRFTVFAQDEGGEGLVGYADVQ-----VNLKDINDNAP  1665

Query  334   FRPASKTFTVQKGISSKKLVDYILGTYQAIDEDTNKAASNVK--YVMGRN------DGGY  385
               P    F     ++       ++ T  A+D D     +N K  Y + +N
Sbjct  1666  IFPQGVYF---GNVTENGTAGMVVMTMTAVDYDDPNEGTNAKLIYSIEKNVIEEETGSPI  1722

Query  386   LMIDSKTAEIK-FVKNMNRDSTFIVNKTITAEVLAIDEYTGKTSTGTVYVRVPDFNDNCP  444
               I++ T  IK  V  ++R+ T       + +V+A+D   G   TGT  +RV D ND  P
Sbjct  1723  FEIEADTGVIKTAVCCLDRERT----PDYSIQVVAMDG-GGLKGTGTASIRVKDINDMPP  1777

 Score = 75.9 bits (185),  Expect = 2e-12, Method: Compositional matrix adjust.
 Identities = 108/429 (25%), Positives = 178/429 (41%), Gaps = 53/429 (12%)

Query  53    VDKNTGDINITAIVDREETPSFLITCRALNAQGLDVEKPLILTVKILDINDNPPVFSQ-Q  111
             +D++  ++N+TA+ D         +C  +N          ++ V I D+NDN PVF
Sbjct  950   LDRDKYELNVTAMDDG--------SC-CVNGDATIHTSTAVVVVFITDVNDNKPVFKDCG  1000

Query  112   IFMGEIEENSASNSLVMILNATDADEPNHLNSKIAFKIVSQEPAGTPMFLLSRNTGEVRT  171
              +  ++EE + + S V+ + ATD D+   +N ++ + IV Q       F +   TGEV T
Sbjct  1001  TYYPKVEEGAPNGSPVIKVQATDEDK--GVNGQVKYSIVQQPNQKGTKFTVDEETGEVST  1058

Query  172   LTNSLDRE--QASSYRLVVSGADKDGEGLSTQCECNIKVKDVNDNFPMFRDSQYSARIEE  229
                  DRE        + V   D+    L   C   +++ DVNDN P+F   +Y   +++
Sbjct  1059  -NKVFDREGDDGKFVSVTVKATDQGEPSLEGVCSFTVEITDVNDNPPLFDRQKYVENVKQ  1117
```

```
Query  230   NI-LSSELLRFQVTDLDEEYTDNWLAVYFFT---SGNEGNWFEIQTDPRTNEGILKVVKA  285
             +  + + +LR  V+  DE+  +N   VY  +   S N+  +FEIQ  P +   +LK  K
Sbjct  1118  DASIGTNILR--VSASDEDADNNGAIVYTLSAPHSPNDLEYFEIQ--PESGWIVLK--KP  1171

Query  286   LD-YEQLQSVKLSIAVK----------NKAEFHQSVISRYRVQSTPV----TIQVINVRE  330
             LD    + SV+    +K          + A+    +    R    P+        I++RE
Sbjct  1172  LDGCYSVSSVRERYRLKVYASDRGDPPSTADVDVELDVVDRNNKPPIWDRNIYGPIHIRE  1231

Query  331   GIAFRPASKTFTVQKGISSKKLVDYILGTYQAIDEDTNKAASNVKYVMGRNDGGYLMIDS  390
              +             GI     V Y L         TNK   +  Y+  R D G
Sbjct  1232  NVTVGTVVTAIKASSGIEGNPTVFYRL--MPGSTAQTNKL--HTFYLQQRPDNG-----D  1282

Query  391   KTAEIKFVKNMNRDSTFIVNKTITAEVLAIDEYTGKTSTGTVYYVRVPDFNDNCPTAV-LE  449
               A+IK    ++ ++    N TI  E     +     S  TVY+++ D ND  P     E
Sbjct  1283  TWADIKVNHPLDFETIKEYNLTIRVENNGAQQL---ASEATVYIQLEDVNDEIPLFTERE  1339

Query  450   KDAVCSSSP  458
             ++ V    P
Sbjct  1340  QETVLEGEP  1348

 Score = 61.2 bits (147),  Expect = 5e-08, Method: Compositional matrix adjust.
 Identities = 87/366 (24%), Positives = 156/366 (43%), Gaps = 38/366 (10%)

Query  51    FVVDKNTGDINITAIVDRE-ETPSFL-ITCRALNAQGLDVEKPLILTVKILDINDNPPVF  108
             F VD+ TG+++   + DRE +   F+ +T +A +     +E     TV+I D+NDNPP+F
Sbjct  1047  FTVDEETGEVSTNKVFDREGDDGKFVSVTVKATDQGEPSLEGVCSFTVEITDVNDNPPLF  1106

Query  109   SQQIFMGEIEENSASNSLVMILNATDADEPNHLNSKIAFKIVS-QEPAGTPMFLLSRNTG  167
              +Q ++  ++++++  + ++ ++A+D D  N  N  I + + +   P     F +   +G
Sbjct  1107  DRQKYVENVKQDASIGTNILRVSASDEDADN--NGAIVYTLSAPHSPNDLEYFEIQPESG  1164

Query  168   EVRTLTNSLDREQASS-----YRLVVSGADKDGEGLSTQCECNIKVKDVNDNFPMFRDSQ  222
              +  L   LD   + S     YRL V  +D+ G+  ST            +N P   D
Sbjct  1165  WI-VLKKPLDGCYSVSSVRERYRLKVYASDR-GDPPSTADVDVELDVVDRNNKPPIWDRN  1222

Query  223   YSA--RIEENILSSELLRFQVTDLDEEYTDNWLAVYFFTSG-----NEGNWFEIQTDPRT  275
                   I EN+   ++              N    Y    G     N+ + F +Q  P
Sbjct  1223  IYGPIHIRENVTVGTVV--TAIKASSGIEGNPTVFYRLMPGSTAQTNKLHTFYLQQRPDN  1280

Query  276   NE--GILKVVKALDYEQLQSVKLSIAVKNKAEFHQSVISRYRVQSTPVTIQVINVREGIA  333
              +    +KV   LD+E ++   L+I V+N         ++       V IQ+ +V + I
Sbjct  1281  GDTWADIKVNHPLDFETIKEYNLTIRVENNG-------AQQLASEATVYIQLEDVNDEIP  1333

Query  334   FRPASKTFTVQKGISSKKLVDYILGTYQAIDEDTNKAASNVKYVM---GRNDG-GYLMID  389
                  +  TV +G      V  +     AID+D     + V Y +    RN+G  +  I+
Sbjct  1334  LFTEREQETVLEGEPIGTKVTQV----NAIDKDGTFPNNQVYYYIVDSPRNEGKDFFEIN  1389

Query  390   SKTAEI  395
              ++ EI
Sbjct  1390  LQSGEI  1395

 Score = 55.8 bits (133),  Expect = 3e-06, Method: Compositional matrix adjust.
 Identities = 49/158 (31%), Positives = 78/158 (49%), Gaps = 24/158 (15%)

Query  102   NDNPPVFSQQIFMGEIEENSASNSLVMILNATDADEPNHLNSKIAFKIVSQEPAGTP--M  159
             ND  P FSQQ++   ++EN+  N+LV  + A+D D  N     + F  V     GT
Sbjct  880   NDEEPKFSQQVYTPNVDENAGPNTLVTTVVASDKDGDN-----VRFGFVG---GGTSSGQ  931

Query  160   FLLSRNTGEVRTLTN--SLDREQASSYRLVVSGADK-------DGEGLSTQCECNIKVKD  210
             F++   TG +R  +   SLDR++   Y L V+  D        D    ++     + + D
Sbjct  932   FVIEEITGVIRLHSKAISLDRDK---YELNVTAMDDGSCCVNGDATIHTSTAVVVVFITD  988

Query  211   VNDNFPMFRD-SQYSARIEENILS-SELLRFQVTDLDE  246
             VNDN P+F+D   Y  ++EE   + S +++ Q TD D+
Sbjct  989   VNDNKPVFKDCGTYYPKVEEGAPNGSPVIKVQATDEDK  1026

 Score = 53.9 bits (128),  Expect = 1e-05, Method: Compositional matrix adjust.
 Identities = 70/306 (23%), Positives = 126/306 (41%), Gaps = 30/306 (10%)
```

```
Query  20    PIAKIT-SDYQATQKITYRI---SGVGIDQPPFGIFVVDKNTGDINITAIVDRE---ETP  72
             PI  +T  D   T K  Y++   SG G D+  F +   +  TG + I   +D E   ++
Sbjct  1800  PILTVTVHDEDETNKFQYKVIDNSGYGADK--FTMVRNNDGTGSLKIVQPLDYEDPLQSN  1857

Query  73    SFLITCRALNAQGLDVEKPLI------LTVKILDINDNPPVFSQQIFMGEIEENSASNSL  126
              F    + +N +G D +           + VK+ DINDN P F +       + EN+
Sbjct  1858  GFRFRIQ-VNDKGEDNDNDKYHVAYSWVVVKLRDINDNKPQFERPNIEVSVYENADVGKT  1916

Query  127   VMILNATDADEPNHLNSKIAFKIVSQEPAGTPMFLLSRNTGEVRTLTNSLDREQASSYRL  186
             +    ATD D+      SK+++ I   + +        S N     T+  +LDRE    +++
Sbjct  1917  LETFKATDPDQGG--KSKVSYAI---DRSSDRQRQFSINQEGTVTIQRNLDREVTPRHQV  1971

Query  187   VVSGADKDGEGLSTQCECNIKVKDVNDNFPMFRDSQYSARIEENILSSELLRFQVTDLDE  246
              +   D      +       + V+D+NDN P F    Y   + E++   +++    TD D+
Sbjct  1972  KILAIDDGIPPKTATATLTVIVQDINDNPPKFL-KDYRPVLPEHVPPRKVVEILATDDDD  2030

Query  247   EYTDNWLAVYFFTSGNEGNW----FEIQTDPRTNEG----ILKVVKALDYEQLQSVKLSI  298
                N     F        +     F+++ D +   G     I+  +++ D EQ +   + I
Sbjct  2031  RSKSNGPPFQFRLDPGADDIIRASFKVEQDQKGANGDGMAIVSSLRSFDREQQKEYLIPI  2090

Query  299   AVKNKA  304
              +K+
Sbjct  2091  VIKDHG  2096

 Score = 51.6 bits (122),  Expect = 5e-05, Method: Compositional matrix adjust.
 Identities = 96/398 (24%), Positives = 163/398 (41%), Gaps = 54/398 (14%)

Query  106   PVFSQQIFMGEIEENSASNSLVMILNATDADEPNHLNSKIAFKIVSQ-EPAGTPMFLLSR  164
             P F    +  EI EN   +S ++ + A       +  + +I + + +Q + AGT  F +
Sbjct  677   PQFYMPSYEAEIPENQKKDSDIISVKAK-----SFADREIRYTLKAQGQGAGT--FNIGP  729

Query  165   NTGEVRTLTNSLDRE---QASSYRLVVSGADKDGEGLSTQCECNIKVKDVNDNFPMFRDS  221
              +G V+ L   LD E   Q   Y LVV+ A +D  G ST  E  I+V DVNDN P F
Sbjct  730   TSGIVK-LAKELDFEDLRQPHVYSLVVT-ATEDSGGFSTSVELTIRVTDVNDNAPKFELP  787

Query  222   QYSAR-IEENI-LSSELLRFQVTDLDEEYTDNWLAVYFFTSGNEGNWFEIQTDPRTNEGI  279
              Y A  ++E+I + + +L+ +  D D          + +  S +    F + ++     GI
Sbjct  788   DYQAHNVDEDIQIGTSILKVKAMDADSGAN---AEIEYLVSDDH---FAVDSN-----GI  836

Query  280   LKVVKALDYEQLQSV-KLSIAVKNKAEFHQSVISRYRVQSTPVTIQVINVREGIAFRPAS  338
             +   K LD +   +  +  +  K+K E  ++  +  RV +              E   F
Sbjct  837   IVNNKQLDADNNNAYYEFVVTAKDKGEPAKTGTATVRVYTKNKN------DEEPKFSQQV  890

Query  339   KTFTVQKGISSKKLVDYILGTYQAIDEDTNKAASNVKY--VMGRNDGGYLMIDSKTAEIK  396
              T  V +       LV  ++ +         +K   NV++  V G    G  +I+  T  I+
Sbjct  891   YTPNVDENAGPNTLVTTVVAS--------DKDGDNVRFGFVGGGTSSGQFVIEEITGVIR  942

Query  397   FVK---NMNRDSTFI-VNKTITAEVLAIDEYTGKTSTGTVYVRVPDFNDN------CPTA  446
                   +++RD   + V               + T  TST  V V + D NDN      C T
Sbjct  943   LHSKAISLDRDKYELNVTAMDDGSCCVNGDATIHTSTAVVVVFITDVNDNKPVFKDCGTY  1002

Query  447   VLE-KDAVCSSSPSVVVSARTLNNRYTGPYTFALEDQP  483
              + ++    + SP + V A    +    G   +++  QP
Sbjct  1003  YPKVEEGAPNGSPVIKVQATDEDKGVNGQVKYSIVQQP  1040

 Score = 44.7 bits (104),  Expect = 0.008, Method: Compositional matrix adjust.
 Identities = 48/199 (24%), Positives = 73/199 (37%), Gaps = 19/199 (10%)

Query  27    DYQATQKITYRISGVGIDQPPFGIFVVDKNTGDINITAIVDREETPSFLITCRALNAQGL  86
             D      K++Y I       Q  F I        G + I   +DRE TP    +    A++
Sbjct  1926  DQGGKSKVSYAIDRSSDRQRQFSI----NQEGTVTIQRNLDREVTPRHQVKILAIDDGIP  1981

Query  87    DVEKPLILTVKILDINDNPPVFSQQIFMGEIEENSASNSLVMILNATDADEPNHLNSKIA  146
                      LTV + DINDNPP F +  +    + E+      +V IL   D D
Sbjct  1982  PKTATATLTVIVQDINDNPPKFLKD-YRPVLPEHVPPRKVVEILATDDDDRSKSNGPPFQ  2040

Query  147   FKIVSQEPAGTPMFLLSRNTGEVRTLTN-----------SLDREQASSYRLVVSGADKDG  195
             F++    +P    +    S    + +    N             S DREQ    Y + +    D
```

```
Sbjct  2041  FRL---DPGADDIIRASFKVEQDQKGANGDGMAIVSSLRSFDREQQKEYLIPIVIKDHGN  2097

Query  196   EGLSTQCECNIKVKDVNDN  214
               ++      + + DVNDN
Sbjct  2098  PAMTGTSTLTVVIGDVNDN  2116

 Score = 40.8 bits (94),  Expect = 0.11, Method: Compositional matrix adjust.
 Identities = 55/242 (23%), Positives = 102/242 (42%), Gaps = 22/242 (9%)

Query  4    KFAKPCREGE--DNSKRNP-IAKITSDYQATQKITYRISGVGIDQPPFGIFVVDKNTGDI  60
            +F  P  E E  +N K++  I  + +   A ++I Y +   G      G F +   +G +
Sbjct  678  QFYMPSYEAEIPENQKKDSDIISVKAKSFADREIRYTLKAQGQGA---GTFNIGPTSGIV  734

Query  61   NITAIVDREE--TPSF--LITCRALNAQGLDVEKPLILTVKILDINDNPPVFSQQIFMGE  116
             +   +D E+   P    L+     ++ G      + LT+++ D+NDN P F    +
Sbjct  735  KLAKELDFEDLRQPHVYSLVVTATEDSGGFSTS--VELTIRVTDVNDNAPKFELPDYQAH  792

Query  117  -IEENSASNSLVMILNATDADEPNHLNSKIAFKIVSQEPAGTPMFLLSRNTGEVRTLTNS  175
             ++E+    + ++ + A DAD  +  N++I + +VS +      F +  N   V
Sbjct  793  NVDEDIQIGTSILKVKAMDAD--SGANAEIEY-LVSDD-----HFAVDSNGIIVNNKQLD  844

Query  176  LDREQASSYRLVVSGADKDGEGLSTQCECNIKVKDVNDNFPMFRDSQYSARIEENILSSE  235
             D   A  Y  VV+  DK     +      +  K+ ND  P F    Y+  ++EN    +
Sbjct  845  ADNNNA-YYEFVVTAKDKGEPAKTGTATVRVYTKNKNDEEPKFSQQVYTPNVDENAGPNT  903

Query  236  LL  237
            L+
Sbjct  904  LV  905

 Score = 39.3 bits (90),  Expect = 0.30, Method: Compositional matrix adjust.
 Identities = 55/225 (24%), Positives = 88/225 (39%), Gaps = 24/225 (11%)

Query  75   LITCRALNAQGLDVEKPLILTVKILDINDNPPVFSQQIF--MGEIEENSASNSLVMILNA  132
            +IT +  NA GL       + + + D+ND PP F  +       ++ N+  N+ V  L A
Sbjct  539  IITNQGHNA-GLKYTDNQRVIIHVKDVNDEPPYFINRPLPMQAVVQLNAPPNTPVFTLQA  597

Query  133  TDADEPNHLNSKIAFKIVSQEPAGTPMFLLSRNTGEVRTLTNSLDREQASSYRLVVSGAD  192
             D D     +  I + IV     G   F +   +G VRT    L  +    Y L V   D
Sbjct  598  RDPD----TDHNIHYFIVRDRTGG--RFEVDERSGVVRTRGTDL-FQLDMEYVLYVKAED  650

Query  193  KDGE----GLSTQCECNIKVKDVNDNFPMFRDSQYSARIEEN-ILSSELLRFQVTDLDEE  247
            ++G+       +  E  + +       P F    Y A I EN    S+++  +      +
Sbjct  651  QNGKVDDRRFQSTPEERLSIVG-GKRAPQFYMPSYEAEIPENQKKDSDIISVKA----KS  705

Query  248  YTDNWLAVYFFTSGNEGNWFEIQTDPRTNEGILKVVKALDYEQLQ  292
            + D  +       G     F I        GI+K+ K LD+E L+
Sbjct  706  FADREIRYTLKAQGQGAGTFNIG----PTSGIVKLAKELDFEDLR  746

>AALB20_033171.P66503 | transcript=AALB20_033171.R66503 | gene=AALB20_033171
| organism=Anopheles_albimanus_STECLA_2020 |
gene_product=neural-cadherin, transcript variant X38 |
transcript_product=neural-cadherin,
transcript variant X10 | location=CM024292:75037560-75208393(+)
| protein_length=3131 | sequence_SO=chromosome
| SO=protein_coding_gene | is_pseudo=false
Length=3131

 Score = 144 bits (364),  Expect = 1e-34, Method: Compositional matrix adjust.
 Identities = 125/437 (29%), Positives = 211/437 (48%), Gaps = 33/437 (8%)

Query  26    SDYQATQKITYRISGVGID--QPPFGIFVVDKNTGDINITAIVDREET---PSFLITCRA  80
              D    Q I Y ++G GID   P    F +++ TG+I +   +DR++    P +  T  A
Sbjct  1580  GDKDRPQNIVYFLTGQGIDPDNPANSKFDINRTTGEIFVLKPLDRDQPNGRPQWRFTVFA  1639

Query  81    LNAQGLDVEKPLILTVKILDINDNPPVFSQQIFMGEIEENSASNSLVMILNATDADEPNH  140
              +  G  +     + V + DINDN P+F Q ++ G + EN  +  +VM + A D D+PN
Sbjct  1640  QDEGGEGLVGYADVQVNLKDINDNAPIFPQGVYFGNVTENGTAGMVVMTMTAVDYDDPNE  1699
```

```
Query  141   -LNSKIAFKI---VSQEPAGTPMFLLSRNTGEVRTLTNSLDREQASSYRLVVSGADKDGE  196
               N+K+ + I   V +E  G+P+F +  +TG ++T    LDRE+   Y + V     DG
Sbjct  1700  GTNAKLIYSIEKNVIEEETGSPIFEIEADTGVIKTAVCCLDRERTPDYSIQVVAM--DGG  1757

Query  197   GLSTQCECNIKVKDVNDNFPMFRDSQYSARIEENILSS--ELLRFQVTDLDEEYTDNWLA  254
             GL      +I+VKD+ND  P F   ++   ++E   ++  E+    VT  DE+ T+ +
Sbjct  1758  GLKGTGTASIRVKDINDMPPQFTKDEWFTEVDETDGTNLPEMPILTVTVHDEDETNKFQY  1817

Query  255   VYFFTSGNEGNWFEIQTDPRTNEGI--LKVVKALDYE---QLQSVKLSIAVKNKAEFHQS  309
                  SG   + F +    R N+G   LK+V+ LDYE   Q    +  I V +K E + +
Sbjct  1818  KVIDNSGYGADKFTMV---RNNDGTGSLKIVQPLDYEDPLQSNGFRFIQVNDKGEDNDN  1874

Query  310   VISRYRVQSTPVTIQVINVREGIAFRPASKTFTVQKGISSKKLVDYILGTYQAIDEDTNK  369
                +Y V  + V +++ ++ +    +P  +   ++  +     V   L T++A D D
Sbjct  1875  --DKYHVAYSWVVVKLRDINDN---KPQFERPNIEVSVYENADVGKTLETFKATDPDQG-  1928

Query  370   AASNVKYVMGR-NDGGYLMIDSKTAEIKFVKNMNRDSTFIVNKTITAEVLAIDE-YTGKT  427
               S V Y + R +D       ++   +   +N++R+    V      ++LAID+    KT
Sbjct  1929  GKSKVSYAIDRSSDRQRQFSINQEGTVTIQRNLDRE----VTPRHQVKILAIDDGIPPKT  1984

Query  428   STGTVYYVRVPDFNDNCP  444
             +T T+ V V D NDN P
Sbjct  1985  ATATLTVIVQDINDNPP  2001

 Score = 85.9 bits (211),  Expect = 1e-15, Method: Compositional matrix adjust.
 Identities = 108/420 (26%), Positives = 177/420 (42%), Gaps = 52/420 (12%)

Query  50    IFVVDKNTGDINITAIVDREETPSFLITCRAL----------NAQGLDVEKPLILTVKIL  99
              F ++  +G+I    + DRE+  ++ +   A           N Q   V K   + + I
Sbjct  1385  FFEINLQSGEIFTKVVFDREKQGAYALEVEARDGAPSARPNSNQQPNSVTK--FIRIGIA  1442

Query  100   DINDNPPVFSQQIFMGEIEENSASNSLVMILNATDADEPNHLNSKIAFKIVSQEPAGTPM  159
             D NDNPP F + ++  E++EN      V+ + A D DE    +S+I ++I S    G
Sbjct  1443  DKNDNPPYFDKALYEAEVDENEDIQHTVLTVTAKDHDE----SSRIRYEITSGNIGGA--  1496

Query  160   FLLSRNTGEVRTLTNSLDREQASSYRLVVSGADKDGEGLSTQCECNIKVKDVNDNFPMFR  219
             F +   TG +  +  +LD E    Y L ++ +D   E   +T    I VKDVNDN P+F
Sbjct  1497  FAVKNMTGAI-YVAGALDYETRKRYELRLAASDNLKENYTTVV---IHVKDVNDNPPVFE  1552

Query  220   DSQYSARIEENI---LSSELLRFQVTDLDEEYTDNWLAVYFFTSG--NEGNWFEIQTDPR  274
                Y  +I E     L   +L+   TD D++   N   VYF T    +  N    + D
Sbjct  1553  RPTYRTQITEEDDRNLPKRVLQVTATDGDKDRPQN--IVYFLTGQGIDPDNPANSKFDIN  1610

Query  275   TNEGILKVVKALDYEQLQS-VKLSIAVKNKAEFHQSVISRYRVQSTPVTIQVINVREGIA  333
                G + V+K LD +Q     +    V  + E  + ++    VQ     + + ++ +
Sbjct  1611  RTTGEIFVLKPLDRDQPNGRPQWRFTVFAQDEGGEGLVGYADVQ-----VNLKDINDNAP  1665

Query  334   FRPASKTFTVQKGISSKKLVDYILGTYQAIDEDTNKAASNVK--YVMGRN------DGGY  385
               P    F     ++       ++ T  A+D D     +N K  Y + +N
Sbjct  1666  IFPQGVYF---GNVTENGTAGMVVMTMTAVDYDDPNEGTNAKLIYSIEKNVIEEETGSPI  1722

Query  386   LMIDSKTAEIK-FVKNMNRDSTFIVNKTITAEVLAIDEYTGKTSTGTVYYVRVPDFNDNCP  444
               I++ T  IK  V  ++R+ T       + +V+A+D   G   TGT  +RV D ND  P
Sbjct  1723  FEIEADTGVIKTAVCCLDRERT----PDYSIQVVAMDG-GGLKGTGTASIRVKDINDMPP  1777

 Score = 75.9 bits (185),  Expect = 2e-12, Method: Compositional matrix adjust.
 Identities = 108/429 (25%), Positives = 178/429 (41%), Gaps = 53/429 (12%)

Query  53    VDKNTGDINITAIVDREETPSFLITCRALNAQGLDVEKPLILTVKILDINDNPPVFSQ-Q  111
             +D++  ++N+TA+ D         +C  +N          ++ V I D+NDN PVF
Sbjct  950   LDRDKYELNVTAMDDG---------SC-CVNGDATIHTSTAVVVVFITDVNDNKPVFKDCG  1000

Query  112   IFMGEIEENSASNSLVMILNATDADEPNHLNSKIAFKIVSQEPAGTPMFLLSRNTGEVRT  171
              +  ++EE + + S V+ + ATD D+   +N ++ + IV Q       F +   TGEV T
Sbjct  1001  TYYPKVEEGAPNGSPVIKVQATDEDK--GVNGQVKYSIVQQPNQKGTKFTVDEETGEVST  1058

Query  172   LTNSLDRE--QASSYRLVVSGADKDGEGLSTQCECNIKVKDVNDNFPMFRDSQYSARIEE  229
                  DRE        + V   D+    L   C   +++ DVNDN P+F   +Y   +++
Sbjct  1059  -NKVFDREGDDGKFVSVTVKATDQGEPSLEGVCSFTVEITDVNDNPPLFDRQKYVENVKQ  1117
```

```
Query  230   NI-LSSELLRFQVTDLDEEYTDNWLAVYFFT---SGNEGNWFEIQTDPRTNEGILKVVKA  285
             +  + + +LR  V+  DE+  +N   VY  +   S N+  +FEIQ  P +   +LK  K
Sbjct  1118  DASIGTNILR--VSASDEDADNNGAIVYTLSAPHSPNDLEYFEIQ--PESGWIVLK--KP  1171

Query  286   LD-YEQLQSVKLSIAVK----------NKAEFHQSVISRYRVQSTPV----TIQVINVRE  330
             LD    + SV+    +K          + A+    +    R    P+        I++RE
Sbjct  1172  LDGCYSVSSVRERYRLKVYASDRGDPPSTADVDVELDVVDRNNKPPIWDRNIYGPIHIRE  1231

Query  331   GIAFRPASKTFTVQKGISSKKLVDYILGTYQAIDEDTNKAASNVKYVMGRNDGGYLMIDS  390
              +              GI     V Y L          TNK   +  Y+  R D G
Sbjct  1232  NVTVGTVVTAIKASSGIEGNPTVFYRL--MPGSTAQTNKL--HTFYLQQRPDNG-----D  1282

Query  391   KTAEIKFVKNMNRDSTFIVNKTITAEVLAIDEYTGKTSTGTVYVRVPDFNDNCPTAV-LE  449
               A+IK    ++ ++    N TI  E     +      S  TVY+++ D ND  P     E
Sbjct  1283  TWADIKVNHPLDFETIKEYNLTIRVENNGAQQL---ASEATVYIQLEDVNDEIPLFTERE  1339

Query  450   KDAVCSSSP  458
             ++ V    P
Sbjct  1340  QETVLEGEP  1348

 Score = 61.2 bits (147),  Expect = 5e-08, Method: Compositional matrix adjust.
 Identities = 87/366 (24%), Positives = 156/366 (43%), Gaps = 38/366 (10%)

Query  51    FVVDKNTGDINITAIVDRE-ETPSFL-ITCRALNAQGLDVEKPLILTVKILDINDNPPVF  108
             F VD+ TG+++    + DRE +   F+ +T +A +     +E     TV+I D+NDNPP+F
Sbjct  1047  FTVDEETGEVSTNKVFDREGDDGKFVSVTVKATDQGEPSLEGVCSFTVEITDVNDNPPLF  1106

Query  109   SQQIFMGEIEENSASNSLVMILNATDADEPNHLNSKIAFKIVS-QEPAGTPMFLLSRNTG  167
              +Q ++  ++++++  + ++ ++A+D D  N  N  I + + +   P     F +   +G
Sbjct  1107  DRQKYVENVKQDASIGTNILRVSASDEDADN--NGAIVYTLSAPHSPNDLEYFEIQPESG  1164

Query  168   EVRTLTNSLDREQASS-----YRLVVSGADKDGEGLSTQCECNIKVKDVNDNFPMFRDSQ  222
              +  L   LD   + S     YRL V  +D+ G+  ST             +N P   D
Sbjct  1165  WI-VLKKPLDGCYSVSSVRERYRLKVYASDR-GDPPSTADVDVELDVVDRNNKPPIWDRN  1222

Query  223   YSA--RIEENILSSELLRFQVTDLDEEYTDNWLAVYFFTSG-----NEGNWFEIQTDPRT  275
                    I EN+    ++               N    Y    G     N+ + F +Q  P
Sbjct  1223  IYGPIHIRENVTVGTVV--TAIKASSGIEGNPTVFYRLMPGSTAQTNKLHTFYLQQRPDN  1280

Query  276   NE--GILKVVKALDYEQLQSVKLSIAVKNKAEFHQSVISRYRVQSTPVTIQVINVREGIA  333
              +    +KV   LD+E ++   L+I V+N          ++       V IQ+ +V + I
Sbjct  1281  GDTWADIKVNHPLDFETIKEYNLTIRVENNG-------AQQLASEATVYIQLEDVNDEIP  1333

Query  334   FRPASKTFTVQKGISSKKLVDYILGTYQAIDEDTNKAASNVKYVM---GRNDG-GYLMID  389
                  +  TV +G      V  +     AID+D     + V Y +    RN+G  +  I+
Sbjct  1334  LFTEREQETVLEGEPIGTKVTQV----NAIDKDGTFPNNQVYYYIVDSPRNEGKDFFEIN  1389

Query  390   SKTAEI  395
              ++ EI
Sbjct  1390  LQSGEI  1395

 Score = 55.8 bits (133),  Expect = 3e-06, Method: Compositional matrix adjust.
 Identities = 49/158 (31%), Positives = 78/158 (49%), Gaps = 24/158 (15%)

Query  102   NDNPPVFSQQIFMGEIEENSASNSLVMILNATDADEPNHLNSKIAFKIVSQEPAGTP--M  159
             ND  P FSQQ++   ++EN+  N+LV  + A+D D  N     + F  V     GT
Sbjct  880   NDEEPKFSQQVYTPNVDENAGPNTLVTTVVASDKDGDN-----VRFGFVG---GGTSSGQ  931

Query  160   FLLSRNTGEVRTLTN--SLDREQASSYRLVVSGADK-------DGEGLSTQCECNIKVKD  210
             F++   TG +R  +   SLDR++   Y L V+  D         D   ++      + + D
Sbjct  932   FVIEEITGVIRLHSKAISLDRDK---YELNVTAMDDGSCCVNGDATIHTSTAVVVVFITD  988

Query  211   VNDNFPMFRD-SQYSARIEENILS-SELLRFQVTDLDE  246
             VNDN P+F+D   Y  ++EE   + S +++ Q TD D+
Sbjct  989   VNDNKPVFKDCGTYYPKVEEGAPNGSPVIKVQATDEDK  1026

 Score = 53.9 bits (128),  Expect = 1e-05, Method: Compositional matrix adjust.
```

```
 Identities = 70/306 (23%), Positives = 126/306 (41%), Gaps = 30/306 (10%)

Query  20    PIAKIT-SDYQATQKITYRI---SGVGIDQPPFGIFVVDKNTGDINITAIVDRE---ETP  72
             PI  +T  D   T K  Y++   SG G D+  F +    +  TG + I   +D E   ++
Sbjct  1800  PILTVTVHDEDETNKFQYKVIDNSGYGADK--FTMVRNNDGTGSLKIVQPLDYEDPLQSN  1857

Query  73    SFLITCRALNAQGLDVEKPLI------LTVKILDINDNPPVFSQQIFMGEIEENSASNSL  126
              F    + +N +G D +           + VK+ DINDN P F +       + EN+
Sbjct  1858  GFRFRIQ-VNDKGEDNDNDKYHVAYSWVVVKLRDINDNKPQFERPNIEVSVYENADVGKT  1916

Query  127   VMILNATDADEPNHLNSKIAFKIVSQEPAGTPMFLLSRNTGEVRTLTNSLDREQASSYRL  186
             +    ATD D+     SK+++ I   + +       S N     T+  +LDRE    +++
Sbjct  1917  LETFKATDPDQGG--KSKVSYAI---DRSSDRQRQFSINQEGTVTIQRNLDREVTPRHQV  1971

Query  187   VVSGADKDGEGLSTQCECNIKVKDVNDNFPMFRDSQYSARIEENILSSELLRFQVTDLDE  246
              +   D      +       + V+D+NDN P F    Y   + E++   +++    TD D+
Sbjct  1972  KILAIDDGIPPKTATATLTVIVQDINDNPPKFL-KDYRPVLPEHVPPRKVVEILATDDDD  2030

Query  247   EYTDNWLAVYFFTSGNEGNW----FEIQTDPRTNEG----ILKVVKALDYEQLQSVKLSI  298
                 N     F       +     F+++ D +   G    I+  +++ D EQ +   + I
Sbjct  2031  RSKSNGPPFQFRLDPGADDIIRASFKVEQDQKGANGDGMAIVSSLRSFDREQQKEYLIPI  2090

Query  299   AVKNKA  304
              +K+
Sbjct  2091  VIKDHG  2096

 Score = 51.6 bits (122),  Expect = 5e-05, Method: Compositional matrix adjust.
 Identities = 96/398 (24%), Positives = 163/398 (41%), Gaps = 54/398 (14%)

Query  106   PVFSQQIFMGEIEENSASNSLVMILNATDADEPNHLNSKIAFKIVSQ-EPAGTPMFLLSR  164
             P F    +  EI EN   +S ++ + A       +  + +I + + +Q + AGT  F +
Sbjct  677   PQFYMPSYEAEIPENQKKDSDIISVKAK-----SFADREIRYTLKAQGQGAGT--FNIGP  729

Query  165   NTGEVRTLTNSLDRE---QASSYRLVVSGADKDGEGLSTQCECNIKVKDVNDNFPMFRDS  221
              +G V+ L   LD E   Q   Y LVV+ A +D  G ST  E  I+V DVNDN P F
Sbjct  730   TSGIVK-LAKELDFEDLRQPHVYSLVVT-ATEDSGGFSTSVELTIRVTDVNDNAPKFELP  787

Query  222   QYSAR-IEENI-LSSELLRFQVTDLDEEYTDNWLAVYFFTSGNEGNWFEIQTDPRTNEGI  279
              Y A  ++E+I + + +L+ +  D D          + +  S +    F + ++     GI
Sbjct  788   DYQAHNVDEDIQIGTSILKVKAMDADSGAN---AEIEYLVSDDH---FAVDSN-----GI  836

Query  280   LKVVKALDYEQLQSV-KLSIAVKNKAEFHQSVISRYRVQSTPVTIQVINVREGIAFRPAS  338
             +   K LD +   +  +  +  K+K E  ++  +  RV +             E   F
Sbjct  837   IVNNKQLDADNNNAYYEFVVTAKDKGEPAKTGTATVRVYTKNKN------DEEPKFSQQV  890

Query  339   KTFTVQKGISSKKLVDYILGTYQAIDEDTNKAASNVKY--VMGRNDGGYLMIDSKTAEIK  396
              T  V +      LV  ++ +         +K   NV++  V G    G  +I+  T  I+
Sbjct  891   YTPNVDENAGPNTLVTTVVAS--------DKDGDNVRFGFVGGGTSSGQFVIEEITGVIR  942

Query  397   FVK---NMNRDSTFI-VNKTITAEVLAIDEYTGKTSTGTVYVRVPDFNDN------CPTA  446
                  +++RD   + V              + T  TST  V V + D NDN      C T
Sbjct  943   LHSKAISLDRDKYELNVTAMDDGSCCVNGDATIHTSTAVVVVFITDVNDNKPVFKDCGTY  1002

Query  447   VLE-KDAVCSSSPSVVVSARTLNNRYTGPYTFALEDQP  483
               + ++   + SP + V A    +    G   +++  QP
Sbjct  1003  YPKVEEGAPNGSPVIKVQATDEDKGVNGQVKYSIVQQP  1040

 Score = 44.7 bits (104),  Expect = 0.008, Method: Compositional matrix adjust.
 Identities = 48/199 (24%), Positives = 73/199 (37%), Gaps = 19/199 (10%)

Query  27    DYQATQKITYRISGVGIDQPPFGIFVVDKNTGDINITAIVDREETPSFLITCRALNAQGL  86
             D     K++Y I       Q  F I        G + I   +DRE TP    +    A++
Sbjct  1926  DQGGKSKVSYAIDRSSDRQRQFSI----NQEGTVTIQRNLDREVTPRHQVKILAIDDGIP  1981

Query  87    DVEKPLILTVKILDINDNPPVFSQQIFMGEIEENSASNSLVMILNATDADEPNHLNSKIA  146
                    LTV + DINDNPP F +  +    + E+      +V IL   D D
Sbjct  1982  PKTATATLTVIVQDINDNPPKFLKD-YRPVLPEHVPPRKVVEILATDDDDRSKSNGPPFQ  2040

Query  147   FKIVSQEPAGTPMFLLSRNTGEVRTLTN-----------SLDREQASSYRLVVSGADKDG  195
```

```
             F++   +P    +   S    + +   N           S DREQ   Y + +   D
Sbjct  2041  FRL---DPGADDIIRASFKVEQDQKGANGDGMAIVSSLRSFDREQQKEYLIPIVIKDHGN  2097

Query  196   EGLSTQCECNIKVKDVNDN  214
               ++      + + DVNDN
Sbjct  2098  PAMTGTSTLTVVIGDVNDN  2116

 Score = 40.8 bits (94),  Expect = 0.11, Method: Compositional matrix adjust.
 Identities = 55/242 (23%), Positives = 102/242 (42%), Gaps = 22/242 (9%)

Query  4    KFAKPCREGE--DNSKRNP-IAKITSDYQATQKITYRISGVGIDQPPFGIFVVDKNTGDI  60
            +F  P  E E  +N K++  I  + +   A ++I Y +   G      G F +   +G +
Sbjct  678  QFYMPSYEAEIPENQKKDSDIISVKAKSFADREIRYTLKAQGQGA---GTFNIGPTSGIV  734

Query  61   NITAIVDREE--TPSF--LITCRALNAQGLDVEKPLILTVKILDINDNPPVFSQQIFMGE  116
             +   +D E+   P    L+     ++ G      + LT+++ D+NDN P F    +
Sbjct  735  KLAKELDFEDLRQPHVYSLVVTATEDSGGFSTS--VELTIRVTDVNDNAPKFELPDYQAH  792

Query  117  -IEENSASNSLVMILNATDADEPNHLNSKIAFKIVSQEPAGTPMFLLSRNTGEVRTLTNS  175
             ++E+    + ++ + A DAD  +  N++I + +VS +      F +  N   V
Sbjct  793  NVDEDIQIGTSILKVKAMDAD--SGANAEIEY-LVSDD-----HFAVDSNGIIVNNKQLD  844

Query  176  LDREQASSYRLVVSGADKDGEGLSTQCECNIKVKDVNDNFPMFRDSQYSARIEENILSSE  235
             D   A  Y  VV+  DK     +      +  K+ ND  P F    Y+  ++EN    +
Sbjct  845  ADNNNA-YYEFVVTAKDKGEPAKTGTATVRVYTKNKNDEEPKFSQQVYTPNVDENAGPNT  903

Query  236  LL  237
            L+
Sbjct  904  LV  905

 Score = 39.3 bits (90),  Expect = 0.30, Method: Compositional matrix adjust.
 Identities = 55/225 (24%), Positives = 88/225 (39%), Gaps = 24/225 (11%)

Query  75   LITCRALNAQGLDVEKPLILTVKILDINDNPPVFSQQIF--MGEIEENSASNSLVMILNA  132
            +IT +  NA GL       + + + D+ND PP F  +       ++ N+  N+ V  L A
Sbjct  539  IITNQGHNA-GLKYTDNQRVIIHVKDVNDEPPYFINRPLPMQAVVQLNAPPNTPVFTLQA  597

Query  133  TDADEPNHLNSKIAFKIVSQEPAGTPMFLLSRNTGEVRTLTNSLDREQASSYRLVVSGAD  192
             D D     +  I + IV     G   F +   +G VRT    L  +    Y L V   D
Sbjct  598  RDPD----TDHNIHYFIVRDRTGG--RFEVDERSGVVRTRGTDL-FQLDMEYVLYVKAED  650

Query  193  KDGE----GLSTQCECNIKVKDVNDNFPMFRDSQYSARIEEN-ILSSELLRFQVTDLDEE  247
            ++G+       +  E  + +       P F    Y A I EN    S+++  +     +
Sbjct  651  QNGKVDDRRFQSTPEERLSIVG-GKRAPQFYMPSYEAEIPENQKKDSDIISVKA----KS  705

Query  248  YTDNWLAVYFFTSGNEGNWFEIQTDPRTNEGILKVVKALDYEQLQ  292
            + D  +       G     F I        GI+K+ K LD+E L+
Sbjct  706  FADREIRYTLKAQGQGAGTFNIG----PTSGIVKLAKELDFEDLR  746

>AALB20_033171.P66502 | transcript=AALB20_033171.R66502 | gene=AALB20_033171
| organism=Anopheles_albimanus_STECLA_2020 |
gene_product=neural-cadherin, transcript variant X38 |
transcript_product=neural-cadherin,
transcript variant X22 | location=CM024292:75037560-75208393(+)
| protein_length=3131 | sequence_SO=chromosome
| SO=protein_coding_gene | is_pseudo=false
Length=3131

 Score = 144 bits (364),  Expect = 1e-34, Method: Compositional matrix adjust.
 Identities = 125/437 (29%), Positives = 211/437 (48%), Gaps = 33/437 (8%)

Query  26    SDYQATQKITYRISGVGID--QPPFGIFVVDKNTGDINITAIVDREET---PSFLITCRA  80
              D    Q I Y ++G GID   P    F +++ TG+I +   +DR++     P +  T  A
Sbjct  1580  GDKDRPQNIVYFLTGQGIDPDNPANSKFDINRTTGEIFVLKPLDRDQPNGRPQWRFTVFA  1639

Query  81    LNAQGLDVEKPLILTVKILDINDNPPVFSQQIFMGEIEENSASNSLVMILNATDADEPNH  140
              +  G  +     + V + DINDN P+F Q ++ G + EN  +  +VM + A D D+PN
Sbjct  1640  QDEGGEGLVGYADVQVNLKDINDNAPIFPQGVYFGNVTENGTAGMVVMTMTAVDYDDPNE  1699
```

```
Query  141   -LNSKIAFKI---VSQEPAGTPMFLLSRNTGEVRTLTNSLDREQASSYRLVVSGADKDGE  196
               N+K+ + I   V +E  G+P+F +  +TG ++T    LDRE+   Y + V     DG
Sbjct  1700  GTNAKLIYSIEKNVIEEETGSPIFEIEADTGVIKTAVCCLDRERTPDYSIQVVAM--DGG  1757

Query  197   GLSTQCECNIKVKDVNDNFPMFRDSQYSARIEENILSS--ELLRFQVTDLDEEYTDNWLA  254
             GL      +I+VKD+ND  P F   ++   ++E   ++  E+    VT  DE+ T+ +
Sbjct  1758  GLKGTGTASIRVKDINDMPPQFTKDEWFTEVDETDGTNLPEMPILTVTVHDEDETNKFQY  1817

Query  255   VYFFTSGNEGNWFEIQTDPRTNEGI--LKVVKALDYE---QLQSVKLSIAVKNKAEFHQS  309
                  SG   + F +    R N+G   LK+V+ LDYE   Q    +  I V +K E + +
Sbjct  1818  KVIDNSGYGADKFTMV---RNNDGTGSLKIVQPLDYEDPLQSNGFRFRIQVNDKGEDNDN  1874

Query  310   VISRYRVQSTPVTIQVINVREGIAFRPASKTFTVQKGISSKKLVDYILGTYQAIDEDTNK  369
               +Y V  + V +++ ++ +    +P  +   ++  +     V   L T++A D D
Sbjct  1875  --DKYHVAYSWVVVKLRDINDN---KPQFERPNIEVSVYENADVGKTLETFKATDPDQG-  1928

Query  370   AASNVKYVMGR-NDGGYLMIDSKTAEIKFVKNMNRDSTFIVNKTITAEVLAIDE-YTGKT  427
               S V Y + R +D       ++   +   +N++R+    V       ++LAID+    KT
Sbjct  1929  GKSKVSYAIDRSSDRQRQFSINQEGTVTIQRNLDRE----VTPRHQVKILAIDDGIPPKT  1984

Query  428   STGTVYVRVPDFNDNCP  444
             +T T+ V V D NDN P
Sbjct  1985  ATATLTVIVQDINDNPP  2001

 Score = 85.9 bits (211),  Expect = 1e-15, Method: Compositional matrix adjust.
 Identities = 108/420 (26%), Positives = 177/420 (42%), Gaps = 52/420 (12%)

Query  50    IFVVDKNTGDINITAIVDREETPSFLITCRAL----------NAQGLDVEKPLILTVKIL  99
              F ++  +G+I    + DRE+  ++ +   A            N Q   V K   + + I
Sbjct  1385  FFEINLQSGEIFTKVVFDREKQGAYALEVEARDGAPSARPNSNQQPNSVTK--FIRIGIA  1442

Query  100   DINDNPPVFSQQIFMGEIEENSASNSLVMILNATDADEPNHLNSKIAFKIVSQEPAGTPM  159
             D NDNPP F + ++  E++EN       V+ + A D DE    +S+I ++I S    G
Sbjct  1443  DKNDNPPYFDKALYEAEVDENEDIQHTVLTVTAKDHDE----SSRIRYEITSGNIGGA--  1496

Query  160   FLLSRNTGEVRTLTNSLDREQASSYRLVVSGADKDGEGLSTQCECNIKVKDVNDNFPMFR  219
             F +   TG +  +  +LD E    Y L ++ +D   E  +T     I VKDVNDN P+F
Sbjct  1497  FAVKNMTGAI-YVAGALDYETRKRYELRLAASDNLKENYTTVV---IHVKDVNDNPPVFE  1552

Query  220   DSQYSARIEENI---LSSELLRFQVTDLDEEYTDNWLAVYFFTSG--NEGNWFEIQTDPR  274
                Y  +I E     L   +L+   TD D++   N   VYF T    +  N    + D
Sbjct  1553  RPTYRTQITEEDDRNLPKRVLQVTATDGDKDRPQN--IVYFLTGQGIDPDNPANSKFDIN  1610

Query  275   TNEGILKVVKALDYEQLQS-VKLSIAVKNKAEFHQSVISRYRVQSTPVTIQVINVREGIA  333
                G + V+K LD +Q     +     V  + E  + ++    VQ     + + ++ +
Sbjct  1611  RTTGEIFVLKPLDRDQPNGRPQWRFTVFAQDEGGEGLVGYADVQ-----VNLKDINDNAP  1665

Query  334   FRPASKTFTVQKGISSKKLVDYILGTYQAIDEDTNKAASNVK--YVMGRN------DGGY  385
               P    F     ++        ++ T  A+D D     +N K  Y + +N
Sbjct  1666  IFPQGVYF---GNVTENGTAGMVVMTMTAVDYDDPNEGTNAKLIYSIEKNVIEEETGSPI  1722

Query  386   LMIDSKTAEIK-FVKNMNRDSTFIVNKTITAEVLAIDEYTGKTSTGTVYVRVPDFNDNCP  444
               I++ T  IK  V  ++R+ T        + +V+A+D   G   TGT  +RV D ND  P
Sbjct  1723  FEIEADTGVIKTAVCCLDRERT----PDYSIQVVAMDG-GGLKGTGTASIRVKDINDMPP  1777

 Score = 75.9 bits (185),  Expect = 2e-12, Method: Compositional matrix adjust.
 Identities = 108/429 (25%), Positives = 178/429 (41%), Gaps = 53/429 (12%)

Query  53    VDKNTGDINITAIVDREETPSFLITCRALNAQGLDVEKPLILTVKILDINDNPPVFSQ-Q  111
             +D++  ++N+TA+ D          +C  +N            ++ V I D+NDN PVF
Sbjct  950   LDRDKYELNVTAMDDG--------SC-CVNGDATIHTSTAVVVVFITDVNDNKPVFKDCG  1000

Query  112   IFMGEIEENSASNSLVMILNATDADEPNHLNSKIAFKIVSQEPAGTPMFLLSRNTGEVRT  171
              +  ++EE + + S V+ + ATD D+    +N ++ + IV Q        F +   TGEV T
Sbjct  1001  TYYPKVEEGAPNGSPVIKVQATDEDK--GVNGQVKYSIVQQPNQKGTKFTVDEETGEVST  1058

Query  172   LTNSLDRE--QASSYRLVVSGADKDGEGLSTQCECNIKVKDVNDNFPMFRDSQYSARIEE  229
                  DRE          + V   D+     L   C    +++ DVNDN P+F    +Y    +++
```

```
Sbjct  1059  -NKVFDREGDDGKFVSVTVKATDQGEPSLEGVCSFTVEITDVNDNPPLFDRQKYVENVKQ  1117

Query  230   NI-LSSELLRFQVTDLDEEYTDNWLAVYFFT---SGNEGNWFEIQTDPRTNEGILKVVKA  285
             +  + + +LR  V+  DE+  +N   VY  +   S N+  +FEIQ  P +   +LK  K
Sbjct  1118  DASIGTNILR--VSASDEDADNNGAIVYTLSAPHSPNDLEYFEIQ--PESGWIVLK--KP  1171

Query  286   LD-YEQLQSVKLSIAVK----------NKAEFHQSVISRYRVQSTPV----TIQVINVRE  330
             LD    + SV+    +K          + A+    +    R    P+        I++RE
Sbjct  1172  LDGCYSVSSVRERYRLKVYASDRGDPPSTADVDVELDVVDRNNKPPIWDRNIYGPIHIRE  1231

Query  331   GIAFRPASKTFTVQKGISSKKLVDYILGTYQAIDEDTNKAASNVKYVMGRNDGGYLMIDS  390
              +             GI     V Y L         TNK   +  Y+  R D G
Sbjct  1232  NVTVGTVVTAIKASSGIEGNPTVFYRL--MPGSTAQTNKL--HTFYLQQRPDNG-----D  1282

Query  391   KTAEIKFVKNMNRDSTFIVNKTITAEVLAIDEYTGKTSTGTVYVRVPDFNDNCPTAV-LE  449
               A+IK    ++ ++    N TI  E     +     S  TVY+++ D ND  P     E
Sbjct  1283  TWADIKVNHPLDFETIKEYNLTIRVENNGAQQL---ASEATVYIQLEDVNDEIPLFTERE  1339

Query  450   KDAVCSSSP  458
             ++ V    P
Sbjct  1340  QETVLEGEP  1348

 Score = 61.2 bits (147),  Expect = 5e-08, Method: Compositional matrix adjust.
 Identities = 87/366 (24%), Positives = 156/366 (43%), Gaps = 38/366 (10%)

Query  51    FVVDKNTGDINITAIVDRE-ETPSFL-ITCRALNAQGLDVEKPLILTVKILDINDNPPVF  108
             F VD+ TG+++   + DRE +   F+ +T +A +     +E     TV+I D+NDNPP+F
Sbjct  1047  FTVDEETGEVSTNKVFDREGDDGKFVSVTVKATDQGEPSLEGVCSFTVEITDVNDNPPLF  1106

Query  109   SQQIFMGEIEENSASNSLVMILNATDADEPNHLNSKIAFKIVS-QEPAGTPMFLLSRNTG  167
              +Q ++  ++++++  + ++ ++A+D D  N  N  I + + +   P     F +   +G
Sbjct  1107  DRQKYVENVKQDASIGTNILRVSASDEDADN--NGAIVYTLSAPHSPNDLEYFEIQPESG  1164

Query  168   EVRTLTNSLDREQASS-----YRLVVSGADKDGEGLSTQCECNIKVKDVNDNFPMFRDSQ  222
              +  L   LD   + S     YRL V  +D+ G+  ST            +N P   D
Sbjct  1165  WI-VLKKPLDGCYSVSSVRERYRLKVYASDR-GDPPSTADVDVELDVVDRNNKPPIWDRN  1222

Query  223   YSA--RIEENILSSELLRFQVTDLDEEYTDNWLAVYFFTSG-----NEGNWFEIQTDPRT  275
                   I EN+    ++              N    Y    G     N+ + F +Q  P
Sbjct  1223  IYGPIHIRENVTVGTVV--TAIKASSGIEGNPTVFYRLMPGSTAQTNKLHTFYLQQRPDN  1280

Query  276   NE--GILKVVKALDYEQLQSVKLSIAVKNKAEFHQSVISRYRVQSTPVTIQVINVREGIA  333
              +    +KV   LD+E ++   L+I V+N          ++       V IQ+ +V + I
Sbjct  1281  GDTWADIKVNHPLDFETIKEYNLTIRVENNG-------AQQLASEATVYIQLEDVNDEIP  1333

Query  334   FRPASKTFTVQKGISSKKLVDYILGTYQAIDEDTNKAASNVKYVM---GRNDG-GYLMID  389
                  +  TV +G      V  +     AID+D     + V Y +    RN+G  +  I+
Sbjct  1334  LFTEREQETVLEGEPIGTKVTQV----NAIDKDGTFPNNQVYYYIVDSPRNEGKDFFEIN  1389

Query  390   SKTAEI  395
              ++ EI
Sbjct  1390  LQSGEI  1395

 Score = 55.8 bits (133),  Expect = 3e-06, Method: Compositional matrix adjust.
 Identities = 49/158 (31%), Positives = 78/158 (49%), Gaps = 24/158 (15%)

Query  102   NDNPPVFSQQIFMGEIEENSASNSLVMILNATDADEPNHLNSKIAFKIVSQEPAGTP--M  159
             ND  P FSQQ++   ++EN+  N+LV  + A+D D  N     + F  V     GT
Sbjct  880   NDEEPKFSQQVYTPNVDENAGPNTLVTTVVASDKDGDN-----VRFGFVG---GGTSSGQ  931

Query  160   FLLSRNTGEVRTLTN--SLDREQASSYRLVVSGADK-------DGEGLSTQCECNIKVKD  210
             F++    TG +R  +   SLDR++   Y L V+  D        D    ++     + + D
Sbjct  932   FVIEEITGVIRLHSKAISLDRDK---YELNVTAMDDGSCCVNGDATIHTSTAVVVVFITD  988

Query  211   VNDNFPMFRD-SQYSARIEENILS-SELLRFQVTDLDE  246
             VNDN P+F+D   Y  ++EE   + S +++ Q TD D+
Sbjct  989   VNDNKPVFKDCGTYYPKVEEGAPNGSPVIKVQATDEDK  1026
```

```
 Score = 53.9 bits (128),  Expect = 1e-05, Method: Compositional matrix adjust.
 Identities = 70/306 (23%), Positives = 126/306 (41%), Gaps = 30/306 (10%)

Query  20    PIAKIT-SDYQATQKITYRI---SGVGIDQPPFGIFVVDKNTGDINITAIVDRE---ETP  72
             PI  +T  D   T K  Y++   SG G D+  F +   +  TG + I   +D E   ++
Sbjct  1800  PILTVTVHDEDETNKFQYKVIDNSGYGADK--FTMVRNNDGTGSLKIVQPLDYEDPLQSN  1857

Query  73    SFLITCRALNAQGLDVEKPLI------LTVKILDINDNPPVFSQQIFMGEIEENSASNSL  126
              F    + +N +G D +          + VK+ DINDN P F +      + EN+
Sbjct  1858  GFRFRIQ-VNDKGEDNDNDKYHVAYSWVVVKLRDINDNKPQFERPNIEVSVYENADVGKT  1916

Query  127   VMILNATDADEPNHLNSKIAFKIVSQEPAGTPMFLLSRNTGEVRTLTNSLDREQASSYRL  186
             +    ATD D+     SK+++ I   + +       S N     T+  +LDRE    +++
Sbjct  1917  LETFKATDPDQGG--KSKVSYAI---DRSSDRQRQFSINQEGTVTIQRNLDREVTPRHQV  1971

Query  187   VVSGADKDGEGLSTQCECNIKVKDVNDNFPMFRDSQYSARIEENILSSELLRFQVTDLDE  246
              +   D      +      + V+D+NDN P F    Y   + E++   +++    TD D+
Sbjct  1972  KILAIDDGIPPKTATATLTVIVQDINDNPPKFL-KDYRPVLPEHVPPRKVVEILATDDDD  2030

Query  247   EYTDNWLAVYFFTSGNEGNW----FEIQTDPRTNEG----ILKVVKALDYEQLQSVKLSI  298
                 N     F       +     F+++ D +   G    I+  +++ D EQ +   + I
Sbjct  2031  RSKSNGPPFQFRLDPGADDIIRASFKVEQDQKGANGDGMAIVSSLRSFDREQQKEYLIPI  2090

Query  299   AVKNKA  304
              +K+
Sbjct  2091  VIKDHG  2096

 Score = 51.6 bits (122),  Expect = 5e-05, Method: Compositional matrix adjust.
 Identities = 96/398 (24%), Positives = 163/398 (41%), Gaps = 54/398 (14%)

Query  106   PVFSQQIFMGEIEENSASNSLVMILNATDADEPNHLNSKIAFKIVSQ-EPAGTPMFLLSR  164
             P F    +  EI EN   +S ++ + A      +  + +I + + +Q + AGT  F +
Sbjct  677   PQFYMPSYEAEIPENQKKDSDIISVKAK-----SFADREIRYTLKAQGQGAGT--FNIGP  729

Query  165   NTGEVRTLTNSLDRE---QASSYRLVVSGADKDGEGLSTQCECNIKVKDVNDNFPMFRDS  221
              +G V+ L   LD E   Q   Y LVV+ A +D  G ST  E  I+V DVNDN P F
Sbjct  730   TSGIVK-LAKELDFEDLRQPHVYSLVVT-ATEDSGGFSTSVELTIRVTDVNDNAPKFELP  787

Query  222   QYSAR-IEENI-LSSELLRFQVTDLDEEYTDNWLAVYFFTSGNEGNWFEIQTDPRTNEGI  279
              Y A  ++E+I + + +L+ +  D D         + +  S +    F + ++     GI
Sbjct  788   DYQAHNVDEDIQIGTSILKVKAMDADSGAN---AEIEYLVSDDH---FAVDSN-----GI  836

Query  280   LKVVKALDYEQLQSV-KLSIAVKNKAEFHQSVISRYRVQSTPVTIQVINVREGIAFRPAS  338
             +   K LD +   +  +  +  K+K E  ++  +  RV +           E   F
Sbjct  837   IVNNKQLDADNNNAYYEFVVTAKDKGEPAKTGTATVRVYTKNKN------DEEPKFSQQV  890

Query  339   KTFTVQKGISSKKLVDYILGTYQAIDEDTNKAASNVKY--VMGRNDGGYLMIDSKTAEIK  396
              T  V+       LV  ++ +        +K   NV++  V G    G  +I+  T  I+
Sbjct  891   YTPNVDENAGPNTLVTTVVAS--------DKDGDNVRFGFVGGGTSSGQFVIEEITGVIR  942

Query  397   FVK---NMNRDSTFI-VNKTITAEVLAIDEYTGKTSTGTVYVRVPDFNDN------CPTA  446
                   +++RD   + V            + T  TST  V V + D NDN      C T
Sbjct  943   LHSKAISLDRDKYELNVTAMDDGSCCVNGDATIHTSTAVVVVFITDVNDNKPVFKDCGTY  1002

Query  447   VLE-KDAVCSSSPSVVVSARTLNNRYTGPYTFALEDQP  483
               + ++   + SP + V A   +    G   +++  QP
Sbjct  1003  YPKVEEGAPNGSPVIKVQATDEDKGVNGQVKYSIVQQP  1040

 Score = 44.7 bits (104),  Expect = 0.008, Method: Compositional matrix adjust.
 Identities = 48/199 (24%), Positives = 73/199 (37%), Gaps = 19/199 (10%)

Query  27    DYQATQKITYRISGVGIDQPPFGIFVVDKNTGDINITAIVDREETPSFLITCRALNAQGL  86
             D     K++Y I      Q  F I       G + I   +DRE TP   +   A++
Sbjct  1926  DQGGKSKVSYAIDRSSDRQRQFSI----NQEGTVTIQRNLDREVTPRHQVKILAIDDGIP  1981

Query  87    DVEKPLILTVKILDINDNPPVFSQQIFMGEIEENSASNSLVMILNATDADEPNHLNSKIA  146
                    LTV + DINDNPP F +  +   + E+     +V IL   D D
Sbjct  1982  PKTATATLTVIVQDINDNPPKFLKD-YRPVLPEHVPPRKVVEILATDDDDRSKSNGPPFQ  2040
```

```
Query  147   FKIVSQEPAGTPMFLLSRNTGEVRTLTN-----------SLDREQASSYRLVVSGADKDG  195
             F++  +P     +   S    + +   N           S DREQ   Y + +   D
Sbjct  2041  FRL---DPGADDIIRASFKVEQDQKGANGDGMAIVSSLRSFDREQQKEYLIPIVIKDHGN  2097

Query  196   EGLSTQCECNIKVKDVNDN  214
               ++      + + DVNDN
Sbjct  2098  PAMTGTSTLTVVIGDVNDN  2116

 Score = 40.8 bits (94),  Expect = 0.11, Method: Compositional matrix adjust.
 Identities = 55/242 (23%), Positives = 102/242 (42%), Gaps = 22/242 (9%)

Query  4    KFAKPCREGE--DNSKRNP-IAKITSDYQATQKITYRISGVGIDQPPFGIFVVDKNTGDI  60
            +F  P  E E  +N K++  I  + +   A ++I Y +   G      G F +   +G +
Sbjct  678  QFYMPSYEAEIPENQKKDSDIISVKAKSFADREIRYTLKAQGQGA---GTFNIGPTSGIV  734

Query  61   NITAIVDREE--TPSF--LITCRALNAQGLDVEKPLILTVKILDINDNPPVFSQQIFMGE  116
             +   +D E+   P    L+     ++ G      + LT+++ D+NDN P F    +
Sbjct  735  KLAKELDFEDLRQPHVYSLVVTATEDSGGFSTS--VELTIRVTDVNDNAPKFELPDYQAH  792

Query  117  -IEENSASNSLVMILNATDADEPNHLNSKIAFKIVSQEPAGTPMFLLSRNTGEVRTLTNS  175
             ++E+    + ++ + A DAD  +  N++I + +VS +      F +  N   V
Sbjct  793  NVDEDIQIGTSILKVKAMDAD--SGANAEIEY-LVSDD-----HFAVDSNGIIVNNKQLD  844

Query  176  LDREQASSYRLVVSGADKDGEGLSTQCECNIKVKDVNDNFPMFRDSQYSARIEENILSSE  235
             D   A  Y  VV+  DK     +      +  K+ ND  P F    Y+  ++EN   +
Sbjct  845  ADNNNA-YYEFVVTAKDKGEPAKTGTATVRVYTKNKNDEEPKFSQQVYTPNVDENAGPNT  903

Query  236  LL  237
            L+
Sbjct  904  LV  905

 Score = 39.3 bits (90),  Expect = 0.30, Method: Compositional matrix adjust.
 Identities = 55/225 (24%), Positives = 88/225 (39%), Gaps = 24/225 (11%)

Query  75   LITCRALNAQGLDVEKPLILTVKILDINDNPPVFSQQIF--MGEIEENSASNSLVMILNA  132
            +IT +  NA GL       + + + D+ND PP F  +       ++ N+  N+ V  L A
Sbjct  539  IITNQGHNA-GLKYTDNQRVIIHVKDVNDEPPYFINRPLPMQAVVQLNAPPNTPVFTLQA  597

Query  133  TDADEPNHLNSKIAFKIVSQEPAGTPMFLLSRNTGEVRTLTNSLDREQASSYRLVVSGAD  192
             D D     +  I + IV     G   F +   +G VRT    L  +    Y L V   D
Sbjct  598  RDPD----TDHNIHYFIVRDRTGG--RFEVDERSGVVRTRGTDL-FQLDMEYVLYVKAED  650

Query  193  KDGE----GLSTQCECNIKVKDVNDNFPMFRDSQYSARIEEN-ILSSELLRFQVTDLDEE  247
            ++G+         +  E  + +       P F    Y A I EN    S+++  +     +
Sbjct  651  QNGKVDDRRFQSTPEERLSIVG-GKRAPQFYMPSYEAEIPENQKKDSDIISVKA----KS  705

Query  248  YTDNWLAVYFFTSGNEGNWFEIQTDPRTNEGILKVVKALDYEQLQ  292
            + D  +       G     F I        GI+K+ K LD+E L+
Sbjct  706  FADREIRYTLKAQGQGAGTFNIG----PTSGIVKLAKELDFEDLR  746

>AALB20_033171.P66500 | transcript=AALB20_033171.R66500 | gene=AALB20_033171
| organism=Anopheles_albimanus_STECLA_2020 |
gene_product=neural-cadherin, transcript variant X38 |
transcript_product=neural-cadherin,
transcript variant X7 | location=CM024292:75037560-75208393(+)
| protein_length=3131 | sequence_SO=chromosome
| SO=protein_coding_gene | is_pseudo=false
Length=3131

 Score = 144 bits (364),  Expect = 1e-34, Method: Compositional matrix adjust.
 Identities = 125/437 (29%), Positives = 211/437 (48%), Gaps = 33/437 (8%)

Query  26    SDYQATQKITYRISGVGID--QPPFGIFVVDKNTGDINITAIVDREET---PSFLITCRA  80
              D    Q I Y ++G GID   P    F +++ TG+I +   +DR++    P +  T  A
Sbjct  1580  GDKDRPQNIVYFLTGQGIDPDNPANSKFDINRTTGEIFVLKPLDRDQPNGRPQWRFTVFA  1639

Query  81    LNAQGLDVEKPLILTVKILDINDNPPVFSQQIFMGEIEENSASNSLVMILNATDADEPNH  140
               +  G  +      + V + DINDN P+F Q ++ G + EN  +  +VM + A D D+PN
```

```
Sbjct  1640  QDEGGEGLVGYADVQVNLKDINDNAPIFPQGVYFGNVTENGTAGMVVMTMTAVDYDDPNE  1699

Query  141   -LNSKIAFKI---VSQEPAGTPMFLLSRNTGEVRTLTNSLDREQASSYRLVVSGADKDGE  196
               N+K+ + I   V +E  G+P+F +  +TG ++T    LDRE+   Y + V     DG
Sbjct  1700  GTNAKLIYSIEKNVIEEETGSPIFEIEADTGVIKTAVCCLDRERTPDYSIQVVAM--DGG  1757

Query  197   GLSTQCECNIKVKDVNDNFPMFRDSQYSARIEENILSS--ELLRFQVTDLDEEYTDNWLA  254
             GL      +I+VKD+ND  P F   ++   ++E   ++  E+    VT  DE+ T+ +
Sbjct  1758  GLKGTGTASIRVKDINDMPPQFTKDEWFTEVDETDGTNLPEMPILTVTVHDEDETNKFQY  1817

Query  255   VYFFTSGNEGNWFEIQTDPRTNEGI--LKVVKALDYE---QLQSVKLSIAVKNKAEFHQS  309
                  SG   + F +    R N+G   LK+V+ LDYE   Q    +  I V +K E + +
Sbjct  1818  KVIDNSGYGADKFTMV---RNNDGTGSLKIVQPLDYEDPLQSNGFRFRIQVNDKGEDNDN  1874

Query  310   VISRYRVQSTPVTIQVINVREGIAFRPASKTFTVQKGISSKKLVDYILGTYQAIDEDTNK  369
                +Y V  + V +++ ++ +    +P  +   ++  +     V   L T++A D D
Sbjct  1875  --DKYHVAYSWVVVKLRDINDN---KPQFERPNIEVSVYENADVGKTLETFKATDPDQG-  1928

Query  370   AASNVKYVMGR-NDGGYLMIDSKTAEIKFVKNMNRDSTFIVNKTITAEVLAIDE-YTGKT  427
               S V Y + R +D       ++   +   +N++R+    V      ++LAID+    KT
Sbjct  1929  GKSKVSYAIDRSSDRQRQFSINQEGTVTIQRNLDRE----VTPRHQVKILAIDDGIPPKT  1984

Query  428   STGTVYYVRVPDFNDNCP  444
             +T T+ V V D NDN P
Sbjct  1985  ATATLTVIVQDINDNPP  2001

 Score = 85.9 bits (211),  Expect = 1e-15, Method: Compositional matrix adjust.
 Identities = 108/420 (26%), Positives = 177/420 (42%), Gaps = 52/420 (12%)

Query  50    IFVVDKNTGDINITAIVDREETPSFLITCRAL-----------NAQGLDVEKPLILTVKIL  99
              F ++  +G+I    + DRE+  ++ +   A           N Q   V K   + + I
Sbjct  1385  FFEINLQSGEIFTKVVFDREKQGAYALEVEARDGAPSARPNSNQQPNSVTK--FIRIGIA  1442

Query  100   DINDNPPVFSQQIFMGEIEENSASNSLVMILNATDADEPNHLNSKIAFKIVSQEPAGTPM  159
             D NDNPP F + ++  E++EN      V+ + A D DE    +S+I ++I S    G
Sbjct  1443  DKNDNPPYFDKALYEAEVDENEDIQHTVLTVTAKDHDE----SSRIRYEITSGNIGGA--  1496

Query  160   FLLSRNTGEVRTLTNSLDREQASSYRLVVSGADKDGEGLSTQCECNIKVKDVNDNFPMFR  219
             F +   TG +  +  +LD E    Y L ++ +D   E  +T     I VKDVNDN P+F
Sbjct  1497  FAVKNMTGAI-YVAGALDYETRKRYELRLAASDNLKENYTTVV---IHVKDVNDNPPVFE  1552

Query  220   DSQYSARIEENI---LSSELLRFQVTDLDEEYTDNWLAVYFFTSG--NEGNWFEIQTDPR  274
                Y  +I E     L   +L+   TD D++   N   VYF T    +  N    + D
Sbjct  1553  RPTYRTQITEEDDRNLPKRVLQVTATDGDKDRPQN--IVYFLTGQGIDPDNPANSKFDIN  1610

Query  275   TNEGILKVVKALDYEQLQS-VKLSIAVKNKAEFHQSVISRYRVQSTPVTIQVINVREGIA  333
                G + V+K LD +Q     +    V  + E  + ++    VQ     + + ++ +
Sbjct  1611  RTTGEIFVLKPLDRDQPNGRPQWRFTVFAQDEGGEGLVGYADVQ-----VNLKDINDNAP  1665

Query  334   FRPASKTFTVQKGISSKKLVDYILGTYQAIDEDTNKAASNVK--YVMGRN------DGGY  385
               P    F     ++       ++ T  A+D D     +N K  Y + +N
Sbjct  1666  IFPQGVYF---GNVTENGTAGMVVMTMTAVDYDDPNEGTNAKLIYSIEKNVIEEETGSPI  1722

Query  386   LMIDSKTAEIK-FVKNMNRDSTFIVNKTITAEVLAIDEYTGKTSTGTVYYVRVPDFNDNCP  444
               I++ T  IK  V  ++R+ T       + +V+A+D   G   TGT  +RV D ND  P
Sbjct  1723  FEIEADTGVIKTAVCCLDRERT----PDYSIQVVAMDG-GGLKGTGTASIRVKDINDMPP  1777

 Score = 75.9 bits (185),  Expect = 2e-12, Method: Compositional matrix adjust.
 Identities = 108/429 (25%), Positives = 178/429 (41%), Gaps = 53/429 (12%)

Query  53    VDKNTGDINITAIVDREETPSFLITCRALNAQGLDVEKPLILTVKILDINDNPPVFSQ-Q  111
             +D++  ++N+TA+ D         +C  +N          ++ V I D+NDN PVF
Sbjct  950   LDRDKYELNVTAMDDG--------SC-CVNGDATIHTSTAVVVVFITDVNDNKPVFKDCG  1000

Query  112   IFMGEIEENSASNSLVMILNATDADEPNHLNSKIAFKIVSQEPAGTPMFLLSRNTGEVRT  171
              +  ++EE + + S V+ + ATD D+   +N ++ + IV Q       F +   TGEV T
Sbjct  1001  TYYPKVEEGAPNGSPVIKVQATDEDK--GVNGQVKYSIVQQPNQKGTKFTVDEETGEVST  1058

Query  172   LTNSLDRE--QASSYRLVVSGADKDGEGLSTQCECNIKVKDVNDNFPMFRDSQYSARIEE  229
```

```
                  DRE        + V   D+    L   C   +++ DVNDN P+F   +Y   +++
Sbjct  1059  -NKVFDREGDDGKFVSVTVKATDQGEPSLEGVCSFTVEITDVNDNPPLFDRQKYVENVKQ  1117

Query  230   NI-LSSELLRFQVTDLDEEYTDNWLAVYFFT---SGNEGNWFEIQTDPRTNEGILKVVKA  285
             +  + + +LR  V+  DE+  +N   VY  +   S N+  +FEIQ  P +   +LK  K
Sbjct  1118  DASIGTNILR--VSASDEDADNNGAIVYTLSAPHSPNDLEYFEIQ--PESGWIVLK--KP  1171

Query  286   LD-YEQLQSVKLSIAVK----------NKAEFHQSVISRYRVQSTPV----TIQVINVRE  330
             LD    + SV+    +K          + A+    +    R    P+        I++RE
Sbjct  1172  LDGCYSVSSVRERYRLKVYASDRGDPPSTADVDVELDVVDRNNKPPIWDRNIYGPIHIRE  1231

Query  331   GIAFRPASKTFTVQKGISSKKLVDYILGTYQAIDEDTNKAASNVKYVMGRNDGGYLMIDS  390
              +             GI     V Y L         TNK   +  Y+  R D G
Sbjct  1232  NVTVGTVVTAIKASSGIEGNPTVFYRL--MPGSTAQTNKL--HTFYLQQRPDNG-----D  1282

Query  391   KTAEIKFVKNMNRDSTFIVNKTITAEVLAIDEYTGKTSTGTVYVRVPDFNDNCPTAV-LE  449
               A+IK    ++ ++    N TI  E     +     S  TVY+++ D ND  P     E
Sbjct  1283  TWADIKVNHPLDFETIKEYNLTIRVENNGAQQL---ASEATVYIQLEDVNDEIPLFTERE  1339

Query  450   KDAVCSSSP  458
             ++ V    P
Sbjct  1340  QETVLEGEP  1348

 Score = 61.2 bits (147),  Expect = 5e-08, Method: Compositional matrix adjust.
 Identities = 87/366 (24%), Positives = 156/366 (43%), Gaps = 38/366 (10%)

Query  51    FVVDKNTGDINITAIVDRE-ETPSFL-ITCRALNAQGLDVEKPLILTVKILDINDNPPVF  108
             F VD+ TG+++   + DRE +   F+ +T +A +     +E     TV+I D+NDNPP+F
Sbjct  1047  FTVDEETGEVSTNKVFDREGDDGKFVSVTVKATDQGEPSLEGVCSFTVEITDVNDNPPLF  1106

Query  109   SQQIFMGEIEENSASNSLVMILNATDADEPNHLNSKIAFKIVS-QEPAGTPMFLLSRNTG  167
              +Q ++  ++++++  + ++ ++A+D D  N  N  I + + +   P     F +   +G
Sbjct  1107  DRQKYVENVKQDASIGTNILRVSASDEDADN--NGAIVYTLSAPHSPNDLEYFEIQPESG  1164

Query  168   EVRTLTNSLDREQASS-----YRLVVSGADKDGEGLSTQCECNIKVKDVNDNFPMFRDSQ  222
              +  L   LD   + S     YRL V  +D+ G+  ST            +N P   D
Sbjct  1165  WI-VLKKPLDGCYSVSSVRERYRLKVYASDR-GDPPSTADVDVELDVVDRNNKPPIWDRN  1222

Query  223   YSA--RIEENILSSELLRFQVTDLDEEYTDNWLAVYFFTSG-----NEGNWFEIQTDPRT  275
                   I EN+    ++              N    Y    G     N+ + F +Q  P
Sbjct  1223  IYGPIHIRENVTVGTVV--TAIKASSGIEGNPTVFYRLMPGSTAQTNKLHTFYLQQRPDN  1280

Query  276   NE--GILKVVKALDYEQLQSVKLSIAVKNKAEFHQSVISRYRVQSTPVTIQVINVREGIA  333
              +    +KV   LD+E ++   L+I V+N         ++        V IQ+ +V + I
Sbjct  1281  GDTWADIKVNHPLDFETIKEYNLTIRVENNG-------AQQLASEATVYIQLEDVNDEIP  1333

Query  334   FRPASKTFTVQKGISSKKLVDYILGTYQAIDEDTNKAASNVKYVM---GRNDG-GYLMID  389
                  +  TV +G      V  +     AID+D     + V Y +    RN+G  +  I+
Sbjct  1334  LFTEREQETVLEGEPIGTKVTQV----NAIDKDGTFPNNQVYYYIVDSPRNEGKDFFEIN  1389

Query  390   SKTAEI  395
              ++ EI
Sbjct  1390  LQSGEI  1395

 Score = 55.8 bits (133),  Expect = 3e-06, Method: Compositional matrix adjust.
 Identities = 49/158 (31%), Positives = 78/158 (49%), Gaps = 24/158 (15%)

Query  102   NDNPPVFSQQIFMGEIEENSASNSLVMILNATDADEPNHLNSKIAFKIVSQEPAGTP--M  159
             ND  P FSQQ++   ++EN+  N+LV  + A+D D  N     + F  V     GT
Sbjct  880   NDEEPKFSQQVYTPNVDENAGPNTLVTTVVASDKDGDN-----VRFGFVG---GGTSSGQ  931

Query  160   FLLSRNTGEVRTLTN--SLDREQASSYRLVVSGADK-------DGEGLSTQCECNIKVKD  210
             F++   TG +R  +   SLDR++   Y L V+  D        D    ++      + + D
Sbjct  932   FVIEEITGVIRLHSKAISLDRDK---YELNVTAMDDGSCCVNGDATIHTSTAVVVVFITD  988

Query  211   VNDNFPMFRD-SQYSARIEENILS-SELLRFQVTDLDE  246
             VNDN P+F+D   Y  ++EE   + S +++ Q TD D+
Sbjct  989   VNDNKPVFKDCGTYYPKVEEGAPNGSPVIKVQATDEDK  1026
```

```
 Score = 53.9 bits (128),  Expect = 1e-05, Method: Compositional matrix adjust.
 Identities = 70/306 (23%), Positives = 126/306 (41%), Gaps = 30/306 (10%)

Query  20    PIAKIT-SDYQATQKITYRI---SGVGIDQPPFGIFVVDKNTGDINITAIVDRE---ETP  72
             PI  +T  D   T K  Y++   SG G D+  F +   +  TG + I   +D E   ++
Sbjct  1800  PILTVTVHDEDETNKFQYKVIDNSGYGADK--FTMVRNNDGTGSLKIVQPLDYEDPLQSN  1857

Query  73    SFLITCRALNAQGLDVEKPLI------LTVKILDINDNPPVFSQQIFMGEIEENSASNSL  126
              F    + +N +G D +          + VK+ DINDN P F +      + EN+
Sbjct  1858  GFRFRIQ-VNDKGEDNDNDKYHVAYSWVVVKLRDINDNKPQFERPNIEVSVYENADVGKT  1916

Query  127   VMILNATDADEPNHLNSKIAFKIVSQEPAGTPMFLLSRNTGEVRTLTNSLDREQASSYRL  186
             +    ATD D+     SK+++ I   + +       S N     T+  +LDRE    +++
Sbjct  1917  LETFKATDPDQGG--KSKVSYAI---DRSSDRQRQFSINQEGTVTIQRNLDREVTPRHQV  1971

Query  187   VVSGADKDGEGLSTQCECNIKVKDVNDNFPMFRDSQYSARIEENILSSELLRFQVTDLDE  246
              +   D      +      + V+D+NDN P F    Y   + E++   +++    TD D+
Sbjct  1972  KILAIDDGIPPKTATATLTVIVQDINDNPPKFL-KDYRPVLPEHVPPRKVVEILATDDDD  2030

Query  247   EYTDNWLAVYFFTSGNEGNW----FEIQTDPRTNEG----ILKVVKALDYEQLQSVKLSI  298
                 N     F       +     F+++ D +   G    I+  +++ D EQ +   + I
Sbjct  2031  RSKSNGPPFQFRLDPGADDIIRASFKVEQDQKGANGDGMAIVSSLRSFDREQQKEYLIPI  2090

Query  299   AVKNKA  304
              +K+
Sbjct  2091  VIKDHG  2096

 Score = 51.6 bits (122),  Expect = 5e-05, Method: Compositional matrix adjust.
 Identities = 96/398 (24%), Positives = 163/398 (41%), Gaps = 54/398 (14%)

Query  106   PVFSQQIFMGEIEENSASNSLVMILNATDADEPNHLNSKIAFKIVSQ-EPAGTPMFLLSR  164
             P F    +  EI EN   +S ++ + A      +  + +I + + +Q + AGT  F +
Sbjct  677   PQFYMPSYEAEIPENQKKDSDIISVKAK-----SFADREIRYTLKAQGQGAGT--FNIGP  729

Query  165   NTGEVRTLTNSLDRE---QASSYRLVVSGADKDGEGLSTQCECNIKVKDVNDNFPMFRDS  221
              +G V+ L   LD E   Q   Y LVV+ A +D  G ST  E  I+V DVNDN P F
Sbjct  730   TSGIVK-LAKELDFEDLRQPHVYSLVVT-ATEDSGGFSTSVELTIRVTDVNDNAPKFELP  787

Query  222   QYSAR-IEENI-LSSELLRFQVTDLDEEYTDNWLAVYFFTSGNEGNWFEIQTDPRTNEGI  279
              Y A  ++E+I + + +L+ +  D D         + +  S +    F + ++     GI
Sbjct  788   DYQAHNVDEDIQIGTSILKVKAMDADSGAN---AEIEYLVSDDH---FAVDSN-----GI  836

Query  280   LKVVKALDYEQLQSV-KLSIAVKNKAEFHQSVISRYRVQSTPVTIQVINVREGIAFRPAS  338
             +   K LD +   +  +  +  K+K E  ++  +  RV +           E   F
Sbjct  837   IVNNKQLDADNNNAYYEFVVTAKDKGEPAKTGTATVRVYTKNKN------DEEPKFSQQV  890

Query  339   KTFTVQKGISSKKLVDYILGTYQAIDEDTNKAASNVKY--VMGRNDGGYLMIDSKTAEIK  396
              T  V +      LV  ++ +        +K   NV++  V G    G  +I+  T  I+
Sbjct  891   YTPNVDENAGPNTLVTTVVAS--------DKDGDNVRFGFVGGGTSSGQFVIEEITGVIR  942

Query  397   FVK---NMNRDSTFI-VNKTITAEVLAIDEYTGKTSTGTVYVRVPDFNDN------CPTA  446
                   +++RD   + V            + T  TST  V V + D NDN      C T
Sbjct  943   LHSKAISLDRDKYELNVTAMDDGSCCVNGDATIHTSTAVVVVFITDVNDNKPVFKDCGTY  1002

Query  447   VLE-KDAVCSSSSPSVVVSARTLNNRYTGPYTFALEDQP  483
               + ++   + SP + V A    +   G   +++  QP
Sbjct  1003  YPKVEEGAPNGSPVIKVQATDEDKGVNGQVKYSIVQQP  1040

 Score = 44.7 bits (104),  Expect = 0.008, Method: Compositional matrix adjust.
 Identities = 48/199 (24%), Positives = 73/199 (37%), Gaps = 19/199 (10%)

Query  27    DYQATQKITYRISGVGIDQPPFGIFVVDKNTGDINITAIVDREETPSFLITCRALNAQGL  86
             D     K++Y I      Q  F I       G + I   +DRE TP   +   A++
Sbjct  1926  DQGGKSKVSYAIDRSSDRQRQFSI----NQEGTVTIQRNLDREVTPRHQVKILAIDDGIP  1981

Query  87    DVEKPLILTVKILDINDNPPVFSQQIFMGEIEENSASNSLVMILNATDADEPNHLNSKIA  146
                    LTV + DINDNPP F +  +   + E+     +V IL   D D
Sbjct  1982  PKTATATLTVIVQDINDNPPKFLKD-YRPVLPEHVPPRKVVEILATDDDDRSKSNGPPFQ  2040
```

```
Query  147   FKIVSQEPAGTPMFLLSRNTGEVRTLTN-----------SLDREQASSYRLVVSGADKDG  195
             F++   +P    +   S    + +   N           S DREQ   Y + +   D
Sbjct  2041  FRL---DPGADDIIRASFKVEQDQKGANGDGMAIVSSLRSFDREQQKEYLIPIVIKDHGN  2097

Query  196   EGLSTQCECNIKVKDVNDN  214
               ++       + + DVNDN
Sbjct  2098  PAMTGTSTLTVVIGDVNDN  2116

 Score = 40.8 bits (94),  Expect = 0.11, Method: Compositional matrix adjust.
 Identities = 55/242 (23%), Positives = 102/242 (42%), Gaps = 22/242 (9%)

Query  4    KFAKPCREGE--DNSKRNP-IAKITSDYQATQKITYRISGVGIDQPPFGIFVVDKNTGDI  60
            +F  P  E E  +N K++  I  + +   A ++I Y +   G       G F +   +G +
Sbjct  678  QFYMPSYEAEIPENQKKDSDIISVKAKSFADREIRYTLKAQGQGA---GTFNIGPTSGIV  734

Query  61   NITAIVDREE--TPSF--LITCRALNAQGLDVEKPLILTVKILDINDNPPVFSQQIFMGE  116
             +   +D E+   P    L+     ++ G       + LT+++ D+NDN P F    +
Sbjct  735  KLAKELDFEDLRQPHVYSLVVTATEDSGGFSTS--VELTIRVTDVNDNAPKFELPDYQAH  792

Query  117  -IEENSASNSLVMILNATDADEPNHLNSKIAFKIVSQEPAGTPMFLLSRNTGEVRTLTNS  175
             ++E+    + ++ + A DAD  +  N++I + +VS +      F +  N   V
Sbjct  793  NVDEDIQIGTSILKVKAMDAD--SGANAEIEY-LVSDD-----HFAVDSNGIIVNNKQLD  844

Query  176  LDREQASSYRLVVSGADKDGEGLSTQCECNIKVKDVNDNFPMFRDSQYSARIEENILSSE  235
             D   A  Y  VV+  DK     +      +  K+ ND  P F    Y+  ++EN   +
Sbjct  845  ADNNNA-YYEFVVTAKDKGEPAKTGTATVRVYTKNKNDEEPKFSQQVYTPNVDENAGPNT  903

Query  236  LL  237
            L+
Sbjct  904  LV  905

 Score = 39.3 bits (90),  Expect = 0.30, Method: Compositional matrix adjust.
 Identities = 55/225 (24%), Positives = 88/225 (39%), Gaps = 24/225 (11%)

Query  75   LITCRALNAQGLDVEKPLILTVKILDINDNPPVFSQQIF--MGEIEENSASNSLVMILNA  132
            +IT +  NA GL       + + + D+ND PP F  +       ++ N+  N+ V  L A
Sbjct  539  IITNQGHNA-GLKYTDNQRVIIHVKDVNDEPPYFINRPLPMQAVVQLNAPPNTPVFTLQA  597

Query  133  TDADEPNHLNSKIAFKIVSQEPAGTPMFLLSRNTGEVRTLTNSLDREQASSYRLVVSGAD  192
             D D     +  I + IV     G   F +   +G VRT    L  +    Y L V   D
Sbjct  598  RDPD----TDHNIHYFIVRDRTGG--RFEVDERSGVVRTRGTDL-FQLDMEYVLYVKAED  650

Query  193  KDGE----GLSTQCECNIKVKDVNDNFPMFRDSQYSARIEEN-ILSSELLRFQVTDLDEE  247
            ++G+        +  E  + +        P F    Y A I EN    S+++  +     +
Sbjct  651  QNGKVDDRRFQSTPEERLSIVG-GKRAPQFYMPSYEAEIPENQKKDSDIISVKA----KS  705

Query  248  YTDNWLAVYFFTSGNEGNWFEIQTDPRTNEGILKVVKALDYEQLQ  292
            + D  +       G     F I        GI+K+ K LD+E L+
Sbjct  706  FADREIRYTLKAQGQGAGTFNIG----PTSGIVKLAKELDFEDLR  746

>AALB20_033171.P66499 | transcript=AALB20_033171.R66499 | gene=AALB20_033171
| organism=Anopheles_albimanus_STECLA_2020 |
gene_product=neural-cadherin, transcript variant X38 |
transcript_product=neural-cadherin,
transcript variant X16 | location=CM024292:75037560-75208393(+)
| protein_length=3131 | sequence_SO=chromosome
| SO=protein_coding_gene | is_pseudo=false
Length=3131

 Score = 144 bits (364),  Expect = 1e-34, Method: Compositional matrix adjust.
 Identities = 125/437 (29%), Positives = 211/437 (48%), Gaps = 33/437 (8%)

Query  26    SDYQATQKITYRISGVGID--QPPFGIFVVDKNTGDINITAIVDREET---PSFLITCRA  80
               D    Q I Y ++G GID   P    F +++ TG+I +   +DR++    P +  T  A
Sbjct  1580  GDKDRPQNIVYFLTGQGIDPDNPANSKFDINRTTGEIFVLKPLDRDQPNGRPQWRFTVFA  1639

Query  81    LNAQGLDVEKPLILTVKILDINDNPPVFSQQIFMGEIEENSASNSLVMILNATDADEPNH  140
```

```
              +  G  +     + V + DINDN P+F Q ++ G + EN  +  +VM + A D D+PN
Sbjct  1640  QDEGGEGLVGYADVQVNLKDINDNAPIFPQGVYFGNVTENGTAGMVVMTMTAVDYDDPNE  1699

Query  141   -LNSKIAFKI---VSQEPAGTPMFLLSRNTGEVRTLTNSLDREQASSYRLVVSGADKDGE  196
               N+K+ + I   V +E  G+P+F +  +TG ++T    LDRE+   Y + V     DG
Sbjct  1700  GTNAKLIYSIEKNVIEEETGSPIFEIEADTGVIKTAVCCLDRERTPDYSIQVVAM--DGG  1757

Query  197   GLSTQCECNIKVKDVNDNFPMFRDSQYSARIEENILSS--ELLRFQVTDLDEEYTDNWLA  254
             GL      +I+VKD+ND  P F   ++   ++E   ++  E+    VT  DE+ T+ +
Sbjct  1758  GLKGTGTASIRVKDINDMPPQFTKDEWFTEVDETDGTNLPEMPILTVTVHDEDETNKFQY  1817

Query  255   VYFFTSGNEGNWFEIQTDPRTNEGI--LKVVKALDYE---QLQSVKLSIAVKNKAEFHQS  309
                  SG   + F +    R N+G   LK+V+ LDYE   Q    +  I V +K E + +
Sbjct  1818  KVIDNSGYGADKFTMV---RNNDGTGSLKIVQPLDYEDPLQSNGFRFRIQVNDKGEDNDN  1874

Query  310   VISRYRVQSTPVTIQVINVREGIAFRPASKTFTVQKGISSKKLVDYILGTYQAIDEDTNK  369
                +Y V  + V +++ ++ +    +P  +   ++  +     V   L T++A D D
Sbjct  1875  --DKYHVAYSWVVVKLRDINDN---KPQFERPNIEVSVYENADVGKTLETFKATDPDQG-  1928

Query  370   AASNVKYVMGR-NDGGYLMIDSKTAEIKFVKNMNRDSTFIVNKTITAEVLAIDE-YTGKT  427
               S V Y + R +D       ++   +   +N++R+    V      ++LAID+    KT
Sbjct  1929  GKSKVSYAIDRSSDRQRQFSINQEGTVTIQRNLDRE----VTPRHQVKILAIDDGIPPKT  1984

Query  428   STGTVYVRVPDFNDNCP  444
             +T T+ V V D NDN P
Sbjct  1985  ATATLTVIVQDINDNPP  2001

 Score = 85.9 bits (211),  Expect = 1e-15, Method: Compositional matrix adjust.
 Identities = 108/420 (26%), Positives = 177/420 (42%), Gaps = 52/420 (12%)

Query  50    IFVVDKNTGDINITAIVDREETPSFLITCRAL----------NAQGLDVEKPLILTVKIL  99
              F ++  +G+I    + DRE+  ++ +   A            N Q   V K   + + I
Sbjct  1385  FFEINLQSGEIFTKVVFDREKQGAYALEVEARDGAPSARPNSNQQPNSVTK--FIRIGIA  1442

Query  100   DINDNPPVFSQQIFMGEIEENSASNSLVMILNATDADEPNHLNSKIAFKIVSQEPAGTPM  159
             D NDNPP F + ++  E++EN      V+ + A D DE    +S+I ++I S    G
Sbjct  1443  DKNDNPPYFDKALYEAEVDENEDIQHTVLTVTAKDHDE----SSRIRYEITSGNIGGA--  1496

Query  160   FLLSRNTGEVRTLTNSLDREQASSYRLVVSGADKDGEGLSTQCECNIKVKDVNDNFPMFR  219
             F +   TG +  +  +LD E    Y L ++ +D   E  +T     I VKDVNDN P+F
Sbjct  1497  FAVKNMTGAI-YVAGALDYETRKRYELRLAASDNLKENYTTVV---IHVKDVNDNPPVFE  1552

Query  220   DSQYSARIEENI---LSSELLRFQVTDLDEEYTDNWLAVYFFTSG--NEGNWFEIQTDPR  274
                Y  +I E     L  +L+   TD D++   N   VYF T    +  N    + D
Sbjct  1553  RPTYRTQITEEDDRNLPKRVLQVTATDGDKDRPQN--IVYFLTGQGIDPDNPANSKFDIN  1610

Query  275   TNEGILKVVKALDYEQLQS-VKLSIAVKNKAEFHQSVISRYRVQSTPVTIQVINVREGIA  333
                G + V+K LD +Q     +    V  + E  + ++    VQ     + + ++ +
Sbjct  1611  RTTGEIFVLKPLDRDQPNGRPQWRFTVFAQDEGGEGLVGYADVQ-----VNLKDINDNAP  1665

Query  334   FRPASKTFTVQKGISSKKLVDYILGTYQAIDEDTNKAASNVK--YVMGRN------DGGY  385
               P    F     ++        ++ T  A+D D     +N K  Y + +N
Sbjct  1666  IFPQGVYF---GNVTENGTAGMVVMTMTAVDYDDPNEGTNAKLIYSIEKNVIEEETGSPI  1722

Query  386   LMIDSKTAEIK-FVKNMNRDSTFIVNKTITAEVLAIDEYTGKTSTGTVYVRVPDFNDNCP  444
               I++ T  IK  V  ++R+ T        + +V+A+D   G   TGT  +RV D ND  P
Sbjct  1723  FEIEADTGVIKTAVCCLDRERT----PDYSIQVVAMDG-GGLKGTGTASIRVKDINDMPP  1777

 Score = 75.9 bits (185),  Expect = 2e-12, Method: Compositional matrix adjust.
 Identities = 108/429 (25%), Positives = 178/429 (41%), Gaps = 53/429 (12%)

Query  53    VDKNTGDINITAIVDREETPSFLITCRALNAQGLDVEKPLILTVKILDINDNPPVFSQ-Q  111
             +D++  ++N+TA+ D          +C  +N           ++ V I D+NDN PVF
Sbjct  950   LDRDKYELNVTAMDDG--------SC-CVNGDATIHTSTAVVVVFITDVNDNKPVFKDCG  1000

Query  112   IFMGEIEENSASNSLVMILNATDADEPNHLNSKIAFKIVSQEPAGTPMFLLSRNTGEVRT  171
              +  ++EE + + S V+ + ATD D+   +N ++ + IV Q       F +   TGEV T
Sbjct  1001  TYYPKVEEGAPNGSPVIKVQATDEDK--GVNGQVKYSIVQQPNQKGTKFTVDEETGEVST  1058
```

```
Query  172   LTNSLDRE--QASSYRLVVSGADKDGEGLSTQCECNIKVKDVNDNFPMFRDSQYSARIEE  229
                  DRE        + V   D+    L   C   +++ DVNDN P+F   +Y   +++
Sbjct  1059  -NKVFDREGDDGKFVSVTVKATDQGEPSLEGVCSFTVEITDVNDNPPLFDRQKYVENVKQ  1117

Query  230   NI-LSSELLRFQVTDLDEEYTDNWLAVYFFT---SGNEGNWFEIQTDPRTNEGILKVVKA  285
             +  + + +LR  V+  DE+  +N   VY  +   S N+  +FEIQ  P +   +LK  K
Sbjct  1118  DASIGTNILR--VSASDEDADNNGAIVYTLSAPHSPNDLEYFEIQ--PESGWIVLK--KP  1171

Query  286   LD-YEQLQSVKLSIAVK----------NKAEFHQSVISRYRVQSTPV----TIQVINVRE  330
             LD    + SV+    +K          + A+    +    R    P+        I++RE
Sbjct  1172  LDGCYSVSSVRERYRLKVYASDRGDPPSTADVDVELDVVDRNNKPPIWDRNIYGPIHIRE  1231

Query  331   GIAFRPASKTFTVQKGISSKKLVDYILGTYQAIDEDTNKAASNVKYVMGRNDGGYLMIDS  390
              +             GI     V Y L         TNK   +  Y+  R D G
Sbjct  1232  NVTVGTVVTAIKASSGIEGNPTVFYRL--MPGSTAQTNKL--HTFYLQQRPDNG-----D  1282

Query  391   KTAEIKFVKNMNRDSTFIVNKTITAEVLAIDEYTGKTSTGTVYVRVPDFNDNCPTAV-LE  449
               A+IK    ++ ++    N TI  E     +     S  TVY+++ D ND  P     E
Sbjct  1283  TWADIKVNHPLDFETIKEYNLTIRVENNGAQQL---ASEATVYIQLEDVNDEIPLFTERE  1339

Query  450   KDAVCSSSP  458
             ++ V    P
Sbjct  1340  QETVLEGEP  1348

 Score = 61.2 bits (147),  Expect = 5e-08, Method: Compositional matrix adjust.
 Identities = 87/366 (24%), Positives = 156/366 (43%), Gaps = 38/366 (10%)

Query  51    FVVDKNTGDINITAIVDRE-ETPSFL-ITCRALNAQGLDVEKPLILTVKILDINDNPPVF  108
             F VD+ TG+++   + DRE +   F+ +T +A +     +E     TV+I D+NDNPP+F
Sbjct  1047  FTVDEETGEVSTNKVFDREGDDGKFVSVTVKATDQGEPSLEGVCSFTVEITDVNDNPPLF  1106

Query  109   SQQIFMGEIEENSASNSLVMILNATDADEPNHLNSKIAFKIVS-QEPAGTPMFLLSRNTG  167
              +Q ++  ++++++  + ++ ++A+D D  N  N  I + + +   P     F +   +G
Sbjct  1107  DRQKYVENVKQDASIGTNILRVSASDEDADN--NGAIVYTLSAPHSPNDLEYFEIQPESG  1164

Query  168   EVRTLTNSLDREQASS-----YRLVVSGADKDGEGLSTQCECNIKVKDVNDNFPMFRDSQ  222
              +  L   LD   + S     YRL V  +D+ G+  ST            +N P   D
Sbjct  1165  WI-VLKKPLDGCYSVSSVRERYRLKVYASDR-GDPPSTADVDVELDVVDRNNKPPIWDRN  1222

Query  223   YSA--RIEENILSSELLRFQVTDLDEEYTDNWLAVYFFTSG-----NEGNWFEIQTDPRT  275
                   I EN+    ++             N    Y    G     N+ + F +Q  P
Sbjct  1223  IYGPIHIRENVTVGTVV--TAIKASSGIEGNPTVFYRLMPGSTAQTNKLHTFYLQQRPDN  1280

Query  276   NE--GILKVVKALDYEQLQSVKLSIAVKNKAEFHQSVISRYRVQSTPVTIQVINVREGIA  333
              +    +KV   LD+E ++   L+I V+N         ++       V IQ+ +V + I
Sbjct  1281  GDTWADIKVNHPLDFETIKEYNLTIRVENNG-------AQQLASEATVYIQLEDVNDEIP  1333

Query  334   FRPASKTFTVQKGISSKKLVDYILGTYQAIDEDTNKAASNVKYVM---GRNDG-GYLMID  389
                  +  TV +G      V  +     AID+D     + V Y +    RN+G  +  I+
Sbjct  1334  LFTEREQETVLEGEPIGTKVTQV----NAIDKDGTFPNNQVYYYIVDSPRNEGKDFFEIN  1389

Query  390   SKTAEI  395
              ++ EI
Sbjct  1390  LQSGEI  1395

 Score = 55.8 bits (133),  Expect = 3e-06, Method: Compositional matrix adjust.
 Identities = 49/158 (31%), Positives = 78/158 (49%), Gaps = 24/158 (15%)

Query  102   NDNPPVFSQQIFMGEIEENSASNSLVMILNATDADEPNHLNSKIAFKIVSQEPAGTP--M  159
             ND  P FSQQ++   ++EN+  N+LV  + A+D D  N     + F  V     GT
Sbjct  880   NDEEPKFSQQVYTPNVDENAGPNTLVTTVVASDKDGDN-----VRFGFVG---GGTSSGQ  931

Query  160   FLLSRNTGEVRTLTN--SLDREQASSYRLVVSGADK-------DGEGLSTQCECNIKVKD  210
             F++   TG +R  +   SLDR++   Y L V+  D        D    ++     + + D
Sbjct  932   FVIEEITGVIRLHSKAISLDRDK---YELNVTAMDDGSCCVNGDATIHTSTAVVVVFITD  988

Query  211   VNDNFPMFRD-SQYSARIEENILS-SELLRFQVTDLDE  246
             VNDN P+F+D   Y  ++EE   + S +++ Q TD D+
Sbjct  989   VNDNKPVFKDCGTYYPKVEEGAPNGSPVIKVQATDEDK  1026
```

```
 Score = 53.9 bits (128),  Expect = 1e-05, Method: Compositional matrix adjust.
 Identities = 70/306 (23%), Positives = 126/306 (41%), Gaps = 30/306 (10%)

Query  20    PIAKIT-SDYQATQKITYRI---SGVGIDQPPFGIFVVDKNTGDINITAIVDRE---ETP  72
             PI  +T  D   T K  Y++   SG G D+  F +   +  TG + I   +D E   ++
Sbjct  1800  PILTVTVHDEDETNKFQYKVIDNSGYGADK--FTMVRNNDGTGSLKIVQPLDYEDPLQSN  1857

Query  73    SFLITCRALNAQGLDVEKPLI------LTVKILDINDNPPVFSQQIFMGEIEENSASNSL  126
              F    + +N +G D +           + VK+ DINDN P F +       + EN+
Sbjct  1858  GFRFRIQ-VNDKGEDNDNDKYHVAYSWVVVKLRDINDNKPQFERPNIEVSVYENADVGKT  1916

Query  127   VMILNATDADEPNHLNSKIAFKIVSQEPAGTPMFLLSRNTGEVRTLTNSLDREQASSYRL  186
             +    ATD D+      SK+++ I   + +        S N     T+  +LDRE    +++
Sbjct  1917  LETFKATDPDQGG--KSKVSYAI---DRSSDRQRQFSINQEGTVTIQRNLDREVTPRHQV  1971

Query  187   VVSGADKDGEGLSTQCECNIKVKDVNDNFPMFRDSQYSARIEENILSSELLRFQVTDLDE  246
              +   D      +       + V+D+NDN P F    Y   + E++   +++     TD D+
Sbjct  1972  KILAIDDGIPPKTATATLTVIVQDINDNPPKFL-KDYRPVLPEHVPPRKVVEILATDDDD  2030

Query  247   EYTDNWLAVYFFTSGNEGNW----FEIQTDPRTNEG----ILKVVKALDYEQLQSVKLSI  298
                 N     F       +     F+++ D +   G    I+  +++ D EQ +   + I
Sbjct  2031  RSKSNGPPFQFRLDPGADDIIRASFKVEQDQKGANGDGMAIVSSLRSFDREQQKEYLIPI  2090

Query  299   AVKNKA  304
              +K+
Sbjct  2091  VIKDHG  2096

 Score = 51.6 bits (122),  Expect = 5e-05, Method: Compositional matrix adjust.
 Identities = 96/398 (24%), Positives = 163/398 (41%), Gaps = 54/398 (14%)

Query  106   PVFSQQIFMGEIEENSASNSLVMILNATDADEPNHLNSKIAFKIVSQ-EPAGTPMFLLSR  164
             P F    +  EI EN   +S ++ + A       +  + +I + + +Q + AGT  F +
Sbjct  677   PQFYMPSYEAEIPENQKKDSDIISVKAK-----SFADREIRYTLKAQGQGAGT--FNIGP  729

Query  165   NTGEVRTLTNSLDRE---QASSYRLVVSGADKDGEGLSTQCECNIKVKDVNDNFPMFRDS  221
              +G V+ L   LD E   Q   Y LVV+ A +D  G ST  E  I+V DVNDN P F
Sbjct  730   TSGIVK-LAKELDFEDLRQPHVYSLVVT-ATEDSGGFSTSVELTIRVTDVNDNAPKFELP  787

Query  222   QYSAR-IEENI-LSSELLRFQVTDLDEEYTDNWLAVYFFTSGNEGNWFEIQTDPRTNEGI  279
              Y A  ++E+I + + +L+ +  D D          + +  S +    F + ++     GI
Sbjct  788   DYQAHNVDEDIQIGTSILKVKAMDADSGAN---AEIEYLVSDDH---FAVDSN-----GI  836

Query  280   LKVVKALDYEQLQSV-KLSIAVKNKAEFHQSVISRYRVQSTPVTIQVINVREGIAFRPAS  338
             +   K LD +   +  +  +  K+K E  ++  +  RV +            E   F
Sbjct  837   IVNNKQLDADNNNAYYEFVVTAKDKGEPAKTGTATVRVYTKNKN------DEEPKFSQQV  890

Query  339   KTFTVQKGISSKKLVDYILGTYQAIDEDTNKAASNVKY--VMGRNDGGYLMIDSKTAEIK  396
              T  V +      LV  ++ +        +K   NV++  V G    G  +I+  T  I+
Sbjct  891   YTPNVDENAGPNTLVTTVVAS--------DKDGDNVRFGFVGGGTSSGQFVIEEITGVIR  942

Query  397   FVK---NMNRDSTFI-VNKTITAEVLAIDEYTGKTSTGTVYVRVPDFNDN------CPTA  446
                  +++RD   + V          + T  TST  V V + D NDN      C T
Sbjct  943   LHSKAISLDRDKYELNVTAMDDGSCCVNGDATIHTSTAVVVVFITDVNDNKPVFKDCGTY  1002

Query  447   VLE-KDAVCSSSPSVVVSARTLNNRYTGPYTFALEDQP  483
               + ++   + SP + V A   +    G   +++  QP
Sbjct  1003  YPKVEEGAPNGSPVIKVQATDEDKGVNGQVKYSIVQQP  1040

 Score = 44.7 bits (104),  Expect = 0.008, Method: Compositional matrix adjust.
 Identities = 48/199 (24%), Positives = 73/199 (37%), Gaps = 19/199 (10%)

Query  27    DYQATQKITYRISGVGIDQPPFGIFVVDKNTGDINITAIVDREETPSFLITCRALNAQGL  86
             D     K++Y I       Q  F I        G + I   +DRE TP   +    A++
Sbjct  1926  DQGGKSKVSYAIDRSSDRQRQFSI----NQEGTVTIQRNLDREVTPRHQVKILAIDDGIP  1981

Query  87    DVEKPLILTVKILDINDNPPVFSQQIFMGEIEENSASNSLVMILNATDADEPNHLNSKIA  146
                    LTV + DINDNPP F +  +    + E+      +V IL   D D
```

```
Sbjct  1982  PKTATATLTVIVQDINDNPPKFLKD-YRPVLPEHVPPRKVVEILATDDDDRSKSNGPPFQ  2040

Query  147   FKIVSQEPAGTPMFLLSRNTGEVRTLTN-----------SLDREQASSYRLVVSGADKDG  195
             F++   +P    +   S    + +   N           S DREQ   Y + +   D
Sbjct  2041  FRL---DPGADDIIRASFKVEQDQKGANGDGMAIVSSLRSFDREQQKEYLIPIVIKDHGN  2097

Query  196   EGLSTQCECNIKVKDVNDN  214
               ++      + + DVNDN
Sbjct  2098  PAMTGTSTLTVVIGDVNDN  2116

 Score = 40.8 bits (94),  Expect = 0.11, Method: Compositional matrix adjust.
 Identities = 55/242 (23%), Positives = 102/242 (42%), Gaps = 22/242 (9%)

Query  4    KFAKPCREGE--DNSKRNP-IAKITSDYQATQKITYRISGVGIDQPPFGIFVVDKNTGDI  60
            +F  P  E E  +N K++  I  + +   A ++I Y +   G      G F +   +G +
Sbjct  678  QFYMPSYEAEIPENQKKDSDIISVKAKSFADREIRYTLKAQGQGA---GTFNIGPTSGIV  734

Query  61   NITAIVDREE--TPSF--LITCRALNAQGLDVEKPLILTVKILDINDNPPVFSQQIFMGE  116
             +   +D E+   P    L+     ++ G      + LT+++ D+NDN P F    +
Sbjct  735  KLAKELDFEDLRQPHVYSLVVTATEDSGGFSTS--VELTIRVTDVNDNAPKFELPDYQAH  792

Query  117  -IEENSASNSLVMILNATDADEPNHLNSKIAFKIVSQEPAGTPMFLLSRNTGEVRTLTNS  175
             ++E+    + ++ + A DAD  +  N++I + +VS +      F +  N   V
Sbjct  793  NVDEDIQIGTSILKVKAMDAD--SGANAEIEY-LVSDD-----HFAVDSNGIIVNNKQLD  844

Query  176  LDREQASSYRLVVSGADKDGEGLSTQCECNIKVKDVNDNFPMFRDSQYSARIEENILSSE  235
             D   A  Y  VV+  DK     +      +  K+ ND  P F    Y+  ++EN   +
Sbjct  845  ADNNNA-YYEFVVTAKDKGEPAKTGTATVRVYTKNKNDEEPKFSQQVYTPNVDENAGPNT  903

Query  236  LL  237
            L+
Sbjct  904  LV  905

 Score = 39.3 bits (90),  Expect = 0.30, Method: Compositional matrix adjust.
 Identities = 55/225 (24%), Positives = 88/225 (39%), Gaps = 24/225 (11%)

Query  75   LITCRALNAQGLDVEKPLILTVKILDINDNPPVFSQQIF--MGEIEENSASNSLVMILNA  132
            +IT +  NA GL       + + + D+ND PP F  +       ++ N+  N+ V  L A
Sbjct  539  IITNQGHNA-GLKYTDNQRVIIHVKDVNDEPPYFINRPLPMQAVVQLNAPPNTPVFTLQA  597

Query  133  TDADEPNHLNSKIAFKIVSQEPAGTPMFLLSRNTGEVRTLTNSLDREQASSYRLVVSGAD  192
             D D     +  I + IV     G   F +   +G VRT    L  +    Y L V   D
Sbjct  598  RDPD----TDHNIHYFIVRDRTGG--RFEVDERSGVVRTRGTDL-FQLDMEYVLYVKAED  650

Query  193  KDGE----GLSTQCECNIKVKDVNDNFPMFRDSQYSARIEEN-ILSSELLRFQVTDLDEE  247
            ++G+       +  E  + +       P F    Y A I EN    S+++  +     +
Sbjct  651  QNGKVDDRRFQSTPEERLSIVG-GKRAPQFYMPSYEAEIPENQKKDSDIISVKA----KS  705

Query  248  YTDNWLAVYFFTSGNEGNWFEIQTDPRTNEGILKVVKALDYEQLQ  292
            + D  +       G     F I        GI+K+ K LD+E L+
Sbjct  706  FADREIRYTLKAQGQGAGTFNIG----PTSGIVKLAKELDFEDLR  746

>AALB20_033171.P66497 | transcript=AALB20_033171.R66497 | gene=AALB20_033171
| organism=Anopheles_albimanus_STECLA_2020 |
gene_product=neural-cadherin, transcript variant X38 |
transcript_product=neural-cadherin,
transcript variant X6 | location=CM024292:75037560-75208393(+)
| protein_length=3131 | sequence_SO=chromosome
| SO=protein_coding_gene | is_pseudo=false
Length=3131

 Score = 144 bits (364),  Expect = 1e-34, Method: Compositional matrix adjust.
 Identities = 125/437 (29%), Positives = 211/437 (48%), Gaps = 33/437 (8%)

Query  26    SDYQATQKITYRISGVGID--QPPFGIFVVDKNTGDINITAIVDREET---PSFLITCRA  80
              D    Q I Y ++G GID   P    F +++ TG+I +   +DR++    P +  T  A
Sbjct  1580  GDKDRPQNIVYFLTGQGIDPDNPANSKFDINRTTGEIFVLKPLDRDQPNGRPQWRFTVFA  1639
```

```
Query  81    LNAQGLDVEKPLILTVKILDINDNPPVFSQQIFMGEIEENSASNSLVMILNATDADEPNH  140
              +  G  +     + V + DINDN P+F Q ++ G + EN  +   +VM + A D D+PN
Sbjct  1640  QDEGGEGLVGYADVQVNLKDINDNAPIFPQGVYFGNVTENGTAGMVVMTMTAVDYDDPNE  1699

Query  141   -LNSKIAFKI---VSQEPAGTPMFLLSRNTGEVRTLTNSLDREQASSYRLVVSGADKDGE  196
               N+K+ + I   V +E  G+P+F +  +TG ++T    LDRE+   Y + V     DG
Sbjct  1700  GTNAKLIYSIEKNVIEEETGSPIFEIEADTGVIKTAVCCLDRERTPDYSIQVVAM--DGG  1757

Query  197   GLSTQCECNIKVKDVNDNFPMFRDSQYSARIEENILSS--ELLRFQVTDLDEEYTDNWLA  254
             GL       +I+VKD+ND  P F   ++   ++E   ++  E+    VT  DE+ T+ +
Sbjct  1758  GLKGTGTASIRVKDINDMPPQFTKDEWFTEVDETDGTNLPEMPILTVTVHDEDETNKFQY  1817

Query  255   VYFFTSGNEGNWFEIQTDPRTNEGI--LKVVKALDYE---QLQSVKLSIAVKNKAEFHQS  309
                  SG   + F +    R N+G   LK+V+ LDYE   Q     +  I V +K E + +
Sbjct  1818  KVIDNSGYGADKFTMV---RNNDGTGSLKIVQPLDYEDPLQSNGFRFRIQVNDKGEDNDN  1874

Query  310   VISRYRVQSTPVTIQVINVREGIAFRPASKTFTVQKGISSKKLVDYILGTYQAIDEDTNK  369
               +Y V  + V +++ ++ +    +P  +   ++  +      V   L T++A D D
Sbjct  1875  --DKYHVAYSWVVVKLRDINDN---KPQFERPNIEVSVYENADVGKTLETFKATDPDQG-  1928

Query  370   AASNVKYVMGR-NDGGYLMIDSKTAEIKFVKNMNRDSTFIVNKTITAEVLAIDE-YTGKT  427
               S V Y + R +D       ++    +   +N++R+    V       ++LAID+    KT
Sbjct  1929  GKSKVSYAIDRSSDRQRQFSINQEGTVTIQRNLDRE----VTPRHQVKILAIDDGIPPKT  1984

Query  428   STGTVYVRVPDFNDNCP  444
             +T T+ V V D NDN P
Sbjct  1985  ATATLTVIVQDINDNPP  2001

 Score = 85.9 bits (211),  Expect = 1e-15, Method: Compositional matrix adjust.
 Identities = 108/420 (26%), Positives = 177/420 (42%), Gaps = 52/420 (12%)

Query  50    IFVVDKNTGDINITAIVDREETPSFLITCRAL----------NAQGLDVEKPLILTVKIL  99
              F ++  +G+I    + DRE+  ++ +   A            N Q   V K   + + I
Sbjct  1385  FFEINLQSGEIFTKVVFDREKQGAYALEVEARDGAPSARPNSNQQPNSVTK--FIRIGIA  1442

Query  100   DINDNPPVFSQQIFMGEIEENSASNSLVMILNATDADEPNHLNSKIAFKIVSQEPAGTPM  159
             D NDNPP F + ++  E++EN      V+ + A D DE    +S+I ++I S    G
Sbjct  1443  DKNDNPPYFDKALYEAEVDENEDIQHTVLTVTAKDHDE----SSRIRYEITSGNIGGA--  1496

Query  160   FLLSRNTGEVRTLTNSLDREQASSYRLVVSGADKDGEGLSTQCECNIKVKDVNDNFPMFR  219
             F +   TG +  +  +LD E    Y L ++ +D   E  +T      I VKDVNDN P+F
Sbjct  1497  FAVKNMTGAI-YVAGALDYETRKRYELRLAASDNLKENYTTVV---IHVKDVNDNPPVFE  1552

Query  220   DSQYSARIEENI---LSSELLRFQVTDLDEEYTDNWLAVYFFTSG--NEGNWFEIQTDPR  274
                Y  +I E     L   +L+   TD D++   N   VYF T    +  N    + D
Sbjct  1553  RPTYRTQITEEDDRNLPKRVLQVTATDGDKDRPQN--IVYFLTGQGIDPDNPANSKFDIN  1610

Query  275   TNEGILKVVKALDYEQLQS-VKLSIAVKNKAEFHQSVISRYRVQSTPVTIQVINVREGIA  333
                G + V+K LD +Q      +    V  + E  + ++    VQ     + + ++ +
Sbjct  1611  RTTGEIFVLKPLDRDQPNGRPQWRFTVFAQDEGGEGLVGYADVQ-----VNLKDINDNAP  1665

Query  334   FRPASKTFTVQKGISSKKLVDYILGTYQAIDEDTNKAASNVK--YVMGRN------DGGY  385
               P    F     ++         ++ T  A+D D     +N K  Y + +N
Sbjct  1666  IFPQGVYF---GNVTENGTAGMVVMTMTAVDYDDPNEGTNAKLIYSIEKNVIEEETGSPI  1722

Query  386   LMIDSKTAEIK-FVKNMNRDSTFIVNKTITAEVLAIDEYTGKTSTGTVYVRVPDFNDNCP  444
               I++ T  IK  V  ++R+ T        + +V+A+D   G   TGT  +RV D ND  P
Sbjct  1723  FEIEADTGVIKTAVCCLDRERT----PDYSIQVVAMDG-GGLKGTGTASIRVKDINDMPP  1777

 Score = 75.9 bits (185),  Expect = 2e-12, Method: Compositional matrix adjust.
 Identities = 108/429 (25%), Positives = 178/429 (41%), Gaps = 53/429 (12%)

Query  53    VDKNTGDINITAIVDREETPSFLITCRALNAQGLDVEKPLILTVKILDINDNPPVFSQ-Q  111
             +D++  ++N+TA+ D           +C  +N           ++ V I D+NDN PVF
Sbjct  950   LDRDKYELNVTAMDDG--------SC-CVNGDATIHTSTAVVVVFITDVNDNKPVFKDCG  1000

Query  112   IFMGEIEENSASNSLVMILNATDADEPNHLNSKIAFKIVSQEPAGTPMFLLSRNTGEVRT  171
              +  ++EE + + S V+ + ATD D+   +N ++ + IV Q        F +   TGEV T
Sbjct  1001  TYYPKVEEGAPNGSPVIKVQATDEDK--GVNGQVKYSIVQQPNQKGTKFTVDEETGEVST  1058
```

```
Query  172   LTNSLDRE--QASSYRLVVSGADKDGEGLSTQCECNIKVKDVNDNFPMFRDSQYSARIEE  229
                  DRE        + V   D+    L   C   +++ DVNDN P+F   +Y   +++
Sbjct  1059  -NKVFDREGDDGKFVSVTVKATDQGEPSLEGVCSFTVEITDVNDNPPLFDRQKYVENVKQ  1117

Query  230   NI-LSSELLRFQVTDLDEEYTDNWLAVYFFT---SGNEGNWFEIQTDPRTNEGILKVVKA  285
             +  + + +LR  V+  DE+  +N   VY  +   S N+  +FEIQ  P +   +LK  K
Sbjct  1118  DASIGTNILR--VSASDEDADNNGAIVYTLSAPHSPNDLEYFEIQ--PESGWIVLK--KP  1171

Query  286   LD-YEQLQSVKLSIAVK----------NKAEFHQSVISRYRVQSTPV----TIQVINVRE  330
             LD    + SV+    +K            + A+    +    R    P+         I++RE
Sbjct  1172  LDGCYSVSSVRERYRLKVYASDRGDPPSTADVDVELDVVDRNNKPPIWDRNIYGPIHIRE  1231

Query  331   GIAFRPASKTFTVQKGISSKKLVDYILGTYQAIDEDTNKAASNVKYVMGRNDGGYLMIDS  390
              +               GI      V Y L          TNK    +  Y+  R D G
Sbjct  1232  NVTVGTVVTAIKASSGIEGNPTVFYRL--MPGSTAQTNKL--HTFYLQQRPDNG-----D  1282

Query  391   KTAEIKFVKNMNRDSTFIVNKTITAEVLAIDEYTGKTSTGTVYYRVPDFNDNCPTAV-LE  449
               A+IK    ++ ++    N TI  E     +      S  TVY+++ D ND  P     E
Sbjct  1283  TWADIKVNHPLDFETIKEYNLTIRVENNGAQQL---ASEATVYIQLEDVNDEIPLFTERE  1339

Query  450   KDAVCSSSP  458
             ++ V    P
Sbjct  1340  QETVLEGEP  1348

 Score = 61.2 bits (147),  Expect = 5e-08, Method: Compositional matrix adjust.
 Identities = 87/366 (24%), Positives = 156/366 (43%), Gaps = 38/366 (10%)

Query  51    FVVDKNTGDINITAIVDRE-ETPSFL-ITCRALNAQGLDVEKPLILTVKILDINDNPPVF  108
             F VD+ TG+++    + DRE +   F+ +T +A +      +E      TV+I D+NDNPP+F
Sbjct  1047  FTVDEETGEVSTNKVFDREGDDGKFVSVTVKATDQGEPSLEGVCSFTVEITDVNDNPPLF  1106

Query  109   SQQIFMGEIEENSASNSLVMILNATDADEPNHLNSKIAFKIVS-QEPAGTPMFLLSRNTG  167
              +Q ++  ++++++  + ++ ++A+D D  N  N  I + + +   P     F +   +G
Sbjct  1107  DRQKYVENVKQDASIGTNILRVSASDEDADN--NGAIVYTLSAPHSPNDLEYFEIQPESG  1164

Query  168   EVRTLTNSLDREQASS-----YRLVVSGADKDGEGLSTQCECNIKVKDVNDNFPMFRDSQ  222
              +  L   LD   + S      YRL V  +D+ G+  ST              +N P   D
Sbjct  1165  WI-VLKKPLDGCYSVSSVRERYRLKVYASDR-GDPPSTADVDVELDVVDRNNKPPIWDRN  1222

Query  223   YSA--RIEENILSSELLRFQVTDLDEEYTDNWLAVYFFTSG-----NEGNWFEIQTDPRT  275
                   I EN+    ++              N    Y    G     N+ + F +Q  P
Sbjct  1223  IYGPIHIRENVTVGTVV--TAIKASSGIEGNPTVFYRLMPGSTAQTNKLHTFYLQQRPDN  1280

Query  276   NE--GILKVVKALDYEQLQSVKLSIAVKNKAEFHQSVISRYRVQSTPVTIQVINVREGIA  333
              +    +KV   LD+E ++   L+I V+N          ++        V IQ+ +V + I
Sbjct  1281  GDTWADIKVNHPLDFETIKEYNLTIRVENNG-------AQQLASEATVYIQLEDVNDEIP  1333

Query  334   FRPASKTFTVQKGISSKKLVDYILGTYQAIDEDTNKAASNVKYVM---GRNDG-GYLMID  389
                  +  TV +G       V  +     AID+D     + V Y +    RN+G  +  I+
Sbjct  1334  LFTEREQETVLEGEPIGTKVTQV----NAIDKDGTFPNNQVYYYIVDSPRNEGKDFFEIN  1389

Query  390   SKTAEI  395
              ++ EI
Sbjct  1390  LQSGEI  1395

 Score = 55.8 bits (133),  Expect = 3e-06, Method: Compositional matrix adjust.
 Identities = 49/158 (31%), Positives = 78/158 (49%), Gaps = 24/158 (15%)

Query  102   NDNPPVFSQQIFMGEIEENSASNSLVMILNATDADEPNHLNSKIAFKIVSQEPAGTP--M  159
             ND  P FSQQ++    ++EN+  N+LV  + A+D D  N      + F  V     GT
Sbjct  880   NDEEPKFSQQVYTPNVDENAGPNTLVTTVVASDKDGDN-----VRFGFVG---GGTSSGQ  931

Query  160   FLLSRNTGEVRTLTN--SLDREQASSYRLVVSGADK-------DGEGLSTQCECNIKVKD  210
             F++    TG +R  +   SLDR++    Y L V+  D         D    ++      + + D
Sbjct  932   FVIEEITGVIRLHSKAISLDRDK---YELNVTAMDDGSCCVNGDATIHTSTAVVVVFITD  988

Query  211   VNDNFPMFRD-SQYSARIEENILS-SELLRFQVTDLDE  246
             VNDN P+F+D   Y  ++EE   + S +++ Q TD D+
```

```
Sbjct  989   VNDNKPVFKDCGTYYPKVEEGAPNGSPVIKVQATDEDK  1026

 Score = 53.9 bits (128),  Expect = 1e-05, Method: Compositional matrix adjust.
 Identities = 70/306 (23%), Positives = 126/306 (41%), Gaps = 30/306 (10%)

Query  20    PIAKIT-SDYQATQKITYRI---SGVGIDQPPFGIFVVDKNTGDINITAIVDRE---ETP  72
             PI  +T  D   T K  Y++   SG G D+  F +   +  TG + I   +D E   ++
Sbjct  1800  PILTVTVHDEDETNKFQYKVIDNSGYGADK--FTMVRNNDGTGSLKIVQPLDYEDPLQSN  1857

Query  73    SFLITCRALNAQGLDVEKPLI------LTVKILDINDNPPVFSQQIFMGEIEENSASNSL  126
              F    + +N +G D +           + VK+ DINDN P F +      + EN+
Sbjct  1858  GFRFRIQ-VNDKGEDNDNDKYHVAYSWVVVKLRDINDNKPQFERPNIEVSVYENADVGKT  1916

Query  127   VMILNATDADEPNHLNSKIAFKIVSQEPAGTPMFLLSRNTGEVRTLTNSLDREQASSYRL  186
             +    ATD D+      SK+++ I   + +        S N     T+  +LDRE    +++
Sbjct  1917  LETFKATDPDQGG--KSKVSYAI---DRSSDRQRQFSINQEGTVTIQRNLDREVTPRHQV  1971

Query  187   VVSGADKDGEGLSTQCECNIKVKDVNDNFPMFRDSQYSARIEENILSSELLRFQVTDLDE  246
              +   D       +       + V+D+NDN P F    Y   + E++   +++     TD D+
Sbjct  1972  KILAIDDGIPPKTATATLTVIVQDINDNPPKFL-KDYRPVLPEHVPPRKVVEILATDDDD  2030

Query  247   EYTDNWLAVYFFTSGNEGNW----FEIQTDPRTNEG----ILKVVKALDYEQLQSVKLSI  298
                N     F        +     F+++ D +   G    I+  +++ D EQ +    + I
Sbjct  2031  RSKSNGPPFQFRLDPGADDIIRASFKVEQDQKGANGDGMAIVSSLRSFDREQQKEYLIPI  2090

Query  299   AVKNKA  304
              +K+
Sbjct  2091  VIKDHG  2096

 Score = 51.6 bits (122),  Expect = 5e-05, Method: Compositional matrix adjust.
 Identities = 96/398 (24%), Positives = 163/398 (41%), Gaps = 54/398 (14%)

Query  106   PVFSQQIFMGEIEENSASNSLVMILNATDADEPNHLNSKIAFKIVSQ-EPAGTPMFLLSR  164
             P F    +  EI EN   +S ++ + A       +  + +I + + +Q + AGT  F +
Sbjct  677   PQFYMPSYEAEIPENQKKDSDIISVKAK-----SFADREIRYTLKAQGQGAGT--FNIGP  729

Query  165   NTGEVRTLTNSLDRE---QASSYRLVVSGADKDGEGLSTQCECNIKVKDVNDNFPMFRDS  221
              +G V+ L   LD E   Q   Y LVV+ A +D  G ST  E  I+V DVNDN P F
Sbjct  730   TSGIVK-LAKELDFEDLRQPHVYSLVVT-ATEDSGGFSTSVELTIRVTDVNDNAPKFELP  787

Query  222   QYSAR-IEENI-LSSELLRFQVTDLDEEYTDNWLAVYFFTSGNEGNWFEIQTDPRTNEGI  279
              Y A  ++E+I + + +L+ +  D D          + +  S +    F + ++     GI
Sbjct  788   DYQAHNVDEDIQIGTSILKVKAMDADSGAN---AEIEYLVSDDH---FAVDSN-----GI  836

Query  280   LKVVKALDYEQLQSV-KLSIAVKNKAEFHQSVISRYRVQSTPVTIQVINVREGIAFRPAS  338
             +   K LD +   +  +  K+K E  ++  +  RV +             E   F
Sbjct  837   IVNNKQLDADNNNAYYEFVVTAKDKGEPAKTGTATVRVYTKNKN------DEEPKFSQQV  890

Query  339   KTFTVQKGISSKKLVDYILGTYQAIDEDTNKAASNVKY--VMGRNDGGYLMIDSKTAEIK  396
              T  V +      LV  ++ +          +K   NV++  V G    G  +I+  T  I+
Sbjct  891   YTPNVDENAGPNTLVTTVVAS--------DKDGDNVRFGFVGGGTSSGQFVIEEITGVIR  942

Query  397   FVK---NMNRDSTFI-VNKTITAEVLAIDEYTGKTSTGTVYVRVPDFNDN------CPTA  446
                   +++RD   + V              + T  TST  V V + D NDN      C T
Sbjct  943   LHSKAISLDRDKYELNVTAMDDGSCCVNGDATIHTSTAVVVVFITDVNDNKPVFKDCGTY  1002

Query  447   VLE-KDAVCSSSPSVVVSARTLNNRYTGPYTFALEDQP  483
               + ++   + SP + V A   +    G   +++  QP
Sbjct  1003  YPKVEEGAPNGSPVIKVQATDEDKGVNGQVKYSIVQQP  1040

 Score = 44.7 bits (104),  Expect = 0.008, Method: Compositional matrix adjust.
 Identities = 48/199 (24%), Positives = 73/199 (37%), Gaps = 19/199 (10%)

Query  27    DYQATQKITYRISGVGIDQPPFGIFVVDKNTGDINITAIVDREETPSFLITCRALNAQGL  86
             D     K++Y I       Q  F I        G + I   +DRE TP    +    A++
Sbjct  1926  DQGGKSKVSYAIDRSSDRQRQFSI----NQEGTVTIQRNLDREVTPRHQVKILAIDDGIP  1981

Query  87    DVEKPLILTVKILDINDNPPVFSQQIFMGEIEENSASNSLVMILNATDADEPNHLNSKIA  146
```

```
                    LTV + DINDNPP F +  +   + E+     +V IL   D D
Sbjct  1982  PKTATATLTVIVQDINDNPPKFLKD-YRPVLPEHVPPRKVVEILATDDDDRSKSNGPPFQ  2040

Query  147   FKIVSQEPAGTPMFLLSRNTGEVRTLTN-----------SLDREQASSYRLVVSGADKDG  195
             F++   +P    +   S    + +   N           S DREQ   Y + +   D
Sbjct  2041  FRL---DPGADDIIRASFKVEQDQKGANGDGMAIVSSLRSFDREQQKEYLIPIVIKDHGN  2097

Query  196   EGLSTQCECNIKVKDVNDN  214
               ++      + + DVNDN
Sbjct  2098  PAMTGTSTLTVVIGDVNDN  2116

 Score = 40.8 bits (94),  Expect = 0.11, Method: Compositional matrix adjust.
 Identities = 55/242 (23%), Positives = 102/242 (42%), Gaps = 22/242 (9%)

Query  4    KFAKPCREGE--DNSKRNP-IAKITSDYQATQKITYRISGVGIDQPPFGIFVVDKNTGDI  60
            +F  P  E E  +N K++  I  + +   A ++I Y +   G      G F +   +G +
Sbjct  678  QFYMPSYEAEIPENQKKDSDIISVKAKSFADREIRYTLKAQGQGA---GTFNIGPTSGIV  734

Query  61   NITAIVDREE--TPSF--LITCRALNAQGLDVEKPLILTVKILDINDNPPVFSQQIFMGE  116
             +   +D E+   P    L+     ++ G      + LT+++ D+NDN P F    +
Sbjct  735  KLAKELDFEDLRQPHVYSLVVTATEDSGGFSTS--VELTIRVTDVNDNAPKFELPDYQAH  792

Query  117  -IEENSASNSLVMILNATDADEPNHLNSKIAFKIVSQEPAGTPMFLLSRNTGEVRTLTNS  175
             ++E+    + ++ + A DAD  +  N++I + +VS +      F +  N   V
Sbjct  793  NVDEDIQIGTSILKVKAMDAD--SGANAEIEY-LVSDD-----HFAVDSNGIIVNNKQLD  844

Query  176  LDREQASSYRLVVSGADKDGEGLSTQCECNIKVKDVNDNFPMFRDSQYSARIEENILSSE  235
             D   A  Y  VV+  DK     +      +  K+ ND  P F    Y+  ++EN   +
Sbjct  845  ADNNNA-YYEFVVTAKDKGEPAKTGTATVRVYTKNKNDEEPKFSQQVYTPNVDENAGPNT  903

Query  236  LL  237
            L+
Sbjct  904  LV  905

 Score = 39.3 bits (90),  Expect = 0.30, Method: Compositional matrix adjust.
 Identities = 55/225 (24%), Positives = 88/225 (39%), Gaps = 24/225 (11%)

Query  75   LITCRALNAQGLDVEKPLILTVKILDINDNPPVFSQQIF--MGEIEENSASNSLVMILNA  132
            +IT +  NA GL       + + + D+ND PP F  +       ++ N+  N+ V  L A
Sbjct  539  IITNQGHNA-GLKYTDNQRVIIHVKDVNDEPPYFINRPLPMQAVVQLNAPPNTPVFTLQA  597

Query  133  TDADEPNHLNSKIAFKIVSQEPAGTPMFLLSRNTGEVRTLTNSLDREQASSYRLVVSGAD  192
             D D     +  I + IV     G   F +   +G VRT    L  +    Y L V   D
Sbjct  598  RDPD----TDHNIHYFIVRDRTGG--RFEVDERSGVVRTRGTDL-FQLDMEYVLYVKAED  650

Query  193  KDGE----GLSTQCECNIKVKDVNDNFPMFRDSQYSARIEEN-ILSSELLRFQVTDLDEE  247
            ++G+       +  E  + +       P F    Y A I EN    S+++  +     +
Sbjct  651  QNGKVDDRRFQSTPEERLSIVG-GKRAPQFYMPSYEAEIPENQKKDSDIISVKA----KS  705

Query  248  YTDNWLAVYFFTSGNEGNWFEIQTDPRTNEGILKVVKALDYEQLQ  292
            + D  +       G     F I        GI+K+ K LD+E L+
Sbjct  706  FADREIRYTLKAQGQGAGTFNIG----PTSGIVKLAKELDFEDLR  746

>AALB20_033171.P66496 | transcript=AALB20_033171.R66496 | gene=AALB20_033171
| organism=Anopheles_albimanus_STECLA_2020 |
gene_product=neural-cadherin, transcript variant X38 |
transcript_product=neural-cadherin,
transcript variant X23 | location=CM024292:75037560-75208393(+)
| protein_length=3131 | sequence_SO=chromosome
| SO=protein_coding_gene | is_pseudo=false
Length=3131

 Score = 144 bits (364),  Expect = 1e-34, Method: Compositional matrix adjust.
 Identities = 125/437 (29%), Positives = 211/437 (48%), Gaps = 33/437 (8%)

Query  26    SDYQATQKITYRISGVGID--QPPFGIFVVDKNTGDINITAIVDREET---PSFLITCRA  80
              D    Q I Y ++G GID   P    F +++ TG+I +   +DR++    P +  T  A
Sbjct  1580  GDKDRPQNIVYFLTGQGIDPDNPANSKFDINRTTGEIFVLKPLDRDQPNGRPQWRFTVFA  1639
```

```
Query  81    LNAQGLDVEKPLILTVKILDINDNPPVFSQQIFMGEIEENSASNSLVMILNATDADEPNH  140
              +  G  +     + V + DINDN P+F Q ++ G + EN  +  +VM + A D D+PN
Sbjct  1640  QDEGGEGLVGYADVQVNLKDINDNAPIFPQGVYFGNVTENGTAGMVVMTMTAVDYDDPNE  1699

Query  141   -LNSKIAFKI---VSQEPAGTPMFLLSRNTGEVRTLTNSLDREQASSYRLVVSGADKDGE  196
               N+K+ + I   V +E  G+P+F +  +TG ++T    LDRE+   Y + V      DG
Sbjct  1700  GTNAKLIYSIEKNVIEEETGSPIFEIEADTGVIKTAVCCLDRERTPDYSIQVVAM--DGG  1757

Query  197   GLSTQCECNIKVKDVNDNFPMFRDSQYSARIEENILSS--ELLRFQVTDLDEEYTDNWLA  254
             GL       +I+VKD+ND  P F   ++   ++E   ++  E+    VT  DE+ T+ +
Sbjct  1758  GLKGTGTASIRVKDINDMPPQFTKDEWFTEVDETDGTNLPEMPILTVTVHDEDETNKFQY  1817

Query  255   VYFFTSGNEGNWFEIQTDPRTNEGI--LKVVKALDYE---QLQSVKLSIAVKNKAEFHQS  309
                  SG   + F +    R N+G   LK+V+ LDYE   Q    +  I V +K E + +
Sbjct  1818  KVIDNSGYGADKFTMV---RNNDGTGSLKIVQPLDYEDPLQSNGFRFRIQVNDKGEDNDN  1874

Query  310   VISRYRVQSTPVTIQVINVREGIAFRPASKTFTVQKGISSKKLVDYILGTYQAIDEDTNK  369
                +Y V  + V +++ ++ +    +P  +   ++  +     V   L T++A D D
Sbjct  1875  --DKYHVAYSWVVVKLRDINDN---KPQFERPNIEVSVYENADVGKTLETFKATDPDQG-  1928

Query  370   AASNVKYVMGR-NDGGYLMIDSKTAEIKFVKNMNRDSTFIVNKTITAEVLAIDE-YTGKT  427
               S V Y + R +D       ++   +   +N++R+    V      ++LAID+    KT
Sbjct  1929  GKSKVSYAIDRSSDRQRQFSINQEGTVTIQRNLDRE----VTPRHQVKILAIDDGIPPKT  1984

Query  428   STGTVYVRVPDFNDNCP  444
             +T T+ V V D NDN P
Sbjct  1985  ATATLTVIVQDINDNPP  2001

 Score = 85.9 bits (211),  Expect = 1e-15, Method: Compositional matrix adjust.
 Identities = 108/420 (26%), Positives = 177/420 (42%), Gaps = 52/420 (12%)

Query  50    IFVVDKNTGDINITAIVDREETPSFLITCRAL----------NAQGLDVEKPLILTVKIL  99
              F ++  +G+I    + DRE+  ++ +   A            N Q   V K   + + I
Sbjct  1385  FFEINLQSGEIFTKVVFDREKQGAYALEVEARDGAPSARPNSNQQPNSVTK--FIRIGIA  1442

Query  100   DINDNPPVFSQQIFMGEIEENSASNSLVMILNATDADEPNHLNSKIAFKIVSQEPAGTPM  159
             D NDNPP F + ++  E++EN      V+ + A D DE    +S+I ++I S    G
Sbjct  1443  DKNDNPPYFDKALYEAEVDENEDIQHTVLTVTAKDHDE----SSRIRYEITSGNIGGA--  1496

Query  160   FLLSRNTGEVRTLTNSLDREQASSYRLVVSGADKDGEGLSTQCECNIKVKDVNDNFPMFR  219
             F +   TG +  +  +LD E    Y L ++ +D   E  +T      I VKDVNDN P+F
Sbjct  1497  FAVKNMTGAI-YVAGALDYETRKRYELRLAASDNLKENYTTVV---IHVKDVNDNPPVFE  1552

Query  220   DSQYSARIEENI---LSSELLRFQVTDLDEEYTDNWLAVYFFTSG--NEGNWFEIQTDPR  274
                Y  +I E     L   +L+   TD D++   N   VYF T    +  N    + D
Sbjct  1553  RPTYRTQITEEDDRNLPKRVLQVTATDGDKDRPQN--IVYFLTGQGIDPDNPANSKFDIN  1610

Query  275   TNEGILKVVKALDYEQLQS-VKLSIAVKNKAEFHQSVISRYRVQSTPVTIQVINVREGIA  333
                G + V+K LD +Q     +    V  + E  + ++    VQ     + + ++ +
Sbjct  1611  RTTGEIFVLKPLDRDQPNGRPQWRFTVFAQDEGGEGLVGYADVQ-----VNLKDINDNAP  1665

Query  334   FRPASKTFTVQKGISSKKLVDYILGTYQAIDEDTNKAASNVK--YVMGRN------DGGY  385
               P    F     ++        ++ T  A+D D     +N K   Y + +N
Sbjct  1666  IFPQGVYF---GNVTENGTAGMVVMTMTAVDYDDPNEGTNAKLIYSIEKNVIEEETGSPI  1722

Query  386   LMIDSKTAEIK-FVKNMNRDSTFIVNKTITAEVLAIDEYTGKTSTGTVYVRVPDFNDNCP  444
               I++ T  IK  V  ++R+ T        + +V+A+D   G   TGT  +RV D ND  P
Sbjct  1723  FEIEADTGVIKTAVCCLDRERT----PDYSIQVVAMDG-GGLKGTGTASIRVKDINDMPP  1777

 Score = 75.9 bits (185),  Expect = 2e-12, Method: Compositional matrix adjust.
 Identities = 108/429 (25%), Positives = 178/429 (41%), Gaps = 53/429 (12%)

Query  53    VDKNTGDINITAIVDREETPSFLITCRALNAQGLDVEKPLILTVKILDINDNPPVFSQ-Q  111
             +D++  ++N+TA+ D          +C  +N           ++ V I D+NDN PVF
Sbjct  950   LDRDKYELNVTAMDDG--------SC-CVNGDATIHTSTAVVVVFITDVNDNKPVFKDCG  1000

Query  112   IFMGEIEENSASNSLVMILNATDADEPNHLNSKIAFKIVSQEPAGTPMFLLSRNTGEVRT  171
              +  ++EE + + S V+ + ATD D+   +N ++ + IV Q        F +   TGEV T
```

```
Sbjct  1001  TYYPKVEEGAPNGSPVIKVQATDEDK--GVNGQVKYSIVQQPNQKGTKFTVDEETGEVST  1058

Query  172   LTNSLDRE--QASSYRLVVSGADKDGEGLSTQCECNIKVKDVNDNFPMFRDSQYSARIEE  229
                   DRE         + V   D+    L   C   +++ DVNDN P+F   +Y   +++
Sbjct  1059  -NKVFDREGDDGKFVSVTVKATDQGEPSLEGVCSFTVEITDVNDNPPLFDRQKYVENVKQ  1117

Query  230   NI-LSSELLRFQVTDLDEEYTDNWLAVYFFT---SGNEGNWFEIQTDPRTNEGILKVVKA  285
             +  + + +LR  V+  DE+  +N   VY  +   S N+  +FEIQ  P +   +LK  K
Sbjct  1118  DASIGTNILR--VSASDEDADNNGAIVYTLSAPHSPNDLEYFEIQ--PESGWIVLK--KP  1171

Query  286   LD-YEQLQSVKLSIAVK----------NKAEFHQSVISRYRVQSTPV----TIQVINVRE  330
             LD    + SV+    +K             + A+    +    R    P+        I++RE
Sbjct  1172  LDGCYSVSSVRERYRLKVYASDRGDPPSTADVDVELDVVDRNNKPPIWDRNIYGPIHIRE  1231

Query  331   GIAFRPASKTFTVQKGISSKKLVDYILGTYQAIDEDTNKAASNVKYVMGRNDGGYLMIDS  390
              +              GI     V Y L          TNK   +  Y+  R D G
Sbjct  1232  NVTVGTVVTAIKASSGIEGNPTVFYRL--MPGSTAQTNKL--HTFYLQQRPDNG-----D  1282

Query  391   KTAEIKFVKNMNRDSTFIVNKTITAEVLAIDEYTGKTSTGTVYVRVPDFNDNCPTAV-LE  449
               A+IK    ++ ++    N TI  E      +      S  TVY+++ D ND  P     E
Sbjct  1283  TWADIKVNHPLDFETIKEYNLTIRVENNGAQQL---ASEATVYIQLEDVNDEIPLFTERE  1339

Query  450   KDAVCSSSP  458
             ++ V    P
Sbjct  1340  QETVLEGEP  1348

 Score = 61.2 bits (147),  Expect = 5e-08, Method: Compositional matrix adjust.
 Identities = 87/366 (24%), Positives = 156/366 (43%), Gaps = 38/366 (10%)

Query  51    FVVDKNTGDINITAIVDRE-ETPSFL-ITCRALNAQGLDVEKPLILTVKILDINDNPPVF  108
             F VD+ TG+++    + DRE +   F+ +T +A +      +E      TV+I D+NDNPP+F
Sbjct  1047  FTVDEETGEVSTNKVFDREGDDGKFVSVTVKATDQGEPSLEGVCSFTVEITDVNDNPPLF  1106

Query  109   SQQIFMGEIEENSASNSLVMILNATDADEPNHLNSKIAFKIVS-QEPAGTPMFLLSRNTG  167
              +Q ++  ++++++  + ++ ++A+D D  N  N  I + + +   P      F +    +G
Sbjct  1107  DRQKYVENVKQDASIGTNILRVSASDEDADN--NGAIVYTLSAPHSPNDLEYFEIQPESG  1164

Query  168   EVRTLTNSLDREQASS-----YRLVVSGADKDGEGLSTQCECNIKVKDVNDNFPMFRDSQ  222
              +  L   LD   + S      YRL V  +D+ G+  ST              +N P   D
Sbjct  1165  WI-VLKKPLDGCYSVSSVRERYRLKVYASDR-GDPPSTADVDVELDVVDRNNKPPIWDRN  1222

Query  223   YSA--RIEENILSSELLRFQVTDLDEEYTDNWLAVYFFTSG-----NEGNWFEIQTDPRT  275
                   I EN+    ++               N    Y    G     N+ + F +Q  P
Sbjct  1223  IYGPIHIRENVTVGTVV--TAIKASSGIEGNPTVFYRLMPGSTAQTNKLHTFYLQQRPDN  1280

Query  276   NE--GILKVVKALDYEQLQSVKLSIAVKNKAEFHQSVISRYRVQSTPVTIQVINVREGIA  333
              +    +KV   LD+E ++   L+I V+N          ++        V IQ+ +V + I
Sbjct  1281  GDTWADIKVNHPLDFETIKEYNLTIRVENNG-------AQQLASEATVYIQLEDVNDEIP  1333

Query  334   FRPASKTFTVQKGISSKKLVDYILGTYQAIDEDTNKAASNVKYVM---GRNDG-GYLMID  389
                  +  TV +G       V  +     AID+D     + V Y +    RN+G  +  I+
Sbjct  1334  LFTEREQETVLEGEPIGTKVTQV----NAIDKDGTFPNNQVYYYIVDSPRNEGKDFFEIN  1389

Query  390   SKTAEI  395
              ++ EI
Sbjct  1390  LQSGEI  1395

 Score = 55.8 bits (133),  Expect = 3e-06, Method: Compositional matrix adjust.
 Identities = 49/158 (31%), Positives = 78/158 (49%), Gaps = 24/158 (15%)

Query  102   NDNPPVFSQQIFMGEIEENSASNSLVMILNATDADEPNHLNSKIAFKIVSQEPAGTP--M  159
             ND  P FSQQ++   ++EN+  N+LV  + A+D D  N     + F  V     GT
Sbjct  880   NDEEPKFSQQVYTPNVDENAGPNTLVTTVVASDKDGDN-----VRFGFVG---GGTSSGQ  931

Query  160   FLLSRNTGEVRTLTN--SLDREQASSYRLVVSGADK-------DGEGLSTQCECNIKVKD  210
             F++    TG +R  +   SLDR++    Y L V+  D         D    ++      + + D
Sbjct  932   FVIEEITGVIRLHSKAISLDRDK---YELNVTAMDDGSCCVNGDATIHTSTAVVVVFITD  988

Query  211   VNDNFPMFRD-SQYSARIEENILS-SELLRFQVTDLDE  246
```

```
                 VNDN P+F+D   Y  ++EE   + S +++ Q TD D+
Sbjct  989   VNDNKPVFKDCGTYYPKVEEGAPNGSPVIKVQATDEDK  1026

 Score = 53.9 bits (128),  Expect = 1e-05, Method: Compositional matrix adjust.
 Identities = 70/306 (23%), Positives = 126/306 (41%), Gaps = 30/306 (10%)

Query  20    PIAKIT-SDYQATQKITYRI---SGVGIDQPPFGIFVVDKNTGDINITAIVDRE---ETP  72
             PI  +T  D   T K  Y++   SG G D+  F +   +  TG + I   +D E   ++
Sbjct  1800  PILTVTVHDEDETNKFQYKVIDNSGYGADK--FTMVRNNDGTGSLKIVQPLDYEDPLQSN  1857

Query  73    SFLITCRALNAQGLDVEKPLI------LTVKILDINDNPPVFSQQIFMGEIEENSASNSL  126
              F    + +N +G D +           + VK+ DINDN P F +      + EN+
Sbjct  1858  GFRFRIQ-VNDKGEDNDNDKYHVAYSWVVVKLRDINDNKPQFERPNIEVSVYENADVGKT  1916

Query  127   VMILNATDADEPNHLNSKIAFKIVSQEPAGTPMFLLSRNTGEVRTLTNSLDREQASSYRL  186
             +    ATD D+     SK+++ I   + +       S N     T+  +LDRE    +++
Sbjct  1917  LETFKATDPDQGG--KSKVSYAI---DRSSDRQRQFSINQEGTVTIQRNLDREVTPRHQV  1971

Query  187   VVSGADKDGEGLSTQCECNIKVKDVNDNFPMFRDSQYSARIEENILSSELLRFQVTDLDE  246
              +   D      +       + V+D+NDN P F    Y   + E++   +++    TD D+
Sbjct  1972  KILAIDDGIPPKTATATLTVIVQDINDNPPKFL-KDYRPVLPEHVPPRKVVEILATDDDD  2030

Query  247   EYTDNWLAVYFFTSGNEGNW----FEIQTDPRTNEG----ILKVVKALDYEQLQSVKLSI  298
                N     F       +     F+++ D +   G    I+  +++ D EQ +   + I
Sbjct  2031  RSKSNGPPFQFRLDPGADDIIRASFKVEQDQKGANGDGMAIVSSLRSFDREQQKEYLIPI  2090

Query  299   AVKNKA  304
              +K+
Sbjct  2091  VIKDHG  2096

 Score = 51.6 bits (122),  Expect = 5e-05, Method: Compositional matrix adjust.
 Identities = 96/398 (24%), Positives = 163/398 (41%), Gaps = 54/398 (14%)

Query  106   PVFSQQIFMGEIEENSASNSLVMILNATDADEPNHLNSKIAFKIVSQ-EPAGTPMFLLSR  164
             P F    +  EI EN   +S ++ + A       +  + +I + + +Q + AGT  F +
Sbjct  677   PQFYMPSYEAEIPENQKKDSDIISVKAK-----SFADREIRYTLKAQGQGAGT--FNIGP  729

Query  165   NTGEVRTLTNSLDRE---QASSYRLVVSGADKDGEGLSTQCECNIKVKDVNDNFPMFRDS  221
              +G V+ L   LD E   Q   Y LVV+ A +D  G ST  E  I+V DVNDN P F
Sbjct  730   TSGIVK-LAKELDFEDLRQPHVYSLVVT-ATEDSGGFSTSVELTIRVTDVNDNAPKFELP  787

Query  222   QYSAR-IEENI-LSSELLRFQVTDLDEEYTDNWLAVYFFTSGNEGNWFEIQTDPRTNEGI  279
              Y A  ++E+I + + +L+ +  D D          + +  S +    F + ++     GI
Sbjct  788   DYQAHNVDEDIQIGTSILKVKAMDADSGAN---AEIEYLVSDDH---FAVDSN-----GI  836

Query  280   LKVVKALDYEQLQSV-KLSIAVKNKAEFHQSVISRYRVQSTPVTIQVINVREGIAFRPAS  338
             +   K LD +   +  +  +  K+K E  ++  +  RV +             E   F
Sbjct  837   IVNNKQLDADNNNAYYEFVVTAKDKGEPAKTGTATVRVYTKNKN------DEEPKFSQQV  890

Query  339   KTFTVQKGISSKKLVDYILGTYQAIDEDTNKAASNVKY--VMGRNDGGYLMIDSKTAEIK  396
              T  V +      LV  ++ +        +K   NV++  V G    G  +I+  T  I+
Sbjct  891   YTPNVDENAGPNTLVTTVVAS--------DKDGDNVRFGFVGGGTSSGQFVIEEITGVIR  942

Query  397   FVK---NMNRDSTFI-VNKTITAEVLAIDEYTGKTSTGTVYVRVPDFNDN------CPTA  446
                   +++RD   + V              + T  TST  V V + D NDN      C T
Sbjct  943   LHSKAISLDRDKYELNVTAMDDGSCCVNGDATIHTSTAVVVVFITDVNDNKPVFKDCGTY  1002

Query  447   VLE-KDAVCSSSPSVVVSARTLNNRYTGPYTFALEDQP  483
              + ++   + SP + V A   +    G   +++  QP
Sbjct  1003  YPKVEEGAPNGSPVIKVQATDEDKGVNGQVKYSIVQQP  1040

 Score = 44.7 bits (104),  Expect = 0.008, Method: Compositional matrix adjust.
 Identities = 48/199 (24%), Positives = 73/199 (37%), Gaps = 19/199 (10%)

Query  27    DYQATQKITYRISGVGIDQPPFGIFVVDKNTGDINITAIVDREETPSFLITCRALNAQGL  86
             D     K++Y I      Q  F I        G + I   +DRE TP   +   A++
Sbjct  1926  DQGGKSKVSYAIDRSSDRQRQFSI----NQEGTVTIQRNLDREVTPRHQVKILAIDDGIP  1981
```

```
Query  87    DVEKPLILTVKILDINDNPPVFSQQIFMGEIEENSASNSLVMILNATDADEPNHLNSKIA  146
                    LTV + DINDNPP F +  +   + E+     +V IL   D D
Sbjct  1982  PKTATATLTVIVQDINDNPPKFLKD-YRPVLPEHVPPRKVVEILATDDDDRSKSNGPPFQ  2040

Query  147   FKIVSQEPAGTPMFLLSRNTGEVRTLTN-----------SLDREQASSYRLVVSGADKDG  195
             F++   +P    +   S    + +   N           S DREQ   Y + +   D
Sbjct  2041  FRL---DPGADDIIRASFKVEQDQKGANGDGMAIVSSLRSFDREQQKEYLIPIVIKDHGN  2097

Query  196   EGLSTQCECNIKVKDVNDN  214
               ++      + + DVNDN
Sbjct  2098  PAMTGTSTLTVVIGDVNDN  2116

 Score = 40.8 bits (94),  Expect = 0.11, Method: Compositional matrix adjust.
 Identities = 55/242 (23%), Positives = 102/242 (42%), Gaps = 22/242 (9%)

Query  4    KFAKPCREGE--DNSKRNP-IAKITSDYQATQKITYRISGVGIDQPPFGIFVVDKNTGDI  60
            +F  P  E E  +N K++  I  + +   A ++I Y +   G      G F +   +G +
Sbjct  678  QFYMPSYEAEIPENQKKDSDIISVKAKSFADREIRYTLKAQGQGA---GTFNIGPTSGIV  734

Query  61   NITAIVDREE--TPSF--LITCRALNAQGLDVEKPLILTVKILDINDNPPVFSQQIFMGE  116
              +   +D E+   P    L+     ++ G      + LT+++ D+NDN P F    +
Sbjct  735  KLAKELDFEDLRQPHVYSLVVTATEDSGGFSTS--VELTIRVTDVNDNAPKFELPDYQAH  792

Query  117  -IEENSASNSLVMILNATDADEPNHLNSKIAFKIVSQEPAGTPMFLLSRNTGEVRTLTNS  175
              ++E+    + ++ + A DAD  +  N++I + +VS +      F +  N   V
Sbjct  793  NVDEDIQIGTSILKVKAMDAD--SGANAEIEY-LVSDD-----HFAVDSNGIIVNNKQLD  844

Query  176  LDREQASSYRLVVSGADKDGEGLSTQCECNIKVKDVNDNFPMFRDSQYSARIEENILSSE  235
              D   A  Y  VV+  DK      +       +  K+ ND  P F    Y+  ++EN   +
Sbjct  845  ADNNNA-YYEFVVTAKDKGEPAKTGTATVRVYTKNKNDEEPKFSQQVYTPNVDENAGPNT  903

Query  236  LL  237
            L+
Sbjct  904  LV  905

 Score = 39.3 bits (90),  Expect = 0.30, Method: Compositional matrix adjust.
 Identities = 55/225 (24%), Positives = 88/225 (39%), Gaps = 24/225 (11%)

Query  75   LITCRALNAQGLDVEKPLILTVKILDINDNPPVFSQQIF--MGEIEENSASNSLVMILNA  132
            +IT +  NA GL       + + + D+ND PP F  +       ++ N+  N+ V  L A
Sbjct  539  IITNQGHNA-GLKYTDNQRVIIHVKDVNDEPPYFINRPLPMQAVVQLNAPPNTPVFTLQA  597

Query  133  TDADEPNHLNSKIAFKIVSQEPAGTPMFLLSRNTGEVRTLTNSLDREQASSYRLVVSGAD  192
             D D     +  I + IV     G   F +   +G VRT    L  +     Y L V   D
Sbjct  598  RDPD----TDHNIHYFIVRDRTGG--RFEVDERSGVVRTRGTDL-FQLDMEYVLYVKAED  650

Query  193  KDGE----GLSTQCECNIKVKDVNDNFPMFRDSQYSARIEEN-ILSSELLRFQVTDLDEE  247
            ++G+        +  E  + +       P F    Y A I EN    S+++  +     +
Sbjct  651  QNGKVDDRRFQSTPEERLSIVG-GKRAPQFYMPSYEAEIPENQKKDSDIISVKA----KS  705

Query  248  YTDNWLAVYFFTSGNEGNWFEIQTDPRTNEGILKVVKALDYEQLQ  292
            + D  +       G     F I         GI+K+ K LD+E L+
Sbjct  706  FADREIRYTLKAQGQGAGTFNIG----PTSGIVKLAKELDFEDLR  746

>AALB20_033171.P66495 | transcript=AALB20_033171.R66495 | gene=AALB20_033171
| organism=Anopheles_albimanus_STECLA_2020 |
gene_product=neural-cadherin, transcript variant X38 |
transcript_product=neural-cadherin,
transcript variant X12 | location=CM024292:75037560-75208393(+)
| protein_length=3131 | sequence_SO=chromosome
| SO=protein_coding_gene | is_pseudo=false
Length=3131

 Score = 144 bits (364),  Expect = 1e-34, Method: Compositional matrix adjust.
 Identities = 125/437 (29%), Positives = 211/437 (48%), Gaps = 33/437 (8%)

Query  26    SDYQATQKITYRISGVGID--QPPFGIFVVDKNTGDINITAIVDREET---PSFLITCRA  80
               D    Q I Y ++G GID   P    F +++ TG+I +   +DR++    P +  T  A
```

```
Sbjct  1580  GDKDRPQNIVYFLTGQGIDPDNPANSKFDINRTTGEIFVLKPLDRDQPNGRPQWRFTVFA  1639

Query  81    LNAQGLDVEKPLILTVKILDINDNPPVFSQQIFMGEIEENSASNSLVMILNATDADEPNH  140
              +  G        + V + DINDN P+F Q ++ G + EN  +  +VM + A D D+PN
Sbjct  1640  QDEGGEGLVGYADVQVNLKDINDNAPIFPQGVYFGNVTENGTAGMVVMTMTAVDYDDPNE  1699

Query  141   -LNSKIAFKI---VSQEPAGTPMFLLSRNTGEVRTLTNSLDREQASSYRLVVSGADKDGE  196
               N+K+ + I   V +E  G+P+F +  +TG ++T    LDRE+   Y + V     DG
Sbjct  1700  GTNAKLIYSIEKNVIEEETGSPIFEIEADTGVIKTAVCCLDRERTPDYSIQVVAM--DGG  1757

Query  197   GLSTQCECNIKVKDVNDNFPMFRDSQYSARIEENILSS--ELLRFQVTDLDEEYTDNWLA  254
             GL      +I+VKD+ND  P F   ++   ++E   ++  E+    VT  DE+ T+ +
Sbjct  1758  GLKGTGTASIRVKDINDMPPQFTKDEWFTEVDETDGTNLPEMPILTVTVHDEDETNKFQY  1817

Query  255   VYFFTSGNEGNWFEIQTDPRTNEGI--LKVVKALDYE---QLQSVKLSIAVKNKAEFHQS  309
                  SG   + F +    R N+G   LK+V+ LDYE   Q    +  I V +K E + +
Sbjct  1818  KVIDNSGYGADKFTMV---RNNDGTGSLKIVQPLDYEDPLQSNGFRFRIQVNDKGEDNDN  1874

Query  310   VISRYRVQSTPVTIQVINVREGIAFRPASKTFTVQKGISSKKLVDYILGTYQAIDEDTNK  369
                +Y V  + V +++ ++ +    +P  +   ++  +     V   L T++A D D
Sbjct  1875  --DKYHVAYSWVVVKLRDINDN---KPQFERPNIEVSVYENADVGKTLETFKATDPDQG-  1928

Query  370   AASNVKYVMGR-NDGGYLMIDSKTAEIKFVKNMNRDSTFIVNKTITAEVLAIDE-YTGKT  427
               S V Y + R +D       ++   +   +N++R+    V      ++LAID+    KT
Sbjct  1929  GKSKVSYAIDRSSDRQRQFSINQEGTVTIQRNLDRE----VTPRHQVKILAIDDGIPPKT  1984

Query  428   STGTVYVRVPDFNDNCP  444
             +T T+ V V D NDN P
Sbjct  1985  ATATLTVIVQDINDNPP  2001

 Score = 85.9 bits (211),  Expect = 1e-15, Method: Compositional matrix adjust.
 Identities = 108/420 (26%), Positives = 177/420 (42%), Gaps = 52/420 (12%)

Query  50    IFVVDKNTGDINITAIVDREETPSFLITCRAL----------NAQGLDVEKPLILTVKIL  99
              F ++  +G+I    + DRE+  ++ +   A           N Q   V K   + + I
Sbjct  1385  FFEINLQSGEIFTKVVFDREKQGAYALEVEARDGAPSARPNSNQQPNSVTK--FIRIGIA  1442

Query  100   DINDNPPVFSQQIFMGEIEENSASNSLVMILNATDADEPNHLNSKIAFKIVSQEPAGTPM  159
             D NDNPP F + ++  E++EN      V+ + A D DE    +S+I ++I S    G
Sbjct  1443  DKNDNPPYFDKALYEAEVDENEDIQHTVLTVTAKDHDE----SSRIRYEITSGNIGGA--  1496

Query  160   FLLSRNTGEVRTLTNSLDREQASSYRLVVSGADKDGEGLSTQCECNIKVKDVNDNFPMFR  219
             F +   TG +  +  +LD E    Y L ++ +D   E  +T     I VKDVNDN P+F
Sbjct  1497  FAVKNMTGAI-YVAGALDYETRKRYELRLAASDNLKENYTTVV---IHVKDVNDNPPVFE  1552

Query  220   DSQYSARIEENI---LSSELLRFQVTDLDEEYTDNWLAVYFFTSG--NEGNWFEIQTDPR  274
                Y  +I E     L   +L+   TD D++   N   VYF T    +  N    + D
Sbjct  1553  RPTYRTQITEEDDRNLPKRVLQVTATDGDKDRPQN--IVYFLTGQGIDPDNPANSKFDIN  1610

Query  275   TNEGILKVVKALDYEQLQS-VKLSIAVKNKAEFHQSVISRYRVQSTPVTIQVINVREGIA  333
                G + V+K LD +Q     +    V  + E  + ++    VQ     + + ++ +
Sbjct  1611  RTTGEIFVLKPLDRDQPNGRPQWRFTVFAQDEGGEGLVGYADVQ-----VNLKDINDNAP  1665

Query  334   FRPASKTFTVQKGISSKKLVDYILGTYQAIDEDTNKAASNVK--YVMGRN------DGGY  385
               P    F     ++       ++ T  A+D D     +N K  Y + +N
Sbjct  1666  IFPQGVYF---GNVTENGTAGMVVMTMTAVDYDDPNEGTNAKLIYSIEKNVIEEETGSPI  1722

Query  386   LMIDSKTAEIK-FVKNMNRDSTFIVNKTITAEVLAIDEYTGKTSTGTVYVRVPDFNDNCP  444
               I++ T  IK  V  ++R+ T       + +V+A+D   G   TGT  +RV D ND  P
Sbjct  1723  FEIEADTGVIKTAVCCLDRERT----PDYSIQVVAMDG-GGLKGTGTASIRVKDINDMPP  1777

 Score = 75.9 bits (185),  Expect = 2e-12, Method: Compositional matrix adjust.
 Identities = 108/429 (25%), Positives = 178/429 (41%), Gaps = 53/429 (12%)

Query  53    VDKNTGDINITAIVDREETPSFLITCRALNAQGLDVEKPLILTVKILDINDNPPVFSQ-Q  111
             +D++  ++N+TA+ D         +C  +N          ++ V I D+NDN PVF
Sbjct  950   LDRDKYELNVTAMDDG--------SC-CVNGDATIHTSTAVVVVFITDVNDNKPVFKDCG  1000

Query  112   IFMGEIEENSASNSLVMILNATDADEPNHLNSKIAFKIVSQEPAGTPMFLLSRNTGEVRT  171
```

```
                 +  ++EE + + S V+ + ATD D+   +N ++ + IV Q       F +   TGEV T
Sbjct  1001  TYYPKVEEGAPNGSPVIKVQATDEDK--GVNGQVKYSIVQQPNQKGTKFTVDEETGEVST  1058

Query  172   LTNSLDRE--QASSYRLVVSGADKDGEGLSTQCECNIKVKDVNDNFPMFRDSQYSARIEE  229
                  DRE        + V   D+    L   C   +++ DVNDN P+F   +Y   +++
Sbjct  1059  -NKVFDREGDDGKFVSVTVKATDQGEPSLEGVCSFTVEITDVNDNPPLFDRQKYVENVKQ  1117

Query  230   NI-LSSELLRFQVTDLDEEYTDNWLAVYFFT---SGNEGNWFEIQTDPRTNEGILKVVKA  285
              +  + + +LR  V+  DE+  +N   VY  +   S N+  +FEIQ  P +   +LK  K
Sbjct  1118  DASIGTNILR--VSASDEDADNNGAIVYTLSAPHSPNDLEYFEIQ--PESGWIVLK--KP  1171

Query  286   LD-YEQLQSVKLSIAVK----------NKAEFHQSVISRYRVQSTPV----TIQVINVRE  330
              LD    + SV+    +K           + A+    +    R    P+        I++RE
Sbjct  1172  LDGCYSVSSVRERYRLKVYASDRGDPPSTADVDVELDVVDRNNKPPIWDRNIYGPIHIRE  1231

Query  331   GIAFRPASKTFTVQKGISSKKLVDYILGTYQAIDEDTNKAASNVKYVMGRNDGGYLMIDS  390
               +             GI     V Y L         TNK   +  Y+  R D G
Sbjct  1232  NVTVGTVVTAIKASSGIEGNPTVFYRL--MPGSTAQTNKL--HTFYLQQRPDNG-----D  1282

Query  391   KTAEIKFVKNMNRDSTFIVNKTITAEVLAIDEYTGKTSTGTVYVRVPDFNDNCPTAV-LE  449
                A+IK    ++ ++    N TI  E     +     S  TVY+++ D ND  P     E
Sbjct  1283  TWADIKVNHPLDFETIKEYNLTIRVENNGAQQL---ASEATVYIQLEDVNDEIPLFTERE  1339

Query  450   KDAVCSSSP  458
              ++ V    P
Sbjct  1340  QETVLEGEP  1348

 Score = 61.2 bits (147),  Expect = 5e-08, Method: Compositional matrix adjust.
 Identities = 87/366 (24%), Positives = 156/366 (43%), Gaps = 38/366 (10%)

Query  51    FVVDKNTGDINITAIVDRE-ETPSFL-ITCRALNAQGLDVEKPLILTVKILDINDNPPVF  108
              F VD+ TG+++   + DRE +   F+ +T +A +     +E     TV+I D+NDNPP+F
Sbjct  1047  FTVDEETGEVSTNKVFDREGDDGKFVSVTVKATDQGEPSLEGVCSFTVEITDVNDNPPLF  1106

Query  109   SQQIFMGEIEENSASNSLVMILNATDADEPNHLNSKIAFKIVS-QEPAGTPMFLLSRNTG  167
               +Q ++  ++++++  + ++ ++A+D D  N  N  I + + +   P     F +   +G
Sbjct  1107  DRQKYVENVKQDASIGTNILRVSASDEDADN--NGAIVYTLSAPHSPNDLEYFEIQPESG  1164

Query  168   EVRTLTNSLDREQASS-----YRLVVSGADKDGEGLSTQCECNIKVKDVNDNFPMFRDSQ  222
               +  L   LD   + S     YRL V  +D+ G+  ST              +N P   D
Sbjct  1165  WI-VLKKPLDGCYSVSSVRERYRLKVYASDR-GDPPSTADVDVELDVVDRNNKPPIWDRN  1222

Query  223   YSA--RIEENILSSELLRFQVTDLDEEYTDNWLAVYFFTSG-----NEGNWFEIQTDPRT  275
                  I EN+    ++               N    Y    G     N+ + F +Q  P
Sbjct  1223  IYGPIHIRENVTVGTVV--TAIKASSGIEGNPTVFYRLMPGSTAQTNKLHTFYLQQRPDN  1280

Query  276   NE--GILKVVKALDYEQLQSVKLSIAVKNKAEFHQSVISRYRVQSTPVTIQVINVREGIA  333
               +    +KV   LD+E ++   L+I V+N          ++        V IQ+ +V + I
Sbjct  1281  GDTWADIKVNHPLDFETIKEYNLTIRVENNG-------AQQLASEATVYIQLEDVNDEIP  1333

Query  334   FRPASKTFTVQKGISSKKLVDYILGTYQAIDEDTNKAASNVKYVM---GRNDG-GYLMID  389
                   +  TV +G      V  +     AID+D     + V Y +    RN+G  +  I+
Sbjct  1334  LFTEREQETVLEGEPIGTKVTQV----NAIDKDGTFPNNQVYYYIVDSPRNEGKDFFEIN  1389

Query  390   SKTAEI  395
               ++ EI
Sbjct  1390  LQSGEI  1395

 Score = 55.8 bits (133),  Expect = 3e-06, Method: Compositional matrix adjust.
 Identities = 49/158 (31%), Positives = 78/158 (49%), Gaps = 24/158 (15%)

Query  102   NDNPPVFSQQIFMGEIEENSASNSLVMILNATDADEPNHLNSKIAFKIVSQEPAGTP--M  159
              ND  P FSQQ++   ++EN+  N+LV  + A+D D  N     + F  V     GT
Sbjct  880   NDEEPKFSQQVYTPNVDENAGPNTLVTTVVASDKDGDN-----VRFGFVG---GGTSSGQ  931

Query  160   FLLSRNTGEVRTLTN--SLDREQASSYRLVVSGADK-------DGEGLSTQCECNIKVKD  210
              F++    TG +R  +    SLDR++    Y L V+  D         D    ++     + + D
Sbjct  932   FVIEEITGVIRLHSKAISLDRDK---YELNVTAMDDGSCCVNGDATIHTSTAVVVVFITD  988
```

```
Query  211   VNDNFPMFRD-SQYSARIEENILS-SELLRFQVTDLDE  246
             VNDN P+F+D   Y  ++EE   + S +++ Q TD D+
Sbjct  989   VNDNKPVFKDCGTYYPKVEEGAPNGSPVIKVQATDEDK  1026

 Score = 53.9 bits (128),  Expect = 1e-05, Method: Compositional matrix adjust.
 Identities = 70/306 (23%), Positives = 126/306 (41%), Gaps = 30/306 (10%)

Query  20    PIAKIT-SDYQATQKITYRI---SGVGIDQPPFGIFVVDKNTGDINITAIVDRE---ETP  72
             PI  +T  D   T K  Y++   SG G D+  F +   +  TG + I   +D E   ++
Sbjct  1800  PILTVTVHDEDETNKFQYKVIDNSGYGADK--FTMVRNNDGTGSLKIVQPLDYEDPLQSN  1857

Query  73    SFLITCRALNAQGLDVEKPLI------LTVKILDINDNPPVFSQQIFMGEIEENSASNSL  126
              F    + +N +G D +          + VK+ DINDN P F +       + EN+
Sbjct  1858  GFRFRIQ-VNDKGEDNDNDKYHVAYSWVVVKLRDINDNKPQFERPNIEVSVYENADVGKT  1916

Query  127   VMILNATDADEPNHLNSKIAFKIVSQEPAGTPMFLLSRNTGEVRTLTNSLDREQASSYRL  186
             +    ATD D+     SK+++ I   + +       S N     T+  +LDRE    +++
Sbjct  1917  LETFKATDPDQGG--KSKVSYAI---DRSSDRQRQFSINQEGTVTIQRNLDREVTPRHQV  1971

Query  187   VVSGADKDGEGLSTQCECNIKVKDVNDNFPMFRDSQYSARIEENILSSELLRFQVTDLDE  246
              +   D      +      + V+D+NDN P F    Y   + E++   +++    TD D+
Sbjct  1972  KILAIDDGIPPKTATATLTVIVQDINDNPPKFL-KDYRPVLPEHVPPRKVVEILATDDDD  2030

Query  247   EYTDNWLAVYFFTSGNEGNW----FEIQTDPRTNEG----ILKVVKALDYEQLQSVKLSI  298
                 N     F       +     F+++ D +   G    I+  +++ D EQ +   + I
Sbjct  2031  RSKSNGPPFQFRLDPGADDIIRASFKVEQDQKGANGDGMAIVSSLRSFDREQQKEYLIPI  2090

Query  299   AVKNKA  304
              +K+
Sbjct  2091  VIKDHG  2096

 Score = 51.6 bits (122),  Expect = 5e-05, Method: Compositional matrix adjust.
 Identities = 96/398 (24%), Positives = 163/398 (41%), Gaps = 54/398 (14%)

Query  106   PVFSQQIFMGEIEENSASNSLVMILNATDADEPNHLNSKIAFKIVSQ-EPAGTPMFLLSR  164
             P F    +  EI EN   +S ++ + A      +  + +I + + +Q + AGT  F +
Sbjct  677   PQFYMPSYEAEIPENQKKDSDIISVKAK-----SFADREIRYTLKAQGQGAGT--FNIGP  729

Query  165   NTGEVRTLTNSLDRE---QASSYRLVVSGADKDGEGLSTQCECNIKVKDVNDNFPMFRDS  221
              +G V+ L   LD E   Q   Y LVV+ A +D  G ST  E  I+V DVNDN P F
Sbjct  730   TSGIVK-LAKELDFEDLRQPHVYSLVVT-ATEDSGGFSTSVELTIRVTDVNDNAPKFELP  787

Query  222   QYSAR-IEENI-LSSELLRFQVTDLDEEYTDNWLAVYFFTSGNEGNWFEIQTDPRTNEGI  279
              Y A  ++E+I + + +L+ +  D D         + +  S +    F + ++     GI
Sbjct  788   DYQAHNVDEDIQIGTSILKVKAMDADSGAN---AEIEYLVSDDH---FAVDSN-----GI  836

Query  280   LKVVKALDYEQLQSV-KLSIAVKNKAEFHQSVISRYRVQSTPVTIQVINVREGIAFRPAS  338
             +   K LD +   +  +  +  K+K E  ++  +  RV +           E   F
Sbjct  837   IVNNKQLDADNNNAYYEFVVTAKDKGEPAKTGTATVRVYTKNKN------DEEPKFSQQV  890

Query  339   KTFTVQKGISSKKLVDYILGTYQAIDEDTNKAASNVKY--VMGRNDGGYLMIDSKTAEIK  396
              T  V +      LV  ++ +        +K   NV++  V G    G  +I+  T  I+
Sbjct  891   YTPNVDENAGPNTLVTTVVAS--------DKDGDNVRFGFVGGGTSSGQFVIEEITGVIR  942

Query  397   FVK---NMNRDSTFI-VNKTITAEVLAIDEYTGKTSTGTVYVRVPDFNDN------CPTA  446
                   +++RD   + V               + T  TST  V V + D NDN      C T
Sbjct  943   LHSKAISLDRDKYELNVTAMDDGSCCVNGDATIHTSTAVVVVFITDVNDNKPVFKDCGTY  1002

Query  447   VLE-KDAVCSSSPSVVVSARTLNNRYTGPYTFALEDQP  483
               + ++   + SP + V A   +    G   +++  QP
Sbjct  1003  YPKVEEGAPNGSPVIKVQATDEDKGVNGQVKYSIVQQP  1040

 Score = 44.7 bits (104),  Expect = 0.008, Method: Compositional matrix adjust.
 Identities = 48/199 (24%), Positives = 73/199 (37%), Gaps = 19/199 (10%)

Query  27    DYQATQKITYRISGVGIDQPPFGIFVVDKNTGDINITAIVDREETPSFLITCRALNAQGL  86
             D     K++Y I      Q  F I       G + I   +DRE TP   +   A++
Sbjct  1926  DQGGKSKVSYAIDRSSDRQRQFSI----NQEGTVTIQRNLDREVTPRHQVKILAIDDGIP  1981
```

```
Query  87    DVEKPLILTVKILDINDNPPVFSQQIFMGEIEENSASNSLVMILNATDADEPNHLNSKIA  146
                    LTV + DINDNPP F +  +   + E+     +V IL   D D
Sbjct  1982  PKTATATLTVIVQDINDNPPKFLKD-YRPVLPEHVPPRKVVEILATDDDDRSKSNGPPFQ  2040

Query  147   FKIVSQEPAGTPMFLLSRNTGEVRTLTN-----------SLDREQASSYRLVVSGADKDG  195
             F++   +P    +   S    + +   N           S DREQ   Y + +   D
Sbjct  2041  FRL---DPGADDIIRASFKVEQDQKGANGDGMAIVSSLRSFDREQQKEYLIPIVIKDHGN  2097

Query  196   EGLSTQCECNIKVKDVNDN  214
               ++      + + DVNDN
Sbjct  2098  PAMTGTSTLTVVIGDVNDN  2116

 Score = 40.8 bits (94),  Expect = 0.11, Method: Compositional matrix adjust.
 Identities = 55/242 (23%), Positives = 102/242 (42%), Gaps = 22/242 (9%)

Query  4    KFAKPCREGE--DNSKRNP-IAKITSDYQATQKITYRISGVGIDQPPFGIFVVDKNTGDI  60
            +F  P  E E  +N K++  I  + +   A ++I Y +   G      G F +   +G +
Sbjct  678  QFYMPSYEAEIPENQKKDSDIISVKAKSFADREIRYTLKAQGQGA---GTFNIGPTSGIV  734

Query  61   NITAIVDREE--TPSF--LITCRALNAQGLDVEKPLILTVKILDINDNPPVFSQQIFMGE  116
             +   +D E+   P    L+     ++ G      + LT+++ D+NDN P F    +
Sbjct  735  KLAKELDFEDLRQPHVYSLVVTATEDSGGFSTS--VELTIRVTDVNDNAPKFELPDYQAH  792

Query  117  -IEENSASNSLVMILNATDADEPNHLNSKIAFKIVSQEPAGTPMFLLSRNTGEVRTLTNS  175
             ++E+    + ++ + A DAD  +  N++I + +VS +      F +  N   V
Sbjct  793  NVDEDIQIGTSILKVKAMDAD--SGANAEIEY-LVSDD-----HFAVDSNGIIVNNKQLD  844

Query  176  LDREQASSYRLVVSGADKDGEGLSTQCECNIKVKDVNDNFPMFRDSQYSARIEENILSSE  235
             D   A  Y  VV+  DK     +      +  K+ ND  P F    Y+  ++EN    +
Sbjct  845  ADNNNA-YYEFVVTAKDKGEPAKTGTATVRVYTKNKNDEEPKFSQQVYTPNVDENAGPNT  903

Query  236  LL  237
            L+
Sbjct  904  LV  905

 Score = 39.3 bits (90),  Expect = 0.30, Method: Compositional matrix adjust.
 Identities = 55/225 (24%), Positives = 88/225 (39%), Gaps = 24/225 (11%)

Query  75   LITCRALNAQGLDVEKPLILTVKILDINDNPPVFSQQIF--MGEIEENSASNSLVMILNA  132
            +IT +  NA GL       + + + D+ND PP F  +       ++ N+  N+ V  L A
Sbjct  539  IITNQGHNA-GLKYTDNQRVIIHVKDVNDEPPYFINRPLPMQAVVQLNAPPNTPVFTLQA  597

Query  133  TDADEPNHLNSKIAFKIVSQEPAGTPMFLLSRNTGEVRTLTNSLDREQASSYRLVVSGAD  192
             D D     +  I + IV     G   F +   +G VRT    L  +    Y L V   D
Sbjct  598  RDPD----TDHNIHYFIVRDRTGG--RFEVDERSGVVRTRGTDL-FQLDMEYVLYVKAED  650

Query  193  KDGE----GLSTQCECNIKVKDVNDNFPMFRDSQYSARIEEN-ILSSELLRFQVTDLDEE  247
            ++G+       +  E  + +       P F    Y A I EN    S+++  +     +
Sbjct  651  QNGKVDDRRFQSTPEERLSIVG-GKRAPQFYMPSYEAEIPENQKKDSDIISVKA----KS  705

Query  248  YTDNWLAVYFFTSGNEGNWFEIQTDPRTNEGILKVVKALDYEQLQ  292
            + D  +       G     F I        GI+K+ K LD+E L+
Sbjct  706  FADREIRYTLKAQGQGAGTFNIG----PTSGIVKLAKELDFEDLR  746

>AALB20_033171.P66494 | transcript=AALB20_033171.R66494 | gene=AALB20_033171
| organism=Anopheles_albimanus_STECLA_2020 |
gene_product=neural-cadherin, transcript variant X38 |
transcript_product=neural-cadherin,
transcript variant X5 | location=CM024292:75037560-75208393(+)
| protein_length=3131 | sequence_SO=chromosome
| SO=protein_coding_gene | is_pseudo=false
Length=3131

 Score = 144 bits (364),  Expect = 1e-34, Method: Compositional matrix adjust.
 Identities = 125/437 (29%), Positives = 211/437 (48%), Gaps = 33/437 (8%)

Query  26    SDYQATQKITYRISGVGID--QPPFGIFVVDKNTGDINITAIVDREET---PSFLITCRA  80
```

```
              D    Q I Y ++G GID   P    F +++ TG+I +   +DR++    P +  T  A
Sbjct  1580  GDKDRPQNIVYFLTGQGIDPDNPANSKFDINRTTGEIFVLKPLDRDQPNGRPQWRFTVFA  1639

Query  81    LNAQGLDVEKPLILTVKILDINDNPPVFSQQIFMGEIEENSASNSLVMILNATDADEPNH  140
              +  G  +     + V + DINDN P+F Q ++ G + EN  +  +VM + A D D+PN
Sbjct  1640  QDEGGEGLVGYADVQVNLKDINDNAPIFPQGVYFGNVTENGTAGMVVMTMTAVDYDDPNE  1699

Query  141   -LNSKIAFKI---VSQEPAGTPMFLLSRNTGEVRTLTNSLDREQASSYRLVVSGADKDGE  196
               N+K+ + I   V +E  G+P+F +  +TG ++T    LDRE+   Y + V     DG
Sbjct  1700  GTNAKLIYSIEKNVIEEETGSPIFEIEADTGVIKTAVCCLDERTPDYSIQVVAM--DGG  1757

Query  197   GLSTQCECNIKVKDVNDNFPMFRDSQYSARIEENILSS--ELLRFQVTDLDEEYTDNWLA  254
             GL      +I+VKD+ND  P F   ++   ++E   ++  E+    VT  DE+ T+ +
Sbjct  1758  GLKGTGTASIRVKDINDMPPQFTKDEWFTEVDETDGTNLPEMPILTVTVHDEDETNKFQY  1817

Query  255   VYFFTSGNEGNWFEIQTDPRTNEGI--LKVVKALDYE---QLQSVKLSIAVKNKAEFHQS  309
                  SG   + F +    R N+G   LK+V+ LDYE   Q    +  I V +K E + +
Sbjct  1818  KVIDNSGYGADKFTMV---RNNDGTGSLKIVQPLDYEDPLQSNGFRFRIQVNDKGEDNDN  1874

Query  310   VISRYRVQSTPVTIQVINVREGIAFRPASKTFTVQKGISSKKLVDYILGTYQAIDEDTNK  369
                +Y V  + V +++ ++ +    +P  +   ++  +     V   L T++A D D
Sbjct  1875  --DKYHVAYSWVVVKLRDINDN---KPQFERPNIEVSVYENADVGKTLETFKATDPDQG-  1928

Query  370   AASNVKYVMGR-NDGGYLMIDSKTAEIKFVKNMNRDSTFIVNKTITAEVLAIDE-YTGKT  427
               S V Y + R +D       ++   +   +N++R+    V      ++LAID+    KT
Sbjct  1929  GKSKVSYAIDRSSDRQRQFSINQEGTVTIQRNLDRE----VTPRHQVKILAIDDGIPPKT  1984

Query  428   STGTVYVRVPDFNDNCP  444
             +T T+ V V D NDN P
Sbjct  1985  ATATLTVIVQDINDNPP  2001

 Score = 85.9 bits (211),  Expect = 1e-15, Method: Compositional matrix adjust.
 Identities = 108/420 (26%), Positives = 177/420 (42%), Gaps = 52/420 (12%)

Query  50    IFVVDKNTGDINITAIVDREETPSFLITCRAL----------NAQGLDVEKPLILTVKIL  99
              F ++  +G+I    + DRE+  ++ +   A           N Q   V K   + + I
Sbjct  1385  FFEINLQSGEIFTKVVFDREKQGAYALEVEARDGAPSARPNSNQQPNSVTK--FIRIGIA  1442

Query  100   DINDNPPVFSQQIFMGEIEENSASNSLVMILNATDADEPNHLNSKIAFKIVSQEPAGTPM  159
             D NDNPP F + ++  E++EN      V+ + A D DE    +S+I ++I S    G
Sbjct  1443  DKNDNPPYFDKALYEAEVDENEDIQHTVLTVTAKDHDE----SSRIRYEITSGNIGGA--  1496

Query  160   FLLSRNTGEVRTLTNSLDREQASSYRLVVSGADKDGEGLSTQCECNIKVKDVNDNFPMFR  219
             F +   TG +  +  +LD E    Y L ++ +D   E  +T     I VKDVNDN P+F
Sbjct  1497  FAVKNMTGAI-YVAGALDYETRKRYELRLAASDNLKENYTTVV---IHVKDVNDNPPVFE  1552

Query  220   DSQYSARIEENI---LSSELLRFQVTDLDEEYTDNWLAVYFFTSG--NEGNWFEIQTDPR  274
                Y  +I E     L   +L+   TD D++   N   VYF T    +  N    + D
Sbjct  1553  RPTYRTQITEEDDRNLPKRVLQVTATDGDKDRPQN--IVYFLTGQGIDPDNPANSKFDIN  1610

Query  275   TNEGILKVVKALDYEQLQS-VKLSIAVKNKAEFHQSVISRYRVQSTPVTIQVINVREGIA  333
                G + V+K LD +Q     +    V  + E  + ++    VQ     + + ++ +
Sbjct  1611  RTTGEIFVLKPLDRDQPNGRPQWRFTVFAQDEGGEGLVGYADVQ-----VNLKDINDNAP  1665

Query  334   FRPASKTFTVQKGISSKKLVDYILGTYQAIDEDTNKAASNVK--YVMGRN------DGGY  385
               P    F     ++       ++ T  A+D D     +N K  Y + +N
Sbjct  1666  IFPQGVYF---GNVTENGTAGMVVMTMTAVDYDDPNEGTNAKLIYSIEKNVIEEETGSPI  1722

Query  386   LMIDSKTAEIK-FVKNMNRDSTFIVNKTITAEVLAIDEYTGKTSTGTVYVRVPDFNDNCP  444
               I++ T  IK  V  ++R+ T       + +V+A+D   G   TGT  +RV D ND  P
Sbjct  1723  FEIEADTGVIKTAVCCLDRERT----PDYSIQVVAMDG-GGLKGTGTASIRVKDINDMPP  1777

 Score = 75.9 bits (185),  Expect = 2e-12, Method: Compositional matrix adjust.
 Identities = 108/429 (25%), Positives = 178/429 (41%), Gaps = 53/429 (12%)

Query  53    VDKNTGDINITAIVDREETPSFLITCRALNAQGLDVEKPLILTVKILDINDNPPVFSQ-Q  111
             +D++  ++N+TA+ D         +C  +N          ++ V I D+NDN PVF
Sbjct  950   LDRDKYELNVTAMDDG--------SC-CVNGDATIHTSTAVVVVFITDVNDNKPVFKDCG  1000
```

```
Query  112   IFMGEIEENSASNSLVMILNATDADEPNHLNSKIAFKIVSQEPAGTPMFLLSRNTGEVRT  171
              +  ++EE + + S V+ + ATD D+   +N ++ + IV Q       F +   TGEV T
Sbjct  1001  TYYPKVEEGAPNGSPVIKVQATDEDK--GVNGQVKYSIVQQPNQKGTKFTVDEETGEVST  1058

Query  172   LTNSLDRE--QASSYRLVVSGADKDGEGLSTQCECNIKVKDVNDNFPMFRDSQYSARIEE  229
                  DRE         + V   D+    L   C    +++ DVNDN P+F    +Y   +++
Sbjct  1059  -NKVFDREGDDGKFVSVTVKATDQGEPSLEGVCSFTVEITDVNDNPPLFDRQKYVENVKQ  1117

Query  230   NI-LSSELLRFQVTDLDEEYTDNWLAVYFFT---SGNEGNWFEIQTDPRTNEGILKVVKA  285
             +  + + +LR  V+  DE+  +N    VY  +   S N+  +FEIQ  P +    +LK  K
Sbjct  1118  DASIGTNILR--VSASDEDADNNGAIVYTLSAPHSPNDLEYFEIQ--PESGWIVLK--KP  1171

Query  286   LD-YEQLQSVKLSIAVK----------NKAEFHQSVISRYRVQSTPV----TIQVINVRE  330
             LD    + SV+    +K            + A+    +    R    P+        I++RE
Sbjct  1172  LDGCYSVSSVRERYRLKVYASDRGDPPSTADVDVELDVVDRNNKPPIWDRNIYGPIHIRE  1231

Query  331   GIAFRPASKTFTVQKGISSKKLVDYILGTYQAIDEDTNKAASNVKYVMGRNDGGYLMIDS  390
              +               GI     V Y L          TNK   +  Y+  R D G
Sbjct  1232  NVTVGTVVTAIKASSGIEGNPTVFYRL--MPGSTAQTNKL--HTFYLQQRPDNG-----D  1282

Query  391   KTAEIKFVKNMNRDSTFIVNKTITAEVLAIDEYTGKTSTGTVYVRVPDFNDNCPTAV-LE  449
               A+IK    ++ ++    N TI  E     +     S  TVY+++ D ND  P     E
Sbjct  1283  TWADIKVNHPLDFETIKEYNLTIRVENNGAQQL---ASEATVYIQLEDVNDEIPLFTERE  1339

Query  450   KDAVCSSSP  458
             ++ V    P
Sbjct  1340  QETVLEGEP  1348

 Score = 61.2 bits (147),  Expect = 5e-08, Method: Compositional matrix adjust.
 Identities = 87/366 (24%), Positives = 156/366 (43%), Gaps = 38/366 (10%)

Query  51    FVVDKNTGDINITAIVDRE-ETPSFL-ITCRALNAQGLDVEKPLILTVKILDINDNPPVF  108
             F VD+ TG+++   + DRE +   F+ +T +A +     +E     TV+I D+NDNPP+F
Sbjct  1047  FTVDEETGEVSTNKVFDREGDDGKFVSVTVKATDQGEPSLEGVCSFTVEITDVNDNPPLF  1106

Query  109   SQQIFMGEIEENSASNSLVMILNATDADEPNHLNSKIAFKIVS-QEPAGTPMFLLSRNTG  167
              +Q ++   ++++++  + ++ ++A+D D  N  N  I + + +    P     F +   +G
Sbjct  1107  DRQKYVENVKQDASIGTNILRVSASDEDADN--NGAIVYTLSAPHSPNDLEYFEIQPESG  1164

Query  168   EVRTLTNSLDREQASS-----YRLVVSGADKDGEGLSTQCECNIKVKDVNDNFPMFRDSQ  222
              +  L   LD   + S      YRL V  +D+ G+  ST              +N P   D
Sbjct  1165  WI-VLKKPLDGCYSVSSVRERYRLKVYASDR-GDPPSTADVDVELDVVDRNNKPPIWDRN  1222

Query  223   YSA--RIEENILSSELLRFQVTDLDEEYTDNWLAVYFFTSG-----NEGNWFEIQTDPRT  275
                   I EN+    ++              N    Y    G     N+ + F +Q  P
Sbjct  1223  IYGPIHIRENVTVGTVV--TAIKASSGIEGNPTVFYRLMPGSTAQTNKLHTFYLQQRPDN  1280

Query  276   NE--GILKVVKALDYEQLQSVKLSIAVKNKAEFHQSVISRYRVQSTPVTIQVINVREGIA  333
              +    +KV   LD+E ++   L+I V+N          ++        V IQ+ +V + I
Sbjct  1281  GDTWADIKVNHPLDFETIKEYNLTIRVENNG-------AQQLASEATVYIQLEDVNDEIP  1333

Query  334   FRPASKTFTVQKGISSKKLVDYILGTYQAIDEDTNKAASNVKYVM---GRNDG-GYLMID  389
                  +  TV +G       V  +     AID+D     + V Y +    RN+G  +  I+
Sbjct  1334  LFTEREQETVLEGEPIGTKVTQV----NAIDKDGTFPNNQVYYYIVDSPRNEGKDFFEIN  1389

Query  390   SKTAEI  395
              ++ EI
Sbjct  1390  LQSGEI  1395

 Score = 55.8 bits (133),  Expect = 3e-06, Method: Compositional matrix adjust.
 Identities = 49/158 (31%), Positives = 78/158 (49%), Gaps = 24/158 (15%)

Query  102   NDNPPVFSQQIFMGEIEENSASNSLVMILNATDADEPNHLNSKIAFKIVSQEPAGTP--M  159
             ND  P FSQQ++   ++EN+  N+LV  + A+D D  N      + F  V      GT
Sbjct  880   NDEEPKFSQQVYTPNVDENAGPNTLVTTVVASDKDGDN-----VRFGFVG---GGTSSGQ  931

Query  160   FLLSRNTGEVRTLTN--SLDREQASSYRLVVSGADK-------DGEGLSTQCECNIKVKD  210
             F++   TG +R  +   SLDR++   Y L V+  D         D    ++     + + D
Sbjct  932   FVIEEITGVIRLHSKAISLDRDK---YELNVTAMDDGSCCVNGDATIHTSTAVVVVFITD  988
```

```
Query  211   VNDNFPMFRD-SQYSARIEENILS-SELLRFQVTDLDE  246
             VNDN P+F+D   Y  ++EE   + S +++ Q TD D+
Sbjct  989   VNDNKPVFKDCGTYYPKVEEGAPNGSPVIKVQATDEDK  1026

 Score = 53.9 bits (128),  Expect = 1e-05, Method: Compositional matrix adjust.
 Identities = 70/306 (23%), Positives = 126/306 (41%), Gaps = 30/306 (10%)

Query  20    PIAKIT-SDYQATQKITYRI---SGVGIDQPPFGIFVVDKNTGDINITAIVDRE---ETP  72
             PI  +T  D   T K  Y++   SG G D+  F +   +  TG + I   +D E   ++
Sbjct  1800  PILTVTVHDEDETNKFQYKVIDNSGYGADK--FTMVRNNDGTGSLKIVQPLDYEDPLQSN  1857

Query  73    SFLITCRALNAQGLDVEKPLI------LTVKILDINDNPPVFSQQIFMGEIEENSASNSL  126
              F    + +N +G D +          + VK+ DINDN P F +      + EN+
Sbjct  1858  GFRFRIQ-VNDKGEDNDNDKYHVAYSWVVVKLRDINDNKPQFERPNIEVSVYENADVGKT  1916

Query  127   VMILNATDADEPNHLNSKIAFKIVSQEPAGTPMFLLSRNTGEVRTLTNSLDREQASSYRL  186
             +    ATD D+     SK+++ I   + +       S N     T+  +LDRE    +++
Sbjct  1917  LETFKATDPDQGG--KSKVSYAI---DRSSDRQRQFSINQEGTVTIQRNLDREVTPRHQV  1971

Query  187   VVSGADKDGEGLSTQCECNIKVKDVNDNFPMFRDSQYSARIEENILSSELLRFQVTDLDE  246
              +   D      +      + V+D+NDN P F    Y   + E++   +++    TD D+
Sbjct  1972  KILAIDDGIPPKTATATLTVIVQDINDNPPKFL-KDYRPVLPEHVPPRKVVEILATDDDD  2030

Query  247   EYTDNWLAVYFFTSGNEGNW----FEIQTDPRTNEG----ILKVVKALDYEQLQSVKLSI  298
                 N     F       +     F+++ D +   G    I+  +++ D EQ +   + I
Sbjct  2031  RSKSNGPPFQFRLDPGADDIIRASFKVEQDQKGANGDGMAIVSSLRSFDREQQKEYLIPI  2090

Query  299   AVKNKA  304
              +K+
Sbjct  2091  VIKDHG  2096

 Score = 51.6 bits (122),  Expect = 5e-05, Method: Compositional matrix adjust.
 Identities = 96/398 (24%), Positives = 163/398 (41%), Gaps = 54/398 (14%)

Query  106   PVFSQQIFMGEIEENSASNSLVMILNATDADEPNHLNSKIAFKIVSQ-EPAGTPMFLLSR  164
             P F    +  EI EN   +S ++ + A      +  + +I + + +Q + AGT  F +
Sbjct  677   PQFYMPSYEAEIPENQKKDSDIISVKAK-----SFADREIRYTLKAQGQGAGT--FNIGP  729

Query  165   NTGEVRTLTNSLDRE---QASSYRLVVSGADKDGEGLSTQCECNIKVKDVNDNFPMFRDS  221
              +G V+ L   LD E   Q   Y LVV+ A +D  G ST  E  I+V DVNDN P F
Sbjct  730   TSGIVK-LAKELDFEDLRQPHVYSLVVT-ATEDSGGFSTSVELTIRVTDVNDNAPKFELP  787

Query  222   QYSAR-IEENI-LSSELLRFQVTDLDEEYTDNWLAVYFFTSGNEGNWFEIQTDPRTNEGI  279
              Y A  ++E+I + + +L+ +  D D         + +  S +    F + ++     GI
Sbjct  788   DYQAHNVDEDIQIGTSILKVKAMDADSGAN---AEIEYLVSDDH---FAVDSN-----GI  836

Query  280   LKVVKALDYEQLQSV-KLSIAVKNKAEFHQSVISRYRVQSTPVTIQVINVREGIAFRPAS  338
             +   K LD +   +  +  +  K+K E  ++  +  RV +           E   F
Sbjct  837   IVNNKQLDADNNNAYYEFVVTAKDKGEPAKTGTATVRVYTKNKN------DEEPKFSQQV  890

Query  339   KTFTVQKGISSKKLVDYILGTYQAIDEDTNKAASNVKY--VMGRNDGGYLMIDSKTAEIK  396
              T  V +      LV  ++ +        +K   NV++  V G    G  +I+  T  I+
Sbjct  891   YTPNVDENAGPNTLVTTVVAS--------DKDGDNVRFGFVGGGTSSGQFVIEEITGVIR  942

Query  397   FVK---NMNRDSTFI-VNKTITAEVLAIDEYTGKTSTGTVYVRVPDFNDN------CPTA  446
                   +++RD   + V            + T  TST  V V + D NDN      C T
Sbjct  943   LHSKAISLDRDKYELNVTAMDDGSCCVNGDATIHTSTAVVVVFITDVNDNKPVFKDCGTY  1002

Query  447   VLE-KDAVCSSSPSVVVSARTLNNRYTGPYTFALEDQP  483
               + ++   + SP + V A   +    G   +++  QP
Sbjct  1003  YPKVEEGAPNGSPVIKVQATDEDKGVNGQVKYSIVQQP  1040

 Score = 44.7 bits (104),  Expect = 0.008, Method: Compositional matrix adjust.
 Identities = 48/199 (24%), Positives = 73/199 (37%), Gaps = 19/199 (10%)

Query  27    DYQATQKITYRISGVGIDQPPFGIFVVDKNTGDINITAIVDREETPSFLITCRALNAQGL  86
             D     K++Y I      Q  F I       G + I   +DRE TP   +   A++
```

```
Sbjct  1926  DQGGKSKVSYAIDRSSDRQRQFSI----NQEGTVTIQRNLDREVTPRHQVKILAIDDGIP  1981

Query  87    DVEKPLILTVKILDINDNPPVFSQQIFMGEIEENSASNSLVMILNATDADEPNHLNSKIA  146
                    LTV + DINDNPP F +  +   + E+      +V IL   D D
Sbjct  1982  PKTATATLTVIVQDINDNPPKFLKD-YRPVLPEHVPPRKVVEILATDDDDRSKSNGPPFQ  2040

Query  147   FKIVSQEPAGTPMFLLSRNTGEVRTLTN-----------SLDREQASSYRLVVSGADKDG  195
             F++   +P    +   S    + +   N           S DREQ   Y + +   D
Sbjct  2041  FRL---DPGADDIIRASFKVEQDQKGANGDGMAIVSSLRSFDREQQKEYLIPIVIKDHGN  2097

Query  196   EGLSTQCECNIKVKDVNDN  214
               ++      + + DVNDN
Sbjct  2098  PAMTGTSTLTVVIGDVNDN  2116

 Score = 40.8 bits (94),  Expect = 0.11, Method: Compositional matrix adjust.
 Identities = 55/242 (23%), Positives = 102/242 (42%), Gaps = 22/242 (9%)

Query  4    KFAKPCREGE--DNSKRNP-IAKITSDYQATQKITYRISGVGIDQPPFGIFVVDKNTGDI  60
            +F  P  E E  +N K++  I  + +   A ++I Y +   G      G F +   +G +
Sbjct  678  QFYMPSYEAEIPENQKKDSDIISVKAKSFADREIRYTLKAQGQGA---GTFNIGPTSGIV  734

Query  61   NITAIVDREE--TPSF--LITCRALNAQGLDVEKPLILTVKILDINDNPPVFSQQIFMGE  116
             +   +D E+   P    L+     ++ G      + LT+++ D+NDN P F    +
Sbjct  735  KLAKELDFEDLRQPHVYSLVVTATEDSGGFSTS--VELTIRVTDVNDNAPKFELPDYQAH  792

Query  117  -IEENSASNSLVMILNATDADEPNHLNSKIAFKIVSQEPAGTPMFLLSRNTGEVRTLTNS  175
             ++E+    + ++ + A DAD  +  N++I + +VS +      F +  N   V
Sbjct  793  NVDEDIQIGTSILKVKAMDAD--SGANAEIEY-LVSDD-----HFAVDSNGIIVNNKQLD  844

Query  176  LDREQASSYRLVVSGADKDGEGLSTQCECNIKVKDVNDNFPMFRDSQYSARIEENILSSE  235
             D   A  Y  VV+  DK     +      +  K+ ND  P F    Y+  ++EN   +
Sbjct  845  ADNNNA-YYEFVVTAKDKGEPAKTGTATVRVYTKNKNDEEPKFSQQVYTPNVDENAGPNT  903

Query  236  LL  237
            L+
Sbjct  904  LV  905

 Score = 39.3 bits (90),  Expect = 0.30, Method: Compositional matrix adjust.
 Identities = 55/225 (24%), Positives = 88/225 (39%), Gaps = 24/225 (11%)

Query  75   LITCRALNAQGLDVEKPLILTVKILDINDNPPVFSQQIF--MGEIEENSASNSLVMILNA  132
            +IT +  NA GL       + + + D+ND PP F  +       ++ N+  N+ V  L A
Sbjct  539  IITNQGHNA-GLKYTDNQRVIIHVKDVNDEPPYFINRPLPMQAVVQLNAPPNTPVFTLQA  597

Query  133  TDADEPNHLNSKIAFKIVSQEPAGTPMFLLSRNTGEVRTLTNSLDREQASSYRLVVSGAD  192
             D D     +  I + IV     G   F +   +G VRT    L  +    Y L V   D
Sbjct  598  RDPD----TDHNIHYFIVRDRTGG--RFEVDERSGVVRTRGTDL-FQLDMEYVLYVKAED  650

Query  193  KDGE----GLSTQCECNIKVKDVNDNFPMFRDSQYSARIEEN-ILSSELLRFQVTDLDEE  247
            ++G+       +  E  + +       P F    Y A I EN    S+++  +      +
Sbjct  651  QNGKVDDRRFQSTPEERLSIVG-GKRAPQFYMPSYEAEIPENQKKDSDIISVKA----KS  705

Query  248  YTDNWLAVYFFTSGNEGNWFEIQTDPRTNEGILKVVKALDYEQLQ  292
            + D  +       G     F I        GI+K+ K LD+E L+
Sbjct  706  FADREIRYTLKAQGQGAGTFNIG----PTSGIVKLAKELDFEDLR  746

>AALB20_033171.P66491 | transcript=AALB20_033171.R66491 | gene=AALB20_033171
| organism=Anopheles_albimanus_STECLA_2020 |
gene_product=neural-cadherin, transcript variant X38 |
transcript_product=neural-cadherin,
transcript variant X1 | location=CM024292:75037560-75208393(+)
| protein_length=3131 | sequence_SO=chromosome
| SO=protein_coding_gene | is_pseudo=false
Length=3131

 Score = 144 bits (364),  Expect = 1e-34, Method: Compositional matrix adjust.
 Identities = 125/437 (29%), Positives = 211/437 (48%), Gaps = 33/437 (8%)
```

```
Query  26    SDYQATQKITYRISGVGID--QPPFGIFVVDKNTGDINITAIVDREET---PSFLITCRA  80
              D    Q I Y ++G GID   P    F +++ TG+I +   +DR++    P +  T  A
Sbjct  1580  GDKDRPQNIVYFLTGQGIDPDNPANSKFDINRTTGEIFVLKPLDRDQPNGRPQWRFTVFA  1639

Query  81    LNAQGLDVEKPLILTVKILDINDNPPVFSQQIFMGEIEENSASNSLVMILNATDADEPNH  140
              +  G  +     + V + DINDN P+F Q ++ G + EN  +  +VM + A D D+PN
Sbjct  1640  QDEGGEGLVGYADVQVNLKDINDNAPIFPQGVYFGNVTENGTAGMVVMTMTAVDYDDPNE  1699

Query  141   -LNSKIAFKI---VSQEPAGTPMFLLSRNTGEVRTLTNSLDREQASSYRLVVSGADKDGE  196
               N+K+ + I   V +E  G+P+F +  +TG ++T    LDRE+   Y + V     DG
Sbjct  1700  GTNAKLIYSIEKNVIEEETGSPIFEIEADTGVIKTAVCCLDRERTPDYSIQVVAM--DGG  1757

Query  197   GLSTQCECNIKVKDVNDNFPMFRDSQYSARIEENILSS--ELLRFQVTDLDEEYTDNWLA  254
             GL      +I+VKD+ND  P F   ++   ++E   ++  E+    VT  DE+ T+ +
Sbjct  1758  GLKGTGTASIRVKDINDMPPQFTKDEWFTEVDETDGTNLPEMPILTVTVHDEDETNKFQY  1817

Query  255   VYFFTSGNEGNWFEIQTDPRTNEGI--LKVVKALDYE---QLQSVKLSIAVKNKAEFHQS  309
                  SG    + F +    R N+G   LK+V+ LDYE   Q    +  I V +K E + +
Sbjct  1818  KVIDNSGYGADKFTMV---RNNDGTGSLKIVQPLDYEDPLQSNGFRFIQVNDKGEDNDN  1874

Query  310   VISRYRVQSTPVTIQVINVREGIAFRPASKTFTVQKGISSKKLVDYILGTYQAIDEDTNK  369
                +Y V  + V +++ ++ +    +P  +   ++  +     V   L T++A D D
Sbjct  1875  --DKYHVAYSWVVVKLRDINDN---KPQFERPNIEVSVYENADVGKTLETFKATDPDQG-  1928

Query  370   AASNVKYVMGR-NDGGYLMIDSKTAEIKFVKNMNRDSTFIVNKTITAEVLAIDE-YTGKT  427
               S V Y + R +D       ++   +   +N++R+    V      ++LAID+    KT
Sbjct  1929  GKSKVSYAIDRSSDRQRQFSINQEGTVTIQRNLDRE----VTPRHQVKILAIDDGIPPKT  1984

Query  428   STGTVYVRVPDFNDNCP  444
             +T T+ V V D NDN P
Sbjct  1985  ATATLTVIVQDINDNPP  2001

 Score = 85.9 bits (211),  Expect = 1e-15, Method: Compositional matrix adjust.
 Identities = 108/420 (26%), Positives = 177/420 (42%), Gaps = 52/420 (12%)

Query  50    IFVVDKNTGDINITAIVDREETPSFLITCRAL----------NAQGLDVEKPLILTVKIL  99
              F ++  +G+I    + DRE+  ++ +   A           N Q   V K   + + I
Sbjct  1385  FFEINLQSGEIFTKVVFDREKQGAYALEVEARDGAPSARPNSNQQPNSVTK--FIRIGIA  1442

Query  100   DINDNPPVFSQQIFMGEIEENSASNSLVMILNATDADEPNHLNSKIAFKIVSQEPAGTPM  159
             D NDNPP F + ++  E++EN      V+ + A D DE    +S+I ++I S    G
Sbjct  1443  DKNDNPPYFDKALYEAEVDENEDIQHTVLTVTAKDHDE----SSRIRYEITSGNIGGA--  1496

Query  160   FLLSRNTGEVRTLTNSLDREQASSYRLVVSGADKDGEGLSTQCECNIKVKDVNDNFPMFR  219
             F +   TG +  +  +LD E    Y L ++ +D   E  +T     I VKDVNDN P+F
Sbjct  1497  FAVKNMTGAI-YVAGALDYETRKRYELRLAASDNLKENYTTVV---IHVKDVNDNPPVFE  1552

Query  220   DSQYSARIEENI---LSSELLRFQVTDLDEEYTDNWLAVYFFTSG--NEGNWFEIQTDPR  274
                Y  +I E     L   +L+   TD D++   N   VYF T    +  N    + D
Sbjct  1553  RPTYRTQITEEDDRNLPKRVLQVTATDGDKDRPQN--IVYFLTGQGIDPDNPANSKFDIN  1610

Query  275   TNEGILKVVKALDYEQLQS-VKLSIAVKNKAEFHQSVISRYRVQSTPVTIQVINVREGIA  333
                G + V+K LD +Q     +    V  + E  + ++    VQ     + + ++ +
Sbjct  1611  RTTGEIFVLKPLDRDQPNGRPQWRFTVFAQDEGGEGLVGYADVQ-----VNLKDINDNAP  1665

Query  334   FRPASKTFTVQKGISSKKLVDYILGTYQAIDEDTNKAASNVK--YVMGRN------DGGY  385
               P    F     ++       ++ T  A+D D     +N K  Y + +N
Sbjct  1666  IFPQGVYF---GNVTENGTAGMVVMTMTAVDYDDPNEGTNAKLIYSIEKNVIEEETGSPI  1722

Query  386   LMIDSKTAEIK-FVKNMNRDSTFIVNKTITAEVLAIDEYTGKTSTGTVYVRVPDFNDNCP  444
               I++ T  IK  V  ++R+ T       + +V+A+D   G   TGT  +RV D ND  P
Sbjct  1723  FEIEADTGVIKTAVCCLDRERT----PDYSIQVVAMDG-GGLKGTGTASIRVKDINDMPP  1777

 Score = 75.9 bits (185),  Expect = 2e-12, Method: Compositional matrix adjust.
 Identities = 108/429 (25%), Positives = 178/429 (41%), Gaps = 53/429 (12%)

Query  53    VDKNTGDINITAIVDREETPSFLITCRALNAQGLDVEKPLILTVKILDINDNPPVFSQ-Q  111
             +D++  ++N+TA+ D         +C  +N          ++ V I D+NDN PVF
Sbjct  950   LDRDKYELNVTAMDDG--------SC-CVNGDATIHTSTAVVVVFITDVNDNKPVFKDCG  1000
```

```
Query  112   IFMGEIEENSASNSLVMILNATDADEPNHLNSKIAFKIVSQEPAGTPMFLLSRNTGEVRT  171
              +  ++EE + + S V+ + ATD D+   +N ++ + IV Q       F +   TGEV T
Sbjct  1001  TYYPKVEEGAPNGSPVIKVQATDEDK--GVNGQVKYSIVQQPNQKGTKFTVDEETGEVST  1058

Query  172   LTNSLDRE--QASSYRLVVSGADKDGEGLSTQCECNIKVKDVNDNFPMFRDSQYSARIEE  229
                   DRE         + V   D+    L   C   +++ DVNDN P+F   +Y   +++
Sbjct  1059  -NKVFDREGDDGKFVSVTVKATDQGEPSLEGVCSFTVEITDVNDNPPLFDRQKYVENVKQ  1117

Query  230   NI-LSSELLRFQVTDLDEEYTDNWLAVYFFT---SGNEGNWFEIQTDPRTNEGILKVVKA  285
             +  + + +LR  V+  DE+  +N   VY  +   S N+  +FEIQ  P +   +LK  K
Sbjct  1118  DASIGTNILR--VSASDEDADNNGAIVYTLSAPHSPNDLEYFEIQ--PESGWIVLK--KP  1171

Query  286   LD-YEQLQSVKLSIAVK----------NKAEFHQSVISRYRVQSTPV----TIQVINVRE  330
             LD    + SV+    +K          + A+    +    R    P+        I++RE
Sbjct  1172  LDGCYSVSSVRERYRLKVYASDRGDPPSTADVDVELDVVDRNNKPPIWDRNIYGPIHIRE  1231

Query  331   GIAFRPASKTFTVQKGISSKKLVDYILGTYQAIDEDTNKAASNVKYVMGRNDGGYLMIDS  390
              +             GI     V Y L         TNK   +  Y+  R D G
Sbjct  1232  NVTVGTVVTAIKASSGIEGNPTVFYRL--MPGSTAQTNKL--HTFYLQQRPDNG-----D  1282

Query  391   KTAEIKFVKNMNRDSTFIVNKTITAEVLAIDEYTGKTSTGTVYVRVPDFNDNCPTAV-LE  449
               A+IK    ++ ++    N TI  E     +     S  TVY+++ D ND  P     E
Sbjct  1283  TWADIKVNHPLDFETIKEYNLTIRVENNGAQQL---ASEATVYIQLEDVNDEIPLFTERE  1339

Query  450   KDAVCSSSP  458
             ++ V    P
Sbjct  1340  QETVLEGEP  1348

 Score = 61.2 bits (147),  Expect = 5e-08, Method: Compositional matrix adjust.
 Identities = 87/366 (24%), Positives = 156/366 (43%), Gaps = 38/366 (10%)

Query  51    FVVDKNTGDINITAIVDRE-ETPSFL-ITCRALNAQGLDVEKPLILTVKILDINDNPPVF  108
             F VD+ TG+++   + DRE +   F+ +T +A +     +E     TV+I D+NDNPP+F
Sbjct  1047  FTVDEETGEVSTNKVFDREGDDGKFVSVTVKATDQGEPSLEGVCSFTVEITDVNDNPPLF  1106

Query  109   SQQIFMGEIEENSASNSLVMILNATDADEPNHLNSKIAFKIVS-QEPAGTPMFLLSRNTG  167
              +Q ++  ++++++  + ++ ++A+D D  N  N  I + + +   P     F +   +G
Sbjct  1107  DRQKYVENVKQDASIGTNILRVSASDEDADN--NGAIVYTLSAPHSPNDLEYFEIQPESG  1164

Query  168   EVRTLTNSLDREQASS-----YRLVVSGADKDGEGLSTQCECNIKVKDVNDNFPMFRDSQ  222
              +  L   LD   + S     YRL V  +D+ G+  ST            +N P   D
Sbjct  1165  WI-VLKKPLDGCYSVSSVRERYRLKVYASDR-GDPPSTADVDVELDVVDRNNKPPIWDRN  1222

Query  223   YSA--RIEENILSSELLRFQVTDLDEEYTDNWLAVYFFTSG-----NEGNWFEIQTDPRT  275
                   I EN+    ++             N    Y    G     N+ + F +Q  P
Sbjct  1223  IYGPIHIRENVTVGTVV--TAIKASSGIEGNPTVFYRLMPGSTAQTNKLHTFYLQQRPDN  1280

Query  276   NE--GILKVVKALDYEQLQSVKLSIAVKNKAEFHQSVISRYRVQSTPVTIQVINVREGIA  333
              +    +KV   LD+E ++   L+I V+N         ++       V IQ+ +V + I
Sbjct  1281  GDTWADIKVNHPLDFETIKEYNLTIRVENNG-------AQQLASEATVYIQLEDVNDEIP  1333

Query  334   FRPASKTFTVQKGISSKKLVDYILGTYQAIDEDTNKAASNVKYVM---GRNDG-GYLMID  389
                  +  TV +G      V  +     AID+D     + V Y +    RN+G  +  I+
Sbjct  1334  LFTEREQETVLEGEPIGTKVTQV----NAIDKDGTFPNNQVYYYIVDSPRNEGKDFFEIN  1389

Query  390   SKTAEI  395
              ++ EI
Sbjct  1390  LQSGEI  1395

 Score = 55.8 bits (133),  Expect = 3e-06, Method: Compositional matrix adjust.
 Identities = 49/158 (31%), Positives = 78/158 (49%), Gaps = 24/158 (15%)

Query  102   NDNPPVFSQQIFMGEIEENSASNSLVMILNATDADEPNHLNSKIAFKIVSQEPAGTP--M  159
             ND  P FSQQ++   ++EN+  N+LV  + A+D D  N     + F  V     GT
Sbjct  880   NDEEPKFSQQVYTPNVDENAGPNTLVTTVVASDKDGDN-----VRFGFVG---GGTSSGQ  931

Query  160   FLLSRNTGEVRTLTN--SLDREQASSYRLVVSGADK-------DGEGLSTQCECNIKVKD  210
             F++   TG +R  +   SLDR++   Y L V+  D        D    ++     + + D
```

```
Sbjct  932   FVIEEITGVIRLHSKAISLDRDK---YELNVTAMDDGSCCVNGDATIHTSTAVVVVFITD  988

Query  211   VNDNFPMFRD-SQYSARIEENILS-SELLRFQVTDLDE  246
             VNDN P+F+D   Y  ++EE   + S +++ Q TD D+
Sbjct  989   VNDNKPVFKDCGTYYPKVEEGAPNGSPVIKVQATDEDK  1026

 Score = 53.9 bits (128),  Expect = 1e-05, Method: Compositional matrix adjust.
 Identities = 70/306 (23%), Positives = 126/306 (41%), Gaps = 30/306 (10%)

Query  20    PIAKIT-SDYQATQKITYRI---SGVGIDQPPFGIFVVDKNTGDINITAIVDRE---ETP  72
             PI  +T  D   T K  Y++   SG G D+  F +   +  TG + I   +D E   ++
Sbjct  1800  PILTVTVHDEDETNKFQYKVIDNSGYGADK--FTMVRNNDGTGSLKIVQPLDYEDPLQSN  1857

Query  73    SFLITCRALNAQGLDVEKPLI------LTVKILDINDNPPVFSQQIFMGEIEENSASNSL  126
              F    + +N +G D +          + VK+ DINDN P F +      + EN+
Sbjct  1858  GFRFRIQ-VNDKGEDNDNDKYHVAYSWVVVKLRDINDNKPQFERPNIEVSVYENADVGKT  1916

Query  127   VMILNATDADEPNHLNSKIAFKIVSQEPAGTPMFLLSRNTGEVRTLTNSLDREQASSYRL  186
             +    ATD D+     SK+++ I   + +       S N     T+  +LDRE    +++
Sbjct  1917  LETFKATDPDQGG--KSKVSYAI---DRSSDRQRQFSINQEGTVTIQRNLDREVTPRHQV  1971

Query  187   VVSGADKDGEGLSTQCECNIKVKDVNDNFPMFRDSQYSARIEENILSSELLRFQVTDLDE  246
              +   D      +      + V+D+NDN P F    Y   + E++   +++    TD D+
Sbjct  1972  KILAIDDGIPPKTATATLTVIVQDINDNPPKFL-KDYRPVLPEHVPPRKVVEILATDDDD  2030

Query  247   EYTDNWLAVYFFTSGNEGNW----FEIQTDPRTNEG----ILKVVKALDYEQLQSVKLSI  298
                 N     F       +     F+++ D +   G    I+  +++ D EQ +   + I
Sbjct  2031  RSKSNGPPFQFRLDPGADDIIRASFKVEQDQKGANGDGMAIVSSLRSFDREQQKEYLIPI  2090

Query  299   AVKNKA  304
              +K+
Sbjct  2091  VIKDHG  2096

 Score = 51.6 bits (122),  Expect = 5e-05, Method: Compositional matrix adjust.
 Identities = 96/398 (24%), Positives = 163/398 (41%), Gaps = 54/398 (14%)

Query  106   PVFSQQIFMGEIEENSASNSLVMILNATDADEPNHLNSKIAFKIVSQ-EPAGTPMFLLSR  164
             P F    +  EI EN   +S ++ + A      +  + +I + + +Q + AGT  F +
Sbjct  677   PQFYMPSYEAEIPENQKKDSDIISVKAK-----SFADREIRYTLKAQGQGAGT--FNIGP  729

Query  165   NTGEVRTLTNSLDRE---QASSYRLVVSGADKDGEGLSTQCECNIKVKDVNDNFPMFRDS  221
              +G V+ L   LD E   Q   Y LVV+ A +D  G ST  E  I+V DVNDN P F
Sbjct  730   TSGIVK-LAKELDFEDLRQPHVYSLVVT-ATEDSGGFSTSVELTIRVTDVNDNAPKFELP  787

Query  222   QYSAR-IEENI-LSSELLRFQVTDLDEEYTDNWLAVYFFTSGNEGNWFEIQTDPRTNEGI  279
              Y A  ++E+I + + +L+ +  D D         + +  S +    F + ++     GI
Sbjct  788   DYQAHNVDEDIQIGTSILKVKAMDADSGAN---AEIEYLVSDDH---FAVDSN-----GI  836

Query  280   LKVVKALDYEQLQSV-KLSIAVKNKAEFHQSVISRYRVQSTPVTIQVINVREGIAFRPAS  338
             +   K LD +   +  +  +  K+K E  ++  +  RV +           E   F
Sbjct  837   IVNNKQLDADNNNAYYEFVVTAKDKGEPAKTGTATVRVYTKNKN------DEEPKFSQQV  890

Query  339   KTFTVQKGISSKKLVDYILGTYQAIDEDTNKAASNVKY--VMGRNDGGYLMIDSKTAEIK  396
              T  V +      LV  ++ +        +K   NV++  V G    G  +I+  T  I+
Sbjct  891   YTPNVDENAGPNTLVTTVVAS--------DKDGDNVRFGFVGGGTSSGQFVIEEITGVIR  942

Query  397   FVK---NMNRDSTFI-VNKTITAEVLAIDEYTGKTSTGTVYVRVPDFNDN------CPTA  446
                   +++RD   + V            + T  TST  V V + D NDN      C T
Sbjct  943   LHSKAISLDRDKYELNVTAMDDGSCCVNGDATIHTSTAVVVVFITDVNDNKPVFKDCGTY  1002

Query  447   VLE-KDAVCSSSPSVVVSARTLNNRYTGPYTFALEDQP  483
               + ++   + SP + V A   +    G   +++  QP
Sbjct  1003  YPKVEEGAPNGSPVIKVQATDEDKGVNGQVKYSIVQQP  1040

 Score = 44.7 bits (104),  Expect = 0.008, Method: Compositional matrix adjust.
 Identities = 48/199 (24%), Positives = 73/199 (37%), Gaps = 19/199 (10%)

Query  27    DYQATQKITYRISGVGIDQPPFGIFVVDKNTGDINITAIVDREETPSFLITCRALNAQGL  86
```

```
             D     K++Y I      Q  F I       G + I   +DRE TP   +   A++
Sbjct  1926  DQGGKSKVSYAIDRSSDRQRQFSI----NQEGTVTIQRNLDREVTPRHQVKILAIDDGIP  1981

Query  87    DVEKPLILTVKILDINDNPPVFSQQIFMGEIEENSASNSLVMILNATDADEPNHLNSKIA  146
                    LTV + DINDNPP F +  +   + E+     +V IL   D D
Sbjct  1982  PKTATATLTVIVQDINDNPPKFLKD-YRPVLPEHVPPRKVVEILATDDDDRSKSNGPPFQ  2040

Query  147   FKIVSQEPAGTPMFLLSRNTGEVRTLTN-----------SLDREQASSYRLVVSGADKDG  195
             F++   +P    +   S    + +   N           S DREQ   Y + +   D
Sbjct  2041  FRL---DPGADDIIRASFKVEQDQKGANGDGMAIVSSLRSFDREQQKEYLIPIVIKDHGN  2097

Query  196   EGLSTQCECNIKVKDVNDN  214
               ++      + + DVNDN
Sbjct  2098  PAMTGTSTLTVVIGDVNDN  2116

 Score = 40.8 bits (94),  Expect = 0.11, Method: Compositional matrix adjust.
 Identities = 55/242 (23%), Positives = 102/242 (42%), Gaps = 22/242 (9%)

Query  4    KFAKPCREGE--DNSKRNP-IAKITSDYQATQKITYRISGVGIDQPPFGIFVVDKNTGDI  60
            +F  P  E E  +N K++  I  + +   A ++I Y +   G      G F +   +G +
Sbjct  678  QFYMPSYEAEIPENQKKDSDIISVKAKSFADREIRYTLKAQGQGA---GTFNIGPTSGIV  734

Query  61   NITAIVDREE--TPSF--LITCRALNAQGLDVEKPLILTVKILDINDNPPVFSQQIFMGE  116
             +   +D E+   P    L+     ++ G      + LT+++ D+NDN P F    +
Sbjct  735  KLAKELDFEDLRQPHVYSLVVTATEDSGGFSTS--VELTIRVTDVNDNAPKFELPDYQAH  792

Query  117  -IEENSASNSLVMILNATDADEPNHLNSKIAFKIVSQEPAGTPMFLLSRNTGEVRTLTNS  175
             ++E+    + ++ + A DAD  +  N++I + +VS +      F +  N   V
Sbjct  793  NVDEDIQIGTSILKVKAMDAD--SGANAEIEY-LVSDD-----HFAVDSNGIIVNNKQLD  844

Query  176  LDREQASSYRLVVSGADKDGEGLSTQCECNIKVKDVNDNFPMFRDSQYSARIEENILSSE  235
             D   A  Y  VV+  DK     +      +  K+ ND  P F    Y+  ++EN   +
Sbjct  845  ADNNNA-YYEFVVTAKDKGEPAKTGTATVRVYTKNKNDEEPKFSQQVYTPNVDENAGPNT  903

Query  236  LL  237
            L+
Sbjct  904  LV  905

 Score = 39.3 bits (90),  Expect = 0.30, Method: Compositional matrix adjust.
 Identities = 55/225 (24%), Positives = 88/225 (39%), Gaps = 24/225 (11%)

Query  75   LITCRALNAQGLDVEKPLILTVKILDINDNPPVFSQQIF--MGEIEENSASNSLVMILNA  132
            +IT +  NA GL       + + + D+ND PP F  +       ++ N+  N+ V  L A
Sbjct  539  IITNQGHNA-GLKYTDNQRVIIHVKDVNDEPPYFINRPLPMQAVVQLNAPPNTPVFTLQA  597

Query  133  TDADEPNHLNSKIAFKIVSQEPAGTPMFLLSRNTGEVRTLTNSLDREQASSYRLVVSGAD  192
             D D     +  I + IV     G   F +   +G VRT    L  +    Y L V   D
Sbjct  598  RDPD----TDHNIHYFIVRDRTGG--RFEVDERSGVVRTRGTDL-FQLDMEYVLYVKAED  650

Query  193  KDGE----GLSTQCECNIKVKDVNDNFPMFRDSQYSARIEEN-ILSSELLRFQVTDLDEE  247
            ++G+       +  E  + +       P F    Y A I EN    S+++  +     +
Sbjct  651  QNGKVDDRRFQSTPEERLSIVG-GKRAPQFYMPSYEAEIPENQKKDSDIISVKA----KS  705

Query  248  YTDNWLAVYFFTSGNEGNWFEIQTDPRTNEGILKVVKALDYEQLQ  292
            + D  +       G     F I        GI+K+ K LD+E L+
Sbjct  706  FADREIRYTLKAQGQGAGTFNIG----PTSGIVKLAKELDFEDLR  746

>AALB20_033171.P66490 | transcript=AALB20_033171.R66490 | gene=AALB20_033171
| organism=Anopheles_albimanus_STECLA_2020 |
gene_product=neural-cadherin, transcript variant X38 |
transcript_product=neural-cadherin,
transcript variant X8 | location=CM024292:75037560-75208393(+)
| protein_length=3131 | sequence_SO=chromosome
| SO=protein_coding_gene | is_pseudo=false
Length=3131

 Score = 144 bits (364),  Expect = 1e-34, Method: Compositional matrix adjust.
 Identities = 125/437 (29%), Positives = 211/437 (48%), Gaps = 33/437 (8%)
```

```
Query  26    SDYQATQKITYRISGVGID--QPPFGIFVVDKNTGDINITAIVDREET---PSFLITCRA  80
              D    Q I Y ++G GID   P    F +++ TG+I +   +DR++    P +  T  A
Sbjct  1580  GDKDRPQNIVYFLTGQGIDPDNPANSKFDINRTTGEIFVLKPLDRDQPNGRPQWRFTVFA  1639

Query  81    LNAQGLDVEKPLILTVKILDINDNPPVFSQQIFMGEIEENSASNSLVMILNATDADEPNH  140
              +  G  +     + V + DINDN P+F Q ++ G + EN  +  +VM + A D D+PN
Sbjct  1640  QDEGGEGLVGYADVQVNLKDINDNAPIFPQGVYFGNVTENGTAGMVVMTMTAVDYDDPNE  1699

Query  141   -LNSKIAFKI---VSQEPAGTPMFLLSRNTGEVRTLTNSLDREQASSYRLVVSGADKDGE  196
               N+K+ + I   V +E  G+P+F +  +TG ++T    LDRE+   Y + V     DG
Sbjct  1700  GTNAKLIYSIEKNVIEEETGSPIFEIEADTGVIKTAVCCLDRERTPDYSIQVVAM--DGG  1757

Query  197   GLSTQCECNIKVKDVNDNFPMFRDSQYSARIEENILSS--ELLRFQVTDLDEEYTDNWLA  254
             GL      +I+VKD+ND  P F   ++   ++E   ++  E+    VT  DE+ T+ +
Sbjct  1758  GLKGTGTASIRVKDINDMPPQFTKDEWFTEVDETDGTNLPEMPILTVTVHDEDETNKFQY  1817

Query  255   VYFFTSGNEGNWFEIQTDPRTNEGI--LKVVKALDYE---QLQSVKLSIAVKNKAEFHQS  309
                  SG   + F +    R N+G   LK+V+ LDYE   Q    +  I V +K E + +
Sbjct  1818  KVIDNSGYGADKFTMV---RNNDGTGSLKIVQPLDYEDPLQSNGFRFRIQVNDKGEDNDN  1874

Query  310   VISRYRVQSTPVTIQVINVREGIAFRPASKTFTVQKGISSKKLVDYILGTYQAIDEDTNK  369
                +Y V  + V +++ ++ +    +P  +   ++  +     V   L T++A D D
Sbjct  1875  --DKYHVAYSWVVVKLRDINDN---KPQFERPNIEVSVYENADVGKTLETFKATDPDQG-  1928

Query  370   AASNVKYVMGR-NDGGYLMIDSKTAEIKFVKNMNRDSTFIVNKTITAEVLAIDE-YTGKT  427
               S V Y + R +D       ++   +   +N++R+    V      ++LAID+    KT
Sbjct  1929  GKSKVSYAIDRSSDRQRQFSINQEGTVTIQRNLDRE----VTPRHQVKILAIDDGIPPKT  1984

Query  428   STGTVYVRVPDFNDNCP  444
             +T T+ V V D NDN P
Sbjct  1985  ATATLTVIVQDINDNPP  2001

 Score = 85.9 bits (211),  Expect = 1e-15, Method: Compositional matrix adjust.
 Identities = 108/420 (26%), Positives = 177/420 (42%), Gaps = 52/420 (12%)

Query  50    IFVVDKNTGDINITAIVDREETPSFLITCRAL----------NAQGLDVEKPLILTVKIL  99
              F ++  +G+I    + DRE+  ++ +   A           N Q   V K   + + I
Sbjct  1385  FFEINLQSGEIFTKVVFDREKQGAYALEVEARDGAPSARPNSNQQPNSVTK--FIRIGIA  1442

Query  100   DINDNPPVFSQQIFMGEIEENSASNSLVMILNATDADEPNHLNSKIAFKIVSQEPAGTPM  159
             D NDNPP F + ++  E++EN      V+ + A D DE    +S+I ++I S    G
Sbjct  1443  DKNDNPPYFDKALYEAEVDENEDIQHTVLTVTAKDHDE----SSRIRYEITSGNIGGA--  1496

Query  160   FLLSRNTGEVRTLTNSLDREQASSYRLVVSGADKDGEGLSTQCECNIKVKDVNDNFPMFR  219
             F +   TG +  +  +LD E    Y L ++ +D   E  +T     I VKDVNDN P+F
Sbjct  1497  FAVKNMTGAI-YVAGALDYETRKRYELRLAASDNLKENYTTVV---IHVKDVNDNPPVFE  1552

Query  220   DSQYSARIEENI---LSSELLRFQVTDLDEEYTDNWLAVYFFTSG--NEGNWFEIQTDPR  274
                Y  +I E     L   +L+   TD D++   N   VYF T    +  N    + D
Sbjct  1553  RPTYRTQITEEDDRNLPKRVLQVTATDGDKDRPQN--IVYFLTGQGIDPDNPANSKFDIN  1610

Query  275   TNEGILKVVKALDYEQLQS-VKLSIAVKNKAEFHQSVISRYRVQSTPVTIQVINVREGIA  333
                G + V+K LD +Q     +    V  + E  + ++    VQ     + + ++ +
Sbjct  1611  RTTGEIFVLKPLDRDQPNGRPQWRFTVFAQDEGGEGLVGYADVQ-----VNLKDINDNAP  1665

Query  334   FRPASKTFTVQKGISSKKLVDYILGTYQAIDEDTNKAASNVK--YVMGRN------DGGY  385
               P    F     ++       ++ T  A+D D     +N K  Y + +N
Sbjct  1666  IFPQGVYF---GNVTENGTAGMVVMTMTAVDYDDPNEGTNAKLIYSIEKNVIEEETGSPI  1722

Query  386   LMIDSKTAEIK-FVKNMNRDSTFIVNKTITAEVLAIDEYTGKTSTGTVYVRVPDFNDNCP  444
               I++ T  IK  V  ++R+ T       + +V+A+D   G   TGT  +RV D ND  P
Sbjct  1723  FEIEADTGVIKTAVCCLDRERT----PDYSIQVVAMDG-GGLKGTGTASIRVKDINDMPP  1777

 Score = 75.9 bits (185),  Expect = 2e-12, Method: Compositional matrix adjust.
 Identities = 108/429 (25%), Positives = 178/429 (41%), Gaps = 53/429 (12%)

Query  53    VDKNTGDINITAIVDREETPSFLITCRALNAQGLDVEKPLILTVKILDINDNPPVFSQ-Q  111
             +D++  ++N+TA+ D          +C  +N          ++ V I D+NDN PVF
```

```
Sbjct  950   LDRDKYELNVTAMDDG--------SC-CVNGDATIHTSTAVVVVFITDVNDNKPVFKDCG  1000

Query  112   IFMGEIEENSASNSLVMILNATDADEPNHLNSKIAFKIVSQEPAGTPMFLLSRNTGEVRT  171
              +  ++EE + + S V+ + ATD D+   +N ++ + IV Q       F +   TGEV T
Sbjct  1001  TYYPKVEEGAPNGSPVIKVQATDEDK--GVNGQVKYSIVQQPNQKGTKFTVDEETGEVST  1058

Query  172   LTNSLDRE--QASSYRLVVSGADKDGEGLSTQCECNIKVKDVNDNFPMFRDSQYSARIEE  229
                   DRE        + V   D+    L   C   +++ DVNDN P+F   +Y   +++
Sbjct  1059  -NKVFDREGDDGKFVSVTVKATDQGEPSLEGVCSFTVEITDVNDNPPLFDRQKYVENVKQ  1117

Query  230   NI-LSSELLRFQVTDLDEEYTDNWLAVYFFT---SGNEGNWFEIQTDPRTNEGILKVVKA  285
             +  + + +LR  V+  DE+  +N   VY  +    S N+  +FEIQ  P +   +LK  K
Sbjct  1118  DASIGTNILR--VSASDEDADNNGAIVYTLSAPHSPNDLEYFEIQ--PESGWIVLK--KP  1171

Query  286   LD-YEQLQSVKLSIAVK----------NKAEFHQSVISRYRVQSTPV----TIQVINVRE  330
             LD    + SV+    +K          + A+    +    R    P+        I++RE
Sbjct  1172  LDGCYSVSSVRERYRLKVYASDRGDPPSTADVDVELDVVDRNNKPPIWDRNIYGPIHIRE  1231

Query  331   GIAFRPASKTFTVQKGISSKKLVDYILGTYQAIDEDTNKAASNVKYVMGRNDGGYLMIDS  390
              +             GI     V Y L         TNK   +  Y+  R D G
Sbjct  1232  NVTVGTVVTAIKASSGIEGNPTVFYRL--MPGSTAQTNKL--HTFYLQQRPDNG-----D  1282

Query  391   KTAEIKFVKNMNRDSTFIVNKTITAEVLAIDEYTGKTSTGTVYVRVPDFNDNCPTAV-LE  449
               A+IK    ++ ++    N TI  E     +     S  TVY+++ D ND  P     E
Sbjct  1283  TWADIKVNHPLDFETIKEYNLTIRVENNGAQQL---ASEATVYIQLEDVNDEIPLFTERE  1339

Query  450   KDAVCSSSP  458
             ++ V    P
Sbjct  1340  QETVLEGEP  1348

 Score = 61.2 bits (147),  Expect = 5e-08, Method: Compositional matrix adjust.
 Identities = 87/366 (24%), Positives = 156/366 (43%), Gaps = 38/366 (10%)

Query  51    FVVDKNTGDINITAIVDRE-ETPSFL-ITCRALNAQGLDVEKPLILTVKILDINDNPPVF  108
             F VD+ TG+++   + DRE +   F+ +T +A +     +E     TV+I D+NDNPP+F
Sbjct  1047  FTVDEETGEVSTNKVFDREGDDGKFVSVTVKATDQGEPSLEGVCSFTVEITDVNDNPPLF  1106

Query  109   SQQIFMGEIEENSASNSLVMILNATDADEPNHLNSKIAFKIVS-QEPAGTPMFLLSRNTG  167
              +Q ++  ++++++  + ++ ++A+D D  N  N  I + + +   P     F +   +G
Sbjct  1107  DRQKYVENVKQDASIGTNILRVSASDEDADN--NGAIVYTLSAPHSPNDLEYFEIQPESG  1164

Query  168   EVRTLTNSLDREQASS-----YRLVVSGADKDGEGLSTQCECNIKVKDVNDNFPMFRDSQ  222
              +  L   LD   + S     YRL V  +D+ G+  ST            +N P   D
Sbjct  1165  WI-VLKKPLDGCYSVSSVRERYRLKVYASDR-GDPPSTADVDVELDVVDRNNKPPIWDRN  1222

Query  223   YSA--RIEENILSSELLRFQVTDLDEEYTDNWLAVYFFTSG-----NEGNWFEIQTDPRT  275
                   I EN+    ++             N    Y    G     N+ + F +Q  P
Sbjct  1223  IYGPIHIRENVTVGTVV--TAIKASSGIEGNPTVFYRLMPGSTAQTNKLHTFYLQQRPDN  1280

Query  276   NE--GILKVVKALDYEQLQSVKLSIAVKNKAEFHQSVISRYRVQSTPVTIQVINVREGIA  333
              +    +KV   LD+E ++   L+I V+N         ++       V IQ+ +V + I
Sbjct  1281  GDTWADIKVNHPLDFETIKEYNLTIRVENNG-------AQQLASEATVYIQLEDVNDEIP  1333

Query  334   FRPASKTFTVQKGISSKKLVDYILGTYQAIDEDTNKAASNVKYVM---GRNDG-GYLMID  389
                  +  TV +G      V  +     AID+D     + V Y +    RN+G  +  I+
Sbjct  1334  LFTEREQETVLEGEPIGTKVTQV----NAIDKDGTFPNNQVYYYIVDSPRNEGKDFFEIN  1389

Query  390   SKTAEI  395
              ++ EI
Sbjct  1390  LQSGEI  1395

 Score = 55.8 bits (133),  Expect = 3e-06, Method: Compositional matrix adjust.
 Identities = 49/158 (31%), Positives = 78/158 (49%), Gaps = 24/158 (15%)

Query  102   NDNPPVFSQQIFMGEIEENSASNSLVMILNATDADEPNHLNSKIAFKIVSQEPAGTP--M  159
             ND  P FSQQ++   ++EN+  N+LV  + A+D D  N     + F  V     GT
Sbjct  880   NDEEPKFSQQVYTPNVDENAGPNTLVTTVVASDKDGDN-----VRFGFVG---GGTSSGQ  931

Query  160   FLLSRNTGEVRTLTN--SLDREQASSYRLVVSGADK-------DGEGLSTQCECNIKVKD  210
```

```
             F++   TG +R  +   SLDR++   Y L V+  D        D    ++     + + D
Sbjct  932   FVIEEITGVIRLHSKAISLDRDK---YELNVTAMDDGSCCVNGDATIHTSTAVVVVFITD  988

Query  211   VNDNFPMFRD-SQYSARIEENILS-SELLRFQVTDLDE  246
             VNDN P+F+D   Y  ++EE   + S +++ Q TD D+
Sbjct  989   VNDNKPVFKDCGTYYPKVEEGAPNGSPVIKVQATDEDK  1026

 Score = 53.9 bits (128),  Expect = 1e-05, Method: Compositional matrix adjust.
 Identities = 70/306 (23%), Positives = 126/306 (41%), Gaps = 30/306 (10%)

Query  20    PIAKIT-SDYQATQKITYRI---SGVGIDQPPFGIFVVDKNTGDINITAIVDRE---ETP  72
             PI  +T  D   T K  Y++   SG G D+  F +   +  TG + I   +D E   ++
Sbjct  1800  PILTVTVHDEDETNKFQYKVIDNSGYGADK--FTMVRNNDGTGSLKIVQPLDYEDPLQSN  1857

Query  73    SFLITCRALNAQGLDVEKPLI------LTVKILDINDNPPVFSQQIFMGEIEENSASNSL  126
              F    + +N +G D +          + VK+ DINDN P F +      + EN+
Sbjct  1858  GFRFRIQ-VNDKGEDNDNDKYHVAYSWVVVKLRDINDNKPQFERPNIEVSVYENADVGKT  1916

Query  127   VMILNATDADEPNHLNSKIAFKIVSQEPAGTPMFLLSRNTGEVRTLTNSLDREQASSYRL  186
             +    ATD D+     SK+++ I   + +       S N     T+  +LDRE    +++
Sbjct  1917  LETFKATDPDQGG--KSKVSYAI---DRSSDRQRQFSINQEGTVTIQRNLDREVTPRHQV  1971

Query  187   VVSGADKDGEGLSTQCECNIKVKDVNDNFPMFRDSQYSARIEENILSSELLRFQVTDLDE  246
              +   D      +      + V+D+NDN P F    Y   + E++   +++    TD D+
Sbjct  1972  KILAIDDGIPPKTATATLTVIVQDINDNPPKFL-KDYRPVLPEHVPPRKVVEILATDDDD  2030

Query  247   EYTDNWLAVYFFTSGNEGNW----FEIQTDPRTNEG----ILKVVKALDYEQLQSVKLSI  298
                 N     F       +     F+++ D +   G    I+  +++ D EQ +   + I
Sbjct  2031  RSKSNGPPFQFRLDPGADDIIRASFKVEQDQKGANGDGMAIVSSLRSFDREQQKEYLIPI  2090

Query  299   AVKNKA  304
              +K+
Sbjct  2091  VIKDHG  2096

 Score = 51.6 bits (122),  Expect = 5e-05, Method: Compositional matrix adjust.
 Identities = 96/398 (24%), Positives = 163/398 (41%), Gaps = 54/398 (14%)

Query  106   PVFSQQIFMGEIEENSASNSLVMILNATDADEPNHLNSKIAFKIVSQ-EPAGTPMFLLSR  164
             P F    +  EI EN   +S ++ + A      +  + +I + + +Q + AGT  F +
Sbjct  677   PQFYMPSYEAEIPENQKKDSDIISVKAK-----SFADREIRYTLKAQGQGAGT--FNIGP  729

Query  165   NTGEVRTLTNSLDRE---QASSYRLVVSGADKDGEGLSTQCECNIKVKDVNDNFPMFRDS  221
              +G V+ L   LD E   Q   Y LVV+ A +D  G ST  E  I+V DVNDN P F
Sbjct  730   TSGIVK-LAKELDFEDLRQPHVYSLVVT-ATEDSGGFSTSVELTIRVTDVNDNAPKFELP  787

Query  222   QYSAR-IEENI-LSSELLRFQVTDLDEEYTDNWLAVYFFTSGNEGNWFEIQTDPRTNEGI  279
              Y A  ++E+I + + +L+ +  D D         + +  S +    F + ++     GI
Sbjct  788   DYQAHNVDEDIQIGTSILKVKAMDADSGAN---AEIEYLVSDDH---FAVDSN-----GI  836

Query  280   LKVVKALDYEQLQSV-KLSIAVKNKAEFHQSVISRYRVQSTPVTIQVINVREGIAFRPAS  338
             +   K LD +   +  +  +  K+K E  ++  +  RV +           E   F
Sbjct  837   IVNNKQLDADNNNAYYEFVVTAKDKGEPAKTGTATVRVYTKNKN------DEEPKFSQQV  890

Query  339   KTFTVQKGISSKKLVDYILGTYQAIDEDTNKAASNVKY--VMGRNDGGYLMIDSKTAEIK  396
              T  V +      LV  ++ +        +K   NV++  V G    G  +I+  T  I+
Sbjct  891   YTPNVDENAGPNTLVTTVVAS--------DKDGDNVRFGFVGGGTSSGQFVIEEITGVIR  942

Query  397   FVK---NMNRDSTFI-VNKTITAEVLAIDEYTGKTSTGTVYVRVPDFNDN------CPTA  446
                   +++RD   + V            + T  TST  V V + D NDN      C T
Sbjct  943   LHSKAISLDRDKYELNVTAMDDGSCCVNGDATIHTSTAVVVVFITDVNDNKPVFKDCGTY  1002

Query  447   VLE-KDAVCSSSPSVVVSARTLNNRYTGPYTFALEDQP  483
               + ++   + SP + V A   +    G   +++  QP
Sbjct  1003  YPKVEEGAPNGSPVIKVQATDEDKGVNGQVKYSIVQQP  1040

 Score = 44.7 bits (104),  Expect = 0.008, Method: Compositional matrix adjust.
 Identities = 48/199 (24%), Positives = 73/199 (37%), Gaps = 19/199 (10%)
```

```
Query  27    DYQATQKITYRISGVGIDQPPFGIFVVDKNTGDINITAIVDREETPSFLITCRALNAQGL  86
             D     K++Y I      Q  F I       G + I   +DRE TP   +   A++
Sbjct  1926  DQGGKSKVSYAIDRSSDRQRQFSI----NQEGTVTIQRNLDREVTPRHQVKILAIDDGIP  1981

Query  87    DVEKPLILTVKILDINDNPPVFSQQIFMGEIEENSASNSLVMILNATDADEPNHLNSKIA  146
                    LTV + DINDNPP F +  +   + E+     +V IL   D D
Sbjct  1982  PKTATATLTVIVQDINDNPPKFLKD-YRPVLPEHVPPRKVVEILATDDDDRSKSNGPPFQ  2040

Query  147   FKIVSQEPAGTPMFLLSRNTGEVRTLTN-----------SLDREQASSYRLVVSGADKDG  195
             F++   +P    +   S    + +   N           S DREQ   Y + +   D
Sbjct  2041  FRL---DPGADDIIRASFKVEQDQKGANGDGMAIVSSLRSFDREQQKEYLIPIVIKDHGN  2097

Query  196   EGLSTQCECNIKVKDVNDN  214
               ++      + + DVNDN
Sbjct  2098  PAMTGTSTLTVVIGDVNDN  2116

 Score = 40.8 bits (94),  Expect = 0.11, Method: Compositional matrix adjust.
 Identities = 55/242 (23%), Positives = 102/242 (42%), Gaps = 22/242 (9%)

Query  4    KFAKPCREGE--DNSKRNP-IAKITSDYQATQKITYRISGVGIDQPPFGIFVVDKNTGDI  60
            +F  P  E E  +N K++  I  +  +   A ++I Y +   G      G F +   +G +
Sbjct  678  QFYMPSYEAEIPENQKKDSDIISVKAKSFADREIRYTLKAQGQGA---GTFNIGPTSGIV  734

Query  61   NITAIVDREE--TPSF--LITCRALNAQGLDVEKPLILTVKILDINDNPPVFSQQIFMGE  116
             +   +D E+   P    L+     ++ G      + LT+++ D+NDN P F    +
Sbjct  735  KLAKELDFEDLRQPHVYSLVVTATEDSGGFSTS--VELTIRVTDVNDNAPKFELPDYQAH  792

Query  117  -IEENSASNSLVMILNATDADEPNHLNSKIAFKIVSQEPAGTPMFLLSRNTGEVRTLTNS  175
             ++E+    + ++ + A DAD  +  N++I + +VS +      F +  N   V
Sbjct  793  NVDEDIQIGTSILKVKAMDAD--SGANAEIEY-LVSDD-----HFAVDSNGIIVNNKQLD  844

Query  176  LDREQASSYRLVVSGADKDGEGLSTQCECNIKVKDVNDNFPMFRDSQYSARIEENILSSE  235
             D   A  Y  VV+  DK     +      +  K+ ND  P F    Y+  ++EN   +
Sbjct  845  ADNNNA-YYEFVVTAKDKGEPAKTGTATVRVYTKNKNDEEPKFSQQVYTPNVDENAGPNT  903

Query  236  LL  237
            L+
Sbjct  904  LV  905

 Score = 39.3 bits (90),  Expect = 0.30, Method: Compositional matrix adjust.
 Identities = 55/225 (24%), Positives = 88/225 (39%), Gaps = 24/225 (11%)

Query  75   LITCRALNAQGLDVEKPLILTVKILDINDNPPVFSQQIF--MGEIEENSASNSLVMILNA  132
            +IT +  NA GL       + + + D+ND PP F  +       ++ N+  N+ V  L A
Sbjct  539  IITNQGHNA-GLKYTDNQRVIIHVKDVNDEPPYFINRPLPMQAVVQLNAPPNTPVFTLQA  597

Query  133  TDADEPNHLNSKIAFKIVSQEPAGTPMFLLSRNTGEVRTLTNSLDREQASSYRLVVSGAD  192
             D D     +  I + IV     G   F +   +G VRT    L  +    Y L V   D
Sbjct  598  RDPD----TDHNIHYFIVRDRTGG--RFEVDERSGVVRTRGTDL-FQLDMEYVLYVKAED  650

Query  193  KDGE----GLSTQCECNIKVKDVNDNFPMFRDSQYSARIEEN-ILSSELLRFQVTDLDEE  247
            ++G+       +  E  + +       P F    Y A I EN    S+++  +     +
Sbjct  651  QNGKVDDRRFQSTPEERLSIVG-GKRAPQFYMPSYEAEIPENQKKDSDIISVKA----KS  705

Query  248  YTDNWLAVYFFTSGNEGNWFEIQTDPRTNEGILKVVKALDYEQLQ  292
            + D  +       G     F I        GI+K+ K LD+E L+
Sbjct  706  FADREIRYTLKAQGQGAGTFNIG----PTSGIVKLAKELDFEDLR  746

>ACMO_007424.P13499 | transcript=ACMO_007424.R13499 | gene=ACMO_007424
| organism=Anopheles_coluzzii_MOPTI | gene_product=neural-cadherin,
transcript variant X4 | transcript_product=neural-cadherin,
transcript variant X12 | location=CM029349:40739311-40945968(+)
| protein_length=3100 | sequence_SO=chromosome
| SO=protein_coding_gene | is_pseudo=false
Length=3100

 Score = 144 bits (364),  Expect = 2e-34, Method: Compositional matrix adjust.
 Identities = 125/437 (29%), Positives = 211/437 (48%), Gaps = 33/437 (8%)
```

```
Query  26    SDYQATQKITYRISGVGID--QPPFGIFVVDKNTGDINITAIVDREET---PSFLITCRA  80
              D    Q I Y ++G GID   P    F +++ TG+I +   +DR++    P +  T  A
Sbjct  1550  GDKDRPQNIVYFLTGQGIDPDNPANSKFDINRTTGEIFVLKPLDRDQPNGRPQWRFTVFA  1609

Query  81    LNAQGLDVEKPLILTVKILDINDNPPVFSQQIFMGEIEENSASNSLVMILNATDADEPNH  140
              +  G  +      + V + DINDN P+F Q ++ G + EN  +  +VM + A D D+PN
Sbjct  1610  QDEGGEGLVGYADVQVNLKDINDNAPIFPQGVYFGNVTENGTAGMVVMTMTAVDYDDPNE  1669

Query  141   -LNSKIAFKI---VSQEPAGTPMFLLSRNTGEVRTLTNSLDREQASSYRLVVSGADKDGE  196
               N+K+ + I   V +E  G+P+F +  +TG ++T    LDRE+   Y + V     DG
Sbjct  1670  GTNAKLIYSIEKNVIEEETGSPIFEIEADTGVIKTAVCCLDRERTPDYSIQVVAM--DGG  1727

Query  197   GLSTQCECNIKVKDVNDNFPMFRDSQYSARIEENILSS--ELLRFQVTDLDEEYTDNWLA  254
             GL      +I+VKD+ND  P F   ++   ++E   ++  E+    VT  DE+ T+ +
Sbjct  1728  GLKGTGTASIRVKDINDMPPQFTKDEWFTEVDETDGTNLPEMPILTVTVHDEDETNKFQY  1787

Query  255   VYFFTSGNEGNWFEIQTDPRTNEGI--LKVVKALDYE---QLQSVKLSIAVKNKAEFHQS  309
                  SG   + F +    R N+G   LK+V+ LDYE   Q    +  I V +K E + +
Sbjct  1788  KVIDNSGYGADKFTMV---RNNDGTGSLKIVQPLDYEDPLQSNGFRFRIQVNDKGEDNDN  1844

Query  310   VISRYRVQSTPVTIQVINVREGIAFRPASKTFTVQKGISSKKLVDYILGTYQAIDEDTNK  369
                +Y V  + V +++ ++ +    +P  +   ++  +     V   L T++A D D
Sbjct  1845  --DKYHVAYSWVVVKLRDINDN---KPQFERPNIEVSVYENADVGKTLETFKATDPDQG-  1898

Query  370   AASNVKYVMGR-NDGGYLMIDSKTAEIKFVKNMNRDSTFIVNKTITAEVLAIDE-YTGKT  427
               S V Y + R +D       ++   +   +N++R+    V      ++LAID+    KT
Sbjct  1899  GKSKVSYAIDRSSDRQRQFSINQEGTVTIQRNLDRE----VTPRHQVKILAIDDGIPPKT  1954

Query  428   STGTVYVRVPDFNDNCP  444
             +T T+ V V D NDN P
Sbjct  1955  ATATLTVIVQDINDNPP  1971

 Score = 85.9 bits (211),  Expect = 1e-15, Method: Compositional matrix adjust.
 Identities = 108/420 (26%), Positives = 177/420 (42%), Gaps = 52/420 (12%)

Query  50    IFVVDKNTGDINITAIVDREETPSFLITCRAL----------NAQGLDVEKPLILTVKIL  99
              F ++  +G+I    + DRE+  ++ +   A           N Q   V K   + + I
Sbjct  1355  FFEINLQSGEIFTKVVFDREKQGAYALEVEARDGAPSARPNSNQQPNSVTK--FIRIGIA  1412

Query  100   DINDNPPVFSQQIFMGEIEENSASNSLVMILNATDADEPNHLNSKIAFKIVSQEPAGTPM  159
             D NDNPP F + ++  E++EN      V+ + A D DE    +S+I ++I S    G
Sbjct  1413  DKNDNPPYFDKALYEAEVDENEDIQHTVLTVTAKDHDE----SSRIRYEITSGNIGGA--  1466

Query  160   FLLSRNTGEVRTLTNSLDREQASSYRLVVSGADKDGEGLSTQCECNIKVKDVNDNFPMFR  219
             F +   TG +  +  +LD E    Y L ++ +D   E  +T     I VKDVNDN P+F
Sbjct  1467  FAVKNMTGAI-YVAGALDYETRKRYELRLAASDNLKENYTTVV---IHVKDVNDNPPVFE  1522

Query  220   DSQYSARIEENI---LSSELLRFQVTDLDEEYTDNWLAVYFFTSG--NEGNWFEIQTDPR  274
                Y  +I E     L   +L+   TD D++   N   VYF T    +  N    + D
Sbjct  1523  RPTYRTQITEEDDRNLPKRVLQVTATDGDKDRPQN--IVYFLTGQGIDPDNPANSKFDIN  1580

Query  275   TNEGILKVVKALDYEQLQS-VKLSIAVKNKAEFHQSVISRYRVQSTPVTIQVINVREGIA  333
                G + V+K LD +Q     +    V  + E  + ++    VQ     + + ++ +
Sbjct  1581  RTTGEIFVLKPLDRDQPNGRPQWRFTVFAQDEGGEGLVGYADVQ-----VNLKDINDNAP  1635

Query  334   FRPASKTFTVQKGISSKKLVDYILGTYQAIDEDTNKAASNVK--YVMGRN------DGGY  385
               P    F     ++       ++ T  A+D D     +N K  Y + +N
Sbjct  1636  IFPQGVYF---GNVTENGTAGMVVMTMTAVDYDDPNEGTNAKLIYSIEKNVIEEETGSPI  1692

Query  386   LMIDSKTAEIK-FVKNMNRDSTFIVNKTITAEVLAIDEYTGKTSTGTVYVRVPDFNDNCP  444
               I++ T  IK  V  ++R+ T       + +V+A+D   G   TGT  +RV D ND  P
Sbjct  1693  FEIEADTGVIKTAVCCLDRERT----PDYSIQVVAMDG-GGLKGTGTASIRVKDINDMPP  1747

 Score = 79.0 bits (193),  Expect = 2e-13, Method: Compositional matrix adjust.
 Identities = 106/424 (25%), Positives = 180/424 (42%), Gaps = 51/424 (12%)

Query  53    VDKNTGDINITAIVDREETPSFLITCRALNAQGLDVEKPLILTVKILDINDNPPVFSQ-Q  111
             +D++  ++N+TA+ D         +C  +N          ++ V I D+NDN PVF
```

```
Sbjct  928   LDRDKYELNVTAMDDG--------SC-CVNGDATIHTSTAVVVVFITDVNDNKPVFKDCG  978

Query  112   IFMGEIEENSASNSLVMILNATDADEPNHLNSKIAFKIVSQEPAGTPMFLLSRNTGEVRT  171
              +  ++EE + + S V+ + ATD D+   +N ++ + IV Q       F +   TGEV T
Sbjct  979   TYYPKVEEGAPNGSPVIKVQATDEDK--GVNGQVKYSIVQQPNQKGTKFTVDEETGEVST  1036

Query  172   LTNSLDRE--QASSYRLVVSGADKDGEGLSTQCECNIKVKDVNDNFPMFRDSQYSARIEE  229
                  DRE        + V   D+    L   C   +++ DVNDN P+F   +Y   +++
Sbjct  1037  -NKVFDREGDDGKFVSVTVKATDQGEPSLEGVCSFTVEITDVNDNPPLFDRQKYVENVKQ  1095

Query  230   NI-LSSELLRFQVTDLDEEYTDNWLAVYFFT---SGNEGNWFEIQTDPRTNEGILKVVKA  285
             +  + + +LR  V+  DE+  +N   VY  +   S N+  +FEIQ  P +   +LK  K
Sbjct  1096  DASIGTNILR--VSASDEDADNNGAIVYTLSAPHSPNDLEYFEIQ--PESGWIVLK--KP  1149

Query  286   LDYEQLQSVKLSIAVKNK------AEFHQSVISRYRVQSTPVTIQV----INVREGIAFR  335
             LD E+    +L +   ++       A+    +    R    P+  +     I++RE +
Sbjct  1150  LDRERY---RLKVYASDRGDPPSTADVDVELDVVDRNNKPPIWDRTIYGPIHIRENVTVG  1206

Query  336   PASKTFTVQKGISSKKLVDYILGTYQAIDEDTNKAASNVKYVMGRNDGGYLMIDSKTAEI  395
                 +     GI     V Y L         TNK   +  Y+  R D G        A+I
Sbjct  1207  TVVTSVKASSGIEGNPTVFYRL--MPGSTAQTNKL--HTFYLQQRPDNG-----DTWADI  1257

Query  396   KFVKNMNRDSTFIVNKTITAEVLAIDEYTGKTSTGTVYYRVPDFNDNCPTAV-LEKDAVC  454
             K    ++ ++    N T+  E     +      S  TVY+++ D ND  P     E++ V
Sbjct  1258  KVNHPLDFETIKEYNLTVRVENNGAQQL---ASEATVYIQLEDVNDEIPLFTEREQETVL  1314

Query  455   SSSP  458
                P
Sbjct  1315  EGEP  1318

 Score = 65.9 bits (159),  Expect = 2e-09, Method: Compositional matrix adjust.
 Identities = 85/359 (24%), Positives = 153/359 (43%), Gaps = 32/359 (9%)

Query  51    FVVDKNTGDINITAIVDRE-ETPSFL-ITCRALNAQGLDVEKPLILTVKILDINDNPPVF  108
             F VD+ TG+++   + DRE +   F+ +T +A +     +E     TV+I D+NDNPP+F
Sbjct  1025  FTVDEETGEVSTNKVFDREGDDGKFVSVTVKATDQGEPSLEGVCSFTVEITDVNDNPPLF  1084

Query  109   SQQIFMGEIEENSASNSLVMILNATDADEPNHLNSKIAFKIVS-QEPAGTPMFLLSRNTG  167
              +Q ++  ++++++  + ++ ++A+D D  N  N  I + + +   P     F +   +G
Sbjct  1085  DRQKYVENVKQDASIGTNILRVSASDEDADN--NGAIVYTLSAPHSPNDLEYFEIQPESG  1142

Query  168   EVRTLTNSLDREQASSYRLVVSGADKDGEGLSTQCECNIKVKDVNDNFPMFRDSQYSARI  227
              +  L   LDRE+   YRL V  +D+ G+  ST             +N P   D      I
Sbjct  1143  WI-VLKKPLDRER---YRLKVYASDR-GDPPSTADVDVELDVVDRNNKPPIWDRTIYGPI  1197

Query  228   EENILSSELLRFQVTDLDEEYTDNWLAVYFFTSG-----NEGNWFEIQTDPRTNE--GIL  280
                   +                N    Y    G     N+ + F +Q  P   +    +
Sbjct  1198  HIRENVTVGTVVTSVKASSGIEGNPTVFYRLMPGSTAQTNKLHTFYLQQRPDNGDTWADI  1257

Query  281   KVVKALDYEQLQSVKLSIAVKNKAEFHQSVISRYRVQSTPVTIQVINVREGIAFRPASKT  340
             KV   LD+E ++   L++ V+N          ++       V IQ+ +V + I      +
Sbjct  1258  KVNHPLDFETIKEYNLTVRVENNG-------AQQLASEATVYIQLEDVNDEIPLFTEREQ  1310

Query  341   FTVQKGISSKKLVDYILGTYQAIDEDTNKAASNVKYVM---GRNDG-GYLMIDSKTAEI  395
              TV +G      V  +     AID+D     + V Y +    RN+G  +  I+ ++ EI
Sbjct  1311  ETVLEGEPIGTKVTQV----NAIDKDGTFPNNQVYYYIVDSPRNEGKDFFEINLQSGEI  1365

 Score = 55.8 bits (133),  Expect = 3e-06, Method: Compositional matrix adjust.
 Identities = 49/158 (31%), Positives = 78/158 (49%), Gaps = 24/158 (15%)

Query  102   NDNPPVFSQQIFMGEIEENSASNSLVMILNATDADEPNHLNSKIAFKIVSQEPAGTP--M  159
             ND  P FSQQ++   ++EN+  N+LV  + A+D D  N     + F  V     GT
Sbjct  858   NDEEPKFSQQVYTPNVDENAGPNTLVTTVVASDKDGDN-----VRFGFVG---GGTSSGQ  909

Query  160   FLLSRNTGEVRTLTN--SLDREQASSYRLVVSGADK-------DGEGLSTQCECNIKVKD  210
             F++   TG +R  +   SLDR++   Y L V+  D       D    ++     + + D
Sbjct  910   FVIEEITGVIRLHSKAISLDRDK---YELNVTAMDDGSCCVNGDATIHTSTAVVVVFITD  966

Query  211   VNDNFPMFRD-SQYSARIEENILS-SELLRFQVTDLDE  246
```

```
             VNDN P+F+D   Y  ++EE   + S +++ Q TD D+
Sbjct  967   VNDNKPVFKDCGTYYPKVEEGAPNGSPVIKVQATDEDK  1004

 Score = 53.9 bits (128),  Expect = 1e-05, Method: Compositional matrix adjust.
 Identities = 70/306 (23%), Positives = 126/306 (41%), Gaps = 30/306 (10%)

Query  20    PIAKIT-SDYQATQKITYRI---SGVGIDQPPFGIFVVDKNTGDINITAIVDRE---ETP  72
             PI  +T  D   T K  Y++   SG G D+  F +   +  TG + I   +D E   ++
Sbjct  1770  PILTVTVHDEDETNKFQYKVIDNSGYGADK--FTMVRNNDGTGSLKIVQPLDYEDPLQSN  1827

Query  73    SFLITCRALNAQGLDVEKPLI------LTVKILDINDNPPVFSQQIFMGEIEENSASNSL  126
              F    + +N +G D +           + VK+ DINDN P F +       + EN+
Sbjct  1828  GFRFRIQ-VNDKGEDNDNDKYHVAYSWVVVKLRDINDNKPQFERPNIEVSVYENADVGKT  1886

Query  127   VMILNATDADEPNHLNSKIAFKIVSQEPAGTPMFLLSRNTGEVRTLTNSLDREQASSYRL  186
             +    ATD D+      SK+++ I    + +        S N      T+  +LDRE    +++
Sbjct  1887  LETFKATDPDQGG--KSKVSYAI---DRSSDRQRQFSINQEGTVTIQRNLDREVTPRHQV  1941

Query  187   VVSGADKDGEGLSTQCECNIKVKDVNDNFPMFRDSQYSARIEENILSSELLRFQVTDLDE  246
              +    D      +      + V+D+NDN  P F    Y   + E++   +++    TD D+
Sbjct  1942  KILAIDDGIPPKTATATLTVIVQDINDNPPKFL-KDYRPVLPEHVPPRKVVEILATDDDD  2000

Query  247   EYTDNWLAVYFFTSGNEGNW----FEIQTDPRTNEG----ILKVVKALDYEQLQSVKLSI  298
                 N     F       +     F+++ D +   G    I+  +++ D EQ +   + I
Sbjct  2001  RSKSNGPPFQFRLDPGADDIIRASFKVEQDQKGANGDGMAIVSSLRSFDREQQKEYLIPI  2060

Query  299   AVKNKA  304
              +K+
Sbjct  2061  VIKDHG  2066

 Score = 52.0 bits (123),  Expect = 4e-05, Method: Compositional matrix adjust.
 Identities = 97/398 (24%), Positives = 163/398 (41%), Gaps = 54/398 (14%)

Query  106   PVFSQQIFMGEIEENSASNSLVMILNATDADEPNHLNSKIAFKIVSQ-EPAGTPMFLLSR  164
             P F    +  EI EN   +S ++ + A       +  + +I + + +Q + AGT  F +
Sbjct  655   PQFYMPSYEAEIPENQKKDSDIISVKAK-----SFADREIRYTLKAQGQGAGT--FNIGP  707

Query  165   NTGEVRTLTNSLDRE---QASSYRLVVSGADKDGEGLSTQCECNIKVKDVNDNFPMFRDS  221
              +G V+ L   LD E   Q   Y LVV+ A +D  G ST  E  I+V DVNDN P F
Sbjct  708   TSGIVK-LAKELDFEDLRQPHVYSLVVT-ATEDSGGFSTSVELTIRVTDVNDNAPKFELP  765

Query  222   QYSAR-IEENI-LSSELLRFQVTDLDEEYTDNWLAVYFFTSGNEGNWFEIQTDPRTNEGI  279
              Y A  ++E+I L + +L+ +  D D          + +  S +    F + ++     GI
Sbjct  766   DYQAHNVDEDIPLGTSILKVKAMDSDSGAN---AEIEYLVSDDH---FAVDSN-----GI  814

Query  280   LKVVKALDYEQLQS-VKLSIAVKNKAEFHQSVISRYRVQSTPVTIQVINVREGIAFRPAS  338
             +   K LD +   +  +  +  K+K E  ++  +  RV +            E   F
Sbjct  815   IVNNKQLDADNNNAYYEFVVTAKDKGEPAKTGTATVRVYTKNKN------DEEPKFSQQV  868

Query  339   KTFTVQKGISSKKLVDYILGTYQAIDEDTNKAASNVK--YVMGRNDGGYLMIDSKTAEIK  396
              T  V +      LV  ++          ++K   NV+  +V G    G  +I+  T  I+
Sbjct  869   YTPNVDENAGPNTLVTTVVA--------SDKDGDNVRFGFVGGGTSSGQFVIEEITGVIR  920

Query  397   FVK---NMNRDSTFI-VNKTITAEVLAIDEYTGKTSTGTVYVRVPDFNDN------CPTA  446
                   +++RD   + V              + T  TST  V V + D NDN      C T
Sbjct  921   LHSKAISLDRDKYELNVTAMDDGSCCVNGDATIHTSTAVVVVFITDVNDNKPVFKDCGTY  980

Query  447   VLE-KDAVCSSSPSVVVSARTLNNRYTGPYTFALEDQP  483
               + ++   + SP + V A    +   G   +++  QP
Sbjct  981   YPKVEEGAPNGSPVIKVQATDEDKGVNGQVKYSIVQQP  1018

 Score = 44.7 bits (104),  Expect = 0.007, Method: Compositional matrix adjust.
 Identities = 48/199 (24%), Positives = 73/199 (37%), Gaps = 19/199 (10%)

Query  27    DYQATQKITYRISGVGIDQPPFGIFVVDKNTGDINITAIVDREETPSFLITCRALNAQGL  86
             D     K++Y I      Q  F I        G + I   +DRE TP   +    A++
Sbjct  1896  DQGGKSKVSYAIDRSSDRQRQFSI----NQEGTVTIQRNLDREVTPRHQVKILAIDDGIP  1951
```

```
Query  87    DVEKPLILTVKILDINDNPPVFSQQIFMGEIEENSASNSLVMILNATDADEPNHLNSKIA  146
                    LTV + DINDNPP F +  +   + E+    +V IL   D D
Sbjct  1952  PKTATATLTVIVQDINDNPPKFLKD-YRPVLPEHVPPRKVVEILATDDDDRSKSNGPPFQ  2010

Query  147   FKIVSQEPAGTPMFLLSRNTGEVRTLTN-----------SLDREQASSYRLVVSGADKDG  195
             F++   +P    +   S    + +   N           S DREQ   Y + +   D
Sbjct  2011  FRL---DPGADDIIRASFKVEQDQKGANGDGMAIVSSLRSFDREQQKEYLIPIVIKDHGN  2067

Query  196   EGLSTQCECNIKVKDVNDN  214
               ++      + + DVNDN
Sbjct  2068  PAMTGTSTLTVVIGDVNDN  2086

 Score = 40.0 bits (92),  Expect = 0.20, Method: Compositional matrix adjust.
 Identities = 89/389 (23%), Positives = 149/389 (38%), Gaps = 62/389 (16%)

Query  75   LITCRALNAQGLDVEKPLILTVKILDINDNPPVFSQQIF--MGEIEENSASNSLVMILNA  132
            +IT +  NA GL       + + + D+ND PP F  +       ++ N+  N+ V  L A
Sbjct  517  IITNQGHNA-GLKYTDNQRVIIHVKDVNDEPPYFINRPLPMQAVVQLNAPPNTPVFTLQA  575

Query  133  TDADEPNHLNSKIAFKIVSQEPAGTPMFLLSRNTGEVRTLTNSLDREQASSYRLVVSGAD  192
             D D     +  I + IV     G   F +   +G VRT    L  +    Y L V   D
Sbjct  576  RDPD----TDHNIHYFIVRDRTGG--RFEVDERSGVVRTRGTDL-FQLDMEYVLYVKAED  628

Query  193  KDGE----GLSTQCECNIKVKDVNDNFPMFRDSQYSARIEEN-ILSSELLRFQVTDLDEE  247
            ++G+       +  E  + +       P F    Y A I EN    S+++  +     +
Sbjct  629  QNGKVDDRRFQSTPEERLSIVG-GKRAPQFYMPSYEAEIPENQKKDSDIISVKA----KS  683

Query  248  YTDNWLAVYFFTSGNEGNWFEIQTDPRTNEGILKVVKALDYEQLQSVKLS----IAVKNK  303
            + D  +       G     F I        GI+K+ K LD+E L+   +      A ++
Sbjct  684  FADREIRYTLKAQGQGAGTFNIGP----TSGIVKLAKELDFEDLRQPHVYSLVVTATEDS  739

Query  304  AEFHQSVISRYRVQSTPVTIQVINVREGIAFRPASKTFTVQKGISSKKLVDYILGT----  359
              F  SV          +TI+V +V +       +  F +    +     D  LGT
Sbjct  740  GGFSTSV---------ELTIRVTDVNDN------APKFELPDYQAHNVDEDIPLGTSILK  784

Query  360  YQAIDEDTNKAASNVKYVMGRN----DGGYLMIDSKTAEIKFVKNMNRDSTFIVNKTITA  415
             +A+D D+  A + ++Y++  +    D   +++++K  +     N N    F+V
Sbjct  785  VKAMDSDSG-ANAEIEYLVSDDHFAVDSNGIIVNNKQLD---ADNNNAYYEFVVTAKDKG  840

Query  416  EVLAIDEYTGKTSTGTVYVRVPDFNDNCP  444
            E         KT T TV V   + ND  P
Sbjct  841  E-------PAKTGTATVRVYTKNKNDEEP  862

 Score = 39.7 bits (91),  Expect = 0.25, Method: Compositional matrix adjust.
 Identities = 54/242 (22%), Positives = 102/242 (42%), Gaps = 22/242 (9%)

Query  4    KFAKPCREGE--DNSKRNP-IAKITSDYQATQKITYRISGVGIDQPPFGIFVVDKNTGDI  60
            +F  P  E E  +N K++  I  + +   A ++I Y +   G      G F +   +G +
Sbjct  656  QFYMPSYEAEIPENQKKDSDIISVKAKSFADREIRYTLKAQGQGA---GTFNIGPTSGIV  712

Query  61   NITAIVDREE--TPSF--LITCRALNAQGLDVEKPLILTVKILDINDNPPVFSQQIFMGE  116
             +   +D E+   P    L+     ++ G      + LT+++ D+NDN P F    +
Sbjct  713  KLAKELDFEDLRQPHVYSLVVTATEDSGGFSTS--VELTIRVTDVNDNAPKFELPDYQAH  770

Query  117  -IEENSASNSLVMILNATDADEPNHLNSKIAFKIVSQEPAGTPMFLLSRNTGEVRTLTNS  175
             ++E+    + ++ + A D+D  +  N++I + +VS +      F +  N   V
Sbjct  771  NVDEDIPLGTSILKVKAMDSD--SGANAEIEY-LVSDD-----HFAVDSNGIIVNNKQLD  822

Query  176  LDREQASSYRLVVSGADKDGEGLSTQCECNIKVKDVNDNFPMFRDSQYSARIEENILSSE  235
             D   A  Y  VV+  DK     +      +  K+ ND  P F    Y+  ++EN   +
Sbjct  823  ADNNNA-YYEFVVTAKDKGEPAKTGTATVRVYTKNKNDEEPKFSQQVYTPNVDENAGPNT  881

Query  236  LL  237
            L+
Sbjct  882  LV  883

Lambda      K        H
```

```
   0.315    0.132    0.378

Gapped
Lambda      K        H
   0.267   0.0410    0.140

Effective search space used: 188825744674

  Database: /eupath/data/apiSiteFilesStaging/VectorBase/49/real/webServices/Vect
orBase/release-
CURRENT/AaegyptiLVP_AGWG/blast/AaegyptiLVP_AGWGAnnotatedProteins
    Posted date:  Dec 12, 2023  10:17 AM
  Number of letters in database: 20,223,379
  Number of sequences in database:  28,391

  Database: /eupath/data/apiSiteFilesStaging/VectorBase/49/real/webServices/Vect
orBase/release-
CURRENT/AalbimanusSTECLA2020/blast/AalbimanusSTECLA2020AnnotatedProt
eins
    Posted date:  Sep 27, 2023  7:35 PM
  Number of letters in database: 19,833,488
  Number of sequences in database:  23,947

  Database: /eupath/data/apiSiteFilesStaging/VectorBase/49/real/webServices/Vect
orBase/release-
CURRENT/AalbimanusSTECLA/blast/AalbimanusSTECLAAnnotatedProteins
    Posted date:  Sep 27, 2023  7:42 PM
  Number of letters in database: 7,382,630
  Number of sequences in database:  12,788

  Database: /eupath/data/apiSiteFilesStaging/VectorBase/49/real/webServices/Vect
orBase/release-CURRENT/AalbopictusC6-36/blast/AalbopictusC6-
36AnnotatedProteins
    Posted date:  Sep 27, 2023  9:21 PM
  Number of letters in database: 27,840,591
  Number of sequences in database:  42,912

  Database: /eupath/data/apiSiteFilesStaging/VectorBase/49/real/webServices/Vect
orBase/release-
CURRENT/AalbopictusFoshan/blast/AalbopictusFoshanAnnotatedProteins
    Posted date:  Sep 27, 2023  11:27 PM
  Number of letters in database: 8,055,056
  Number of sequences in database:  17,535

  Database: /eupath/data/apiSiteFilesStaging/VectorBase/49/real/webServices/Vect
orBase/release-
CURRENT/AalbopictusFoshanFPA/blast/AalbopictusFoshanFPAAnnotatedProt
eins
    Posted date:  Sep 27, 2023  10:46 PM
  Number of letters in database: 24,517,600
  Number of sequences in database:  40,073

  Database: /eupath/data/apiSiteFilesStaging/VectorBase/49/real/webServices/Vect
orBase/release-
CURRENT/AaquasalisAaquGF1/blast/AaquasalisAaquGF1AnnotatedProteins
    Posted date:  Sep 27, 2023  11:10 AM
  Number of letters in database: 15,770,026
  Number of sequences in database:  20,271

  Database: /eupath/data/apiSiteFilesStaging/VectorBase/49/real/webServices/Vect
orBase/release-
CURRENT/AarabiensisDONGOLA2021/blast/AarabiensisDONGOLA2021Annotated
Proteins
    Posted date:  Jan 4, 2024  3:41 PM
  Number of letters in database: 20,363,936
  Number of sequences in database:  25,532

  Database: /eupath/data/apiSiteFilesStaging/VectorBase/49/real/webServices/Vect
orBase/release-
CURRENT/AarabiensisDongola/blast/AarabiensisDongolaAnnotatedProteins
```

```
    Posted date:  Dec 29, 2023  9:40 PM
  Number of letters in database: 7,979,927
  Number of sequences in database:  14,037

  Database: /eupath/data/apiSiteFilesStaging/VectorBase/49/real/webServices/Vect
orBase/release-
CURRENT/AatroparvusEBRO/blast/AatroparvusEBROAnnotatedProteins
    Posted date:  Sep 27, 2023  8:02 PM
  Number of letters in database: 7,248,879
  Number of sequences in database:  13,776

  Database: /eupath/data/apiSiteFilesStaging/VectorBase/49/real/webServices/Vect
orBase/release-
CURRENT/AbellatorAbelBR1/blast/AbellatorAbelBR1AnnotatedProteins
    Posted date:  Dec 31, 2023  10:23 AM
  Number of letters in database: 8,360,873
  Number of sequences in database:  14,231

  Database: /eupath/data/apiSiteFilesStaging/VectorBase/49/real/webServices/Vect
orBase/release-
CURRENT/AchristyiACHKN1017/blast/AchristyiACHKN1017AnnotatedProteins
    Posted date:  Sep 27, 2023  9:27 PM
  Number of letters in database: 5,990,400
  Number of sequences in database:  10,960

  Database: /eupath/data/apiSiteFilesStaging/VectorBase/49/real/webServices/Vect
orBase/release-
CURRENT/AcoluzziiAcolN3/blast/AcoluzziiAcolN3AnnotatedProteins
    Posted date:  Jan 22, 2024  12:39 PM
  Number of letters in database: 19,503,377
  Number of sequences in database:  24,025

  Database: /eupath/data/apiSiteFilesStaging/VectorBase/49/real/webServices/Vect
orBase/release-
CURRENT/AcoluzziiMOPTI/blast/AcoluzziiMOPTIAnnotatedProteins
    Posted date:  Jan 4, 2024  10:32 PM
  Number of letters in database: 18,309,293
  Number of sequences in database:  23,396

  Database: /eupath/data/apiSiteFilesStaging/VectorBase/49/real/webServices/Vect
orBase/release-CURRENT/AcoluzziiMali-NIH/blast/AcoluzziiMali-
NIHAnnotatedProteins
    Posted date:  Dec 30, 2023  1:58 AM
  Number of letters in database: 6,730,302
  Number of sequences in database:  14,510

  Database: /eupath/data/apiSiteFilesStaging/VectorBase/49/real/webServices/Vect
orBase/release-
CURRENT/AcoluzziiNgousso/blast/AcoluzziiNgoussoAnnotatedProteins
    Posted date:  Jan 11, 2024  6:34 PM
  Number of letters in database: 8,705,208
  Number of sequences in database:  14,705

  Database: /eupath/data/apiSiteFilesStaging/VectorBase/49/real/webServices/Vect
orBase/release-
CURRENT/AcoustaniAcouGA1/blast/AcoustaniAcouGA1AnnotatedProteins
    Posted date:  Jan 22, 2024  12:28 PM
  Number of letters in database: 9,404,970
  Number of sequences in database:  15,049

  Database: /eupath/data/apiSiteFilesStaging/VectorBase/49/real/webServices/Vect
orBase/release-
CURRENT/AcruziiAcruBR1/blast/AcruziiAcruBR1AnnotatedProteins
    Posted date:  Mar 9, 2024  8:57 PM
  Number of letters in database: 8,316,424
  Number of sequences in database:  14,301

  Database: /eupath/data/apiSiteFilesStaging/VectorBase/49/real/webServices/Vect
orBase/release-CURRENT/AculicifaciesA-37/blast/AculicifaciesA-
37AnnotatedProteins
    Posted date:  Sep 27, 2023  10:28 PM
```

```
  Number of letters in database: 7,103,499
  Number of sequences in database:  14,385

  Database: /eupath/data/apiSiteFilesStaging/VectorBase/49/real/webServices/VectorBase/release-CURRENT/AdarlingiAdarGF1/blast/AdarlingiAdarGF1AnnotatedProteins
    Posted date:  Oct 2, 2023  12:53 AM
  Number of letters in database: 16,032,793
  Number of sequences in database:  20,247

  Database: /eupath/data/apiSiteFilesStaging/VectorBase/49/real/webServices/VectorBase/release-CURRENT/AdarlingiCoari/blast/AdarlingiCoariAnnotatedProteins
    Posted date:  Sep 28, 2023  5:48 PM
  Number of letters in database: 6,124,718
  Number of sequences in database:  10,602

  Database: /eupath/data/apiSiteFilesStaging/VectorBase/49/real/webServices/VectorBase/release-CURRENT/AdirusWRAIR2/blast/AdirusWRAIR2AnnotatedProteins
    Posted date:  Sep 27, 2023  10:38 PM
  Number of letters in database: 7,179,458
  Number of sequences in database:  13,172

  Database: /eupath/data/apiSiteFilesStaging/VectorBase/49/real/webServices/VectorBase/release-CURRENT/AepiroticusEpiroticus2/blast/AepiroticusEpiroticus2AnnotatedProteins
    Posted date:  Sep 27, 2023  10:53 PM
  Number of letters in database: 6,787,400
  Number of sequences in database:  11,998

  Database: /eupath/data/apiSiteFilesStaging/VectorBase/49/real/webServices/VectorBase/release-CURRENT/AfarautiFAR1/blast/AfarautiFAR1AnnotatedProteins
    Posted date:  Sep 27, 2023  11:01 PM
  Number of letters in database: 7,069,663
  Number of sequences in database:  13,029

  Database: /eupath/data/apiSiteFilesStaging/VectorBase/49/real/webServices/VectorBase/release-CURRENT/AfunestusAfunGA1/blast/AfunestusAfunGA1AnnotatedProteins
    Posted date:  Jan 11, 2024  3:35 PM
  Number of letters in database: 21,034,589
  Number of sequences in database:  25,342

  Database: /eupath/data/apiSiteFilesStaging/VectorBase/49/real/webServices/VectorBase/release-CURRENT/AfunestusFUMOZ/blast/AfunestusFUMOZAnnotatedProteins
    Posted date:  Jan 22, 2024  1:13 PM
  Number of letters in database: 8,537,875
  Number of sequences in database:  14,621

  Database: /eupath/data/apiSiteFilesStaging/VectorBase/49/real/webServices/VectorBase/release-CURRENT/AgambiaeIfakara/blast/AgambiaeIfakaraAnnotatedProteins
    Posted date:  Mar 14, 2024  2:41 PM
  Number of letters in database: 25,895,024
  Number of sequences in database:  30,505

  Database: /eupath/data/apiSiteFilesStaging/VectorBase/49/real/webServices/VectorBase/release-CURRENT/AgambiaePEST/blast/AgambiaePESTAnnotatedProteins
    Posted date:  Sep 27, 2023  11:21 PM
  Number of letters in database: 9,076,656
  Number of sequences in database:  15,328

  Database: /eupath/data/apiSiteFilesStaging/VectorBase/49/real/webServices/VectorBase/release-CURRENT/Amaculatusmaculatus3/blast/Amaculatusmaculatus3AnnotatedProteins
```

```
    Posted date:  Sep 27, 2023  11:33 PM
  Number of letters in database: 5,487,961
  Number of sequences in database:  14,865

  Database: /eupath/data/apiSiteFilesStaging/VectorBase/49/real/webServices/Vect
orBase/release-
CURRENT/AmaculipalpisAmacGA1/blast/AmaculipalpisAmacGA1AnnotatedProt
eins
    Posted date:  Dec 28, 2023  1:45 AM
  Number of letters in database: 10,143,829
  Number of sequences in database:  14,422

  Database: /eupath/data/apiSiteFilesStaging/VectorBase/49/real/webServices/Vect
orBase/release-
CURRENT/AmarshalliiAmarGA1/blast/AmarshalliiAmarGA1AnnotatedProteins
    Posted date:  Mar 22, 2024  5:41 AM
  Number of letters in database: 7,453,026
  Number of sequences in database:  12,038

  Database: /eupath/data/apiSiteFilesStaging/VectorBase/49/real/webServices/Vect
orBase/release-
CURRENT/AmelasCM1001059_A/blast/AmelasCM1001059_AAnnotatedProteins
    Posted date:  Sep 27, 2023  11:39 PM
  Number of letters in database: 7,072,729
  Number of sequences in database:  15,389

  Database: /eupath/data/apiSiteFilesStaging/VectorBase/49/real/webServices/Vect
orBase/release-
CURRENT/AmerusMAF2021/blast/AmerusMAF2021AnnotatedProteins
    Posted date:  Jan 2, 2024  2:08 AM
  Number of letters in database: 22,819,858
  Number of sequences in database:  28,438

  Database: /eupath/data/apiSiteFilesStaging/VectorBase/49/real/webServices/Vect
orBase/release-CURRENT/AmerusMAF/blast/AmerusMAFAnnotatedProteins
    Posted date:  Dec 30, 2023  7:59 AM
  Number of letters in database: 7,016,177
  Number of sequences in database:  13,343

  Database: /eupath/data/apiSiteFilesStaging/VectorBase/49/real/webServices/Vect
orBase/release-
CURRENT/AminimusMINIMUS1/blast/AminimusMINIMUS1AnnotatedProteins
    Posted date:  Sep 27, 2023  11:56 PM
  Number of letters in database: 7,103,778
  Number of sequences in database:  13,056

  Database: /eupath/data/apiSiteFilesStaging/VectorBase/49/real/webServices/Vect
orBase/release-
CURRENT/AmouchetiAmouCM1/blast/AmouchetiAmouCM1AnnotatedProteins
    Posted date:  Mar 11, 2024  3:46 AM
  Number of letters in database: 9,772,858
  Number of sequences in database:  15,541

  Database: /eupath/data/apiSiteFilesStaging/VectorBase/49/real/webServices/Vect
orBase/release-
CURRENT/AniliAnilCM1/blast/AniliAnilCM1AnnotatedProteins
    Posted date:  Mar 9, 2024  8:33 PM
  Number of letters in database: 7,186,097
  Number of sequences in database:  11,869

  Database: /eupath/data/apiSiteFilesStaging/VectorBase/49/real/webServices/Vect
orBase/release-
CURRENT/AquadriannulatusSANGWE/blast/AquadriannulatusSANGWEAnnotated
Proteins
    Posted date:  Sep 28, 2023  12:01 AM
  Number of letters in database: 7,602,541
  Number of sequences in database:  13,926

  Database: /eupath/data/apiSiteFilesStaging/VectorBase/49/real/webServices/Vect
orBase/release-
CURRENT/AsinensisChina/blast/AsinensisChinaAnnotatedProteins
```

Posted date:  Mar 7, 2024  2:57 PM
Number of letters in database: 7,281,440
Number of sequences in database:  19,352

Database: /eupath/data/apiSiteFilesStaging/VectorBase/49/real/webServices/VectorBase/release-CURRENT/AsinensisSINENSIS/blast/AsinensisSINENSISAnnotatedProteins
Posted date:  Mar 7, 2024  7:51 PM
Number of letters in database: 5,985,446
Number of sequences in database:  12,950

Database: /eupath/data/apiSiteFilesStaging/VectorBase/49/real/webServices/VectorBase/release-CURRENT/AstephensiIndian/blast/AstephensiIndianAnnotatedProteins
Posted date:  Jan 11, 2024  7:20 PM
Number of letters in database: 6,748,567
Number of sequences in database:  11,789

Database: /eupath/data/apiSiteFilesStaging/VectorBase/49/real/webServices/VectorBase/release-CURRENT/AstephensiSDA-500/blast/AstephensiSDA-500AnnotatedProteins
Posted date:  Jan 11, 2024  7:28 PM
Number of letters in database: 7,115,611
Number of sequences in database:  13,561

Database: /eupath/data/apiSiteFilesStaging/VectorBase/49/real/webServices/VectorBase/release-CURRENT/AstephensiUCISS2018/blast/AstephensiUCISS2018AnnotatedProteins
Posted date:  Jan 11, 2024  7:09 PM
Number of letters in database: 24,355,569
Number of sequences in database:  29,673

Database: /eupath/data/apiSiteFilesStaging/VectorBase/49/real/webServices/VectorBase/release-CURRENT/AziemanniAzieGA1/blast/AziemanniAzieGA1AnnotatedProteins
Posted date:  Jan 2, 2024  12:47 AM
Number of letters in database: 7,965,337
Number of sequences in database:  12,900

Database: /eupath/data/apiSiteFilesStaging/VectorBase/49/real/webServices/VectorBase/release-CURRENT/CquinquefasciatusJHB2020/blast/CquinquefasciatusJHB2020AnnotatedProteins
Posted date:  Dec 12, 2023  10:40 AM
Number of letters in database: 16,471,461
Number of sequences in database:  24,676

Database: /eupath/data/apiSiteFilesStaging/VectorBase/49/real/webServices/VectorBase/release-CURRENT/CquinquefasciatusJohannesburg/blast/CquinquefasciatusJohannesburgAnnotatedProteins
Posted date:  Oct 1, 2023  2:05 PM
Number of letters in database: 8,339,467
Number of sequences in database:  19,038

Database: /eupath/data/apiSiteFilesStaging/VectorBase/49/real/webServices/VectorBase/release-CURRENT/ScyaneusScyaPA1/blast/ScyaneusScyaPA1AnnotatedProteins
Posted date:  Jan 24, 2024  3:16 PM
Number of letters in database: 12,570,107
Number of sequences in database:  17,957

Matrix: BLOSUM62
Gap Penalties: Existence: 11, Extension: 1
Neighboring words threshold: 11
Window for multiple hits: 40

| Run on Tue May 13 17:52:44 2025
|
|gmx_MMPBSA Version=1.6.4 based on MMPBSA.py v.16.0
|Complex Structure file: md.tpr
|Complex (AMBER) topology file: COM.prmtop
|Receptor (AMBER) topology file: REC.prmtop
|Ligand (AMBER) topology file: LIG.prmtop
|Initial trajectories: COM_traj_0.xtc
|
|Receptor mask: ":1-412"
|Ligand mask: ":413-428"
|
|Calculations performed using 10 complex frames
|
|Generalized Born ESURF calculated using 'LCPO' surface areas
|
|Using temperature = 298.15 K
|All units are reported in kcal/mol
|
|SD - Sample standard deviation, SEM - Sample standard error of the mean
|SD(Prop.), SEM(Prop.) - SD and SEM obtained with propagation of uncertainty formula
|https://en.wikipedia.org/wiki/Propagation_of_uncertainty#Example_formulae
|
gmx_mmpbsa_hDSG3

-------------------------------------------------------------------------------

-------------------------------------------------------------------------------

GENERALIZED BORN:

Complex:

| Energy Component | Average | SD(Prop.) | SD | SEM(Prop.) | SEM |
|---|---|---|---|---|---|
| BOND | 26255492.85 | 221417.00 | 221417.00 | 73805.67 | 73805.67 |
| ANGLE | 3569.63 | 139.04 | 139.04 | 46.35 | 46.35 |
| DIHED | 4510.50 | 18.06 | 18.06 | 6.02 | 6.02 |
| VDWAALS | -3352.99 | 25.82 | 25.82 | 8.61 | 8.61 |
| EEL | -27033.55 | 51.06 | 51.06 | 17.02 | 17.02 |
| 1-4 VDW | 1772.87 | 22.24 | 22.24 | 7.41 | 7.41 |
| 1-4 EEL | 15968.07 | 41.28 | 41.28 | 13.76 | 13.76 |
| EGB | -7700.48 | 24.32 | 24.32 | 8.11 | 8.11 |
| ESURF | 170.94 | 1.09 | 1.09 | 0.36 | 0.36 |
| GGAS | 26250927.37 | 221417.06 | 221330.10 | 73805.69 | 73776.70 |
| GSOLV | -7529.54 | 24.35 | 24.53 | 8.12 | 8.18 |
| TOTAL | 26243397.83 | 221417.06 | 221338.45 | 73805.69 | 73779.48 |

-------------------------------------------------------------------------------

-------------------------------------------------------------------------------

Receptor:

| Energy Component | Average | SD(Prop.) | SD | SEM(Prop.) | SEM |
|---|---|---|---|---|---|
| BOND | 26255491.26 | 221417.01 | 221417.01 | 73805.67 | 73805.67 |
| ANGLE | 3460.74 | 140.08 | 140.08 | 46.69 | 46.69 |
| DIHED | 4349.56 | 20.99 | 20.99 | 7.00 | 7.00 |
| VDWAALS | -3231.38 | 24.71 | 24.71 | 8.24 | 8.24 |
| EEL | -26067.04 | 43.53 | 43.53 | 14.51 | 14.51 |
| 1-4 VDW | 1710.21 | 21.62 | 21.62 | 7.21 | 7.21 |
| 1-4 EEL | 15363.66 | 42.00 | 42.00 | 14.00 | 14.00 |
| EGB | -7360.47 | 21.32 | 21.32 | 7.11 | 7.11 |
| ESURF | 167.79 | 1.00 | 1.00 | 0.33 | 0.33 |
| GGAS | 26251077.01 | 221417.07 | 221331.19 | 73805.69 | 73777.06 |
| GSOLV | -7192.68 | 21.35 | 21.54 | 7.12 | 7.18 |
| TOTAL | 26243884.33 | 221417.07 | 221336.24 | 73805.69 | 73778.75 |

-------------------------------------------------------------------------------

-------------------------------------------------------------------------------

Ligand:

| Energy Component | Average | SD(Prop.) | SD | SEM(Prop.) | SEM |
|---|---|---|---|---|---|
| BOND | 1.59 | 0.11 | 0.11 | 0.04 | 0.04 |
| ANGLE | 108.89 | 4.73 | 4.73 | 1.58 | 1.58 |

```
DIHED                    160.94          4.08        4.08          1.36        1.36
VDWAALS                  -54.82          2.84        2.84          0.95        0.95
EEL                    -1004.57          9.69        9.69          3.23        3.23
1-4 VDW                   62.65          1.79        1.79          0.60        0.60
1-4 EEL                  604.41          4.26        4.26          1.42        1.42
EGB                     -343.55          7.69        7.69          2.56        2.56
ESURF                     13.00          0.16        0.16          0.05        0.05

GGAS                    -120.91         12.74        7.96          4.25        2.65
GSOLV                   -330.55          7.69        7.57          2.56        2.52

TOTAL                   -451.46         14.88        4.38          4.96        1.46

-------------------------------------------------------------------------------
-------------------------------------------------------------------------------
Delta (Complex - Receptor - Ligand):
Energy Component        Average     SD(Prop.)          SD   SEM(Prop.)         SEM
-------------------------------------------------------------------------------
ΔBOND                     -0.00          0.10        0.00          0.03        0.00
ΔANGLE                    -0.00          3.69        0.00          1.23        0.00
ΔDIHED                    -0.00          1.14        0.00          0.38        0.00
ΔVDWAALS                 -66.79          1.72        3.61          0.57        1.20
ΔEEL                      38.06          2.17       11.28          0.72        3.76
Δ1-4 VDW                   0.00          1.18        0.00          0.39        0.00
Δ1-4 EEL                   0.00          3.55        0.00          1.18        0.00
ΔEGB                       3.54          4.69        8.65          1.56        2.88
ΔESURF                    -9.85          0.07        0.13          0.02        0.04

ΔGGAS                    -28.73          2.77       10.40          0.92        3.47
ΔGSOLV                    -6.31          4.69        8.61          1.56        2.87

ΔTOTAL                   -35.04          5.45        4.26          1.82        1.42
-------------------------------------------------------------------------------
-------------------------------------------------------------------------------
|
Run on Sat May 17 00:43:05 2025
|
|gmx_MMPBSA Version=1.6.4 based on MMPBSA.py v.16.0
|Complex Structure file:                                     last_frame_amber.pdb
|Complex (AMBER) topology file:                                      COM.prmtop
|Receptor (AMBER) topology file:                                     REC.prmtop
|Ligand (AMBER) topology file:                                       LIG.prmtop
|Initial trajectories:                                           COM_traj_0.xtc
|
|Receptor mask:                  ":1-412"
|Ligand mask:                    ":413-428"
|
|Calculations performed using 10 complex frames
|
|Generalized Born ESURF calculated using 'LCPO' surface areas
|
|Using temperature = 298.15 K
|All units are reported in kcal/mol
|
|SD - Sample standard deviation, SEM - Sample standard error of the mean
|SD(Prop.), SEM(Prop.) - SD and SEM obtained with propagation of uncertainty formula
|https://en.wikipedia.org/wiki/Propagation_of_uncertainty#Example_formulae
|
gmx_mmpbsa_Aa_p
-----------------------------------------------------------------------------
-------------------------------------------------------------------------------

GENERALIZED BORN:

Complex:
Energy Component        Average     SD(Prop.)          SD   SEM(Prop.)         SEM
-------------------------------------------------------------------------------
BOND                24359742.88   13377772.18 13377772.18    4230423.01 4230423.01
ANGLE                   3634.03        493.90      493.90        156.18      156.18
DIHED                   4475.04         33.53       33.53         10.60       10.60
VDWAALS                -3395.21         24.52       24.52          7.76        7.76
EEL                   -26851.57         67.84       67.84         21.45       21.45
1-4 VDW                 1763.98         10.52       10.52          3.33        3.33
```

```
1-4 EEL              15847.29        42.02        42.02        13.29        13.29
EGB                  -7910.68        79.58        79.58        25.16        25.16
ESURF                  166.42         0.94         0.94         0.30         0.30

GGAS              24355216.44  13377772.19  13378307.98   4230423.01   4230592.45
GSOLV                -7744.26        79.58        79.17        25.17        25.04

TOTAL             24347472.19  13377772.19  13378234.73   4230423.01   4230569.28

-------------------------------------------------------------------------------
-------------------------------------------------------------------------------
Receptor:
Energy Component       Average     SD(Prop.)           SD   SEM(Prop.)          SEM
-------------------------------------------------------------------------------
BOND              24359741.34  13377772.13  13377772.13   4230423.00   4230423.00
ANGLE                 3545.09       493.78       493.78       156.15       156.15
DIHED                 4332.07        35.19        35.19        11.13        11.13
VDWAALS              -3246.61        23.98        23.98         7.58         7.58
EEL                 -25981.02        65.75        65.75        20.79        20.79
1-4 VDW               1702.47         9.46         9.46         2.99         2.99
1-4 EEL              15226.11        38.82        38.82        12.27        12.27
EGB                  -7427.36        78.58        78.58        24.85        24.85
ESURF                  166.18         0.82         0.82         0.26         0.26

GGAS              24355319.45  13377772.14  13378307.68   4230423.00   4230592.35
GSOLV                -7261.19        78.59        78.20        24.85        24.73

TOTAL             24348058.26  13377772.14  13378236.54   4230423.00   4230569.86

-------------------------------------------------------------------------------
-------------------------------------------------------------------------------
Ligand:
Energy Component       Average     SD(Prop.)           SD   SEM(Prop.)          SEM
-------------------------------------------------------------------------------
BOND                     1.54         0.12         0.12         0.04         0.04
ANGLE                   88.94         5.37         5.37         1.70         1.70
DIHED                  142.98         4.11         4.11         1.30         1.30
VDWAALS                -38.93         1.43         1.43         0.45         0.45
EEL                   -907.48         7.62         7.62         2.41         2.41
1-4 VDW                 61.52         2.17         2.17         0.69         0.69
1-4 EEL                621.18         6.01         6.01         1.90         1.90
EGB                   -483.28         3.79         3.79         1.20         1.20
ESURF                   14.97         0.08         0.08         0.03         0.03

GGAS                   -30.27        12.11         9.91         3.83         3.13
GSOLV                 -468.32         3.79         3.81         1.20         1.21

TOTAL                 -498.58        12.69         9.10         4.01         2.88

-------------------------------------------------------------------------------
-------------------------------------------------------------------------------
Delta (Complex - Receptor - Ligand):
Energy Component       Average     SD(Prop.)           SD   SEM(Prop.)          SEM
-------------------------------------------------------------------------------
ΔBOND                   -0.00         0.00         0.00         0.00         0.00
ΔANGLE                  -0.00         5.25         0.00         1.66         0.00
ΔDIHED                   0.00         2.46         0.00         0.78         0.00
ΔVDWAALS              -109.68         0.89         3.35         0.28         1.06
ΔEEL                    36.93         5.53         7.89         1.75         2.50
Δ1-4 VDW                -0.00         1.11         0.00         0.35         0.00
Δ1-4 EEL                 0.00         2.80         0.00         0.89         0.00
ΔEGB                    -0.03         2.79         7.35         0.88         2.32
ΔESURF                 -14.72         0.05         0.19         0.01         0.06

ΔGGAS                  -72.74         5.61         7.83         1.77         2.48
ΔGSOLV                 -14.75         2.79         7.38         0.88         2.33

ΔTOTAL                 -87.50         6.26         3.43         1.98         1.08
-------------------------------------------------------------------------------
-------------------------------------------------------------------------------
```
